# Bibliometric analysis of emerging trends and research foci in brainstem tumor field over thirty years (1992-2023)

**DOI:** 10.1101/2023.09.16.23295661

**Authors:** Yibo Geng, Luyang Xie, Jinping Li, Yang Wang, Xiong Li

**Author notes:** Yibo Geng and Luyang Xie contributed equally to this work. Corresponding author Xiong Li, Department of Neurosurgery, Beijing Chaoyang Hospital, Capital Medical University, No.8, South Road of Workers Stadium, Chaoyang District, Beijing, 100020, China.

## Abstract

**Background:** Over the past several decades, numerous articles have been published on brainstem tumors. However, there has been limited bibliometric analysis in this field. Therefore, we conducted a bibliometric analysis to elucidate the evolution and current status of brainstem tumor research.

**Methods:** We retrieved 5,927 studies published in English between 1992 and 2023 from the Web of Science Core Collection database. We employed bibliometric tools and VOSviewer to conduct the analysis.

**Results:** We identified and included a total of 5,927 publications for further analysis. The number of annual publications has exhibited steady growth over time. The United States accounted for the highest number of publications and total citations. The St. Jude Children’s Research Hospital emerged as the most productive institution, while the Journal of Neurosurgery was the leading journal in terms of publication output. Among individual researchers, Liwei Zhang had the highest number of publications, while Eric Bouffet received the most citations in this field. The study titled "Somatic Histone H3 Alterations in Pediatric Diffuse Intrinsic Pontine Gliomas and Non-brainstem Glioblastomas" stood out as the most cited work in this field. Keyword analysis revealed that immune therapy and epigenetic research are the focal points of this field.

**Conclusion:** Our bibliometric analysis underscores the enduring significance of brainstem tumors in the realm of neuro-oncology research. The field’s hotspots have transitioned from surgery and radiochemotherapy to investigating epigenetic mechanisms and immune therapy.

## 1. Introduction

Brainstem tumors constitute a heterogeneous group of predominantly pediatric tumors[1]. Despite remarkable strides in reducing brainstem tumor-related mortality in recent decades, the precise pathological mechanisms, treatment modalities, and prognosis remain unsatisfactorily understood[2]. Consequently, this field has garnered substantial attention from scientists worldwide. Over the past thirty years, more than 5,927 publications have emerged in this field. Given the volume of literature, a bibliometric analysis becomes essential to synthesize valuable publications, identify research trends, and pinpoint potential frontiers in brainstem tumor research.

Bibliometric analysis is an emerging method for systematically examining scientific literature, offering both quantitative and qualitative insights into evolving trends[3]. It encompasses the analysis of citations within a specific research domain, as well as an extensive examination of cited references associated with these studies. Additionally, it explores various characteristics such as journal distribution, collaborative relationships among countries, institutions, and authors, and the identification of keywords and temporal patterns, all of which are invaluable for researchers seeking to discern the key focal points of a study[4]. Bibliometric analysis has found widespread application across a spectrum of studies, encompassing diverse areas like liver disease, acute myocardial infarction, and chordoma[5-7]. Surprisingly, to date, no bibliometric analysis has been conducted to scrutinize brainstem tumors. Thus, the objective of our study is to furnish a comprehensive overview of current research trends and promote a deeper comprehension of this field.

## 2. Methods

### 2.1. Data acquisition

We conducted a comprehensive search in the Web of Science Core Collection (WoSCC) database, spanning from January 1992 to August 2023. Our search terms included "brainstem tumor," "midbrain tumor," "medulla tumor," "pontine tumor," and "medulla tumor." We focused on articles and review articles and restricted the search to publications in English. After this rigorous selection process, we identified and included a total of 5,927 records for our analysis. (Supplementary file 1).

### 2.2. Data analysis

The analysis and visualization of bibliometric data were conducted using a suite of specialized software tools, including the R Bibliometrix package (R, Version 4.3.1), VOSviewer (Version 1.6.19), and Microsoft Office Excel 2016.

VOSviewer, a Java-based bibliometric analysis tool[8], played a central role in our study. It was employed to extract and visually represent essential insights from the comprehensive body of literature. Our utilization of VOSviewer encompassed several critical analyses: the examination of co-authorship relationships among countries, authors, and institutions, the exploration of co-citations involving journals and references, and the identification of keyword co-occurrence patterns. Within the visualizations generated by VOSviewer, node size signified the volume of entities, while color coding categorized these entities. Furthermore, the thickness of connecting lines in the visualizations conveyed the extent of collaboration or co-citation among the entities.

The R package known as "bibliometrix" was instrumental in our study[9]. It facilitated the calculation of the annual growth rate of publications and the establishment of a global distribution map of scientific production by country. Microsoft Office Excel 2016 was applied to perform quantitative data analysis and publication trends curve.

## 3. Results

### 3.1. Trends and annual publications

The volume of global publications in the field of brainstem tumors exhibited consistent growth, rising from 73 in 1992 to 312 in 2022 (Figure 1A), as illustrated by the smooth upward trajectory depicted in Figure 1B. The zenith of production was observed in 2021, with a total of 348 articles published.

**Figure 1.**
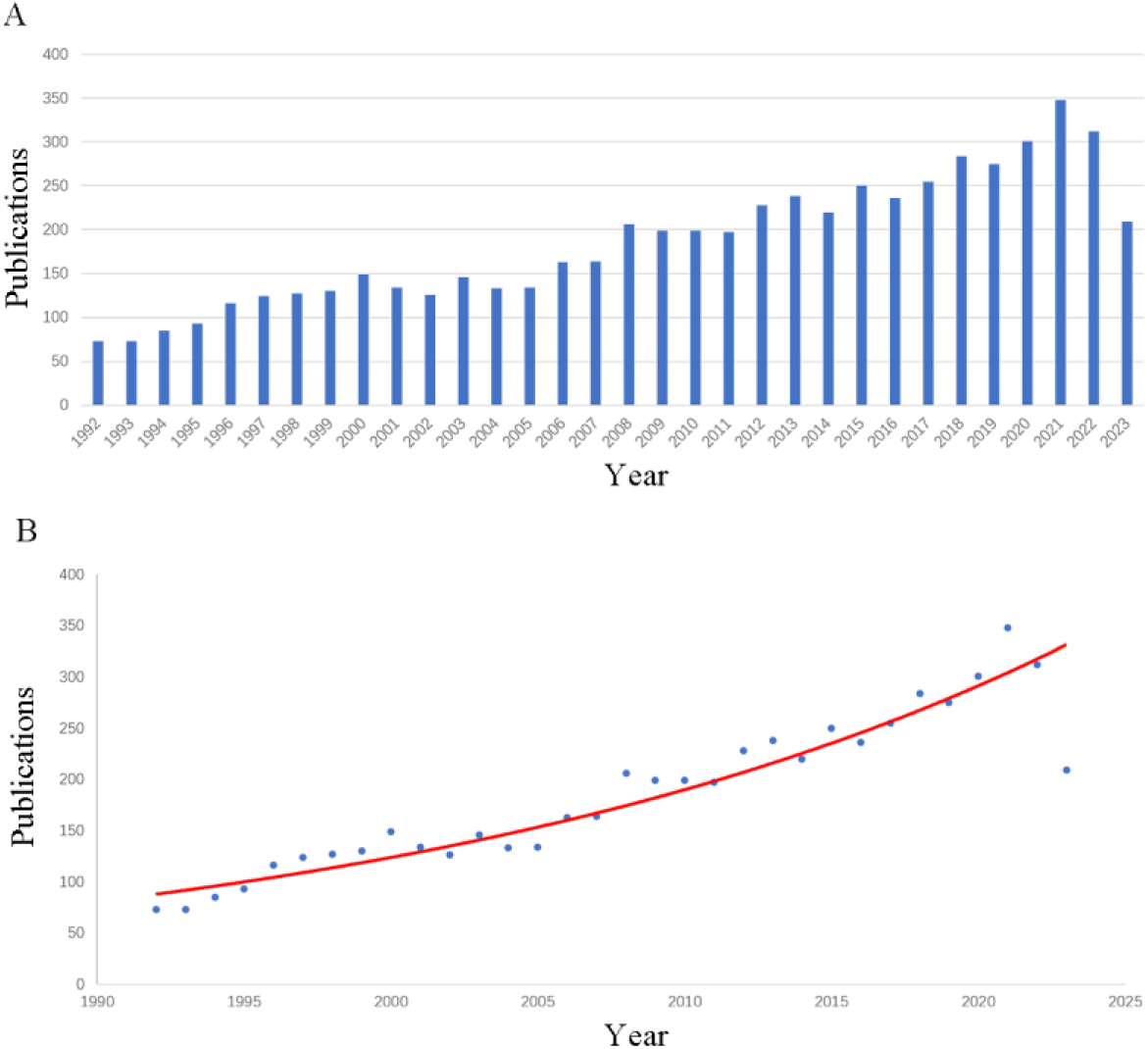
Analysis of publication trend in brainstem tumor field. **(A)** Annual global publications between 1992 to 2023. **(B)** The publication consistently increased suggested by time curve.

### 3.2. Countries and organizations

In terms of geographical distribution, a total of 5,927 documents originated from 84 different countries and regions (Figure 2A). The United States led in publications, contributing 1,987 papers, constituting 33.5% of the total. This was followed by China with 555 papers (9.4%), Japan with 511 papers (8.6%), Germany with 382 papers (6.4%), and Italy with 244 papers (4.1%) (Figure 2B). Regarding citations, the United States commanded the highest citation frequency, amassing an impressive 89,538 citations. Germany followed with 11,558 citations, Japan with 11,085 citations, and Canada with 8,389 citations (Figure 2C). A comprehensive analysis of total link strength among 43 countries unveiled that the top five nations were the United States (1,096 times), Germany (485 times), England (415 times), Canada and France, both with 341 times (Figure 3A).

**Figure 2.**
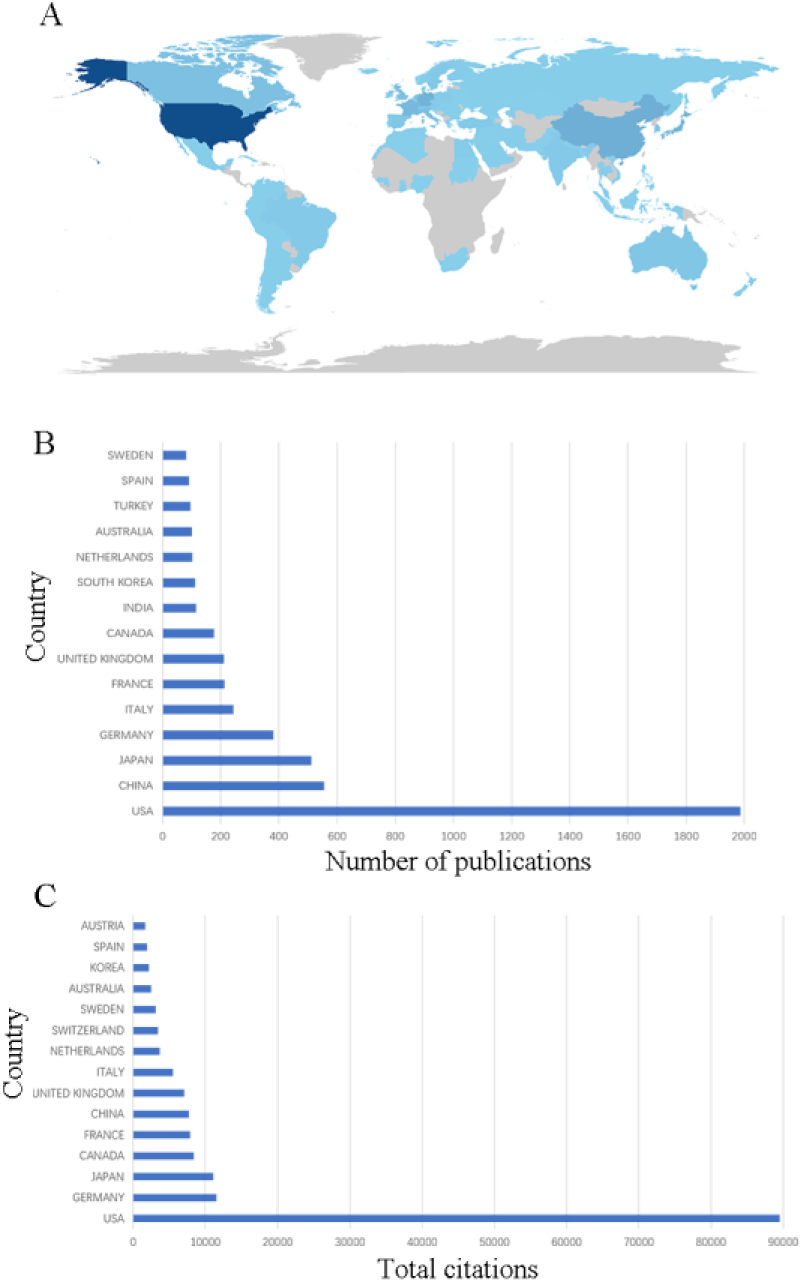
Global countries investigating brainstem tumors. **(A)** The heatmap of global publication distribution. **(B)** Publications of top fifteen countries. **(C)** Citations of top fifteen countries.

**Figure 3.**
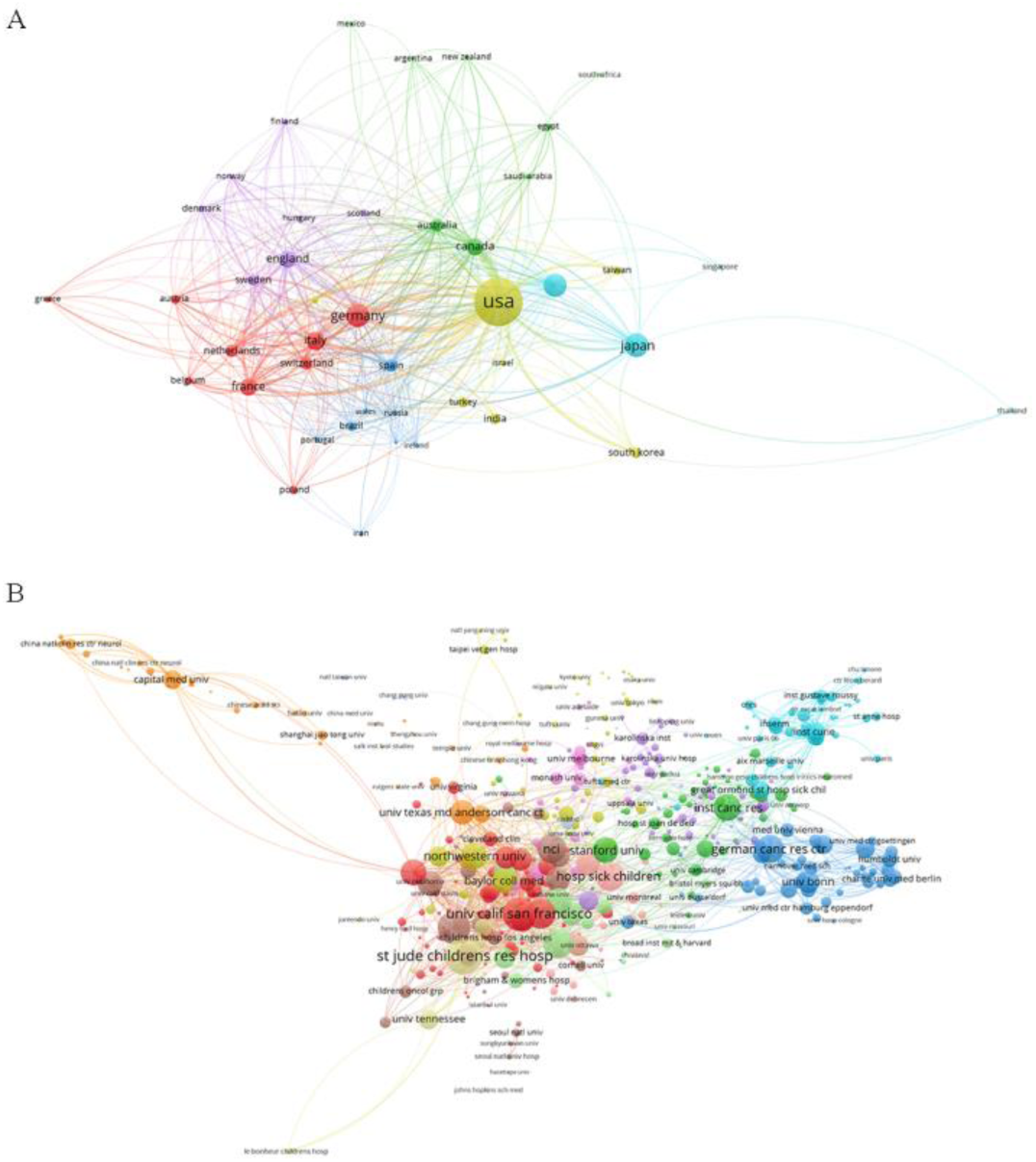
Total link strength analysis of countries and organizations. A network map revealing correlation between countries **(A)** and organizations **(B)**.

In terms of organizational contributions, a noteworthy total of 4,886 institutions participated in brainstem tumor research. Leading the pack, St. Jude Children’s Research Hospital made the most substantial impact with 129 records, representing 2.2% of all articles. Following closely were the University of California San Francisco (119 records, 2.0%), Capital Medical University (109 records, 1.8%), Memorial Sloan Kettering Cancer Center (101 records, 1.7%), and Duke University (87 records, 1.5%). Moreover, the assessment of total link strength among institutions encompassed 283 institutions, with only one organization excluded due to its lack of collaboration with other institutions (Figure 3B). Among the 209 entities considered, the top five institutions in terms of link strength were St. Jude Children’s Research Hospital (329), Children’s National Medical Center (273), University of California San Francisco (272), Memorial Sloan Kettering Cancer Center (235), and Dana-Farber Cancer Institute (234).

### 3.3. Sources of the publications

The top 10 journals from a pool of 1,285 journals, representing the highest number of publications related to brainstem tumors, are summarized in Table 1. The Journal of Neurosurgery led the pack with 200 records, accounting for 3.4% of all articles, followed by the Journal of Neuro-Oncology (169 records, 2.9%), Childs Nervous System (151 records, 2.5%), World Neurosurgery (129 records, 2.2%), and the International Journal of Radiation Oncology Biology Physics (96 records, 1.6%).

**Table 1.** Top 10 journals that have the outmost number of publications and cited journals have the largest number of citations.

| Journals | Number | Impact<br>Factor<br>(partition) | Cited journals | Number | Impact<br>Factor<br>(partition) |
| --- | --- | --- | --- | --- | --- |
| Journal of<br>Neurosurgery | 200 | 4.1 (Q1) | Journal of<br>Neurosurgery | **8301** | 4.1 (Q1) |
| Journal of<br>Neuro-Oncology | 169 | 3.9 (Q2) | Neurosurgery | **5767** | 4.8 (Q1) |
| Childs Nervous<br>System | 151 | 1.4 (Q3) | IJROBP | **4886** | 7.0 (Q1) |
| World<br>Neurosurgery | 129 | 2.0 (Q2) | Journal of<br>Neuro-Oncology | **3752** | 3.9 (Q2) |
| IJROBP | 96 | 7.0 (Q1) | **Journal of Clinical**<br>**Oncology** | **3507** | 45.3 (Q1) |
| Neurosurgery | 94 | 4.8 (Q1) | **Nature** | **3496** | 64.8 (Q1) |
| Journal of<br>Neurosurgery:<br>Pediatrics | 93 | 1.9 (Q2) | Neuro-Oncology | **3463** | 15.9 (Q1) |
| Neuro-Oncology | 83 | 15.9 (Q1) | **PNAS** | **3303** | 11.1 (Q1) |
| Journal of<br>Clinical<br>Neuroscience | 82 | 2.0 (Q4) | **Cancer Research** | **2726** | 11.2 (Q1) |
| Acta<br>Neurochirurgica | 78 | 2.4 (Q2) | **Acta**<br>**Neuropathologica** | **2711** | 12.7 (Q1) |

Figure 4 illustrates that the analysis of co-citations among journals included a subset of 640 journals that had published more than 50 articles related to brainstem tumor. Furthermore, Table 1 presents the top 10 journals with the highest number of cited brainstem tumor-related publications. Specifically, the Journal of Neurosurgery secured the highest number of citations (8,301), followed by Neurosurgery (5,767), International Journal of Radiation Oncology Biology Physics (4,886), Journal of Neuro-Oncology (3,752), and Journal of Clinical Oncology (3,507).

**Figure 4.**
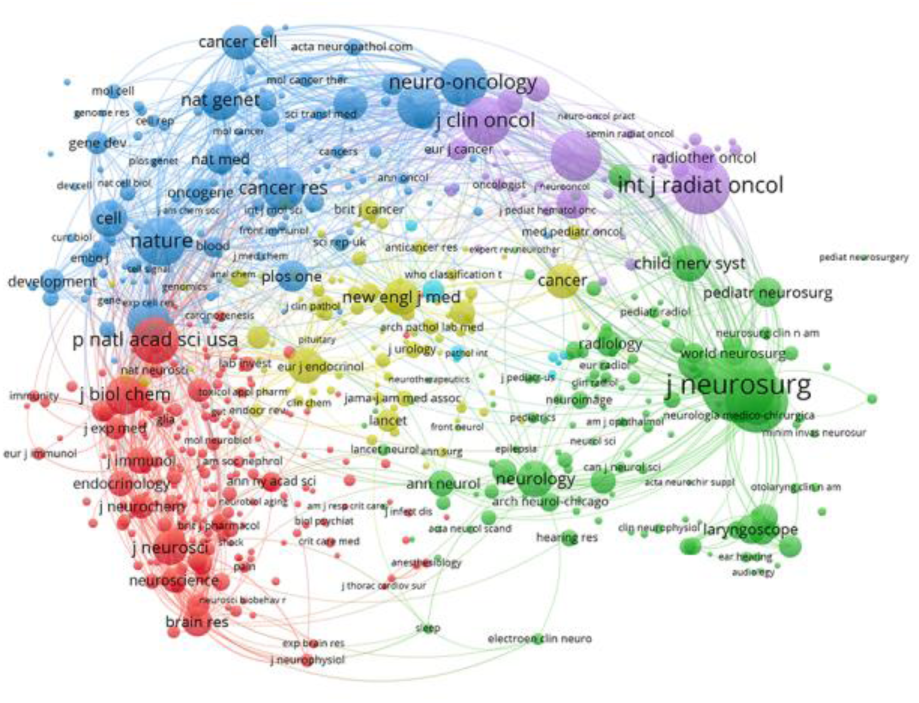
A network map of 640 journals used in the co-cited analysis.

### 3.4. Authors and co-cited authors

A total of 26,809 authors have contributed articles related to brainstem tumors. As depicted in Figure 5A, Liwei Zhang from Capital Medical University emerged as the most prolific author with the highest number of published papers, totaling 65 articles (Figure 5B). He was followed closely by Amar Gajjar from St. Jude Children’s Research Hospital with 51 publications, Alberto Broniscer from the University of Pittsburgh with 48, Junting Zhang from Capital Medical University with 47, and Jacques Grill from Gustave Roussy with 45. Furthermore, in terms of citations, Eric Bouffet secured the top position in this field with 1,551 citations, closely followed by Cynthia Hawkins with 1,550 citations, Chris Jones with 1,448 citations, Jacques Grill with 1,428 citations, and Ute Bartels with 1,302 citations (Figure 5C).

**Figure 5.**
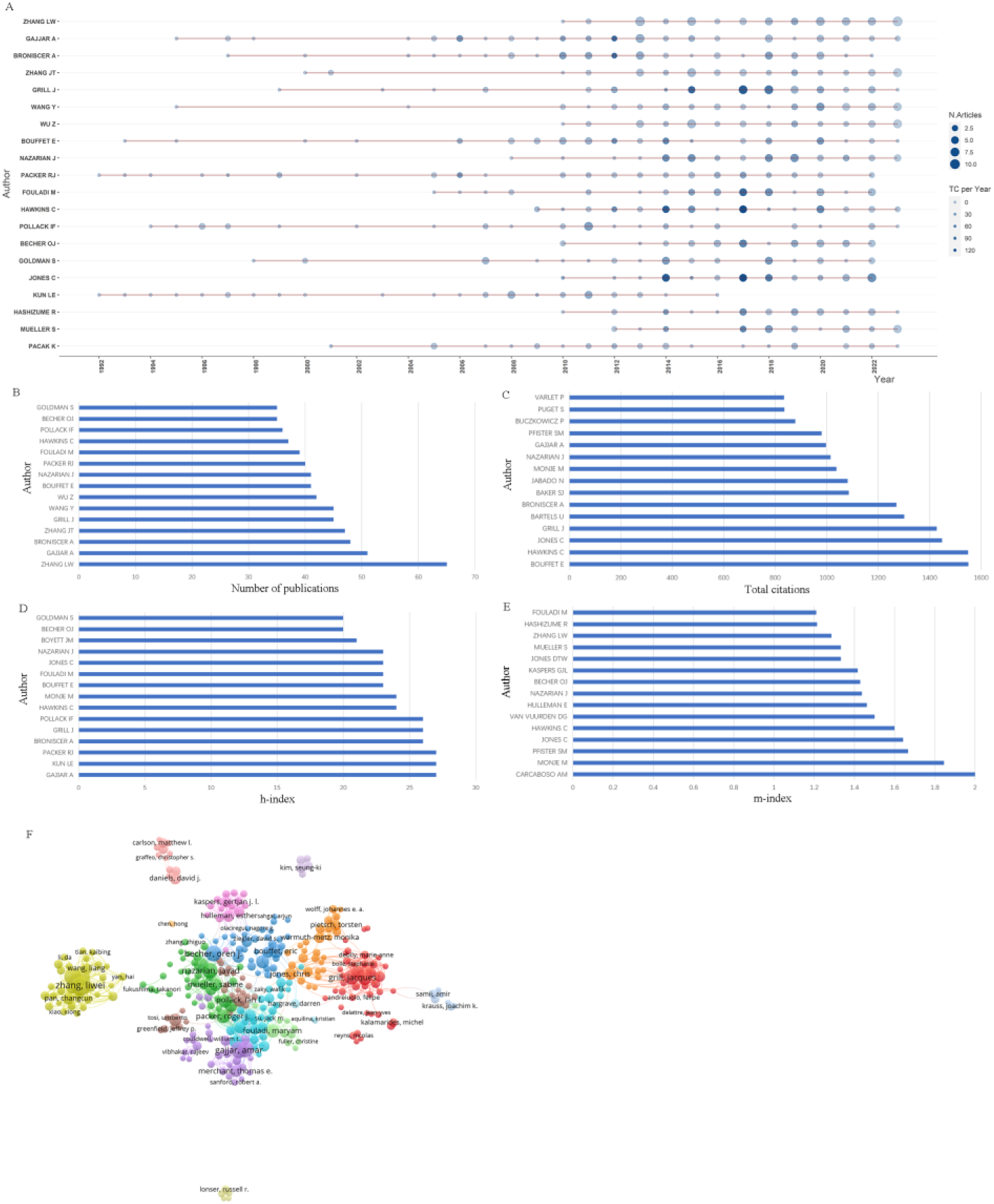
Authors analysis. **(A)** Top fifteen authors’ production over time. Node size indicated annual number of publications, and node color indicated annual total citation. **(B)** The top fifteen productive authors. **(C)** The top fifteen citation author. **(D)** H-index rank list. **(E)** M-index rank list. **(F)** Co-occurrence analysis of 318 authors.

On the other hand, the h-index serves as an author-level metric, assessing both the productivity and scientific impact of researchers. The top six authors with the highest h-index values were as follows: Amar Gajjar, LE Kun, and Roger J. Packer, all with an h-index of 27, followed by Alberto Broniscer, Jacques Grill, and Ian F. Pollack, each with an h-index of 26 (Figure 5D). Similarly, the m-index is inclined to highlight young and promising academics. Topping the m-index rankings was Angel Montero Carcaboso with a score of 2.00, followed by Michelle Monje (1.85), Stefan M Pfister (1.67), Chris Jones (1.64), and Cynthia Hawkins (1.60) (Figure 5E).

Among the 493 authors considered for co-author analysis, 175 authors were excluded due to a lack of connections with others. Among the 493 authors considered for co-author analysis, 175 authors were excluded due to a lack of connections with others (Figure 5F).

### 3.5. Citations analysis

The results reveal that there were 535 publications garnering over 80 citations (Figure 6A). In Table 2, we present the top 10 publications with the highest global citations. Notably, the publications titled "CBTRUS Statistical Report: Primary Brain and Other Central Nervous System Tumors Diagnosed in the United States in 2011-2015," "Single-dose radiosurgical treatment of recurrent previously irradiated primary brain tumors and brain metastases: final report of RTOG protocol 90-05," and "Somatic histone H3 alterations in pediatric diffuse intrinsic pontine gliomas and non-brainstem glioblastomas" claimed the top three positions.

**Figure 6.**
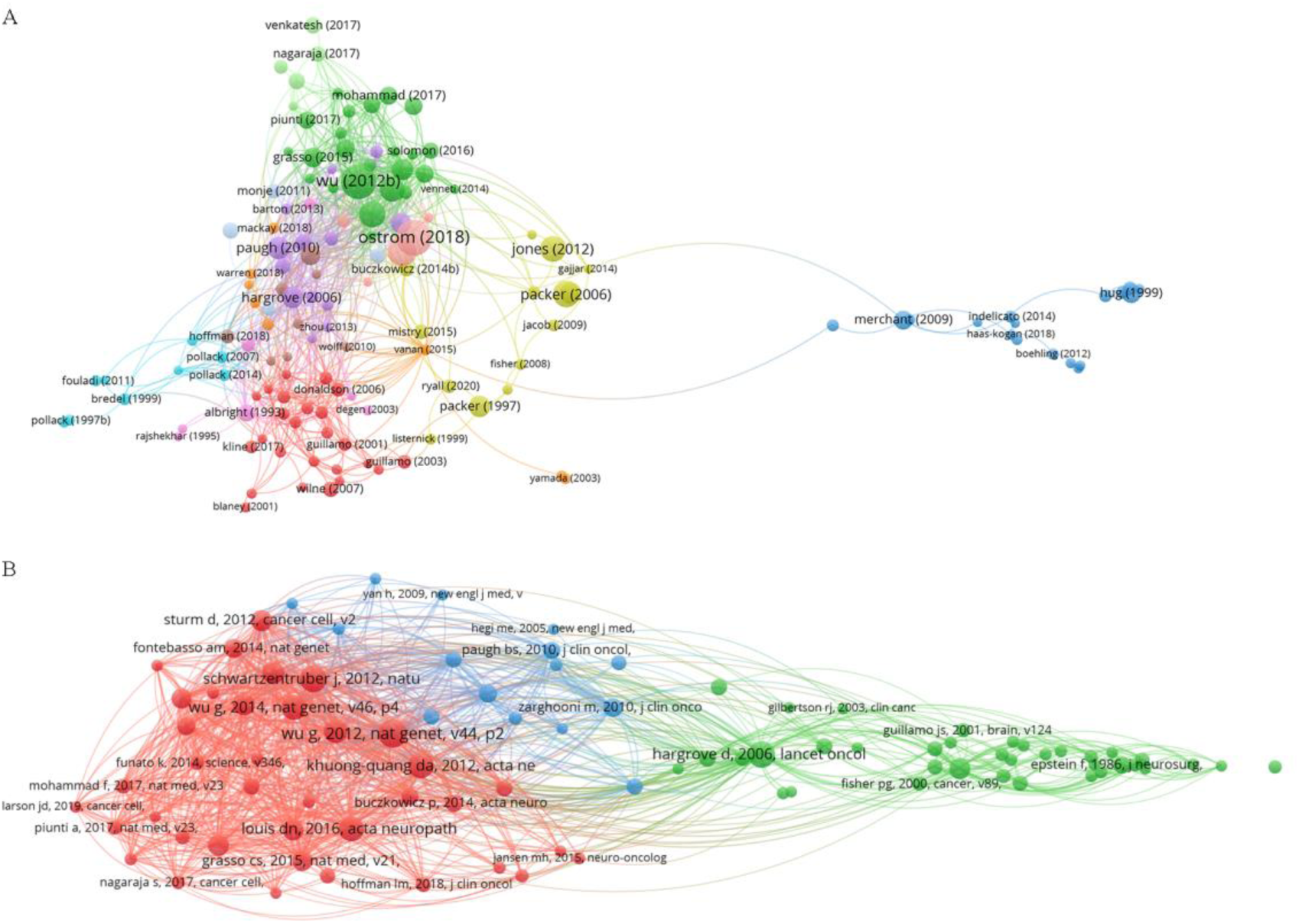
Citation analysis. **(A)** Citation analysis of documents. **(B)** Co-citation analysis of publications.

**Table 2.**
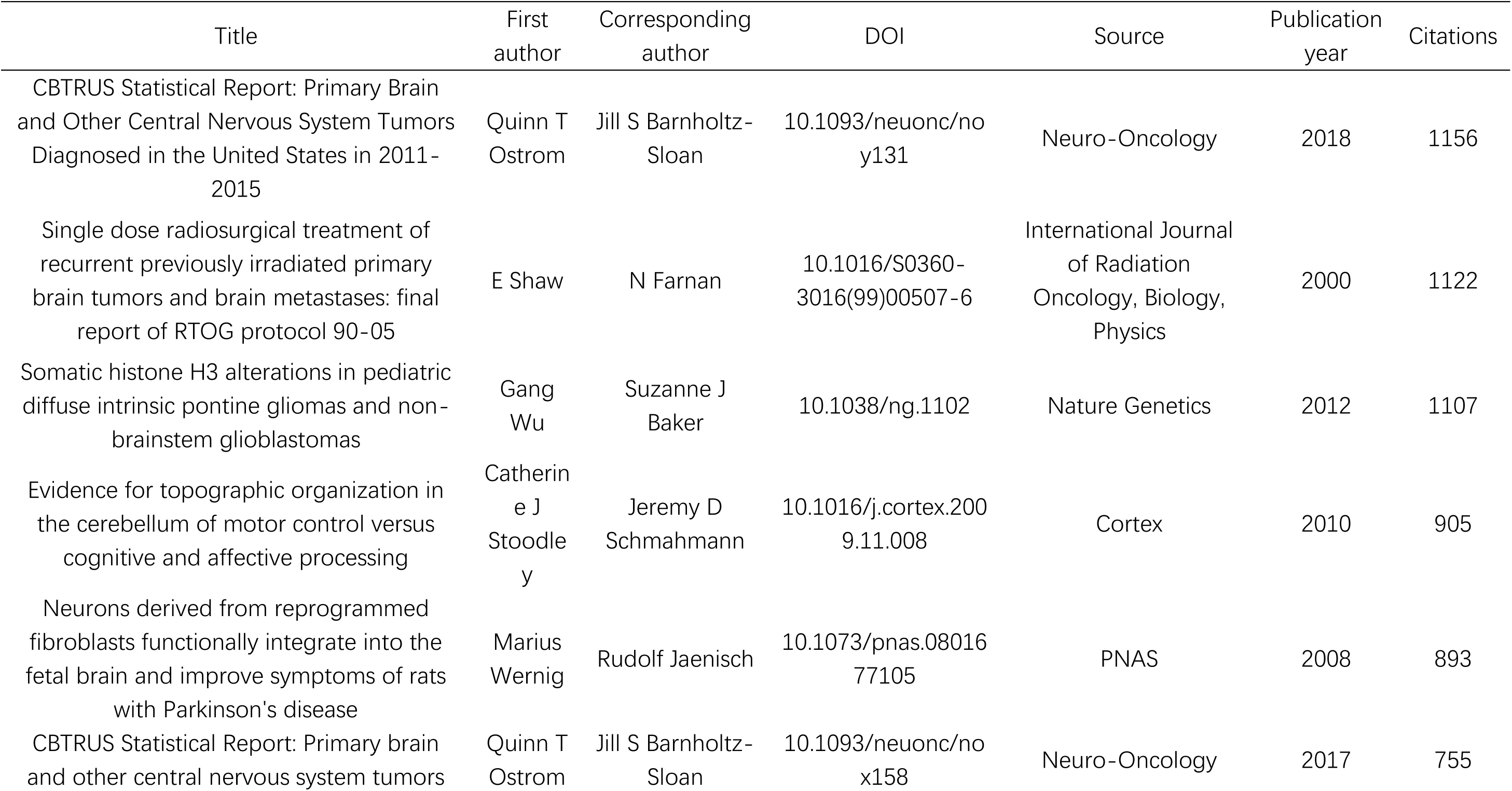
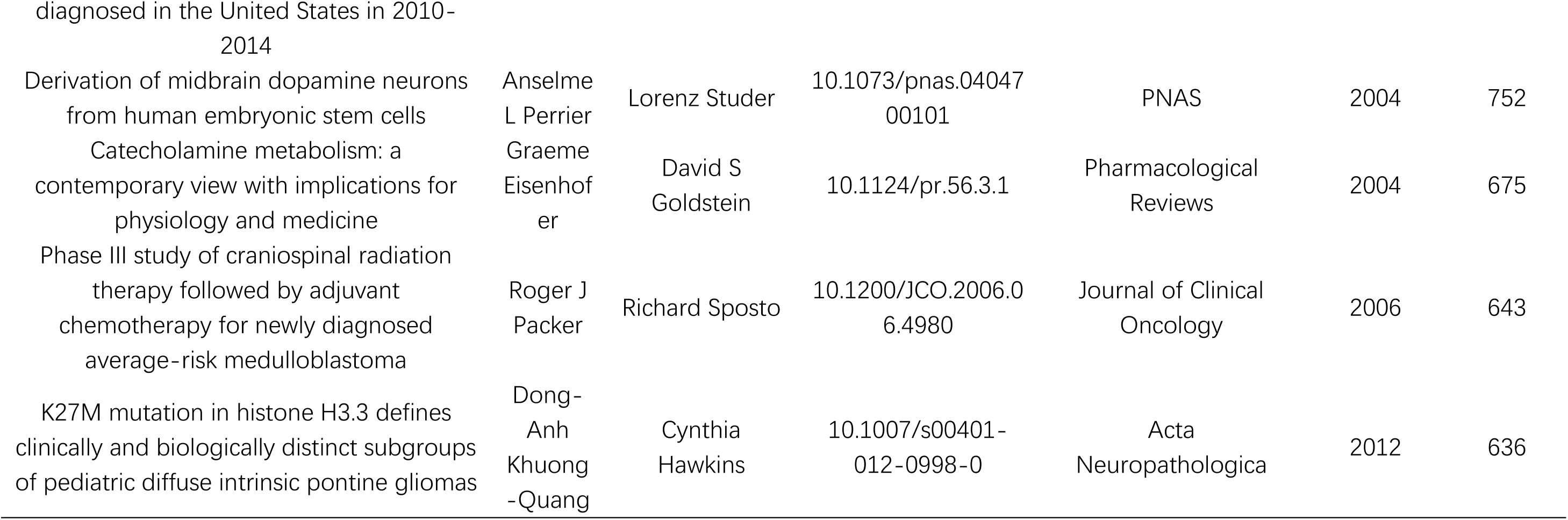
Top ten publication of have the outmost number of citations in global documents.

In the realm of co-cited analysis, we included 93 publications (Figure 6B), and among them, we present the top 10 publications with the highest number of citations in Table 3. The top three publications, based on citation rank, were "Somatic histone H3 alterations in pediatric diffuse intrinsic pontine gliomas and non-brainstem glioblastomas" (313 citations), "Driver mutations in histone H3.3 and chromatin remodeling genes in pediatric glioblastoma" (283 citations), and "Diffuse brainstem glioma in children: critical review of clinical trials" (274 citations).

**Table 3.**
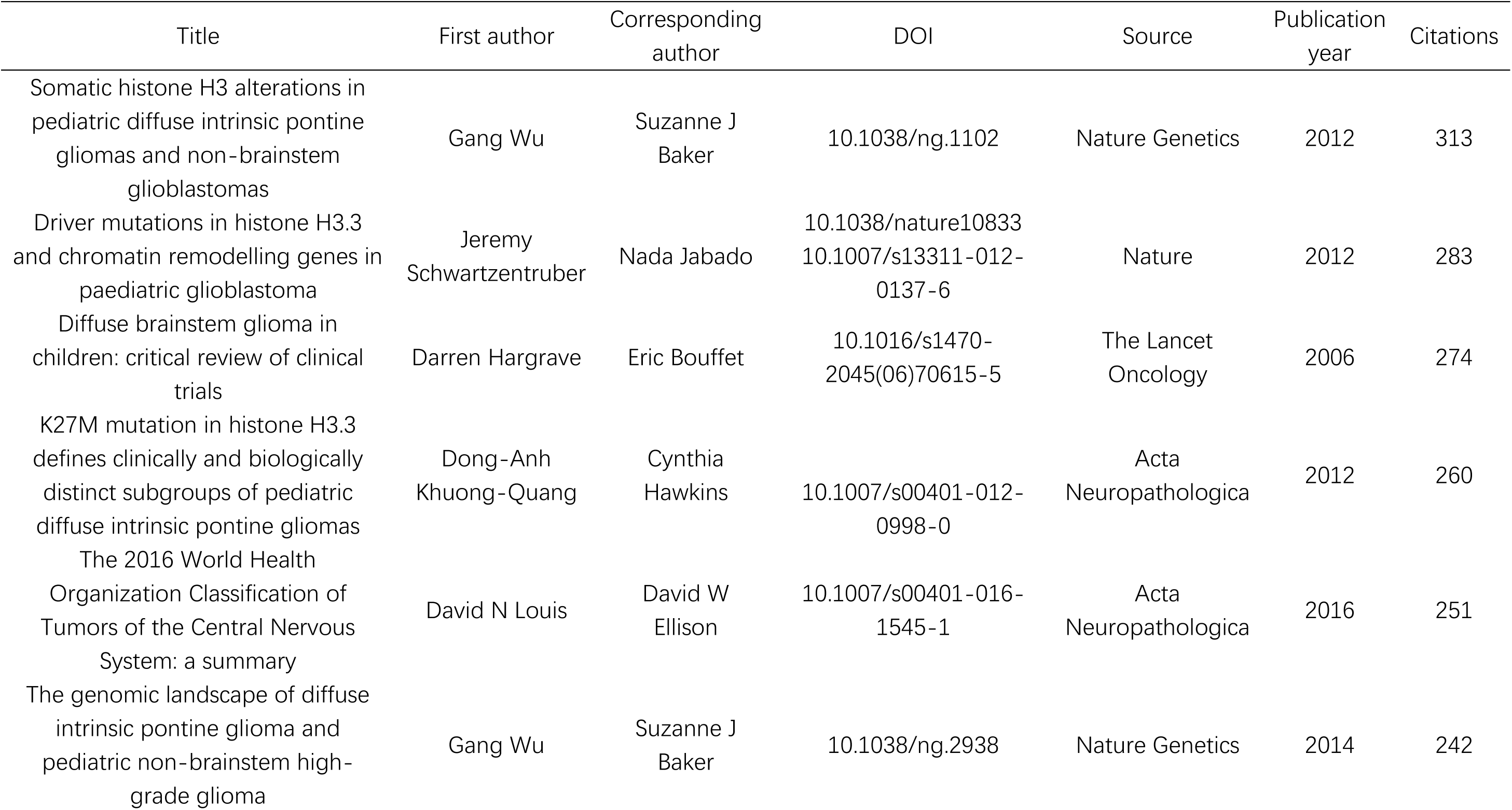
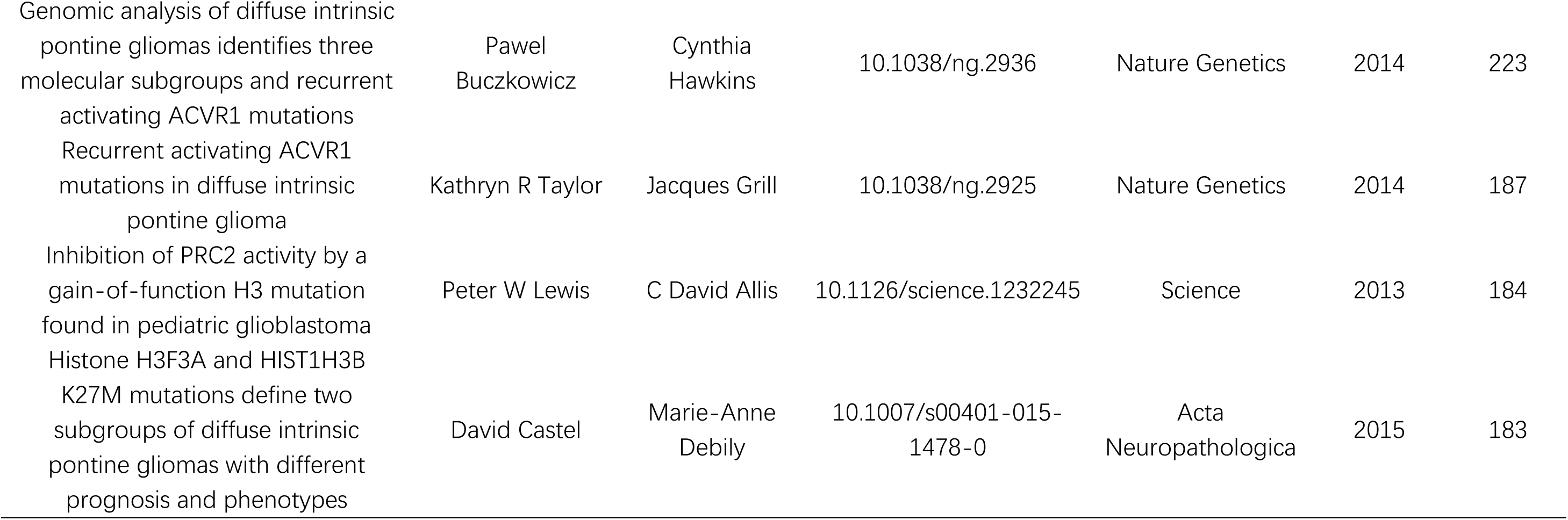
Top ten publications that have the most cited counts from collaborative analysis in brainstem tumor research.

| Title | First author | Corresponding author | DOI | Source | Publication year | Citations |
| --- | --- | --- | --- | --- | --- | --- |
| Somatic histone H3 alterations in pediatric diffuse intrinsic pontine gliomas and non-brainstem glioblastomas | Gang Wu | Suzanne J Baker | 10.1038/ng.1102 | Nature Genetics | 2012 | 313 |
| Driver mutations in histone H3.3 and chromatin remodelling genes in paediatric glioblastoma | Jeremy Schwartzentruber | Nada Jabado | 10.1038/nature10833<br>10.1007/s13311-012-0137-6 | Nature | 2012 | 283 |
| Diffuse brainstem glioma in children: critical review of clinical trials | Darren Hargrave | Eric Bouffet | 10.1016/s1470-2045(06)70615-5 | The Lancet Oncology | 2006 | 274 |
| K27M mutation in histone H3.3 defines clinically and biologically distinct subgroups of pediatric diffuse intrinsic pontine gliomas | Dong-Anh Khuong-Quang | Cynthia Hawkins | 10.1007/s00401-012-0998-0 | Acta Neuropathologica | 2012 | 260 |
| The 2016 World Health Organization Classification of Tumors of the Central Nervous System: a summary | David N Louis | David W Ellison | 10.1007/s00401-016-1545-1 | Acta Neuropathologica | 2016 | 251 |
| The genomic landscape of diffuse intrinsic pontine glioma and pediatric non-brainstem high-grade glioma | Gang Wu | Suzanne J Baker | 10.1038/ng.2938 | Nature Genetics | 2014 | 242 |
| Genomic analysis of diffuse intrinsic pontine gliomas identifies three molecular subgroups and recurrent activating ACVR1 mutations | Pawel Buczkowicz | Cynthia Hawkins | 10.1038/ng.2936 | Nature Genetics | 2014 | 223 |
| Recurrent activating ACVR1 mutations in diffuse intrinsic pontine glioma | Kathryn R Taylor | Jacques Grill | 10.1038/ng.2925 | Nature Genetics | 2014 | 187 |
| Inhibition of PRC2 activity by a gain-of-function H3 mutation found in pediatric glioblastoma | Peter W Lewis | C David Allis | 10.1126/science.1232245 | Science | 2013 | 184 |
| Histone H3F3A and HIST1H3B K27M mutations define two subgroups of diffuse intrinsic pontine gliomas with different prognosis and phenotypes | David Castel | Marie-Anne Debily | 10.1007/s00401-015-1478-0 | Acta Neuropathologica | 2015 | 183 |

### 3.6. Hotspots and frontiers

A total of 19,419 keywords were extracted from the titles and abstracts of all publications. We employed VOSviewer to visualize the network and temporal patterns of 1,011 keywords that exceeded the threshold of 10 occurrences (Figure 7A, B). The most prominent research field, represented by the keyword network, revolved around "tumors" (856 records, constituting 14.4% of all articles), followed closely by "children" (704 records, 11.9%), "management" (446 records, 7.5%), "expression" (433 records, 7.3%), and "radiotherapy" (378 records, 6.4%). These keywords formed four distinct clusters: "children tumor," "gene expression," "radiotherapy," and "surgery management" (Figure 7A). Recent trends in brainstem tumor research have witnessed a notable surge in epigenetic studies and investigations into histone-related aspects (Figure 7B). This indicates a growing focus on the epigenetic mechanisms and histone alterations within the realm of brainstem tumor research.

**Figure 7.**
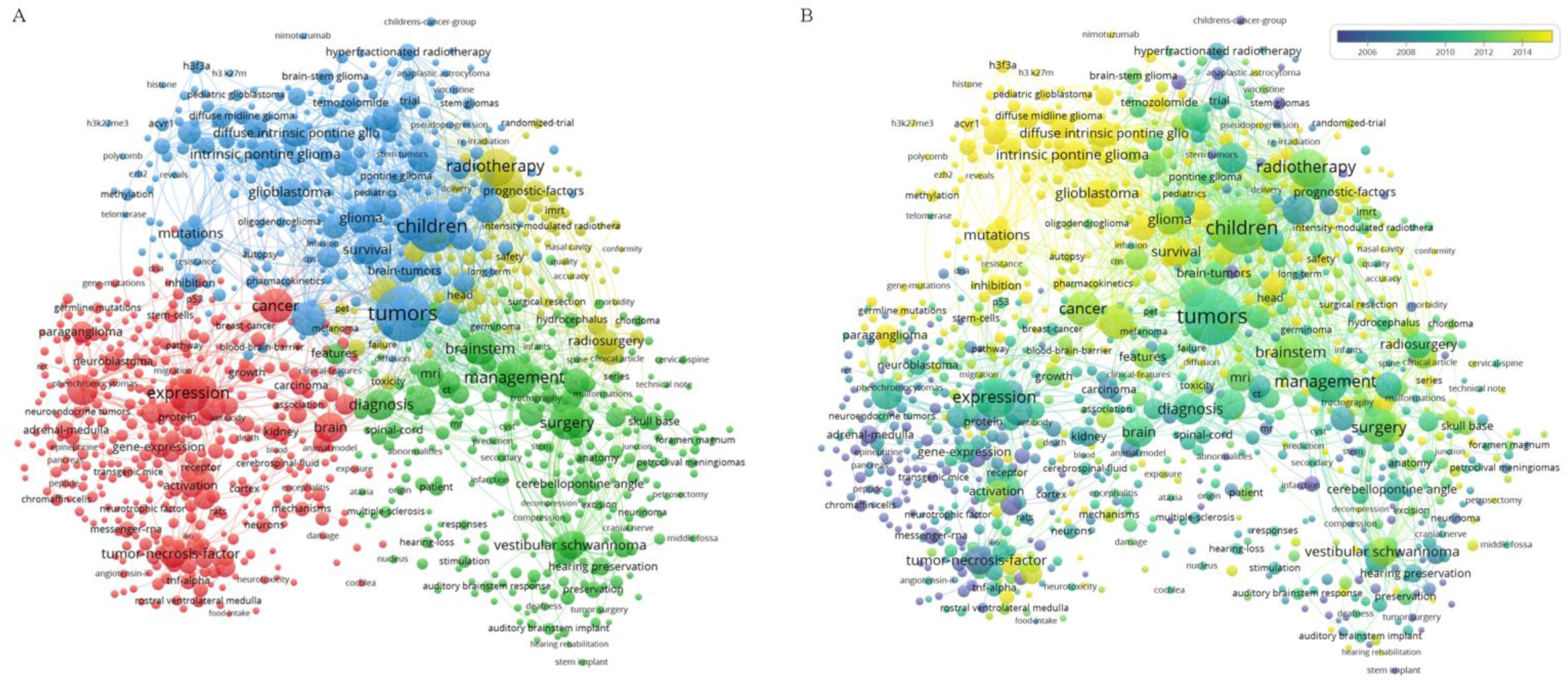
Co-occurrence analysis of keywords. **(A)** Cluster visualization of keywords. **(B)** Timeline visualization of keywords.

## 4. Discussion

### 4.1. General information

In the era of information proliferation, bibliometric analysis serves as a valuable tool for researchers, enabling them to efficiently manage and visually depict knowledge structures[10,11]. Brainstem tumors, characterized by their high malignancy rates[12], remain a challenging aspect of central nervous system pathology, with incomplete understanding of their underlying mechanisms and treatment strategies. In response to this knowledge gap, we undertook an extensive bibliometric analysis of 5,927 articles concerning brainstem tumor diseases published between 1992 and 2023.

Employing robust scientific quantitative analysis and visualization techniques, we delved into various facets of this research domain, including countries of origin, journals, prolific authors, citation patterns, and emerging hotspots. Our findings unveiled a steady and persistent annual growth in publications related to brainstem tumors since 1992. This continuous upward trend signifies that the brainstem tumor field retains numerous enigmas and remains a prominent area of study over the past few decades.

In terms of national contributions, the United States (USA), China, Japan, Germany, and Italy emerged as the leading contributors in terms of publications, with the USA taking the top spot, nearly tripling the output of the second-ranked country. Notably, while the number of publications from China accounted for only one-fourth of that from the USA, the total citations from China were merely one-tenth of those from the USA. This observation suggests that the USA maintains a more substantial research depth and produces high-quality publications compared to other countries. It is worth highlighting that two articles authored by Gang Wu and Jeremy Schwartzentruber in 2012 emerged as the two most highly cited references, both focusing on histone H3 (to be discussed in the following paragraph). Among the top five productive institutions in this field, four are located in the USA, namely St. Jude Children’s Research Hospital, University of California San Francisco, Memorial Sloan Kettering Cancer Center, and Duke University, with Capital Medical University representing China. Interestingly, among the authors contributing to this field, Liwei Zhang stood out as the most prolific author with 65 articles, while no Chinese author appeared in the top ten list of most cited authors.

The h-index serves as a valuable metric for evaluating the academic impact of researchers, shedding light on their contributions to the field[13]. In the case of brainstem tumor research, notable figures like Amar Gajjar, LE Kun, and Roger J. Packer have left significant imprints. Amar Gajjar’s work has primarily revolved around pioneering therapies and clinical trials for childhood brain tumors, with a particular focus on medulloblastomas, PNET, and Rhabdoid tumors. LE Kun has dedicated attention to radiation therapy for pediatric brain tumors, establishing pediatric brain tumor treatment programs in the 1990s. Roger J. Packer’s contributions span clinical and applied basic science research, with a focus on pediatric brain tumors, neurogenetic diseases, and NF1.

While the h-index effectively gauges academic impact over one’s scientific career, it tends to disadvantage junior and potentially promising scientists due to their shorter career lengths. To address this, the m-index, introduced by Leo Egghe, was created to provide a more balanced assessment. Notably, the most promising authors based on m-index rankings include Angel Montero Carcaboso, Michelle Monje, and Stefan M Pfister. These individuals are all under 50 years of age, indicating their potential for long-lasting contributions to brainstem tumor-related research. Angel Montero Carcaboso’s work spans multidisciplinary and translational efforts in the field of pediatric oncology, including the development of patient-derived tumor cell models and xenografts. Michelle Monje has delved into the mechanisms and consequences of neuron-glial interactions in malignant glioma, conducting translational studies and advancing novel therapeutics for pediatric glioma. Stefan M Pfister has made significant strides in the genetic and epigenetic characterization of childhood brain tumors, with a focus on identifying novel clinical options.

The analysis of journals and co-cited journals offers valuable insights into the landscape of publications, aiding scholars in identifying suitable journals for their research endeavors[14]. Since 1992, a total of 1,285 journals have disseminated articles in the field of brainstem tumors. It’s noteworthy that the top four journals with the highest number of publications all belong to the neuro-specialized category. This observation underscores that brainstem tumor research continues to occupy a niche within the academic landscape, making it challenging to publish research in high-impact factor journals. Conversely, an intriguing revelation emerges when examining the most cited journals. Four out of the top five most cited journals reside in the JCR Q1 category, denoting their substantial influence and commitment to maintaining high-quality publications in this field.

### 4.2. Research focus and future research directions

As surgical techniques and molecular biology have advanced, the focal points of brainstem tumor research have shifted from surgery and chemoradiotherapy to the realms of abnormal gene expression, epigenomics, and immune-based treatments. The primary goal of surgery in brainstem tumors has transitioned from seeking a cure to obtaining tumor samples and mitigating tumor volume. Neuro-navigation and fiber reconstruction have been instrumental in pushing the boundaries of surgical intervention. Genetic and epigenetic alterations have emerged as pivotal drivers of dysregulated gene expression, exerting a profound influence on the initiation and progression of brainstem tumors. These molecular aberrations are central in understanding the underlying mechanisms of tumorigenesis and hold significant promise for the development of innovative therapeutic approaches[15]. As depicted in Figure 7b, recent studies have prominently featured recurring keywords such as "diffuse midline glioma," "h3f3a," "methylation," "nimotuzumab," and "ezh2." These keywords underscore the growing focus on various aspects of brainstem tumor research. Key areas of interest encompass tumorigenesis, driver mutations, epigenetic alterations, neuron-glial-tumor interactions, immune evasion by tumors, and targeted therapies. These avenues of research are likely to shape the future of brainstem tumor investigation and potential therapeutic directions. Furthermore, the integration of nanomaterials and interdisciplinary approaches is poised to revolutionize drug delivery systems. This innovation holds the promise of providing precise and low-toxicity treatments, thereby advancing the field of precision medicine.

### 4.3. Clinical trial for diffuse midline glioma

We summarized the ongoing clinical trial for diffuse midline glioma to figure out the expectation in the recent future and encourage more bench-to-bed research (Table 4). Generally, 28 clinical trials were registered in official organization (https://classic.clinicaltrials.gov/). Remarkably, more than a quarter of these trials (9 out of 28) are directly targeting the immune system, emphasizing the growing prominence of immune therapies as a potentially promising avenue for treatment. Conversely, only one-tenth of the trials (3 out of 28) are centered on traditional radiochemotherapy. Additionally, it is noteworthy that eight out of the 28 clinical trials specifically focus on histone H3, a star mutation and a well-established hotspot within the domain of brainstem tumor research. This underscores the significance of histone H3-related investigations in the ongoing pursuit of effective treatments for diffuse midline glioma.

**Table 4.** Summary of clinical trials of diffuse midline glioma (Recruiting and Not yet recruiting)

| Identifier | Status | Begin<br>year<br>/Phase | Title | Disease | Treatment |
| --- | --- | --- | --- | --- | --- |
| NCT05762419 | Recruiting | 2023/I | FUS Etoposide for DMG - A Feasibility Study | DIPG and DMG,<br>H3K27M | Ultrasound with neuro-<br>navigator-controlled<br>sonication+Etoposide |
| NCT05544526 | Not yet<br>recruiting | 2022/I | CAR T Cells to Target GD2 for DMG<br>(CARMIGO) | DMG | GD2 CAR T cells |
| NCT05478837 | Recruiting | 2022/I | Genetically Modified Cells (KIND T Cells) for<br>the Treatment of HLA-A*0201-Positive<br>Patients With H3.3K27M-Mutated Glioma | DMG, HLA-A*0201-positive | Autologous Anti-H3.3K27M TCR-expressing T-cells |
| NCT04943848 | Recruiting | 2021/I | rHSC-DIPGVax Plus Checkpoint Blockade for<br>the Treatment of Newly Diagnosed DIPG<br>and DMG | DIPG and DMG,<br>H3K27M | rHSC-DIPGVax + BALSTILIMAB +<br>ZALIFRELIMAB |
| NCT05768880 | Recruiting | 2023/I | Study of B7-H3, EGFR806, HER2, And IL13-Zetakine (Quad) CAR T Cell Locoregional<br>Immunotherapy For Pediatric Diffuse<br>Intrinsic Pontine Glioma, Diffuse Midline<br>Glioma, And Recurrent Or Refractory Central<br>Nervous System Tumors | DIPG, DMG,<br>Recurrent CNS<br>Tumor | SC-CAR4BRAIN |
| NCT04185038 | Recruiting | 2019/I | Study of B7-H3-Specific CAR T Cell<br>Locoregional Immunotherapy for Diffuse<br>Intrinsic Pontine Glioma/Diffuse Midline | DIPG, DMG,<br>Pediatric Central<br>Nervous System<br>Tumors | B7-H3-Specific CAR T Cell |

| Glioma and Recurrent or Refractory Pediatric Central Nervous System Tumors |  |  |  |  |  |
| --- | --- | --- | --- | --- | --- |
| NCT04196413 | Recruiting | 2019/I | GD2 CAR T Cells in Diffuse Intrinsic Pontine Gliomas(DIPG) & Spinal Diffuse Midline Glioma(DMG) | DIPG | GD2 CAR T cells |
| NCT05298995 | Not yet recruiting | 2022/I | GD2-CAR T Cells for Pediatric Brain Tumours | Pediatric Brain Tumor | iC9-GD2-CAR T-cells |
| NCT05835687 | Recruiting | 2023/I | Loc3CAR: Locoregional Delivery of B7-H3-CAR T Cells for Pediatric Patients With Primary CNS Tumors | DMG | B7-H3-CAR T cells |
| NCT04808245 | Recruiting | 2021/I | A MulticENTER Phase I Peptide VaCcine Trial for the Treatment of H3-Mutated Gliomas | H3-mutated Glioma | H3K27M peptide vaccine |
| NCT04732065 | Recruiting | 2021/I | ONC206 for Treatment of Newly Diagnosed, or Recurrent Diffuse Midline Gliomas, and Other Recurrent Malignant CNS Tumors | DMG, Recurrent Malignant CNS Tumors | ONC206+Radiation |
| NCT04541082 | Recruiting | 2020/I | Phase I Study of Oral ONC206 in Recurrent and Rare Primary Central Nervous System Neoplasms | Glioma | ONC206 |
| NCT05009992 | Recruiting | 2021/II | Combination Therapy for the Treatment of Diffuse Midline Gliomas | DIPG, DMG, Glioma (WHO III) | ONC201, Radiation, Paxalisib combination |
| NCT05476939 | Recruiting | 2022/III | Biological Medicine for Diffuse Intrinsic Pontine Glioma (DIPG) Eradication 2.0 | DIPG, DMG | ONC201 vs everolimus |
| NCT04771897 | Recruiting | 2021/I | A Study of BXQ-350 in Children With Newly Diagnosed Diffuse Intrinsic Pontine Glioma (DIPG) or Diffuse Midline Glioma (DMG) | DIPG and DMG | BXQ-350 |
| NCT05077735 | Recruiting | 2021/II | Stereotactic Biopsy Split-Course Radiation Therapy in Diffuse Midline Glioma, SPORT-DMG Study | DMG | Hypofractionated Radiation Therapy |
| NCT06000787 | Not yet recruiting | 2023 | MCT for the Harvard/UCSF ROBIN Center | DMG | External beam radiotherapy |
| NCT03101813 | Recruiting | 2017 | International Diffuse Intrinsic Pontine Glioma (DIPG)/Diffuse Midline Glioma (DMG) Registry and Repository | DIPG and DMG | NA |
| NCT05259605 | Recruiting | 2022 | Observational Study for Assessing Treatment and Outcome of Patients With Primary Brain Tumours Using cIMPACT-NOW and 2021 WHO Classification | Glioma | NA |
| NCT05099003 | Recruiting | 2021/I+II | A Study of the Drug Selinexor With Radiation Therapy in Patients With Newly-Diagnosed Diffuse Intrinsic Pontine (DIPG) Glioma and High-Grade Glioma (HGG) | DIPG, HGG | Selinexor + Radiation Therapy |
| NCT05500508 | Recruiting | 2022/I+II | Oral AMXT 1501 Dicaprate in Combination With IV DFMO | Malignant tumor | AMXT<br>1501+Difluoromethylornithine |
| NCT03243461 | Recruiting | 2017/III | International Cooperative Phase III Trial of the HIT-HGG Study Group (HIT-HGG-2013) | DIPG, DMG, GBM, anaplastic astrocytoma | Temozolomide + Valproic Acid vs Temozolomide |
| NCT05839379 | Recruiting | 2023 | Targeted Pediatric High-Grade Glioma Therapy | HGG | NA |
| NCT04264143 | Recruiting | 2020/I | CED of MTX110 Newly Diagnosed Diffuse Midline Gliomas | DIPG/DTG/DMG | CED infusate MTX110 and gadolinium |
| NCT04908176 | Recruiting | 2021/I | A Drug-drug Interaction Study of Avapritinib and Midazolam | CNS tumor | Avapritinib+midazolam |
| NCT04870944 | Recruiting | 2021/I+II | CBL0137 for the Treatment of Relapsed or Refractory Solid Tumors, Including CNS Tumors and Lymphoma | DMG, Metastasis, Refractory CNS tumor | CBL0137 |
| NCT05413304 | Recruiting | 2022/I | Abemaciclib Neuropharmacokinetics of Diffuse Midline Glioma Using Intratumoral Microdialysis | DMG | abemaciclib + temozolomide |
| NCT05843253 | Not yet recruiting | 2023/II | Study of Ribociclib and Everolimus in HGG and DIPG | DIPG | ribociclib+everolimus vs traditional |

### 4.4. Limitation

Our study, while informative, does present certain limitations that warrant consideration. Firstly, our analysis was solely based on data from the Web of Science Core Collection (WOSCC) database. A more comprehensive view could have been achieved by incorporating additional databases such as Scopus and PubMed, thereby potentially capturing a broader spectrum of research output. Secondly, it’s important to acknowledge that cutting-edge research and emerging technologies may be underrepresented in our study, as they may not yet have accumulated substantial citation counts. This could result in a potential underestimation of the latest advancements in the field. Lastly, it’s worth noting that some of the papers included in our analysis may not have been specifically focused on brainstem tumors, which could introduce a degree of bias into our findings. However, we believe that this bias is unlikely to significantly impact the overarching trends identified in our study.

## 5. Conclusion

In summary, our comprehensive bibliometric analysis underscores the enduring significance of brainstem tumors within the realm of cancer research. The quantity and caliber of brainstem tumor studies exhibit consistent growth trends, reflecting the sustained interest and commitment of the scientific community. The United States plays a prominent leadership role in this field, while the contributions of a notable group of Chinese neuroscientists have become increasingly prominent in the past decade. Their collective efforts have enriched the global body of knowledge in brainstem tumor research. Furthermore, our analysis illuminates a significant transformation in the research frontiers of brainstem tumors, with a shift from traditional approaches like surgery and chemoradiotherapy towards a greater emphasis on epigenomics and immune-based therapies. This evolving landscape underscores the dynamic nature of scientific inquiry in this field.

In conclusion, our study has conducted a rigorous and systematic bibliometric analysis of the existing literature on brainstem tumors, providing a data-driven reference point for future investigations and guiding the direction of research endeavors in this vital area of study.

## Data Availability

All data produced in the present work are contained in the manuscript

## 6. Acknowledgements

This work was supported by the Beijing Chao-yang Hospital Golden Seed foundation (No. CYJZ202201), the inner-hospital foundation of Beijing Chao-yang Hospital, and the National Natural Science Foundation of China (No. 81960330).

## 7. Author contributions

YG and XL contributed to conception and design of the study. YG and LX performed the bibliometric analysis. YG and JL wrote the first draft of the manuscript. XL and YW supervised the study. All authors contributed to manuscript revision, read, and approved the submitted version.

