## Supplemental Material 1 for "Bibliometric analysis of emerging trends and research foci in brainstem tumor field over thirty years (1992-2023)": savedrecs (1).txt

FN Clarivate Analytics Web of Science
VR 1.0
PT J
AU Darken, RS
   Bogitch, R
   Leonard, J
   Perry, A
   McKinstry, RC
   Gutmann, DH
   Rubin, JB
AF Darken, Rachel S.
   Bogitch, Ray
   Leonard, Jeffrey
   Perry, Arie
   McKinstry, Robert C.
   Gutmann, David H.
   Rubin, Joshua B.
TI Brainstem Glioma Presenting as Pruritus in Children With
   Neurofibromatosis-1
SO JOURNAL OF PEDIATRIC HEMATOLOGY ONCOLOGY
LA English
DT Article
DE NF1; brainstem glioma; pruritus; fibrillary astrocytoma
ID UNILATERAL NEUROGENIC PRURITUS; SPINAL-CORD TUMOR; MULTIPLE-SCLEROSIS;
   ITCH; SYMPTOM; PATHWAY
AB Children with neurofibromatosis type 1 (NF1) are at increased risk of developing tumors of the nervous system. The most common intracranial tumors are low-grade astrocytomas, which most frequently involve the optic pathway, and less often, the brainstem. In cases of brainstem glioma in NF1, treatment decisions are frequently complicated by the paucity of symptoms referable to the tumor. Here, we describe 2 children with NF1 whose initial presentation of a growing brainstem glioma was localized pruritus. This unusual presentation emphasizes the importance of appreciating subtle, often hard to localize, symptoms in this high-risk population.
C1 [Rubin, Joshua B.] Washington Univ, Sch Med, Dept Pediat, Div Hematol Oncol, St Louis, MO 63119 USA.
   [Darken, Rachel S.; Bogitch, Ray; Gutmann, David H.] Washington Univ, Sch Med, Dept Neurol, St Louis, MO 63119 USA.
   [Leonard, Jeffrey] Washington Univ, Sch Med, Dept Neurosurg, St Louis, MO 63119 USA.
   [Perry, Arie] Washington Univ, Sch Med, Dept Pathol Neuropathol, St Louis, MO 63119 USA.
   [McKinstry, Robert C.] Washington Univ, Sch Med, Dept Radiol Neuroradiol, St Louis, MO 63119 USA.
C3 Washington University (WUSTL); Washington University (WUSTL); Washington
   University (WUSTL); Washington University (WUSTL); Washington University
   (WUSTL)
RP Rubin, JB (通讯作者)，Washington Univ, Sch Med, Dept Pediat, Div Hematol Oncol, Campus Box 8208,660 S Euclid Ave, St Louis, MO 63119 USA.
EM
OI Gutmann, David/0000-0002-3127-5045; Perry, Arie/0000-0002-8300-7261
CR Andreev V C, 1971, Z Haut Geschlechtskr, V46, P544
   Andrew D, 2001, NAT NEUROSCI, V4, P72, DOI 10.1038/82924
   Dey DD, 2005, PAIN, V113, P233, DOI 10.1016/j.pain.2004.09.032
   Friedman JM, 1999, NEUROFIBROMATOSIS PH, V3e, P29
   HABIBY R, 1995, J PEDIATR-US, V126, P364, DOI 10.1016/S0022-3476(95)70449-3
   Johnson RE, 2000, J AM ACAD DERMATOL, V43, P958, DOI 10.1067/mjd.2000.104000
   Khaw SL, 2007, PEDIATR BLOOD CANCER, V49, P808, DOI 10.1002/pbc.21270
   KING CA, 1982, ANN INTERN MED, V97, P222, DOI 10.7326/0003-4819-97-2-222
   Leonard JR, 2006, NEUROLOGY, V67, P1509, DOI 10.1212/01.wnl.0000240076.31298.47
   LISTERNICK R, 1994, J PEDIATR-US, V125, P63, DOI 10.1016/S0022-3476(94)70122-9
   Magilner D, 2006, PEDIATR EMERG CARE, V22, P746, DOI 10.1097/01.pec.0000238735.14532.2e
   MASSEY EW, 1984, STROKE, V15, P901, DOI 10.1161/01.STR.15.5.901
   Matthews WB, 1991, MC ALPINES MULTIPLE, P43
   OSTERMAN PO, 1975, BRAIN, V98, P189, DOI 10.1093/brain/98.2.189
   Pollack IF, 1996, NEUROLOGY, V46, P1652, DOI 10.1212/WNL.46.6.1652
   RICCARDI VM, 1981, BIRTH DEFECTS-ORIG, V17, P129
   Shabtai H, 1996, NEUROLOGY, V46, P940, DOI 10.1212/WNL.46.4.940
   SULLIVAN MJ, 1984, NEUROLOGY, V34, P828, DOI 10.1212/WNL.34.6.828
   SUMMERS CG, 1988, J CHILD NEUROL, V3, P189, DOI 10.1177/088307388800300308
   Via PD, 2007, NEURO-ONCOLOGY, V9, P430, DOI 10.1215/15228517-2007-031
   Wallengren Joanna, 2005, Dermatol Ther, V18, P292
   YAMAMOTO M, 1981, J NEUROL NEUROSUR PS, V44, P19, DOI 10.1136/jnnp.44.1.19
NR 22
TC 13
Z9 13
U1 0
U2 4
PU LIPPINCOTT WILLIAMS & WILKINS
PI PHILADELPHIA
PA TWO COMMERCE SQ, 2001 MARKET ST, PHILADELPHIA, PA 19103 USA
SN 1077-4114
EI 1536-3678
J9 J PEDIAT HEMATOL ONC
JI J. Pediatr. Hematol. Oncol.
PD DEC
PY 2009
VL 31
IS 12
BP 972
EP 976
DI 10.1097/MPH.0b013e3181b8701f
PG 5
WC Oncology; Hematology; Pediatrics
WE Science Citation Index Expanded (SCI-EXPANDED)
SC Oncology; Hematology; Pediatrics
GA 531IQ
UT WOS:000272658700020
PM 19935099
DA 2023-08-27
ER

PT J
AU Ramos, A
   Hilario, A
   Lagares, A
   Salvador, E
   Perez-Nunez, A
   Sepulveda, J
AF Ramos, Ana
   Hilario, Amaya
   Lagares, Alfonso
   Salvador, Elena
   Perez-Nunez, Angel
   Sepulveda, Juan
TI Brainstem Gliomas
SO SEMINARS IN ULTRASOUND CT AND MRI
LA English
DT Article
ID MAGNETIC-RESONANCE SPECTROSCOPY; GAMMA-KNIFE RADIOSURGERY;
   PEDIATRIC-ONCOLOGY-GROUP; LOW-GRADE GLIOMAS; NEUROFIBROMATOSIS TYPE-1;
   PROGNOSTIC-FACTORS; NATURAL-HISTORY; HYPERFRACTIONATED RADIOTHERAPY; MR
   SPECTROSCOPY; PONTINE TUMORS
AB Historically, brainstem gliomas have been considered as a single entity. Since the introduction of magnetic resonance (MR) imaging in the late 1980s, these tumors are now regarded as a heterogeneous group of neoplasms with different age of onset, clinical and radiologic presentation, and varying behavior and natural history. This article describes the different subtypes of brainstem gliomas in children and adults. We focus on recent advances in MR such as MR spectroscopy, MR perfusion, and diffusion tensor imaging that often strongly suggest the histopathologic diagnosis of the lesion. Semin Ultrasound CT MRI 34:104-112 (C) 2013 Elsevier Inc. All rights reserved.
C1 [Ramos, Ana; Hilario, Amaya; Salvador, Elena] Hosp 12 Octubre, Dept Radiol, E-28041 Madrid, Spain.
   [Lagares, Alfonso; Perez-Nunez, Angel] Hosp 12 Octubre, Dept Neurosurg, E-28041 Madrid, Spain.
   [Sepulveda, Juan] Hosp 12 Octubre, Dept Med Oncol, E-28041 Madrid, Spain.
C3 Hospital Universitario 12 de Octubre; Hospital Universitario 12 de
   Octubre; Hospital Universitario 12 de Octubre
RP Ramos, A (通讯作者)，Hosp 12 Octubre, Dept Radiol, E-28041 Madrid, Spain.
EM
RI Perez-Nuñez, Angel/AAG-4422-2020; Hilario, Amaya/IUO-1589-2023; Lagares,
   Alfonso/B-2969-2011
OI Perez-Nuñez, Angel/0000-0002-2391-6586; Lagares,
   Alfonso/0000-0003-3996-0554; Salvador, Elena/0000-0002-8324-6502
CR Albers AC, 2009, EXPERT REV NEUROTHER, V9, P535, DOI [10.1586/ern.09.4, 10.1586/ERN.09.4]
   ALBRIGHT AL, 1986, J NEUROSURG, V65, P751, DOI 10.3171/jns.1986.65.6.0751
   Barajas RF, 2009, AM J NEURORADIOL, V30, P367, DOI 10.3174/ajnr.A1362
   Bilaniuk LT, 1997, NEURORADIOLOGY, V39, P642, DOI 10.1007/s002340050484
   Bonawitz C, 1998, AM J NEURORADIOL, V19, P541
   Bowers DC, 2000, PEDIATR NEUROSURG, V32, P24, DOI 10.1159/000028893
   Broniscer A, 1997, NEUROSURGERY, V40, P331, DOI 10.1097/00006123-199702000-00018
   Constantini S, 1996, J NEURO-ONCOL, V28, P193
   Daglioglu E, 2003, PEDIATR NEUROSURG, V38, P223, DOI 10.1159/000069823
   Donaldson SS, 2006, J CLIN ONCOL, V24, P1266, DOI 10.1200/JCO.2005.04.6599
   EIFEL PJ, 1987, INT J RADIAT ONCOL, V13, P847, DOI 10.1016/0360-3016(87)90098-8
   Frazier JL, 2009, J NEUROSURG-PEDIATR, V3, P259, DOI 10.3171/2008.11.PEDS08281
   FREEMAN CR, 1993, INT J RADIAT ONCOL, V27, P197, DOI 10.1016/0360-3016(93)90228-N
   Freeman CR, 1998, INT J RADIAT ONCOL, V40, P265, DOI 10.1016/S0360-3016(97)00572-5
   Fuchs I, 2002, ACT NEUR S, V84, P85
   Goncalves-Ferreira AJ, 2003, SURG NEUROL, V60, P311, DOI 10.1016/S0090-3019(03)00379-3
   Guillamo JS, 2001, CURR OPIN NEUROL, V14, P711, DOI 10.1097/00019052-200112000-00006
   Guillamo JS, 2001, BRAIN, V124, P2528, DOI 10.1093/brain/124.12.2528
   Gururangan S, 2002, J CLIN ONCOL, V20, P2951, DOI 10.1200/JCO.2002.12.008
   Helton KJ, 2008, J NEUROSURG-PEDIATR, V1, P270, DOI 10.3171/PED/2008/1/4/270
   Helton KJ, 2006, AM J NEURORADIOL, V27, P786
   Jallo GI, 2004, CHILD NERV SYST, V20, P143, DOI 10.1007/s00381-003-0870-6
   Kaplan AM, 1996, PEDIATR NEUROSURG, V24, P185, DOI 10.1159/000121036
   Kesari S, 2008, J NEURO-ONCOL, V88, P175, DOI 10.1007/s11060-008-9545-1
   KHATIB ZA, 1994, PEDIATR NEUROSURG, V20, P2, DOI 10.1159/000120759
   KIHLSTROM L, 1994, ACT NEUR S, V62, P55
   Laigle-Donadey F, 2008, CURR OPIN ONCOL, V20, P662, DOI 10.1097/CCO.0b013e32831186e0
   Landolfi JC, 1998, NEUROLOGY, V51, P1136, DOI 10.1212/WNL.51.4.1136
   Laprie A, 2005, INT J RADIAT ONCOL, V62, P20, DOI 10.1016/j.ijrobp.2004.09.027
   Lashford LS, 2002, J CLIN ONCOL, V20, P4684, DOI 10.1200/JCO.2002.08.141
   Lesniak MS, 2003, PEDIATR NEUROSURG, V39, P314, DOI 10.1159/000075260
   Mandell LR, 1999, INT J RADIAT ONCOL, V43, P959, DOI 10.1016/S0360-3016(98)00501-X
   Moller-Hartmann W, 2002, NEURORADIOLOGY, V44, P371, DOI 10.1007/s00234-001-0760-0
   MOLLOY PT, 1995, NEUROLOGY, V45, P1897, DOI 10.1212/WNL.45.10.1897
   Nafe R, 2003, J NEURO-ONCOL, V63, P233, DOI 10.1023/A:1024249232454
   Pincus DW, 2006, J NEUROSURG, V104, P108, DOI 10.3171/ped.2006.104.2.108
   Pirotte B, 2007, CHILD NERV SYST, V23, P739, DOI 10.1007/s00381-007-0307-8
   POLLACK IF, 1993, J NEUROSURG, V78, P859, DOI 10.3171/jns.1993.78.6.0859
   Pollack IF, 1996, NEUROLOGY, V46, P1652, DOI 10.1212/WNL.46.6.1652
   Porto L, 2007, CHILD NERV SYST, V23, P305, DOI 10.1007/s00381-006-0221-5
   Poussaint TY, 1999, PEDIATR RADIOL, V29, P662, DOI 10.1007/s002470050671
   Rachinger W, 2009, J NEUROL NEUROSUR PS, V80, P1134, DOI 10.1136/jnnp.2009.174250
   Salgado JV, 2007, CLIN NEUROL NEUROSUR, V109, P379, DOI 10.1016/j.clineuro.2007.01.002
   Salmaggi A, 2008, J NEUROL, V255, P171, DOI 10.1007/s00415-008-0589-0
   Schumacher M, 2007, J NEUROSURG, V106, P111, DOI 10.3171/ped.2007.106.2.111
   Selvapandian S, 1999, ACTA NEUROCHIR, V141, P721, DOI 10.1007/s007010050367
   Sousa P, 2004, NEUROCIRUGIA, V15, P56
   Teo C, 2008, CHILD NERV SYST, V24, P1307, DOI 10.1007/s00381-008-0647-z
   Thakur SB, 2006, AM J NEURORADIOL, V27, P806
   Turner CD, 2007, J NEURO-ONCOL, V82, P95, DOI 10.1007/s11060-006-9251-9
   Walker DA, 1999, ARCH DIS CHILD, V80, P558, DOI 10.1136/adc.80.6.558
   Weiner HL, 1997, PEDIATR NEUROSURG, V27, P12, DOI 10.1159/000121219
   Yohay K, 2009, CURR NEUROL NEUROSCI, V9, P247, DOI 10.1007/s11910-009-0036-3
NR 53
TC 20
Z9 22
U1 0
U2 8
PU W B SAUNDERS CO-ELSEVIER INC
PI PHILADELPHIA
PA 1600 JOHN F KENNEDY BOULEVARD, STE 1800, PHILADELPHIA, PA 19103-2899 USA
SN 0887-2171
J9 SEMIN ULTRASOUND CT
JI Semin. Ultrasound CT MRI
PD APR
PY 2013
VL 34
IS 2
BP 104
EP 112
DI 10.1053/j.sult.2013.01.001
PN 2
PG 9
WC Radiology, Nuclear Medicine & Medical Imaging
WE Science Citation Index Expanded (SCI-EXPANDED)
SC Radiology, Nuclear Medicine & Medical Imaging
GA 117AA
UT WOS:000316923300002
PM 23522775
DA 2023-08-27
ER

PT J
AU Wu, WH
   Tian, YJ
   Wan, H
   Ma, JY
   Song, YM
   Wang, Y
   Zhang, LW
AF Wu, Wenhao
   Tian, Yongji
   Wan, Hong
   Ma, Junyan
   Song, Yongmei
   Wang, Yu
   Zhang, Liwei
TI Expression of beta-catenin and E- and N-cadherin in human brainstem
   gliomas and clinicopathological correlations
SO INTERNATIONAL JOURNAL OF NEUROSCIENCE
LA English
DT Article
DE brainstem; gliomas; beta-catenin; E-cadherin; N-cadherin
ID HIGH-GRADE GLIOMAS; PONTINE GLIOMAS; WNT PATHWAY; TUMOR; DISSEMINATION;
   CHILDREN
AB Brainstem gliomas are usually associated with serious dysfunction and poor prognosis especially for diffuse intrinsic brainstem gliomas; however, the reasons are still unclear. Some clinical studies have suggested that the invasive ability may be different among brainstem gliomas, and the dysfunction of beta-catenin and E-and N-cadherin appears to be connected with tumor invasion and progression. In this study, the expression of beta-catenin and E-and N-cadherin was detected in 40 brainstem glioma samples using immunochemistry and was further analyzed in 18 samples using reverse transcription-polymerase chain reaction. The clinicopathological characteristics were also analyzed. The results show that there was no obvious staining for E-cadherin, but weak expression at the messenger RNA (mRNA) level could be seen in a few samples. The protein and mRNA expression levels of beta-catenin and N-cadherin were significantly associated with the pathological grades of brainstem gliomas. No significant differences in the expression levels of beta-catenin and N-cadherin were observed for age, sex, location or diffuse growing pattern. The overall survival of patients with low beta-catenin expression was longer than that with high beta-catenin expression, and there was a trend toward increased expression of N-cadherin with shorter survival; however, both of them had no statistical differences. These results demonstrate that expression of beta-catenin and N-cadherin is associated with the malignant progression of brainstem gliomas but not correlated with the diffuse and invasive growing pattern. beta-catenin and N-cadherin are potential therapeutic targets and prognostic markers for brainstem glioma, which need to be validated in a larger patient cohort.
C1 [Wu, Wenhao; Tian, Yongji; Wang, Yu; Zhang, Liwei] Capital Med Univ, Beijing Tiantan Hosp, Dept Neurosurg, Beijing 100050, Peoples R China.
   [Wan, Hong; Ma, Junyan] Capital Med Univ, Beijing Neurosurg Inst, Dept Biochem, Beijing 100050, Peoples R China.
   [Song, Yongmei] Chinese Acad Med Sci, Canc Inst & Hosp, State Key Lab Mol Oncol, Beijing 100730, Peoples R China.
   [Song, Yongmei] Peking Union Med Coll, Beijing 100021, Peoples R China.
C3 Capital Medical University; Beijing Neurosurgical Institute; Capital
   Medical University; Chinese Academy of Medical Sciences - Peking Union
   Medical College; Cancer Institute & Hospital - CAMS; Chinese Academy of
   Medical Sciences - Peking Union Medical College; Peking Union Medical
   College
RP Zhang, LW (通讯作者)，Capital Med Univ, Beijing Tiantan Hosp, Dept Neurosurg, 6 Tiantan Xili, Beijing 100050, Peoples R China.
EM
OI song, yongmei/0000-0002-7789-0158
FU National Natural Science Foundation of China [30772237, 30900479];
   Capital Medical Development Foundation [2009-1040]; Beijing Science and
   Technology New Star Program [2010B121]
FX This work was supported by the National Natural Science Foundation of
   China (No. 30772237 and 30900479), the Capital Medical Development
   Foundation (No. 2009-1040) and Beijing Science and Technology New Star
   Program (No. 2010B121).
CR Asano K, 2000, NEUROSURG REV, V23, P39, DOI 10.1007/s101430050030
   Asano K, 2004, J NEURO-ONCOL, V70, P3, DOI 10.1023/B:NEON.0000040811.14908.f2
   Barami Kaveh, 2006, Neurosurg Focus, V21, pE13
   D'Urso PI, 2011, J NEURO-ONCOL, V104, P113, DOI 10.1007/s11060-010-0481-5
   Fangusaro J, 2009, J CHILD NEUROL, V24, P1409, DOI 10.1177/0883073809338960
   Huang D, 2008, WORLD J GASTROENTERO, V14, P1823, DOI 10.3748/wjg.14.1823
   Kang H, 2011, ANN SURG ONCOL, V18, P711, DOI 10.1245/s10434-010-1338-z
   Kesari S, 2008, J NEURO-ONCOL, V88, P175, DOI 10.1007/s11060-008-9545-1
   Laigle-Donadey F, 2008, CURR OPIN ONCOL, V20, P662, DOI 10.1097/CCO.0b013e32831186e0
   Lewis-Tuffin LJ, 2010, PLOS ONE, V5, DOI 10.1371/journal.pone.0013665
   Liu Q, 2008, J NEUROSURG, V109, P849, DOI 10.3171/JNS/2008/109/11/0849
   MacDonald BT, 2009, DEV CELL, V17, P9, DOI 10.1016/j.devcel.2009.06.016
   Massimino M, 2008, J NEURO-ONCOL, V87, P355, DOI 10.1007/s11060-008-9525-5
   Matsubayashi S, 2009, PATHOL RES PRACT, V205, P626, DOI 10.1016/j.prp.2009.02.011
   Matsuoka T, 2011, J SURG ONCOL, V103, P674, DOI 10.1002/jso.21854
   Nelson WJ, 2008, BIOCHEM SOC T, V36, P149, DOI 10.1042/BST0360149
   Onder TT, 2008, CANCER RES, V68, P3645, DOI 10.1158/0008-5472.CAN-07-2938
   Pu P, 2009, CANCER GENE THER, V16, P351, DOI 10.1038/cgt.2008.78
   Rosenthal MA, 2008, J CLIN NEUROSCI, V15, P237, DOI 10.1016/j.jocn.2007.04.008
   Rossi M, 2011, CANCER BIOL THER, V11, P753, DOI 10.4161/cbt.11.8.14894
   Utsuki S, 2002, J NEURO-ONCOL, V57, P187, DOI 10.1023/A:1015720220602
NR 21
TC 20
Z9 21
U1 0
U2 10
PU INFORMA HEALTHCARE
PI LONDON
PA TELEPHONE HOUSE, 69-77 PAUL STREET, LONDON EC2A 4LQ, ENGLAND
SN 0020-7454
J9 INT J NEUROSCI
JI Int. J. Neurosci.
PD MAY
PY 2013
VL 123
IS 5
BP 318
EP 323
DI 10.3109/00207454.2012.758123
PG 6
WC Neurosciences
WE Science Citation Index Expanded (SCI-EXPANDED)
SC Neurosciences & Neurology
GA 126YW
UT WOS:000317660400004
PM 23240619
DA 2023-08-27
ER

PT J
AU Alaqeel, AM
   Sabbagh, AJ
AF Alaqeel, Ahmed M.
   Sabbagh, Abdulrahman J.
TI Pediatric brainstem tumors Classifications, investigations, and growth
   patterns
SO NEUROSCIENCES
LA English
DT Article
ID PROGNOSTIC-FACTORS; NATURAL-HISTORY; TECTAL GLIOMAS; CHILDREN;
   CHILDHOOD; MANAGEMENT; DIAGNOSIS; MRI
AB Brainstem gliomas occur in 10-20% of brain tumors in pediatrics. Over the past 3 decades, the treatment of brainstem gliomas has significantly progressed as a result of the gradual advancements in microsurgical techniques, sophisticated imaging technology and, most importantly, the availability of MRI. In this article, we review the current literature on brainstem gliomas and cover diagnosis, imaging, classification, and management. Surgical approaches and intraoperative modalities to tackle operable cases of brainstem gliomas will be discussed in a follow up article.
C1 [Alaqeel, Ahmed M.] King Saud Univ, Coll Med, Div Neurosurg, Riyadh 11461, Saudi Arabia.
   [Sabbagh, Abdulrahman J.] King Saud Univ, Coll Med, Dept Surg, Riyadh 11461, Saudi Arabia.
   [Sabbagh, Abdulrahman J.] King Fahd Med City, Natl Neurosci Inst, Dept Neurosurg & Pediat Neurosurg, Riyadh 11525, Saudi Arabia.
   [Sabbagh, Abdulrahman J.] King Fahd Med City, Natl Neurosci Inst, Dept Neurosurg, Riyadh 11525, Saudi Arabia.
   King Saud Bin Abdulaziz Univ Hlth Sci, Riyadh, Saudi Arabia.
   [Alaqeel, Ahmed M.] Univ Calgary, Dept Clin Neurosci, Calgary, AB, Canada.
C3 King Saud University; King Saud University; King Saud Bin Abdulaziz
   University for Health Sciences; University of Calgary
RP Sabbagh, AJ (通讯作者)，King Fahd Med City, Natl Neurosci Inst, Dept Neurosurg & Pediat Neurosurg, POB 59046, Riyadh 11525, Saudi Arabia.
EM
OI Sabbagh, Abdulrahman J/0000-0003-4849-2166
CR Abbott R, 1996, PEDIATR NEUROSURG, V25, P41, DOI 10.1159/000121095
   Adeolu AA, 2005, NEUROL INDIA, V53, P121, DOI 10.4103/0028-3886.15081
   ALBRIGHT AL, 1993, NEUROSURGERY, V33, P1026
   Albright AL, 1996, YOUMANS NEUROLOGICAL, P2603
   [Anonymous], 2000, OPERATIVE TECHNIQUES
   Astrakas LG, 2004, CLIN CANCER RES, V10, P8220, DOI 10.1158/1078-0432.CCR-04-0603
   BARKOVICH AJ, 1991, PEDIATR NEUROSURG, V16, P73, DOI 10.1159/000120511
   Behnke J, 1997, CHILD NERV SYST, V13, P135, DOI 10.1007/s003810050059
   Beltramello A, 2000, OPERATIVE TECHNIQUES, V3, P87
   Bowers DC, 2000, PEDIATR NEUROSURG, V32, P24, DOI 10.1159/000028893
   BOYDSTON WR, 1992, PEDIATR NEUROSURG, V17, P234, DOI 10.1159/000120603
   Choux M, 2000, PEDIAT NEUROSURGERY, P471
   Daglioglu E, 2003, PEDIATR NEUROSURG, V38, P223, DOI 10.1159/000069823
   Dellaretti M, 2012, J NEUROSURG, V117, P810, DOI 10.3171/2012.7.JNS111992
   Di Maio S, 2009, CHILD NERV SYST, V25, P1401, DOI 10.1007/s00381-009-0956-x
   Donaldson SS, 2006, J CLIN ONCOL, V24, P1266, DOI 10.1200/JCO.2005.04.6599
   EPSTEIN F, 1988, J NEURO-ONCOL, V6, P309
   EPSTEIN F, 1986, J NEUROSURG, V64, P11, DOI 10.3171/jns.1986.64.1.0011
   Epstein F, 1996, PEDIATR NEUROSURG, V24, P24, DOI 10.1159/000121011
   EPSTEIN FJ, 1993, J NEUROSURG, V78, P408, DOI 10.3171/jns.1993.78.3.0408
   Farmer JP, 2001, PEDIATR NEUROSURG, V34, P206, DOI 10.1159/000056021
   FARWELL JR, 1977, CANCER, V40, P3123, DOI 10.1002/1097-0142(197712)40:6<3123::AID-CNCR2820400656>3.0.CO;2-6
   Fischbein NJ, 1996, PEDIATR NEUROSURG, V24, P9, DOI 10.1159/000121010
   Fisher PG, 2000, CANCER, V89, P1569, DOI 10.1002/1097-0142(20001001)89:7<1569::AID-CNCR22>3.0.CO;2-0
   Guillamo JS, 2001, BRAIN, V124, P2528, DOI 10.1093/brain/124.12.2528
   Hargrove D, 2006, LANCET ONCOL, V7, P241, DOI 10.1016/S1470-2045(06)70615-5
   HOFFMAN HJ, 1980, NEUROSURGERY, V7, P243, DOI 10.1227/00006123-198009000-00007
   Hong Semie, 2005, Cancer Res Treat, V37, P109, DOI 10.4143/crt.2005.37.2.109
   Jallo GI, 2004, CHILD NERV SYST, V20, P143, DOI 10.1007/s00381-003-0870-6
   Kaplan AM, 1996, PEDIATR NEUROSURG, V24, P185, DOI 10.1159/000121036
   Kesari S, 2008, J NEURO-ONCOL, V88, P175, DOI 10.1007/s11060-008-9545-1
   KHATIB ZA, 1994, PEDIATR NEUROSURG, V20, P2, DOI 10.1159/000120759
   Korones DN, 2008, PEDIATR BLOOD CANCER, V50, P227, DOI 10.1002/pbc.21154
   Kwon JW, 2006, PEDIATR RADIOL, V36, P959, DOI 10.1007/s00247-006-0256-5
   Laigle-Donadey F, 2008, CURR OPIN ONCOL, V20, P662, DOI 10.1097/CCO.0b013e32831186e0
   Louis DN, 2007, ACTA NEUROPATHOL, V114, P547, DOI 10.1007/s00401-007-0278-6
   Matson DD, 1969, NEUROSURGERY INFANCY, P469
   Mauffrey C, 2006, J CLIN NEUROSCI, V13, P431, DOI 10.1016/j.jocn.2005.05.015
   Moghrabi A, 1995, PEDIATR NEUROSURG, V23, P293, DOI 10.1159/000120974
   Nadvi SS, 1998, PEDIATR RADIOL, V28, P575, DOI 10.1007/s002470050418
   NISHIO S, 1988, J NEURO-ONCOL, V6, P245, DOI 10.1007/BF00163708
   PIERREKAHN A, 1993, J NEUROSURG, V79, P845, DOI 10.3171/jns.1993.79.6.0845
   Pollack IF, 2007, NEURO-ONCOLOGY, V9, P145, DOI 10.1215/15228517-2006-031
   POLLACK IF, 1993, J NEUROSURG, V78, P859, DOI 10.3171/jns.1993.78.6.0859
   POOL JL, 1968, J NEUROSURG, V29, P164, DOI 10.3171/jns.1968.29.2.0164
   Poussaint TY, 1999, PEDIATR RADIOL, V29, P662, DOI 10.1007/s002470050671
   ROBERTSON PL, 1995, PEDIATR NEUROSURG, V22, P65, DOI 10.1159/000120879
   ROBERTSON PL, 1994, NEUROLOGY, V44, P1798, DOI 10.1212/WNL.44.10.1798
   Rubin G, 1998, CHILD NERV SYST, V14, P167, DOI 10.1007/s003810050205
   SANFORD RA, 1988, SURG NEUROL, V30, P276, DOI 10.1016/0090-3019(88)90299-6
   Senft C, 2012, J NEURO-ONCOL, V109, P81, DOI 10.1007/s11060-012-0864-x
   STROINK AR, 1987, NEUROSURGERY, V20, P439, DOI 10.1227/00006123-198703000-00014
   Sun B, 1999, NEURORADIOLOGY, V41, P158, DOI 10.1007/s002340050723
   Walker DA, 1999, ARCH DIS CHILD, V80, P558, DOI 10.1136/adc.80.6.558
   Warren KE, 2000, J CLIN ONCOL, V18, P1020, DOI 10.1200/JCO.2000.18.5.1020
   Zimmerman RA, 1996, PEDIATR NEUROSURG, V25, P45, DOI 10.1159/000121096
NR 56
TC 2
Z9 4
U1 0
U2 0
PU RIYADH ARMED FORCES HOSPITAL
PI RIYADH
PA PO BOX 7897, RIYADH, 11159, SAUDI ARABIA
SN 1319-6138
EI 1658-3183
J9 NEUROSCIENCES
JI Neurosciences
PD APR
PY 2014
VL 19
IS 2
BP 93
EP 99
PG 7
WC Clinical Neurology
WE Science Citation Index Expanded (SCI-EXPANDED)
SC Neurosciences & Neurology
GA AF9AI
UT WOS:000335006800003
PM 24739404
DA 2023-08-27
ER

PT J
AU Dey, M
   Lin, Y
   Melkonian, S
   Lam, S
AF Dey, Mahua
   Lin, Yimo
   Melkonian, Stephanie
   Lam, Sandi
TI Prognostic factors and survival in primary adult high grade brainstem
   astrocytoma: A population based study from 1973-2008
SO JOURNAL OF CLINICAL NEUROSCIENCE
LA English
DT Article
DE Brainstem astrocytoma; Brainstem glioma; Brainstem tumor; Malignant
   glioma; SEER
ID GLIOMAS
AB Adult brainstem astrocytomas are a rare and heterogeneous group of malignancies. Most reports represent low-grade gliomas. This study used the Surveillance, Epidemiology and End Results (SEER) database to analyze the association between survival and demographic factors, tumor histology, and treatment characteristics among adult patients with high-grade brainstem astrocytoma (HGBSA). Adult patients with histologically confirmed diagnoses of primary HGBSA were studied. In univariate and multivariate analysis, we investigated the effect of demographics, tumor histology and treatment modality on survival. Overall median survival in the cohort of 240 adult patients was 7 months, with 1, 2, 5 and 10 year survival rates of 33.2%, 19.7%, 10.1%, and 8.3%, respectively. Age >50 years (hazard ratio [HR] 1.98, 95% confidence interval [CI] 1.45-2.70, p < 0.001) and grade IV versus grade III tumor (HR 1.61, 95% CI 1.15-2.26, p = 0.006) were associated with statistically significant increased mortality in multivariate analyses. Surgical intervention trended toward association with lower mortality (HR 0.68, 95% CI 0.47-1.01, p = 0.055). Our findings suggest that in patients with HGBSA, younger age and lower-grade histology are associated with better prognosis. Surgical intervention trended towards a significant association with better outcome, while radiation treatment was not associated with a statistically significant benefit in survival. (C) 2014 Elsevier Ltd. All rights reserved.
C1 [Dey, Mahua] Univ Chicago, Neurosurg Sect, Chicago, IL 60637 USA.
   [Lin, Yimo; Lam, Sandi] Baylor Coll Med, Texas Childrens Hosp, Dept Neurosurg, Houston, TX 77030 USA.
   [Melkonian, Stephanie] Univ Texas Houston, MD Anderson Canc Ctr, Dept Canc Epidemiol, Houston, TX 77030 USA.
C3 University of Chicago; Baylor College of Medicine; University of Texas
   System; University of Texas Health Science Center Houston; UTMD Anderson
   Cancer Center
RP Lam, S (通讯作者)，Baylor Coll Med, Texas Childrens Hosp, Dept Neurosurg, 6701 Fannin St,CCC Suite 1230-01, Houston, TX 77030 USA.
EM
OI Dey, Mahua/0000-0002-7024-5164; Lam, Sandi/0000-0003-3294-6637
CR Abbott R, 1996, PEDIATR NEUROSURG, V25, P41, DOI 10.1159/000121095
   Adams H, 2012, SPINE, V37, pE727, DOI 10.1097/BRS.0b013e31824584c0
   [Anonymous], 2010, CANCER
   Commission on Cancer, 2002, FORDS FAC ONC REG DA
   Guillamo JS, 2001, BRAIN, V124, P2528, DOI 10.1093/brain/124.12.2528
   Hong Semie, 2005, Cancer Res Treat, V37, P109, DOI 10.4143/crt.2005.37.2.109
   Kesari S, 2008, J NEURO-ONCOL, V88, P175, DOI 10.1007/s11060-008-9545-1
   Landolfi JC, 1998, NEUROLOGY, V51, P1136, DOI 10.1212/WNL.51.4.1136
   LINSTADT DE, 1991, INT J RADIAT ONCOL, V20, P757, DOI 10.1016/0360-3016(91)90019-Z
   Mehta VS, 2009, NEUROL INDIA, V57, P274, DOI 10.4103/0028-3886.53272
   RAJSHEKHAR V, 1995, J NEUROSURG, V82, P976, DOI 10.3171/jns.1995.82.6.0976
   Reyes-Botero G, 2012, ONCOLOGIST, V17, P388, DOI 10.1634/theoncologist.2011-0335
   Salmaggi A, 2008, J NEUROL, V255, P171, DOI 10.1007/s00415-008-0589-0
   Sanai N, 2011, J NEUROSURG, V115, P3, DOI 10.3171/2011.2.JNS10998
   Selvapandian S, 1999, ACTA NEUROCHIR, V141, P721, DOI 10.1007/s007010050367
   TOKURIKI Y, 1986, ACTA NEUROCHIR, V79, P67, DOI 10.1007/BF01407447
NR 16
TC 16
Z9 18
U1 0
U2 6
PU ELSEVIER SCI LTD
PI OXFORD
PA THE BOULEVARD, LANGFORD LANE, KIDLINGTON, OXFORD OX5 1GB, OXON, ENGLAND
SN 0967-5868
EI 1532-2653
J9 J CLIN NEUROSCI
JI J. Clin. Neurosci.
PD AUG
PY 2014
VL 21
IS 8
BP 1298
EP 1303
DI 10.1016/j.jocn.2013.12.011
PG 6
WC Clinical Neurology; Neurosciences
WE Science Citation Index Expanded (SCI-EXPANDED)
SC Neurosciences & Neurology
GA AM1JE
UT WOS:000339601400005
PM 24674698
DA 2023-08-27
ER

PT J
AU Gopalakrishnan, CV
   Dhakoji, A
   Nair, S
AF Gopalakrishnan, C. V.
   Dhakoji, Amit
   Nair, Suresh
TI Epidermoid Cyst of the Brainstem in Children: Case-Based Update
SO JOURNAL OF CHILD NEUROLOGY
LA English
DT Article
DE brainstem; epidermoid; cyst; magnetic resonance imaging; surgical
   treatment
ID TUMORS; MANAGEMENT; EXCISION
AB Epidermoid cysts are rare, slow growing, space-occupying lesions of early adulthood. Among the various locations, involvement of the brainstem is quite exceptional. In children, these tumors are extremely uncommon; only 4 purely intrinsic brainstem epidermoids are reported in the literature. The pathogenesis of true intraaxial brainstem epidermoid is unclear and is usually not considered in the preoperative differential diagnosis of brainstem tumors. We report 2 additional cases of brainstem epidermoid cysts occurring in children, underlining their clinical characteristics, the difficulties faced in the diagnostic work-up, and the surgical treatment adopted. These tumors pose a surgical challenge and attempts at aggressive removal of the cyst wall are fraught with increased risks of morbidity and mortality. On the other hand, cases treated more conservatively show good or excellent results with minimal complications. Neurosurgical judgment at the time of surgery is warranted to ensure maximum resection while minimizing postoperative neurologic deficits.
C1 [Gopalakrishnan, C. V.; Dhakoji, Amit; Nair, Suresh] SCTIMST, Dept Neurosurg, Trivandrum 695011, Kerala, India.
C3 Department of Science & Technology (India); Sree Chitra Tirunal
   Institute for Medical Sciences Technology (SCTIMST)
RP Gopalakrishnan, CV (通讯作者)，SCTIMST, Dept Neurosurg, Trivandrum 695011, Kerala, India.
EM
CR Ahmed I, 2009, J NEUROSURG-PEDIATR, V4, P91, DOI 10.3171/2009.4.PEDS08489
   ALTSCHULER EM, 1990, NEUROSURGERY, V26, P606, DOI 10.1227/00006123-199004000-00008
   ALVORD EC, 1977, ANN NEUROL, V2, P367, DOI 10.1002/ana.410020504
   BERGER MS, 1985, J NEUROSURG, V62, P214, DOI 10.3171/jns.1985.62.2.0214
   BHATIA R, 1978, NEUROL INDIA, V26, P76
   BRAUN IF, 1977, RADIOLOGY, V122, P717, DOI 10.1148/122.3.717
   Caldarelli M, 2004, J NEUROSURG, V100, P473, DOI 10.3171/ped.2004.100.5.0473
   Caldarelli M, 2001, SURG NEUROL, V56, P97, DOI 10.1016/S0090-3019(01)00542-0
   Chatterjee S, 2010, CLIN NEURORADIOL, V20, P66, DOI 10.1007/s00062-010-0031-8
   Cobbs C S, 1997, Clin Neurosurg, V44, P511
   Cruveilhier J., 1829, ANATOMIE PATHOLOGIQU, V1
   DUNN LT, 1994, CHILD NERV SYST, V10, P529, DOI 10.1007/BF00335076
   Dutt SN, 2002, OTOL NEUROTOL, V23, P84, DOI 10.1097/00129492-200201000-00019
   EEKHOF JLA, 1985, SURG NEUROL, V23, P189, DOI 10.1016/0090-3019(85)90344-1
   FOURNIER D, 1992, CHILD NERV SYST, V8, P471, DOI 10.1007/BF00274412
   GRANT FC, 1950, J NEUROSURG, V7, P190, DOI 10.3171/jns.1950.7.3.0190
   IIHARA K, 1989, SURG NEUROL, V32, P377, DOI 10.1016/0090-3019(89)90143-2
   Kachhara R, 2004, ACTA NEUROCHIR WIEN, V142, P97
   Kaylie DM, 2005, LARYNGOSCOPE, V115, P1082, DOI 10.1097/01.MLG.0000163103.36739.AC
   Kuzeyli K, 1996, NEUROSURG REV, V19, P179, DOI 10.1007/BF00512049
   LEAL O, 1978, J NEUROSURG, V48, P811, DOI 10.3171/jns.1978.48.5.0811
   Malcolm GP, 1996, NEUROSURGERY, V38, P579, DOI 10.1097/00006123-199603000-00032
   NAGASHIMA C, 1982, SURG NEUROL, V17, P172, DOI 10.1016/0090-3019(82)90269-5
   OBANA WG, 1991, J NEUROSURG, V74, P123, DOI 10.3171/jns.1991.74.1.0123
   OGAWA T, 1985, Neurologia Medico-Chirurgica, V25, P393, DOI 10.2176/nmc.25.393
   Radha Krishnan V V, 1992, Indian J Cancer, V29, P215
   Recinos PF, 2006, J NEUROSURG, V104, P285, DOI 10.3171/ped.2006.104.4.285
   RUBIN G, 1989, ACTA NEUROCHIR, V97, P1, DOI 10.1007/BF01577734
   Russel DS, 1989, PATHOLOGY TUMOURS NE, P690
   SABIN HI, 1987, NEUROSURGERY, V21, P798, DOI 10.1227/00006123-198712000-00004
   SALAZAR J, 1987, ACTA NEUROCHIR, V85, P34, DOI 10.1007/BF01402367
   Samii M, 1996, J NEUROSURG, V84, P14, DOI 10.3171/jns.1996.84.1.0014
   SCHWARTZ JF, 1978, NEUROLOGY, V28, P124, DOI 10.1212/WNL.28.2.124
   Sinha AK, 1998, NEUROL INDIA, V46, P333
   Takahashi M, 2007, NEUROL MED-CHIR, V47, P140, DOI 10.2176/nmc.47.140
   TAMPIERI D, 1989, AM J NEURORADIOL, V10, P351
   Tancredi A, 2003, ACTA NEUROCHIR, V145, P905, DOI 10.1007/s00701-003-0008-3
   ULRICH J, 1964, J NEUROSURG, V21, P1051, DOI 10.3171/jns.1964.21.12.1051
   WEAVER EN, 1979, J NEUROSURG, V51, P254, DOI 10.3171/jns.1979.51.2.0254
   Wilson CB, 1998, SURG NEUROL, V45, P540
   YAMAKAWA K, 1989, NEUROSURGERY, V24, P568, DOI 10.1227/00006123-198904000-00013
   YASARGIL MG, 1989, NEUROSURGERY, V24, P561
   Yoshizato K, 1996, SURG NEUROL, V45, P537, DOI 10.1016/0090-3019(95)00470-X
   ZHOU L-F, 1990, British Journal of Neurosurgery, V4, P211, DOI 10.3109/02688699008992726
   Ziyal IM, 2005, CHILD NERV SYST, V21, P1025, DOI 10.1007/s00381-005-1172-y
NR 45
TC 10
Z9 10
U1 0
U2 2
PU SAGE PUBLICATIONS INC
PI THOUSAND OAKS
PA 2455 TELLER RD, THOUSAND OAKS, CA 91320 USA
SN 0883-0738
EI 1708-8283
J9 J CHILD NEUROL
JI J. Child Neurol.
PD JAN
PY 2012
VL 27
IS 1
BP 105
EP 112
DI 10.1177/0883073811414709
PG 8
WC Clinical Neurology; Pediatrics
WE Science Citation Index Expanded (SCI-EXPANDED)
SC Neurosciences & Neurology; Pediatrics
GA 873LP
UT WOS:000298885100016
PM 21862831
DA 2023-08-27
ER

PT J
AU Warren, K
   Bent, R
   Wolters, PL
   Prager, A
   Hanson, R
   Packer, R
   Shih, J
   Camphausen, K
AF Warren, Katherine
   Bent, Robyn
   Wolters, Pamela L.
   Prager, Alisa
   Hanson, Ryan
   Packer, Roger
   Shih, Joanna
   Camphausen, Kevin
TI A phase 2 study of pegylated interferon a-2b (PEG-Intron (R)) in
   children with diffuse intrinsic pontine glioma
SO CANCER
LA English
DT Article
DE brainstem; glioma; interferon; pontine; children
ID HYPERFRACTIONATED RADIATION-THERAPY; FIBROBLAST-GROWTH-FACTOR;
   BRAIN-STEM GLIOMAS; I-II TRIAL; CELL-PROLIFERATION; CANCER-PATIENTS;
   ALPHA; TUMORS; ANGIOGENESIS; BETA
AB BACKGROUND: Interferon-a is a cytokine that has demonstrated activity in patients with supratentorial gliomas, but its ideal dose and schedule of administration is unknown. Studies suggest that low-dose, continuous exposure is more efficacious than intermittent, high doses. The authors performed a phase 2 study of recombinant interferon a-2b with monomethoxy polyethylene glycol (PEG-Intron (R)) in children with diffuse intrinsic pontine glioma (DIPG), a population with dismal survival despite decades of clinical investigation. The primary objective was to compare 2-year survival with a historic cohort that received radiation therapy alone. METHODS: Patients received weekly subcutaneous PEG-Intron (R) at a dose of 0.3 mu g/kg beginning 2 to 10 weeks after the completion of radiation therapy until they developed disease progression. Patients were evaluated clinically and radiographically at regular intervals. Serum and urine were assayed for biomarkers before each cycle. Quality-of-life (QOL) evaluations were administered at baseline and before every other cycle of therapy to the parents of patients ages 6 to 18 years. RESULTS: Thirty-two patients (median age, 5.3 years; range, 1.8-14.8 years) were enrolled and received a median of 7 cycles of therapy (range, from 1 cycle to =70 cycles). PEG-Intron (R) was well tolerated, and no decrease in QOL scores was noted in the subset of patients tested. The 2-year survival rate was 14%, which was not significantly improved compared with the historic cohort. However, the median time to progression was 7.8 months, which compared favorably with recent trials reporting a time to progression of 5 months in a similar population. CONCLUSIONS: Although low-dose PEG-Intron (R) therapy did not significantly improve 2-year survival in children with DIPG compared with an historic control population, it did delay the time to progression. Cancer 2012;36073613. (C) 2011 American Cancer Society.
C1 [Warren, Katherine; Bent, Robyn; Wolters, Pamela L.; Hanson, Ryan] NCI, Pediat Oncol Branch, Bethesda, MD 20892 USA.
   [Prager, Alisa; Camphausen, Kevin] NCI, Radiat Oncol Branch, Bethesda, MD 20892 USA.
   [Packer, Roger] Childrens Natl Med Ctr, Washington, DC 20010 USA.
   [Shih, Joanna] NCI, Biometr Res Branch, Div Canc Treatment & Diag, Bethesda, MD 20892 USA.
C3 National Institutes of Health (NIH) - USA; NIH National Cancer Institute
   (NCI); National Institutes of Health (NIH) - USA; NIH National Cancer
   Institute (NCI); Children's National Health System; National Institutes
   of Health (NIH) - USA; NIH National Cancer Institute (NCI)
RP Warren, K (通讯作者)，NCI, Pediat Oncol Branch, Bldg 10,Room 1-5750,9000 Rockville Pike, Bethesda, MD 20892 USA.
EM
OI Hanson, Ryan/0000-0002-2564-3710
FU Intramural Research Program of the National Institutes of Health,
   National Cancer Institute, Center for Cancer Research
FX This research was supported by the Intramural Research Program of the
   National Institutes of Health, National Cancer Institute, Center for
   Cancer Research.
CR ALLEN J, 1991, J CLIN ONCOL, V9, P783, DOI 10.1200/JCO.1991.9.5.783
   BARON S, 1994, ANTIVIR RES, V24, P97, DOI 10.1016/0166-3542(94)90058-2
   Brown AP, 2008, CANCER METAST REV, V27, P415, DOI 10.1007/s10555-008-9143-x
   Chan LW, 2004, J CLIN ONCOL, V22, P499, DOI 10.1200/JCO.2004.07.022
   CLEMENS MJ, 1985, BIOCHEM J, V226, P345, DOI 10.1042/bj2260345
   Cohen KJ, 2011, NEURO-ONCOLOGY, V13, P410, DOI 10.1093/neuonc/noq205
   EDWARDS MSB, 1989, J NEUROSURG, V70, P691, DOI 10.3171/jns.1989.70.5.0691
   EZEKOWITZ A, 1991, BRIT J HAEMATOL, V79, P67, DOI 10.1111/j.1365-2141.1991.tb08123.x
   Fischbein NJ, 1996, PEDIATR NEUROSURG, V24, P9, DOI 10.1159/000121010
   Freeman CR, 1998, INT J RADIAT ONCOL, V40, P265, DOI 10.1016/S0360-3016(97)00572-5
   Fujita M, 2010, CLIN CANCER RES, V16, P3409, DOI 10.1158/1078-0432.CCR-10-0644
   Gerber SA, 2008, J IMMUNOL, V181, P1052, DOI 10.4049/jimmunol.181.2.1052
   Haas-Kogan DA, 2011, NEURO-ONCOLOGY, V13, P298, DOI 10.1093/neuonc/noq202
   Hargrove D, 2006, LANCET ONCOL, V7, P241, DOI 10.1016/S1470-2045(06)70615-5
   HERTZOG PJ, 1994, MOL REPROD DEV, V39, P226
   Jakacki RI, 2000, J NEUROSURG, V92, P255, DOI 10.3171/jns.2000.92.2.0255
   JENKIN RDT, 1987, J NEUROSURG, V66, P227, DOI 10.3171/jns.1987.66.2.0227
   Lee HR, 2009, J INTERF CYTOK RES, V29, P621, DOI 10.1089/jir.2009.0067
   MAHALEY MS, 1985, J NEUROSURG, V63, P719, DOI 10.3171/jns.1985.63.5.0719
   Moschos S, 2007, CYTOKINE GROWTH F R, V18, P451, DOI 10.1016/j.cytogfr.2007.06.020
   Moses MA, 1998, CANCER RES, V58, P1395
   Nagai M, 1984, Neurosurg Rev, V7, P55, DOI 10.1007/BF01743290
   NGUYEN M, 1994, J NATL CANCER I, V86, P356, DOI 10.1093/jnci/86.5.356
   Packer RJ, 1996, CANCER-AM CANCER SOC, V77, P2150, DOI 10.1002/(SICI)1097-0142(19960515)77:10<2150::AID-CNCR28>3.0.CO;2-T
   PESTKA S, 1987, ANNU REV BIOCHEM, V56, P727, DOI 10.1146/annurev.bi.56.070187.003455
   Pollack IF, 2011, NEURO-ONCOLOGY, V13, P290, DOI 10.1093/neuonc/noq199
   REAL FX, 1986, J CLIN ONCOL, V4, P544, DOI 10.1200/JCO.1986.4.4.544
   ROMEO G, 1989, TRENDS GENET, V5, P19, DOI 10.1016/0168-9525(89)90007-3
   SINGH RK, 1995, P NATL ACAD SCI USA, V92, P4562, DOI 10.1073/pnas.92.10.4562
   Slaton JW, 1999, CLIN CANCER RES, V5, P2726
   von Marschall Z, 2003, J NATL CANCER I, V95, P437, DOI 10.1093/jnci/95.6.437
   Wolters P, 2004, NEURO-ONCOLOGY, V6, P448
   Zeuzem S, 2003, SEMIN LIVER DIS, V23, P23
NR 33
TC 27
Z9 32
U1 0
U2 4
PU WILEY-BLACKWELL
PI HOBOKEN
PA 111 RIVER ST, HOBOKEN 07030-5774, NJ USA
SN 0008-543X
J9 CANCER-AM CANCER SOC
JI Cancer
PD JUL 15
PY 2012
VL 118
IS 14
BP 3607
EP 3613
DI 10.1002/cncr.26659
PG 7
WC Oncology
WE Science Citation Index Expanded (SCI-EXPANDED)
SC Oncology
GA 967FD
UT WOS:000305890900020
PM 22086404
OA Bronze, Green Accepted
DA 2023-08-27
ER

PT J
AU Cohen, AR
   Stone, SSD
AF Cohen, Alan R.
   Stone, Scellig S. D.
TI Endoscope-assisted microsurgical resection of intrinsic brainstem
   epidermoid: technical note and review of the literature
SO JOURNAL OF NEUROSURGERY-PEDIATRICS
LA English
DT Review
DE brainstem; brain tumor; epidermoid; microsurgery; MRI; neuroendoscopy;
   diagnosis; oncology; surgical technique
ID CYST; EXCISION; TUMOR
AB Intrinsic epidermoid tumors of the brainstem are rare, histologically benign lesions associated with high surgical morbidity and mortality due to their eloquent location. The authors report a child with progressive severe neurological deterioration from a large midline intrinsic brainstem epidermoid at the pontomedullary junction. The mass was removed through a posterior fossa craniotomy and midline endoscope-assisted microsurgical corridor through the floor of the fourth ventricle, using neurophysiological monitoring. Postoperatively, there was dramatic improvement in the patient's symptoms. Early recurrence of the mass necessitated reoperation with more aggressive resection of the cyst capsule, which led to complete radiographic reconstitution of the brainstem. The patient remains well with a durable recovery 7 years after presentation. The authors review the literature on brainstem epidermoids and discuss the differential diagnosis and management strategies for approaching these lesions, advocating for conservative surgery with resection of as much of the tumor capsule as is safely possible.
C1 [Cohen, Alan R.] Johns Hopkins Univ, Sch Med, Dept Neurosurg, Baltimore, MD 21205 USA.
   [Stone, Scellig S. D.] Harvard Med Sch, Boston Childrens Hosp, Dept Neurosurg, Boston, MA 02115 USA.
C3 Johns Hopkins University; Harvard University; Boston Children's
   Hospital; Harvard Medical School
RP Cohen, AR (通讯作者)，Johns Hopkins Univ, Sch Med, Baltimore, MD 21205 USA.
EM
OI Stone, Scellig/0000-0002-4546-6790
CR BAILEY P, 1920, SURG GYNECOL OBSTET, V31, P390
   Balogun JA, 2018, WORLD NEUROSURG, V112, P153, DOI 10.1016/j.wneu.2018.01.128
   Bambakidis NC., 2009, SURG CEREBELLOPONTIN
   BERGER MS, 1985, J NEUROSURG, V62, P214, DOI 10.3171/jns.1985.62.2.0214
   Caldarelli M, 2001, SURG NEUROL, V56, P97, DOI 10.1016/S0090-3019(01)00542-0
   Dandy W., 1945, THE BRAIN
   DUNN LT, 1994, CHILD NERV SYST, V10, P529, DOI 10.1007/BF00335076
   Forbes JA, 2019, J NEUROSURG, V130, P1599, DOI 10.3171/2017.12.JNS172575
   FOURNIER D, 1992, CHILD NERV SYST, V8, P471, DOI 10.1007/BF00274412
   Gopalakrishnan CV, 2012, J CHILD NEUROL, V27, P105, DOI 10.1177/0883073811414709
   IIHARA K, 1989, SURG NEUROL, V32, P377, DOI 10.1016/0090-3019(89)90143-2
   Kachhara R, 2000, ACTA NEUROCHIR, V142, P97, DOI 10.1007/s007010050013
   Kansal R, 2012, TURK NEUROSURG, V22, P452, DOI 10.5137/1019-5149.JTN.3096-10.2
   LEAL O, 1978, J NEUROSURG, V48, P811, DOI 10.3171/jns.1978.48.5.0811
   Malcolm GP, 1996, NEUROSURGERY, V38, P579, DOI 10.1097/00006123-199603000-00032
   Mankotia DS, 2016, J PEDIATR NEUROSCI, V11, P230, DOI 10.4103/1817-1745.193379
   Mask-Bull L, 2013, CLIN NEUROL NEUROSUR, V115, P1888, DOI 10.1016/j.clineuro.2013.05.002
   Mishra SS, 2014, J PEDIATR NEUROSCI, V9, P52, DOI 10.4103/1817-1745.131487
   Morshed RA, 2019, J NEURO-ONCOL, V144, P89, DOI 10.1007/s11060-019-03202-2
   Mortini P, 2016, ACTA NEUROCHIR, V158, P1197, DOI 10.1007/s00701-016-2813-5
   OBANA WG, 1991, J NEUROSURG, V74, P123, DOI 10.3171/jns.1991.74.1.0123
   Patibandla M R, 2016, Asian J Neurosurg, V11, P194, DOI 10.4103/1793-5482.145163
   Recinos PF, 2006, J NEUROSURG, V104, P285, DOI 10.3171/ped.2006.104.4.285
   ROSENBLUTH PR, 1960, J NEUROSURG, V17, P35, DOI 10.3171/jns.1960.17.1.0035
   SCHWARTZ JF, 1978, NEUROLOGY, V28, P124, DOI 10.1212/WNL.28.2.124
   Shimamoto Y, 1999, SKULL BASE SURG, V9, P75, DOI 10.1055/s-2008-1058152
   Shtaya A, 2017, WORLD NEUROSURG, V104, DOI 10.1016/j.wneu.2017.04.164
   Singh SK, 2018, CHILD NERV SYST, V34, P1589, DOI 10.1007/s00381-018-3774-1
   Takahashi M, 2007, NEUROL MED-CHIR, V47, P140, DOI 10.2176/nmc.47.140
   TSURUDA JS, 1990, AM J ROENTGENOL, V155, P1059, DOI 10.2214/ajr.155.5.2120936
   ULRICH J, 1964, J NEUROSURG, V21, P1051, DOI 10.3171/jns.1964.21.12.1051
   WEAVER EN, 1979, J NEUROSURG, V51, P254, DOI 10.3171/jns.1979.51.2.0254
   YASARGIL MG, 1989, NEUROSURGERY, V24, P561
   Yoshizato K, 1996, SURG NEUROL, V45, P537, DOI 10.1016/0090-3019(95)00470-X
   You E, 2019, WORLD NEUROSURG, V131, P275, DOI 10.1016/j.wneu.2019.07.072
   Ziyal IM, 2005, CHILD NERV SYST, V21, P1025, DOI 10.1007/s00381-005-1172-y
NR 36
TC 0
Z9 0
U1 1
U2 4
PU AMER ASSOC NEUROLOGICAL SURGEONS
PI ROLLING MEADOWS
PA 5550 MEADOWBROOK DRIVE, ROLLING MEADOWS, IL 60008 USA
SN 1933-0707
EI 1933-0715
J9 J NEUROSURG-PEDIATR
JI J. Neurosurg.-Pediatr.
PD DEC
PY 2020
VL 26
IS 6
BP 654
EP 660
DI 10.3171/2020.6.PEDS20482
PG 7
WC Clinical Neurology; Pediatrics; Surgery
WE Science Citation Index Expanded (SCI-EXPANDED)
SC Neurosciences & Neurology; Pediatrics; Surgery
GA PB5ZG
UT WOS:000596398300008
PM 33007748
OA Bronze
DA 2023-08-27
ER

PT J
AU Zhao, XL
   Du, C
   Tian, Y
AF Zhao, Xingli
   Du, Chao
   Tian, Yu
TI Results of Repeat Gamma Knife Treatment: Stereotactic Aspiration Surgery
   Can Improve the Effect of Gamma Knife Treatment for Cystic Small
   Brainstem Glioma Case Report
SO NEUROSURGERY QUARTERLY
LA English
DT Article
DE repeat gamma knife; stereotactic aspiration; treatment efficacy; Cystic
   brainstem glioma
ID RADIOSURGERY; TUMORS; MICROSURGERY; MIDBRAIN
AB A 40-year-old Chinese woman presented with a cystic mass (size of 23 x 23 x 27 mm, volume 7.9 cm(3)) with tumor nodule in the pons. The patient received initial gamma knife surgery (GKS) treatment of dose 18.0 Gy, and the mass increased in volume to 12.8 cm(3) after 7 weeks. Stereotactic aspiration surgery of the intratumoral cyst was performed, yielding 10.5 mL of yellowish-white fluid; biopsy confirmed the diagnosis of glioma. A second GKS of dose 10.5 Gy was performed 1 week later. Follow-up magnetic resonance imaging revealed a cyst volume of 1.0 cm(3), with an obvious decrease in size of the tumor nodule at 18 months after the second GKS. The patient is alive and well 36 months after the combined therapy. To the best of our knowledge, cystic brainstem glioma treated by repeat gamma knife treatment combined with stereotactic aspiration surgery in a single patient has not been reported previously in the English-language literature. The findings of this case suggest that cyst drainage by stereotactic aspiration surgery can improve the effect of GKS for small brainstem glioma, even if the diameter of tumor is less than 30 mm.
C1 [Zhao, Xingli; Du, Chao; Tian, Yu] Jilin Univ, China Japan Union Hosp, Dept Neurosurg, Changchun 130033, Peoples R China.
C3 Jilin University
RP Tian, Y (通讯作者)，Jilin Univ, China Japan Union Hosp, Dept Neurosurg, Changchun 130033, Peoples R China.
EM
FU National Science Foundation of China [30672159]; New Century Excellent
   Talents in University of China [NCET-06-0306]
FX Supported by grants from the National Science Foundation of China (No.
   30672159) and New Century Excellent Talents in University of China
   (NCET-06-0306).
CR Davidson L, 2009, J CLIN NEUROSCI, V16, P1139, DOI 10.1016/j.jocn.2009.03.003
   Fuchs I, 2002, ACT NEUR S, V84, P85
   Hadjipanayis CG, 2002, J NEUROSURG, V97, P56, DOI 10.3171/jns.2002.97.1.0056
   Hadjipanayis Costas G, 2003, Neurosurg Focus, V14, pe15
   HIRATO M, 1995, STEREOT FUNCT NEUROS, V64, P32, DOI 10.1159/000098762
   HOOD TW, 1989, NEUROSURGERY, V24, P373, DOI 10.1227/00006123-198903000-00011
   Kano H, 2009, J NEURO-ONCOL, V95, P211, DOI 10.1007/s11060-009-9913-5
   Kim MS, 1999, STEREOT FUNCT NEUROS, V72, P38, DOI 10.1159/000056437
   Matsumoto K, 1998, J NEUROSURG, V88, P141, DOI 10.3171/jns.1998.88.1.0141
   Niranjan A, 2000, CAN J NEUROL SCI, V27, P229, DOI 10.1017/S031716710000086X
   Peraud A, 2007, CHILD NERV SYST, V23, P39, DOI 10.1007/s00381-006-0203-7
   Petti PL, 2008, J NEUROSURG, V109, P34, DOI 10.3171/JNS/2008/109/12/S7
   Sharma MS, 2008, NEUROSURGERY, V63, P728, DOI 10.1227/01.NEU.0000325726.72815.22
   Tsuboi K, 2006, PEDIATR NEUROSURG, V42, P311, DOI 10.1159/000094069
   Vesper J, 2009, EUR J MED RES, V14, P93, DOI 10.1186/2047-783X-14-3-93
   Yen CP, 2007, J NEUROSURG, V106, P8, DOI 10.3171/jns.2007.106.1.8
NR 16
TC 39
Z9 40
U1 0
U2 1
PU LIPPINCOTT WILLIAMS & WILKINS
PI PHILADELPHIA
PA TWO COMMERCE SQ, 2001 MARKET ST, PHILADELPHIA, PA 19103 USA
SN 1050-6438
EI 1534-4916
J9 NEUROSURG QUART
JI Neurosurg. Q.
PD FEB
PY 2012
VL 22
IS 1
BP 22
EP 25
DI 10.1097/WNQ.0b013e3182277d55
PG 4
WC Neurosciences; Surgery
WE Science Citation Index Expanded (SCI-EXPANDED)
SC Neurosciences & Neurology; Surgery
GA 894DZ
UT WOS:000300408500006
DA 2023-08-27
ER

PT J
AU de la Pena, PPG
   Rodriiguez-Romero, R
AF Pinero-Gonzalez de la Pena, Pilar
   Rodriguez-Romero, Rafael
TI Nonglial Tumors of the Brainstem
SO SEMINARS IN ULTRASOUND CT AND MRI
LA English
DT Article
ID CAVERNOUS MALFORMATIONS; MEDULLOBLASTOMA; MANAGEMENT; FEATURES; CNS
AB Nonglial tumors of the brainstem constitute a histologically heterogeneous group of lesions with quite a different behavior and aggressiveness. Therefore, the diverse therapeutic options depend on a correct and prompt diagnosis. We can limit their differential diagnosis by using clinical and demographic data and imaging findings, which in most cases will be a translation of their histologic characteristics. The main clinical, neuroimaging, and pathologic features of these lesions are described according to the last updated classification of the World Health Organization for central nervous system tumors. We provide some useful clues, based on the direct correlation of the imaging appearance with its gross pathologic and histologic appearance, for a comprehensive diagnostic approach.
   Embryonic tumors (medulloblastoma and primitive neuroectodermal tumor), cavernoma, lymphoma, hemangioblastoma, and ganglionic and mixed tumors as long as lesions affecting the central nervous system by external compression (arising from the skull, cerebrospinal fluid spaces, or extraaxial nervous and vascular elements) are included. All cases presented belong to the archive data of our hospital. Semin Ultrasound CT MRI 34:113-122 (C) 2013 Elsevier Inc. All rights reserved.
C1 [Pinero-Gonzalez de la Pena, Pilar; Rodriguez-Romero, Rafael] Virgen del Rocio Hosp, Dept Neuroradiol, Seville, Spain.
C3 Virgen del Rocio University Hospital
RP de la Pena, PPG (通讯作者)，C Gerardo Diego 9,P1,2 D, Seville 41013, Spain.
EM
RI IBIS, NEURORRADIOLOGÍA/C-2563-2016
CR Abe T, 1999, Brain Tumor Pathol, V16, P49, DOI 10.1007/BF02478902
   [Anonymous], 2007, WHO HISTOLOGICAL CLA
   BARKOVICH AJ, 1991, PEDIATR NEUROSURG, V16, P73, DOI 10.1159/000120511
   Bonneville F, 2001, RADIOGRAPHICS, V21, P419, DOI 10.1148/radiographics.21.2.g01mr13419
   BOURGOUIN PM, 1992, AM J ROENTGENOL, V159, P609, DOI 10.2214/ajr.159.3.1503035
   Bradac GB, 1990, CRANIAL MENINGIOMAS, P1
   David KM, 1997, J NEUROSURG, V86, P13, DOI 10.3171/jns.1997.86.1.0013
   DONATI F, 1992, NEUROPEDIATRICS, V23, P32, DOI 10.1055/s-2008-1071309
   Dorai Z, 2006, NEUROONCOLOGY CNS TU, P303
   FRITSCHI JA, 1994, ACTA NEUROCHIR, V130, P35, DOI 10.1007/BF01405501
   GIOMBINI S, 1978, ACTA NEUROCHIR, V40, P61, DOI 10.1007/BF01773116
   HUNTER KMF, 1968, CAN MED ASSOC J, V98, P1
   Kaal ECA, 2005, J NEURO-ONCOL, V75, P15, DOI 10.1007/s11060-004-8094-5
   Koeller KK, 2003, RADIOGRAPHICS, V23, P1613, DOI 10.1148/rg.236035168
   Koerbel A, 2003, ARQ NEURO-PSIQUIAT, V61, P274, DOI 10.1590/S0004-282X2003000200022
   LEE SR, 1989, RADIOLOGY, V171, P463, DOI 10.1148/radiology.171.2.2704812
   Leung RS, 2008, RADIOGRAPHICS, V28, P65, DOI 10.1148/rg.281075052
   Maher EA, 1999, SEMIN ONCOL, V26, P346
   Mathiesen T, 2003, J NEUROSURG, V99, P31, DOI 10.3171/jns.2003.99.1.0031
   MIKHAEL MA, 1985, J COMPUT ASSIST TOMO, V9, P852, DOI 10.1097/00004728-198509000-00002
   Moriarity JL, 1999, NEUROSURGERY, V44, P1166, DOI 10.1097/00006123-199906000-00003
   Ng SH, 2002, CHIN J RADIOL, V27, P21
   OTTEN P, 1989, NEUROCHIRURGIE, V35, P82
   PACKER RJ, 1994, J NEUROSURG, V81, P690, DOI 10.3171/jns.1994.81.5.0690
   PARK TS, 1983, J NEUROSURG, V58, P543, DOI 10.3171/jns.1983.58.4.0543
   Pozzati E, 1998, J NEUROSURG, V89, P498
   Ragheb J, 2000, INT PEDIAT, V15, P15
   Reifenberg G, 2006, NEUROONCOLOGY CNS TU, P3
   RIGAMONTI D, 1987, J NEUROSURG, V67, P518, DOI 10.3171/jns.1987.67.4.0518
   ROBERTS RO, 1991, J NEUROPATH EXP NEUR, V50, P134, DOI 10.1097/00005072-199103000-00005
   Rodallec M, 2004, J NEURORADIOLOGY, V31, P234, DOI 10.1016/S0150-9861(04)97000-9
   ROHRINGER M, 1989, J NEUROSURG, V71, P665, DOI 10.3171/jns.1989.71.5.0665
   Stavrou T, 2001, J PEDIAT HEMATOL ONC, V23, P431, DOI 10.1097/00043426-200110000-00008
   TRUWIT CL, 1990, AM J NEURORADIOL, V11, P665
   Wang CC, 2003, SURG NEUROL, V59, P444, DOI 10.1016/S0090-3019(03)00187-3
   ZIMMERMAN RD, 1985, AM J NEURORADIOL, V6, P149
NR 36
TC 2
Z9 2
U1 1
U2 1
PU W B SAUNDERS CO-ELSEVIER INC
PI PHILADELPHIA
PA 1600 JOHN F KENNEDY BOULEVARD, STE 1800, PHILADELPHIA, PA 19103-2899 USA
SN 0887-2171
EI 1558-5034
J9 SEMIN ULTRASOUND CT
JI Semin. Ultrasound CT MRI
PD APR
PY 2013
VL 34
IS 2
BP 113
EP 122
DI 10.1053/j.sult.2013.01.002
PN 2
PG 10
WC Radiology, Nuclear Medicine & Medical Imaging
WE Science Citation Index Expanded (SCI-EXPANDED)
SC Radiology, Nuclear Medicine & Medical Imaging
GA 117AA
UT WOS:000316923300003
PM 23522776
DA 2023-08-27
ER

PT J
AU Lin, GL
   Monje, M
AF Lin, Grant L.
   Monje, Michelle
TI A Protocol for Rapid Post-mortem Cell Culture of Diffuse Intrinsic
   Pontine Glioma (DIPG)
SO JOVE-JOURNAL OF VISUALIZED EXPERIMENTS
LA English
DT Article
DE Cancer Research; Issue 121; DIPG; glioma; patient-derived cell culture;
   rapid autopsy; brain; tissue dissociation; cancer
ID ACVR1; MUTATIONS; BIOPSY; MODEL
AB Diffuse Intrinsic Pontine Glioma (DIPG) is a childhood brainstem tumor that carries a universally fatal prognosis. Because surgical resection is not a viable treatment strategy and biopsy is not routinely performed, the availability of patient samples for research is limited. Consequently, efforts to study this disease have been challenged by a paucity of faithful disease models. To address this need, we describe here a protocol for the rapid processing of post-mortem autopsy tissue samples in order to generate durable patient-derived cell culture models that can be used in in vitro assays or in vivo orthotopic xenograft experiments. These models can be used to screen for potential drug targets and to study fundamental pathobiological processes within DIPG. This protocol can further be extended to analyze and isolate tumor and microenvironmental cells using Fluorescence-activated Cell Sorting (FACS), which enables subsequent analysis of gene expression, protein expression, or epigenetic modifications of DNA at the bulk cell or single cell level. Finally, this protocol can also be adapted to generate patient-derived cultures for other central nervous system tumors.
C1 [Lin, Grant L.] Stanford Univ, Sch Med, Inst Stem Cell Biol & Regenerat Med, Dept Neurol,Grad Program Neurosci, Stanford, CA 94305 USA.
   [Monje, Michelle] Stanford Univ, Sch Med, Inst Stem Cell Biol & Regenerat Med, Depts Neurol, Stanford, CA 94305 USA.
   [Monje, Michelle] Stanford Univ, Sch Med, Inst Stem Cell Biol & Regenerat Med, Dept Neurosurg, Stanford, CA 94305 USA.
   [Monje, Michelle] Stanford Univ, Sch Med, Inst Stem Cell Biol & Regenerat Med, Dept Pathol & Pediat, Stanford, CA 94305 USA.
C3 Stanford University; Stanford University; Stanford University; Stanford
   University
RP Monje, M (通讯作者)，Stanford Univ, Sch Med, Inst Stem Cell Biol & Regenerat Med, Depts Neurol, Stanford, CA 94305 USA.; Monje, M (通讯作者)，Stanford Univ, Sch Med, Inst Stem Cell Biol & Regenerat Med, Dept Neurosurg, Stanford, CA 94305 USA.; Monje, M (通讯作者)，Stanford Univ, Sch Med, Inst Stem Cell Biol & Regenerat Med, Dept Pathol & Pediat, Stanford, CA 94305 USA.
EM
OI Lin, Grant/0000-0001-6888-2140
FU McKenna Claire Foundation; National Institute of Neurological Disorders
   and Stroke [NINDS K08NS070926, R01NS092597]; Department of Defense
   [NF140075]; California Institute for Regenerative Medicine [CIRM
   RB4-06093, RN3-06510]; Alex's Lemonade Stand Foundation; Cure Starts Now
   Foundation; DIPG Collaborative; Lyla Nsouli Foundation; Unravel
   Pediatric Cancer; Childhood Brain Tumor Foundation; Matthew Larson
   Foundation; V Foundation; Godfrey Family Fund in Memory of Fiona
   Penelope; Wayland Villars DIPG Foundation; Dylan Jewett Memorial Fund;
   Connor Johnson Memorial Fund; Zoey Ganesh Memorial Fund; Dylan Frick
   Memorial Fund; Abigail Jensen Memorial Fund; Jennifer Kranz Memorial
   Fund; N8 Foundation; Virginia and D.K. Ludwig Fund for Cancer Research;
   Child Health Research Institute at Stanford Anne T. and Robert M. Bass
   Endowed Faculty Scholarship in Pediatric Cancer and Blood Diseases
FX The authors gratefully acknowledge support from the McKenna Claire
   Foundation, National Institute of Neurological Disorders and Stroke
   (NINDS K08NS070926 and R01NS092597), Department of Defense (NF140075),
   California Institute for Regenerative Medicine (CIRM RB4-06093 and
   RN3-06510), Alex's Lemonade Stand Foundation, The Cure Starts Now
   Foundation and DIPG Collaborative, Lyla Nsouli Foundation, Unravel
   Pediatric Cancer, Childhood Brain Tumor Foundation, Matthew Larson
   Foundation, V Foundation, Godfrey Family Fund in Memory of Fiona
   Penelope, the Wayland Villars DIPG Foundation, the Dylan Jewett, Connor
   Johnson, Zoey Ganesh, Dylan Frick, Abigail Jensen, and Jennifer Kranz
   Memorial Funds, N8 Foundation, Virginia and D.K. Ludwig Fund for Cancer
   Research, Child Health Research Institute at Stanford Anne T. and Robert
   M. Bass Endowed Faculty Scholarship in Pediatric Cancer and Blood
   Diseases.
CR Aoki Y, 2012, J NEURO-ONCOL, V108, P29, DOI 10.1007/s11060-011-0796-x
   Becher OJ, 2010, CANCER RES, V70, P2548, DOI 10.1158/0008-5472.CAN-09-2503
   Buczkowicz P, 2014, NAT GENET, V46, P451, DOI 10.1038/ng.2936
   Cage TA, 2013, CHILD NERV SYST, V29, P1313, DOI 10.1007/s00381-013-2101-0
   Fisher PG, 2000, CANCER, V89, P1569, DOI 10.1002/1097-0142(20001001)89:7<1569::AID-CNCR22>3.0.CO;2-0
   Fontebasso AM, 2014, NAT GENET, V46, P462, DOI 10.1038/ng.2950
   Funato K, 2014, SCIENCE, V346, P1529, DOI 10.1126/science.1253799
   Grasso CS, 2015, NAT MED, V21, P555, DOI 10.1038/nm.3855
   Johung T. B., 2016, CURR NEUROPHARMACOL
   Jones C, 2014, NAT REV CANC
   Kieran MW, 2015, PEDIATR BLOOD CANCER, V62, P3, DOI 10.1002/pbc.25266
   Monje M, 2011, P NATL ACAD SCI USA, V108, P4453, DOI 10.1073/pnas.1101657108
   Olah M, 2012, GLIA, V60, P96, DOI 10.1002/glia.21251
   Puget S, 2015, CHILD NERV SYST, V31, P1773, DOI 10.1007/s00381-015-2832-1
   Shu Q, 2008, STEM CELLS, V26, P1414, DOI 10.1634/stemcells.2007-1009
   Sturm D, 2014, NAT REV CANCER, V14, P92, DOI 10.1038/nrc3655
   Taylor KR, 2014, NAT GENET, V46, P457, DOI 10.1038/ng.2925
   Wu G, 2014, NAT GENET, V46, P444, DOI 10.1038/ng.2938
NR 18
TC 30
Z9 30
U1 0
U2 5
PU JOURNAL OF VISUALIZED EXPERIMENTS
PI CAMBRIDGE
PA 1 ALEWIFE CENTER, STE 200, CAMBRIDGE, MA 02140 USA
SN 1940-087X
J9 JOVE-J VIS EXP
JI J. Vis. Exp.
PD MAR
PY 2017
IS 121
AR e55360
DI 10.3791/55360
PG 8
WC Multidisciplinary Sciences
WE Science Citation Index Expanded (SCI-EXPANDED)
SC Science & Technology - Other Topics
GA EQ1RW
UT WOS:000397848300065
PM 28362421
OA Green Published, Bronze, Green Submitted
DA 2023-08-27
ER

PT J
AU Ruivo, J
   Antunes, JL
AF Ruivo, Joana
   Antunes, Joao Lobo
TI Maffucci syndrome associated with a pituitary adenoma and a probable
   brainstem tumor
SO JOURNAL OF NEUROSURGERY
LA English
DT Article
DE enchondromatosis; glioma; Maffucci syndrome; pituitary adenoma; tumor
ID SPINDLE-CELL HEMANGIOENDOTHELIOMA; INTRACRANIAL CHONDROSARCOMA;
   OLLIERS-DISEASE; SKULL BASE; DYSCHONDROPLASIA; PATIENT;
   ENCHONDROMATOSIS; HEMANGIOMAS; CARCINOMA; VARIANT
AB Malignancies are a common feature of Maffucci syndrome, with chondrosarcomas being the most common tumor type. The authors present the first case of Maffucci syndrome associated with a pituitary adenoma and a probable brainstem glioma and review the literature concerning intracranial tumors related to this disease.
   They report the case of a 35-year-old woman with Marfucci syndrome (diagnosed when she was 22 years old) who presented with complaints of decreased visual acuity and visual field defect. Neuroimaging revealed a pituitary macroadenoma and a suspected brainstem tumor. The macroadenoma was partially removed. There were no postoperative complications and the patient experienced rapid improvement in visual acuity. On follow-up examination 2 years later, the lesion in the pons showed the same dimensions. No sarcomatous changes of enchondromas or hemangiomas occurred.
   To the authors' knowledge, including the present case, only 7 cases of Maffucci syndrome associated with glioma and 7 cases associated with pituitary adenoma have been reported in the literature. This report emphasizes that patients with this disease are at a higher risk for primary intracranial tumors and reinforces the concept of the multiplicity of tumors that may arise in this syndrome. It also underscores the importance of early diagnosis, regular clinical surveillance, and follow-up studies of these patients. (DOI: 10.3171/2008.8.JNS08150)
C1 [Ruivo, Joana] Hosp Santa Maria, Dept Neuroradiol, P-1600190 Lisbon, Portugal.
   [Antunes, Joao Lobo] Hosp Santa Maria, Dept Neurosurg, P-1600190 Lisbon, Portugal.
C3 Universidade de Lisboa; Hospital Santa Maria; Universidade de Lisboa;
   Hospital Santa Maria
RP Ruivo, J (通讯作者)，Hosp Santa Maria, Dept Neuroradiol, Piso 2,Ave Prof Egas Moniz, P-1600190 Lisbon, Portugal.
EM
CR Abdelmalek M, 2008, AM J CLIN DERMATOL, V9, P61, DOI 10.2165/00128071-200809010-00008
   ARMSTRONG EA, 1981, J NEUROSURG, V55, P479, DOI 10.3171/jns.1981.55.3.0479
   ASHENHURST EM, 1960, ARCH NEUROL-CHICAGO, V2, P552, DOI 10.1001/archneur.1960.03840110066008
   BARADNAY G, 1960, ORVOSI HETILAP, V4, P1753
   BECKER W, 1979, ARCH ORTHOP TRAUM SU, V93, P141, DOI 10.1007/BF00389685
   BERTUCCI V, 1995, PEDIATR DERMATOL, V12, P55, DOI 10.1111/j.1525-1470.1995.tb00127.x
   Boinet E, 1904, ARCH GEN MED, V194, P2689
   BUSHE KA, 1990, NEUROSURGERY, V27, P625, DOI 10.1227/00006123-199010000-00020
   CAMERON JM, 1957, BRIT J SURG, V44, P596
   Carleton A, 1942, Q J MED, V11, P203
   CHAKRABORTTY S, 1991, SURG NEUROL, V36, P216, DOI 10.1016/0090-3019(91)90116-Q
   Cheng F C, 1981, J R Coll Surg Edinb, V26, P181
   CHRISTMAN JE, 1990, GYNECOL ONCOL, V37, P290, DOI 10.1016/0090-8258(90)90350-T
   COOK PL, 1977, BRIT J RADIOL, V50, P833, DOI 10.1259/0007-1285-50-599-833
   CREMER H, 1981, J CANCER RES CLIN, V101, P231, DOI 10.1007/BF00413318
   DAVIDSON TI, 1985, EUR J SURG ONCOL, V11, P381
   Dini LI, 2007, ARQ NEURO-PSIQUIAT, V65, P816, DOI 10.1590/S0004-282X2007000500016
   Faik A, 2006, CLIN RHEUMATOL, V25, P88, DOI 10.1007/s10067-005-1101-1
   FANBURG JC, 1995, AM J SURG PATHOL, V19, P1029, DOI 10.1097/00000478-199509000-00006
   Fernandez-Aguilar S, 2004, BREAST, V13, P247, DOI 10.1016/j.breast.2003.06.001
   Flach HZ, 2001, RADIOGRAPHICS, V21, P1311
   GOTO H, 1987, SHINKEI GEKA, V15, P971
   Hachi H, 2002, Gynecol Obstet Fertil, V30, P692, DOI 10.1016/S1297-9589(02)00416-2
   HALL BD, 1972, ARCH DERMATOL, V105, P608, DOI 10.1001/archderm.1972.01620070078030
   Hisaoka M, 1997, SKELETAL RADIOL, V26, P191, DOI 10.1007/s002560050219
   HOWIE FMC, 1988, SKELETAL RADIOL, V17, P368, DOI 10.1007/BF00367186
   JIRARATTANAPHOCHAI K, 1990, Journal of the Medical Association of Thailand, V73, P288
   KAPLAN RP, 1993, J AM ACAD DERMATOL, V29, P894, DOI 10.1016/0190-9622(93)70265-U
   KERR HD, 1991, SOUTHERN MED J, V84, P1039, DOI 10.1097/00007611-199108000-00024
   KESSLER HB, 1983, SKELETAL RADIOL, V10, P95, DOI 10.1007/BF00360792
   Kitamura Eiji, 2006, Brain and Nerve (Tokyo), V58, P514
   Kurian S, 2004, SKELETAL RADIOL, V33, P609, DOI 10.1007/s00256-004-0802-x
   KUZMA JF, 1948, ARCH PATHOL, V46, P74
   LAWSON JP, 1990, SKELETAL RADIOL, V19, P158
   LEWIS RJ, 1973, J BONE JOINT SURG AM, VA 55, P1465, DOI 10.2106/00004623-197355070-00014
   LOEWINGER RJ, 1977, BRIT J DERMATOL, V96, P317, DOI 10.1111/j.1365-2133.1977.tb06145.x
   LOWELL SH, 1979, ARCH OTOLARYNGOL, V105, P427
   Maffucci A, 1881, MOV MED CHIR NAP, V3, P399
   MARYMONT JV, 1987, SOUTHERN MED J, V80, P1429, DOI 10.1097/00007611-198711000-00022
   MATSUMOTO N, 1986, Neurological Surgery, V14, P403
   McDermott AL, 2001, J LARYNGOL OTOL, V115, P845
   MIKI K, 1987, ACTA NEUROCHIR, V87, P79, DOI 10.1007/BF02076022
   Miyake Mototaka, 2005, Radiat Med, V23, P508
   MOTOMOCHI M, 1979, SHINKEI GEKA, V7, P1011
   Nakamura K, 1999, ARCH ORTHOP TRAUM SU, V119, P470, DOI 10.1007/s004020050024
   NAKAYAMA Y, 1994, NEUROSURGERY, V34, P907, DOI 10.1227/00006123-199405000-00019
   Noel G, 2004, ACTA ONCOL, V43, P705, DOI 10.1080/02841860410002770
   Palacios E, 2001, Ear Nose Throat J, V80, P75
   PELLEGRINI AE, 1995, J CUTAN PATHOL, V22, P173, DOI 10.1111/j.1600-0560.1995.tb01403.x
   Ramina R, 1997, NEUROSURGERY, V41, P269, DOI 10.1097/00006123-199707000-00046
   Ramirez-Bollas Julio, 2005, Cir Cir, V73, P217
   RAWLINGS CE, 1987, NEUROSURGERY, V21, P400, DOI 10.1227/00006123-198709000-00023
   RECTOR JT, 1993, AM J PEDIAT HEMATOL, V15, P427
   Rozeman LB, 2004, HUM MUTAT, V24, P466, DOI 10.1002/humu.20095
   SAKURANE HF, 1967, ARCH DERMATOL, V95, P28, DOI 10.1001/archderm.95.1.28
   SATO K, 1989, TO SHINKEI, V41, P631
   SCHNALL AM, 1976, AM J MED, V61, P952, DOI 10.1016/0002-9343(76)90421-6
   SCHWARTZ HS, 1987, J BONE JOINT SURG AM, V69A, P269, DOI 10.2106/00004623-198769020-00016
   Stenger RD, 1998, PEDIATR HEMAT ONCOL, V15, P179, DOI 10.3109/08880019809167233
   STRANG C, 1950, J BONE JOINT SURG BR, V32, P376, DOI 10.1302/0301-620X.32B3.376
   SUN TC, 1985, J BONE JOINT SURG AM, V67A, P1214, DOI 10.2106/00004623-198567080-00011
   SURINGA DWR, 1970, ARCH DERMATOL, V101, P472, DOI 10.1001/archderm.101.4.472
   Tachibana E, 2000, SURG NEUROL, V54, P165, DOI 10.1016/S0090-3019(00)00252-4
   TANAKA Y, 1992, AM J CLIN PATHOL, V97, P523, DOI 10.1093/ajcp/97.4.523
   Tibbs RE, 1997, SKULL BASE SURG, V7, P49, DOI 10.1055/s-2008-1058625
   Yazidi A, 1998, ANN DERMATOL VENER, V125, P50
   Yuan JQ, 2004, CHINESE MED J-PEKING, V117, P1592
NR 67
TC 8
Z9 8
U1 0
U2 5
PU AMER ASSOC NEUROLOGICAL SURGEONS
PI ROLLING MEADOWS
PA 5550 MEADOWBROOK DRIVE, ROLLING MEADOWS, IL 60008 USA
SN 0022-3085
EI 1933-0693
J9 J NEUROSURG
JI J. Neurosurg.
PD FEB
PY 2009
VL 110
IS 2
BP 363
EP 368
DI 10.3171/2008.8.JNS08150
PG 6
WC Clinical Neurology; Surgery
WE Science Citation Index Expanded (SCI-EXPANDED)
SC Neurosciences & Neurology; Surgery
GA 399GI
UT WOS:000262788000024
PM 18976063
DA 2023-08-27
ER

PT J
AU Lin, CS
   Selch, MT
   Lee, SP
   Wu, JK
   Xiao, FR
   Hong, DS
   Chen, CH
   Hussain, A
   Lee, PP
   De Salles, AA
AF Lin, Chun-Shu
   Selch, Michael T.
   Lee, Steve P.
   Wu, Jeffrey K.
   Xiao, Furen
   Hong, David S.
   Chen, Chien-Hua
   Hussain, Aamir
   Lee, Percy P.
   De Salles, Antonio A.
TI Accelerator-Based Stereotactic Radiosurgery for Brainstem Metastases
SO NEUROSURGERY
LA English
DT Article
DE Brainstem metastases; Radiosurgery; Stereotactic radiosurgery
ID GAMMA-KNIFE RADIOSURGERY; PROGNOSTIC-FACTORS; SURVIVAL; RADIOTHERAPY;
   TUMORS
AB BACKGROUND: Stereotactic radiosurgery represents a noninvasive alternative treatment for intracranial metastases.
   OBJECTIVE: To investigate the treatment outcome of linear accelerator-based stereotactic radiosurgery (linac-SRS) for brainstem metastases.
   METHODS: We retrospectively reviewed our database of patients who were diagnosed with brainstem metastases and underwent linac-SRS between 1997 and 2008 at the University of California, Los Angeles.
   RESULTS: A total of 45 patients with 48 brainstem metastases were treated. The median target volume was 0.40 mL (range, 0.02-5.70 mL), and median prescription dose was 14 Gy (range, 10-17 Gy) at 90% isodose curve. The median survival time was 11.6 months. Longer survival time was associated with higher Karnofsky performance status. The local control rate was 92% at 6 months and 88% at 1 year. Univariate analysis demonstrated a significant relationship between local control and tumor volume (<= 0.4 mL vs > 0.4 mL, P = .023) and SRS mode (conventional circular arc vs dynamic conformal arc, P = .044). There was a trend toward improved local control and prescription dose > 14 Gy (P = .059). Two patients had brainstem complications following treatment, and the complication rate was 4.7% at 2 years. Serious morbidity occurred with 17 Gy.
   CONCLUSION: Linac-SRS using a median dose of 14 Gy provided excellent local control in patients with brainstem metastases less than 0.4 mL with relatively low serious morbidity. The results of the study support the use of linac-SRS for patients with brainstem metastases. We advocate 14 to 16 Gy, given the high local control rate and low complication rate with this dose.
C1 [Lin, Chun-Shu] Natl Def Med Ctr, Dept Radiat Oncol, Triserv Gen Hosp, Taipei 114, Taiwan.
   [Lin, Chun-Shu; Selch, Michael T.; Lee, Steve P.; Wu, Jeffrey K.; Hong, David S.; Hussain, Aamir; Lee, Percy P.] Univ Calif Los Angeles, David Geffen Sch Med, Dept Radiat Oncol, Los Angeles, CA 90095 USA.
   [Xiao, Furen; De Salles, Antonio A.] Univ Calif Los Angeles, David Geffen Sch Med, Dept Neurosurg, Los Angeles, CA 90095 USA.
   [Chen, Chien-Hua] Taichung Vet Gen Hosp, Dept Neurosurg, Taichung, Taiwan.
C3 National Defense Medical Center; Tri-Service General Hospital;
   University of California System; University of California Los Angeles;
   University of California Los Angeles Medical Center; David Geffen School
   of Medicine at UCLA; University of California System; University of
   California Los Angeles; University of California Los Angeles Medical
   Center; David Geffen School of Medicine at UCLA; Taichung Veterans
   General Hospital
RP Lin, CS (通讯作者)，Natl Def Med Ctr, Dept Radiat Oncol, Triserv Gen Hosp, 325,Sec 2,Chenggong Rd, Taipei 114, Taiwan.
EM
RI Lee, Steve Pai-hsun/AAU-8313-2020
OI Lee, Steve Pai-hsun/0000-0001-7168-1542; Xiao, Furen/0000-0001-9683-3979
CR BORGELT B, 1980, INT J RADIAT ONCOL, V6, P1, DOI 10.1016/0360-3016(80)90195-9
   COIA LR, 1988, INT J RADIAT ONCOL, V14, P1261, DOI 10.1016/0360-3016(88)90405-1
   DEANGELIS LM, 1989, NEUROSURGERY, V24, P798, DOI 10.1227/00006123-198906000-00002
   Fuentes S, 2006, NEUROSURGERY, V58, P37, DOI 10.1227/01.NEU.0000190655.95669.5C
   Gaspar L, 1997, INT J RADIAT ONCOL, V37, P745, DOI 10.1016/S0360-3016(96)00619-0
   Huang CF, 1999, J NEUROSURG, V91, P563, DOI 10.3171/jns.1999.91.4.0563
   Hussain A, 2007, INT J RADIAT ONCOL, V67, P521, DOI 10.1016/j.ijrobp.2006.08.081
   Kased N, 2008, J NEURO-ONCOL, V86, P195, DOI 10.1007/s11060-007-9458-4
   Kim DG, 2000, J NEUROSURG, V93, P23, DOI 10.3171/jns.2000.93.supplement_3.0023
   Lorenzoni JG, 2009, SURG NEUROL, V71, P188, DOI 10.1016/j.surneu.2008.01.029
   Marsh JC, 2010, J ONCOL, V2010, DOI 10.1155/2010/198208
   Rutten I, 2007, RADIOTHER ONCOL, V82, P83, DOI 10.1016/j.radonc.2006.11.019
   Shaw E, 2000, INT J RADIAT ONCOL, V47, P291, DOI 10.1016/S0360-3016(99)00507-6
   Sheehan JP, 2002, J NEUROSURG, V97, P1276, DOI 10.3171/jns.2002.97.6.1276
   Shiau CY, 1997, INT J RADIAT ONCOL, V37, P375, DOI 10.1016/S0360-3016(96)00497-X
   Shuto T, 2003, ACTA NEUROCHIR, V145, P755, DOI 10.1007/s00701-003-0034-1
   Solberg TD, 2001, INT J RADIAT ONCOL, V49, P1481, DOI 10.1016/S0360-3016(00)01537-6
   Steinvorth S, 2002, INT J RADIAT ONCOL, V54, P1430, DOI 10.1016/S0360-3016(02)03800-2
   Welzel G, 2008, INT J RADIAT ONCOL, V72, P1311, DOI 10.1016/j.ijrobp.2008.03.009
   Yen CP, 2006, J NEUROSURG, V105, P213, DOI 10.3171/jns.2006.105.2.213
NR 20
TC 24
Z9 24
U1 0
U2 4
PU OXFORD UNIV PRESS INC
PI CARY
PA JOURNALS DEPT, 2001 EVANS RD, CARY, NC 27513 USA
SN 0148-396X
EI 1524-4040
J9 NEUROSURGERY
JI Neurosurgery
PD APR
PY 2012
VL 70
IS 4
BP 953
EP 958
DI 10.1227/NEU.0b013e31823c40fe
PG 6
WC Clinical Neurology; Surgery
WE Science Citation Index Expanded (SCI-EXPANDED)
SC Neurosciences & Neurology; Surgery
GA 914FF
UT WOS:000301934000060
PM 21997541
DA 2023-08-27
ER

PT J
AU Ahn, MS
   Jackler, RK
AF Ahn, MS
   Jackler, RK
TI Exophytic brain tumors mimicking primary lesions of the cerebellopontine
   angle
SO LARYNGOSCOPE
LA English
DT Article; Proceedings Paper
CT Meeting of the Western Section of the
   American-Laryngological-Rhinological-and-Otological-Society
CY JAN 13-14, 1996
CL LOS ANGELES, CA
SP Amer Laryngol Rhinol Otol Soc Inc, W Sect
AB The vast majority of cerebellopontine angle (CPA) tumors are extraaxial masses arising from either the eighth nerve (acoustic neuroma) or meninges (meningioma). Rarely, a tumor that arises from the brain parenchyma may protrude laterally to present with a clinical and radiographic picture simulating that of the much more common extraaxial lesions. Three individuals with CPA lesions that ultimately proved at operation to be exophytic brain tumors (pontine medulloblastoma, cerebellar astrocytoma, and fourth ventricular ependymoma) are described. The clinical manifestations of these lesions, although not entirely typical, fell well within the possible range of presentation of benign tumors primary to the CPA. In two of these cases the tumor actually penetrated into the internal auditory canal. As the optimal management strategy for treating parenchymal tumors differs substantially from that for extraaxial lesions, it is essential that the surgeon have a preoperative awareness of the lesion's nature before embarking on a surgical endeavor. Critical to arriving at the correct diagnosis is the close examination of preoperative imaging studies. The clinician should be alerted to the possibility that a CPA tumor is of intraaxial origin when the preoperative magnetic resonance imaging scan shows 1) blurring of the margin between the tumor and brainstem or cerebellum; 2) a degree of peritumoral hyperintensity on Ta-weighted scans disproportionate to the size of the extraaxial mass; and 3) dilation of the lateral recess of the fourth ventricle.
C1 UNIV SAN FRANCISCO,DEPT OTOLARYNGOL HEAD & NECK SURG,SAN FRANCISCO,CA 94117.
   UNIV SAN FRANCISCO,DEPT NEUROL SURG,SAN FRANCISCO,CA 94117.
C3 University of San Francisco; University of San Francisco
OI jackler, robert/0000-0003-2525-8377
CR ALBRIGHT AL, 1996, NEUROLOGICAL SURGERY
   AMPIL F, 1992, J NEURO-ONCOL, V12, P125
   BARTELS LJ, 1994, NEUROTOLOGY, P835
   BERGER MS, 1983, NEUROSURGERY, V12, P298, DOI 10.1227/00006123-198303000-00008
   BERGER MS, 1996, NEUROLOGICAL SURGERY
   BRACKMANN DE, 1980, OTOLARYNG HEAD NECK, V88, P555, DOI 10.1177/019459988008800508
   COBBS CS, 1996, NEUROLOGICAL SURGERY, P2532
   DUVOISIN B, 1991, EUR J RADIOL, V13, P96, DOI 10.1016/0720-048X(91)90088-D
   EPSTEIN FJ, 1993, J NEUROSURG, V78, P408, DOI 10.3171/jns.1993.78.3.0408
   HENNEBERG R, 1902, ARCH F PSYCHIAT, V36, P251
   HITSELBE.WE, 1968, ARCH OTOLARYNGOL, V88, P712
   HOFFMAN HJ, 1980, NEUROSURGERY, V7, P408
   HOUSE JL, 1985, AM J OTOL S, V6, P147
   Kawase T, 1993, Noshuyo Byori, V10, P53
   KERNOHAN JW, 1948, J NEUROPATH EXP NEUR, V7, P349, DOI 10.1097/00005072-194810000-00001
   LALWANI AK, 1992, OTOLARYNG CLIN N AM, V25, P707
   MOFFAT DA, 1993, J LARYNGOL OTOL, V107, P1087, DOI 10.1017/S0022215100125393
   POLLACK IF, 1994, NEW ENGL J MED, V331, P1500, DOI 10.1056/NEJM199412013312207
   SELESNICK SH, 1993, LARYNGOSCOPE, V103, P431, DOI 10.1002/lary.5541030412
   SMIRNIOTOPOULOS JG, 1993, RADIOGRAPHICS, V13, P1131, DOI 10.1148/radiographics.13.5.8210595
   STROINK AR, 1986, J NEUROSURG, V65, P745, DOI 10.3171/jns.1986.65.6.0745
   Tomita T, 1996, NEUROLOGICAL SURG, P2570
NR 22
TC 22
Z9 22
U1 0
U2 0
PU LIPPINCOTT-RAVEN PUBL
PI PHILADELPHIA
PA 227 EAST WASHINGTON SQ, PHILADELPHIA, PA 19106
SN 0023-852X
J9 LARYNGOSCOPE
JI Laryngoscope
PD APR
PY 1997
VL 107
IS 4
BP 466
EP 471
DI 10.1097/00005537-199704000-00008
PG 6
WC Medicine, Research & Experimental; Otorhinolaryngology
WE Conference Proceedings Citation Index - Science (CPCI-S); Science Citation Index Expanded (SCI-EXPANDED)
SC Research & Experimental Medicine; Otorhinolaryngology
GA WU277
UT WOS:A1997WU27700007
PM 9111375
DA 2023-08-27
ER

PT J
AU Wind, JJ
   Bakhtian, KD
   Sweet, JA
   Mehta, GU
   Thawani, JP
   Asthagiri, AR
   Oldfield, EH
   Lonser, RR
AF Wind, Joshua J.
   Bakhtian, Kamran D.
   Sweet, Jennifer A.
   Mehta, Gautam U.
   Thawani, Jayesh P.
   Asthagiri, Ashok R.
   Oldfield, Edward H.
   Lonser, Russell R.
TI Long-term outcome after resection of brainstem hemangioblastomas in von
   Hippel-Lindau disease
SO JOURNAL OF NEUROSURGERY
LA English
DT Article
DE brainstem; hemangioblastoma; outcome; surgical treatment; von
   Hippel-Lindau disease
ID CENTRAL-NERVOUS-SYSTEM; STEREOTACTIC RADIOSURGERY; SURGICAL-MANAGEMENT;
   MEDULLA-OBLONGATA; NATURAL-HISTORY; SPINAL-CORD
AB Object. Brainstem hemangioblastomas are frequently encountered in patients with von Hippel-Lindau (VHL) disease. These tumors can cause significant morbidity, and their optimal management has not been defined. To better define the outcome and management of these tumors, the authors analyzed the long-term results in patients who underwent resection of brainstem hemangioblastomas.
   Methods. Consecutive patients with VHL disease who underwent resection of brainstem hemangioblastomas with a follow-up of 12 months or more were included in this study. Serial functional assessments, radiographic examinations, and operative records were analyzed.
   Results. Forty-four patients (17 male and 27 female) underwent 51 operations for resection of 71 brainstem hemangioblastomas. The most common presenting symptoms were headache, swallowing difficulties, singultus, gait difficulties, and sensory abnormalities. The mean follow-up was 5.9 +/- 5.0 years (range 1.0-20.8 years). Immediately after 34 operations (66.7%), the patients remained at their preoperative functional status; they improved after 8 operations (15.7%) and worsened after 9 operations (17.6%) as measured by the McCormick scale. Eight (88.9%) of the 9 patients who were worse immediately after resection returned to their preoperative status within 6 months. Two patients experienced functional decline during long-term follow-up (beginning at 2.5 and 5 years postoperatively) caused by extensive VHL disease associated CNS disease.
   Conclusions. Generally, resection of symptomatic brainstem hemangioblastomas is a safe and effective management strategy in patients with VHL disease. Most patients maintain their preoperative functional status, although long-term decline in functional status may occur due to VHL disease associated progression. (DOI: 10.3171/2010.9.JNS10839)
C1 [Wind, Joshua J.; Bakhtian, Kamran D.; Sweet, Jennifer A.; Mehta, Gautam U.; Thawani, Jayesh P.; Asthagiri, Ashok R.; Oldfield, Edward H.; Lonser, Russell R.] Natl Inst Neurol Disorders & Stroke, Surg Neurol Branch, NIH, Bethesda, MD 20892 USA.
   [Wind, Joshua J.; Sweet, Jennifer A.] George Washington Univ, Med Ctr, Dept Neurol Surg, Washington, DC 20037 USA.
   [Oldfield, Edward H.] Univ Virginia Hlth Syst, Dept Neurol Surg, Charlottesville, VA USA.
C3 National Institutes of Health (NIH) - USA; NIH National Institute of
   Neurological Disorders & Stroke (NINDS); George Washington University;
   University of Virginia
RP Lonser, RR (通讯作者)，Natl Inst Neurol Disorders & Stroke, Surg Neurol Branch, NIH, Bldg 10,Room 3D20, Bethesda, MD 20892 USA.
EM
RI Mehta, Gautam/AAP-6600-2020; Wind, Joshua J/D-5480-2013
OI Mehta, Gautam/0000-0002-8009-6430; Wind, Joshua J/0000-0003-3443-8476
FU National Institute of Neurological Disorders and Stroke at the National
   Institutes of Health
FX This research was supported by the Intramural Research Program of the
   National Institute of Neurological Disorders and Stroke at the National
   Institutes of Health.
CR Ammerman JM, 2006, J NEUROSURG, V105, P248, DOI 10.3171/jns.2006.105.2.248
   Asthagiri AR, 2010, NEURO-ONCOLOGY, V12, P80, DOI 10.1093/neuonc/nop018
   Baumgartner JE, 1992, NEUROSURGICAL PROCED, P188
   FILLINGKATZ MR, 1991, NEUROLOGY, V41, P41, DOI 10.1212/WNL.41.1.41
   Fukushima T, 1998, NEUROL MED-CHIR, V38, P489, DOI 10.2176/nmc.38.489
   Glasker S, 2006, CANCER BIOL THER, V5, P549
   Jawahar A, 2000, ACTA NEUROCHIR, V142, P641, DOI 10.1007/s007010070107
   Kano H, 2008, NEUROSURGERY, V63, P443, DOI 10.1227/01.NEU.0000313120.81565.D7
   Karnofsky D.A., 1949, EVALUATION CHEMOTHER, P196
   LAMIELL JM, 1989, MEDICINE, V68, P1
   LATIF F, 1993, SCIENCE, V260, P1317, DOI 10.1126/science.8493574
   LEWIS EA, 1973, BRIT J SURG, V60, P279, DOI 10.1002/bjs.1800600407
   Lonser RR, 2005, ANN NEUROL, V58, P392, DOI 10.1002/ana.20584
   Lonser RR, 2003, LANCET, V361, P2059, DOI 10.1016/S0140-6736(03)13643-4
   LUNDIN P, 1992, J COMPUT ASSIST TOMO, V16, P519, DOI 10.1097/00004728-199207000-00004
   MCCORMICK PC, 1990, J NEUROSURG, V72, P523, DOI 10.3171/jns.1990.72.4.0523
   Moss JM, 2009, NEUROSURGERY, V65, P79, DOI 10.1227/01.NEU.0000348015.51685.D2
   NEUMANN HPH, 1992, J NEUROL NEUROSUR PS, V55, P898, DOI 10.1136/jnnp.55.10.898
   NEUMANN HPH, 1989, J NEUROSURG, V70, P24, DOI 10.3171/jns.1989.70.1.0024
   Park DM, 2007, PLOS MED, V4, P333, DOI 10.1371/journal.pmed.0040060
   Parker F, 2009, J NEURO-ONCOL, V93, P133, DOI 10.1007/s11060-009-9861-0
   Patrice SJ, 1996, INT J RADIAT ONCOL, V35, P493
   Rachinger J, 2009, NEUROSURG REV, V32, P37, DOI 10.1007/s10143-008-0166-0
   Song DK, 2008, J NEUROSURG-PEDIATR, V2, P397, DOI 10.3171/PED.2008.2.12.397
   Tago M, 2005, J NEUROSURG, V102, P171, DOI 10.3171/jns.2005.102.s_supplement.0171
   Wanebo JE, 2003, J NEUROSURG, V98, P82, DOI 10.3171/jns.2003.98.1.0082
   Wang CC, 2001, SURG NEUROL, V56, P218, DOI 10.1016/S0090-3019(01)00590-0
   Wang EM, 2005, J NEUROSURG, V102, P225, DOI 10.3171/jns.2005.102.s_supplement.0225
   Weil RJ, 2003, J NEUROSURG, V98, P95, DOI 10.3171/jns.2003.98.1.0095
   Zhou LF, 2005, SURG NEUROL, V63, P307, DOI 10.1016/j.surneu.2004.07.038
NR 30
TC 35
Z9 37
U1 0
U2 7
PU AMER ASSOC NEUROLOGICAL SURGEONS
PI ROLLING MEADOWS
PA 5550 MEADOWBROOK DRIVE, ROLLING MEADOWS, IL 60008 USA
SN 0022-3085
EI 1933-0693
J9 J NEUROSURG
JI J. Neurosurg.
PD MAY
PY 2011
VL 114
IS 5
BP 1312
EP 1318
DI 10.3171/2010.9.JNS10839
PG 7
WC Clinical Neurology; Surgery
WE Science Citation Index Expanded (SCI-EXPANDED)
SC Neurosciences & Neurology; Surgery
GA 755FW
UT WOS:000289913900018
PM 20932100
OA Green Accepted
DA 2023-08-27
ER

PT J
AU Yanagawa, Y
   Miyazawa, T
   Ishihara, S
   Takiguchi, H
   Shima, K
   Terahata, S
   Tamai, S
   Chigasaki, H
AF Yanagawa, Y
   Miyazawa, T
   Ishihara, S
   Takiguchi, H
   Shima, K
   Terahata, S
   Tamai, S
   Chigasaki, H
TI Pontine glioma with osteoblastic skeletal metastases in a child
SO SURGICAL NEUROLOGY
LA English
DT Article
DE pontine glioma; osteoblastic metastases; extracranial metastases; GFAP;
   child
ID BONY METASTASES; GLIOBLASTOMA; ASTROCYTOMAS
AB BACKGROUND The development of systemic metastases from primary intracranial gliomas is rare. We report here a rare case of pontine glioma with osteoblastic skeletal metastases.
   CASE This 12-year-old boy presented with a 4-month history of hoarseness, dysphagia, and a progressively ataxic gait. Cranial computed tomography (CT) and magnetic resonance imaging (MRI) revealed a brain stem tumor that was diagnosed as a low grade glioma by stereotactic biopsy. Twelve months later following chemotherapy and radiotherapy, neurologic examination and neuroradiologic studies disclosed a recurrence of the pontine glioma. Skeletal roentgenograms revealed widespread osteoblastic metastases in the skull, vertebral bodies, pelvis, and long bones. A specimen from the iliac bone demonstrated cells that were immunoreactive glial fibrillary acidic protein (GFAP).
   DISCUSSION The mechanism of how glioma cells determine their biologic behavior at bony metastatic sites is not known. Infratentorial gliomas, which occur frequently in young patients and demonstrate active bony metabolism, may stimulate osteoblastic cells, and induce osteoblastic changes. (C) 1996 by Elsevier Science Inc.
C1 NATL DEF MED COLL,DEPT NEUROSURG & LAB MED,TOKOROZAWA,SAITAMA 359,JAPAN.
C3 National Defense Medical College - Japan
CR CHOI BH, 1981, ACTA NEUROPATHOL, V54, P269, DOI 10.1007/BF00696999
   GAMIS AS, 1990, CANCER, V66, P180, DOI 10.1002/1097-0142(19900701)66:1<180::AID-CNCR2820660132>3.0.CO;2-M
   HORNSBY VPL, 1985, NEURORADIOLOGY, V27, P426, DOI 10.1007/BF00327607
   KINGSTON JE, 1986, CHILD NERV SYST, V2, P219, DOI 10.1007/BF00706817
   KOSUDA S, 1992, J MED IMAGINGS, V12, P338
   LONGEE DC, 1991, MED PEDIATR ONCOL, V19, P318, DOI 10.1002/mpo.2950190419
   LORUSSO PM, 1988, J NEURO-ONCOL, V6, P53, DOI 10.1007/BF00163541
   MARTIN O, 1979, CANC TREATMENT REP, V63, P1417
   NAKAMURA K, 1986, JAPANESE J CANC CLIN, V32, P281
   PASQUIER B, 1980, CANCER, V45, P112, DOI 10.1002/1097-0142(19800101)45:1<112::AID-CNCR2820450121>3.0.CO;2-9
   SCHATZKI SC, 1977, AM J ROENTGENOL, V128, P321, DOI 10.2214/ajr.128.2.321
   SILVERMAN FN, 1985, CAFFEYS PEDIATRIC XR, P882
   SMITH DR, 1969, J NEUROSURG, V31, P50, DOI 10.3171/jns.1969.31.1.0050
   WEISS L, 1955, CANCER, V8, P161, DOI 10.1002/1097-0142(1955)8:1<161::AID-CNCR2820080123>3.0.CO;2-K
   YUNG WKA, 1983, ANN NEUROL, V14, P581, DOI 10.1002/ana.410140514
NR 15
TC 9
Z9 9
U1 0
U2 0
PU ELSEVIER SCIENCE INC
PI NEW YORK
PA 655 AVENUE OF THE AMERICAS, NEW YORK, NY 10010
SN 0090-3019
J9 SURG NEUROL
JI Surg. Neurol.
PD NOV
PY 1996
VL 46
IS 5
BP 481
EP 484
DI 10.1016/S0090-3019(96)00223-6
PG 4
WC Clinical Neurology; Surgery
WE Science Citation Index Expanded (SCI-EXPANDED)
SC Neurosciences & Neurology; Surgery
GA VN703
UT WOS:A1996VN70300032
PM 8874551
DA 2023-08-27
ER

PT J
AU Vitanza, NA
   Wilson, AL
   Huang, WJ
   Seidel, K
   Brown, C
   Gustafson, JA
   Yokoyama, JK
   Johnson, AJ
   Baxter, BA
   Koning, RW
   Reid, AN
   Meechan, M
   Biery, MC
   Myers, C
   Rawlings-Rhea, SD
   Albert, CM
   Browd, SR
   Hauptman, JS
   Lee, AMY
   Ojemann, JG
   Berens, ME
   Dun, MD
   Foster, JB
   Crotty, EE
   Leary, SES
   Cole, BL
   Perez, FA
   Wright, JN
   Orentas, RJ
   Chour, T
   Newell, EW
   Whiteaker, JR
   Zhao, L
   Paulovich, AG
   Pinto, N
   Gust, J
   Gardner, RA
   Jensen, MC
   Park, JR
AF Vitanza, Nicholas A.
   Wilson, Ashley L.
   Huang, Wenjun
   Seidel, Kristy
   Brown, Christopher
   Gustafson, Joshua A.
   Yokoyama, Jason K.
   Johnson, Adam J.
   Baxter, Blake A.
   Koning, Ryan W.
   Reid, Aquene N.
   Meechan, Michael
   Biery, Matthew C.
   Myers, Carrie
   Rawlings-Rhea, Stephanie D.
   Albert, Catherine M.
   Browd, Samuel R.
   Hauptman, Jason S.
   Lee, Amy
   Ojemann, Jeffrey G.
   Berens, Michael E.
   Dun, Matthew D.
   Foster, Jessica B.
   Crotty, Erin E.
   Leary, Sarah E. S.
   Cole, Bonnie L.
   Perez, Francisco A.
   Wright, Jason N.
   Orentas, Rimas J.
   Chour, Tony
   Newell, Evan W.
   Whiteaker, Jeffrey R.
   Zhao, Lei
   Paulovich, Amanda G.
   Pinto, Navin
   Gust, Juliane
   Gardner, Rebecca A.
   Jensen, Michael C.
   Park, Julie R.
TI Intraventricular B7-H3 CAR T Cells for Diffuse Intrinsic Pontine Glioma:
   Preliminary First-in-Human Bioactivity and Safety
SO CANCER DISCOVERY
LA English
DT Article
ID TANDEM MASS-SPECTROMETRY; CENTRAL-NERVOUS-SYSTEM; THERAPEUTIC TARGET;
   QUANTIFICATION; TUMORS; THYROGLOBULIN; BIOMARKERS; SELECTION; ANTIBODY;
   HEALTH
AB Diffuse intrinsic pontine glioma (DIPG) remains a fatal brainstem tumor demanding innovative therapies. As B7-H3 (CD276) is expressed on central nervous system (CNS) tumors, we designed B7-H3-specifi c chimeric antigen receptor (CAR) T cells, confi rmed their preclinical effi cacy, and opened BrainChild-03 (NCT04185038), a first-in-human phase I trial adminis-tering repeated locoregional B7-H3 CAR T cells to children with recurrent/refractory CNS tumors and DIPG. Here, we report the results of the first three evaluable patients with DIPG (including two who enrolled after progression), who received 40 infusions with no dose-limiting toxicities. One patient had sustained clinical and radiographic improvement through 12 months on study. Patients exhibited cor-relative evidence of local immune activation and persistent cerebrospinal fluid (CSF) B7-H3 CAR T cells. Targeted mass spectrometry of CSF biospecimens revealed modulation of B7-H3 and critical immune analytes (CD14, CD163, CSF-1, CXCL13, and VCAM-1). Our data suggest the feasibility of repeated intracranial B7-H3 CAR T-cell dosing and that intracranial delivery may induce local immune activation.SIGNIFICANCE: This is the first report of repeatedly dosed intracranial B7-H3 CAR T cells for patients with DIPG and includes preliminary tolerability, the detection of CAR T cells in the CSF, CSF cytokine elevations supporting locoregional immune activation, and the feasibility of serial mass spectrometry from both serum and CSF.
C1 [Vitanza, Nicholas A.; Meechan, Michael; Biery, Matthew C.; Myers, Carrie; Albert, Catherine M.; Crotty, Erin E.; Leary, Sarah E. S.; Orentas, Rimas J.; Pinto, Navin; Gardner, Rebecca A.; Park, Julie R.] Seattle Childrens Res Inst, Ben Towne Ctr Childhood Canc Res, Seattle, WA USA.
   [Vitanza, Nicholas A.; Albert, Catherine M.; Crotty, Erin E.; Leary, Sarah E. S.; Orentas, Rimas J.; Pinto, Navin; Gardner, Rebecca A.; Park, Julie R.] Univ Washington, Seattle Childrens Hosp, Dept Pediat, Seattle, WA USA.
   [Wilson, Ashley L.; Huang, Wenjun; Seidel, Kristy; Brown, Christopher; Gustafson, Joshua A.; Yokoyama, Jason K.; Johnson, Adam J.; Baxter, Blake A.; Koning, Ryan W.; Reid, Aquene N.; Rawlings-Rhea, Stephanie D.; Gardner, Rebecca A.; Jensen, Michael C.; Park, Julie R.] Seattle Childrens Therapeut, Seattle, WA USA.
   [Brown, Christopher] Seattle Childrens Res Inst, Therapeut Cell Prod Core, Seattle, WA 98101 USA.
   [Browd, Samuel R.; Hauptman, Jason S.; Lee, Amy; Ojemann, Jeffrey G.] Seattle Childrens Hosp, Div Neurosurg, Seattle, WA USA.
   [Browd, Samuel R.; Hauptman, Jason S.; Lee, Amy; Ojemann, Jeffrey G.] Univ Washington, Dept Neurol Surg, Seattle, WA USA.
   [Berens, Michael E.] Translat Genom Res Inst TGen, Canc & Cell Biol Div, Phoenix, AZ USA.
   [Dun, Matthew D.] Coll Hlth Med & Wellbeing, Sch Biomed Sci & Pharm, Callaghan, NSW, Australia.
   [Dun, Matthew D.] Hunter Med Res Inst, Precis Med Res Program, New Lambton Hts, Australia.
   [Foster, Jessica B.] Childrens Hosp Philadelphia, Div Oncol, Philadelphia, PA USA.
   [Foster, Jessica B.] Univ Penn, Perelman Sch Med, Dept Pediat, Philadelphia, PA USA.
   [Cole, Bonnie L.] Seattle Childrens Hosp, Dept Labs, Seattle, WA USA.
   [Cole, Bonnie L.; Chour, Tony; Newell, Evan W.] Univ Washington, Sch Med, Dept Lab Med & Pathol, Seattle, WA USA.
   [Perez, Francisco A.; Wright, Jason N.] Seattle Childrens Hosp, Dept Radiol, Seattle, WA USA.
   [Chour, Tony; Newell, Evan W.] Fred Hutchinson Canc Res Ctr, Vaccine & Infect Dis Div, Seattle, WA USA.
   [Whiteaker, Jeffrey R.; Zhao, Lei; Paulovich, Amanda G.] Fred Hutchinson Canc Ctr, Clin Res Div, Seattle, WA USA.
   [Gust, Juliane] Seattle Childrens Res Inst, Ctr Integrat Brain Res, Seattle, WA 98101 USA.
   [Gust, Juliane] Univ Washington, Dept Neurol, Div Pediat Neurol, Seattle, WA USA.
C3 Seattle Children's Hospital; Seattle Children's Hospital; University of
   Washington; University of Washington Seattle; Seattle Children's
   Hospital; Seattle Children's Hospital; University of Washington;
   University of Washington Seattle; Translational Genomics Research
   Institute; Hunter Medical Research Institute; University of Newcastle;
   University of Pennsylvania; Pennsylvania Medicine; Childrens Hospital of
   Philadelphia; University of Pennsylvania; Pennsylvania Medicine; Seattle
   Children's Hospital; University of Washington; University of Washington
   Seattle; Seattle Children's Hospital; Fred Hutchinson Cancer Center;
   Fred Hutchinson Cancer Center; Seattle Children's Hospital; University
   of Washington; University of Washington Seattle
RP Vitanza, NA (通讯作者)，Seattle Childrens Res Inst, MS JMB-8,1900 9th Ave, Seattle, WA 98101 USA.
EM
RI Newell, Evan W/F-9711-2012; Park, Julie Ruggieri/IWD-6773-2023
OI Newell, Evan W/0000-0002-2889-243X; Brown,
   Christopher/0000-0003-0834-4003; Wilson, Ashley/0000-0002-4440-001X;
   Johnson, Adam/0000-0002-9967-8959; Chour, Tony/0000-0003-3273-5549;
   Baxter, Blake/0000-0002-6491-1802; Gustafson,
   Joshua/0000-0002-0735-7235; Vitanza, Nicholas/0000-0002-3966-4985;
   Yokoyama, Jason/0000-0003-2520-8134
FU Amazon; Aven Foundation; The Avery Huffman DIPG Foundation; Julianna
   Sayler Foundation; The Kellen Joyce Heart of a Warrior Research Fund;
   Liv Like a Unicorn; Hope for Harlee Foundation; Kick Childhood Cancer;
   Live Gray's Way; The Andrew McDonough B+ Foundation; Love for Lucy;
   Unravel Pediatric Cancer; the McKenna Claire Foundation; Pediatric Brain
   Tumor Research Fund Guild of Seattle Children's; Cookies for Kid's
   Cancer Young Investigator Grant; DIPG All-In; the Run of Hope Seattle;
   Matthew Larson Research Grant; Sam Day Foundation; Alex's Lemonade Stand
   Foundation for Childhood Cancer; Starbucks; the National Center for
   Advancing Translational Sciences of the NIH; the Seattle Sounders; St.
   Baldrick's Stand Up To Cancer Dream Team Translational Cancer Research;
   Team Beans Infant Brain Tumor Fund; Stand Up To Cancer is a division of
   the Entertainment Industry Foundation; Team Cozzi Foundation; American
   Association for Cancer Research; Tommy Strong Foundation; NCI, NIH,
   under the NCI Beau Biden National Cancer Moonshot [HHSN26100025,
   HHSN261201500003I]; Top Pot Doughnuts; Applied Proteogenomics
   Organizational Learning and Outcomes; NCI Academic Industrial
   Partnership; NCI Research Specialist Program [R50CA211499]; NCI
   [R01CA235575]; NCI Clinical Proteomics Tumor Analysis Consortium
   [U01TR002487]; St. Baldrick's Stand Up To Cancer Dream Team
   Translational Cancer Research Grants [SU2C-AACR-DT-27-17]
FX We thank the children and families who bravely shoulder the burden of
   their disease and place their trust in Seattle Children's for their
   care. We are indebted to A. Thomsen for her expert immuno-therapy
   clinical coordination. We thank our clinical research team, including H.
   Ullom, E. Accornero, S. Bagchi, K. Cilluffo, D. Chen, C. Krein, Z.
   Maino, M. MacQuivey, L. McCann, G. Mun, K. Sharf, E. Stowe, C. Verda, Q.
   Wang, and V. Weiss. We thank our neuro-oncology team, including R.
   Geyer, J. Olson, N. Millard, A. Sato, C. Hoeppner, S. Holtzclaw, A.
   Wein, C. Anderson, O. Cook, A. Laurine, W. Iwata, V. Klein, Z. Reinke,
   C. Henson, and E. Grace. We thank J. Stevens, the Seattle Children's
   Hospital's Department of Anatomic Pathology, and the TTS Brain Tumor
   Committee. We thank A. Kong and the Investigational Drug Service team.
   We thank E. Curinga and the Seattle Children's Therapeutics team. We
   thank the Therapeutic Cell Production Core for their tireless efforts to
   manufacture cellular products and the Correlative Studies Lab for
   assistance in correlative sample processing and analysis. We are
   grateful for generous support from Amazon, The Andrew McDonough B+
   Foundation, the Aven Foundation, The Avery Huffman DIPG Foundation,
   Kristie and Joe Berg, Erin Cordry and Eric Hanson, Freckles from Heaven,
   the Julianna Sayler Foundation, The Kellen Joyce Heart of a Warrior
   Research Fund, Liv Like a Unicorn, Hope for Harlee Foundation, Kick
   Childhood Cancer, Live Gray's Way, Love for Lucy, the McKenna Claire
   Foundation, the Pediatric Brain Tumor Research Fund Guild of Seattle
   Children's, Kate and Tom Peters, the Run of Hope Seattle, Sam Day
   Foundation, Starbucks, the Seattle Sounders, Team Beans Infant Brain
   Tumor Fund, Team Cozzi Foundation, Tommy Strong Foundation, Top Pot
   Doughnuts, Unravel Pediatric Cancer, and Jessica and Jared Wray. Funding
   has been provided by Cookies for Kid's Cancer Young Investigator Grant
   (N.A. Vitanza) , DIPG All-In (N.A. Vitanza), Matthew LarsonResearch
   Grant (N.A. Vitanza) , Alex's Lemonade Stand Foundation for Childhood
   Cancer (R.A. Gardner), the National Center for Advancing Translational
   Sciences of the NIH (U01TR002487; A.L. Wilson, W. Huang, R.A. Gardner,
   J.R. Park), and St. Baldrick's Stand Up To Cancer Dream Team
   Translational Cancer Research Grants (SU2C-AACR-DT-27-17; N.A. Vitanza,
   R.J. Orentas, R.A. Gardner, M.C. Jensen, and J.R. Park) . Stand Up To
   Cancer is a division of the Entertainment Industry Foundation. The
   indicated SU2C research grant is administered by the American
   Association for Cancer Research, the scientific partner of SU2C. This
   research has been funded in part with federal funds from the NCI, NIH,
   under the NCI Beau Biden National Cancer Moonshot (A.G. Paulovich, Task
   Order No. HHSN26100025, Applied Proteogenomics Organizational Learning
   and Outcomes, under Contract No. HHSN261201500003I) , the NCI Clinical
   Proteomics Tumor Analysis Consortium (A.G. Paulovich, U01CA214114) , the
   NCI Academic Industrial Partnership (A.G. Paulovich, R01CA235575) , and
   the NCI Research Specialist Program (J.R. Whiteaker, R50CA211499) .
CR Abbatiello SE, 2015, MOL CELL PROTEOMICS, V14, P2357, DOI 10.1074/mcp.M114.047050
   Amezquita RA, 2020, NAT METHODS, V17, P137, DOI 10.1038/s41592-019-0654-x
   Biery Matt C, 2020, J Exp Neurol, V1, P158, DOI 10.33696//Neurol.1.025
   Brown CE, 2016, NEW ENGL J MED, V375, P2561, DOI 10.1056/NEJMoa1610497
   Brun V, 2009, J PROTEOMICS, V72, P740, DOI 10.1016/j.jprot.2009.03.007
   Ceppi F, 2022, CANCER IMMUNOL RES, V10, P856, DOI 10.1158/2326-6066.CIR-21-0501
   Collins CJ, 2020, FRONT IMMUNOL, V11, DOI 10.3389/fimmu.2020.00464
   Collins CJ, 2018, FRONT IMMUNOL, V9, DOI 10.3389/fimmu.2018.02756
   Cooney T, 2017, NEURO-ONCOLOGY, V19, P1279, DOI 10.1093/neuonc/nox107
   Do M, 2020, CLIN CHEM, V66, P1339, DOI 10.1093/clinchem/hvaa178
   Donovan LK, 2020, NAT MED, V26, P720, DOI 10.1038/s41591-020-0827-2
   Dorshkind K, 2009, CURR OPIN IMMUNOL, V21, P404, DOI 10.1016/j.coi.2009.07.001
   Du HW, 2019, CANCER CELL, V35, P221, DOI 10.1016/j.ccell.2019.01.002
   ErcikanAbali EA, 1996, CANCER RES, V56, P4142
   Erskine CL, 2012, JOVE-J VIS EXP, DOI 10.3791/3683
   Perez-Larraya JG, 2022, NEW ENGL J MED, V386, P2471, DOI 10.1056/NEJMoa2202028
   Gardner RA, 2017, BLOOD, V129, P3322, DOI 10.1182/blood-2017-02-769208
   Gregorio A, 2008, HISTOPATHOLOGY, V53, P73, DOI 10.1111/j.1365-2559.2008.03070.x
   Gruver AL, 2007, J PATHOL, V211, P144, DOI 10.1002/path.2104
   Guedan S, 2019, MOL THER-METH CLIN D, V12, P145, DOI 10.1016/j.omtm.2018.12.009
   Guedan S, 2018, JCI INSIGHT, V3, DOI 10.1172/jci.insight.96976
   Haydar D, 2021, NEURO-ONCOLOGY, V23, P999, DOI 10.1093/neuonc/noaa278
   Hembrough T, 2013, J MOL DIAGN, V15, P454, DOI 10.1016/j.jmoldx.2013.03.002
   Hudecek M, 2013, CLIN CANCER RES, V19, P3153, DOI 10.1158/1078-0432.CCR-13-0330
   Jain MD, 2021, BLOOD, V137, P2621, DOI 10.1182/blood.2020007445
   Jin LC, 2019, NAT COMMUN, V10, DOI 10.1038/s41467-019-11869-4
   Jonnalagadda M, 2013, GENE THER, V20, P853, DOI 10.1038/gt.2012.97
   Jonnalagadda M, 2013, PLOS ONE, V8, DOI 10.1371/journal.pone.0065519
   Kennedy JJ, 2021, CLIN CHEM, V67, P1008, DOI 10.1093/clinchem/hvab047
   Kennedy JJ, 2016, J PROTEOME RES, V15, P2717, DOI 10.1021/acs.jproteome.6b00245
   Krishna S, 2020, SCIENCE, V370, P1328, DOI 10.1126/science.abb9847
   Kunkele A, 2015, CANCER IMMUNOL RES, V3, P368, DOI 10.1158/2326-6066.CIR-14-0200
   Kuhn E, 2012, MOL CELL PROTEOMICS, V11, DOI 10.1074/mcp.M111.013854
   Kushnir MM, 2013, CLIN CHEM, V59, P982, DOI 10.1373/clinchem.2012.195594
   Lieberman NAP, 2019, NEURO-ONCOLOGY, V21, P83, DOI 10.1093/neuonc/noy145
   Loo D, 2012, CLIN CANCER RES, V18, P3834, DOI 10.1158/1078-0432.CCR-12-0715
   Louis DN, 2016, ACTA NEUROPATHOL, V131, P803, DOI 10.1007/s00401-016-1545-1
   MacLean B, 2010, BIOINFORMATICS, V26, P966, DOI 10.1093/bioinformatics/btq054
   Majzner RG, 2022, NATURE, V603, P934, DOI 10.1038/s41586-022-04489-4
   Majzner RG, 2019, CLIN CANCER RES, V25, P2560, DOI 10.1158/1078-0432.CCR-18-0432
   Mrdjen D, 2018, IMMUNITY, V48, P380, DOI 10.1016/j.immuni.2018.01.011
   Nehama D, 2019, EBIOMEDICINE, V47, P33, DOI 10.1016/j.ebiom.2019.08.030
   Netzel BC, 2015, J CLIN ENDOCR METAB, V100, pE1074, DOI 10.1210/jc.2015-1967
   Neubert H, 2020, CLIN CHEM, V66, P282, DOI 10.1093/clinchem/hvz022
   Pino LK, 2020, MASS SPECTROM REV, V39, P229, DOI 10.1002/mas.21540
   Ravanpay Ali C, 2019, Oncotarget, V10, P7080, DOI 10.18632/oncotarget.27389
   Talbot LJ, 2021, FRONT IMMUNOL, V12, DOI 10.3389/fimmu.2021.691741
   Tang X, 2021, SIGNAL TRANSDUCT TAR, V6, DOI 10.1038/s41392-021-00505-7
   Tang X, 2020, CLIN TRANSL IMMUNOL, V9, DOI 10.1002/cti2.1137
   Tang X, 2019, MOL THER-ONCOLYTICS, V14, P279, DOI 10.1016/j.omto.2019.07.002
   Theruvath J, 2020, NAT MED, V26, P712, DOI 10.1038/s41591-020-0821-8
   Van Oekelen O, 2021, NAT MED, V27, P2099, DOI 10.1038/s41591-021-01564-7
   Vitanza NA, 2021, NAT MED, V27, P1544, DOI 10.1038/s41591-021-01404-8
   Vitanza NA, 2021, NEURO-ONCOLOGY, V23, P376, DOI 10.1093/neuonc/noaa249
   Vitanza NA, 2019, CURR TREAT OPTION NE, V21, DOI 10.1007/s11940-019-0577-y
   Wang XL, 2011, BLOOD, V118, P1255, DOI 10.1182/blood-2011-02-337360
   Watanabe N, 2016, ONCOIMMUNOLOGY, V5, DOI 10.1080/2162402X.2016.1253656
   Whiteaker JR, 2007, ANAL BIOCHEM, V362, P44, DOI 10.1016/j.ab.2006.12.023
   Whiteaker JR, 2021, FRONT IMMUNOL, V12, DOI 10.3389/fimmu.2021.765898
   Whiteaker JR, 2018, DNA REPAIR, V65, P47, DOI 10.1016/j.dnarep.2018.03.003
   Whiteaker JR, 2018, RADIAT RES, V189, P505, DOI 10.1667/RR14963.1
   Whiteaker JR, 2014, NAT METHODS, V11, P703, DOI 10.1038/nmeth.3002
   Whiteaker JR, 2012, MOL CELL PROTEOMICS, V11, DOI 10.1074/mcp.M111.015347
   Whiteaker JR, 2011, NAT BIOTECHNOL, V29, P625, DOI 10.1038/nbt.1900
   Wickham H., 2016, GGPLOT2, DOI DOI 10.1007/978-3-319-24277-4
   Zhang ZL, 2020, MOL THER-ONCOLYTICS, V17, P180, DOI 10.1016/j.omto.2020.03.019
   Zhao L, 2011, JOVE-J VIS EXP, DOI 10.3791/2812
   Zhou ZP, 2013, J NEURO-ONCOL, V111, P257, DOI 10.1007/s11060-012-1021-2
NR 68
TC 20
Z9 20
U1 6
U2 7
PU AMER ASSOC CANCER RESEARCH
PI PHILADELPHIA
PA 615 CHESTNUT ST, 17TH FLOOR, PHILADELPHIA, PA 19106-4404 USA
SN 2159-8274
EI 2159-8290
J9 CANCER DISCOV
JI Cancer Discov.
PD JAN
PY 2023
VL 13
IS 1
BP 114
EP 131
DI 10.1158/2159-8290.CD-22-0750
PG 18
WC Oncology
WE Science Citation Index Expanded (SCI-EXPANDED)
SC Oncology
GA 8E3OY
UT WOS:000918888200001
PM 36259971
OA hybrid, Green Published
HC Y
HP N
DA 2023-08-27
ER

PT J
AU Grundy, RG
   Wilne, SH
   Robinson, KJ
   Ironside, JW
   Cox, T
   Chong, WK
   Michalski, A
   Campbell, RHA
   Bailey, CC
   Thorp, N
   Pizer, B
   Punt, J
   Walker, DA
   Ellison, DW
   Machin, D
AF Grundy, R. G.
   Wilne, S. H.
   Robinson, K. J.
   Ironside, J. W.
   Cox, T.
   Chong, W. K.
   Michalski, A.
   Campbell, R. H. A.
   Bailey, C. C.
   Thorp, N.
   Pizer, B.
   Punt, J.
   Walker, D. A.
   Ellison, D. W.
   Machin, D.
CA Children's Canc Leukaemia Grp
   Brain Tumour Comm
TI Primary postoperative chemotherapy without radiotherapy for treatment of
   brain tumours other than ependymoma in children under 3 years: Results
   of the first UKCCSG/SIOP CNS 9204 trial
SO EUROPEAN JOURNAL OF CANCER
LA English
DT Article
DE Brain tumours; Infants; Chemotherapy; Radiotherapy; Medulloblastoma;
   Astrocytoma high-grade glioma; Diffuse intrinsic pontine glioma; Choroid
   plexus carcinoma; Central nervous system primitive neuroectodermal
   tumour; Atypical teratoid/rhabdoid tumours
ID CHOROID-PLEXUS CARCINOMA; CENTRAL-NERVOUS-SYSTEM; YOUNG-CHILDREN; CANCER
   GROUP; MULTIAGENT CHEMOTHERAPY; STEM GLIOMAS; MEDULLOBLASTOMA;
   CHILDHOOD; INFANTS; PROGNOSIS
AB Background: Radiotherapy is an effective adjuvant treatment for brain tumours arising in very young children, but it has the potential to damage the child's developing nervous system at a crucial time - with a resultant reduction in IQ leading to cognitive impairment, associated endocrinopathy and risk of second malignancy. we aimed to assess the role of a primary chemotherapy strategy in avoiding or delaying radiotherapy in children younger than 3 years with malignant brain tumours other than ependymoma, the results of which have already been published.
   Methods: Ninety-seven children were enrolled between March 1993 and July 2003 and, following diagnostic review, comprised: medulloblastoma (n = 31), astrocytoma (26), choroid plexus carcinoma [CPC] (15), CNS PNET (11), atypical teratoid/rhabdoid tumours [AT/RT] (6) and ineligible (6). Following maximal surgical resection, chemotherapy was delivered every 14 d for 1 year or until disease progression. Radiotherapy was withheld in the absence of progression.
   Findings: Over all diagnostic groups the cumulative progression rate was 80.9% at 5 years while the corresponding need-for-radiotherapy rate for progression was 54.6%, but both rates varied by tumour type. There was no clear relationship between chemotherapy dose intensity and outcome. Patients with medulloblastoma presented as a high-risk group, 83.9% having residual disease and/or metastases at diagnosis. For these patients, outcome was related to histology. The 5-year OS for desmoplastic/nodular medulloblastoma was 52.9% (95% confidence interval (CI): 27.6-73.0) and for classic medulloblastoma. 33.3% (CI: 4.6-67.6); the 5-year EFS were 35.3% (CI: 14.5-57.0) and 33.3% (CI: 4.6-67.6), respectively.
   All children with large cell or anaplastic variants of medulloblastoma died within 2 years of diagnosis. The 5-year EFS for non-brainstem high-grade gliomas [HGGs] was 13.0% (CI: 2.2-33.4) and the OS was 30.9% (CI: 11.5-52.8). For CPC the 5-year OS was 26.67% (CI: 8.3-49.6) without RT. This treatment strategy was less effective for AT/RT with 3-year OS of 16.7% (CI: 0.8-51.7) and CNS PNET with 1-year OS of 9.1% (CI: 0.5-33.3).
   Interpretation: The outcome for very young children with brain tumours is dictated by degree of surgical resection and histological tumour type and underlying biology as an indicator of treatment sensitivity. Overall, the median age at radiotherapy was 3 years and radiotherapy was avoided in 45% of patients. Desmoplastic/nodular sub-type of medulloblastoma has a better prognosis than classic histology, despite traditional adverse clinical features of metastatic disease and incomplete surgical resection. A subgroup with HGG and CPC are long-term survivors without RT. This study highlights the differing therapeutic challenges presented by the malignant brain tumours of early childhood, the importance of surgical approaches and the need to explore individualised brain sparing approaches to the range of malignant brain tumours that present in early childhood. (C) 2009 Elsevier Ltd. All rights reserved.
C1 [Grundy, R. G.] Univ Nottingham, Sch Med, Childrens Brain Tumour Res Ctr, Queens Med Ctr, Nottingham NG7 2UH, England.
   [Robinson, K. J.; Campbell, R. H. A.; Bailey, C. C.; Machin, D.] Ctr Data, Childrens Canc & Leukaemia Grp, Leicester, Leics, England.
   [Ironside, J. W.] Univ Edinburgh, Western Gen Hosp, Edinburgh, Midlothian, Scotland.
   [Cox, T.; Chong, W. K.; Michalski, A.] Great Ormond St Hosp Sick Children, London, England.
   [Thorp, N.; Pizer, B.] Alder Hey Childrens Hosp, Liverpool L12 2AP, Merseyside, England.
   [Ellison, D. W.] St Jude Childrens Hosp, Dept Pathol, Memphis, TN 38105 USA.
C3 University of Nottingham; University of Leicester; University of
   Edinburgh; University of London; University College London; Great Ormond
   Street Hospital for Children NHS Foundation Trust; Alder Hey Children's
   NHS Foundation Trust; Alder Hey Children's Hospital; St Jude Children's
   Research Hospital
RP Grundy, RG (通讯作者)，Univ Nottingham, Sch Med, Childrens Brain Tumour Res Ctr, Queens Med Ctr, Nottingham NG7 2UH, England.
EM
RI Ellison, David/N-4806-2018; Lien, Brian/AAI-7882-2020
OI Ironside, James/0000-0001-5869-2108; THORP, NICOLA/0000-0003-3754-9075
FU Cancer Research-UK; Samantha Dickson Brain Tumour Trust
FX The Children's Cancer and Leukaemia Group (CCLG) is supported by Cancer
   Research-UK and this study was generously supported by the Samantha
   Dickson Brain Tumour Trust. The sponsors have taken no role in study
   design, collection, analysis and interpretation of the data or in the
   writing of the report.; The following investigators and institutions
   participated in this study: Coordinating Centre: CCLG Data Centre,
   University of Leicester, Leicester, UK; Clinical Centres: (the number of
   patients enrolled from each Centre is given in parentheses); Denmark:
   University Hospital, Copenhagen (1); Eire: our Lady's Hospital for Sick
   Children, Dublin (6); England: Birmingham Children's Hospital,
   Birmingham (7); Bristol Children's Hospital, Bristol (6); Addenbrooke's
   Hospital, Cambridge (2); St James' University Hospital, Leeds (7); Great
   Ormond Street Hospital for Children, London (11); Guy's Hospital, London
   (2); The Royal Manchester Children's Hospital, Manchester (12); Royal
   Victoria infirmary, Newcastle upon Tyne (6); Queen's Medical Centre,
   Nottingham (12); John Radcliffe Hospital, Oxford (2); Sheffield
   Children's Hospital, Sheffield (1); Southampton General Hospital,
   Southampton (1); Royal Marsden Hospital, Sutton (4); Northern Ireland:
   The Royal Hospital for Sick Children, Belfast (4); Scotland: Royal
   Hospital for Sick Children, Edinburgh (4); Spain: Centro Medico La
   Zarzuelo, Madrid (2); The Netherlands: Emma Kinderziekenhuis, Amsterdam
   (6); Wales: The Children's Hospital for Wales, Cardiff (1).; We are
   grateful to Drs. Martin English and Heidi Traunecker for reviewing and
   commenting on the manuscript.
CR ALLEN J, 1992, J NEURO-ONCOL, V12, P69
   Allen JC, 1996, PEDIATR NEUROSURG, V24, P98, DOI 10.1159/000121024
   [Anonymous], 2007, SURVIVAL ANAL PRACTI
   Berger C, 1998, NEUROSURGERY, V42, P470, DOI 10.1097/00006123-199803000-00006
   Duffner P K, 1999, Neuro Oncol, V1, P152, DOI 10.1093/neuonc/1.2.152
   DUFFNER PK, 1985, CANCER-AM CANCER SOC, V56, P1841, DOI 10.1002/1097-0142(19851001)56:7+<1841::AID-CNCR2820561325>3.0.CO;2-C
   DUFFNER PK, 1993, NEW ENGL J MED, V328, P1725, DOI 10.1056/NEJM199306173282401
   DUFFNER PK, 1989, ANN NEUROL, V26, P460
   Dufour C, 2006, EUR J CANCER, V42, P2939, DOI 10.1016/j.ejca.2006.06.021
   GEYER JR, 1995, CANCER-AM CANCER SOC, V75, P1045, DOI 10.1002/1097-0142(19950215)75:4<1045::AID-CNCR2820750422>3.0.CO;2-K
   Geyer JR, 2005, J CLIN ONCOL, V23, P7621, DOI 10.1200/JCO.2005.09.095
   Giangaspero F, 1999, J NEUROSURG, V91, P971, DOI 10.3171/jns.1999.91.6.0971
   GIANGASPERO F, WHO CLASSIFICATION T
   Grill J, 2005, LANCET ONCOL, V6, P573, DOI 10.1016/S1470-2045(05)70252-7
   Grundy RG, 2007, LANCET ONCOL, V8, P696, DOI 10.1016/S1470-2045(07)70208-5
   HOROWITZ ME, 1988, NEUROSURGERY, V22, P687, DOI 10.1227/00006123-198804000-00011
   HYDER D, 2007, J NEUROONCOL
   Kaplan AM, 1996, PEDIATR NEUROSURG, V24, P185, DOI 10.1159/000121036
   Kellie SJ, 1999, CHILD NERV SYST, V15, P592, DOI 10.1007/s003810050548
   Lashford LS, 1996, ARCH DIS CHILD, V74, P219, DOI 10.1136/adc.74.3.219
   LOUIS LN, 2007, WHO CLASSIFICATION T
   MICHALSKI AG, 2004, BRAIN SPINAL TUMOURS, P359
   Mulhern RK, 2004, LANCET ONCOL, V5, P399, DOI 10.1016/S1470-2045(04)01507-4
   PACKER R, 1991, CANCER, V69, P580
   Pollack IF, 2003, NEURO-ONCOLOGY, V5, P197, DOI 10.1215/S1152851703000097
   Pomeroy SL, 2002, NATURE, V415, P436, DOI 10.1038/415436a
   Romer J, 2005, CANCER RES, V65, P4975, DOI 10.1158/0008-5472.CAN-05-0481
   Rutkowski S, 2005, NEW ENGL J MED, V352, P978, DOI 10.1056/NEJMoa042176
   Sanders RP, 2007, PEDIATR BLOOD CANCER, V49, P888, DOI 10.1002/pbc.21272
   TAI BC, 2009, INT J RAD ONCOL BIOL
   Thompson MC, 2006, J CLIN ONCOL, V24, P1924, DOI 10.1200/JCO.2005.04.4974
   van der Knaap MS, 2005, MAGNETIC RESONANCE M, P1, DOI DOI 10.1007/3-540-27660-2_1
   VANEYS J, 1985, J NEURO-ONCOL, V3, P237, DOI 10.1007/BF00165184
   Wrede B, 2005, ANTICANCER RES, V25, P4429
NR 34
TC 76
Z9 85
U1 0
U2 6
PU ELSEVIER SCI LTD
PI OXFORD
PA THE BOULEVARD, LANGFORD LANE, KIDLINGTON, OXFORD OX5 1GB, OXON, ENGLAND
SN 0959-8049
J9 EUR J CANCER
JI Eur. J. Cancer
PD JAN
PY 2010
VL 46
IS 1
BP 120
EP 133
DI 10.1016/j.ejca.2009.09.013
PG 14
WC Oncology
WE Science Citation Index Expanded (SCI-EXPANDED)
SC Oncology
GA 550BZ
UT WOS:000274103600016
PM 19818598
DA 2023-08-27
ER

PT J
AU Ramos, AA
   Vega, MAA
   Valencia, HS
   Garcia, JC
   Perez, VC
AF Antuna Ramos, Aida
   Alvarez Vega, Marco Antonio
   Sandoval Valencia, Hernan
   Campos Garcia, Juan
   Calatayud Perez, Vicente
TI Intraparenchymal Schwannoma Involving the Brainstem in a Young Woman
SO PEDIATRIC NEUROLOGY
LA English
DT Article
ID OF-THE-LITERATURE; INTRACEREBRAL SCHWANNOMA
AB Schwannomas are tumors derived from the Schwann cells, which form the myelin sheath of the peripheral nerves. Fewer than 1% of these tumors occur within the brain parenchyma without arising from the cranial nerves. Only 55 cases have been published after the first recorded case. We report a 17-year-old girl with a 3-month history of unspecific dizziness, unsteadiness, and headache. Magnetic resonance imaging showed a heterogeneous cystic lesion involving midbrain, pons, and left cerebellar peduncle. The patient underwent a retromastoid craniotomy with complete resection of the tumor. Pathologic examination was compatible with intraparenchymal schwannoma. Since the first case of intraparenchymal schwannoma involving the brainstem was described in 1980, only seven others have been reported. Diagnosis of intraparenchymal schwannoma is almost never made preoperatively. Immunohistochemical staining is crucial in distinguishing a Schwannoma from a meningioma, glial tumor, or metastatic tumor. Pathologic findings are those typical of acoustic neurinomas. Histogenesis of intraparenchymal schwannoma remains unclear, and several theories have been proposed to explain their origin. The recognition of this curable tumor and its differentiation from brainstem glioma, which generally has a less favorable outcome, is of obvious importance. (c) 2013 Elsevier Inc. All rights reserved.
C1 [Antuna Ramos, Aida; Sandoval Valencia, Hernan; Campos Garcia, Juan; Calatayud Perez, Vicente] Gen Hosp Albacete, Dept Neurosurg, Albacete, Spain.
   [Alvarez Vega, Marco Antonio] Univ Cent Hosp Asturias, Dept Neurosurg, Oviedo, Spain.
RP Ramos, AA (通讯作者)，Gen Hosp Albacete, Dept Neurosurg, C Fernandez Ladreda 30,6 B, Oviedo 33011, Asturias, Spain.
EM
RI Alvarez-Vega, Marco Antonio/AAB-8257-2019
OI Alvarez-Vega, Marco Antonio/0000-0001-8183-5963
CR Anton T, 2006, J NEURO-ONCOL, V76, P307, DOI 10.1007/s11060-005-7286-y
   Beskonakli E, 1997, NEUROSURG REV, V20, P139, DOI 10.1007/BF01138200
   CASADEI GP, 1995, CANCER, V75, P1109, DOI 10.1002/1097-0142(19950301)75:5<1109::AID-CNCR2820750510>3.0.CO;2-M
   GIBSON AAM, 1966, J NEUROSURG, V24, P552, DOI 10.3171/jns.1966.24.2.0552
   Lee SH, 1999, ACTA NEUROCHIR, V141, P779, DOI 10.1007/s007010050375
   Lin JK, 2003, J NEUROSURG, V98, P621, DOI 10.3171/jns.2003.98.3.0621
   Long DM, 1998, J NEUROSURG, V64, P446
   Muzzafar S, 2010, AM J NEURORADIOL, V31, P567, DOI 10.3174/ajnr.A1758
   Paredes I, 2012, CLIN NEUROL NEUROSUR, V114, P42, DOI 10.1016/j.clineuro.2011.07.003
   PRAKASH B, 1980, J NEUROSURG, V53, P121, DOI 10.3171/jns.1980.53.1.0121
   SARKAR C, 1987, J NEUROSURG, V67, P120, DOI 10.3171/jns.1987.67.1.0120
   Sharma MC, 1996, J NEUROL NEUROSUR PS, V60, P200, DOI 10.1136/jnnp.60.2.200
   Srivastav AK, 2011, NEUROL INDIA, V59, P924, DOI 10.4103/0028-3886.91389
   Tanabe Michiharu, 1996, Neurologia Medico-Chirurgica, V36, P880, DOI 10.2176/nmc.36.880
   Tsuiki H, 1997, ACTA NEUROCHIR, V139, P756, DOI 10.1007/BF01420049
   Zagardo MT, 1998, AM J NEURORADIOL, V19, P1290
NR 16
TC 8
Z9 8
U1 0
U2 5
PU ELSEVIER SCIENCE INC
PI NEW YORK
PA 360 PARK AVE SOUTH, NEW YORK, NY 10010-1710 USA
SN 0887-8994
EI 1873-5150
J9 PEDIATR NEUROL
JI Pediatr. Neurol.
PD JUN
PY 2013
VL 48
IS 6
BP 472
EP 474
DI 10.1016/j.pediatrneurol.2013.02.009
PG 3
WC Clinical Neurology; Pediatrics
WE Science Citation Index Expanded (SCI-EXPANDED)
SC Neurosciences & Neurology; Pediatrics
GA 152QG
UT WOS:000319545600012
PM 23668875
DA 2023-08-27
ER

PT J
AU Ebner, FH
   Roser, F
   Shiozawa, T
   Ruetschlin, S
   Kirschniak, A
   Koerbel, A
   Tatagiba, M
AF Ebner, F. H.
   Roser, F.
   Shiozawa, T.
   Ruetschlin, S.
   Kirschniak, A.
   Koerbel, A.
   Tatagiba, M.
TI Petrosal vein occlusion in cerebello-pontine angle tumour surgery: An
   anatomical study of alternative draining pathways
SO EJSO
LA English
DT Article
DE Petrosal vein occlusion; Posterior fossa surgery; Petroclival meningioma
ID MR-ANGIOGRAPHY; MICROVASCULAR DECOMPRESSION; MICROSURGICAL ANATOMY;
   VENOGRAPHY; BRAIN; FOSSA; TESLA
AB Objective: The present anatomic study investigates alternative draining pathways of the petrosal vein territory, which allow compensation in case of surgical sacrifice.
   Methods: In eight (four formaldehyde fixed and four alcohol fixed) specimens the petrosal vein complex has been dissected and studied. Three heads have been selectively injected via the superior petrous sinus with colored silicon in two different colors. Thereafter the posterior Fossa content was removed epidurally from the skull and further fixed in 4% formaldehyde. The nervous and vascular structures were dissected under microscopic control. measured and photographed. 3D-photographs were elaborated.
   Results: The petrosal vein was present in all cases and joined the superior petrous sinus always lateral to the trigeminal nerve as a single trunk. In the selectively injected specimens no passage of the colored silicon mixture to the contralateral venous brainstem territory could be discerned. However, the ipsilateral anastomoses to the deep supratentorial venous system - peduncular, anterolateral pontomesencephalic. lateral mesencephalic veins. and the rectal veins in connection with the pontotrigeminal vein - filled in all cases.
   Conclusion: Although the present anatomical model does not reflect physiological aspects of vascular dynamics. we document an apparently compensatory venous blood drainage occurring via anastomotic pathways directed to the ipsilateral supratentorial venous system. These findings represent an interesting aspect for preoperative image-guided planning in cerebello-pontine angle surgery. (C) 2008 Elsevier Ltd. All rights reserved.
C1 [Ebner, F. H.; Roser, F.; Koerbel, A.; Tatagiba, M.] Univ Tubingen, Dept Neurosurg, D-72076 Tubingen, Germany.
   [Shiozawa, T.; Ruetschlin, S.; Kirschniak, A.] Univ Tubingen, Inst Anat, Tubingen, Germany.
   [Kirschniak, A.] Univ Tubingen, Dept Gen Visceral & Transplant Surg, Tubingen, Germany.
C3 Eberhard Karls University of Tubingen; Eberhard Karls University of
   Tubingen; Eberhard Karls University of Tubingen
RP Ebner, FH (通讯作者)，Univ Tubingen, Dept Neurosurg, Morgenstelle 1, D-72076 Tubingen, Germany.
EM
RI Shiozawa, Thomas/AAF-6083-2020
OI Roser, Florian/0000-0002-7854-6789
FU Eberhard-Karls University Tubingen
FX This study was supported by a grant of the fortune-program,
   Eberhard-Karls University Tubingen. The authors are grateful to Mr. Gerd
   Pfister for professional help in elaborathig 3D-photographs.
CR Ardeshiri A, 2006, NEUROSURG REV, V29, P154, DOI 10.1007/s10143-005-0016-2
   ARDESHIRI A, 2006, NEUROSURG REV, V29, P58
   Dandy WE, 1929, ARCH SURG-CHICAGO, V18, P687, DOI 10.1001/archsurg.1929.04420030081005
   Deda H, 2005, SURG NEUROL, V64, P67, DOI 10.1016/j.surneu.2005.07.056
   Duvernoy HM., 1975, SUPERFICIAL VEINS HU, DOI [10.1007/978-3-642-65929-4, DOI 10.1007/978-3-642-65929-4]
   Fujimaki T, 1999, J NEUROSURG, V90, P1148
   Gharabaghi A, 2006, NEUROSURGERY, V59, P68, DOI 10.1227/01.NEU.0000219821.34450.59
   Koerbel A, 2009, EJSO-EUR J SURG ONC, V35, P773, DOI 10.1016/j.ejso.2008.02.015
   KOROSEC FR, 1993, MAGN RESON MED, V30, P704, DOI 10.1002/mrm.1910300608
   Lee YH, 2005, BRIT J RADIOL, V78, P948, DOI 10.1259/bjr/71475327
   MATSUSHIMA T, 1983, J NEUROSURG, V59, P63, DOI 10.3171/jns.1983.59.1.0063
   McLaughlin MR, 1999, J NEUROSURG, V90, P1, DOI 10.3171/jns.1999.90.1.0001
   Nakamura M, 2005, NEUROSURGERY, V57, P77, DOI 10.1227/01.NEU.0000154699.29796.34
   NISHIMURA DG, 1990, MAGNET RESON MED, V14, P194, DOI 10.1002/mrm.1910140206
   Rauscher A, 2003, J MAGN RESON IMAGING, V18, P175, DOI 10.1002/jmri.10346
   Reichenbach JR, 1997, RADIOLOGY, V204, P272, DOI 10.1148/radiology.204.1.9205259
   Reichenbach JR, 2000, J COMPUT ASSIST TOMO, V24, P949, DOI 10.1097/00004728-200011000-00023
   Rhoton AL, 2000, NEUROSURGERY, V47, pS69, DOI 10.1097/00006123-200009001-00012
   RUNGE VM, 1993, JMRI-J MAGN RESON IM, V3, P233, DOI 10.1002/jmri.1880030135
   Samii M, 1999, J CLIN NEUROSCI, V6, P27, DOI 10.1016/S0967-5868(99)90598-2
   Samii M, 2000, J NEUROSURG, V92, P235, DOI 10.3171/jns.2000.92.2.0235
   Samii M, 1997, NEUROSURGERY, V40, P11, DOI 10.1097/00006123-199701000-00002
   Samii M, 1997, NEUROSURGERY, V40, P29
   Sehgal V, 2005, J MAGN RESON IMAGING, V22, P439, DOI 10.1002/jmri.20404
   Strauss C, 2000, J NEUROL NEUROSUR PS, V69, P679, DOI 10.1136/jnnp.69.5.679
NR 25
TC 18
Z9 20
U1 0
U2 6
PU ELSEVIER SCI LTD
PI OXFORD
PA THE BOULEVARD, LANGFORD LANE, KIDLINGTON, OXFORD OX5 1GB, OXON, ENGLAND
SN 0748-7983
EI 1532-2157
J9 EJSO-EUR J SURG ONC
JI EJSO
PD MAY
PY 2009
VL 35
IS 5
BP 552
EP 556
DI 10.1016/j.ejso.2008.06.011
PG 5
WC Oncology; Surgery
WE Science Citation Index Expanded (SCI-EXPANDED)
SC Oncology; Surgery
GA 441XJ
UT WOS:000265803500018
PM 18653307
DA 2023-08-27
ER

PT J
AU Lu, VM
   Brown, DA
   Daniels, DJ
AF Lu, Victor M.
   Brown, Desmond A.
   Daniels, David J.
TI Rare Diffuse Intrinsic Pontine Glioma Metastasis Throughout the Brain
   and Spine
SO WORLD NEUROSURGERY
LA English
DT Article
DE Diffuse intrinsic pontine glioma; DIPG; H3 K27M; Metastasis; Pediatric
AB Diffuse intrinsic pontine glioma with H3 K27M mutation is a rare brain tumor that primarily affects children. It is extremely lethal, and our understanding of the natural course of this disease, and how it progresses, is lacking. We have presented a case that demonstrates how aggressive this disease can be after progression, with remarkable spread throughout the brain and spine despite upfront radiotherapy. Although rarely reported, widespread dissemination of metastatic diffuse intrinsic pontine glioma throughout the brain and spine is possible.
C1 [Lu, Victor M.; Brown, Desmond A.; Daniels, David J.] Mayo Clin, Dept Neurol Surg, Rochester, MN 55905 USA.
C3 Mayo Clinic
RP Daniels, DJ (通讯作者)，Mayo Clin, Dept Neurol Surg, Rochester, MN 55905 USA.
EM
RI Brown, Desmond/AAP-7304-2020
OI Brown, Desmond/0000-0002-2896-1722
CR [Anonymous], 2017, J NEUROONCOL, DOI DOI 10.1007/S11060-016-2363-Y
   Belien ATJ, 1999, J CELL BIOL, V144, P373, DOI 10.1083/jcb.144.2.373
   BRYAN P, 1974, CLIN RADIOL, V25, P355, DOI 10.1016/S0009-9260(74)80168-6
   Buczkowicz P, 2014, ACTA NEUROPATHOL, V128, P573, DOI 10.1007/s00401-014-1319-6
   Sethi R, 2011, J NEURO-ONCOL, V102, P121, DOI 10.1007/s11060-010-0301-y
NR 5
TC 5
Z9 5
U1 0
U2 2
PU ELSEVIER SCIENCE INC
PI NEW YORK
PA STE 800, 230 PARK AVE, NEW YORK, NY 10169 USA
SN 1878-8750
EI 1878-8769
J9 WORLD NEUROSURG
JI World Neurosurg.
PD AUG
PY 2020
VL 140
BP 301
EP 302
DI 10.1016/j.wneu.2020.05.205
PG 2
WC Clinical Neurology; Surgery
WE Science Citation Index Expanded (SCI-EXPANDED)
SC Neurosciences & Neurology; Surgery
GA NA7WY
UT WOS:000560029600026
PM 32474088
DA 2023-08-27
ER

PT J
AU Zhang, Y
   Pan, CC
   Wang, JM
   Cao, JL
   Liu, YH
   Wang, YJ
   Zhang, LW
AF Zhang, Yang
   Pan, Changcun
   Wang, Junmei
   Cao, Jingli
   Liu, Yuhan
   Wang, Yajie
   Zhang, Liwei
TI Genetic and immune features of resectable malignant brainstem gliomas
SO ONCOTARGET
LA English
DT Article
DE brainstem gliomas; genetic features; programmed death ligand 1; CD8+T
   cell infiltration; prognosis
ID GRADE GLIOMAS; PD-1 BLOCKADE; CHILDREN; SAFETY; B7-H1; TEMOZOLOMIDE;
   LYMPHOCYTES; EXPRESSION; MUTATIONS; PROGNOSIS
AB We surveyed common genetic mutations (IDH1, H3F3A, PPM1D, and TP53) and immune features (PD-L1 expression and CD8(+) T cell tumor infiltration) in a series of 62 malignant brainstem gliomas that were resected via microsurgery. IDH1 mutations were mutually exclusive with H3F3A mutations. IDH1 mutations appeared only in adults and occurred more frequently in tumors larger than 10cm(3) (8/29 vs 1/32, Fisher's exact test, p=0.010). H3F3A mutations occurred more frequently in children and adolescent patients (19/24 vs 18/38, chi-square test, p=0.013), low preoperative Karnofsky Performance Scale (KPS) patients (10/11 vs 20/43, chi-square test, p=0.021), and higher grade brainstem gliomas (8/21 in grade II vs 16/24 in grade III vs 13/17 in grade IV; chi-square test, p=0.038). PPM1D mutations clustered in H3F3A-mutated tumors (12/37), whereas were rare in H3F3A wildtype tumors (1/25). MGMT promoter methylations clustered in IDH1-mutated tumors (4/9), but were rare in H3F3A-mutated tumors (1/37). PD-L1 staining was detected in 59.7% of brainstem glioma specimens (37/62). High intra-tumoral CD8(+) T cell density was less frequent in the H3F3A-mutated than H3F3A-wild-type tumors (4/37 vs. 11/25, p=0.005). Patients with H3F3A-mutated tumors (13.8 months overall survival) had much worse prognoses than those with IDH1-mutated (54.9 months, p=0.001) or H3F3A-IDH1 co-wildtype tumors (38.4 months, p=0.001). H3F3A mutations independently increased the relative risk of death as much as 4.19-fold according to a multivariate Cox regression model. Taken together, resectable malignant brainstem gliomas can be classified into three subtypes: H3F3A-mutated, IDH1 mutated and H3F3A-IDH1 co-wildtype tumors, which have distinct clinical characteristics, prognoses, genetic and immune features.
C1 [Zhang, Yang; Pan, Changcun; Liu, Yuhan; Zhang, Liwei] Capital Med Univ, China Natl Clin Res Ctr Neurol Dis, Beijing Tiantan Hosp, Dept Neurosurg, Beijing 100050, Peoples R China.
   [Wang, Junmei] Capital Med Univ, Beijing Neurosurg Inst, Dept Pathol, Beijing 100050, Peoples R China.
   [Cao, Jingli; Wang, Yajie] Capital Med Univ, Beijing Tiantan Hosp, Core Lab Clin Med Res, Beijing 100050, Peoples R China.
C3 Capital Medical University; Beijing Neurosurgical Institute; Capital
   Medical University; Capital Medical University
RP Zhang, LW (通讯作者)，Capital Med Univ, China Natl Clin Res Ctr Neurol Dis, Beijing Tiantan Hosp, Dept Neurosurg, Beijing 100050, Peoples R China.
EM
RI liu, jiajia/ISS-0316-2023; Yu, Kun/IAP-9807-2023; liu,
   jiajia/IUN-0901-2023; liu, jia/HKE-9796-2023; Li, JW/HNC-1743-2023; li,
   jiawei/HOA-5023-2023
FU Beijing Natural Science Foundation [7164253]; National Key Technology
   Research and Development Program of the Ministry of Science and
   Technology of China [2013BAI09B01]; Key Control Technology Research for
   Ischemic Cerebrovascular Disease and Brain Tumor [2015BAI12B04]; Beijing
   Talents Fund [2014000021469G257]
FX This work was supported by grants from Beijing Natural Science
   Foundation [7164253], National Key Technology Research and Development
   Program of the Ministry of Science and Technology of China
   [2013BAI09B01], The Key Control Technology Research for Ischemic
   Cerebrovascular Disease and Brain Tumor [2015BAI12B04] and Beijing
   Talents Fund [2014000021469G257].
CR Ansell SM, 2015, NEW ENGL J MED, V372, P311, DOI 10.1056/NEJMoa1411087
   Babu R, 2014, J NEURO-ONCOL, V119, P177, DOI 10.1007/s11060-014-1471-9
   Berghoff AS, 2015, NEURO-ONCOLOGY, V17, P1064, DOI 10.1093/neuonc/nou307
   Blank C, 2004, CANCER RES, V64, P1140, DOI 10.1158/0008-5472.CAN-03-3259
   Brahmer JR, 2012, NEW ENGL J MED, V366, P2455, DOI 10.1056/NEJMoa1200694
   Chiang KL, 2010, CHILD NERV SYST, V26, P1035, DOI 10.1007/s00381-010-1106-1
   Dahlin AM, 2011, MODERN PATHOL, V24, P671, DOI 10.1038/modpathol.2010.234
   Dong HD, 2002, NAT MED, V8, P793, DOI 10.1038/nm730
   Durand-Panteix S, 2012, J IMMUNOL, V189, P181, DOI 10.4049/jimmunol.1102277
   Dutoit V, 2016, FRONT ONCOL, V6, DOI 10.3389/fonc.2016.00256
   Eisenstat DD, 2015, J NEURO-ONCOL, V121, P573, DOI 10.1007/s11060-014-1669-x
   Goodwin CR, 2016, CLIN NEUROL NEUROSUR, V142, P120, DOI 10.1016/j.clineuro.2016.01.007
   Hamid O, 2013, NEW ENGL J MED, V369, P134, DOI 10.1056/NEJMoa1305133
   Han S, 2014, BRIT J CANCER, V110, P2560, DOI 10.1038/bjc.2014.162
   Hu J, 2016, FRONT ONCOL, V6, DOI 10.3389/fonc.2016.00180
   Hu SQ, 2015, PLOS GENET, V11, DOI 10.1371/journal.pgen.1005367
   Johung TB, 2017, CURR NEUROPHARMACOL, V15, P88, DOI 10.2174/1570159X14666160509123229
   Kim YH, 2012, BRIT J NEUROSURG, V26, P21, DOI 10.3109/02688697.2011.584986
   Lachi Pavan Kumar, 2015, Asian J Neurosurg, V10, P298, DOI 10.4103/1793-5482.162709
   Louis DN, 2016, ACTA NEUROPATHOL, V131, P803, DOI 10.1007/s00401-016-1545-1
   Ludwig K, 2017, J NEUROONCOL
   Madore J, 2015, PIGM CELL MELANOMA R, V28, DOI 10.1111/pcmr.12340
   Ostrom QT, 2014, NEURO-ONCOLOGY, V16, P1, DOI [10.1093/neuonc/nov189, 10.1093/neuonc/nou327, 10.1093/neuonc/nou223]
   Powles T, 2014, NATURE, V515, P558, DOI 10.1038/nature13904
   Reardon DA, 2014, NEURO-ONCOLOGY, V16, P1441, DOI 10.1093/neuonc/nou212
   Reithmeier T, 2014, BMC CANCER, V14, DOI 10.1186/1471-2407-14-115
   Reitman ZJ, 2014, PHARMACOGENOMICS, V15, P1639, DOI [10.2217/PGS.14.123, 10.2217/pgs.14.123]
   Reyes-Botero G, 2012, ONCOLOGIST, V17, P388, DOI 10.1634/theoncologist.2011-0335
   Rizzo D, 2015, J CHEMOTHERAPY, V27, P106, DOI 10.1179/1973947814Y.0000000228
   Robert C, 2014, LANCET, V384, P1109, DOI 10.1016/S0140-6736(14)60958-2
   Sharma P, 2015, SCIENCE, V348, P56, DOI 10.1126/science.aaa8172
   Smith MA, 1998, J NATL CANCER I, V90, P1269, DOI 10.1093/jnci/90.17.1269
   Sunshine J, 2015, CURR OPIN PHARMACOL, V23, P32, DOI 10.1016/j.coph.2015.05.011
   Topalian SL, 2016, NAT REV CANCER, V16, P275, DOI 10.1038/nrc.2016.36
   Topalian SL, 2012, NEW ENGL J MED, V366, P2443, DOI 10.1056/NEJMoa1200690
   Tumeh PC, 2014, NATURE, V515, P568, DOI 10.1038/nature13954
   Vanan MI, 2015, FRONT ONCOL, V5, DOI 10.3389/fonc.2015.00237
   Wu G, 2014, NAT GENET, V46, P444, DOI 10.1038/ng.2938
   Yamasaki F, 2011, NEURO-ONCOLOGY, V13, P791, DOI 10.1093/neuonc/nor038
   Yan H, 2009, NEW ENGL J MED, V360, P765, DOI 10.1056/NEJMoa0808710
   Yao Y, 2009, NEURO-ONCOLOGY, V11, P757, DOI 10.1215/15228517-2009-014
   Zhang LW, 2014, NAT GENET, V46, P726, DOI 10.1038/ng.2995
NR 42
TC 9
Z9 9
U1 0
U2 5
PU IMPACT JOURNALS LLC
PI ORCHARD PARK
PA 6666 E QUAKER ST, STE 1, ORCHARD PARK, NY 14127 USA
EI 1949-2553
J9 ONCOTARGET
JI Oncotarget
PD OCT 10
PY 2017
VL 8
IS 47
BP 82571
EP 82582
DI 10.18632/oncotarget.19653
PG 12
WC Oncology; Cell Biology
WE Science Citation Index Expanded (SCI-EXPANDED)
SC Oncology; Cell Biology
GA FJ4DA
UT WOS:000412683900077
PM 29137285
OA Green Published, gold
DA 2023-08-27
ER

PT J
AU Komune, N
   Yagmurlu, K
   Matsuo, S
   Miki, K
   Abe, H
   Rhoton, AL
AF Komune, Noritaka
   Yagmurlu, Kaan
   Matsuo, Satoshi
   Miki, Koichi
   Abe, Hiroshi
   Rhoton, Albert L., Jr.
TI Auditory Brainstem Implantation: Anatomy and Approaches
SO OPERATIVE NEUROSURGERY
LA English
DT Article
DE Auditory brainstem implantation; Auditory pathways; Brainstem; Cochlear
   nuclei; Inferior colliculus; Retrosigmoid; Translabyrinthine
ID INFRATENTORIAL SUPRACEREBELLAR APPROACH; VESTIBULAR SCHWANNOMA SURGERY;
   NEUROFIBROMATOSIS TYPE-2; ELECTRICAL-STIMULATION; COCHLEAR IMPLANTATION;
   MIDBRAIN IMPLANT; INFERIOR COLLICULUS; CLINICAL-EXPERIENCE; SURGICAL
   ANATOMY; NF2 PATIENTS
AB BACKGROUND: Auditory brainstem implantation at the cochlear nuclei used mainly for neurofibromatosis type 2 patients with bilateral loss of the cochlear nerves has more recently been extended to the inferior colliculus.
   OBJECTIVE: To examine the microsurgical and endoscopic anatomy of the cochlear nuclei and inferior colliculus as seen through the translabyrinthine and retrosigmoid approaches used for cochlear nuclei and inferior collicular implantation. METHODS: Ten cerebellopontine angles of formalin-fixed adult cadaveric heads were examined with the aid of the surgical microscope and endoscope. The ascending auditory pathways between the cochlear nuclei and inferior colliculi and above were examined by the fiber dissection technique.
   RESULTS: Both the translabyrinthine and retrosigmoid routes provide sufficient exposure for concurrent tumor removal and implantation at either the cochlear nuclei or inferior colliculus. The position of the inferior colliculus in the auditory pathways and its accessibility in the infratentorial supracerebellar exposure directed through either the translabyrinthine or retrosigmoid approach makes it an alternative site for electrode placement if the cochlear nuclei are not functionally or structurally suitable for implantation. Endoscopic assistance may aid the exposure and electrode placement at either site.
   CONCLUSION: The translabyrinthine or retrosigmoid approaches provide access to the cochlear nuclei for implantation and also to the inferior colliculus through the translabyrinthine or retrosigmoid infratentorial supracerebellar route. The endoscope may aid in exposing either site.
C1 [Komune, Noritaka; Yagmurlu, Kaan; Matsuo, Satoshi; Rhoton, Albert L., Jr.] Univ Florida, Coll Med, Dept Neurosurg, Gainesville, FL USA.
   [Miki, Koichi; Abe, Hiroshi] Fukuoka Univ, Dept Neurosurg, Fukuoka 81401, Japan.
C3 State University System of Florida; University of Florida; Fukuoka
   University
RP Rhoton, AL (通讯作者)，Dept Neurosurg, POB 100265, Gainesville, FL 32610 USA.
EM
RI Matsuo, Satoshi/AAE-3849-2020
OI Matsuo, Satoshi/0000-0002-2052-0471; YAGMURLU, KAAN/0000-0002-7635-2809
FU University of Florida Foundation
FX The authors have no personal, financial, or institutional interest in
   any of the drugs, materials, or devices described in this article. This
   work was supported by a University of Florida Foundation.
CR Abe H, 2006, NEUROSURGERY, V58, P728, DOI 10.1227/01.NEU.0000204870.83778.A1
   Ammirati M, 2002, J NEUROSURG, V97, P922, DOI 10.3171/jns.2002.97.4.0922
   ARRIAGA MMA, 1995, OTOLARYNG HEAD NECK, V112, P325, DOI 10.1016/S0194-5998(95)70257-1
   Behr R, 2007, SKULL BASE-INTERD AP, V17, P91, DOI 10.1055/s-2006-950390
   Behr R, 2014, OTOL NEUROTOL, V35, P1844, DOI 10.1097/MAO.0000000000000584
   BRACKMANN DE, 1993, OTOLARYNG HEAD NECK, V108, P624, DOI 10.1177/019459989310800602
   Cai SQ, 2009, JARO-J ASSOC RES OTO, V10, P5, DOI 10.1007/s10162-008-0142-y
   Calixto R, 2013, PLOS ONE, V8, DOI 10.1371/journal.pone.0082148
   Choi JY, 2011, LARYNGOSCOPE, V121, P2610, DOI 10.1002/lary.22137
   Colletti L, 2012, CURR OPIN OTOLARYNGO, V20, P353, DOI 10.1097/MOO.0b013e328357613d
   Colletti V, 2000, AM J OTOL, V21, P826
   Colletti V, 2005, LARYNGOSCOPE, V115, P1974, DOI 10.1097/01.mlg.0000178327.42926.ec
   Colletti V, 2004, AUDIOL NEURO-OTOL, V9, P247, DOI 10.1159/000078394
   Colletti V, 2007, OTOL NEUROTOL, V28, P39, DOI 10.1097/01.mao.0000247808.47712.02
   Colletti V, 2009, OTOL NEUROTOL, V30, P614, DOI 10.1097/MAO.0b013e3181a864f2
   DOBELLE WH, 1973, ANN OTO RHINOL LARYN, V82, P445, DOI 10.1177/000348947308200404
   Eisenberg L S, 1987, J Rehabil Res Dev, V24, P9, DOI 10.1682/JRRD.1987.07.0009
   Friedland DR, 1999, LARYNGOSCOPE, V109, P175, DOI 10.1097/00005537-199902000-00001
   Geniec P, 1971, Acta Otolaryngol Suppl, V295, P1
   Grayeli AB, 2008, OTOL NEUROTOL, V29, P1140, DOI 10.1097/MAO.0b013e31818b6238
   Heller JW, 1996, AM J OTOL, V17, P724
   House WF, 2001, ANN OTO RHINOL LARYN, V110, P103, DOI 10.1177/000348940111000201
   LABORDE G, 1992, ACTA NEUROCHIR, V114, P135, DOI 10.1007/BF01400602
   Lenarz T, 2001, OTOL NEUROTOL, V22, P823, DOI 10.1097/00129492-200111000-00019
   Lenarz T, 2006, OTOL NEUROTOL, V27, P838, DOI 10.1097/01.mao.0000232010.01116.e9
   Lim HH, 2008, HEARING RES, V242, P74, DOI 10.1016/j.heares.2008.02.003
   Lim Hubert H, 2009, Trends Amplif, V13, P149, DOI 10.1177/1084713809348372
   Lloyd SKW, 2014, OTOL NEUROTOL, V35, P43, DOI 10.1097/MAO.0000000000000185
   Longatti P, 2009, NEUROSURGERY, V65, P1078, DOI 10.1227/01.NEU.0000360133.29217.44
   Merkus P, 2014, EUR ARCH OTO-RHINO-L, V271, P3, DOI 10.1007/s00405-013-2378-3
   Nakatomi Hirofumi, 2011, Nihon Jibiinkoka Gakkai Kaiho, V114, P851
   Ogata N, 1997, NEUROSURGERY, V40, P101, DOI 10.1097/00006123-199701000-00023
   Otto SR, 2002, J NEUROSURG, V96, P1063, DOI 10.3171/jns.2002.96.6.1063
   Otto SR, 2008, OTOL NEUROTOL, V29, P1147, DOI 10.1097/MAO.0b013e31818becb4
   Peris-Celda M, 2014, OPER NEUROSURG, V10, P121, DOI 10.1227/NEU.0000000000000205
   Rosahl SK, 2013, NEUROSURGERY, V72, P58, DOI 10.1227/NEU.0b013e31826cde82
   Rozet Irene, 2007, Anesthesiol Clin, V25, P631, DOI 10.1016/j.anclin.2007.05.009
   Samii A, 2007, OTOL NEUROTOL, V28, P31, DOI 10.1097/01.mao.0000247819.16325.7d
   Samii M, 1996, J NEUROSURG, V84, P375, DOI 10.3171/jns.1996.84.3.0375
   Sanna M, 2012, OTOL NEUROTOL, V33, P154, DOI 10.1097/MAO.0b013e318241bc71
   Schwartz MS, 2008, NEUROTHERAPEUTICS, V5, P128, DOI 10.1016/j.nurt.2007.10.068
   Schwartz MS, 2003, STEREOT FUNCT NEUROS, V81, P110, DOI 10.1159/000075113
   Seki Y, 2003, ACTA NEUROCHIR SUPPL, V87, P85
   Shannon RV, 2012, CURR OPIN NEUROL, V25, P61, DOI 10.1097/WCO.0b013e32834ef878
   Sharifi M, 2009, SURG RADIOL ANAT, V31, P431, DOI 10.1007/s00276-009-0464-4
   Siegbahn M, 2014, ACTA OTO-LARYNGOL, V134, P1052, DOI 10.3109/00016489.2014.909051
   SIMMONS FB, 1964, ARCHIV OTOLARYNGOL, V79, P559, DOI 10.1001/archotol.79.6.559
   STEIN BM, 1971, J NEUROSURG, V35, P197, DOI 10.3171/jns.1971.35.2.0197
   Straka MM, 2014, J NEURAL ENG, V11, DOI 10.1088/1741-2560/11/4/046021
   Sun JQ, 2010, ACTA OTO-LARYNGOL, V130, P1125, DOI 10.3109/00016481003699674
   Takemura Y, 2014, WORLD NEUROSURG, V82, P427, DOI 10.1016/j.wneu.2013.07.013
   TERR LI, 1985, ARCH OTOLARYNGOL, V111, P495
   Tysome JR, 2013, OTOL NEUROTOL, V34, P1743, DOI 10.1097/MAO.0b013e3182a1a8b4
   Ulm AJ, 2004, NEUROSURGERY, V54, P1313, DOI 10.1227/01.NEU.0000126129.68707.E7
   Vince GH, 2010, J CLIN NEUROSCI, V17, P107, DOI 10.1016/j.jocn.2009.06.034
   Vincent C, 2012, ANAT REC, V295, P1981, DOI 10.1002/ar.22588
   Vishteh AG, 2000, NEUROSURGERY, V46, P384, DOI 10.1097/00006123-200002000-00022
   Vollmer M, 2007, J NEUROPHYSIOL, V98, P2588, DOI 10.1152/jn.00011.2007
   Yagmurlu K, 2014, OPER NEUROSURG, V10, P602, DOI 10.1227/NEU.0000000000000466
NR 59
TC 14
Z9 15
U1 0
U2 2
PU OXFORD UNIV PRESS INC
PI CARY
PA JOURNALS DEPT, 2001 EVANS RD, CARY, NC 27513 USA
SN 2332-4252
EI 2332-4260
J9 OPER NEUROSURG
JI Oper. Neurosurg.
PD JUN
PY 2015
VL 11
IS 2
BP 306
EP 320
DI 10.1227/NEU.0000000000000736
PG 15
WC Clinical Neurology; Surgery
WE Science Citation Index Expanded (SCI-EXPANDED)
SC Neurosciences & Neurology; Surgery
GA CV4DE
UT WOS:000364215100032
PM 25793729
DA 2023-08-27
ER

PT J
AU Thong, PM
   Duc, NM
AF Minh Thong, Pham
   Minh Duc, Nguyen
TI A Rare Case of Intra-Fourth-Ventricular Germinoma, Derived from the
   Medulla Oblongata
SO PEDIATRIC NEUROSURGERY
LA English
DT Article
DE Germinoma; Fourth ventricle; Medulla oblongata; Posterior fossa tumor
ID GERM-CELL TUMORS; INTRACRANIAL GERMINOMA
AB Introduction: Extragonadal germinomas rarely emerge from the brain stem; however, proper diagnosis and treatment can result in favorable prognosis. Unfortunately, the preoperative diagnosis of medulla oblongata germinoma is difficult due to insufficient clinical signs and symptoms that are specific to this diagnosis. Case Representation: We present a 12-year-old male patient with an intra-fourth-ventricular germinoma, derived from the medulla oblongata, with no abnormalities in the supratentorial region. The germinoma was initially assessed by advance MRI sequences, including diffusion-weighted imaging, T1 perfusion, and spectroscopy. Conclusion: In summary, although existing imaging technologies cannot completely distinguish germinomas from other primary brain neoplasms in the fourth ventricle, in patients aged between 12 and 40 years, a small mass on the dorsal side of medulla oblongata that emerges into the fourth ventricle and is characterized by homogeneous contrast enhancement, the absence of calcification and hemorrhage, and the lack of hydrocephalus should be considered for a potential medulla oblongata germinoma diagnosis.
C1 [Minh Thong, Pham; Minh Duc, Nguyen] Hanoi Med Univ, Dept Radiol, Hanoi, Vietnam.
   [Minh Duc, Nguyen] Pham Ngoc Thach Univ Med, Dept Radiol, 2 Duong Quang Trung Ward 12 Dist 10, Ho Chi Minh City 700000, Vietnam.
   [Minh Duc, Nguyen] Childrens Hosp 2, Dept Radiol, Ho Chi Minh City, Vietnam.
C3 Hanoi Medical University
RP Duc, NM (通讯作者)，Pham Ngoc Thach Univ Med, Dept Radiol, 2 Duong Quang Trung Ward 12 Dist 10, Ho Chi Minh City 700000, Vietnam.
EM
OI , Pham Minh Thong/0000-0003-1294-2743; Minh Duc,
   Nguyen/0000-0001-5411-1492
CR Akimoto J, 2009, J CLIN NEUROSCI, V16, P705, DOI 10.1016/j.jocn.2008.06.009
   Arora RS, 2009, NEURO-ONCOLOGY, V11, P403, DOI 10.1215/15228517-2008-097
   Budohoski KP, 2016, BRIT J NEUROSURG, V30, P348, DOI 10.3109/02688697.2015.1096907
   Chen YW, 2012, CANCER-AM CANCER SOC, V118, P2752, DOI 10.1002/cncr.26564
   Fangusaro J, 2008, PEDIATR BLOOD CANCER, V50, P312, DOI 10.1002/pbc.21555
   Hao SY, 2013, WORLD J SURG ONCOL, V11, DOI 10.1186/1477-7819-11-274
   HASHIMOTO M, 1992, SURG NEUROL, V37, P384, DOI 10.1016/0090-3019(92)90009-C
   Hayashi S, 2015, J NEUROL SCI, V358, P512, DOI 10.1016/j.jns.2015.09.369
   HOFFMAN HJ, 1991, J NEUROSURG, V74, P545, DOI 10.3171/jns.1991.74.4.0545
   Hoque Romy, 2008, J La State Med Soc, V160, P319
   Hu YW, 2012, INT J RADIAT ONCOL, V84, P639, DOI 10.1016/j.ijrobp.2011.12.052
   Israel Z, 1996, ACTA NEUROCHIR, V138, P1252, DOI 10.1007/BF01809757
   IWASAKI I, 1984, Neurologia Medico-Chirurgica, V24, P51, DOI 10.2176/nmc.24.51
   Khan AA, 2013, J CLIN NEUROSCI, V20, P1620, DOI 10.1016/j.jocn.2012.11.025
   MASUZAWA T, 1986, CLIN NEUROPATHOL, V5, P190
   Nakajima H, 2000, SURG NEUROL, V53, P448, DOI 10.1016/S0090-3019(00)00224-X
   Nakatsuka S, 2012, INT J SURG PATHOL, V20, P276, DOI 10.1177/1066896911424489
   Neelima R, 2010, NEUROL INDIA, V58, P768, DOI 10.4103/0028-3886.72190
   Ostrom QT, 2013, NEURO-ONCOLOGY, V15, P1, DOI 10.1093/neuonc/not151
   POUNGVARIN N, 1991, Journal of the Medical Association of Thailand, V74, P55
   Rosenblum MK, 2000, PATHOLOGY GENETICS T, P207
   Sato K, 2009, PROG NEUROL SURG, V23, P59, DOI 10.1159/000210053
   Seifert K, 2020, WORLD NEUROSURG, V138, P137, DOI 10.1016/j.wneu.2020.02.147
   Shuto T, 2012, J CLIN NEUROSCI, V19, P769, DOI 10.1016/j.jocn.2011.06.036
   Sugiyama K, 1994, Neurol Med Chir (Tokyo), V34, P291, DOI 10.2176/nmc.34.291
   Tashiro Takashi, 1993, Neurologia Medico-Chirurgica, V33, P251, DOI 10.2176/nmc.33.251
   Yang DT, 2009, J CLIN NEUROSCI, V16, P321, DOI 10.1016/j.jocn.2007.08.026
   Yasuhara T, 2011, NEUROL MED-CHIR, V51, P326, DOI 10.2176/nmc.51.326
   Yen PS, 2003, J NEURO-ONCOL, V62, P339, DOI 10.1023/A:1023331700995
   Yip Chi-Man, 2014, Surg Neurol Int, V5, pS170, DOI 10.4103/2152-7806.134815
   Yoshida K, 2003, ACTA NEUROCHIR, V145, P789, DOI 10.1007/s00701-003-0043-0
NR 31
TC 0
Z9 0
U1 0
U2 0
PU KARGER
PI BASEL
PA ALLSCHWILERSTRASSE 10, CH-4009 BASEL, SWITZERLAND
SN 1016-2291
EI 1423-0305
J9 PEDIATR NEUROSURG
JI Pediatr. Neurosurg.
PD JAN
PY 2021
VL 55
IS 6
BP 426
EP 431
DI 10.1159/000512345
EA DEC 2020
PG 6
WC Clinical Neurology; Pediatrics; Surgery
WE Science Citation Index Expanded (SCI-EXPANDED)
SC Neurosciences & Neurology; Pediatrics; Surgery
GA QC3XC
UT WOS:000601392100001
PM 33352572
DA 2023-08-27
ER

PT J
AU Mhatre, R
   Bopanna, KM
   Ganigi, P
   Anita, M
AF Mhatre, Radhika
   Bopanna, K. M.
   Ganigi, Praveen
   Anita, Mahadevan
TI Rare occurrence of gangliocytoma in the medulla oblongata
SO CHILDS NERVOUS SYSTEM
LA English
DT Article
DE Gangliocytoma; Medulla oblongata; Hamartoma; Cerebellar seizures
ID HAMARTOMA; SEIZURES
AB Gangliocytoma is an uncommon low-grade neuroepithelial tumour arising in the cerebral hemispheres, with medulla oblongata being extremely rare site of occurrence. We report a case of an 8-year-old female with recurrent vomiting, hypertension and left hemifacial spasms with a dorsally exophytic lesion arising from the medulla projecting into the fourth ventricle. She underwent excision of the lesion with intra operative cranial nerve monitoring guidance. Histopathology revealed clusters of large mature ganglion cells which were labelled by synaptophysin and CD34. Extreme rarity in this location makes clinical diagnosis and management of gangliocytoma challenging, although prompt diagnosis and total excision has a successful outcome. There are only six previous reports of gangliocytoma in the medulla oblongata.
C1 [Mhatre, Radhika; Anita, Mahadevan] NIMHANS, Dept Neuropathol, Bangalore 560029, Karnataka, India.
   [Bopanna, K. M.; Ganigi, Praveen] Manipal Hosp, Bangalore, Karnataka, India.
C3 National Institute of Mental Health & Neurosciences - India
RP Anita, M (通讯作者)，NIMHANS, Dept Neuropathol, Bangalore 560029, Karnataka, India.
EM;
  
CR ALTMAN NR, 1988, AM J NEURORADIOL, V9, P917
   Awa R, 2012, J NEURO-ONCOL, V106, P201, DOI 10.1007/s11060-011-0641-2
   Capper D, 2016, WHO CLASSIFICATION T, P136
   Choi YH, 2001, PEDIATR RADIOL, V31, P377, DOI 10.1007/s002470100445
   Delalande O, 2001, NEUROSURGERY, V49, P726, DOI 10.1097/00006123-200109000-00037
   Koerbel A, 2003, ARQ NEURO-PSIQUIAT, V61, P274, DOI 10.1590/S0004-282X2003000200022
   Makary R, 2008, INTERNET J NEUROL, V11, P1
   McCrory PR, 1999, NEUROLOGY, V52, P1888, DOI 10.1212/WNL.52.9.1888
   Miller JH, 2016, TRANSLATIONAL RES AN, V5, P1, DOI [10.1016/j.tria.2016.09.001, DOI 10.1016/J.TRIA.2016.09.001]
   Odia Y, 2016, CLIN ONCOL, V1, P1
   Prayson RA, 2006, RUSSELL RUBINSTEINS, P321
   Sharma MS, 2005, J NEURO-ONCOL, V74, P325, DOI 10.1007/s11060-004-8266-3
   SOUTHALL DP, 1987, DEV MED CHILD NEUROL, V29, P789
   STEFANI F H, 1971, Archiv fuer Psychiatrie und Nervenkrankheiten, V214, P107, DOI 10.1007/BF00342909
   Takahashi M, 2016, LEGAL MED-TOKYO, V19, P119, DOI 10.1016/j.legalmed.2015.07.009
   Wakao N, 2012, NEURORADIOLOGY, V54, P893, DOI 10.1007/s00234-012-1041-9
   Zulch KJ, 1979, INT HISTOLOGICAL CLA
NR 17
TC 0
Z9 0
U1 0
U2 1
PU SPRINGER
PI NEW YORK
PA ONE NEW YORK PLAZA, SUITE 4600, NEW YORK, NY, UNITED STATES
SN 0256-7040
EI 1433-0350
J9 CHILD NERV SYST
JI Childs Nerv. Syst.
PD FEB
PY 2020
VL 36
IS 2
BP 447
EP 450
DI 10.1007/s00381-019-04392-w
EA OCT 2019
PG 4
WC Clinical Neurology; Pediatrics; Surgery
WE Science Citation Index Expanded (SCI-EXPANDED)
SC Neurosciences & Neurology; Pediatrics; Surgery
GA KI9HJ
UT WOS:000492339700002
PM 31650219
DA 2023-08-27
ER

PT J
AU Lee, TT
   Galarza, M
   Petito, CK
   Heros, RC
AF Lee, TT
   Galarza, M
   Petito, CK
   Heros, RC
TI Exophytic malignant brainstem mixed glioma in an adult: A case report
SO JOURNAL OF NEURO-ONCOLOGY
LA English
DT Article
DE anaplastic mixed glioma; brainstem glioma; posterior fossa tumor
ID STEM GLIOMA; INTRACRANIAL GLIOMAS; TUMORS; CHILDREN; DIAGNOSIS; THERAPY
AB Anaplastic mixed gliomas are rare tumors that occur mostly in the cerebral hemispheres. They have a distinctive histological appearance characterized by the presence of two or more glial cellular constituents. The incidence of malignant mixed glioma of the brainstem and posterior fossa is extremely low. The authors report an unusual case of an exophytic malignant mixed glioma. Following subtotal resection, the patient received conventional radiotherapy, but continued to deteriorate, and died five months after surgery. The extensive literature review focuses on histopathology, clinical features, natural history, and possible treatment modalities of this unusual neoplasm.
C1 Univ Miami, Sch Med, Dept Neurol Surg, Miami, FL 33101 USA.
   Univ Miami, Sch Med, Dept Pathol, Miami, FL 33101 USA.
C3 University of Miami; University of Miami
RP Lee, TT (通讯作者)，Univ Miami, Sch Med, Dept Neurol Surg, POB 016960 M813, Miami, FL 33101 USA.
CR ALBRIGHT AL, 1996, NEUROLOGICAL SURG, P2603
   APUZZO LJ, 1987, NEUROL RES, V9, P91
   BARKOVICH AJ, 1991, PEDIATR NEUROSURG, V16, P73, DOI 10.1159/000120511
   BIGNER SH, 1988, CANCER RES, V48, P405
   CHALLA VR, 1983, NEUROSURGERY, V12, P18, DOI 10.1227/00006123-198301000-00004
   DAVIS R, 1983, CLIN NEUROSCIENCES, P106
   DIRR LY, 1989, NEUROLOGY, V39, P865, DOI 10.1212/WNL.39.6.865
   EPSTEIN FJ, 1993, J NEUROSURG, V78, P408, DOI 10.3171/jns.1993.78.3.0408
   Franzini A, 1988, Acta Neurochir Suppl (Wien), V42, P170
   GEROSA MA, 1995, STEREOT FUNCT NEUROS, V64, P56, DOI 10.1159/000098764
   GRABB PA, 1992, NEUROSURGERY, V30, P64, DOI 10.1227/00006123-199201000-00012
   HART MN, 1974, CANCER, V33, P134, DOI 10.1002/1097-0142(197401)33:1<134::AID-CNCR2820330120>3.0.CO;2-I
   HELSETH A, 1989, APMIS, V97, P547, DOI 10.1111/j.1699-0463.1989.tb00829.x
   HOFMAN H, 1996, NEUROSURGERY, P1184
   KAWANO N, 1983, ACTA NEUROPATHOL, V62, P141, DOI 10.1007/BF00684931
   KEDAR A, 1994, MED PEDIATR ONCOL, V23, P428, DOI 10.1002/mpo.2950230507
   KURIMOTO M, 1987, Neurological Surgery, V15, P167
   KYRITSIS AP, 1993, NEUROSURGERY, V32, P365, DOI 10.1227/00006123-199303000-00005
   LANDY HJ, 1994, SOUTH MED J, V87, P211, DOI 10.1097/00007611-199402000-00013
   MARIA BL, 1993, J CHILD NEUROL, V8, P112, DOI 10.1177/088307389300800203
   MILLEN SJ, 1985, AM J OTOL, V6, P503
   MULLER PJ, 1995, SEMIN SURG ONCOL, V11, P346, DOI 10.1002/ssu.2980110504
   OSULLIVAN C, 1994, J NEUROSURG, V81, P507, DOI 10.3171/jns.1994.81.4.0507
   POLLACK IF, 1993, J NEUROSURG, V78, P859, DOI 10.3171/jns.1993.78.6.0859
   ROLLINS NK, 1995, PEDIATR NEUROSURG, V22, P8, DOI 10.1159/000121293
   SARKAR C, 1990, INDIAN J MED RES-B, V92, P324
   SARWAR M, 1977, RADIOLOGY, V124, P403, DOI 10.1148/124.2.403
   SHUANGSHOTI S, 1984, J NEUROSURG, V61, P777, DOI 10.3171/jns.1984.61.4.0777
   SHUANGSHOTI S, 1986, J SURG ONCOL, V31, P148, DOI 10.1002/jso.2930310215
   SPECHT CS, 1986, NEUROSURGERY, V19, P278, DOI 10.1227/00006123-198608000-00019
   STROINK AR, 1986, J NEUROSURG, V65, P745, DOI 10.3171/jns.1986.65.6.0745
   TOYURIKI Y, 1986, ACTANEUROCHIR, V79, P67
   VERTOSICK FT, 1992, SURG NEUROL, V38, P359, DOI 10.1016/0090-3019(92)90022-F
   YONG WH, 1995, J NEUROPATH EXP NEUR, V54, P622, DOI 10.1097/00005072-199509000-00002
NR 34
TC 6
Z9 6
U1 0
U2 0
PU KLUWER ACADEMIC PUBL
PI DORDRECHT
PA SPUIBOULEVARD 50, PO BOX 17, 3300 AA DORDRECHT, NETHERLANDS
SN 0167-594X
J9 J NEURO-ONCOL
JI J. Neuro-Oncol.
PD APR
PY 1998
VL 37
IS 2
BP 123
EP 129
DI 10.1023/A:1005969429013
PG 7
WC Oncology; Clinical Neurology
WE Science Citation Index Expanded (SCI-EXPANDED)
SC Oncology; Neurosciences & Neurology
GA ZB144
UT WOS:000072440800004
PM 9524090
DA 2023-08-27
ER

PT J
AU Lopez, JI
   Larrinaga, G
   Kuroda, N
   Angulo, JC
AF Lopez, Jose I.
   Larrinaga, Gorka
   Kuroda, Naoto
   Angulo, Javier C.
TI The normal and pathologic renal medulla: A comprehensive overview
SO PATHOLOGY RESEARCH AND PRACTICE
LA English
DT Review
DE Kidney; Renal medulla; Embryology; Anatomy; Physiology; Pathology
ID COLLECTING-DUCT CARCINOMA; ANGIOTENSIN-CONVERTING ENZYME; CELL
   CARCINOMA; TUBULOCYSTIC CARCINOMA; EPITHELIAL NEOPLASMS; KIDNEY;
   NEPHRON; TUMORS; DIFFERENTIATION; PHYSIOLOGY
AB The renal medulla comprises an intricate system of tubules, blood vessels and interstitium that is not well understood by most general pathologists. We conducted an extensive review of the literature on the renal medulla, in both normal and pathologic conditions. We set out in detail the points of key interest to pathologists: normal and pathological development, physiology, microscopic anatomy, histology and immunohistochemistry; and the specific and most common other types of disease associated with this part of the kidney: developmental abnormalities, (multicystic dysplastic kidney, autosomal dominant and recessive polycystic kidney diseases, medullary cystic kidney disease), inflammatory conditions (xanthogranulomatous pyelonephritis, malakoplakia), hyperplasia and dysplasia, and neoplastic processes (oncocytoma, atypical oncocytic tumors, chromophobe cell carcinoma, collecting duct carcinoma, urothelial carcinoma, other carcinomas, renal medullary fibroma and metastatic tumors). This condensed overview of the origin, function and pathology of the renal medulla, both in terms of development, inflammation and neoplastic processes, should help focus the interest of clinical pathologists on this widely overlooked part of the kidney. (C) 2014 Elsevier GmbH. All rights reserved.
C1 [Lopez, Jose I.] Univ Basque Country UPV EHU, Dept Anat Pathol, Cruces Univ Hosp, Biocruces Res Inst, Baracaldo, Bizkaia, Spain.
   [Larrinaga, Gorka] Univ Basque Country UPV EHU, Dept Physiol, Biocruces Res Inst, Leioa, Bizkaia, Spain.
   [Larrinaga, Gorka] Univ Basque Country UPV EHU, Dept Nursing, Biocruces Res Inst, Leioa, Bizkaia, Spain.
   [Kuroda, Naoto] Kochi Red Cross Hosp, Dept Diagnost Pathol, Kochi, Japan.
   [Angulo, Javier C.] European Univ Madrid, Getafe Univ Hosp, Dept Urol, Madrid, Spain.
C3 Hospital Universitario Cruces; University of Basque Country; University
   of Basque Country; University of Basque Country; European University of
   Madrid; Hospital Universitario de Getafe
RP Lopez, JI (通讯作者)，Hosp Univ Cruces, Serv Anat Patol, Plaza Cruces S-N, Baracaldo 48903, Bizkaia, Spain.
EM
RI Lopez, Jose Ignacio/L-3884-2019; Lopez, Jose Ignacio/E-2973-2012; Lopez,
   Jose/HZL-8909-2023
OI Lopez, Jose Ignacio/0000-0003-0842-5348; Lopez, Jose
   Ignacio/0000-0003-0842-5348; Angulo, Javier/0000-0002-1735-8792;
   LARRINAGA ENBEITA, GORKA/0000-0002-6744-818X
CR Amin MB, 2008, AM J SURG PATHOL, V32, P1822, DOI 10.1097/PAS.0b013e3181831e68
   [Anonymous], 2019, HISTOLOGY CELL BIOL
   [Anonymous], 1958, BORDERLAND EMBRYOLOG
   Aydin H, 2010, MODERN PATHOL, V23, p176A
   Balinski B. I., 1983, INTRO EMBRIOLOGIA
   BIGGAR WD, 1981, TRANSPLANTATION, V31, P109, DOI 10.1097/00007890-198102000-00003
   Blanco L, 2008, AM J PHYSIOL-RENAL, V294, pF850, DOI 10.1152/ajprenal.00469.2007
   BRACKEN RB, 1979, SOUTH MED J, V72, P806, DOI 10.1097/00007611-197907000-00013
   Cheng L, 2009, HUM PATHOL, V40, P10, DOI 10.1016/j.humpath.2008.09.009
   Clapp W. L., 2007, HISTOLOGY PATHOLOGIS, P839
   Danilczyk U, 2006, CIRC RES, V98, P463, DOI 10.1161/01.RES.0000205761.22353.5f
   De la Fuente IM, 2013, PLOS ONE, V8, DOI 10.1371/journal.pone.0058284
   De la Fuente IM, 2011, PLOS ONE, V6, DOI 10.1371/journal.pone.0027224
   Deshayes F, 2005, TRENDS ENDOCRIN MET, V16, P293, DOI 10.1016/j.tem.2005.07.009
   Donoghue M, 2000, CIRC RES, V87, pE1, DOI 10.1161/01.res.87.5.e1
   Farreras-Rozman F.P., 2009, MED INTERNA
   Hara N, 2004, INT J UROL, V11, P68, DOI 10.1111/j.1442-2042.2004.00754.x
   Herring JC, 1997, J UROLOGY, V157, P2246, DOI 10.1016/S0022-5347(01)64734-7
   Hes O, 2004, Cas Lek Cesk, V143, P169
   Holmes RP, 2012, MOL ASPECTS MED, V33, P547, DOI 10.1016/j.mam.2012.01.001
   Holthofer H, 1983, LAB INVEST, V49, P317
   Janech MG, 2007, AM J PHYSIOL-RENAL, V292, pF501, DOI 10.1152/ajprenal.00298.2006
   Kaissling B, 2008, HISTOCHEM CELL BIOL, V130, P247, DOI 10.1007/s00418-008-0452-5
   Kobayashi N, 2008, HUM PATHOL, V39, P1350, DOI 10.1016/j.humpath.2007.11.020
   Kobori H, 2007, PHARMACOL REV, V59, P251, DOI 10.1124/pr.59.3.3
   Koeppen B, 2009, BERNE LEVI FISIOLOGI
   Kumar V., 2008, ROBBINS PATOLOGIA HU
   Kuroda N, 2014, POL J PATHOL, V65, P93, DOI 10.5114/PJP.2014.43958
   Kuroda N, 2012, HISTOL HISTOPATHOL, V27, P1407, DOI 10.14670/HH-27.1407
   Kuroda N, 2011, PATHOL INT, V61, P552, DOI 10.1111/j.1440-1827.2011.02689.x
   Lacson A., 2007, POTTERS PATHOLOGY FE, V2, P1281
   Larrinaga G, 2007, REGUL PEPTIDES, V144, P56, DOI 10.1016/j.regpep.2007.06.002
   Larrinaga G, 2013, CLIN BIOCHEM, V46, P638, DOI 10.1016/j.clinbiochem.2012.12.023
   Lely AT, 2004, J PATHOL, V204, P587, DOI 10.1002/path.1670
   Leroy X, 2002, HUM PATHOL, V33, P574, DOI 10.1053/hupa.2002.124010
   Li M, 2001, MODERN PATHOL, V14, P623, DOI 10.1038/modpathol.3880361
   Lindgren V, 2004, ARCH PATHOL LAB MED, V128, P1274
   LOPEZ JI, 1991, TUMORI, V77, P358, DOI 10.1177/030089169107700414
   Lopez JI, 2013, PATHOL RES PRACT, V209, P137, DOI 10.1016/j.prp.2013.01.007
   Lopez-Beltran A, 2006, EUR UROL, V49, P798, DOI 10.1016/j.eururo.2005.11.035
   MacLennan GT, 1997, UROLOGY, V50, P679, DOI 10.1016/S0090-4295(97)00335-X
   Maestroni U, 2006, TUMORI, V92, P545, DOI 10.1177/030089160609200615
   MALEK RS, 1978, J UROLOGY, V119, P589, DOI 10.1016/S0022-5347(17)57559-X
   Metzger R, 1999, KIDNEY INT, V56, P1442, DOI 10.1046/j.1523-1755.1999.00660.x
   Michal M, 1998, Ann Diagn Pathol, V2, P149, DOI 10.1016/S1092-9134(98)80001-4
   Mizuiri S, 2008, AM J KIDNEY DIS, V51, P613, DOI 10.1053/j.ajkd.2007.11.022
   Nagashima Y, 2000, PATHOL RES PRACT, V196, P647, DOI 10.1016/S0344-0338(00)80008-4
   Nese N, 2009, ANN DIAGN PATHOL, V13, P1, DOI 10.1016/j.anndiagpath.2008.10.002
   Orsola A, 2005, UROLOGY, V65, P49, DOI 10.1016/j.urology.2004.08.012
   Otani M, 2001, PATHOL INT, V51, P954, DOI 10.1046/j.1440-1827.2001.01297.x
   Parwani AV, 2001, HUM PATHOL, V32, P506, DOI 10.1053/hupa.2001.24320
   Paul M, 2006, PHYSIOL REV, V86, P747, DOI 10.1152/physrev.00036.2005
   Petersson F, 2014, VIRCHOWS ARCH, V465, P215, DOI 10.1007/s00428-014-1600-z
   Petersson F, 2010, INT J CLIN EXP PATHO, V3, P534
   Pote N, 2013, VIRCHOWS ARCH, V462, P633, DOI 10.1007/s00428-013-1422-4
   Rakozy C, 2002, MODERN PATHOL, V15, P1162, DOI 10.1097/01.MP.0000031709.40712.46
   Sands JM, 2009, SEMIN NEPHROL, V29, P178, DOI 10.1016/j.semnephrol.2009.03.008
   THOENES W, 1988, J PATHOL, V155, P277, DOI 10.1002/path.1711550402
   THOENES W, 1985, VIRCHOWS ARCH B, V48, P207, DOI 10.1007/BF02890129
   Tipnis SR, 2000, J BIOL CHEM, V275, P33238, DOI 10.1074/jbc.M002615200
   Tokuda N, 2006, J UROLOGY, V176, P40, DOI 10.1016/S0022-5347(06)00502-7
   Trivedi PP, 2013, INT J SURG PATHOL, V21, P615, DOI 10.1177/1066896913491322
   Ugalde A, 2008, ACTAS UROL ESP, V32, P799, DOI 10.1016/S0210-4806(08)73938-X
   Varona A, 2007, AM J PHYSIOL-RENAL, V292, pF780, DOI 10.1152/ajprenal.00148.2006
   Williams P. L, 1996, ANATOMIA DE GRAY, V2
   Williams PA, 2013, PATHOL RES PRACT, V209, P654, DOI 10.1016/j.prp.2013.07.007
   Yang XJ, 2008, AM J SURG PATHOL, V32, P177, DOI 10.1097/PAS.0b013e318150df1d
   Zhou M, 2009, AM J SURG PATHOL, V33, P1840, DOI 10.1097/PAS.0b013e3181be22d1
NR 68
TC 5
Z9 5
U1 0
U2 10
PU ELSEVIER GMBH
PI MUNICH
PA HACKERBRUCKE 6, 80335 MUNICH, GERMANY
SN 0344-0338
J9 PATHOL RES PRACT
JI Pathol. Res. Pract.
PY 2015
VL 211
IS 4
BP 271
EP 280
DI 10.1016/j.prp.2014.12.009
PG 10
WC Pathology
WE Science Citation Index Expanded (SCI-EXPANDED)
SC Pathology
GA CG5DS
UT WOS:000353311900001
PM 25595996
DA 2023-08-27
ER

PT J
AU Chastkofsky, MI
   Pituch, KC
   Katagi, H
   Zannikou, M
   Ilut, L
   Xiao, T
   Han, Y
   Sonabend, AM
   Curiel, DT
   Bonner, ER
   Nazarian, J
   Horbinski, CM
   James, CD
   Saratsis, AM
   Hashizume, R
   Lesniak, MS
   Balyasnikova, IV
AF Chastkofsky, Michael, I
   Pituch, Katarzyna C.
   Katagi, Hiroaki
   Zannikou, Markella
   Ilut, Liliana
   Xiao, Ting
   Han, Yu
   Sonabend, Adam M.
   Curiel, David T.
   Bonner, Erin R.
   Nazarian, Javad
   Horbinski, Craig M.
   James, C. David
   Saratsis, Amanda M.
   Hashizume, Rintaro
   Lesniak, Maciej S.
   Balyasnikova, Irina, V
TI Mesenchymal Stem Cells Successfully Deliver Oncolytic Virotherapy to
   Diffuse Intrinsic Pontine Glioma
SO CLINICAL CANCER RESEARCH
LA English
DT Article
ID INTRANASAL DELIVERY; EXTENDS SURVIVAL; BRAIN-TUMORS; ADENOVIRUS;
   EXPRESSION; MUTATIONS; THERAPY; MODEL; REIRRADIATION; MODULATION
AB Purpose: Diffuse intrinsic pontine glioma (DIPG) is among the deadliest of pediatric brain tumors. Radiotherapy is the standard-of-care treatment for DIPG, but offers only transient relief of symptoms for patients with DIPG without providing significant survival benefit. Oncolytic virotherapy is an anticancer treatment that has been investigated for treating various types of brain tumors.
   Experimental Design: Here, we have explored the use of mesenchymal stem cells (MSC) for oncolytic virus (OV) delivery and evaluated treatment efficacy using preclinical models of DIPG. The survivin promoter drives the conditional replication of OV used in our studies. The efficiency of OV entry into the cells is mediated by fiber modification with seven lysine residues (CRAd.S.pK7). Patients' samples and cell lines were analyzed for the expression of viral entry proteins and survivin. The ability of MSCs to deliver OV to DIPG was studied in the context of a low dose of irradiation.
   Results: Our results show that DIPG cells and tumors exhibit robust expression of cell surface proteins and survivin that enable efficient OV entry and replication in DIPG cells. MSCs loaded with OV disseminate within a tumor and release OV throughout the DIPG brainstem xenografts in mice. Administration of OV-loaded MSCs with radiotherapy to mice bearing brainstem DIPG xenografts results in more prolonged survival relative to that conferred by either therapy alone (P < 0.01).
   Conclusions: Our study supports OV, CRAd.S.pK7, encapsulated within MSCs as a therapeutic strategy that merits further investigation and potential translation for DIPG treatment.
C1 [Chastkofsky, Michael, I; Pituch, Katarzyna C.; Katagi, Hiroaki; Zannikou, Markella; Ilut, Liliana; Xiao, Ting; Han, Yu; Sonabend, Adam M.; Horbinski, Craig M.; James, C. David; Saratsis, Amanda M.; Hashizume, Rintaro; Lesniak, Maciej S.; Balyasnikova, Irina, V] Northwestern Univ, Feinberg Sch Med, Dept Neurol Surg, Chicago, IL 60611 USA.
   [Curiel, David T.] Washington Univ, Dept Radiat Oncol, St Louis, MO USA.
   [Bonner, Erin R.; Nazarian, Javad] Childrens Natl Med Ctr, Ctr Genom & Precis Med, Washington, DC 20010 USA.
   [Bonner, Erin R.] George Washington Univ, Sch Med & Hlth Sci, Inst Biomed Sci, Washington, DC 20052 USA.
   [Nazarian, Javad] George Washington Univ, Sch Med & Hlth Sci, Dept Integrat Syst Biol, Washington, DC 20052 USA.
   [Horbinski, Craig M.] Northwestern Univ, Dept Pathol, Feinberg Sch Med, Chicago, IL 60611 USA.
   [Saratsis, Amanda M.] Ann & Robert H Lurie Childrens Hosp Chicago, Div Neurosurg, Dept Pediat Surg, Chicago, IL USA.
   [Saratsis, Amanda M.; Hashizume, Rintaro] Northwestern Univ, Feinberg Sch Med, Dept Biochem & Mol Genet, Chicago, IL 60611 USA.
C3 Northwestern University; Feinberg School of Medicine; Washington
   University (WUSTL); Children's National Health System; George Washington
   University; George Washington University; Northwestern University;
   Feinberg School of Medicine; Ann & Robert H. Lurie Children's Hospital
   of Chicago; Northwestern University; Feinberg School of Medicine
RP Balyasnikova, IV (通讯作者)，Northwestern Univ, Feinberg Sch Med, 303 E Super St,SQ Room 6-520, Chicago, IL 60611 USA.
EM
RI Han, Yu/GZA-9220-2022
OI Katagi, Hiroaki/0000-0003-3620-9116; Sonabend, Adam
   M/0000-0002-8347-1945; XIAO, TING/0000-0002-7502-4395
FU NIH [R01NS087990, R01NS106379, R33 NS101150, P50CA221747]; ORIP of the
   NIH [P40OD011050]; Northwestern University Flow Cytometry Core Facility
   - NCI [P30-CA060553]; Center for Advanced Microscopy - NCI
   [P30-CA060553]; Mouse Histology and Phenotyping Laboratory - NCI
   [P30-CA060553]
FX This work was supported by NIH R01NS087990, R01NS106379, R33 NS101150,
   and P50CA221747 SPORE for Translational Approaches to Brain Cancer. We
   are also grateful to the Nora Redman Endowment Fund of the Community
   Foundation of Louisville, the IDP Foundation Inc., and to the Isabella
   Kerr Molina Foundation for generous gifts to support the study. Some of
   the materials employed in this work were provided by the Texas A&M
   Health Science Center College of Medicine Institute for Regenerative
   Medicine at Scott & White through a grant from ORIP of the NIH, grant
   no., P40OD011050. This work was also supported by the Northwestern
   University Flow Cytometry Core Facility, the Center for Advanced
   Microscopy, and the Mouse Histology and Phenotyping Laboratory, all
   supported by NCI P30-CA060553 awarded to the Robert H Lurie
   Comprehensive Cancer Center. The authors are also grateful to the
   Pathology Core Facility of the Robert H Lurie Comprehensive Cancer
   Center for its excellent service.
CR Ahmed AU, 2013, JNCI-J NATL CANCER I, V105, P968, DOI 10.1093/jnci/djt141
   Ahmed AU, 2011, MOL PHARMACEUT, V8, P1559, DOI 10.1021/mp200161f
   Ahmed AU, 2010, MOL THER, V18, P1846, DOI 10.1038/mt.2010.131
   Altieri DC, 1999, LAB INVEST, V79, P1327
   Balyasnikova IV, 2014, MOL THER, V22, P140, DOI 10.1038/mt.2013.199
   BERGER MS, 1983, NEUROSURGERY, V12, P298, DOI 10.1227/00006123-198303000-00008
   Bonora M, 2015, ONCOGENE, V34, P1608, DOI [10.1038/onc.2014.462, 10.1038/onc.2014.96]
   Bowman RL, 2017, NEURO-ONCOLOGY, V19, P139, DOI 10.1093/neuonc/now247
   Breitbach CJ, 2007, MOL THER, V15, P1686, DOI 10.1038/sj.mt.6300215
   Cerami E, 2012, CANCER DISCOV, V2, P401, DOI 10.1158/2159-8290.CD-12-0095
   Choi SA, 2012, EUR J CANCER, V48, P129, DOI 10.1016/j.ejca.2011.04.033
   Danielyan L, 2011, REJUV RES, V14, P3, DOI 10.1089/rej.2010.1130
   Danielyan L, 2009, EUR J CELL BIOL, V88, P315, DOI 10.1016/j.ejcb.2009.02.001
   Dey M, 2016, STEM CELL REP, V7, P471, DOI 10.1016/j.stemcr.2016.07.024
   Evgin L, 2016, MOL THER-ONCOLYTICS, V3, DOI 10.1038/mto.2016.27
   Fontanilla HP, 2012, AM J CLIN ONCOL-CANC, V35, P51, DOI 10.1097/COC.0b013e318201a2b7
   Freese C, 2017, PRACT RADIAT ONCOL, V7, P86, DOI 10.1016/j.prro.2016.11.005
   Fulci G, 2006, P NATL ACAD SCI USA, V103, P12873, DOI 10.1073/pnas.0605496103
   Gao JJ, 2013, SCI SIGNAL, V6, DOI 10.1126/scisignal.2004088
   Gwak HS, 2017, CRIT REV ONCOL HEMAT, V120, P111, DOI 10.1016/j.critrevonc.2017.10.013
   Hammer K, 2015, INT J CANCER, V137, P978, DOI 10.1002/ijc.29442
   Han JF, 2015, CANCER RES, V75, P5273, DOI 10.1158/0008-5472.CAN-15-0894
   Hashizume R, 2008, NEURO-ONCOLOGY, V10, P112, DOI 10.1215/15228517-2007-052
   Hashizume R, 2012, J NEURO-ONCOL, V110, P305, DOI 10.1007/s11060-012-0973-6
   Hashizume R, 2010, J NEURO-ONCOL, V96, P151, DOI 10.1007/s11060-009-9954-9
   Hendricks BK, 2015, NEUROSURG FOCUS, V38, DOI 10.3171/2015.1.FOCUS14767
   Huang TY, 2017, ACTA NEUROPATHOL COM, V5, DOI 10.1186/s40478-017-0436-6
   Jansen MHA, 2012, CANCER TREAT REV, V38, P27, DOI 10.1016/j.ctrv.2011.06.007
   Jones C, 2017, NEURO-ONCOLOGY, V19, P153, DOI 10.1093/neuonc/now101
   Jung BK, 2017, BIOMATERIALS, V147, P26, DOI 10.1016/j.biomaterials.2017.09.009
   Kurozumi K, 2007, JNCI-J NATL CANCER I, V99, P1768, DOI 10.1093/jnci/djm229
   Layek B, 2018, MOL CANCER THER, V17, P1196, DOI 10.1158/1535-7163.MCT-17-0682
   Moyano AL, 2018, MOL THER, V26, P730, DOI 10.1016/j.ymthe.2018.01.008
   Martinez-Velez N, 2019, NAT COMMUN, V10, DOI 10.1038/s41467-019-10043-0
   Martinez-Velez N, 2019, ACTA NEUROPATHOL COM, V7, DOI 10.1186/s40478-019-0714-6
   Meel MH, 2019, DRUG RESIST UPDATE, V44, P15, DOI 10.1016/j.drup.2019.06.001
   Mendes D, 2018, NEUROLOGIST, V23, P141, DOI 10.1097/NRL.0000000000000188
   Mendez F, 2020, CLIN CANCER RES, V26, P4080, DOI 10.1158/1078-0432.CCR-19-3714
   Morshed RA, 2015, CANCER GENE THER, V22, P55, DOI 10.1038/cgt.2014.72
   Parker Kerrigan Brittany C, 2018, Prog Neurol Surg, V32, P124, DOI 10.1159/000469686
   Paugh BS, 2013, CANCER RES, V73, P6219, DOI 10.1158/0008-5472.CAN-13-1491
   Paugh BS, 2010, J CLIN ONCOL, V28, P3061, DOI 10.1200/JCO.2009.26.7252
   Qi J, 2019, ACTA NEUROPATHOL COM, V7, DOI 10.1186/s40478-019-0727-1
   Ribas A, 2017, CELL, V170, P1109, DOI 10.1016/j.cell.2017.08.027
   Ruano D, 2020, MOL THER, V28, P1033, DOI 10.1016/j.ymthe.2020.01.019
   Saratsis AM, 2014, ACTA NEUROPATHOL, V127, P881, DOI 10.1007/s00401-013-1218-2
   Sonabend AM, 2009, CANCER GENE THER, V16, P362, DOI 10.1038/cgt.2008.80
   Sonabend AM, 2008, STEM CELLS, V26, P831, DOI 10.1634/stemcells.2007-0758
   Spencer D, 2019, THERANOSTICS, V9, P2071, DOI 10.7150/thno.29581
   Takai N, 2002, INT J MOL MED, V10, P211
   Takami H, 2015, BRAIN PATHOL, V25, P256, DOI 10.1111/bpa.12173
   Timin AS, 2019, ACS APPL MATER INTER, V11, P13091, DOI 10.1021/acsami.8b22685
   Ulasov IV, 2007, HUM GENE THER, V18, P589, DOI 10.1089/hum.2007.002
   van Houdt WJ, 2006, J NEUROSURG, V104, P583, DOI 10.3171/jns.2006.104.4.583
   Wang XL, 2018, INT J NANOMED, V13, P5231, DOI 10.2147/IJN.S167142
   Wu G, 2012, NAT GENET, V44, P251, DOI 10.1038/ng.1102
   Yong RL, 2009, CANCER RES, V69, P8932, DOI 10.1158/0008-5472.CAN-08-3873
   Yoo JY, 2019, NEURO-ONCOLOGY, V21, P1131, DOI 10.1093/neuonc/noz079
   Young JS, 2014, EXPERT OPIN DRUG DEL, V11, P1733, DOI 10.1517/17425247.2014.937420
   Zhao DH, 2008, MOL CANCER RES, V6, P1819, DOI 10.1158/1541-7786.MCR-08-0146
NR 60
TC 28
Z9 30
U1 1
U2 7
PU AMER ASSOC CANCER RESEARCH
PI PHILADELPHIA
PA 615 CHESTNUT ST, 17TH FLOOR, PHILADELPHIA, PA 19106-4404 USA
SN 1078-0432
EI 1557-3265
J9 CLIN CANCER RES
JI Clin. Cancer Res.
PD MAR 15
PY 2021
VL 27
IS 6
BP 1766
EP 1777
DI 10.1158/1078-0432.CCR-20-1499
PG 12
WC Oncology
WE Science Citation Index Expanded (SCI-EXPANDED)
SC Oncology
GA QY6IV
UT WOS:000630142100018
PM 33272983
OA Green Accepted, Bronze, Green Submitted
DA 2023-08-27
ER

PT J
AU Clymer, J
   Kieran, MW
AF Clymer, Jessica
   Kieran, Mark W.
TI The Integration of Biology Into the Treatment of Diffuse Intrinsic
   Pontine Glioma: A Review of the North American Clinical Trial
   Perspective
SO FRONTIERS IN ONCOLOGY
LA English
DT Review
DE diffuse intrinsic pontine gliomas; brainstem glioma; targeted therapy;
   convection-enhanced delivery; immunotherapy; clinical trials
ID TEMOZOLOMIDE
AB Dramatic advances in the molecular analysis of diffuse intrinsic pontine glioma have occurred over the last decade and resulted in the identification of potential therapeutic targets. In spite of these advances, no significant improvement in the outcome has been achieved and median survival remains approximately 10 months. An understanding of the approaches that have been taken to date, why they failed, and how that information can lead the field forward is critical if we are to change the status quo. In this review, we will discuss the clinical trial landscape in North America with an overview of historical approaches that failed and what might account for this failure. We will then provide a discussion of how our understanding of the genotype of this disease has led to the development of a number of trials targeting the mutations and epigenome of diffuse intrinsic pontine gliomas and the issues related to these trials. Similarly, the introduction of methodologies to address penetration across the blood-brain barrier will be considered in the context of both targeted approaches, epigenetic modification, and immune surveillance of these tumors. The comprehensive analysis of these data, generated through cooperative groups, collaborative clinical trials, and pilot studies in North America will be the focus of the IVth Memorial Alicia Pueyo international symposium in Barcelona on March 12th, 2018 and will be compared and contrasted with a similar comprehensive analysis of the European data with the goal of bringing all of these data together to develop a uniform platform on which new rational trials can be based.
C1 [Clymer, Jessica; Kieran, Mark W.] Dana Farber Canc Inst, Dept Pediat Oncol, Boston, MA 02115 USA.
   [Clymer, Jessica; Kieran, Mark W.] Boston Childrens Hosp, Div Pediat Hematol Oncol, Boston, MA 02115 USA.
   [Kieran, Mark W.] Harvard Med Sch, Boston, MA USA.
C3 Harvard University; Dana-Farber Cancer Institute; Harvard University;
   Boston Children's Hospital; Harvard University; Harvard Medical School
RP Clymer, J (通讯作者)，Dana Farber Canc Inst, Dept Pediat Oncol, Boston, MA 02115 USA.; Clymer, J (通讯作者)，Boston Childrens Hosp, Div Pediat Hematol Oncol, Boston, MA 02115 USA.
EM
FU Ellie Kavalieros DIPG Fund
FX This article is funded by Ellie Kavalieros DIPG Fund.
CR Aguilera TA, 2015, INT J RADIAT ONCOL, V91, P895, DOI 10.1016/j.ijrobp.2015.01.041
   Barua NU, 2013, ACTA NEUROCHIR WIEN, V155, P1459, DOI 10.1007/s00701-013-1700-6
   Bobo RH, 1994, P NATL ACAD SCI USA, V91, P2076, DOI 10.1073/pnas.91.6.2076
   Bouffet E, 2000, CANCER, V88, P685, DOI 10.1002/(SICI)1097-0142(20000201)88:3<685::AID-CNCR27>3.0.CO;2-K
   Cage TA, 2013, CHILDS NERV SYST, V29, P1313, DOI 10.1007/s00381-013-2101-0
   Castel D, 2015, ACTA NEUROPATHOL, V130, P815, DOI 10.1007/s00401-015-1478-0
   Chassot A, 2012, J NEUROONCOL, V106, P399, DOI 10.1007/s11060-011-0681-7
   Chittiboina P, 2013, J NEUROSURG-PEDIATR, V11, P37, DOI 10.3171/2012.8.PEDS11296
   Cohen KJ, 2011, NEUROONCOL, V13, P410, DOI 10.1093/neuonc/noq205
   EDWARDS MSB, 1989, J NEUROSURG, V70, P691, DOI 10.3171/jns.1989.70.5.0691
   Ellis JA, 2012, J NEUROSURG-PEDIATR, V10, P14, DOI 10.3171/2012.3.PEDS11228
   Freeman CR, 1988, INT J RADIAT ONCOL, V15, P311, DOI 10.1016/S0360-3016(98)90010-4
   Freeman CR, 1991, CANCER, V68, P474, DOI 10.1002/1097-0142(19910801)68:3<474
   Geyer MB, 2016, CYTOTHERAPY, V18, P1393, DOI 10.1016/j.jcyt.2016.07.003
   Gobin YP, 2001, RADIOLOGY, V218, P724, DOI 10.1148/radiology.218.3.r01mr41724
   Goodwin CR, 2016, CLIN NEUROL NEUROSUR, V142, P120, DOI 10.1016/j.clineuro.2016.01.007
   Grasso CS, 2015, NAT MED, V21, P555, DOI 10.1038/nm.3855
   Grimm SA, 2013, CURR NEUROL NEUROSCI, V13, P346, DOI 10.1007/s11910-013-0346-3
   Gupta N, 2017, NEURO ONCOL S6, V19, pvi188, DOI 10.1093/neuonc/nox168.762
   Gururangan S, 2010, J CLIN ONCOL, V28, P3069, DOI 10.1200/JCO.2009.26.8789
   Hamisch C, 2017, J NEUROSURG-PEDIATR, V20, P261, DOI 10.3171/2017.2.PEDS1665
   Hao C, 2017, MEDICINE BALTIMORE, V96, pe7325, DOI 10.1097/MD.0000000000007325
   Hargrave D, 2006, LANCET ONCOL, V7, P241, DOI 10.1016/S1470-2045(06)70615-5
   Hennika T, 2017, PLOS ONE, V12, DOI 10.1371/journal.pone.0169485
   Hu Z, 2017, NAT REV IMMUNOL, V18, P168, DOI 10.1038/nri.2017.131
   Hummel TR, 2016, J NEUROONCOL, V127, P53, DOI 10.1007/s11060-015-2008-6
   Janssens GO, 2013, INT J RADIAT ONCOL, V85, P315, DOI 10.1016/j.ijrobp.2012.04.006
   Janssens GO, 2009, INT J RADIAT ONCOL, V73, P722, DOI 10.1016/j.ijrobp.2008.05.030
   Khuong-Quang DA, 2012, ACTA NEUROPATHOL, V124, P439, DOI 10.1007/s00401-012-0998-0
   Killick-Cole CL, 2017, PLOS ONE, V12, DOI 10.1371/journal.pone.0176855
   Lakin N, 2017, FRONT ONCOL, V7, DOI 10.3389/fonc.2017.00141
   Lewis PW, 2013, SCIENCE, V340, P857, DOI 10.1126/science.1232245
   Lonser RR, 2007, J NEUROSURG, V107, P190, DOI 10.3171/JNS-07/07/0190
   Louis DN, 2016, ACTA NEUROPATHOL, V131, P803, DOI 10.1007/s00401-016-1545-1
   Marcus KJ, 2003, INT J RADIAT ONCOL, V55, P1182, DOI 10.1016/S0360-3016(02)04391-2
   Negretti L, 2011, J NEUROONCOL, V104, P773, DOI 10.1007/s11060-011-0542-4
   Neuwelt EA, 2006, PEDIAT BLOOD CANC, V47, P174, DOI 10.1002/pbc.20529
   Neuwelt EA, 1985, NEUROSURGERY, V17, P419, DOI 10.1227/00006123-198509000-00004
   Packer RJ, 1994, CANCER, V74, P1827, DOI 10.1002/1097-0142(19940915)74:6<1827::AID-CNCR2820740628>3.0.CO;2-Q
   Packer RJ, 1987, INT J RADIAT ONCOL, V13, P1647, DOI 10.1016/0360-3016(87)90160-X
   Poussaint TY, 2011, NEUROONCOL, V13, P417, DOI 10.1093/neuonc/noq200
   Puget S, 2015, CHILDS NERV SYST, V31, P1773, DOI 10.1007/s00381-015-2832-1
   Rajappa P, 2011, INTERV NEURORADIOL, V17, P459, DOI 10.1177/159101991101700410
   Riina HA, 2010, INTERV NEURORADIOL, V16, P71, DOI 10.1177/159101991001600109
   Sandberg DI, 2002, J NEUROSURG, V96, P885, DOI 10.3171/jns.2002.96.5.0885
   Sayour EJ, 2017, BRAIN SCI, V7, pE137, DOI 10.3390/brainsci7100137
   Schwartzentruber J, 2012, NATURE, V482, P226, DOI 10.1038/nature10833
   Sharp JR, 2010, EUR J CANCER, V46, P3271, DOI 10.1016/j.ejca.2010.06.115
   Vanan MI, 2015, FRONT ONCOL, V5, P237, DOI 10.3389/fonc.2015.00237
   Warren KE, 2013, CNS ONCOL, V2, P427, DOI 10.2217/cns.13.37
   Wu G, 2012, NAT GENET, V44, P251, DOI 10.1038/ng.1102
   Zaky W, 2013, PEDIATR HEMAT ONCOL, V30, P623, DOI 10.3109/08880018.2013.829895
NR 52
TC 11
Z9 12
U1 0
U2 2
PU FRONTIERS MEDIA SA
PI LAUSANNE
PA AVENUE DU TRIBUNAL FEDERAL 34, LAUSANNE, CH-1015, SWITZERLAND
SN 2234-943X
J9 FRONT ONCOL
JI Front. Oncol.
PD MAY 18
PY 2018
VL 8
AR 169
DI 10.3389/fonc.2018.00169
PG 8
WC Oncology
WE Science Citation Index Expanded (SCI-EXPANDED)
SC Oncology
GA GG3FU
UT WOS:000432578500001
PM 29868485
OA Green Published, gold
DA 2023-08-27
ER

PT J
AU Szmuda, T
   Sloniewski, P
   Ali, S
   Pereira, PMG
   Pacholski, M
   Timemy, F
   Sabisz, A
   Szurowska, E
   Kieronska, S
AF Szmuda, Tomasz
   Sloniewski, Pawel
   Ali, Shan
   Goncalves Pereira, Pedro M.
   Pacholski, Mateusz
   Timemy, Fanar
   Sabisz, Agnieszka
   Szurowska, Edyta
   Kieronska, Sara
TI Reliability of diffusion tensor tractography of facial nerve in
   cerebello-pontine angle tumours
SO NEUROLOGIA I NEUROCHIRURGIA POLSKA
LA English
DT Article
DE cerebello-pontine angle; facial nerve; magnetic resonance imaging;
   diffusion tensor imaging; vestibular schwannoma
ID VESTIBULAR SCHWANNOMA; PREOPERATIVE IDENTIFICATION; IMAGING
   TRACTOGRAPHY; CRANIAL NERVES; PREDICTION; SURGERY; PRESERVATION;
   POSITION
AB Aim of the study. This study aimed to verify the accuracy of preoperative visualisation of the facial nerve (FN) by magnetic resonance-based (MR) diffusion tensor imaging-fibre tracking (DTI-FT) with neuronavigation system integration in patients with cerebello-pontine angle (CPA) tumours.
   Clinical rationale for the study. Complete excision with preservation of the FN remains the critical goal of today's vestibular schwannoma (VS) surgery. DTI-FT of the FN with neuronavigation is yet to be fully evaluated, and could make surgery safer.
   Materials and methods. This was a prospective cohort study in which 38 consecutive patients with a CPA tumour (32 VSs, five meningiomas and one epidermoid cyst) were operated on via the retrosigmoid route from 2013 to 2019. The course of the FN was simulated before surgery using StealthViz and the images were transferred to the Medtronic S7 neuronavigation system. The FN location reconstructed by DTI-FT was verified during the surgery.
   Results. MR acquisition was inappropriate in three patients (7.9%). DTI-FT correctly predicted the course of the FN in 31 of the 38 patients; the discordance rate was 18.4%. The accuracy of DTI-FT was 81.6% (95% CI: 65.67-92.26), sensitivity 88.57% (95%CI: 73.26-96.80) and positive predictive value was 91.18% (95% CI: 90.17-92.09). The reliability of the neuronavigation-integrated visualisation of the FN did not depend on the tumour size (p = 0.85), but the method was more accurate when the nerve was compact in shape (p = 0.03, area under curve (AUC) 0.87, 95% CI: 0.60-1.00) and in females (p = 0.04, AUC 0.78, 95% CI: 0.56-1.00). Following surgery, 86.5% ofthe patients presented with useful facial function (House-Brackmann grades I-III). Correct simulation of the FN did not prevent postoperative facial palsy (p = 0.35).
   Conclusions. The accuracy of DTI-FT of the FN integrated with neuronavigation remains unsatisfactory. This method does not provide any clinical benefit over non-integrated DTI-FT in terms of nerve function preservation.
   Clinical implications. Due to the low reliability of the predictions, further technical advances in predicting the course of the FN are awaited by clinicians. However, DTI-FT images in the operating theatre would make tumour excision more comfortable for the surgeon.
C1 [Szmuda, Tomasz; Sloniewski, Pawel; Kieronska, Sara] Med Univ Gdansk, Neurosurg Dept, Debinki 7 Str, PL-80952 Gdansk, Poland.
   [Ali, Shan; Pacholski, Mateusz; Timemy, Fanar] Med Univ Gdansk, Students Sci Circle Neurol & Neurosurg, Gdansk, Poland.
   [Goncalves Pereira, Pedro M.] Univ Algarve, Algarve Biomed Ctr, Faro, Portugal.
   [Sabisz, Agnieszka; Szurowska, Edyta] Med Univ Gdansk, Radiol Dept, Gdansk, Poland.
C3 Fahrenheit Universities; Medical University Gdansk; Fahrenheit
   Universities; Medical University Gdansk; Universidade do Algarve;
   Fahrenheit Universities; Medical University Gdansk
RP Szmuda, T (通讯作者)，Med Univ Gdansk, Neurosurg Dept, Debinki 7 Str, PL-80952 Gdansk, Poland.
EM
RI Kierońska-Siwak, Sara/HNP-2184-2023; Ali, Shan/AAC-3931-2020; Pereira,
   Pedro Miguel M.S. Gonçalves/AAG-5903-2020
OI Kierońska-Siwak, Sara/0000-0003-0290-0822; Ali,
   Shan/0000-0002-0005-8830; Goncalves Pereira, Pedro
   M./0000-0002-5369-489X; Szurowska, Edyta/0000-0002-7042-4381;
   Sloniewski, Pawel/0000-0001-8295-2336; Sabisz,
   Agnieszka/0000-0002-6653-5425
CR Baro V, 2019, WORLD NEUROSURG, V125, P24, DOI 10.1016/j.wneu.2019.01.099
   Borkar SA, 2016, NEUROL INDIA, V64, P965, DOI 10.4103/0028-3886.190270
   Breun M, 2019, WORLD NEUROSURG, V129, pE114, DOI 10.1016/j.wneu.2019.05.056
   Choi KS, 2014, J KOREAN NEUROSURG S, V56, P11, DOI 10.3340/jkns.2014.56.1.11
   Churi ON, 2019, WORLD NEUROSURG, V127, pE509, DOI 10.1016/j.wneu.2019.03.190
   Gerganov VM, 2011, J NEUROSURG, V115, P1087, DOI 10.3171/2011.7.JNS11495
   Jacquesson T, 2019, NEUROSURGERY, V84, P313, DOI 10.1093/neuros/nyy229
   Jacquesson T, 2018, WORLD NEUROSURG, V117, P363, DOI 10.1016/j.wneu.2018.06.182
   Koos WT, 1998, J NEUROSURG, V88, P506, DOI 10.3171/jns.1998.88.3.0506
   Li H, 2017, WORLD NEUROSURG, V107, P669, DOI 10.1016/j.wneu.2017.08.048
   Pereira P, 2019, J NUCL MED RAD THERA, V10, DOI [10.4172/2155-9619.1000396, DOI 10.4172/2155-9619.1000396]
   Rinaldi V, 2012, J NEUROL SURG PART B, V73, P21, DOI 10.1055/s-0032-1304559
   Roundy N, 2012, J NEUROSURG, V116, P697, DOI 10.3171/2011.12.JNS111404
   Samala R, 2019, NEUROL INDIA, V67, P149, DOI 10.4103/0028-3886.253631
   Sampath P, 2000, J NEUROSURG, V92, P70, DOI 10.3171/jns.2000.92.1.0070
   Savardekar AR, 2018, NEUROSURG FOCUS, V44, DOI 10.3171/2017.12.FOCUS17672
   Shapey J, 2019, FRONT NEUROSCI-SWITZ, V13, DOI 10.3389/fnins.2019.00023
   Simon NG, 2014, J NEUROSURG, V120, P1113, DOI 10.3171/2014.2.JNS131975
   Starnoni D, 2018, NEUROSURG FOCUS, V44, DOI 10.3171/2017.12.FOCUS17669
   Sun MZ, 2012, NEUROSURG FOCUS, V33, DOI 10.3171/2012.6.FOCUS12198
   Szmuda T, 2015, OTORYNOLARYNGOLOGIA, V13, P42
   Taoka T, 2006, J MAGN RESON IMAGING, V24, P1005, DOI 10.1002/jmri.20725
   von Elm E, 2008, J CLIN EPIDEMIOL, V61, P344, DOI [10.1016/j.jclinepi.2007.11.008, 10.2471/BLT.07.045120]
   Wei PH, 2015, ACTA NEUROCHIR, V157, P1239, DOI 10.1007/s00701-015-2431-7
   Yoshino M, 2015, J NEUROSURG, V123, P1480, DOI 10.3171/2014.11.JNS14988
   Yoshino M, 2015, ACTA NEUROCHIR, V157, P939, DOI 10.1007/s00701-015-2411-y
   Yoshino M, 2015, INT J COMPUT ASS RAD, V10, P383, DOI 10.1007/s11548-014-1129-2
   Zhang YH, 2013, ACTA NEUROCHIR, V155, P1857, DOI 10.1007/s00701-013-1815-9
   Zolal A, 2017, WORLD NEUROSURG, V107, P47, DOI [10.1016/J.WNEU.2017.07.102, 10.1016/j.wneu.2017.07.102]
NR 29
TC 6
Z9 6
U1 0
U2 4
PU VIA MEDICA
PI GDANSK
PA UL SWIETOKRZYSKA 73, 80-180 GDANSK, POLAND
SN 0028-3843
EI 1897-4260
J9 NEUROL NEUROCHIR POL
JI Neurol. Neurochir. Pol.
PY 2020
VL 54
IS 1
BP 73
EP 82
DI 10.5603/PJNNS.a2020.0001
PG 10
WC Clinical Neurology
WE Science Citation Index Expanded (SCI-EXPANDED)
SC Neurosciences & Neurology
GA KT1NI
UT WOS:000518777900014
PM 31956974
OA Bronze, Green Published
DA 2023-08-27
ER

PT J
AU Baugh, J
   Bartels, U
   Leach, J
   Jones, B
   Chaney, B
   Warren, KE
   Kirkendall, J
   Doughman, R
   Hawkins, C
   Miles, L
   Fuller, C
   Hassall, T
   Bouffet, E
   Lane, A
   Hargrave, D
   Grill, J
   Hoffman, LM
   Jones, C
   Towbin, A
   Savage, SA
   Monje, M
   Li, XN
   Ziegler, DS
   van Zanten, SV
   Kramm, CM
   van Vuurden, DG
   Fouladi, M
AF Baugh, Joshua
   Bartels, Ute
   Leach, James
   Jones, Blaise
   Chaney, Brooklyn
   Warren, Katherine E.
   Kirkendall, Jenavieve
   Doughman, Renee
   Hawkins, Cynthia
   Miles, Lili
   Fuller, Christine
   Hassall, Tim
   Bouffet, Eric
   Lane, Adam
   Hargrave, Darren
   Grill, Jacques
   Hoffman, Lindsey M.
   Jones, Chris
   Towbin, Alex
   Savage, Sharon A.
   Monje, Michelle
   Li, Xiao-Nan
   Ziegler, David S.
   van Zanten, Sophie Veldhuijzen
   Kramm, Christof M.
   van Vuurden, Dannis G.
   Fouladi, Maryam
TI The international diffuse intrinsic pontine glioma registry: an
   infrastructure to accelerate collaborative research for an orphan
   disease
SO JOURNAL OF NEURO-ONCOLOGY
LA English
DT Article
DE Brain tumor; Diffuse intrinsic pontine glioma; Registry
ID ACTIVATING ACVR1 MUTATIONS; CLINICAL-TRIALS; DRIVER MUTATIONS; CHILDREN;
   BIOPSY; DIPG; BIOLOGY; AUTOPSY; GRADE
AB Diffuse intrinsic pontine glioma (DIPG), a rare, often fatal childhood brain tumor, remains a major therapeutic challenge. In 2012, investigators, funded by the DIPG Collaborative (a philanthropic partnership among 29 private foundations), launched the International DIPG Registry (IDIPGR) to advance understanding of DIPG. Comprised of comprehensive deidentified but linked clinical, imaging, histopathological, and genomic repositories, the IDIPGR uses standardized case report forms for uniform data collection; serial imaging and histopathology are centrally reviewed by IDIPGR neuro-radiologists and neuro-pathologists, respectively. Tissue and genomic data, and cell cultures derived from autopsies coordinated by the IDIPGR are available to investigators for studies approved by the Scientific Advisory Committee. From April 2012 to December 2016, 670 patients diagnosed with DIPG have been enrolled from 55 participating institutions in the US, Canada, Australia and New Zealand. The radiology repository contains 3558 studies from 448 patients. The pathology repository contains tissue on 81 patients with another 98 samples available for submission. Fresh DIPG tissue from seven autopsies has been sent to investigators to develop primary cell cultures. The bioinformatics repository contains next-generation sequencing data on 66 tumors. Nine projects using data/tissue from the IDIPGR by 13 principle investigators from around the world are now underway. The IDIPGR, a successful alliance among philanthropic agencies and investigators, has developed and maintained a highly collaborative, hypothesis-driven research infrastructure for interdisciplinary and translational projects in DIPG to improve diagnosis, response assessment, treatment and outcome for patients.
C1 [Baugh, Joshua; Leach, James; Jones, Blaise; Chaney, Brooklyn; Kirkendall, Jenavieve; Doughman, Renee; Miles, Lili; Fuller, Christine; Lane, Adam; Hoffman, Lindsey M.; Towbin, Alex; Fouladi, Maryam] Cincinnati Childrens Hosp Med Ctr, Brain Tumor Ctr, Canc & Blood Dis Inst, 3333 Burnet Ave, Cincinnati, OH 45229 USA.
   [Bartels, Ute; Hawkins, Cynthia; Bouffet, Eric] Hosp Sick Children, Toronto, ON, Canada.
   [Warren, Katherine E.; Savage, Sharon A.] NCI, NIH, Bethesda, MD 20892 USA.
   [Hassall, Tim] Lady Cilento Childrens Hosp, Brisbane, Qld, Australia.
   [Hargrave, Darren] Great Ormond St Hosp Sick Children, London, England.
   [Grill, Jacques] Gustave Roussy Inst, Villejuif, France.
   [Jones, Chris] Inst Canc Res, Sutton, Surrey, England.
   [Monje, Michelle] Stanford Univ, Palo Alto, CA 94304 USA.
   [Monje, Michelle] Lucile Packard Childrens Hosp, Palo Alto, CA 94304 USA.
   [Li, Xiao-Nan] Texas Childrens Hosp, Houston, TX 77030 USA.
   [Ziegler, David S.] Sydney Childrens Hosp, Randwick, NSW, Australia.
   [Ziegler, David S.] Univ New South Wales, Childrens Canc Inst, Kensington, NSW, Australia.
   [van Zanten, Sophie Veldhuijzen; van Vuurden, Dannis G.] Vrije Univ Amsterdam Med Ctr, Amsterdam, Netherlands.
   [Kramm, Christof M.] Univ Gottingen, Gottingen, Germany.
C3 Cincinnati Children's Hospital Medical Center; University of Toronto;
   Hospital for Sick Children (SickKids); National Institutes of Health
   (NIH) - USA; NIH National Cancer Institute (NCI); University of London;
   University College London; Great Ormond Street Hospital for Children NHS
   Foundation Trust; UNICANCER; Gustave Roussy; University of London;
   Institute of Cancer Research - UK; Stanford University; Lucile Packard
   Children's Hospital (LPCH); Baylor College of Medicine; Children's
   Cancer Institute; University of New South Wales Sydney; Vrije
   Universiteit Amsterdam; VU UNIVERSITY MEDICAL CENTER; University of
   Gottingen
RP Fouladi, M (通讯作者)，Cincinnati Childrens Hosp Med Ctr, Brain Tumor Ctr, Canc & Blood Dis Inst, 3333 Burnet Ave, Cincinnati, OH 45229 USA.
EM
RI Grill, Jacques/R-8093-2019; van Vuurden, Dannis/ACA-6952-2022; Savage,
   Sharon/AAB-5160-2019; Towbin, Alexander/AAA-1343-2021; Baugh,
   Joshua/AAH-7710-2020; Hargrave, Darren R/H-1066-2014; Jones,
   Chris/ABE-6059-2021; Bouffet, Eric/AAB-8999-2020
OI Savage, Sharon/0000-0001-6006-0740; Towbin,
   Alexander/0000-0003-1729-5071; Hargrave, Darren R/0000-0001-8219-9807;
   Jones, Chris/0000-0001-8118-2296; Veldhuijzen van Zanten, Sophie
   E.M./0000-0002-3888-0977; Hassall, Timothy/0000-0002-7883-3710; Lane,
   Adam/0000-0002-5522-8112; Ziegler, David/0000-0001-7451-7916; Baugh,
   Joshua/0000-0001-7281-6306
FU DIPG Collaborative; Cancer Research UK [13982] Funding Source:
   researchfish; Great Ormond Street Hospital Childrens Charity [W1075F]
   Funding Source: researchfish; The Brain Tumour Charity [16/193] Funding
   Source: researchfish
FX This research was financially supported by the DIPG Collaborative.
CR Angelini P, 2011, J NEURO-ONCOL, V101, P75, DOI 10.1007/s11060-010-0224-7
   Broniscer A, 2010, CANCER-AM CANCER SOC, V116, P4632, DOI 10.1002/cncr.25405
   Buczkowicz P, 2014, NAT GENET, V46, P451, DOI 10.1038/ng.2936
   Grasso CS, 2015, NAT MED, V21, P555, DOI 10.1038/nm.3855
   Hargrove D, 2006, LANCET ONCOL, V7, P241, DOI 10.1016/S1470-2045(06)70615-5
   Hoffman LM, 2016, ACTA NEUROPATHOL COM, V4, DOI 10.1186/s40478-015-0269-0
   Jansen MHA, 2012, CANCER TREAT REV, V38, P27, DOI 10.1016/j.ctrv.2011.06.007
   Kambhampati M, 2015, ONCOTARGET, V6, P12740, DOI 10.18632/oncotarget.3374
   Kieran MW, 2015, PEDIATR BLOOD CANCER, V62, P3, DOI 10.1002/pbc.25266
   La Madrid AM, 2015, FRONT ONCOL, V5, DOI 10.3389/fonc.2015.00148
   Nikbakht H, 2016, NAT COMMUN, V7, DOI 10.1038/ncomms11185
   Puget S, 2015, CHILD NERV SYST, V31, P1773, DOI 10.1007/s00381-015-2832-1
   Schwartzentruber J, 2012, NATURE, V482, P226, DOI [10.1038/nature10833, 10.1007/s13311-012-0137-6]
   Taylor KR, 2014, NAT GENET, V46, P457, DOI 10.1038/ng.2925
   Walker DA, 2013, NEURO-ONCOLOGY, V15, P462, DOI 10.1093/neuonc/nos330
   Wang ZHJ, 2015, PEDIATR BLOOD CANCER, V62, P163, DOI 10.1002/pbc.25224
   Warren KE, 2012, FRONT ONCOL, V2, DOI 10.3389/fonc.2012.00205
   Wu G, 2014, NAT GENET, V46, P444, DOI 10.1038/ng.2938
   Wu G, 2012, NAT GENET, V44, P251, DOI 10.1038/ng.1102
NR 19
TC 22
Z9 23
U1 0
U2 5
PU SPRINGER
PI NEW YORK
PA 233 SPRING ST, NEW YORK, NY 10013 USA
SN 0167-594X
EI 1573-7373
J9 J NEURO-ONCOL
JI J. Neuro-Oncol.
PD APR
PY 2017
VL 132
IS 2
BP 323
EP 331
DI 10.1007/s11060-017-2372-5
PG 9
WC Oncology; Clinical Neurology
WE Science Citation Index Expanded (SCI-EXPANDED)
SC Oncology; Neurosciences & Neurology
GA ER7TD
UT WOS:000399015500015
PM 28093680
OA Green Submitted, Green Accepted
DA 2023-08-27
ER

PT J
AU Xu, QW
   Bao, WM
   Mao, RL
AF Xu, QW
   Bao, WM
   Mao, RL
TI Diagnosis and treatment of solid brainstem tumors in adults
SO CHINESE MEDICAL JOURNAL
LA English
DT Article
ID STEM; MANAGEMENT
AB Objective To evaluate the diagnosis and aggressive surgical approach for brainstem tumors.
   Methods A series of 24 adult patients with brainstem tumors were involved. Magnetic resonance (MR) imaging was performed on all patients. Surgical technique, potential risk and management of complications were described.
   Results Diagnosis was confirmed by MR imaging and histological examination in all the patients. Tumors were totally or subtotally removed in twenty-two patients. Signs and symptoms were improved in 19 patients on discharge.
   Conclusions Our study shows that MR imaging has an important value for localizing tumors, identifying their nature and selecting appropriate surgical approaches. Most brainstem rumors, except for high-grade gliomas and small ventral tumors, are amenable to an aggressive surgical approach. The favorable results depend upon the appropriate surgical approach, microsurgical technique and intensive care after operation. Total removal can provide both high-quality survival and favorable long-term prognosis.
RP Xu, QW (通讯作者)，SHANGHAI MED UNIV,INST NEUROL,DEPT NEUROSURG,SHANGHAI 200040,PEOPLES R CHINA.
RI Mao, Rong/Q-2595-2019
OI Mao, Rong/0000-0002-0469-2281
CR EDWARDS MSB, 1994, J NEUROSURG, V80, P20, DOI 10.3171/jns.1994.80.1.0020
   EPSTEIN F, 1987, J NEUROSURG, V67, P483, DOI 10.3171/jns.1987.67.4.0483
   EPSTEIN FJ, 1993, J NEUROSURG, V78, P408, DOI 10.3171/jns.1993.78.3.0408
   FASANO VA, 1986, SURG NEUROL, V25, P219, DOI 10.1016/0090-3019(86)90231-4
   HOOD TW, 1989, NEUROSURGERY, V24, P373, DOI 10.1227/00006123-198903000-00011
   LI MH, 1991, ACTA RADIOL, V32, P505
   VANDERTOP WP, 1992, NEUROSURGERY, V31, P186, DOI 10.1227/00006123-199208000-00003
   ZHU DH, 1987, CHIN J NEUROSURG, V3, P11
NR 8
TC 1
Z9 2
U1 0
U2 1
PU CHINESE MEDICAL ASSOCIATION
PI BEIJING
PA 42 DONGSI XIDAJIE, BEIJING 100710, PEOPLES R CHINA
SN 0366-6999
J9 CHINESE MED J-PEKING
JI Chin. Med. J.
PD FEB
PY 1997
VL 110
IS 2
BP 109
EP 113
PG 5
WC Medicine, General & Internal
WE Science Citation Index Expanded (SCI-EXPANDED)
SC General & Internal Medicine
GA WT710
UT WOS:A1997WT71000007
PM 9594280
DA 2023-08-27
ER

PT J
AU Levesque, MF
   Parker, F
AF Levesque, MF
   Parker, F
TI MKM-guided resection of diffuse brainstem neoplasms
SO STEREOTACTIC AND FUNCTIONAL NEUROSURGERY
LA English
DT Article; Proceedings Paper
CT Meeting of the
   American-Society-for-Stereotactic-and-Functional-Neurosurgery
CY JUL 08-10, 1999
CL SNOWBIRD, UTAH
SP Amer Soc Stereotact & Funct Neurosurg
DE brain neoplasm; brainstem/surgery; hemangioblastoma; ependymoma;
   image-guided neurosurgery
AB Objectives: Some primary brainstem tumors, when extensive, are considered inoperable, We wanted to assess the value of robotic image-guided microscopic surgery in the resection of these tumors and to improve survival and quality of life for these patients. Methods: Two patients with extensive brainstem tumors were evaluated at our center. They previously underwent several biopsies, attempted partial resections, radiotherapy and shunting. They presented with progressive neurological deterioration,'coma vigil' for several months, and required life-supporting measures prior to surgery, Both patients underwent frameless stereotactic craniotomy using a MKM robotic microscope, intraoperative neurophysiological monitoring, and extensive resection of their recurrent brainstem tumors. Results: In the immediate weeks after surgery, both patients became interactive and regained major motor and cranial nerve deficits present prior to surgery. Nine months after surgery, 1 patient succumbed to pneumonia. At 2 years after the operation, 1 patient has maintained his neurological status and showed no recurrence on imaging studies. Conclusions: Image-guided surgery with an MKM microscope allows surgical outlines to be injected in the microscope viewer and facilitates resection of extensive brainstem tumors previously considered inoperable. Copyright (C) 2000 S. Karger AG, Basel.
C1 Cedars Sinai Med Ctr, Funct Neurosurg Unit, Los Angeles, CA 90048 USA.
C3 Cedars Sinai Medical Center
CR Parker F, 1999, INTERV NEURORADIOL, V5, P19, DOI 10.1177/159101999900500103
NR 1
TC 4
Z9 4
U1 0
U2 0
PU KARGER
PI BASEL
PA ALLSCHWILERSTRASSE 10, CH-4009 BASEL, SWITZERLAND
SN 1011-6125
J9 STEREOT FUNCT NEUROS
JI Stereotact. Funct. Neurosurg.
PY 1999
VL 73
IS 1-4
BP 15
EP 18
DI 10.1159/000029744
PG 4
WC Neurosciences; Neuroimaging; Surgery
WE Conference Proceedings Citation Index - Science (CPCI-S); Science Citation Index Expanded (SCI-EXPANDED)
SC Neurosciences & Neurology; Surgery
GA 326HN
UT WOS:000087727900003
PM 10853091
DA 2023-08-27
ER

PT J
AU Allen, J
   Siffert, J
   Donahue, B
   Nirenberg, A
   Jakacki, R
   Robertson, P
   DaRosso, R
   Thoron, L
   Rosovsky, M
   Pinto, R
AF Allen, J
   Siffert, J
   Donahue, B
   Nirenberg, A
   Jakacki, R
   Robertson, P
   DaRosso, R
   Thoron, L
   Rosovsky, M
   Pinto, R
TI A phase I/II study of carboplatin combined with hyperfractionated
   radiotherapy for brainstem gliomas
SO CANCER
LA English
DT Article
DE brainstem glioma; radiosensitizers; radiotherapy; chemotherapy; clinical
   trial; Phase I/II trials
ID STEM GLIOMAS; RADIATION-THERAPY; CHILDHOOD; CANCER; TUMORS; SURVIVAL;
   TRIAL; CIS-DIAMMINEDICHLOROPLATINUM(II); IRRADIATION; CISPLATIN
AB BACKGROUND. Brainstem gliomas often respond to radiotherapy but long term disease control is exceptional. The concomitant administration of a chemotherapy agent with radiosensitizing properties such as carboplatin may increase the efficacy of radiotherapy.
   METHODS. A dose escalation schedule of carboplatin was devised to determine the maximum tolerated dose (MTD) of intravenous carboplatin when given on a twice-weekly schedule during a course of hyperfractionated, involved field radiotherapy (100 centigrays [cGy] twice daily to 7200 cGy). The starting dose was 20 mg/m(2) and the dose was increased by 15 mg/m(2) after every 3 patients provided no Grade 3 or 4 (according to the National Institutes of Health Common Toxicity Criteria) toxicity occurred. Magnetic resonance imaging (MRI) scans (brain and spine) were obtained before treatment and at the time of disease progression. Clinical entry criteria included an MRI scan demonstrating a diffuse intrinsic pontine tumor and a typical 2-3-month history of evolving cranial neuropathies and a gait disorder. Biopsy-confirmed evidence of a high grade glioma was required for nonpontine brain stem tumors.
   RESULTS. A total of 34 patients were enrolled. The median age of the patients was 7.8 years (range, 3.6-15.4 years) and the median prodrome duration was 1.5 months (range, 0.25-36 months). The MTD was 110 mg/m(2) or a total cumulative dose of 1540 mg/m(2) over 7 weeks. The dose-limiting toxicity was hematologic. The median progression free survival was 8 months (range, 0-104+ months) and the overall survival was 12 months (range, 5-104+ months). At last follow-up there were 5 long term survivors (15%) who remained in continuous remission after a mean follow-up period of 79 months (range, 46-104 months). Fifteen of the 29 patients (52%) with recurrence and or disease progression developed leptomeningeal/intraaxial tumor spread beyond the local radiation field.
   CONCLUSIONS. The cumulative MTD for carboplatin is 1540 mg/m(2) when administered concomitantly with involved field, hyperfractionated radiotherapy in a twice-weekly schedule for 7 weeks. Subsequent Phase II and III clinical trials can be conducted safely at this level. (C) 1999 American Cancer Society.
C1 Beth Israel Med Ctr, Dept Neurol, New York, NY 10003 USA.
   Beth Israel Med Ctr, Dept Radiol, New York, NY 10003 USA.
   NYU, Med Ctr, Dept Neurol, New York, NY 10016 USA.
   NYU, Med Ctr, Dept Radiol, New York, NY 10016 USA.
   James Whitcomb Riley Hosp Children, Dept Pediat, Indianapolis, IN USA.
   Univ Michigan, Ctr Med, Dept Neurol, Ann Arbor, MI 48109 USA.
C3 Harvard University; Beth Israel Deaconess Medical Center; Harvard
   University; Beth Israel Deaconess Medical Center; New York University;
   New York University; James Whitcomb Riley Hospital Children; University
   of Michigan System; University of Michigan
RP Allen, J (通讯作者)，Beth Israel Med Ctr Singer, 170 East End Ave, New York, NY 10128 USA.
OI Allen, Jeffrey/0000-0001-9957-6664
CR ALBERTS DS, 1989, SEMIN ONCOL, V16, P19
   ALLEN JC, 1987, J CLIN ONCOL, V5, P459, DOI 10.1200/JCO.1987.5.3.459
   BARKOVICH AJ, 1991, PEDIATR NEUROSURG, V16, P73, DOI 10.1159/000120511
   BARTELINK H, 1986, RADIOTHER ONCOL, V6, P61, DOI 10.1016/S0167-8140(86)80110-4
   DEWIT L, 1987, INT J RADIAT ONCOL, V13, P403, DOI 10.1016/0360-3016(87)90015-0
   Donahue B, 1998, INT J RADIAT ONCOL, V40, P677, DOI 10.1016/S0360-3016(97)00846-8
   DOUPLE EB, 1985, CANCER TREAT REV, V12, P111, DOI 10.1016/0305-7372(85)90026-X
   Freeman CR, 1996, CANCER-AM CANCER SOC, V77, P555
   Freeman CR, 1996, PEDIATR NEUROSURG, V24, P103, DOI 10.1159/000121025
   GAYNON PS, 1990, CANCER, V66, P2465, DOI 10.1002/1097-0142(19901215)66:12<2465::AID-CNCR2820661204>3.0.CO;2-N
   HIBI T, 1992, NEUROSURGERY, V31, P643
   JENKIN RDT, 1987, J NEUROSURG, V66, P227, DOI 10.3171/jns.1987.66.2.0227
   Jeremic B, 1996, J CLIN ONCOL, V14, P1065, DOI 10.1200/JCO.1996.14.4.1065
   Matsukado K, 1996, NEUROSURGERY, V39, P125, DOI 10.1097/00006123-199607000-00025
   PACKER RJ, 1994, CANCER-AM CANCER SOC, V74, P1827, DOI 10.1002/1097-0142(19940915)74:6<1827::AID-CNCR2820740628>3.0.CO;2-Q
   PATEL M, 1986, P AACR, V37, P403
   SCHWACHOFER JHM, 1991, INT J RADIAT ONCOL, V20, P1235, DOI 10.1016/0360-3016(91)90233-T
   SKOV K, 1991, INT J RADIAT ONCOL, V20, P221, DOI 10.1016/0360-3016(91)90094-K
   Turrisi AT, 1999, NEW ENGL J MED, V340, P265, DOI 10.1056/NEJM199901283400403
   Volling P, 1992, Semin Oncol, V19, P66
   ZELTZER PM, 1991, CANCER, V67, P43, DOI 10.1002/1097-0142(19910101)67:1<43::AID-CNCR2820670109>3.0.CO;2-7
NR 21
TC 49
Z9 51
U1 0
U2 1
PU WILEY-LISS
PI NEW YORK
PA DIV JOHN WILEY & SONS INC, 605 THIRD AVE, NEW YORK, NY 10158-0012 USA
SN 0008-543X
J9 CANCER
JI Cancer
PD SEP 15
PY 1999
VL 86
IS 6
BP 1064
EP 1069
DI 10.1002/(SICI)1097-0142(19990915)86:6<1064::AID-CNCR24>3.0.CO;2-1
PG 6
WC Oncology
WE Science Citation Index Expanded (SCI-EXPANDED)
SC Oncology
GA 236DL
UT WOS:000082582900024
PM 10491535
DA 2023-08-27
ER

PT J
AU BRUGGERS, CS
   FRIEDMAN, HS
   FULLER, GN
   TIEN, RD
   MARKS, LB
   HALPERIN, EC
   HOCKENBERGER, B
   OAKES, WJ
   HOFFMAN, JM
AF BRUGGERS, CS
   FRIEDMAN, HS
   FULLER, GN
   TIEN, RD
   MARKS, LB
   HALPERIN, EC
   HOCKENBERGER, B
   OAKES, WJ
   HOFFMAN, JM
TI COMPARISON OF SERIAL PET AND MRI SCANS IN A PEDIATRIC-PATIENT WITH A
   BRAIN-STEM GLIOMA
SO MEDICAL AND PEDIATRIC ONCOLOGY
LA English
DT Article
DE BRAIN-STEM GLIOMA; PET SCAN; PONTINE GLIOMA
ID HYPERFRACTIONATED RADIATION-THERAPY; STEM TUMORS; RADIOTHERAPY;
   CHILDREN; CHILDHOOD; CRITERIA; LESIONS; CGY
AB Profound clinical deficits may be associated with insults to the brainstem, making management of patients with brainstem gliomas very complex. Small changes in the radiographic appearance of a brainstem tumor may be associated with significant clinical deterioration. Furthermore, both magnetic resonance imaging and computed tomography are frequently unable to differentiate between therapy-related tissue reactions and progressive tumor. Two clinical scenarios particularly difficult to resolve include: (1) transient radiographic and clinical deterioration following hyperfractionated radiotherapy, and (2) clinical deterioration in a patient who has failed initial therapy, but has stable radiographic findings following a second therapy. We report a child with a pontine glioma whose tumor progression was demonstrated more convincingly with a F-18-deoxyglucose positron emission scan than with magnetic resonance imaging. PET scans may be helpful in confirming that tumor progression is responsible for clinical deterioration in a patient whose MRI scans remain stable.
C1 DUKE UNIV,MED CTR,DEPT PATHOL,DURHAM,NC 27710.
   DUKE UNIV,MED CTR,DEPT RADIOL,DURHAM,NC 27710.
   DUKE UNIV,MED CTR,DEPT SURG,DURHAM,NC 27710.
   DUKE UNIV,MED CTR,DEPT RADIAT ONCOL,DURHAM,NC 27710.
C3 Duke University; Duke University; Duke University; Duke University
RP BRUGGERS, CS (通讯作者)，DUKE UNIV,MED CTR,DEPT PEDIAT,DIV HEMATOL ONCOL,BOX 2916,DURHAM,NC 27710, USA.
OI Fuller, Gregory/0000-0001-9447-2647
CR ALBRIGHT AL, 1983, CANCER, V52, P2313, DOI 10.1002/1097-0142(19831215)52:12<2313::AID-CNCR2820521226>3.0.CO;2-I
   BYRNE JV, 1989, NEURORADIOLOGY, V31, P129, DOI 10.1007/BF00698839
   COHEN ME, 1984, BRAIN TUMORS CHILDRE
   COLEMAN RE, 1991, CANCER, V67, P1261
   DICHIRO G, 1987, AM J NEURORADIOL, V8, P1083
   EDWARDS MSB, 1989, J NEUROSURG, V70, P691, DOI 10.3171/jns.1989.70.5.0691
   FRANCAVILLA TL, 1989, NEUROSURGERY, V24, P1
   FREEMAN CR, 1986, INT J RADIAT ONCOL, V12, P1823, DOI 10.1016/0360-3016(86)90325-1
   FREEMAN CR, 1991, CANCER-AM CANCER SOC, V68, P474, DOI 10.1002/1097-0142(19910801)68:3<474::AID-CNCR2820680305>3.0.CO;2-7
   FREEMAN CR, 1988, INT J RADIAT ONCOL, V15, P311, DOI 10.1016/S0360-3016(98)90010-4
   GRIEBEL M, 1991, MED PEDIATR ONCOL, V19, P182, DOI 10.1002/mpo.2950190307
   HALPERIN EC, 1989, MED PEDIATR ONCOL, V17, P116, DOI 10.1002/mpo.2950170209
   HALPERIN EC, 1989, PEDIATRIC RAD ONCOLO
   HERHOLZ K, 1990, CEREBROVAS BRAIN MET, V2, P240
   HOFFMAN JM, 1992, J COMPUT ASSIST TOMO, V16, P62, DOI 10.1097/00004728-199201000-00011
   HUEFTLE MG, 1985, J COMPUT ASSIST TOMO, V91, P263
   KIM TH, 1980, INT J RADIAT ONCOL, V6, P51, DOI 10.1016/0360-3016(80)90203-5
   LONGEE DC, 1988, NEUROSURGERY, V23, P248, DOI 10.1227/00006123-198808000-00024
   PACKER RJ, 1985, NEUROLOGY, V35, P397, DOI 10.1212/WNL.35.3.397
   PACKER RJ, 1985, PEDIATRICS, V76, P84
   PACKER RJ, 1990, ANN NEUROL, V27, P167, DOI 10.1002/ana.410270212
   PIZZO PA, 1989, PRINCIPLES PRACTICE
   RAEFFELL C, 1988, PEDIATRICS, V82, P733
   VALK PE, 1988, J NEUROSURG, V69, P830, DOI 10.3171/jns.1988.69.6.0830
   WHYTE TR, RADIOLOGY, V93, P413
   ZELTZER PM, 1985, CANCER, V56, P1824, DOI 10.1002/1097-0142(19851001)56:7+<1824::AID-CNCR2820561321>3.0.CO;2-H
NR 26
TC 31
Z9 31
U1 0
U2 0
PU WILEY-LISS
PI NEW YORK
PA DIV JOHN WILEY & SONS INC 605 THIRD AVE, NEW YORK, NY 10158-0012
SN 0098-1532
J9 MED PEDIATR ONCOL
JI Med. Pediatr. Oncol.
PY 1993
VL 21
IS 4
BP 301
EP 306
DI 10.1002/mpo.2950210414
PG 6
WC Oncology; Pediatrics
WE Science Citation Index Expanded (SCI-EXPANDED)
SC Oncology; Pediatrics
GA KV298
UT WOS:A1993KV29800013
PM 8469227
DA 2023-08-27
ER

PT J
AU Carceller, F
   Aleu, A
   Casasco, A
   Guimaraens, L
   Lopez-Pino, MA
   Madero, L
   Ramirez, M
AF Carceller, Fernando
   Aleu, Aitziber
   Casasco, Alfredo
   Guimaraens, Leopoldo
   Lopez-Pino, Migel A.
   Madero, Luis
   Ramirez, Manuel
TI Superselective Intracerebral Catheterization for Administration of
   Oncolytic Virotherapy in a Case of Diffuse Intrinsic Pontine Glioma
SO JOURNAL OF PEDIATRIC HEMATOLOGY ONCOLOGY
LA English
DT Article
DE diffuse intrinsic pontine glioma; virotherapy; angiography
ID BRAIN-STEM TUMORS; BCNU
AB New therapies are needed to improve current results in diffuse intrinsic pontine glioma. We present here the initial experience of administering Celyvir, autologous mesenchymal stem cells infected with ICOVIR-5, an oncolytic adenovirus that selectively replicates in cancer cells, by means of superselective intra-arterial delivery, in a patient diagnosed of diffuse intrinsic pontine glioma. Feasibility, safety, and morbidity rates of the superselective catheterization technique are comparable with those of diagnostic angiography. The intra-arterial approach warrants a greater contact of the mesenchymal stem cells with the tumor mass, and minimizes hemorrhages or vascular disruption. The tolerance to the 2 administrations was excellent, with no acute or delayed adverse effect, underscoring the feasibility of this technique for the delivery of virotherapies and/or cellular therapies in this location.
C1 [Carceller, Fernando; Madero, Luis; Ramirez, Manuel] Hosp Univ Nino Jesus, Dept Oncohematol, Madrid 28009, Spain.
   [Lopez-Pino, Migel A.] Hosp Univ Nino Jesus, Dept Radiol, Madrid 28009, Spain.
   [Aleu, Aitziber; Casasco, Alfredo; Guimaraens, Leopoldo] Clin Nuestra Senora del Rosario, Dept Endovasc Therapies, Madrid, Spain.
RP Ramirez, M (通讯作者)，Hosp Univ Nino Jesus, Serv Oncohematol, Menendez Pelayo 65, Madrid 28009, Spain.
EM
RI Ramirez, Manuel/H-7710-2015
OI Ramirez, Manuel/0000-0003-0332-6973
CR Bartels U, 2011, J NEURO-ONCOL, V105, P119, DOI 10.1007/s11060-011-0704-4
   FREEMAN CR, 1986, INT J RADIAT ONCOL, V12, P1823, DOI 10.1016/0360-3016(86)90325-1
   Garcia-Castro J, 2010, CANCER GENE THER, V17, P476, DOI 10.1038/cgt.2010.4
   Maes W, 2011, CANCER IMMUNOL IMMUN, V60, P153, DOI 10.1007/s00262-010-0946-6
   Nduom EK, 2012, J NEUROSURG, V117, P1150, DOI 10.3171/2012.9.JNS12506
   Okada H, 2011, EXPERT REV NEUROTHER, V11, P619, DOI [10.1586/ern.11.49, 10.1586/ERN.11.49]
   Recinos PF, 2007, PEDIATR NEUROSURG, V43, P192, DOI 10.1159/000098831
   THERON J, 1986, NEURORADIOLOGY, V28, P118, DOI 10.1007/BF00327882
   TYLER JL, 1986, J NUCL MED, V27, P775
NR 9
TC 5
Z9 5
U1 0
U2 5
PU LIPPINCOTT WILLIAMS & WILKINS
PI PHILADELPHIA
PA 530 WALNUT ST, PHILADELPHIA, PA 19106-3621 USA
SN 1077-4114
EI 1536-3678
J9 J PEDIAT HEMATOL ONC
JI J. Pediatr. Hematol. Oncol.
PD OCT
PY 2014
VL 36
IS 7
BP E430
EP E432
DI 10.1097/MPH.0000000000000084
PG 3
WC Oncology; Hematology; Pediatrics
WE Science Citation Index Expanded (SCI-EXPANDED)
SC Oncology; Hematology; Pediatrics
GA AQ8BF
UT WOS:000343045300007
DA 2023-08-27
ER

PT J
AU Osorio, DS
   Patel, N
   Ji, LY
   Sposto, R
   Stanek, J
   Gardner, SL
   Allen, JC
   Cornelius, A
   McCowage, GB
   Termuhlen, A
   Dunkel, IJ
   Comito, M
   Garvin, J
   Finlay, JL
AF Osorio, Diana S.
   Patel, Neha
   Ji, Lingyun
   Sposto, Richard
   Stanek, Joseph
   Gardner, Sharon L.
   Allen, Jeffrey C.
   Cornelius, Albert
   McCowage, Geoffrey B.
   Termuhlen, Amanda
   Dunkel, Ira J.
   Comito, Melanie
   Garvin, James
   Finlay, Jonathan L.
TI Pre-irradiation intensive induction and marrow-ablative consolidation
   chemotherapy in young children with newly diagnosed high-grade brainstem
   gliomas: report of the "head-start" I and II clinical trials
SO JOURNAL OF NEURO-ONCOLOGY
LA English
DT Article
DE Marrow-ablative therapy; High-dose chemotherapy; High-grade glioma;
   Brainstem; Brainstem glioma; Diffuse intrinsic pontine glioma; DIPG
ID HIGH-DOSE-CHEMOTHERAPY; HYPERFRACTIONATED RADIATION-THERAPY; PHASE-II;
   PRERADIATION CHEMOTHERAPY; CEREBROSPINAL-FLUID; PEDIATRIC-PATIENTS;
   ONCOLOGY-GROUP; THIOTEPA; RESCUE; TUMORS
AB Background The dismal outcome in children with high-grade brainstem gliomas (BSG) accentuates the need for effective therapeutic strategies. We investigated the role of intensive, including marrow-ablative, chemotherapy regimens in the treatment of young children with newly-diagnosed high-grade BSG.
   Methods Between 1991-and-2002, 15 eligible children less than 10 years of age with a diagnosis of high-grade BSG were treated on "Head-Start" I and II protocols (HSI and HSII). Treatment included Induction with 4-5 cycles of one of three intensive chemotherapy regimens followed by Consolidation with one cycle of marrow-ablative chemotherapy (thiotepa, carboplatin and etoposide) with autologous hematopoietic cell rescue (AHCR). Irradiation was required for children over 6 years of age or for those with residual tumor at the end of Consolidation.
   Results We had two long-term survivors who were found retrospectively to harbor low-grade glial tumors and thus were not included in the survival analysis. Of the remaining 13 patients, the 1-year event-free (EFS) and overall (OS) survival for these children were 31% (95% CI 9-55%) and 38% (95% CI 14-63%), respectively. Median EFS and OS were 6.6 (95% CI 2.7, 12.7) and 8.7 months (95% CI 6.9, 20.9), respectively. Eight patients developed progressive disease during study treatment (seven during Induction and one at the end of Consolidation). Ten children received focal irradiation, five for residual tumor (three following Induction and two following Consolidation) and five due to disease progression.
   Conclusions Children with high-grade BSG did not benefit from this intensive chemotherapy strategy administered prior to irradiation.
C1 [Osorio, Diana S.; Stanek, Joseph; Finlay, Jonathan L.] Ohio State Univ, Div Pediat Hematol Oncol, Nationwide Childrens Hosp, Columbus, OH 43210 USA.
   [Patel, Neha] Univ Wisconsin, Div Pediat Hematol Oncol, Madison, WI USA.
   [Ji, Lingyun; Sposto, Richard; Termuhlen, Amanda] Univ Southern Calif, Dept Prevent Med, Los Angeles, CA USA.
   [Gardner, Sharon L.; Allen, Jeffrey C.] NYU, Div Pediat Oncol, Langone Med Ctr, New York, NY USA.
   [Cornelius, Albert] Helen DeVos Childrens Hosp, Div Pediat Hematol Oncol, Grand Rapids, MI USA.
   [McCowage, Geoffrey B.] Childrens Hosp Westmead, Dept Oncol, Sydney, NSW, Australia.
   [Dunkel, Ira J.] Mem Sloan Kettering Canc Ctr, Dept Pediat, 1275 York Ave, New York, NY 10021 USA.
   [Dunkel, Ira J.] Weill Cornell Med Coll, New York, NY USA.
   [Comito, Melanie] Penn State Hershey Med Ctr, Div Pediat Hematol Oncol, Hershey, PA USA.
   [Garvin, James] Columbia Univ, Med Ctr, Div Pediat Hematol Oncol & Stem Cell Transplant, New York, NY USA.
C3 University System of Ohio; Ohio State University; Nationwide Childrens
   Hospital; University of Wisconsin System; University of Wisconsin
   Madison; University of Southern California; New York University; NYU
   Langone Medical Center; University of Sydney; Memorial Sloan Kettering
   Cancer Center; Cornell University; Weill Cornell Medicine; Pennsylvania
   Commonwealth System of Higher Education (PCSHE); Pennsylvania State
   University; Penn State Health; Columbia University
RP Osorio, DS (通讯作者)，Ohio State Univ, Div Pediat Hematol Oncol, Nationwide Childrens Hosp, Columbus, OH 43210 USA.
EM;
  ;
  ;
  ;
  ;
  ;
  
RI McCowage, Geoffrey/ABB-5072-2020
OI McCowage, Geoffrey/0000-0003-3471-8443; gardner,
   sharon/0000-0002-8857-5487; Allen, Jeffrey/0000-0001-9957-6664
FU NCI NIH HHS [P30 CA008748] Funding Source: Medline
CR Ascendo JAT, 1989, P AM ASSOC CANC RES, V9, P90
   BERNIERCHASTAGNER, 2005, CANCER, V104, P2792, DOI DOI 10.1002/CNCR.21534
   Bouffet E, 1997, EUR J CANCER, V33, P91, DOI 10.1016/S0959-8049(96)00369-3
   Bouffet E, 2000, CANCER, V88, P685, DOI 10.1002/(SICI)1097-0142(20000201)88:3<685::AID-CNCR27>3.0.CO;2-K
   Broniscer A, 2005, CANCER-AM CANCER SOC, V103, P133, DOI 10.1002/cncr.20741
   Broniscer A, 2000, J CLIN ONCOL, V18, P1246, DOI 10.1200/JCO.2000.18.6.1246
   CALVERT AH, 1989, J CLIN ONCOL, V7, P1748, DOI 10.1200/JCO.1989.7.11.1748
   Cartmill M, 1999, CHILD NERV SYST, V15, P235
   Cohen KJ, 2011, NEURO-ONCOLOGY, V13, P410, DOI 10.1093/neuonc/noq205
   DONELLI MG, 1992, CANCER CHEMOTH PHARM, V30, P251, DOI 10.1007/BF00686291
   Doz F, 2002, EUR J CANCER, V38, P815, DOI 10.1016/S0959-8049(02)00029-1
   DUFFNER PK, 1993, NEW ENGL J MED, V328, P1725, DOI 10.1056/NEJM199306173282401
   Dunkel IJ, 1998, J NEURO-ONCOL, V37, P67, DOI 10.1023/A:1005874508975
   EDWARDS MS, 1979, CANCER TREAT REP, V63, P1419
   EIFEL PJ, 1987, INT J RADIAT ONCOL, V13, P847, DOI 10.1016/0360-3016(87)90098-8
   Espinoza JC, 2016, PEDIATR BLOOD CANCER, V63, P1806, DOI 10.1002/pbc.26118
   Estlin EJ, 1998, BRIT J CANCER, V78, P652, DOI 10.1038/bjc.1998.555
   Fagioli F, 2004, CANCER, V100, P2215, DOI 10.1002/cncr.20227
   FINLAY JL, 1990, J NEURO-ONCOL, V9, P239, DOI 10.1007/BF02341155
   Finlay JL, 1996, J CLIN ONCOL, V14, P2495, DOI 10.1200/JCO.1996.14.9.2495
   Fouladi M, 2007, CANCER-AM CANCER SOC, V110, P2535, DOI 10.1002/cncr.23078
   Frappaz D, 2008, NEURO-ONCOLOGY, V10, P599, DOI 10.1215/15228517-2008-029
   FREEMAN CR, 1986, INT J RADIAT ONCOL, V12, P1823, DOI 10.1016/0360-3016(86)90325-1
   Gokce-Samar Z, 2016, CHILD NERV SYST, V32, P1415, DOI 10.1007/s00381-016-3153-8
   Gururangan S, 1998, J CLIN ONCOL, V16, P2486, DOI 10.1200/JCO.1998.16.7.2486
   Hargrove D, 2006, LANCET ONCOL, V7, P241, DOI 10.1016/S1470-2045(06)70615-5
   HEIDEMAN RL, 1993, CANCER-AM CANCER SOC, V72, P271, DOI 10.1002/1097-0142(19930701)72:1<271::AID-CNCR2820720147>3.0.CO;2-K
   HEIDEMAN RL, 1989, CANCER RES, V49, P736
   HEIDEMAN RL, 1993, J CLIN ONCOL, V11, P1458, DOI 10.1200/JCO.1993.11.8.1458
   HOCHBERG FH, 1981, J NEUROSURG, V54, P455, DOI 10.3171/jns.1981.54.4.0455
   Jakacki RI, 1999, J NEURO-ONCOL, V44, P77, DOI 10.1023/A:1006360222643
   Jansen MHA, 2012, CANCER TREAT REV, V38, P27, DOI 10.1016/j.ctrv.2011.06.007
   JENKIN RDT, 1987, J NEUROSURG, V66, P227, DOI 10.3171/jns.1987.66.2.0227
   Jennings MT, 2002, J CLIN ONCOL, V20, P3431, DOI 10.1200/JCO.2002.04.109
   JOHNSON DB, 1987, J CLIN ONCOL, V5, P783, DOI 10.1200/JCO.1987.5.5.783
   Jones C, 2017, NEURO-ONCOLOGY, V19, P153, DOI 10.1093/neuonc/now101
   KALIFA C, 1992, BONE MARROW TRANSPL, V9, P227
   KEDAR A, 1994, MED PEDIATR ONCOL, V23, P428, DOI 10.1002/mpo.2950230507
   KRETSCHMAR CS, 1993, CANCER-AM CANCER SOC, V72, P1404, DOI 10.1002/1097-0142(19930815)72:4<1404::AID-CNCR2820720441>3.0.CO;2-G
   MARIA BL, 1993, J CHILD NEUROL, V8, P112, DOI 10.1177/088307389300800203
   Mason WP, 1998, J CLIN ONCOL, V16, P210, DOI 10.1200/JCO.1998.16.1.210
   MBIDDE EK, 1988, BRIT J CANCER, V58, P779, DOI 10.1038/bjc.1988.308
   Packer RJ, 1996, CANCER-AM CANCER SOC, V77, P2150, DOI 10.1002/(SICI)1097-0142(19960515)77:10<2150::AID-CNCR28>3.0.CO;2-T
   PACKER RJ, 1994, CANCER-AM CANCER SOC, V74, P1827, DOI 10.1002/1097-0142(19940915)74:6<1827::AID-CNCR2820740628>3.0.CO;2-Q
   Papadakis V, 2000, BONE MARROW TRANSPL, V26, P153, DOI 10.1038/sj.bmt.1702475
   POSTMUS PE, 1984, J CLIN ONCOL, V2, P215, DOI 10.1200/JCO.1984.2.3.215
   Sharp JR, 2010, EUR J CANCER, V46, P3271, DOI 10.1016/j.ejca.2010.06.115
   TARTAGLIA RL, 1989, P AM ASSOC CANC RES, V30, P1833
   Walker DA, 1999, ARCH DIS CHILD, V80, P558, DOI 10.1136/adc.80.6.558
   Zimmerman RA, 1996, PEDIATR NEUROSURG, V25, P45, DOI 10.1159/000121096
NR 50
TC 5
Z9 5
U1 1
U2 4
PU SPRINGER
PI NEW YORK
PA 233 SPRING ST, NEW YORK, NY 10013 USA
SN 0167-594X
EI 1573-7373
J9 J NEURO-ONCOL
JI J. Neuro-Oncol.
PD DEC
PY 2018
VL 140
IS 3
BP 717
EP 725
DI 10.1007/s11060-018-03003-z
PG 9
WC Oncology; Clinical Neurology
WE Science Citation Index Expanded (SCI-EXPANDED)
SC Oncology; Neurosciences & Neurology
GA HC2MG
UT WOS:000451635500025
PM 30392092
OA Green Accepted
DA 2023-08-27
ER

PT J
AU Liu, AK
   Brandon, J
   Foreman, NK
   Fenton, LZ
AF Liu, Arthur K.
   Brandon, Jonathan
   Foreman, Nicholas K.
   Fenton, Laura Z.
TI Conventional MRI at presentation does not predict clinical response to
   radiation therapy in children with diffuse pontine glioma
SO PEDIATRIC RADIOLOGY
LA English
DT Article
DE Diffuse pontine glioma; MRI; Children
ID BRAIN-STEM GLIOMA; PEDIATRIC-ONCOLOGY-GROUP; HYPERFRACTIONATED
   RADIOTHERAPY; TUMORS; TRIAL; PET
AB Diffuse pontine glioma is a pediatric brain tumor with a median survival time of 1 year and few long-term survivors. Conventional MRI has not been shown to predict overall survival. With this dismal prognosis, clinical improvement after radiation therapy might be a more relevant issue.
   To determine whether initial imaging would predict clinical response.
   We performed a retrospective review of children with diffuse pontine glioma treated at The Children's Hospital, Denver, and the University of Colorado-Denver from 1995 to 2008. The 30 children were divided into two groups based on acute clinical response to radiation therapy. Quantitative measurements on presenting MRIs were compared between the groups.
   Seven of 30 children did not respond to radiation therapy. There were no statistically significant differences in initial imaging characteristics between responders and non-responders. Necrosis and enhancement were more common in responders. Tumor surrounding the basilar artery, compression of the 4th ventricle and extension to the cerebellum were more common in non-responders.
   Conventional MRI at presentation does not predict clinical response in children with diffuse pontine glioma. Additional studies with larger numbers of patients and other imaging modalities are needed.
C1 [Liu, Arthur K.] Univ Colorado Denver, Dept Radiat Oncol, Aurora, CO 80045 USA.
   [Brandon, Jonathan; Fenton, Laura Z.] Childrens Hosp, Dept Radiol, Aurora, CO USA.
   [Foreman, Nicholas K.] Childrens Hosp, Dept Neurooncol, Aurora, CO USA.
C3 Children's Hospital Colorado; University of Colorado System; University
   of Colorado Anschutz Medical Campus; Children's Hospital Colorado;
   Children's Hospital Colorado
RP Liu, AK (通讯作者)，Univ Colorado Denver, Dept Radiat Oncol, MS F706,1665 Aurora Court, Aurora, CO 80045 USA.
EM
OI Foreman, Nicholas/0000-0003-4848-4801
CR BRUGGERS CS, 1993, MED PEDIATR ONCOL, V21, P301, DOI 10.1002/mpo.2950210414
   Fenton LZ, 2003, MED PEDIATR ONCOL, V40, P260, DOI 10.1002/mpo.10227
   FREEMAN CR, 1993, INT J RADIAT ONCOL, V27, P197, DOI 10.1016/0360-3016(93)90228-N
   Hargrave D, 2008, J NEURO-ONCOL, V86, P313, DOI 10.1007/s11060-007-9473-5
   Janssens GORJ, 2009, INT J RADIAT ONCOL, V73, P722, DOI 10.1016/j.ijrobp.2008.05.030
   Korones DN, 2008, PEDIATR BLOOD CANCER, V50, P227, DOI 10.1002/pbc.21154
   Laprie A, 2005, INT J RADIAT ONCOL, V62, P20, DOI 10.1016/j.ijrobp.2004.09.027
   Mandell LR, 1999, INT J RADIAT ONCOL, V43, P959, DOI 10.1016/S0360-3016(98)00501-X
   Moghrabi A, 1995, PEDIATR NEUROSURG, V23, P293, DOI 10.1159/000120974
   PACKER RJ, 1993, CANCER-AM CANCER SOC, V72, P1414, DOI 10.1002/1097-0142(19930815)72:4<1414::AID-CNCR2820720442>3.0.CO;2-C
   Panigrahy A, 2008, NEURO-ONCOLOGY, V10, P32, DOI 10.1215/15228517-2007-042
   Pirotte BJM, 2007, J NEUROSURG, V107, P392, DOI 10.3171/PED-07/11/392
   SMITH RR, 1990, NEURORADIOLOGY, V32, P265, DOI 10.1007/BF00593044
NR 13
TC 20
Z9 20
U1 0
U2 5
PU SPRINGER
PI NEW YORK
PA 233 SPRING ST, NEW YORK, NY 10013 USA
SN 0301-0449
J9 PEDIATR RADIOL
JI Pediatr. Radiol.
PD DEC
PY 2009
VL 39
IS 12
BP 1317
EP 1320
DI 10.1007/s00247-009-1368-5
PG 4
WC Pediatrics; Radiology, Nuclear Medicine & Medical Imaging
WE Science Citation Index Expanded (SCI-EXPANDED)
SC Pediatrics; Radiology, Nuclear Medicine & Medical Imaging
GA 519AA
UT WOS:000271736200007
PM 19657635
DA 2023-08-27
ER

PT J
AU Machetanz, K
   Grimm, F
   Wang, S
   Schuhmann, MU
   Tatagiba, M
   Gharabaghi, A
   Naros, G
AF Machetanz, Kathrin
   Grimm, Florian
   Wang, Sophie
   Schuhmann, Martin U.
   Tatagiba, Marcos
   Gharabaghi, Alireza
   Naros, Georgios
TI Rediscovery of the transcerebellar approach: improving the risk-benefit
   ratio in robot-assisted brainstem biopsies
SO NEUROSURGICAL FOCUS
LA English
DT Article
DE brainstem tumors; cerebellar anatomy; robot-assisted biopsies;
   transcerebellar approach; transfrontal approach
ID INTRINSIC PONTINE GLIOMA; STEREOTACTIC BIOPSY; CHILDREN; FRAMELESS;
   SURGERY; LESIONS; SAFETY; SERIES; EXPERIENCE; EFFICACY
AB OBJECTIVE Conventional frame-based stereotaxy through a transfrontal approach (TFA) is the gold standard in brainstem biopsies. Because of the high surgical morbidity and limited impact on therapy, brainstem biopsies are contro-versial. The introduction of robot-assisted stereotaxy potentially improves the risk-benefit ratio by simplifying a transcerebellar approach (TCA). The aim of this single-center cohort study was to evaluate the risk-benefit ratio of transcerebellar brainstem biopsies performed by 2 different robotic systems. In addition to standard quality indicators, a special focus was set on trajectory selection for reducing surgical morbidity.
   METHODS This study included 25 pediatric (n = 7) and adult (n = 18) patients who underwent 26 robot-assisted biopsies via a TCA. The diagnostic yield, complication rate, trajectory characteristics (i.e., length, anatomical entry, and target-point location), and skin-to-skin (STS) time were evaluated. Transcerebellar and hypothetical transfrontal trajectories were reconstructed and transferred into a common MR space for further comparison with anatomical atlases.
   RESULTS Robot-assisted, transcerebellar biopsies demonstrated a high diagnostic yield (96.2%) while exerting no sur-gical mortality and no permanent morbidity in both pediatric and adult patients. Only 3.8% of cases involved a transient neurological deterioration. Transcerebellar trajectories had a length of 48.4 +/- 7.3 mm using a wide stereotactic corridor via crus I or II of the cerebellum and the middle cerebellar peduncle. The mean STS time was 49.5 +/- 23.7 minutes and differed significantly between the robotic systems (p = 0.017). The TFA was characterized by longer trajectories (107.4 +/- 11.8 mm, p < 0.001) and affected multiple eloquent structures. Transfrontal target points were located significantly more medial (-3.4 +/- 7.2 mm, p = 0.042) and anterior (-3.9 +/- 8.4 mm, p = 0.048) in comparison with the transcerebellar trajectories.
   CONCLUSIONS Robot-assisted, transcerebellar stereotaxy can improve the risk-benefit ratio of brainstem biopsies by avoiding the restrictions of a TFA and conventional frame-based stereotaxy. Profound registration and anatomical-functional trajectory selection were essential to reduce mortality and morbidity.
C1 [Machetanz, Kathrin; Grimm, Florian; Wang, Sophie; Schuhmann, Martin U.; Tatagiba, Marcos; Gharabaghi, Alireza; Naros, Georgios] Eberhard Karls Univ Tubingen, Dept Neurosurg & Neurotechnol, Neurosurg Clin, Tubingen, Germany.
   [Machetanz, Kathrin; Grimm, Florian; Gharabaghi, Alireza; Naros, Georgios] Eberhard Karls Univ Tubingen, Inst Neuromodulat & Neurotechnol, Dept Neurosurg & Neurotechnol, Tubingen, Germany.
C3 Eberhard Karls University of Tubingen; Eberhard Karls University of
   Tubingen
RP Naros, G (通讯作者)，Eberhard Karls Univ Tubingen, Tubingen, Germany.
EM
RI Naros, Georgios/ABR-8074-2022
OI Naros, Georgios/0000-0003-4807-6234; Gharabaghi,
   Alireza/0000-0002-9782-5281; Wang, Sophie Shih-Yung/0000-0001-9632-2909
CR ABERNATHEY CD, 1989, J NEUROSURG, V70, P195, DOI 10.3171/jns.1989.70.2.0195
   ALBRIGHT AL, 1993, NEUROSURGERY, V33, P1026
   APUZZO MLJ, 1987, NEUROSURGERY, V20, P930, DOI 10.1227/00006123-198706000-00019
   BARKOVICH AJ, 1991, PEDIATR NEUROSURG, V16, P73, DOI 10.1159/000120511
   Carai A, 2017, WORLD NEUROSURG, V101, P584, DOI 10.1016/j.wneu.2017.02.088
   Coca HA, 2016, NEUROCHIRURGIE, V62, P327, DOI 10.1016/j.neuchi.2016.07.005
   Dammers R, 2008, ACTA NEUROCHIR, V150, P23, DOI 10.1007/s00701-007-1473-x
   Dawes W, 2019, J ROBOT SURG, V13, P575, DOI 10.1007/s11701-018-0899-x
   De Benedictis A, 2017, NEUROSURG FOCUS, V42, DOI 10.3171/2017.2.FOCUS16579
   Dellaretti M, 2020, SCI REP-UK, V10, DOI 10.1038/s41598-020-68063-6
   Dellaretti M, 2012, STEREOT FUNCT NEUROS, V90, P79, DOI 10.1159/000335502
   Diedrichsen J, 2006, NEUROIMAGE, V33, P127, DOI 10.1016/j.neuroimage.2006.05.056
   Grimaldi G, 2012, CEREBELLUM, V11, P336, DOI 10.1007/s12311-011-0247-4
   Grimm F, 2015, J NEUROSURG, V123, P737, DOI 10.3171/2014.12.JNS141781
   Grossman R, 2005, ACTA NEUROCHIR, V147, P627, DOI 10.1007/s00701-005-0495-5
   Gupta M, 2021, J NEUROSURG-PEDIATR, V27, P317, DOI 10.3171/2020.7.PEDS20373
   Haegelen C, 2010, NEUROCHIRURGIE, V56, P363, DOI 10.1016/j.neuchi.2010.05.006
   Hankinson TC, 2011, J NEUROSURG-PEDIATR, V8, P97, DOI 10.3171/2011.4.PEDS1180
   Hargrove D, 2006, LANCET ONCOL, V7, P241, DOI 10.1016/S1470-2045(06)70615-5
   Horisawa S, 2017, WORLD NEUROSURG, V107, P1, DOI [10.1016/J.WNEU.2017.07.087, 10.1016/j.wneu.2017.07.087]
   Horn A, 2015, NEUROIMAGE, V107, P127, DOI 10.1016/j.neuroimage.2014.12.002
   Jaradat A, 2021, ACTA NEUROCHIR, V163, P1957, DOI 10.1007/s00701-021-04733-2
   Kickingereder P, 2013, NEUROSURGERY, V72, P873, DOI 10.1227/NEU.0b013e31828bf445
   KONDZIOLKA D, 1995, SURG NEUROL, V43, P558, DOI 10.1016/0090-3019(95)00009-7
   Lefranc M, 2015, J NEUROSURG, V122, P342, DOI 10.3171/2014.9.JNS14107
   Machetanz K, 2021, INT J MED ROBOT COMP, V17, DOI 10.1002/rcs.2288
   Machetanz K, 2021, J NEUROSURG, V135, P1477, DOI 10.3171/2020.10.JNS201843
   Machetanz K, 2021, STEREOT FUNCT NEUROS, V99, P25, DOI 10.1159/000510107
   Mader MMD, 2019, J NEURO-ONCOL, V141, P183, DOI 10.1007/s11060-018-03024-8
   Manoj N, 2014, J NEUROSCI RURAL PRA, V5, P32, DOI 10.4103/0976-3147.127869
   Massager N, 2000, J NEUROSURG, V93, P951, DOI 10.3171/jns.2000.93.6.0951
   Merchant TE, 2010, SEMIN RADIAT ONCOL, V20, P58, DOI 10.1016/j.semradonc.2009.09.005
   Naros G, 2021, INT J MED ROBOT COMP, V17, DOI 10.1002/rcs.2282
   Payne CJ, NEUROMETHODS, V2021, P3
   Rachinger W, 2009, J NEUROL NEUROSUR PS, V80, P1134, DOI 10.1136/jnnp.2009.174250
   RAJSHEKHAR V, 1995, J NEUROSURG, V82, P976, DOI 10.3171/jns.1995.82.6.0976
   Sawin PD, 1998, SURG NEUROL, V49, P640, DOI 10.1016/S0090-3019(97)00435-7
   Schaller B, 1999, J NEUROSURG, V90, P215, DOI 10.3171/jns.1999.90.2.0215
   Schmahmann JD, 2019, NEUROSCI LETT, V688, P62, DOI 10.1016/j.neulet.2018.07.005
   Smith JA, 2016, ANN BIOMED ENG, V44, P836, DOI 10.1007/s10439-015-1475-4
   Spyrantis A, 2019, EPILEPSY BEHAV, V91, P38, DOI 10.1016/j.yebeh.2018.11.002
   STECK J, 1995, SURG NEUROL, V43, P563, DOI 10.1016/0090-3019(95)00156-5
   Tang YC, 2018, NEUROIMAGE, V169, P227, DOI 10.1016/j.neuroimage.2017.12.042
   Williams JR, 2020, NEUROSURG FOCUS, V48, DOI 10.3171/2019.9.FOCUS19745
   Xiao YM, 2017, DATA BRIEF, V12, P370, DOI 10.1016/j.dib.2017.04.013
NR 45
TC 3
Z9 3
U1 2
U2 4
PU AMER ASSOC NEUROLOGICAL SURGEONS
PI ROLLING MEADOWS
PA 5550 MEADOWBROOK DRIVE, ROLLING MEADOWS, IL 60008 USA
SN 1092-0684
J9 NEUROSURG FOCUS
JI Neurosurg. Focus
PD JAN
PY 2022
VL 52
IS 1
AR E12
DI 10.3171/2021.10.FOCUS21359
PG 8
WC Clinical Neurology; Surgery
WE Science Citation Index Expanded (SCI-EXPANDED)
SC Neurosciences & Neurology; Surgery
GA YE1XP
UT WOS:000740923400002
PM 34973665
OA gold
DA 2023-08-27
ER

PT J
AU Kim, SD
   Kim, JH
   Lee, CY
   Kim, HW
AF Kim, Sung-Duk
   Kim, Jong Hyun
   Lee, Cheol-Young
   Kim, Hyun-Woo
TI Ganglioglioma in Brainstem : Case Report and a Review of Literatures
SO JOURNAL OF KOREAN NEUROSURGICAL SOCIETY
LA English
DT Review
DE Ganglioglioma; Brainstem
ID CEREBELLAR GANGLIOGLIOMA; CEREBELLOPONTINE ANGLE; RARE TUMOR; LOCATION;
   PATIENT
AB Ganglioglioma is an infrequent tumor of the central nervous system (CNS); mostly supratentorial region. But, they can occur anywhere in the central nervous system such as brainstem, cerebellopontine angle (CPA), thalamus, optic nerve and spinal cord. Although it occurs rarely, ganglioglioma should be included in the differential diagnosis of a posterior fossa mass because early recognition is important for treatment and patient counseling.
C1 [Kim, Sung-Duk; Kim, Jong Hyun; Lee, Cheol-Young; Kim, Hyun-Woo] Konyang Univ Hosp, Dept Neurosurg, Taejon 302718, South Korea.
   [Lee, Cheol-Young] Kyung Hee Univ, Grad Sch Med, Seoul, South Korea.
C3 Konyang University; Konyang University Hospital; Kyung Hee University
RP Kim, JH (通讯作者)，Konyang Univ Hosp, Dept Neurosurg, 158 Gwanjeodong Ro, Taejon 302718, South Korea.
EM
CR Chae JH, 2001, EPILEPSIA, V42, P1204, DOI 10.1046/j.1528-1157.2001.43398.x
   Chan MH, 2012, PEDIATR BLOOD CANCER, V59, P1173, DOI 10.1002/pbc.24232
   Jang EW, 2007, J KOREAN NEUROSURG S, V42, P53
   Kwon JW, 2001, AM J NEURORADIOL, V22, P1377
   Lagares A, 2001, SURG NEUROL, V56, P315, DOI 10.1016/S0090-3019(01)00618-8
   LANG FF, 1993, J NEUROSURG, V79, P867, DOI 10.3171/jns.1993.79.6.0867
   Lobel U, 2011, CLIN RADIOL, V66, P194, DOI 10.1016/j.crad.2010.10.005
   Manning HL, 2000, AM J RESP CRIT CARE, V161, P2100, DOI 10.1164/ajrccm.161.6.9904087
   Milligan BD, 2007, J NEUROSURG, V107, P292, DOI 10.3171/PED-07/10/292
   Mpairamidis E, 2008, J CHILD NEUROL, V23, P1481, DOI 10.1177/0883073808319316
   Park SH, 2008, J KOREAN NEUROSURG S, V43, P165, DOI 10.3340/jkns.2008.43.3.165
   Shin JJ, 2001, J KOREAN NEUROSURG S, V30, P239
   Shin MJ, 2002, J KOREAN NEUROSURG S, V32, P279
   Westwood DA, 2009, J CLIN NEUROSCI, V16, P108, DOI 10.1016/j.jocn.2007.11.016
   Wetmore C, 2012, PEDIATR BLOOD CANCER, V59, P1153, DOI 10.1002/pbc.24287
   Zhang S, 2013, J NEUROSURG, V118, P884, DOI 10.3171/2013.1.JNS121323
NR 16
TC 8
Z9 8
U1 1
U2 3
PU KOREAN NEUROSURGICAL SOC
PI SEOUL
PA #402 POSCO # OFFICE BUILDING 151 SUNHWA-DONG, JUNG-GU, SEOUL, 100-130,
   SOUTH KOREA
SN 2005-3711
EI 1598-7876
J9 J KOREAN NEUROSURG S
JI J. Korean. Neurosurg. Soc.
PD MAR
PY 2014
VL 55
IS 3
BP 164
EP 166
DI 10.3340/jkns.2014.55.3.164
PG 3
WC Clinical Neurology; Surgery
WE Science Citation Index Expanded (SCI-EXPANDED)
SC Neurosciences & Neurology; Surgery
GA AG0RJ
UT WOS:000335123000010
PM 24851154
OA Green Published, gold
DA 2023-08-27
ER

PT J
AU Ni, C
   Qin, DM
   Cheng, H
   Zhou, M
   Luo, DD
AF Ni, Cheng
   Qin, Daming
   Cheng, Hong
   Zhou, Meng
   Luo, Dandan
TI Effect Evaluation of Combined Application of Magnetic Resonance
   Diffusion Tensor Imaging and Brain Function Imaging in Radiation Therapy
   of Brain Tumours Involving Motor Pathways
SO JOURNAL OF MEDICAL IMAGING AND HEALTH INFORMATICS
LA English
DT Article
DE Functional MRI; Diffuse Tensor Imaging; Brain Tumour; Radiotherapy; Cone
   Beam Imaging
ID RESPONSE EVALUATION CRITERIA; SOLID TUMORS; NECROSIS; CANCER; MRI
AB This study is an attempt to find a way for functional imaging information to be applied clinically in radiation therapy. The basal nucleus is a collective term for a group of neural nucleus in the central nervous system that connects the pontine, brainstem, and cerebral cortex, including the caudate nucleus, the bean-shaped nucleus, the screen-shaped nucleus, and the amygdala. It is difficult to find the exact position of these neural nuclei on the computed tomography (CT) image or the T1 or T2 sequence of magnetic resonance. However, the development of neurosurgery has partially confirmed that these functional nuclei are involved in advanced cognitive functions such as memory, emotion, and learning. Neurosurgery has tried to avoid damaging these nucleus groups during surgery to improve the quality of life of patients, and there is currently no clear strategy for this in radiotherapy. Because CT and magnetic resonance spin echo (SE) sequences are difficult to find the anatomical location of the nucleus, it is difficult to have any strategy to protect these functions in radiotherapy planning. This article uses diffusion tensor imaging (DTI) images and fiber bundle tracking to obtain a more accurate anatomical position of the nerve nucleus on the image, and provides some available strategies for radiotherapy to protect patients' brain function. The conclusion of this paper is that the combined application of DTI and functional magnetic resonance imaging (fMRI) can better observe the relationship among tumours, functional areas and white matter fibers, and guide the designation of radiotherapy plans.
C1 [Ni, Cheng; Qin, Daming; Cheng, Hong; Zhou, Meng; Luo, Dandan] Cent Hosp Enshi Tujia & Miao Autonomous Prefectur, En Shi 445000, Peoples R China.
RP Luo, DD (通讯作者)，Cent Hosp Enshi Tujia & Miao Autonomous Prefectur, En Shi 445000, Peoples R China.
RI Luo, Dan/HZM-2412-2023
CR Alexander AL, 2007, NEUROTHERAPEUTICS, V4, P316, DOI 10.1016/j.nurt.2007.05.011
   [Anonymous], 2018, EUROPEAN RADIOLOGY, DOI DOI 10.6111/JKCGCT.2018.28.1.001
   Apostolopolou E, 2010, J CLIN NURS, V19, P1560, DOI 10.1111/j.1365-2702.2009.03011.x
   Becker CO, 2018, SCI REP-UK, V8, DOI 10.1038/s41598-017-18769-x
   Bojaxhiu B, 2018, INT J RADIAT ONCOL, V100, P987, DOI 10.1016/j.ijrobp.2017.11.037
   Bondy ML, 2008, CANCER-AM CANCER SOC, V113, P1953, DOI 10.1002/cncr.23741
   Chakhoyan A, 2018, AM J NEURORADIOL, V39, P43, DOI 10.3174/ajnr.A5433
   Edeline J, 2012, CANCER-AM CANCER SOC, V118, P147, DOI 10.1002/cncr.26255
   Esteban O, 2019, NAT METHODS, V16, P111, DOI 10.1038/s41592-018-0235-4
   Freyschlag C F, 2017, Brain Inform, V4, P201, DOI 10.1007/s40708-017-0064-8
   Holtsche MM, 2018, BRIT J DERMATOL, V179, P918, DOI 10.1111/bjd.16553
   Jacobson LA, 2019, CHILD NEUROPSYCHOL, V25, P217, DOI 10.1080/09297049.2018.1456517
   Koch SBJ, 2017, J PSYCHIATR NEUROSCI, V42, P331, DOI 10.1503/jpn.160129
   Lequin M, 2017, NEUROIMAG CLIN N AM, V27, P167, DOI 10.1016/j.nic.2016.08.007
   Liu C.-B., 2004, COMPUT MED IMAGING G, V10, P404
   Liu CB, 2018, NEURAL REGEN RES, V13, P877, DOI 10.4103/1673-5374.232485
   Mardor Y, 2004, NEOPLASIA, V6, P136, DOI 10.1593/neo.03349
   Park K, 2016, JAMA ONCOL, V2, P305, DOI 10.1001/jamaoncol.2015.4921
   Popp Ilinca, 2019, Top Magn Reson Imaging, V28, P63, DOI 10.1097/RMR.0000000000000198
   Scarabino T, 2003, EUR J RADIOL, V48, P154, DOI 10.1016/j.ejrad.2003.08.008
   Stavinoha Peter L., 2018, Bioengineering-Basel, V5, P73, DOI 10.3390/bioengineering5030073
   Tan HB, 2011, CLIN NUCL MED, V36, P978, DOI 10.1097/RLU.0b013e31822f68a6
   Tummala Subhash, 2017, Int J MS Care, V19, P158, DOI 10.7224/1537-2073.2016-068
   Williams L.J., 2010, ENCY RES DESIGN, V218, P840, DOI DOI 10.4135/9781412961288.N154
   Wnuk N, 2019, INT J GASTROINT INT, V8, P41, DOI 10.18528/ijgii180037
   Yanagawa M, 2012, J NUCL MED, V53, P872, DOI 10.2967/jnumed.111.098699
NR 26
TC 1
Z9 2
U1 0
U2 19
PU AMER SCIENTIFIC PUBLISHERS
PI VALENCIA
PA 26650 THE OLD RD, STE 208, VALENCIA, CA 91381-0751 USA
SN 2156-7018
EI 2156-7026
J9 J MED IMAG HEALTH IN
JI J. Med. Imaging Health Inform.
PD MAR
PY 2021
VL 11
IS 3
BP 894
EP 902
DI 10.1166/jmihi.2021.3329
PG 9
WC Mathematical & Computational Biology; Radiology, Nuclear Medicine &
   Medical Imaging
WE Science Citation Index Expanded (SCI-EXPANDED)
SC Mathematical & Computational Biology; Radiology, Nuclear Medicine &
   Medical Imaging
GA PL4EZ
UT WOS:000603078500031
DA 2023-08-27
ER

PT J
AU Morais, BA
   Solla, DJF
   Matushita, H
   Teixeira, MJ
   Monaco, BA
AF Morais, Barbara Albuquerque
   Fontoura Solla, Davi Jorge
   Matushita, Hamilton
   Teixeira, Manoel Jacobsen
   Monaco, Bernardo A.
TI Pediatric intrinsic brainstem lesions: clinical, imaging, histological
   characterization, and predictors of survival
SO CHILDS NERVOUS SYSTEM
LA English
DT Article
DE Stereotactic biopsy; Brainstem; Children; Histopathological diagnosis;
   Survival
ID PONTINE GLIOMA; DIAGNOSIS; BIOPSY; CHILDHOOD; MUTATIONS; SUBGROUPS;
   CHILDREN; TUMORS
AB Purpose Brainstem lesions comprise 10-20% of all pediatric brain tumors. Over the past years, the benefits of stereotactic biopsy versus the use alone of MRI features to guide treatment have been controversial. Methods Retrospective study with pediatric patients submitted to stereotactic brainstem biopsies between 2008 and 2018. Demographic, clinical, imaging, and surgical characteristics were recorded, as well as the histological diagnosis, complications, and survival. Predictors of survival were evaluated through Cox regression models after multivariate adjustment. Results Twenty-six patients (mean age of 8.8 +/- 4.3 years and 14 female). Diagnosis was reached on 84.6% (95% CI 65.1-95.6%) of the patients. Glioma was diagnosed on 20 cases (11 high-grade and 9 low-grade lesions). There was no association between age and gender and the dichotomized histological diagnosis. Contrast enhancement, diffuse distribution, invasion of adjacent structures, and remote injury were present on 62.5%, 75.0%, 62.5%, and 25.0% of the cases. Hydrocephalus at admission was present on almost half of the patients (46.2%). Only radiological invasion of adjacent structures had a possible association with high-grade lesions (p = 0.057). Surgical trajectory was trans-cerebellar in most of the cases (79.9%). There were no major complications and only two minor/transitory complications. Poorer survival was independently associated with high-grade lesions (HR 32.14, 95% CI 1.40-735.98, p = 0.030) and contrast enhancement at MRI (HR 36.54, 95% CI 1.40-952.26, p = 0.031). Conclusions Stereotactic biopsy was safe and allows successful tissue sampling for a definite diagnosis. Poorer survival was independently associated with high-grade and contrast-enhancing lesions.
C1 [Morais, Barbara Albuquerque; Fontoura Solla, Davi Jorge; Matushita, Hamilton; Teixeira, Manoel Jacobsen; Monaco, Bernardo A.] Univ Sao Paulo, Sch Med, Dept Neurosurg, St Eneas de Carvalho 155, Sao Paulo, SP, Brazil.
C3 Universidade de Sao Paulo
RP Morais, BA (通讯作者)，Univ Sao Paulo, Sch Med, Dept Neurosurg, St Eneas de Carvalho 155, Sao Paulo, SP, Brazil.
EM
RI Solla, Davi/GWU-9718-2022; Solla, Davi Jorge Fontoura/E-6988-2013;
   Assumpcao de Monaco, Bernardo/AHD-3228-2022
OI Solla, Davi Jorge Fontoura/0000-0002-5092-6595; Assumpcao de Monaco,
   Bernardo/0000-0002-2475-9228
CR ALBRIGHT AL, 1993, NEUROSURGERY, V33, P1026
   Buczkowicz P, 2014, ACTA NEUROPATHOL, V128, P573, DOI 10.1007/s00401-014-1319-6
   Buczkowicz P, 2014, NAT GENET, V46, P451, DOI 10.1038/ng.2936
   Castel D, 2015, ACTA NEUROPATHOL, V130, P815, DOI 10.1007/s00401-015-1478-0
   Dellaretti M, 2011, J NEUROSURG-PEDIATR, V8, P539, DOI 10.3171/2011.9.PEDS1167
   EPSTEIN F, 1986, J NEUROSURG, V64, P11, DOI 10.3171/jns.1986.64.1.0011
   Furlanetti LL, 2015, CLIN NEUROL NEUROSUR, V130, P42, DOI 10.1016/j.clineuro.2014.12.011
   GLEASON CA, 1978, NEUROSURGERY, V2, P217, DOI 10.1227/00006123-197805000-00006
   Hamisch C, 2017, J NEUROSURG-PEDIATR, V20, P261, DOI 10.3171/2017.2.PEDS1665
   Hankinson TC, 2011, J NEUROSURG-PEDIATR, V8, P97, DOI 10.3171/2011.4.PEDS1180
   Hennika T, 2016, J CHILD NEUROL, V31, P1377, DOI 10.1177/0883073815601495
   Jallo G, 2006, CHILD NERV SYST, V22, P1, DOI 10.1007/s00381-005-1267-5
   Pierre-Aurelien B, 2016, WORLD NEUROSURG, V93, DOI 10.1016/j.wneu.2016.07.011
   Poussaint TY, 2011, NEURO-ONCOLOGY, V13, P417, DOI 10.1093/neuonc/noq200
   Puget S, 2015, CHILD NERV SYST, V31, P1773, DOI 10.1007/s00381-015-2832-1
   Schumacher M, 2007, J NEUROSURG, V106, P111, DOI 10.3171/ped.2007.106.2.111
   STROINK AR, 1986, J NEUROSURG, V65, P745, DOI 10.3171/jns.1986.65.6.0745
   Taylor KR, 2014, NAT GENET, V46, P457, DOI 10.1038/ng.2925
NR 18
TC 2
Z9 2
U1 0
U2 1
PU SPRINGER
PI NEW YORK
PA ONE NEW YORK PLAZA, SUITE 4600, NEW YORK, NY, UNITED STATES
SN 0256-7040
EI 1433-0350
J9 CHILD NERV SYST
JI Childs Nerv. Syst.
PD MAY
PY 2020
VL 36
IS 5
BP 933
EP 939
DI 10.1007/s00381-019-04453-0
PG 7
WC Clinical Neurology; Pediatrics; Surgery
WE Science Citation Index Expanded (SCI-EXPANDED)
SC Neurosciences & Neurology; Pediatrics; Surgery
GA LD5JM
UT WOS:000526065900012
PM 31836906
DA 2023-08-27
ER

PT J
AU Bhatia, R
   Desai, S
   Garg, A
   Padma, MV
   Prasad, K
   Tripathi, M
AF Bhatia, Rohit
   Desai, Soaham
   Garg, Ajay
   Padma, Madakasira V.
   Prasad, Kameshwar
   Tripathi, Manjari
TI Isolated facial myokymia as a presenting feature of pontine
   neurocysticercosis
SO MOVEMENT DISORDERS
LA English
DT Article
DE myokymia; neurocysticercosis; brainstem
ID MULTIPLE-SCLEROSIS; GLIOMA; CONTRACTURE; SYMPTOM; TUMOR
AB A 45-year-old healthy man presented with 2 weeks history of continuous rippling and quivering movements of his right side of face and neck suggestive of myokymia. MRI scan of the head revealed neurocysticercus in the pons. Treatment with steroids and carbamezapine produced a significant benefit. This is the first report of pontine neurocysticercosis presenting as an isolated facial myokymia. (c) 2007 Movement Disorder Society.
C1 [Bhatia, Rohit; Desai, Soaham; Padma, Madakasira V.; Prasad, Kameshwar; Tripathi, Manjari] All India Inst Med Sci, Dept Neurol, New Delhi 110029, India.
   [Garg, Ajay] All India Inst Med Sci, Dept Neuroradiol, New Delhi 110029, India.
C3 All India Institute of Medical Sciences (AIIMS) New Delhi; All India
   Institute of Medical Sciences (AIIMS) New Delhi
RP Bhatia, R (通讯作者)，All India Inst Med Sci, Dept Neurol, New Delhi 110029, India.
EM
RI Garg, Ajay/AAM-2603-2020; Desai, Soaham/I-6857-2013
OI Garg, Ajay/0000-0002-9385-291X; Desai, Soaham/0000-0003-0378-1888
CR BEYDOUN SR, 1994, MUSCLE NERVE, V17, P1060, DOI 10.1002/mus.880170914
   Blunt SB, 1997, MOVEMENT DISORD, V12, P235, DOI 10.1002/mds.870120215
   BOGHEN D, 1977, NEUROLOGY, V27, P270, DOI 10.1212/WNL.27.3.270
   CHALK CH, 1988, NEUROLOGY, V38, P1811, DOI 10.1212/WNL.38.11.1811
   ESPINOSA RE, 1967, MAYO CLIN PROC, V42, P258
   Garcia HH, 2005, LANCET NEUROL, V4, P653, DOI 10.1016/S1474-4422(05)70194-0
   GUTMANN L, 1994, MUSCLE NERVE, V17, P1461, DOI 10.1002/mus.880171216
   GUTMANN L, 1969, J AMER MED ASSOC, V209, P389, DOI 10.1001/jama.209.3.389
   Gutmann L, 2001, NEUROLOGY, V57, P1707, DOI 10.1212/WNL.57.9.1707
   Jimenez-Jimenez F J, 1992, Acta Neurol (Napoli), V14, P56
   Joseph BV, 2002, NEUROL INDIA, V50, P369
   JUVARRA G, 1981, ITAL J NEUROL SCI, V2, P35, DOI 10.1007/BF02351685
   KRAUSS JK, 1993, NEUROSURGERY, V32, P1031, DOI 10.1227/00006123-199306000-00027
   NEGRI S, 1976, EUR NEUROL, V14, P108, DOI 10.1159/000114733
   RADU EW, 1975, EUR NEUROL, V13, P499, DOI 10.1159/000114706
   SANDYK R, 1985, EUR NEUROL, V24, P112, DOI 10.1159/000115770
   Sedano MJ, 2000, EUR NEUROL, V43, P137, DOI 10.1159/000008152
   Selvapandian S, 1999, NEUROL INDIA, V47, P241
   SETHI PK, 1974, J NEUROL NEUROSUR PS, V37, P745, DOI 10.1136/jnnp.37.6.745
   SETHI PK, 1975, BRIT MED J, V1, P476
   Sharma R R, 1992, J Postgrad Med, V38, P37
   WASSERSTROM WR, 1977, ARCH NEUROL-CHICAGO, V34, P576, DOI 10.1001/archneur.1977.00500210078016
NR 22
TC 11
Z9 12
U1 0
U2 0
PU WILEY-LISS
PI HOBOKEN
PA DIV JOHN WILEY & SONS INC, 111 RIVER ST, HOBOKEN, NJ 07030 USA
SN 0885-3185
J9 MOVEMENT DISORD
JI Mov. Disord.
PD JAN 15
PY 2008
VL 23
IS 1
BP 135
EP 137
DI 10.1002/mds.21792
PG 3
WC Clinical Neurology
WE Science Citation Index Expanded (SCI-EXPANDED)
SC Neurosciences & Neurology
GA 258WS
UT WOS:000252901400023
PM 17987648
DA 2023-08-27
ER

PT J
AU Walter, AW
   Gajjar, A
   Ochs, JS
   Langston, JW
   Sanford, RA
   Kun, LE
   Heideman, R
AF Walter, AW
   Gajjar, A
   Ochs, JS
   Langston, JW
   Sanford, RA
   Kun, LE
   Heideman, R
TI Carboplatin and etoposide with hyperfractionated radiotherapy in
   children with newly diagnosed diffuse pontine gliomas: A phase I/II
   study
SO MEDICAL AND PEDIATRIC ONCOLOGY
LA English
DT Article
DE brainstem; glioma; brain neoplasms; antineoplastic agents; child
ID BRAIN-STEM TUMORS; PEDIATRIC-ONCOLOGY-GROUP; RADIATION-THERAPY; II
   TRIAL; CHEMOTHERAPY; CHILDHOOD; CISPLATIN; GY; IRRADIATION; IPROPLATIN
AB Background. Diffuse pontine gliomas remain one of the most lethal oi pediatric malignancies despite the use of increasingly intensive therapies. We delivered intensive chemotherapy during and following 70.2 Gy of hyperfractionated radiation therapy in an attempt to improve survival.
   Procedure. Nine consecutive children with diffuse pontine gliomas were treated on this single arm study. Carboplatin, given in combination with fixed dose etoposide, was escalated in successive cohorts to determine its maximum tolerated systemic exposure (AUG). Outcome was coded based on imaging characteristics and clinical status.
   Results. Eight of the nine children on this-study died of their disease at a median of 44 weeks, essentially the same survival as those treated on a previous Pediatric Oncology Group study using hyperfractionated radiation therapy alone. Toxicity was almost exclusively hematologic and not associated with significant morbidity.
   Conclusions. The use of concurrent carboplatin and etoposide with hyperfractionated radiation therapy did not appear to improve the survival in this group of children with diffuse pontine gliomas. The toxicity of this chemotherapy during radiation therapy was primarily hematologic and well tolerated. New approaches to the treatment of these tumors need to be investigated. (C) 1998 Wiley-Liss. Inc.
C1 ST JUDE CHILDRENS HOSP,DEPT RADIAT ONCOL,MEMPHIS,TN 38105.
   ST JUDE CHILDRENS HOSP,DEPT DIAGNOST IMAGING,MEMPHIS,TN 38105.
   UNIV TENNESSEE,CTR HLTH SCI,DEPT PEDIAT,MEMPHIS,TN 38163.
   UNIV TENNESSEE,DEPT RADIAT ONCOL,MEMPHIS,TN 38163.
   UNIV TENNESSEE,CTR HLTH SCI,DEPT RADIOL,MEMPHIS,TN 38163.
   UNIV TENNESSEE,DEPT NEUROSURG,MEMPHIS,TN 38163.
   UNIV ARKANSAS MED SCI,DEPT PEDIAT,DIV PEDIAT,DIV PEDIAT HEMATOL ONCOL,LITTLE ROCK,AR 72205.
   UNIV ARKANSAS MED SCI,DEPT NEUROSURG,LITTLE ROCK,AR 72205.
C3 St Jude Children's Research Hospital; St Jude Children's Research
   Hospital; University of Tennessee System; University of Tennessee Health
   Science Center; University of Tennessee System; University of Tennessee
   Health Science Center; University of Tennessee System; University of
   Tennessee Health Science Center; University of Tennessee System;
   University of Tennessee Health Science Center; University of Arkansas
   System; University of Arkansas Medical Sciences; University of Arkansas
   System; University of Arkansas Medical Sciences
RP Walter, AW (通讯作者)，ST JUDE CHILDRENS HOSP,DEPT HEMATOL ONCOL,332 N LAUDERDALE ST,MEMPHIS,TN 38105, USA.
RI Gajjar, Amar/N-8059-2018
FU NCI NIH HHS [P01CA-23099, P30CA-21765] Funding Source: Medline
CR Albright AL, 1996, PEDIATR NEUROSURG, V24, P252
   ALLEN JC, 1987, J CLIN ONCOL, V5, P459, DOI 10.1200/JCO.1987.5.3.459
   ALLEN JC, 1983, CANCER, V52, P2001, DOI 10.1002/1097-0142(19831201)52:11<2001::AID-CNCR2820521105>3.0.CO;2-D
   CALVERT AH, 1989, J CLIN ONCOL, V7, P1748, DOI 10.1200/JCO.1989.7.11.1748
   DROPCHO EJ, 1992, J CLIN ONCOL, V10, P452, DOI 10.1200/JCO.1992.10.3.452
   EDWARDS MSB, 1989, J NEUROSURG, V70, P691, DOI 10.3171/jns.1989.70.5.0691
   FEUN LG, 1984, CANCER, V54, P794, DOI 10.1002/1097-0142(19840901)54:5<794::AID-CNCR2820540503>3.0.CO;2-F
   FREEMAN C, 1995, INT J RADIAT ONCOL, V32, P85
   FREEMAN CR, 1986, INT J RADIAT ONCOL, V12, P1823, DOI 10.1016/0360-3016(86)90325-1
   FREEMAN CR, 1993, INT J RADIAT ONCOL, V27, P197, DOI 10.1016/0360-3016(93)90228-N
   FREEMAN CR, 1991, CANCER-AM CANCER SOC, V68, P474, DOI 10.1002/1097-0142(19910801)68:3<474::AID-CNCR2820680305>3.0.CO;2-7
   Freeman CR, 1996, PEDIATR NEUROSURG, V24, P103, DOI 10.1159/000121025
   FREEMAN CR, 1988, INT J RADIAT ONCOL, V15, P311, DOI 10.1016/S0360-3016(98)90010-4
   FREEMAN CR, 1995, INT J RADIAT ONCOL, V32, P549, DOI 10.1016/0360-3016(95)00141-K
   FRIEDMAN HS, 1992, J CLIN ONCOL, V10, P249, DOI 10.1200/JCO.1992.10.2.249
   GAYNON PS, 1990, CANCER, V66, P2465, DOI 10.1002/1097-0142(19901215)66:12<2465::AID-CNCR2820661204>3.0.CO;2-N
   GAYNON PS, 1987, CANCER TREAT REP, V71, P1039
   JENKIN RDT, 1987, J NEUROSURG, V66, P227, DOI 10.3171/jns.1987.66.2.0227
   KEDAR A, 1994, MED PEDIATR ONCOL, V23, P428, DOI 10.1002/mpo.2950230507
   KNOX SJ, 1994, INT J RADIAT ONCOL, V28, P445, DOI 10.1016/0360-3016(94)90069-8
   Kolotas C, 1996, INT J RADIAT ONCOL, V35, P137, DOI 10.1016/S0360-3016(96)85022-X
   KRETSCHMAR CS, 1993, CANCER-AM CANCER SOC, V72, P1404, DOI 10.1002/1097-0142(19930815)72:4<1404::AID-CNCR2820720441>3.0.CO;2-G
   MAHALEY MS, 1989, J NEUROSURG, V70, P371, DOI 10.3171/jns.1989.70.3.0371
   MARINA NM, 1993, J CLIN ONCOL, V11, P554, DOI 10.1200/JCO.1993.11.3.554
   *NAT CANC I DIV CA, 1993, INV HDB MAN PART CLI
   NEWTON HB, 1989, J NEURO-ONCOL, V7, P39, DOI 10.1007/BF00149377
   OHARA JA, 1986, INT J RADIAT ONCOL, V12, P1419, DOI 10.1016/0360-3016(86)90185-9
   Packer RJ, 1996, CANCER-AM CANCER SOC, V77, P2150, DOI 10.1002/(SICI)1097-0142(19960515)77:10<2150::AID-CNCR28>3.0.CO;2-T
   PACKER RJ, 1993, CANCER-AM CANCER SOC, V72, P1414, DOI 10.1002/1097-0142(19930815)72:4<1414::AID-CNCR2820720442>3.0.CO;2-C
   PACKER RJ, 1994, CANCER-AM CANCER SOC, V74, P1827, DOI 10.1002/1097-0142(19940915)74:6<1827::AID-CNCR2820740628>3.0.CO;2-Q
   PACKER RJ, 1990, ANN NEUROL, V27, P167, DOI 10.1002/ana.410270212
   PFEFFER MR, 1990, INT J RADIAT ONCOL, V19, P1439, DOI 10.1016/0360-3016(90)90356-O
   PIERREKAHN A, 1993, J NEUROSURG, V79, P845, DOI 10.3171/jns.1993.79.6.0845
   PRADOS MD, 1995, INT J RADIAT ONCOL, V32, P85, DOI 10.1016/0360-3016(95)00563-E
   SCHABEL FM, 1979, CANCER TREAT REP, V63, P1459
   WITHERS HR, 1982, INT J RADIAT ONCOL, V8, P1807, DOI 10.1016/0360-3016(82)90307-8
   YUNG WKA, 1991, J CLIN ONCOL, V9, P860, DOI 10.1200/JCO.1991.9.5.860
   ZELTZER PM, 1991, CANCER, V67, P43, DOI 10.1002/1097-0142(19910101)67:1<43::AID-CNCR2820670109>3.0.CO;2-7
NR 38
TC 49
Z9 49
U1 0
U2 0
PU WILEY-LISS
PI NEW YORK
PA DIV JOHN WILEY & SONS INC, 605 THIRD AVE, NEW YORK, NY 10158-0012
SN 0098-1532
J9 MED PEDIATR ONCOL
JI Med. Pediatr. Oncol.
PD JAN
PY 1998
VL 30
IS 1
BP 28
EP 33
DI 10.1002/(SICI)1096-911X(199801)30:1<28::AID-MPO9>3.0.CO;2-2
PG 6
WC Oncology; Pediatrics
WE Science Citation Index Expanded (SCI-EXPANDED)
SC Oncology; Pediatrics
GA YD979
UT WOS:A1998YD97900009
PM 9371386
DA 2023-08-27
ER

PT J
AU Larson, JD
   Kasper, LH
   Paugh, BS
   Jin, H
   Wu, G
   Kwon, CH
   Fan, Y
   Shaw, TI
   Silveira, AB
   Qu, C
   Xu, R
   Zhu, X
   Zhang, J
   Russell, HR
   Peters, JL
   Finkelstein, D
   Xu, B
   Lin, T
   Tinkle, CL
   Patay, Z
   Onar-Thomas, A
   Pounds, SB
   Mckinnon, PJ
   Ellison, DW
   Zhang, J
   Baker, SJ
AF Larson, Jon D.
   Kasper, Lawryn H.
   Paugh, Barbara S.
   Jin, Hongjian
   Wu, Gang
   Kwon, Chang-Hyuk
   Fan, Yiping
   Shaw, Timothy, I
   Silveira, Andre B.
   Qu, Chunxu
   Xu, Raymond
   Zhu, Xiaoyan
   Zhang, Junyuan
   Russell, Helen R.
   Peters, Jennifer L.
   Finkelstein, David
   Xu, Beisi
   Lin, Tong
   Tinkle, Christopher L.
   Patay, Zoltan
   Onar-Thomas, Arzu
   Pounds, Stanley B.
   Mckinnon, Peter J.
   Ellison, David W.
   Zhang, Jinghui
   Baker, Suzanne J.
TI Histone H3.3 K27M Accelerates Spontaneous Brainstem Glioma and Drives
   Restricted Changes in Bivalent Gene Expression
SO CANCER CELL
LA English
DT Article
ID PEDIATRIC HIGH-GRADE; NEURAL PROGENITOR CELLS; SET ENRICHMENT ANALYSIS;
   READ ALIGNMENT; NERVOUS-SYSTEM; MUTATIONS; TUMORS; MODEL; ACVR1; EZH2
AB Diffuse intrinsic pontine gliomas (DIPGs) are incurable childhood brainstem tumors with frequent histone H3 K27M mutations and recurrent alterations in PDGFRA and TP53. We generated genetically engineered inducible mice and showed that H3.3 K27M enhanced neural stem cell self-renewal while preserving regional identity. Neonatal induction of H3.3 K27M cooperated with activating platelet-derived growth factor receptor alpha (PDGFR alpha) mutant and Trp53 loss to accelerate development of diffuse brainstem gliomas that recapitulated human DIPG gene expression signatures and showed global changes in H3K27 posttranslational modifications, but relatively restricted gene expression changes. Genes upregulated in H3.3 K27M tumors were enriched for those associated with neural development where H3K27me3 loss released the poised state of apparently bivalent promoters, whereas downregulated genes were enriched for those encoding homeodomain transcription factors.
C1 [Larson, Jon D.; Kasper, Lawryn H.; Paugh, Barbara S.; Kwon, Chang-Hyuk; Silveira, Andre B.; Xu, Raymond; Zhu, Xiaoyan; Zhang, Junyuan; Baker, Suzanne J.] St Jude Childrens Res Hosp, Dept Dev Neurobiol, 332 N Lauderdale St, Memphis, TN 38105 USA.
   [Jin, Hongjian; Wu, Gang; Fan, Yiping; Shaw, Timothy, I; Finkelstein, David; Xu, Beisi; Zhang, Jinghui] St Jude Childrens Res Hosp, Dept Computat Biol, 332 N Lauderdale St, Memphis, TN 38105 USA.
   [Russell, Helen R.; Mckinnon, Peter J.] St Jude Childrens Res Hosp, Dept Genet, 332 N Lauderdale St, Memphis, TN 38105 USA.
   [Peters, Jennifer L.] St Jude Childrens Res Hosp, Cellular Imaging Shared Resource, 332 N Lauderdale St, Memphis, TN 38105 USA.
   [Qu, Chunxu; Ellison, David W.] St Jude Childrens Res Hosp, Dept Pathol, 332 N Lauderdale St, Memphis, TN 38105 USA.
   [Tinkle, Christopher L.] St Jude Childrens Res Hosp, Dept Radiat Oncol, 332 N Lauderdale St, Memphis, TN 38105 USA.
   [Patay, Zoltan] St Jude Childrens Res Hosp, Dept Diagnost Imaging, 332 N Lauderdale St, Memphis, TN 38105 USA.
   [Lin, Tong; Onar-Thomas, Arzu; Pounds, Stanley B.] St Jude Childrens Res Hosp, Dept Biostat, 332 N Lauderdale St, Memphis, TN 38105 USA.
C3 St Jude Children's Research Hospital; St Jude Children's Research
   Hospital; St Jude Children's Research Hospital; St Jude Children's
   Research Hospital; St Jude Children's Research Hospital; St Jude
   Children's Research Hospital; St Jude Children's Research Hospital; St
   Jude Children's Research Hospital
RP Baker, SJ (通讯作者)，St Jude Childrens Res Hosp, Dept Dev Neurobiol, 332 N Lauderdale St, Memphis, TN 38105 USA.
EM
RI Wu, Gang/M-8012-2014; Pounds, Stanley B/N-2480-2018; Silveira, André
   B/I-8859-2012; Xu, Beisi/C-8560-2009; Jin, Hong-jian/M-9543-2013;
   Ellison, David/N-4806-2018; Baker, Suzanne J/N-8167-2018; Kasper,
   Lawryn/GWV-5700-2022; Kwon, Chang-hyuk/E-3450-2011; Patay,
   Zoltan/N-6994-2018; Shaw, Timothy I/AAE-8523-2019
OI Wu, Gang/0000-0002-1678-5864; Pounds, Stanley B/0000-0002-9167-2114;
   Silveira, André B/0000-0003-3375-1081; Xu, Beisi/0000-0003-0099-858X;
   Patay, Zoltan/0000-0003-1479-9864; Shaw, Timothy I/0000-0002-9316-1924;
   Jin, Hongjian/0000-0003-3833-7170
FU NIH [CA096832, CA188516, CA211481]; Swim Across America; NCI Cancer
   Center Support Grant [CA21765]; ALSAC; Musicians Against Childhood
   Cancer
FX 0 This work was supported by NIH grants CA096832 (to S.J.B., P.J.McK.,
   D.W.E., and J.Z.), CA188516 (to S.J.B.), CA211481 (CIVIT), Swim Across
   America (to S.J.B.), the NCI Cancer Center Support Grant CA21765,
   Musicians Against Childhood Cancer and ALSAC.
CR Anders S, 2015, BIOINFORMATICS, V31, P166, DOI 10.1093/bioinformatics/btu638
   Bayliss J, 2016, SCI TRANSL MED, V8, DOI 10.1126/scitranslmed.aah6904
   Bender S, 2013, CANCER CELL, V24, P660, DOI 10.1016/j.ccr.2013.10.006
   Buczkowicz P, 2014, NAT GENET, V46, P451, DOI 10.1038/ng.2936
   Carbon S, 2017, NUCLEIC ACIDS RES, V45, pD331, DOI 10.1093/nar/gkw1108
   Chan KM, 2013, GENE DEV, V27, P985, DOI 10.1101/gad.217778.113
   Chow LML, 2011, CANCER CELL, V19, P305, DOI 10.1016/j.ccr.2011.01.039
   Cordero FJ, 2017, MOL CANCER RES, V15, P1243, DOI 10.1158/1541-7786.MCR-16-0389
   Fontebasso AM, 2014, NAT GENET, V46, P462, DOI 10.1038/ng.2950
   Fujii Y, 2013, GENES CELLS, V18, P79, DOI 10.1111/gtc.12024
   Funato K, 2014, SCIENCE, V346, P1529, DOI 10.1126/science.1253799
   Gritti A, 1999, J NEUROSCI, V19, P3287
   Heinz S, 2010, MOL CELL, V38, P576, DOI 10.1016/j.molcel.2010.05.004
   Herz HM, 2014, SCIENCE, V345, P1065, DOI 10.1126/science.1255104
   Huse JT, 2011, GLIA, V59, P1190, DOI 10.1002/glia.21165
   Jones C, 2014, NAT REV CANCER, V14, P651, DOI 10.1038/nrc3811
   Jonkers J, 2001, NAT GENET, V29, P418, DOI 10.1038/ng747
   Kuleshov MV, 2016, NUCLEIC ACIDS RES, V44, pW90, DOI 10.1093/nar/gkw377
   Larson JD, 2019, METHODS MOL BIOL, V1869, P207, DOI 10.1007/978-1-4939-8805-1_18
   Le S, 2008, J STAT SOFTW, V25, P1, DOI 10.18637/jss.v025.i01
   Lewis PW, 2013, SCIENCE, V340, P857, DOI 10.1126/science.1232245
   Li H, 2009, BIOINFORMATICS, V25, P1754, DOI 10.1093/bioinformatics/btp324
   Li H, 2009, BIOINFORMATICS, V25, P1094, DOI [10.1093/bioinformatics/btp100, 10.1093/bioinformatics/btp324]
   Lindquist RA, 2016, NAT COMMUN, V7, DOI 10.1038/ncomms11628
   Liu PT, 2003, GENOME RES, V13, P476, DOI 10.1101/gr.749203
   Louis DN, 2016, ACTA NEUROPATHOL, V131, P803, DOI 10.1007/s00401-016-1545-1
   Lu TTH, 2018, CELL METAB, V27, P1294, DOI 10.1016/j.cmet.2018.04.013
   Mackay A, 2017, CANCER CELL, V32, P520, DOI 10.1016/j.ccell.2017.08.017
   Margueron R, 2011, NATURE, V469, P343, DOI 10.1038/nature09784
   Mohammad F, 2017, NAT MED, V23, P483, DOI 10.1038/nm.4293
   Nishino J, 2013, ELIFE, V2, DOI 10.7554/eLife.00924
   Orlando DA, 2014, CELL REP, V9, P1163, DOI 10.1016/j.celrep.2014.10.018
   Pajtler KW, 2018, ACTA NEUROPATHOL, V136, P211, DOI 10.1007/s00401-018-1877-0
   Pathania M, 2017, CANCER CELL, V32, P684, DOI 10.1016/j.ccell.2017.09.014
   Paugh BS, 2013, CANCER RES, V73, P6219, DOI 10.1158/0008-5472.CAN-13-1491
   Paugh BS, 2011, J CLIN ONCOL, V29, P3999, DOI 10.1200/JCO.2011.35.5677
   Paugh BS, 2010, J CLIN ONCOL, V28, P3061, DOI 10.1200/JCO.2009.26.7252
   Phillips HS, 2006, CANCER CELL, V9, P157, DOI 10.1016/j.ccr.2006.02.019
   Piunti A, 2017, NAT MED, V23, P493, DOI 10.1038/nm.4296
   Puget S, 2012, PLOS ONE, V7, DOI 10.1371/journal.pone.0030313
   Quinlan AR, 2010, BIOINFORMATICS, V26, P841, DOI 10.1093/bioinformatics/btq033
   Schwartzentruber J, 2012, NATURE, V482, P226, DOI [10.1038/nature10833, 10.1007/s13311-012-0137-6]
   Shechter D, 2007, NAT PROTOC, V2, P1445, DOI 10.1038/nprot.2007.202
   Subramanian A, 2005, P NATL ACAD SCI USA, V102, P15545, DOI 10.1073/pnas.0506580102
   Taylor KR, 2014, NAT GENET, V46, P457, DOI 10.1038/ng.2925
   Thompson CL, 2014, NEURON, V83, P309, DOI 10.1016/j.neuron.2014.05.033
   Thorvaldsdottir H, 2013, BRIEF BIOINFORM, V14, P178, DOI 10.1093/bib/bbs017
   Tronche F, 1999, NAT GENET, V23, P99, DOI 10.1038/12703
   Venneti S, 2013, BRAIN PATHOL, V23, P558, DOI 10.1111/bpa.12042
   Vo BT, 2017, CELL REP, V18, P2907, DOI 10.1016/j.celrep.2017.02.073
   Wu G, 2016, MODERN PATHOL, V29, P359, DOI 10.1038/modpathol.2016.37
   Wu G, 2014, NAT GENET, V46, P444, DOI 10.1038/ng.2938
   Yang M, 2015, DEVELOPMENT, V142, P1616, DOI 10.1242/dev.120543
   Zang CZ, 2009, BIOINFORMATICS, V25, P1952, DOI 10.1093/bioinformatics/btp340
   Zhang Y, 2014, J NEUROSCI, V34, P11929, DOI [10.1523/JNEUROSCI.1860-14.2014, 10.11772/j.issn.1001-9081.2014.07.1929]
   Zhang Y, 2008, GENOME BIOL, V9, DOI 10.1186/gb-2008-9-9-r137
   Zhou VW, 2011, NAT REV GENET, V12, P7, DOI 10.1038/nrg2905
   Zhu G, 2012, DEVELOPMENT, V139, P3422, DOI 10.1242/dev.083154
NR 58
TC 150
Z9 156
U1 1
U2 29
PU CELL PRESS
PI CAMBRIDGE
PA 50 HAMPSHIRE ST, FLOOR 5, CAMBRIDGE, MA 02139 USA
SN 1535-6108
EI 1878-3686
J9 CANCER CELL
JI Cancer Cell
PD JAN 14
PY 2019
VL 35
IS 1
BP 140
EP +
DI 10.1016/j.ccell.2018.11.015
PG 23
WC Oncology; Cell Biology
WE Science Citation Index Expanded (SCI-EXPANDED)
SC Oncology; Cell Biology
GA HH4TV
UT WOS:000455719400014
PM 30595505
OA Green Accepted, Bronze
DA 2023-08-27
ER

PT J
AU Walter, J
   Kapitza, S
   Krayenbuhl, N
   Tarnutzer, AA
AF Walter, Johannes
   Kapitza, Sandra
   Krayenbuehl, Niklaus
   Tarnutzer, Alexander A.
TI Clival Chondrosarcoma Associated With an Intra-Axial Cystic Medullary
   Lesion Responsive to Steroids
SO FRONTIERS IN NEUROLOGY
LA English
DT Article
DE medulla oblongata; inflammation; brainstem; surgery; clivus;
   chondrosarcoma; dexamethasone
ID OBLONGATA; TUMOR
AB Introduction: Here we present a 75-year-old patient who was admitted with acute-onset right-sided hemiparesis, dysphagia, dysarthria and nystagmus. Repeated MRI scans showed two lesions with contact to one another: one solid stationary extra-axial lesion at the caudal part of the clivus and a rapidly growing intra-axial cystic lesion at the level of the medulla oblongata. Biopsy of the solid lesion demonstrated a low-grade chondrosarcoma, while no tissue sample of the cystic lesion could be retrieved. After initiation of dexamethasone therapy the cystic lesion markedly regressed.
   Background: A literature search on published cases with the same combination of a stationary solid extra-axial mass at the caudal part of the clivus and a growing intra-axial cystic mass in the medulla oblongata was negative, indicating that the case described here is both unique and novel.
   Discussion: Considering the rapid progression of symptoms and growth on MR-imaging in combination with the marked response to steroids, an inflammatory response linked to the chondrosarcoma is most likely. At the same time other possible explanations as a second neoplasm, an abscess or an ischemic lesion seem unlikely.
   Concluding remarks: This case underlines an unusual complication of a rare brainstem tumor and outlines both the differential diagnosis and potential treatment options. For such cystic lesions in combination with chondrosarcoma, a treatment course with steroids should be considered along with surgical exploration necessary to obtain the diagnosis and for potential reduction of mass-effect on the medulla oblongata.
C1 [Walter, Johannes; Kapitza, Sandra; Krayenbuehl, Niklaus; Tarnutzer, Alexander A.] Univ Hosp Zurich, Dept Neurosurg, Zurich, Switzerland.
   [Krayenbuehl, Niklaus; Tarnutzer, Alexander A.] Univ Zurich, Fac Med, Zurich, Switzerland.
C3 University of Zurich; University Zurich Hospital; University of Geneva;
   University of Zurich
RP Tarnutzer, AA (通讯作者)，Univ Hosp Zurich, Dept Neurosurg, Zurich, Switzerland.; Tarnutzer, AA (通讯作者)，Univ Zurich, Fac Med, Zurich, Switzerland.
EM
RI Tarnutzer, Alexander/H-3236-2019
OI Tarnutzer, Alexander/0000-0002-6984-6958; Krayenbuehl,
   Niklaus/0000-0003-1089-221X
CR Arvinius C, 2016, SICOT-J, V2, DOI 10.1051/sicotj/2015042
   Cho JM, 2010, CHILD NERV SYST, V26, P853, DOI 10.1007/s00381-010-1109-y
   Fukushima T, 1998, NEUROL MED-CHIR, V38, P489, DOI 10.2176/nmc.38.489
   Hoosien M, 2011, J NEURO-ONCOL, V101, P135, DOI 10.1007/s11060-010-0216-7
   Inagaki Y, 2016, CLIN SARCOMA RES, V6, DOI 10.1186/s13569-016-0053-3
   Kalinski T, 2014, PATHOLOGE, V35, P249, DOI 10.1007/s00292-014-1968-1
   Prakkamakul S, 2017, J NEUROIMAGING, V27, P135, DOI 10.1111/jon.12361
   Preece MT, 2006, AM J NEURORADIOL, V27, P1211
   Royo A, 2013, SEMIN ULTRASOUND CT, V34, P153, DOI 10.1053/j.sult.2013.01.006
   Watanabe T, 2014, ACTA NEUROCHIR, V156, P1145, DOI 10.1007/s00701-014-2085-x
   Yoon W, 2007, EUR RADIOL, V17, P983, DOI 10.1007/s00330-006-0272-8
   Zhang D, 2010, KOREAN J RADIOL, V11, P269, DOI 10.3348/kjr.2010.11.3.269
NR 12
TC 0
Z9 0
U1 0
U2 2
PU FRONTIERS MEDIA SA
PI LAUSANNE
PA AVENUE DU TRIBUNAL FEDERAL 34, LAUSANNE, CH-1015, SWITZERLAND
SN 1664-2295
J9 FRONT NEUROL
JI Front. Neurol.
PD JUN 26
PY 2018
VL 9
AR 502
DI 10.3389/fneur.2018.00502
PG 4
WC Clinical Neurology; Neurosciences
WE Science Citation Index Expanded (SCI-EXPANDED)
SC Neurosciences & Neurology
GA GK6PM
UT WOS:000436310600001
PM 29997571
OA Green Published, gold, Green Accepted
DA 2023-08-27
ER

PT J
AU Zhang, S
   Wang, X
   Liu, XS
   Ju, Y
   Hui, XH
AF Zhang, Si
   Wang, Xiang
   Liu, Xuesong
   Ju, Yan
   Hui, Xuhui
TI Brainstem gangliogliomas: a retrospective series Clinical article
SO JOURNAL OF NEUROSURGERY
LA English
DT Article
DE brainstem; ganglioglioma; diagnosis; microsurgery; prognosis; oncology
ID SUPRATENTORIAL GANGLIOGLIOMAS; TUMOR RECURRENCE
AB Object. The authors retrospectively analyzed data on brainstem gangliogliomas treated in their department and reviewed the pertinent literature to foster understanding of the preoperative characteristics, management, and clinical outcomes of this disease.
   Methods. In 2006, the authors established a database of treated lesions of the posterior fossa. The epidemiology findings, clinical presentations, radiological investigations, pathological diagnoses, management, and prognosis for brainstem gangliogliomas were retrospectively analyzed.
   Results. Between 2006 and 2012, 7 patients suffering from brainstem ganglioglioma were treated at the West China Hospital of Sichuan University. The mean age of the patients, mean duration of symptoms prior to diagnosis, and mean duration of follow-up were 28.6 years, 19.4 months, and 38.1 months, respectively. The main presentations were progressive cranial nerve deficits and cerebellar signs. Subtotal resection was achieved in 2 patients, and partial resection in 5. All tumors were pathologically diagnosed as WHO Grade I or II ganglioglioma. Radiotherapy and adjuvant chemotherapy were not administered. After 21-69 months of follow-up, patient symptoms were resolved or stable without aggravation, and MRI showed that the size of residual lesions was unchanged without progression or recurrence.
   Conclusions. The diagnosis of brainstem ganglioglioma is of great importance given its favorable prognosis. The authors recommend the maximal safe resection followed by close observation without adjuvant therapy as the optimal treatment for this disease. (http://thejns.org/doi/abs/10.3171/2013.1.JNS121323)
C1 [Zhang, Si; Wang, Xiang; Liu, Xuesong; Ju, Yan; Hui, Xuhui] Sichuan Univ, West China Hosp, Dept Neurosurg, Chengdu, Peoples R China.
C3 Sichuan University
RP Hui, XH (通讯作者)，Sichuan Univ, West China Hosp, Dept Neurosurg, 37 Guo Xue Xiang, Chengdu, Peoples R China.
EM
CR Akiyama H, 2006, NEUROL SURG TOKYO, V34, P1255
   Baussard B, 2007, J NEUROSURG, V107, P286, DOI 10.3171/PED-07/10/286
   Cultrera F, 2006, J CLIN NEUROSCI, V13, P963, DOI 10.1016/j.jocn.2005.09.011
   Gleckman AM, 1998, AM J FOREN MED PATH, V19, P303, DOI 10.1097/00000433-199812000-00002
   Im SH, 2002, J NEURO-ONCOL, V59, P173, DOI 10.1023/A:1019661528350
   Johnson JH, 1997, PEDIATR NEUROSURG, V27, P203, DOI 10.1159/000121252
   Lagares A, 2001, SURG NEUROL, V56, P315, DOI 10.1016/S0090-3019(01)00618-8
   LANG FF, 1993, J NEUROSURG, V79, P867, DOI 10.3171/jns.1993.79.6.0867
   Lobel U, 2011, CLIN RADIOL, V66, P194, DOI 10.1016/j.crad.2010.10.005
   Luyken C, 2004, CANCER, V101, P146, DOI 10.1002/cncr.20332
   Majores M, 2008, CANCER-AM CANCER SOC, V113, P3355, DOI 10.1002/cncr.23965
   Mpairamidis E, 2008, J CHILD NEUROL, V23, P1481, DOI 10.1177/0883073808319316
   Pandita A, 2007, NEURO-ONCOLOGY, V9, P124, DOI 10.1215/15228517-2006-029
   Rades D, 2010, CANCER-AM CANCER SOC, V116, P432, DOI 10.1002/cncr.24716
   Rumana CS, 1999, ACTA NEUROCHIR, V141, P63, DOI 10.1007/s007010050267
   Rumana CS, 1998, NEUROSURGERY, V42, P1038, DOI 10.1097/00006123-199805000-00049
   Safavi-Abbasi S, 2007, SKULL BASE-INTERD AP, V17, P253, DOI 10.1055/s-2007-984486
   Siddique K, 2002, NEUROSURGERY, V50, P1133, DOI 10.1097/00006123-200205000-00034
   Westwood DA, 2009, J CLIN NEUROSCI, V16, P108, DOI 10.1016/j.jocn.2007.11.016
   ZENTNER J, 1994, J NEUROL NEUROSUR PS, V57, P1497, DOI 10.1136/jnnp.57.12.1497
NR 20
TC 26
Z9 29
U1 0
U2 4
PU AMER ASSOC NEUROLOGICAL SURGEONS
PI ROLLING MEADOWS
PA 5550 MEADOWBROOK DRIVE, ROLLING MEADOWS, IL 60008 USA
SN 0022-3085
J9 J NEUROSURG
JI J. Neurosurg.
PD APR
PY 2013
VL 118
IS 4
BP 884
EP 888
DI 10.3171/2013.1.JNS121323
PG 5
WC Clinical Neurology; Surgery
WE Science Citation Index Expanded (SCI-EXPANDED)
SC Neurosciences & Neurology; Surgery
GA 114BT
UT WOS:000316715700033
PM 23373800
DA 2023-08-27
ER

PT J
AU Zhang, L
   Yu, HW
   Xue, YX
   Liu, YH
AF Zhang, Lei
   Yu, Hongwei
   Xue, Yixue
   Liu, Yunhui
TI Decreased natural killer cells in diffuse intrinsic pontine glioma
   patients
SO CHILDS NERVOUS SYSTEM
LA English
DT Article
DE Natural killer cell; NK; Diffuse intrinsic pontine glioma; DIPG
AB Purpose Diffuse intrinsic pontine glioma (DIPG) remains the leading cause of death among pediatric brain tumor patients. Its pontine location and aggressive nature make developing effective treatment an ongoing challenge in pediatric oncology. Although studies have found that one of the hallmark features of glioma is immunosuppression, the immune status of DIPG patient is not clearly understood. Methods We tested the lymphocyte profile in four radiologically diagnosed DIPG patients. All the four patients did not receive any steroids, radiotherapy, and chemotherapy before the collection of blood. Results We found decreased natural killer (NK) cell level and increased B cell level in all four cases. Conclusion These findings suggested that decreased NK cells and increased B cells may aid the tumorigenesis and growth seen in DIPG patient. Increased NK and decreased B cells may be the future direction for the treatment of DIPG patient.
C1 [Zhang, Lei; Yu, Hongwei; Liu, Yunhui] China Med Univ, Dept Neurosurg, Shengjing Hosp, 36 Heping Dist, Shenyang 110004, Liaoning, Peoples R China.
   [Zhang, Lei; Liu, Yunhui] Liaoning Res Ctr Translat Med Nervous Syst Dis, Shenyang, Peoples R China.
   [Zhang, Lei; Liu, Yunhui] Key Lab Neurooncol Liaoning Prov, Shenyang, Peoples R China.
   [Xue, Yixue] China Med Univ, Sch Life Sci, Dept Neurobiol, Shenyang, Peoples R China.
C3 China Medical University; China Medical University
RP Liu, YH (通讯作者)，China Med Univ, Dept Neurosurg, Shengjing Hosp, 36 Heping Dist, Shenyang 110004, Liaoning, Peoples R China.; Liu, YH (通讯作者)，Liaoning Res Ctr Translat Med Nervous Syst Dis, Shenyang, Peoples R China.; Liu, YH (通讯作者)，Key Lab Neurooncol Liaoning Prov, Shenyang, Peoples R China.
EM
CR Lieberman NAP, 2019, NEURO-ONCOLOGY, V21, P83, DOI 10.1093/neuonc/noy145
   Qin ZH, 1998, NAT MED, V4, P627, DOI 10.1038/nm0598-627
NR 2
TC 4
Z9 4
U1 0
U2 1
PU SPRINGER
PI NEW YORK
PA ONE NEW YORK PLAZA, SUITE 4600, NEW YORK, NY, UNITED STATES
SN 0256-7040
EI 1433-0350
J9 CHILD NERV SYST
JI Childs Nerv. Syst.
PD JUL
PY 2020
VL 36
IS 7
BP 1345
EP 1346
DI 10.1007/s00381-020-04665-9
EA MAY 2020
PG 2
WC Clinical Neurology; Pediatrics; Surgery
WE Science Citation Index Expanded (SCI-EXPANDED)
SC Neurosciences & Neurology; Pediatrics; Surgery
GA LY8TW
UT WOS:000533176100001
PM 32418048
DA 2023-08-27
ER

PT J
AU Turner, CD
   Gururangan, S
   Eastwood, J
   Bottom, K
   Watral, M
   Beason, R
   McLendon, RE
   Friedman, AH
   Tourt-Uhlig, S
   Miller, LL
   Friedman, HS
AF Turner, CD
   Gururangan, S
   Eastwood, J
   Bottom, K
   Watral, M
   Beason, R
   McLendon, RE
   Friedman, AH
   Tourt-Uhlig, S
   Miller, LL
   Friedman, HS
TI Phase II study of irinotecan (CPT-11) in children with high-risk
   malignant brain tumors: The Duke experience
SO NEURO-ONCOLOGY
LA English
DT Article
ID STEM-CELL RESCUE; DRUG-RESISTANCE; CLINICAL-TRIALS; ACTIVE EFFLUX;
   RECURRENT; XENOGRAFT; SCHEDULE; SN-38; MEDULLOBLASTOMA; INHIBITION
AB A phase II study of irinotecan (CPT-11) was conducted at Duke University Medical Center, Durham, NC, to evaluate the activity of this agent in children with high-risk malignant brain tumors. A total of 22 children were enrolled in this study, including 13 with histologically verified recurrent malignant brain tumors (glioblastoma multiforme [GBM] 4, anaplastic astrocytoma 1, ependymoma 5, and medulloblastoma/primitive neuroectodermal tumor 3), 5 with recurrent diffuse pontine glioma, and 4 with newly diagnosed GBM. All patients with recurrent tumor had prior chemotherapy and/or irradiation. Each course of CPT-11 consisted of 125 mg/m(2) per week given i.v. for 4 weeks followed by a 2-week rest period. Patients with recurrent tumors received therapy until disease progression or unacceptable toxicity. Patients with newly diagnosed tumors initially received 3 cycles of treatment to assess tumor response and then were allowed radiotherapy at physician's choice; patients who demonstrated a response to CPT-11 prior to radiotherapy were allowed to continue the drug after radiation until disease progression or unacceptable toxicity. A 25% to 50% close reduction was made for grade III-IV toxicity. Responses were assessed after every course by gadolinium-enhanced MRI of the brain and spine. Twenty-two patients received a median of 2 courses of CPT-11 (range, 1-16). Responses were seen in 4 of 9 patients with GBM or anaplastic astrocytoma (44%; 95% confidence interval, 11%-82%) (complete response in 2 patients with recurrent GBM lasting 9 months and 48+ months; partial response in one patient with a newly diagnosed midbrain GBM lasting 18 months prior to radiotherapy; and partial response lasting 11 months in I patient with recurrent anaplastic astrocytoma), 1 of 5 patients with recurrent ependymoma (partial response initially followed by stable disease lasting 11 months), and none of 5 patients with recurrent diffuse pontine glioma. Two of 3 patients with medulloblastoma/primitive neuroectodermal tumor had stable disease for 9 and 13 months. Toxicity was mainly myelosuppression, with 12 of 22 patients (50%) suffering grade II-IV neutropenia. Seven patients required dose reduction secondary to neutropenia. CPT-11, given in this schedule, appears to be active in children with malignant glioma, medulloblastoma, and ependymoma with acceptable toxicity. Ongoing studies will demonstrate if activity of CPT-11 can be enhanced when combined with alkylating agents, including carmustine and temozolomide.
C1 Duke Univ, Med Ctr, Brain Tumor Ctr, Durham, NC 27710 USA.
   Duke Univ, Med Ctr, Div Neuroradiol, Durham, NC 27710 USA.
   Duke Univ, Med Ctr, Div Pathol, Durham, NC 27710 USA.
   Duke Univ, Med Ctr, Div Neurosurg, Durham, NC 27710 USA.
   Pharmacia & Upjohn Inc, Kalamazoo, MI 49001 USA.
C3 Duke University; Duke University; Duke University; Duke University;
   Pfizer
RP Gururangan, S (通讯作者)，Duke Univ, Med Ctr, Brain Tumor Ctr, Box 3624, Durham, NC 27710 USA.
CR Benedetti MS, 2000, FUND CLIN PHARMACOL, V14, P301, DOI 10.1111/j.1472-8206.2000.tb00411.x
   Blaney S, 2001, CLIN CANCER RES, V7, P32
   Chen ZS, 1999, CANCER LETT, V138, P13, DOI 10.1016/S0304-3835(98)00367-X
   Chen ZS, 1999, MOL PHARMACOL, V55, P921
   Chu XY, 1999, J PHARMACOL EXP THER, V288, P735
   Cogger CG, 1998, J PROD AGRIC, V11, P41, DOI 10.2134/jpa1998.0041
   Dunkel IJ, 1998, J CLIN ONCOL, V16, P222, DOI 10.1200/JCO.1998.16.1.222
   FEUN LG, 1994, J NEURO-ONCOL, V20, P165, DOI 10.1007/BF01052726
   Finlay J L, 1999, Pediatr Transplant, V3 Suppl 1, P87
   Finlay JL, 1996, J CLIN ONCOL, V14, P2495, DOI 10.1200/JCO.1996.14.9.2495
   Friedman HS, 1999, J CLIN ONCOL, V17, P1516, DOI 10.1200/JCO.1999.17.5.1516
   Furman WL, 1999, J CLIN ONCOL, V17, P1815, DOI 10.1200/JCO.1999.17.6.1815
   Graham ML, 1997, J CLIN ONCOL, V15, P1814, DOI 10.1200/JCO.1997.15.5.1814
   Hare CB, 1997, CANCER CHEMOTH PHARM, V39, P187
   HENSON JW, 1992, J NEURO-ONCOL, V14, P37
   Houghton PJ, 2000, CLIN CANCER RES, V6, P4110
   Houghton PJ, 1998, ONCOLOGY-NY, V12, P84
   KAWATO Y, 1991, CANCER RES, V51, P4187
   Kono A., 1991, JPN J CANC CHEMOTHER, V18, P2175
   LEVY RH, 1995, EPILEPSIA, V36, pS8, DOI 10.1111/j.1528-1157.1995.tb06007.x
   Patel VJ, 2000, CLIN CANCER RES, V6, P4154
   PHILLIPS PC, 1991, NEUROL CLIN, V9, P383, DOI 10.1016/S0733-8619(18)30292-5
   Pommier Y, 1994, Adv Pharmacol, V29B, P73
   Pourquier P, 2001, CANCER RES, V61, P53
   Ratain MJ, 2000, CLIN CANCER RES, V6, P3393
   Rothenberg ML, 1998, ONCOLOGY-NY, V12, P68
   Santos A, 2000, CLIN CANCER RES, V6, P2012
   SLICHENMYER WJ, 1994, CANC CHEMOTHER PHA S, V34, P53
   Vassal G, 1998, BIOCHIMIE, V80, P271, DOI 10.1016/S0300-9084(98)80009-6
   Vassal G, 1997, INT J CANCER, V73, P156, DOI 10.1002/(SICI)1097-0215(19970926)73:1<156::AID-IJC24>3.0.CO;2-D
   VonHoff D, 1996, EUR J CANCER, V32A, pS9, DOI 10.1016/0959-8049(96)00291-2
NR 31
TC 60
Z9 60
U1 0
U2 3
PU CARDEN JENNINGS PUBL CO LTD
PI CHARLOTTESVILLE
PA BLAKE CTR, STE 200, 1224 W MAIN ST, CHARLOTTESVILLE, VA 22903 USA
SN 1522-8517
J9 NEURO-ONCOLOGY
JI Neuro-Oncology
PD APR
PY 2002
VL 4
IS 2
BP 102
EP 108
PG 7
WC Oncology; Clinical Neurology
WE Science Citation Index Expanded (SCI-EXPANDED)
SC Oncology; Neurosciences & Neurology
GA 534RJ
UT WOS:000174600300004
PM 11916501
OA Green Published, Bronze
DA 2023-08-27
ER

PT J
AU JACKLER, RK
   SIM, DW
   GUTIN, PH
   PITTS, LH
AF JACKLER, RK
   SIM, DW
   GUTIN, PH
   PITTS, LH
TI SYSTEMATIC-APPROACH TO INTRADURAL TUMORS VENTRAL TO THE BRAIN-STEM
SO AMERICAN JOURNAL OF OTOLOGY
LA English
DT Article
ID RETROLABYRINTHINE TRANSTENTORIAL APPROACH; CEREBELLOPONTINE ANGLE; SKULL
   BASE; TRANSCOCHLEAR APPROACH; TRANSLABYRINTHINE APPROACH; PETROCLIVAL
   MENINGIOMAS; TRANSCONDYLAR APPROACH; HEARING PRESERVATION; PETROSAL
   APPROACH; ANTERIOR PORTION
AB Intradural tumors that are situated anterior to the midbrain, pons, and medulla have historically been among the most inaccessible of all intracranial lesions.  The classic approaches to the posterior fossa (e.g., suboccipita, retrosigmoid) provide only limited access to the anterior midline, primarily due to interposition of the cerebellum, brain stem, and numerous cranial nerves between the tumor and the viewpoint of the surgeon.  A variety of techniques have been developed in recent years that create a craniotomy by removal of a portion of the lateral skull base.  These procedures enhance exposure of the ventral surface of the brain stem while markedly reducing the need for brain retraction.  An underlying theme of transbasal craniotomy is judicious removal of a portion of the petrous pyramid.  The most radical form of petrosectomy, the extended transcochlear approach, involves removal of the entire petrous pyramid along with the lateral aspect of the clivus.  This provides an unimpeded view of the ventral surface of the pons, including the basilar artery, vertebrobasilar junction, and both abducens nerves.  Whereas this technique provides splendid exposure along the midsegment of the brain stem, it carries substantial morbidity, including hearing loss and transient facial palsy, which typically recovers incompletely and with synkinesis.  Over the past few years transcochlear procedures have been gradually supplanted, at the University of California Medical Center, by techniques that involve creating a simultaneous craniotomy of both the middle and posterior fossae fashioned around a more limited petrosectomy.  These versatile procedures, in particular the middle fossa/retrolabyrinthine approach, provide excellent exposure of the region ventral to the midbrain and pons with less morbidity than the transcochlear approach.  When tumors extend inferiorly, ventral to the lower medulla and/or upper cervical spinal cord, augmented inferior exposure is required.  Approaches to ventrally situated lesions at the craniovertebral junction include the far lateral (transcondylar) approach to the foramen magnum and the transjugular approach, both of which involve removal of the inferior portion of the petrous bone.  To efficiently utilize these innovative surgical options the surgeon must decide which of the potential approaches optimizes resection while minimizing morbidity.  An analysis of the anatomy of the tumor, the functional integrity of cranial nerves, and the extent of resection planned provides the surgeon with the information needed to arrive at a rational choice.
C1 UNIV CALIF SAN FRANCISCO,MED CTR,DEPT OTOLARYNGOL HEAD & NECK SURG,SAN FRANCISCO,CA 94143.
   UNIV CALIF SAN FRANCISCO,MED CTR,DEPT NEUROL SURG,SAN FRANCISCO,CA 94143.
C3 University of California System; University of California San Francisco;
   University of California System; University of California San Francisco
OI jackler, robert/0000-0003-2525-8377
CR Al-Mefty O, 1991, MENINGIOMAS, P517
   ALMEFTY O, 1988, NEUROSURGERY, V22, P510, DOI 10.1227/00006123-198803000-00010
   AMMIRATI M, 1993, SKULL BASE SURG, V3, P193, DOI 10.1055/s-2008-1060584
   AMMIRATI M, 1992, SKULL BASE SURG, V2, P124, DOI 10.1055/s-2008-1057122
   ANDREWS RJ, 1993, NEUROSURGERY, V33, P1052
   ARRIAGA MA, 1993, LARYNGOSCOPE, V103, P693
   BERTALANFFY H, 1991, NEUROSURGERY, V29, P815, DOI 10.1227/00006123-199112000-00002
   CANALIS RF, 1991, LARYNGOSCOPE, V101, P6
   CANNALIS RF, 1993, LARYNGOSCOPE, V103, P343
   CHEUNG S, 1995, IN PRESS AM J OTOL
   DASPIT CP, 1991, OTOLARYNG HEAD NECK, V105, P788, DOI 10.1177/019459989110500604
   De la Cruz A, 1981, Rev Laryngol Otol Rhinol (Bord), V102, P33
   GEORGE B, 1988, SURG NEUROL, V29, P484, DOI 10.1016/0090-3019(88)90145-0
   HAKUBA A, 1988, SURG NEUROL, V30, P108, DOI 10.1016/0090-3019(88)90095-X
   HEROS RC, 1986, J NEUROSURG, V64, P559, DOI 10.3171/jns.1986.64.4.0559
   HERZOG JA, 1991, OTOLARYNGOL HEAD NEC, V104, P103
   HIRSCH BE, 1993, AM J OTOL, V14, P533
   HORN KL, 1991, OTOLARYNG HEAD NECK, V104, P37, DOI 10.1177/019459989110400108
   HOUSE WF, 1986, AM J OTOL, V7, P1
   HOUSE WF, 1976, ARCH OTOLARYNGOL, V102, P334
   KAWASE T, 1991, NEUROSURGERY, V28, P869, DOI 10.1227/00006123-199106000-00014
   KING WA, 1993, J NEUROSURG, V79, P508, DOI 10.3171/jns.1993.79.4.0508
   KINNEY SE, 1992, AM J OTOL, V13, P426
   LANSER MJ, 1992, NEUROMONITORING OTOL, P131
   MAYBERG MR, 1986, J NEUROSURG, V65, P160, DOI 10.3171/jns.1986.65.2.0160
   MCELVEEN JT, 1991, J LARYNGOL OTOL, V105, P34, DOI 10.1017/S0022215100114768
   MILLER CG, 1993, NEUROSURGERY, V33, P461
   PELLET W, 1988, J NEUROSURG, V69, P887, DOI 10.3171/jns.1988.69.6.0887
   SAKAKI S, 1987, SURG NEUROL, V28, P245, DOI 10.1016/0090-3019(87)90301-6
   SAMII M, 1988, ACTA NEUROCHIR, V95, P6, DOI 10.1007/BF01793075
   SASAKI CT, 1977, ARCH OTOLARYNGOL, V103, P730
   SEN CN, 1990, NEUROSURGERY, V27, P197, DOI 10.1227/00006123-199008000-00004
   SHIMAMURA Y, 1982, Neurological Surgery, V10, P263
   SPETZLER RF, 1991, SKULL BASE SURG, V1, P226, DOI 10.1055/s-2008-1057102
   Spetzler RF, 1990, BNI Q, V6, P35
   SPETZLER RF, 1992, J NEUROSURG, V76, P6588
   THEDINGER BA, 1992, AM J OTOL, V13, P408
   YINGLING CD, 1994, NEUROTOLOGY, P967
NR 38
TC 19
Z9 20
U1 0
U2 1
PU LIPPINCOTT-RAVEN PUBL
PI PHILADELPHIA
PA 227 EAST WASHINGTON SQ, PHILADELPHIA, PA 19106
SN 0192-9763
J9 AM J OTOL
JI Am. J. Otol.
PD JAN
PY 1995
VL 16
IS 1
BP 39
EP 51
PG 13
WC Otorhinolaryngology
WE Science Citation Index Expanded (SCI-EXPANDED)
SC Otorhinolaryngology
GA QA480
UT WOS:A1995QA48000009
PM 8579176
DA 2023-08-27
ER

PT J
AU Stanton, SG
   Cashman, MZ
AF Stanton, SG
   Cashman, MZ
TI Auditory brainstem response - A comparison of different interpretation
   strategies for detection of cerebellopontine angle tumors
SO SCANDINAVIAN AUDIOLOGY
LA English
DT Article
DE ABR; auditory brainstem response; BAER; correction factor; evoked
   potential
ID SENSORINEURAL HEARING-LOSS; STEM RESPONSE; EVOKED-POTENTIALS; ACOUSTIC
   NEUROMAS; 2000-4000 HZ; DIAGNOSIS; AGE; ABR; SENSITIVITY; AUDIOMETRY
AB In this retrospective study (111 tumor; 1370 non-tumor patients), the effectiveness of the auditory brainstem response for identifying cerebellopontine angle tumors was studied using different criteria to interpret the test. Individual ABR features were examined. The I-V was best (sensitivity = 82%; specificity = 97%) but present for only 44% of the population I-III (sensitivity = 50%; specificity = 96%) and III-V (sensitivity = 30%; specificity = 97%) were worst. Ninety-three percent of patients had V and ILDV, with good results (sensitivity = 74% and 81% respectively; specificity = 93% each). Three interpretation strategies were compared: (I)included the latencies of V, ILDV, I-V, III-V and I-III and ILDI-V, (2) excluded III-V and I-III, (3) used I-V and ILDI-V when available; V and ILDV otherwise. When corrected for hearing loss, strategy 3 provided significantly better specificity (91%) than strategy 1 (87%), with no significant difference in sensitivity (92% and 93%, respectively).
RP Stanton, SG (通讯作者)，SUNNYBROOK HLTH SCI CTR,AUDIOL SERV,ROOM A210,2075 BAYVIEW AVE,TORONTO,ON M4N 3M5,CANADA.
CR ALLISON T, 1983, ELECTROEN CLIN NEURO, V55, P619, DOI 10.1016/0013-4694(83)90272-9
   BARRS DM, 1985, ARCH OTOLARYNGOL, V111, P17
   BARRS DM, 1987, OTOLARYNG HEAD NECK, V96, P523, DOI 10.1177/019459988709600601
   BAUCH CD, 1986, EAR HEARING, V7, P314, DOI 10.1097/00003446-198610000-00004
   BAUCH CD, 1988, AUDIOLOGY, V27, P156
   BAUCH CD, 1987, EAR HEARING, V8, P184, DOI 10.1097/00003446-198706000-00010
   BAUCH CD, 1982, EAR HEARING, V3, P83, DOI 10.1097/00003446-198203000-00006
   CASHMAN MZ, 1983, J OTOLARYNGOL, V12, P180
   CASHMAN MZ, 1983, SCAND AUDIOL, V12, P35, DOI 10.3109/01050398309076222
   CASHMAN MZ, 1993, SCAND AUDIOL, V22, P153, DOI 10.3109/01050399309047461
   CHU NS, 1985, ELECTROEN CLIN NEURO, V62, P431, DOI 10.1016/0168-5597(85)90053-X
   CLEMIS JD, 1979, LARYNGOSCOPE, V89, P31
   EGGERMONT JJ, 1980, ANN OTO RHINOL LARYN, V89, P1
   ELBERLING C, 1987, SCAND AUDIOL, V16, P49, DOI 10.3109/01050398709042155
   FERRARO JA, 1989, EAR HEARING, V10, P161, DOI 10.1097/00003446-198906000-00004
   GSTOETTNER W, 1992, EUR ARCH OTO-RHINO-L, V249, P253, DOI 10.1007/BF00714487
   HART RG, 1983, NEUROLOGY, V33, P211, DOI 10.1212/WNL.33.2.211
   HYDE ML, 1981, J OTOLARYNGOL, V10, P117
   HYDE ML, 1994, CURR OPIN OTOLARYNGO, V2, P177
   JACOBSON GP, 1994, PRINCIPLES APPL AUDI
   JERGER J, 1988, EAR HEARING, V9, P168, DOI 10.1097/00003446-198808000-00002
   MOLLER MB, 1983, ANN OTO RHINOL LARYN, V92, P645, DOI 10.1177/000348948309200622
   MUSIEK FE, 1986, EAR HEARING, V7, P100, DOI 10.1097/00003446-198604000-00008
   MUSIEK FE, 1983, AM J OTOL, V4, P281
   Prasher D K, 1983, Br J Audiol, V17, P163, DOI 10.3109/03005368309107881
   PROSSER S, 1987, AUDIOLOGY, V25, P179
   ROBINSON K, 1983, J NEUROL SCI, V60, P1, DOI 10.1016/0022-510X(83)90123-5
   ROSENHALL U, 1986, SCAND AUDIOL, V15, P179, DOI 10.3109/01050398609042141
   ROSENHALL U, 1985, ELECTROEN CLIN NEURO, V62, P426, DOI 10.1016/0168-5597(85)90052-8
   ROSENHAMER HJ, 1981, SCAND AUDIOL, V10, P3, DOI 10.3109/01050398109076156
   SELTERS WA, 1977, ARCH OTOLARYNGOL, V103, P181
   SELTERS WA, 1979, ACOUSTIC TUMORS
   STOCKARD J J, 1978, American Journal of EEG Technology, V18, P177
   STURZEBECHER E, 1985, SCAND AUDIOL, V14, P83, DOI 10.3109/01050398509045927
   STURZEBECHER E, 1987, SCAND AUDIOL, V16, P153, DOI 10.3109/01050398709042170
   TERKILDSEN K, 1981, SCAND AUDIOL S13, V10, P103
   ZOLLNER C, 1987, ORL J OTO-RHINO-LARY, V49, P123
   ZOLLNER C, 1981, ARCH OTO-RHINO-LARYN, V230, P135, DOI 10.1007/BF00456143
NR 38
TC 5
Z9 5
U1 0
U2 2
PU SCANDINAVIAN UNIVERSITY PRESS
PI OSLO
PA PO BOX 2959 TOYEN, JOURNAL DIVISION CUSTOMER SERVICE, N-0608 OSLO,
   NORWAY
SN 0105-0397
J9 SCAND AUDIOL
JI Scand. Audiol.
PY 1996
VL 25
IS 2
BP 109
EP 120
DI 10.3109/01050399609047992
PG 12
WC Otorhinolaryngology
WE Science Citation Index Expanded (SCI-EXPANDED)
SC Otorhinolaryngology
GA UK172
UT WOS:A1996UK17200005
PM 8738636
DA 2023-08-27
ER

PT J
AU Hankinson, TC
   Campagna, EJ
   Foreman, NK
   Handler, MH
AF Hankinson, Todd C.
   Campagna, Elizabeth J.
   Foreman, Nicholas K.
   Handler, Michael H.
TI Interpretation of magnetic resonance images in diffuse intrinsic pontine
   glioma: a survey of pediatric neurosurgeons Clinical article
SO JOURNAL OF NEUROSURGERY-PEDIATRICS
LA English
DT Article
DE diffuse intrinsic pontine glioma; pediatric brain tumor; surgical
   practice survey
ID BRAIN-STEM GLIOMAS; SHOULD-NOT; CHILDREN; BIOPSY
AB Object. The current management paradigm for children with newly diagnosed diffuse intrinsic pontine glioma (DIPG) is to establish a diagnosis and begin therapy based on MR imaging findings correlated with an appropriate clinical presentation, and without a tissue diagnosis. This strategy assumes that pediatric neurosurgeons and neurooncologists uniformly interpret MR imaging findings in this population. This study sought to examine the consistency of North American pediatric neurosurgeons in assessing MR images in this patient population, and in their surgical plans based on the interpretation of those images.
   Methods. The authors created an online survey and invited all members of the American Society of Pediatric Neurosurgeons and the American Association of Neurological Surgeons/Congress of Neurological Surgeons Joint Section for Pediatric Neurosurgery to participate. The survey consisted of 58 questions, and 48 pertained to representative MR images from 16 children who presented to The Children's Hospital of Colorado with diffuse pontine tumors. Based on the imaging presented and a standard clinical scenario, the respondent was asked if she or he believed a lesion to be "typical" or "atypical," whether she or he would biopsy the lesion, and what surgical approach would be chosen. The remaining 10 questions pertained to respondent demographics and his or her practice regarding tissue preservation and interest in participating in a multicenter trial that included tumor biopsy in selected cases. Rates at which each lesion was considered to be typical or atypical and rates of recommended biopsy were calculated.
   Results. Surveys were received by 269 individuals. Eighty-six responses were received (32.0%). No tumor was judged to be either typical or atypical by all respondents. Individual surgeons varied widely regarding how many of the tumors were judged as typical or warranted a biopsy. The percentage of respondents who disagreed with the majority opinion regarding whether a tumor qualified as typical ranged from 2.3% to 48.8%, with a median of 28.6%. More than 75% agreement regarding whether a tumor was typical or atypical was found in 7 (43.8%) of 16 cases. The K statistic regarding typicality was 0.297 +/- 0.0004 (mean +/- SEM), implying only fair agreement. For every tumor, at least 1 respondent who believed the lesion to be typical in appearance would nevertheless biopsy the lesion. Of those respondents who considered a lesion to be typical, a median of 5.1% (range 1.2%-66.7%) would choose to biopsy this lesion. Of those respondents who considered a lesion atypical, a median of 18.3% (range 3.7%-100%) would choose not to biopsy the lesion. Of 85 responses to the question, "Would you be willing to biopsy a typical diffuse pontine glioma as a part of a multicenter trial?," 59 (69.4%) of 85 respondents answered that they would.
   Conclusions. Although making a diagnosis based on radiographic evidence alone represents a well-established management paradigm for children with suspected DIPG, this study demonstrates considerable inconsistency on the part of pediatric neurosurgeons in the application of this strategy to individual patients. As such, the practice of diagnosing DIPG based on imaging characteristics and clinical history alone does not reach the appropriate threshold to be considered a standard of care. (DOI: 10.3171/2011.4.PEDS1180)
C1 [Hankinson, Todd C.; Handler, Michael H.] Univ Colorado, Childrens Hosp Colorado, Div Pediat Neurosurg, Denver, CO 80202 USA.
   [Hankinson, Todd C.; Campagna, Elizabeth J.] Univ Colorado, Childrens Hosp Colorado, Childrens Outcomes Res Program, Denver, CO 80202 USA.
   [Foreman, Nicholas K.] Univ Colorado, Dept Pediat, Div Neurooncol, Childrens Hosp Colorado, Denver, CO 80202 USA.
C3 Children's Hospital Colorado; University of Colorado System; University
   of Colorado Denver; Children's Hospital Colorado; University of Colorado
   System; University of Colorado Denver; Children's Hospital Colorado;
   University of Colorado System; University of Colorado Denver
RP Handler, MH (通讯作者)，Univ Colorado Denver, Childrens Hosp Colorado, Dept Pediat Neurosurg, 13123 E 16th Ave,Box 330, Aurora, CO 80045 USA.
EM
OI Foreman, Nicholas/0000-0003-4848-4801
CR ALBRIGHT AL, 1993, NEUROSURGERY, V33, P1030
   BARKOVICH AJ, 1991, PEDIATR NEUROSURG, V16, P73, DOI DOI 10.1159/000120511]
   Brodbelt A, 2008, BRIT J NEUROSURG, V22, P625, DOI 10.1080/02688690802482938
   de Leon FCP, 2003, CHILD NERV SYST, V19, P305, DOI 10.1007/s00381-003-0737-x
   Dunkel IJ, 2009, J NEUROS-PEDIATR, V3, P257, DOI 10.3171/2008.12.PEDS08388
   Fangusaro J, 2009, J CHILD NEUROL, V24, P1409, DOI 10.1177/0883073809338960
   Frazier JL, 2009, J NEUROSURG-PEDIATR, V3, P259, DOI 10.3171/2008.11.PEDS08281
   Handler MH, 2010, J NEUROS-PEDIATR, V5, P140, DOI 10.3171/2009.5.PEDS09265
   Hargrave D, 2008, J NEURO-ONCOL, V86, P313, DOI 10.1007/s11060-007-9473-5
   Hargrave D, 2008, BRIT J NEUROSURG, V22, P624, DOI 10.1080/02688690802484405
   Hargrove D, 2006, LANCET ONCOL, V7, P241, DOI 10.1016/S1470-2045(06)70615-5
   Hayward RM, 2008, J NEURO-ONCOL, V90, P57, DOI 10.1007/s11060-008-9631-4
   Leach PA, 2008, BRIT J NEUROSURG, V22, P619, DOI 10.1080/02688690802366198
   Liu AK, 2009, PEDIATR RADIOL, V39, P1317, DOI 10.1007/s00247-009-1368-5
   Roujeau T, 2007, J NEUROSURG, V107, P1, DOI 10.3171/PED-07/07/001
   Wilkinson R, 2008, BRIT J NEUROSURG, V22, P617, DOI 10.1080/02688690802482896
   Zimmerman RA, 1996, PEDIATR NEUROSURG, V25, P45, DOI 10.1159/000121096
NR 17
TC 46
Z9 48
U1 0
U2 2
PU AMER ASSOC NEUROLOGICAL SURGEONS
PI ROLLING MEADOWS
PA 5550 MEADOWBROOK DRIVE, ROLLING MEADOWS, IL 60008 USA
SN 1933-0707
J9 J NEUROSURG-PEDIATR
JI J. Neurosurg.-Pediatr.
PD JUL
PY 2011
VL 8
IS 1
BP 97
EP 102
DI 10.3171/2011.4.PEDS1180
PG 6
WC Clinical Neurology; Pediatrics; Surgery
WE Science Citation Index Expanded (SCI-EXPANDED)
SC Neurosciences & Neurology; Pediatrics; Surgery
GA 782JQ
UT WOS:000292005000015
PM 21721895
DA 2023-08-27
ER

PT J
AU Furtak, J
   Sledzinska, P
   Bebyn, MG
   Szylberg, T
   Krajewski, S
   Birski, M
   Harat, M
AF Furtak, Jacek
   Sledzinska, Paulina
   Bebyn, Marek G.
   Szylberg, Tadeusz
   Krajewski, Stanislaw
   Birski, Marcin
   Harat, Marek
TI Infratentorial Stereotactic Biopsy of Brainstem and Cerebellar Lesions
SO BRAIN SCIENCES
LA English
DT Article
DE brainstem tumors; infratentorial approach; molecular analyses;
   procedural complications; stereotactic biopsy; Toxoplasma gondii
ID SURGICAL RESECTION; CONTRAST ENHANCEMENT; TUMORS; DIAGNOSIS;
   TRANSCEREBELLAR; GLIOMAS
AB Stereotactic biopsy of posterior fossa lesions is often regarded as hazardous due to the critical structures in that area. Therefore, the aim of the study was to evaluate the diagnostic accuracy and safety of infratentorial stereotactic biopsy of brainstem or cerebellar lesions and its associations with other clinical, laboratory, and radiological parameters. From January 2000 to May 2021, 190 infratentorial stereotactic biopsies of posterior fossa tumors, including 108 biopsies of brainstem lesions, were performed. Moreover, 63 supratentorial biopsies of cerebral peduncle lesions were analyzed to compare the safety and efficacy of both approaches. Additionally, the presence of antibodies against Toxoplasma gondii and Epstein-Barr Virus (EBV) were documented in 67 and 66 patients, respectively, and magnetic resonance imaging (MRI) scans were evaluated in 114 patients. Only 4% of patients had minor complications and 1.5% had major complications, including one patient who died from intracranial bleeding. Nine (4.7%) biopsies were non-diagnostic. Isocitrate dehydrogenase 1 (IDH1) mutation, 1p/19q codeletion, and O6-methylguanine-DNA methyltransferase (MGMT) promoter methylation status were assessed in 29 patients, and were non-diagnostic in only 3 (10.3%) cases. Patients with high-grade gliomas (HGG) were more frequently seropositive for T. gondii than individuals with low-grade gliomas (LGG; p < 0.001). A total of 27% of HGG and 41% of LGG were non-enhancing on MRI. The infratentorial approach is generally safe and reliable for biopsy of brainstem and cerebellar lesions. In our study, the safety and efficacy of supratentorial biopsy of the cerebral peduncle and infratentorial biopsy of lesions below the cerebral peduncle were comparably high. Moreover, patients with HGG were more frequently seropositive for T. gondii than patients with LGG, and the relationship between toxoplasmosis and gliomagenesis requires further investigation.
C1 [Furtak, Jacek; Krajewski, Stanislaw; Birski, Marcin; Harat, Marek] 10th Mil Res Hosp & Polyclin, Dept Neurosurg, PL-85681 Bydgoszcz, Poland.
   [Furtak, Jacek] Franciszek Lukaszczyk Oncol Ctr, Dept Neurooncol & Radiosurg, PL-85796 Bydgoszcz, Poland.
   [Sledzinska, Paulina] Nicolaus Copernicus Univ, Coll Med, Fac Med, PL-85067 Bydgoszcz, Poland.
   [Bebyn, Marek G.] Med Univ Gdansk, Fac Med, PL-80210 Gdansk, Poland.
   [Szylberg, Tadeusz] 10th Mil Res Hosp, Dept Pathomorphol, PL-85681 Bydgoszcz, Poland.
   [Krajewski, Stanislaw] Univ Bydgoszcz, Dept Physiotherapy, PL-85059 Bydgoszcz, Poland.
   [Harat, Marek] Nicolaus Copernicus Univ, Dept Neurosurg & Neurol, Coll Med Bydgoszcz, PL-87100 Torun, Poland.
C3 Nicolaus Copernicus University; Fahrenheit Universities; Medical
   University Gdansk; Kazimierz Wielki University; University of Bydgoszcz;
   Nicolaus Copernicus University
RP Furtak, J (通讯作者)，10th Mil Res Hosp & Polyclin, Dept Neurosurg, PL-85681 Bydgoszcz, Poland.; Furtak, J (通讯作者)，Franciszek Lukaszczyk Oncol Ctr, Dept Neurooncol & Radiosurg, PL-85796 Bydgoszcz, Poland.
EM;
  ;
  
RI Bebyn, Marek Grzegorz/GQP-0013-2022; Śledzińska, Paulina/ACP-3063-2022;
   Furtak, Jacek/GQR-0467-2022
OI Bebyn, Marek Grzegorz/0000-0002-8650-6402; Śledzińska,
   Paulina/0000-0002-8151-8332; Birski, Marcin/0000-0001-6011-7559
CR [Anonymous], ACCURACY FRAMELESS F, DOI [10.1179/016164105X40057, DOI 10.1179/016164105X40057]
   [Anonymous], 2021, WHO CLASSIFICATION T, V5th
   Carruthers VB, 2007, SCHIZOPHRENIA BULL, V33, P745, DOI 10.1093/schbul/sbm008
   Cavalheiro S, 2015, CHILD NERV SYST, V31, P1815, DOI 10.1007/s00381-015-2799-y
   Chen SY, 2011, J CHIN MED ASSOC, V74, P110, DOI 10.1016/j.jcma.2011.01.024
   Corcino YL, 2019, SCI REP-UK, V9, DOI 10.1038/s41598-018-36724-2
   Dellaretti M, 2012, STEREOT FUNCT NEUROS, V90, P79, DOI 10.1159/000335502
   Faulkner H, 2021, WORLD NEUROSURG, V146, pE639, DOI 10.1016/j.wneu.2020.10.147
   Fritsch MJ, 1998, STEREOT FUNCT NEUROS, V71, P36, DOI 10.1159/000029645
   Fujimaki T, 2019, NEURO-ONCOLOGY, V21, P244
   Furtak J, 2019, NEUROL NEUROCHIR POL, V53, P435, DOI 10.5603/PJNNS.a2019.0056
   Ginsberg LE, 1998, SURG NEUROL, V49, P436, DOI 10.1016/S0090-3019(97)00360-1
   Goncalves-Ferreira AJ, 2003, SURG NEUROL, V60, P311, DOI 10.1016/S0090-3019(03)00379-3
   Hodge JM, 2021, INT J CANCER, V148, P2449, DOI 10.1002/ijc.33443
   Jackson RJ, 2001, NEURO-ONCOLOGY, V3, P193, DOI 10.1093/neuonc/3.3.193
   Jung BK, 2016, KOREAN J PARASITOL, V54, P201, DOI 10.3347/kjp.2016.54.2.201
   Kickingereder P, 2013, NEUROSURGERY, V72, P873, DOI 10.1227/NEU.0b013e31828bf445
   KONDZIOLKA D, 1995, SURG NEUROL, V43, P558, DOI 10.1016/0090-3019(95)00009-7
   Laigle-Donadey F, 2008, CURR OPIN ONCOL, V20, P662, DOI 10.1097/CCO.0b013e32831186e0
   Mathon B, 2019, CLIN NEUROL NEUROSUR, V186, DOI 10.1016/j.clineuro.2019.105544
   McGirt MJ, 2003, SURG NEUROL, V59, P277, DOI 10.1016/S0090-3019(03)00048-X
   Muniz-Feliciano L, 2013, PLOS PATHOG, V9, DOI 10.1371/journal.ppat.1003809
   Pallud J, 2009, NEURO-ONCOLOGY, V11, P176, DOI 10.1215/15228517-2008-066
   Pincus DW, 2006, J NEUROSURG, V104, P108, DOI 10.3171/ped.2006.104.2.108
   Portillo JAC, 2017, PLOS PATHOG, V13, DOI 10.1371/journal.ppat.1006671
   Rachinger W, 2009, J NEUROL NEUROSUR PS, V80, P1134, DOI 10.1136/jnnp.2009.174250
   Ramakonar HH, 2017, NEUROSURGERY, V64, P260, DOI 10.1093/neuros/nyx417.220
   Reithmeier T, 2013, CLIN NEUROL NEUROSUR, V115, P1955, DOI 10.1016/j.clineuro.2013.05.019
   Roujeau T, 2007, J NEUROSURG, V107, P1, DOI 10.3171/PED-07/07/001
   RYAN P, 1993, INT J EPIDEMIOL, V22, P412, DOI 10.1093/ije/22.3.412
   Sabbagh AJ, 2015, NEUROSCIENCES, V20, P98, DOI 10.17712/nsj.2015.2.20140621
   Samadani U, 2003, STEREOT FUNCT NEUROS, V81, P5, DOI 10.1159/000075097
   Sanai N, 2008, NEUROSURGERY, V63, P460, DOI 10.1227/01.NEU.0000324731.68843.74
   SCHUMAN LM, 1967, AM J PUBLIC HEALTH N, V57, P848, DOI 10.2105/AJPH.57.5.848
   Sledzinska P, 2021, INT J MOL SCI, V22, DOI 10.3390/ijms221910373
   STECK J, 1995, SURG NEUROL, V43, P563, DOI 10.1016/0090-3019(95)00156-5
   Tilgner J, 2005, NEUROSURGERY, V56, P257, DOI 10.1227/01.neu.0000148899.39020.87
   Vittecoq M, 2012, INFECT GENET EVOL, V12, P496, DOI 10.1016/j.meegid.2012.01.013
   Weller M, 2021, NAT REV CLIN ONCOL, V18, P170, DOI 10.1038/s41571-020-00447-z
NR 39
TC 8
Z9 8
U1 0
U2 1
PU MDPI
PI BASEL
PA ST ALBAN-ANLAGE 66, CH-4052 BASEL, SWITZERLAND
EI 2076-3425
J9 BRAIN SCI
JI Brain Sci.
PD NOV
PY 2021
VL 11
IS 11
AR 1432
DI 10.3390/brainsci11111432
PG 12
WC Neurosciences
WE Science Citation Index Expanded (SCI-EXPANDED)
SC Neurosciences & Neurology
GA 2I9KK
UT WOS:000815289600001
PM 34827431
OA gold, Green Published
DA 2023-08-27
ER

PT J
AU Matthies, C
   Brill, S
   Varallyay, C
   Solymosi, L
   Gelbrich, G
   Roosen, K
   Ernestus, RI
   Helms, J
   Hagen, R
   Mlynski, R
   Shehata-Dieler, W
   Muller, J
AF Matthies, Cordula
   Brill, Stefan
   Varallyay, Csanad
   Solymosi, Laszlo
   Gelbrich, Goetz
   Roosen, Klaus
   Ernestus, Ralf-Ingo
   Helms, Jan
   Hagen, Rudolf
   Mlynski, Robert
   Shehata-Dieler, Wafaa
   Mueller, Joachim
TI Auditory brainstem implants in neurofibromatosis Type 2: is open speech
   perception feasible? Clinical article
SO JOURNAL OF NEUROSURGERY
LA English
DT Article
DE acoustic neuroma; auditory brainstem implant; auditory midbrain implant;
   neurofibromatosis; open-set sentence recognition; speech perception;
   vestibular schwannoma; oncology; skull base
ID COCHLEAR IMPLANTATION; ELECTRICAL-STIMULATION; MIDBRAIN IMPLANT;
   VESTIBULAR SCHWANNOMAS; AUDIOLOGICAL PERFORMANCE; HEARING
   REHABILITATION; NUCLEUS; RESTORATION; DEAFNESS; ADULTS
AB Object. Patients with bilateral auditory nerve destruction may perceive some auditory input with auditory brainstem implants (ABIs). Despite technological developments and trials in new stimulation sites, hearing is very variable and of limited quality. The goal of this study was to identify advantageous and critical factors that influence the quality of auditory function, especially speech perception.
   Methods. The authors conducted a prospective study on ABI operations performed with the aid of multimodality neuromonitoring between 2005 and 2009 in 18 patients with neurofibromatosis Type 2. Outcome was evaluated by testing word recognition (monotrochee-polysyllabic word test at auditory-only mode [MTPa]) and open speech perception (Hochmair-Schulz-Moser [HSM] sentence test), both in pure auditory mode. The primary outcome was the HSM score at 24 months. The predictive meaning of general clinical data, tumor volume, number of active electrodes, duration of deafness, and early hearing data was examined.
   Results. In 16 successful ABI activations the average score for MTPa was 89% (SD 13%), and for HSM it was 41% (SD 32%) at 24 months. There were 2 nonresponders, 1 after radiosurgery and the other in an anatomical variant. Direct facial nerve reconstruction during the same surgery was followed by successful nerve recovery in 2 patients, with a simultaneous very good HSM result. Patients' age, tumor extension, and tumor volume were not negative predictors. There was an inverse relationship between HSM scores and deafness duration; 50% or higher HSM scores were found only in patients with ipsilateral deafness duration up to 24 months. The higher the deafness sum of both sides, the less likely that any HSM score will be achieved (p = 0.034). In patients with total deafness duration of less than 240 months, higher numbers of active electrodes were significantly associated with better outcomes. The strongest cross-correlation was identified between early MTPa score at 3 months and 24-month HSM outcome.
   Conclusions. This study documents that open-set speech recognition in pure auditory mode is feasible in patients with ABIs. Large tumor volumes do not prevent good outcome. Positive preconditions are short ipsilateral and short bilateral deafness periods and high number of auditory electrodes. Early ability in pure auditory word recognition tests indicates long-term capability of open speech perception.
C1 [Matthies, Cordula; Roosen, Klaus; Ernestus, Ralf-Ingo] Univ Wurzburg Hosp, Dept Neurosurg, D-97080 Wurzburg, Germany.
   [Brill, Stefan; Helms, Jan; Hagen, Rudolf; Mlynski, Robert; Shehata-Dieler, Wafaa; Mueller, Joachim] Univ Wurzburg Hosp, Dept Otorhinolaryngol, D-97080 Wurzburg, Germany.
   [Varallyay, Csanad; Solymosi, Laszlo] Univ Wurzburg Hosp, Dept Neuroradiol, D-97080 Wurzburg, Germany.
   [Gelbrich, Goetz] Univ Wurzburg Hosp, Dept Epidemiol & Biometr, D-97080 Wurzburg, Germany.
C3 University of Wurzburg; University of Wurzburg; University of Wurzburg;
   University of Wurzburg
RP Matthies, C (通讯作者)，Univ Wurzburg Hosp, Dept Neurosurg, Josef Schneider Str 11, D-97080 Wurzburg, Germany.
EM
RI Matthies, Cordula/AAE-5708-2019; Solymosi, László/E-6449-2018
OI Solymosi, László/0000-0001-9647-8647; Mlynski,
   Robert/0000-0001-6404-6364
CR Arndt S, 2011, OTOL NEUROTOL, V32, P39, DOI 10.1097/MAO.0b013e3181fcf271
   Behr R, 2007, SKULL BASE-INTERD AP, V17, P91, DOI 10.1055/s-2006-950390
   Bouccara D, 2007, Ann Otolaryngol Chir Cervicofac, V124, P148
   BRACKMANN DE, 1993, OTOLARYNG HEAD NECK, V108, P624, DOI 10.1177/019459989310800602
   Clair E G, 2006, COCHLEAR IMPLANTS, P222
   Colletti V, 2005, OTOLARYNG HEAD NECK, V133, P126, DOI 10.1016/j.otohns.2005.03.022
   Colletti Vittorio, 2006, Adv Otorhinolaryngol, V64, P167, DOI 10.1159/000094651
   Colletti V, 2007, OTOL NEUROTOL, V28, P39, DOI 10.1097/01.mao.0000247808.47712.02
   Colletti V, 2009, PROG BRAIN RES, V175, P333, DOI 10.1016/S0079-6123(09)17523-4
   Crea KN, 2010, AUDIOL NEURO-OTOL, V15, P18, DOI 10.1159/000218359
   Durisin M, 2008, EUR ARCH OTO-RHINO-L, V265, P381, DOI 10.1007/s00405-008-0584-1
   EDGERTON BJ, 1982, ANN OTO RHINOL LARYN, V91, P117
   Evans DGR, 2011, ADV OTO-RHINO-LARYNG, V70, P91, DOI 10.1159/000322482
   Evans DGR, 2005, OTOL NEUROTOL, V26, P93, DOI 10.1097/00129492-200501000-00016
   Formanek M, 1998, EUR ARCH OTO-RHINO-L, V255, P175, DOI 10.1007/s004050050038
   Grayeli AB, 2008, OTOL NEUROTOL, V29, P1140, DOI 10.1097/MAO.0b013e31818b6238
   Hanemann CO, 2008, BRAIN, V131, P606, DOI 10.1093/brain/awm249
   Helms J, 1997, ORL J OTO-RHINO-LARY, V59, P23, DOI 10.1159/000276901
   Hirschfelder A, 2008, OTOLARYNG HEAD NECK, V138, P357, DOI 10.1016/j.otohns.2007.10.019
   HITSELBERGER WE, 1984, OTOLARYNG HEAD NECK, V92, P52, DOI 10.1177/019459988409200111
   HOCHMAIRDESOYER IJ, 1980, ARCH OTO-RHINO-LARYN, V229, P81, DOI 10.1007/BF00454232
   Huy PTB, 2009, ACTA OTO-LARYNGOL, V129, P971, DOI 10.1080/00016480802510202
   Jackson Kim B, 2002, Am J Audiol, V11, P128, DOI 10.1044/1059-0889(2002/015)
   Kalamarides M, 2001, J NEUROSURG, V95, P1028, DOI 10.3171/jns.2001.95.6.1028
   Kanowitz SJ, 2004, LARYNGOSCOPE, V114, P2135, DOI 10.1097/01.mlg.0000149447.52888.f6
   Kuchta J, 2004, J NEUROSURG, V100, P16, DOI 10.3171/jns.2004.100.1.0016
   Laszig R, 2004, OTOL NEUROTOL, V25, P958, DOI 10.1097/00129492-200411000-00016
   Lenarz M, 2002, OTOL NEUROTOL, V23, P694, DOI 10.1097/00129492-200209000-00015
   Lenarz M, 2006, JARO-J ASSOC RES OTO, V7, P383, DOI 10.1007/s10162-006-0056-5
   Lenarz T, 2001, OTOL NEUROTOL, V22, P823, DOI 10.1097/00129492-200111000-00019
   Lenarz T, 2006, OTOL NEUROTOL, V27, P838, DOI 10.1097/01.mao.0000232010.01116.e9
   Lesinski-Schiedat A, 2000, J LARYNGOL OTOL, V114, P15
   Lim HH, 2008, NEUROSCIENCE, V154, P370, DOI 10.1016/j.neuroscience.2008.02.041
   Lim HH, 2008, HEARING RES, V242, P74, DOI 10.1016/j.heares.2008.02.003
   Lim HH, 2007, J NEUROSCI, V27, P13541, DOI 10.1523/JNEUROSCI.3123-07.2007
   Lustig LR, 2006, OTOL NEUROTOL, V27, P512, DOI 10.1097/00129492-200606000-00013
   Maini Sangeeta, 2009, Cochlear Implants Int, V10 Suppl 1, P33, DOI 10.1179/cim.2009.10.Supplement-1.33
   Matthies C, 2000, J LARYNGOL OTOL, V114, P32
   Mautner VF, 1996, NEUROSURGERY, V38, P880, DOI 10.1097/00006123-199605000-00004
   McCreery DB, 2008, HEARING RES, V242, P64, DOI 10.1016/j.heares.2007.11.014
   Moller AR, 2006, COCHLEAR BRAINSTEM I
   Neff BA, 2007, LARYNGOSCOPE, V117, P1069, DOI 10.1097/MLG.0b013e31804b1ae7
   Nevison B, 2002, EAR HEARING, V23, P170, DOI 10.1097/00003446-200206000-00002
   Oh SH, 2003, ACTA OTO-LARYNGOL, V123, P148, DOI 10.1080/0036554021000028111
   Otto SR, 2004, ARCH OTOLARYNGOL, V130, P656, DOI 10.1001/archotol.130.5.656
   Otto SR, 1998, OTOLARYNG HEAD NECK, V118, P291, DOI 10.1016/S0194-5998(98)70304-3
   Otto SR, 2002, J NEUROSURG, V96, P1063, DOI 10.3171/jns.2002.96.6.1063
   OTTO SR, 1990, ANN OTO RHINOL LARYN, V99, P789, DOI 10.1177/000348949009901006
   Otto SR, 2008, OTOL NEUROTOL, V29, P1147, DOI 10.1097/MAO.0b013e31818becb4
   Peasgood Alison, 2003, Cochlear Implants Int, V4, P171, DOI 10.1179/cim.2003.4.4.171
   Samii M, 1997, NEUROSURGERY, V40, P696, DOI 10.1097/00006123-199704000-00007
   Samii M, 2008, Prog Neurol Surg, V21, P169, DOI 10.1159/000156905
   Samii M, 1997, NEUROSURGERY, V40, P248, DOI 10.1097/00006123-199702000-00005
   Schwartz M, 2003, J ADV MATER-COVINA, V35, P2
   Schwartz MS, 2008, NEUROTHERAPEUTICS, V5, P128, DOI 10.1016/j.nurt.2007.10.068
   Sekiya T, 2009, EXP NEUROL, V218, P117, DOI 10.1016/j.expneurol.2009.04.014
   SHANNON RV, 1989, HEARING RES, V40, P173, DOI 10.1016/0378-5955(89)90110-X
   SHANNON RV, 1993, OTOLARYNG HEAD NECK, V108, P634, DOI 10.1177/019459989310800603
   Sollman WP, 1997, 11TH INTERNATIONAL CONGRESS OF NEUROLOGICAL SURGERY, VOLS 1 AND 2, P1569
   Sughrue ME, 2011, J NEUROSURG, V114, P381, DOI 10.3171/2010.4.JNS091962
   Vincent C, 2002, OTOL NEUROTOL, V23, P56, DOI 10.1097/00129492-200201000-00014
   Vincenti V, 2008, AUDIOL NEURO-OTOL, V13, P273, DOI 10.1159/000115437
   WILSON BS, 1991, NATURE, V352, P236, DOI 10.1038/352236a0
NR 63
TC 33
Z9 37
U1 0
U2 11
PU AMER ASSOC NEUROLOGICAL SURGEONS
PI ROLLING MEADOWS
PA 5550 MEADOWBROOK DRIVE, ROLLING MEADOWS, IL 60008 USA
SN 0022-3085
EI 1933-0693
J9 J NEUROSURG
JI J. Neurosurg.
PD FEB
PY 2014
VL 120
IS 2
BP 546
EP 558
DI 10.3171/2013.9.JNS12686
PG 13
WC Clinical Neurology; Surgery
WE Science Citation Index Expanded (SCI-EXPANDED)
SC Neurosciences & Neurology; Surgery
GA 301UA
UT WOS:000330556400041
PM 24329026
DA 2023-08-27
ER

PT J
AU Hargrave, DR
   Mabbott, DJ
   Bouffet, E
AF Hargrave, DR
   Mabbott, DJ
   Bouffet, E
TI Pathological laughter and behavioural change in childhood pontine glioma
SO JOURNAL OF NEURO-ONCOLOGY
LA English
DT Article
DE anxiety; behaviour; cerebellum; glioma; pathological laughter; pontine;
   recurrence
ID POSTERIOR-FOSSA TUMORS; FOU RIRE PRODROMIQUE; CHILDREN
AB Children with pontine glioma usually present classically with ataxia, motor deficits and cranial nerve palsies. The pons has generally not been regarded as a structure that mediates complex affective behaviour. However, we report nine children who either at the time of presentation or progression demonstrated marked behavioural changes manifesting as either "pathological laughter" or separation anxiety in the form of school refusal. A mechanism of how pontine lesions can cause such complex affective and cognitive behaviour has been suggested to consist of the disruption of a network of cerebro-ponto-cerebellar pathways and the evidence for this mechanism is discussed.
C1 Royal Marsden Hosp, Paediat Oncol Unit, Sutton SM2 5PT, Surrey, England.
   Hosp Sick Children, Paediat Brain Tumour Program, Toronto, ON M5G 1X8, Canada.
C3 Royal Marsden NHS Foundation Trust; University of Toronto; Hospital for
   Sick Children (SickKids)
RP Hargrave, DR (通讯作者)，Royal Marsden Hosp, Paediat Oncol Unit, Downs Rd, Sutton SM2 5PT, Surrey, England.
EM
RI Hargrave, Darren R/H-1066-2014; Bouffet, Eric/AAB-8999-2020
OI Hargrave, Darren R/0000-0001-8219-9807; 
CR ACHARI AN, 1976, JAMA-J AM MED ASSOC, V235, P1469
   ALBRIGHT AL, 1993, NEUROSURGERY, V33, P1026
   ARSENI C, 1959, ACTA NEUROCHIR SUPP, P292
   Barrow-Ferreira M, 1975, ELECTROENCEPHALOGR C, V38, P203
   CAIRNS H, 1950, Folia Psychiatr Neurol Neurochir Neerl, V53, P193
   CANTU RC, 1966, J NEUROSURG, V24, P1024, DOI 10.3171/jns.1966.24.6.1024
   Dabby R, 2004, BEHAV NEUROL, V15, P73, DOI 10.1155/2004/409248
   Gondim FDA, 2001, J NEUROL NEUROSUR PS, V71, P802, DOI 10.1136/jnnp.71.6.802
   IRONSIDE R, 1956, BRAIN, V79, P589, DOI 10.1093/brain/79.4.589
   LAL AP, 1992, J NEUROL NEUROSUR PS, V55, P628, DOI 10.1136/jnnp.55.7.628-a
   LASSMAN LP, 1967, LANCET, V1, P913
   LEPESTRE F, 1828, ARCH GEN MED, V18, P19
   Levisohn L, 2000, BRAIN, V123, P1041, DOI 10.1093/brain/123.5.1041
   MATSUOKA S, 1993, J NEUROSURG, V79, P428, DOI 10.3171/jns.1993.79.3.0428
   PANITCH HS, 1970, AM J DIS CHILD, V119, P465, DOI 10.1001/archpedi.1970.02100050467001
   Parvizi J, 2001, BRAIN, V124, P1708, DOI 10.1093/brain/124.9.1708
   Pecker J, 1970, Rev Otoneuroophtalmol, V42, P236
   Riva D, 2000, BRAIN, V123, P1051, DOI 10.1093/brain/123.5.1051
   RODRIGUEZ, 2003, NEUROLOGIA, V18, P225
   SANO K, 1967, CLIN NEUROL, P359
   Shafqat S, 1998, NEUROLOGY, V50, P1918, DOI 10.1212/WNL.50.6.1918
   Stein PD, 1996, SEM RESP CRIT CARE M, V17, P3, DOI 10.1055/s-2007-1009866
   WALI GM, 1993, J NEUROL NEUROSUR PS, V56, P209, DOI 10.1136/jnnp.56.2.209
NR 23
TC 18
Z9 19
U1 0
U2 1
PU SPRINGER
PI NEW YORK
PA 233 SPRING STREET, NEW YORK, NY 10013 USA
SN 0167-594X
J9 J NEURO-ONCOL
JI J. Neuro-Oncol.
PD MAY
PY 2006
VL 77
IS 3
BP 267
EP 271
DI 10.1007/s11060-005-9034-8
PG 5
WC Oncology; Clinical Neurology
WE Science Citation Index Expanded (SCI-EXPANDED)
SC Oncology; Neurosciences & Neurology
GA 060QY
UT WOS:000238817900006
PM 16314950
DA 2023-08-27
ER

PT J
AU Mahadewa, TGB
   Nakagawa, H
   Watabe, T
   Inoue, T
AF Mahadewa, TGB
   Nakagawa, H
   Watabe, T
   Inoue, T
TI Intramedullary neurosarcoidosis in the medulla oblongata: A case report
SO SURGICAL NEUROLOGY
LA English
DT Article
DE medulla oblongata; neurosarcoidosis; biopsy; steroid
ID FOLLOW-UP; SARCOIDOSIS
AB OBJECTIVE We present a rare case of neurosarcoidosis mimicking an intramedullary tumor in the medulla oblongata. The features of the clinical presentation, magnetic resonance (MRI) appearances, and management strategy are discussed.
   CASE PRESENTATION A 59-year-old man without evidence of systemic sarcoidosis was presented with a history of progressive numbness and deep sensation disturbance in bilateral lower extremities. MR imaging revealed an enhanced intra-axial mass lesion on the dorsal side of medulla. Under neurophysiological monitoring, tumor biopsy was performed. Pathologic evaluation revealed noncaseating granuloma composed of large epithelioid cells with multinucleated giant cells, suggesting sarcoidosis. Findings of comprehensive hematologic laboratory studies; cerebrospinal fluid examination; and examinations for bacteria, fungi, and acid fast bacilli were all negative. This mass lesion was diagnosed as medullary neurosarcoidosis, and then high-dose steroid therapy was tried. On follow-up, nearly complete resolution of the neurosarcoidosis on MRI was revealed.
   CONCLUSION To our knowledge, this is the first reported case of neurosarcoidosis manifested in the medulla oblongata. A biopsy is sufficient for a diagnosis and high-dose steroid is recommended. (C) 2004 Elsevier Inc. All rights reserved.
C1 Aichi Med Univ, Dept Neurol Surg, Nagakute, Aichi 4801195, Japan.
C3 Aichi Medical University
RP Nakagawa, H (通讯作者)，Aichi Med Univ, Dept Neurol Surg, 21 Karimata Yazato Nagakute, Nagakute, Aichi 4801195, Japan.
RI Tjokorda, Mahadewa Gde Bagus/K-3792-2016
OI Tjokorda, Mahadewa Gde Bagus/0000-0002-4445-4085
CR AMOS OD, 2001, J NEUROSURG, V94, P849
   ANTHONY AG, 2002, CURR OPIN PULM MED, V8, P445
   Baughman RP, 2002, CURR OPIN PULM MED, V8, P464, DOI 10.1097/00063198-200209000-00021
   DENNIS AN, 2000, J NEUROSURG, V93, P897
   Elias WJ, 1999, SURG NEUROL, V51, P185, DOI 10.1016/S0090-3019(97)00497-7
   Ferriby D, 2001, NEUROLOGY, V57, P927, DOI 10.1212/WNL.57.5.927
   JACK WJ, 2002, AM J NEURORADIOL, V23, P660
   Jarnier D, 1999, NEUROCHIRURGIE, V45, P214
   LEXA FJ, 1994, AM J NEURORADIOL, V15, P973
   Manterola-Burgaleta A, 2001, REV NEUROLOGIA, V32, P57, DOI 10.33588/rn.3201.2000027
   MARK L, 1998, NEUROSURGERY, V43, P788
   MARK N, 1996, NEUROSURGERY, V39, P179
   MARK SG, 2001, HDB NEUROSURGERY, P79
   MARK SG, 2001, HDB NEUROSURGERY, P404
   Murialdo G, 2002, J ENDOCRINOL INVEST, V25, P650, DOI 10.1007/BF03345093
   NOBU M, 1995, NEUROSURGERY, V37, P922
   Nowak DA, 2001, J NEUROL, V248, P363, DOI 10.1007/s004150170175
   Radziwill A, 1995, NERVENARZT, V66, P915
   Randeva HS, 2002, ENDOCRINE, V17, P241, DOI 10.1385/ENDO:17:3:241
   ROBERT JJ, 1998, NEUROSURGERY, V42, P635
   ROBERT JW, 2001, J NEUROSURG, V94, P1013
   STEPHAN AM, 1993, J NEUROSURG, V78, P994
   Uchino M, 2001, NEUROL MED-CHIR, V41, P48, DOI 10.2176/nmc.41.48
   Vinas FC, 2001, J CLIN NEUROSCI, V8, P505, DOI 10.1054/jocn.2000.0950
   Walker Aljoeson, 2001, Curr Treat Options Neurol, V3, P529, DOI 10.1007/s11940-001-0015-8
   Wang P Y, 1999, Zhonghua Yi Xue Za Zhi (Taipei), V62, P250
   Waubant E, 1997, NEURORADIOLOGY, V39, P357, DOI 10.1007/s002340050424
   Zajicek JP, 2000, CURR OPIN NEUROL, V13, P323, DOI 10.1097/00019052-200006000-00016
NR 28
TC 8
Z9 8
U1 0
U2 2
PU ELSEVIER SCIENCE INC
PI NEW YORK
PA STE 800, 230 PARK AVE, NEW YORK, NY 10169 USA
SN 0090-3019
J9 SURG NEUROL
JI Surg. Neurol.
PD MAR
PY 2004
VL 61
IS 3
BP 283
EP 287
DI 10.1016/S0090-3019(03)00398-7
PG 5
WC Clinical Neurology; Surgery
WE Science Citation Index Expanded (SCI-EXPANDED)
SC Neurosciences & Neurology; Surgery
GA 777TC
UT WOS:000189192800015
PM 14985005
DA 2023-08-27
ER

PT J
AU Xiao, AQ
   Xu, JG
   He, X
   You, C
AF Xiao, Anqi
   Xu, Jianguo
   He, Xin
   You, Chao
TI Extraventricular choroid plexus papilloma in the brainstem
SO JOURNAL OF NEUROSURGERY-PEDIATRICS
LA English
DT Article
DE choroid plexus papilloma; brainstem; MRI; Virchow-Robin space; oncology
ID OF-THE-LITERATURE; TUMORS
AB Choroid plexus papilloma (CPP) is extremely rare in the brainstem. The authors report the case of a 10-year-old boy with a lesion in the pons that was misdiagnosed as a glioma preoperatively. The boy underwent partial resection of the lesion, which was diagnosed as a CPP based on histopathological findings. The authors review the MRI findings in this case and conclude that the presence of a well-defined boundary and no obvious cerebral edema are valuable features for distinguishing brainstem CPP from glioma. Although previous reports of parenchymal CPPs have described enhancement on contrast-enhanced T1-weighted MR images, the lesion in this case did not demonstrate significant enhancement. The authors note that the diagnosis of extraventricular CPP cannot be ruled out in a case of brainstem tumor without marked enhancement.
C1 [Xiao, Anqi; Xu, Jianguo; You, Chao] Sichuan Univ, Dept Neurosurg, West China Hosp, Chengdu 610041, Sichuan, Peoples R China.
   [He, Xin] Sichuan Univ, Dept Pathol, West China Hosp, Chengdu 610041, Sichuan, Peoples R China.
C3 Sichuan University; Sichuan University
RP You, C (通讯作者)，Sichuan Univ, Dept Neurosurg, West China Hosp, 37 Guo Xue Xiang St, Chengdu 610041, Sichuan, Peoples R China.
EM
RI xiao, an/HZL-5077-2023
CR AZZAM NI, 1981, J NEUROSURG, V55, P651, DOI 10.3171/jns.1981.55.4.0651
   Garcia-Valtuille R, 2000, J NEUROSURG, V92, P870, DOI 10.3171/jns.2000.92.5.0870
   Gelabert-Gonzalez M, 2001, REV NEUROLOGIA, V33, P177, DOI 10.33588/rn.3302.2000554
   Imai Masaaki, 2011, Surg Neurol Int, V2, P151, DOI 10.4103/2152-7806.86228
   Karim A, 2006, CLIN NEUROL NEUROSUR, V108, P586, DOI 10.1016/j.clineuro.2005.04.008
   KIMURA M, 1992, NEUROSURGERY, V31, P563, DOI 10.1227/00006123-199209000-00020
   McIver JI, 2003, SURG NEUROL, V60, P360, DOI 10.1016/S0090-3019(03)00157-5
   Meyers SP, 2004, NEURORADIOLOGY, V46, P770, DOI 10.1007/s00234-004-1238-7
   Nomura Hiroaki, 1997, Neurologia Medico-Chirurgica, V37, P685, DOI 10.2176/nmc.37.685
   Okay O, 2009, TURK NEUROSURG, V19, P281
   Peyre M, 2012, ACTA NEUROCHIR, V154, P295, DOI 10.1007/s00701-011-1172-5
   Pillai A, 2004, J NEUROSURG, V100, P1076, DOI 10.3171/jns.2004.100.6.1076
   Rickert CH, 2001, MICROSC RES TECHNIQ, V52, P104
   Shin JH, 2001, CLIN IMAG, V25, P154, DOI 10.1016/S0899-7071(01)00284-4
   Strojan P, 2004, NEOPLASMA, V51, P306
   Tacconi L, 1996, ACTA NEUROCHIR, V138, P802, DOI 10.1007/BF01411257
   Zhang TJ, 2011, CLIN IMAG, V35, P64, DOI 10.1016/j.clinimag.2010.02.010
NR 17
TC 6
Z9 8
U1 1
U2 6
PU AMER ASSOC NEUROLOGICAL SURGEONS
PI ROLLING MEADOWS
PA 5550 MEADOWBROOK DRIVE, ROLLING MEADOWS, IL 60008 USA
SN 1933-0707
J9 J NEUROSURG-PEDIATR
JI J. Neurosurg.-Pediatr.
PD SEP
PY 2013
VL 12
IS 3
BP 247
EP 250
DI 10.3171/2013.6.PEDS137
PG 4
WC Clinical Neurology; Pediatrics; Surgery
WE Science Citation Index Expanded (SCI-EXPANDED)
SC Neurosciences & Neurology; Pediatrics; Surgery
GA 204IK
UT WOS:000323358800008
PM 23889355
DA 2023-08-27
ER

PT J
AU Cao, ZK
   Lv, JP
   Wei, XH
   Quan, W
AF Cao, ZhiKai
   Lv, JianPing
   Wei, Xinhua
   Quan, Wei
TI Appliance of preoperative diffusion tensor imaging and fiber
   tractography in patients with brainstem lesions
SO NEUROLOGY INDIA
LA English
DT Article
DE Brainstem; diffusion tensor imaging; lesion; surgery
ID CAVERNOUS MALFORMATIONS; WHITE-MATTER; TECHNICAL CONSIDERATIONS; 3 T;
   EXPERIENCE; RESECTION; TUMORS; RESOLUTION; GLIOMAS; SYSTEM
AB Background: Surgical resection of brainstem lesions has a high risk of morbidity, because vital fasciculi in the brainstem can be damaged along the entry routes. Diffusion tensor imaging (DTI) is an in vivo method for mapping white matter fiber tracts in the brain. Objective: To summarize the experience of surgical treatment of brainstem lesions with the assistance of DTI and fiber tractography. Materials and Methods: A retrospective analysis clinical data of nine patients with brainstem lesions were investigated between July 2007 and September 2009. The analysis included age distribution, clinical presentation, pre- and postoperative modified Rankin score (mRS), and surgical approach. DTI and fiber tractography were used to reconstruct the corticospinal tracts and the medial lemnisci. Results: DTI and fiber tractography showed that most of the corticospinal tracts were compressed anteriorly or anterolaterally, except for one case (posteriorly). All the medial lemnisci were displaced posteriorly or posterolaterally. Individualized surgical approaches were designed according to the information provided by DTI and fiber tractography. Total resection was achieved in two patients with brainstem cavernomas and two patients with pilocytic astrocytoma. Partial resection was performed in the other patients. The neurological functional status was better than preoperative period in eight patients, one patient with medulla oblongata astrocytoma deteriorated. The preoperative average mRS score was 2.22 points. At the time of the last follow-up, the average postoperative score had improved by 0.56 to 1.66 points. Conclusions: DTI and fiber tractography can provide valuable information regarding the relationship between the principal fiber tracts and brainstem lesions, which is useful in neurosurgical planning.
C1 [Cao, ZhiKai; Lv, JianPing; Wei, Xinhua; Quan, Wei] Guangzhou Med Coll, Guangzhou Municipal Peoples Hosp 1, Dept Neurosurg, Guangzhou 510180, Guangdong, Peoples R China.
C3 Guangzhou Medical University
RP Lv, JP (通讯作者)，Guangzhou Med Coll, Guangzhou Municipal Peoples Hosp 1, Dept Neurosurg, Guangzhou 510080, Guangdong, Peoples R China.
EM
CR Bricolo A, 1991, Acta Neurochir Suppl (Wien), V53, P148
   Chen X, 2007, ACTA NEUROCHIR, V149, P1117, DOI 10.1007/s00701-007-1282-2
   Chen XL, 2007, SURG NEUROL, V68, P285, DOI 10.1016/j.surneu.2007.05.005
   Constantini S, 1996, J NEURO-ONCOL, V28, P193
   de Oliveira JG, 2010, NEUROSURGERY, V66, P389, DOI 10.1227/01.NEU.0000363702.67016.5D
   DUKATZ T, 2007, CLIN NEUROPHYSIOL, V118, pE28
   Ferroli P, 2005, NEUROSURGERY, V56, P1203, DOI 10.1227/01.NEU.0000159644.04757.45
   FRITSCHI JA, 1994, ACTA NEUROCHIR, V130, P35, DOI 10.1007/BF01405501
   Habas C, 2007, NEURORADIOLOGY, V49, P849, DOI 10.1007/s00234-007-0267-4
   HEFFEZ DS, 1990, NEUROSURGERY, V27, P789, DOI 10.1227/00006123-199011000-00018
   Helton KJ, 2008, J NEUROSURG-PEDIATR, V1, P270, DOI 10.3171/PED/2008/1/4/270
   Helton KJ, 2006, AM J NEURORADIOL, V27, P786
   Jallo GI, 2003, ANN NEUROSURG, V3, P1
   Jissendi P, 2008, J NEURORADIOLOGY, V35, P42, DOI 10.1016/j.neurad.2007.11.001
   Kamali A, 2009, EUR RADIOL, V19, P1480, DOI 10.1007/s00330-009-1305-x
   Kansal Somil, 1999, Indian Journal of Cancer, V36, P99
   Kawashima M, 2009, NEUROSURG REV, V32, P457, DOI 10.1007/s10143-009-0211-7
   KYOSHIMA K, 1993, J NEUROSURG, V78, P987, DOI 10.3171/jns.1993.78.6.0987
   Li H, 2009, NEUROL INDIA, V57, P269, DOI 10.4103/0028-3886.53269
   Mathiesen T, 2003, J NEUROSURG, V99, P31, DOI 10.3171/jns.2003.99.1.0031
   Mehta VS, 2009, NEUROL INDIA, V57, P274, DOI 10.4103/0028-3886.53272
   Nagae-Poetscher LM, 2004, AM J NEURORADIOL, V25, P1325
   Porter RW, 1999, J NEUROSURG, V90, P50, DOI 10.3171/jns.1999.90.1.0050
   REQUENA I, 1991, J NEUROL NEUROSUR PS, V54, P590, DOI 10.1136/jnnp.54.7.590
   Salamon N, 2005, NEURORADIOLOGY, V47, P895, DOI 10.1007/s00234-005-1439-8
   Sandalcioglu IE, 2002, J NEUROL NEUROSUR PS, V72, P351, DOI 10.1136/jnnp.72.3.351
   Wedeen VJ, 2008, NEUROIMAGE, V41, P1267, DOI 10.1016/j.neuroimage.2008.03.036
   Xu QW, 1997, SURG NEUROL, V48, P30, DOI 10.1016/S0090-3019(97)00125-0
NR 28
TC 0
Z9 0
U1 0
U2 2
PU WOLTERS KLUWER MEDKNOW PUBLICATIONS
PI MUMBAI
PA WOLTERS KLUWER INDIA PVT LTD , A-202, 2ND FLR, QUBE, C T S  NO 1498A-2
   VILLAGE MAROL, ANDHERI EAST, MUMBAI, 400059, INDIA
SN 0028-3886
EI 1998-4022
J9 NEUROL INDIA
JI Neurol. India
PD NOV-DEC
PY 2010
VL 58
IS 6
BP 72
EP 76
DI 10.4103/0028-3886.73736
PG 5
WC Neurosciences
WE Science Citation Index Expanded (SCI-EXPANDED)
SC Neurosciences & Neurology
GA 697XF
UT WOS:000285552400013
DA 2023-08-27
ER

PT J
AU Barton, KL
   Misuraca, K
   Cordero, F
   Dobrikova, E
   Min, HD
   Gromeier, M
   Kirsch, DG
   Becher, OJ
AF Barton, Kelly L.
   Misuraca, Katherine
   Cordero, Francisco
   Dobrikova, Elena
   Min, Hooney D.
   Gromeier, Matthias
   Kirsch, David G.
   Becher, Oren J.
TI PD-0332991, a CDK4/6 Inhibitor, Significantly Prolongs Survival in a
   Genetically Engineered Mouse Model of Brainstem Glioma
SO PLOS ONE
LA English
DT Article
ID INTRINSIC PONTINE GLIOMAS; DEPENDENT KINASE 4/6; PD 0332991;
   GLIOBLASTOMA; MUTATION; GROWTH; CELLS; PHOSPHORYLATION; INACTIVATION;
   SENSITIVITY
AB Diffuse intrinsic pontine glioma (DIPG) is an incurable tumor that arises in the brainstem of children. To date there is not a single approved drug to effectively treat these tumors and thus novel therapies are desperately needed. Recent studies suggest that a significant fraction of these tumors contain alterations in cell cycle regulatory genes including amplification of the D-type cyclins and CDK4/6, and less commonly, loss of Ink4a-ARF leading to aberrant cell proliferation. In this study, we evaluated the therapeutic approach of targeting the cyclin-CDK-Retinoblastoma (Rb) pathway in a genetically engineered PDGF-B-driven brainstem glioma (BSG) mouse model. We found that PD-0332991 (PD), a CDK4/6 inhibitor, induces cell-cycle arrest in our PDGF-B; Ink4a-ARF deficient model both in vitro and in vivo. By contrast, the PDGF-B; p53 deficient model was mostly resistant to treatment with PD. We noted that a 7-day treatment course with PD significantly prolonged survival by 12% in the PDGF-B; Ink4a-ARF deficient BSG model. Furthermore, a single dose of 10 Gy radiation therapy (RT) followed by 7 days of treatment with PD increased the survival by 19% in comparison to RT alone. These findings provide the rationale for evaluating PD in children with Ink4a-ARF deficient gliomas.
C1 [Barton, Kelly L.; Cordero, Francisco; Becher, Oren J.] Duke Univ, Dept Pediat, Durham, NC 27706 USA.
   [Cordero, Francisco; Becher, Oren J.] Duke Univ, Dept Pathol, Durham, NC 27706 USA.
   [Barton, Kelly L.; Cordero, Francisco; Becher, Oren J.] Duke Univ, Preston Robert Tisch Brain Tumor Ctr, Durham, NC 27706 USA.
   [Misuraca, Katherine] Duke Univ, Grad Program Mol Canc Biol, Durham, NC 27706 USA.
   [Dobrikova, Elena; Gromeier, Matthias] Duke Univ, Dept Surg, Durham, NC 27706 USA.
   [Min, Hooney D.; Kirsch, David G.] Duke Univ, Dept Radiat Oncol, Durham, NC 27706 USA.
   [Kirsch, David G.] Duke Univ, Dept Pharmacol & Canc Biol, Durham, NC 27706 USA.
C3 Duke University; Duke University; Duke University; Duke University; Duke
   University; Duke University; Duke University
RP Becher, OJ (通讯作者)，Duke Univ, Dept Pediat, Durham, NC 27706 USA.
EM
RI Becher, Oren/AAI-7495-2020; Becher, Oren/GXN-3269-2022
OI Becher, Oren/0000-0003-4016-8492
FU Duke Cancer Institute; Rory David Deutsch Foundation
FX This work was supported by a Duke Cancer Institute seed grant to OJB and
   DGK. OJB is also supported by the Rory David Deutsch Foundation.
   http://www.roryd.org/. The funders had no role in study design, data
   collection and analysis, decision to publish, or preparation of the
   manuscript.
CR Anders L, 2011, CANCER CELL, V20, P620, DOI 10.1016/j.ccr.2011.10.001
   Aoki Y, 2012, J NEURO-ONCOL, V108, P29, DOI 10.1007/s11060-011-0796-x
   Becher OJ, 2010, CANCER RES, V70, P2548, DOI 10.1158/0008-5472.CAN-09-2503
   Cen L, 2012, NEURO-ONCOLOGY, V14, P870, DOI 10.1093/neuonc/nos114
   Chan KM, 2013, GENE DEV, V27, P985, DOI 10.1101/gad.217778.113
   Khuong-Quang DA, 2012, ACTA NEUROPATHOL, V124, P439, DOI 10.1007/s00401-012-0998-0
   Flaherty KT, 2012, CLIN CANCER RES, V18, P568, DOI 10.1158/1078-0432.CCR-11-0509
   Fry DW, 2004, MOL CANCER THER, V3, P1427
   Harbour JW, 1999, CELL, V98, P859, DOI 10.1016/S0092-8674(00)81519-6
   Hargrove D, 2006, LANCET ONCOL, V7, P241, DOI 10.1016/S1470-2045(06)70615-5
   Huillard E, 2012, P NATL ACAD SCI USA, V109, P8710, DOI 10.1073/pnas.1117255109
   Katsumi Y, 2011, BIOCHEM BIOPH RES CO, V413, P62, DOI 10.1016/j.bbrc.2011.08.047
   Leonard JP, 2012, BLOOD, V119, P4597, DOI 10.1182/blood-2011-10-388298
   Lewis PW, 2013, SCIENCE, V340, P857, DOI 10.1126/science.1232245
   Lundberg AS, 1998, MOL CELL BIOL, V18, P753, DOI 10.1128/MCB.18.2.753
   Michaud K, 2010, CANCER RES, V70, P3228, DOI 10.1158/0008-5472.CAN-09-4559
   MOSMANN T, 1983, J IMMUNOL METHODS, V65, P55, DOI 10.1016/0022-1759(83)90303-4
   Newton J, 2011, MED PHYS, V38, P6754, DOI 10.1118/1.3663675
   Paugh BS, 2011, J CLIN ONCOL, V29, P3999, DOI 10.1200/JCO.2011.35.5677
   Paugh BS, 2010, J CLIN ONCOL, V28, P3061, DOI 10.1200/JCO.2009.26.7252
   Puget S, 2012, PLOS ONE, V7, DOI 10.1371/journal.pone.0030313
   Warren KE, 2012, NEURO-ONCOLOGY, V14, P326, DOI 10.1093/neuonc/nor190
   Wiedemeyer WR, 2010, P NATL ACAD SCI USA, V107, P11501, DOI 10.1073/pnas.1001613107
   Zarghooni M, 2010, J CLIN ONCOL, V28, P1337, DOI 10.1200/JCO.2009.25.5463
NR 24
TC 119
Z9 122
U1 1
U2 14
PU PUBLIC LIBRARY SCIENCE
PI SAN FRANCISCO
PA 1160 BATTERY STREET, STE 100, SAN FRANCISCO, CA 94111 USA
SN 1932-6203
J9 PLOS ONE
JI PLoS One
PD OCT 2
PY 2013
VL 8
IS 10
AR e77639
DI 10.1371/journal.pone.0077639
PG 7
WC Multidisciplinary Sciences
WE Science Citation Index Expanded (SCI-EXPANDED)
SC Science & Technology - Other Topics
GA 231RG
UT WOS:000325434500103
PM 24098593
OA gold, Green Published, Green Submitted
DA 2023-08-27
ER

PT J
AU Kossatz, S
   Carney, B
   Schweitzer, M
   Carlucci, G
   Miloushev, VZ
   Maachani, UB
   Rajappa, P
   Keshari, KR
   Pisapia, D
   Weber, WA
   Souweidane, MM
   Reiner, T
AF Kossatz, Susanne
   Carney, Brandon
   Schweitzer, Melanie
   Carlucci, Giuseppe
   Miloushev, Vesselin Z.
   Maachani, Uday B.
   Rajappa, Prajwal
   Keshari, Kayvan R.
   Pisapia, David
   Weber, Wolfgang A.
   Souweidane, Mark M.
   Reiner, Thomas
TI Biomarker-Based PET Imaging of Diffuse Intrinsic Pontine Glioma in Mouse
   Models
SO CANCER RESEARCH
LA English
DT Article
ID ADP-RIBOSE POLYMERASE; BRAIN-STEM GLIOMA; POLY(ADP-RIBOSE) POLYMERASE;
   DNA-DAMAGE; RESPONSE ASSESSMENT; ONCOLOGY-GROUP; SINGLE-CELL; PHASE-II;
   PARP1; EXPRESSION
AB Diffuse intrinsic pontine glioma (DIPG) is a childhood brainstem tumor with a universally poor prognosis. Here, we characterize a positron emission tomography (PET) probe for imaging DIPG in vivo. In human histological tissues, the probes target, PARP1, was highly expressed in DIPG compared to normal brain. PET imaging allowed for the sensitive detection of DIPG in a genetically engineered mouse model, and probe uptake correlated to histologically determined tumor infiltration. Imaging with the sister fluorescence agent revealed that uptake was confined to proliferating, PARP1-expressing cells. Comparison with other imaging technologies revealed remarkable accuracy of our biomarker approach. We subsequently demonstrated that serial imaging of DIPG in mouse models enables monitoring of tumor growth, as shown in modeling of tumor progression. Overall, this validated method for quantifying DIPG burden would serve useful in monitoring treatment response in early phase clinical trials. (C) 2017 AACR.
C1 [Kossatz, Susanne; Carney, Brandon; Carlucci, Giuseppe; Miloushev, Vesselin Z.; Keshari, Kayvan R.; Weber, Wolfgang A.; Reiner, Thomas] Mem Sloan Kettering Canc Ctr, Dept Radiol, 1275 York Ave, New York, NY 10021 USA.
   [Carney, Brandon] CUNY, Hunter Coll, Dept Chem, New York, NY 10021 USA.
   [Carney, Brandon] CUNY, Grad Ctr, PhD Program Chem, New York, NY USA.
   [Schweitzer, Melanie; Maachani, Uday B.; Rajappa, Prajwal; Souweidane, Mark M.] Weill Cornell Med Coll, Dept Neurol Surg, New York, NY USA.
   [Miloushev, Vesselin Z.; Keshari, Kayvan R.; Weber, Wolfgang A.; Reiner, Thomas] Weill Cornell Med Coll, Dept Radiol, New York, NY USA.
   [Keshari, Kayvan R.] Mem Sloan Kettering Canc Ctr, Mol Pharmacol Program, 1275 York Ave, New York, NY 10021 USA.
   [Pisapia, David] Weill Cornell Med Coll, Dept Pathol & Lab Med, New York, NY USA.
   [Souweidane, Mark M.] Mem Sloan Kettering Canc Ctr, Dept Neurosurg, 1275 York Ave, New York, NY 10021 USA.
C3 Memorial Sloan Kettering Cancer Center; City University of New York
   (CUNY) System; Hunter College (CUNY); City University of New York (CUNY)
   System; Cornell University; Weill Cornell Medicine; Cornell University;
   Weill Cornell Medicine; Memorial Sloan Kettering Cancer Center; Cornell
   University; Weill Cornell Medicine; Memorial Sloan Kettering Cancer
   Center
RP Reiner, T (通讯作者)，Mem Sloan Kettering Canc Ctr, 1275 York Ave, New York, NY 10044 USA.
EM
RI Kossatz, Susanne/Q-7453-2017
OI Kossatz, Susanne/0000-0002-1908-1782; Reiner,
   Thomas/0000-0002-7819-5480; Rahimi-Keshari, Kayvan/0000-0002-5181-8035;
   Weber, Wolfgang/0000-0002-7854-4345; Maachani, Uday
   Bhanu/0000-0003-4089-0979; Carlucci, Giuseppe/0000-0001-7494-335X
FU National Institutes of Health [NIH1 R01 HL125703, R01CA204441,
   R21CA191679, K25 EB016673, P30 CA008748-50]; Center for Molecular
   Imaging and Nanotechnology (CMINT); Tow Fellowship Program in Molecular
   Imaging and Nanotechnology; MSKCC Radiology Development Project Grant;
   National Science Foundation [IGERT 0965983]; German Research Foundation
FX This work was supported by National Institutes of Health grants NIH1 R01
   HL125703 to T. Reiner, R01CA204441 to T. Reiner, R21CA191679 to T.
   Reiner and W.A. Weber, K25 EB016673 to T. Reiner, and P30 CA008748-50.
   For financial support, the authors also thank the Center for Molecular
   Imaging and Nanotechnology (CMINT; T. Reiner), the Tow Fellowship
   Program in Molecular Imaging and Nanotechnology (B. Carney and S.
   Kossatz), the MSKCC Radiology Development Project Grant (V.Z. Miloushe)
   the National Science Foundation's Integrative Graduate Education and
   Research Traineeship (IGERT 0965983 for B. Carney) and the German
   Research Foundation (S. Kossatz).
CR ALBRIGHT AL, 1993, NEUROSURGERY, V33, P1026
   Aleskandarany M, 2015, BREAST CANCER RES TR, V150, P81, DOI 10.1007/s10549-015-3306-6
   Becher OJ, 2010, CANCER RES, V70, P2548, DOI 10.1158/0008-5472.CAN-09-2503
   Bieche I, 1996, CLIN CANCER RES, V2, P1163
   Bouchard WJ, 2003, EXP HEMATOL, V31, P446, DOI 10.1016/S0301-472X(03)00083-3
   Buczkowicz P, 2015, FRONT ONCOL, V5, DOI 10.3389/fonc.2015.00147
   Buczkowicz P, 2014, ACTA NEUROPATHOL, V128, P573, DOI 10.1007/s00401-014-1319-6
   Carney B, 2016, MOL IMAGING BIOL, V18, P386, DOI 10.1007/s11307-015-0904-y
   Chornenkyy Y, 2015, MOL CANCER THER, V14, P2560, DOI 10.1158/1535-7163.MCT-15-0282
   Chow JPH, 2013, MOL CANCER THER, V12, P2517, DOI 10.1158/1535-7163.MCT-13-0010
   Cohen KJ, 2011, NEURO-ONCOLOGY, V13, P410, DOI 10.1093/neuonc/noq205
   deMurcia JM, 1997, P NATL ACAD SCI USA, V94, P7303, DOI 10.1073/pnas.94.14.7303
   Deorah Sundeep, 2006, Neurosurg Focus, V20, pE1, DOI 10.3171/foc.2006.20.4.E1
   Khuong-Quang DA, 2012, ACTA NEUROPATHOL, V124, P439, DOI 10.1007/s00401-012-0998-0
   Galia A, 2012, EUR J HISTOCHEM, V56, P45, DOI 10.4081/ejh.2012.e9
   Grasso CS, 2015, NAT MED, V21, P827, DOI 10.1038/nm0715-827a
   Green AR, 2015, BREAST CANCER RES TR, V149, P353, DOI 10.1007/s10549-014-3230-1
   Haas-Kogan DA, 2011, NEURO-ONCOLOGY, V13, P298, DOI 10.1093/neuonc/noq202
   Hambardzumyan D, 2009, TRANSL ONCOL, V2, P89, DOI 10.1593/tlo.09100
   Hargrave D, 2008, J NEURO-ONCOL, V86, P313, DOI 10.1007/s11060-007-9473-5
   Hargrove D, 2006, LANCET ONCOL, V7, P241, DOI 10.1016/S1470-2045(06)70615-5
   Hashizume R, 2012, J NEURO-ONCOL, V110, P305, DOI 10.1007/s11060-012-0973-6
   Irwin CP, 2014, NEOPLASIA, V16, P432, DOI 10.1016/j.neo.2014.05.005
   Jalali R, 2010, INT J RADIAT ONCOL, V77, P113, DOI 10.1016/j.ijrobp.2009.04.031
   Kohler BA, 2011, JNCI-J NATL CANCER I, V103, P714, DOI 10.1093/jnci/djr077
   Korones DN, 2008, PEDIATR BLOOD CANCER, V50, P227, DOI 10.1002/pbc.21154
   Kossatz S, 2016, SCI REP-UK, V6, DOI 10.1038/srep21371
   KRETSCHMAR CS, 1993, CANCER-AM CANCER SOC, V72, P1404, DOI 10.1002/1097-0142(19930815)72:4<1404::AID-CNCR2820720441>3.0.CO;2-G
   Liu AK, 2009, PEDIATR RADIOL, V39, P1317, DOI 10.1007/s00247-009-1368-5
   Lobel U, 2016, NEURORADIOLOGY, V58, P1027, DOI 10.1007/s00234-016-1724-8
   Martin SJ, 2002, NUCL MED BIOL, V29, P263, DOI 10.1016/S0969-8051(01)00289-X
   Michalski A, 2010, J NEURO-ONCOL, V100, P81, DOI 10.1007/s11060-010-0141-9
   Michels J, 2015, ANN ONCOL, V26, P2470, DOI 10.1093/annonc/mdv393
   Mosdzianowski C, 2001, EUR J NUCL MED, V28, P1228
   Nikbakht H, 2016, NAT COMMUN, V7, DOI 10.1038/ncomms11185
   Nosho K, 2006, EUR J CANCER, V42, P2374, DOI 10.1016/j.ejca.2006.01.061
   Oh J, 2005, J MAGN RESON IMAGING, V21, P701, DOI 10.1002/jmri.20335
   Ossovskaya Valeria, 2010, Genes Cancer, V1, P812, DOI 10.1177/1947601910383418
   Ostrom QT, 2014, NEURO-ONCOLOGY, V16, P1, DOI [10.1093/neuonc/nov189, 10.1093/neuonc/nou327, 10.1093/neuonc/nou223]
   Park SH, 2015, TRANSL ONCOL, V8, P239, DOI 10.1016/j.tranon.2015.04.004
   Patel S, 2014, CHILD NERV SYST, V30, P147, DOI 10.1007/s00381-013-2307-1
   Poussaint TY, 2011, NEURO-ONCOLOGY, V13, P417, DOI 10.1093/neuonc/noq200
   Reiner T, 2012, NEOPLASIA, V14, P169, DOI 10.1593/neo.12414
   Rojo F, 2012, ANN ONCOL, V23, P1156, DOI 10.1093/annonc/mdr361
   Scott CL, 2015, J CLIN ONCOL, V33, P1397, DOI 10.1200/JCO.2014.58.8848
   Sharp JR, 2010, EUR J CANCER, V46, P3271, DOI 10.1016/j.ejca.2010.06.115
   Smith MA, 2010, J CLIN ONCOL, V28, P2625, DOI 10.1200/JCO.2009.27.0421
   Smith SJ, 2011, NEURO-ONCOLOGY, V13, P1171, DOI 10.1093/neuonc/nor115
   Staibano S, 2005, HUM PATHOL, V36, P724, DOI 10.1016/j.humpath.2005.04.017
   Sutinen E, 2004, EUR J NUCL MED MOL I, V31, P317, DOI 10.1007/s00259-003-1377-9
   Thurber GM, 2014, MOL CANCER THER, V13, P986, DOI 10.1158/1535-7163.MCT-13-0801
   Thurber GM, 2013, NAT COMMUN, V4, DOI 10.1038/ncomms2506
   Tolvanen T, 2010, EUR J NUCL MED MOL I, V37, P874, DOI 10.1007/s00259-009-1346-z
   Warren KE, 2012, FRONT ONCOL, V2, DOI 10.3389/fonc.2012.00205
   Wen PY, 2010, J CLIN ONCOL, V28, P1963, DOI 10.1200/JCO.2009.26.3541
NR 55
TC 19
Z9 19
U1 1
U2 13
PU AMER ASSOC CANCER RESEARCH
PI PHILADELPHIA
PA 615 CHESTNUT ST, 17TH FLOOR, PHILADELPHIA, PA 19106-4404 USA
SN 0008-5472
EI 1538-7445
J9 CANCER RES
JI Cancer Res.
PD APR 15
PY 2017
VL 77
IS 8
BP 2112
EP 2123
DI 10.1158/0008-5472.CAN-16-2850
PG 12
WC Oncology
WE Science Citation Index Expanded (SCI-EXPANDED)
SC Oncology
GA ER9ZQ
UT WOS:000399186100027
PM 28108511
OA Green Submitted, Green Accepted, Bronze
DA 2023-08-27
ER

PT J
AU Dellaretti, M
   Touzet, G
   Reyns, N
   Dubois, F
   Gusmao, S
   Pereira, JLB
   Blond, S
AF Dellaretti, Marcos
   Touzet, Gustavo
   Reyns, Nicolas
   Dubois, Francois
   Gusmao, Sebastiao
   Pereira, Julio Leonardo Barbosa
   Blond, Serge
TI Correlation among magnetic resonance imaging findings, prognostic
   factors for survival, and histological diagnosis of intrinsic brainstem
   lesions in children Clinical article
SO JOURNAL OF NEUROSURGERY-PEDIATRICS
LA English
DT Article
DE brainstem tumor; biopsy; children
ID STEREOTACTIC BIOPSY; GLIOMAS; MULTICENTER; SURGERY; MASSES; TUMORS; RISK
AB Object. The aim of this study was to compare MR imaging characteristics with histopathological findings of intrinsic brainstem lesions and also to show the prognostic factors in patients with diffuse brainstem glioma.
   Methods. Between February 1988 and August 2007, 44 brainstem biopsies were performed at the Roger Salengro Hospital in Lille. France, in children with intrinsic brainstem lesions not amenable to excision. Twenty-six were female and 18 male, and the mean age was 6 years.
   Results. Histological evaluation revealed diffuse brainstem glioma in all patients with diffuse nonenhancing brainstem lesions. Diffuse brainstem glioma was found in 18 patients (90%) with diffuse enhancing brainstem lesions. Pathological entities different from diffuse glioma were verified in 2 patients (10%)-1 with ependymoma and 1 with ganglioglioma.
   In 4 of 5 patients with a focal nonenhancing brainstem lesion, the histopathological diagnosis was diffuse low-grade glioma. In 6 of 10 patients with focal enhancing brainstem lesion, the diagnosis was diffuse brainstem glioma, and pathological entities different from diffuse brainstem glioma were verified in 2 (20%), both with pilocytic astrocytoma.
   The mean 1-year actuarial survival rates for patients classified with low-grade and high-grade glioma were 80.4% +/- 0.08% and 48.6% +/- 0.14%, respectively.
   Conclusions. The impact of stereotactic biopsy on intrinsic brainstem lesions was greater in patients with MR imaging-documented enhancing lesions in whom the diagnosis of diffuse glioma was less frequent. Patients with low-grade glioma seem to have longer survival than those with high-grade glioma. (DOI: 10.3171/2011.9.PEDS1167)
C1 [Dellaretti, Marcos; Pereira, Julio Leonardo Barbosa] Santa Casa Hosp, Dept Neurosurg, Belo Horizonte, MG, Brazil.
   [Dellaretti, Marcos; Touzet, Gustavo; Reyns, Nicolas; Dubois, Francois; Blond, Serge] CHRU Hop Roger Salengro, Dept Neurosurg, Lille, France.
   [Dellaretti, Marcos; Gusmao, Sebastiao] Univ Fed Minas Gerais, Fac Med, Belo Horizonte, MG, Brazil.
C3 Universite de Lille - ISITE; CHU Lille; Universidade Federal de Minas
   Gerais
RP Dellaretti, M (通讯作者)，Santa Casa Hosp, Dept Neurosurg, Belo Horizonte, MG, Brazil.
EM
RI Dellaretti, Marcos/G-4664-2012
CR Aker FV, 2005, NEUROPATHOLOGY, V25, P207, DOI 10.1111/j.1440-1789.2005.00634.x
   ALBRIGHT AL, 1993, NEUROSURGERY, V33, P1026
   Donaldson SS, 2006, J CLIN ONCOL, V24, P1266, DOI 10.1200/JCO.2005.04.6599
   Guillamo JS, 2001, CURR OPIN NEUROL, V14, P711, DOI 10.1097/00019052-200112000-00006
   Hargrove D, 2006, LANCET ONCOL, V7, P241, DOI 10.1016/S1470-2045(06)70615-5
   Jeuken JWM, 2006, TECHNOL CANCER RES T, V5, P215, DOI 10.1177/153303460600500305
   Kesari S, 2008, J NEURO-ONCOL, V88, P175, DOI 10.1007/s11060-008-9545-1
   KONDZIOLKA D, 1995, SURG NEUROL, V43, P558, DOI 10.1016/0090-3019(95)00009-7
   Kreth FW, 2001, ACTA NEUROCHIR, V143, P539, DOI 10.1007/s007010170058
   Levivier M, 1998, CRIT REV NEUROSURG, V8, P338, DOI 10.1007/s003290050099
   Massager N, 2000, NEUROSURG FOCUS, V8
   Pincus DW, 2006, J NEUROSURG, V104, P108, DOI 10.3171/ped.2006.104.2.108
   Pirotte BJM, 2007, J NEUROSURG, V107, P392, DOI 10.3171/PED-07/11/392
   Rachinger W, 2009, J NEUROL NEUROSUR PS, V80, P1134, DOI 10.1136/jnnp.2009.174250
   RAJSHEKHAR V, 1995, J NEUROSURG, V82, P976, DOI 10.3171/jns.1995.82.6.0976
   RANJAN A, 1993, J NEUROSURG, V79, P839, DOI 10.3171/jns.1993.79.6.0839
   Roujeau T, 2007, J NEUROSURG, V107, P1, DOI 10.3171/PED-07/07/001
   Schumacher M, 2007, J NEUROSURG, V106, P111, DOI 10.3171/ped.2007.106.2.111
   Yen CP, 2007, J NEUROSURG, V106, P8, DOI 10.3171/jns.2007.106.1.8
NR 19
TC 28
Z9 28
U1 0
U2 3
PU AMER ASSOC NEUROLOGICAL SURGEONS
PI ROLLING MEADOWS
PA 5550 MEADOWBROOK DRIVE, ROLLING MEADOWS, IL 60008 USA
SN 1933-0707
J9 J NEUROSURG-PEDIATR
JI J. Neurosurg.-Pediatr.
PD DEC
PY 2011
VL 8
IS 6
BP 539
EP 543
DI 10.3171/2011.9.PEDS1167
PG 5
WC Clinical Neurology; Pediatrics; Surgery
WE Science Citation Index Expanded (SCI-EXPANDED)
SC Neurosciences & Neurology; Pediatrics; Surgery
GA 853UC
UT WOS:000297450200003
PM 22132909
DA 2023-08-27
ER

PT J
AU Curless, RG
   Schatz, NJ
   Bowen, BC
   Rodriguez, Z
   Ruiz, A
AF Curless, RG
   Schatz, NJ
   Bowen, BC
   Rodriguez, Z
   Ruiz, A
TI Lyme neuroborreliosis masquerading as a brainstem tumor in a 15-year-old
SO PEDIATRIC NEUROLOGY
LA English
DT Article
ID NERVOUS-SYSTEM MANIFESTATIONS; NEUROLOGIC MANIFESTATIONS;
   MULTIPLE-SCLEROSIS; DISEASE; BORRELIOSIS; CHILDREN
AB A 15-year-old boy had onset of unilateral facial weakness. A few days later, he experienced mild vertigo, double vision, and headache. Examination confirmed a peripheral right seventh nerve weakness in addition to an internuclear ophthalmoplegia, The neurologic features suggested a pontine glioma. A T-2-weighted MRI scan revealed demyelinating lesions in the pens and in several areas of the cerebrum, including the periventricular region. Subsequent history revealed that he had been diagnosed with Lyme arthritis 7 years earlier while living in Connecticut. The radiographic studies favored a diagnosis of multiple sclerosis. However, studies of blood and cerebrospinal fluid established a diagnosis of Lyme neuroborreliosis. (C) 1996 by Elsevier Science Inc. All rights reserved.
C1 BASCOM PALMER EYE INST,DEPT OPHTHALMOL,DIV NEUROOPHTHALMOL,MIAMI,FL 33101.
   BASCOM PALMER EYE INST,DEPT OPHTHALMOL,DIV NEUROOPHTHALMOL,MIAMI,FL 33101.
   UNIV MIAMI,SCH MED,DEPT RADIOL,DIV NEURORADIOL,MIAMI,FL.
C3 Bascom Palmer Eye Institute; Bascom Palmer Eye Institute; University of
   Miami
RP Curless, RG (通讯作者)，BASCOM PALMER EYE INST,DEPT NEUROL D45,DIV PEDIAT NEUROL,POB 016960,MIAMI,FL 33101, USA.
CR BELMAN AL, 1992, PEDIATR NEUROL, V8, P428, DOI 10.1016/0887-8994(92)90003-H
   *CDC, 1991, MMWR-MORBID MORTAL W, V40, P471
   DRESSLER F, 1993, J INFECT DIS, V167, P392, DOI 10.1093/infdis/167.2.392
   FERNANDEZ RE, 1990, AM J NEURORADIOL, V11, P479
   FINKEL MJ, 1992, ARCH NEUROL-CHICAGO, V49, P102, DOI 10.1001/archneur.1992.00530250106024
   GEANMARTON AD, 1991, RADIOLOGY, V180, P215, DOI 10.1148/radiology.180.1.2052698
   HALPERIN JJ, 1989, NEUROLOGY, V39, P753, DOI 10.1212/WNL.39.6.753
   HALPERIN JJ, 1991, NEUROLOGY, V41, P1571, DOI 10.1212/WNL.41.10.1571
   HOROWITZ AL, 1989, AM J NEURORADIOL, V10, P303
   LOGIGIAN EL, 1990, NEW ENGL J MED, V323, P1438, DOI 10.1056/NEJM199011223232102
   PACHNER AR, 1989, ARCH NEUROL-CHICAGO, V46, P790, DOI 10.1001/archneur.1989.00520430086023
   PACHNER AR, 1993, ANN NEUROL, V34, P544, DOI 10.1002/ana.410340407
   PFISTER HW, 1989, ARCH NEUROL-CHICAGO, V46, P1190, DOI 10.1001/archneur.1989.00520470044025
   SZER IS, 1991, NEW ENGL J MED, V325, P159, DOI 10.1056/NEJM199107183250304
NR 14
TC 17
Z9 17
U1 0
U2 0
PU ELSEVIER SCIENCE INC
PI NEW YORK
PA 655 AVENUE OF THE AMERICAS, NEW YORK, NY 10010
SN 0887-8994
J9 PEDIATR NEUROL
JI Pediatr. Neurol.
PD OCT
PY 1996
VL 15
IS 3
BP 258
EP 260
DI 10.1016/S0887-8994(96)00172-5
PG 3
WC Clinical Neurology; Pediatrics
WE Science Citation Index Expanded (SCI-EXPANDED)
SC Neurosciences & Neurology; Pediatrics
GA VR143
UT WOS:A1996VR14300015
PM 8916168
DA 2023-08-27
ER

PT J
AU Kwon, JW
   Kim, IO
   Cheon, JE
   Kim, WS
   Moon, SG
   Kim, TJ
   Chi, JG
   Wang, KC
   Chung, JK
   Yeon, KM
AF Kwon, Jong Won
   Kim, In-One
   Cheon, Jung-Eun
   Kim, Woo Sun
   Moon, Sung Gyu
   Kim, Tae Jung
   Chi, Je Geun
   Wang, Kyu-Chang
   Chung, June Key
   Yeon, Kyung Mo
TI Paediatric brain-stem gliomas: MRI, FDG-PET and histological grading
   correlation
SO PEDIATRIC RADIOLOGY
LA English
DT Article
DE brain stem; brain neoplasms; MRI; PET; children
ID CHILDREN; ASTROCYTOMA; CHILDHOOD; DIAGNOSIS; TUMORS
AB Background: MRI and FDG-PET may predict the histological grading of paediatric brain-stem gliomas. Objective: To assess MRI findings and metabolic imaging using FDG-PET of brain-stem gliomas based on histological grading. Materials and methods: Included in the study were 20 paediatric patients (age 3-14 years, mean 8.2 years) with brain-stem glioma (five glioblastomas, ten anaplastic astrocytomas and five low-grade astrocytomas). MR images were assessed for the anatomical site of tumour origin, focality, pattern of tumour growth, and enhancement. Results: All glioblastomas were located in the pons and showed diffuse pontine enlargement with focally exophytic features. Eight anaplastic astrocytomas were located in the pons and demonstrated diffuse pontine enlargement without exophytic features. Low-grade astrocytomas were located in the pons, midbrain or medulla and showed focally exophytic growth features and peripheral enhancement. In 12 patients in whom FDG-PET was undertaken, glioblastomas showed hypermetabolic or hypometabolic lesions, anaplastic astrocytomas showed no metabolic change or hypometabolic lesions and low-grade astrocytomas showed hypometabolism compared with the cerebellum. Conclusion: MRI findings correlated well with histological grading of brain-stem gliomas and MRI may therefore predict the histological grading. FDG-PET may be helpful in differentiating between anaplastic astrocytoma and glioblastomas among high-grade tumours.
C1 Seoul Natl Univ Hosp, Dept Radiol, Seoul 110744, South Korea.
   Seoul Natl Univ, Coll Med, Dept Pathol, Seoul 151, South Korea.
   Seoul Natl Univ, Coll Med, Dept Neurosurg, Seoul, South Korea.
   Seoul Natl Univ, Coll Med, Dept Nucl Med, Seoul, South Korea.
C3 Seoul National University (SNU); Seoul National University Hospital;
   Seoul National University (SNU); Seoul National University (SNU); Seoul
   National University (SNU)
RP Kim, IO (通讯作者)，Seoul Natl Univ Hosp, Dept Radiol, 28 Yongon Dong, Seoul 110744, South Korea.
EM
RI Kim, Woo Sun/D-6511-2012; Geun, Je/G-4989-2011; Wang,
   Kyu-Chang/J-5545-2012; Cheon, Jung-Eun/J-5684-2012; Kim,
   In-One/J-5427-2012
OI Chi, Je-Geun/0000-0002-9950-2072
CR ALBRIGHT AL, 1983, CANCER, V52, P2313, DOI 10.1002/1097-0142(19831215)52:12<2313::AID-CNCR2820521226>3.0.CO;2-I
   BARKOVICH AJ, 1991, PEDIATR NEUROSURG, V16, P73, DOI 10.1159/000120511
   Bowers DC, 2001, PEDIATR NEUROSURG, V34, P229, DOI 10.1159/000056027
   COHEN ME, 1986, NEUROLOGY, V36, P602, DOI 10.1212/WNL.36.5.602
   De Witte O, 2000, J NEURO-ONCOL, V49, P157, DOI 10.1023/A:1026518002800
   DELBEKE D, 1995, RADIOLOGY, V195, P47, DOI 10.1148/radiology.195.1.7892494
   EPSTEIN FJ, 1993, J NEUROSURG, V78, P408, DOI 10.3171/jns.1993.78.3.0408
   FARWELL JR, 1977, CANCER, V40, P3123, DOI 10.1002/1097-0142(197712)40:6<3123::AID-CNCR2820400656>3.0.CO;2-6
   Fenton LZ, 2003, MED PEDIATR ONCOL, V40, P260, DOI 10.1002/mpo.10227
   Fischbein NJ, 1996, PEDIATR NEUROSURG, V24, P9, DOI 10.1159/000121010
   Freeman CR, 1996, CANCER-AM CANCER SOC, V77, P555
   Freeman CR, 1998, INT J RADIAT ONCOL, V40, P265, DOI 10.1016/S0360-3016(97)00572-5
   Jallo GI, 2004, CHILD NERV SYST, V20, P143, DOI 10.1007/s00381-003-0870-6
   KAHN AP, 1993, J NEUROSURG, V79, P845
   KANE AG, 1993, AM J NEURORADIOL, V14, P941
   Kaplan AM, 1996, PEDIATR NEUROSURG, V24, P185, DOI 10.1159/000121036
   Lesniak MS, 2003, PEDIATR NEUROSURG, V39, P314, DOI 10.1159/000075260
   Moghrabi A, 1995, PEDIATR NEUROSURG, V23, P293, DOI 10.1159/000120974
   Padma MV, 2003, J NEURO-ONCOL, V64, P227, DOI 10.1023/A:1025665820001
   POLLACK IF, 1993, J NEUROSURG, V78, P859, DOI 10.3171/jns.1993.78.6.0859
   Poussaint TY, 1999, PEDIATR RADIOL, V29, P662, DOI 10.1007/s002470050671
   Rubin G, 1998, CHILD NERV SYST, V14, P167, DOI 10.1007/s003810050205
   STROINK AR, 1987, NEUROSURGERY, V20, P439, DOI 10.1227/00006123-198703000-00014
   STROINK AR, 1986, J NEUROSURG, V65, P745, DOI 10.3171/jns.1986.65.6.0745
NR 24
TC 35
Z9 39
U1 0
U2 3
PU SPRINGER
PI NEW YORK
PA 233 SPRING STREET, NEW YORK, NY 10013 USA
SN 0301-0449
J9 PEDIATR RADIOL
JI Pediatr. Radiol.
PD SEP
PY 2006
VL 36
IS 9
BP 959
EP 964
DI 10.1007/s00247-006-0256-5
PG 6
WC Pediatrics; Radiology, Nuclear Medicine & Medical Imaging
WE Science Citation Index Expanded (SCI-EXPANDED)
SC Pediatrics; Radiology, Nuclear Medicine & Medical Imaging
GA 071HU
UT WOS:000239591400009
PM 16847598
DA 2023-08-27
ER

PT J
AU Schumacher, M
   Schulte-Monting, J
   Stoeter, P
   Warmuth-Metz, M
   Solymosi, L
AF Schumacher, Martin
   Schulte-Moenting, Juergen
   Stoeter, Peter
   Warmuth-Metz, Monika
   Solymosi, Laszlo
TI Magnetic resonance imaging compared with biopsy in the diagnosis of
   brainstem diseases of childhood: a multicenter review
SO JOURNAL OF NEUROSURGERY
LA English
DT Article
DE brainstem tumor; magnetic resonance imaging; biopsy; pediatric
   neurosurgery
ID CHILDRENS CANCER GROUP; CERVICOMEDULLARY TUMORS; GLIOMAS; THERAPY;
   ASTROCYTOMAS; CHEMOTHERAPY; PATHOLOGY; NMR; MRI
AB Object. Data analysis was performed in a multicenter study to evaluate magnetic resonance (MR) imaging for classification of brain tumors, prognosis, and prediction of tumor histological diagnosis.
   Methods. The clinical, MR imaging, and histological findings obtained in 142 pediatric cases of brainstem disease were assessed in a multicenter study performed as a blinded review. Clinical data were available in 142 cases, histopathological findings in 126, and MR images in 131. The subgroup of cases involving brainstem gliomas (78 cases) was analyzed separately. Images that met criteria for evaluation were reviewed in random order by three experienced observers who were initially blinded to clinical data as well as histopathological diagnoses. Subsequently, the images were randomized again and provided to the observers for review together with the clinical symptoms of the specific patients.
   The three observers were able to correctly identify lesions as tumors or nontumorous disease on MR images in 99, 96, and 95% of cases, resulting in an overall sensitivity of 0.94, a specificity of 0.43, a positive predictive value of 0.96, and a negative predictive value of 0.45. Awareness of clinical symptoms did not change the results.
   Conclusions. Based on 14 imaging criteria together with the patient's clinical history and symptoms, laboratory data (results of cerebrospinal fluid analysis as well as infectious and immunological parameters), and imaging follow up, a diagnosis of brainstem tumor, as opposed to demyelination, encephalitis, or granuloma, could generally be made. Given these findings, there is only rarely a need for biopsy, and in those patients in whom it is considered, the potential costs and benefits must be carefully assessed on a case-by-case basis.
C1 Univ Freiburg Klinikum, Dept Neuroradiol, D-29106 Freiburg, Germany.
   Inst Med Biometrie & Med Informat, Dept Med iometry & Stat, Freiburg, Germany.
   Univ Mainz Klinikum, Inst Neuroradiol, Mainz, Germany.
   Univ Klinikum Wurzburg, Dept Neuroradiol, Wurzburg, Germany.
C3 University of Freiburg; Johannes Gutenberg University of Mainz;
   University of Wurzburg
RP Schumacher, M (通讯作者)，Univ Freiburg Klinikum, Dept Neuroradiol, Breisacher Str 64, D-29106 Freiburg, Germany.
EM
RI Solymosi, László/E-6449-2018
OI Solymosi, László/0000-0001-9647-8647
CR Albright AL, 1996, PEDIATR NEUROSURG, V24, P252
   ALBRIGHT AL, 1986, J NEUROSURG, V65, P751, DOI 10.3171/jns.1986.65.6.0751
   ALBRIGHT AL, 1993, NEUROSURGERY, V33, P1026
   BARKOVICH AJ, 1991, PEDIATR NEUROSURG, V16, P73, DOI 10.1159/000120511
   Behnke J, 1997, CHILD NERV SYST, V13, P122, DOI 10.1007/s003810050058
   BRADAC GB, 1985, NEURORADIOLOGY, V27, P208, DOI 10.1007/BF00344489
   Bricolo A, 1991, Acta Neurochir Suppl (Wien), V53, P148
   BRUNO L, 1982, PEDIATRIC NEUROSURGE, P361
   Burger PC, 1996, PEDIATR NEUROSURG, V24, P35, DOI 10.1159/000121012
   BYRNE JV, 1989, NEURORADIOLOGY, V31, P129, DOI 10.1007/BF00698839
   CHANG T, 1993, NEURORADIOLOGY, V35, P274, DOI 10.1007/BF00602613
   Colosimo C, 1997, RIV NEURORADIOL, V10, P41, DOI 10.1177/197140099701000104
   Constantini S, 1996, J NEURO-ONCOL, V28, P193
   Dunkel IJ, 1996, PEDIATR NEUROSURG, V24, P263, DOI 10.1159/000121049
   EDWARDS MSB, 1989, J NEUROSURG, V70, P691, DOI 10.3171/jns.1989.70.5.0691
   EPSTEIN F, 1985, CANCER, V56, P1804, DOI 10.1002/1097-0142(19851001)56:7+<1804::AID-CNCR2820561316>3.0.CO;2-5
   EPSTEIN F, 1986, J NEUROSURG, V64, P11, DOI 10.3171/jns.1986.64.1.0011
   Epstein F, 1996, PEDIATR NEUROSURG, V24, P24, DOI 10.1159/000121011
   EPSTEIN FJ, 1993, J NEUROSURG, V78, P408, DOI 10.3171/jns.1993.78.3.0408
   Fischbein NJ, 1996, PEDIATR NEUROSURG, V24, P9, DOI 10.1159/000121010
   Freeman CR, 1996, PEDIATR NEUROSURG, V24, P103, DOI 10.1159/000121025
   FULTON DS, 1981, J NEUROSURG, V54, P721, DOI 10.3171/jns.1981.54.6.0721
   GOLDEN GS, 1972, J NEUROL NEUROSUR PS, V35, P732, DOI 10.1136/jnnp.35.5.732
   HAN JS, 1984, RADIOLOGY, V150, P705, DOI 10.1148/radiology.150.3.6695071
   Hoffman HJ, 1996, PEDIATR NEUROSURG, V24, P256, DOI 10.1159/000121048
   HOFFMAN HJ, 1987, CONCEPTS PEDIATR NEU, V7, P105
   KANE AG, 1993, AM J NEURORADIOL, V14, P941
   Kaplan AM, 1996, PEDIATR NEUROSURG, V24, P185, DOI 10.1159/000121036
   LEE BCP, 1985, AM J NEURORADIOL, V6, P159
   LITTMAN P, 1980, CANCER, V45, P2787, DOI 10.1002/1097-0142(19800601)45:11<2787::AID-CNCR2820451113>3.0.CO;2-V
   MARIA BL, 1993, J CHILD NEUROL, V8, P112, DOI 10.1177/088307389300800203
   OSTERTAG CB, 1980, SURG NEUROL, V14, P275
   Packer RJ, 1996, PEDIATR NEUROSURG, V24, P217, DOI 10.1159/000121041
   PETERMAN SB, 1985, NEURORADIOLOGY, V27, P202, DOI 10.1007/BF00344488
   ROBERTSON PL, 1994, NEUROLOGY, V44, P1798, DOI 10.1212/WNL.44.10.1798
   Rubin G, 1998, CHILD NERV SYST, V14, P167, DOI 10.1007/s003810050205
   SANFORD RA, 1988, SURG NEUROL, V30, P276, DOI 10.1016/0090-3019(88)90299-6
   STROINK AR, 1987, NEUROSURGERY, V20, P439, DOI 10.1227/00006123-198703000-00014
   Tilgner J, 2005, NEUROSURGERY, V56, P257, DOI 10.1227/01.neu.0000148899.39020.87
   Valentino V, 2004, RIV NEURORADIOL, V17, P539, DOI 10.1177/197140090401700406
   vonKummer R, 1996, AM J NEURORADIOL, V17, P1743
   WARNKE PC, 1998, NEUROONKOLOGIE DIAGN, P75
NR 42
TC 53
Z9 57
U1 0
U2 1
PU AMER ASSOC NEUROLOGICAL SURGEONS
PI ROLLING MEADOWS
PA 5550 MEADOWBROOK DRIVE, ROLLING MEADOWS, IL 60008 USA
SN 0022-3085
EI 1933-0693
J9 J NEUROSURG
JI J. Neurosurg.
PD FEB
PY 2007
VL 106
IS 2
SU S
BP 111
EP 119
DI 10.3171/ped.2007.106.2.111
PG 9
WC Clinical Neurology; Surgery
WE Science Citation Index Expanded (SCI-EXPANDED)
SC Neurosciences & Neurology; Surgery
GA 140QH
UT WOS:000244520100009
PM 17330536
DA 2023-08-27
ER

PT J
AU Haas-Kogan, DA
   Banerjee, A
   Poussaint, TY
   Kocak, M
   Prados, MD
   Geyer, JR
   Fouladi, M
   Broniscer, A
   Minturn, JE
   Pollack, IF
   Packer, RJ
   Boyett, JM
   Kun, LE
AF Haas-Kogan, Daphne A.
   Banerjee, Anuradha
   Poussaint, Tina Young
   Kocak, Mehmet
   Prados, Michael D.
   Geyer, J. Russell
   Fouladi, Maryam
   Broniscer, Alberto
   Minturn, Jane E.
   Pollack, Ian F.
   Packer, Roger J.
   Boyett, James M.
   Kun, Larry E.
TI Phase II trial of tipifarnib and radiation in children with newly
   diagnosed diffuse intrinsic pontine gliomas
SO NEURO-ONCOLOGY
LA English
DT Article
DE diffuse intrinsic pontine glioma; farnesyltransferase inhibitors;
   pediatric
ID BRAIN-STEM GLIOMA; RECURRENT MALIGNANT GLIOMA; HIGH-GRADE GLIOMAS;
   FARNESYLTRANSFERASE INHIBITORS; ANTIEPILEPTIC DRUGS; CLINICAL PROSPECTS;
   ASTROCYTIC TUMORS; R115777; PSEUDOPROGRESSION; RADIOTHERAPY
AB We performed a phase II study to assess the efficacy and toxicity of tipifarnib, a farnesyltransferase inhibitor, administered with radiation therapy (RT) in children with newly diagnosed diffuse intrinsic pontine gliomas. Children 3-21 years old with pontine gliomas (BSGs) were treated with concurrent tipifarnib and RT, followed by adjuvant tipifarnib. Tipifarnib was taken orally twice daily (125 mg/m(2)/dose) during RT; after RT, it was taken at 200 mg/m(2) twice daily for 21 days, in 28-day cycles. Initial and follow-up neuroimaging was centrally reviewed. Forty eligible patients (median age, 5.5 years; range, 3.3-16.5 years) had a median progression-free survival of 6.8 months (range, 0.2-18.6 months) and median overall survival of 8.3 months (range, 0.2-18.6 months). Kaplan-Meier estimates ( standard error) of 1-year progression-free and overall survival were 12.9% +/- 4.9% and 34.3% +/- 7.4%, respectively. A single patient remained on tipifarnib without progression at the completion of the study, two years after initiation of treatment. Seven patients were without disease progression for at least six months, three of whom remained controlled for more than a year. The most frequent toxicity was grade 3 lymphopenia. We documented a single instance of "pseudoprogression" by neuroimaging review. We found no discordance among 3 approaches to defining disease progression: as interpreted by treating institutions (based on clinical status and/or imaging) and by central review (using bi-dimensional tumor "area" versus volumetric measurements). For children with diffuse BSGs, tipifarnib administered with irradiation offered no clinical advantage over historical controls. Biopsies and molecular analyses of pediatric BSGs are vital for identification of new agents and for rational use of targeted agents.
C1 [Haas-Kogan, Daphne A.] Univ Calif San Francisco, Dept Radiat Oncol, San Francisco, CA 94115 USA.
   [Poussaint, Tina Young] Childrens Hosp, Boston, MA 02115 USA.
   [Kocak, Mehmet; Boyett, James M.; Kun, Larry E.] St Jude Childrens Hosp, Memphis, TN 38105 USA.
   [Geyer, J. Russell] Seattle Childrens Hosp, Seattle, WA USA.
   [Minturn, Jane E.] Childrens Hosp Philadelphia, Philadelphia, PA 19104 USA.
   [Pollack, Ian F.] Childrens Hosp Pittsburgh, Pittsburgh, PA 15213 USA.
   [Packer, Roger J.] Childrens Natl Med Ctr, Washington, DC 20010 USA.
C3 University of California System; University of California San Francisco;
   Harvard University; Boston Children's Hospital; St Jude Children's
   Research Hospital; Seattle Children's Hospital; University of
   Pennsylvania; Pennsylvania Medicine; Childrens Hospital of Philadelphia;
   Pennsylvania Commonwealth System of Higher Education (PCSHE); University
   of Pittsburgh; Children's National Health System
RP Haas-Kogan, DA (通讯作者)，Univ Calif San Francisco, Dept Radiat Oncol, 1600 Divisadero St,Suite H1031, San Francisco, CA 94115 USA.
EM
RI Prados, Michael/N-5220-2019
OI Broniscer, Alberto/0000-0001-9646-0463; Kocak,
   Mehmet/0000-0002-3386-1734
FU National Institutes of Health [U01 CA81457]; Brain Tumor SPORE grant
   [P50 CA097257]; National Center for Research Resources [M01 RR00188];
   American Lebanese Syrian Associated Charities; Nancy and Stephen Grand
   Philanthropic Fund
FX National Institutes of Health (U01 CA81457 for the Pediatric Brain Tumor
   Consortium), Brain Tumor SPORE grant (P50 CA097257 to D.H.K.), National
   Center for Research Resources (M01 RR00188), American Lebanese Syrian
   Associated Charities, and The Nancy and Stephen Grand Philanthropic Fund
   (to D.H.K.).
CR Brandsma D, 2008, LANCET ONCOL, V9, P453, DOI 10.1016/S1470-2045(08)70125-6
   Burzynski Stanislaw R, 2006, Paediatr Drugs, V8, P167, DOI 10.2165/00148581-200608030-00003
   Cloughesy TF, 2005, J CLIN ONCOL, V23, P6647, DOI 10.1200/JCO.2005.10.068
   Cloughesy TF, 2006, J CLIN ONCOL, V24, P3651, DOI 10.1200/JCO.2006.06.2323
   Delmas C, 2002, INT J CANCER, V100, P43, DOI 10.1002/ijc.10439
   Fangusaro J, 2009, J CHILD NEUROL, V24, P1409, DOI 10.1177/0883073809338960
   Fouladi M, 2007, CANCER-AM CANCER SOC, V110, P2535, DOI 10.1002/cncr.23078
   Freeman CR, 1998, INT J RADIAT ONCOL, V40, P265, DOI 10.1016/S0360-3016(97)00572-5
   Gahramanov S, 2011, INT J RADIAT ONCOL, V79, P514, DOI 10.1016/j.ijrobp.2009.10.072
   Gilbertson RJ, 2003, CLIN CANCER RES, V9, P3620
   Haas-Kogan DA, 2008, NEURO-ONCOLOGY, V10, P341, DOI 10.1215/15228517-2008-004
   Hayward RM, 2008, J NEURO-ONCOL, V90, P57, DOI 10.1007/s11060-008-9631-4
   Henson JW, 2008, AM J NEURORADIOL, V29, P419, DOI 10.3174/ajnr.A0963
   Jones HA, 2001, SEMIN RADIAT ONCOL, V11, P328, DOI 10.1053/srao.2001.26020
   Korones DN, 2008, PEDIATR BLOOD CANCER, V50, P227, DOI 10.1002/pbc.21154
   Lustig R, 2008, NEURO-ONCOLOGY, V10, P1004, DOI 10.1215/15228517-2008-070
   Moyal ECJ, 2007, INT J RADIAT ONCOL, V68, P1396, DOI 10.1016/j.ijrobp.2007.02.043
   Okada H, 2008, J NEURO-ONCOL, V88, P245, DOI 10.1007/s11060-008-9566-9
   Pollack IF, 2006, J NEUROSURG, V105, P418, DOI 10.3171/ped.2006.105.5.418
   Prendergast GC, 2000, CURR OPIN CELL BIOL, V12, P166, DOI 10.1016/S0955-0674(99)00072-1
   Prendergast GC, 2000, SEMIN CANCER BIOL, V10, P443, DOI 10.1006/scbi.2000.0335
   Sorensen AG, 2008, NAT CLIN PRACT ONCOL, V5, P634, DOI 10.1038/ncponc1204
   Sorensen AG, 2001, J CLIN ONCOL, V19, P551, DOI 10.1200/JCO.2001.19.2.551
   Szybka M, 2003, CLIN NEUROPATHOL, V22, P180
   Wen PY, 2010, J CLIN ONCOL, V28, P1963, DOI 10.1200/JCO.2009.26.3541
NR 25
TC 57
Z9 59
U1 0
U2 2
PU OXFORD UNIV PRESS INC
PI CARY
PA JOURNALS DEPT, 2001 EVANS RD, CARY, NC 27513 USA
SN 1522-8517
J9 NEURO-ONCOLOGY
JI Neuro-Oncology
PD MAR
PY 2011
VL 13
IS 3
BP 298
EP 306
DI 10.1093/neuonc/noq202
PG 9
WC Oncology; Clinical Neurology
WE Science Citation Index Expanded (SCI-EXPANDED)
SC Oncology; Neurosciences & Neurology
GA 732AA
UT WOS:000288151400004
PM 21339191
OA Bronze, Green Published
DA 2023-08-27
ER

PT J
AU Mursch, K
   Buhre, W
   Behnke-Mursch, J
   Markakis, E
AF Mursch, K
   Buhre, W
   Behnke-Mursch, J
   Markakis, E
TI Peroperative cardiovascular stability during brainstem surgery. The use
   of high-dose methylprednisolone compared to dexamethasone - A
   retrospective analysis
SO ACTA ANAESTHESIOLOGICA SCANDINAVICA
LA English
DT Article
DE brainstem tumors; brainstem surgery; dexamethasone; methylprednisolone;
   hemodynamic stability; blood pressure; heart rate
ID SPINAL-CORD INJURY; CONTROLLED TRIAL; TUMORS; CHILDREN; NALOXONE;
   GLIOMAS; PONS; STEM
AB Background: In neurosurgical procedures within brainstem structures, corticosteroids are routinely administered to prevent oedema and to reduce intraoperative trauma, After replacing the routine administration of dexamethasone (DX) by high-dose methylprednisolone (MP) during surgery for tumours within brainstem structures, a decreased incidence of intraoperative haemodynamic instability events was observed. To test this hypothesis, a retrospective analysis was performed.
   Methods: Peroperative data of 62 surgical procedures of brainstem tumours were retrospectively analysed with respect to haemodynamic instability requiring changes in surgical strategy and/or emergence medication with vasoactive drugs. Severe changes in haemodynamic parameters were defined as a significant increase or decrease in heart rate and/or mean arterial blood pressure greater than 30% compared to baseline values. From 1988 to 1994, intravenous dexamethasone was given peroperatively in 33 patients. After a bolus of 1 mg kg(-1) body weight (BW) 30 min preoperatively, 0.2 mg kg(-1) were given every 4 h. From 1994 until now, methylprednisolone was administered instead of dexamethasone in 29 patients. After an initial bolus of 30 mg kg(-1) BW immediately before surgery, 5.4 mg kg(-1) h(-1) were given 23 h postoperatively.
   Results: The results of this retrospective analysis suggest that the number of operations with episodes of bradycardia, arterial hypotension (P<0.05), tachycardia and arterial hypertension (P<0.005) was significantly decreased in the group of patients treated with high-dose methylprednisolone.
   Conclusion: The retrospective analysis of the clinical data showed that the routine use of high-dose methylprednisolone was associated with a decreased incidence of haemodynamic instability in a selected group of patients undergoing brainstem surgery. This finding has to be proven in prospective double-blind controlled studies.
   (C) Acta Anaesthesiologica Scandinavica 44 (2000).
C1 Univ Gottingen, Dept Neurosurg, Klin & Poliklin Neurochirurg, D-37075 Gottingen, Germany.
   Univ Gottingen, Zentrum Anaesthesiol Rettungs & Intens Med, D-37075 Gottingen, Germany.
C3 University of Gottingen; University of Gottingen
RP Mursch, K (通讯作者)，Univ Gottingen, Dept Neurosurg, Klin & Poliklin Neurochirurg, Robert Koch Str 40, D-37075 Gottingen, Germany.
RI Buhre, Wolfgang FFA/X-9192-2018
OI Buhre, Wolfgang/0000-0002-8296-9426
CR Behnke J, 1996, CHILD NERV SYST, V12, P125, DOI 10.1007/BF00266812
   Behnke J, 1997, CHILD NERV SYST, V13, P135, DOI 10.1007/s003810050059
   BEHNKE J, 1997, CHILD NERV SYST, V13, P125
   BRACKEN MB, 1993, J NEUROSURG, V79, P500, DOI 10.3171/jns.1993.79.4.0500
   BRACKEN MB, 1990, NEW ENGL J MED, V322, P1405, DOI 10.1056/NEJM199005173222001
   BRACKEN MB, 1992, PARAPLEGIA, V30, P102, DOI 10.1038/sc.1992.34
   Bracken MB, 1997, JAMA-J AM MED ASSOC, V277, P1597, DOI 10.1001/jama.277.20.1597
   BRAUGHLER JM, 1982, J NEUROSURG, V56, P838, DOI 10.3171/jns.1982.56.6.0838
   Bricolo A, 1995, Adv Tech Stand Neurosurg, V22, P261
   EPSTEIN F, 1985, CANCER, V56, P1804, DOI 10.1002/1097-0142(19851001)56:7+<1804::AID-CNCR2820561316>3.0.CO;2-5
   EPSTEIN F, 1986, J NEUROSURG, V64, P11, DOI 10.3171/jns.1986.64.1.0011
   EPSTEIN F, 1987, J NEUROSURG, V67, P483, DOI 10.3171/jns.1987.67.4.0483
   HALL ED, 1992, J NEUROSURG, V76, P13, DOI 10.3171/jns.1992.76.1.0013
   HALL ED, 1984, J NEUROSURG, V61, P124, DOI 10.3171/jns.1984.61.1.0124
   HALL ED, 1992, J NEUROTRAUMA S1, V9, P165
   MURSCH K, 1999, ZENTRALBL NEUROCHI S, P42
   WHITE HH, 1963, NEUROLOGY, V13, P292, DOI 10.1212/WNL.13.4.292
   YOUNG W, 1982, J NEUROSURG, V57, P667, DOI 10.3171/jns.1982.57.5.0667
NR 18
TC 8
Z9 9
U1 0
U2 0
PU MUNKSGAARD INT PUBL LTD
PI COPENHAGEN
PA 35 NORRE SOGADE, PO BOX 2148, DK-1016 COPENHAGEN, DENMARK
SN 0001-5172
J9 ACTA ANAESTH SCAND
JI Acta Anaesthesiol. Scand.
PD APR
PY 2000
VL 44
IS 4
BP 378
EP 382
DI 10.1034/j.1399-6576.2000.440404.x
PG 5
WC Anesthesiology
WE Science Citation Index Expanded (SCI-EXPANDED)
SC Anesthesiology
GA 296TG
UT WOS:000086041400004
PM 10757568
DA 2023-08-27
ER

PT J
AU Dawes, W
   Marcus, HJ
   Tisdall, M
   Aquilina, K
AF Dawes, William
   Marcus, Hani J.
   Tisdall, Martin
   Aquilina, Kristian
TI Robot-assisted stereotactic brainstem biopsy in children: prospective
   cohort study
SO JOURNAL OF ROBOTIC SURGERY
LA English
DT Article
DE Surgery; Robotics; Stereotaxy; Diffuse intrinsic brainstem glioma; DIPG
ID METAANALYSIS; EXPERIENCE; KEYHOLE; GLIOMAS; SERIES; TUMORS
AB Tumours located within the brainstem comprise approximately a tenth of all paediatric brain tumours. Surgical biopsy of these tumours is technically challenging and has historically been associated with considerable risk. To this end, robot-assisted surgery theoretically allows for increased accuracy and precision. In this study we report our experience using the Neuromate robot (Renishaw, Gloucestershire, UK) to perform robot-assisted stereotactic biopsy in children with tumours located within the brainstem. An uncontrolled prospective cohort study was performed (phase II) according to the IDEAL model for safe surgical innovation. All cases were recorded on a prospectively maintained database. The database was searched over a 2-year period between the 1st December 2015 and the 31st November 2017 to identify all children with brainstem tumours that underwent robot-assisted stereotactic brain biopsy. When accessible, the post-operative MRI scans and pre-operative plans were compared to assess the target point localisation error (TPLE). Adverse events were recorded prospectively according to whether they resulted in increased hospital stay, caused neurological injury, or lead to death. In all, 11 consecutive children were identified with brain tumours located within the brainstem. In 10/11 cases specimens were diagnostic; in the remaining case a further biopsy was successful. The most frequent pathology was DIPG (7/15). Seven patients underwent an early post-operative volumetric MRI; the calculated median TPLE was 2.7mm (range 0.5-4.2mm). There were no surgical complications noted. Robot-assisted stereotactic biopsy in children appears to be feasible and safe. Research databases and comparative studies are warranted to further assess the technique.
C1 [Dawes, William; Marcus, Hani J.; Tisdall, Martin; Aquilina, Kristian] Great Ormond St Hosp Sick Children, Dept Neurosurg, London, England.
   [Marcus, Hani J.] UCL, Wellcome EPSRC Ctr Intervent & Surg Sci, 8-02 Malet Pl Bldg,Gower St, London WC1E 6BT, England.
C3 University of London; University College London; Great Ormond Street
   Hospital for Children NHS Foundation Trust; UK Research & Innovation
   (UKRI); Engineering & Physical Sciences Research Council (EPSRC);
   University of London; University College London
RP Marcus, HJ (通讯作者)，Great Ormond St Hosp Sick Children, Dept Neurosurg, London, England.; Marcus, HJ (通讯作者)，UCL, Wellcome EPSRC Ctr Intervent & Surg Sci, 8-02 Malet Pl Bldg,Gower St, London WC1E 6BT, England.
EM
RI Marcus, Hani/E-2885-2013; Aquilina, Kristian/AAF-5334-2019
OI Dawes, William/0000-0003-1828-4542; Tisdall, Martin/0000-0001-8880-8386;
   Aquilina, Kristian/0000-0001-7015-5196
CR ALBRIGHT AL, 1993, NEUROSURGERY, V33, P1026
   Bekelis K, 2012, J NEUROSURG, V116, P1002, DOI 10.3171/2012.1.JNS111746
   Carai A, 2017, WORLD NEUROSURG, V101, P584, DOI 10.1016/j.wneu.2017.02.088
   Coca HA, 2016, NEUROCHIRURGIE, V62, P327, DOI 10.1016/j.neuchi.2016.07.005
   De Benedictis A, 2017, NEUROSURG FOCUS, V42, DOI 10.3171/2017.2.FOCUS16579
   Dellaretti M, 2012, STEREOT FUNCT NEUROS, V90, P79, DOI 10.1159/000335502
   Dlaka D, 2018, INT J MED ROBOT COMP, V14, DOI 10.1002/rcs.1884
   DRAKE JM, 1991, NEUROSURGERY, V29, P27, DOI 10.1227/00006123-199107000-00005
   Glauser D, 1995, J Image Guid Surg, V1, P266, DOI 10.3109/10929089509106332
   Haegelen C, 2010, NEUROCHIRURGIE, V56, P363, DOI 10.1016/j.neuchi.2010.05.006
   Hamisch C, 2017, J NEUROSURG-PEDIATR, V20, P261, DOI 10.3171/2017.2.PEDS1665
   Hargrove D, 2006, LANCET ONCOL, V7, P241, DOI 10.1016/S1470-2045(06)70615-5
   Johnson KJ, 2014, CANCER EPIDEM BIOMAR, V23, P2716, DOI 10.1158/1055-9965.EPI-14-0207
   KWOH YS, 1988, IEEE T BIO-MED ENG, V35, P153, DOI 10.1109/10.1354
   Lefranc M, 2015, J NEUROSURG, V122, P342, DOI 10.3171/2014.9.JNS14107
   Mackay A, 2017, CANCER CELL, V32, P520, DOI 10.1016/j.ccell.2017.08.017
   Marcus H, 2013, IEEE T BIO-MED ENG, V60, P674, DOI 10.1109/TBME.2013.2243731
   Marcus HJ, 2014, OPER NEUROSURG, V10, P84, DOI 10.1227/NEU.0000000000000123
   Marcus HJ, 2018, CHILDS NERV SYST
   Miller BA, 2017, J NEUROSURG-PEDIATR, V20, P364, DOI 10.3171/2017.5.PEDS1782
   Minchev G, 2017, J NEUROSURG, V126, P985, DOI 10.3171/2016.1.JNS152005
   Perez-Gomez JL, 2010, CHILD NERV SYST, V26, P29, DOI 10.1007/s00381-009-1000-x
   Puget S, 2015, CHILD NERV SYST, V31, P1773, DOI 10.1007/s00381-015-2832-1
   Quick-Weller J, 2017, J CLIN NEUROSCI, V44, P254, DOI 10.1016/j.jocn.2017.06.028
   Willems PWA, 2003, ACTA NEUROCHIR, V145, P889, DOI 10.1007/s00701-003-0112-4
NR 25
TC 14
Z9 17
U1 3
U2 4
PU SPRINGERNATURE
PI LONDON
PA CAMPUS, 4 CRINAN ST, LONDON, N1 9XW, ENGLAND
SN 1863-2483
EI 1863-2491
J9 J ROBOT SURG
JI J. Robot. Surg.
PD AUG
PY 2019
VL 13
IS 4
BP 575
EP 579
DI 10.1007/s11701-018-0899-x
PG 5
WC Surgery
WE Science Citation Index Expanded (SCI-EXPANDED)
SC Surgery
GA IJ0WK
UT WOS:000475620500006
PM 30523502
OA Green Published, hybrid
DA 2023-08-27
ER

PT J
AU Caldarelli, M
   Colosimo, C
   Di Rocco, C
AF Caldarelli, M
   Colosimo, C
   Di Rocco, C
TI Intra-axial dermoid/epidermoid tumors of the brainstem in children
SO SURGICAL NEUROLOGY
LA English
DT Article
DE brain stem tumour; dermoid cysts; epidermoid cysts; MRI; surgical
   treatment
ID EPIDERMOID CYSTS; MR
AB BACKGROUND Epidermoid and dermoid cysts are rare intracranial space-occupying lesions, which account for about 1% of all intracranial tumors; they are even rarer in the pediatric age group. Among the varius locations, that inside the brain stem is quite exceptional (only 12 cases reported to date in the literature). We report two additional cases occurring in children, underlining their clinical characteristics, the difficulties faced in the diagnostic work-up, and the surgical treatment adopted (which consisted of removal of the cyst contents and partial resection of the cyst membrane, because of its adherence to the surrounding nervous and vascular structures).
   METHODS We report two cases of intraaxial dermoid/epidermoid cysts observed within the last 5 years. Both patients complained of cervico-nuchal pain, with a remittent/intermittent character in one of them; this patient also exhibited transient 6th and 7th cranial nerve deficit. One lesion was approached through the floor of the fourth ventricle, the second one through the ventrolateral aspect of the brainstem.
   RESULTS Subtotal tumor resection was achieved in both cases. Both patients had a smooth postoperative course without permanent neurological deficits. In the first patient residual tumor remains stable on follow-up MRI. In the second one, early tumor regrowth necessitated a second operation, after which the residual tumor has remained stable.
   CONCLUSIONS Based on our experience we suggest that a cautious surgical approach ("conservative" resection) to these lesions is the best choice. In fact, attempts at radical removal carry unacceptably high morbidity and mortality rates. (C) 2001 by Elsevier Science Inc.
C1 Univ Cattolica Sacro Cuore, Sch Med, Sect Pediat Neurosurg, Dept Neurosurg, I-00168 Rome, Italy.
   Univ Cattolica Sacro Cuore, Sch Med, Dept Radiol, I-00168 Rome, Italy.
C3 Catholic University of the Sacred Heart; IRCCS Policlinico Gemelli;
   Catholic University of the Sacred Heart; IRCCS Policlinico Gemelli
RP Caldarelli, M (通讯作者)，Univ Cattolica Sacro Cuore, Sch Med, Sect Pediat Neurosurg, Dept Neurosurg, Largo A Gemelli 8, I-00168 Rome, Italy.
CR ALVORD EC, 1977, ANN NEUROL, V2, P367, DOI 10.1002/ana.410020504
   Baxter JW, 1985, NEUROSURGERY, V655, P61
   BERGER MS, 1985, J NEUROSURG, V62, P214, DOI 10.3171/jns.1985.62.2.0214
   BHATIA R, 1978, NEUROL INDIA, V26, P76
   CALDARELLI M, 1999, PEDIAT NEUROSURGERY, P486
   Conley FK, 1985, NEUROSURGERY, P668
   DUNN LT, 1994, CHILD NERV SYST, V10, P529, DOI 10.1007/BF00335076
   EEKHOF JLA, 1985, SURG NEUROL, V23, P189, DOI 10.1016/0090-3019(85)90344-1
   FORD K, 1981, J COMPUT ASSIST TOMO, V5, P895, DOI 10.1097/00004728-198112000-00021
   FOURNIER D, 1992, CHILD NERV SYST, V8, P471, DOI 10.1007/BF00274412
   GUALDI GF, 1991, AM J NEURORADIOL, V12, P771
   GUIDETTI B, 1977, J NEUROSURG, V47, P12, DOI 10.3171/jns.1977.47.1.0012
   GUY G, 1989, NEUROCHIRURGIE    S1, P1
   IIHARA K, 1989, SURG NEUROL, V32, P377, DOI 10.1016/0090-3019(89)90143-2
   LEAL O, 1978, J NEUROSURG, V48, P811, DOI 10.3171/jns.1978.48.5.0811
   NETSKY MG, 1988, SURG NEUROL, V29, P477, DOI 10.1016/0090-3019(88)90144-9
   OBANA WG, 1991, J NEUROSURG, V74, P123, DOI 10.3171/jns.1991.74.1.0123
   OBRADOR S, 1969, J NEUROL NEUROSUR PS, V32, P450, DOI 10.1136/jnnp.32.5.450
   OGAWA T, 1985, Neurologia Medico-Chirurgica, V25, P393, DOI 10.2176/nmc.25.393
   Russel DS, 1989, PATHOLOGY TUMOURS NE, P690
   SABIN HI, 1987, NEUROSURGERY, V21, P798, DOI 10.1227/00006123-198712000-00004
   SCHWARTZ JF, 1978, NEUROLOGY, V28, P124, DOI 10.1212/WNL.28.2.124
   TOGLIA JU, 1965, J NEUROSURG, V23, P384, DOI 10.3171/jns.1965.23.4.0384
   TSURUDA JS, 1990, AM J NEURORADIOL, V11, P925
   ULRICH J, 1964, J NEUROSURG, V21, P1051, DOI 10.3171/jns.1964.21.12.1051
   VIONDURY J, 1987, NEURORADIOLOGY, V29, P333, DOI 10.1007/BF00348910
   WEAVER EN, 1979, J NEUROSURG, V51, P254, DOI 10.3171/jns.1979.51.2.0254
   Wilson CB, 1996, SURG NEUROL, V45, P540, DOI 10.1016/S0090-3019(96)90749-1
   YAMAKAWA K, 1989, NEUROSURGERY, V24, P568, DOI 10.1227/00006123-198904000-00013
   YASARGIL MG, 1989, NEUROSURGERY, V24, P561
   Yoshizato K, 1996, SURG NEUROL, V45, P537, DOI 10.1016/0090-3019(95)00470-X
NR 31
TC 54
Z9 64
U1 0
U2 0
PU ELSEVIER SCIENCE INC
PI NEW YORK
PA STE 800, 230 PARK AVE, NEW YORK, NY 10169 USA
SN 0090-3019
J9 SURG NEUROL
JI Surg. Neurol.
PD AUG
PY 2001
VL 56
IS 2
BP 97
EP 105
DI 10.1016/S0090-3019(01)00542-0
PG 9
WC Clinical Neurology; Surgery
WE Science Citation Index Expanded (SCI-EXPANDED)
SC Neurosciences & Neurology; Surgery
GA 479VG
UT WOS:000171423800012
PM 11580945
DA 2023-08-27
ER

PT J
AU Postovsky, S
   Eran, A
   Ben Arush, MW
AF Postovsky, Sergey
   Eran, Ayelet
   Ben Arush, Myriam Weyl
TI Unusual case of leptomeningeal dissemination of a diffuse pontine
   high-grade astrocytoma in a child
SO PEDIATRIC NEUROSURGERY
LA English
DT Article
DE astrocytoma; brain stem; diffuse pontine glioma
ID BRAIN-STEM GLIOMAS; HIGH-DOSE-CHEMOTHERAPY; HYPERFRACTIONATED
   RADIOTHERAPY; PHASE-I/II; TUMORS; METASTASES; DIAGNOSIS; THERAPY
AB A 12-year-old girl was seen in our hospital and diagnosed with diffuse pontine glioma. Treatment with radiotherapy was suggested, but the parents rejected this plan and took the child to another hospital where partial resection of the tumor was performed. The histological diagnosis was anaplastic astrocytoma. Returning to our hospital, the parents agreed to radiotherapy which was given in a total dose of 54 Gy. However, 3 months after completion of radiotherapy, the child developed widespread leptomeningeal dissemination of her disease and died 12 months after the initial diagnosis. Since no effective curative methods of treatment exist for patients with diffuse pontine gliomas and surgical intervention is obsolete in this situation and may facilitate leptomeningeal dissemination of the disease, further innovative methods of treatment are needed to improve the bleak prognosis of these patients. Copyright (C) 2008 S. Karger AG, Basel.
C1 [Postovsky, Sergey; Ben Arush, Myriam Weyl] Technion Israel Inst Technol, Dept Pediat Hematol Oncol, Haifa, Israel.
   [Eran, Ayelet] Technion Israel Inst Technol, Dept Med Imaging, Haifa, Israel.
   [Eran, Ayelet] Technion Israel Inst Technol, Fac Med, Haifa, Israel.
C3 Technion Israel Institute of Technology; Technion Israel Institute of
   Technology; Technion Israel Institute of Technology; Rappaport Faculty
   of Medicine
RP Postovsky, S (通讯作者)，Meyer Childrens Hosp, Rambam Med Ctr, Dept Pediat Hematol Oncol, POB 9602, IL-31096 Haifa, Israel.
EM
CR ALBRIGHT AL, 1993, NEUROSURGERY, V33, P1026
   Allen J, 1999, CANCER, V86, P1064, DOI 10.1002/(SICI)1097-0142(19990915)86:6<1064::AID-CNCR24>3.0.CO;2-1
   Allen JC, 1996, PEDIATR NEUROSURG, V24, P98, DOI 10.1159/000121024
   ATAC MS, 1979, CLIN NEUROL NEUROSUR, V81, P281, DOI 10.1016/0303-8467(79)90032-5
   Bouffet E, 2000, CANCER, V88, P685, DOI 10.1002/(SICI)1097-0142(20000201)88:3<685::AID-CNCR27>3.0.CO;2-K
   Broniscer A, 2005, CANCER-AM CANCER SOC, V103, P133, DOI 10.1002/cncr.20741
   Carrie Christian, 2004, Bull Cancer, V91, pE167
   Dunkel IJ, 1996, PEDIATR NEUROSURG, V24, P263, DOI 10.1159/000121049
   EPSTEIN F, 1988, J NEURO-ONCOL, V6, P309
   Epstein F, 1990, Neurosurg Clin N Am, V1, P111
   Gururangan S, 2006, J NEURO-ONCOL, V77, P207, DOI 10.1007/s11060-005-9029-5
   Kaplan AM, 1996, PEDIATR NEUROSURG, V24, P185, DOI 10.1159/000121036
   KEPES JJ, 1976, J NEUROL NEUROSUR PS, V39, P66, DOI 10.1136/jnnp.39.1.66
   Mandell LR, 1999, INT J RADIAT ONCOL, V43, P959, DOI 10.1016/S0360-3016(98)00501-X
   Motoyama Y, 2002, NEUROL MED-CHIR, V42, P309, DOI 10.2176/nmc.42.309
   O'Brien DF, 2006, J NEUROSURG, V105, P219, DOI 10.3171/ped.2006.105.3.219
   Packer R J, 1992, Neurosurg Clin N Am, V3, P863
   PACKER RJ, 1985, NEUROLOGY, V35, P397, DOI 10.1212/WNL.35.3.397
   PACKER RJ, 1983, ANN NEUROL, V14, P177, DOI 10.1002/ana.410140204
   PACKER RJ, 1994, SEMIN ONCOL, V21, P260
   PACKER RJ, 1993, CANCER-AM CANCER SOC, V72, P1414, DOI 10.1002/1097-0142(19930815)72:4<1414::AID-CNCR2820720442>3.0.CO;2-C
   PACKER RJ, 1994, CANCER-AM CANCER SOC, V74, P1827, DOI 10.1002/1097-0142(19940915)74:6<1827::AID-CNCR2820740628>3.0.CO;2-Q
   RENIER WO, 1980, NEUROPEDIATRICS, V11, P262, DOI 10.1055/s-2008-1071395
   ROBERTSON PL, 1994, NEUROLOGY, V44, P1798, DOI 10.1212/WNL.44.10.1798
   Shuper A, 1998, CHILD NERV SYST, V14, P578, DOI 10.1007/s003810050275
   Wagner S, 2006, BRIT J CANCER, V95, P991, DOI 10.1038/sj.bjc.6603402
   Wolff JEA, 2002, BRIT J CANCER, V87, P945, DOI 10.1038/sj.bjc.6600552
NR 27
TC 2
Z9 2
U1 0
U2 1
PU KARGER
PI BASEL
PA ALLSCHWILERSTRASSE 10, CH-4009 BASEL, SWITZERLAND
SN 1016-2291
EI 1423-0305
J9 PEDIATR NEUROSURG
JI Pediatr. Neurosurg.
PY 2008
VL 44
IS 3
BP 208
EP 211
DI 10.1159/000120152
PG 4
WC Clinical Neurology; Pediatrics; Surgery
WE Science Citation Index Expanded (SCI-EXPANDED)
SC Neurosciences & Neurology; Pediatrics; Surgery
GA 311AF
UT WOS:000256571600006
PM 18334845
DA 2023-08-27
ER

PT J
AU Van Poppel, K
   Broniscer, A
   Patay, Z
   Morris, EB
AF Van Poppel, Katherine
   Broniscer, Alberto
   Patay, Zoltan
   Morris, E. Brannon
TI Alexander Disease: An Important Mimicker of Focal Brainstem Glioma
SO PEDIATRIC BLOOD & CANCER
LA English
DT Article
DE brainstem glioma; Alexander's disease; brain tumor mimic; neuro-oncology
ID FIBRILLARY ACIDIC PROTEIN; JUVENILE; MUTATIONS; DIAGNOSIS; GFAP
AB We report the case of a 6-year-old male who was referred to a tertiary oncology center with a focal brainstem lesion which was presumed to be neoplastic. Due to the symmetric nature of the lesion oil magnetic resonance imaging, the evaluation was expanded to investigate other possible Causes and eventual diagnosis of Alexander's disease (AD) was made. AD is a neurodegenerative disease which Must be included in the differential for tumor-like lesions within the posterior fossa. Pediatr Blood Cancer 2009;53: 1355-1356. (C) 2009 Wiley-Liss, Inc.
C1 [Van Poppel, Katherine; Morris, E. Brannon] St Jude Childrens Hosp, Div Neurol, Memphis, TN 38105 USA.
   [Broniscer, Alberto; Morris, E. Brannon] St Jude Childrens Hosp, Div Neurooncol, Memphis, TN 38105 USA.
   [Patay, Zoltan] St Jude Childrens Hosp, Div Diagnost Imaging, Memphis, TN 38105 USA.
C3 St Jude Children's Research Hospital; St Jude Children's Research
   Hospital; St Jude Children's Research Hospital
RP Morris, EB (通讯作者)，St Jude Childrens Hosp, Div Neurol, 262 Danny Thomas Pl, Memphis, TN 38105 USA.
EM
RI Patay, Zoltan/N-6994-2018
OI Patay, Zoltan/0000-0003-1479-9864; Broniscer,
   Alberto/0000-0001-9646-0463
FU National Institutes of Health [P30 CA 21765]; National Institute of
   Child Health and Human Development [HD49888]; American Lebanese Syrian
   Associated Charities (ALSAC)
FX Grant sponsor: National Institutes of Health; Grant number: P30 CA
   21765;; Grant sponsor: National Institute of Child Health and Human
   Development;; Grant number: HD49888;; Grant sponsor: American Lebanese
   Syrian Associated Charities (ALSAC).
CR Brenner M, 2001, NAT GENET, V27, P117, DOI 10.1038/83679
   Franzoni E, 2006, J CHILD NEUROL, V21, P1075, DOI 10.1177/7010.2006.00235
   KAYE EM, 2006, PEDIAT NEUROLOGY PRI, P1356
   Li R, 2005, ANN NEUROL, V57, P310, DOI 10.1002/ana.20406
   Mignot C, 2009, BRAIN DEV-JPN, V31, P244, DOI 10.1016/j.braindev.2008.05.005
   Probst EN, 2003, ANN NEUROL, V53, P118, DOI 10.1002/ana.10382
   Quinlan RA, 2007, EXP CELL RES, V313, P2077, DOI 10.1016/j.yexcr.2007.04.004
   Rutka JT, 1997, J NEUROSURG, V87, P420, DOI 10.3171/jns.1997.87.3.0420
   van der Knaap MS, 2006, NEUROLOGY, V66, P494, DOI 10.1212/01.wnl.0000198770.80743.37
   van der Knaap MS, 2005, ANN NEUROL, V57, P327, DOI 10.1002/ana.20381
   van der Knaap MS, 2001, AM J NEURORADIOL, V22, P541
NR 11
TC 10
Z9 10
U1 0
U2 0
PU WILEY
PI HOBOKEN
PA 111 RIVER ST, HOBOKEN 07030-5774, NJ USA
SN 1545-5009
EI 1545-5017
J9 PEDIATR BLOOD CANCER
JI Pediatr. Blood Cancer
PD DEC 15
PY 2009
VL 53
IS 7
BP 1355
EP 1356
DI 10.1002/pbc.22232
PG 2
WC Oncology; Hematology; Pediatrics
WE Science Citation Index Expanded (SCI-EXPANDED)
SC Oncology; Hematology; Pediatrics
GA 513ZV
UT WOS:000271363800039
PM 19672978
OA Green Accepted
DA 2023-08-27
ER

PT J
AU Deligne, C
   Hachani, J
   Duban-Deweer, S
   Meignan, S
   Leblond, P
   Carcaboso, AM
   Sano, Y
   Shimizu, F
   Kanda, T
   Gosselet, F
   Dehouck, MP
   Mysiorek, C
AF Deligne, Clemente
   Hachani, Johan
   Duban-Deweer, Sophie
   Meignan, Samuel
   Leblond, Pierre
   Carcaboso, Angel M.
   Sano, Yasuteru
   Shimizu, Fumitaka
   Kanda, Takashi
   Gosselet, Fabien
   Dehouck, Marie-Pierre
   Mysiorek, Caroline
TI Development of a human in vitro blood-brain tumor barrier model of
   diffuse intrinsic pontine glioma to better understand the
   chemoresistance
SO FLUIDS AND BARRIERS OF THE CNS
LA English
DT Article
DE Blood-brain barrier; Blood-brain tumor barrier; Diffuse intrinsic
   pontine glioma; Chemoresistance; Pediatric brain cancer
ID TIGHT JUNCTIONAL MOLECULES; CENTRAL-NERVOUS-SYSTEM; TEMOZOLOMIDE;
   PERICYTES; PHARMACOKINETICS; TRANSPORTERS; ENZYMES
AB Background Pediatric diffuse intrinsic pontine glioma (DIPG) represents one of the most devastating and lethal brain tumors in children with a median survival of 12 months. The high mortality rate can be explained by the ineligibility of patients to surgical resection due to the diffuse growth pattern and midline localization of the tumor. While the therapeutic strategies are unfortunately palliative, the blood-brain barrier (BBB) is suspected to be responsible for the treatment inefficiency. Located at the brain capillary endothelial cells (ECs), the BBB has specific properties to tightly control and restrict the access of molecules to the brain parenchyma including chemotherapeutic compounds. However, these BBB specific properties can be modified in a pathological environment, thus modulating brain exposure to therapeutic drugs. Hence, this study aimed at developing a syngeneic human blood-brain tumor barrier model to understand how the presence of DIPG impacts the structure and function of brain capillary ECs. Methods A human syngeneic in vitro BBB model consisting of a triple culture of human (ECs) (differentiated from CD34(+)-stem cells), pericytes and astrocytes was developed. Once validated in terms of BBB phenotype, this model was adapted to develop a blood-brain tumor barrier (BBTB) model specific to pediatric DIPG by replacing the astrocytes by DIPG-007, -013 and -014 cells. The physical and metabolic properties of the BBTB ECs were analyzed and compared to the BBB ECs. The permeability of both models to chemotherapeutic compounds was evaluated. Results In line with clinical observation, the integrity of the BBTB ECs remained intact until 7 days of incubation. Both transcriptional expression and activity of efflux transporters were not strongly modified by the presence of DIPG. The permeability of ECs to the chemotherapeutic drugs temozolomide and panobinostat was not affected by the DIPG environment. Conclusions This original human BBTB model allows a better understanding of the influence of DIPG on the BBTB ECs phenotype. Our data reveal that the chemoresistance described for DIPG does not come from the development of a "super BBB". These results, validated by the absence of modification of drug transport through the BBTB ECs, point out the importance of understanding the implication of the different protagonists in the pathology to have a chance to significantly improve treatment efficiency.
C1 [Deligne, Clemente; Gosselet, Fabien; Dehouck, Marie-Pierre; Mysiorek, Caroline] Univ Artois, Lab Barriere Hematoencephal LBHE, UR 2465, F-62300 Lens, France.
   [Hachani, Johan; Duban-Deweer, Sophie] Univ Artois, Lab Barriere Hematoencephal LBHE, Plateau Spectrometrie Masse ARTois SMART, UR 2465, F-62300 Lens, France.
   [Meignan, Samuel] INSERM, U908, F-59000 Lille, France.
   [Meignan, Samuel] Inst Rech Canc Lille IRCL, F-59000 Lille, France.
   [Meignan, Samuel] Ctr Oscar Lambret, Unite Tumorigenese & Resistance Traitements, 3 Rue Frederic Combemale, F-59000 Lille, France.
   [Leblond, Pierre] Inst Hematol & Oncol Pediat, Dept Cancerol Pediat, F-69000 Lyon, France.
   [Carcaboso, Angel M.] Inst Recerca St Joan Deu, Barcelona 08950, Spain.
   [Sano, Yasuteru; Shimizu, Fumitaka; Kanda, Takashi] Yamaguchi Univ, Dept Neurol & Clin Neurosci, Grad Sch Med, Ube, Yamaguchi, Japan.
C3 Universite d'Artois; Universite d'Artois; Institut National de la Sante
   et de la Recherche Medicale (Inserm); Universite de Lille - ISITE;
   Universite de Lille; UNICANCER; Centre Oscar Lambret; Yamaguchi
   University
RP Mysiorek, C (通讯作者)，Univ Artois, Lab Barriere Hematoencephal LBHE, UR 2465, F-62300 Lens, France.
EM
RI Meignan, Samuel/E-9628-2017; Gosselet, Fabien/U-8427-2019
OI Gosselet, Fabien/0000-0002-0481-5026; Carcaboso,
   Angel/0000-0002-8485-426X; Mysiorek, Caroline/0000-0003-2338-8821
FU Conseil regional du Nord-Pas-de-Calais; Conseil Regional Hauts de
   France; European Regional Development Fund (ERDF); Artois University;
   "Societe Francaise de lutte contre les Cancers et les leucemies de
   l'Enfant et de l'adolescent"(SFCE); Association"l'etoile de Martin";
   Association "Cassandra contre la leucemie"
FX This study is granted by the `Conseil regional du Nord-Pas-de-Calais'
   (Fellowship to C. Deligne). The funders had no role in study design,
   data collection and analysis, decision to publish, or preparation of the
   manuscript. The mass spectrometer of the Spectrometrie de Masse de
   l'Artois (SMART) core facilities used in this study was funded by the
   European Regional Development Fund (ERDF), the Conseil Regional Hauts de
   France and Artois University. This work was granted by the"Societe
   Francaise de lutte contre les Cancers et les leucemies de l'Enfant et de
   l'adolescent"(SFCE), the Association"l'etoile de Martin"and the
   Association "Cassandra contre la leucemie".
CR Abbott NJ, 2013, J INHERIT METAB DIS, V36, P437, DOI 10.1007/s10545-013-9608-0
   Abbott NJ, 2006, NAT REV NEUROSCI, V7, P41, DOI 10.1038/nrn1824
   Arvanitis CD, 2020, NAT REV CANCER, V20, P26, DOI 10.1038/s41568-019-0205-x
   Cecchelli R, 2014, PLOS ONE, V9, DOI 10.1371/journal.pone.0099733
   Chaves C, 2020, PHARMACEUTICS, V12, DOI 10.3390/pharmaceutics12050399
   Decleves X, 2011, CURR DRUG METAB, V12, P732, DOI 10.2174/138920011798357024
   Drolez A, 2016, INT J MOL SCI, V17, DOI 10.3390/ijms17081309
   Drolez A, 2016, PLOS ONE, V11, DOI 10.1371/journal.pone.0151155
   Dutheil F, 2008, BIOCHIMIE, V90, P426, DOI 10.1016/j.biochi.2007.10.007
   El-Khouly FE, 2019, J NEURO-ONCOL, V145, P177, DOI 10.1007/s11060-019-03287-9
   Ghosh C, 2016, DRUG DISCOV TODAY, V21, P1609, DOI 10.1016/j.drudis.2016.06.004
   Gomez-Zepeda D, 2020, PHARMACEUTICS, V12, DOI 10.3390/pharmaceutics12010020
   Grasso CS, 2015, NAT MED, V21, P555, DOI 10.1038/nm.3855
   Gwak HS, 2017, CRIT REV ONCOL HEMAT, V120, P111, DOI 10.1016/j.critrevonc.2017.10.013
   Halvorson KG, 2015, PLOS ONE, V10, DOI 10.1371/journal.pone.0118926
   Hennika T, 2017, PLOS ONE, V12, DOI 10.1371/journal.pone.0169485
   Hennika T, 2016, J CHILD NEUROL, V31, P1377, DOI 10.1177/0883073815601495
