## Supplemental Material 1 for "Bibliometric analysis of emerging trends and research foci in brainstem tumor field over thirty years (1992-2023)": savedrecs (2).txt

FN Clarivate Analytics Web of Science
VR 1.0
PT J
AU Du, C
   Li, ZH
   Wang, ZJ
   Wang, LP
   Tian, Y
AF Du, Chao
   Li, Zhaohui
   Wang, Zhijia
   Wang, Liping
   Tian, Yu
TI Stereotactic aspiration combined with gamma knife radiosurgery for the
   treatment of cystic brainstem metastasis originating from lung
   adenosquamous carcinoma: A case report
SO ONCOLOGY LETTERS
LA English
DT Article
DE adenosquamous carcinoma; gamma knife radiosurgery; brainstem metastasis;
   stereotactic aspiration
ID PROGNOSTIC-FACTORS; CLINICAL ARTICLE; SURVIVAL; SURGERY; OUTCOMES;
   TUMORS; EXPERIENCE; MANAGEMENT; RESECTION
AB Brainstem metastases have a poor prognosis and are difficult to manage. The present study describes the first case of histopathologically-confirmed brainstem metastasis originating from lung adenosquamous carcinoma, and discusses the outcomes of treatment by stereotactic aspiration combined with gamma knife radiosurgery (GKRS). A 59-year-old female presented with a cystic mass (15x12x13 mm; volume, 1.3 cm(3)) located in the pons, two years following surgical treatment for adenosquamous carcinoma of the lung. The patient received initial GKRS for the lesion in the pons with a total dose of 54.0 Gy, however, the volume of the mass subsequently increased to 3.9 cm(3) over a period of three months. Computed tomography-guided stereotactic biopsy and aspiration of the intratumoral cyst were performed, yielding 2.0 cm(3) of yellow-white fluid. Histology confirmed the diagnosis of adenosquamous carcinoma. Aspiration provided immediate symptomatic relief, and was followed one week later by repeat GKRS with a dose of 12.0 Gy. The patient survived for 12 months following the repeat GKRS; however, later succumbed to the disease after lapsing into a two-week coma. The findings of this case suggest that stereotactic aspiration of cysts may improve the effects of GKRS for the treatment of cystic brainstem metastasis; the decrease in tumor volume allowed a higher radiation dose to be administered with a lower risk of radiation-induced side effects. Therefore, stereotactic aspiration combined with GKRS may be an effective treatment for brainstem metastasis originating from adenosquamous carcinoma.
C1 [Du, Chao; Li, Zhaohui; Tian, Yu] Jilin Univ, China Japan Union Hosp, Dept Neurosurg, Changchun 130033, Jilin, Peoples R China.
   [Wang, Zhijia] Jilin Univ, China Japan Union Hosp, Dept Radiol, Changchun 130033, Jilin, Peoples R China.
   [Wang, Liping] Jilin Univ, China Japan Union Hosp, Dept Pathol, Changchun 130033, Jilin, Peoples R China.
C3 Jilin University; Jilin University; Jilin University
RP Tian, Y (通讯作者)，Jilin Univ, China Japan Union Hosp, Dept Neurosurg, 126 Xiantai St, Changchun 130033, Jilin, Peoples R China.
EM
CR Aung T. H., 1997, Hong Kong Medical Journal, V3, P232
   Cooke DT, 2010, ANN THORAC SURG, V90, P943, DOI 10.1016/j.athoracsur.2010.05.025
   Filosso PL, 2011, LUNG CANCER, V74, P25, DOI 10.1016/j.lungcan.2011.01.030
   Fuentes S, 2006, NEUROSURGERY, V58, P37, DOI 10.1227/01.NEU.0000190655.95669.5C
   Gawrychowski J, 2005, EUR J CARDIO-THORAC, V27, P686, DOI 10.1016/j.ejcts.2004.12.030
   Hatiboglu MA, 2011, NEUROSURGERY, V69, P796, DOI 10.1227/NEU.0b013e31821d31de
   Higuchi F, 2012, J NEUROSURG, V117, P17, DOI 10.3171/2012.7.GKS121001
   Hofmann BM, 2012, J NEUROSURG, V116, P373, DOI 10.3171/2011.6.JNS081451
   Huang CF, 1999, J NEUROSURG, V91, P563, DOI 10.3171/jns.1999.91.4.0563
   Hussain A, 2007, INT J RADIAT ONCOL, V67, P521, DOI 10.1016/j.ijrobp.2006.08.081
   Jung EW, 2013, CLIN NEUROL NEUROSUR, V115, P2023, DOI 10.1016/j.clineuro.2013.06.012
   Kased N, 2008, J NEURO-ONCOL, V86, P195, DOI 10.1007/s11060-007-9458-4
   Kawabe T, 2012, J NEUROSURG, V117, P23, DOI 10.3171/2012.7.GKS12977
   Kelly PJ, 2011, J NEURO-ONCOL, V104, P553, DOI 10.1007/s11060-010-0514-0
   Kilburn JM, 2014, J NEURO-ONCOL, V117, P167, DOI 10.1007/s11060-014-1373-x
   Kim IY, 2014, J CLIN NEUROSCI, V21, P694, DOI 10.1016/j.jocn.2013.05.027
   Koyfman SA, 2010, INT J RADIAT ONCOL, V78, P409, DOI 10.1016/j.ijrobp.2009.07.1750
   Lamm AF, 2013, ISRN SURG, V2013, DOI DOI 10.1155/2013/652895236913653649612
   Li YH, 2012, J NEUROSURG, V117, P13, DOI 10.3171/2012.7.GKS121020
   Lin CS, 2012, NEUROSURGERY, V70, P953, DOI 10.1227/NEU.0b013e31823c40fe
   Lorenzoni JG, 2009, SURG NEUROL, V71, P188, DOI 10.1016/j.surneu.2008.01.029
   Lu YB, 2014, ACTA RADIOL, V55, P121, DOI 10.1177/0284185113493272
   Maranzano E, 2005, TUMORI J, V91, P325, DOI 10.1177/030089160509100408
   Matsumoto K, 1998, J NEUROSURG, V88, P141, DOI 10.3171/jns.1998.88.1.0141
   Nakagawa K, 2003, ANN THORAC SURG, V75, P1740, DOI 10.1016/S0003-4975(03)00022-5
   Park WH, 2009, J KOREAN NEUROSURG S, V46, P360, DOI 10.3340/jkns.2009.46.4.360
   Peterson HE, 2014, INT J MOL SCI, V15, P9748, DOI 10.3390/ijms15069748
   Samblas JM, 2009, CLIN TRANSL ONCOL, V11, P677, DOI 10.1007/s12094-009-0423-x
   Sengoz M, 2013, J NEURO-ONCOL, V113, P33, DOI 10.1007/s11060-013-1086-6
   Server A, 2010, ACTA RADIOL, V51, P316, DOI 10.3109/02841850903482901
   Shuto T, 2003, ACTA NEUROCHIR, V145, P755, DOI 10.1007/s00701-003-0034-1
   Valery CA, 2011, INT J RADIAT ONCOL, V80, P362, DOI 10.1016/j.ijrobp.2010.02.028
   Yen CP, 2006, J NEUROSURG, V105, P213, DOI 10.3171/jns.2006.105.2.213
   Yoo TW, 2011, J KOREAN NEUROSURG S, V50, P299, DOI 10.3340/jkns.2011.50.4.299
   Zhao XL, 2012, NEUROSURG QUART, V22, P22, DOI 10.1097/WNQ.0b013e3182277d55
NR 35
TC 5
Z9 6
U1 0
U2 3
PU SPANDIDOS PUBL LTD
PI ATHENS
PA POB 18179, ATHENS, 116 10, GREECE
SN 1792-1074
EI 1792-1082
J9 ONCOL LETT
JI Oncol. Lett.
PD APR
PY 2015
VL 9
IS 4
BP 1607
EP 1613
DI 10.3892/ol.2015.2968
PG 7
WC Oncology
WE Science Citation Index Expanded (SCI-EXPANDED)
SC Oncology
GA CE2GF
UT WOS:000351631900021
PM 25789009
OA Green Submitted, gold, Green Published
DA 2023-08-27
ER

PT J
AU Fung, MM
   Viveros, OH
   O'Connor, DT
AF Fung, M. M.
   Viveros, O. H.
   O'Connor, D. T.
TI Diseases of the adrenal medulla
SO ACTA PHYSIOLOGICA
LA English
DT Review
DE adrenal; catecholamine; chromaffin; phaeochromocytoma
ID CHROMOGRANIN-A; CLINICAL PRESENTATION; PHEOCHROMOCYTOMA; DIAGNOSIS;
   MUTATIONS; NEUROBLASTOMA; STORAGE
AB The adrenal glands are vital in the organism's response to environmental stress. The outer cortex releases steroid hormones: glucocorticoids, mineralocorticoids and sex hormones, which are crucial to metabolism, inflammatory reactions and fluid homeostasis. The medulla is different developmentally, functionally and structurally. It co-releases catecholamines (primarily adrenaline and to some extent noradrenaline) as well as peptides by the all-or-none process of exocytosis from chromaffin granules, to aid in blood pressure and blood flow regulation, with regulated increments during the activation of the sympathetic nervous system. The co-released peptides function to regulate catecholamine release, blood vessel contraction and innate immune responses. Pathology within the adrenal medulla and the autonomic nervous system is primarily because of neoplasms. The most common tumour, called phaeochromocytoma when located in the adrenal medulla, originates from chromaffin cells and excretes catecholamines, but may be referred to as secreting paragangliomas when found in extra-adrenal chromaffin cells. Neoplasms, such as neuroblastomas and ganglioneuromas, may also be of neuronal lineage. We will also briefly discuss the catecholamine deficiency state.
C1 [Fung, M. M.; Viveros, O. H.; O'Connor, D. T.] Univ Calif San Diego, CHGG, Dept Med & Pharmacol, La Jolla, CA 92093 USA.
C3 University of California System; University of California San Diego
RP O'Connor, DT (通讯作者)，Univ Calif San Diego, CHGG, Dept Med & Pharmacol, Skaggs SSPPS Room4256,9500 Gilman Dr, La Jolla, CA 92093 USA.
EM
FU NHLBI NIH HHS [P01 HL058120-090004, P01 HL058120-099006, P01
   HL058120-09, P01 HL058120] Funding Source: Medline; NIDDK NIH HHS [R01
   DK060702, R01 DK060702-05] Funding Source: Medline
CR Benn DE, 2006, J CLIN ENDOCR METAB, V91, P827, DOI 10.1210/jc.2005-1862
   BRAVO EL, 1981, NEW ENGL J MED, V305, P623, DOI 10.1056/NEJM198109103051107
   Brouwers FM, 2006, ENDOCRIN METAB CLIN, V35, P699, DOI 10.1016/j.ecl.2006.09.014
   Erickson D, 2001, J CLIN ENDOCR METAB, V86, P5210, DOI 10.1210/jc.86.11.5210
   Geoerger B, 2001, CANCER, V91, P1905, DOI 10.1002/1097-0142(20010515)91:10<1905::AID-CNCR1213>3.0.CO;2-4
   Gimenez-Roqueplo AP, 2006, CLIN ENDOCRINOL, V65, P699, DOI 10.1111/j.1365-2265.2006.02714.x
   GOZES I, 1983, REGUL PEPTIDES, V6, P111, DOI 10.1016/0167-0115(83)90004-6
   HAVEKES B, 2007, J CLIN ENDOCRINOL ME
   HSIAO RJ, 1991, MEDICINE, V70, P33, DOI 10.1097/00005792-199101000-00003
   HSIAO RJ, 1990, AM J MED, V88, P607, DOI 10.1016/0002-9343(90)90526-J
   HSIAO RJ, 1990, J CLIN INVEST, V85, P1555, DOI 10.1172/JCI114604
   HSIAO RJ, 1990, KIDNEY INT, V37, P955, DOI 10.1038/ki.1990.71
   ILIAS L, 2003, J CLIN ENDOCR METAB, V88, P4080
   KAILASAM MT, 1995, AM J HYPERTENS, V8, P651, DOI 10.1016/0895-7061(95)00076-2
   Ketch T, 2002, CIRCULATION, V105, P2518, DOI 10.1161/01.CIR.0000017186.52382.F4
   Kim CH, 2002, AM J MED GENET, V108, P140, DOI 10.1002/ajmg.10196
   Lau SK, 2006, AM J SURG PATHOL, V30, P268, DOI 10.1097/01.pas.0000178095.07513.38
   Lenders JWM, 2002, JAMA-J AM MED ASSOC, V287, P1427, DOI 10.1001/jama.287.11.1427
   Maxwell PH, 2005, CANCER CELL, V8, P91, DOI 10.1016/j.ccr.2005.07.012
   Neumann HPH, 2002, NEW ENGL J MED, V346, P1459, DOI 10.1056/NEJMoa020152
   O'Connor DT, 2003, CECILS TXB MED, P1419
   OCONNOR DT, 1983, JAMA-J AM MED ASSOC, V249, P383
   OCONNOR DT, 1983, J CLIN ENDOCR METAB, V56, P582, DOI 10.1210/jcem-56-3-582
   Opocher G, 2006, ANN NY ACAD SCI, V1073, P149, DOI 10.1196/annals.1353.015
   Pacak K, 2007, NAT CLIN PRACT ENDOC, V3, P92, DOI 10.1038/ncpendmet0396
   PARMER RJ, 1988, J HYPERTENS, V6, P187, DOI 10.1097/00004872-198803000-00002
   Rao FW, 2000, HYPERTENSION, V36, P1045, DOI 10.1161/01.HYP.36.6.1045
   Reisch N, 2006, J HYPERTENS, V24, P2331, DOI 10.1097/01.hjh.0000251887.01885.54
   Sawka AM, 2003, J CLIN ENDOCR METAB, V88, P553, DOI 10.1210/jc.2002-021251
   Schiavi F, 2005, JAMA-J AM MED ASSOC, V294, P2057, DOI 10.1001/jama.294.16.2057
   Scholz T, 2007, J CLIN ENDOCR METAB, V92, P1217, DOI 10.1210/jc.2006-1544
   Singh RJ, 2004, CLIN LAB MED, V24, P85, DOI 10.1016/j.cll.2004.01.015
   Taupenot L, 2003, NEW ENGL J MED, V348, P1134, DOI 10.1056/NEJMra021405
NR 33
TC 32
Z9 34
U1 0
U2 8
PU WILEY
PI HOBOKEN
PA 111 RIVER ST, HOBOKEN 07030-5774, NJ USA
SN 1748-1708
EI 1748-1716
J9 ACTA PHYSIOL
JI Acta Physiol.
PD FEB
PY 2008
VL 192
IS 2
BP 325
EP 335
DI 10.1111/j.1748-1716.2007.01809.x
PG 11
WC Physiology
WE Science Citation Index Expanded (SCI-EXPANDED)
SC Physiology
GA 256DI
UT WOS:000252707500017
PM 18021328
OA Green Accepted
DA 2023-08-27
ER

PT J
AU Hoffman, HJ
AF Hoffman, HJ
TI Management of brainstem tumors
SO CRITICAL REVIEWS IN NEUROSURGERY
LA English
DT Article
ID HYPERFRACTIONATED RADIOTHERAPY; STEM GLIOMAS; CHILDREN; CHEMOTHERAPY
C1 HOSP SICK CHILDREN,DEPT SURG,DIV NEUROSURG,TORONTO,ON M5G 1X8,CANADA.
C3 University of Toronto; Hospital for Sick Children (SickKids)
CR Allen JC, 1996, PEDIATR NEUROSURG, V24, P98, DOI 10.1159/000121024
   Alvarez JA, 1996, J NEUROSURG, V85, P1165, DOI 10.3171/jns.1996.85.6.1165
   Burger PC, 1996, PEDIATR NEUROSURG, V24, P35, DOI 10.1159/000121012
   Freeman CR, 1996, PEDIATR NEUROSURG, V24, P103, DOI 10.1159/000121025
   Giovanini MA, 1996, NEUROSURGERY, V39, P404, DOI 10.1097/00006123-199608000-00039
   Hoffman HJ, 1996, PEDIATR NEUROSURG, V24, P256, DOI 10.1159/000121048
   Jennings MT, 1996, J NEURO-ONCOL, V28, P207
   Packer RJ, 1996, PEDIATR NEUROSURG, V24, P217, DOI 10.1159/000121041
NR 8
TC 0
Z9 0
U1 0
U2 0
PU SPRINGER VERLAG
PI NEW YORK
PA 175 FIFTH AVE, NEW YORK, NY 10010
SN 0939-0146
J9 CRIT REV NEUROSURG
JI Crit. Rev. Neurosurg.
PD SEP
PY 1997
VL 7
IS 5
BP 310
EP 314
DI 10.1007/s003290050040
PG 5
WC Neurosciences; Surgery
WE Science Citation Index Expanded (SCI-EXPANDED)
SC Neurosciences & Neurology; Surgery
GA XZ243
UT WOS:A1997XZ24300004
DA 2023-08-27
ER

PT J
AU Qin, B
   Tabbara, AK
   Delalle, I
   Holsapple, J
   Hohler, A
AF Qin, BaiJing
   Tabbara, Abdul Kader
   Delalle, Ivana
   Holsapple, James
   Hohler, Anna
TI An enigmatic brainstem posterior fossa ganglioglioma in an adult
SO INTERNATIONAL JOURNAL OF NEUROSCIENCE
LA English
DT Article
DE gangliogliomatosis; posterior fossa tumor; primary central nervous
   system neoplasm
ID TUMOR
AB Ganglioglioma is a rare central nervous system neoplasm representing 0.4% to 1.7% of all brain tumors and most frequently occurs in the pediatric population with an incidence of 7.6%. These tumors are usually slow-growing and well-circumscribed solid or cystic lesions. Gangliogliomatosis infrequently occurs in the frontal lobe, pineal gland, basal ganglia, hypothalamus, and optic chiasm, with very few reports of brainstem ganglioglioma. We report a case of a 35-year-old female who initially presented with headache, vertigo, ataxia, saccadic dysfunction, dysarthria, and dysmetria for several years due to an unknown etiology. Her brain imaging showed multiple lesions in the pons and the cerebellum with cystic changes and size reduction and enlargement over the next few years while her neurological symptoms continued to worsen. The patient received courses of steroid treatment that improved her neurological symptoms, suggesting an inflammatory component of her disease. Extensive workup for an inflammatory or infectious etiology was unfruitful and two brain biopsies were inconclusive. A third biopsy showed atypical glial nuclei, binucleated cells, and Rosenthal fibers and the presence of BRAF V600E mutation was detected. The diagnosis of gangliogliomatosis was consequently established. This case illustrates that gangliogliomatosis may present with the waxing-and-waning neurological signs and symptoms. It can masquerade inflammatory processes in the central nervous system on brain imaging and deserves careful consideration in the diagnosis of patients with an indolent course of neurological deterioration.
C1 [Qin, BaiJing] Boston Univ, Sch Med, Boston, MA 02118 USA.
   [Tabbara, Abdul Kader; Holsapple, James] Boston Med Ctr, Dept Neurosurg, Boston, MA USA.
   [Delalle, Ivana] Boston Med Ctr, Dept Pathol, Boston, MA USA.
   [Hohler, Anna] Boston Med Ctr, Dept Neurol, Boston, MA USA.
C3 Boston University; Boston Medical Center; Boston Medical Center; Boston
   Medical Center
RP Qin, B (通讯作者)，Boston Med Ctr, Boston, MA 02118 USA.
EM
OI Delalle, Ivana/0000-0002-1873-3064
CR Baussard B, 2007, J NEUROSURG, V107, P286, DOI 10.3171/PED-07/10/286
   Blumcke I, 2002, J NEUROPATH EXP NEUR, V61, P575
   Castillo M, 1998, AM J NEURORADIOL, V19, P807
   Courville CB, 1930, ARCH NEURO PSYCHIATR, V24, P439, DOI 10.1001/archneurpsyc.1930.02220150002001
   Lagares A, 2001, SURG NEUROL, V56, P315, DOI 10.1016/S0090-3019(01)00618-8
   Lantos PL, 1997, GREENFIELDS NEUROPAT, P663
   Louis DN, 2007, ACTA NEUROPATHOL, V114, P547, DOI 10.1007/s00401-007-0278-6
   Provenzale JM, 2000, AM J ROENTGENOL, V174, P859, DOI 10.2214/ajr.174.3.1740859
   Westwood DA, 2009, J CLIN NEUROSCI, V16, P108, DOI 10.1016/j.jocn.2007.11.016
   ZENTNER J, 1994, J NEUROL NEUROSUR PS, V57, P1497, DOI 10.1136/jnnp.57.12.1497
NR 10
TC 5
Z9 5
U1 0
U2 3
PU TAYLOR & FRANCIS LTD
PI ABINGDON
PA 2-4 PARK SQUARE, MILTON PARK, ABINGDON OR14 4RN, OXON, ENGLAND
SN 0020-7454
EI 1563-5279
J9 INT J NEUROSCI
JI Int. J. Neurosci.
PD SEP
PY 2014
VL 124
IS 9
BP 704
EP 706
DI 10.3109/00207454.2013.877901
PG 3
WC Neurosciences
WE Science Citation Index Expanded (SCI-EXPANDED)
SC Neurosciences & Neurology
GA AN3BW
UT WOS:000340460900010
PM 24405263
DA 2023-08-27
ER

PT J
AU Wang, M
   Zhang, Y
   Shi, W
   Zhu, RQ
   Li, H
   Zhao, R
AF Wang, Min
   Zhang, Yi
   Shi, Wei
   Zhu, Renqing
   Li, Hao
   Zhao, Rui
TI Frameless robot-assisted stereotactic biopsy: an effective and minimally
   invasive technique for pediatric diffuse intrinsic pontine gliomas
SO JOURNAL OF NEURO-ONCOLOGY
LA English
DT Article
DE Pediatric; Diffuse intrinsic pontine glioma; Robot; Stereotactic biopsy;
   Diagnosis
ID BRAIN-STEM GLIOMAS; PREDICTORS; ADVANTAGES; CHILDHOOD; ACCURACY
AB Purpose Diffuse intrinsic pontine gliomas (DIPGs) are prone to high surgical risks, and they could even lead to death due to their specific sites. To determine the value of frameless robot-assisted stereotactic biopsies of DIPGs, when compared it with microsurgical biopsies. Methods We conducted a retrospective study of 71 pediatric patients who underwent biopsies from January 2016 to January 2021. (i) group 1: microsurgical biopsies, and (ii) group 2: frameless robot-assisted stereotactic biopsies. Demographic information, neuroimaging characteristics, pathological diagnoses, operation time, postoperative intensive care unit (ICU) stay time, postoperative hospitalization time, complications, cost, and perioperative mortality rate (POMR) were collected for analyses. Results 32 Cases underwent microsurgical biopsies (group 1) and 39 cases underwent frameless robot-assisted stereotactic biopsies (group 2). All cases were accurately diagnosed after surgery. There was no significant difference in gender, age, symptom times and tumor volumes between the two groups (p > 0.05); operation time, postoperative ICU, stay time and postoperative hospitalization time were longer in group 1 than in group 2 (p < 0.001); the intraoperative bleeding volumes and cost were higher in group 1 than in group 2 (p < 0.001). Group 1 patients required more perioperative blood transfusion than group 2 (p = 0.001), and the new neurological impairments were more frequent in group 1 than in group 2 (p = 0.003). The POMR was 9.38% (3/32) in group 1 and 0 in group 2 (p = 0.087). Conclusions Frameless robot-assisted stereotactic biopsy was an effective and minimally invasive technique for pediatric DIPGs.
C1 [Wang, Min; Zhang, Yi; Shi, Wei; Zhu, Renqing; Li, Hao; Zhao, Rui] Fudan Univ, Natl Childrens Med Ctr, Dept Neurosurg, Childrens Hosp, 399 Wan Yuan Ave, Shanghai 201102, Peoples R China.
C3 Fudan University
RP Li, H; Zhao, R (通讯作者)，Fudan Univ, Natl Childrens Med Ctr, Dept Neurosurg, Childrens Hosp, 399 Wan Yuan Ave, Shanghai 201102, Peoples R China.
EM
FU Medical Innovation Research Project, Shanghai Science and Technology
   Innovation Plan of Action [22Y11904400]
FX This study was supported by Medical Innovation Research Project,
   Shanghai Science and Technology Innovation Plan of Action (Grant No.
   22Y11904400).
CR ALBRIGHT AL, 1993, NEUROSURGERY, V33, P1026
   Bjartmarz H, 2007, STEREOT FUNCT NEUROS, V85, P235, DOI 10.1159/000103262
   Carai A, 2017, WORLD NEUROSURG, V101, P584, DOI 10.1016/j.wneu.2017.02.088
   Cavalheiro S, 2015, CHILD NERV SYST, V31, P1815, DOI 10.1007/s00381-015-2799-y
   Chen JG, 2020, BIOORG CHEM, V99, DOI 10.1016/j.bioorg.2020.103847
   Chheda ZS, 2018, J EXP MED, V215, P141, DOI 10.1084/jem.20171046
   Dawes W, 2019, J ROBOT SURG, V13, P575, DOI 10.1007/s11701-018-0899-x
   Dorward NL, 2002, BRIT J NEUROSURG, V16, P110, DOI 10.1080/02688690220131705
   DRAKE JM, 1991, NEUROSURGERY, V29, P27, DOI 10.1227/00006123-199107000-00005
   Epstein F, 1996, PEDIATR NEUROSURG, V24, P24, DOI 10.1159/000121011
   EUHUS DM, 1986, J SURG ONCOL, V31, P229, DOI 10.1002/jso.2930310402
   Gralla J, 2003, ZBL NEUROCHIR, V64, P166
   Grill J, 2012, PEDIATR BLOOD CANCER, V58, P489, DOI 10.1002/pbc.24060
   Gupta M, 2021, J NEUROSURG-PEDIATR, V27, P317, DOI 10.3171/2020.7.PEDS20373
   Hassan H, 2017, J NEURO-ONCOL, V135, P13, DOI 10.1007/s11060-017-2546-1
   Johnson KJ, 2014, CANCER EPIDEM BIOMAR, V23, P2716, DOI 10.1158/1055-9965.EPI-14-0207
   Lefranc M, 2015, J NEUROSURG, V122, P342, DOI 10.3171/2014.9.JNS14107
   MacDonald TJ, 2012, PEDIATR BLOOD CANCER, V58, P487, DOI 10.1002/pbc.24090
   Malone H, 2015, WORLD NEUROSURG, V84, P1084, DOI 10.1016/j.wneu.2015.05.025
   McGirt MJ, 2005, J NEUROSURG, V102, P897, DOI 10.3171/jns.2005.102.5.0897
   Minchev G, 2020, J NEUROSURG, V132, P150, DOI 10.3171/2018.8.JNS182096
   Minchev G, 2017, J NEUROSURG, V126, P985, DOI 10.3171/2016.1.JNS152005
   Moriarty TM, 2000, NEUROSURGERY, V47, P1138, DOI 10.1097/00006123-200011000-00023
   Smith JS, 2005, J NEURO-ONCOL, V73, P173, DOI 10.1007/s11060-004-4208-3
   Taweesomboonyat C, 2019, J NEUROSCI RURAL PRA, V10, P78, DOI 10.4103/jnrp.jnrp_166_18
   Tejada S, 2020, CHILD NERV SYST, V36, P705, DOI 10.1007/s00381-020-04523-8
   Watters DA, 2015, WORLD J SURG, V39, P856, DOI 10.1007/s00268-014-2638-4
   Yasid H, 2019, WORLD NEUROSURG, V123, pE450, DOI 10.1016/j.wneu.2018.11.187
NR 28
TC 0
Z9 0
U1 4
U2 5
PU SPRINGER
PI NEW YORK
PA ONE NEW YORK PLAZA, SUITE 4600, NEW YORK, NY, UNITED STATES
SN 0167-594X
EI 1573-7373
J9 J NEURO-ONCOL
JI J. Neuro-Oncol.
PD OCT
PY 2022
VL 160
IS 1
BP 107
EP 114
DI 10.1007/s11060-022-04122-4
EA AUG 2022
PG 8
WC Oncology; Clinical Neurology
WE Science Citation Index Expanded (SCI-EXPANDED)
SC Oncology; Neurosciences & Neurology
GA 5V1XE
UT WOS:000846702600002
PM 35997920
DA 2023-08-27
ER

PT J
AU Colletti, V
   Carner, M
   Miorelli, V
   Guida, M
   Colletti, L
   Fiorino, F
AF Colletti, V
   Carner, M
   Miorelli, V
   Guida, M
   Colletti, L
   Fiorino, F
TI Cochlear implant failure: Is an auditory brainstem implant the answer?
SO ACTA OTO-LARYNGOLOGICA
LA English
DT Article
DE auditory brainstem implant; auditory neuropathy; cochlear implant;
   cochlear nerve aplasia; cochlear ossification
ID SENSORINEURAL HEARING-LOSS; SPEECH-PERCEPTION; NEUROPATHY;
   LABYRINTHITIS; DEAFNESS; NERVE
AB Objective-To investigate the auditory rehabilitative results achieved in five patients with cochlear implants (CIs) who subsequently received, due to poor results, auditory brainstem implants (ABIs).
   Material and Methods--Between April 1997 and March 2003, 37 patients (age range 14 months to 70 years) were fitted with ABIs in our ENT Department. Fourteen subjects had neurofibromatosis type 2 and 23 were non-tumor patients who had cochlea or cochlear nerve disease. Five subjects had previously been treated with a CI and received an ABI owing to the poor results achieved. One child had bilateral undiagnosed cochlear nerve aplasia and one was suffering from auditory neuropathy; three adults had total cochlear ossification.
   Results-The open-set sentence recognition score (auditory-only mode) 6-8 months after ABI activation ranged from 0% to 100% in adults. In I subject the speech-tracking score was 56 words/min with the ABI. The two children who had achieved no hearing ability with their CI were able to detect sounds and words as early as 3 months after activation of the ABI.
   Conclusion-CI failure as a result of anatomical abnormalities can be remedied by an ABI.
C1 Univ Verona, Policlin GB Rossi, ENT Dept, IT-37100 Verona, Italy.
C3 University of Verona; Azienda Ospedaliera Universitaria Integrata Verona
RP Colletti, V (通讯作者)，Univ Verona, Policlin GB Rossi, ENT Dept, Ple LA Scuro 10, IT-37100 Verona, Italy.
EM
OI COLLETTI, Liliana/0000-0001-8089-5681
CR BALKANY T, 1988, ANN OTOL RHINOL LA S, V135, P3
   Camilleri AE, 1999, J LARYNGOL OTOL, V113, P454, DOI 10.1017/S0022215100144202
   COLIGADO EJ, 1993, ARCH PHYS MED REHAB, V74, P653, DOI 10.1016/0003-9993(93)90166-8
   Colletti V, 2000, AM J OTOL, V21, P826
   Colletti V, 2002, OTOLARYNG HEAD NECK, V127, P84, DOI 10.1067/mhn.2002.126723
   GANTZ BJ, 1988, OTOLARYNG HEAD NECK, V98, P72, DOI 10.1177/019459988809800113
   Gibson WPR, 2000, ADV OTO-RHINO-LARYNG, V57, P250
   Gray RF, 1998, J LARYNGOL OTOL, V112, P646, DOI 10.1017/S0022215100141349
   HINOJOSA R, 1991, AM J OTOL, V12, P3
   Lenarz T, 2001, OTOL NEUROTOL, V22, P24, DOI 10.1097/00129492-200101000-00006
   Mawman DJ, 2000, ADV OTO-RHINO-LARYNG, V57, P364
   Maxwell AP, 1999, AM J OTOL, V20, P335
   Miyamoto RT, 2000, ADV OTO-RHINO-LARYNG, V57, P160
   Miyamoto RT, 1999, LARYNGOSCOPE, V109, P181, DOI 10.1097/00005537-199902000-00002
   Moore A, 1999, J LARYNGOL OTOL, V113, P34, DOI 10.1017/S0022215100146079
   NADOL JB, 1991, ANN OTO RHINOL LARYN, V100, P712, DOI 10.1177/000348949110000904
   Proops DW, 1999, J LARYNGOL OTOL, V113, P5, DOI 10.1017/S0022215100146018
   Rauch SD, 1997, LARYNGOSCOPE, V107, P1606, DOI 10.1097/00005537-199712000-00005
   Reed Charlotte M., 1995, Seminars in Hearing, V16, P305, DOI 10.1055/s-0028-1083728
   Richardson HC, 1999, J LARYNGOL OTOL, V113, P811, DOI 10.1017/S002221510014527X
   SHELTON C, 1989, OTOLARYNG HEAD NECK, V100, P227, DOI 10.1177/019459988910000310
   Starr A, 1996, BRAIN, V119, P741, DOI 10.1093/brain/119.3.741
   Toh EH, 2002, OTOLARYNG CLIN N AM, V35, P325, DOI 10.1016/S0030-6665(02)00016-6
   Valimaa TT, 2001, SCAND AUDIOL, V30, P112, DOI 10.1080/010503901300112220
   Wu H, 2000, ADV OTO-RHINO-LARYNG, V57, P382
NR 25
TC 22
Z9 26
U1 0
U2 8
PU TAYLOR & FRANCIS LTD
PI ABINGDON
PA 2-4 PARK SQUARE, MILTON PARK, ABINGDON OR14 4RN, OXON, ENGLAND
SN 0001-6489
EI 1651-2251
J9 ACTA OTO-LARYNGOL
JI Acta Oto-Laryngol.
PD MAY
PY 2004
VL 124
IS 4
BP 353
EP 357
DI 10.1080/00016480410016441
PG 5
WC Otorhinolaryngology
WE Science Citation Index Expanded (SCI-EXPANDED)
SC Otorhinolaryngology
GA 829GN
UT WOS:000222035500004
PM 15224852
DA 2023-08-27
ER

PT J
AU Porkholm, M
   Valanne, L
   Lonnqvist, T
   Holm, S
   Lannering, B
   Riikonen, P
   Wojcik, D
   Sehested, A
   Clausen, N
   Harila-Saari, A
   Schomerus, E
   Thorarinsdottir, HK
   Lahteenmaki, P
   Arola, M
   Thomassen, H
   Saarinen-Pihkala, UM
   Kivivuori, SM
AF Porkholm, Mikaela
   Valanne, Leena
   Lonnqvist, Tuula
   Holm, Stefan
   Lannering, Birgitta
   Riikonen, Pekka
   Wojcik, Dorota
   Sehested, Astrid
   Clausen, Niels
   Harila-Saari, Arja
   Schomerus, Eckhard
   Thorarinsdottir, Halldora K.
   Lahteenmaki, Paivi
   Arola, Mikko
   Thomassen, Harald
   Saarinen-Pihkala, Ulla M.
   Kivivuori, Sanna-Maria
TI Radiation Therapy and Concurrent Topotecan Followed by Maintenance
   Triple Anti-Angiogenic Therapy With Thalidomide, Etoposide, and
   Celecoxib for Pediatric Diffuse Intrinsic Pontine Glioma
SO PEDIATRIC BLOOD & CANCER
LA English
DT Article
DE anti-angiogenic therapy; celecoxib; etoposide; pediatric; pontine
   glioma; thalidomide
ID BRAIN-STEM GLIOMAS; PHASE-II; CHILDREN; RADIOSENSITIZER; RADIOTHERAPY;
   CHILDHOOD; CANCER
AB Background. Despite major treatment attempts, the prognosis for pediatric diffuse intrinsic pontine gliomas (DIPGs) remains dismal. Gliomas are highly vascularized tumors, suggesting that the prevention of vessel formation by anti-angiogenic treatment might be effective. Procedure. Forty-one pediatric patients with DIPG were treated according to the Angiocomb protocol, starting with radiotherapy combined with topotecan and followed by anti-angiogenic triple medication consisting of thalidomide, etoposide, and celecoxib. Overall survival, radiological response, quality of life, requirement of corticosteroids, and adverse effects were monitored. Eight patients treated with only radiotherapy were used as controls. Results. For study patients, the 12 and 24 months overall survival was 61% and 17%, respectively. The median overall survival was 12 months (range 4-60 months). Four radiological complete responses were seen, of which two were transient. Radiologically, 56% of the tumors reduced in size and 78% in signal intensity. Study patients were able to visit school or daycare and walk for a significantly longer time compared to controls (Log Rank 0.036 and 0.008, respectively). Adverse effects were generally minor. Conclusions. The Angiocomb protocol created a noticeable share of long-term survivors and was well tolerated, suggesting that anti-angiogenic therapy for patients with DIPG should be studied more in the future. (c) 2014 Wiley Periodicals, Inc.
C1 [Porkholm, Mikaela; Saarinen-Pihkala, Ulla M.; Kivivuori, Sanna-Maria] Univ Helsinki, Cent Hosp, Childrens Hosp, Div Hematol Oncol & Stem Cell Transplantat, FIN-00028 Helsinki, Finland.
   [Valanne, Leena] Univ Helsinki, Cent Hosp, Helsinki Med Imaging Ctr, FIN-00028 Helsinki, Finland.
   [Lonnqvist, Tuula] Univ Helsinki, Cent Hosp, Div Child Neurol, FIN-00028 Helsinki, Finland.
   [Holm, Stefan] Karolinska Univ Hosp, Astrid Lindgren Childrens Hosp, Dept Pediat Hematol Oncol, Stockholm, Sweden.
   [Lannering, Birgitta] Univ Gothenburg, Queen Silvia Childrens Hosp, Dept Pediat Hematol Oncol, Gothenburg, Sweden.
   [Riikonen, Pekka] Kuopio Univ Hosp, Dept Pediat, SF-70210 Kuopio, Finland.
   [Wojcik, Dorota] Haukeland Hosp, N-5021 Bergen, Norway.
   [Sehested, Astrid] Rigshosp, Dept Pediat Hematol Oncol, DK-2100 Copenhagen, Denmark.
   [Clausen, Niels] Aarhus Univ Hosp, Dept Pediat, DK-8000 Aarhus, Denmark.
   [Harila-Saari, Arja] Oulu Univ Hosp, Dept Pediat, Pediat Oncol Hematol Sect, Oulu, Finland.
   [Schomerus, Eckhard] Odense Univ Hosp, HC Andersen Childrens Hosp, Dept Pediat Hematol & Oncol, DK-5000 Odense, Denmark.
   [Thorarinsdottir, Halldora K.] Childrens Hosp, Reykjavik, Iceland.
   [Lahteenmaki, Paivi] Turku Univ, Cent Hosp, Dept Pediat, Turku, Finland.
   [Arola, Mikko] Tampere Univ, Cent Hosp, Dept Pediat, SF-33520 Tampere, Finland.
   [Thomassen, Harald] Univ Trondheim Hosp, Trondheim, Norway.
C3 University of Helsinki; Helsinki University Central Hospital; University
   of Helsinki; Helsinki University Central Hospital; University of
   Helsinki; Helsinki University Central Hospital; Karolinska Institutet;
   Karolinska University Hospital; Queen Silvia Children's Hospital;
   University of Gothenburg; Kuopio University Hospital; University of
   Eastern Finland; Rigshospitalet; University of Copenhagen; Aarhus
   University; University of Oulu; University of Southern Denmark; Odense
   University Hospital; University of Turku; Tampere University; Tampere
   University Hospital; Norwegian University of Science & Technology (NTNU)
RP Kivivuori, SM (通讯作者)，Univ Helsinki, Cent Hosp, Childrens Hosp, POB 281, FIN-00028 Helsinki, Finland.
EM
RI Harila-Saari, Arja/E-2124-2014
OI Harila-Saari, Arja/0000-0003-2767-5828; Lahteenmaki, Paivi
   Maria/0000-0002-5500-9606; Valanne, Leena/0000-0002-8572-829X
CR Bernier-Chastagner V, 2005, CANCER-AM CANCER SOC, V104, P2792, DOI 10.1002/cncr.21534
   Chen HJ, 2010, AM J NEURORADIOL, V31, P1879, DOI 10.3174/ajnr.A2179
   Cohen KJ, 2011, NEURO-ONCOLOGY, V13, P410, DOI 10.1093/neuonc/noq205
   Grimm SA, 2013, CURR NEUROL NEUROSCI, V13, DOI 10.1007/s11910-013-0346-3
   Hargrave D, 2008, J NEURO-ONCOL, V86, P313, DOI 10.1007/s11060-007-9473-5
   Heath JA, 2012, ASIA-PAC J CLIN ONCO, V8, P223, DOI 10.1111/j.1743-7563.2012.01558.x
   Jansen MHA, 2012, CANCER TREAT REV, V38, P27, DOI 10.1016/j.ctrv.2011.06.007
   Janssens GO, 2013, INT J RADIAT ONCOL, V85, P315, DOI 10.1016/j.ijrobp.2012.04.006
   Kebudi R, 2013, CHILD NERV SYST, V29, P583, DOI 10.1007/s00381-012-1986-3
   Khatua S, 2011, CHILD NERV SYST, V27, P1391, DOI 10.1007/s00381-011-1468-z
   Kim CY, 2010, J NEURO-ONCOL, V100, P193, DOI 10.1007/s11060-010-0157-1
   Kivivuori SM, 2011, ACTA PAEDIATR, V100, P134, DOI 10.1111/j.1651-2227.2010.01961.x
   Korones DN, 2008, PEDIATR BLOOD CANCER, V50, P227, DOI 10.1002/pbc.21154
   Lien K, 2013, EUR J CANCER, V49, P3387, DOI 10.1016/j.ejca.2013.06.038
   Pasquier E, 2011, TRANSL ONCOL, V4, P189, DOI 10.1593/tlo.11124
   Pollack IF, 2011, NEURO-ONCOLOGY, V13, P290, DOI 10.1093/neuonc/noq199
   Puget S, 2012, PLOS ONE, V7, DOI 10.1371/journal.pone.0030313
   Robison NJ, 2014, PEDIATR BLOOD CANCER, V61, P636, DOI 10.1002/pbc.24794
   Sanghavi SN, 2003, NEURO-ONCOLOGY, V5, P8, DOI 10.1093/neuonc/5.1.8
   Sirachainan N, 2008, NEURO-ONCOLOGY, V10, P577, DOI 10.1215/15228517-2008-025
   Takano Shingo, 2004, Brain Tumor Pathol, V21, P69, DOI 10.1007/BF02484513
   Veringa SJE, 2013, PLOS ONE, V8, DOI 10.1371/journal.pone.0061512
   Warren KE, 2012, FRONT ONCOL, V2, DOI 10.3389/fonc.2012.00205
NR 23
TC 20
Z9 20
U1 0
U2 5
PU WILEY
PI HOBOKEN
PA 111 RIVER ST, HOBOKEN 07030-5774, NJ USA
SN 1545-5009
EI 1545-5017
J9 PEDIATR BLOOD CANCER
JI Pediatr. Blood Cancer
PD SEP
PY 2014
VL 61
IS 9
BP 1603
EP 1609
DI 10.1002/pbc.25045
PG 7
WC Oncology; Hematology; Pediatrics
WE Science Citation Index Expanded (SCI-EXPANDED)
SC Oncology; Hematology; Pediatrics
GA AN4EO
UT WOS:000340540600015
PM 24692119
DA 2023-08-27
ER

PT J
AU Jansen, MHA
   Lagerweij, T
   Sewing, ACP
   Vugts, DJ
   van Vuurden, DG
   Molthoff, CFM
   Caretti, V
   Veringa, SJE
   Petersen, N
   Carcaboso, AM
   Noske, DP
   Vandertop, WP
   Wesseling, P
   van Dongen, GAMS
   Kaspers, GJL
   Hulleman, E
AF Jansen, Marc H. A.
   Lagerweij, Tonny
   Sewing, A. Charlotte P.
   Vugts, Danielle J.
   van Vuurden, Dannis G.
   Molthoff, Carla F. M.
   Caretti, Viola
   Veringa, Susanna J. E.
   Petersen, Naomi
   Carcaboso, Angel M.
   Noske, David P.
   Vandertop, W. Peter
   Wesseling, Pieter
   van Dongen, Guus A. M. S.
   Kaspers, Gertjan J. L.
   Hulleman, Esther
TI Bevacizumab Targeting Diffuse Intrinsic Pontine Glioma: Results of
   Zr-89-Bevacizumab PET Imaging in Brain Tumor Models
SO MOLECULAR CANCER THERAPEUTICS
LA English
DT Article
ID ENDOTHELIAL GROWTH-FACTOR; NEWLY-DIAGNOSED GLIOBLASTOMA; RECURRENT
   MALIGNANT GLIOMA; HIGH-GRADE GLIOMAS; ZR-89 IMMUNO-PET; VEGF-A;
   MONOCLONAL-ANTIBODIES; PLUS IRINOTECAN; CLINICAL-TRIALS; STEM GLIOMA
AB The role of the VEGF inhibitor bevacizumab in the treatment of diffuse intrinsic pontine glioma (DIPG) is unclear. We aim to study the biodistribution and uptake of zirconium-89 (Zr-89)labeled bevacizumab in DIPG mouse models. Human E98-FM, U251-FM glioma cells, and HSJD-DIPG-007-FLUC primary DIPG cells were injected into the subcutis, pons, or striatum of nude mice. Tumor growth was monitored by bioluminescence imaging (BLI) and visualized by MRI. Seventy-two to 96 hours after Zr-89-bevacizumab injections, mice were imaged by positron emission tomography (PET), and biodistribution was analyzed ex vivo. High VEGF expression in human DIPG was confirmed in a publically available mRNA database, but no significant (89)-Zrbevacizumab uptake could be detected in xenografts located in the pons and striatum at an early or late stage of the disease. E98-FM, and to a lesser extent the U251-FM and HSJD-DIPG-007 subcutaneous tumors, showed high accumulation of Zr-89-bevacizumab. VEGF expression could not be demonstrated in the intracranial tumors by in situ hybridization (ISH) but was clearly present in the perinecrotic regions of subcutaneous E98-FM tumors. The poor uptake of Zr-89-bevacizumab in xenografts located in the brain suggests that VEGF targeting with bevacizumab has limited efficacy for diffuse infiltrative parts of glial brain tumors in mice. Translating these results to the clinic would imply that treatment with bevacizumab in patients with DIPG is only justified after targeting of VEGF has been demonstrated by Zr-89-bevacizumab immuno-PET. We aim to confirm this observation in a clinical PET study with patients with DIPG. (C)2016 AACR.
C1 [Jansen, Marc H. A.; Sewing, A. Charlotte P.; van Vuurden, Dannis G.; Caretti, Viola; Veringa, Susanna J. E.; Kaspers, Gertjan J. L.; Hulleman, Esther] Ctr Canc, Dept Pediat Pediat Hematol & Oncol, Amsterdam, Netherlands.
   [Lagerweij, Tonny; Sewing, A. Charlotte P.; van Vuurden, Dannis G.; Caretti, Viola; Veringa, Susanna J. E.; Petersen, Naomi; Noske, David P.; Wesseling, Pieter; Hulleman, Esther] Ctr Canc, Neurooncol Res Grp, Amsterdam, Netherlands.
   [Lagerweij, Tonny; Noske, David P.; Vandertop, W. Peter] Vrije Univ Amsterdam, Med Ctr, Dept Neurosurg, Amsterdam, Netherlands.
   [Lagerweij, Tonny; Noske, David P.; Vandertop, W. Peter] Acad Med Ctr, Amsterdam, Netherlands.
   [Vugts, Danielle J.; Molthoff, Carla F. M.; van Dongen, Guus A. M. S.] Vrije Univ Amsterdam, Med Ctr, Dept Radiol & Nucl Med, Amsterdam, Netherlands.
   [Caretti, Viola] Stanford Univ, Dept Neurol, Sch Med, Stanford, CA 94305 USA.
   [Caretti, Viola] Stanford Univ, Dept Pediat, Sch Med, Stanford, CA 94305 USA.
   [Caretti, Viola] Stanford Univ, Dept Neurosurg, Sch Med, Stanford, CA 94305 USA.
   [Carcaboso, Angel M.] Hosp St Joan de Deu, Dept Oncol, Preclin Therapeut & Drug Delivery Res Program, Barcelona, Spain.
   [Wesseling, Pieter] Vrije Univ Amsterdam, Med Ctr, Dept Pathol, Amsterdam, Netherlands.
   [Wesseling, Pieter] Radboud Univ Nijmegen, Med Ctr, Dept Pathol, Nijmegen, Netherlands.
C3 Vrije Universiteit Amsterdam; University of Amsterdam; Academic Medical
   Center Amsterdam; Vrije Universiteit Amsterdam; Stanford University;
   Stanford University; Stanford University; University of Barcelona; Vrije
   Universiteit Amsterdam; Radboud University Nijmegen
RP Hulleman, E (通讯作者)，Vrije Univ Amsterdam, Med Ctr, Canc Ctr Amsterdam, De Boelelaan 1117, NL-1081 HV Amsterdam, Netherlands.
EM
RI Wesseling, P./H-8114-2014; Vandertop, William P./K-9288-2018; Kaspers,
   Gertjan/ABC-5334-2020; Lagerweij, Tonny/AAY-7128-2020; van Vuurden,
   Dannis/ACA-6952-2022; Hulleman, Esther/AAD-7073-2021
OI Wesseling, P./0000-0001-5453-5201; Kaspers, Gertjan/0000-0001-7716-8475;
   Vugts, Danielle/0000-0001-7068-8152; Noske, David/0000-0001-5992-4468;
   Hulleman, Esther/0000-0001-9256-947X; Vandertop,
   William/0000-0001-5417-0265; Carcaboso, Angel/0000-0002-8485-426X; Van
   Vuurden, Dannis/0000-0002-1364-9007
FU Semmy Foundation; Stichting Kika (Children-Cancer-free) [69];
   ISCIII-FEDER [CP13/00189]
FX E. Hulleman received funding from Semmy Foundation and Stichting Kika
   (Children-Cancer-free, project 69). M.H. Jansen received funding from
   Stichting Egbers. A.M. Carcaboso acknowledges funding from ISCIII-FEDER
   (CP13/00189).
CR Aguilera Dolly G, 2013, J Pediatr Hematol Oncol, V35, pe42, DOI 10.1097/MPH.0b013e318279aed8
   Bernsen HJJA, 1998, J NEURO-ONCOL, V38, P51, DOI 10.1023/A:1005957201431
   Boult JKR, 2015, CANCER RES, V75, DOI 10.1158/1538-7445.AM2015-3271
   Caretti V, 2011, BRAIN PATHOL, V21, P441, DOI 10.1111/j.1750-3639.2010.00468.x
   Chinot OL, 2014, NEW ENGL J MED, V370, P709, DOI 10.1056/NEJMoa1308345
   Claes A, 2008, BRAIN PATHOL, V18, P423, DOI 10.1111/j.1750-3639.2008.00141.x
   de Jong HWAM, 2007, PHYS MED BIOL, V52, P1505, DOI 10.1088/0031-9155/52/5/019
   de Weger RA, 2000, BRIT J HAEMATOL, V110, P647, DOI 10.1046/j.1365-2141.2000.02233.x
   Ferrara N, 2004, ENDOCR REV, V25, P581, DOI 10.1210/er.2003-0027
   Frankin K.B.J., 2008, MOUSE BRAIN STEREOTA, V3rd ed.
   Friedman HS, 2009, J CLIN ONCOL, V27, P4733, DOI 10.1200/JCO.2008.19.8721
   Gagner JP, 2005, BRAIN PATHOL, V15, P342
   Gilbert MR, 2014, NEW ENGL J MED, V370, P699, DOI 10.1056/NEJMoa1308573
   Gomez-Manzano C, 2008, NEURO-ONCOLOGY, V10, P940, DOI 10.1215/15228517-2008-061
   Grasso CS, 2015, NAT MED, V21, P555, DOI 10.1038/nm.3855
   Gururangan S, 2010, J CLIN ONCOL, V28, P3069, DOI 10.1200/JCO.2009.26.8789
   Hargrave D, 2008, J NEURO-ONCOL, V86, P313, DOI 10.1007/s11060-007-9473-5
   Hargrove D, 2006, LANCET ONCOL, V7, P241, DOI 10.1016/S1470-2045(06)70615-5
   Hummel TR, 2016, J NEURO-ONCOL, V127, P53, DOI 10.1007/s11060-015-2008-6
   Jacobs VL, 2011, ASN NEURO, V3, DOI 10.1042/AN20110014
   Jansen MHA, 2012, CANCER TREAT REV, V38, P27, DOI 10.1016/j.ctrv.2011.06.007
   Jansen MH, 2015, NEURO-ONCOLOGY, V17, P160, DOI 10.1093/neuonc/nou104
   Jansen MHA, 2012, EXPERT REV ANTICANC, V12, P1109, DOI [10.1586/ERA.12.95, 10.1586/era.12.95]
   Johansson M, 2002, J NEURO-ONCOL, V59, P1, DOI 10.1023/A:1016317407043
   Jubb AM, 2010, LANCET ONCOL, V11, P1172, DOI 10.1016/S1470-2045(10)70232-1
   Jubb AM, 2006, J CLIN ONCOL, V24, P217, DOI 10.1200/JCO.2005.01.5388
   Khasraw M, 2014, COCHRANE DB SYST REV, DOI 10.1002/14651858.CD008218.pub3
   Loening Andreas Markus, 2003, Mol Imaging, V2, P131, DOI 10.1162/153535003322556877
   Mir SE, 2010, CANCER CELL, V18, P244, DOI 10.1016/j.ccr.2010.08.011
   Narayana A, 2010, NEURO-ONCOLOGY, V12, P985, DOI 10.1093/neuonc/noq033
   Navis AC, 2011, J PATHOL, V223, P626, DOI 10.1002/path.2836
   Paez-Ribes M, 2009, CANCER CELL, V15, P220, DOI 10.1016/j.ccr.2009.01.027
   Papadopoulos N, 2012, ANGIOGENESIS, V15, P171, DOI 10.1007/s10456-011-9249-6
   Parekh C, 2011, J NEURO-ONCOL, V103, P673, DOI 10.1007/s11060-010-0444-x
   Paugh BS, 2011, J CLIN ONCOL, V29, P3999, DOI 10.1200/JCO.2011.35.5677
   Paugh BS, 2010, J CLIN ONCOL, V28, P3061, DOI 10.1200/JCO.2009.26.7252
   Roodink I, 2006, INT J CANCER, V119, P2054, DOI 10.1002/ijc.22072
   Salloum R, 2015, J NEURO-ONCOL, V121, P591, DOI 10.1007/s11060-014-1671-3
   Shibuya M, 2006, J BIOCHEM MOL BIOL, V39, P469
   Shih T, 2006, CLIN THER, V28, P1779, DOI 10.1016/j.clinthera.2006.11.015
   Taylor KR, 2014, NAT GENET, V46, P457, DOI 10.1038/ng.2925
   Verel I, 2003, J NUCL MED, V44, P1663
   Verel I, 2003, J NUCL MED, V44, P1271
   Vredenburgh JJ, 2007, J CLIN ONCOL, V25, P4722, DOI 10.1200/JCO.2007.12.2440
   Vredenburgh JJ, 2007, CLIN CANCER RES, V13, P1253, DOI 10.1158/1078-0432.CCR-06-2309
   Wesseling P, 1997, J NEURO-ONCOL, V32, P253, DOI 10.1023/A:1005746320099
   Yu LL, 2008, INVEST OPHTH VIS SCI, V49, P522, DOI 10.1167/iovs.07-1175
   Zaky W, 2013, PEDIATR HEMAT ONCOL, V30, P623, DOI 10.3109/08880018.2013.829895
   Zhao LN, 2011, J MOL NEUROSCI, V44, P122, DOI 10.1007/s12031-010-9487-x
NR 49
TC 45
Z9 46
U1 1
U2 18
PU AMER ASSOC CANCER RESEARCH
PI PHILADELPHIA
PA 615 CHESTNUT ST, 17TH FLOOR, PHILADELPHIA, PA 19106-4404 USA
SN 1535-7163
EI 1538-8514
J9 MOL CANCER THER
JI Mol. Cancer Ther.
PD SEP 1
PY 2016
VL 15
IS 9
BP 2166
EP 2174
DI 10.1158/1535-7163.MCT-15-0558
PG 9
WC Oncology
WE Science Citation Index Expanded (SCI-EXPANDED)
SC Oncology
GA DV6MQ
UT WOS:000383050800015
PM 27325687
OA Green Submitted
DA 2023-08-27
ER

PT J
AU Zhang, JQ
   Liu, Q
   Yuan, ZY
   Zhao, LJ
   Wang, XG
   Wang, P
AF Zhang, Jiaqi
   Liu, Qun
   Yuan, Zhiyong
   Zhao, Lujun
   Wang, Xiaoguang
   Wang, Ping
TI Clinical Efficacy of CyberKnife Radiosurgery for Adult Brainstem Glioma:
   10 Years Experience at Tianjin CyberKnife Center and Review of the
   Literature
SO FRONTIERS IN ONCOLOGY
LA English
DT Review
DE brainstem gliomas; stereotactic body radiotherapy; CyberKnife;
   radiosurgery; prognostic factor
ID GAMMA-KNIFE RADIOSURGERY; MANAGEMENT; DIAGNOSIS; PSEUDOPROGRESSION;
   CHILDHOOD
AB Background: Brainstemglioma is a rare brain tumor with poor prognosis and difficulty for surgical resection. We sought to retrospectively analyze and evaluate the clinical efficacy of CyberKnife for brainstem gliomas.
   Methods: From 2006 to 2015, a total of 21 brainstem gliomas patients who received CyberKnife radiosurgery treatment enrolled in this study and 18 patients with follow up. CyberKnife image-guided radiosurgical system were applied consecutively with the median prescribed total dose of 26Gy (14-33Gy) at two to six fractions on days utilizing CyberKnife system, and the median biological equivalent doses of 59.8Gy (33.6-76.56Gy). The clinic pathlogical features, survival were analyzed to explore the efficacy of CyberKnife radiosurgery in treatment of brainstem glioma.
   Results: With median follow-up of 54.5 months, patients with brainstem gliomas had median overall survival of 19 months, five patients still alive. The primary endpoints of the 1- and 2-year overall survival rates were 87.5 and 52.4%, respectively. During the treatment course, six patients were observed to have pseudoprogression with mass effect onMRI. Four patients developed radiation complications. Grade 2 radiation-related toxicity were observed in three patients and one patient with grade 3.
   Conclusion: The efficacy of brainstem gliomas-treated with CyberKnife is efficacious with mild toxicity.
C1 [Zhang, Jiaqi; Liu, Qun; Yuan, Zhiyong; Zhao, Lujun; Wang, Xiaoguang; Wang, Ping] Tianjin Med Univ, Dept Radiotherapy, Natl Clin Res Ctr Canc, Canc Inst & Hosp, Tianjin, Peoples R China.
C3 Tianjin Medical University
RP Wang, P (通讯作者)，Tianjin Med Univ, Dept Radiotherapy, Natl Clin Res Ctr Canc, Canc Inst & Hosp, Tianjin, Peoples R China.
EM
CR ALBRIGHT AL, 1993, NEUROSURGERY, V33, P1026
   Babu R, 2014, J NEURO-ONCOL, V119, P177, DOI 10.1007/s11060-014-1471-9
   Boothe D, 2013, NEURO-ONCOLOGY, V15, P1257, DOI 10.1093/neuonc/not085
   Brandsma D, 2008, LANCET ONCOL, V9, P453, DOI 10.1016/S1470-2045(08)70125-6
   Chamberlain MC, 2008, J CLIN ONCOL, V26, P4359, DOI 10.1200/JCO.2008.18.4440
   El-Shehaby AMN, 2015, ACTA NEUROCHIR, V157, P247, DOI 10.1007/s00701-014-2299-y
   EPSTEIN F, 1986, J NEUROSURG, V64, P11, DOI 10.3171/jns.1986.64.1.0011
   Fuchs I, 2002, ACT NEUR S, V84, P85
   Guillamo JS, 2001, BRAIN, V124, P2528, DOI 10.1093/brain/124.12.2528
   Hong Semie, 2005, Cancer Res Treat, V37, P109, DOI 10.4143/crt.2005.37.2.109
   Jansen MHA, 2012, CANCER TREAT REV, V38, P27, DOI 10.1016/j.ctrv.2011.06.007
   Jung EW, 2013, CLIN NEUROL NEUROSUR, V115, P2023, DOI 10.1016/j.clineuro.2013.06.012
   Lin CS, 2012, NEUROSURGERY, V70, P953, DOI 10.1227/NEU.0b013e31823c40fe
   Liu SH, 2016, J CLIN NEUROSCI, V25, P105, DOI 10.1016/j.jocn.2015.10.013
   Lopez WOC, 2013, CLIN NEUROL NEUROSUR, V115, P1451, DOI 10.1016/j.clineuro.2013.01.012
   Miltenburg D, 1996, CAN J NEUROL SCI, V23, P118, DOI 10.1017/S031716710003883X
   Purohit B, 2015, EUR J RADIOL, V84, P709, DOI 10.1016/j.ejrad.2014.12.025
   Ramos A, 2013, SEMIN ULTRASOUND CT, V34, P104, DOI 10.1053/j.sult.2013.01.001
   Reyes-Botero G, 2012, ONCOLOGIST, V17, P388, DOI 10.1634/theoncologist.2011-0335
   Sabbagh AJ, 2015, NEUROSCIENCES, V20, P98, DOI 10.17712/nsj.2015.2.20140621
   Salmaggi A, 2008, J NEUROL, V255, P171, DOI 10.1007/s00415-008-0589-0
   Sharma MS, 2008, NEUROSURGERY, V63, P728, DOI 10.1227/01.NEU.0000325726.72815.22
   SHRIEVE DC, 1992, INT J RADIAT ONCOL, V24, P599, DOI 10.1016/0360-3016(92)90704-L
   STROINK AR, 1986, J NEUROSURG, V65, P745, DOI 10.3171/jns.1986.65.6.0745
   Taal W, 2008, CANCER, V113, P405, DOI 10.1002/cncr.23562
   Theeler BJ, 2015, J NEUROL SCI, V353, P92, DOI 10.1016/j.jns.2015.04.014
   Timmerman RD, 2008, SEMIN RADIAT ONCOL, V18, P215, DOI 10.1016/j.semradonc.2008.04.001
   Valery CA, 2011, INT J RADIAT ONCOL, V80, P362, DOI 10.1016/j.ijrobp.2010.02.028
   Yen CP, 2007, J NEUROSURG, V106, P8, DOI 10.3171/jns.2007.106.1.8
   Yen CP, 2006, J NEUROSURG, V105, P213, DOI 10.3171/jns.2006.105.2.213
   Yin LX, 2013, CAN J NEUROL SCI, V40, P348, DOI 10.1017/S0317167100014293
   Yoshikawa K, 2006, MINIM INVAS NEUROSUR, V49, P110, DOI 10.1055/s-2006-932183
NR 32
TC 2
Z9 3
U1 1
U2 6
PU FRONTIERS MEDIA SA
PI LAUSANNE
PA AVENUE DU TRIBUNAL FEDERAL 34, LAUSANNE, CH-1015, SWITZERLAND
SN 2234-943X
J9 FRONT ONCOL
JI Front. Oncol.
PD APR 12
PY 2019
VL 9
AR 257
DI 10.3389/fonc.2019.00257
PG 7
WC Oncology
WE Science Citation Index Expanded (SCI-EXPANDED)
SC Oncology
GA HT2DM
UT WOS:000464373400001
PM 31032223
OA Green Published, gold
DA 2023-08-27
ER

PT J
AU Sufianov, R
   Pitskhelauri, D
   Bykanov, A
AF Sufianov, Rinat
   Pitskhelauri, David
   Bykanov, Andrey
TI Fourth Ventricle Tumors: A Review of Series Treated With Microsurgical
   Technique
SO FRONTIERS IN SURGERY
LA English
DT Review
DE IV ventricle; tumors of the IV ventricle; median suboccipital approach;
   minimally invasive suboccipital approach; posterior fossa; keyhole
   approach; brainstem
ID TELOVELAR APPROACH; CEREBELLOMEDULLARY FISSURE; TRANSCEREBELLOMEDULLARY
   FISSURE; FORAMEN; ANATOMY; LESIONS
AB Tumors of the IV ventricle represent 1-5% of all intracranial lesions; they are implicated in 2/3 of the tumors of the ventricular system. According to modern standards, the first treatment stage for this pathology is microsurgical removal. Currently, for the removal of neoplasms of the IV ventricle and brainstem, the median suboccipital approach is widely used, followed by one of the microapproaches. Moreover, with the development of microsurgical techniques, keyhole approaches are now beginning to be utilized. However, surgical treatment of these tumors remains a challenge for neurosurgeons due to the proximity of functionally important anatomical structures (the brainstem, the cerebellum, pathways, vessels, etc.) of the posterior cranial fossa. Therefore, surgery in this area is associated with the possible occurrence of a wide range of postoperative complications. The authors provide a review of series of fourth ventricle tumors treated with microsurgical technique.
C1 [Sufianov, Rinat; Pitskhelauri, David; Bykanov, Andrey] Minist Hlth Russian Federat, NN Burdenko Natl Med Res Ctr Neurosurg, Dept Neurooncol, Moscow, Russia.
   [Sufianov, Rinat] Sechenov Univ, Sechenov Moscow State Med Univ 1, Dept Neurosurg, Moscow, Russia.
C3 Ministry of Health of the Russian Federation; Russian Academy of Medical
   Sciences; N.N.Burdenko National Medical Research Center of Neurosurgery;
   Sechenov First Moscow State Medical University
RP Sufianov, R (通讯作者)，Minist Hlth Russian Federat, NN Burdenko Natl Med Res Ctr Neurosurg, Dept Neurooncol, Moscow, Russia.; Sufianov, R (通讯作者)，Sechenov Univ, Sechenov Moscow State Med Univ 1, Dept Neurosurg, Moscow, Russia.
EM
RI Bykanov, Andrey Egorovich/R-1370-2017
OI Bykanov, Andrey Egorovich/0000-0002-0588-4779
CR [Anonymous], 2008, KEYHOLE APPROACHES N
   [Anonymous], 1996, MICRONEUROSURGERY 4
   [Anonymous], 1945, SURG BRAIN
   Aydin I, 2018, WORLD NEUROSURG, V114, pE1107, DOI 10.1016/j.wneu.2018.03.157
   [Бабичев К.Н. Babichev K.N.], 2018, [Нейрохирургия, Neirokhirurgiya], V20, P10, DOI 10.17650/1683-3295-2018-20-4-10-19
   Bergsneider M, 1999, J NEUROSURG, V91, P340, DOI 10.3171/jns.1999.91.2.0340
   Cohen AR., 1996, SURG DISORDERS 4 VEN, P94
   Corniola MV, 2022, ACTA NEUROCHIR, V164, P767, DOI 10.1007/s00701-020-04601-5
   El-Bahy K, 2005, ACTA NEUROCHIR, V147, P137, DOI 10.1007/s00701-004-0407-0
   Ferguson SD, 2018, J NEUROSURG, V128, P339, DOI 10.3171/2016.11.JNS161167
   Gallo P, 2012, NEUROL INDIA, V60, P271, DOI 10.4103/0028-3886.98507
   Gnanalingham KK, 2002, J NEUROSURG, V97, P821, DOI 10.3171/jns.2002.97.4.0821
   Gok A, 2004, NEUROSURG REV, V27, P50, DOI 10.1007/s10143-003-0286-5
   Hamada H, 1999, NEUROL MED-CHIR, V39, P950, DOI 10.2176/nmc.39.950
   Han S, 2013, ACTA NEUROCHIR, V155, P1011, DOI 10.1007/s00701-013-1689-x
   Helmut Bertalanffy OB., 2011, YOUMANS NEUROLOGICAL, V6th
   Jamshidi AO, 2019, OPER NEUROSURG, V16, P167, DOI 10.1093/ons/opy175
   Jean WC, 2003, NEUROSURGERY, V52, P860, DOI 10.1227/01.NEU.0000053146.83780.74
   [Калиновский А.В. Kalinovskiy A.V.], 2018, [Нейрохирургия, Neirokhirurgiya], V20, P8, DOI 10.17650/1683-3295-2018-20-2-8-16
   Karakhan VB., 2015, HEAD NECK TUMORS, DOI [10.17650/2222-1468-2012-0-4-10-18, DOI 10.17650/2222-1468-2012-0-4-10-18]
   Kellogg JX, 1997, PEDIATR NEUROSURG, V27, P28, DOI 10.1159/000121221
   Kushel Y, 2019, WORLD NEUROSURG, V129, pE171, DOI 10.1016/j.wneu.2019.05.091
   Lawton MT, 2006, NEUROSURGERY, V59, P244, DOI 10.1227/01.NEU.0000232767.16809.68
   MATSUSHIMA T, 1992, NEUROSURGERY, V30, P325, DOI 10.1227/00006123-199203000-00003
   Matsushima T, 2001, J NEUROSURG, V94, P257, DOI 10.3171/jns.2001.94.2.0257
   Matsushima T, 2014, WORLD NEUROSURG, V82, pE615, DOI 10.1016/j.wneu.2014.04.064
   Meybodi AT, 2017, J NEUROSURG, V127, P768, DOI 10.3171/2016.8.JNS16855
   Mussi ACM, 2000, J NEUROSURG, V92, P812, DOI 10.3171/jns.2000.92.5.0812
   Ostrom QT, 2021, NEURO-ONCOLOGY, V23, P1, DOI 10.1093/neuonc/noab200
   Pitskhelauri D, 2019, WORLD NEUROSURG, V126, pE1257, DOI 10.1016/j.wneu.2019.02.239
   Pitskhelauri D, 2018, ACTA NEUROCHIR, V160, P1079, DOI 10.1007/s00701-018-3507-y
   Prell J, 2011, ACTA NEUROCHIR, V153, P541, DOI 10.1007/s00701-010-0924-y
   Qiu B, 2016, ONCOL LETT, V11, P4099, DOI 10.3892/ol.2016.4507
   Rajesh BJ, 2007, CHILD NERV SYST, V23, P555, DOI 10.1007/s00381-006-0295-0
   Raouf A, 2013, ACTA NEUROCHIR, V155, P1957, DOI 10.1007/s00701-013-1843-5
   Rappoport MY., 1935, KLINIKA KLINIKO ANAT, P232
   Razdolsky IY., 1940, OPUKHOLI 4 ZHELUDOCH, P5
   REKATE HL, 1985, ARCH NEUROL-CHICAGO, V42, P697, DOI 10.1001/archneur.1985.04060070091023
   Rhoton AL, 2000, NEUROSURGERY, V47, pS7, DOI 10.1097/00006123-200009001-00007
   Tanriover N, 2004, J NEUROSURG, V101, P484, DOI 10.3171/jns.2004.101.3.0484
   Teo C, 2015, PRINCIPLES PRACTICE
   Tomasello F, 2015, WORLD NEUROSURG, V83, P1141, DOI 10.1016/j.wneu.2015.01.039
   Toyota S, 2004, MINIM INVAS NEUROSUR, V47, P312, DOI 10.1055/s-2004-830070
   Tubbs RS, 2013, J NEUROSURG, V118, P315, DOI 10.3171/2012.10.JNS12794
   Yasargil MG., 1994, MICRONEUROSURGERY 4, V1st
   Zaheer SN, 2010, PEDIATR NEUROSURG, V46, P340, DOI 10.1159/000321539
   Ziyal IM, 1999, BRIT J NEUROSURG, V13, P276
NR 47
TC 1
Z9 1
U1 0
U2 3
PU FRONTIERS MEDIA SA
PI LAUSANNE
PA AVENUE DU TRIBUNAL FEDERAL 34, LAUSANNE, CH-1015, SWITZERLAND
SN 2296-875X
J9 FRONT SURG
JI Front. Surg.
PD JUN 6
PY 2022
VL 9
AR 915253
DI 10.3389/fsurg.2022.915253
PG 7
WC Surgery
WE Science Citation Index Expanded (SCI-EXPANDED)
SC Surgery
GA 2F6GZ
UT WOS:000813007200001
PM 35733438
OA gold, Green Published
DA 2023-08-27
ER

PT J
AU Janssens, GORJ
   Gidding, CEM
   Van Lindert, EJ
   Oldenburger, FR
   Erasmus, CE
   Schouten-Meeteren, AYN
   Kaanders, JHAM
AF Janssens, Geert O. R. J.
   Gidding, Corrie E. M.
   Van Lindert, Erik J.
   Oldenburger, Foppe R.
   Erasmus, Corrie E.
   Schouten-Meeteren, Antoinette Y. N.
   Kaanders, Johannes H. A. M.
TI THE ROLE OF HYPOFRACTIONATION RADIOTHERAPY FOR DIFFUSE INTRINSIC
   BRAINSTEM GLIOMA IN CHILDREN: A PILOT STUDY
SO INTERNATIONAL JOURNAL OF RADIATION ONCOLOGY BIOLOGY PHYSICS
LA English
DT Article
DE Hypofractionation; Radiotherapy; Brainstem Glioma
ID PEDIATRIC-ONCOLOGY-GROUP; HYPERFRACTIONATED RADIOTHERAPY;
   GLIOBLASTOMA-MULTIFORME; RADIATION-THERAPY; PHASE-II; TUMORS;
   CHEMOTHERAPY; MANAGEMENT; TRIAL
AB Purpose: Most children with a diffuse intrinsic brainstem glioma will die within 1 year after diagnosis. To reduce patient burden, we investigated the feasibility of a radical hypofractionation radiotherapy schedule, given over 3 weeks, as an alternative to the standard regimen (30 fractions over 6 weeks).
   Methods and Materials: Nine children, ages 3-13, were treated by 13 fractions of 3 Gy (n = 8) or 6 fractions of 5.5 Gy (n = 1) given over 3 weeks. All patients had symptoms for <= 3 months and <= 2 signs of the neurologic triad (long tract signs, ataxia, cranial nerve deficit). Bilateral involvement of the pons (n = 8), encasement of the basilar artery (n = 7) and extension into the cerebellar peduncle (n = 6) was visible on magnetic resonance imaging.
   Results: Symptom improvement occurred in all patients within 2 weeks after start of radiotherapy. At a mean follow-up time of 15 months, 7 patients have died. Median time to progression and overall survival was 4.9 and 8.6 months, respectively. Median time to death after progression was 3.6 months. No Grade 3 or 4 toxicity was observed. In a recently published review of clinical trials, median time to progression, overall survival, and time between progression and death ranged from 5.0-8.8, 7.0-16, and 1.0-4.5 months, respectively, with more aggressive regimens.
   Conclusion: This radical hypofractionation radiotherapy regimen for children with diffuse intrinsic brainstem glioma is feasible and associated with no Grade 3 or 4 toxicities. With a minimal overall treatment time, it offers quick symptom relief and outcome results within the range of published data. (c) 2009 Elsevier Inc.
C1 [Janssens, Geert O. R. J.; Kaanders, Johannes H. A. M.] Radboud Univ Nijmegen, Med Ctr, Dept Radiat Oncol, NL-6500 HB Nijmegen, Netherlands.
   [Gidding, Corrie E. M.] Radboud Univ Nijmegen, Med Ctr, Dept Pediat Oncol, NL-6500 HB Nijmegen, Netherlands.
   [Van Lindert, Erik J.] Radboud Univ Nijmegen, Med Ctr, Dept Neurosurg, NL-6500 HB Nijmegen, Netherlands.
   [Erasmus, Corrie E.] Radboud Univ Nijmegen, Med Ctr, Dept Pediat Neurol, NL-6500 HB Nijmegen, Netherlands.
   [Oldenburger, Foppe R.] Univ Amsterdam, Acad Med Ctr, Dept Radiat Oncol, NL-1105 AZ Amsterdam, Netherlands.
   [Schouten-Meeteren, Antoinette Y. N.] Univ Amsterdam, Acad Med Ctr, Dept Pediat Oncol, NL-1105 AZ Amsterdam, Netherlands.
C3 Radboud University Nijmegen; Radboud University Nijmegen; Radboud
   University Nijmegen; Radboud University Nijmegen; University of
   Amsterdam; Academic Medical Center Amsterdam; University of Amsterdam;
   Academic Medical Center Amsterdam
RP Janssens, GORJ (通讯作者)，Radboud Univ Nijmegen, Med Ctr, Dept Radiat Oncol, Post Box 9101, NL-6500 HB Nijmegen, Netherlands.
EM
RI Erasmus, C.E./L-4295-2015; Janssens, G.O.R.J./L-4412-2015; Kaanders,
   Hans/A-7432-2014; Gidding, C.E.M./L-4323-2015; van Lindert,
   E.J./L-4491-2015
OI Erasmus, C.E./0000-0002-6657-941X; Kaanders, Hans/0000-0001-5374-9558; 
CR Bernier-Chastagner V, 2005, CANCER-AM CANCER SOC, V104, P2792, DOI 10.1002/cncr.21534
   Bouffet E, 2000, CANCER, V88, P685, DOI 10.1002/(SICI)1097-0142(20000201)88:3<685::AID-CNCR27>3.0.CO;2-K
   Burger PC, 1998, AM J SURG PATHOL, V22, P1083, DOI 10.1097/00000478-199809000-00007
   Donaldson SS, 2006, J CLIN ONCOL, V24, P1266, DOI 10.1200/JCO.2005.04.6599
   Freeman CR, 1996, CANCER-AM CANCER SOC, V77, P555
   Freeman CR, 2000, INT J RADIAT ONCOL, V47, P561, DOI 10.1016/S0360-3016(00)00471-5
   FREEMAN CR, 1993, INT J RADIAT ONCOL, V27, P197, DOI 10.1016/0360-3016(93)90228-N
   Haas-Kogan DA, 1999, INT J RADIAT ONCOL, V43, P399, DOI 10.1016/S0360-3016(98)00393-9
   Hargrove D, 2006, LANCET ONCOL, V7, P241, DOI 10.1016/S1470-2045(06)70615-5
   Hulshof MCCM, 2000, RADIOTHER ONCOL, V54, P143, DOI 10.1016/S0167-8140(99)00183-8
   Korones DN, 2007, EXPERT REV ANTICANC, V7, P663, DOI 10.1586/14737140.7.5.663
   KRETSCHMAR CS, 1993, CANCER-AM CANCER SOC, V72, P1404, DOI 10.1002/1097-0142(19930815)72:4<1404::AID-CNCR2820720441>3.0.CO;2-G
   LANGMOEN IA, 1991, CHILD NERV SYST, V7, P13, DOI 10.1007/BF00263825
   Lewis J, 1997, INT J RADIAT ONCOL, V38, P925, DOI 10.1016/S0360-3016(97)00134-X
   Mandell LR, 1999, INT J RADIAT ONCOL, V43, P959, DOI 10.1016/S0360-3016(98)00501-X
   PACKER RJ, 1994, CANCER-AM CANCER SOC, V74, P1827, DOI 10.1002/1097-0142(19940915)74:6<1827::AID-CNCR2820740628>3.0.CO;2-Q
   Slotman BJ, 1996, INT J RADIAT ONCOL, V34, P895, DOI 10.1016/0360-3016(95)02038-1
   Smith MA, 1998, J NATL CANCER I, V90, P1269, DOI 10.1093/jnci/90.17.1269
   Zagzag D, 2000, PEDIATRICS, V106, P1045, DOI 10.1542/peds.106.5.1045
   Zimmerman RA, 1996, PEDIATR NEUROSURG, V25, P45, DOI 10.1159/000121096
NR 20
TC 36
Z9 41
U1 0
U2 3
PU ELSEVIER SCIENCE INC
PI NEW YORK
PA 360 PARK AVE SOUTH, NEW YORK, NY 10010-1710 USA
SN 0360-3016
J9 INT J RADIAT ONCOL
JI Int. J. Radiat. Oncol. Biol. Phys.
PD MAR 1
PY 2009
VL 73
IS 3
BP 722
EP 726
DI 10.1016/j.ijrobp.2008.05.030
PG 5
WC Oncology; Radiology, Nuclear Medicine & Medical Imaging
WE Science Citation Index Expanded (SCI-EXPANDED)
SC Oncology; Radiology, Nuclear Medicine & Medical Imaging
GA 408NE
UT WOS:000263440900014
PM 18990510
DA 2023-08-27
ER

PT J
AU Kalamarides, M
   Grayeli, AB
   Bouccara, D
   Ambert, E
   Sollmann, WP
   Sterkers, O
   Rey, A
AF Kalamarides, M
   Grayeli, AB
   Bouccara, D
   Ambert, E
   Sollmann, WP
   Sterkers, O
   Rey, A
TI Hearing restoration with auditory brainstem implants after radiosurgery
   for neurofibromatosis Type 2 - Report of three cases
SO JOURNAL OF NEUROSURGERY
LA English
DT Article
DE acoustic neuroma; neurofibromatosis Type 2; auditory brainstem implant;
   vestibular schwannoma; radiosurgery
ID VESTIBULAR SCHWANNOMA MANAGEMENT; COCHLEAR NUCLEAR-COMPLEX; ACOUSTIC
   NEUROMAS; STEREOTACTIC RADIOSURGERY; STEM IMPLANT; PERFORMANCE; SURGERY;
   ISSUES
AB The auditory brainstem implant (ABI) is designed to restore useful auditory sensations in patients, with neurofibromatosis Type 2 (NF2). The implantation is usually performed at the time of tumor removal in patients who do not undergo radiation treatment. The authors evaluated the performance of ABIs in three patients with NF2 in whom vestibular schwannoma continued to grow after radiation treatment. These three patients with NF2 received a 21-channel ABI; a translabyrinthine approach was used for both the tumor removal and the ABI placement. The interval between radiosurgery and the tumor removal plus device implantation ranged from 2 to 11 years. In all cases, the tumor was growing and the patients presented with total deafness. The mean number of active electrodes in these three patients was equivalent to the average results reported in other patients who received ABIs. The patients in this study used the ABI regularly for everyday life and obtained useful levels of environmental sound recognition. It is concluded that hearing function can be rehabilitated using ABIs in patients with NF2, even if radiosurgery fails to control the tumor growth.
C1 Univ Paris 07, Hop Beaujon, Fac Xavier Bichat, Serv Neurochirurg,Dept Neurosurg, F-92100 Clichy, France.
   Univ Paris 07, Hop Beaujon, Fac Xavier Bichat, Dept Otolaryngol, F-92100 Clichy, France.
   Stadt Klinikum Braunschweig, Neurochirurg Klin, Braunschweig, Germany.
C3 UDICE-French Research Universities; Universite Paris Cite; Assistance
   Publique Hopitaux Paris (APHP); Hopital Universitaire Beaujon - APHP;
   UDICE-French Research Universities; Universite Paris Cite; Assistance
   Publique Hopitaux Paris (APHP); Hopital Universitaire Beaujon - APHP
RP Kalamarides, M (通讯作者)，Univ Paris 07, Hop Beaujon, Fac Xavier Bichat, Serv Neurochirurg,Dept Neurosurg, 100 Blvd Gen Leclerc, F-92100 Clichy, France.
CR Battista RA, 2000, AM J OTOL, V21, P371, DOI 10.1016/S0196-0709(00)80047-2
   BRACKMANN DE, 1993, OTOLARYNG HEAD NECK, V108, P624, DOI 10.1177/019459989310800602
   Chang SD, 1998, J NEUROSURG, V89, P321, DOI 10.3171/jns.1998.89.2.0321
   EDGERTON BJ, 1982, ANN OTO RHINOL LARYN, V91, P117
   HITSELBERGER WE, 1984, OTOLARYNG HEAD NECK, V92, P52, DOI 10.1177/019459988409200111
   HOUSE JW, 1985, OTOLARYNG HEAD NECK, V93, P146, DOI 10.1177/019459988509300202
   Ito K, 2000, INT J RADIAT ONCOL, V48, P75, DOI 10.1016/S0360-3016(00)00570-8
   Kida Y, 2000, SURG NEUROL, V53, P383, DOI 10.1016/S0090-3019(00)00174-9
   KUROKI A, 1995, J NEUROSURG, V82, P933, DOI 10.3171/jns.1995.82.6.0933
   Laszig R, 1995, Ann Otol Rhinol Laryngol Suppl, V166, P28
   Laszig R, 1997, AM J OTOL, V18, pS160
   Mautner VF, 1996, NEUROSURGERY, V38, P880, DOI 10.1097/00006123-199605000-00004
   MCELVEEN JT, 1987, OTOLARYNG HEAD NECK, V96, P135, DOI 10.1177/019459988709600204
   MONSELL EM, 1987, AM J OTOL, V8, P450
   Otto SR, 1998, OTOLARYNG HEAD NECK, V118, P291, DOI 10.1016/S0194-5998(98)70304-3
   Pollock BE, 1998, J NEUROSURG, V89, P949, DOI 10.3171/jns.1998.89.6.0949
   Pollock BE, 1998, NEUROSURGERY, V43, P475, DOI 10.1097/00006123-199809000-00041
   Prasad D, 2000, J NEUROSURG, V92, P745, DOI 10.3171/jns.2000.92.5.0745
   Samii M, 1997, NEUROSURGERY, V40, P696, DOI 10.1097/00006123-199704000-00007
   SHANNON RV, 1993, OTOLARYNG HEAD NECK, V108, P634, DOI 10.1177/019459989310800603
   SLATTERY WH, 1995, AM J OTOL, V16, P315
   Slattery WH, 1998, AM J OTOL, V19, P638
   Sterkers O, 1997, Ann Otolaryngol Chir Cervicofac, V114, P176
   Subach BR, 1999, J NEUROSURG, V90, P815, DOI 10.3171/jns.1999.90.5.0815
   Suh JH, 2000, INT J CANCER, V90, P145, DOI 10.1002/1097-0215(20000620)90:3<145::AID-IJC4>3.0.CO;2-V
   TERR LI, 1985, ARCH OTOLARYNGOL, V111, P495
   Waring M D, 1995, Ann Otol Rhinol Laryngol Suppl, V166, P33
   Wu H, 2000, AUDIOLOGY, V39, P247
NR 28
TC 20
Z9 21
U1 0
U2 1
PU AMER ASSOC NEUROLOGICAL SURGEONS
PI CHARLOTTESVILLE
PA UNIV VIRGINIA, 1224 WEST MAIN ST, STE 450, CHARLOTTESVILLE, VA 22903 USA
SN 0022-3085
J9 J NEUROSURG
JI J. Neurosurg.
PD DEC
PY 2001
VL 95
IS 6
BP 1028
EP 1033
DI 10.3171/jns.2001.95.6.1028
PG 6
WC Clinical Neurology; Surgery
WE Science Citation Index Expanded (SCI-EXPANDED)
SC Neurosciences & Neurology; Surgery
GA 499HA
UT WOS:000172564600017
PM 11765818
DA 2023-08-27
ER

PT J
AU van Zanten, SEMV
   Cruz, O
   Kaspers, GJL
   Hargrave, DR
   van Vuurden, DG
AF van Zanten, Sophie E. M. Veldhuijzen
   Cruz, Ofelia
   Kaspers, Gertjan J. L.
   Hargrave, Darren R.
   van Vuurden, Dannis G.
CA SIOPE DIPG Network
TI State of affairs in use of steroids in diffuse intrinsic pontine glioma:
   an international survey and a review of the literature
SO JOURNAL OF NEURO-ONCOLOGY
LA English
DT Review
DE Diffuse Intrinsic Pontine Glioma (DIPG); Pediatric oncology; Quality of
   life (QoL); Steroids; Side effects
ID PERITUMORAL BRAIN EDEMA; STEM TUMORS; PHASE-I; CHILDREN; DEXAMETHASONE;
   RADIATION; THERAPY; RADIOTHERAPY; MANAGEMENT
AB Children diagnosed with diffuse intrinsic pontine glioma (DIPG) face a dismal prognosis, with severe neurologic deterioration and inevitable death at a median of 9 months from diagnosis. Steroids are widely prescribed as supportive or palliative treatment although they are known to cause severe side effects that may reduce the quality of life. This study aims to review the current knowledge on, and use of, steroids in DIPG patients. A global questionnaire-study among health care professionals was performed to ascertain information on the current (multi-)institutional and (multi-)national use of steroids, the availability of clinical guidelines, and the need for improvements in prescribing steroids to DIPG patients. In addition, an extensive literature search was performed to review studies investigating steroids in pediatric brain tumor patients. From 150 responding health care professionals, only 7 % had clinical guidelines. The use of steroids was heterogeneous and over 85 % of respondents reported serious side effects. Fourteen articles, with low level of evidence, described the use of steroids in pediatric brain tumor patients. Clinical trials investigating optimal dose or regimen were lacking. This study is a first inventory of the availability of evidence-based information and clinical guidelines, and the current attitude towards the use of steroids in DIPG patients. To date, the risk-benefit ratio of steroids in this disease is yet to be determined. We emphasize the need for clinical trials resulting in guidelines on steroids, and possibly alternative drugs, to optimize the quality of care and quality of life of DIPG patients.
C1 [van Zanten, Sophie E. M. Veldhuijzen; Kaspers, Gertjan J. L.; van Vuurden, Dannis G.] Vrije Univ Amsterdam Med Ctr, Div Oncol Haematol, Dept Paediat, De Boelelaan 1118,Room KTC4-027, NL-1081 HZ Amsterdam, Netherlands.
   [Cruz, Ofelia] HSJD, Dept Paediat Oncol, Passeig St Joan de Deu 2, Barcelona 08950, Spain.
   [Kaspers, Gertjan J. L.] Princess Maxima Ctr Pediat Oncol, Lundlaan 6, NL-3584 EA Utrecht, Netherlands.
   [Hargrave, Darren R.] GOSH, Paediat Oncol Unit, Great Ormond St, London WC1N 3JH, England.
C3 Vrije Universiteit Amsterdam; VU UNIVERSITY MEDICAL CENTER; University
   of London; University College London; Great Ormond Street Hospital for
   Children NHS Foundation Trust
RP van Zanten, SEMV (通讯作者)，Vrije Univ Amsterdam Med Ctr, Div Oncol Haematol, Dept Paediat, De Boelelaan 1118,Room KTC4-027, NL-1081 HZ Amsterdam, Netherlands.
EM
RI Hargrave, Darren R/H-1066-2014; van Vuurden, Dannis/ACA-6952-2022;
   Kaspers, Gertjan/ABC-5334-2020
OI Hargrave, Darren R/0000-0001-8219-9807; Kaspers,
   Gertjan/0000-0001-7716-8475; cruz, ofelia/0000-0001-7102-3283;
   Veldhuijzen van Zanten, Sophie E.M./0000-0002-3888-0977; Van Vuurden,
   Dannis/0000-0002-1364-9007
FU Semmy Foundation (Stichting Semmy); NIHR Biomedical Research Centre at
   Great Ormond Street Hospital; Great Ormond Street Hospital Childrens
   Charity [W1075F] Funding Source: researchfish; The Brain Tumour Charity
   [16/193] Funding Source: researchfish
FX DIPG research in VU University Medical Center is possible thanks to
   financial support by the Semmy Foundation (Stichting Semmy). Dr Hargrave
   is supported by the NIHR Biomedical Research Centre at Great Ormond
   Street Hospital. We thank Fatma El-Khouly, pharmacy student, for
   checking the converted drug dosages. Special thanks go out to the
   members of the SIOPE DIPG Network for their input in creating the
   international survey and their guidance in interpreting the results.
CR [Anonymous], 2009, LEVELS EVIDENCE
   Beltran Chris, 2011, J Appl Clin Med Phys, V12, P3421
   Broniscer A, 2010, J CLIN ONCOL, V28, P4762, DOI 10.1200/JCO.2010.30.3545
   Chemotherapy Standardisation Group, 2008, EST BOD SURF AR INF
   Edelbauer M, 2002, PEDIATR ALLERGY IMMU, V13, P375, DOI 10.1034/j.1399-3038.2002.02039.x
   Fontanilla HP, 2012, AM J CLIN ONCOL-CANC, V35, P51, DOI 10.1097/COC.0b013e318201a2b7
   FREEMAN CR, 1991, CANCER-AM CANCER SOC, V68, P474, DOI 10.1002/1097-0142(19910801)68:3<474::AID-CNCR2820680305>3.0.CO;2-7
   FREEMAN CR, 1988, INT J RADIAT ONCOL, V15, P311, DOI 10.1016/S0360-3016(98)90010-4
   Glaser AW, 1997, ARCH DIS CHILD, V76, P76, DOI 10.1136/adc.76.1.76
   Grossman SA, 2011, CLIN CANCER RES, V17, P5473, DOI 10.1158/1078-0432.CCR-11-0774
   Hargrove D, 2006, LANCET ONCOL, V7, P241, DOI 10.1016/S1470-2045(06)70615-5
   Janssen G, 2000, KLIN PADIATR, V212, P189, DOI 10.1055/s-2000-9676
   Liu AK, 2009, INT J RADIAT ONCOL, V75, P1148, DOI 10.1016/j.ijrobp.2008.12.032
   Mallur PS, 2008, INT J PEDIATR OTORHI, V72, P529, DOI 10.1016/j.ijporl.2007.12.010
   Meyzer C, 2010, RADIAT ONCOL, V5, DOI 10.1186/1748-717X-5-9
   Moliterno JA, 2009, EXPERT OPIN INV DRUG, V18, P1413, DOI 10.1517/13543780903190689
   Mursch K, 2000, ACTA ANAESTH SCAND, V44, P378, DOI 10.1034/j.1399-6576.2000.440404.x
   SCHMID UD, 1986, J NEUROSURG, V65, P649, DOI 10.3171/jns.1986.65.5.0649
   TOFTEGAARD M, 1995, INTENS CARE MED, V21, P238, DOI 10.1007/BF01701480
   Villalona-Calero MA, 1998, ANN ONCOL, V9, P71, DOI 10.1023/A:1008251426425
   Warren KE, 2012, FRONT ONCOL, V2, DOI 10.3389/fonc.2012.00205
   Wheeler H, 2012, J CHILD NEUROL, V27, P927, DOI 10.1177/0883073811428007
   Wolff JEA, 1998, ANTICANCER RES, V18, P2895
   Yamasaki Fumiyuki, 2012, No Shinkei Geka, V40, P607
NR 24
TC 19
Z9 19
U1 0
U2 5
PU SPRINGER
PI NEW YORK
PA 233 SPRING ST, NEW YORK, NY 10013 USA
SN 0167-594X
EI 1573-7373
J9 J NEURO-ONCOL
JI J. Neuro-Oncol.
PD JUL
PY 2016
VL 128
IS 3
BP 387
EP 394
DI 10.1007/s11060-016-2141-x
PG 8
WC Oncology; Clinical Neurology
WE Science Citation Index Expanded (SCI-EXPANDED)
SC Oncology; Neurosciences & Neurology
GA DO6TB
UT WOS:000377915200002
PM 27177627
OA Green Published, hybrid
DA 2023-08-27
ER

PT J
AU Wang, X
   Zhang, HW
   Zhang, AL
   Han, L
   Wang, K
   Liu, R
   Yang, SH
   Pu, PY
   Shen, CH
   Kang, CS
   Yu, CJ
AF Wang, Xuan
   Zhang, Hongwei
   Zhang, Anling
   Han, Lei
   Wang, Kun
   Liu, Ran
   Yang, Shaohua
   Pu, Peiyu
   Shen, Changhong
   Kang, Chunsheng
   Yu, Chunjiang
TI Upregulation of miR-20a and miR-106b is involved in the acquisition of
   malignancy of pediatric brainstem gliomas
SO ONCOLOGY REPORTS
LA English
DT Article
DE miR-20a; miR-106b; microRNA; brainstem glioma; pediatric tumor
ID GLIOBLASTOMA CELLS; PROGNOSTIC-FACTORS; MICRORNAS; ADULTS; PROGRESSION;
   CHILDREN; IDENTIFICATION; PATHWAY; GROWTH; TUMORS
AB Brainstem glioma (BSG) is an entity which commonly occurs in pediatric patients and carries a dismal prognosis. However, the category of adult BSG has displayed considerably benign biological behavior, thus providing a unique perspective to comparatively understand the malignant features of pediatric BSG. microRNAs (miRNAs) have been associated with cancer development and progression. To identify miRNAs specifically involved in the acquisition of malignant progression of pediatric BSG, we analyzed the miRNA expression profiles in orthotopic models which could simulate the BSG heterogeneity by microarrays. Our research revealed that miR-20a and miR-106b (known to be closely related) were the two most robust upregulated miRNAs in pediatric BSG compared to adult subtype. Furthermore, the two types of human BSG tissue were utilized to verify the microarray data by qRT-PCR and in situ hybridization. The results indicated good consistency with that of the microarray method. In conclusion, our studies provide evidence that miR-20a and miR-106b may serve as putative causative involvement of malignant progression of pediatric BSG, thereby serving as likely novel targets for constraining the rapid fatal course of pediatric BSG.
C1 [Wang, Xuan; Zhang, Hongwei; Yu, Chunjiang] Capital Med Univ, Beijing Sanbo Brain Hosp, Dept Neurosurg, Beijing 100093, Peoples R China.
   [Zhang, Anling; Han, Lei; Pu, Peiyu; Shen, Changhong; Kang, Chunsheng] Tianjin Med Univ, Gen Hosp, Dept Neurosurg, Tianjin 300052, Peoples R China.
   [Zhang, Anling; Han, Lei; Pu, Peiyu; Kang, Chunsheng] Tianjin Neurol Inst, Lab Neurooncol, Tianjin 300052, Peoples R China.
   [Wang, Kun] Zhejiang Univ, Sch Med, Dept Neurosurg, Sir Run Run Show Hosp, Hangzhou 310016, Zhejiang, Peoples R China.
   [Liu, Ran; Yang, Shaohua] Univ N Texas Hlth Sci Ctr, Dept Pharmacol & Neurosci, Ft Worth, TX 76107 USA.
C3 Capital Medical University; Tianjin Medical University; Zhejiang
   University; University of North Texas System; University of North Texas
   Health Science Center
RP Yu, CJ (通讯作者)，Capital Med Univ, Beijing Sanbo Brain Hosp, Dept Neurosurg, 50 Xiangshan Yikesong Rd, Beijing 100093, Peoples R China.
EM
RI Zhang, An/GRS-5852-2022; Kang, Chunsheng/AAM-5979-2021
OI Kang, Chunsheng/0000-0002-3255-3369; Liu, Ran/0000-0002-3882-0616
FU China National Natural Scientific Found [81172399, 30872647]; Beijing
   Natural Science Found [7092041]
FX This study was supported by the China National Natural Scientific Found
   (81172399, 30872647), and the Beijing Natural Science Found (7092041).
CR Bhatti I, 2009, J GASTROINTEST SURG, V13, P1379, DOI 10.1007/s11605-009-0887-6
   Bottoni A, 2007, J CELL PHYSIOL, V210, P370, DOI 10.1002/jcp.20832
   Ciafre SA, 2005, BIOCHEM BIOPH RES CO, V334, P1351, DOI 10.1016/j.bbrc.2005.07.030
   Cui QH, 2006, MOL SYST BIOL, V2, DOI 10.1038/msb4100089
   Epstein F, 1990, Neurosurg Clin N Am, V1, P111
   Gilbertson RJ, 2003, CLIN CANCER RES, V9, P3620
   Gokhale A, 2010, J CANCER RES THER, V6, P521, DOI 10.4103/0973-1482.77072
   Guillamo JS, 2001, CURR OPIN NEUROL, V14, P711, DOI 10.1097/00019052-200112000-00006
   Guillamo JS, 2001, BRAIN, V124, P2528, DOI 10.1093/brain/124.12.2528
   Hargrove D, 2006, LANCET ONCOL, V7, P241, DOI 10.1016/S1470-2045(06)70615-5
   Hashizume R, 2010, J NEURO-ONCOL, V96, P151, DOI 10.1007/s11060-009-9954-9
   Inomata M, 2009, BLOOD, V113, P396, DOI 10.1182/blood-2008-07-163907
   Ivanovska I, 2008, MOL CELL BIOL, V28, P2167, DOI 10.1128/MCB.01977-07
   Jallo G, 2006, CHILD NERV SYST, V22, P1, DOI 10.1007/s00381-005-1267-5
   Karginov FV, 2007, P NATL ACAD SCI USA, V104, P19291, DOI 10.1073/pnas.0709971104
   Kesari S, 2008, J NEURO-ONCOL, V88, P175, DOI 10.1007/s11060-008-9545-1
   Laigle-Donadey F, 2008, CURR OPIN ONCOL, V20, P662, DOI 10.1097/CCO.0b013e32831186e0
   LEE RC, 1993, CELL, V75, P843, DOI 10.1016/0092-8674(93)90529-Y
   Liu Q, 2008, J NEUROSURG, V109, P849, DOI 10.3171/JNS/2008/109/11/0849
   Malzkorn B, 2010, BRAIN PATHOL, V20, P539, DOI 10.1111/j.1750-3639.2009.00328.x
   Mischel PS, 2003, BRAIN PATHOL, V13, P52
   Miwa T, 2009, J NEURO-ONCOL, V91, P251, DOI 10.1007/s11060-008-9714-2
   Mursch K, 2005, BRIT J NEUROSURG, V19, P128, DOI 10.1080/02688690500145530
   Pang JCS, 2009, ACTA NEUROPATHOL, V117, P599, DOI 10.1007/s00401-009-0525-0
   Peck B, 2011, AGING-US, V3, P329, DOI 10.18632/aging.100302
   Petrocca F, 2008, CANCER RES, V68, P8191, DOI 10.1158/0008-5472.CAN-08-1768
   Rutka JT, 2008, NEOPLASIA, V10, P149, DOI 10.1593/neo.07909
   Salmaggi A, 2008, J NEUROL, V255, P171, DOI 10.1007/s00415-008-0589-0
   Selvapandian S, 1999, ACTA NEUROCHIR, V141, P721, DOI 10.1007/s007010050367
   Song YJ, 2011, BRAIN RES, V1387, P134, DOI 10.1016/j.brainres.2011.02.073
   Wang X, 2011, ONCOL REP, V25, P97, DOI 10.3892/or_00001047
   Yaz G, 2004, AM J CLIN ONCOL-CANC, V27, P384, DOI 10.1097/01.COC.0000071945.15623.C6
   Yin LX, 2010, ZHONG GUO WEI QIN XI, V15, P158
   Zhou X, 2010, LAB INVEST, V90, P144, DOI 10.1038/labinvest.2009.126
NR 34
TC 13
Z9 16
U1 0
U2 20
PU SPANDIDOS PUBL LTD
PI ATHENS
PA POB 18179, ATHENS, 116 10, GREECE
SN 1021-335X
EI 1791-2431
J9 ONCOL REP
JI Oncol. Rep.
PD OCT
PY 2012
VL 28
IS 4
BP 1293
EP 1300
DI 10.3892/or.2012.1927
PG 8
WC Oncology
WE Science Citation Index Expanded (SCI-EXPANDED)
SC Oncology
GA 013OT
UT WOS:000309315400025
PM 22825541
OA Bronze
DA 2023-08-27
ER

PT J
AU Rais-Bahrami, S
   Argani, P
AF Rais-Bahrami, Soroush
   Argani, Pedram
TI Unusual renal pathology associated with a Wilms tumor in a 15-month-old
   infant
SO INTERNATIONAL JOURNAL OF SURGICAL PATHOLOGY
LA English
DT Article
DE Beckwith-Wiedemann syndrome; dysplastic medullary ray nodule; Beckwith
   medulla
ID BECKWITH-WIEDEMANN-SYNDROME
AB Beckwith-Wiedemann syndrome is an overgrowth disorder that predisposes to the development of a variety of malignancies including Wilms tumor. Pathologists may suspect the presence of Beckwith-Wiedemann syndrome, which may be clinically subtle, on the basis of the histopathology of the renal parenchyma adjacent to a Wilms tumor. These kidneys typically harbor multiple nephrogenic rests (especially perilobar nephrogenic rests), and their medullas have blunted papillae resulting from excessive fibromyxoid stroma, decreased collecting ducts, and decreased Henle loops. This complex of findings constitutes the so-called Beckwith Medulla. Extensions of this stroma-rich medulla into the cortex results in characteristic dysplastic medullary-ray nodules. Identification of these histologic abnormalities by the pathologist should prompt the clinician to evaluate the child for Beckwith-Wiedemann syndrome. Proper screening of these children can allow for early detection of cancers and treatment while these cancers are curable.
C1 Johns Hopkins Med Inst, Baltimore, MD 21205 USA.
C3 Johns Hopkins University; Johns Hopkins Medicine
RP Argani, P (通讯作者)，Johns Hopkins Univ Hosp, Div Surg Pathol, Weinberg Bldg,401 N Broadway Room 2242, Baltimore, MD 21231 USA.
EM
OI Rais-Bahrami, Soroush/0000-0001-9466-9925
CR ARGANI P, 2004, STERNBERGS DIAGNOSTI, P2001
   BECKWITH J B, 1990, Pediatric Pathology, V10, P1
   BECKWITH J B, 1969, Birth Defects Original Article Series, V5, P188
   Beckwith JB, 1998, J PEDIATR-US, V132, P377, DOI 10.1016/S0022-3476(98)70001-0
   Beckwith JB, 1963, ANN M W SOC PED RES
   Cohen MM, 2005, PEDIATR DEVEL PATHOL, V8, P287, DOI 10.1007/s10024-005-1154-9
   Li M, 1998, AM J MED GENET, V79, P253
   Weksberg R, 2003, HUM MOL GENET, V12, pR61, DOI 10.1093/hmg/ddg067
   WENG EY, 1995, CLIN PEDIATR, V34, P317, DOI 10.1177/000992289503400605
   WIEDEMANN H. R., 1964, J GENET HUM, V13, P223
NR 10
TC 1
Z9 1
U1 0
U2 0
PU SAGE PUBLICATIONS INC
PI THOUSAND OAKS
PA 2455 TELLER RD, THOUSAND OAKS, CA 91320 USA
SN 1066-8969
J9 INT J SURG PATHOL
JI Int. J. Surg. Pathol.
PD JUL
PY 2006
VL 14
IS 3
BP 218
EP 220
DI 10.1177/1066896906290144
PG 3
WC Pathology; Surgery
WE Science Citation Index Expanded (SCI-EXPANDED)
SC Pathology; Surgery
GA 073XP
UT WOS:000239776300006
PM 16959702
DA 2023-08-27
ER

PT J
AU Prasad, SR
   Humphrey, PA
   Menias, CO
   Middleton, WD
   Siegel, MJ
   Bae, KT
   Heiken, JP
AF Prasad, SR
   Humphrey, PA
   Menias, CO
   Middleton, WD
   Siegel, MJ
   Bae, KT
   Heiken, JP
TI Neoplasms of the renal medulla: Radiologic-pathologic correlation
SO RADIOGRAPHICS
LA English
DT Article
ID RHABDOID TUMOR; KIDNEY; CARCINOMA; LYMPHOMA; CT; CHILDREN
AB Tumors of the renal medulla cover a wide spectrum, with characteristic histomorphology and variable biologic profiles. Renal medullary tumors can be categorized into benign and malignant neoplasms based on histologic features and clinico-biologic behavior. They can be further classified into pediatric and adult tumors based on the patient age group. When small, renal medullary tumors may be differentiated from the more common renal adenocarcinomas by their central location and certain demographic characteristics. Although most large malignant medullary tumors demonstrate imaging findings that are indistinguishable from those of other renal malignancies, some tumors demonstrate imaging findings that may suggest a specific diagnosis. (c) RSNA, 2005.
C1 Washington Univ, Sch Med, Mallinckrodt Inst Radiol, St Louis, MO 63110 USA.
   Univ Texas, Hlth Sci Ctr, Dept Radiol, San Antonio, TX 78285 USA.
C3 Washington University (WUSTL); University of Texas System; University of
   Texas Health Science Center at San Antonio
RP Heiken, JP (通讯作者)，Washington Univ, Sch Med, Mallinckrodt Inst Radiol, 510 S Kingshighway Blvd,9th Fl, St Louis, MO 63110 USA.
EM
RI Prasad, Srinivasa/AAJ-8654-2020
OI Prasad, Srinivasa/0000-0003-4594-2117
CR Adeniran A, 2003, ARCH PATHOL LAB MED, V127, P1637
   Agrons GA, 1997, AM J ROENTGENOL, V168, P447, DOI 10.2214/ajr.168.2.9016225
   Argani P, 2000, AM J SURG PATHOL, V24, P4, DOI 10.1097/00000478-200001000-00002
   CATALONA WJ, 1998, CAMPBELLS UROLOGY, P2383
   Chao D, 2002, J UROLOGY, V167, P71, DOI 10.1016/S0022-5347(05)65385-2
   CHUNG CJ, 1995, AM J ROENTGENOL, V164, P697, DOI 10.2214/ajr.164.3.7863897
   Daneshmand S, 2002, J UROLOGY, V167, P488, DOI 10.1016/S0022-5347(01)69070-0
   DAVIDSON AJ, 1995, RADIOLOGY, V195, P83, DOI 10.1148/radiology.195.1.7892499
   DAVIS CJ, 1995, AM J SURG PATHOL, V19, P1, DOI 10.1097/00000478-199501000-00001
   FERRY JA, 1995, AM J SURG PATHOL, V19, P134, DOI 10.1097/00000478-199502000-00002
   GRIGNON DJ, 1990, CANCER, V65, P1611, DOI 10.1002/1097-0142(19900401)65:7<1611::AID-CNCR2820650727>3.0.CO;2-G
   Han TI, 2001, PEDIATR RADIOL, V31, P233, DOI 10.1007/s002470000417
   KIDD JM, 1970, AM J PATHOL, V59, pA16
   Lee H S, 2000, Korean J Radiol, V1, P60
   Lowe LH, 2000, RADIOGRAPHICS, V20, P1585, DOI 10.1148/radiographics.20.6.g00nv051585
   ORDONEZ NG, 1996, ACKERMANS SURG PATHO, P1140
   Pickhardt PJ, 2000, RADIOGRAPHICS, V20, P215, DOI 10.1148/radiographics.20.1.g00ja08215
   Pickhardt PJ, 2001, AM J ROENTGENOL, V176, P627, DOI 10.2214/ajr.176.3.1760627
   Sheeran SR, 1998, AM J ROENTGENOL, V171, P1067, DOI 10.2214/ajr.171.4.9762998
   Swartz MA, 2002, UROLOGY, V60, P1083, DOI 10.1016/S0090-4295(02)02154-4
   Urban BA, 2000, RADIOGRAPHICS, V20, P197, DOI 10.1148/radiographics.20.1.g00ja09197
NR 21
TC 40
Z9 44
U1 0
U2 1
PU RADIOLOGICAL SOC NORTH AMERICA
PI OAK BROOK
PA 820 JORIE BLVD, OAK BROOK, IL 60523 USA
SN 0271-5333
J9 RADIOGRAPHICS
JI Radiographics
PD MAR-APR
PY 2005
VL 25
IS 2
BP 369
EP 380
DI 10.1148/rg.252045073
PG 12
WC Radiology, Nuclear Medicine & Medical Imaging
WE Science Citation Index Expanded (SCI-EXPANDED)
SC Radiology, Nuclear Medicine & Medical Imaging
GA 034OU
UT WOS:000236940200009
PM 15798056
DA 2023-08-27
ER

PT J
AU Yakupoglu, H
   Civelek, E
   Onal, B
   Kircelli, A
   Ulubay, H
   Diren, B
AF Yakupoglu, Hakan
   Civelek, Erdinc
   Onal, Bulent
   Kircelli, Attila
   Ulubay, Hakan
   Diren, Baris
TI Surgery Decision in Brainstem Mass Resection by Diffusion Tensor Based
   Fiber Tracking Assistance: A Report of Four Cases
SO JOURNAL OF NEUROLOGICAL SCIENCES-TURKISH
LA English
DT Article
DE Brainstem; Diffusion tensor imaging; White matter tractography
ID NEURONAVIGATION; TRACT
AB Brainstem is surrounded by noteworthy dense packed white matter fibers which must be preserved during surgery. Brain stem surgery carries serious risks. It is very important to know about the localization of the tumor and the eloquent white matter tracts for surgery decision and for preoperative neurosurgical planning. In this report, four cases with brain stem mass were discussed. The three of them were operated and the one of the four cases was referred to Oncology department for radiotherapy in which major fibers could not be preserved based on White matter tractography (WMT) and Diffusion tensor imaging (DTI) findings.
C1 [Civelek, Erdinc] Baskent Univ, Istanbul Hosp, Dept Neurosurg, Istanbul, Turkey.
   [Yakupoglu, Hakan] Medicana Ankara Hosp, Dept Neurosurg, Ankara, Turkey.
   [Onal, Bulent] Siirt State Hosp, Siirt, Turkey.
   [Kircelli, Attila] Goztepe Safak Hosp, Dept Neurosurg, Istanbul, Turkey.
   [Ulubay, Hakan; Diren, Baris] Medicana Ankara Hosp, Dept Radiol, Ankara, Turkey.
C3 Baskent University; Private Istanbul Hospital; Medicana Hospital; Siirt
   State Hospital; Medicana Hospital
RP Civelek, E (通讯作者)，Baskent Univ, Istanbul Hosp, Dept Neurosurg, Istanbul, Turkey.
EM
RI Civelek, Erdinc/ABI-6105-2020; DIREN, HALDUN BARIS/O-7235-2018
OI Civelek, Erdinc/0000-0002-3988-4064; DIREN, HALDUN
   BARIS/0000-0001-8958-2040
CR Chen X, 2007, ACTA NEUROCHIR, V149, P1117, DOI 10.1007/s00701-007-1282-2
   Chen XL, 2007, SURG NEUROL, V68, P285, DOI 10.1016/j.surneu.2007.05.005
   FLORENCE JM, 1992, PHYS THER, V72, P115, DOI 10.1093/ptj/72.2.115
   Helton KJ, 2006, AM J NEURORADIOL, V27, P786
   Laundre BJ, 2005, AM J NEURORADIOL, V26, P791
   Price SJ, 2006, AM J NEURORADIOL, V27, P1969
   Wang SM, 2009, NEUROIMAGE, V44, P653, DOI 10.1016/j.neuroimage.2008.09.027
   Wang W, 2009, AM J NEURORADIOL, V30, P203, DOI 10.3174/ajnr.A1303
   Wu JS, 2007, NEUROSURGERY, V61, P935, DOI 10.1227/01.neu.0000303189.80049.ab
NR 9
TC 1
Z9 1
U1 0
U2 0
PU JOURNAL NEUROLOGICAL SCIENCES
PI BORNOVA-IZMIR
PA EGE UNIV HOSP. FAC MED, DEPT NEUROSURGERY, BORNOVA-IZMIR, TR35100,
   TURKEY
SN 1302-1664
J9 J NEUROL SCI-TURK
JI J. Neurol. Sci.-Turk.
PY 2012
VL 29
IS 1
BP 117
EP 121
PG 5
WC Neurosciences
WE Science Citation Index Expanded (SCI-EXPANDED)
SC Neurosciences & Neurology
GA 946BX
UT WOS:000304324800013
DA 2023-08-27
ER

PT J
AU Jaimes, C
   Vajapeyam, S
   Brown, D
   Kao, PC
   Ma, C
   Greenspan, L
   Gupta, N
   Goumnerova, L
   Bandopahayay, P
   Dubois, F
   Greenwald, NF
   Zack, T
   Shapira, O
   Beroukhim, R
   Ligon, KL
   Chi, S
   Kieran, MW
   Wright, KD
   Poussaint, TY
AF Jaimes, C.
   Vajapeyam, S.
   Brown, D.
   Kao, P. -C.
   Ma, C.
   Greenspan, L.
   Gupta, N.
   Goumnerova, L.
   Bandopahayay, P.
   Dubois, F.
   Greenwald, N. F.
   Zack, T.
   Shapira, O.
   Beroukhim, R.
   Ligon, K. L.
   Chi, S.
   Kieran, M. W.
   Wright, K. D.
   Poussaint, T. Y.
TI MR Imaging Correlates for Molecular and Mutational Analyses in Children
   with Diffuse Intrinsic Pontine Glioma
SO AMERICAN JOURNAL OF NEURORADIOLOGY
LA English
DT Article
ID COEFFICIENT HISTOGRAM METRICS; APPARENT DIFFUSION; MIDLINE GLIOMAS;
   DISTINCT SUBGROUPS; SURVIVAL; CLASSIFICATION; PREDICTORS; SURROGATES;
   BIOPSY; TUMORS
AB BACKGROUND AND PURPOSE:
   Recent advances in molecular techniques have characterized distinct subtypes of diffuse intrinsic pontine gliomas. Our aim was the identification of MR imaging correlates of these subtypes.
   MATERIALS AND METHODS:
   Initial MRIs from subjects with diffuse intrinsic pontine gliomas recruited for a prospective clinical trial before treatment were analyzed. Retrospective imaging analyses included FLAIR/T2 tumor volume, tumor volume enhancing, the presence of cyst and/or necrosis, median, mean, mode, skewness, kurtosis of ADC tumor volume based on FLAIR, and enhancement at baseline. Molecular subgroups based on EGFR and MGMT mutations were established. Histone mutations were also determined (H3F3A, HIST1H3B, HIST1H3C). Univariate Cox proportional hazards regression was used to test the association of imaging predictors with overall and progression-free survival. Wilcoxon rank sum, Kruskal-Wallis, and Fisher exact tests were used to compare imaging measures among groups.
   RESULTS:
   Fifty patients had biopsy and MR imaging. The median age at trial registration was 6?years (range, 3.3?17.5?years); 52% were female. On the basis of immunohistochemical results, 48 patients were assigned to 1 of 4 subgroups: 28 in MGMT?/epidermal growth factor receptor (EGFR)?, 14 in MGMT?/EGFR+, 3 in MGMT+/EGFR?, and 3 in MGMT+/EGFR+. Twenty-three patients had histone mutations in H3F3A, 8 in HIST1H3B, and 3 in HIST1H3C. Enhancing tumor volume was near-significantly different across molecular subgroups (P?=?.04), after accounting for the false discovery rate. Tumor volume enhancing, median, mode, skewness, and kurtosis ADC T2-FLAIR/T2 were significantly different (P???.048) between patients with H3F3A and HIST1H3B/C mutations.
   CONCLUSIONS:
   MR imaging features including enhancement and ADC histogram parameters are correlated with molecular subgroups and mutations in children with diffuse intrinsic pontine gliomas.
   Initial MRIs from 50 subjects with diffuse intrinsic pontine gliomas recruited for a prospective clinical trial before treatment were analyzed. Retrospective imaging analyses included FLAIR/T2 tumor volume, tumor volume enhancing, the presence of cyst and/or necrosis, median, mean, mode, skewness, kurtosis of ADC tumor volume based on FLAIR, and enhancement at baseline. Molecular subgroups based on EGFR and MGMT mutations were established. Histone mutations were also determined (H3F3A, HIST1H3B, HIST1H3C). Enhancing tumor volume was near-significantly different across molecular subgroups, after accounting for the false discovery rate. Tumor volume enhancing, median, mode, skewness, and kurtosis ADC T2-FLAIR/T2 were significantly different between patients with H3F3A and HIST1H3B/C mutations.
C1 [Jaimes, C.; Vajapeyam, S.; Poussaint, T. Y.] Boston Childrens Hosp, Dept Radiol, 300 Longwood Ave, Boston, MA 02115 USA.
   [Kao, P. -C.; Ma, C.; Greenspan, L.; Bandopahayay, P.; Beroukhim, R.; Chi, S.; Wright, K. D.] Boston Childrens Hosp, Dept Pediat, Div Oncol, Boston, MA USA.
   [Jaimes, C.] Boston Childrens Hosp, Div Newborn Med, Fetal Neonatal Neuroimaging & Dev Sci Ctr, Boston, MA USA.
   [Brown, D.] Massachusetts Gen Hosp, Tumor Imaging Metr Core, Boston, MA 02114 USA.
   [Kao, P. -C.; Ma, C.; Greenspan, L.; Bandopahayay, P.; Dubois, F.; Shapira, O.; Beroukhim, R.; Ligon, K. L.; Chi, S.; Wright, K. D.] Dana Farber Canc Inst, Boston, MA 02115 USA.
   [Gupta, N.] Univ Calif San Francisco, Benioff Childrens Hosp, Dept Pediat Neurosurg, San Francisco, CA 94143 USA.
   [Kieran, M. W.] Bristol Myers Squibb, Clin Trials Div, New York, NY USA.
   [Ligon, K. L.] Brigham & Womens Hosp, Dept Pathol, 75 Francis St, Boston, MA 02115 USA.
   [Jaimes, C.; Vajapeyam, S.; Ma, C.; Bandopahayay, P.; Dubois, F.; Beroukhim, R.; Ligon, K. L.; Chi, S.; Wright, K. D.; Poussaint, T. Y.] Harvard Med Sch, Boston, MA 02115 USA.
   [Gupta, N.; Zack, T.] Univ Calif San Francisco, Sch Med, San Francisco, CA USA.
   [Greenwald, N. F.] Stanford Univ, Sch Med, Palo Alto, CA 94304 USA.
   [Shapira, O.] Broad Inst Massachusetts Inst Technol & Harvard U, Cambridge, MA USA.
C3 Harvard University; Boston Children's Hospital; Harvard University;
   Boston Children's Hospital; Harvard University; Boston Children's
   Hospital; Harvard University; Massachusetts General Hospital; Harvard
   University; Dana-Farber Cancer Institute; University of California
   System; University of California San Francisco; Bristol-Myers Squibb;
   Harvard University; Brigham & Women's Hospital; Harvard University;
   Harvard Medical School; University of California System; University of
   California San Francisco; Stanford University; Harvard University;
   Massachusetts Institute of Technology (MIT); Broad Institute
RP Poussaint, TY (通讯作者)，Boston Childrens Hosp, Dept Radiol, 300 Longwood Ave, Boston, MA 02115 USA.
EM
RI Greenwald, Noah F./GNH-3982-2022
OI Greenwald, Noah F./0000-0002-7836-4379; Kao,
   Pei-Chi/0000-0003-0702-8324; Gupta, Nalin/0000-0001-9539-7052;
   Vajapeyam, Sridhar/0000-0002-1782-1919; Jaimes,
   Camilo/0000-0002-7169-4244; Ma, Clement/0000-0002-8188-9600
FU Schlaeger Fellowship for Neuroscience Research
FX C. Jaimes was partially supported by the Schlaeger Fellowship for
   Neuroscience Research.
CR Aboian MS, 2019, AM J NEURORADIOL, V40, P1804, DOI 10.3174/ajnr.A6302
   Aboian MS, 2017, AM J NEURORADIOL, V38, P795, DOI 10.3174/ajnr.A5076
   Bailey S, 2013, EUR J CANCER, V49, P3856, DOI 10.1016/j.ejca.2013.08.006
   BENJAMINI Y, 1995, J R STAT SOC B, V57, P289, DOI 10.1111/j.2517-6161.1995.tb02031.x
   Buczkowicz P, 2014, ACTA NEUROPATHOL, V128, P573, DOI 10.1007/s00401-014-1319-6
   Castel D, 2015, ACTA NEUROPATHOL, V130, P815, DOI 10.1007/s00401-015-1478-0
   Chen HJ, 2010, AM J NEURORADIOL, V31, P1879, DOI 10.3174/ajnr.A2179
   Chen H, 2019, EUR J RADIOL, V114, P152, DOI 10.1016/j.ejrad.2019.03.006
   Khuong-Quang DA, 2012, ACTA NEUROPATHOL, V124, P439, DOI 10.1007/s00401-012-0998-0
   Fischbein NJ, 1996, PEDIATR NEUROSURG, V24, P9, DOI 10.1159/000121010
   Goya-Outi JC, 2019, P I EL EL ENG ENG ME, P1
   Goya-Outi J, 2018, PHYS MED BIOL, V63, DOI 10.1088/1361-6560/aabd21
   Gupta N, 2018, NEURO-ONCOLOGY, V20, P1547, DOI 10.1093/neuonc/noy070
   Gutman DA, 2013, RADIOLOGY, V267, P560, DOI 10.1148/radiol.13120118
   Hassan H, 2017, J NEURO-ONCOL, V135, P13, DOI 10.1007/s11060-017-2546-1
   Hoffman LM, 2018, J CLIN ONCOL, V36, P1963, DOI 10.1200/JCO.2017.75.9308
   Infinger LK, 2017, CURR NEUROPHARMACOL, V15, P129, DOI 10.2174/1570159X14666160425114024
   Jansen MH, 2015, NEURO-ONCOLOGY, V17, P160, DOI 10.1093/neuonc/nou104
   Jenkinson M, 2012, NEUROIMAGE, V62, P782, DOI 10.1016/j.neuroimage.2011.09.015
   Kieran MW, 2016, J NEUROSURG-PEDIATR, V18, P390, DOI 10.3171/2015.6.PEDS15374
   Lober RM, 2014, J NEURO-ONCOL, V117, P175, DOI 10.1007/s11060-014-1375-8
   Lobel U, 2011, AM J NEURORADIOL, V32, P315, DOI 10.3174/ajnr.A2277
   Louis DN, 2016, ACTA NEUROPATHOL, V131, P803, DOI 10.1007/s00401-016-1545-1
   Nowak J, 2018, NEURO-ONCOLOGY, V20, P1672, DOI 10.1093/neuonc/noy111
   Nowosielski M, 2011, NEURORADIOLOGY, V53, P291, DOI 10.1007/s00234-010-0808-0
   Ostrom QT, 2018, NEURO-ONCOLOGY, V20, P1, DOI 10.1093/neuonc/noy131
   Perreault S, 2014, AM J NEURORADIOL, V35, P1263, DOI 10.3174/ajnr.A3990
   Pope WB, 2008, RADIOLOGY, V249, P268, DOI 10.1148/radiol.2491072000
   Poussaint TY, 2016, NEURO-ONCOLOGY, V18, P725, DOI 10.1093/neuonc/nov256
   Poussaint TY, 2011, NEURO-ONCOLOGY, V13, P417, DOI 10.1093/neuonc/noq200
   Schindelin J, 2012, NAT METHODS, V9, P676, DOI [10.1038/NMETH.2019, 10.1038/nmeth.2019]
   Yan H, 2009, NEW ENGL J MED, V360, P765, DOI 10.1056/NEJMoa0808710
   Zukotynski KA, 2017, J NUCL MED, V58, P1264, DOI 10.2967/jnumed.116.185389
NR 33
TC 12
Z9 12
U1 0
U2 10
PU AMER SOC NEURORADIOLOGY
PI DENVILLE
PA PO BOX 3000, DENVILLE, NJ 07834-9349 USA
SN 0195-6108
EI 1936-959X
J9 AM J NEURORADIOL
JI Am. J. Neuroradiol.
PD MAY
PY 2020
VL 41
IS 5
BP 874
EP 881
DI 10.3174/ajnr.A6546
PG 8
WC Clinical Neurology; Neuroimaging; Radiology, Nuclear Medicine & Medical
   Imaging
WE Science Citation Index Expanded (SCI-EXPANDED)
SC Neurosciences & Neurology; Radiology, Nuclear Medicine & Medical Imaging
GA LS1FU
UT WOS:000536138400028
PM 32381545
OA hybrid, Green Published
DA 2023-08-27
ER

PT J
AU Matthies, C
   Samii, M
AF Matthies, C
   Samii, M
TI Direct brainstem recording of auditory evoked potentials during
   vestibular schwannoma resection: Nuclear BAEP recording - Technical note
   and preliminary results
SO JOURNAL OF NEUROSURGERY
LA English
DT Article; Proceedings Paper
CT 2nd International Skull Base Congress / 7th Annual Meeting of the
   North-American-Skull-Base-Society
CY JUN 29-JUL 04, 1996
CL SAN DIEGO, CA
SP N Amer Skull Base Soc
DE acoustic nerve tumor; auditory nucleus; cerebellopontine angle;
   brainstem auditory evoked potential; direct eighth cranial nerve
   recording; hearing preservation
ID ACOUSTIC NEUROMA SURGERY; HEARING PRESERVATION; CEREBELLOPONTINE ANGLE;
   NEURINOMA SURGERY; RESPONSES
AB The usefulness of intraoperative monitoring in cerebellopontine angle surgery should be improved by obtaining faster and stronger brainstem auditory evoked potential (BAEP) responses. A new technique of direct recording at thee brainstem has been developed, which is applicable to all tumor sizes. By placing a retractor with electrodes attached to its tip at the cerebellomedullary junction, the authors have recorded BAEP amplitudes that are 10 times greater than those recorded using the conventional technique. Only small sampling numbers (64-256 recordings) are required and are obtained in 5 to 15 seconds. The technique has been applied successfully in 34 patients who underwent vestibular schwannoma resections. It has also been tested in patients with intrameatal-extrameatal meningiomas and in those with vascular compressive disorders; there have been no false results. The advantages of this new technique are: 1) identification of BAEP components is easier and faster; 2) reliable BAEP responses are obtained in some cases in which conventional BAEP responses are lost or severely deformed; and 3) BAEP response deterioration and improvement are recognized earlier than would occur using the conventional technique. This last advantage provides the surgeon with a useful warning at a stage of surgery at which BAEP changes are still temporary and can be reversed. This method is different from other trials of intradural BAEP recordings in three respects: its use is not limited to particular tumor sizes; there is no interference with the surgical process; and, most important, the obtained responses correlate well with those of conventional BAEP responses, probably because the recording site is in the vicinity of the anterior cochlear nucleus. In conclusion, the chances of useful monitoring feedback with adequate adaptation of the microsurgical strategy are improved considerably.
C1 HANNOVER MED SCH,TEACHING HOSP,D-3000 HANNOVER,GERMANY.
C3 Hannover Medical School
RP Matthies, C (通讯作者)，NORDSTADT HOSP,DEPT NEUROSURG,HALTENHOFFSTR 41,D-30167 HANNOVER,GERMANY.
CR ABRAMSON M, 1985, LARYNGOSCOPE, V95, P1318
   COHEN NL, 1986, ANN OTO RHINOL LARYN, V95, P21, DOI 10.1177/000348948609500105
   COLLETTI V, 1993, ACTA OTO-LARYNGOL, V113, P746, DOI 10.3109/00016489309135895
   FISCHER G, 1992, J NEUROSURG, V76, P910, DOI 10.3171/jns.1992.76.6.0910
   FISHER RS, 1995, J CLIN NEUROPHYSIOL, V12, P97
   GLASSCOCK ME, 1993, J NEUROSURG, V78, P864, DOI 10.3171/jns.1993.78.6.0864
   HAINES SJ, 1993, J NEUROSURG, V79, P515, DOI 10.3171/jns.1993.79.4.0515
   LANDI A, 1991, STEREOT FUNCT NEUROS, V57, P167, DOI 10.1159/000099569
   Matthies C, 1997, NEUROSURGERY, V40, P459, DOI 10.1097/00006123-199703000-00007
   MATTHIES C, 1994, SKULL BASE SURGERY, P1177
   MOLLER AR, 1994, NEUROSURGERY, V34, P688, DOI 10.1227/00006123-199404000-00018
   MOLLER AR, 1983, SCAND AUDIOL, V12, P125, DOI 10.3109/01050398309076235
   MOLLER AR, 1994, ELECTROEN CLIN NEURO, V92, P215, DOI 10.1016/0168-5597(94)90065-5
   OJEMANN RG, 1984, J NEUROSURG, V61, P938, DOI 10.3171/jns.1984.61.5.0938
   POST KD, 1995, J NEUROSURG, V83, P191, DOI 10.3171/jns.1995.83.2.0191
   Samii M, 1995, Adv Tech Stand Neurosurg, V22, P343
   Samii M., 1996, CURRENT TECHNIQUES N, P93
   SILVERSTEIN H, 1985, AM J OTOL, P99
   SILVERSTEIN H, 1993, OTOLARYNG HEAD NECK, V108, P1
   SILVERSTEIN H, 1986, OTOLARYNG HEAD NECK, V95, P285, DOI 10.1177/01945998860953P104
   Symon L, 1989, Br J Neurosurg, V3, P343, DOI 10.3109/02688698909002814
   TANIGUCHI M, 1992, HNO, V40, P186
   TORRENS M, 1994, BRIT J NEUROSURG, V8, P655, DOI 10.3109/02688699409101179
   WIGAND ME, 1989, ARCH OTO-RHINO-LARYN, V246, P299, DOI 10.1007/BF00463580
   YAMAMOTO T, 1994, BRAIN INJURY, V8, P623, DOI 10.3109/02699059409151015
   YINGLING CD, 1992, OTOLARYNG CLIN N AM, V25, P413
NR 26
TC 28
Z9 38
U1 0
U2 0
PU AMER ASSOC NEUROLOGICAL SURGEONS
PI CHARLOTTESVILLE
PA UNIV VIRGINIA, 1224 WEST MAIN ST, STE 450, CHARLOTTESVILLE, VA 22903
SN 0022-3085
J9 J NEUROSURG
JI J. Neurosurg.
PD JUN
PY 1997
VL 86
IS 6
BP 1057
EP 1062
DI 10.3171/jns.1997.86.6.1057
PG 6
WC Clinical Neurology; Surgery
WE Conference Proceedings Citation Index - Science (CPCI-S); Science Citation Index Expanded (SCI-EXPANDED)
SC Neurosciences & Neurology; Surgery
GA XA676
UT WOS:A1997XA67600023
PM 9171191
DA 2023-08-27
ER

PT J
AU van Zanten, SEMV
   El-Khouly, FE
   Jansen, MHA
   Bakker, DP
   Aliaga, ES
   Haasbeek, CJA
   Wolf, NI
   Zwaan, CM
   Vandertop, WP
   van Vuurden, DG
   Kaspers, GJL
AF van Zanten, Sophie E. M. Veldhuijzen
   El-Khouly, Fatma E.
   Jansen, Marc H. A.
   Bakker, Dewi P.
   Aliaga, Esther Sanchez
   Haasbeek, Cornelis J. A.
   Wolf, Nicole I.
   Zwaan, C. Michel
   Vandertop, W. Peter
   van Vuurden, Dannis G.
   Kaspers, Gertjan J. L.
TI A phase I/II study of gemcitabine during radiotherapy in children with
   newly diagnosed diffuse intrinsic pontine glioma
SO JOURNAL OF NEURO-ONCOLOGY
LA English
DT Article
DE Diffuse intrinsic pontine glioma (DIPG); Radiotherapy; Radiosensitizer;
   Gemcitabine
ID ONCOLOGY-GROUP-REPORT; CLINICAL-TRIALS; DOSE-ESCALATION; RADIATION;
   LEUKEMIA; STEROIDS; THERAPY
AB The purpose of this phase I/II, open-label, single-arm trial is to investigate the safety, tolerability, maximum tolerated dose and preliminary efficacy of the potential radiosensitizer gemcitabine, administered concomitantly to radiotherapy, in children with newly diagnosed diffuse intrinsic pontine glioma (DIPG). Six doses of weekly gemcitabine were administered intravenously, concomitantly to 6 weeks of hyperfractionated radiotherapy. Successive cohorts received increasing doses of 140, 175 and 200 mg/m(2) gemcitabine, respectively, following a 3 + 3 dose-escalation schedule without expansion cohort. Dose-limiting toxicities (DLT) were monitored during treatment period. Clinical response was assessed using predefined case report forms and radiological response was assessed using the modified RANO criteria. Quality of life (QoL) was assessed using PedsQL questionnaires. Between June 2012 and December 2016, nine patients were enrolled. Treatment was well tolerated, and no DLTs were observed up to the maximum dose of 200 mg/m(2). All patients experienced reduction of tumor-related symptoms. QoL tended to improve during treatment. PFS and MOS were 4.8 months (95% CI 4.0-5.7) and 8.7 months (95% CI 7.0-10.4). Classifying patients according to the recently developed DIPG survival prediction model, intermediate risk patients (n = 4), showed a PFS and MOS of 6.4 and 12.4 months, respectively, versus a PFS and MOS of 4.5 and 8.1 months, respectively, in high risk patient (n = 5). Gemcitabine up to 200 mg/m(2)/once weekly, added to radiotherapy, is safe and well tolerated in children with newly diagnosed DIPG. PFS and MOS were not significantly different from literature.
C1 [van Zanten, Sophie E. M. Veldhuijzen; El-Khouly, Fatma E.; Jansen, Marc H. A.; van Vuurden, Dannis G.; Kaspers, Gertjan J. L.] Vrije Univ Amsterdam Med Ctr, Dept Pediat Oncol Hematol, Amsterdam, Netherlands.
   [El-Khouly, Fatma E.] Vrije Univ Amsterdam Med Ctr, Dept Clin Pharmacol & Pharm, Amsterdam, Netherlands.
   [Bakker, Dewi P.; Wolf, Nicole I.] Vrije Univ Amsterdam Med Ctr, Dept Child Neurol, Amsterdam, Netherlands.
   [Aliaga, Esther Sanchez] Vrije Univ Amsterdam Med Ctr, Dept Radiol & Nucl Med, Amsterdam, Netherlands.
   [Haasbeek, Cornelis J. A.] Vrije Univ Amsterdam Med Ctr, Dept Radiotherapy, Amsterdam, Netherlands.
   [Wolf, Nicole I.] Neurosci Amsterdam, Amsterdam, Netherlands.
   [Zwaan, C. Michel] Sophia Childrens Univ Hosp, Dept Pediat Oncol Hematol, Erasmus MC, Rotterdam, Netherlands.
   [Vandertop, W. Peter] Vrije Univ Amsterdam Med Ctr, Neurosurg Ctr Amsterdam, Amsterdam, Netherlands.
   [Vandertop, W. Peter] Acad Med Ctr, Neurosurg Ctr Amsterdam, Amsterdam, Netherlands.
   [Kaspers, Gertjan J. L.] Princess Maxima Ctr Pediat Oncol, Utrecht, Netherlands.
   [van Vuurden, Dannis G.] Vrije Univ Amsterdam Med Ctr, Div Hematol Oncol, Dept Pediat, De Boelelaan 1117,Room 9D36, NL-1081 HV Amsterdam, Netherlands.
C3 Vrije Universiteit Amsterdam; VU UNIVERSITY MEDICAL CENTER; Vrije
   Universiteit Amsterdam; VU UNIVERSITY MEDICAL CENTER; Vrije Universiteit
   Amsterdam; VU UNIVERSITY MEDICAL CENTER; Vrije Universiteit Amsterdam;
   VU UNIVERSITY MEDICAL CENTER; Vrije Universiteit Amsterdam; VU
   UNIVERSITY MEDICAL CENTER; Erasmus University Rotterdam; Erasmus MC;
   Erasmus MC - Sophia Children's Hospital; Vrije Universiteit Amsterdam;
   VU UNIVERSITY MEDICAL CENTER; University of Amsterdam; Academic Medical
   Center Amsterdam; Vrije Universiteit Amsterdam; VU UNIVERSITY MEDICAL
   CENTER
RP van Vuurden, DG (通讯作者)，Vrije Univ Amsterdam Med Ctr, Dept Pediat Oncol Hematol, Amsterdam, Netherlands.; van Vuurden, DG (通讯作者)，Vrije Univ Amsterdam Med Ctr, Div Hematol Oncol, Dept Pediat, De Boelelaan 1117,Room 9D36, NL-1081 HV Amsterdam, Netherlands.
EM
RI van Vuurden, Dannis/ACA-6952-2022; Wolf, Nicole I/H-3943-2014; Kaspers,
   Gertjan/ABC-5334-2020; Vandertop, William P./K-9288-2018
OI Wolf, Nicole I/0000-0003-1721-0728; Kaspers,
   Gertjan/0000-0001-7716-8475; Vandertop, William/0000-0001-5417-0265;
   El-Khouly, Fatma E./0000-0003-1451-8516; Veldhuijzen van Zanten, Sophie
   E.M./0000-0002-3888-0977
FU Semmy Foundation (Stichting Semmy)
FX DIPG research in VU University Medical Center is financially supported
   by the Semmy Foundation (Stichting Semmy). We would like to thank Drs.
   Lindsay M.H. Steur (Department of Pediatric Oncology-Hematology, VU
   University Medical Center, Amsterdam) for her support in the statistical
   analysis of the quality of life data.
CR Angiolillo AL, 2006, PEDIATR BLOOD CANCER, V46, P193, DOI 10.1002/pbc.20419
   [Anonymous], 2010, COMM TERM CRIT ADV E
   Bergman AM, 2001, EUR J PHARMACOL, V416, P19, DOI 10.1016/S0014-2999(01)00858-5
   Cole PD, 2009, J CLIN ONCOL, V27, P1456, DOI 10.1200/JCO.2008.20.3778
   EPSTEIN F, 1985, CANCER, V56, P1804, DOI 10.1002/1097-0142(19851001)56:7+<1804::AID-CNCR2820561316>3.0.CO;2-5
   Fabi A, 2008, J NEURO-ONCOL, V87, P79, DOI 10.1007/s11060-007-9489-x
   Gassler N, 2005, BRIT J CANCER, V92, P1084, DOI 10.1038/sj.bjc.6602453
   Hargrove D, 2006, LANCET ONCOL, V7, P241, DOI 10.1016/S1470-2045(06)70615-5
   Jansen MHA, 2012, CANCER TREAT REV, V38, P27, DOI 10.1016/j.ctrv.2011.06.007
   Jansen MH, 2015, NEURO-ONCOLOGY, V17, P160, DOI 10.1093/neuonc/nou104
   Jaspan T, 2016, AM J NEURORADIOL, V37, P1581, DOI 10.3174/ajnr.A4782
   Kim MM, 2016, INT J RADIAT ONCOL, V94, P305, DOI 10.1016/j.ijrobp.2015.10.032
   LANGMOEN IA, 1991, CHILD NERV SYST, V7, P13, DOI 10.1007/BF00263825
   Le Tourneau C, 2009, JNCI-J NATL CANCER I, V101, P708, DOI 10.1093/jnci/djp079
   Mandrell BN, 2016, J NEURO-ONCOL, V129, P373, DOI 10.1007/s11060-016-2187-9
   Metro G, 2010, CANCER CHEMOTH PHARM, V65, P391, DOI 10.1007/s00280-009-1155-x
   Pauwels B, 2005, ONCOLOGIST, V10, P34, DOI 10.1634/theoncologist.10-1-34
   Sigmond J, 2008, GEMCITABINE UPTAKE G, DOI [10.1093/annonc/mdn543, DOI 10.1093/ANNONC/MDN543]
   Steinherz PG, 2002, LEUKEMIA LYMPHOMA, V43, P1945, DOI 10.1080/1042819021000015880
   van Vulpen M, 2002, ONCOL REP, V9, P683
   van Zanten SEMV, 2017, J NEURO-ONCOL, V134, P231, DOI 10.1007/s11060-017-2514-9
   van Zanten SEMV, 2016, J NEURO-ONCOL, V128, P387, DOI 10.1007/s11060-016-2141-x
   van Zanten SEMV, 2015, EXPERT REV ANTICANC, V15, P157, DOI 10.1586/14737140.2015.974563
   Varni JW, 2016, PEDIAT QUALITY LIFE
   Varni JW, 2016, PEDSQL MEASUREMENT M
   Wagner S, 2006, J NEURO-ONCOL, V79, P281, DOI 10.1007/s11060-006-9133-1
   Wagner-Bohn A, 2006, ANTI-CANCER DRUG, V17, P859, DOI 10.1097/01.cad.0000217426.82702.74
NR 27
TC 18
Z9 19
U1 0
U2 2
PU SPRINGER
PI NEW YORK
PA 233 SPRING ST, NEW YORK, NY 10013 USA
SN 0167-594X
EI 1573-7373
J9 J NEURO-ONCOL
JI J. Neuro-Oncol.
PD NOV
PY 2017
VL 135
IS 2
BP 307
EP 315
DI 10.1007/s11060-017-2575-9
PG 9
WC Oncology; Clinical Neurology
WE Science Citation Index Expanded (SCI-EXPANDED)
SC Oncology; Neurosciences & Neurology
GA FL4PH
UT WOS:000414212300010
PM 28748343
OA Green Published
DA 2023-08-27
ER

PT J
AU Di Nardo, W
   Fetoni, A
   Buldrini, S
   Di Girolamo, S
AF Di Nardo, W
   Fetoni, A
   Buldrini, S
   Di Girolamo, S
TI Auditory brainstem and cochlear implants: functional results obtained
   after one year of rehabilitation
SO EUROPEAN ARCHIVES OF OTO-RHINO-LARYNGOLOGY
LA English
DT Article
DE auditory brainstem implant; cochlear implant; speech perception
ID STEM IMPLANT; RESTORATION; HEARING
AB Very little information has been published on the clinical outcome of auditory brainstem implants (ABI). The present paper evaluates results obtained in a patient affected by a bilateral acoustic neuroma in type II neurofibromatosis who received an implant during removal of the residual tumor. One year later surgical revision of the ABI was necessary because no auditory sensation was obtained after ABI activation. Twelve months after the surgical revision, 12 electrodes out of 15 evoked auditory sensation. The results of rehabilitation were compared with those obtained in a group of eight postlingually deaf patients with cochlear implants (CI), Twelve months post operatively the CI patients identified 97.7 +/- 5.1% of bisyllabic words in a closed set while the ABI patient identified 86%. CI patients recognized 87.1 +/- 11.3% of sentences and 81.3 +/- 14.8% of words with contextual cues while the ABI patient recognized 75% and 65% respectively. Speech recognition improved more slowly in the ABI patient than in the CI patients and his scores for open-set words and sentences without lip reading and contextual cues were poorer. Although the results obtained in the ABT patient were not as goad as those obtained in the CI patients, the ABT patient said his quality of life was improved.
C1 Univ Cattolica Sacro Cuore, Policlin A Gemelli, Ist Clin Otorinolaringoiatr, I-00168 Rome, Italy.
C3 Catholic University of the Sacred Heart; IRCCS Policlinico Gemelli
RP Di Nardo, W (通讯作者)，Univ Cattolica Sacro Cuore, Policlin A Gemelli, Ist Clin Otorinolaringoiatr, Largo Agostino Gemelli 8, I-00168 Rome, Italy.
EM
RI Di Girolamo, Stefano/F-7820-2011; di girolamo, stefano/C-9518-2013
OI di girolamo, stefano/0000-0002-6419-9704
CR AMIGONI E, 1997, IMPIANTO COCLEARE TE
   BRACKMANN DE, 1993, OTOLARYNG HEAD NECK, V108, P624, DOI 10.1177/019459989310800602
   Clark GM, 1999, AM J OTOL, V20, P4
   Geier L, 1999, ANN OTO RHINOL LARYN, V108, P80, DOI 10.1177/00034894991080S416
   LASZIG R, 1995, J LARYNGOL OTOL, V109, P385, DOI 10.1017/S0022215100130257
   Laszig R, 1999, Ear Nose Throat J, V78, P110
   LASZIG R, 1991, EUR ARCH OTO-RHINO-L, V248, P420, DOI 10.1007/BF01463568
   MOORE JK, 1987, HEARING RES, V29, P1, DOI 10.1016/0378-5955(87)90202-4
   Otto SR, 1998, OTOLARYNG HEAD NECK, V118, P291, DOI 10.1016/S0194-5998(98)70304-3
   Otto SR, 1997, ADV OTO-RHINO-LARYNG, V52, P1
   ROSSI M, 1995, MAT LINGUISTICO ED P
   Shin YJ, 1998, AM J OTOL, V19, P649
   STRUTZ J, 1995, 5 INT S COCHL IMPL S, P68
   Waltzman SB, 1999, AM J OTOL, V20, P435
NR 14
TC 11
Z9 11
U1 0
U2 4
PU SPRINGER
PI NEW YORK
PA ONE NEW YORK PLAZA, SUITE 4600, NEW YORK, NY, UNITED STATES
SN 0937-4477
EI 1434-4726
J9 EUR ARCH OTO-RHINO-L
JI Eur. Arch. Oto-Rhino-Laryn.
PD JAN
PY 2001
VL 258
IS 1
BP 5
EP 8
DI 10.1007/PL00007518
PG 4
WC Otorhinolaryngology
WE Science Citation Index Expanded (SCI-EXPANDED)
SC Otorhinolaryngology
GA 425GN
UT WOS:000168283000002
PM 11271437
DA 2023-08-27
ER

PT S
AU Lu, VM
   Daniels, DJ
AF Lu, Victor M.
   Daniels, David J.
BE Fang, D
   Han, J
TI Epigenetic-Targeted Treatments for H3K27M-Mutant Midline Gliomas
SO HISTONE MUTATIONS AND CANCER
SE Advances in Experimental Medicine and Biology
LA English
DT Article; Book Chapter
DE Diffuse intrinsic pontine glioma; DIPG; H3; K27M; DMG; Trimethylation;
   Phosphorylation; Acetylation; Demethylase; HDAC; BET
ID DEACETYLASE INHIBITOR PANOBINOSTAT; PHASE-I TRIAL; HISTONE H3;
   GENE-EXPRESSION; BRAIN-TUMORS; SOLID TUMORS; EZH2; METHYLATION;
   POLYCOMB; MUTATION
AB Diffuse intrinsic pontine glioma (DIPG) is a lethal midline brainstem tumor that most commonly occurs in children and is genetically defined by substitution of methionine for lysine at site 27 of histone 3 (H3K27M) in the majority of cases. This mutation has since been shown to exert an influence on the post-translational epigenetic landscape of this disease, with the loss of trimethylation at lysine 27 (H3K27me3) the most common alteration. Based on these findings, a number of drugs targeting these epigenetic changes have been proposed, specifically that alter histone trimethylation, acetylation, or phosphorylation. Various mechanisms have been explored, including inhibition of H327 demethylase and methyltransferase to target trimethylation, inhibition of histone deacetylase (HDAC) and bromodomain and extraterminal (BET) to target acetylation, and inhibition of phosphatase-related enzymes to target phosphorylation. This chapter reviews the current rationales and progress made to date in epigenetically targeting DIPG via these mechanisms.
C1 [Lu, Victor M.; Daniels, David J.] Mayo Clin, Dept Neurosurg, Rochester, MN 55905 USA.
C3 Mayo Clinic
RP Daniels, DJ (通讯作者)，Mayo Clin, Dept Neurosurg, Rochester, MN 55905 USA.
EM
CR Agger K, 2007, NATURE, V449, P731, DOI 10.1038/nature06145
   Alimova I, 2012, INT J CANCER, V131, P1800, DOI 10.1002/ijc.27455
   Bachmann IM, 2006, J CLIN ONCOL, V24, P268, DOI 10.1200/JCO.2005.01.5180
   Bender S, 2013, CANCER CELL, V24, P660, DOI 10.1016/j.ccr.2013.10.006
   Brown ZZ, 2014, J AM CHEM SOC, V136, P13498, DOI 10.1021/ja5060934
   Buczkowicz P, 2014, ACTA NEUROPATHOL, V128, P573, DOI 10.1007/s00401-014-1319-6
   Cao R, 2002, SCIENCE, V298, P1039, DOI 10.1126/science.1076997
   Castel D, 2018, ACTA NEUROPATHOL COM, V6, DOI 10.1186/s40478-018-0614-1
   Castel D, 2015, ACTA NEUROPATHOL, V130, P815, DOI 10.1007/s00401-015-1478-0
   Chan KM, 2013, GENE DEV, V27, P985, DOI 10.1101/gad.217778.113
   Churchman LS, 2011, NATURE, V469, P368, DOI 10.1038/nature09652
   Cordero FJ, 2017, MOL CANCER RES, V15, P1243, DOI 10.1158/1541-7786.MCR-16-0389
   de Blank PM, 2015, CANCER MED-US, V4, P608, DOI 10.1002/cam4.410
   De Souza C, 2015, RECENT PAT ANTI-CANC, V10, P145
   El-Khouly FE, 2017, FRONT ONCOL, V7, DOI 10.3389/fonc.2017.00254
   Ellis L, 2008, CLIN CANCER RES, V14, P4500, DOI 10.1158/1078-0432.CCR-07-4262
   Fang D, 2018, ELIFE, V7, DOI 10.7554/eLife.36696
   Figueroa JM, 2017, NEURO-ONCOLOGY, V19, P1494, DOI 10.1093/neuonc/nox085
   Filippakopoulos P, 2014, NAT REV DRUG DISCOV, V13, P339, DOI 10.1038/nrd4286
   Funato K, 2014, SCIENCE, V346, P1529, DOI 10.1126/science.1253799
   Furey TS, 2013, SCIENCE, V342, P705, DOI 10.1126/science.1246755
   Gehani SS, 2010, MOL CELL, V39, P886, DOI 10.1016/j.molcel.2010.08.020
   Gezer U, 2015, INT J MOL SCI, V16, P29654, DOI 10.3390/ijms161226180
   Grasso CS, 2015, NAT MED, V21, P555, DOI 10.1038/nm.3855
   Hake SB, 2005, P NATL ACAD SCI USA, V102, P6344, DOI 10.1073/pnas.0502413102
   Hashizume R, 2017, NEUROL MED-CHIR, V57, P331, DOI 10.2176/nmc.ra.2017-0018
   Hashizume R, 2014, NAT MED, V20, P1394, DOI 10.1038/nm.3716
   Hennika T, 2017, PLOS ONE, V12, DOI 10.1371/journal.pone.0169485
   Herz HM, 2014, SCIENCE, V345, P1065, DOI 10.1126/science.1255104
   Hodges C, 2009, SCIENCE, V325, P626, DOI 10.1126/science.1172926
   Kaelin WG, 2013, CELL, V153, P56, DOI 10.1016/j.cell.2013.03.004
   Knutson SK, 2014, MOL CANCER THER, V13, P842, DOI 10.1158/1535-7163.MCT-13-0773
   Krug B, 2019, CANCER CELL, V35, P782, DOI 10.1016/j.ccell.2019.04.004
   Kruidenier L, 2012, NATURE, V488, P404, DOI 10.1038/nature11262
   Lewis PW, 2013, SCIENCE, V340, P857, DOI 10.1126/science.1232245
   Li YX, 2016, CSH PERSPECT MED, V6, DOI 10.1101/cshperspect.a026831
   Louis DN, 2016, ACTA NEUROPATHOL, V131, P803, DOI 10.1007/s00401-016-1545-1
   Lu VM, 2019, BIOMARK MED, V13, P597, DOI 10.2217/bmm-2019-0139
   Lu VM, 2019, J NEUROSURG-PEDIATR, V23, P308, DOI 10.3171/2018.9.PEDS18419
   Lund AH, 2004, CURR OPIN CELL BIOL, V16, P239, DOI 10.1016/j.ceb.2004.03.010
   MARUSHIGE K, 1976, P NATL ACAD SCI USA, V73, P3937, DOI 10.1073/pnas.73.11.3937
   Maury E, 2017, EPIGENETICS-US, V12, P353, DOI 10.1080/15592294.2016.1278095
   Merchant TE, 2010, SEMIN RADIAT ONCOL, V20, P58, DOI 10.1016/j.semradonc.2009.09.005
   Mohammad F, 2017, NAT MED, V23, P483, DOI 10.1038/nm.4293
   Moreno L, 2015, CLIN CANCER RES, V21, P267, DOI 10.1158/1078-0432.CCR-14-1592
   Nowak SJ, 2003, MOL CELL BIOL, V23, P6129, DOI 10.1128/MCB.23.17.6129-6138.2003
   Ntziachristos P, 2014, NATURE, V514, P513, DOI 10.1038/nature13605
   Pathak P, 2015, J NEURO-ONCOL, V121, P489, DOI 10.1007/s11060-014-1675-z
   Piunti A, 2017, NAT MED, V23, P493, DOI 10.1038/nm.4296
   Qi J, 2014, CSH PERSPECT BIOL, V6, DOI 10.1101/cshperspect.a018663
   Richardson P, 2017, EXPERT REV HEMATOL, V10, P933, DOI 10.1080/17474086.2017.1369399
   Ruthenburg AJ, 2007, MOL CELL, V25, P15, DOI 10.1016/j.molcel.2006.12.014
   Schramm K, 2019, NEURO-ONCOLOGY, DOI [10.1093/neuonc/noz057, DOI 10.1093/NEUONC/N0Z057]
   Seet BT, 2006, NAT REV MOL CELL BIO, V7, P473, DOI 10.1038/nrm1960
   Silveira AB, 2019, ACTA NEUROPATHOL, V137, P637, DOI 10.1007/s00401-019-01975-4
   Suva ML, 2009, CANCER RES, V69, P9211, DOI 10.1158/0008-5472.CAN-09-1622
   Syren P, 2017, SCAND J GASTROENTERO, V52, P668, DOI 10.1080/00365521.2017.1301989
   Thomas S, 2016, ANN ONCOL, V27, P947, DOI 10.1093/annonc/mdw044
   van Zanten SEMV, 2017, J NEURO-ONCOL, V132, P255, DOI 10.1007/s11060-016-2363-y
   Vanan MI, 2015, FRONT ONCOL, V5, DOI 10.3389/fonc.2015.00237
   Venneti S, 2013, BRAIN PATHOL, V23, P558, DOI 10.1111/bpa.12042
   Verma SK, 2012, ACS MED CHEM LETT, V3, P1091, DOI 10.1021/ml3003346
   Wadhwa E, 2016, CUREUS J MED SCIENCE, V8, DOI 10.7759/cureus.620
   Wang Z, 2015, BIOMED REP, V3, P543, DOI 10.3892/br.2015.462
   Wiese M, 2016, KLIN PADIATR, V228, P113, DOI 10.1055/s-0042-105292
   Wu G, 2012, NAT GENET, V44, P251, DOI 10.1038/ng.1102
   Yang XJ, 2008, MOL CELL, V31, P449, DOI 10.1016/j.molcel.2008.07.002
   Zhang YQ, 2017, CELL BIOSCI, V7, DOI 10.1186/s13578-017-0184-0
NR 68
TC 3
Z9 3
U1 0
U2 4
PU SPRINGER-VERLAG SINGAPORE PTE LTD
PI SINGAPORE
PA 152 BEACH ROAD, #21-01/04 GATEWAY EAST, SINGAPORE, 189721, SINGAPORE
SN 0065-2598
EI 2214-8019
BN 978-981-15-8104-5; 978-981-15-8103-8
J9 ADV EXP MED BIOL
JI Adv.Exp.Med.Biol.
PY 2021
VL 1283
BP 73
EP 84
DI 10.1007/978-981-15-8104-5_6
D2 10.1007/978-981-15-8104-5
PG 12
WC Biochemistry & Molecular Biology; Oncology
WE Book Citation Index– Science (BKCI-S); Science Citation Index Expanded (SCI-EXPANDED)
SC Biochemistry & Molecular Biology; Oncology
GA BQ5AC
UT WOS:000595857500006
PM 33155139
DA 2023-08-27
ER

PT J
AU Nadvi, SS
   Ebrahim, FS
   Corr, P
AF Nadvi, SS
   Ebrahim, FS
   Corr, P
TI The value of (201)thallium-SPECT imaging in childhood brainstem gliomas
SO PEDIATRIC RADIOLOGY
LA English
DT Article
ID EMISSION COMPUTED-TOMOGRAPHY; TL-201 SPECT; CEREBRAL-TUMORS; DIAGNOSIS;
   GRADE
AB Objective. To compare (201)thallium (T1) uptake and SPECT with MRI in children with brainstem gliomas,
   Materials and methods. Ten children with brainstem gliomas were prospectively evaluated by Tl-201-SPECT and MRI, Histological verification was obtained in eight children - two died prior to surgery, Quantitative thallium uptake index (UI) was obtainable in five cases and was compared to tumour grade. In addition, two patients with known benign brainstem lesions (neurofibromatosis and tuberculoma) were similarly prospectively evaluated.
   Results, All children with brainstem glioma accumulated thallium, (Mean U1 3.23, 100% sensitivity), The single patient with brainstem tuberculoma also accumulated thallium (UI 2.80, 91.7% specificity). There was no correlation between thallium uptake and tumour grade. Uptake could not be conclusively correlated with the following MR features: gadolinium enhancement, exophytic or intrinsic gliomas, necrosis and location of glioma within the brainstem.
   Conclusions. Tl-201-SPECT is a promising imaging adjunct in the assessment of childhood brainstem gliomas.
C1 Univ Natal, Fac Med, Wentworth Hosp, Dept Neurosurg, ZA-4026 Durban, South Africa.
   Univ Natal, Fac Med, Addington & Wentworth Hosp, Dept Radiol, Durban, South Africa.
C3 University of Kwazulu Natal; University of Kwazulu Natal
RP Nadvi, SS (通讯作者)，Univ Natal, Fac Med, Wentworth Hosp, Dept Neurosurg, Private Bag Jacobs, ZA-4026 Durban, South Africa.
RI Corr, Peter/AEZ-9176-2022; CORR, PETER/A-5156-2010
CR ABDELDAYEM H, 1994, NUCL MED CLIN DIAGNO, P727
   ATKINS HL, 1977, J NUCL MED, V18, P133
   BARKOVICH AJ, 1991, PEDIATR NEUROSURG, V16, P73, DOI 10.1159/000120511
   Barzen G., 1992, Strahlentherapie und Onkologie, V168, P732
   BATIER HL, 1985, MAYO CLIN P, V60, P75
   BIERSACK HJ, 1991, SEMIN NUCL MED, V21, P2, DOI 10.1016/S0001-2998(05)80075-3
   BLACK KL, 1994, NEUROL RES, V16, P93
   BLACK KL, 1989, J NEUROSURG, V71, P342, DOI 10.3171/jns.1989.71.3.0342
   BRISMAR T, 1989, BRAIN RES, V500, P30, DOI 10.1016/0006-8993(89)90296-5
   BROOKS DJ, 1986, SEMIN ONCOL, V13, P83
   DUFFNER PJ, 1986, CANCER, V15, P561
   FINLAY JL, 1987, AM J PEDIAT HEMATOL, V9, P246
   HOLMAN BL, 1995, J NEUROIMAGING, V5, pS34
   JOHNSON PC, 1989, RADIOLOGY, V170, P211, DOI 10.1148/radiology.170.1.2535765
   KAHN D, 1994, AM J ROENTGENOL, V163, P1459, DOI 10.2214/ajr.163.6.7992747
   KAPLAN WD, 1987, J NUCL MED, V28, P47
   KERNOHAN JW, 1949, P STAFF M MAYO CLIN, V24, P71
   KIM KT, 1990, J NUCL MED, V31, P965
   KOSUDA S, 1994, INT J RADIAT ONCOL, V30, P1201, DOI 10.1016/0360-3016(94)90329-8
   Leeds Norman E., 1994, Current Opinion in Oncology, V6, P254, DOI 10.1097/00001622-199405000-00006
   MARIA BL, 1994, PEDIATR NEUROSURG, V20, P11, DOI 10.1159/000120760
   MOUNTZ JM, 1988, J NEUROSURG, V68, P705, DOI 10.3171/jns.1988.68.5.0705
   NADEL HR, 1993, SEMIN NUCL MED, V23, P243, DOI 10.1016/S0001-2998(05)80105-9
   OMALLEY JP, 1994, AM J ROENTGENOL, V163, P417, DOI 10.2214/ajr.163.2.8037042
   OTUAMA LA, 1991, PEDIATR NEUROL, V7, P249, DOI 10.1016/0887-8994(91)90040-R
   RAJSHEKHAR V, 1995, J NEUROSURG, V82, P976, DOI 10.3171/jns.1995.82.6.0976
   SJOHOLM H, 1995, ACTA NEUROL SCAND, V91, P66
   TONAMI N, 1990, CLIN NUCL MED, V15, P397, DOI 10.1097/00003072-199006000-00007
   UEDA T, 1993, EUR J NUCL MED, V20, P138
NR 29
TC 6
Z9 6
U1 0
U2 1
PU SPRINGER VERLAG
PI NEW YORK
PA 175 FIFTH AVE, NEW YORK, NY 10010 USA
SN 0301-0449
J9 PEDIATR RADIOL
JI Pediatr. Radiol.
PD AUG
PY 1998
VL 28
IS 8
BP 575
EP 579
DI 10.1007/s002470050418
PG 5
WC Pediatrics; Radiology, Nuclear Medicine & Medical Imaging
WE Science Citation Index Expanded (SCI-EXPANDED)
SC Pediatrics; Radiology, Nuclear Medicine & Medical Imaging
GA 119CM
UT WOS:000075877800004
PM 9716624
DA 2023-08-27
ER

PT J
AU Guillamo, JS
   Creange, A
   Kalifa, C
   Grill, J
   Rodriguez, D
   Doz, F
   Barbarot, S
   Zerah, M
   Sanson, M
   Bastuji-Garin, S
   Wolkenstein, P
AF Guillamo, JS
   Creange, A
   Kalifa, C
   Grill, J
   Rodriguez, D
   Doz, F
   Barbarot, S
   Zerah, M
   Sanson, M
   Bastuji-Garin, S
   Wolkenstein, P
CA Reseau NF France
TI Prognostic factors of CNS tumours in Neurofibromatosis 1 (NF1)
SO BRAIN
LA English
DT Article
DE brainstem glioma; Neurorbromatosis 1; optic pathway glioma
ID OPTIC PATHWAY TUMORS; BRAIN-STEM GLIOMAS; VONRECKLINGHAUSEN
   NEUROFIBROMATOSIS; TYPE-1 NEUROFIBROMATOSIS; CHILDREN; MANAGEMENT;
   CARBOPLATIN; LESIONS; MRI
AB In addition to multiple peripheral neurofibromas, Neurofibromatosis 1 (NF1) predisposes to CNS tumours. Most of them are pilocytic astrocytomas, arise in children and are located in the optic pathways or in the brainstem. The majority are indolent, but factors predictive of poor prognosis have yet to be identified. Furthermore, the incidence and natural history of gliomas of a higher grade, arising in adults or involving other locations are largely unknown in NF1. In order to address these issues, we performed a retrospective study of 104 patients followed in seven French centres between 1982 and 2000. Inclusion criteria were a diagnosis of NF1, according to the National Institutes of Health criteria, and the presence of a CNS tumour, regardless of type, location or age of onset. The series included 88 children (age range 3 months to 17 years) and 16 adults (age range 19-52 years). The median follow-up was 5.6 years. One hundred and twenty-seven CNS tumours were observed in the 104 patients. Eighty-four (66%) were optic pathway tumours (OPT) and 43 (34%) extra-optic pathway tumours (extra-OPT) (brainstem: n=21; other locations: n=22). Twenty-one patients (20%) had multiple CNS tumours. OPT were symptomatic in 50 patients and extra-OPT in 19. Main clinical findings at presentation included visual loss (n=29; 58%) and precocious puberty (n=5; 10%) for OPT, increased intracranial pressure (n=9; 48%) for extra-OPT. Fourteen out of the 27 symptomatic tumours with histology were pilocytic astrocytomas. The overall survival rate was 90% at 5 years (95% confidence interval 82-95%). Extra-optic location, tumour diagnosis in adulthood and symptomatic tumours were independent factors associated with shorter survival time (P<0.05, Cox model). Radiotherapy for OPT was associated with vascular complications (ischaemic strokes) and growth hormone deficiency in 32 and 46% of patients, respectively. In conclusion, mortality is high in extra-OPT, particularly in adults, whereas OPT are only exceptionally life-threatening. Radiotherapy of OPT is associated with an important morbidity in NF1.
C1 Hop Henri Mondor, Serv Neurol, F-94010 Creteil, France.
   Hop Henri Mondor, INSERM, Unite 421, Fac Med, F-94010 Creteil, France.
   Hop Henri Mondor, Serv St Publ, F-94010 Creteil, France.
   Hop Henri Mondor, Serv Dermatol, F-94010 Creteil, France.
   Inst Gustave Roussy, Serv Oncol Pediat, Villejuif, France.
   Hop St Vincent De Paul, Serv Neurol Pediat, Paris, France.
   Inst Curie, Serv Oncol Pediat, Paris, France.
   Hop Necker Enfants Malad, Serv Neurochirurg Pediat, Paris, France.
   Grp Hosp Pitie Salpetriere, Serv Neurol, F-75634 Paris, France.
   Hop Hotel Dieu, Serv Dermatol, Nantes, France.
C3 Assistance Publique Hopitaux Paris (APHP); Universite
   Paris-Est-Creteil-Val-de-Marne (UPEC); Hopital Universitaire
   Henri-Mondor - APHP; Assistance Publique Hopitaux Paris (APHP);
   Universite Paris-Est-Creteil-Val-de-Marne (UPEC); Hopital Universitaire
   Henri-Mondor - APHP; Institut National de la Sante et de la Recherche
   Medicale (Inserm); Assistance Publique Hopitaux Paris (APHP); Universite
   Paris-Est-Creteil-Val-de-Marne (UPEC); Hopital Universitaire
   Henri-Mondor - APHP; Assistance Publique Hopitaux Paris (APHP);
   Universite Paris-Est-Creteil-Val-de-Marne (UPEC); Hopital Universitaire
   Henri-Mondor - APHP; UNICANCER; Gustave Roussy; UDICE-French Research
   Universities; Universite Paris Cite; Assistance Publique Hopitaux Paris
   (APHP); Hopital Universitaire Cochin - APHP; UDICE-French Research
   Universities; Universite PSL; UNICANCER; Institut Curie; UDICE-French
   Research Universities; Universite Paris Cite; Assistance Publique
   Hopitaux Paris (APHP); Hopital Universitaire Cochin - APHP; Hopital
   Universitaire Necker-Enfants Malades - APHP; Assistance Publique
   Hopitaux Paris (APHP); Hopital Universitaire Pitie-Salpetriere - APHP;
   UDICE-French Research Universities; Sorbonne Universite; Nantes
   Universite; CHU de Nantes
RP Creange, A (通讯作者)，Hop Henri Mondor, Serv Neurol, 51 Ave Marechal Lattre Tassigny, F-94010 Creteil, France.
EM
RI Bastuji-Garin, Sylvie/R-3479-2018; Guillamo,
   Jean-Sebastien/AAC-9525-2021; Grill, Jacques/R-8093-2019
OI Bastuji-Garin, Sylvie/0000-0001-9855-5183; Guillamo,
   Jean-Sebastien/0000-0003-0165-8333; Doz, Francois/0000-0001-9286-4831;
   Barbarot, Sebastien/0000-0002-6629-9100
CR [Anonymous], 1988, ARCH NEUROL-CHICAGO, V45, P575, DOI 10.1097/00005537-199606000-00007
   [Anonymous], 2000, WORLD HLTH ORG CLASS
   AOKI S, 1989, RADIOLOGY, V172, P527, DOI 10.1148/radiology.172.2.2501822
   BATAINI JP, 1991, INT J RADIAT ONCOL, V21, P615, DOI 10.1016/0360-3016(91)90678-W
   Bilaniuk LT, 1997, NEURORADIOLOGY, V39, P642, DOI 10.1007/s002340050484
   Bonawitz C, 1998, AM J NEURORADIOL, V19, P541
   COX DR, 1972, J R STAT SOC B, V34, P187
   Creange A, 1999, BRAIN, V122, P473, DOI 10.1093/brain/122.3.473
   DeBella K, 2000, NEUROLOGY, V54, P1646, DOI 10.1212/WNL.54.8.1646
   Deliganis AV, 1996, NEUROSURGERY, V38, P1114, DOI 10.1097/00006123-199606000-00010
   DiMario FJ, 1998, ARCH NEUROL-CHICAGO, V55, P500, DOI 10.1001/archneur.55.4.500
   ES SV, 1996, PEDIATR RADIOL, V26, P478
   FERNER RE, 1993, J NEUROL NEUROSUR PS, V56, P492, DOI 10.1136/jnnp.56.5.492
   Friedman JM, 1999, AM J MED GENET, V89, P1
   Friedman JM, 1997, AM J MED GENET, V70, P138, DOI 10.1002/(SICI)1096-8628(19970516)70:2<138::AID-AJMG7>3.0.CO;2-U
   Grill J, 1999, ANN NEUROL, V45, P393, DOI 10.1002/1531-8249(199903)45:3<393::AID-ANA17>3.0.CO;2-B
   Grill J, 2000, EUR J PEDIATR, V159, P692, DOI 10.1007/s004310000531
   Guillamo JS, 2001, BRAIN, V124, P2528, DOI 10.1093/brain/124.12.2528
   Gutmann DH, 1997, JAMA-J AM MED ASSOC, V278, P51, DOI 10.1001/jama.278.1.51
   HUSON SM, 1988, BRAIN, V111, P1355, DOI 10.1093/brain/111.6.1355
   JENKIN D, 1993, INT J RADIAT ONCOL, V25, P215, DOI 10.1016/0360-3016(93)90342-S
   KAPLAN EL, 1958, J AM STAT ASSOC, V53, P457, DOI 10.2307/2281868
   Kornreich L, 2001, AM J NEURORADIOL, V22, P1963
   LELLOUCHTUBIANA A, 1995, ACTA NEUROPATHOL, V90, P319
   Leroy K, 2001, ARCH DERMATOL, V137, P908
   LEWIS RA, 1984, OPHTHALMOLOGY, V91, P929
   LISTERNICK R, 1994, J PEDIATR-US, V125, P63, DOI 10.1016/S0022-3476(94)70122-9
   Listernick R, 1997, ANN NEUROL, V41, P143, DOI 10.1002/ana.410410204
   Listernick R, 1999, AM J MED GENET, V89, P38, DOI 10.1002/(SICI)1096-8628(19990326)89:1<38::AID-AJMG8>3.0.CO;2-M
   Listernick R, 1999, J NEURO-ONCOL, V45, P185, DOI 10.1023/A:1006338322266
   LISTERNICK R, 1995, J PEDIATR-US, V127, P718, DOI 10.1016/S0022-3476(95)70159-1
   MATSUI I, 1993, CANCER-AM CANCER SOC, V72, P2746, DOI 10.1002/1097-0142(19931101)72:9<2746::AID-CNCR2820720936>3.0.CO;2-W
   McGaughran JM, 1999, J MED GENET, V36, P197
   MOLLOY PT, 1995, NEUROLOGY, V45, P1897, DOI 10.1212/WNL.45.10.1897
   Packer RJ, 1997, J NEUROSURG, V86, P747, DOI 10.3171/jns.1997.86.5.0747
   Pinson S, 2001, ANN DERMATOL VENER, V128, P567
   Pollack IF, 1996, J NEURO-ONCOL, V28, P257
   Pollack IF, 1996, NEUROLOGY, V46, P1652, DOI 10.1212/WNL.46.6.1652
   RAFFEL C, 1989, NEUROSURGERY, V25, P959, DOI 10.1227/00006123-198912000-00018
   Rasmussen SA, 2000, AM J EPIDEMIOL, V151, P33, DOI 10.1093/oxfordjournals.aje.a010118
   RICCARDI VM, 1991, NEW ENGL J MED, V324, P1283, DOI 10.1056/NEJM199105023241812
   SEIZINGER BR, 1993, NAT GENET, V3, P97, DOI 10.1038/ng0293-97
   SORENSEN SA, 1986, NEW ENGL J MED, V314, P1010, DOI 10.1056/NEJM198604173141603
   STERN J, 1980, ARCH OPHTHALMOL-CHIC, V98, P505
   Tao ML, 1997, INT J RADIAT ONCOL, V39, P579, DOI 10.1016/S0360-3016(97)00359-3
NR 45
TC 187
Z9 192
U1 0
U2 9
PU OXFORD UNIV PRESS
PI OXFORD
PA GREAT CLARENDON ST, OXFORD OX2 6DP, ENGLAND
SN 0006-8950
EI 1460-2156
J9 BRAIN
JI Brain
PD JAN
PY 2003
VL 126
BP 152
EP 160
DI 10.1093/brain/awg016
PN 1
PG 9
WC Clinical Neurology; Neurosciences
WE Science Citation Index Expanded (SCI-EXPANDED)
SC Neurosciences & Neurology
GA 625NF
UT WOS:000179822000011
PM 12477702
OA Bronze
DA 2023-08-27
ER

PT J
AU SHRIEVE, DC
   WARA, WM
   EDWARDS, MSB
   SNEED, PK
   PRADOS, MD
   COGEN, PH
   LARSON, DA
   LEVIN, VA
AF SHRIEVE, DC
   WARA, WM
   EDWARDS, MSB
   SNEED, PK
   PRADOS, MD
   COGEN, PH
   LARSON, DA
   LEVIN, VA
TI HYPERFRACTIONATED RADIATION-THERAPY FOR GLIOMAS OF THE BRAIN-STEM IN
   CHILDREN AND IN ADULTS
SO INTERNATIONAL JOURNAL OF RADIATION ONCOLOGY BIOLOGY PHYSICS
LA English
DT Article
DE BRAIN-STEM GLIOMA; RADIOTHERAPY; HYPERFRACTIONATION; PEDIATRIC BRAIN
   TUMORS
ID RAT SPINAL-CORD; STEM GLIOMAS; PROGNOSTIC FACTORS; MALIGNANT GLIOMAS;
   TUMORS; RADIOTHERAPY; IRRADIATION; FRACTIONATION; EXPERIENCE; MANAGEMENT
AB Between February 1984 and September 1990, 60 patients with brainstem gliomas were treated with hyperfractionated radiotherapy in the Department of Radiation Oncology at the University of California, San Francisco. Forty-one children (less-than-or-equal-to 18 years) and 19 adults were treated with 100 cGy twice daily with 4-8 hr between doses. Thirty-one patients (21 children and 10 adults) received total doses of 66-72 Gy and 29 patients (20 children and nine adults) received 74-78 Gy. Median follow-up was 208 weeks for all patients (214 weeks for children, 157 weeks for adults). Twenty-three patients (14 children and nine adults) were alive at the time of analysis, surviving 59-359 weeks following treatment. Median actuarial survival was 73.6 weeks overall (72 weeks for children, 190 weeks for adults; p = 0.43). Survival at 12 and 24 months was 65% and 38%, respectively (63% and 32%, for children; 68% and 53% for adults). All patients had pretreatment magnetic resonance imaging by which tumors were classified as either focal or diffuse. No significant pretreatment prognostic factors for adults were identified. In children, significant favorable prognostic factors on univariate analysis were older age (p = 0.001), tumor location in thalamus or midbrain (p = 0.002), focal appearance on MRI scan (p < 0.001 ) and duration of symptoms > 2 months prior to treatment (p < 0.001 ). Thirty-five patients had tumor biopsies, leading to a diagnosis in 33 (22 children and 11 adults). Children with moderately anaplastic astrocytomas survived significantly longer than those with glioblastoma multiforme or unbiopsied tumors (p < 0.001). Only duration of symptoms > 2 months remained significant as a favorable prognostic indicator for children on multivariate analysis (p < 0.001). Survival was not significantly different for patients receiving less-than-or-equal-to 72 Gy and those receiving > 72 Gy (p = 0.18). No subgroup of patients showed significantly better survival with the higher dose. These findings indicate that hyperfractionated radiotherapy is effective treatment for adults and a subgroup of better prognosis children with brainstem gliomas. There is a subgroup of pediatric patients with extremely poor prognosis for whom even this aggressive treatment does little to extend survival. We conclude that there is no benefit to increasing total dose above 72 Gy for any of the groups analyzed.
C1 UNIV CALIF SAN FRANCISCO, SCH MED, DEPT NEUROL SURG, SAN FRANCISCO, CA 94143 USA.
C3 University of California System; University of California San Francisco
RP SHRIEVE, DC (通讯作者)，UNIV CALIF SAN FRANCISCO, SCH MED, DEPT RADIAT ONCOL, BOX 0226, SAN FRANCISCO, CA 94143 USA.
RI Prados, Michael/N-5220-2019
CR ALBRIGHT AL, 1986, J NEUROSURG, V65, P751, DOI 10.3171/jns.1986.65.6.0751
   ANG KK, 1985, INT J RADIAT ONCOL, V11, P105, DOI 10.1016/0360-3016(85)90368-2
   BARKOVICH AJ, 1991, PEDIATR NEUROSURG, V16, P73, DOI DOI 10.1159/000120511]
   BERGER MS, 1983, NEUROSURGERY, V12, P298, DOI 10.1227/00006123-198303000-00008
   BLOOM HJG, 1990, INT J RADIAT ONCOL, V18, P723, DOI 10.1016/0360-3016(90)90392-W
   COHEN ME, 1984, BRAIN TUMORS CHILDRE
   COX DR, 1972, J R STAT SOC B, V34, P187
   DAVIS RL, 1984, J NEURO-ONCOL, V2, P267
   EDWARDS MSB, 1987, PEDIATR NEUROSCI, V13, P309, DOI 10.1159/000120349
   EDWARDS MSB, 1989, J NEUROSURG, V70, P691, DOI 10.3171/jns.1989.70.5.0691
   EIFEL PJ, 1987, INT J RADIAT ONCOL, V13, P847, DOI 10.1016/0360-3016(87)90098-8
   EPSTEIN F, 1985, CANCER, V56, P1804, DOI 10.1002/1097-0142(19851001)56:7+<1804::AID-CNCR2820561316>3.0.CO;2-5
   EPSTEIN F, 1988, J NEURO-ONCOL, V6, P309
   FREEMAN CR, 1986, INT J RADIAT ONCOL, V12, P1823, DOI 10.1016/0360-3016(86)90325-1
   FREEMAN CR, 1991, CANCER-AM CANCER SOC, V68, P474, DOI 10.1002/1097-0142(19910801)68:3<474::AID-CNCR2820680305>3.0.CO;2-7
   FREEMAN CR, 1988, INT J RADIAT ONCOL, V15, P311, DOI 10.1016/S0360-3016(98)90010-4
   GREENBERGER JS, 1977, RADIOLOGY, V122, P463, DOI 10.1148/122.2.463
   GRIGSBY PW, 1989, INT J RADIAT ONCOL, V16, P649, DOI 10.1016/0360-3016(89)90480-X
   GRIGSBY PW, 1987, CANCER, V60, P2901, DOI 10.1002/1097-0142(19871215)60:12<2901::AID-CNCR2820601210>3.0.CO;2-W
   HALPERIN EC, 1989, MED PEDIATR ONCOL, V17, P116, DOI 10.1002/mpo.2950170209
   HALPERIN EC, 1985, INT J RADIAT ONCOL, V11, P1293, DOI 10.1016/0360-3016(85)90244-5
   KAPLAN EL, 1958, J AM STAT ASSOC, V53, P457, DOI 10.2307/2281868
   KIM TH, 1980, INT J RADIAT ONCOL, V6, P51, DOI 10.1016/0360-3016(80)90203-5
   LEVIN VA, 1984, NEUROSURGERY, V14, P679, DOI 10.1227/00006123-198406000-00006
   LINSTADT DE, 1991, INT J RADIAT ONCOL, V20, P757, DOI 10.1016/0360-3016(91)90019-Z
   LITTMAN P, 1980, CANCER, V45, P2787, DOI 10.1002/1097-0142(19800601)45:11<2787::AID-CNCR2820451113>3.0.CO;2-V
   MANTEL NATHAN, 1966, CANCERCHEMOTHERAP REP, V50, P163
   PACKER RJ, 1990, ANN NEUROL, V27, P167, DOI 10.1002/ana.410270212
   PACKER RJ, 1987, INT J RADIAT ONCOL, V13, P1647, DOI 10.1016/0360-3016(87)90160-X
   SANFORD RA, 1988, SURG NEUROL, V30, P276, DOI 10.1016/0090-3019(88)90299-6
   SMITH RR, 1990, NEURORADIOLOGY, V32, P265, DOI 10.1007/BF00593044
   STEWART AM, 1973, BRIT J CANCER, V28, P568, DOI 10.1038/bjc.1973.187
   STRANGE P, 1982, ACTA NEUROCHIR, V62, P219, DOI 10.1007/BF01403627
   STROINK AR, 1986, J NEUROSURG, V65, P745, DOI 10.3171/jns.1986.65.6.0745
   THAMES HD, 1982, INT J RADIAT ONCOL, V8, P219, DOI 10.1016/0360-3016(82)90517-X
   VANDERSCHUEREN E, 1988, INT J RADIAT ONCOL, V14, P297, DOI 10.1016/0360-3016(88)90436-1
   WALKER MD, 1979, INT J RADIAT ONCOL, V5, P1725, DOI 10.1016/0360-3016(79)90553-4
   WITHERS HR, 1985, CANCER, V55, P2086, DOI 10.1002/1097-0142(19850501)55:9+<2086::AID-CNCR2820551409>3.0.CO;2-1
NR 38
TC 52
Z9 55
U1 0
U2 0
PU ELSEVIER SCIENCE INC
PI NEW YORK
PA 360 PARK AVE SOUTH, NEW YORK, NY 10010-1710 USA
SN 0360-3016
EI 1879-355X
J9 INT J RADIAT ONCOL
JI Int. J. Radiat. Oncol. Biol. Phys.
PY 1992
VL 24
IS 4
BP 599
EP 610
DI 10.1016/0360-3016(92)90704-L
PG 12
WC Oncology; Radiology, Nuclear Medicine & Medical Imaging
WE Science Citation Index Expanded (SCI-EXPANDED)
SC Oncology; Radiology, Nuclear Medicine & Medical Imaging
GA JY089
UT WOS:A1992JY08900003
PM 1429081
OA Bronze
DA 2023-08-27
ER

PT J
AU Ross, JL
   Vega, JV
   Plant, A
   MacDonald, TJ
   Becher, OJ
   Hambardzumyan, D
AF Ross, James L.
   Vega, Jose Velazquez
   Plant, Ashley
   MacDonald, Tobey J.
   Becher, Oren J.
   Hambardzumyan, Dolores
TI Tumour immune landscape of paediatric high-grade gliomas
SO BRAIN
LA English
DT Review
DE paediatric glioma; diffuse intrinsic pontine glioma; tumour associated
   macrophage; inflammation; histone
ID MALIGNANT GLIOMAS; DENDRITIC CELLS; BRAIN-STEM; PONTINE GLIOMA;
   GLIOBLASTOMA; MANAGEMENT; CHILDREN; REVEALS; TEMOZOLOMIDE; INHIBITION
AB Over the past decade, remarkable progress has been made towards elucidating the origin and genomic landscape of childhood high-grade brain tumours. It has become evident that paediatric high-grade gliomas differ from those in adults with respect to multiple defining aspects including: DNA copy number, gene expression profiles, tumour locations within the CNS and genetic alterations such as somatic histone mutations.
   Despite these advances, clinical trials for children with gliomas have historically been based on ineffective adult regimens that fail to take into consideration the fundamental biological differences between the two. Additionally, although our knowledge of the intrinsic cellular mechanisms driving tumour progression has considerably expanded, little is known about the dynamic tumour immune microenvironment in paediatric high-grade gliomas. In this review, we explore the genetic and epigenetic landscape of these gliomas and how this drives the creation of specific tumour subgroups with meaningful survival outcomes. Further, we provide a comprehensive analysis of the paediatric high-grade glioma tumour immune microenvironment and discuss emerging therapeutic efforts aimed at exploiting the immune functions of these tumours.
C1 [Ross, James L.] Emory Univ, Emory Vaccine Ctr, Dept Microbiol & Immunol, Sch Med, Atlanta, GA 30322 USA.
   [Vega, Jose Velazquez] Emory Univ, Sch Med, Dept Pathol & Lab Med, Atlanta, GA 30322 USA.
   [Plant, Ashley; Becher, Oren J.] Ann & Robert H Lurie Childrens Hosp Chicago, Div Hematol Oncol & Stem Cell Transplant, Chicago, IL 60611 USA.
   [MacDonald, Tobey J.] Emory Univ, Aflac Canc & Blood Disorders Ctr, Dept Pediat, Childrens Healthcare Atlanta,Sch Med, Atlanta, GA 30322 USA.
   [Hambardzumyan, Dolores] Mt Sinai Icahn Sch Med, Tisch Canc Inst, Dept Oncol Sci, New York, NY 10029 USA.
   [Hambardzumyan, Dolores] Mt Sinai Icahn Sch Med, Dept Neurosurg, New York, NY 10029 USA.
C3 Emory University; Emory University; Ann & Robert H. Lurie Children's
   Hospital of Chicago; Children's Healthcare of Atlanta (CHOA); Emory
   University; Icahn School of Medicine at Mount Sinai; Icahn School of
   Medicine at Mount Sinai
RP Hambardzumyan, D (通讯作者)，1425 Madison Ave,15th Floor,Room 15-20B, New York, NY 10029 USA.; Ross, JL (通讯作者)，1510 Clifton Rd,Room G-211, Atlanta, GA 30322 USA.
EM
RI Becher, Oren/GXN-3269-2022; Plant, Ashley/GVS-9394-2022
OI Becher, Oren/0000-0003-4016-8492; Hambardzumyan,
   Dolores/0000-0003-1975-4665; Ross, James/0000-0002-7311-416X
FU National Cancer Institute [F31CA232531]; Cancer Research Instute;
   National Institutes of Health [NS106554]
FX This study was funded by the National Cancer Institute F31CA232531
   (J.L.R.), Cancer Research Instute (J.L.R.), and National Institutes of
   Health NS106554 (D.H.).
CR Ahmed N, 2017, JAMA ONCOL, V3, P1094, DOI 10.1001/jamaoncol.2017.0184
   Ardon H, 2010, PEDIATR BLOOD CANCER, V54, P519, DOI 10.1002/pbc.22319
   Aziz-Bose R, 2019, CURR OPIN ONCOL, V31, P522, DOI 10.1097/CCO.0000000000000577
   Bailey CP, 2020, NEURO-ONCOLOGY, V22, P1302, DOI 10.1093/neuonc/noaa058
   Bender S, 2013, CANCER CELL, V24, P660, DOI 10.1016/j.ccr.2013.10.006
   Benitez-Ribas D, 2018, FRONT ONCOL, V8, DOI 10.3389/fonc.2018.00127
   Bernstock JD, 2020, HUM GENE THER, V31, P1132, DOI 10.1089/hum.2020.101
   Bouffet E, 2016, J CLIN ONCOL, V34, P2206, DOI 10.1200/JCO.2016.66.6552
   Braunstein S, 2017, J NEURO-ONCOL, V134, P541, DOI 10.1007/s11060-017-2393-0
   Brown NF, 2018, BRIT J CANCER, V119, P1171, DOI 10.1038/s41416-018-0258-8
   Buczkowicz P, 2014, NAT GENET, V46, P451, DOI 10.1038/ng.2936
   Butowski N, 2009, J NEURO-ONCOL, V91, P175, DOI 10.1007/s11060-008-9693-3
   Butowski N, 2009, J NEURO-ONCOL, V91, P183, DOI 10.1007/s11060-008-9705-3
   Caretti V, 2014, ACTA NEUROPATHOL, V127, P897, DOI 10.1007/s00401-014-1272-4
   Caruso DA, 2005, CANCER-AM CANCER SOC, V103, P1280, DOI 10.1002/cncr.20911
   Chen ZH, 2020, GLIA, V68, P2148, DOI 10.1002/glia.23883
   Chen ZH, 2017, CANCER RES, V77, P2266, DOI 10.1158/0008-5472.CAN-16-2310
   Chheda ZS, 2018, J EXP MED, V215, P141, DOI 10.1084/jem.20171046
   Cockle JV, 2017, MOL THER-ONCOLYTICS, V5, P75, DOI 10.1016/j.omto.2017.04.002
   Cohen KJ, 2017, NEURO-ONCOLOGY, V19, P1025, DOI 10.1093/neuonc/nox021
   Cohen KJ, 2011, NEURO-ONCOLOGY, V13, P317, DOI 10.1093/neuonc/noq191
   Cordero FJ, 2017, MOL CANCER RES, V15, P1243, DOI 10.1158/1541-7786.MCR-16-0389
   Curtin JF, 2006, J IMMUNOL, V176, P3566, DOI 10.4049/jimmunol.176.6.3566
   De Haas AH, 2008, GLIA, V56, P888, DOI 10.1002/glia.20663
   De Vleeschouwer S, 2008, CLIN CANCER RES, V14, P3098, DOI 10.1158/1078-0432.CCR-07-4875
   Desjardins A, 2018, NEW ENGL J MED, V379, P150, DOI 10.1056/NEJMoa1716435
   Diwanji TP, 2017, ADOLESC HEALTH MED T, V8, P99, DOI 10.2147/AHMT.S53391
   Doorn KJ, 2015, FRONT CELL NEUROSCI, V9, DOI 10.3389/fncel.2015.00084
   Dorfer C, 2021, ACTA NEUROCHIR, V163, P2025, DOI 10.1007/s00701-020-04589-y
   Ebrahimi A, 2016, ACTA NEUROPATHOL COM, V4, DOI 10.1186/s40478-016-0331-6
   Faria CMC, 2011, J NEUROSURG-PEDIATR, V8, P119, DOI 10.3171/2011.5.PEDS1140
   Filbin MG, 2018, SCIENCE, V360, P331, DOI 10.1126/science.aao4750
   Finlay JL, 1999, CHILD NERV SYST, V15, P786, DOI 10.1007/s003810050471
   Fischer U, 2005, ONCOGENE, V24, P1231, DOI 10.1038/sj.onc.1208290
   Foster JB, 2019, NEURO-ONCOLOGY, V21, P1226, DOI 10.1093/neuonc/noz077
   Friedman GK, 2018, SCI REP-UK, V8, DOI 10.1038/s41598-018-32353-x
   Garcia-Fabiani MB, 2020, METHOD ENZYMOL, V632, P369, DOI 10.1016/bs.mie.2019.05.023
   Geletneky K, 2010, NEURO-ONCOLOGY, V12, P804, DOI 10.1093/neuonc/noq023
   Ginhoux F, 2010, SCIENCE, V330, P841, DOI 10.1126/science.1194637
   Goldberg AD, 2010, CELL, V140, P678, DOI 10.1016/j.cell.2010.01.003
   Grabert K, 2016, NAT NEUROSCI, V19, P504, DOI 10.1038/nn.4222
   Grabovska Y, 2020, NAT COMMUN, V11, DOI 10.1038/s41467-020-18070-y
   Griesinger AM, 2013, J IMMUNOL, V191, P4880, DOI 10.4049/jimmunol.1301966
   Guerra-Garcia P, 2020, PEDIATR BLOOD CANCER, V67, DOI 10.1002/pbc.28011
   Guerrini-Rousseau L, 2019, NEURO-ONCOL ADV, V1, DOI 10.1093/noajnl/vdz033
   Hartman LLR, 2014, J PEDIAT HEMATOL ONC, V36, P451, DOI 10.1097/MPH.0000000000000047
   Hegi ME, 2005, NEW ENGL J MED, V352, P997, DOI 10.1056/NEJMoa043331
   Herting CJ, 2019, BRAIN, V142, P3834, DOI 10.1093/brain/awz331
   Herz J, 2017, IMMUNITY, V46, P943, DOI 10.1016/j.immuni.2017.06.007
   Hilf N, 2019, NATURE, V565, P240, DOI 10.1038/s41586-018-0810-y
   Hoeman CM, 2019, NAT COMMUN, V10, DOI 10.1038/s41467-019-08823-9
   Jakacki RI, 2016, NEURO-ONCOLOGY, V18, P1442, DOI 10.1093/neuonc/now038
   Jha P, 2019, NEUROPATHOLOGY, V39, P413, DOI 10.1111/neup.12594
   Jiang H, 2014, PLOS ONE, V9, DOI 10.1371/journal.pone.0097407
   Johanns TM, 2016, CANCER DISCOV, V6, P1230, DOI 10.1158/2159-8290.CD-16-0575
   Jones C, 2014, NAT REV CANCER, V14, P651, DOI 10.1038/nrc3811
   Jones C, 2017, NEURO-ONCOLOGY, V19, P153, DOI 10.1093/neuonc/now101
   Josupeit R, 2016, VIRUSES-BASEL, V8, DOI 10.3390/v8050138
   Juratli TA, 2018, PHARMACOL THERAPEUT, V182, P70, DOI 10.1016/j.pharmthera.2017.08.006
   Kaffes I, 2019, ONCOIMMUNOLOGY, V8, DOI 10.1080/2162402X.2019.1655360
   Keskin DB, 2019, NATURE, V565, P234, DOI 10.1038/s41586-018-0792-9
   Kiprianova I, 2011, CLIN CANCER RES, V17, P5333, DOI 10.1158/1078-0432.CCR-10-3124
   Korin B, 2017, NAT NEUROSCI, V20, P1300, DOI 10.1038/nn.4610
   Korshunov A, 2015, ACTA NEUROPATHOL, V129, P669, DOI 10.1007/s00401-015-1405-4
   Koschmann C, 2016, ONCOTARGET, V7, P65696, DOI 10.18632/oncotarget.11602
   Kyritsis AP, 2010, NEURO-ONCOLOGY, V12, P104, DOI 10.1093/neuonc/nop011
   Lang FF, 2018, J CLIN ONCOL, V36, P1419, DOI 10.1200/JCO.2017.75.8219
   Larson JD, 2019, CANCER CELL, V35, P140, DOI 10.1016/j.ccell.2018.11.015
   Lasky JL, 2013, ANTICANCER RES, V33, P2047
   Lewis PW, 2013, SCIENCE, V340, P857, DOI 10.1126/science.1232245
   Lieberman NAP, 2019, NEURO-ONCOLOGY, V21, P83, DOI 10.1093/neuonc/noy145
   Lin GL, 2018, ACTA NEUROPATHOL COM, V6, DOI 10.1186/s40478-018-0553-x
   Liu XY, 2014, CSH PERSPECT BIOL, V6, DOI 10.1101/cshperspect.a018689
   Liu ZG, 2013, NEURO-ONCOLOGY, V15, P1173, DOI 10.1093/neuonc/not065
   Louis DN, 2016, ACTA NEUROPATHOL, V131, P803, DOI 10.1007/s00401-016-1545-1
   Lukas RV, 2019, ONCOLOGY-NY, V33, P91
   Mackay A, 2018, CANCER CELL, V33, P829, DOI 10.1016/j.ccell.2018.04.004
   Mackay A, 2017, CANCER CELL, V32, P520, DOI 10.1016/j.ccell.2017.08.017
   Majzner RG, 2019, CLIN CANCER RES, V25, P2560, DOI 10.1158/1078-0432.CCR-18-0432
   Margueron R, 2011, NATURE, V469, P343, DOI 10.1038/nature09784
   Martinez-Velez N, 2019, NAT COMMUN, V10, DOI 10.1038/s41467-019-10043-0
   Martinez-Velez N, 2019, ACTA NEUROPATHOL COM, V7, DOI 10.1186/s40478-019-0714-6
   Mendez F, 2020, CLIN CANCER RES, V26, P4080, DOI 10.1158/1078-0432.CCR-19-3714
   Mosaab A, 2020, SCI REP-UK, V10, DOI 10.1038/s41598-020-65272-x
   Mount CW, 2018, NAT MED, V24, P572, DOI 10.1038/s41591-018-0006-x
   Mueller S, 2020, J CLIN INVEST, V130, P6325, DOI 10.1172/JCI140378
   Nazha B, 2020, FRONT ONCOL, V10, DOI 10.3389/fonc.2020.01000
   Ochs K, 2017, ONCOIMMUNOLOGY, V6, DOI 10.1080/2162402X.2017.1328340
   Ostrom Quinn T, 2019, Neuro Oncol, V21, pv1, DOI 10.1093/neuonc/noz150
   Ostrom QT, 2014, NEURO-ONCOLOGY, V16, P1, DOI [10.1093/neuonc/nov189, 10.1093/neuonc/nou327, 10.1093/neuonc/nou223]
   Pathania M, 2017, CANCER CELL, V32, P684, DOI 10.1016/j.ccell.2017.09.014
   Patil N, 2021, NEURO-ONCOLOGY, V23, P990, DOI 10.1093/neuonc/noaa295
   Patterson JD, 2020, FRONT ONCOL, V10, DOI 10.3389/fonc.2020.01582
   Paugh BS, 2011, J CLIN ONCOL, V29, P3999, DOI 10.1200/JCO.2011.35.5677
   Paugh BS, 2010, J CLIN ONCOL, V28, P3061, DOI 10.1200/JCO.2009.26.7252
   Plant AS, 2018, J NEURO-ONCOL, V137, P269, DOI 10.1007/s11060-017-2737-9
   Pollack IF, 2006, J CLIN ONCOL, V24, P3431, DOI 10.1200/JCO.2006.05.7265
   Pollack IF, 2014, J CLIN ONCOL, V32, P2050, DOI 10.1200/JCO.2013.54.0526
   Robinson MH, 2020, J IMMUNOTHER CANCER, V8, DOI 10.1136/jitc-2020-001066
   Rong Y, 2006, J NEUROPATH EXP NEUR, V65, P529, DOI 10.1097/00005072-200606000-00001
   Ross JL, 2021, BRAIN, V144, P53, DOI 10.1093/brain/awaa382
   Ross JL, 2017, SCI REP-UK, V7, DOI 10.1038/s41598-017-15849-w
   Salazar AM, 1996, NEUROSURGERY, V38, P1096, DOI 10.1097/00006123-199606000-00006
   Sarthy JF, 2020, ELIFE, V9, DOI [10.7554/eLife.61090, 10.7554/elife.61090]
   Schwartzentruber J, 2012, NATURE, V482, P226, DOI [10.1038/nature10833, 10.1007/s13311-012-0137-6]
   Shemer A, 2015, SEMIN IMMUNOPATHOL, V37, P613, DOI 10.1007/s00281-015-0519-z
   Shum T, 2017, CANCER DISCOV, V7, P1238, DOI 10.1158/2159-8290.CD-17-0538
   Silvin A, 2018, GLIA, V66, P2045, DOI 10.1002/glia.23458
   Solomon DA, 2016, BRAIN PATHOL, V26, P569, DOI 10.1111/bpa.12336
   Stucklin ASG, 2019, NAT COMMUN, V10, DOI 10.1038/s41467-019-12187-5
   Sturm D, 2012, CANCER CELL, V22, P425, DOI 10.1016/j.ccr.2012.08.024
   Subashi E, 2016, J NEURO-ONCOL, V126, P243, DOI 10.1007/s11060-015-1969-9
   Tang X, 2019, MOL THER-ONCOLYTICS, V14, P279, DOI 10.1016/j.omto.2019.07.002
   Tasic B, 2016, NAT NEUROSCI, V19, P335, DOI 10.1038/nn.4216
   Tejada S, 2018, FRONT ONCOL, V8, DOI 10.3389/fonc.2018.00061
   Tejada S, 2018, NEUROSURGERY, V83, P1050, DOI 10.1093/neuros/nyx507
   Thewissen K, 2014, MULT SCLER J, V20, P548, DOI 10.1177/1352458513505352
   Vanan MI, 2014, NEURO-ONCOL PRACT, V1, P145, DOI 10.1093/nop/npu022
   Vinci M, 2018, NAT MED, V24, P1204, DOI 10.1038/s41591-018-0086-7
   Wang LM, 2018, HUM PATHOL, V78, P89, DOI 10.1016/j.humpath.2018.04.015
   Wang SS, 2019, TRENDS IMMUNOL, V40, P748, DOI 10.1016/j.it.2019.05.009
   Wu G, 2014, NAT GENET, V46, P444, DOI 10.1038/ng.2938
   Wu G, 2012, NAT GENET, V44, P251, DOI 10.1038/ng.1102
   Yang T, 2013, WORLD NEUROSURG, V79, P537, DOI 10.1016/j.wneu.2012.09.015
   Yona S, 2013, IMMUNITY, V38, P79, DOI 10.1016/j.immuni.2012.12.001
   Yu LT, 2011, NEURO-ONCOLOGY, V13, P14, DOI 10.1093/neuonc/noq148
   Zhu XM, 2010, CANCER IMMUNOL IMMUN, V59, P1401, DOI 10.1007/s00262-010-0876-3
NR 127
TC 10
Z9 11
U1 2
U2 3
PU OXFORD UNIV PRESS
PI OXFORD
PA GREAT CLARENDON ST, OXFORD OX2 6DP, ENGLAND
SN 0006-8950
EI 1460-2156
J9 BRAIN
JI Brain
PD SEP
PY 2021
VL 144
BP 2594
EP 2609
DI 10.1093/brain/awab155
EA APR 2021
PN 9
PG 16
WC Clinical Neurology; Neurosciences
WE Science Citation Index Expanded (SCI-EXPANDED)
SC Neurosciences & Neurology
GA XT6VH
UT WOS:000733722200021
PM 33856022
OA Green Published, Bronze
DA 2023-08-27
ER

PT J
AU Ohno, N
   Terada, N
   Komada, M
   Saitoh, S
   Costantini, F
   Pace, V
   Germann, PG
   Weber, K
   Yamakawa, H
   Ohara, O
   Ohno, S
AF Ohno, Nobuhiko
   Terada, Nobuo
   Komada, Masayuki
   Saitoh, Sei
   Costantini, Frank
   Pace, Virgilio
   Germann, Paul-Georg
   Weber, Klaus
   Yamakawa, Hisashi
   Ohara, Osamu
   Ohno, Shinichi
TI Dispensable role of protein 4.1B/DAL-1 in rodent adrenal medulla
   regarding generation of pheochromocytoma and plasmalemmal localization
   of TSLC1
SO BIOCHIMICA ET BIOPHYSICA ACTA-MOLECULAR CELL RESEARCH
LA English
DT Article
DE Adrenal medulla; Chromaffin cell; Pheochromocytoma; Protein 4.1B; TSLC1
ID IN-VIVO CRYOTECHNIQUE; CELL-ADHESION MOLECULE; BREAST-CANCER CELLS;
   TUMOR-SUPPRESSOR; CHROMAFFIN CELLS; LUNG-CANCER; HEMODYNAMIC CONDITIONS;
   EPITHELIAL-CELLS; MALE-INFERTILITY; MESSENGER-RNA
AB Protein 4.1B is a membrane skeletal protein expressed in various organs, and is associated with tumor suppressor in lung cancer-1 (TSLC1) in vitro. Although involvement of 4.1B in the intercellular junctions and tumor-suppression was suggested, some controversial results posed questions to the general tumor-suppressive function of 4.1B and its relation to TSLC1 in vivo. In this study, the expression of 4.1B and its interaction with TSLC1 were examined in rodent adrenal gland, and the involvement of 4.1B in tumorigenesis and the effect of 4.1B deficiency on TSLC1 distribution were also investigated using rodent pheochromocytoma and 4.1B-knockout mice. Although plasmalemmal immunolocalization of 4.1B was shown in chromaffin cells of rodent adrenal medulla, expression of 4.1B was maintained in developed pheochromocytoma, and morphological abnormality or pheochromocytoma generation could not be found in 4.1B-deficient mice. Furthermore, molecular interaction and colocalization of 4.1B and TSLC1 were observed in mouse adrenal gland, but the immunolocalization of TSLC1 along chromaffin cell membranes was not affected in the 4.1B-deficient mice. These results suggest that the function of 4.1B as tumor suppressor might significantly differ among organs and species, and that plasmalemmal retention of TSLC1 would be maintained by molecules other than 4.1B interacting in rodent chromaffin cells. (c) 2009 Elsevier B.V. All rights reserved.
C1 [Ohno, Nobuhiko; Terada, Nobuo; Saitoh, Sei; Ohno, Shinichi] Univ Yamanashi, Dept Anat & Mol Histol, Interdisciplinary Grad Sch Med & Engn, Chuo Ku, Yamanashi 4093898, Japan.
   [Komada, Masayuki] Tokyo Inst Technol, Dept Biol Sci, Kanagawa 2268501, Japan.
   [Costantini, Frank] Columbia Univ, Med Ctr, Dept Genet & Dev, New York, NY 10032 USA.
   [Pace, Virgilio] Dompe Pharma SpA, R&D, I-67100 Laquila, Italy.
   [Germann, Paul-Georg] Nycomed GmbH, D-75467 Constance, Germany.
   [Weber, Klaus] RCC Ltd, Dept Pathol, CH-4452 Itingen, Switzerland.
   [Yamakawa, Hisashi; Ohara, Osamu] Kazusa DNA Res Inst, Dept Human Gene Res, Chiba 2920818, Japan.
C3 University of Yamanashi; Tokyo Institute of Technology; Columbia
   University; Dompe; Nycomed GmbH; Kazusa DNA Research Institute
RP Ohno, S (通讯作者)，Univ Yamanashi, Dept Anat & Mol Histol, Interdisciplinary Grad Sch Med & Engn, Chuo Ku, Yamanashi 4093898, Japan.
EM
RI Ohara, Osamu/A-9119-2012; Weber, Klaus/P-8241-2019; Ohara,
   Osamu/G-5448-2015
OI Weber, Klaus/0000-0002-2421-6178; Ohara, Osamu/0000-0002-3328-9571
FU Japan Society for the Promotion of Science [18590181]; National Cancer
   Institute [5P01CA023767]; Grants-in-Aid for Scientific Research
   [18590181] Funding Source: KAKEN
FX This work was partially supported by a grant from Japan Society for the
   Promotion of Science (#18590181) to N.T and a grant from the National
   Cancer Institute (5P01CA023767) to F.C. We thank Dr. Christopher Nelson
   for his careful editing of this manuscript.
CR BENEDECZKY I, 1975, CELL TISSUE RES, V162, P541
   Braga VMM, 2002, CURR OPIN CELL BIOL, V14, P546, DOI 10.1016/S0955-0674(02)00373-3
   Cavanna T, 2007, J CELL SCI, V120, P606, DOI 10.1242/jcs.000273
   Chan SA, 2005, J NEUROPHYSIOL, V94, P1037, DOI 10.1152/jn.01213.2004
   Charboneau AL, 2002, INT J CANCER, V100, P181, DOI 10.1002/ijc.10470
   COUPLAND RE, 1965, J ANAT, V99, P231
   DEISENKONEHRBAS.N, 2003, EUR J NEUROSCI, V17, P411
   Delhommeau F, 2002, BLOOD, V100, P2629, DOI 10.1182/blood.V100.7.2629
   Fritz A, 2002, CANCER RES, V62, P3048
   Fukuhara H, 2003, ONCOGENE, V22, P6160, DOI 10.1038/sj.onc.1206744
   Garcia AG, 2006, PHYSIOL REV, V86, P1093, DOI 10.1152/physrev.00039.2005
   Gascard P, 2004, BBA-GENE STRUCT EXPR, V1680, P71, DOI 10.1016/j.bbaexp.2004.08.006
   Gollan L, 2002, J CELL BIOL, V157, P1247, DOI 10.1083/jcb.200203050
   Gutmann DH, 2000, HUM MOL GENET, V9, P1495, DOI 10.1093/hmg/9.10.1495
   Heller G, 2006, ONCOGENE, V25, P959, DOI 10.1038/sj.onc.1209115
   Heller G, 2007, BREAST CANCER RES TR, V103, P283, DOI 10.1007/s10549-006-9377-7
   Houshmandi SS, 2006, NEUROLOGY, V67, P1863, DOI 10.1212/01.wnl.0000244472.56198.84
   Jamora C, 2002, NAT CELL BIOL, V4, pE101, DOI 10.1038/ncb0402-e101
   Jiang W, 2006, MOL CANCER, V5, DOI 10.1186/1476-4598-5-4
   Jimenez C, 2006, J CLIN ENDOCR METAB, V91, P2851, DOI 10.1210/jc.2005-2178
   Kannan R, 1996, FEBS LETT, V385, P159, DOI 10.1016/0014-5793(96)00350-X
   Karagiannis A, 2007, ENDOCR-RELAT CANCER, V14, P935, DOI 10.1677/ERC-07-0142
   Kikuchi S, 2006, CANCER-AM CANCER SOC, V106, P1751, DOI 10.1002/cncr.21800
   Komada M, 1999, GENE DEV, V13, P1475, DOI 10.1101/gad.13.11.1475
   Kuns R, 2005, ONCOGENE, V24, P6502, DOI 10.1038/sj.onc.1208813
   Kuramochi M, 2001, NAT GENET, V27, P427, DOI 10.1038/86934
   LACY PE, 1959, ANN NY ACAD SCI, V82, P287, DOI 10.1111/j.1749-6632.1959.tb44909.x
   Li ZL, 2006, HISTOCHEM CELL BIOL, V126, P399, DOI 10.1007/s00418-006-0175-4
   McCarty JH, 2005, P NATL ACAD SCI USA, V102, P13479, DOI 10.1073/pnas.0506068102
   Murakami Y, 2005, CANCER SCI, V96, P543, DOI 10.1111/j.1349-7006.2005.00089.x
   Ohara R, 2000, MOL BRAIN RES, V85, P41, DOI 10.1016/S0169-328X(00)00233-3
   Ohno N, 2004, HISTOCHEM CELL BIOL, V122, P579, DOI 10.1007/s00418-004-0716-7
   Ohno N, 2007, J COMP NEUROL, V505, P292, DOI 10.1002/cne.21498
   Ohno N, 2006, HISTOCHEM CELL BIOL, V126, P389, DOI 10.1007/s00418-006-0173-6
   Pace V, 2002, TOXICOL PATHOL, V30, P492, DOI 10.1080/01926230290105668
   Parra M, 2004, GENOMICS, V84, P637, DOI 10.1016/j.ygeno.2004.06.004
   Parra M, 2000, J BIOL CHEM, V275, P3247, DOI 10.1074/jbc.275.5.3247
   Perez-Moreno M, 2003, CELL, V112, P535, DOI 10.1016/S0092-8674(03)00108-9
   Ramez M, 2003, KIDNEY INT, V63, P1321, DOI 10.1046/j.1523-1755.2003.00870.x
   Revenu C, 2004, NAT REV MOL CELL BIO, V5, P635, DOI 10.1038/nrm1437
   Robb VA, 2005, ONCOGENE, V24, P1946, DOI 10.1038/sj.onc.1208335
   Shi ZT, 1999, J CLIN INVEST, V103, P331, DOI 10.1172/JCI3858
   Shingai T, 2003, J BIOL CHEM, V278, P35421, DOI 10.1074/jbc.M305387200
   Singh PK, 2002, MODERN PATHOL, V15, P526, DOI 10.1038/modpathol.3880558
   Smith-Hicks CL, 2000, EMBO J, V19, P612, DOI 10.1093/emboj/19.4.612
   Sun CX, 2002, J CELL SCI, V115, P3991, DOI 10.1242/jcs.00094
   Terada N, 2005, HISTOCHEM CELL BIOL, V124, P303, DOI 10.1007/s00418-005-0031-y
   Terada N, 2004, NEUROSCI LETT, V366, P15, DOI 10.1016/j.neulet.2004.05.005
   Terada N, 2004, J HISTOCHEM CYTOCHEM, V52, P769, DOI 10.1369/jhc.3A6192.2004
   Terada N, 2003, HISTOCHEM CELL BIOL, V120, P277, DOI 10.1007/s00418-003-0573-9
   Terada N, 2007, J HISTOCHEM CYTOCHEM, V55, P1199, DOI 10.1369/jhc.7A7266.2007
   Terada Nobuo, 2004, Medical Electron Microscopy, V37, P45
   Tran YK, 1999, CANCER RES, V59, P35
   Tsujiuchi T, 2007, CANCER SCI, V98, P943, DOI 10.1111/j.1349-7006.2007.00480.x
   van der Weyden L, 2006, MOL CELL BIOL, V26, P3595, DOI 10.1128/MCB.26.9.3595-3609.2006
   Wakayama T, 2001, MOL REPROD DEV, V60, P158, DOI 10.1002/mrd.1072
   Walensky LD, 1999, J NEUROSCI, V19, P6457
   Wong SY, 2007, P NATL ACAD SCI USA, V104, P12784, DOI 10.1073/pnas.0705499104
   Yageta M, 2002, CANCER RES, V62, P5129
   Yamada D, 2006, MOL CELL BIOL, V26, P3610, DOI 10.1128/MCB.26.9.3610-3624.2006
   Yamakawa H, 2000, GENE, V248, P137, DOI 10.1016/S0378-1119(00)00130-X
   Yi CL, 2005, MOL CELL BIOL, V25, P10052, DOI 10.1128/MCB.25.22.10052-10059.2005
NR 62
TC 6
Z9 6
U1 0
U2 3
PU ELSEVIER SCIENCE BV
PI AMSTERDAM
PA PO BOX 211, 1000 AE AMSTERDAM, NETHERLANDS
SN 0167-4889
EI 0006-3002
J9 BBA-MOL CELL RES
JI Biochim. Biophys. Acta-Mol. Cell Res.
PD MAR
PY 2009
VL 1793
IS 3
BP 506
EP 515
DI 10.1016/j.bbamcr.2009.01.005
PG 10
WC Biochemistry & Molecular Biology; Cell Biology
WE Science Citation Index Expanded (SCI-EXPANDED)
SC Biochemistry & Molecular Biology; Cell Biology
GA 420EK
UT WOS:000264271500008
PM 19321127
DA 2023-08-27
ER

PT J
AU Miyahara, H
   Yadavilli, S
   Natsumeda, M
   Rubens, JA
   Rodgers, L
   Kambhampati, M
   Taylor, IC
   Kaur, H
   Asnaghi, L
   Eberhart, CG
   Warren, KE
   Nazarian, J
   Raabe, EH
AF Miyahara, Hiroaki
   Yadavilli, Sridevi
   Natsumeda, Manabu
   Rubens, Jeffrey A.
   Rodgers, Louis
   Kambhampati, Madhuri
   Taylor, Isabella C.
   Kaur, Harpreet
   Asnaghi, Laura
   Eberhart, Charles G.
   Warren, Katherine E.
   Nazarian, Javad
   Raabe, Eric H.
TI The dual mTOR kinase inhibitor TAK228 inhibits tumorigenicity and
   enhances radiosensitization in diffuse intrinsic pontine glioma
SO CANCER LETTERS
LA English
DT Article
DE Pediatric brain tumor; Sapanisertib; mTOR; INK128; MLN0128
ID NEURAL STEM-CELLS; ANTITUMOR-ACTIVITY; K27M MUTATION; GRADE GLIOMA;
   GROWTH; ACTIVATION; RADIATION; INVASION; MLN0128; TRANSFORMATION
AB Diffuse intrinsic pontine glioma (DIPG) is an invasive and treatment-refractory pediatric brain tumor. Primary DIPG tumors harbor a number of mutations including alterations in PTEN, AKT, and PI3K and exhibit activation of mammalian Target of Rapamycin Complex 1 and 2 (mTORC1/2). mTORC1/2 regulate protein translation, cell growth, survival, invasion, and metabolism. Pharmacological inhibition of mTORC1 is minimally effective in DIPG. However, the activity of dual TORC kinase inhibitors has not been examined in this tumor type.
   Nanomolar levels of the mTORC1/2 inhibitor TAK228 reduced expression of p-AKT(S473) and p-S6(S240/244) and suppressed the growth of DIPG lines JHH-DIPG1, SF7761, and SU-DIPG-XIII. TAK228 induced apoptosis in DIPG cells and cooperated with radiation to further block proliferation and enhance apoptosis.
   TAK228 monotherapy inhibited the tumorigenicity of a murine orthotopic model of DIPG, more than doubling median survival (p = 0.0017) versus vehicle. We conclude that dual mTOR inhibition is a promising potential candidate for DIPG treatment. (C) 2017 Elsevier B.V. All rights reserved.
C1 [Miyahara, Hiroaki; Natsumeda, Manabu; Asnaghi, Laura; Eberhart, Charles G.; Raabe, Eric H.] Johns Hopkins Univ, Sch Med, Dept Pathol, Div Neuropathol, 1800 Orleans, Baltimore, MD 21287 USA.
   [Yadavilli, Sridevi; Kambhampati, Madhuri; Nazarian, Javad] Childrens Natl Hlth Syst, Med Genet Res Ctr, Washington, DC 20010 USA.
   [Rubens, Jeffrey A.; Taylor, Isabella C.; Kaur, Harpreet; Raabe, Eric H.] Johns Hopkins Univ, Sch Med, Sidney Kimmel Comprehens Canc Ctr, Div Pediat Oncol, 1800 Orleans, Baltimore, MD 21287 USA.
   [Rodgers, Louis; Warren, Katherine E.] NCI, NIH, Bethesda, MD 20892 USA.
   [Nazarian, Javad] George Washington Univ, Sch Med & Hlth Sci, Dept Integrat Syst Biol, Washington, DC 20052 USA.
C3 Johns Hopkins University; Children's National Health System; Johns
   Hopkins University; Johns Hopkins Medicine; National Institutes of
   Health (NIH) - USA; NIH National Cancer Institute (NCI); George
   Washington University
RP Raabe, EH (通讯作者)，Johns Hopkins Univ, Sch Med, 1800 Orleans, Baltimore, MD 21287 USA.
EM
RI Natsumeda, Manabu/Z-2072-2019; Kaur, Harpreet/HTN-0358-2023
OI Natsumeda, Manabu/0000-0003-3098-8323; Nazarian,
   Javad/0000-0002-1951-9828
FU Cure Starts Now Foundation; Giant Food Foundation; NCI [P30 CA006973];
   National Institute of Health/National Cancer Institute [T32CA60441];
   Smashing Walnuts/Piedmont community; Goldwin Foundation
FX Funding was provided by the Cure Starts Now Foundation to the
   Mid-Atlantic DIPG consortium (Drs. Nazarian, Raabe, Warren). Additional
   funding by the Giant Food Foundation, NCI Core Grant to the Johns
   Hopkins Sidney Kimmel Comprehensive Cancer Center P30 CA006973, National
   Institute of Health/National Cancer Institute (T32CA60441 to J.A.R),
   Smashing Walnuts/Piedmont community (Dr. Nazarian), and the Goldwin
   Foundation (Dr. Nazarian). EHR is a St. Baldrick's Scholar.
CR Buczkowicz P, 2014, ACTA NEUROPATHOL, V128, P573, DOI 10.1007/s00401-014-1319-6
   Buczkowicz P, 2014, NAT GENET, V46, P451, DOI 10.1038/ng.2936
   Choi EJ, 2014, BMC CANCER, V14, DOI 10.1186/1471-2407-14-17
   Khuong-Quang DA, 2012, ACTA NEUROPATHOL, V124, P439, DOI 10.1007/s00401-012-0998-0
   Eun SY, 2015, ONCOL REP, V34, P195, DOI 10.3892/or.2015.3972
   Garcia-Garcia C, 2012, CLIN CANCER RES, V18, P2603, DOI 10.1158/1078-0432.CCR-11-2750
   Gielen GH, 2013, AM J CLIN PATHOL, V139, P345, DOI 10.1309/AJCPABOHBC33FVMO
   Gokmen-Polar Y, 2012, BREAST CANCER RES TR, V136, P673, DOI 10.1007/s10549-012-2298-8
   Grasso CS, 2015, NAT MED, V21, P555, DOI 10.1038/nm.3855
   Guertin DA, 2005, TRENDS MOL MED, V11, P353, DOI 10.1016/j.molmed.2005.06.007
   Guertin DA, 2009, SCI SIGNAL, V2, DOI 10.1126/scisignal.267pe24
   Halvorson KG, 2015, PLOS ONE, V10, DOI 10.1371/journal.pone.0118926
   Hashizume R, 2012, J NEURO-ONCOL, V110, P305, DOI 10.1007/s11060-012-0973-6
   Hassan B, 2014, ONCOTARGET, V5, P8544, DOI 10.18632/oncotarget.2337
   Heikamp EB, 2014, NAT IMMUNOL, V15, P457, DOI 10.1038/ni.2867
   Hoffman LM, 2016, ACTA NEUROPATHOL COM, V4, DOI 10.1186/s40478-015-0269-0
   Hou J, 2015, PLOS ONE, V10, DOI 10.1371/journal.pone.0122179
   Hsieh AC, 2012, NATURE, V485, P55, DOI 10.1038/nature10912
   Hutt-Cabezas M, 2013, NEURO-ONCOLOGY, V15, P1604, DOI 10.1093/neuonc/not132
   Kallappagoudar S, 2015, CHROMOSOMA, V124, P177, DOI 10.1007/s00412-015-0510-4
   Kang MH, 2014, PEDIATR BLOOD CANCER, V61, P1486, DOI 10.1002/pbc.24989
   Kaur H, 2016, CANCER LETT, V377, P55, DOI 10.1016/j.canlet.2016.04.020
   Kaur H, 2015, J NEUROPATH EXP NEUR, V74, P177, DOI 10.1097/NEN.0000000000000161
   Kaur H, 2012, MOL CANCER RES, V10, P293, DOI 10.1158/1541-7786.MCR-11-0457
   Kuo Y, 2015, SCI REP-UK, V5, DOI 10.1038/srep10339
   Laplante M, 2012, CELL, V149, P274, DOI 10.1016/j.cell.2012.03.017
   Lou HZ, 2014, BIOCHEM BIOPH RES CO, V450, P973, DOI 10.1016/j.bbrc.2014.06.081
   Mao XG, 2013, ONCOTARGET, V4, P1050, DOI 10.18632/oncotarget.1131
   Masri J, 2007, CANCER RES, V67, P11712, DOI 10.1158/0008-5472.CAN-07-2223
   Matsuoka H, 2009, EXP CELL RES, V315, P2022, DOI 10.1016/j.yexcr.2009.04.009
   Mazan-Mamczarz K, 2015, CELL COMMUN SIGNAL, V13, DOI 10.1186/s12964-015-0091-0
   Mehta M, 2015, MOL CANCER THER, V14, P1171, DOI 10.1158/1535-7163.MCT-14-0708
   Millet P, 2013, INT J ONCOL, V43, P375, DOI 10.3892/ijo.2013.1970
   Nikbakht H, 2016, NAT COMMUN, V7, DOI 10.1038/ncomms11185
   Noh EM, 2015, ONCOL REP, V34, P803, DOI 10.3892/or.2015.4027
   Puget S, 2015, CHILD NERV SYST, V31, P1773, DOI 10.1007/s00381-015-2832-1
   Raabe EH, 2011, CLIN CANCER RES, V17, P3590, DOI 10.1158/1078-0432.CCR-10-3349
   Rasband W.S., 1997, IMAGEJ
   Rispal D, 2015, J BIOL CHEM, V290, P14963, DOI 10.1074/jbc.M114.627794
   Schenone S, 2011, CURR MED CHEM, V18, P2995, DOI 10.2174/092986711796391651
   Slotkin EK, 2015, MOL CANCER THER, V14, P395, DOI 10.1158/1535-7163.MCT-14-0711
   Taylor IC, 2015, J NEUROPATH EXP NEUR, V74, P778, DOI 10.1097/NEN.0000000000000216
   Warren KE, 2012, FRONT ONCOL, V2, DOI 10.3389/fonc.2012.00205
   Warren KE, 2012, NEURO-ONCOLOGY, V14, P326, DOI 10.1093/neuonc/nor190
   Weingart MF, 2015, ONCOTARGET, V6, P3165, DOI 10.18632/oncotarget.3078
   Yadavilli S, 2015, ONCOTARGET, V6, P12141, DOI 10.18632/oncotarget.3716
   Yang MY, 2010, J AGR FOOD CHEM, V58, P5806, DOI 10.1021/jf100718b
   Yue PYK, 2010, J BIOMOL SCREEN, V15, P427, DOI 10.1177/1087057110361772
   Zarghooni M, 2010, J CLIN ONCOL, V28, P1337, DOI 10.1200/JCO.2009.25.5463
   Zhang HY, 2015, APOPTOSIS, V20, P50, DOI 10.1007/s10495-014-1066-0
NR 50
TC 42
Z9 42
U1 0
U2 17
PU ELSEVIER IRELAND LTD
PI CLARE
PA ELSEVIER HOUSE, BROOKVALE PLAZA, EAST PARK SHANNON, CO, CLARE, 00000,
   IRELAND
SN 0304-3835
EI 1872-7980
J9 CANCER LETT
JI Cancer Lett.
PD AUG 1
PY 2017
VL 400
BP 110
EP 116
DI 10.1016/j.canlet.2017.04.019
PG 7
WC Oncology
WE Science Citation Index Expanded (SCI-EXPANDED)
SC Oncology
GA EY0NO
UT WOS:000403655200012
PM 28450157
OA Green Accepted
DA 2023-08-27
ER

PT J
AU Shahar, E
   Postovsky, S
   Bennett, O
AF Shahar, E
   Postovsky, S
   Bennett, O
TI Central neurogenic hyperventilation in a conscious child associated with
   glioblastoma multiforme
SO PEDIATRIC NEUROLOGY
LA English
DT Article
ID AWAKE PATIENT; LYMPHOMA; TUMOR
AB Central neurogenic hyperventilation refers to progressive tachypnea leading to hypocarbia and respiratory alkalosis caused by cortical disorders, initially reported in comatose patients with mainly pontine infarction. Central neurogenic hyperventilation in conscious patients is even rarer, numbering around 30 reported cases including seven children, mainly associated with infiltrative gliomas and lymphomas of the brainstem and pons. We report the evolution of central neurogenic hyperventilation in a conscious child associated with an infiltrative glioblastoma multiforme diagnosed 1 year before admission. He presented with progressive tachypnea and dyspnea of 1 week duration. On examination he was fully alert and aware of his respiratory disorder. Respiratory rate was 56 breaths per minute using accessory respiratory muscles. Hyperventilation was unchanged during sleep. Arterial blood gases disclosed marked hypocarbia: Pco(2) of 8 mm Hg resulting in severe respiratory alkalosis at pH of 7.8. Central neurogenic hyperventilation was therefore suggested after exclusion of other respiratory or cardiac disorders. The exaggerated tachypnea persisted along with respiratory alkalosis. Over a period of 2 months his overall state markedly deteriorated; he lapsed into coma, and finally succumbed after involvement of medullary cardiovascular centers. Although extremely rare in the pediatric age group, central neurogenic hyperventilation should be suspected in any alert child presenting with unexplained increasing tachypnea and hypocarbia leading to respiratory alkalosis. The evolution of such a disorder may be an alarming sign of ensuing deterioration in patients with tumors of the brainstem and medulla before cardiovascullar derangement. (C) 2004 by Elsevier Inc. All rights reserved.
C1 Meyer Children Hosp, Rambam Med Ctr, Child Neurol Unit, Rappaport Sch Med, IL-31096 Haifa, Israel.
   Meyer Children Hosp, Rambam Med Ctr, Epilepsy Serv, IL-31096 Haifa, Israel.
   Meyer Children Hosp, Rambam Med Ctr, Rappaport Sch Med, Hematol Oncol Dept, IL-31096 Haifa, Israel.
C3 Rambam Health Care Campus; Technion Israel Institute of Technology;
   Rappaport Faculty of Medicine; Rambam Health Care Campus; Rambam Health
   Care Campus; Technion Israel Institute of Technology; Rappaport Faculty
   of Medicine
RP Shahar, E (通讯作者)，Meyer Children Hosp, Rambam Med Ctr, Child Neurol Unit, Rappaport Sch Med, IL-31096 Haifa, Israel.
CR Alibhoy A. T., 2001, Ceylon Medical Journal, V46, P99
   BATEMAN DE, 1985, ANN NEUROL, V17, P402, DOI 10.1002/ana.410170418
   COHN FS, 1985, ANN NEUROL, V18, P163
   DUBAYBO BA, 1991, CHEST, V99, P767, DOI 10.1378/chest.99.3.767
   GOTTLIEB D, 1987, EUR NEUROL, V27, P51, DOI 10.1159/000116129
   GOULON M, 1969, REV NEUROL-FRANCE, V121, P636
   HOCHBERG FH, 1988, J NEUROSURG, V68, P835, DOI 10.3171/jns.1988.68.6.0835
   HOOL GJ, 1993, J PAEDIATR CHILD H, V29, P154
   JAECKLE KA, 1990, NEUROLOGY, V40, P1715, DOI 10.1212/WNL.40.11.1715
   Johnston SC, 2001, NEUROLOGY, V57, P2131, DOI 10.1212/WNL.57.11.2131
   KRENDEL DA, 1991, NEUROLOGY, V41, P1156, DOI 10.1212/WNL.41.7.1156-a
   Kubota M, 1996, ACTA PAEDIATR JAPON, V38, P57
   LANGE LS, 1965, J NEUROL NEUROSUR PS, V28, P317, DOI 10.1136/jnnp.28.4.317
   Laskey AL, 2000, PEDIATR NEUROL, V22, P305, DOI 10.1016/S0887-8994(99)00143-5
   Madrid HS, 2000, REV INVEST CLIN, V52, P584
   MOHAMED EEH, 1990, BRIT J CLIN PRACT, V44, P201
   Nakasu Y, 1988, Nihon Geka Hokan, V57, P165
   NGAI SH, 1957, AM J PHYSIOL, V190, P343, DOI 10.1152/ajplegacy.1957.190.2.343
   PAUZNER R, 1989, ARCH NEUROL-CHICAGO, V46, P510, DOI 10.1001/archneur.1989.00520410044021
   PLUM F, 1982, ANN NEUROL, V11, P636, DOI 10.1002/ana.410110615
   PLUM F, 1959, ARCH NEURO PSYCHIATR, V81, P535, DOI 10.1001/archneurpsyc.1959.02340170001001
   RODRIGUEZ M, 1982, ANN NEUROL, V11, P625, DOI 10.1002/ana.410110612
   Sakamoto T, 2001, Radiat Med, V19, P209
   SALVESEN R, 1989, J NEUROL NEUROSUR PS, V52, P1441, DOI 10.1136/jnnp.52.12.1441
   SCIALDONE AM, 1990, ANN EMERG MED, V19, P817, DOI 10.1016/S0196-0644(05)81711-1
   Shams PN, 2002, J NEUROL NEUROSUR PS, V72, P271, DOI 10.1136/jnnp.72.2.271
   SHIBATA Y, 1992, J NEUROSURG, V76, P696, DOI 10.3171/jns.1992.76.4.0696
   Siderowf AD, 1996, NEUROLOGY, V46, P1160, DOI 10.1212/WNL.46.4.1160
   Sugama S, 1990, Rinsho Shinkeigaku, V30, P994
   SUNDERRAJAN EV, 1984, CHEST, V86, P634, DOI 10.1378/chest.86.4.634
   Tinaztepe B, 1981, Turk J Pediatr, V23, P269
   TOBIAS JD, 1991, PEDIATRICS, V88, P818
NR 32
TC 10
Z9 10
U1 0
U2 3
PU ELSEVIER SCIENCE INC
PI NEW YORK
PA STE 800, 230 PARK AVE, NEW YORK, NY 10169 USA
SN 0887-8994
EI 1873-5150
J9 PEDIATR NEUROL
JI Pediatr. Neurol.
PD APR
PY 2004
VL 30
IS 4
BP 287
EP 290
DI 10.1016/j.pediatrneurol.2003.10.003
PG 4
WC Clinical Neurology; Pediatrics
WE Science Citation Index Expanded (SCI-EXPANDED)
SC Neurosciences & Neurology; Pediatrics
GA 816VZ
UT WOS:000221138700012
PM 15087110
DA 2023-08-27
ER

PT J
AU Goya-Outi, J
   Orlhac, F
   Calmon, R
   Alentorn, A
   Nioche, C
   Philippe, C
   Puget, S
   Boddaert, N
   Buvat, I
   Grill, J
   Frouin, V
   Frouin, F
AF Goya-Outi, Jessica
   Orlhac, Fanny
   Calmon, Raphael
   Alentorn, Agusti
   Nioche, Christophe
   Philippe, Cathy
   Puget, Stephanie
   Boddaert, Nathalie
   Buvat, Irene
   Grill, Jacques
   Frouin, Vincent
   Frouin, Frederique
TI Computation of reliable textural indices from multimodal brain MRI:
   suggestions based on a study of patients with diffuse intrinsic pontine
   glioma
SO PHYSICS IN MEDICINE AND BIOLOGY
LA English
DT Article
DE radiomics; texture analysis; diffuse intrinsic pontine glioma; 3D MRI
ID MAGNETIC-RESONANCE IMAGES; RADIOMICS; NORMALIZATION; HETEROGENEITY;
   REGISTRATION; INFORMATION; PERFORMANCE; PARAMETERS; FEATURES
AB Few methodological studies regarding widely used textural indices robustness in MRI have been reported. In this context, this study aims to propose some rules to compute reliable textural indices from multimodal 3D brain MRI. Diagnosis and post-biopsy MR scans including T1, post-contrast T1, T2 and FLAIR images from thirty children with diffuse intrinsic pontine glioma (DIPG) were considered. The hybrid white stripe method was adapted to standardize MR intensities. Sixty textural indices were then computed for each modality in different regions of interest (ROI), including tumor and white matter (WM). Three types of intensity binning were compared d(i): constant bin width and relative bounds; d(ii) : constant number of bins and relative bounds; d(iii) : constant number of bins and absolute bounds. The impact of the volume of the region was also tested within the WM. First, the mean Hellinger distance between patient-based intensity distributions decreased by a factor greater than 10 in WM and greater than 2.5 in gray matter after standardization. Regarding the binning strategy, the ranking of patients was highly correlated for 188/240 features when comparing d(i) with d(iii), but for only 20 when comparing d(i) with d(ii), and nine when comparing d(ii) with d(iii). Furthermore, when using d(i) or d(iii), texture indices reflected tumor heterogeneity as assessed visually by experts. Last, 41 features presented statistically significant differences between contralateral WM regions when ROI size slightly varies across patients, and none when using ROI of the same size. For regions with similar size, 224 features were significantly different between WM and tumor. Valuable information from texture indices can be biased by methodological choices. Recommendations are to standardize intensities in MR brain volumes, to use intensity binning with constant bin width, and to define regions with the same volumes to get reliable textural indices.
C1 [Goya-Outi, Jessica; Orlhac, Fanny; Nioche, Christophe; Buvat, Irene; Frouin, Frederique] Univ Paris Sud, Univ Paris Saclay, CNRS, CEA,IMIV,INSERM, Orsay, France.
   [Calmon, Raphael; Boddaert, Nathalie] Hop Necker Enfants Malad, AP HP, Pediat Radiol Dept, Paris, France.
   [Calmon, Raphael; Boddaert, Nathalie] INSERM, U1000, Paris, France.
   [Calmon, Raphael; Boddaert, Nathalie] Imagine Inst Malad Genet, UMR 1163, Paris, France.
   [Calmon, Raphael; Boddaert, Nathalie] Univ Paris 05, ComUE Sorbonne Paris Cite, Paris, France.
   [Alentorn, Agusti] Grp Hosp Pitie Salpetriere, AP HP, Neurol Dept, Paris, France.
   [Alentorn, Agusti] UPMC Univ Paris 6, CNRS, Sorbonne Univ, INSERM,ICM, Paris 6, France.
   [Alentorn, Agusti; Philippe, Cathy; Frouin, Vincent] Univ Paris Saclay, CEA, Neurospin, UNATI, Gif Sur Yvette, France.
   [Puget, Stephanie] Hop Necker Enfants Malad, AP HP, Pediat Neurosurg Dept, Paris, France.
   [Grill, Jacques] Gustave Roussy, Dept Cancerol Enfant & Adolescent, Villejuif, France.
   [Grill, Jacques] Univ Paris Saclay, CNRS UMR 8203, Villejuif, France.
C3 CEA; Centre National de la Recherche Scientifique (CNRS); Institut
   National de la Sante et de la Recherche Medicale (Inserm); UDICE-French
   Research Universities; Universite Paris Saclay; Assistance Publique
   Hopitaux Paris (APHP); Hopital Universitaire Paul-Brousse - APHP;
   UDICE-French Research Universities; Universite Paris Cite; Hopital
   Universitaire Cochin - APHP; Hopital Universitaire Hotel-Dieu - APHP;
   Hopital Universitaire Necker-Enfants Malades - APHP; Hopital
   Universitaire Saint-Louis - APHP; Institut National de la Sante et de la
   Recherche Medicale (Inserm); UDICE-French Research Universities;
   Universite Paris Cite; UDICE-French Research Universities; Universite
   Paris Cite; UDICE-French Research Universities; Universite Paris Cite;
   Assistance Publique Hopitaux Paris (APHP); Hopital Universitaire
   Paul-Brousse - APHP; Hopital Universitaire Pitie-Salpetriere - APHP;
   UDICE-French Research Universities; Sorbonne Universite; Universite
   Paris Cite; Hopital Universitaire Cochin - APHP; Hopital Universitaire
   Hotel-Dieu - APHP; Hopital Universitaire Saint-Louis - APHP; Centre
   National de la Recherche Scientifique (CNRS); Institut National de la
   Sante et de la Recherche Medicale (Inserm); UDICE-French Research
   Universities; Sorbonne Universite; CEA; UDICE-French Research
   Universities; Universite Paris Saclay; Assistance Publique Hopitaux
   Paris (APHP); Hopital Universitaire Paul-Brousse - APHP; UDICE-French
   Research Universities; Universite Paris Cite; Hopital Universitaire
   Cochin - APHP; Hopital Universitaire Hotel-Dieu - APHP; Hopital
   Universitaire Necker-Enfants Malades - APHP; Hopital Universitaire
   Saint-Louis - APHP; UNICANCER; Gustave Roussy; Centre National de la
   Recherche Scientifique (CNRS); CNRS - National Institute for Biology
   (INSB); UDICE-French Research Universities; Universite Paris Saclay
RP Goya-Outi, J (通讯作者)，Univ Paris Sud, Univ Paris Saclay, CNRS, CEA,IMIV,INSERM, Orsay, France.
EM
RI Grill, Jacques/R-8093-2019; Buvat, Irene/U-8447-2018; boddaert,
   nathalie/GWV-2865-2022; Orlhac, Fanny/AAC-6428-2019; Frouin,
   Frédérique/N-7709-2015; Alentorn, Agustí/Q-3610-2018; PHILIPPE,
   Cathy/GRY-4381-2022
OI Buvat, Irene/0000-0002-7053-6471; Orlhac, Fanny/0000-0002-5588-1867;
   Frouin, Frédérique/0000-0002-7288-1543; Alentorn,
   Agustí/0000-0002-7220-2686; Goya Outi, Jessica/0000-0003-2031-5774;
   PHILIPPE, Cathy/0000-0002-3262-4390; Nioche,
   Christophe/0000-0002-6247-2025
FU 'Lidex-PIM' project - IDEX Paris-Saclay [ANR-11-IDEX-0003-02]
FX This work was partly supported by the 'Lidex-PIM' project funded by the
   IDEX Paris-Saclay, ANR-11-IDEX-0003-02.
CR Aerts HJWL, 2016, JAMA ONCOL, V2, P1636, DOI 10.1001/jamaoncol.2016.2631
   Ashburner J, 2005, NEUROIMAGE, V26, P839, DOI 10.1016/j.neuroimage.2005.02.018
   Avants BB, 2008, MED IMAGE ANAL, V12, P26, DOI 10.1016/j.media.2007.06.004
   Castel D, 2015, ACTA NEUROPATHOL, V130, P815, DOI 10.1007/s00401-015-1478-0
   Christensen JD, 2003, MAGN RESON IMAGING, V21, P817, DOI 10.1016/S0730-725X(03)00102-4
   Cohen KJ, 2017, NEURO-ONCOLOGY, V19, P1025, DOI 10.1093/neuonc/nox021
   Gevaert O, 2014, RADIOLOGY, V273, P168, DOI 10.1148/radiol.14131731
   Gillies RJ, 2016, RADIOLOGY, V278, P563, DOI 10.1148/radiol.2015151169
   HARALICK RM, 1973, IEEE T SYST MAN CYB, VSMC3, P610, DOI 10.1109/TSMC.1973.4309314
   Hatt M, 2015, J NUCL MED, V56, P38, DOI 10.2967/jnumed.114.144055
   Jenkinson M, 2002, NEUROIMAGE, V17, P825, DOI 10.1006/nimg.2002.1132
   Kickingereder P, 2016, RADIOLOGY, V280, P880, DOI 10.1148/radiol.2016160845
   Lambin P, 2012, EUR J CANCER, V48, P441, DOI 10.1016/j.ejca.2011.11.036
   Larroza A, 2016, TEXTURE ANAL MAGNETI, P75
   Leijenaar RTH, 2015, SCI REP-UK, V5, DOI 10.1038/srep11075
   Madabhushi A, 2006, MED PHYS, V33, P3426, DOI 10.1118/1.2335487
   Mayerhoefer ME, 2009, MED PHYS, V36, P1236, DOI 10.1118/1.3081408
   Michoux N, 2015, BMC CANCER, V15, DOI 10.1186/s12885-015-1563-8
   Molina D, 2016, BRIT J RADIOL, V89, P1064
   Molina D, 2017, PLOS ONE, V12, DOI 10.1371/journal.pone.0178843
   Nikulin MS, 2001, ENCY MATH
   Nyul LG, 2000, IEEE T MED IMAGING, V19, P143, DOI 10.1109/42.836373
   Orlhac F, 2015, PLOS ONE, V10, DOI 10.1371/journal.pone.0145063
   Orlhac F, 2014, J NUCL MED, V55, P414, DOI 10.2967/jnumed.113.129858
   Shinohara RT, 2014, NEUROIMAGE-CLIN, V6, P9, DOI 10.1016/j.nicl.2014.08.008
   Sikio M, 2015, INT SYMP IMAGE SIG, P149
   Skogen K, 2016, EUR J RADIOL, V85, P824, DOI 10.1016/j.ejrad.2016.01.013
   Tang XO, 1998, IEEE T IMAGE PROCESS, V7, P1602, DOI 10.1109/83.725367
   Thibault G, 2009, 10 INT C PATT REC IN
   Tustison NJ, 2010, IEEE T MED IMAGING, V29, P1310, DOI 10.1109/TMI.2010.2046908
   van Griethuysen JJM, 2017, CANCER RES, V77, pE104, DOI 10.1158/0008-5472.CAN-17-0339
   Yan JH, 2015, J NUCL MED, V56, P1667, DOI 10.2967/jnumed.115.156927
   Yang DL, 2015, MED PHYS, V42, P6725, DOI 10.1118/1.4934373
NR 33
TC 28
Z9 28
U1 1
U2 19
PU IOP PUBLISHING LTD
PI BRISTOL
PA TEMPLE CIRCUS, TEMPLE WAY, BRISTOL BS1 6BE, ENGLAND
SN 0031-9155
EI 1361-6560
J9 PHYS MED BIOL
JI Phys. Med. Biol.
PD MAY
PY 2018
VL 63
IS 10
AR 105003
DI 10.1088/1361-6560/aabd21
PG 12
WC Engineering, Biomedical; Radiology, Nuclear Medicine & Medical Imaging
WE Science Citation Index Expanded (SCI-EXPANDED)
SC Engineering; Radiology, Nuclear Medicine & Medical Imaging
GA GF4QY
UT WOS:000431949400003
PM 29633962
OA Green Published
DA 2023-08-27
ER

PT J
AU Choi, SK
   Rhee, BA
   Lim, YJ
AF Choi, Seok-Keun
   Rhee, Bong-Arm
   Lim, Young Jin
TI Hemifacial Spasm Caused by Epidermoid Tumor at Cerebello Pontine Angle
SO JOURNAL OF KOREAN NEUROSURGICAL SOCIETY
LA English
DT Article
DE Hemifacial spasm; Facial nerve; Epidermal cyst
ID NERVE
AB Hemifacial spasm (HFS) is almost always induced by vascular compression but in some cases the cause of HFS are tumors at cerebellopontine angle (CPA) or vascular malformations. We present a rare case of hemifacial spasm caused by epidermoid tumors and the possible pathogenesis of HFS is discussed. A 36-year-old female patient presented with a 27-month history of progressive involuntary facial twitching and had been treated with acupuncture and herb medication. On imaging study, a mass lesion was seen at right CPA. Microvascular decompression combined with mass removal was undertaken through retrosigmoid approach. The lesion was avascular mass and diagnosed with an epidermoid tumor pathologically. Eventually, we found a offending vessel (AICA : anterior inferior cerebellar artery) compressing facial nerve root exit zone (REZ). In case of HFS caused by tumor compression on the facial nerve REZ, surgeons should try to find an offending vessel under the mass. This case supports the vascular compression theory as a pathogenesis of HFS.
C1 [Choi, Seok-Keun; Rhee, Bong-Arm; Lim, Young Jin] Kyung Hee Univ, Dept Neurosurg, Sch Med, Seoul 130701, South Korea.
C3 Kyung Hee University
RP Choi, SK (通讯作者)，Kyung Hee Univ, Dept Neurosurg, Sch Med, Seoul 130701, South Korea.
EM
RI Lim, Young Jin/J-5621-2012
CR Chung SS, 2000, ACTA NEUROCHIR, V142, P901, DOI 10.1007/s007010070076
   CHUNG SY, 2001, J KOREAN NEUROSURG S, V30, P352
   Elgamal EA, 2005, NEUROSURG REV, V28, P330, DOI 10.1007/s10143-005-0392-7
   Ferroli P, 2005, NEUROL SCI, V26, P3, DOI 10.1007/s10072-005-0375-z
   Fukuda Masafumi, 1997, Neurologia Medico-Chirurgica, V37, P771, DOI 10.2176/nmc.37.771
   Galvez-Jimenez N, 2001, SEMIN NEUROL, V21, P75, DOI 10.1055/s-2001-13122
   Harada A, 2002, J NEUROSURG, V97, P482, DOI 10.3171/jns.2002.97.2.0482
   INOUE T, 1995, NEUROSURGERY, V36, P846, DOI 10.1227/00006123-199504000-00029
   Iwai Y, 2001, NEUROL MED-CHIR, V41, P87, DOI 10.2176/nmc.41.87
   JANNETTA PJ, 1984, NEUROSURGERY, V14, P89, DOI 10.1227/00006123-198401000-00021
   Kobata H, 2002, NEUROSURGERY, V50, P276, DOI 10.1097/00006123-200202000-00008
   NAGATA S, 1992, SURG NEUROL, V38, P204, DOI 10.1016/0090-3019(92)90170-R
   NISHI T, 1987, NEUROLOGY, V37, P339, DOI 10.1212/WNL.37.2.339
   NUSSBAUM M, 1977, ANN OTO RHINOL LARYN, V86, P73, DOI 10.1177/000348947708600111
   Revuelta R, 2003, CLIN NEUROL NEUROSUR, V106, P19, DOI 10.1016/j.clineuro.2003.07.001
   RHEE BA, 1995, NEUROSURGERY, V36, P393, DOI 10.1227/00006123-199502000-00021
   Sandberg DI, 1999, PEDIATR NEUROL, V21, P754, DOI 10.1016/S0887-8994(99)00077-6
   YEH HS, 1981, NEUROSURGERY, V9, P383, DOI 10.1227/00006123-198110000-00006
NR 18
TC 9
Z9 9
U1 0
U2 0
PU KOREAN NEUROSURGICAL SOC
PI SEOUL
PA BLDG B, #402, 27, CHILPAE-RO, JUNG-GU, SEOUL, 04511, SOUTH KOREA
SN 2005-3711
EI 1598-7876
J9 J KOREAN NEUROSURG S
JI J. Korean. Neurosurg. Soc.
PD MAR
PY 2009
VL 45
IS 3
BP 196
EP 198
DI 10.3340/jkns.2009.45.3.196
PG 3
WC Clinical Neurology; Surgery
WE Science Citation Index Expanded (SCI-EXPANDED)
SC Neurosciences & Neurology; Surgery
GA 430PD
UT WOS:000265000100013
PM 19352486
OA gold, Green Published
DA 2023-08-27
ER

PT J
AU Skarzynski, H
   Szuchnik, J
   Lorens, A
   Zawadzki, R
AF Skarzynski, H
   Szuchnik, J
   Lorens, A
   Zawadzki, R
TI First auditory brainstem implantation in Poland: auditory perception
   results over 12 months
SO JOURNAL OF LARYNGOLOGY AND OTOLOGY
LA English
DT Article; Proceedings Paper
CT 2nd International Auditory Brainstem Implant Symposium
CY APR 22-24, 1999
CL FREIBERG, GERMANY
DE brain stem; prosthesis implantation; neurofibromatosis 2; rehabilitation
   of hearing impaired
AB Auditory brainstem implants (ABIs) are a modern method of treatment of total bilateral deafness in cases of extracochlear origin. In most cases therapy is applied in patients with neurofibromatosis type 2 (NF2). This paper presents the results of surgical treatment and rehabilitation in a 28-year-old woman with bilateral, multiple tumours of the central nervous system causing total deafness. Simultaneous removal of the tumours and implantation of ABI allowed treatment of the potentially lethal pathology and hearing restoration. Improving auditory skills and excellent tests results were noted in the year following implantation.
C1 Inst Physiol & Pathol Hearing, Warsaw, Poland.
C3 Institute of Physiology & Hearing Pathology
RP Skarzynski, H (通讯作者)，Pstrowskiego 1, PL-01943 Warsaw, Poland.
RI Lorens, Artur/AAO-7553-2020; Skarzynski, Piotr H./A-9642-2012
OI Skarzynski, Piotr H./0000-0002-4978-1915; Skarzynski,
   Henryk/0000-0001-7141-9851
CR EDGERTON BJ, 1982, ANN OTO RHINOL LARYN, V91, P117
   LASZIG R, 1991, EUR ARCH OTO-RHINO-L, V248, P420, DOI 10.1007/BF01463568
   SKARZYNSKI H, 1998, MED SCI MONITOR S2, V4, P40
NR 3
TC 7
Z9 8
U1 0
U2 1
PU HEADLEY BROTHERS LTD
PI ASHFORD
PA INVICTA PRESS, ASHFORD, KENT, ENGLAND TN24 8HH
SN 0022-2151
J9 J LARYNGOL OTOL
JI J. Laryngol. Otol.
PD DEC
PY 2000
VL 114
SU 27
BP 44
EP 45
PG 2
WC Otorhinolaryngology
WE Conference Proceedings Citation Index - Science (CPCI-S); Science Citation Index Expanded (SCI-EXPANDED)
SC Otorhinolaryngology
GA 403LM
UT WOS:000167043400013
PM 11211438
DA 2023-08-27
ER

PT J
AU Chassot, A
   Canale, S
   Varlet, P
   Puget, S
   Roujeau, T
   Negretti, L
   Dhermain, F
   Rialland, X
   Raquin, MA
   Grill, J
   Dufour, C
AF Chassot, Andrea
   Canale, Sandra
   Varlet, Pascale
   Puget, Stephanie
   Roujeau, Thomas
   Negretti, Laura
   Dhermain, Frederic
   Rialland, Xavier
   Raquin, Marie Anne
   Grill, Jacques
   Dufour, Christelle
TI Radiotherapy with concurrent and adjuvant temozolomide in children with
   newly diagnosed diffuse intrinsic pontine glioma
SO JOURNAL OF NEURO-ONCOLOGY
LA English
DT Article
DE Brainstem glioma; Child; Radiation therapy; Temozolomide
ID BRAIN-STEM GLIOMAS; HYPERFRACTIONATED RADIATION-THERAPY;
   PEDIATRIC-ONCOLOGY-GROUP; TUMORS; MANAGEMENT; GY; CHEMOTHERAPY;
   CONCOMITANT; PATTERNS; FEATURES
AB The purpose of this study is to evaluate the efficacy and toxicity of radiation therapy (RT) with concurrent temozolomide (TMZ) chemotherapy followed by adjuvant TMZ in children with diffuse intrinsic pontine glioma (DIPG). Newly diagnosed patients younger than 18 years with histologically proven DIPG were treated with focal radiotherapy to a dose of 54 Gy in 30 fractions along with concurrent daily TMZ (75 mg/m(2)/day). Four weeks after completing the initial RT-TMZ schedule, adjuvant TMZ (200 mg/m(2)/day, days 1-5) was given every 28 days up to six cycles. Responses/progressions were assessed by clinical and 2-monthly MRI follow-up studies. Between September 2005 and September 2009, 21 patients with newly diagnosed histologically confirmed DIPG were eligible for this study. Median age at diagnosis was 6.4 years (range 4-16 years). At last update in August 2010, 17 children have died, 1 child was alive with progressive disease and 3 with stable disease. Metastatic relapse was documented in the cerebral site in two patients and in spinal cord in two cases. The median time to progression was 7.5 months (range 28 days-14.5 months) and the median survival was 11.7 months (range 26 days-17.5 months). The 1-year PFS and the 1-year OS were 33 and 50%, respectively. Five patients presented radiological findings compatible with pseudoprogression during the treatment. Haematological toxicity (Grade III/IV thrombocytopenia and leucopenia) was the most commonly found and led to dose reductions of TMZ in 58% of the patients. TMZ with radiation therapy has not yielded any significant improvement in outcome of children with DIPG and is associated with higher toxicity compared with radiotherapy alone. Novel treatment modalities are needed to improve the outcome of these patients.
C1 [Chassot, Andrea; Raquin, Marie Anne; Grill, Jacques; Dufour, Christelle] Inst Gustave Roussy, Dept Paediat & Adolescent Oncol, F-94800 Villejuif, France.
   [Canale, Sandra] Inst Gustave Roussy, Dept Radiol, Villejuif, France.
   [Varlet, Pascale] Hosp St Anne, Dept Pathol, Paris, France.
   [Puget, Stephanie; Roujeau, Thomas] Hosp Necker Enfants Malad, Dept Neurosurg, Paris, France.
   [Negretti, Laura; Dhermain, Frederic] Inst Gustave Roussy, Dept Radiat Oncol, Villejuif, France.
   [Rialland, Xavier] CHU Angers, Dept Paediat Oncol, Angers, France.
C3 UNICANCER; Gustave Roussy; UNICANCER; Gustave Roussy; UDICE-French
   Research Universities; Universite Paris Cite; GHU PARIS Psychiatrie
   Neurosciences; UDICE-French Research Universities; Universite Paris
   Cite; Assistance Publique Hopitaux Paris (APHP); Hopital Universitaire
   Necker-Enfants Malades - APHP; UNICANCER; Gustave Roussy; Universite
   d'Angers; Centre Hospitalier Universitaire d'Angers
RP Dufour, C (通讯作者)，Inst Gustave Roussy, Dept Paediat & Adolescent Oncol, F-94800 Villejuif, France.
EM
RI Grill, Jacques/R-8093-2019
OI VARLET, pascale/0000-0002-8164-1517
CR Bouffet E, 2000, CANCER, V88, P685, DOI 10.1002/(SICI)1097-0142(20000201)88:3<685::AID-CNCR27>3.0.CO;2-K
   Brandsma D, 2008, LANCET ONCOL, V9, P453, DOI 10.1016/S1470-2045(08)70125-6
   Broniscer A, 2005, CANCER-AM CANCER SOC, V103, P133, DOI 10.1002/cncr.20741
   Broniscer A, 2000, J CLIN ONCOL, V18, P1246, DOI 10.1200/JCO.2000.18.6.1246
   Chiang KL, 2010, CHILD NERV SYST, V26, P1035, DOI 10.1007/s00381-010-1106-1
   Cohen KJ, 2011, NEURO-ONCOLOGY, V13, P410, DOI 10.1093/neuonc/noq205
   Danson S J, 2001, Expert Rev Anticancer Ther, V1, P13, DOI 10.1586/14737140.1.1.13
   Donahue B, 1998, INT J RADIAT ONCOL, V40, P677, DOI 10.1016/S0360-3016(97)00846-8
   EDWARDS MSB, 1989, J NEUROSURG, V70, P691, DOI 10.3171/jns.1989.70.5.0691
   FREEMAN CR, 1993, INT J RADIAT ONCOL, V27, P197, DOI 10.1016/0360-3016(93)90228-N
   Freeman CR, 1998, INT J RADIAT ONCOL, V40, P265, DOI 10.1016/S0360-3016(97)00572-5
   Gururangan S, 2006, J NEURO-ONCOL, V77, P207, DOI 10.1007/s11060-005-9029-5
   Hargrove D, 2006, LANCET ONCOL, V7, P241, DOI 10.1016/S1470-2045(06)70615-5
   Jalali R, 2010, INT J RADIAT ONCOL, V77, P113, DOI 10.1016/j.ijrobp.2009.04.031
   KAPLAN EL, 1958, J AM STAT ASSOC, V53, P457, DOI 10.2307/2281868
   Korones DN, 2008, PEDIATR BLOOD CANCER, V50, P227, DOI 10.1002/pbc.21154
   KRETSCHMAR CS, 1993, CANCER-AM CANCER SOC, V72, P1404, DOI 10.1002/1097-0142(19930815)72:4<1404::AID-CNCR2820720441>3.0.CO;2-G
   LANGMOEN IA, 1991, CHILD NERV SYST, V7, P13, DOI 10.1007/BF00263825
   Lewis J, 1997, INT J RADIAT ONCOL, V38, P925, DOI 10.1016/S0360-3016(97)00134-X
   Louis DN, 2007, ACTA NEUROPATHOL, V114, P547, DOI 10.1007/s00401-007-0278-6
   Mandell LR, 1999, INT J RADIAT ONCOL, V43, P959, DOI 10.1016/S0360-3016(98)00501-X
   PACKER RJ, 1993, CANCER-AM CANCER SOC, V72, P1414, DOI 10.1002/1097-0142(19930815)72:4<1414::AID-CNCR2820720442>3.0.CO;2-C
   PACKER RJ, 1994, CANCER-AM CANCER SOC, V74, P1827, DOI 10.1002/1097-0142(19940915)74:6<1827::AID-CNCR2820740628>3.0.CO;2-Q
   PRADOS MD, 1995, INT J RADIAT ONCOL, V32, P85, DOI 10.1016/0360-3016(95)00563-E
   Qaddoumi I, 2009, CANCER-AM CANCER SOC, V115, P5761, DOI 10.1002/cncr.24663
   Roujeau T, 2007, J NEUROSURG, V107, P1, DOI 10.3171/PED-07/07/001
   Stupp R, 2002, J CLIN ONCOL, V20, P1375, DOI 10.1200/JCO.20.5.1375
   Stupp R, 2005, NEW ENGL J MED, V352, P987, DOI 10.1056/NEJMoa043330
   Wagner S, 2006, BRIT J CANCER, V95, P991, DOI 10.1038/sj.bjc.6603402
   Walter AW, 1998, MED PEDIATR ONCOL, V30, P28, DOI 10.1002/(SICI)1096-911X(199801)30:1<28::AID-MPO9>3.0.CO;2-2
NR 30
TC 77
Z9 80
U1 0
U2 14
PU SPRINGER
PI NEW YORK
PA ONE NEW YORK PLAZA, SUITE 4600, NEW YORK, NY, UNITED STATES
SN 0167-594X
EI 1573-7373
J9 J NEURO-ONCOL
JI J. Neuro-Oncol.
PD JAN
PY 2012
VL 106
IS 2
BP 399
EP 407
DI 10.1007/s11060-011-0681-7
PG 9
WC Oncology; Clinical Neurology
WE Science Citation Index Expanded (SCI-EXPANDED)
SC Oncology; Neurosciences & Neurology
GA 858KI
UT WOS:000297795200019
PM 21858607
DA 2023-08-27
ER

PT J
AU Tscherpel, C
   Dunkl, V
   Ceccon, G
   Stoffels, G
   Judov, N
   Rapp, M
   Meyer, PT
   Kops, ER
   Ermert, J
   Fink, GR
   Shah, NJ
   Langen, KJ
   Galldiks, N
AF Tscherpel, Caroline
   Dunkl, Veronika
   Ceccon, Garry
   Stoffels, Gabriele
   Judov, Natalie
   Rapp, Marion
   Meyer, Philipp T.
   Kops, Elena Rota
   Ermert, Johannes
   Fink, Gereon R.
   Shah, Nadim J.
   Langen, Karl-Josef
   Galldiks, Norbert
TI The use of O-(2-F-18-fluoroethyl)-L-tyrosine PET in the diagnosis of
   gliomas located in the brainstem and spinal cord
SO NEURO-ONCOLOGY
LA English
DT Article
DE amino acid PET; brainstem glioma; dynamic acquisition; FET;
   infratentorial
ID POSITRON-EMISSION-TOMOGRAPHY; STEREOTACTIC BIOPSY; MR-PET; TUMORS;
   NEUROONCOLOGY; GLITTERS; LESIONS; TOOL
AB Despite an increasing number of O-(2-F-18-fluoroethyl)-L-tyrosine (F-18-FET) PET studies in supratentorial gliomas, studies regarding the usefulness of F-18-FET PET in brainstem and spinal cord gliomas to date remain scarce.
   Thirty-six F-18-FET PET scans were performed in 29 patients with brainstem (n = 29 scans) or spinal cord glioma (n = 7 scans). In 32 of 36 PET scans, a dynamic acquisition was performed. Fifteen scans in 15 patients were performed to assess newly diagnosed lesions, and 21 scans were obtained during follow-up: for diagnosing tumor progression (n = 15 scans in 14 patients) as well as for treatment monitoring (n = 6 scans in 3 patients). Four patients underwent additional serial scans (range, 1-2), and 3 of these 4 patients were examined for more than one indication. Maximum and mean tumor/brain ratios (TBRmax/mean) of F-18-FET uptake (20-40 min post injection) as well as kinetic F-18-FET uptake parameters were determined. Final diagnoses were confirmed histologically (54%) or by clinical follow-up (46%).
   In all newly diagnosed high-grade (n = 3 patients) and in 5 of 11 patients with low-grade gliomas, F-18-FET uptake was increased (TBRmax a parts per thousand 2.5 and/or TBRmean a parts per thousand 1.9). In 2 patients with newly diagnosed gliomas without MR contrast enhancement, F-18-FET PET nevertheless showed increased metabolism. At suspected progression, the combination of TBRs with kinetic F-18-FET parameters correctly identified presence or absence of progressive disease in 9 of 11 patients (82%).
   This preliminary study suggests that F-18-FET PET adds valuable diagnostic information in brainstem and spinal cord glioma, particularly when the diagnostic information derived from MRI is equivocal.
C1 [Tscherpel, Caroline; Dunkl, Veronika; Ceccon, Garry; Fink, Gereon R.; Galldiks, Norbert] Univ Cologne, Dept Neurol, Cologne, Germany.
   [Tscherpel, Caroline; Stoffels, Gabriele; Judov, Natalie; Kops, Elena Rota; Ermert, Johannes; Fink, Gereon R.; Shah, Nadim J.; Langen, Karl-Josef; Galldiks, Norbert] Forschungszentrum Julich, Inst Neurosci & Med, D-52425 Julich, Germany.
   [Rapp, Marion] Univ Dusseldorf, Dept Neurosurg, Dusseldorf, Germany.
   [Meyer, Philipp T.] Univ Freiburg, Fac Med, Med Ctr, Dept Nucl Med, Freiburg, Germany.
   [Shah, Nadim J.] Univ Aachen, Dept Neurol, Aachen, Germany.
   [Langen, Karl-Josef] Univ Aachen, Dept Nucl Med, Aachen, Germany.
   [Shah, Nadim J.; Langen, Karl-Josef] JARA, Sect JARA Brain, Julich, Germany.
   [Galldiks, Norbert] Univ Cologne, CIO, Cologne, Germany.
   [Galldiks, Norbert] Univ Bonn, CIO, Bonn, Germany.
C3 University of Cologne; Helmholtz Association; Research Center Julich;
   Heinrich Heine University Dusseldorf; University of Freiburg; RWTH
   Aachen University; RWTH Aachen University; University of Cologne;
   University of Bonn
RP Galldiks, N (通讯作者)，Forschungszentrum Julich, Inst Neurosci & Med, D-52425 Julich, Germany.
EM
RI Fink, Gereon R./E-1616-2012; Fink, Gereon R./AFW-9377-2022; Galldiks,
   Norbert/AAG-6896-2019; Dunkl, Veronika/HZJ-9483-2023; Langen,
   Karl-Josef/H-4396-2013; Shah, N. Jon/H-5849-2013; Ermert,
   Johannes/H-9684-2013
OI Fink, Gereon R./0000-0002-8230-1856; Fink, Gereon
   R./0000-0002-8230-1856; Galldiks, Norbert/0000-0002-2485-1796; Langen,
   Karl-Josef/0000-0003-1101-5075; Shah, N. Jon/0000-0002-8151-6169;
   Ermert, Johannes/0000-0002-2561-7766; Rapp, Marion/0000-0003-2343-244X;
   Stoffels, Gabriele/0000-0001-7114-1941; Dunkl,
   Veronika/0000-0001-6885-7666; Judov, Natalie/0000-0002-3909-9306
CR Calcagni ML, 2011, CLIN NUCL MED, V36, P841, DOI 10.1097/RLU.0b013e3182291b40
   Chamberlain MC, 2011, CURR NEUROL NEUROSCI, V11, P320, DOI 10.1007/s11910-011-0190-2
   Galldiks N, 2015, NEURO-ONCOLOGY, V17, P1434, DOI 10.1093/neuonc/nov118
   Galldiks N, 2015, NEURO-ONCOLOGY, V17, P1293, DOI 10.1093/neuonc/nov088
   Galldiks N, 2015, EUR J NUCL MED MOL I, V42, P685, DOI 10.1007/s00259-014-2959-4
   Galldiks N, 2012, J NUCL MED, V53, P1048, DOI 10.2967/jnumed.111.098590
   Grimm SA, 2013, CURR NEUROL NEUROSCI, V13, DOI 10.1007/s11910-013-0346-3
   Grosu AL, 2011, INT J RADIAT ONCOL, V81, P1049, DOI 10.1016/j.ijrobp.2010.07.002
   Hamacher K, 2002, APPL RADIAT ISOTOPES, V57, P853, DOI 10.1016/S0969-8043(02)00225-7
   Herzog H, 2011, NUKLEARMED-NUCL MED, V50, P74, DOI 10.3413/Nukmed-0347-10-09
   HIGANO S, 1990, J COMPUT ASSIST TOMO, V14, P297, DOI 10.1097/00004728-199003000-00027
   Hutterer M, 2013, NEURO-ONCOLOGY, V15, P341, DOI 10.1093/neuonc/nos300
   Jacobs AH, 2005, J NUCL MED, V46, P1948
   Langen KJ, 2011, NUKLEARMED-NUCL MED, V50, P167, DOI 10.3413/nuk-2011041
   Langen KJ, 2013, NEURO-ONCOLOGY, V15, P816, DOI 10.1093/neuonc/not059
   Langen KJ, 2006, NUCL MED BIOL, V33, P287, DOI 10.1016/j.nucmedbio.2006.01.002
   Langen KJ, 2003, NUCL MED BIOL, V30, P501, DOI 10.1016/S0969-8051(03)00023-4
   Louis DN, 2007, ACTA NEUROPATHOL, V114, P547, DOI 10.1007/s00401-007-0278-6
   Massager N, 2000, J NEUROSURG, V93, P951, DOI 10.3171/jns.2000.93.6.0951
   Neuner I, 2012, EUR RADIOL, V22, P2568, DOI 10.1007/s00330-012-2543-x
   Parsa Andrew T, 2005, Clin Neurosurg, V52, P76
   Pauleit D, 2005, BRAIN, V128, P678, DOI 10.1093/brain/awh399
   Pirotte BJM, 2007, J NEUROSURG, V107, P392, DOI 10.3171/PED-07/11/392
   Purohit B, 2015, EUR J RADIOL, V84, P709, DOI 10.1016/j.ejrad.2014.12.025
   Rapp M, 2013, J NUCL MED, V54, P229, DOI 10.2967/jnumed.112.109603
   Reithmeier T, 2013, CLIN NEUROL NEUROSUR, V115, P165, DOI 10.1016/j.clineuro.2012.04.027
   Rosenfeld A, 2011, J PEDIAT HEMATOL ONC, V33, P369, DOI 10.1097/MPH.0b013e31820ad915
   Sasajima T, 1996, NEURORADIOLOGY, V38, P53, DOI 10.1007/BF00593221
   Shah NJ, 2013, J MAGN RESON, V229, P101, DOI 10.1016/j.jmr.2012.11.027
   Stupp R, 2005, NEW ENGL J MED, V352, P987, DOI 10.1056/NEJMoa043330
   Swissmedic, 2014, J SWISSMEDIC, V13, P651
   Tomura N, 2013, AM J NEURORADIOL, V34, P1278, DOI 10.3174/ajnr.A3374
   Weber WA, 2000, EUR J NUCL MED, V27, P542, DOI 10.1007/s002590050541
   Wen PY, 2010, J CLIN ONCOL, V28, P1963, DOI 10.1200/JCO.2009.26.3541
   Wester HJ, 1999, J NUCL MED, V40, P205
   Wilmshurst JM, 2000, J CHILD NEUROL, V15, P465, DOI 10.1177/088307380001500708
   Wong TZ, 2002, NEUROIMAG CLIN N AM, V12, P615, DOI 10.1016/S1052-5149(02)00033-3
   Yamaguchi S, 2009, J NEUROSURG-PEDIATR, V3, P270, DOI 10.3171/2008.12.PEDS08323
NR 38
TC 12
Z9 12
U1 0
U2 3
PU OXFORD UNIV PRESS INC
PI CARY
PA JOURNALS DEPT, 2001 EVANS RD, CARY, NC 27513 USA
SN 1522-8517
EI 1523-5866
J9 NEURO-ONCOLOGY
JI Neuro-Oncology
PD MAY
PY 2017
VL 19
IS 5
BP 710
EP 718
DI 10.1093/neuonc/now243
PG 9
WC Oncology; Clinical Neurology
WE Science Citation Index Expanded (SCI-EXPANDED)
SC Oncology; Neurosciences & Neurology
GA EU2YT
UT WOS:000400896500013
PM 28039366
OA Green Published, Bronze
DA 2023-08-27
ER

PT J
AU Buczkowicz, P
   Zarghooni, M
   Bartels, U
   Morrison, A
   Misuraca, KL
   Chan, T
   Bouffet, E
   Huang, A
   Becher, O
   Hawkins, C
AF Buczkowicz, Pawel
   Zarghooni, Maryam
   Bartels, Ute
   Morrison, Andrew
   Misuraca, Katherine L.
   Chan, Tiffany
   Bouffet, Eric
   Huang, Annie
   Becher, Oren
   Hawkins, Cynthia
TI Aurora Kinase B Is a Potential Therapeutic Target in Pediatric Diffuse
   Intrinsic Pontine Glioma
SO BRAIN PATHOLOGY
LA English
DT Article
DE astrocytoma; Aurora kinase; DIPG; glioma; pediatric
ID BRAIN-STEM GLIOMAS; GLIOBLASTOMA-MULTIFORME; CANCER-CELLS; EXPRESSION;
   INHIBITOR; CHECKPOINT; REVEALS; DISEASE
AB Pediatric high-grade astrocytomas (HGAs) account for 1520% of all pediatric central nervous system tumors. These neoplasms predominantly involve the supratentorial hemispheres or the ponsdiffuse intrinsic pontine gliomas (DIPG). Assumptions that pediatric HGAs are biologically similar to adult HGAs have recently been challenged, and the development of effective therapeutic modalities for DIPG and supratentorial HGA hinges on a better understanding of their biological properties. Here, 20 pediatric HGAs (9 DIPGs and 11 supratentorial HGAs) were subject to gene expression profiling following approval by the research ethics board at our institution. Many of these tumors showed expression signatures composed of genes that promote G1/S and G2/M cell cycle progression. In particular, Aurora kinase B (AURKB) was consistently and highly overexpressed in 6/9 DIPGs and 8/11 HGAs. Array data were validated using quantitative real-time PCR and immunohistochemistry, as well as cross-validation of our data set with previously published series. Inhibition of Aurora B activity in DIPG and in pediatric HGA cell lines resulted in growth arrest accompanied by morphological changes, cell cycle aberrations, nuclear fractionation and polyploidy as well as a reduction in colony formation. Our data highlight Aurora B as a potential therapeutic target in DIPG.
C1 [Buczkowicz, Pawel; Zarghooni, Maryam; Morrison, Andrew; Hawkins, Cynthia] Hosp Sick Children, Div Pathol, Toronto, ON M5G 1X8, Canada.
   [Bartels, Ute; Bouffet, Eric; Huang, Annie] Hosp Sick Children, Div Haematol & Oncol, Toronto, ON M5G 1X8, Canada.
   [Buczkowicz, Pawel; Zarghooni, Maryam; Morrison, Andrew; Chan, Tiffany; Huang, Annie; Hawkins, Cynthia] Hosp Sick Children, Arthur & Sonia Labatt Brain Tumour Res Ctr, Toronto, ON M5G 1X8, Canada.
   [Becher, Oren] Duke Univ, Med Ctr, Div Pediat Hematol Oncol, Durham, NC USA.
   [Misuraca, Katherine L.] Duke Univ, Dept Pharmacol & Canc Biol, Durham, NC USA.
   [Buczkowicz, Pawel; Chan, Tiffany; Huang, Annie; Hawkins, Cynthia] Univ Toronto, Fac Med, Dept Lab Med & Pathobiol, Toronto, ON, Canada.
C3 University of Toronto; Hospital for Sick Children (SickKids); University
   of Toronto; Hospital for Sick Children (SickKids); University of
   Toronto; Hospital for Sick Children (SickKids); Duke University; Duke
   University; University of Toronto
RP Hawkins, C (通讯作者)，Hosp Sick Children, Dept Paediat Lab Med, 555 Univ Ave, Toronto, ON M5G 1X8, Canada.
EM
RI Becher, Oren/AAI-7495-2020; Becher, Oren/GXN-3269-2022; Bouffet,
   Eric/AAB-8999-2020
OI Becher, Oren/0000-0003-4016-8492; Buczkowicz, Pawel/0000-0002-6469-1475
FU National Brain Tumor Society
FX We would like to thank the National Brain Tumor Society for providing
   funding for this research.
CR Aihara A, 2010, J HEPATOL, V52, P63, DOI 10.1016/j.jhep.2009.10.013
   Angelini P, 2011, J NEURO-ONCOL, V101, P75, DOI 10.1007/s11060-010-0224-7
   Bax DA, 2009, PLOS ONE, V4, DOI 10.1371/journal.pone.0005209
   Becher OJ, 2010, CANCER RES, V70, P2548, DOI 10.1158/0008-5472.CAN-09-2503
   Bischoff JR, 1998, EMBO J, V17, P3052, DOI 10.1093/emboj/17.11.3052
   Broniscer A, 2004, ONCOLOGIST, V9, P197, DOI 10.1634/theoncologist.9-2-197
   Chen YJ, 2009, VIRCHOWS ARCH, V455, P431, DOI 10.1007/s00428-009-0838-3
   Cheung CHA, 2009, EXPERT OPIN INV DRUG, V18, P379, DOI [10.1517/13543780902806392 , 10.1517/13543780902806392]
   D'Alise AM, 2008, MOL CANCER THER, V7, P1140, DOI 10.1158/1535-7163.MCT-07-2051
   Dar AA, 2010, MOL CANCER THER, V9, P268, DOI 10.1158/1535-7163.MCT-09-0765
   Ditchfield C, 2003, J CELL BIOL, V161, P267, DOI 10.1083/jcb.200208091
   Donaldson SS, 2006, J CLIN ONCOL, V24, P1266, DOI 10.1200/JCO.2005.04.6599
   Freeman CR, 1999, CHILD NERV SYST, V15, P545, DOI 10.1007/s003810050542
   Hargrove D, 2006, LANCET ONCOL, V7, P241, DOI 10.1016/S1470-2045(06)70615-5
   Harrington EA, 2004, NAT MED, V10, P262, DOI 10.1038/nm1003
   Hauf S, 2003, J CELL BIOL, V161, P281, DOI 10.1083/jcb.200208092
   Hodgson JG, 2009, NEURO-ONCOLOGY, V11, P477, DOI 10.1215/15228517-2008-113
   Hosoi H, 1998, MOL PHARMACOL, V54, P815, DOI 10.1124/mol.54.5.815
   Kitzen JJEM, 2010, CRIT REV ONCOL HEMAT, V73, P99, DOI 10.1016/j.critrevonc.2009.03.009
   Lens SMA, 2010, NAT REV CANCER, V10, P825, DOI 10.1038/nrc2964
   Li Y, 2010, AM J TRANSL RES, V2, P296
   Paugh BS, 2011, J CLIN ONCOL, V29, P3999, DOI 10.1200/JCO.2011.35.5677
   Paugh BS, 2010, J CLIN ONCOL, V28, P3061, DOI 10.1200/JCO.2009.26.7252
   Phillips HS, 2006, CANCER CELL, V9, P157, DOI 10.1016/j.ccr.2006.02.019
   Rafehi H, 2011, J VIS EXP, V13, P25731
   Rao B, 2010, ONCOTARGET, V1, P639
   Stolz A, 2009, CANCER RES, V69, P3874, DOI 10.1158/0008-5472.CAN-08-3597
   Valera ET, 2009, CHILD NERV SYST, V25, P39, DOI 10.1007/s00381-008-0740-3
   Zarghooni M, 2010, J CLIN ONCOL, V28, P1337, DOI 10.1200/JCO.2009.25.5463
   Zeng WFF, 2007, J CLIN PATHOL, V60, P218, DOI 10.1136/jcp.2006.036806
NR 30
TC 25
Z9 27
U1 0
U2 7
PU WILEY-BLACKWELL
PI HOBOKEN
PA 111 RIVER ST, HOBOKEN 07030-5774, NJ USA
SN 1015-6305
J9 BRAIN PATHOL
JI Brain Pathol.
PD MAY
PY 2013
VL 23
IS 3
BP 244
EP 253
DI 10.1111/j.1750-3639.2012.00633.x
PG 10
WC Clinical Neurology; Neurosciences; Pathology
WE Science Citation Index Expanded (SCI-EXPANDED)
SC Neurosciences & Neurology; Pathology
GA 129TP
UT WOS:000317865800002
PM 22971244
OA Green Published
DA 2023-08-27
ER

PT J
AU Saeger, W
AF Saeger, W
TI Histopathological classification of adrenal tumours
SO EUROPEAN JOURNAL OF CLINICAL INVESTIGATION
LA English
DT Article; Proceedings Paper
CT Adrenal 2000 Conference - From Molecular Basics to New Clinical
   Dimensions
CY 2000
CL Toronto, CANADA
DE adrenal tumours; histopathology
ID HUMAN ADRENOCORTICAL NEOPLASMS; DIFFERENTIAL-DIAGNOSIS; CORTICAL
   CARCINOMA; TUMORS; IMMUNOHISTOCHEMISTRY; EXPRESSION; HYBRIDIZATION;
   ADENOMAS; MARKERS; LESIONS
AB Tumours in the adrenals originate from the adrenal cortex, the adrenal medulla or as metastases from extra-adrenal primaries. Differentiating between these three groups is the first task a pathologist has to tackle when dealing with specimens from the adrenal region. Whereas this is possible in every case with total removal of the adrenal tumour it may be impossible in fine needle biopsies of such tumours. The second great problem is the dignity of adrenal rumours, which cannot be determined in many adrenomedullary and some adrenocortical rumours. Immunostainings are helpful but the basic method remains the histopathological examination of paraffin sections. This review gives an update of pathological findings in several adrenal tumour entities, and provides guidelines for the diagnosis of these rumours in the light of recently published data.
C1 Marien Hosp, Inst Pathol, D-22087 Hamburg, Germany.
C3 St. Marien Hospital
RP Saeger, W (通讯作者)，Marien Hosp, Inst Pathol, Alfredstr 9, D-22087 Hamburg, Germany.
EM
CR ABRAMS HL, 1950, CANCER, V3, P74, DOI 10.1002/1097-0142(1950)3:1<74::AID-CNCR2820030111>3.0.CO;2-7
   BODIE B, 1989, J UROLOGY, V141, P257, DOI 10.1016/S0022-5347(17)40734-8
   Busam KJ, 1998, AM J SURG PATHOL, V22, P57, DOI 10.1097/00000478-199801000-00007
   GICQUEL C, 1994, CLIN ENDOCRINOL, V40, P465, DOI 10.1111/j.1365-2265.1994.tb02485.x
   GOLDBLUM JR, 1993, MODERN PATHOL, V6, P663
   HENLEY DJ, 1983, SURGERY, V94, P926
   HOUGH AJ, 1979, AM J CLIN PATHOL, V72, P390
   Iino K, 1997, MODERN PATHOL, V10, P901
   KOMMINOTH P, 1995, LAB INVEST, V72, P424
   Lack EE., 1997, TUMORS ADRENAL GLAND
   Li Q, 1998, APMIS, V106, P807, DOI 10.1111/j.1699-0463.1998.tb00227.x
   LUTON JP, 1990, NEW ENGL J MED, V322, P1195, DOI 10.1056/NEJM199004263221705
   MARX C, 1998, J CLIN ENDOCR METAB, V81, P4488
   McNicol AM, 1997, J PATHOL, V181, P146
   NAITO M, 1991, J PEDIATR SURG, V26, P834, DOI 10.1016/0022-3468(91)90150-R
   Neville A. M., 1982, HUMAN ADRENAL CORTEX
   Reininger AJ, 1998, PLATELETS, V9, P241, DOI 10.1080/09537109876771
   Saeger W, 1995, Minerva Endocrinol, V20, P1
   Saeger W, 1998, ENDOCR PATHOL, V9, P235, DOI 10.1007/BF02739963
   Saeger W., 1993, Zentralblatt fuer Pathologie, V139, P157
   SASANO H, 1994, MODERN PATHOL, V7, P741
   SASANO H, 1993, HUM PATHOL, V24, P397, DOI 10.1016/0046-8177(93)90088-X
   Sautner D, 1990, UROL B, V30, P44
   SCHRODER S, 1995, PATHOLOGE, V16, P307, DOI 10.1007/s002920050107
   SCHRODER S, 1990, VIRCHOWS ARCH A, V417, P89, DOI 10.1007/BF02190525
   SUZUKI T, 1992, MODERN PATHOL, V5, P224
   Vargas MP, 1997, AM J SURG PATHOL, V21, P556, DOI 10.1097/00000478-199705000-00008
   VONSLOOTEN H, 1985, CANCER, V55, P766
   WEISS LM, 1984, AM J SURG PATHOL, V8, P163, DOI 10.1097/00000478-198403000-00001
NR 29
TC 28
Z9 28
U1 0
U2 7
PU WILEY
PI HOBOKEN
PA 111 RIVER ST, HOBOKEN 07030-5774, NJ USA
SN 0014-2972
EI 1365-2362
J9 EUR J CLIN INVEST
JI Eur. J. Clin. Invest.
PD DEC
PY 2000
VL 30
SU 3
BP 58
EP 62
DI 10.1046/j.1365-2362.2000.0300s3058.x
PG 5
WC Medicine, General & Internal; Medicine, Research & Experimental
WE Science Citation Index Expanded (SCI-EXPANDED); Conference Proceedings Citation Index - Science (CPCI-S)
SC General & Internal Medicine; Research & Experimental Medicine
GA 405RD
UT WOS:000167172200013
PM 11281370
DA 2023-08-27
ER

PT J
AU Nakada, T
   Sasagawa, I
   Kubota, Y
   Suzuki, H
   Ishigooka, M
   Watanabe, M
AF Nakada, T
   Sasagawa, I
   Kubota, Y
   Suzuki, H
   Ishigooka, M
   Watanabe, M
TI Dihydroxyphenylglycol in pheochromocytoma: Its diagnostic use for
   norepinephrine dominant tumor
SO JOURNAL OF UROLOGY
LA English
DT Article
DE adrenal medulla; pheochromocytoma; norepinephrine; epinephrine
ID PLASMA DIHYDROXYPHENYLGLYCOL; NEURONAL REMOVAL;
   3,4-DIHYDROXYPHENYLGLYCOL; EPINEPHRINE; METABOLISM; URINARY; ASSAY
AB Purpose: To discern whether the 3,4-dihydroxyphenylglycol produced in adrenal medulla is associated with altered urinary excretion, we compared the tissue and urinary levels of this catecholamine metabolite, epinephrine and norepinephrine in 23 patients with pheochromocytoma and 26 subjects with renal cell carcinoma.
   Materials and Methods: Tissue and urine concentrations of dihydroxyphenylglycol, epinephrine and norepinephrine were determined by catechol-O-methyl-transferase-based radioenzymatic method and high performance liquid chromatography with electrochemical detection.
   Results: Contents of dihydroxyphenylglycol, epinephrine and norepinephrine in pheochromocytomas were higher than those in normal adrenal medullae. Removal of pheochromocytomas lowered urinary excretion of these catecholamines and dihydroxyphenylglycol. Norepinephrine content correlated closely with dihydroxyphenylglycol content in normal adrenal medullae and pheochromocytomas but not with epinephrine content in either tissue. Contents of norepinephrine and dihydroxyphenylglycol correlated significantly with urinary norepinephrine excretion and dihydroxyphenylglycol excretion in patients with pheochromocytoma but not in subjects with renal cell carcinoma. The norepinephrine-to-dihydroxyphenyl glycol ratio in a 24-hour preoperative urine collection from patients with pheochromocytoma was higher than that from subjects with renal cell carcinoma, which was almost similar to that of epinephrine dominant pheochromocytomas.
   Conclusions: Our study suggests that dihydroxyphenylglycol in urine in subjects with renal cell carcinoma is predominantly of a neuronal origin and urinary dihydroxyphenylglycol in patients with pheochromocytoma is of tumor origin. A high level of norepinephrine-to-dihydroxyphenyl glycol ratio in urine can be used to determine the diagnosis of the norepinephrine dominant type pheochromocytoma. Measurement of at least a few markers is preferable for precise biochemical diagnosis of pheochromocytoma.
RP Nakada, T (通讯作者)，YAMAGATA UNIV,SCH MED,DEPT UROL,YAMAGATA 99023,JAPAN.
CR BRAVO EL, 1991, KIDNEY INT, V40, P544, DOI 10.1038/ki.1991.244
   BROWN MJ, 1984, EUR J CLIN INVEST, V14, P67, DOI 10.1111/j.1365-2362.1984.tb00706.x
   DEQUATTRO V, 1979, ENDOCRINOLOGY, V2, P1261
   DUNCAN MW, 1988, NEW ENGL J MED, V319, P136, DOI 10.1056/NEJM198807213190303
   EISENHOFER G, 1990, J CLIN ENDOCR METAB, V70, P1710, DOI 10.1210/jcem-70-6-1710
   ESLER M, 1981, CLIN PHARMACOL THER, V29, P12, DOI 10.1038/clpt.1981.3
   GOLDSTEIN DS, 1988, J CLIN INVEST, V81, P213, DOI 10.1172/JCI113298
   GOLDSTEIN DS, 1985, J CLIN INVEST, V76, P15, DOI 10.1172/JCI111939
   HOFFMANN JJML, 1986, CLIN CHIM ACTA, V156, P221, DOI 10.1016/0009-8981(86)90156-7
   HOFFMANN JJML, 1982, CLIN CHIM ACTA, V125, P319, DOI 10.1016/0009-8981(82)90263-7
   HORTNAGL H, 1983, KLIN WOCHENSCHR, V61, P601, DOI 10.1007/BF01487338
   IZZO JL, 1985, LIFE SCI, V37, P1033, DOI 10.1016/0024-3205(85)90593-4
   LENDERS JWM, 1992, AM J MED, V92, P147, DOI 10.1016/0002-9343(92)90105-K
   NAKADA T, 1989, J UROLOGY, V141, P998, DOI 10.1016/S0022-5347(17)41085-8
   NAKADA T, 1988, J UROLOGY, V140, P1348, DOI 10.1016/S0022-5347(17)42041-6
   PLOUIN PF, 1981, BRIT MED J, V282, P853, DOI 10.1136/bmj.282.6267.853
   SMYTHE GA, 1992, CLIN CHEM, V38, P486
   WAGNER J, 1982, J NEUROCHEM, V38, P1241, DOI 10.1111/j.1471-4159.1982.tb07897.x
   YAMAGUCHI T, 1992, EXPERIENTIA, V48, P667, DOI 10.1007/BF02118315
NR 19
TC 3
Z9 3
U1 0
U2 0
PU WILLIAMS & WILKINS
PI BALTIMORE
PA 351 WEST CAMDEN ST, BALTIMORE, MD 21201-2436
SN 0022-5347
J9 J UROLOGY
JI J. Urol.
PD JAN
PY 1996
VL 155
IS 1
BP 14
EP 18
DI 10.1016/S0022-5347(01)66526-1
PG 5
WC Urology & Nephrology
WE Science Citation Index Expanded (SCI-EXPANDED)
SC Urology & Nephrology
GA TJ434
UT WOS:A1996TJ43400004
PM 7490813
DA 2023-08-27
ER

PT J
AU Rameh, V
   Vajapeyam, S
   Ziaei, A
   Kao, P
   London, WB
   Baker, SJ
   Chiang, J
   Lucas, J
   Tinkle, CL
   Wright, KD
   Poussaint, TY
AF Rameh, V.
   Vajapeyam, S.
   Ziaei, A.
   Kao, P.
   London, W. B.
   Baker, S. J.
   Chiang, J.
   Lucas, J.
   Tinkle, C. L.
   Wright, K. D.
   Poussaint, T. Y.
TI Correlation between Multiparametric MR Imaging and Molecular Genetics in
   Pontine Pediatric High-Grade Glioma
SO AMERICAN JOURNAL OF NEURORADIOLOGY
LA English
DT Article
ID DIFFUSE MIDLINE GLIOMAS; CENTRAL-NERVOUS-SYSTEM; APPARENT DIFFUSION;
   TUMORS; SURVIVAL; FEATURES; CLASSIFICATION; MUTATIONS; SUBGROUPS; BIOPSY
AB BACKGROUND AND PURPOSE: Molecular profiling is a crucial feature in the "integrated diagnosis" of CNS tumors. We aimed to determine whether radiomics could distinguish molecular types of pontine pediatric high-grade gliomas that have similar/overlapping phenotypes on conventional anatomic MR images.MATERIALS AND METHODS: Baseline MR images from children with pontine pediatric high-grade gliomas were analyzed. Retrospective imaging studies included standard precontrast and postcontrast sequences and DTI. Imaging analyses included median, mean, mode, skewness, and kurtosis of the ADC histogram of the tumor volume based on T2 FLAIR and enhancement at baseline. Histone H3 mutations were identified through immunohistochemistry and/or Sanger or next-generation DNA sequencing. The log-rank test identified imaging factors prognostic of survival from the time of diagnosis. Wilcoxon rank-sum and Fisher exact tests compared imaging predictors among groups.RESULTS: Eighty-three patients had pretreatment MR imaging and evaluable tissue sampling. The median age was 6 years (range, 0.7-17 years); 50 tumors had a K27M mutation in H3-3A, and 11, in H3C2/3. Seven tumors had histone H3 K27 alteration, but the specific gene was unknown. Fifteen were H3 wild-type. Overall survival was significantly higher in H3C2/3-compared with H3-3A -mutant tumors (P = .003) and in wild-type tumors compared with any histone mutation (P = .001). Lower overall survival was observed in patients with enhancing tumors (P = .02) compared with those without enhancement. H3C2/3-mutant tumors showed higher mean, median, and mode ADC_total values (P<.001) and ADC_enhancement (P<.004), with lower ADC_total skewness and kurtosis (P<.003) relative to H3-3A-mutant tumors. CONCLUSIONS: ADC histogram parameters are correlated with histone H3 mutation status in pontine pediatric high-grade glioma.
C1 [Rameh, V.; Vajapeyam, S.; Ziaei, A.; Poussaint, T. Y.] Harvard Med Sch, Boston Childrens Hosp, Dept Radiol, Boston, MA USA.
   [Kao, P.; London, W. B.; Wright, K. D.] Harvard Med Sch, Dana Farber Canc Inst, Dept Pediat Oncol, Boston, MA USA.
   [Baker, S. J.] St Jude Childrens Res Hosp, Dept Dev Neurobiol, Memphis, TN USA.
   [Chiang, J.] St Jude Childrens Res Hosp, Dept Pathol, Memphis, TN USA.
   [Chiang, J.; Tinkle, C. L.] St Jude Childrens Res Hosp, Dept Radiat Oncol, Memphis, TN USA.
   [Poussaint, T. Y.] Boston Childrens Hosp, Dept Radiol, 300 Longwood Ave, Boston, MA 02115 USA.
C3 Harvard University; Boston Children's Hospital; Harvard Medical School;
   Harvard University; Dana-Farber Cancer Institute; Harvard Medical
   School; St Jude Children's Research Hospital; St Jude Children's
   Research Hospital; St Jude Children's Research Hospital; Harvard
   University; Boston Children's Hospital
RP Poussaint, TY (通讯作者)，Boston Childrens Hosp, Dept Radiol, 300 Longwood Ave, Boston, MA 02115 USA.
EM
RI Lucas, John T./K-5243-2013; Chiang, Jason/N-2766-2018; Baker, Suzanne
   J/N-8167-2018
OI Lucas, John T./0000-0002-4139-4160; Chiang, Jason/0000-0003-3623-8996;
   Kao, Pei-Chi/0000-0003-0702-8324; Vajapeyam,
   Sridhar/0000-0002-1782-1919; Ziaei, Alireza/0000-0003-1005-8997; Wright,
   Karen/0000-0002-9578-1545; Baker, Suzanne/0000-0002-5833-472X; Rameh,
   vanessa/0000-0002-6007-7216
FU American Lebanese Syrian Associated Charities; National Cancer Institute
   [P30 CA021765, P01 CA096832]
FX This work was supported, in part, by the American Lebanese Syrian
   Associated Charities and the National Cancer Institute grant P30
   CA021765 (St. Jude Cancer Center support grant) and P01 CA096832.
CR Aboian MS, 2019, AM J NEURORADIOL, V40, P1804, DOI 10.3174/ajnr.A6302
   Aboian MS, 2017, AM J NEURORADIOL, V38, P795, DOI 10.3174/ajnr.A5076
   ALBRIGHT AL, 1993, NEUROSURGERY, V33, P1026
   Azad TD, 2020, NEUROSURG FOCUS, V48, DOI 10.3171/2019.9.FOCUS19699
   BARKOVICH AJ, 1991, PEDIATR NEUROSURG, V16, P73, DOI 10.1159/000120511
   Buczkowicz P, 2015, FRONT ONCOL, V5, DOI 10.3389/fonc.2015.00147
   Calmon R, 2021, EUR RADIOL, V31, P8913, DOI 10.1007/s00330-021-07991-x
   Castel D, 2020, ACTA NEUROPATHOL, V139, P1109, DOI 10.1007/s00401-020-02142-w
   Castel D, 2016, ACTA NEUROPATHOL, V131, P795, DOI 10.1007/s00401-016-1568-7
   Castel D, 2015, ACTA NEUROPATHOL, V130, P815, DOI 10.1007/s00401-015-1478-0
   Chen HJ, 2010, AM J NEURORADIOL, V31, P1879, DOI 10.3174/ajnr.A2179
   Chenevert TL, 2009, NEUROIMAG CLIN N AM, V19, P559, DOI 10.1016/j.nic.2009.08.009
   Chiang J, 2020, ACTA NEUROPATHOL COM, V8, DOI 10.1186/s40478-020-00930-9
   Khuong-Quang DA, 2012, ACTA NEUROPATHOL, V124, P439, DOI 10.1007/s00401-012-0998-0
   Gauvain KM, 2001, AM J ROENTGENOL, V177, P449, DOI 10.2214/ajr.177.2.1770449
   Giagnacovo M, 2020, CHILD NERV SYST, V36, P697, DOI 10.1007/s00381-019-04463-y
   Gupta N, 2018, NEURO-ONCOLOGY, V20, P1547, DOI 10.1093/neuonc/noy070
   Hoffman LM, 2018, J CLIN ONCOL, V36, P1963, DOI 10.1200/JCO.2017.75.9308
   Jaimes C, 2020, AM J NEURORADIOL, V41, P874, DOI 10.3174/ajnr.A6546
   Jenkinson M, 2012, NEUROIMAGE, V62, P782, DOI 10.1016/j.neuroimage.2011.09.015
   Kfoury-Beaumont N, 2022, BMC BIOL, V20, DOI 10.1186/s12915-022-01324-0
   Khalid F, 2023, FRONT MED-LAUSANNE, V10, DOI 10.3389/fmed.2023.1071447
   Koral K, 2013, RADIOLOGY, V268, P532, DOI 10.1148/radiol.13121362
   Lapin DH, 2017, FRONT ONCOL, V7, DOI 10.3389/fonc.2017.00057
   Leach JL, 2020, NEURO-ONCOLOGY, V22, P1647, DOI 10.1093/neuonc/noaa140
   Louis DN, 2021, NEURO-ONCOLOGY, V23, P1231, DOI 10.1093/neuonc/noab106
   Louis DN, 2016, ACTA NEUROPATHOL, V131, P803, DOI 10.1007/s00401-016-1545-1
   Lu VM, 2019, J NEUROSURG-PEDIATR, V24, P593, DOI 10.3171/2019.6.PEDS19259
   Mackay A, 2017, CANCER CELL, V32, P520, DOI 10.1016/j.ccell.2017.08.017
   Merhemic Z, 2018, EUR J RADIOL, V108, P249, DOI 10.1016/j.ejrad.2018.10.010
   Mulkern RV, 2015, ACAD RADIOL, V22, P363, DOI 10.1016/j.acra.2014.10.006
   Ostrom QT, 2020, NEURO-ONCOLOGY, V22, P1, DOI 10.1093/neuonc/noaa200
   Piccardo A, 2019, EUR J NUCL MED MOL I, V46, P1685, DOI 10.1007/s00259-019-04333-4
   Poussaint TY, 2016, NEURO-ONCOLOGY, V18, P725, DOI 10.1093/neuonc/nov256
   Puget S, 2015, CHILD NERV SYST, V31, P1773, DOI 10.1007/s00381-015-2832-1
   Schindelin J, 2012, NAT METHODS, V9, P676, DOI [10.1038/NMETH.2019, 10.1038/nmeth.2019]
   Varlet P., 2021, WHO CLASSIFICATION T, V5th, P69
   Wagner MW, 2023, CAN ASSOC RADIOL J, V74, P119, DOI 10.1177/08465371221109921
   Wu G, 2014, NAT GENET, V46, P444, DOI 10.1038/ng.2938
   Wu G, 2012, NAT GENET, V44, P251, DOI 10.1038/ng.1102
   Zhang P, 2023, FRONT ONCOL, V13, DOI 10.3389/fonc.2023.1007393
NR 41
TC 0
Z9 0
U1 0
U2 0
PU AMER SOC NEURORADIOLOGY
PI DENVILLE
PA PO BOX 3000, DENVILLE, NJ 07834-9349 USA
SN 0195-6108
EI 1936-959X
J9 AM J NEURORADIOL
JI Am. J. Neuroradiol.
PD JUL
PY 2023
VL 44
IS 7
BP 833
EP 840
DI 10.3174/ajnr.A7910
PG 8
WC Clinical Neurology; Neuroimaging; Radiology, Nuclear Medicine & Medical
   Imaging
WE Science Citation Index Expanded (SCI-EXPANDED)
SC Neurosciences & Neurology; Radiology, Nuclear Medicine & Medical Imaging
GA L6MQ3
UT WOS:001024387800008
PM 37321859
OA hybrid
DA 2023-08-27
ER

PT J
AU Sun, Y
   Sun, Y
   Yan, K
   Li, ZX
   Xu, C
   Geng, YB
   Pan, CC
   Chen, X
   Zhang, LW
   Xi, QR
AF Sun, Yu
   Sun, Ye
   Yan, Kun
   Li, Zhuxuan
   Xu, Cheng
   Geng, Yibo
   Pan, Changcun
   Chen, Xin
   Zhang, Liwei
   Xi, Qiaoran
TI Potent anti-tumor efficacy of palbociclib in treatment-naive
   H3.3K27M-mutant diffuse intrinsic pontine glioma
SO EBIOMEDICINE
LA English
DT Article
DE DIPG; Cell cycle; Palbocidib; H3.3-K27 M mutation
ID DEPENDENT KINASE 4/6; PEDIATRIC HIGH-GRADE; PHARMACOLOGICAL INHIBITION;
   CDK4/6 INHIBITION; ACQUIRED-RESISTANCE; THERAPEUTIC TARGET; CANCER;
   GLIOBLASTOMA; GROWTH; CELL
AB Background: Diffuse intrinsic pontine glioma (DIPG) is a rare and fatal pediatric brain cancer without cure. Seeking therapeutic strategies is still a major challenge in DIPG research. Previous study has shown that dysregulation of G1/S cell cycle checkpoint was common in DIPG and this dysregulation is even more enriched in the H3.3K27 M mutant subgroup. Here we assess potential anti-tumor efficacy of palbociclib, a specific and cytostalic inhibitor of CDK4/6, on high grade H3.3-K27 M-mutant DIPGs in vitro and in vivo.
   Methods: We established patient-derived cell lines from treatment-naive specimens. All the lines have H3.3K27 M mutation. We used a range of biological in vitro assays to assess the effect of palbociclib on growth of DIPGs. Palbociclib activity was also assayed in vivo against three independent DIPG orthotropic xenografts model.
   Findings: Dysregulation of G1/S cell cycle checkpoint is enriched in these DIPGs. Then, we showed that depletion of CDK4 or CDK6 inhibits DIPG cells growth and blocks G1/S transition. Furthermore, palbocidib effectively repressed DIPG growth in vitro. Transcriptome analysis showed that palbociclib not only blocks G1/S transition, it also blocks other oncogenic targets such as MYC. Finally, palbocidib activity was assayed in vivo against DIPG orthotropic xenografts to demonstrate the high efficiency of blocking tumor growth.
   Interpretation: Our findings thus revealed that palbocidib could be the therapeutic strategy for treatment-naive DIPG with H3.3K27 M mutation. (C) 2019 The Authors. Published by Elsevier B.V.
C1 [Sun, Yu; Xu, Cheng; Geng, Yibo; Pan, Changcun; Chen, Xin; Zhang, Liwei] Capital Med Univ, Beijing Tiantan Hosp, Dept Neurosurg, Beijing 100070, Peoples R China.
   [Sun, Ye; Li, Zhuxuan; Xi, Qiaoran] Tsinghua Univ, Sch Life Sci, MOE Key Lab Prot Sci, Beijing 100084, Peoples R China.
   [Yan, Kun] Tsinghua Univ, Sch Life Sci, Tsinghua Peking Ctr Life Sci, Beijing 100084, Peoples R China.
C3 Capital Medical University; Tsinghua University; Tsinghua University
RP Zhang, LW (通讯作者)，Capital Med Univ, Beijing Tiantan Hosp, Dept Neurosurg, Beijing 100070, Peoples R China.; Xi, QR (通讯作者)，Tsinghua Univ, Sch Life Sci, MOE Key Lab Prot Sci, Beijing 100084, Peoples R China.
EM
OI Yan, Kun/0000-0002-1738-9708; Sun, Yu/0000-0002-3568-2238
FU Beijing municipal administration of Hospitals Clinical Medicine
   Development of Special Funding Support [ZYLX201608]; National Natural
   Science Foundation of China [31471229, 31771622, 91540108, 81872048];
   Center for Life Sciences; Ministry of Science and Technology of China
   [2018YFA0107702]; Beijing Municipal Natural Science Foundation [7161004]
FX Liwei Zhang is supported by Beijing municipal administration of
   Hospitals Clinical Medicine Development of Special Funding Support
   (ZYLX201608) to L.Z. Beijing Municipal Natural Science Foundation
   (7161004) to L.Z. and National Natural Science Foundation of China
   (81872048) to L.Z. Qiaoran Xi is supported by National Natural Science
   Foundation of China (31471229, 31771622, and 91540108) to Q.X., Center
   for Life Sciences (to Q.X.), and Ministry of Science and Technology of
   China (2018YFA0107702) to Q.X.
CR Anders L, 2011, CANCER CELL, V20, P620, DOI 10.1016/j.ccr.2011.10.001
   Asby DJ, 2018, CANCER MANAG RES, V10, P3483, DOI 10.2147/CMAR.S167095
   Barton KL, 2013, PLOS ONE, V8, DOI 10.1371/journal.pone.0077639
   Baughn LB, 2006, CANCER RES, V66, P7661, DOI 10.1158/0008-5472.CAN-06-1098
   Beaver JA, 2015, CLIN CANCER RES, V21, P4760, DOI 10.1158/1078-0432.CCR-15-1185
   Bender S, 2013, CANCER CELL, V24, P660, DOI 10.1016/j.ccr.2013.10.006
   Buczkowicz P, 2014, NAT GENET, V46, P451, DOI 10.1038/ng.2936
   Cen L, 2012, NEURO-ONCOLOGY, V14, P870, DOI 10.1093/neuonc/nos114
   Cordero FJ, 2017, MOL CANCER RES, V15, P1243, DOI 10.1158/1541-7786.MCR-16-0389
   Dean JL, 2010, ONCOGENE, V29, P4018, DOI 10.1038/onc.2010.154
   Khuong-Quang DA, 2012, ACTA NEUROPATHOL, V124, P439, DOI 10.1007/s00401-012-0998-0
   Eilers G, 2015, MOL CANCER THER, V14, P1346, DOI 10.1158/1535-7163.MCT-14-0793
   Finn RS, 2009, BREAST CANCER RES, V11, DOI 10.1186/bcr2419
   Freeman CR, 1999, CHILD NERV SYST, V15, P545, DOI 10.1007/s003810050542
   Fry DW, 2004, MOL CANCER THER, V3, P1427
   Garber K, 2014, SCIENCE, V345, P865, DOI 10.1126/science.345.6199.865
   Goel S, 2017, NATURE, V548, P471, DOI 10.1038/nature23465
   Grasso CS, 2015, NAT MED, V21, P555, DOI 10.1038/nm.3855
   Grobner SN, 2018, NATURE, V555, P321, DOI 10.1038/nature25480
   Hashizume R, 2014, NAT MED, V20, P1394, DOI 10.1038/nm.3716
   Hashizume R, 2012, J NEURO-ONCOL, V110, P305, DOI 10.1007/s11060-012-0973-6
   Herrera-Abreu MT, 2016, CANCER RES, V76, P2301, DOI 10.1158/0008-5472.CAN-15-0728
   Huang DW, 2009, NAT PROTOC, V4, P44, DOI 10.1038/nprot.2008.211
   Jones C, 2017, NEURO-ONCOLOGY, V19, P153, DOI 10.1093/neuonc/now101
   Kovatcheva M, 2015, ONCOTARGET, V6, P8226, DOI 10.18632/oncotarget.3364
   Liu SL, 2018, INVEST NEW DRUG, V36, P961, DOI 10.1007/s10637-018-0575-z
   Mackay A, 2017, CANCER CELL, V32, P520, DOI 10.1016/j.ccell.2017.08.017
   MARIA BL, 1993, J CHILD NEUROL, V8, P112, DOI 10.1177/088307389300800203
   Michaud K, 2010, CANCER RES, V70, P3228, DOI 10.1158/0008-5472.CAN-09-4559
   Mohammad F, 2017, NAT MED, V23, P483, DOI 10.1038/nm.4293
   Monje M, 2011, P NATL ACAD SCI USA, V108, P4453, DOI 10.1073/pnas.1101657108
   Mount CW, 2018, NAT MED, V24, P572, DOI 10.1038/s41591-018-0006-x
   Nagaraja S, 2017, CANCER CELL, V31, P635, DOI 10.1016/j.ccell.2017.03.011
   Nemoto A, 2016, MOL CANCER THER, V15, P94, DOI 10.1158/1535-7163.MCT-14-1065
   O'Leary B, 2016, NAT REV CLIN ONCOL, V13, P417, DOI 10.1038/nrclinonc.2016.26
   Olmez I, 2017, CLIN CANCER RES, V23, P6958, DOI 10.1158/1078-0432.CCR-17-0803
   Otto T, 2017, NAT REV CANCER, V17, P93, DOI 10.1038/nrc.2016.138
   Paugh BS, 2011, J CLIN ONCOL, V29, P3999, DOI 10.1200/JCO.2011.35.5677
   Piunti A, 2017, NAT MED, V23, P493, DOI 10.1038/nm.4296
   Root DE, 2006, NAT METHODS, V3, P715, DOI 10.1038/NMETH924
   Saab R, 2006, MOL CANCER THER, V5, P1299, DOI 10.1158/1535-7163.MCT-05-0383
   Sangar MLC, 2017, CLIN CANCER RES, V23, P5802, DOI 10.1158/1078-0432.CCR-16-2943
   Sapkota G, 2007, MOL CELL, V25, P441, DOI 10.1016/j.molcel.2007.01.006
   Shechter D, 2007, NAT PROTOC, V2, P1445, DOI 10.1038/nprot.2007.202
   Subramanian A, 2005, P NATL ACAD SCI USA, V102, P15545, DOI 10.1073/pnas.0506580102
   Sun Y, 2019, NEUROSCI BULL, V35, P216, DOI 10.1007/s12264-018-00329-6
   Taylor IC, 2015, J NEUROPATH EXP NEUR, V74, P778, DOI 10.1097/NEN.0000000000000216
   Taylor KR, 2014, NAT GENET, V46, P457, DOI 10.1038/ng.2925
   Thangavel C, 2011, ENDOCR-RELAT CANCER, V18, P333, DOI 10.1530/ERC-10-0262
   Vanan MI, 2015, FRONT ONCOL, V5, DOI 10.3389/fonc.2015.00237
   Veringa SJE, 2013, PLOS ONE, V8, DOI 10.1371/journal.pone.0061512
   Wang L, 2007, BLOOD, V110, P2075, DOI 10.1182/blood-2007-02-071266
   Wiedemeyer WR, 2010, P NATL ACAD SCI USA, V107, P11501, DOI 10.1073/pnas.1001613107
   Wu YLD, 2017, TRANSL ONCOL, V10, P221, DOI 10.1016/j.tranon.2016.12.008
   Xu C, 2017, ONCOTARGET, V8, P76644, DOI 10.18632/oncotarget.19656
   Zhang N, 2016, AM J CANCER RES, V6, P97
   Zheng HQ, 2017, CANCER CELL, V32, P731, DOI 10.1016/j.ccell.2017.11.002
NR 57
TC 19
Z9 19
U1 0
U2 8
PU ELSEVIER SCIENCE BV
PI AMSTERDAM
PA PO BOX 211, 1000 AE AMSTERDAM, NETHERLANDS
SN 2352-3964
J9 EBIOMEDICINE
JI EBioMedicine
PD MAY
PY 2019
VL 43
BP 171
EP 179
DI 10.1016/j.ebiom.2019.04.043
PG 9
WC Medicine, General & Internal; Medicine, Research & Experimental
WE Science Citation Index Expanded (SCI-EXPANDED)
SC General & Internal Medicine; Research & Experimental Medicine
GA IB2HZ
UT WOS:000470091600030
PM 31060906
OA Green Published, gold
DA 2023-08-27
ER

PT J
AU Rosen, A
   Anderson, V
   Bercovici, E
   Laperriere, N
   D'Souza, R
AF Rosen, Adam
   Anderson, Valerie
   Bercovici, Eduard
   Laperriere, Normand
   D'Souza, Rohan
TI Brainstem gliomas in pregnancy: a systematic review dagger
SO JOURNAL OF MATERNAL-FETAL & NEONATAL MEDICINE
LA English
DT Article
DE Brain stem; glioma; neoplasms; pregnancy; pregnancy complications
ID PROGNOSTIC-FACTORS; CELL-GROWTH; ASTROCYTOMA; TUMOR; MANAGEMENT;
   RADIOTHERAPY; INCREASES
AB Introduction:Although brainstem gliomas are a rare group of neoplasias, when they affect pregnant women, there can be challenges with diagnosis and management. This study describes a case of brainstem glioma diagnosed in pregnancy and systematically reviews the literature on brainstem gliomas in pregnancy to provide guidance for management. Material and methods:We searched five databases from inception until October 2016 using subject headings and keywords related to pregnancy and brainstem glioma, and included original research articles that described pregnancy outcomes in women with brainstem glioma. Data extraction and quality assessment using the Joanna Briggs Institute Critical Appraisal Checklist for case reports were performed in duplicate. Outcomes were reported as proportions. The study protocol was registered with the Prospero International Prospective Register of Systematic Reviews (CRD42017060196). Results:We screened 2737 titles and abstracts, and 89 full-texts. Twelve articles describing 17 pregnancies in 16 women were included in the analysis. The median gestational age at presentation was 23 weeks. All but one case presented with neurologic deficit. Magnetic resonance (MRI) imaging conclusively diagnosed all cases. Surgical tumor resection (n = 4) and radiation therapy (n = 3) were successfully undertaken during pregnancy. There were no reported sequelae of maternal oncological management on neonatal wellbeing. Maternal mortality was high (8/16, 50%) both during (n = 5) and within 4 weeks (n = 3) of pregnancy. Pregnancy losses included one pregnancy termination and four miscarriages (associated with maternal mortality). Of the 12 live-born babies, five were premature. Two of these were the result of spontaneous preterm labor and three were delivered prematurely to facilitate glioma management. There was one case of fetal growth restriction. Conclusions:Although the symptoms of brainstem gliomas often mimic those commonly encountered in pregnancy, neurologic deficits warrant urgent investigation. MRI is the diagnostic modality of choice in pregnancy. Brainstem gliomas are associated with high maternal mortality and appropriate management, including surgical tumor resection and radiation therapy, should not be delayed on account of pregnancy. Pregnancy outcomes are favorable although there is a risk of preterm birth.Key message Brainstem gliomas are associated with high maternal mortality and timely diagnosis using magnetic resonance imaging and treatment including surgical resection and radiation therapy should not be delayed during pregnancy. Pregnancy outcomes are generally favorable except for risk of preterm birth.
C1 [Rosen, Adam; D'Souza, Rohan] Univ Toronto, Mt Sinai Hosp, Dept Obstet & Gynaecol, Div Maternal & Fetal Med, Toronto, ON, Canada.
   [Rosen, Adam] Univ Toronto, Fac Med, Toronto, ON, Canada.
   [Anderson, Valerie] Univ Ottawa, Fac Med, Ottawa, ON, Canada.
   [Bercovici, Eduard] Univ Toronto, Toronto Western Hosp, Krembil Neurosci Ctr, Dept Med,Div Neurol, Toronto, ON, Canada.
   [Laperriere, Normand] Univ Toronto, Princess Margaret Canc Ctr, Dept Radiat Oncol, Toronto, ON, Canada.
C3 University of Toronto; Sinai Health System Toronto; Lunenfeld Tanenbaum
   Research Institute; University of Toronto; University of Ottawa; Krembil
   Research Institute; University of Toronto; University Health Network
   Toronto; University of Toronto; University Health Network Toronto;
   Princess Margaret Cancer Centre
RP D'Souza, R (通讯作者)，Mt Sinai Hosp, Dept Obstet & Gynaecol, Div Maternal Fetal Med, Room 3-908,700 Univ Ave, Toronto, ON, Canada.
EM
RI D'Souza, Rohan/F-7997-2016
OI D'Souza, Rohan/0000-0002-4049-2017; Laperriere,
   Normand/0000-0001-8930-9650
CR Aromataris E, 2020, JBI MANUAL EVIDENCE
   ATANASSOFF PG, 1994, CAN J ANAESTH, V41, P163, DOI 10.1007/BF03009818
   Blumenthal DT, 2008, CANCER-AM CANCER SOC, V113, P3349, DOI 10.1002/cncr.23973
   Cohen-Gadol AA, 2009, J NEUROSURG, V111, P2009, DOI 10.3171/2009.3.JNS081160
   Dey M, 2014, J CLIN NEUROSCI, V21, P1298, DOI 10.1016/j.jocn.2013.12.011
   Eisele SC, 2016, CANCER-AM CANCER SOC, V122, P2799, DOI 10.1002/cncr.29920
   Gonzalez-Aguero G, 2007, ENDOCRINE, V32, P129, DOI 10.1007/s12020-007-9023-0
   Gonzalez-Arenas A, 2012, BBA-MOL CELL RES, V1823, P379, DOI 10.1016/j.bbamcr.2011.11.004
   HAAS JF, 1986, J NEUROL NEUROSUR PS, V49, P874, DOI 10.1136/jnnp.49.8.874
   Hu J, 2016, FRONT ONCOL, V6, DOI 10.3389/fonc.2016.00180
   Isla A, 1997, OBSTET GYNECOL, V89, P19, DOI 10.1016/S0029-7844(96)00381-X
   Kal HB, 2005, LANCET ONCOL, V6, P328, DOI 10.1016/S1470-2045(05)70169-8
   Kesari S, 2008, J NEURO-ONCOL, V88, P175, DOI 10.1007/s11060-008-9545-1
   Liberati A, 2009, ANN INTERN MED, V151, pW65, DOI [10.1371/journal.pmed.1000097, 10.7326/0003-4819-151-4-200908180-00136, 10.1136/bmj.b2700, 10.1136/bmj.b4037]
   MARX GF, 1964, OBSTET GYNECOL, V24, P122
   Mehta VS, 2009, NEUROL INDIA, V57, P274, DOI 10.4103/0028-3886.53272
   MICHELSEN JJ, 1969, J NEUROL NEUROSUR PS, V32, P305, DOI 10.1136/jnnp.32.4.305
   Mishanina E, 2014, CAN MED ASSOC J, V186, P665, DOI 10.1503/cmaj.130925
   Murji A, 2015, CAN MED ASSOC J, V187, P1309, DOI 10.1503/cmaj.140901
   OPESKIN K, 1995, AM J FOREN MED PATH, V16, P168, DOI 10.1097/00000433-199506000-00020
   Pallud J, 2010, ANN NEUROL, V67, P398, DOI 10.1002/ana.21888
   RAND CW, 1950, ARCH NEURO PSYCHIATR, V63, P1
   Reyes-Botero G, 2012, ONCOLOGIST, V17, P388, DOI 10.1634/theoncologist.2011-0335
   Riffaud L, 2006, OBSTET GYNECOL, V108, P801, DOI 10.1097/01.AOG.0000214676.16825.85
   Rosen A, 2017, DIAGNOSIS MANAGEMENT
   Ryskeldiyev N, 2016, J NEUROL SURG REP, V77, pE86, DOI 10.1055/s-0036-1583311
   Salmaggi A, 2008, J NEUROL, V255, P171, DOI 10.1007/s00415-008-0589-0
   Scarrott LJ, 2012, J OBSTET GYNAECOL, V32, P704, DOI 10.3109/01443615.2012.697226
   Selvapandian S, 1999, ACTA NEUROCHIR, V141, P721, DOI 10.1007/s007010050367
   SNEED PK, 1995, INT J RADIAT ONCOL, V32, P823, DOI 10.1016/0360-3016(94)00456-U
   Sterne JAC, 2011, BMJ-BRIT MED J, V343, DOI 10.1136/bmj.d4002
   Tewari KS, 2000, AM J OBSTET GYNECOL, V182, P1215, DOI 10.1016/S0002-9378(00)70188-8
   Umehara T, 2016, MOL CLIN ONCOL, V4, P567, DOI 10.3892/mco.2016.773
NR 33
TC 1
Z9 1
U1 0
U2 1
PU TAYLOR & FRANCIS LTD
PI ABINGDON
PA 2-4 PARK SQUARE, MILTON PARK, ABINGDON OR14 4RN, OXON, ENGLAND
SN 1476-7058
EI 1476-4954
J9 J MATERN-FETAL NEO M
JI J. Matern.-Fetal Neonatal Med.
PD AUG 17
PY 2020
VL 33
IS 16
BP 2841
EP 2851
DI 10.1080/14767058.2018.1560410
PG 11
WC Obstetrics & Gynecology
WE Science Citation Index Expanded (SCI-EXPANDED)
SC Obstetrics & Gynecology
GA ME6ZI
UT WOS:000544802600021
PM 30563399
OA Green Submitted
DA 2023-08-27
ER

PT J
AU Ramsden, R
   Khwaja, S
   Green, K
   O'Driscoll, M
   Mawman, T
AF Ramsden, R
   Khwaja, S
   Green, K
   O'Driscoll, M
   Mawman, T
TI Vestibular schwannoma in the only hearing ear: Cochlear implant or
   auditory brainstem implant?
SO OTOLOGY & NEUROTOLOGY
LA English
DT Article
DE auditory brainstem implant; cochlear implant; plasticity; vestibular
   schwannoma
ID NEURONS; DEATH
AB Objective: To explore the dilemma faced by neurotologists confronted with the patient who develops a vestibular schwannoma in the only hearing ear, the other having been deaf from birth, and to consider the choice between auditory rehabilitation using a cochlear implant (CI) on the congenitally deaf side and an auditory brainstem implant (ABI) on the tumor side. Study
   Design: A record review of two patients born deaf in one ear and who developed a vestibular schwannoma in the contra lateral ear, who then received a CI in the congenitally deaf ear.
   Setting: Tertiary referral center with special experience in vestibular schwannoma surgery, neurofibromatosis type 2 management, and cochlear implantation.
   Results: Neither patient was a good Cl user. At I year postimplant, they both scored 0% on abbreviated words and 0% and 7%, respectively, on Bench Kowal Bamford sentences. They scored 54% and 57%, respectively, on City-University of New York sentences with lip-reading, and both had fair access to environmental sound scoring at 45% each.
   Conclusions: The results from cochlear implantation in the congenitally deaf ear in these patients were poor and suggest that stimulus deprivation in the early stages of the maturation of the auditory pathways is important even for a unilateral hearing loss. Unfortunately, the factors that predict a good ABI result are not known. In these circumstances, the authors advocate the insertion of an ABI at the time of tumor removal, retaining the option of CI in the congenitally deaf ear in the event of a poor outcome with the ABI.
C1 Manchester Royal Infirm, Dept Otolaryngol, Manchester M13 9WL, Lancs, England.
   Manchester Royal Infirm, Dept Audiol, Manchester M13 9WL, Lancs, England.
   Univ Manchester, Manchester Adult Cochlear Implant Programme, Manchester, Lancs, England.
C3 University of Manchester; University of Manchester; University of
   Manchester
RP Khwaja, S (通讯作者)，74 Kerscott Rd, Manchester M23 OFN, Lancs, England.
EM
OI Khwaja, Sadie/0000-0001-7129-8789
CR Clark GM, 1999, AM J OTOL, V20, P4
   Govaerts PJ, 2002, OTOL NEUROTOL, V23, P885, DOI 10.1097/00129492-200211000-00013
   Lenarz M, 2002, OTOL NEUROTOL, V23, P694, DOI 10.1097/00129492-200209000-00015
   Mostafapour SP, 2002, J NEUROSCI, V22, P4670, DOI 10.1523/JNEUROSCI.22-11-04670.2002
   Tierney TS, 1997, J COMP NEUROL, V378, P295, DOI 10.1002/(SICI)1096-9861(19970210)378:2<295::AID-CNE11>3.0.CO;2-R
NR 5
TC 23
Z9 26
U1 0
U2 1
PU LIPPINCOTT WILLIAMS & WILKINS
PI PHILADELPHIA
PA 530 WALNUT ST, PHILADELPHIA, PA 19106-3261 USA
SN 1531-7129
J9 OTOL NEUROTOL
JI Otol. Neurotol.
PD MAR
PY 2005
VL 26
IS 2
BP 261
EP 264
DI 10.1097/00129492-200503000-00023
PG 4
WC Clinical Neurology; Otorhinolaryngology
WE Science Citation Index Expanded (SCI-EXPANDED)
SC Neurosciences & Neurology; Otorhinolaryngology
GA 957ZI
UT WOS:000231411700023
PM 15793416
DA 2023-08-27
ER

PT J
AU Karadag, A
   Senoglu, M
   Middlebrooks, EH
   Kinali, B
   Guvencer, M
   Icke, C
   Sayhan, S
   Karabay, N
   Camlar, M
   Olomu, OU
   Tanriover, N
AF Karadag, Ali
   Senoglu, Mehmet
   Middlebrooks, Erik H.
   Kinali, Burak
   Guvencer, Mustafa
   Icke, Cigdem
   Sayhan, Salih
   Karabay, Nuri
   Camlar, Mahmut
   Olomu, Osarenoma U.
   Tanriover, Necmettin
TI Endoscopic endonasal transclival approach to the ventral brainstem:
   Radiologic, anatomic feasibility and nuances, surgical limitations and
   future directions
SO JOURNAL OF CLINICAL NEUROSCIENCE
LA English
DT Article
DE Endonasal endoscopic surgery; Ventral; Brainstem; Vascular; Tumor
ID SAFE ENTRY ZONES; CAVERNOUS MALFORMATIONS; CEREBELLOPONTINE ANGLE;
   MICROSURGICAL ANATOMY; TRANSNASAL APPROACH; VERTEBRAL ARTERY; SPHENOID
   SINUS; SKULL BASE; RESECTION; ANEURYSM
AB The endoscopic endonasal transclival approach (EETA) is less invasive than traditional open approaches; however, there is currently limited data on the pre-operative features that may predict success of the EETA. Surgical landmarks and exposure of the EETA and expanded EETA are assessed. We retrospectively measured multiple anatomic features in 154 adult dry skulls, 22 C1 and C2 dry bone vertebrae, and 500 paranasal sinus computed tomography (CT) scans. We also dissected 13 formalin-fixed, silicone-injected adult cadaveric heads. Relevant qualitative and quantitative data were obtained with CT (n = 10) and cadaveric dissection (n = 13). They are expressed as mean (SD), as appropriate and compared with unpaired t tests. Categorical variables were compared with chi(2) tests. We determined the prevalence of clival and sphenoid sinus anatomic variations and assessed potential exposure of the craniovertebral region, based on the relative position of the nasal bone tip, posterior point of the hard palate, and their relation to the position of C1 and C2. Depending on anatomic features, EETA should allow visualization of the ventral brainstem. The mean (SD) dimensions of the surgical window for EETA were 43.2 (5.1) x 18.3 (1.1) mm, in the craniocaudad and horizontal dimensions, respectively. The craniocaudad dimension enlarged to 60.2 (4.7) mm with expanded EETA. The EETA allowed satisfactory exposure for odontoid and C1 anterior arch resection (expanded EETA) in all specimens, regardless of the orientation of the palate. The combination of preoperative radiographic assessment and intraoperative considerations allows safe and effective application and facilitate selection of the most appropriate approach. (C) 2020 Elsevier Ltd. All rights reserved.
C1 [Karadag, Ali; Senoglu, Mehmet; Kinali, Burak; Camlar, Mahmut] Hlth Sci Univ, Tepecik Res & Training Hosp, Dept Neurosurg, Izmir, Turkey.
   [Middlebrooks, Erik H.] Mayo Clin, Div Neuroradiol, Jacksonville, FL 32224 USA.
   [Middlebrooks, Erik H.] Mayo Clin, Dept Neurol Surg, Jacksonville, FL 32224 USA.
   [Guvencer, Mustafa; Icke, Cigdem; Sayhan, Salih] Dokuz Eylul Univ, Fac Med, Dept Anat, Izmir, Turkey.
   [Karabay, Nuri] Dokuz Eylul Univ, Fac Med, Dept Radiol, Izmir, Turkey.
   [Olomu, Osarenoma U.] Mayo Clin, Dept Otorhinolaryngol Audiol, Jacksonville, FL 32224 USA.
   [Tanriover, Necmettin] Istanbul Univ, Cerrahpasa Fac Med, Dept Neurosurg, Istanbul, Turkey.
C3 Izmir Tepecik Training & Research Hospital; University of Health
   Sciences Turkey; Mayo Clinic; Mayo Clinic; Dokuz Eylul University; Dokuz
   Eylul University; Mayo Clinic; Istanbul University; Istanbul University
   - Cerrahpasa
RP Senoglu, M (通讯作者)，Tepecik Res & Training Hosp, Izmir, Turkey.
EM
RI Olomu, Osarenoma/AAA-2479-2022; Senoglu, Şenoğlu /
   Prof.Mehmet/AAV-3867-2020; TANRIOVER, NECMETTIN/HGC-5383-2022;
   Tanrıöver, Necmettin/AAD-3212-2020; Güvençer, Mustafa/AAV-5277-2021
OI Tanrıöver, Necmettin/0000-0001-7628-9443; Middlebrooks,
   Erik/0000-0002-4418-9605
CR Aktas U, 2013, J CRANIO MAXILL SURG, V41, P457, DOI 10.1016/j.jcms.2012.11.011
   Alikhani P, 2017, BR J NEUROSURG, P1
   Almefty R, 2014, J NEUROSURG, V120, P40, DOI 10.3171/2013.8.JNS13535
   Benet A, 2014, WORLD NEUROSURG, V81, P385, DOI 10.1016/j.wneu.2013.01.091
   Bozkurt B, 2018, WORLD NEUROSURG, V111, P311, DOI 10.1016/j.wneu.2017.12.064
   Cavalcanti DD, 2016, J NEUROSURG, V124, P1359, DOI 10.3171/2015.4.JNS141945
   Cavallo Luigi M, 2005, Neurosurg Focus, V19, pE2
   Dalian I, 2015, INT J PEDIATR OTORHI, V79, P1584, DOI 10.1016/j.ijporl.2015.06.024
   Deopujari CE, 2014, WORLD NEUROSURG, V82, pS49, DOI 10.1016/j.wneu.2014.07.025
   Drazin D, 2012, J CLIN NEUROSCI, V19, P144, DOI 10.1016/j.jocn.2011.07.013
   Duntze J, 2014, ORTHOP TRAUMATOL-SUR, V100, P775, DOI 10.1016/j.otsr.2014.07.017
   El-Sayed IH, 2010, J CRANIOVERTEBRAL JU, V1, P44, DOI 10.4103/0974-8237.65481
   Eloy JA, 2008, NEUROSURGERY, V62, P142
   Eloy Jean A, 2008, Neurosurgery, V62, P142, DOI 10.1227/01.neu.0000317385.91432.df
   Ensenat J, 2011, NEUROSURGERY, V69, pE121, DOI 10.1227/NEU.0b013e318223b637
   Fernandez-Miranda JC, 2012, NEUROSURGERY, V146-58, P71
   Fernandez-Miranda JC, 2012, NEUROSURGERY, V71, P146, DOI 10.1227/NEU.0b013e3182438915
   Fomichev D, 2017, NEUROSURGERY, V97, pe7
   Froelich S, 2011, NEUROSURGERY, V68, P310, DOI 10.1227/NEU.0b013e3182117063
   Funaki T, 2013, NEUROSURGERY, V73, P155, DOI 10.1227/01.neu.0000431469.82215.93
   Gardner PA, 2015, WORLD NEUROSURG, V84, P1380, DOI 10.1016/j.wneu.2015.06.032
   Goldschlager T, 2015, J NEUROSURG, V122, P511, DOI 10.3171/2014.9.JNS14733
   Gonzalez-Quarante LH, 2016, WORLD NEUROSURG, V91, pe5
   GRANICK MS, 1985, ANN OTO RHINOL LARYN, V94, P34, DOI 10.1177/000348948509400108
   Gunaldi O, 2016, TURK NEUROSURG
   Har-El G, 2005, ANN OTO RHINOL LARYN, V114, P439, DOI 10.1177/000348940511400605
   He SM, 2016, WORLD NEUROSURG, V90, DOI 10.1016/j.wneu.2016.02.064
   Jean WC, 2016, CUREUS J MED SCIENCE, V8, DOI 10.7759/cureus.641
   Jurinovic Pavao, 2016, Acta Inform Med, V24, P74, DOI 10.5455/aim.2016.24.74-77
   Kassam AB, 2005, NEUROSURGERY, V57, P213, DOI 10.1227/01.NEU.0000163687.64774.E4
   Kassam Amin, 2005, Neurosurg Focus, V19, pE4
   Kassam AB, 2006, NEUROSURGERY, V59, P162
   Kooshkabadi A, 2015, NEUROSURGERY, V77, P888, DOI 10.1227/NEU.0000000000000922
   Koutourousiou M, 2012, NEUROSURGERY, V71, P614, DOI 10.1227/NEU.0b013e31825ea3e0
   Labib MA, 2014, NEUROSURGERY S3, V10, P71
   Labib MA, 2014, OPER NEUROSURG, V10, P448, DOI 10.1227/NEU.0000000000000362
   Lega BC, 2011, SKULL BASE-INTERD AP, V21, P65, DOI 10.1055/s-0030-1265825
   Linsler S, 2015, WORLD NEUROSURG, V84, P2064, DOI 10.1016/j.wneu.2015.08.029
   Gomez-Amador JL, 2017, J NEUROSURG, V127, P553, DOI 10.3171/2016.8.JNS161137
   Maroon Joseph C, 2005, Neurosurg Focus, V19, pE1, DOI 10.3171/foc.2005.19.1.2
   Messina A, 2007, NEUROSURG REV, V30, P189, DOI 10.1007/s10143-007-0084-6
   Morales-Valero SF, 2015, NEUROSURG FOCUS, V38, DOI 10.3171/2015.1.FOCUS14831
   Morera VA, 2010, NEUROSURGERY, V66, pONS211, DOI 10.1227/01.NEU.0000369926.01891.5D
   Mukherjee D, 2016, NEUROSURGERY S1, V63, P205
   Porter RW, 1999, J NEUROSURG, V90, P50, DOI 10.3171/jns.1999.90.1.0050
   Ramina R, 2011, NEUROL SCI, V32, P1013, DOI 10.1007/s10072-011-0477-8
   Recalde RJ, 2008, NEUROSURGERY, V63, P9, DOI [10.1227/01.NEU.0000297062.52433.3F, 10.1227/01.neu.0000317368.69523.40]
   Rhoton Jr AL, 2002, NEUROSURGERY, pS341
   Rhoton Jr AL, 2002, NEUROSURGERY, pS359
   Saito K, 2012, ACTA NEUROCHIR, V154, P879, DOI 10.1007/s00701-012-1317-1
   Sanmillan JL, 2016, WORLD NEUROSURG, V89, P368, DOI 10.1016/j.wneu.2016.01.081
   Seker A, 2010, WORLD NEUROSURG, V74, P583, DOI 10.1016/j.wneu.2010.06.033
   Vellutini EDS, 2014, WORLD NEUROSURG, V82, pS106, DOI 10.1016/j.wneu.2014.07.031
   Singh H, 2013, J NEUROL SURG PART B, V74, P176, DOI 10.1055/s-0033-1342923
   Somanna S, 2015, ACTA NEUROCHIR, V157, P2077, DOI 10.1007/s00701-015-2610-6
   Tubbs RS, 2016, CHILD NERV SYST, V32, P55, DOI 10.1007/s00381-015-2864-6
   VINCHON M, 1995, NEUROSURGERY, V36, P52, DOI 10.1227/00006123-199501000-00006
   Vinchon M, 1995, NEUROSURGERY, V36, P52
   Wang J, 2010, NEUROSURGERY, V66, P797, DOI 10.1227/01.NEU.0000367619.24800.B1
   Yagmurlu K, 2014, NEUROSURGERY S4, V10, P19
   Yagmurlu K, 2014, OPER NEUROSURG, V10, P602, DOI 10.1227/NEU.0000000000000466
   Yamada M, 2003, SURG NEUROL, V60, P431, DOI 10.1016/S0090-3019(03)00324-0
   Zhang S, 2017, J CLIN NEUROSCI, V37, P63, DOI 10.1016/j.jocn.2016.11.014
   Zwagerman NT, 2016, J NEURO-ONCOL, V130, P319, DOI 10.1007/s11060-016-2274-y
NR 64
TC 5
Z9 5
U1 0
U2 3
PU ELSEVIER SCI LTD
PI OXFORD
PA THE BOULEVARD, LANGFORD LANE, KIDLINGTON, OXFORD OX5 1GB, OXON, ENGLAND
SN 0967-5868
EI 1532-2653
J9 J CLIN NEUROSCI
JI J. Clin. Neurosci.
PD MAR
PY 2020
VL 73
BP 264
EP 279
DI 10.1016/j.jocn.2020.01.012
PG 16
WC Clinical Neurology; Neurosciences
WE Science Citation Index Expanded (SCI-EXPANDED)
SC Neurosciences & Neurology
GA LF3GN
UT WOS:000527309400047
PM 32005412
DA 2023-08-27
ER

PT J
AU Kshettry, VR
   Chotai, S
   Chen, W
   Zhang, J
   Ammirati, M
AF Kshettry, Varun R.
   Chotai, Silky
   Chen, William
   Zhang, Jun
   Ammirati, Mario
TI Quantitative analysis of the effect of brainstem shift on surgical
   approaches to anterolateral tumors at the craniovertebral junction
SO JOURNAL OF CLINICAL NEUROSCIENCE
LA English
DT Article
DE Craniovertebral junction; Far lateral approach; Neurosurgery; Skull base
   neoplasms
ID FORAMEN MAGNUM MENINGIOMAS; LATERAL TRANSCONDYLAR APPROACH; INTRADURAL
   LESIONS; CRANIOCERVICAL JUNCTION; SUBOCCIPITAL APPROACH;
   MORPHOMETRIC-ANALYSIS; ANTERIOR PORTION; CERVICAL-SPINE; MANAGEMENT;
   RESECTION
AB Many anterolateral craniovertebral junction (CVJ) tumors can safely be resected using a simple posterolateral approach given the surgical corridor provided by brainstem shift. We sought to study how increasing anterolateral CVJ lesion size affects exposure in the posterolateral and far lateral approaches. Six cadaveric heads were used. A posterolateral approach was performed on one side and a far lateral with one-third condyle resection on the other side. Clival and brainstem exposure and surgical freedom were measured. A balloon catheter was used to simulate 10, 15, and 20 mm anterolateral mass lesions. Mean clival exposure was significantly greater with the far lateral approach (197.4 versus [vs] 135.0 mm(2), p = 0.03) with no balloon, but this difference disappeared with lesion sizes of 10 mm (246.8 vs 237.9 mm(2), p = 0.79), 15 mm (306.7 vs 262.4 mm(2), p = 0.25), and 20 mm (360.0 vs 332.7 mm(2), p = 0.64). Mean brainstem exposure was significantly greater with the far lateral approach for 0 mm (127.8 vs 65.8 mm(2), p < 0.01), 10 mm (129.5 vs 87.5 mm(2), p = 0.045), and 15 mm (140.1 vs 97.8 mm(2), p = 0.01) lesions. There was no difference at 20 mm (146.7 vs 147.8 mm(2), p = 0.97). Medial-lateral surgical freedom was greater with the far lateral approach for all sizes. The results of this study provide insight on one important variable in the decision-making process to select the optimal approach for anterolateral CVJ tumors. (C) 2013 Elsevier Ltd. All rights reserved.
C1 [Kshettry, Varun R.; Chotai, Silky; Chen, William; Ammirati, Mario] Ohio State Univ, Med Ctr, Dept Neurol Surg, Dardinger Microneurosurg Skull Base Lab, Columbus, OH 43210 USA.
   [Kshettry, Varun R.] Cleveland Clin, Dept Neurol Surg, Cleveland, OH 44195 USA.
   [Zhang, Jun] Ohio State Univ, Dept Radiol, Columbus, OH 43210 USA.
   [Zhang, Jun] Ohio State Univ, Wright Ctr Innovat Biomed Imaging, Columbus, OH 43210 USA.
C3 University System of Ohio; Ohio State University; Cleveland Clinic
   Foundation; University System of Ohio; Ohio State University; University
   System of Ohio; Ohio State University
RP Kshettry, VR (通讯作者)，Ohio State Univ, Med Ctr, Dept Neurol Surg, Dardinger Microneurosurg Skull Base Lab, Columbus, OH 43210 USA.
EM
RI Kshettry, Varun/ABD-4745-2021; Zhang, Jun/E-4367-2011
OI Kshettry, Varun/0000-0003-3217-8279; zhang, jun/0000-0002-6515-601X
CR Acikbas SC, 1997, ACTA NEUROCHIR, V139, P546, DOI 10.1007/BF02750998
   Arnautovic KI, 1997, J NEUROSURG, V86, P252, DOI 10.3171/jns.1997.86.2.0252
   Arnautovic KI, 2000, J NEUROSURG, V92, P71, DOI 10.3171/spi.2000.92.1.0071
   BABU RP, 1994, J NEUROSURG, V81, P49, DOI 10.3171/jns.1994.81.1.0049
   Barut N, 2009, BRIT J NEUROSURG, V23, P276, DOI 10.1080/02688690902814725
   Bassiouni H, 2006, NEUROSURGERY, V59, P1177, DOI 10.1227/01.NEU.0000245629.77968.37
   BERTALANFFY H, 1991, NEUROSURGERY, V29, P815, DOI 10.1227/00006123-199112000-00002
   Bertalanffy H, 1996, Acta Neurochir Suppl, V65, P82
   Bruneau M, 2006, NEUROSURGERY, V58, P29, DOI 10.1227/01.NEU.0000193930.74183.42
   Bruneau M, 2008, NEUROSURG REV, V31, P19, DOI 10.1007/s10143-007-0097-1
   CROCKARD HA, 1991, NEUROSURGERY, V28, P88
   David C A, 1997, Clin Neurosurg, V44, P467
   Dowd GC, 1999, NEUROSURGERY, V45, P95, DOI 10.1097/00006123-199907000-00023
   Fernandez-Miranda JC, 2012, NEUROSURGERY, V71, P158
   Fernandez-Miranda JC, 2012, NEUROSURGERY, V71, P146, DOI 10.1227/NEU.0b013e3182438915
   GEORGE B, 1995, SKULL BASE SURG, V5, P9, DOI 10.1055/s-2008-1058945
   George B, 1997, SURG NEUROL, V47, P371, DOI 10.1016/S0090-3019(96)00204-2
   GEORGE B, 1988, SURG NEUROL, V29, P484, DOI 10.1016/0090-3019(88)90145-0
   Goel A, 2001, NEUROSURGERY, V49, P102, DOI 10.1097/00006123-200107000-00016
   GUIDETTI B, 1980, SURG NEUROL, V13, P9
   Gupta SK, 2004, BRIT J NEUROSURG, V18, P164, DOI 10.1080/02688690410001681028
   HEROS RC, 1986, J NEUROSURG, V64, P559, DOI 10.3171/jns.1986.64.4.0559
   Kassam Amin, 2005, Neurosurg Focus, V19, pE4
   Koutourousiou M, 2012, NEUROSURGERY, V71, P614, DOI 10.1227/NEU.0b013e31825ea3e0
   KRATIMENOS GP, 1993, BRIT J NEUROSURG, V7, P129, DOI 10.3109/02688699309103469
   Kshettry V, J CLIN NEUR IN PRESS
   Lot G, 1999, ACTA NEUROCHIR, V141, P111, DOI 10.1007/s007010050274
   LOVE J G, 1954, J Int Coll Surg, V22, P1
   M Samii, 1996, NEUROSURGERY, V39, P1094
   Margalit Nevo S, 2005, Neurosurgery, V56, P324, DOI 10.1227/01.NEU.0000156796.28536.6D
   MILLER E, 1987, NEUROSURGERY, V20, P966, DOI 10.1227/00006123-198706000-00026
   Morera VA, 2010, NEUROSURGERY, V66, pONS211, DOI 10.1227/01.NEU.0000369926.01891.5D
   MULLAN S, 1966, J NEUROSURG, V24, P536, DOI 10.3171/jns.1966.24.2.0536
   Muthukumar N, 2005, ACTA NEUROCHIR, V147, P889, DOI 10.1007/s00701-005-0555-x
   Nanda A, 2002, J NEUROSURG, V96, P302, DOI 10.3171/jns.2002.96.2.0302
   Parlato C, 2003, ACTA NEUROCHIR, V145, P31, DOI 10.1007/s00701-002-1024-4
   PRITZ MB, 1991, ACTA NEUROCHIR, V113, P74, DOI 10.1007/BF01402118
   Rhoton AL, 2000, NEUROSURGERY, V47, pS195, DOI 10.1097/00006123-200009001-00020
   Safavi-Abbasi S, 2007, J NEUROSURG, V107, P797, DOI 10.3171/JNS-07/10/0797
   Salas E, 1999, J NEUROSURG, V90, P206, DOI 10.3171/spi.1999.90.2.0206
   Samii M, 1996, NEUROSURGERY, V39, P1086, DOI 10.1097/00006123-199612000-00003
   Seeger W, 1978, ATLAS TOPOGRAPHICAL, P486
   SEN CN, 1990, NEUROSURGERY, V27, P197, DOI 10.1227/00006123-199008000-00004
   SEN CN, 1991, ACTA NEUROCHIR, V108, P70, DOI 10.1007/BF01407670
   SMOLIK EA, 1954, J NEUROSURG, V11, P161, DOI 10.3171/jns.1954.11.2.0161
   Spektor S, 2000, J NEUROSURG, V92, P824, DOI 10.3171/jns.2000.92.5.0824
   STEIN BM, 1963, J NEUROSURG, V20, P740, DOI 10.3171/jns.1963.20.9.0740
   Tubbs RS, 2010, NEUROSURGERY, V66, P385, DOI 10.1227/01.NEU.0000363407.78399.BA
   Vishteh AG, 1999, J NEUROSURG, V90, P91, DOI 10.3171/spi.1999.90.1.0091
   Wanebo JE, 2001, NEUROSURGERY, V49, P934, DOI 10.1097/00006123-200110000-00027
   Wu Z, 2009, SURG NEUROL, V72, P376, DOI 10.1016/j.surneu.2009.05.006
   Yasargil MG, 1996, MICRONEUROSURGERY B, VIVB, P134
   YASUOKA S, 1978, J NEUROSURG, V49, P828, DOI 10.3171/jns.1978.49.6.0828
   ZOLTAN L, 1974, ACTA NEUROCHIR, V30, P217, DOI 10.1007/BF01405581
NR 54
TC 5
Z9 5
U1 0
U2 1
PU ELSEVIER SCI LTD
PI OXFORD
PA THE BOULEVARD, LANGFORD LANE, KIDLINGTON, OXFORD OX5 1GB, OXON, ENGLAND
SN 0967-5868
EI 1532-2653
J9 J CLIN NEUROSCI
JI J. Clin. Neurosci.
PD APR
PY 2014
VL 21
IS 4
BP 644
EP 650
DI 10.1016/j.jocn.2013.08.003
PG 7
WC Clinical Neurology; Neurosciences
WE Science Citation Index Expanded (SCI-EXPANDED)
SC Neurosciences & Neurology
GA AE1IS
UT WOS:000333723600019
PM 24290905
DA 2023-08-27
ER

PT J
AU Schipani, S
   Jain, R
   Shah, K
   Rock, JP
   Movsas, B
   Rosenblum, M
   Ryu, S
AF Schipani, Stefano
   Jain, Rajan
   Shah, Keyur
   Rock, Jack P.
   Movsas, Benjamin
   Rosenblum, Mark
   Ryu, Samuel
TI Clinical, dosimetric, and radiographic correlation of radiation injury
   involving the brainstem and the medial temporal lobes following
   stereotactic radiotherapy for neoplasms of central skull base
SO JOURNAL OF NEURO-ONCOLOGY
LA English
DT Article
DE Radionecrosis; Brainstem; Radiosurgery; Radiation damage; Stereotactic
   radiation
ID GAMMA-KNIFE RADIOSURGERY; HYPERBARIC-OXYGEN THERAPY;
   ARTERIOVENOUS-MALFORMATIONS; MALIGNANT GLIOMAS; OPTIC NEUROPATHY;
   NECROSIS; RADIONECROSIS; TUMOR; RISK; PET
AB Stereotactic Radiotherapy (SRT) is more commonly used for skull base tumors in conjunction with the technical development of radiation intensity modulation. Purpose of this study is to correlate clinical and radiographic characteristics of delayed radiation injury (RI) occurring around central skull base following SRT with SRT dosimetric data. Total of six patients were identified to have developed RI in the vicinity of SRT target volume out of 141 patients who received SRT in he center or near-center of the skull base. The images and medical records were retrospectively reviewed. The analysis was performed for RI location, time of development, imaging and clinical characteristics and evolution of RI and correlated with SRT dosimetric analysis using image fusion with follow-up MRI scans. Mean follow-up time was 24 +/- 9 months. During the follow-up period, twelve sites of RI were found in 6 patients. They were clinically symptomatic in 4/6 patients (66.6%) at median 12.5 months after SRT. Mean time interval between SRT and detection of RI was 9 +/- 3, 18.5 +/- 5, and 13.5 months for brainstem, temporal lobe, and cerebellum/labyrinth lesions, respectively. All RI lesions were included in the region of high SRT doses. After steroid and symptomatic treatment, 50% of RI lesions showed complete response, and 40% showed partial response. RI can occur around the skull base because of irregular shape of target tumor, its close proximity to normal brain parenchyma, and inhomogeneity of dose distribution. Brainstem lesions occurred earlier than temporal lobe RI. The majority of the RI lesions, not mixed with the tumor in this study, showed radiographic and clinical improvement with steroid and symptomatic treatments.
C1 [Schipani, Stefano; Movsas, Benjamin; Ryu, Samuel] Henry Ford Hlth Syst, Dept Radiat Oncol, Detroit, MI 48202 USA.
   [Jain, Rajan; Shah, Keyur] Henry Ford Hosp, Dept Neuroradiol, Detroit, MI 48202 USA.
   [Rock, Jack P.; Rosenblum, Mark; Ryu, Samuel] Henry Ford Hosp, Dept Neurosurg, Detroit, MI 48202 USA.
C3 Henry Ford Health System; Henry Ford Hospital; Henry Ford Health System;
   Henry Ford Hospital; Henry Ford Health System; Henry Ford Hospital
RP Ryu, S (通讯作者)，Henry Ford Hlth Syst, Dept Radiat Oncol, 2799 W Grand Blvd, Detroit, MI 48202 USA.
EM
OI Jain, Rajan/0000-0002-4879-0457; Schipani, Stefano/0000-0002-2898-9928
CR ALMEFTY O, 1990, J NEUROSURG, V73, P502, DOI 10.3171/jns.1990.73.4.0502
   Cha Soonmee, 2003, Magn Reson Imaging Clin N Am, V11, P403, DOI 10.1016/S1064-9689(03)00066-7
   Chao ST, 2001, INT J CANCER, V96, P191, DOI 10.1002/ijc.1016.abs
   Chen JL, 2003, ANN NEUROL, V53, P743, DOI 10.1002/ana.10555
   Chin LS, 2001, J NEUROSURG, V94, P899, DOI 10.3171/jns.2001.94.6.0899
   Chong VFH, 1999, INT J RADIAT ONCOL, V45, P699, DOI 10.1016/S0360-3016(99)00236-9
   Chuba PJ, 1997, CANCER-AM CANCER SOC, V80, P2005, DOI 10.1002/(SICI)1097-0142(19971115)80:10<2005::AID-CNCR19>3.0.CO;2-0
   Delanian S, 2008, J NEUROL SCI, V275, P164, DOI 10.1016/j.jns.2008.08.004
   DILORENZO N, 1978, SURG NEUROL, V10, P281
   Diomede L, 2001, ARTERIOSCL THROM VAS, V21, P1327, DOI 10.1161/hq0801.094222
   Djalilian HR, 2007, AM J OTOLARYNG, V28, P338, DOI 10.1016/j.amjoto.2006.09.008
   Flickinger JC, 2000, INT J RADIAT ONCOL, V46, P1143, DOI 10.1016/S0360-3016(99)00513-1
   Flickinger JC, 1997, INT J RADIAT ONCOL, V38, P485, DOI 10.1016/S0360-3016(97)89481-3
   Flickinger JC, 1998, INT J RADIAT ONCOL, V40, P273, DOI 10.1016/S0360-3016(97)00718-9
   Gonzalez J, 2007, INT J RADIAT ONCOL, V67, P323, DOI 10.1016/j.ijrobp.2006.10.010
   Graves EE, 2001, AM J NEURORADIOL, V22, P613
   *HYP UND MED SOC, HYP UND MED SOC IND
   Jain R, 2007, NEUROSURGERY, V61, P778, DOI 10.1227/01.NEU.0000298906.48388.26
   Karlsson B, 1997, RADIOTHER ONCOL, V43, P275, DOI 10.1016/S0167-8140(97)00060-1
   Kim JH, 2004, RADIAT RES, V161, P137, DOI 10.1667/RR3124
   Kumar AJ, 2000, RADIOLOGY, V217, P377, DOI 10.1148/radiology.217.2.r00nv36377
   Langleben DD, 2000, J NUCL MED, V41, P1861
   Leber KA, 1998, STEREOT FUNCT NEUROS, V70, P229, DOI 10.1159/000056426
   Levegrun S, 2004, INT J RADIAT ONCOL, V59, P796, DOI 10.1016/j.ijrobp.2003.11.033
   Lynn M, 2007, NEUROSURGERY, V60, P579, DOI 10.1227/01.NEU.0000255343.41206.6F
   MACDONALD DR, 1990, J CLIN ONCOL, V8, P1277, DOI 10.1200/JCO.1990.8.7.1277
   Monheit BE, 2004, J NEURO-OPHTHALMOL, V24, P195, DOI 10.1097/00041327-200409000-00002
   Mullins ME, 2005, AM J NEURORADIOL, V26, P1967
   Rogers LR, 2006, NEUROLOGY, V66, pA335
   Ryu S, 2004, OTOLARYNG HEAD NECK, V130, P690, DOI 10.1016/j.otohns.2003.10.009
   Ryu S, 2003, CANCER, V97, P2013, DOI 10.1002/cncr.11296
   Ryu S, 2007, J NEURO-ONCOL, V82, P119, DOI 10.1007/s11060-006-9256-4
   Stokkel M, 1999, NUCL MED COMMUN, V20, P411, DOI 10.1097/00006231-199905000-00003
   Takenaka N, 2003, NEUROL MED-CHIR, V43, P391, DOI 10.2176/nmc.43.391
   Voges J, 1996, INT J RADIAT ONCOL, V36, P1055, DOI 10.1016/S0360-3016(96)00422-1
   Wanebo JE, 2009, SURG NEUROL, V72, P162, DOI 10.1016/j.surneu.2008.03.037
   Williamson R, 2008, STEREOT FUNCT NEUROS, V86, P359, DOI 10.1159/000163557
NR 37
TC 4
Z9 4
U1 0
U2 1
PU SPRINGER
PI NEW YORK
PA 233 SPRING ST, NEW YORK, NY 10013 USA
SN 0167-594X
EI 1573-7373
J9 J NEURO-ONCOL
JI J. Neuro-Oncol.
PD JUN
PY 2010
VL 98
IS 2
SI SI
BP 177
EP 184
DI 10.1007/s11060-010-0171-3
PG 8
WC Oncology; Clinical Neurology
WE Science Citation Index Expanded (SCI-EXPANDED)
SC Oncology; Neurosciences & Neurology
GA 609VF
UT WOS:000278684900005
PM 20376551
DA 2023-08-27
ER

PT J
AU Michalski, A
   Bouffet, E
   Taylor, RE
   Hargrave, D
   Walker, D
   Picton, S
   Robinson, K
   Pizer, B
   Bujkiewicz, S
AF Michalski, Antony
   Bouffet, Eric
   Taylor, Roger E.
   Hargrave, Darren
   Walker, David
   Picton, Susan
   Robinson, Kathryn
   Pizer, Barry
   Bujkiewicz, Sylwia
TI The addition of high-dose tamoxifen to standard radiotherapy does not
   improve the survival of patients with diffuse intrinsic pontine glioma
SO JOURNAL OF NEURO-ONCOLOGY
LA English
DT Article
DE Diffuse intrinsic pontine glioma; Tamoxifen; Progression free survival;
   Overall survival; Corticosteroids
ID BRAIN-STEM TUMORS; PROTEIN-KINASE-C; PEDIATRIC-ONCOLOGY-GROUP;
   HYPERFRACTIONATED RADIATION-THERAPY; MALIGNANT GLIOMAS; CHILDREN;
   CHEMOTHERAPY; CARBOPLATIN; MANAGEMENT; INHIBITOR
AB The study aimed to examine the tolerability of the combination of radiotherapy and tamoxifen and the effect on median and event free survival as well as collecting data on the use of steroids in this population. 31 patients with diffuse intrinsic pontine glioma, diagnosed on clinical and radiological criteria, were treated with high-dose oral tamoxifen (120 mg/m(2)/day) given concomitantly with standard dose radiotherapy (54 Gy in 1.8 Gy fractions over 6 weeks). Results Tamoxifen was well tolerated with no grade 3 or 4 CTC toxicity reported. At 1 year, the progression free and event free survival were 3.2% (95% CI: 0.2-14.1%), and at 6 months 19.4% (CI: 7.9% to 34.6%). The overall survival at 1 year was 16.1% (CI: 5.9-30.9%) with median survival 6.32 months. In this study, in which tamoxifen was used in conjunction with radiotherapy, progression free survival was shown to be less good when compared with historical data HR = 3.1 (CI: 1.7-5.7). There was no significant reduction in overall survival. The addition of high-dose tamoxifen, although well tolerated, confers no clinical benefit to patients treated with diffuse intrinsic pontine glioma treated with standard radiotherapy.
C1 [Michalski, Antony] Great Ormond St Hosp Sick Children, Dept Oncol, London WC1N 3JH, England.
   [Bouffet, Eric] Hosp Sick Children, Div Haematol Oncol, Toronto, ON M5G 1X8, Canada.
   [Taylor, Roger E.] Singleton Hosp, S W Wales Canc Ctr, Swansea SA2 8QA, W Glam, Wales.
   [Hargrave, Darren] Royal Marsden Hosp, Paediat Oncol Unit, Sutton SM2 5PT, Surrey, England.
   [Walker, David] Queens Med Ctr, Childrens Brain Tumour Res Ctr, Div Child Hlth, Nottingham NG7 2UH, England.
   [Picton, Susan] St James Univ Hosp, Reg Paediat Oncol Unit, Leeds LS9 7TF, W Yorkshire, England.
   [Robinson, Kathryn; Bujkiewicz, Sylwia] Univ Leicester, CCLG Data Ctr, Leicester LE1 6TH, Leics, England.
   [Pizer, Barry] Alder Hey Childrens Hosp, Liverpool L12 2AP, Merseyside, England.
C3 University of London; University College London; Great Ormond Street
   Hospital for Children NHS Foundation Trust; University of Toronto;
   Hospital for Sick Children (SickKids); Singleton Hospital; Royal Marsden
   NHS Foundation Trust; University of Nottingham; Saint James's University
   Hospital; University of Leicester; Alder Hey Children's NHS Foundation
   Trust; Alder Hey Children's Hospital
RP Michalski, A (通讯作者)，Great Ormond St Hosp Sick Children, Dept Oncol, London WC1N 3JH, England.
EM
RI Bouffet, Eric/AAB-8999-2020; Hargrave, Darren R/H-1066-2014
OI Hargrave, Darren R/0000-0001-8219-9807; Bujkiewicz,
   Sylwia/0000-0002-3003-9403
FU Cancer Research UK; Samantha Dickson Brain Tumour Trust
FX Authors wish to thank Charles Stiller for providing very useful data
   from the National Registry of Childhood Tumours. The authors wish to
   acknowledge Claire Weston for preliminary statistical analysis and Prof
   David Machin for statistical input into the data analysis and
   manuscript. The CCLG is supported by Cancer Research UK. The CNS
   Division is additionally supported by a grant from the Samantha Dickson
   Brain Tumour Trust.
CR [Anonymous], 2007, SURVIVAL ANAL PRACTI
   BALTUCH GH, 1993, NEUROSURGERY, V33, P495
   Bouffet E, 2000, CANCER, V88, P685, DOI 10.1002/(SICI)1097-0142(20000201)88:3<685::AID-CNCR27>3.0.CO;2-K
   Broniscer A, 2000, J CLIN ONCOL, V18, P1246, DOI 10.1200/JCO.2000.18.6.1246
   Couldwell WT, 1996, CLIN CANCER RES, V2, P619
   Doz F, 2002, EUR J CANCER, V38, P815, DOI 10.1016/S0959-8049(02)00029-1
   Dunkel IJ, 1998, J NEURO-ONCOL, V37, P67, DOI 10.1023/A:1005874508975
   Fine HA, 2005, J CLIN ONCOL, V23, p115S
   Freeman A., 1994, Proceedings of the American Association for Cancer Research Annual Meeting, V35, P470
   FREEMAN CR, 1991, CANCER-AM CANCER SOC, V68, P474, DOI 10.1002/1097-0142(19910801)68:3<474::AID-CNCR2820680305>3.0.CO;2-7
   Glaser AW, 1997, ARCH DIS CHILD, V76, P76, DOI 10.1136/adc.76.1.76
   Graff JR, 2005, CANCER RES, V65, P7462, DOI 10.1158/0008-5472.CAN-05-0071
   Hargrove D, 2006, LANCET ONCOL, V7, P241, DOI 10.1016/S1470-2045(06)70615-5
   JENKIN RDT, 1987, J NEUROSURG, V66, P227, DOI 10.3171/jns.1987.66.2.0227
   Kaplan AM, 1996, PEDIATR NEUROSURG, V24, P185, DOI 10.1159/000121036
   KRETSCHMAR CS, 1993, CANCER-AM CANCER SOC, V72, P1404, DOI 10.1002/1097-0142(19930815)72:4<1404::AID-CNCR2820720441>3.0.CO;2-G
   Lewis J, 1997, INT J RADIAT ONCOL, V38, P925, DOI 10.1016/S0360-3016(97)00134-X
   Mandell LR, 1999, INT J RADIAT ONCOL, V43, P959, DOI 10.1016/S0360-3016(98)00501-X
   Nowak A, 2004, COCHRANE DB SYST REV, DOI DOI 10.1002/14651858.CD001024.PUB2
   Pollack IF, 1997, CLIN CANCER RES, V3, P1109
   POLLACK IF, 1990, CANCER RES, V50, P7134
   *STATACORP LP, 2007, STAT SOFTW REL 10
   Teicher BA, 2001, CLIN CANCER RES, V7, P634
   Teicher BA, 2006, CLIN CANCER RES, V12, P5336, DOI 10.1158/1078-0432.CCR-06-0945
   Walter AW, 1998, MED PEDIATR ONCOL, V30, P28, DOI 10.1002/(SICI)1096-911X(199801)30:1<28::AID-MPO9>3.0.CO;2-2
NR 25
TC 25
Z9 27
U1 0
U2 2
PU SPRINGER
PI NEW YORK
PA ONE NEW YORK PLAZA, SUITE 4600, NEW YORK, NY, UNITED STATES
SN 0167-594X
EI 1573-7373
J9 J NEURO-ONCOL
JI J. Neuro-Oncol.
PD OCT
PY 2010
VL 100
IS 1
BP 81
EP 88
DI 10.1007/s11060-010-0141-9
PG 8
WC Oncology; Clinical Neurology
WE Science Citation Index Expanded (SCI-EXPANDED)
SC Oncology; Neurosciences & Neurology
GA 661CF
UT WOS:000282699900009
PM 20238235
DA 2023-08-27
ER

PT J
AU Negretti, L
   Bouchireb, K
   Levy-Piedbois, C
   Habrand, JL
   Dhermain, F
   Kalifa, C
   Grill, J
   Dufour, C
AF Negretti, Laura
   Bouchireb, Karim
   Levy-Piedbois, Christine
   Habrand, Jean Louis
   Dhermain, Frederic
   Kalifa, Chantal
   Grill, Jacques
   Dufour, Christelle
TI Hypofractionated radiotherapy in the treatment of diffuse intrinsic
   pontine glioma in children: a single institution's experience
SO JOURNAL OF NEURO-ONCOLOGY
LA English
DT Article
DE Brainstem glioma; Child; Hypofractionated radiation therapy
ID BRAIN-STEM TUMORS; PEDIATRIC-ONCOLOGY-GROUP; HYPERFRACTIONATED
   RADIOTHERAPY; RADIATION-THERAPY; MANAGEMENT; TRIALS
AB We report herein our institutional experience in the treatment of diffuse intrinsic pontine glioma (DIPG) with a hypofractionated external-beam radiotherapy schedule. Between April 1996 and January 2004, 22 patients (age 2.9-12.5 years) with newly diagnosed DIPG were treated by hypofractionated radiation therapy delivering a total dose of 45 Gy in daily fractions of 3 Gy, given over 3 weeks. No other treatment was applied concomitantly. Fourteen of the 22 patients received the prescribed dose of 45 Gy in 15 fractions of 3 Gy, and 2 patients received a total dose of 60 and 45 Gy with a combination of two different beams (photons and neutrons). In five cases the daily fraction was modified to 2 Gy due to intolerance, and one patient died due to serious intracranial hypertension after two fractions of 3 Gy and one of 2 Gy. Among 22 children, 14 patients showed clinical improvement, usually starting in the second week of treatment. No grade 3 or 4 acute toxicity from radiotherapy was observed. No treatment interruption was needed. In six patients, steroids could be discontinued within 1 month after the end of radiotherapy. Median time to progression and median overall survival were 5.7 months and 7.6 months, respectively. External radiotherapy with a radical hypofractionated regimen is feasible and well tolerated in children with newly diagnosed DIPG. However, this regimen does not seem to change overall survival in this setting. It could represent a short-duration alternative to more protracted regimens.
C1 [Dufour, Christelle] Inst Gustave Roussy, Dept Pediat & Adolescent, F-94805 Villejuif, France.
   [Negretti, Laura; Habrand, Jean Louis; Dhermain, Frederic] Inst Gustave Roussy, Dept Radiat Oncol, F-94805 Villejuif, France.
   [Bouchireb, Karim; Kalifa, Chantal; Grill, Jacques; Dufour, Christelle] Inst Gustave Roussy, Dept Pediat & Adolescent Oncol, F-94805 Villejuif, France.
   [Levy-Piedbois, Christine] Radiat Oncol Inst, Bobigny, France.
C3 UNICANCER; Gustave Roussy; UNICANCER; Gustave Roussy; UNICANCER; Gustave
   Roussy
RP Dufour, C (通讯作者)，Inst Gustave Roussy, Dept Pediat & Adolescent, 114 Rue Edouard Vaillant, F-94805 Villejuif, France.
EM
CR ALBRIGHT AL, 1993, NEUROSURGERY, V33, P1026
   Donaldson SS, 2006, J CLIN ONCOL, V24, P1266, DOI 10.1200/JCO.2005.04.6599
   EIFEL PJ, 1987, INT J RADIAT ONCOL, V13, P847, DOI 10.1016/0360-3016(87)90098-8
   FREEMAN CR, 1986, INT J RADIAT ONCOL, V12, P1823, DOI 10.1016/0360-3016(86)90325-1
   FREEMAN CR, 1993, INT J RADIAT ONCOL, V27, P197, DOI 10.1016/0360-3016(93)90228-N
   GNEKOW AK, 1995, MED PEDIATR ONCOL, V24, P104, DOI 10.1002/mpo.2950240209
   HALPERIN EC, 1985, INT J RADIAT ONCOL, V11, P1293, DOI 10.1016/0360-3016(85)90244-5
   Hargrove D, 2006, LANCET ONCOL, V7, P241, DOI 10.1016/S1470-2045(06)70615-5
   Janssens GORJ, 2009, INT J RADIAT ONCOL, V73, P722, DOI 10.1016/j.ijrobp.2008.05.030
   KRETSCHMAR CS, 1993, CANCER-AM CANCER SOC, V72, P1404, DOI 10.1002/1097-0142(19930815)72:4<1404::AID-CNCR2820720441>3.0.CO;2-G
   LANGMOEN IA, 1991, CHILD NERV SYST, V7, P13, DOI 10.1007/BF00263825
   LEE F, 1975, J NEUROSURG, V43, P65, DOI 10.3171/jns.1975.43.1.0065
   LITTMAN P, 1980, CANCER, V45, P2787, DOI 10.1002/1097-0142(19800601)45:11<2787::AID-CNCR2820451113>3.0.CO;2-V
   Mandell LR, 1999, INT J RADIAT ONCOL, V43, P959, DOI 10.1016/S0360-3016(98)00501-X
   Smith MA, 1998, J NATL CANCER I, V90, P1269, DOI 10.1093/jnci/90.17.1269
NR 15
TC 27
Z9 28
U1 0
U2 5
PU SPRINGER
PI NEW YORK
PA 233 SPRING ST, NEW YORK, NY 10013 USA
SN 0167-594X
J9 J NEURO-ONCOL
JI J. Neuro-Oncol.
PD SEP
PY 2011
VL 104
IS 3
BP 773
EP 777
DI 10.1007/s11060-011-0542-4
PG 5
WC Oncology; Clinical Neurology
WE Science Citation Index Expanded (SCI-EXPANDED)
SC Oncology; Neurosciences & Neurology
GA 818FK
UT WOS:000294736700017
PM 21327862
DA 2023-08-27
ER

PT J
AU Sethi, R
   Allen, J
   Donahue, B
   Karajannis, M
   Gardner, S
   Wisoff, J
   Kunnakkat, S
   Mathew, J
   Zagzag, D
   Newman, K
   Narayana, A
AF Sethi, Rajni
   Allen, Jeffrey
   Donahue, Bernadine
   Karajannis, Matthias
   Gardner, Sharon
   Wisoff, Jeffrey
   Kunnakkat, Saroj
   Mathew, Jeena
   Zagzag, David
   Newman, Kia
   Narayana, Ashwatha
TI Prospective neuraxis MRI surveillance reveals a high risk of
   leptomeningeal dissemination in diffuse intrinsic pontine glioma
SO JOURNAL OF NEURO-ONCOLOGY
LA English
DT Article
DE Diffuse intrinsic pontine glioma; Leptomeningeal dissemination;
   Radiotherapy
ID BRAIN-STEM GLIOMAS; HYPERFRACTIONATED RADIATION-THERAPY;
   PEDIATRIC-ONCOLOGY-GROUP; GROUP PHASE-I/II; HIGH-GRADE GLIOMAS;
   CHILDREN; RADIOTHERAPY; TUMORS; TRIAL; CARBOPLATIN
AB Prognosis of diffuse intrinsic pontine gliomas (DIPGs) remains poor. Failure has been predominantly local, with leptomeningeal dissemination (LD) occurring in 4-33% of patients in pre-MRI era series. Routine craniospinal imaging after initial treatment may reveal other relapse patterns relapse. Sixteen consecutive pediatric patients with DIPG treated between 2006 and 2009 were retrospectively reviewed. Treatment regimens, recurrence patterns, survival, and pathologic diagnosis were recorded. Fourteen patients received involved-field radiotherapy to 54 Gy, and two patients received craniospinal irradiation for LD at presentation. Neuraxis MRI was performed at diagnosis and at 4 month intervals following radiotherapy. Fifteen patients have had progression of disease (median progression-free survival 5.0 +/- A 1.2 months), and 13 patients have died (median survival 9.0 +/- A 1.4 months). Local failure occurred in 12 patients (75%). LD occurred in nine patients (56%). LD was present at diagnosis in three patients, after initial staging and treatment in six patients, and during autopsy in two patients. Median overall survival was 12.0 +/- A 3.3 months without LD and 8.0 +/- A 2.1 months with LD (P = 0.059, log rank test). Median progression-free survival was 9.5 +/- A 3.9 months without LD and 3.0 +/- A 2.1 months with LD (P = 0.012, log rank test). The high incidence of LD probably reflects liberal use of spine MRI surveillance. All patients should undergo routine craniospinal imaging at diagnosis and follow-up. Central nervous system prophylaxis should be considered in future clinical trials.
C1 [Sethi, Rajni; Donahue, Bernadine; Kunnakkat, Saroj; Narayana, Ashwatha] NYU, Dept Radiat Oncol, Langone Med Ctr, New York, NY 10014 USA.
   [Allen, Jeffrey; Karajannis, Matthias; Gardner, Sharon; Mathew, Jeena] NYU, Dept Pediat, Langone Med Ctr, New York, NY 10014 USA.
   [Allen, Jeffrey] NYU, Dept Neurol, Langone Med Ctr, New York, NY 10014 USA.
   [Wisoff, Jeffrey; Narayana, Ashwatha] NYU, Dept Neurosurg, Langone Med Ctr, New York, NY 10014 USA.
   [Zagzag, David; Newman, Kia] NYU, Dept Pathol, Langone Med Ctr, New York, NY 10014 USA.
   [Donahue, Bernadine] Maimonides Hosp, Dept Radiat Oncol, Brooklyn, NY 11219 USA.
C3 New York University; NYU Langone Medical Center; New York University;
   NYU Langone Medical Center; New York University; NYU Langone Medical
   Center; New York University; NYU Langone Medical Center; New York
   University; NYU Langone Medical Center; Maimonides Medical Center
RP Sethi, R (通讯作者)，NYU, Dept Radiat Oncol, Langone Med Ctr, 566 1st Ave, New York, NY 10014 USA.
EM
RI Wisoff, Jeffrey/AAW-1500-2020; Karajannis, Matthias A/H-8845-2019
OI Karajannis, Matthias A/0000-0002-7151-6528; Allen,
   Jeffrey/0000-0001-9957-6664; gardner, sharon/0000-0002-8857-5487;
   Wisoff, Jeffrey/0000-0003-3303-2220
CR ALBRIGHT AL, 1986, J NEUROSURG, V65, P751, DOI 10.3171/jns.1986.65.6.0751
   Allen J, 1999, CANCER, V86, P1064, DOI 10.1002/(SICI)1097-0142(19990915)86:6<1064::AID-CNCR24>3.0.CO;2-1
   Becher OJ, 2010, CANCER RES, V70, P2548, DOI 10.1158/0008-5472.CAN-09-2503
   Benesch M, 2005, J NEURO-ONCOL, V72, P179, DOI 10.1007/s11060-004-3546-5
   Broniscer A, 2005, CANCER-AM CANCER SOC, V103, P133, DOI 10.1002/cncr.20741
   Donahue B, 1998, INT J RADIAT ONCOL, V40, P677, DOI 10.1016/S0360-3016(97)00846-8
   Donaldson SS, 2006, J CLIN ONCOL, V24, P1266, DOI 10.1200/JCO.2005.04.6599
   EDWARDS MSB, 1987, PEDIATR NEUROSCI, V13, P309, DOI 10.1159/000120349
   EDWARDS MSB, 1989, J NEUROSURG, V70, P691, DOI 10.3171/jns.1989.70.5.0691
   EPSTEIN F, 1988, J NEURO-ONCOL, V6, P309
   Freeman CR, 2000, INT J RADIAT ONCOL, V47, P561, DOI 10.1016/S0360-3016(00)00471-5
   FREEMAN CR, 1993, INT J RADIAT ONCOL, V27, P197, DOI 10.1016/0360-3016(93)90228-N
   FREEMAN CR, 1991, CANCER-AM CANCER SOC, V68, P474, DOI 10.1002/1097-0142(19910801)68:3<474::AID-CNCR2820680305>3.0.CO;2-7
   FREEMAN CR, 1988, INT J RADIAT ONCOL, V15, P311, DOI 10.1016/S0360-3016(98)90010-4
   FREEMAN CR, 1986, IJROBP, V12, P1923
   Gururangan S, 2006, J NEURO-ONCOL, V77, P207, DOI 10.1007/s11060-005-9029-5
   Hargrove D, 2006, LANCET ONCOL, V7, P241, DOI 10.1016/S1470-2045(06)70615-5
   Hayward RM, 2008, J NEURO-ONCOL, V90, P57, DOI 10.1007/s11060-008-9631-4
   Jakacki RI, 1999, J NEURO-ONCOL, V44, P77, DOI 10.1023/A:1006360222643
   KRETSCHMAR CS, 1993, CANCER-AM CANCER SOC, V72, P1404, DOI 10.1002/1097-0142(19930815)72:4<1404::AID-CNCR2820720441>3.0.CO;2-G
   LITTMAN P, 1980, CANCER, V45, P2787, DOI 10.1002/1097-0142(19800601)45:11<2787::AID-CNCR2820451113>3.0.CO;2-V
   Mandell LR, 1999, INT J RADIAT ONCOL, V43, P959, DOI 10.1016/S0360-3016(98)00501-X
   MANTRAVADI RVP, 1982, CANCER, V49, P1294, DOI 10.1002/1097-0142(19820315)49:6<1294::AID-CNCR2820490636>3.0.CO;2-V
   ONDA K, 1989, NEUROSURGERY, V25, P533, DOI 10.1227/00006123-198910000-00005
   Packer RJ, 2005, CANCER-AM CANCER SOC, V104, P1281, DOI 10.1002/cncr.21301
   Packer RJ, 1996, CANCER-AM CANCER SOC, V77, P2150, DOI 10.1002/(SICI)1097-0142(19960515)77:10<2150::AID-CNCR28>3.0.CO;2-T
   PACKER RJ, 1993, CANCER-AM CANCER SOC, V72, P1414, DOI 10.1002/1097-0142(19930815)72:4<1414::AID-CNCR2820720442>3.0.CO;2-C
   PACKER RJ, 1994, CANCER-AM CANCER SOC, V74, P1827, DOI 10.1002/1097-0142(19940915)74:6<1827::AID-CNCR2820740628>3.0.CO;2-Q
   PACKER RJ, 1990, ANN NEUROL, V27, P167, DOI 10.1002/ana.410270212
   PACKER RJ, 1987, INT J RADIAT ONCOL, V13, P1647, DOI 10.1016/0360-3016(87)90160-X
   PACKER RJ, 1983, ANN NEUROL, V72, P1414
   PIERREKAHN A, 1993, J NEUROSURG, V79, P845, DOI 10.3171/jns.1993.79.6.0845
   PRADOS MD, 1995, INT J RADIAT ONCOL, V32, P85, DOI 10.1016/0360-3016(95)00563-E
   Stark AM, 2005, SURG NEUROL, V63, P162, DOI 10.1016/j.surneu.2004.01.028
   Wagner S, 2006, BRIT J CANCER, V95, P991, DOI 10.1038/sj.bjc.6603402
   Walter AW, 1998, MED PEDIATR ONCOL, V30, P28, DOI 10.1002/(SICI)1096-911X(199801)30:1<28::AID-MPO9>3.0.CO;2-2
   Zarghooni M, 2010, J CLIN ONCOL, V28, P1337, DOI 10.1200/JCO.2009.25.5463
NR 37
TC 39
Z9 39
U1 0
U2 4
PU SPRINGER
PI NEW YORK
PA 233 SPRING ST, NEW YORK, NY 10013 USA
SN 0167-594X
J9 J NEURO-ONCOL
JI J. Neuro-Oncol.
PD MAR
PY 2011
VL 102
IS 1
BP 121
EP 127
DI 10.1007/s11060-010-0301-y
PG 7
WC Oncology; Clinical Neurology
WE Science Citation Index Expanded (SCI-EXPANDED)
SC Oncology; Neurosciences & Neurology
GA 723KC
UT WOS:000287504400015
PM 20623246
DA 2023-08-27
ER

PT J
AU Singh, S
   Bhutani, R
   Jalali, R
AF Singh, Suruchi
   Bhutani, Ritu
   Jalali, Rakesh
TI Leptomeninges as a site of relapse in locally controlled, diffuse
   pontine glioma with review of literature
SO CHILDS NERVOUS SYSTEM
LA English
DT Review
DE brain stem glioma; patterns of failure; leptomeningeal spread
ID BRAIN-STEM GLIOMAS; DISSEMINATION; PATTERNS; FAILURE; TUMORS
AB Case report Leptomeningeal involvement in diffuse intrinsic brain stem gliomas is rarely diagnosed clinically and in majority of the instances diagnosed only on postmortem examination. We report two cases of diffuse pontine glioma diagnosed clinicoradiologically and treated with conventional radiotherapy.
   Observations On follow-up, both patients showed clinical features suggestive of meningeal spread although imaging for pontine tumor showed stable disease. Leptomeningeal disease in the spine was confirmed on imaging and in one case by cerebrospinal fluid examination also. During the follow-up of patients with pontine glioma, the possibility of leptomeningeal involvement must be borne in mind.
C1 Tata Mem Hosp, Dept Radiat Oncol, Bombay 400012, Maharashtra, India.
C3 Tata Memorial Centre (TMC); Tata Memorial Hospital
RP Jalali, R (通讯作者)，Tata Mem Hosp, Dept Radiat Oncol, 116, Bombay 400012, Maharashtra, India.
EM
CR Donahue B, 1998, INT J RADIAT ONCOL, V40, P677, DOI 10.1016/S0360-3016(97)00846-8
   EPSTEIN F, 1985, CANCER, V56, P1804, DOI 10.1002/1097-0142(19851001)56:7+<1804::AID-CNCR2820561316>3.0.CO;2-5
   HALPERIN EC, 1985, INT J RADIAT ONCOL, V11, P1293, DOI 10.1016/0360-3016(85)90244-5
   Kanai R, 2005, BRAIN DEV-JPN, V27, P58, DOI 10.1016/j.braindev.2004.03.007
   LITTMAN P, 1980, CANCER, V45, P2787, DOI 10.1002/1097-0142(19800601)45:11<2787::AID-CNCR2820451113>3.0.CO;2-V
   MONTRAVADI R, 1982, CANCER, V49, P1294
   PACKER RJ, 1983, ANN NEUROL, V14, P177, DOI 10.1002/ana.410140204
   Tarnaris A, 2005, J NEUROSURG, V102, P224, DOI 10.3171/jns.2005.102.2.0224
   Yoshimura J, 2003, NEUROL MED-CHIR, V43, P375, DOI 10.2176/nmc.43.375
   Zagzag D, 2000, PEDIATRICS, V106, P1045, DOI 10.1542/peds.106.5.1045
NR 10
TC 5
Z9 5
U1 0
U2 1
PU SPRINGER
PI NEW YORK
PA 233 SPRING STREET, NEW YORK, NY 10013 USA
SN 0256-7040
J9 CHILD NERV SYST
JI Childs Nerv. Syst.
PD JAN
PY 2007
VL 23
IS 1
BP 117
EP 121
DI 10.1007/s00381-006-0168-6
PG 5
WC Clinical Neurology; Pediatrics; Surgery
WE Science Citation Index Expanded (SCI-EXPANDED)
SC Neurosciences & Neurology; Pediatrics; Surgery
GA 112CA
UT WOS:000242501600019
PM 16983574
DA 2023-08-27
ER

PT J
AU Warren, KE
   Gururangan, S
   Geyer, JR
   McLendon, RE
   Poussaint, TY
   Wallace, D
   Balis, FM
   Berg, SL
   Packer, RJ
   Goldman, S
   Minturn, JE
   Pollack, IF
   Boyett, JM
   Kun, LE
AF Warren, Katherine E.
   Gururangan, Sri
   Geyer, J. Russell
   McLendon, Roger E.
   Poussaint, Tina Young
   Wallace, Dana
   Balis, Frank M.
   Berg, Stacey L.
   Packer, Roger J.
   Goldman, Stewart
   Minturn, Jane E.
   Pollack, Ian F.
   Boyett, James M.
   Kun, Larry E.
TI A phase II study of O6-benzylguanine and temozolomide in pediatric
   patients with recurrent or progressive high-grade gliomas and brainstem
   gliomas: a Pediatric Brain Tumor Consortium study
SO JOURNAL OF NEURO-ONCOLOGY
LA English
DT Article
DE Glioma; Pediatric; Resistance; Alkylating agent; Brainstem glioma; AGT;
   MGMT
ID MGMT PROMOTER METHYLATION; O-6-ALKYLGUANINE-DNA ALKYLTRANSFERASE;
   MALIGNANT GLIOMA; ANTITUMOR IMIDAZOTETRAZINES; PLUS O-6-BENZYLGUANINE;
   MISMATCH REPAIR; CHILDREN; TRIAL; METHYLTRANSFERASE; PHARMACOKINETICS
AB To estimate the sustained (a parts per thousand yen8 weeks) objective response rate in pediatric patients with recurrent or progressive high-grade gliomas (HGG, Stratum A) or brainstem gliomas (BSG, Stratum B) treated with the combination of O6-benzylguanine (O6BG) and temozolomide(A (R)) (TMZ). Patients received O6BG 120 mg/m(2)/d IV followed by TMZ 75 mg/m(2)/d orally daily for 5 consecutive days of each 28-day course. The target objective response rate to consider the combination active was 17%. A two-stage design was employed. Forty-three patients were enrolled; 41 were evaluable for response, including 25 patients with HGG and 16 patients with BSG. The combination of O6BG and TMZ was tolerable, and the primary toxicities were myelosuppression and gastrointestinal symptoms. One sustained (a parts per thousand yen8 weeks) partial response was observed in the HGG cohort; no sustained objective responses were observed in the BSG cohort. Long-term (a parts per thousand yen6 courses) stable disease (SD) was observed in 4 patients in Stratum A and 1 patient in Stratum B. Of the 5 patients with objective response or long-term SD, 3 underwent central review with 2 reclassified as low-grade gliomas. The combination of O6BG and TMZ did not achieve the target response rate for activity in pediatric patients with recurrent or progressive HGG and BSG.
C1 [Warren, Katherine E.] NCI, Pediat Oncol Branch, Bethesda, MD 20892 USA.
   [Gururangan, Sri; McLendon, Roger E.] Duke Univ, Med Ctr, Durham, NC 27706 USA.
   [Geyer, J. Russell] Seattle Childrens Hosp, Seattle, WA USA.
   [Poussaint, Tina Young] Childrens Hosp, Boston, MA 02115 USA.
   [Wallace, Dana; Boyett, James M.] Operat & Biostat Ctr Pediat Brain Tumor Consortiu, Memphis, TN USA.
   [Balis, Frank M.; Minturn, Jane E.] Childrens Hosp, Philadelphia, PA 19104 USA.
   [Berg, Stacey L.] Texas Childrens Canc Ctr, Houston, TX USA.
   [Packer, Roger J.] Childrens Natl Med Ctr, Washington, DC 20010 USA.
   [Goldman, Stewart] Childrens Mem Hosp, Chicago, IL 60614 USA.
   [Pollack, Ian F.] Childrens Hosp Pittsburgh, Pittsburgh, PA 15213 USA.
   [Kun, Larry E.] St Jude Childrens Res Hosp, Memphis, TN 38105 USA.
C3 National Institutes of Health (NIH) - USA; NIH National Cancer Institute
   (NCI); Duke University; Seattle Children's Hospital; Harvard University;
   Boston Children's Hospital; University of Pennsylvania; Pennsylvania
   Medicine; Childrens Hospital of Philadelphia; Texas Children's Cancer
   Center; Children's National Health System; Ann & Robert H. Lurie
   Children's Hospital of Chicago; Pennsylvania Commonwealth System of
   Higher Education (PCSHE); University of Pittsburgh; St Jude Children's
   Research Hospital
RP Warren, KE (通讯作者)，NCI, Pediat Oncol Branch, Bldg 10 CRC,Room 1-5750, Bethesda, MD 20892 USA.
EM
OI Balis, Frank/0000-0002-8117-6926
FU NIH [U01 CA81457, 5M01RR000188]; American Lebanese Syrian Associated
   Charities
FX This work was supported in part by NIH grant U01 CA81457 for the
   Pediatric Brain Tumor Consortium, NIH grant 5M01RR000188, and the
   American Lebanese Syrian Associated Charities, and the intramural
   program of the NIH. The authors and the PBTC acknowledge the protocol
   management of Mr. Christopher Smith, the central pathology review
   participation of Dr. Adesina Adekunle and Dr. Veena Rajaram and the
   statistical support of Mr. Shawn Lesh.
CR BERG SL, 1995, CANCER RES, V55, P4606
   Bobola MS, 2005, CLIN CANCER RES, V11, P2747, DOI 10.1158/1078-0432.CCR-04-2045
   Brada M, 2001, ANN ONCOL, V12, P259, DOI 10.1023/A:1008382516636
   Brandes AA, 2002, ONCOLOGY-BASEL, V63, P38, DOI 10.1159/000065718
   Brandes AA, 2006, J CLIN ONCOL, V24, P4746, DOI 10.1200/JCO.2006.06.3891
   Broniscer A, 2005, CANCER-AM CANCER SOC, V103, P133, DOI 10.1002/cncr.20741
   Broniscer A, 2007, CLIN CANCER RES, V13, P6712, DOI 10.1158/1078-0432.CCR-07-1016
   Chibbaro S, 2004, J NEURO-ONCOL, V67, P77, DOI 10.1023/B:NEON.0000021728.36747.93
   Cohen KJ, 2011, NEURO-ONCOLOGY, V13, P410, DOI 10.1093/neuonc/noq205
   Cohen KJ, 2011, NEURO-ONCOLOGY, V13, P317, DOI 10.1093/neuonc/noq191
   DENNY BJ, 1994, BIOCHEMISTRY-US, V33, P9045, DOI 10.1021/bi00197a003
   Donson AM, 2007, PEDIATR BLOOD CANCER, V48, P403, DOI 10.1002/pbc.20803
   FRIEDMAN HS, 1995, CANCER RES, V55, P2853
   Friedman HS, 2002, MOL CANCER THER, V1, P943
   Friedman HS, 1998, J CLIN ONCOL, V16, P3570, DOI 10.1200/JCO.1998.16.11.3570
   Friedman HS, 1998, J CLIN ONCOL, V16, P3851, DOI 10.1200/JCO.1998.16.12.3851
   Gerson SL, 2002, J CLIN ONCOL, V20, P2388, DOI 10.1200/JCO.2002.06.110
   Hegi ME, 2008, J CLIN ONCOL, V26, P4189, DOI 10.1200/JCO.2007.11.5964
   Ishii D, 2007, NEUROL MED-CHIR, V47, P341, DOI 10.2176/nmc.47.341
   KARRAN P, 1994, BIOESSAYS, V16, P833, DOI 10.1002/bies.950161110
   Kokkinakis DM, 2001, CLIN CANCER RES, V7, P421
   Lashford LS, 2002, J CLIN ONCOL, V20, P4684, DOI 10.1200/JCO.2002.08.141
   Maxwell JA, 2006, MOL CANCER THER, V5, P2531, DOI 10.1158/1535-7163.MCT-06-0106
   NEWLANDS ES, 1992, BRIT J CANCER, V65, P287, DOI 10.1038/bjc.1992.57
   Nicholson HS, 2007, CANCER, V110, P1542, DOI 10.1002/cncr.22961
   Panetta JC, 2003, CANCER CHEMOTH PHARM, V52, P435, DOI 10.1007/s00280-003-0670-4
   Patel M, 2003, J NEURO-ONCOL, V61, P203, DOI 10.1023/A:1022592913323
   Quinn JA, 2002, J CLIN ONCOL, V20, P2277, DOI 10.1200/JCO.2002.09.084
   Quinn JA, 2009, J CLIN ONCOL, V27, P1262, DOI 10.1200/JCO.2008.18.8417
   Rich JN, 2005, CLIN CANCER RES, V11, P8145, DOI 10.1158/1078-0432.CCR-05-0319
   Roos WP, 2007, ONCOGENE, V26, P186, DOI 10.1038/sj.onc.1209785
   Sankar A, 1999, ANTI-CANCER DRUG, V10, P179, DOI 10.1097/00001813-199902000-00006
   STEVENS MFG, 1987, CANCER RES, V47, P5846
   Stupp R, 2002, J CLIN ONCOL, V20, P1375, DOI 10.1200/JCO.20.5.1375
   Stupp R, 2007, J CLIN ONCOL, V25, P4127, DOI 10.1200/JCO.2007.11.8554
   Warren KE, 2005, J CLIN ONCOL, V23, P7646, DOI 10.1200/JCO.2005.02.0024
   Wedge SR, 1996, BRIT J CANCER, V73, P1049, DOI 10.1038/bjc.1996.203
   Yung WKA, 1999, J CLIN ONCOL, V17, P2762, DOI 10.1200/JCO.1999.17.9.2762
NR 38
TC 52
Z9 55
U1 0
U2 12
PU SPRINGER
PI NEW YORK
PA ONE NEW YORK PLAZA, SUITE 4600, NEW YORK, NY, UNITED STATES
SN 0167-594X
EI 1573-7373
J9 J NEURO-ONCOL
JI J. Neuro-Oncol.
PD FEB
PY 2012
VL 106
IS 3
BP 643
EP 649
DI 10.1007/s11060-011-0709-z
PG 7
WC Oncology; Clinical Neurology
WE Science Citation Index Expanded (SCI-EXPANDED)
SC Oncology; Neurosciences & Neurology
GA 879LX
UT WOS:000299331800022
PM 21968943
OA Green Accepted
DA 2023-08-27
ER

PT J
AU Yin, LX
   Zhang, LW
AF Yin, Luxin
   Zhang, Liwei
TI Correlation Between MRI Findings and Histological Diagnosis of Brainstem
   Glioma
SO CANADIAN JOURNAL OF NEUROLOGICAL SCIENCES
LA English
DT Article
ID MANAGEMENT; TUMORS; CHILDREN; ADULT
AB Objective: In most studies, treatment decisions of brainstem glioma are based solely on MRI features and do not incorporate a histopathological diagnosis. In the current study, we sought to compare MRI characteristics with histopathological findings of bainstem glioma. Methods: From April 2003 through April 2012, 150 patients were diagnosed with brainstem gliomas by MRI and microsurgically treated in Tiantan Hospital, Beijing, China. All the MRI and histopathological findings of these patients were respectively reviewed. Results: Of the 150 patients, 65 were female and 85 were male, 120 were adults and 30 were children (age < 18 years), 108 were low-grade glioma (72.0%), 35 were high-grade glioma (23.3%). The accuracy of the MRI diagnosis for brainstem glioma was 95.3%. Data analysis of the MRI findings revealed that a focal lesion was associated with a more favorable histopathological diagnosis in intrinsic (P=0.005) and exophytic (P=0.001) brainstem glioma patients. In the intrinsic diffuse type, tumors without enhancement had more favorable pathological findings (P=0.009). Conclusions: To our knowledge, this is the largest case series of this nature reported in the literature to date. The results of this study suggest that MRI features of brainstem gliomas could predict some pathological features and guide prognosis, choice of biopsy and treatment modalities. The pathology of tumors with a focal appearance on MRI was associated with a prognosis that was significantly better than their diffuse counterparts. For the intrinsic diffuse gliomas, non-enhancing tumors had pathology suggestive of a favorable prognosis.
C1 [Yin, Luxin; Zhang, Liwei] Capital Med Univ, Beijing Tiantan Hosp, Dept Neurosurg, TiantanXili 6, Beijing 100050, Peoples R China.
C3 Capital Medical University
RP Yin, LX (通讯作者)，Capital Med Univ, Beijing Tiantan Hosp, Dept Neurosurg, TiantanXili 6, Beijing 100050, Peoples R China.
EM
CR ALBRIGHT AL, 1986, J NEUROSURG, V65, P751, DOI 10.3171/jns.1986.65.6.0751
   Bowers DC, 2000, PEDIATR NEUROSURG, V32, P24, DOI 10.1159/000028893
   BOYDSTON WR, 1992, PEDIATR NEUROSURG, V17, P234, DOI 10.1159/000120603
   Chaddad Neto C, 2007, ARQ NEURO-PSIQUIAT, V65, P996, DOI 10.1590/S0004-282X2007000600015
   Choux M, 2000, PEDIAT NEUROSURGERY, P471
   Dellaretti M, 2012, NEURO-ONCOLOGY, V14, P381, DOI 10.1093/neuonc/nor215
   EPSTEIN FJ, 1993, J NEUROSURG, V78, P408, DOI 10.3171/jns.1993.78.3.0408
   Fischbein NJ, 1996, PEDIATR NEUROSURG, V24, P9, DOI 10.1159/000121010
   Freeman CR, 1998, INT J RADIAT ONCOL, V40, P265, DOI 10.1016/S0360-3016(97)00572-5
   Grant GA, 1999, PEDIATR NEUROSURG, V31, P170, DOI 10.1159/000028857
   Guillamo JS, 2001, CURR OPIN NEUROL, V14, P711, DOI 10.1097/00019052-200112000-00006
   Jallo GI, 2004, CHILD NERV SYST, V20, P143, DOI 10.1007/s00381-003-0870-6
   Kaplan AM, 1996, PEDIATR NEUROSURG, V24, P185, DOI 10.1159/000121036
   Kim BS, 2004, SURG NEUROL, V62, P64, DOI 10.1016/j.surneu.2003.07.001
   Koziarski A, 2004, ACTA NEUROCHIR, V146, P169, DOI 10.1007/s00701-003-0158-3
   Liu Q, 2008, J NEUROSURG, V109, P849, DOI 10.3171/JNS/2008/109/11/0849
   Rachinger W, 2009, J NEUROL NEUROSUR PS, V80, P1134, DOI 10.1136/jnnp.2009.174250
   Tobias ME, 2008, CHILD NERV SYST, V24, P219, DOI 10.1007/s00381-007-0405-7
   VANDERTOP WP, 1992, NEUROSURGERY, V31, P186, DOI 10.1227/00006123-199208000-00003
   Walker DA, 1999, ARCH DIS CHILD, V80, P558, DOI 10.1136/adc.80.6.558
   Yeh DD, 2002, NEUROSURGERY, V50, P735, DOI 10.1097/00006123-200204000-00009
   Zagzag D, 2000, PEDIATRICS, V106, P1045, DOI 10.1542/peds.106.5.1045
NR 22
TC 12
Z9 15
U1 0
U2 4
PU CAMBRIDGE UNIV PRESS
PI NEW YORK
PA 32 AVENUE OF THE AMERICAS, NEW YORK, NY 10013-2473 USA
SN 0317-1671
EI 2057-0155
J9 CAN J NEUROL SCI
JI Can. J. Neurol. Sci.
PD MAY
PY 2013
VL 40
IS 3
BP 348
EP 354
DI 10.1017/S0317167100014293
PG 7
WC Clinical Neurology
WE Science Citation Index Expanded (SCI-EXPANDED)
SC Neurosciences & Neurology
GA V43UT
UT WOS:000209707000014
PM 23603170
OA Bronze
DA 2023-08-27
ER

PT J
AU Fric, R
   Eide, PK
AF Fric, Radek
   Eide, Per Kristian
TI The presigmoid approach for removal of tumours causing ventral
   compression of the brainstem. Surgical results and postoperative quality
   of life
SO BRITISH JOURNAL OF NEUROSURGERY
LA English
DT Article
ID TRANSPETROSAL-TRANSTENTORIAL APPROACH; PETROCLIVAL MENINGIOMAS; FOSSA
   APPROACH; EXPERIENCE; CLIVUS
AB Methods. A review of all our patients operated on with removal of tumours causing ventral compression of the brainstem in whom either the presigmoid or combined presigmoid/retrosigmoid approaches were used. Post-operative quality of life was assessed using the SF-36 questionnaire.
   Results. A total of 12 consecutive patients were included. A favourable complication profile was found. Cerebrospinal fluid leakage and secondary hydrocephalus were the most frequent complications. Post-operative scores from Karnofsky Performance Scale and ECOG after median 24 months were median 85 and 1, respectively, representing ability to carry on daily activities. Nevertheless, quality of life as assessed subjectively by SF-36 showed significantly reduced values as compared with normative values from Norwegian population.
   Conclusions. We found favourable surgical results when using the presigmoid approach alone or combined with the retrosigmoid route for large-sized extrinsic tumours causing ventral compression of the brainstem, both regarding complication profile, degree of tumour removal and functional outcome. However, impact of disease and surgery on quality of life appears to be more significant when assessed subjectively by the patients.
C1 [Fric, Radek; Eide, Per Kristian] Natl Hosp Norway, Oslo Univ Hosp, Dept Neurosurg, N-0027 Oslo, Norway.
C3 University of Oslo; National Hospital Norway
RP Fric, R (通讯作者)，Natl Hosp Norway, Oslo Univ Hosp, Dept Neurosurg, Sognsvannsveien 20, N-0027 Oslo, Norway.
EM
CR [Anonymous], 2000, MANUAL INTERPRETATIO
   Bambakidis NC, 2007, NEUROSURGERY, V61, P202, DOI [10.1227/01.neu.0000303218.61230.39, 10.1227/01.NIEU.0000280119.36270.BA]
   BRICOLO AP, 1992, NEUROSURGERY, V31, P813, DOI 10.1227/00006123-199211000-00001
   Cho CW, 2002, NEUROSURGERY, V51, P708, DOI 10.1097/00006123-200209000-00017
   Couldwell WT, 1996, J NEUROSURG, V84, P20, DOI 10.3171/jns.1996.84.1.0020
   Goel A, 2004, SURG NEUROL, V62, P332, DOI 10.1016/j.surneu.2003.12.008
   Goel A, 1999, BRIT J NEUROSURG, V13, P270, DOI 10.1080/02688699943673
   GOEL A, 1995, ACTA NEUROCHIR, V135, P78, DOI 10.1007/BF02307418
   HAKUBA A, 1988, SURG NEUROL, V30, P108, DOI 10.1016/0090-3019(88)90095-X
   Jung HW, 2000, NEUROSURGERY, V46, P567, DOI 10.1097/00006123-200003000-00008
   KARNOFSKY DA, 1948, CANCER-AM CANCER SOC, V1, P634, DOI 10.1002/1097-0142(194811)1:4<634::AID-CNCR2820010410>3.0.CO;2-L
   KAWASE T, 1991, NEUROSURGERY, V28, P869, DOI 10.1227/00006123-199106000-00014
   Krisht Ali F, 2005, Neurosurg Focus, V19, pE2
   Little KM, 2005, NEUROSURGERY, V56, P546, DOI 10.1227/01.NEU.0000153906.12640.62
   Loge JH, 1998, SCAND J SOC MED, V26, P250
   Mathiesen T, 2007, NEUROSURGERY, V60, P982, DOI 10.1227/01.NEU.0000255476.06247.F1
   Natarajan SK, 2007, NEUROSURGERY, V60, P965, DOI 10.1227/01.NEU.0000255472.52882.D6
   Neil-Dwyer G, 2000, ACTA NEUROCHIR, V142, P367, DOI 10.1007/s007010050444
   OKEN MM, 1982, AM J CLIN ONCOL-CANC, V5, P649, DOI 10.1097/00000421-198212000-00014
   Park CK, 2006, SURG NEUROL, V66, P160, DOI 10.1016/j.surneu.2005.12.024
   Roberti F, 2001, SURG NEUROL, V56, P8, DOI 10.1016/S0090-3019(01)00479-7
   SAMII M, 1992, ACTA NEUROCHIR, V118, P27, DOI 10.1007/BF01400723
   Seifert V, 2003, ACTA NEUROCHIR, V145, P631, DOI 10.1007/s00701-003-0086-2
   SEKHAR LN, 1994, J NEUROSURG, V81, P860, DOI 10.3171/jns.1994.81.6.0860
   SPETZLER RF, 1992, J NEUROSURG, V76, P588, DOI 10.3171/jns.1992.76.4.0588
   Tahara A, 2009, J CLIN NEUROSCI, V16, P655, DOI 10.1016/j.jocn.2008.08.011
   Zentner J, 1997, J NEUROL NEUROSUR PS, V62, P341, DOI 10.1136/jnnp.62.4.341
NR 27
TC 8
Z9 8
U1 0
U2 2
PU TAYLOR & FRANCIS LTD
PI ABINGDON
PA 2-4 PARK SQUARE, MILTON PARK, ABINGDON OR14 4RN, OXON, ENGLAND
SN 0268-8697
EI 1360-046X
J9 BRIT J NEUROSURG
JI Br. J. Neurosurg.
PD FEB
PY 2011
VL 25
IS 1
BP 86
EP 93
DI 10.3109/02688697.2010.525266
PG 8
WC Clinical Neurology; Surgery
WE Science Citation Index Expanded (SCI-EXPANDED)
SC Neurosciences & Neurology; Surgery
GA 722BO
UT WOS:000287405000012
PM 20979440
DA 2023-08-27
ER

PT J
AU Nguyen, TT
   Wray, AC
   Laidlaw, JD
AF Nguyen, TT
   Wray, AC
   Laidlaw, JD
TI Midbrain and thalamic haemorrhage as first presentation of intracerebral
   glioma
SO JOURNAL OF CLINICAL NEUROSCIENCE
LA English
DT Article
ID INTRA-CEREBRAL HEMORRHAGE; BRAIN-TUMOR; INTRACRANIAL TUMORS; NEOPLASMS;
   HEMATOMA; TOMOGRAPHY; STROKE
AB A case of spontaneous intracerebral haemorrhage (midbrain and thalamic, with intraventricular extension) as the first presentation of an anaplastic astrocytoma is presented. Multiple CT scans and cerebral angiography failed to identify any vascular or neoplastic cause for the haemorrhage, and a presumptive diagnosis of hypertensive haemorrhage was made. Shunting of hydrocephalus was followed by early clinical improvement. However, delayed progressive deterioration necessitated MRI scan, which demonstrated a mass lesion in the basal ganglia and midbrain. This was subsequently found to be anaplastic astrocytoma on biopsy. The literature regarding this uncommon presentation of spontaneous intracerebral haemorrhage from an occult brain tumour is reviewed. The need for investigation and close follow-up of presumed hypertensive haemorrhage is emphasised by this case. (c) 2005 Elsevier Ltd. All rights reserved.
C1 Univ Melbourne, Private Med Ctr, Royal Melbourne Hosp, Dept Neurosurg, Parkville, Vic 3050, Australia.
   Univ Melbourne, Dept Surg, Royal Melbourne Hosp, Parkville, Vic 3050, Australia.
C3 Royal Melbourne Hospital; University of Melbourne; Royal Melbourne
   Hospital; University of Melbourne
RP Laidlaw, JD (通讯作者)，Univ Melbourne, Private Med Ctr, Royal Melbourne Hosp, Dept Neurosurg, Suite 13, Parkville, Vic 3050, Australia.
EM
CR ALBERT FK, 1986, NEUROCHIRURGIA, V29, P67
   BARTH H, 1994, NERVENARZT, V65, P854
   BITOH S, 1984, SURG NEUROL, V22, P57, DOI 10.1016/0090-3019(84)90230-1
   Bromowicz J, 1983, Neurol Neurochir Pol, V17, P471
   BROTT T, 1986, STROKE, V17, P1078, DOI 10.1161/01.STR.17.6.1078
   DRURY I, 1984, NEUROLOGY, V34, P653, DOI 10.1212/WNL.34.5.653
   FUJITA K, 1980, Neurological Surgery, V8, P929
   FURLAN AJ, 1979, ANN NEUROL, V5, P367, DOI 10.1002/ana.410050410
   GARRAWAY WM, 1979, NEW ENGL J MED, V300, P449, DOI 10.1056/NEJM197903013000901
   HAMILTON MG, 1994, NEUROVASCULAR SURG, P477
   HEROLD S, 1982, J NEUROL, V228, P267, DOI 10.1007/BF00313417
   KONDZIOLKA D, 1987, J NEUROSURG, V67, P852, DOI 10.3171/jns.1987.67.6.0852
   KOTHBAUER P, 1979, ACTA NEUROCHIR, V49, P35, DOI 10.1007/BF01809172
   Lieu AS, 1999, J FORMOS MED ASSOC, V98, P365
   Little JR, 1979, STROKE, V10, P238
   Morgenstern LB, 1999, J NEURO-ONCOL, V44, P47, DOI 10.1023/A:1006237421731
   Niedermayer I, 1999, RADIOLOGE, V39, P821, DOI 10.1007/s001170050718
   SACCO RL, 1984, NEUROLOGY, V34, P847, DOI 10.1212/WNL.34.7.847
   SALCMAN M, 1999, INTRACEREBRAL HEMATO, P65
   Schrader B, 2000, ACTA NEUROCHIR, V142, P979, DOI 10.1007/s007010070052
   SCHUETZ H, 1992, STROKE, V23, P653, DOI 10.1161/01.STR.23.5.653
   SCOTT M, 1975, J NEUROSURG, V42, P338, DOI 10.3171/jns.1975.42.3.0338
   Tung GA, 2003, NEURORADIOLOGY, V45, P357, DOI 10.1007/s00234-003-0994-0
   WAKAI S, 1982, NEUROSURGERY, V10, P437, DOI 10.1227/00006123-198204000-00004
   Young N, 1993, Australas Radiol, V37, P315, DOI 10.1111/j.1440-1673.1993.tb00086.x
   Zhu XL, 1997, STROKE, V28, P1406, DOI 10.1161/01.STR.28.7.1406
NR 26
TC 5
Z9 5
U1 0
U2 2
PU ELSEVIER SCI LTD
PI OXFORD
PA THE BOULEVARD, LANGFORD LANE, KIDLINGTON, OXFORD OX5 1GB, OXON, ENGLAND
SN 0967-5868
EI 1532-2653
J9 J CLIN NEUROSCI
JI J. Clin. Neurosci.
PD NOV
PY 2005
VL 12
IS 8
BP 946
EP 949
DI 10.1016/j.jocn.2004.10.015
PG 4
WC Clinical Neurology; Neurosciences
WE Science Citation Index Expanded (SCI-EXPANDED)
SC Neurosciences & Neurology
GA 998WX
UT WOS:000234351700022
PM 16326274
DA 2023-08-27
ER

PT J
AU Wang, ZH
   Xu, C
   Diplas, BH
   Moure, CJ
   Chen, CPJ
   Chen, LH
   Du, CZ
   Zhu, HS
   Greer, PK
   Zhang, LW
   He, YP
   Waitkus, MS
   Yan, H
AF Wang, Zhaohui
   Xu, Cheng
   Diplas, Bill H.
   Moure, Casey J.
   Chen, Chin-Pu Jason
   Chen, Lee H.
   Du, Changzheng
   Zhu, Huishan
   Greer, Paula K.
   Zhang, Liwei
   He, Yiping
   Waitkus, Matthew S.
   Yan, Hai
TI Targeting Mutant PPM1D Sensitizes Diffuse Intrinsic Pontine Glioma Cells
   to the PARP Inhibitor Olaparib
SO MOLECULAR CANCER RESEARCH
LA English
DT Article
ID HOMOLOGOUS RECOMBINATION; WIP1 PHOSPHATASE; MUTATIONS; ACVR1; PLATFORM
AB Diffuse intrinsic pontine glioma (DIPG) is an invariably fatal brain tumor occurring predominantly in children. Up to 90% of pediatric DIPGs harbor a somatic heterozygous mutation resulting in the replacement of lysine 27 with methionine (K27M) in genes encoding histone H3.3 (H3F3A, 65%) or H3.1 (HIST1H3B, 25%). Several studies have also identified recurrent truncating mutations in the gene encoding protein phosphatase 1D, PPM1D, in 9%-23% of DIPGs. Here, we sought to investigate the therapeutic potential of targeting PPM1D, alone or in combination with inhibitors targeting specific components of DNA damage response pathways in patient-derived DIPG cell lines. We found that GSK2830371, an allosteric PPM1D inhibitor, suppressed the proliferation of PPM1D-mutant, but not PPM1D wild-type DIPG cells. We further observed that PPM1D inhibition sensitized PPM1D-mutant DIPG cells to PARP inhibitor (PARPi) treatment. Mechanistically, combined PPM1D and PARP inhibition show synergistic effects on suppressing a p53-dependent RAD51 expression and the formation of RAD51 nuclear foci, possibly leading to impaired homologous recombination (HR)-mediated DNA repair in PPM1D-mutant DIPG cells. Collectively, our findings reveal the potential role of the PPM1D-p53 signaling axis in the regulation of HR-mediated DNA repair and provide preclinical evidence demonstrating that combined inhibition of PPM1D and PARP1/2 may be a promising therapeutic combination for targeting PPM1D-mutant DIPG tumors.
C1 [Wang, Zhaohui; Xu, Cheng; Diplas, Bill H.; Moure, Casey J.; Chen, Chin-Pu Jason; Chen, Lee H.; Du, Changzheng; Zhu, Huishan; Greer, Paula K.; He, Yiping; Waitkus, Matthew S.; Yan, Hai] Duke Univ, Dept Pathol, Durham, NC 27706 USA.
   [Wang, Zhaohui; Xu, Cheng; Diplas, Bill H.; Moure, Casey J.; Chen, Chin-Pu Jason; Chen, Lee H.; Du, Changzheng; Zhu, Huishan; Greer, Paula K.; He, Yiping; Waitkus, Matthew S.; Yan, Hai] Duke Univ, Med Ctr, Preston Robert Tisch Brain Tumor Ctr, Durham, NC 27710 USA.
   [Zhang, Liwei] Capital Med Univ, Beijing Tiantan Hosp, Dept Neurosurg, Beijing, Peoples R China.
C3 Duke University; Duke University; Capital Medical University
RP Waitkus, MS; Yan, H (通讯作者)，Duke Med Ctr, DUMC-3156,199B MSRB Res Dr, Durham, NC 27710 USA.
EM
RI Waitkus, Matthew/ISS-3412-2023
OI Waitkus, Matthew/0000-0002-7116-6978; Diplas, Bill/0000-0001-9248-7351
FU American Brain Tumor Association in honor of the Bradley Benton Davis
   Memorial Foundation [BRF1600002]; Southeastern Brain Tumor Foundation
FX Z. Wang received a fellowship (BRF1600002) grant from The American Brain
   Tumor Association in honor of the Bradley Benton Davis Memorial
   Foundation. Z. Wang and H. Yan received a research grant (SBTF-2015)
   from the Southeastern Brain Tumor Foundation. The authors would like to
   thank Drs. C. David James and Angel Montero Carcaboso for the generous
   gifts of DIPG cell lines. We also thank Dr. Jiaoti Huang for the
   generous gift of the RAD51 antibody. The authors would like to thank the
   core facilities used in this study, including the Duke Cancer Institute
   Flow Cytometry Shared Resource (Nancy Martin, Lynn Martinek, and Michael
   Cook), the Light Microscopy Core Facility (Lisa Cameron, Benjamin
   Carlson, and Yasheng Gao), and the Functional Genomics Core Facility
   (Sufeng Li and So Young Kim).
CR Adimoolam S, 2007, P NATL ACAD SCI USA, V104, P19482, DOI 10.1073/pnas.0707828104
   Akamandisa MP, 2019, NEURO-ONCOLOGY, V21, P786, DOI 10.1093/neuonc/noz053
   Arias-Lopez C, 2006, EMBO REP, V7, P219, DOI 10.1038/sj.embor.7400587
   Brandsma I, 2017, EXPERT OPIN INV DRUG, V26, P1341, DOI 10.1080/13543784.2017.1389895
   Buczkowicz P, 2015, FRONT ONCOL, V5, DOI 10.3389/fonc.2015.00147
   Buczkowicz P, 2014, NAT GENET, V46, P451, DOI 10.1038/ng.2936
   Burdova K, 2019, CELLS-BASEL, V8, DOI 10.3390/cells8101258
   Castroviejo Bermejo M, 2018, EMBO MOL MED, V10, pe9172
   Clymer J, 2018, FRONT ONCOL, V8, DOI 10.3389/fonc.2018.00169
   Cruz C, 2018, ANN ONCOL, V29, P1203, DOI 10.1093/annonc/mdy099
   Di Veroli GY, 2016, BIOINFORMATICS, V32, P2866, DOI 10.1093/bioinformatics/btw230
   Diplas BH, 2018, NAT COMMUN, V9, DOI 10.1038/s41467-018-04448-6
   Dudgeon C, 2013, CELL CYCLE, V12, P2656, DOI 10.4161/cc.25694
   El Ayoubi R, 2017, J NEURO-ONCOL, V134, P465, DOI 10.1007/s11060-017-2523-8
   Fons NR, 2019, NAT COMMUN, V10, DOI 10.1038/s41467-019-11732-6
   Fontebasso AM, 2014, NAT GENET, V46, P462, DOI 10.1038/ng.2950
   Gilmartin AG, 2014, NAT CHEM BIOL, V10, P181, DOI [10.1038/NCHEMBIO.1427, 10.1038/nchembio.1427]
   Gupta A, 2014, RADIAT RES, V181, P1, DOI 10.1667/RR13572.1
   Hashizume R, 2012, J NEURO-ONCOL, V110, P305, DOI 10.1007/s11060-012-0973-6
   Hoffman LM, 2018, J CLIN ONCOL, V36, P1963, DOI 10.1200/JCO.2017.75.9308
   Hsu JI, 2018, CELL STEM CELL, V23, P700, DOI 10.1016/j.stem.2018.10.004
   Jiang JH, 2011, CANCER RES, V71, P5546, DOI 10.1158/0008-5472.CAN-10-3423
   Kahn JD, 2018, BLOOD, V132, P1095, DOI 10.1182/blood-2018-05-850339
   Kalev P, 2012, CANCER RES, V72, P6414, DOI 10.1158/0008-5472.CAN-12-1667
   Kim H, 2017, CLIN CANCER RES, V23, P3097, DOI 10.1158/1078-0432.CCR-16-2273
   Kleiblova P, 2013, J CELL BIOL, V201, P511, DOI 10.1083/jcb.201210031
   Litton JK, 2018, NEW ENGL J MED, V379, P753, DOI 10.1056/NEJMoa1802905
   Lord CJ, 2017, SCIENCE, V355, P1152, DOI 10.1126/science.aam7344
   Lord CJ, 2016, NAT REV CANCER, V16, P110, DOI 10.1038/nrc.2015.21
   Lu XB, 2004, MOL CELL, V15, P621, DOI 10.1016/j.molcel.2004.08.007
   Maranto C, 2018, CLIN CANCER RES, V24, P1917, DOI 10.1158/1078-0432.CCR-17-2768
   Mendes-Pereira AM, 2009, EMBO MOL MED, V1, P315, DOI 10.1002/emmm.200900041
   Moon SH, 2010, J BIOL CHEM, V285, P12935, DOI 10.1074/jbc.M109.071696
   Mueller S, 2019, INT J CANCER, V145, P1889, DOI 10.1002/ijc.32258
   Nguyen TA, 2010, DNA REPAIR, V9, P813, DOI 10.1016/j.dnarep.2010.04.005
   Nikbakht H, 2016, NAT COMMUN, V7, DOI 10.1038/ncomms11185
   Pechackova S, 2017, J MOL MED, V95, P589, DOI 10.1007/s00109-017-1536-2
   Pilie PG, 2019, NAT REV CLIN ONCOL, V16, P81, DOI 10.1038/s41571-018-0114-z
   Ran FA, 2013, NAT PROTOC, V8, P2281, DOI 10.1038/nprot.2013.143
   Schindelin J, 2012, NAT METHODS, V9, P676, DOI [10.1038/NMETH.2019, 10.1038/nmeth.2019]
   Sizemore ST, 2018, MOL CANCER RES, V16, P1092, DOI 10.1158/1541-7786.MCR-18-0106
   Taylor KR, 2014, NAT GENET, V46, P457, DOI 10.1038/ng.2925
   Tsoli Maria, 2018, Oncotarget, V9, P7541, DOI 10.18632/oncotarget.24045
   Vinci M, 2015, JOVE-J VIS EXP, V2015, P1
   Waitkus MS, 2018, CANCER RES, V78, P36, DOI 10.1158/0008-5472.CAN-17-1352
   Wu G, 2014, NAT GENET, V46, P444, DOI 10.1038/ng.2938
   Wu G, 2012, NAT GENET, V44, P251, DOI 10.1038/ng.1102
   Xu C, 2017, ONCOTARGET, V8, P76644, DOI 10.18632/oncotarget.19656
   Zhang LW, 2014, NAT GENET, V46, P726, DOI 10.1038/ng.2995
   Zhu YH, 2012, PROG MOL BIOL TRANSL, V106, P307, DOI 10.1016/B978-0-12-396456-4.00001-8
NR 50
TC 18
Z9 18
U1 1
U2 4
PU AMER ASSOC CANCER RESEARCH
PI PHILADELPHIA
PA 615 CHESTNUT ST, 17TH FLOOR, PHILADELPHIA, PA 19106-4404 USA
SN 1541-7786
EI 1557-3125
J9 MOL CANCER RES
JI Mol. Cancer Res.
PD JUL
PY 2020
VL 18
IS 7
BP 968
EP 980
DI 10.1158/1541-7786.MCR-19-0507
PG 13
WC Oncology; Cell Biology
WE Science Citation Index Expanded (SCI-EXPANDED)
SC Oncology; Cell Biology
GA MG4PZ
UT WOS:000546016600003
PM 32229503
OA Green Submitted
DA 2023-08-27
ER

PT J
AU Aotsuka, Y
   Uzawa, A
   Nishimura, K
   Kojima, K
   Yamaguchi, M
   Makino, T
   Nakamichi, K
   Saijo, M
   Kuwabara, S
AF Aotsuka, Yuya
   Uzawa, Akiyuki
   Nishimura, Kazutaka
   Kojima, Kazuho
   Yamaguchi, Mika
   Makino, Takahiro
   Nakamichi, Kazuo
   Saijo, Masayuki
   Kuwabara, Satoshi
TI Progressive Multifocal Leukoencephalopathy Localized in the Cerebellum
   and Brainstem Associated with Idiopathic CD4(+) T Lymphocytopenia
SO INTERNAL MEDICINE
LA English
DT Article
DE brainstem; cerebellum; crescent-shaped; idiopathic CD4(+) T
   lymphocytopenia; progressive multifocal leukoencephalopathy
ID PATIENT; FORM
AB Progressive multifocal leukoencephalopathy (PML) is a demyelinating disease that favors the cerebrum and typically occurs in immunosuppressed patients. We herein report the case of a 66-year-old man with PML, idiopathic CD4(+) T lymphocytopenia (ICL), and chronic renal failure. Cranial magnetic resonance imaging (MRI) showed a crescent-shaped lesion in the left cerebellum, brainstem, and middle cerebellar peduncle. Although the patient did not present with HIV infection, collagen diseases, or tumors, JC virus DNA was detected in the cerebrospinal fluid. Clinicians should consider PML and ICL in the differential diagnosis if the patient develops progressive ataxia and a crescent-shaped cerebellar lesion on MRI.
C1 [Aotsuka, Yuya; Uzawa, Akiyuki; Nishimura, Kazutaka; Kojima, Kazuho; Kuwabara, Satoshi] Chiba Univ, Grad Sch Med, Dept Neurol, Chiba, Japan.
   [Yamaguchi, Mika; Makino, Takahiro] Chiba Med Ctr, Dept Neurol, Chiba, Japan.
   [Nakamichi, Kazuo; Saijo, Masayuki] Natl Inst Infect Dis, Dept Virol 1, Chiba, Japan.
C3 Chiba University; National Institute of Infectious Diseases (NIID)
RP Uzawa, A (通讯作者)，Chiba Univ, Grad Sch Med, Dept Neurol, Chiba, Japan.
EM
RI Saijo, Masayuki/L-2697-2019; Nakamichi, Kazuo/ACD-1764-2022
OI Nakamichi, Kazuo/0000-0001-7594-2607; Aotsuka, Yuya/0000-0003-4232-2772;
   Kuwabara, Satoshi/0000-0002-4716-8578; Saijo,
   Masayuki/0000-0001-5458-7298
FU JSPS KAKENHI Grant [26461286]; Research Committee of Prion Disease and
   Slow Virus Infection, Research on Policy Planning and Evaluation for
   Rare and Intractable Diseases from the Ministry of Health, Labour and
   Welfare of Japan; Research Committee of Molecular Pathogenesis and
   Therapies for Prion Disease and Slow Virus Infection; Practical Research
   Project for Rare/Intractable Disease from Japan Agency for Medical
   Research and Development, AMED;  [H24-AIDS-Wakate-002]; Grants-in-Aid
   for Scientific Research [26461286] Funding Source: KAKEN
FX This work was partly supported by the following: JSPS KAKENHI Grant
   Number 26461286, Grants-in-Aid for the Research on HIV/AIDS
   (H24-AIDS-Wakate-002), the Research Committee of Prion Disease and Slow
   Virus Infection, Research on Policy Planning and Evaluation for Rare and
   Intractable Diseases from the Ministry of Health, Labour and Welfare of
   Japan, Research Committee of Molecular Pathogenesis and Therapies for
   Prion Disease and Slow Virus Infection, and The Practical Research
   Project for Rare/Intractable Disease from Japan Agency for Medical
   Research and Development, AMED.
CR Boster A, 2009, ARCH NEUROL-CHICAGO, V66, P593, DOI 10.1001/archneurol.2009.31
   Delgado-Alvarado M, 2013, J NEUROL SCI, V327, P75, DOI 10.1016/j.jns.2013.02.002
   Gheuens S, 2010, J NEUROL NEUROSUR PS, V81, P247, DOI 10.1136/jnnp.2009.187666
   IRIE T, 1992, INTERNAL MED, V31, P218, DOI 10.2169/internalmedicine.31.218
   Iwase T, 1998, J NEUROL NEUROSUR PS, V64, P788, DOI 10.1136/jnnp.64.6.788
   Izaki S, 2015, RINSHO SHINKEIGAKU, V55, P345
   Kobayashi Z, 2013, J NEUROL SCI, V324, P190, DOI 10.1016/j.jns.2012.11.004
   Molloy ES, 2009, ARTHRITIS RHEUM-US, V60, P3761, DOI 10.1002/art.24966
   Nakamichi K, 2011, JPN J INFECT DIS, V64, P211
   Post MJD, 1999, AM J NEURORADIOL, V20, P1896
   Rueger MA, 2006, CLIN INFECT DIS, V42, pE53, DOI 10.1086/500400
   Sahraian MA, 2012, EUR J NEUROL, V19, P1060, DOI 10.1111/j.1468-1331.2011.03597.x
   TAKAHASHI H, 1992, ACTA NEUROPATHOL, V83, P105, DOI 10.1007/BF00308469
NR 13
TC 9
Z9 9
U1 0
U2 1
PU JAPAN SOC INTERNAL MEDICINE
PI TOKYO
PA 34-3 3-CHOME HONGO BUNKYO-KU, TOKYO, 113, JAPAN
SN 0918-2918
EI 1349-7235
J9 INTERNAL MED
JI Intern. Med.
PY 2016
VL 55
IS 12
BP 1645
EP 1647
DI 10.2169/internalmedicine.55.6649
PG 3
WC Medicine, General & Internal
WE Science Citation Index Expanded (SCI-EXPANDED)
SC General & Internal Medicine
GA DO9NX
UT WOS:000378114500025
PM 27301521
OA gold
DA 2023-08-27
ER

PT J
AU Fujimaki, M
   Takanashi, M
   Kobayashi, M
   Wada, K
   Machida, Y
   Kondo, A
   Hattori, N
   Miwa, H
AF Fujimaki, Motoki
   Takanashi, Masashi
   Kobayashi, Manami
   Wada, Kei-ichiro
   Machida, Yutaka
   Kondo, Akihide
   Hattori, Nobutaka
   Miwa, Hideto
TI Cerebrospinal fluid dissemination of anaplastic intraventricular
   meningioma: report of a case presenting with progressive brainstem
   dysfunction and multiple cranial nerve palsies
SO BMC NEUROLOGY
LA English
DT Article
DE Brainstem dysfunction; Multiple cranial nerve palsies; Cerebrospinal
   fluid dissemination; Anaplastic meningioma
AB Background: It is extremely rare to see cerebrospinal fluid dissemination of intraventricular meningioma, particularly with the development of acute, progressive brainstem/cerebellar dysfunction with an absence of mass formation in the corresponding anatomical sites.
   Case presentation: An 81-year-old man was admitted because of double vision, right facial nerve palsy and truncal ataxia. Brain magnetic resonance imaging showed normal findings except for a tumor mass in the left lateral ventricle, which had been noted over 6 months previously. The patient developed hiccups, hyperventilation, and drowsiness, which worsened progressively, and did not respond to corticosteroid or intraventricular immunoglobulin therapy. Cerebrospinal fluid study revealed a mild elevation of protein, and cytology was negative. The patient died and an autopsy was performed. Postmortem investigation disclosed a malignant transformation of benign fibroid meningioma with cerebrospinal fluid dissemination of the malignant cells, diversely involving the surface of brainstem, cerebellum, and spinal cords, secondarily resulting in extensive ischemia in the brain parenchyma by vessel occlusion.
   Conclusion: If a patient with an intraventricular tumor develops acute, progressive neurological symptoms, the possibility that it is be caused by cerebrospinal fluid dissemination of tumor cells, after malignant transformation, should be considered.
C1 [Fujimaki, Motoki; Takanashi, Masashi; Kobayashi, Manami; Wada, Kei-ichiro; Machida, Yutaka; Miwa, Hideto] Juntendo Univ, Nerima Hosp, Dept Neurol, 3-1-10 Takanodai, Nerimaku, Tokyo 1778521, Japan.
   [Fujimaki, Motoki; Kobayashi, Manami; Hattori, Nobutaka] Juntendo Univ, Sch Med, Dept Neurol, Bunkyo Ku, Tokyo 1138431, Japan.
   [Kondo, Akihide] Juntendo Univ, Sch Med, Dept Neurosurg, Bunkyo Ku, 2-1-1 Hongo, Tokyo 1138431, Japan.
   [Fujimaki, Motoki] 2-1-1 Hongo,Bunkyouku, Tokyo 1138431, Japan.
C3 Juntendo University; Juntendo University; Juntendo University
RP Fujimaki, M (通讯作者)，Juntendo Univ, Nerima Hosp, Dept Neurol, 3-1-10 Takanodai, Nerimaku, Tokyo 1778521, Japan.; Fujimaki, M (通讯作者)，Juntendo Univ, Sch Med, Dept Neurol, Bunkyo Ku, Tokyo 1138431, Japan.; Fujimaki, M (通讯作者)，2-1-1 Hongo,Bunkyouku, Tokyo 1138431, Japan.
EM
RI Hattori, Nobutaka/ACR-0069-2022
OI Hattori, Nobutaka/0000-0002-2034-2556
CR Chuan-yuan T, 2014, WORLD J SURG ONCOL, V12, P238
   Clarke JL, 2010, NEUROLOGY, V74, P1449, DOI 10.1212/WNL.0b013e3181dc1a69
   KLEINSCHMIDTDEMASTERS BK, 1985, CLIN NEUROPATHOL, V4, P214
   Liu YL, 2004, MODERN PATHOL, V17, P1129, DOI 10.1038/modpathol.3800162
   Marc CC, 2005, CANCER, V103, P1427
   Maria GP, 2015, NEUROL SCI, V36, P117
   Michael B, 1996, ARCH NEUROL-CHICAGO, V53, P626
   Michael JG, 1998, CANCER, V82, P733
   Nortina S, 2013, J NEUROL NEUROSUR PS, V84, P576
   Odegaard KM, 2013, NEUROSURG REV, V36, P57, DOI 10.1007/s10143-012-0410-5
   Riemenschneider MJ, 2006, LANCET NEUROL, V5, P1045, DOI 10.1016/S1474-4422(06)70625-1
   Toyooka T, 2005, J CLIN NEUROSCI, V12, P834, DOI 10.1016/j.jocn.2004.09.027
   William RW, 1982, CANCER, V49, P759
NR 13
TC 4
Z9 4
U1 0
U2 4
PU BIOMED CENTRAL LTD
PI LONDON
PA 236 GRAYS INN RD, FLOOR 6, LONDON WC1X 8HL, ENGLAND
SN 1471-2377
J9 BMC NEUROL
JI BMC Neurol.
PD MAY 31
PY 2016
VL 16
AR 82
DI 10.1186/s12883-016-0611-6
PG 7
WC Clinical Neurology
WE Science Citation Index Expanded (SCI-EXPANDED)
SC Neurosciences & Neurology
GA DO2DQ
UT WOS:000377590300001
PM 27245327
OA Green Published, gold
DA 2023-08-27
ER

PT J
AU Amato, VG
   Assietti, R
   Morosi, M
   Arienta, C
AF Amato, VG
   Assietti, R
   Morosi, M
   Arienta, C
TI Acute brainstem dissection of syringomyelia associated with cervical
   intramedullary neurinoma
SO NEUROSURGICAL REVIEW
LA English
DT Article
DE acute presentation; intramedullary neurinoma; intramedullary tumors;
   syringobulbia; syringomyelia
ID CEREBROSPINAL-FLUID FLOW; CENTRAL-NERVOUS-SYSTEM; SPINAL-CORD;
   PATHOPHYSIOLOGY; PRESSURE; PRESENTATIONS; EDEMA; SPACE; MR
AB Intramedullary tumors and syringomyelia typically present with slowly progressing deficits. More rarely, they are characterized by acute presentation or worsening, at times mimicking other more common etiologies. The acute onset of syringomyelia is most likely attributable to an acute increase in cerebrospinal fluid and epidural venous pressure that results in impulsive fluid movement and, ultimately, in the rupture of the syrinx and dissection into the spinal cord or brainstem. Reported here is a case of acute presentation of a small cervical intramedullary neurinoma due to the upward dissection of its associated syrinx. Critical questions are: (1) how can a small tumor produce a large syrinx? and (2) in the absence of craniospinal interferences, which mechanism underlies the acute expansion of the cavity, resulting in a rapid onset? The authors examined the pathophysiology of syrinx formation and enlargement in intramedullary tumors and reviewed the literature, emphasizing the relationship between spinal cord movements and intramedullary pressure. On the basis of current pathogenetic concepts, the authors concluded that tumor-related syringomyelia might be caused by an association of mechanisms, both from within (obstruction of perivascular spaces; increase in extracellular fluid viscosity due to the tumor itself; intramedullary pressure gradients among different cord levels and between the cord and the subarachnoid space) and from without (the cerebrospinal fluid entering the tissue). All these factors may be amplified, as in the reported case, by a tumor located dorsally at the cervical level. Abnormal postures of the spine, such as a prolonged and excessive flexed neck position, may ultimately contribute to the acute dissection of the syrinx.
C1 Fatebenefratelli Hosp, Dept Neurosurg, I-20121 Milan, Italy.
   Univ Pavia, Neurosurg Clin, I-27100 Pavia, Italy.
C3 University of Pavia
RP Amato, VG (通讯作者)，Via Saldini 38, I-20133 Milan, Italy.
EM
CR Anwer UE, 1996, EUR NEUROL, V36, P215, DOI 10.1159/000117251
   BARNETT HJM, 1973, SYRINGOMYELIA MAJOR, V1, P261
   BATZDORF U, 1991, SYRINGOMYELIA CURREN
   CSERR HF, 1974, EXP NEUROL, V45, P50, DOI 10.1016/0014-4886(74)90099-5
   Delpirou C, 2001, REV NEUROL-FRANCE, V157, P692
   DICHIRO G, 1964, NATURE, V204, P290, DOI 10.1038/204290a0
   ELLERTSS.AB, 1970, LANCET, V1, P1234
   ELLERTSSON AB, 1969, ACTA NEUROL SCAND, V45, P418
   Fischbein NJ, 1999, AM J NEURORADIOL, V20, P7
   Fuller R, 2000, LANCET, V355, P284, DOI 10.1016/S0140-6736(99)08502-5
   HALL P, 1980, J NEUROSURG, V52, P812, DOI 10.3171/jns.1980.52.6.0812
   Harrison DE, 1999, J MANIP PHYSIOL THER, V22, P399, DOI 10.1016/S0161-4754(99)70086-2
   Heiss JD, 1999, J NEUROSURG, V91, P553, DOI 10.3171/jns.1999.91.4.0553
   IKATA T, 1988, SPINE, V13, P737, DOI 10.1097/00007632-198807000-00005
   Jarzem PF, 1992, SPINE, V17, P227, DOI 10.1097/00007632-199208001-00003
   Josephson A, 2001, NEUROSURGERY, V48, P636, DOI 10.1097/00006123-200103000-00039
   Kitahara Yukio, 1995, Neurologia Medico-Chirurgica, V35, P285, DOI 10.2176/nmc.35.285
   Kiwit JCW, 1988, MODERN METHODS NEURO, P211
   Klekamp J, 2002, ACTA NEUROCHIR, V144, P649, DOI 10.1007/s00701-002-0944-3
   Klekamp J, 2001, NEUROSURGERY, V48, P174, DOI 10.1097/00006123-200101000-00031
   Levy EI, 2000, J NEUROSURG, V92, P93, DOI 10.3171/spi.2000.92.1.0093
   Levy L M, 1999, Magn Reson Imaging Clin N Am, V7, P573
   LEVY LM, 1990, NEURORADIOLOGY, V32, P399, DOI 10.1007/BF00588473
   Meves SH, 2000, EUR NEUROL, V43, P47, DOI 10.1159/000008128
   Milhorat TH, 1997, NEUROSURGERY, V41, P1102, DOI 10.1097/00006123-199711000-00014
   Milhorat TH, 2003, NEUROSURGERY, V53, P1220, DOI 10.1227/01.NEU.0000088809.14965.00
   MILHORAT TH, 1995, J NEUROSURG, V82, P802, DOI 10.3171/jns.1995.82.5.0802
   OLDFIELD EH, 1994, J NEUROSURG, V80, P3, DOI 10.3171/jns.1994.80.1.0003
   Oldfield EH, 2001, J NEUROSURG, V95, P153, DOI 10.3171/spi.2001.95.1.0153
   QUENCER RM, 1990, NEURORADIOLOGY, V32, P371, DOI 10.1007/BF00588471
   RENNELS ML, 1985, BRAIN RES, V326, P47, DOI 10.1016/0006-8993(85)91383-6
   SAMII M, 1994, NEUROSURGERY, V35, P865, DOI 10.1227/00006123-199411000-00010
   SCHLESINGER EB, 1981, NEUROSURGERY, V9, P356
   Shiojiri T, 1993, No To Shinkei, V45, P1075
   Stoodley M.A., 2000, NEUROSURG FOCUS, V8, P1
   TACHIBANA S, 1992, J NEUROSURG, V76, P67, DOI 10.3171/jns.1992.76.1.0067
   TACHIBANA S, 1994, SPINE, V19, P2174, DOI 10.1097/00007632-199410000-00008
   WILLIAMS B, 1986, Neurological Research, V8, P130
   Williams B, 1993, Acta Neurochir (Wien), V123, P190
   Yuan Q, 1998, SPINE, V23, P1677, DOI 10.1097/00007632-199808010-00012
   ZAGER EL, 1990, J NEUROSURG, V72, P133, DOI 10.3171/jns.1990.72.1.0133
NR 41
TC 3
Z9 5
U1 0
U2 4
PU SPRINGER
PI NEW YORK
PA 233 SPRING STREET, NEW YORK, NY 10013 USA
SN 0344-5607
J9 NEUROSURG REV
JI Neurosurg. Rev.
PD APR
PY 2005
VL 28
IS 2
BP 163
EP 167
DI 10.1007/s10143-004-0362-5
PG 5
WC Clinical Neurology; Surgery
WE Science Citation Index Expanded (SCI-EXPANDED)
SC Neurosciences & Neurology; Surgery
GA 910NZ
UT WOS:000227939900016
PM 15789252
DA 2023-08-27
ER

PT J
AU Uh, J
   Merchant, TE
   Li, YM
   Feng, TS
   Gajjar, A
   Ogg, RJ
   Hua, CH
AF Uh, Jinsoo
   Merchant, Thomas E.
   Li, Yimei
   Feng, Tianshu
   Gajjar, Amar
   Ogg, Robert J.
   Hua, Chiaho
TI Differences in Brainstem Fiber Tract Response to Radiation: A
   Longitudinal Diffusion Tensor Imaging Study
SO INTERNATIONAL JOURNAL OF RADIATION ONCOLOGY BIOLOGY PHYSICS
LA English
DT Article
ID APPEARING WHITE-MATTER; RADIOSENSITIVITY; MEDULLOBLASTOMA; INJURY;
   TUMORS
AB Purpose: To determine whether radiation-induced changes in white matter tracts are uniform across the brainstem.
   Methods and Materials: We analyzed serial diffusion tensor imaging data, acquired before radiation therapy and over 48 to 72 months of follow-up, from 42 pediatric patients (age 6-20 years) with medulloblastoma. FSL software (FMRIB, Oxford, UK) was used to calculate fractional anisotropy (FA) and axial, radial, and mean diffusivities. For a consistent identification of volumes of interest (VOIs), the parametric maps of each patient were transformed to a standard brain space (MNI152), on which we identified VOIs including corticospinal tract (CST), medial lemniscus (ML), transverse pontine fiber (TPF), and middle cerebellar peduncle (MCP) at the level of pons. Temporal changes of DTI parameters in VOIs were compared using a linear mixed effect model.
   Results: Radiation-induced white matter injury was marked by a decline in FA after treatment. The decline was often accompanied by decreased axial diffusivity, increased radial diffusivity, or both. This implied axonal damage and demyelination. We observed that the magnitude of the changes was not always uniform across substructures of the brainstem. Specifically, the changes in DTI parameters for TPF were more pronounced than in other regions (P<.001 for FA) despite similarities in the distribution of dose. We did not find a significant difference among CST, ML, and MCP in these patients (P>.093 for all parameters).
   Conclusions: Changes in the structural integrity of white matter tracts, assessed by DTI, were not uniform across the brainstem after radiation therapy. These results support a role for tract-based assessment in radiation treatment planning and determination of brainstem tolerance. (C) 2013 Elsevier Inc.
C1 [Uh, Jinsoo; Merchant, Thomas E.; Ogg, Robert J.; Hua, Chiaho] St Jude Childrens Res Hosp, Dept Radiol Sci, Memphis, TN 38105 USA.
   [Li, Yimei; Feng, Tianshu] St Jude Childrens Res Hosp, Dept Biostat, Memphis, TN 38105 USA.
   [Gajjar, Amar] St Jude Childrens Res Hosp, Dept Oncol, Memphis, TN 38105 USA.
C3 St Jude Children's Research Hospital; St Jude Children's Research
   Hospital; St Jude Children's Research Hospital
RP Uh, J (通讯作者)，St Jude Childrens Res Hosp, Dept Radiol Sci, 262 Danny Thomas Pl, Memphis, TN 38105 USA.
EM
RI Gajjar, Amar/N-8059-2018; Hua, Chia-ho/D-4220-2018; Merchant, Thomas
   E/N-3344-2018
OI Merchant, Thomas E/0000-0002-0412-6255; Uh, Jinsoo/0000-0001-6837-8681
FU American Lebanese Syrian Associated Charities; NIH [HD049888]
FX Supported in part by the funding from the American Lebanese Syrian
   Associated Charities and NIH R01 grant HD049888.
CR Alexander DC, 2001, IEEE T MED IMAGING, V20, P1131, DOI 10.1109/42.963816
   Bijl HP, 2005, INT J RADIAT ONCOL, V61, P543, DOI 10.1016/j.ijrobp.2004.10.018
   CALVO W, 1988, BRIT J RADIOL, V61, P1043, DOI 10.1259/0007-1285-61-731-1043
   Chapman CH, 2012, INT J RADIAT ONCOL, V82, P2033, DOI 10.1016/j.ijrobp.2011.01.068
   Debus J, 1997, INT J RADIAT ONCOL, V39, P967, DOI 10.1016/S0360-3016(97)00364-7
   Hua CH, 2012, INT J RADIAT ONCOL, V82, P2047, DOI 10.1016/j.ijrobp.2011.03.057
   Karampinos DC, 2009, MAGN RESON MED, V62, P1007, DOI 10.1002/mrm.22105
   Khong PL, 2003, AM J NEURORADIOL, V24, P734
   Kumar R, 2012, J NEUROSCI RES, V90, P346, DOI 10.1002/jnr.22757
   Mayo C, 2010, INT J RADIAT ONCOL, V76, pS36, DOI 10.1016/j.ijrobp.2009.08.078
   Nagesh V, 2008, INT J RADIAT ONCOL, V70, P1002, DOI 10.1016/j.ijrobp.2007.08.020
   Qiu D, 2007, INT J RADIAT ONCOL, V69, P846, DOI 10.1016/j.ijrobp.2007.04.041
   Rueckriegel SM, 2010, INT J RADIAT ONCOL, V76, P859, DOI 10.1016/j.ijrobp.2009.02.054
   SCHULTHEISS TE, 1995, INT J RADIAT ONCOL, V31, P1093, DOI 10.1016/0360-3016(94)00655-5
   Wang SL, 2009, CANCER RES, V69, P1190, DOI 10.1158/0008-5472.CAN-08-2661
NR 15
TC 31
Z9 34
U1 0
U2 5
PU ELSEVIER SCIENCE INC
PI NEW YORK
PA 360 PARK AVE SOUTH, NEW YORK, NY 10010-1710 USA
SN 0360-3016
J9 INT J RADIAT ONCOL
JI Int. J. Radiat. Oncol. Biol. Phys.
PD JUN 1
PY 2013
VL 86
IS 2
BP 292
EP 297
DI 10.1016/j.ijrobp.2013.01.028
PG 6
WC Oncology; Radiology, Nuclear Medicine & Medical Imaging
WE Science Citation Index Expanded (SCI-EXPANDED)
SC Oncology; Radiology, Nuclear Medicine & Medical Imaging
GA 151ZZ
UT WOS:000319500200020
PM 23474114
OA Green Accepted
DA 2023-08-27
ER

PT J
AU Sharp, JR
   Bouffet, E
   Stempak, D
   Gammon, J
   Stephens, D
   Johnston, DL
   Eisenstat, D
   Hukin, J
   Samson, Y
   Bartels, U
   Tabori, U
   Huang, A
   Baruchel, S
AF Sharp, Julia R.
   Bouffet, Eric
   Stempak, Diana
   Gammon, Janet
   Stephens, Derek
   Johnston, Donna L.
   Eisenstat, David
   Hukin, Juliette
   Samson, Yvan
   Bartels, Ute
   Tabori, Uri
   Huang, Annie
   Baruchel, Sylvain
TI A multi-centre Canadian pilot study of metronomic temozolomide combined
   with radiotherapy for newly diagnosed paediatric brainstem glioma
SO EUROPEAN JOURNAL OF CANCER
LA English
DT Article
DE Temozolomide; Radiotherapy; Multimodal treatment; Anti-angiogenic;
   Paediatrics; Brainstem tumour; Glioma; Drug toxicity; Angiogenic
   proteins
ID ANTI-ANGIOGENIC THERAPY; PHASE-I TRIAL; ADJUVANT TEMOZOLOMIDE;
   HYPERFRACTIONATED RADIOTHERAPY; MALIGNANT GLIOMAS; CLINICAL-TRIALS;
   GROWTH-FACTOR; CHILDREN; TUMORS; GLIOBLASTOMA
AB Purpose: Survival rates for paediatric diffuse intrinsic brainstem glioma (DIBSG) are dismal. Metronomic dosing of temozolomide (TMZ) combined with standard radiotherapy may improve survival by increasing the therapeutic index and anti-angiogenic effect of TMZ. This study aimed to evaluate the safety and efficacy of this regimen in paediatric DIBSG patients.
   Methods: Children aged 18 years or younger with newly diagnosed DIBSG were treated with standard radiotherapy and concomitant metronomic TMZ at 85 mg/m(2)/day for 6 weeks, followed by metronomic TMZ monotherapy at the same dose. Treatment was continued until tumour progression or unacceptable toxicity occurred. Primary endpoints included overall survival and toxicities. For patients who consented, plasma and urine samples were collected at diagnosis, post-induction and prior to each course of maintenance therapy for the quantification of angiogenesis markers.
   Results: Fifteen eligible patients were enrolled, with a median age of 6.4 years. The most common toxicities were myelosuppression, most notably prolonged lymphopaenia and thrombocytopaenia. The only dose-limiting toxicity was thrombocytopaenia. Intratumoural haemorrhage was confirmed in one patient. Median time to progression was 5.13 months (95% CI = 6.4, 10.8) and median overall survival (OS) was 9.8 months (95% CI = 6.4, 10.8). Six-months OS was 80% +/- 10.3%, with a 1-year OS of 20% +/- 10.3%. Serum levels of both VEGF and endoglin tended to decrease during the first two cycles of therapy.
   Conclusion: Chemoradiotherapy with metronomic dosing of TMZ showed similar toxicity to previous TMZ regimens, and does not appear to improve survival in paediatric DIBSG. (C) 2010 Elsevier Ltd. All rights reserved.
C1 [Sharp, Julia R.; Bouffet, Eric; Stempak, Diana; Gammon, Janet; Bartels, Ute; Tabori, Uri; Huang, Annie; Baruchel, Sylvain] Hosp Sick Children, Dept Paediat, Div Haematol Oncol, New Agent & Innovat Therapy Program, Toronto, ON M5G 1X8, Canada.
   [Stephens, Derek] SickKids Res Inst, Toronto, ON M5G 1E2, Canada.
   [Johnston, Donna L.] Childrens Hosp Eastern Ontario, Div Haematol Oncol, Ottawa, ON K1H 8L1, Canada.
   [Eisenstat, David] Canc Care Manitoba & Childrens Hosp, Sect Paediat Haematol Oncol BMT, Winnipeg, MB R3E 0V9, Canada.
   [Hukin, Juliette] British Columbia Childrens Hosp, Dept Haematol Oncol, Vancouver, BC V6H 3V4, Canada.
   [Samson, Yvan] Ctr Hosp Univ Quebec, Dept Hematol Oncol, Quebec City, PQ G1R 2J6, Canada.
C3 University of Toronto; Hospital for Sick Children (SickKids); University
   of Toronto; Hospital for Sick Children (SickKids); University of Ottawa;
   Children's Hospital of Eastern Ontario; BC Childrens Hospital;
   University of British Columbia; Laval University
RP Baruchel, S (通讯作者)，Hosp Sick Children, Dept Paediat, Div Haematol Oncol, New Agent & Innovat Therapy Program, 555 Univ Ave, Toronto, ON M5G 1X8, Canada.
EM
RI Bouffet, Eric/AAB-8999-2020; Eisenstat, David/AAA-4653-2020; Eisenstat,
   David/HZH-9682-2023
OI Eisenstat, David/0000-0002-5976-0798; Hukin,
   Juliette/0000-0002-8727-8456; Tabori, Uri/0000-0002-5019-2683
FU Brain Tumour Research Assistance and Information Network;
   Schering-Plough
FX The study was supported by Schering-Plough and b.r.a.i.n.child, and was
   sponsored by the principal investigator. Decision to submit to EJC was
   made by the principal investigator.The study was funded by
   b.r.a.i.n.child (Brain Tumour Research Assistance and Information
   Network). Temozolomide was provided by Schering-Plough Canada.
CR Baruchel S, 2006, EUR J CANCER, V42, P2335, DOI 10.1016/j.ejca.2006.03.023
   Bernier-Chastagner V, 2005, CANCER-AM CANCER SOC, V104, P2792, DOI 10.1002/cncr.21534
   Brock CS, 1998, CANCER RES, V58, P4363
   Broniscer A, 2006, CANCER, V106, P1364, DOI 10.1002/cncr.21749
   Chiang KL, CHILDS NERV SYST
   Clarke JL, 2009, J CLIN ONCOL, V27, P3861, DOI 10.1200/JCO.2008.20.7944
   Doz F, 2002, EUR J CANCER, V38, P815, DOI 10.1016/S0959-8049(02)00029-1
   Emmanuel J, 2008, NEUROSURGERY, V62, P31
   Estlin EJ, 1998, BRIT J CANCER, V78, P652, DOI 10.1038/bjc.1998.555
   Fox SB, 1997, INVEST NEW DRUG, V15, P15, DOI 10.1023/A:1005714527315
   Freeman CR, 2000, INT J RADIAT ONCOL, V47, P561, DOI 10.1016/S0360-3016(00)00471-5
   Hargrove D, 2006, LANCET ONCOL, V7, P241, DOI 10.1016/S1470-2045(06)70615-5
   Jalali R, 2009, INT J RAD ONCOL 0731
   Khan RB, 2002, NEURO-ONCOLOGY, V4, P39, DOI 10.1093/neuonc/4.1.39
   Kieran MW, 2005, J NEURO-ONCOL, V75, P327, DOI 10.1007/s11060-005-6764-6
   Lashford LS, 2002, J CLIN ONCOL, V20, P4684, DOI 10.1200/JCO.2002.08.141
   Lewis J, 1997, INT J RADIAT ONCOL, V38, P925, DOI 10.1016/S0360-3016(97)00134-X
   Mandell LR, 1999, INT J RADIAT ONCOL, V43, P959, DOI 10.1016/S0360-3016(98)00501-X
   Marcus KJ, 2003, INT J RADIAT ONCOL, V55, P1182, DOI 10.1016/S0360-3016(02)04391-2
   Merchant TE, 2009, INT J RADIAT ONCOL, V74, P159, DOI 10.1016/j.ijrobp.2008.07.058
   Ostermann S, 2004, CLIN CANCER RES, V10, P3728, DOI 10.1158/1078-0432.CCR-03-0807
   Poon RTP, 2001, J CLIN ONCOL, V19, P1207, DOI 10.1200/JCO.2001.19.4.1207
   Sebille V, 2003, FUND CLIN PHARMACOL, V17, P505, DOI 10.1046/j.1472-8206.2003.00192.x
   Sirachainan N, 2008, NEURO-ONCOLOGY, V10, P577, DOI 10.1215/15228517-2008-025
   Sterba J, 2002, NEOPLASMA, V49, P117
   Stupp R, 2005, NEW ENGL J MED, V352, P987, DOI 10.1056/NEJMoa043330
   Stupp R, 2009, LANCET ONCOL, V10, P459, DOI 10.1016/S1470-2045(09)70025-7
   Su YB, 2004, J CLIN ONCOL, V22, P610, DOI 10.1200/JCO.2004.07.060
   Takano S, 1996, CANCER RES, V56, P2185
   van Rijn J, 2000, INT J RADIAT ONCOL, V47, P779, DOI 10.1016/S0360-3016(99)00539-8
   Wedge SR, 1997, ANTI-CANCER DRUG, V8, P92, DOI 10.1097/00001813-199701000-00013
NR 31
TC 33
Z9 33
U1 0
U2 2
PU ELSEVIER SCI LTD
PI OXFORD
PA THE BOULEVARD, LANGFORD LANE, KIDLINGTON, OXFORD OX5 1GB, OXON, ENGLAND
SN 0959-8049
EI 1879-0852
J9 EUR J CANCER
JI Eur. J. Cancer
PD DEC
PY 2010
VL 46
IS 18
BP 3271
EP 3279
DI 10.1016/j.ejca.2010.06.115
PG 9
WC Oncology
WE Science Citation Index Expanded (SCI-EXPANDED)
SC Oncology
GA 701IY
UT WOS:000285812800026
PM 20656474
DA 2023-08-27
ER

PT J
AU Peciu-Florianu, I
   Legrand, V
   Monfilliette-Djelad, A
   Maurage, CA
   Vannod-Michel, Q
   Blond, S
   Touzet, G
   Reyns, N
AF Peciu-Florianu, Iulia
   Legrand, Victor
   Monfilliette-Djelad, Apolline
   Maurage, Claude-Alain
   Vannod-Michel, Quentin
   Blond, Serge
   Touzet, Gustavo
   Reyns, Nicolas
TI Frameless robot-assisted stereotactic biopsies for lesions of the
   brainstem-a series of 103 consecutive biopsies
SO JOURNAL OF NEURO-ONCOLOGY
LA English
DT Article
DE Brainstem; Frameless; Robot; Stereotactic; Biopsy; Case series
ID SAFE ENTRY ZONES; CENTRAL-NERVOUS-SYSTEM; IN-VIVO MEASUREMENT;
   MICROSURGICAL ANATOMY; DIAGNOSTIC-VALUE; POSTERIOR-FOSSA; TUMORS;
   ACCURACY; METAANALYSIS; CONSORTIUM
AB Purpose Targeted treatment for brainstem lesions requires above all a precise histopathological and molecular diagnosis. In the current technological era, robot-assisted stereotactic biopsies represent an accurate and safe procedure for tissue diagnosis. We present our center's experience in frameless robot-assisted biopsies for brainstem lesions. Methods We performed a retrospective analysis of all patients benefitting from a frameless robot-guided stereotactic biopsy at our University Hospital, from 2001 to 2017. Patients consented to the use of data and/or images. The NeuroMate (R) robot (Renishaw (TM), UK) was used. We report on lesion location, trajectory strategy, histopathological diagnosis and procedure safety. Results Our series encompasses 96 patients (103 biopsies) treated during a 17 years period. Mean age at biopsy: 34.0 years (range 1-78). Most common location: pons (62.1%). Transcerebellar approach: 61 procedures (59.2%). Most common diagnoses: diffuse glioma (67.0%), metastases (7.8%) and lymphoma (6.8%). Non conclusive diagnosis: 10 cases (9.7%). After second biopsy this decreased to 4 cases (4.1%). Overall biopsy diagnostic yield: 95.8%. Permanent disability was recorded in 3 patients (2.9%, all adults), while transient complications in 17 patients (17.7%). Four cases of intra-tumoral hematoma were recorded (one case with rapid decline and fatal issue). Adjuvant targeted treatment was performed in 72.9% of patients. Mean follow-up (in the Neurosurgery Department): 2.2 years. Conclusion Frameless robot-assisted stereotactic biopsies can provide the initial platform towards a safe and accurate management for brainstem lesions, offering a high diagnostic yield with low permanent morbidity.
C1 [Peciu-Florianu, Iulia; Legrand, Victor; Blond, Serge; Touzet, Gustavo; Reyns, Nicolas] CHU Lille, Neurosurg Dept, Roger Salengro Hosp, 1 Rue Emile Laine, F-59000 Lille, France.
   [Reyns, Nicolas] Univ Lille, U1189 ONCO THAI Assisted Laser Therapy & Immunoth, INSERM, CHU Lille, F-59000 Lille, France.
   [Monfilliette-Djelad, Apolline] CHU Lille, Neurol Dept, Neurooncol Unit, F-59000 Lille, France.
   [Maurage, Claude-Alain] CHU Lille, Neuropathol Dept, F-59000 Lille, France.
   [Vannod-Michel, Quentin] CHU Lille, Neuroradiol Dept, F-59000 Lille, France.
C3 Universite de Lille - ISITE; CHU Lille; Institut National de la Sante et
   de la Recherche Medicale (Inserm); Universite de Lille - ISITE; CHU
   Lille; Universite de Lille; Universite de Lille - ISITE; CHU Lille;
   Universite de Lille - ISITE; CHU Lille; Universite de Lille - ISITE; CHU
   Lille
RP Peciu-Florianu, I (通讯作者)，CHU Lille, Neurosurg Dept, Roger Salengro Hosp, 1 Rue Emile Laine, F-59000 Lille, France.
EM
CR Agha RA, 2020, INT J SURG, V84, P231, DOI 10.1016/j.ijsu.2020.11.005
   ALBRIGHT AL, 1993, NEUROSURGERY, V33, P1026
   Amundson EW, 2005, J NEUROSURG, V102, P565, DOI 10.3171/jns.2005.102.3.0565
   Aziz-Bose R, 2019, CURR OPIN ONCOL, V31, P522, DOI 10.1097/CCO.0000000000000577
   BAHRAMI E, 2020, WORLD NEUROSURG
   BUCHANAN D, 1957, AM J SURG, V93, P935, DOI 10.1016/0002-9610(57)90670-0
   Bush NAO, 2017, NEUROSURG REV, V40, P1, DOI 10.1007/s10143-016-0709-8
   Capitanio JF, 2019, J NEUROSURG SCI, V63, P194, DOI 10.23736/S0390-5616.16.03637-7
   Carai A, 2017, WORLD NEUROSURG, V101, P584, DOI 10.1016/j.wneu.2017.02.088
   Cavalcanti DD, 2016, J NEUROSURG, V124, P1359, DOI 10.3171/2015.4.JNS141945
   Cavalcanti DD, 2019, WORLD NEUROSURG, V122, pE1285, DOI 10.1016/j.wneu.2018.11.033
   Chen SY, 2011, J CHIN MED ASSOC, V74, P110, DOI 10.1016/j.jcma.2011.01.024
   Coca HA, 2016, NEUROCHIRURGIE, V62, P327, DOI 10.1016/j.neuchi.2016.07.005
   Dawes W, 2019, J ROBOT SURG, V13, P575, DOI 10.1007/s11701-018-0899-x
   Dorward NL, 1999, J NEUROSURG, V90, P160, DOI 10.3171/jns.1999.90.1.0160
   Gass A, 2000, J NEUROL SCI, V172, pS43, DOI 10.1016/S0022-510X(99)00278-6
   Goncalves-Ferreira AJ, 2003, SURG NEUROL, V60, P311, DOI 10.1016/S0090-3019(03)00379-3
   Gonzalez-Martinez J, 2016, NEUROSURGERY, V78, P169, DOI 10.1227/NEU.0000000000001034
   Gupta M, 2021, J NEUROSURG-PEDIATR, V27, P317, DOI 10.3171/2020.7.PEDS20373
   Gupta N, 2018, NEURO-ONCOLOGY, V20, P1547, DOI 10.1093/neuonc/noy070
   Haegelen C, 2010, NEUROCHIRURGIE, V56, P363, DOI 10.1016/j.neuchi.2010.05.006
   Hamisch C, 2017, J NEUROSURG-PEDIATR, V20, P261, DOI 10.3171/2017.2.PEDS1665
   Haralur Y, 2020, NEUROL CLIN, V38, P149, DOI 10.1016/j.ncl.2019.09.002
   HOOD TW, 1986, J NEUROSURG, V65, P172, DOI 10.3171/jns.1986.65.2.0172
   Hughes-Hallett A, 2014, ANN SURG, V260, P205, DOI 10.1097/SLA.0000000000000662
   Johnson KJ, 2014, CANCER EPIDEM BIOMAR, V23, P2716, DOI 10.1158/1055-9965.EPI-14-0207
   Kickingereder P, 2013, NEUROSURGERY, V72, P873, DOI 10.1227/NEU.0b013e31828bf445
   KWOH YS, 1988, IEEE T BIO-MED ENG, V35, P153, DOI 10.1109/10.1354
   KYOSHIMA K, 1993, J NEUROSURG, V78, P987, DOI 10.3171/jns.1993.78.6.0987
   Law LY, 2019, NEUROLOGY, V93, P390, DOI 10.1212/WNL.0000000000008015
   Lefranc M, 2014, STEREOT FUNCT NEUROS, V92, P242, DOI 10.1159/000362936
   Li QH, 2002, CARS 2002: COMPUTER ASSISTED RADIOLOGY AND SURGERY, PROCEEDINGS, P167
   Louis DN, 2020, BRAIN PATHOL, V30, P844, DOI 10.1111/bpa.12832
   Louis DN, 2018, ACTA NEUROPATHOL, V135, P639, DOI 10.1007/s00401-018-1826-y
   Louis DN, 2017, BRAIN PATHOL, V27, P851, DOI 10.1111/bpa.12457
   Louis DN, 2016, ACTA NEUROPATHOL, V131, P803, DOI 10.1007/s00401-016-1545-1
   Machetanz K, 2022, NEUROSURG FOCUS, V52, DOI 10.3171/2021.10.FOCUS21359
   Marcus HJ, 2018, CHILD NERV SYST, V34, P1299, DOI 10.1007/s00381-018-3821-y
   Ostrom QT, 2014, NEURO-ONCOLOGY, V16, P1, DOI [10.1093/neuonc/nov189, 10.1093/neuonc/nou327, 10.1093/neuonc/nou223]
   Pillai A, 2019, OPER NEUROSURG, V17, P164, DOI 10.1093/ons/opy229
   Prakkamakul S, 2017, J NEUROIMAGING, V27, P135, DOI 10.1111/jon.12361
   Quick-Weller J, 2016, J NEURO-ONCOL, V129, P243, DOI 10.1007/s11060-016-2166-1
   Quick-Weller J, 2016, WORLD NEUROSURG, V86, P466, DOI 10.1016/j.wneu.2015.11.036
   Recalde RJ, 2008, NEUROSURGERY, V63, P9, DOI [10.1227/01.NEU.0000297062.52433.3F, 10.1227/01.neu.0000317368.69523.40]
   SPIEGELMANN R, 1991, J NEUROSURG, V75, P486, DOI 10.3171/jns.1991.75.3.0486
   Ughratdar I, 2015, J PARKINSON DIS, V5, P483, DOI 10.3233/JPD-150579
   Varma TRK, 2006, INT J MED ROBOT COMP, V2, P107, DOI 10.1002/rcs.88
   von Langsdorff D, 2015, J NEUROSURG, V122, P191, DOI 10.3171/2014.9.JNS14256
   Widmann G, 2012, INT J MED ROBOT COMP, V8, P1, DOI 10.1002/rcs.441
   Williams JR, 2020, NEUROSURG FOCUS, V48, DOI 10.3171/2019.9.FOCUS19745
NR 50
TC 6
Z9 6
U1 0
U2 2
PU SPRINGER
PI NEW YORK
PA ONE NEW YORK PLAZA, SUITE 4600, NEW YORK, NY, UNITED STATES
SN 0167-594X
EI 1573-7373
J9 J NEURO-ONCOL
JI J. Neuro-Oncol.
PD MAR
PY 2022
VL 157
IS 1
BP 109
EP 119
DI 10.1007/s11060-022-03952-6
EA JAN 2022
PG 11
WC Oncology; Clinical Neurology
WE Science Citation Index Expanded (SCI-EXPANDED)
SC Oncology; Neurosciences & Neurology
GA ZW9CI
UT WOS:000748671200001
PM 35083580
OA Green Submitted
DA 2023-08-27
ER

PT J
AU Frappaz, D
   Schell, M
   Thiesse, P
   Marec-Berard, P
   Mottolese, C
   Perol, D
   Bergeron, C
   Philip, T
   Ricci, AC
   Galand-Desme, S
   Szathmari, A
   Carrie, C
AF Frappaz, Didier
   Schell, Matthias
   Thiesse, Philippe
   Marec-Berard, Perrine
   Mottolese, Carmine
   Perol, David
   Bergeron, Christophe
   Philip, Thierry
   Ricci, Anne Claire
   Galand-Desme, Sophie
   Szathmari, Alexandru
   Carrie, Christian
TI Preradiation chemotherapy may improve survival in pediatric diffuse
   intrinsic brainstem gliomas: Final results of BSG 98 prospective trial
SO NEURO-ONCOLOGY
LA English
DT Article
DE brainstem; chemotherapy; glioma; radiotherapy
ID HIGH-DOSE CHEMOTHERAPY; HYPERFRACTIONATED RADIOTHERAPY;
   RADIATION-THERAPY; PHASE-II; POSTOPERATIVE CHEMOTHERAPY; ONCOLOGY-GROUP;
   CHILDREN; TUMORS; IRRADIATION; CARBOPLATIN
AB Radiation therapy remains the only treatment that provides clinical benefit to children with diffuse brainstem tumors. Their median survival, however, rarely exceeds 9 months. The authors report a prospective trial of frontline chemotherapy aimed at delaying radiation until time of clinical progression. The aim was to investigate the possibility that radiotherapy would maintain its activity in children whose disease progressed after chemotherapy. Twenty-three patients took part in this protocol, the BSG 98 protocol, which consisted of frontline chemotherapy alternating hematotoxic and nonhematotoxic schedules. Each cycle included three courses delivered monthly; the first course was 1,3-bis(2-chloroethyl)-1-nitrosourea-cisplatin, and the second and third were high-dose methotrexate. Three patients underwent one cycle; 5 patients each, two and three cycles; and 10 patients, four cycles. Twenty of the 23 patients eventually received local radiation therapy. A historical cohort of 14 patients who received at least local radiation therapy served as controls. Four patients experienced severe iatrogenic infections, and 11 patients required platelet transfusions. Median survival increased significantly in patients participating in the protocol compared to that in the historical controls (17 months, 95% confidence interval [CI], 10-23 months, vs. 9 months, 95% CI, 8-10 months; p = 0.022), though hospitalization was prolonged (57 vs. 25 days, p = 0.001). Although frontline chemotherapy alternating hematotoxic and nonhematotoxic schedules significantly increases overall median survival, its cost from infection and hospitalization deserves honest discussion with the children and their parents.
C1 [Frappaz, Didier; Schell, Matthias; Thiesse, Philippe; Marec-Berard, Perrine; Perol, David; Bergeron, Christophe; Philip, Thierry; Galand-Desme, Sophie; Carrie, Christian] Ctr Leon Berard, Dept Pediat, Dept Radiotherapy, F-69373 Lyon 08, France.
   [Frappaz, Didier; Schell, Matthias; Thiesse, Philippe; Marec-Berard, Perrine; Perol, David; Bergeron, Christophe; Philip, Thierry; Galand-Desme, Sophie; Carrie, Christian] Ctr Leon Berard, Biostat Unit, F-69373 Lyon 08, France.
   [Mottolese, Carmine; Ricci, Anne Claire; Szathmari, Alexandru] Hop Pierre Wertheimer, Pediat Neurosurg Unit, Lyon, France.
C3 UNICANCER; Centre Leon Berard; UNICANCER; Centre Leon Berard; CHU Lyon
RP Frappaz, D (通讯作者)，Ctr Leon Berard, Dept Pediat, Dept Radiotherapy, 28 Rue Laennec, F-69373 Lyon 08, France.
EM
RI d, p/GWC-4283-2022; carrie, christian/K-5488-2014; Marec-Berard,
   Perrine/HZL-6119-2023
OI carrie, christian/0000-0002-0766-2547; 
CR Afra D, 2002, LANCET, V359, P1011
   Allen J, 1999, CANCER, V86, P1064, DOI 10.1002/(SICI)1097-0142(19990915)86:6<1064::AID-CNCR24>3.0.CO;2-1
   ALLEN JC, 1988, J CLIN ONCOL, V6, P649, DOI 10.1200/JCO.1988.6.4.649
   BALTUCH GH, 1993, NEUROSURGERY, V33, P495
   BELLISSANT E, 1990, STAT MED, V9, P907, DOI 10.1002/sim.4780090807
   Bouffet E, 1997, CANCER CHEMOTH PHARM, V39, P376, DOI 10.1007/s002800050586
   Broniscer A, 2005, CANCER-AM CANCER SOC, V103, P133, DOI 10.1002/cncr.20741
   Broniscer A, 2000, J CLIN ONCOL, V18, P1246, DOI 10.1200/JCO.2000.18.6.1246
   DJERASSI I, 1985, CANCER, V55, P2741, DOI 10.1002/1097-0142(19850615)55:12<2741::AID-CNCR2820551202>3.0.CO;2-2
   Doz F, 2002, EUR J CANCER, V38, P815, DOI 10.1016/S0959-8049(02)00029-1
   DUFFNER PK, 1993, NEW ENGL J MED, V328, P1725, DOI 10.1056/NEJM199306173282401
   Dunkel IJ, 1998, J NEURO-ONCOL, V37, P67, DOI 10.1023/A:1005874508975
   EPSTEIN F, 1986, J NEUROSURG, V64, P11, DOI 10.3171/jns.1986.64.1.0011
   ETTINGER LJ, 1985, J NEURO-ONCOL, V3, P263, DOI 10.1007/BF00165188
   FINE HA, 1993, CANCER, V71, P2585, DOI 10.1002/1097-0142(19930415)71:8<2585::AID-CNCR2820710825>3.0.CO;2-S
   Frappaz D, 2003, BRIT J CANCER, V89, pS73, DOI 10.1038/sj.bjc.6601087
   Frappaz D, 1999, CHILD NERV SYST, V15, P222, DOI 10.1007/s003810050377
   Freeman CR, 1996, CANCER-AM CANCER SOC, V77, P555
   Freeman CR, 2000, INT J RADIAT ONCOL, V47, P561, DOI 10.1016/S0360-3016(00)00471-5
   FREEMAN CR, 1993, INT J RADIAT ONCOL, V27, P197, DOI 10.1016/0360-3016(93)90228-N
   FUJIWARA T, 1994, NEURORADIOLOGY, V36, P74, DOI 10.1007/BF00599203
   GENTET JC, 1999, MED PEDIATR ONCOL, V33, P205
   Grill J, 2005, LANCET ONCOL, V6, P573, DOI 10.1016/S1470-2045(05)70252-7
   Grossman SA, 1997, J CLIN ONCOL, V15, P2596, DOI 10.1200/JCO.1997.15.7.2596
   Hargrove D, 2006, LANCET ONCOL, V7, P241, DOI 10.1016/S1470-2045(06)70615-5
   Jakacki RI, 1998, MED PEDIATR ONCOL, V31, P483, DOI 10.1002/(SICI)1096-911X(199812)31:6<483::AID-MPO4>3.3.CO;2-0
   JENKIN RDT, 1987, J NEUROSURG, V66, P227, DOI 10.3171/jns.1987.66.2.0227
   KADOTA RP, 1994, PEDIATR NEUROSURG, V20, P221, DOI 10.1159/000120794
   KALIFA C, 1994, PEDIATR NEUROSURG, V21, P214
   KEDAR A, 1994, MED PEDIATR ONCOL, V23, P428, DOI 10.1002/mpo.2950230507
   KRETSCHMAR CS, 1993, CANCER-AM CANCER SOC, V72, P1404, DOI 10.1002/1097-0142(19930815)72:4<1404::AID-CNCR2820720441>3.0.CO;2-G
   LEVIN VA, 1979, J NEUROSURG, V51, P526, DOI 10.3171/jns.1979.51.4.0526
   LEVIN VA, 1984, NEUROSURGERY, V14, P679, DOI 10.1227/00006123-198406000-00006
   Mandell L, 1997, INT J RADIAT ONCOL, V39, P143, DOI 10.1016/S0360-3016(97)80575-5
   PACKER RJ, 1993, CANCER-AM CANCER SOC, V72, P1414, DOI 10.1002/1097-0142(19930815)72:4<1414::AID-CNCR2820720442>3.0.CO;2-C
   PACKER RJ, 1990, ANN NEUROL, V27, P167, DOI 10.1002/ana.410270212
   PACKER RJ, 1994, CANCER, V74, P1872
   PAKISCH B, 1992, CHILD NERV SYST, V8, P215, DOI 10.1007/BF00262849
   PENDERGRASS TW, 1987, J CLIN ONCOL, V5, P1221, DOI 10.1200/JCO.1987.5.8.1221
   POLLACK IF, 1990, CANCER RES, V50, P7134
   PRADOS MD, 1995, INT J RADIAT ONCOL, V32, P85, DOI 10.1016/0360-3016(95)00563-E
   Puchner MJA, 2000, J NEURO-ONCOL, V49, P147, DOI 10.1023/A:1026533016912
   See SJ, 2004, NEURO-ONCOLOGY, V6, P253, DOI 10.1215/S1152851703000607
   Walter AW, 1998, MED PEDIATR ONCOL, V30, P28, DOI 10.1002/(SICI)1096-911X(199801)30:1<28::AID-MPO9>3.0.CO;2-2
   WOLFF JEE, 1997, MED PEDIATR ONCOL, V29, P413
NR 45
TC 37
Z9 39
U1 0
U2 2
PU OXFORD UNIV PRESS INC
PI CARY
PA JOURNALS DEPT, 2001 EVANS RD, CARY, NC 27513 USA
SN 1522-8517
EI 1523-5866
J9 NEURO-ONCOLOGY
JI Neuro-Oncology
PD AUG
PY 2008
VL 10
IS 4
BP 599
EP 607
DI 10.1215/15228517-2008-029
PG 9
WC Oncology; Clinical Neurology
WE Science Citation Index Expanded (SCI-EXPANDED)
SC Oncology; Neurosciences & Neurology
GA 340XS
UT WOS:000258678900010
PM 18577561
OA Green Published
DA 2023-08-27
ER

PT J
AU Jansen, MHA
   Kloet, RW
   van Vuurden, DG
   van Zanten, SEMV
   Witte, BI
   Goldman, S
   Vandertop, WP
   Comans, EFI
   Hoekstra, OS
   Boellaard, R
   Kaspers, GJJL
AF Jansen, Marc H. A.
   Kloet, Reina W.
   van Vuurden, Dannis G.
   van Zanten, Sophie E. M. Veldhuijzen
   Witte, Birgit I.
   Goldman, Serge
   Vandertop, W. Peter
   Comans, Emile F. I.
   Hoekstra, Otto S.
   Boellaard, Ronald
   Kaspers, Gert-Jan J. L.
TI F-18-FDG PET standard uptake values of the normal pons in children:
   establishing a reference value for diffuse intrinsic pontine glioma
SO EJNMMI RESEARCH
LA English
DT Article
DE Positron emission tomography; [F-18]fluorodeoxyglucose; Pontine glioma;
   Brain neoplasms; Reference values; Pons
ID POSITRON-EMISSION-TOMOGRAPHY; BRAIN-STEM GLIOMAS; HIGH-GRADE GLIOMAS;
   FDG-PET; GLUCOSE-METABOLISM; MRI; SURVIVAL; SCANS
AB Background: Positron emission tomography (PET) scanning with [F-18]fluorodeoxyglucose (F-18-FDG) is a useful diagnostic and prediction tool in brain tumors, but its value in childhood diffuse intrinsic pontine glioma (DIPG) is still unclear. For interpretation of F-18-FDG PET results in DIPG, uptake values of the normal pons of children of increasing ages are mandatory. The aim of this study was to determine F-18-FDG standard uptake value ratios (SUVr) of the normal pons and to compare these to those of DIPG.
   Methods: We studied 36 subjects with a normal, non-affected pons (aged 5 to 23 years) and 6 patients with DIPG (aged 4 to 17 years) who underwent F-18-FDG PET scanning. Magnetic resonance imaging (MRI) was co-registered to define the regions of interest. SUVr and SUVrmax for the pons/cerebellum (SUVr(p/c)) and the pons/occipital lobe (SUVr(p/o)) were calculated. Independent-samples t tests and Mann-Whitney U tests were used to compare the mean SUVr and Pearson's test for correlations.
   Results: For the normal pons, mean SUVr(p/c) and SUVr(p/o) were 0.65 (+/- 0.054) and 0.51 (+/- 0.056), respectively. No significant correlations were found between the SUVr of the normal pons and sex, age, nor pontine volume. A modest but statistically significant correlation was found between SUVr and post-injection time acquisition timing. For DIPG, mean SUVr(p/c) and SUVr(p/o) were 0.74 (+/- 0.20) and 0.65 (+/- 0.30), respectively, while mean SUVr(p(max)/c) and SUVr(p(max)/o) were 1.95 (+/- 0.48) and 1.81 (+/- 0.20), respectively.
   Conclusion: The SUVr of the unaffected pons are strikingly constant between children, irrespective of sex and age, and can therefore be well used as a reference value for F-18-FDG PET studies in DIPG.
C1 [Jansen, Marc H. A.; van Vuurden, Dannis G.; van Zanten, Sophie E. M. Veldhuijzen; Kaspers, Gert-Jan J. L.] Vrije Univ Amsterdam Med Ctr, Dept Pediat, Div Hematol & Oncol, NL-1007 MB Amsterdam, Netherlands.
   [Kloet, Reina W.; Comans, Emile F. I.; Hoekstra, Otto S.; Boellaard, Ronald] Vrije Univ Amsterdam Med Ctr, Dept Radiol & Nucl Med, NL-1081 HV Amsterdam, Netherlands.
   [van Vuurden, Dannis G.] Canc Ctr Amsterdam, Neurooncol Res Grp, NL-1081 HV Amsterdam, Netherlands.
   [Witte, Birgit I.] Vrije Univ Amsterdam Med Ctr, Dept Epidemiol & Biostat, NL-1081 HV Amsterdam, Netherlands.
   [Goldman, Serge] ULB Hop Erasme Brussels, Dept Nucl Med, B-1070 Brussels, Belgium.
   [Vandertop, W. Peter] Vrije Univ Amsterdam Med Ctr, Neurosurg Ctr Amsterdam, NL-1081 HV Amsterdam, Netherlands.
C3 Vrije Universiteit Amsterdam; VU UNIVERSITY MEDICAL CENTER; Vrije
   Universiteit Amsterdam; VU UNIVERSITY MEDICAL CENTER; University of
   Amsterdam; Vrije Universiteit Amsterdam; Vrije Universiteit Amsterdam;
   VU UNIVERSITY MEDICAL CENTER; Universite Catholique Louvain; Cliniques
   Universitaires Saint-Luc; Universite Libre de Bruxelles; Vrije
   Universiteit Brussel; Vrije Universiteit Amsterdam; VU UNIVERSITY
   MEDICAL CENTER
RP Jansen, MHA (通讯作者)，Vrije Univ Amsterdam Med Ctr, Dept Pediat, Div Hematol & Oncol, Boelelaan 1118, NL-1007 MB Amsterdam, Netherlands.
EM
RI Kaspers, Gertjan/ABC-5334-2020; Vandertop, William P./K-9288-2018; van
   Vuurden, Dannis/ACA-6952-2022
OI Kaspers, Gertjan/0000-0001-7716-8475; Lissenberg-Witte,
   Birgit/0000-0001-9448-1826; Hoekstra, Otto S/0000-0003-0767-2734;
   Goldman, Serge/0000-0002-9917-8735; Boellaard,
   Ronald/0000-0002-0313-5686; Vandertop, William/0000-0001-5417-0265; Van
   Vuurden, Dannis/0000-0002-1364-9007; Veldhuijzen van Zanten, Sophie
   E.M./0000-0002-3888-0977
FU Semmy foundation; Egbers foundation
FX DIPG research is funded by the Semmy and Egbers foundations. The
   sponsors had no role in the preparation and execution of the study
   and/or manuscript.
CR Boellaard R, 2004, IEEE NUCL SCI CONF R, P2553
   Boellaard R, 2011, J NUCL MED, V52, p93S, DOI 10.2967/jnumed.110.085662
   Brix G, 1997, J NUCL MED, V38, P1614
   BRUGGERS CS, 1993, MED PEDIATR ONCOL, V21, P301, DOI 10.1002/mpo.2950210414
   Caretti V, 2013, NEUROPATH APPL NEURO, V39, P426, DOI 10.1111/j.1365-2990.2012.01294.x
   Colavolpe C, 2012, NEURO-ONCOLOGY, V14, P649, DOI 10.1093/neuonc/nos012
   Colavolpe C, 2012, J NEURO-ONCOL, V107, P527, DOI 10.1007/s11060-011-0771-6
   DICHIRO G, 1983, J COMPUT ASSIST TOMO, V7, P937
   Ferrie CD, 1997, J NEUROL NEUROSUR PS, V63, P181, DOI 10.1136/jnnp.63.2.181
   Ferrie CD, 1997, J NUCL MED, V38, P1891
   Goldman S, 1996, CANCER, V78, P1098, DOI 10.1002/(SICI)1097-0142(19960901)78:5<1098::AID-CNCR21>3.3.CO;2-A
   Goldman S, 1997, J NUCL MED, V38, P1459
   Hargrave D, 2008, J NEURO-ONCOL, V86, P313, DOI 10.1007/s11060-007-9473-5
   Jansen MHA, 2012, CANCER TREAT REV, V38, P27, DOI 10.1016/j.ctrv.2011.06.007
   Jansen MHA, 2012, EXPERT REV ANTICANC, V12, P1109, DOI [10.1586/ERA.12.95, 10.1586/era.12.95]
   Kwon JW, 2006, PEDIATR RADIOL, V36, P959, DOI 10.1007/s00247-006-0256-5
   LEIDERMAN DB, 1991, EPILEPSIA, V32, P417, DOI 10.1111/j.1528-1157.1991.tb04671.x
   Pirotte B, 2004, J NEUROSURG, V101, P476, DOI 10.3171/jns.2004.101.3.0476
   Pirotte BJM, 2007, J NEUROSURG, V107, P392, DOI 10.3171/PED-07/11/392
   Roelcke U, 1998, J NUCL MED, V39, P879
   Rosenfeld A, 2011, J PEDIAT HEMATOL ONC, V33, P369, DOI 10.1097/MPH.0b013e31820ad915
   THEODORE WH, 1987, EPILEPSIA, V28, P319, DOI 10.1111/j.1528-1157.1987.tb03650.x
   THEODORE WH, 1988, EPILEPSIA, V29, pS48, DOI 10.1111/j.1528-1157.1988.tb05797.x
   Zukotynski KA, 2011, J NUCL MED, V52, P188, DOI 10.2967/jnumed.110.081463
NR 24
TC 4
Z9 4
U1 0
U2 2
PU SPRINGEROPEN
PI LONDON
PA CAMPUS, 4 CRINAN ST, LONDON, N1 9XW, ENGLAND
SN 2191-219X
J9 EJNMMI RES
JI EJNMMI Res.
PD JAN 28
PY 2014
VL 4
AR 8
DI 10.1186/2191-219X-4-8
PG 7
WC Radiology, Nuclear Medicine & Medical Imaging
WE Science Citation Index Expanded (SCI-EXPANDED)
SC Radiology, Nuclear Medicine & Medical Imaging
GA CM7GI
UT WOS:000357858400001
PM 24472395
OA gold, Green Published
DA 2023-08-27
ER

PT J
AU Zarghooni, M
   Bartels, U
   Lee, E
   Buczkowicz, P
   Morrison, A
   Huang, A
   Bouffet, E
   Hawkins, C
AF Zarghooni, Maryam
   Bartels, Ute
   Lee, Eric
   Buczkowicz, Pawel
   Morrison, Andrew
   Huang, Annie
   Bouffet, Eric
   Hawkins, Cynthia
TI Whole-Genome Profiling of Pediatric Diffuse Intrinsic Pontine Gliomas
   Highlights Platelet-Derived Growth Factor Receptor alpha and Poly
   (ADP-ribose) Polymerase As Potential Therapeutic Targets
SO JOURNAL OF CLINICAL ONCOLOGY
LA English
DT Article; Proceedings Paper
CT 85th Annual Meeting of the American-Association-of-Neuropathologists
CY JUN 11-14, 2009
CL San Antonio, TX
SP Amer Assoc Neuropathol
ID BRAIN-STEM GLIOMAS; POLY(ADP-RIBOSE) POLYMERASE; EGFR AMPLIFICATION;
   EXPRESSION; GLIOBLASTOMA; REPAIR; INHIBITION; CHILDHOOD; CELLS;
   RADIOSENSITIZATION
AB Purpose Diffuse intrinsic pontine glioma (DIPG) is one of the most devastating of pediatric malignancies and one for which no effective therapy exists. A major contributor to the failure of therapeutic trials is the assumption that biologic properties of brainstem tumors in children are identical to cerebral high-grade gliomas of adults. A better understanding of the biology of DIPG itself is needed in order to develop agents targeted more specifically to these children's disease. Herein, we address this lack of knowledge by performing the first high-resolution single nucleotide polymorphism (SNP) -based DNA microarray analysis of a series of DIPGs.
   Patients and Methods Eleven samples (nine postmortem and two pretreatment surgical samples), the largest series thus far examined, were hybridized to SNP arrays (250 k or 6.0). The study was approved by the research ethics board at our institution. All array findings were validated using quantitative polymerase chain reaction, fluorescence in situ hybridization, immunohistochemistry, and/or microsatellite analysis.
   Results Analysis of DIPG copy number alterations showed recurrent changes distinct from those of pediatric supratentorial high-grade astrocytomas. Thirty-six percent of DIPGs had gains in platelet-derived growth factor receptor alpha (PDGFRA; 4 to 18 copies) and all showed PDGFR-alpha expression. Low-level gains in poly (ADP-ribose) polymerase (PARP) -1 were identified in three cases. Pathway analysis revealed genes with loss of heterozygosity were enriched for DNA repair pathways.
   Conclusion To our knowledge, our data provides the first, comprehensive high-resolution genomic analysis of pediatric DIPG. Our findings of recurrent involvement of the PDGFR pathway as well as defects in DNA repair pathways coupled with gain of PARP-1 highlight two potential, biologically based, therapeutic targets directed specifically at this devastating disease.
C1 [Hawkins, Cynthia] Hosp Sick Children, Div Pathol, Labatt Brain Tumor Res Ctr, Toronto, ON M5G 1X8, Canada.
   Hosp Sick Children, Div Haematol & Oncol, Labatt Brain Tumor Res Ctr, Toronto, ON M5G 1X8, Canada.
   Univ Toronto, Dept Lab Med & Pathobiol, Toronto, ON, Canada.
C3 University of Toronto; Hospital for Sick Children (SickKids); University
   of Toronto; Hospital for Sick Children (SickKids); University of Toronto
RP Hawkins, C (通讯作者)，Hosp Sick Children, Div Pathol, Labatt Brain Tumor Res Ctr, 555 Univ Ave, Toronto, ON M5G 1X8, Canada.
EM
RI Bouffet, Eric/AAB-8999-2020
FU Canadian Institutes of Health Research [MOP 82727] Funding Source:
   Medline
CR Ame JC, 2004, BIOESSAYS, V26, P882, DOI 10.1002/bies.20085
   Ashworth A, 2008, J CLIN ONCOL, V26, P3785, DOI 10.1200/JCO.2008.16.0812
   Bredel M, 1999, CLIN CANCER RES, V5, P1786
   Cheng CL, 2005, MOL CANCER THER, V4, P1364, DOI 10.1158/1535-7163.MCT-05-0128
   Cheng Y, 1999, HUM PATHOL, V30, P1284, DOI 10.1016/S0046-8177(99)90057-6
   Donaldson SS, 2006, J CLIN ONCOL, V24, P1266, DOI 10.1200/JCO.2005.04.6599
   Dungey FA, 2008, INT J RADIAT ONCOL, V72, P1188, DOI 10.1016/j.ijrobp.2008.07.031
   Faury D, 2007, J CLIN ONCOL, V25, P1196, DOI 10.1200/JCO.2006.07.8626
   Freeman CR, 1999, CHILD NERV SYST, V15, P545, DOI 10.1007/s003810050542
   Gilbertson RJ, 2003, CLIN CANCER RES, V9, P3620
   Hargrove D, 2006, LANCET ONCOL, V7, P241, DOI 10.1016/S1470-2045(06)70615-5
   Iwanaga Y, 2002, CANCER RES, V62, P2618
   Leung SY, 1998, AM J PATHOL, V153, P1181, DOI 10.1016/S0002-9440(10)65662-3
   Liu LL, 2006, CLIN CANCER RES, V12, P328, DOI 10.1158/1078-0432.CCR-05-2543
   LOUIS DN, 1993, J NEUROPATH EXP NEUR, V52, P507, DOI 10.1097/00005072-199309000-00009
   MARIA BL, 1993, J CHILD NEUROL, V8, P112, DOI 10.1177/088307389300800203
   Muscal JA, 2010, CANCER CHEMOTH PHARM, V65, P419, DOI 10.1007/s00280-009-1044-3
   Nicholas MK, 2006, CLIN CANCER RES, V12, P7261, DOI 10.1158/1078-0432.CCR-06-0874
   Ohgaki H, 2007, AM J PATHOL, V170, P1445, DOI 10.2353/ajpath.2007.070011
   Pollack IF, 2006, J NEUROSURG, V105, P418, DOI 10.3171/ped.2006.105.5.418
   Rabbee N, 2006, BIOINFORMATICS, V22, P7, DOI 10.1093/bioinformatics/bti741
   Ratnam K, 2007, CLIN CANCER RES, V13, P1383, DOI 10.1158/1078-0432.CCR-06-2260
   Ruscetti T, 1998, J BIOL CHEM, V273, P14461, DOI 10.1074/jbc.273.23.14461
   Russo AL, 2009, CLIN CANCER RES, V15, P607, DOI 10.1158/1078-0432.CCR-08-2079
   Schreiber V, 2002, J BIOL CHEM, V277, P23028, DOI 10.1074/jbc.M202390200
   STILLER CA, 1994, BRIT MED J, V309, P1612, DOI 10.1136/bmj.309.6969.1612
   Sung T, 2000, BRAIN PATHOL, V10, P249, DOI 10.1111/j.1750-3639.2000.tb00258.x
   Tabori U, 2006, J CLIN ONCOL, V24, P1522, DOI 10.1200/JCO.2005.04.2127
   Tentori L, 2002, GLIA, V40, P44, DOI 10.1002/glia.10113
   Wharton SB, 2000, NEUROPATH APPL NEURO, V26, P528, DOI 10.1046/j.0305-1846.2000.00288.x
   Wong KK, 2006, CANCER RES, V66, P11172, DOI 10.1158/0008-5472.CAN-06-2438
NR 31
TC 232
Z9 233
U1 0
U2 20
PU AMER SOC CLINICAL ONCOLOGY
PI ALEXANDRIA
PA 2318 MILL ROAD, STE 800, ALEXANDRIA, VA 22314 USA
SN 0732-183X
EI 1527-7755
J9 J CLIN ONCOL
JI J. Clin. Oncol.
PD MAR 10
PY 2010
VL 28
IS 8
BP 1337
EP 1344
DI 10.1200/JCO.2009.25.5463
PG 8
WC Oncology
WE Science Citation Index Expanded (SCI-EXPANDED); Conference Proceedings Citation Index - Science (CPCI-S)
SC Oncology
GA 565RP
UT WOS:000275312300012
PM 20142589
DA 2023-08-27
ER

PT J
AU Bettegowda, C
   Agrawal, N
   Jiao, YC
   Wang, YX
   Wood, LD
   Rodriguez, FJ
   Hruban, RH
   Gallia, GL
   Binder, ZA
   Riggins, CJ
   Salmasi, V
   Riggins, GJ
   Reitman, ZJ
   Rasheed, A
   Keir, S
   Shinjo, S
   Marie, S
   McLendon, R
   Jallo, G
   Vogelstein, B
   Bigner, D
   Yan, H
   Kinzler, KW
   Papadopoulos, N
AF Bettegowda, Chetan
   Agrawal, Nishant
   Jiao, Yuchen
   Wang, Yuxuan
   Wood, Laura D.
   Rodriguez, Fausto J.
   Hruban, Ralph H.
   Gallia, Gary L.
   Binder, Zev A.
   Riggins, Callen J.
   Salmasi, Vafi
   Riggins, Gregory J.
   Reitman, Zachary J.
   Rasheed, Ahmed
   Keir, Stephen
   Shinjo, Sueli
   Marie, Suely
   McLendon, Roger
   Jallo, George
   Vogelstein, Bert
   Bigner, Darell
   Yan, Hai
   Kinzler, Kenneth W.
   Papadopoulos, Nickolas
TI Exomic Sequencing of Four Rare Central Nervous System Tumor Types
SO ONCOTARGET
LA English
DT Article
DE Central nervous system (CNS) tumors; cancer genetics; exome sequencing;
   pediatric tumors; brain tumors
ID PLEOMORPHIC XANTHOASTROCYTOMA; DRIVER MUTATIONS; GENOMIC ANALYSIS; IDH1;
   EVEROLIMUS; SUBGROUPS; FUBP1; GENE
AB A heterogeneous population of uncommon neoplasms of the central nervous system (CNS) cause significant morbidity and mortality. To explore their genetic origins, we sequenced the exomes of 12 pleomorphic xanthoastrocytomas (PXA), 17 non-brainstem pediatric glioblastomas (PGBM), 8 intracranial ependymomas (IEP) and 8 spinal cord ependymomas (SCEP). Analysis of the mutational spectra revealed that the predominant single base pair substitution was a C: G>T: A transition in each of the four tumor types. Our data confirm the critical roles of several known driver genes within CNS neoplasms, including TP53 and ATRX in PGBM, and NF2 in SCEPs. Additionally, we show that activating BRAF mutations play a central role in both low and high grade glial tumors. Furthermore, alterations in genes coding for members of the mammalian target of rapamycin (mTOR) pathway were observed in 33% of PXA. Our study supports the hypothesis that pathologically similar tumors arising in different age groups and from different compartments may represent distinct disease processes with varied genetic composition.
C1 [Bettegowda, Chetan; Gallia, Gary L.; Binder, Zev A.; Riggins, Callen J.; Riggins, Gregory J.; Jallo, George] Johns Hopkins Univ, Sch Med, Dept Neurosurg, Baltimore, MD 21205 USA.
   [Bettegowda, Chetan; Agrawal, Nishant; Jiao, Yuchen; Wang, Yuxuan; Vogelstein, Bert; Kinzler, Kenneth W.; Papadopoulos, Nickolas] Johns Hopkins Univ, Sch Med, Ludwig Ctr Canc Genet, Baltimore, MD USA.
   [Agrawal, Nishant] Johns Hopkins Univ, Sch Med, Dept Otolaryngol Head & Neck Surg, Baltimore, MD 21205 USA.
   [Wood, Laura D.; Rodriguez, Fausto J.; Hruban, Ralph H.] Johns Hopkins Univ, Sch Med, Dept Pathol, Baltimore, MD 21205 USA.
   [Salmasi, Vafi] Cleveland Clin Hosp, Dept Anesthesiol, Cleveland, OH USA.
   [Reitman, Zachary J.; Rasheed, Ahmed; Keir, Stephen; McLendon, Roger; Bigner, Darell; Yan, Hai] Duke Univ, Med Ctr, Preston Robert Tisch Brain Tumor Ctr, Durham, NC USA.
   [Shinjo, Sueli; Marie, Suely] Univ Sao Paulo, Dept Pathol, Sch Med, Sao Paulo, Brazil.
C3 Johns Hopkins University; Johns Hopkins University; Johns Hopkins
   University; Johns Hopkins University; Cleveland Clinic Foundation; Duke
   University; Universidade de Sao Paulo
RP Bettegowda, C (通讯作者)，Johns Hopkins Univ, Sch Med, Dept Neurosurg, Baltimore, MD 21205 USA.
EM
RI Salmasi, Vafi/E-9160-2014; Wood, Laura/ABF-1531-2020; Marie, Suely Kazue
   Nagahashi/D-1870-2012; jiao, yuchen/H-6313-2011; Oba-Shinjo, Sueli
   M./C-4017-2012; Salmasi, Vafi/O-9908-2019
OI Marie, Suely Kazue Nagahashi/0000-0003-4419-7928; Oba-Shinjo, Sueli
   M./0000-0002-1650-5973; Salmasi, Vafi/0000-0002-1288-4531; Reitman,
   Zachary/0000-0002-9122-9550
FU Virginia and D.K. Ludwig Fund for Cancer Research; Pediatric Brain Tumor
   Foundation; Malia's Cord Foundation; Burroughs Wellcome Career Award for
   Medical Scientists; Johns Hopkins Clinical Scientist Award [RC2DE020957,
   5R01-CA140316, CA057345]; American Cancer Society Research Scholar Award
   [RSG-10-126-01-CCE]; FAPESP [01/12898-4, 04/12133-6]; Fundacao de Amparo
   a Pesquisa do Estado de Sao Paulo (FAPESP) [04/12133-6] Funding Source:
   FAPESP
FX We thank our patients for their courage and generosity. We also thank J.
   Ptak, N. Silliman, L. Dobbyn, J. Schaeffer and D. Singh. The work was
   supported by the Virginia and D.K. Ludwig Fund for Cancer Research,
   Pediatric Brain Tumor Foundation, Malia's Cord Foundation, Burroughs
   Wellcome Career Award for Medical Scientists, Johns Hopkins Clinical
   Scientist Award, RC2DE020957, 5R01-CA140316, CA057345, American Cancer
   Society Research Scholar Award RSG-10-126-01-CCE, FAPESP 01/12898-4 and
   FAPESP 04/12133-6.
CR Agrawal N, 2012, J CLIN ENDOCRINOL ME
   Agrawal N, 2012, CANCER DISCOV, V2, P899, DOI 10.1158/2159-8290.CD-12-0189
   Agrawal N, 2011, SCIENCE, V333, P1154, DOI 10.1126/science.1206923
   [Anonymous], SCIENCE
   [Anonymous], 2012, CANC FACTS FIG 2012
   Bettegowda C, 2011, SCIENCE, V333, P1453, DOI 10.1126/science.1210557
   Broniscer A, 2004, ONCOLOGIST, V9, P197, DOI 10.1634/theoncologist.9-2-197
   Chin L, 2008, NATURE, V455, P1061, DOI 10.1038/nature07385
   Dias-Santagata D, 2011, PLOS ONE, V6, DOI 10.1371/journal.pone.0017948
   Dunn GP, 2012, GENE DEV, V26, P756, DOI 10.1101/gad.187922.112
   Giannini C, 1999, CANCER, V85, P2033, DOI 10.1002/(SICI)1097-0142(19990501)85:9<2033::AID-CNCR22>3.3.CO;2-Q
   Horbinski C, 2010, ACTA NEUROPATHOL, V119, P641, DOI 10.1007/s00401-009-0634-9
   Iyer G, 2012, SCIENCE, V338, P221, DOI 10.1126/science.1226344
   Jiao YC, 2012, ONCOTARGET, V3, P709, DOI 10.18632/oncotarget.588
   Johnson RA, 2010, NATURE, V466, P632, DOI 10.1038/nature09173
   Jones DTW, 2008, CANCER RES, V68, P8673, DOI 10.1158/0008-5472.CAN-08-2097
   Jones DTW, 2012, NATURE, V488, P100, DOI 10.1038/nature11284
   Kilday JP, 2009, MOL CANCER RES, V7, P765, DOI 10.1158/1541-7786.MCR-08-0584
   Krueger DA, 2010, NEW ENGL J MED, V363, P1801, DOI 10.1056/NEJMoa1001671
   MacDonald TJ, NEUROONCOL, V13, P1049
   Marton E, 2007, J NEUROL SCI, V252, P144, DOI 10.1016/j.jns.2006.11.008
   Milano MT, 2010, J NEURO-ONCOL, V98, P83, DOI 10.1007/s11060-009-0054-7
   Parsons DW, 2008, SCIENCE, V321, P1807, DOI 10.1126/science.1164382
   Parsons DW, 2011, SCIENCE, V331, P435, DOI 10.1126/science.1198056
   Pugh TJ, 2012, NATURE, V488, P106, DOI 10.1038/nature11329
   Rao AAN, 2010, PEDIATR BLOOD CANCER, V55, P290, DOI 10.1002/pbc.22490
   Rausch T, 2012, CELL, V148, P59, DOI 10.1016/j.cell.2011.12.013
   Reddy AT, 2003, CANCER J, V9, P107, DOI 10.1097/00130404-200303000-00006
   Robinson G, 2012, NATURE, V488, P43, DOI 10.1038/nature11213
   Schiffman JD, 2010, CANCER RES, V70, P512, DOI 10.1158/0008-5472.CAN-09-1851
   Schwartzentruber J, 2012, NATURE, V482, P226, DOI [10.1038/nature10833, 10.1007/s13311-012-0137-6]
   Sturm D, 2012, CANCER CELL, V22, P425, DOI 10.1016/j.ccr.2012.08.024
   Witt H, 2011, CANCER CELL, V20, P143, DOI 10.1016/j.ccr.2011.07.007
   WONG AJ, 1992, P NATL ACAD SCI USA, V89, P2965, DOI 10.1073/pnas.89.7.2965
   Wu G, 2012, NAT GENET, V44, P251, DOI 10.1038/ng.1102
   Yan H, 2009, NEW ENGL J MED, V360, P765, DOI 10.1056/NEJMoa0808710
   Yip S, 2012, J PATHOL, V226, P7, DOI 10.1002/path.2995
NR 37
TC 56
Z9 57
U1 0
U2 8
PU IMPACT JOURNALS LLC
PI ORCHARD PARK
PA 6666 E QUAKER ST, STE 1, ORCHARD PARK, NY 14127 USA
EI 1949-2553
J9 ONCOTARGET
JI Oncotarget
PD APR
PY 2013
VL 4
IS 4
BP 572
EP 583
DI 10.18632/oncotarget.964
PG 12
WC Oncology; Cell Biology
WE Science Citation Index Expanded (SCI-EXPANDED)
SC Oncology; Cell Biology
GA 142GA
UT WOS:000318783800010
PM 23592488
OA hybrid, Green Published, Green Submitted
DA 2023-08-27
ER

PT J
AU Lindblad, C
   Neumann, S
   Kolbeinsdottir, S
   Zachariadis, V
   Thelin, EP
   Enge, M
   Thams, S
   Brundin, L
   Svensson, M
AF Lindblad, Caroline
   Neumann, Susanne
   Kolbeinsdottir, Solrun
   Zachariadis, Vasilios
   Thelin, Eric P.
   Enge, Martin
   Thams, Sebastian
   Brundin, Lou
   Svensson, Mikael
TI Stem cell-derived brainstem mouse astrocytes obtain a neurotoxic
   phenotype in vitro upon neuroinflammation
SO JOURNAL OF INFLAMMATION-LONDON
LA English
DT Article
DE Embryonic stem cells; Disease modelling; Traumatic axonal injury;
   Ventral brainstem- or rostroventral spinal astrocytes; Astrocytes;
   Neuroinflammation
ID CENTRAL-NERVOUS-SYSTEM; DIFFUSE AXONAL INJURY; MESSENGER-RNA;
   GENE-EXPRESSION; ACTIVATION; PROTEIN; DIFFERENTIATION; SUBTYPES;
   DATABASE; NEURONS
AB BackgroundAstrocytes respond to injury and disease through a process known as reactive astrogliosis, of which inflammatory signaling is one subset. This inflammatory response is heterogeneous with respect to the inductive stimuli and the afflicted central nervous system region. This is of plausible importance in e.g. traumatic axonal injury (TAI), where lesions in the brainstem carries a particularly poor prognosis. In fact, astrogliotic forebrain astrocytes were recently suggested to cause neuronal death following axotomy. We therefore sought to assess if ventral brainstem- or rostroventral spinal astrocytes exert similar effects on motor neurons in vitro.MethodsWe derived brainstem/rostroventral spinal astrocyte-like cells (ES-astrocytes) and motor neurons using directed differentiation of mouse embryonic stem cells (ES). We activated the ES-astrocytes using the neurotoxicity-eliciting cytokines interleukin- (IL-) 1 & alpha; and tumor necrosis factor-(TNF-)& alpha; and clinically relevant inflammatory mediators. In co-cultures with reactive ES-astrocytes and motor neurons, we assessed neurotoxic ES-astrocyte activity, similarly to what has previously been shown for other central nervous system (CNS) regions.ResultsWe confirmed the brainstem/rostroventral ES-astrocyte identity using RNA-sequencing, immunocytochemistry, and by comparison with primary subventricular zone-astrocytes. Following cytokine stimulation, the c-Jun N-terminal kinase pathway down-stream product phosphorylated c-Jun was increased, thus demonstrating ES-astrocyte reactivity. These reactive ES-astrocytes conferred a contact-dependent neurotoxic effect upon co-culture with motor neurons. When exposed to IL-1 & beta; and IL-6, two neuroinflammatory cytokines found in the cerebrospinal fluid and serum proteome following human severe traumatic brain injury (TBI), ES-astrocytes exerted similar effects on motor neurons. Activation of ES-astrocytes by these cytokines was associated with pathways relating to endoplasmic reticulum stress and altered regulation of MYC.ConclusionsVentral brainstem and rostroventral spinal cord astrocytes differentiated from mouse ES can exert neurotoxic effects in vitro. This highlights how neuroinflammation following CNS lesions can exert region- and cell-specific effects. Our in vitro model system, which uniquely portrays astrocytes and neurons from one niche, allows for a detailed and translationally relevant model system for future studies on how to improve neuronal survival in particularly vulnerable CNS regions following e.g. TAI.
C1 [Lindblad, Caroline; Neumann, Susanne; Thelin, Eric P.; Thams, Sebastian; Brundin, Lou; Svensson, Mikael] Karolinska Inst, Karolinska Univ Sjukhuset Solna, Dept Clin Neurosci, J520 Svensson Grp, S-17177 Stockholm, Sweden.
   [Kolbeinsdottir, Solrun; Zachariadis, Vasilios; Enge, Martin] Karolinska Inst, Dept Oncol Pathol, Stockholm, Sweden.
   [Thelin, Eric P.; Thams, Sebastian; Brundin, Lou] Karolinska Univ Hosp, Dept Neurol, Stockholm, Sweden.
   [Svensson, Mikael] Karolinska Univ Hosp, Dept Neurosurg, Stockholm, Sweden.
C3 Karolinska Institutet; Karolinska Institutet; Karolinska Institutet;
   Karolinska University Hospital; Karolinska Institutet; Karolinska
   University Hospital
RP Lindblad, C (通讯作者)，Karolinska Inst, Karolinska Univ Sjukhuset Solna, Dept Clin Neurosci, J520 Svensson Grp, S-17177 Stockholm, Sweden.
EM
FU Karolinska Institute [K825715022, K825715052]; Strategic Research Area
   Neuroscience (StratNeuro); Erling-Persson Family Foundation; Region
   Stockholm (Clinical Research Appointment) [FoUI-955376]; Swedish Brain
   Foundation [FO2019-0006]; SFO StratRegen; Swedish Cancer Society;
   Swedish Childhood Cancer Fund; Radiumhemmets forskningsfonder; Swedish
   Research Council [202002940, 201901284]; Cancer Research KI; Region
   Stockholm; Karolinska Institutet
FX Open access funding provided by Karolinska Institute. The authors would
   like to acknowledge funders who enabled this work. CL received funding
   for this study through the Karolinska Institute Funds Clinical Scientist
   Training Program and Research Internship Program (grant number
   K825715022, K825715052). EPT acknowledges funding support from Strategic
   Research Area Neuroscience (StratNeuro), The Erling-Persson Family
   Foundation, Region Stockholm (Clinical Research Appointment,
   FoUI-955376) and the Swedish Brain Foundation (#FO2019-0006). The Enge
   laboratory is supported by SFO StratRegen, The Swedish Cancer Society,
   The Swedish Childhood Cancer Fund, Radiumhemmets forskningsfonder, The
   Swedish Research Council (2020-02940) and Cancer Research KI. The
   Brundin laboratory is supported by The Swedish Research Council
   (2019-01284), the Swedish Brain Foundation, Region Stockholm, and
   Karolinska Institutet. The funders did not participate in study
   design/conceptualization, data acquisition, data
   analysis/interpretation, manuscript compilation, or submission decision.
CR Abbott NJ, 2006, NAT REV NEUROSCI, V7, P41, DOI 10.1038/nrn1824
   Abu Hamdeh S, 2018, SCI REP-UK, V8, DOI 10.1038/s41598-018-25060-0
   Abu Hamdeh S, 2017, J NEUROTRAUM, V34, P341, DOI 10.1089/neu.2016.4426
   ADAMS JH, 1989, HISTOPATHOLOGY, V15, P49, DOI 10.1111/j.1365-2559.1989.tb03040.x
   Adey A, 2010, GENOME BIOL, V11, DOI 10.1186/gb-2010-11-12-r119
   Allodi I, 2019, STEM CELL REP, V12, P1329, DOI 10.1016/j.stemcr.2019.04.009
   Allodi I, 2014, FRONT NEUROSCI-SWITZ, V8, DOI 10.3389/fnins.2014.00109
   Anderson MA, NEUROSCI LETT 2013 1
   Anderson MA, 2016, NATURE, V532, P195, DOI 10.1038/nature17623
   Anderson MA, 2014, NEUROSCI LETT, V565, P23, DOI 10.1016/j.neulet.2013.12.030
   [Anonymous], 2017, GRAPHICS
   Ashburner M, 2000, NAT GENET, V25, P25, DOI 10.1038/75556
   Barbar L, 2020, NEURON, V107, P436, DOI 10.1016/j.neuron.2020.05.014
   Berglund R, 2020, SCI IMMUNOL, V5, DOI 10.1126/sciimmunol.abb5077
   Bonni A, 1997, SCIENCE, V278, P477, DOI 10.1126/science.278.5337.477
   Burda JE, 2016, EXP NEUROL, V275, P305, DOI 10.1016/j.expneurol.2015.03.020
   Bush TG, 1999, NEURON, V23, P297, DOI 10.1016/S0896-6273(00)80781-3
   Cahoy JD, 2008, J NEUROSCI, V28, P264, DOI 10.1523/JNEUROSCI.4178-07.2008
   Canals I, 2018, NAT METHODS, V15, P693, DOI 10.1038/s41592-018-0103-2
   Carlson M, 2021, SET ANNOTATION MAPS
   Clarke BE, 2020, BRAIN, V143, pE85, DOI 10.1093/brain/awaa258
   Covacu R, 2014, J NEUROSCI, V34, P11571, DOI 10.1523/JNEUROSCI.0231-14.2014
   Covacu R, 2006, STEM CELLS, V24, P2792, DOI 10.1634/stemcells.2005-0640
   Croft D, 2014, NUCLEIC ACIDS RES, V42, pD472, DOI [10.1093/nar/gkt1102, 10.1093/nar/gkz1031]
   Csuka E, 2000, NEUROREPORT, V11, P2587, DOI 10.1097/00001756-200008030-00047
   Dag O, ONEWAYTESTS R PACKAG
   Dobin A, 2013, BIOINFORMATICS, V29, P15, DOI 10.1093/bioinformatics/bts635
   Doetsch F, 1999, CELL, V97, P703, DOI 10.1016/S0092-8674(00)80783-7
   Dolgalev I., 2021, MSIGDBR MSIGDB GENE
   Durinck S, 2005, BIOINFORMATICS, V21, P3439, DOI 10.1093/bioinformatics/bti525
   Durinck S, 2009, NAT PROTOC, V4, P1184, DOI 10.1038/nprot.2009.97
   Edgar R, 2002, NUCLEIC ACIDS RES, V30, P207, DOI 10.1093/nar/30.1.207
   Ekmark-Lewen S, 2013, J NEUROINFLAMM, V10, DOI 10.1186/1742-2094-10-44
   Eng LF, 2000, NEUROCHEM RES, V25, P1439, DOI 10.1023/A:1007677003387
   Escartin C, 2021, NAT NEUROSCI, V24, P312, DOI 10.1038/s41593-020-00783-4
   Foo LC, 2011, NEURON, V71, P799, DOI 10.1016/j.neuron.2011.07.022
   Fox J. WS., 2018, R COMPANION APPL REG
   Freeman MR, SPECIFICATION MORPHO
   Gaetani L, 2019, J NEUROL NEUROSUR PS, V90, P870, DOI 10.1136/jnnp-2018-320106
   Gao K, TRAUMATIC SCRATCH IN
   Guttenplan KA, 2021, NATURE, V599, P102, DOI 10.1038/s41586-021-03960-y
   Hans VHJ, 1999, J CEREBR BLOOD F MET, V19, P184, DOI 10.1097/00004647-199902000-00010
   Hasel P, 2021, NAT NEUROSCI, V24, P1475, DOI 10.1038/s41593-021-00905-6
   Hedlund E, 2008, STEM CELLS, V26, P1526, DOI 10.1634/stemcells.2007-0996
   Helmy A, 2011, PROG NEUROBIOL, V95, P352, DOI 10.1016/j.pneurobio.2011.09.003
   Hochstim C, 2008, CELL, V133, P510, DOI 10.1016/j.cell.2008.02.046
   Iram T, 2016, J NEUROSCI, V36, P5185, DOI 10.1523/JNEUROSCI.3850-15.2016
   Jochum W, 2001, ONCOGENE, V20, P2401, DOI 10.1038/sj.onc.1204389
   Johansson CB, 1999, CELL, V96, P25, DOI 10.1016/S0092-8674(00)80956-3
   John CR, 2020, SCI REP-UK, V10, DOI 10.1038/s41598-020-58766-1
   jordbruksverket, 2019, STAT JORDBR FORT
   Kanehisa M, 2000, NUCLEIC ACIDS RES, V28, P27, DOI 10.1093/nar/28.1.27
   Khalil M, 2018, NAT REV NEUROL, V14, P577, DOI 10.1038/s41582-018-0058-z
   Khan Shaukat A, 2012, BMC Res Notes, V5, P636, DOI 10.1186/1756-0500-5-636
   Kim BJ, 2004, EXP MOL MED, V36, P52, DOI 10.1038/emm.2004.7
   Kitchen P, 2020, CELL, V181, P784, DOI 10.1016/j.cell.2020.03.037
   Kleiderman S, 2016, GLIA, V64, P695, DOI 10.1002/glia.22954
   Korotkevich G., 2019, FAST GENE SET ENRICH, DOI [DOI 10.1101/060012, 10.1101/060012v3]
   Krencik R, 2011, NAT BIOTECHNOL, V29, P528, DOI 10.1038/nbt.1877
   Kumar A, 2012, BRAIN BEHAV IMMUN, V26, P1191, DOI 10.1016/j.bbi.2012.06.008
   Li JW, 2021, NAT COMMUN, V12, DOI 10.1038/s41467-021-24232-3
   Li ZY, 2021, SCI ADV, V7, DOI 10.1126/sciadv.abe4458
   Liberzon A, 2015, CELL SYST, V1, P417, DOI 10.1016/j.cels.2015.12.004
   Liberzon A, 2011, BIOINFORMATICS, V27, P1739, DOI 10.1093/bioinformatics/btr260
   Liddelow SA, 2017, NATURE, V541, P481, DOI 10.1038/nature21029
   Lindblad C, 2021, BIOMED CENTRAL, P1, DOI 10
   Love M. I., 2017, BIOCONDUCTOR, V2, P1, DOI DOI 10.1186/S13059-014-0550-8
   Love MI, 2014, GENOME BIOL, V15, DOI 10.1186/s13059-014-0550-8
   Lozzi B, 2020, FRONT NEUROSCI-SWITZ, V14, DOI 10.3389/fnins.2020.00061
   MacDonald JM, 2013, CELL DEATH DIFFER, V20, P1140, DOI 10.1038/cdd.2013.30
   McManus MF, 1999, J NEUROSCI, V19, P9004, DOI 10.1523/JNEUROSCI.19-20-09004.1999
   Medana IM, 2003, BRAIN, V126, P515, DOI 10.1093/brain/awg061
   Middeldorp J, 2011, PROG NEUROBIOL, V93, P421, DOI 10.1016/j.pneurobio.2011.01.005
   Molofsky AV, 2012, GENE DEV, V26, P891, DOI 10.1101/gad.188326.112
   Molofsky AV, 2015, GLIA, V63, P1320, DOI 10.1002/glia.22836
   Morel L, 2017, J NEUROSCI, V37, P8706, DOI 10.1523/JNEUROSCI.3956-16.2017
   Moulson AJ, 2021, FRONT CELL NEUROSCI, V15, DOI 10.3389/fncel.2021.703810
   Nagai M, 2007, NAT NEUROSCI, V10, P615, DOI 10.1038/nn1876
   Neuwirth E., 2014, 2014 COLORBREWER PAL, DOI DOI 10.1007/978-3-642-12772-4
   Ogle D. H., 2021, FSA FISHERIES STOCK
   Oida T, 2010, J IMMUNOL METHODS, V362, P195, DOI 10.1016/j.jim.2010.09.008
   Okusawa BYS, 1988, C5A STIMULATES SECRE, P168
   Pekny M, 2014, NEUROSCI LETT, V565, P30, DOI 10.1016/j.neulet.2013.12.071
   Perriot S, 2018, STEM CELL REP, V11, P1199, DOI 10.1016/j.stemcr.2018.09.015
   Pfaff SL, 1996, CELL
   Philippidou P, 2013, NEURON, V80, P12, DOI 10.1016/j.neuron.2013.09.020
   Picelli S, 2014, NAT PROTOC, V9, P171, DOI 10.1038/nprot.2014.006
   R Development Core Team, 2014, R LANG ENV STAT COMP
   Raivich G, 2008, J NEUROCHEM, V107, P898, DOI 10.1111/j.1471-4159.2008.05684.x
   Rajan P, 1998, J NEUROSCI, V18, P3620
   Ramskold D, 2012, NAT BIOTECHNOL, V30, P777, DOI 10.1038/nbt.2282
   Regan MR, 2007, J NEUROSCI, V27, P6607, DOI 10.1523/JNEUROSCI.0790-07.2007
   Rowitch DH, 2010, NATURE, V468, P214, DOI 10.1038/nature09611
   Roybon L, 2013, CELL REP
   Roybon L, 2013, CELL REP, V4, P1035, DOI 10.1016/j.celrep.2013.06.021
   Schreck I, 2011, BIOCHEM BIOPH RES CO, V407, P735, DOI 10.1016/j.bbrc.2011.03.092
   Skiljan I., IRFANVIEW OFFICIAL H
   Sloan SA, 2017, NEURON, V95, P779, DOI 10.1016/j.neuron.2017.07.035
   Sofroniew MV, 2020, TRENDS IMMUNOL, V41, P758, DOI 10.1016/j.it.2020.07.004
   Sofroniew MV, 2015, NAT REV NEUROSCI, V16, P249, DOI 10.1038/nrn3898
   Stahel PF, 1997, MOL BRAIN RES, V50, P205, DOI 10.1016/S0169-328X(97)00189-7
   Subramanian A, 2005, P NATL ACAD SCI USA, V102, P15545, DOI 10.1073/pnas.0506580102
   Thams S, 2019, MOL THER, V27, P87, DOI 10.1016/j.ymthe.2018.10.010
   Thelin EP, 2018, J NEUROTRAUM, V35, P341, DOI 10.1089/neu.2017.5155
   Thelin EP, 2019, J NEUROTRAUM, DOI 10.1089/neu.2019.6480
   Tsai HH, 2012, SCIENCE, V337, P358, DOI 10.1126/science.1222381
   Weber A, 2010, SCI SIGNAL, V3, DOI 10.1126/scisignal.3105cm1
   WEBSTER RO, 1980, IMMUNOPHARMACOLOGY, V2, P201, DOI 10.1016/0162-3109(80)90050-8
   Werner C, 2007, BRIT J ANAESTH, V99, P4, DOI 10.1093/bja/aem131
   Westergard T, 2020, NEUROCHEM RES, V45, P1298, DOI 10.1007/s11064-020-02959-7
   Wichterle H, 2002, CELL, V110, P385, DOI 10.1016/S0092-8674(02)00835-8
   Wickham H., 2019, J OPEN SOURCE SOFTW, V4, P1686, DOI DOI 10.21105/JOSS.01686
   Wilke CO., 2020, PACKAGECOWPLOTSTREAM, DOI DOI 10.1038/S41598-021-83135-X
   Witten D., 2019, POICLACLU CLASSIFICA
   Woodcock T, 2013, FRONT NEUROL, V4, DOI 10.3389/fneur.2013.00018
   Yang SH, 2013, SHOCK, V40, P471, DOI 10.1097/SHK.0000000000000037
   Ye ZC, 1998, GLIA, V22, P237, DOI 10.1002/(SICI)1098-1136(199803)22:3<237::AID-GLIA3>3.0.CO;2-2
   Zachariadis V, 2020, DIRECT NUCL TAGMENTA
   Zachariadis V, 2020, MOL CELL, V80, P541, DOI 10.1016/j.molcel.2020.09.025
   Zamanian JL, 2012, J NEUROSCI, V32, P6391, DOI 10.1523/JNEUROSCI.6221-11.2012
   Zeke A, 2016, MICROBIOL MOL BIOL R, V80, P793, DOI 10.1128/MMBR.00043-14
   Zhang PS, 1996, J NEUROSCI RES, V46, P114, DOI 10.1002/(SICI)1097-4547(19961001)46:1<114::AID-JNR14>3.0.CO;2-5
   Zhang Y, 2016, NEURON, V89, P37, DOI 10.1016/j.neuron.2015.11.013
   Zhang Y, 2010, CURR OPIN NEUROBIOL, V20, P588, DOI 10.1016/j.conb.2010.06.005
   Zhu AQ, 2019, BIOINFORMATICS, V35, P2084, DOI 10.1093/bioinformatics/bty895
   Zhu YY, 2001, BIOTECHNIQUES, V30, P892, DOI 10.2144/01304pf02
   Zuchero JB, 2015, DEVELOPMENT, V142, P3805, DOI 10.1242/dev.129304
NR 127
TC 0
Z9 0
U1 2
U2 2
PU BMC
PI LONDON
PA CAMPUS, 4 CRINAN ST, LONDON N1 9XW, ENGLAND
SN 1476-9255
J9 J INFLAMM-LOND
JI J. Inflamm.-Lond.
PD JUN 27
PY 2023
VL 20
IS 1
AR 22
DI 10.1186/s12950-023-00349-8
PG 18
WC Immunology
WE Science Citation Index Expanded (SCI-EXPANDED)
SC Immunology
GA L1RN7
UT WOS:001021099900001
PM 37370141
OA Green Published, gold
DA 2023-08-27
ER

PT J
AU Baxter, PA
   Su, JM
   Onar-Thomas, A
   Billups, CA
   Li, XN
   Poussaint, TY
   Smith, ER
   Thompson, P
   Adesina, A
   Ansell, P
   Giranda, V
   Paulino, A
   Kilburn, L
   Quaddoumi, I
   Broniscer, A
   Blaney, SM
   Dunkel, IJ
   Fouladi, M
AF Baxter, Patricia A.
   Su, Jack M.
   Onar-Thomas, Arzu
   Billups, Catherine A.
   Li, Xiao-Nan
   Poussaint, Tina Young
   Smith, Edward R.
   Thompson, Patrick
   Adesina, Adekunle
   Ansell, Pete
   Giranda, Vincent
   Paulino, Arnold
   Kilburn, Lindsey
   Quaddoumi, Ibrahim
   Broniscer, Alberto
   Blaney, Susan M.
   Dunkel, Ira J.
   Fouladi, Maryam
TI A phase I/II study of veliparib (ABT-888) with radiation and
   temozolomide in newly diagnosed diffuse pontine glioma: a Pediatric
   Brain Tumor Consortium study
SO NEURO-ONCOLOGY
LA English
DT Article
DE ABT-888; CNS tumors; DIPG; PARP inhibition; veliparib
ID ADP-RIBOSE POLYMERASE; HIGH-GRADE GLIOMA; POLY(ADP-RIBOSE) POLYMERASE;
   CLINICAL-TRIALS; INHIBITION; MEDULLOBLASTOMA; LETHALITY; CHILDREN;
   THERAPY; METRICS
AB Background. A Pediatric Brain Tumor Consortium (PBTC) phase I/II trial of veliparib and radiation followed by veliparib and temozolomide (TMZ) was conducted in children with newly diagnosed diffuse intrinsic pontine glioma (DIPG).The objectives were to: (i) estimate the recommended phase II dose (RP2D) of veliparib with concurrent radiation; (ii) evaluate the pharmacokinetic parameters of veliparib during radiation; (iii) evaluate feasibility of intrapatient TMZ dose escalation; (iv) describe toxicities of protocol therapy; and (v) estimate the overall survival distribution compared with historical series.
   Methods. Veliparib was given Monday through Friday b.i.d. during radiation followed by a 4-week rest. Patients then received veliparib at 25 mg/m(2) b.i.d. and TMZ 135 mg/m 2 daily for 5 days every 28 days. Intrapatient dose escalation of TMZ was investigated for patients experiencing minimal toxicity.
   Results. Sixty-six patients (65 eligible) were enrolled.The RP2D of veliparib was 65 mg/m 2 b.i.d. with radiation. Dose-limiting toxicities during radiation with veliparib therapy included: grade 2 intratumoral hemorrhage (n = 1), grade 3 maculopapular rash (n = 2), and grade 3 nervous system disorder (generalized neurologic deterioration) (n =1). Intrapatient TMZ dose escalation during maintenance was not tolerated. Following a planned interim analysis, it was concluded that this treatment did not show a survival benefit compared with PBTC historical controls, and accrual was stopped for futility.The 1- and 2-year overall survival rates were 37.2% (SE 7%) and 5.3% (SE 3%), respectively.
   Conclusion. Addition of veliparib to radiation followed by TMZ and veliparib was tolerated but did not improve survival for patients with newly diagnosed DIPG.
C1 [Baxter, Patricia A.; Su, Jack M.; Li, Xiao-Nan; Adesina, Adekunle; Blaney, Susan M.] Texas Childrens Hosp, Baylor Coll Med, Houston, TX 77030 USA.
   [Onar-Thomas, Arzu; Billups, Catherine A.; Quaddoumi, Ibrahim; Broniscer, Alberto] St Jude Childrens Res Hosp, 332 N Lauderdale St, Memphis, TN 38105 USA.
   [Poussaint, Tina Young; Smith, Edward R.] Boston Childrens Hosp, Boston, MA USA.
   [Thompson, Patrick] Univ N Carolina, Childrens Hosp, Chapel Hill, NC 27515 USA.
   [Ansell, Pete; Giranda, Vincent] AbbVie Labs, Abbvie Pk, IL USA.
   [Paulino, Arnold] Univ Texas MD Anderson Canc Ctr, Houston, TX 77030 USA.
   [Kilburn, Lindsey] Childrens Natl Med Ctr, Washington, DC 20010 USA.
   [Dunkel, Ira J.] Mem Sloan Kettering Canc Ctr, 1275 York Ave, New York, NY 10021 USA.
   [Fouladi, Maryam] Cincinnati Childrens Hosp, Cincinnati, OH USA.
C3 Baylor College of Medicine; St Jude Children's Research Hospital;
   Harvard University; Boston Children's Hospital; University of North
   Carolina; University of North Carolina Chapel Hill; AbbVie; University
   of Texas System; UTMD Anderson Cancer Center; Children's National Health
   System; Memorial Sloan Kettering Cancer Center; Cincinnati Children's
   Hospital Medical Center
RP Baxter, PA (通讯作者)，1102 Bates St,Suite 1590, Houston, TX 77030 USA.
EM
OI Smith, Edward/0000-0002-6317-8450; Kilburn, Lindsay/0000-0003-1478-3619
FU National Cancer Institute CTEP PBTC U01 grant [2UM1CA081457 [UM1]];
   American Lebanese Syrian Associated Charities; NIH/NCI Cancer Center
   Support Grant [P30 CA008748]
FX The trial was funded by a National Cancer Institute CTEP PBTC U01 grant
   (2UM1CA081457 [UM1]) and the American Lebanese Syrian Associated
   Charities. This research was funded in part through the NIH/NCI Cancer
   Center Support Grant P30 CA008748.
CR Akino T, 2014, CANCER RES, V74, P3716, DOI 10.1158/0008-5472.CAN-13-3116
   Albert JM, 2007, CLIN CANCER RES, V13, P3033, DOI 10.1158/1078-0432.CCR-06-2872
   Barazzuol L, 2013, RADIAT ONCOL, V8, DOI 10.1186/1748-717X-8-65
   Castel D, 2015, ACTA NEUROPATHOL, V130, P815, DOI 10.1007/s00401-015-1478-0
   Chalmers AJ, 2010, SEMIN RADIAT ONCOL, V20, P274, DOI 10.1016/j.semradonc.2010.06.001
   Chornenkyy Y, 2015, MOL CANCER THER, V14, P2560, DOI 10.1158/1535-7163.MCT-15-0282
   Cohen KJ, 2011, NEURO-ONCOLOGY, V13, P410, DOI 10.1093/neuonc/noq205
   Conway AE, 2014, NEURORADIOLOGY, V56, P405, DOI 10.1007/s00234-014-1348-9
   Donawho CK, 2007, CLIN CANCER RES, V13, P2728, DOI 10.1158/1078-0432.CCR-06-3039
   Fehnel KP, 2016, J NEUROSURG-PEDIATR, V18, P396, DOI 10.3171/2015.12.PEDS15448
   Gupta N, 2018, NEURO-ONCOLOGY, V20, P1547, DOI 10.1093/neuonc/noy070
   Gupta SK, 2016, JNCI-J NATL CANCER I, V108, DOI 10.1093/jnci/djv369
   Han HS, 2018, ANN ONCOL, V29, P154, DOI 10.1093/annonc/mdx505
   Hipp SJ, 2011, NEURO-ONCOLOGY, V13, P904, DOI 10.1093/neuonc/nor076
