## Supplemental Material 1 for "Bibliometric analysis of emerging trends and research foci in brainstem tumor field over thirty years (1992-2023)": savedrecs (3).txt

FN Clarivate Analytics Web of Science
VR 1.0
PT J
AU Kumar, A
   Sardhara, J
   Verma, PK
   Bhaisora, KS
   Srivastav, AK
   Jaiswal, A
   Behari, S
   Kumar, R
AF Kumar, Ashutosh
   Sardhara, Jayesh
   Verma, Pawan Kumar
   Bhaisora, Kamlesh Singh
   Srivastav, Arun Kumar
   Jaiswal, Awadhesh
   Behari, Sanjay
   Kumar, Raj
TI Posterior Fossa Midline Epidermoid Tumors: Role of the Inferior
   Medullary Velum and a Proposed New Imaging Classification
SO NEUROSURGERY
LA English
DT Article
DE Fourth ventricle; Cisterna magna; Epidermoid tumor; Inferior medullary
   velum
ID CYSTS; VENTRICLE
AB BACKGROUND: Posterior fossa midline epidermoid tumors (PFMETs) include the epidermoid tumors of the cisterna magna (CM) and fourth ventricle (FV).
   OBJECTIVE: To report tumor epicenter-based classification of PFMETs and its clinical and surgical implications with outcome.
   METHODS: On retrospective analysis of operated cases of intracranial epidermoid tumors, 19 (N = 19) patients having tumor epicenter within FV, CM, or both were included. Cerebellopontine and prepontine cistern epidermoid were excluded. Tumor location was decided based on preoperative MRI and intraoperative findings. Major complication was defined as new onset or worsening of cranial nerve (CN) deficit, sensory motor impairment, or tracheostomy.
   RESULTS: The mean (+/- SD) age of the patients was 42.0 +/- 11.6 years (range 25-61 years), with no sex predilection (male: female: 1:0.9). The most common symptoms were cerebellar dysfunction, headache, vomiting, and diplopia. Common CNs affected were VII, V, lower cranial nerve, and VI. The PFMETs were classified based on tumor epicenter as type 1 (tumor epicenter in CM, n = 4/21.1%), type 2 (FV, n = 5/26.3%), and type 3 (involved CM and FV, n = 10/52.6%). Type 2 tumors had a higher incidence of raised intracranial pressure and only facial nerve palsy as preoperative CN deficit. Type 1 tumors had the least incidence of postoperative major complications. Type 3 tumors were the largest and had a greater incidence of brainstem adhesion and postoperative complications. The tumor size, duration of symptoms, and patient age were higher in patients with brainstem adhesion (5.3 +/- 1.0 cm, 21 +/- 16 months, 44.1 +/- 9.2 years) as against its absence (4.8 +/- 1.3 cm, 11.2 +/- 7.3 months, 38.2 +/- 11.7 years). Inferior medullary velum and tela choroidea have a critical role in tumorogenesis, tumor extension, and brainstem adhesion.
   CONCLUSION: PFMETs can be classified into 3 subtypes based on tumor epicenter having clinical and surgical implications. Less aggressive dissection and near total excision in the presence of brainstemadhesion yield favorable outcomes.
C1 [Kumar, Ashutosh; Sardhara, Jayesh; Verma, Pawan Kumar; Bhaisora, Kamlesh Singh; Srivastav, Arun Kumar; Jaiswal, Awadhesh; Behari, Sanjay; Kumar, Raj] Sanjay Gandhi Post Grad Inst Med Sci, Dept Neurosurg, Rae Barely Rd, Lucknow, India.
C3 Sanjay Gandhi Postgraduate Institute of Medical Sciences
RP Kumar, A (通讯作者)，Sanjay Gandhi Post Grad Inst Med Sci, Dept Neurosurg, Rae Barely Rd, Lucknow, India.
EM;
  ;
  ;
  
CR Bini W., 1993, NEUROCIRUGIA, V4, P205
   Carmel P., 1990, BRAIN TUMORS DISORDE
   Chung LK, 2017, WORLD NEUROSURG, V105, P689, DOI 10.1016/j.wneu.2017.06.037
   Cobbs C S, 1997, Clin Neurosurg, V44, P511
   DAVID M, 1965, NEURO-CHIR, V11, P363
   FIUME D, 1988, SURG NEUROL, V29, P178, DOI 10.1016/0090-3019(88)90002-X
   Forghani R, 2007, RADIOGRAPHICS, V27, DOI 10.1148/rg.275075011
   Gopalakrishnan CV, 2014, CLIN NEUROL NEUROSUR, V117, P93, DOI 10.1016/j.clineuro.2013.11.025
   Kumar A, 2021, J NEUROSCI RURAL PRA, V12, P571, DOI 10.1055/s-0041-1730125
   Kumar Sanjeev, 2021, Asian J Neurosurg, V16, P470, DOI 10.4103/ajns.AJNS_539_20
   Li XL., 2014, NEUROSURG QUART, V24, P73
   Liu M, 2006, J CLIN NEUROSCI, V13, P428, DOI 10.1016/j.jocn.2005.04.022
   NASSAR SI, 1995, SURG NEUROL, V43, P246, DOI 10.1016/0090-3019(95)80006-3
   Rhoton AL, 2000, NEUROSURGERY, V47, pS7, DOI 10.1097/00006123-200009001-00007
   RUBIN G, 1989, ACTA NEUROCHIR, V97, P1, DOI 10.1007/BF01577734
   SCOTT M, 1974, J NEUROL NEUROSUR PS, V37, P1329, DOI 10.1136/jnnp.37.12.1329
   Tancredi A, 2003, ACTA NEUROCHIR, V145, P905, DOI 10.1007/s00701-003-0008-3
   TOGLIA JU, 1965, J NEUROSURG, V23, P384, DOI 10.3171/jns.1965.23.4.0384
   ULRICH J, 1964, J NEUROSURG, V21, P1051, DOI 10.3171/jns.1964.21.12.1051
   YASARGIL MG, 1989, NEUROSURGERY, V24, P561
NR 20
TC 0
Z9 0
U1 0
U2 0
PU LIPPINCOTT WILLIAMS & WILKINS
PI PHILADELPHIA
PA TWO COMMERCE SQ, 2001 MARKET ST, PHILADELPHIA, PA 19103 USA
SN 0148-396X
EI 1524-4040
J9 NEUROSURGERY
JI Neurosurgery
PD JUL
PY 2023
VL 93
IS 1
BP 112
EP 119
DI 10.1227/neu.0000000000002383
PG 8
WC Clinical Neurology; Surgery
WE Science Citation Index Expanded (SCI-EXPANDED)
SC Neurosciences & Neurology; Surgery
GA J6JM3
UT WOS:001010656700023
PM 36735515
DA 2023-08-27
ER

PT J
AU Allen, JC
AF Allen, JC
TI Initial management of children with hypothalamic and thalamic tumors and
   the modifying role of neurofibromatosis-1
SO PEDIATRIC NEUROSURGERY
LA English
DT Article
DE optic pathway glioma; diencephalic tumor; thalamic tumor;
   neurofibromatosis
ID LOW-GRADE GLIOMAS; DIENCEPHALIC SYNDROME; OPTIC PATHWAY; CHILDHOOD;
   ASTROCYTOMA; VINCRISTINE; DISSEMINATION; CHEMOTHERAPY; CARBOPLATIN;
   CHIASM
AB Diencephalic gliomas may be grouped into 2 clinical categories. Optic pathway/hypothalamus gliomas (OPG) arise primarily from a slower-growing juvenile pilocytic astrocytoma, and thalamic gliomas arise primarily from a fibrillary astrocytoma which can become clinically and histologically more aggressive. Children with OPG have an excellent long-term prognosis with a 10-year survival of over 85%, The major therapeutic challenge for these patients is to maximize their quality of life by preserving visual and endocrine function while minimizing treatment-related morbidity. Treatment is often initiated at diagnosis in infants and toddlers who have a major visual impairment or the diencephalic syndrome. The judicious application of chemotherapy may serve to forestall the need for radiotherapy or surgery. Children with neurofibromatosis-1 (NF-1) usually have a more indolent course. Tumors may grow more slowly or occasionally regress spontaneously. However, over 90% of children with OPG without NF-1 will require some form of therapy. Patients with thalamic gliomas present with a shorter history, often with hydrocephalus, Surgical intervention is often required to relieve intracranial pressure and establish the histologic identity of the tumor. Over 75% of these tumors will become locally aggressive. Current multimodality therapy is relatively ineffective. The bithalamic variant behaves similarly to a pontine glioma, Copyright (C) 2000 S.Karger AG, Basel.
C1 Beth Israel Med Ctr, Div Pediat Neurol, New York, NY 10128 USA.
C3 Harvard University; Beth Israel Deaconess Medical Center
RP Allen, JC (通讯作者)，Beth Israel Med Ctr, Div Pediat Neurol, 170 E End Ave, New York, NY 10128 USA.
OI Allen, Jeffrey/0000-0001-9957-6664
CR ALVORD EC, 1988, J NEUROSURG, V68, P85, DOI 10.3171/jns.1988.68.1.0085
   ARNOLDI K, 1995, J PEDIATR OPHTHALMOL, V32, P347
   ARNOLDI KA, 1995, J PEDIATR OPHTHALMOL, V32, P296
   BURR IM, 1976, J PEDIATR-US, V88, P439, DOI 10.1016/S0022-3476(76)80260-0
   CIVITELLO LA, 1988, NEUROLOGY, V38, P562, DOI 10.1212/WNL.38.4.562
   Deliganis AV, 1996, NEUROSURGERY, V38, P1114, DOI 10.1097/00006123-199606000-00010
   DELIGANIS AV, 1996, NEUROSURGERY, V38, P1118
   DESOUSA AL, 1979, NEUROSURGERY, V4, P207, DOI 10.1227/00006123-197903000-00002
   FINLAY JL, 1995, J CLIN ONCOL, V13, P112, DOI 10.1200/JCO.1995.13.1.112
   GAJJAR A, 1995, J NEUROSURG, V83, P67, DOI 10.3171/jns.1995.83.1.0067
   Gropman AL, 1998, CANCER-AM CANCER SOC, V83, P166, DOI 10.1002/(SICI)1097-0142(19980701)83:1<166::AID-CNCR22>3.0.CO;2-U
   Hjalmars U, 1999, CANCER, V85, P2077, DOI 10.1002/(SICI)1097-0142(19990501)85:9<2077::AID-CNCR27>3.0.CO;2-4
   HOUSEPIAN EM, 1993, J NEURO-ONCOL, V15, P51, DOI 10.1007/BF01050263
   JANSS AJ, 1995, CANCER-AM CANCER SOC, V75, P1051, DOI 10.1002/1097-0142(19950215)75:4<1051::AID-CNCR2820750423>3.0.CO;2-S
   LISTERNICK R, 1994, J PEDIATR-US, V125, P63, DOI 10.1016/S0022-3476(94)70122-9
   LISTERNICK R, 1989, Journal of Pediatrics, V114, P788, DOI 10.1016/S0022-3476(89)80137-4
   PACKER RJ, 1993, J CLIN ONCOL, V11, P850, DOI 10.1200/JCO.1993.11.5.850
   Packer RJ, 1997, J NEUROSURG, V86, P747, DOI 10.3171/jns.1997.86.5.0747
   Poussaint TY, 1997, AM J NEURORADIOL, V18, P1499
   Reardon DA, 1998, PEDIATR NEUROSURG, V29, P29, DOI 10.1159/000028681
   STILLER CA, 1994, INT J EPIDEMIOL, V23, P458, DOI 10.1093/ije/23.3.458
   Tao ML, 1997, INT J RADIAT ONCOL, V39, P579, DOI 10.1016/S0360-3016(97)00359-3
   Wisoff J H, 1992, Neurosurg Clin N Am, V3, P791
NR 23
TC 45
Z9 46
U1 0
U2 3
PU KARGER
PI BASEL
PA ALLSCHWILERSTRASSE 10, CH-4009 BASEL, SWITZERLAND
SN 1016-2291
J9 PEDIATR NEUROSURG
JI Pediatr. Neurosurg.
PD MAR
PY 2000
VL 32
IS 3
BP 154
EP 162
DI 10.1159/000028922
PG 9
WC Clinical Neurology; Pediatrics; Surgery
WE Science Citation Index Expanded (SCI-EXPANDED)
SC Neurosciences & Neurology; Pediatrics; Surgery
GA 327ZK
UT WOS:000087824200010
PM 10867564
DA 2023-08-27
ER

PT J
AU Yao, K
   Duan, ZJ
   Wang, Y
   Zhang, MS
   Fan, T
   Wu, B
   Qi, XL
AF Yao, Kun
   Duan, Zejun
   Wang, Yin
   Zhang, Mingshan
   Fan, Tao
   Wu, Bin
   Qi, Xueling
TI Detection of H3K27M mutation in cases of brain stem subependymoma
SO HUMAN PATHOLOGY
LA English
DT Article
DE H3K27M mutation; Subependymoma; Brain stem neoplasm; Sanger sequencing
ID H3F3A; FEATURES; TUMOR; DNA
AB Subependymomas are rare, slow-growing, grade I glial tumors of the central nervous system. Recently, diffuse midline gliomas with mutations in the H3.1 or H3.3 genes at the position of amino acid 27, resulting in the replacement of lysine by methionine (K27M), were defined as the new grade IV entity. As H3K27M mutations have been reported in midline gliomas, gangliogliomas, and pilocytic astrocytomas, whether they occur in midline subependymomas has been unclear. We determined whether any such mutations can be found in them and analyzed the prognostic relevance of any such mutations in subependymomas. Four subependymomas, all in the brain stem, harbored H3K27M mutations. No such mutation was found in any of the subependymomas from other locations. The mutations were identified by immunohistochemical stains and confirmed with Sanger sequencing. The median follow-up of the patients with the mutations in their tumors was 3.2 years, and 3 are still alive, having received no adjuvant therapy. We demonstrate that H3K27M mutation can occur in brainstem subependymomas; despite the presence of H3K27M mutation, these cases should not be diagnosed or treated as grade IV tumors because they showed a better outcome than the outcome of diffuse midline H3K27M mutant glioma. Our conclusion is not only that brainstem subependymomas can have H3K27M mutations but that they do not carry the rapidly lethal prognosis with which these mutations are usually associated because of their discovery in diffuse intrinsic pontine gliomas. (C) 2018 Elsevier Inc. All rights reserved.
C1 [Yao, Kun; Duan, Zejun; Qi, Xueling] Capital Med Univ, Dept Pathol, San Bo Brain Hosp, Beijing, Peoples R China.
   [Wang, Yin] Fudan Univ, Dept Pathol, Huashan Hosp, Shanghai Med Coll, Shanghai 200040, Peoples R China.
   [Zhang, Mingshan; Fan, Tao; Wu, Bin] Capital Med Univ, Dept Neurosurg, San Bo Brain Hosp, Beijing, Peoples R China.
C3 Capital Medical University; Fudan University; Capital Medical University
RP Qi, XL (通讯作者)，Capital Med Univ, Dept Pathol, San Bo Brain Hosp, Beijing, Peoples R China.
EM;
  ;
  
CR Bender S, 2013, CANCER CELL, V24, P660, DOI 10.1016/j.ccr.2013.10.006
   Bernal Garcia LM, 2015, NEUROSURGERY S2, pE364
   Bi ZY, 2015, J NEUROSURG, V122, P49, DOI 10.3171/2014.9.JNS14155
   Chan KM, 2013, GENE DEV, V27, P985, DOI 10.1101/gad.217778.113
   Cure LM, 2014, SPINE J, V14, pE9, DOI 10.1016/j.spinee.2013.10.056
   Gessi M, 2016, ACTA NEUROPATHOL, V132, P635, DOI 10.1007/s00401-016-1608-3
   Hochart A, 2015, ANN CLIN TRANSL NEUR, V2, P439, DOI 10.1002/acn3.184
   Hou ZG, 2013, J CLIN NEUROSCI, V20, P84, DOI 10.1016/j.jocn.2012.05.026
   Huang TY, 2017, ACTA NEUROPATHOL COM, V5, DOI 10.1186/s40478-017-0436-6
   Kong LY, 2014, J NEUROIMMUNOL, V277, P168, DOI 10.1016/j.jneuroim.2014.10.008
   Lewis PW, 2013, SCIENCE, V340, P857, DOI 10.1126/science.1232245
   Nguyen AT, 2015, NEUROPATH APPL NEURO, V41, P403, DOI 10.1111/nan.12196
   Orillac C, 2016, ACTA NEUROPATHOL COM, V4, DOI 10.1186/s40478-016-0361-0
   Pages M, 2018, BRAIN PATHOL, V28, P103, DOI 10.1111/bpa.12473
   Rushing EJ, 2007, J NEURO-ONCOL, V85, P297, DOI 10.1007/s11060-007-9411-6
   SCHEITHAUER BW, 1978, J NEUROSURG, V49, P689, DOI 10.3171/jns.1978.49.5.0689
   Schwartzentruber J, 2012, NATURE, V484, P130, DOI 10.1038/nature11026
   Sturm D, 2012, CANCER CELL, V22, P425, DOI 10.1016/j.ccr.2012.08.024
   Venneti S, 2013, BRAIN PATHOL, V23, P558, DOI 10.1111/bpa.12042
   Wiestler OD., 2016, WHO CLASSIFICATION T, P102
NR 20
TC 14
Z9 15
U1 0
U2 3
PU W B SAUNDERS CO-ELSEVIER INC
PI PHILADELPHIA
PA 1600 JOHN F KENNEDY BOULEVARD, STE 1800, PHILADELPHIA, PA 19103-2899 USA
SN 0046-8177
EI 1532-8392
J9 HUM PATHOL
JI Hum. Pathol.
PD FEB
PY 2019
VL 84
BP 262
EP 269
DI 10.1016/j.humpath.2018.10.011
PG 8
WC Pathology
WE Science Citation Index Expanded (SCI-EXPANDED)
SC Pathology
GA HP5NZ
UT WOS:000461725100030
PM 30389438
DA 2023-08-27
ER

PT J
AU Cheng, X
   Gao, PY
AF Cheng, Xu
   Gao, Pei-Yi
TI Abnormal Neural Activity in Children With Diffuse Intrinsic Pontine
   Glioma Had Manifested Deficit in Behavioral Inhibition: A Resting-State
   Functional MRI Study
SO JOURNAL OF COMPUTER ASSISTED TOMOGRAPHY
LA English
DT Article
DE diffuse intrinsic pontine glioma; behavior control; resting state;
   functional magnetic resonance imaging; ALFF
ID ATTENTION-DEFICIT/HYPERACTIVITY DISORDER; CEREBRAL-BLOOD-FLOW;
   BRAIN-STEM TUMORS; HYPERACTIVITY DISORDER; EXECUTIVE FUNCTIONS;
   METHYLPHENIDATE; CONNECTIVITY; METAANALYSIS; DYSFUNCTION; CORTEX
AB Purpose The purpose of this study was to investigate whether alterations of regional neural function in children with diffuse intrinsic pontine glioma (DIPG) had manifested deficit in behavioral inhibition using resting-state functional MRI (rs-fMRI). Methods There were 17 participants with DIPG who took part in the study. Eight children were with deficit in behavioral inhibition, whereas the other 9 children did not obtain deficit in behavioral inhibition. Five healthy children with age, sex, and education matched to the study group also participated as the control group. These 3 groups underwent rs-fMRI, and the results were then converted to amplitude of low-frequency fluctuation (ALFF) data. Amplitude of low-frequency fluctuation data were further analyzed by single-factor analysis of variance comparing among 3 groups based on the whole brain levels. Amplitude of low-frequency fluctuation results were subjected to t test of voxel-wised comparison to derive the rs-fMRI brain function differences between the 2 DIPG groups. The Pearson correlation between ALFF values of abnormal regions found in 3 groups and the scores obtained according to the Child Behavior Checklist were analyzed. Results The 3 groups had shown significant differences in terms of the ALFF results, with the ALFF increased in several brain regions (P < 0.05, corrected with AlphaSim, clusters >59 voxels), which include left supramarginal gyrus, left dorsolateral superior frontal gyrus, right precentral gyrus, and right middle frontal gyrus. Participants with deficit in behavioral inhibition had shown significant differences (ALFF decreased) in several brain regions, including left dorsolateral superior frontal gyrus and right fusiform gyrus (P < 0.05, corrected with AlphaSim, clusters >123 voxels), whereas other brain regions had shown ALFF increased, including left supramarginal gyrus, left middle frontal gyrus, and right medial superior frontal gyrus (P < 0.05, corrected with AlphaSim, clusters >123 voxels). There was no significant correlation between ALFF values and Child Behavior Checklist scores (P > 0.05). Conclusions These findings of focal spontaneous hyperfunction and hypofunction, which correlate with deficit in behavioral inhibition processing, and the abnormal brain regions are considered to be inefficient (in regions of the brain that may relate to compensatory brain and behavioral functioning, and it may be that the brain region needs to exert extra energy to perform a task to the same degree as the control group) or inability (inability in a certain region, or underpowered), pointing to a pathophysiologic process in executive dysfunction.
C1 [Cheng, Xu] Capital Med Univ, Beijing Friendship Hosp, Dept Radiol, Beijing, Peoples R China.
   [Cheng, Xu; Gao, Pei-Yi] Capital Med Univ, Beijing Tiantan Hosp, Dept Radiol, Tiantan West 6, Beijing 100050, Peoples R China.
   [Gao, Pei-Yi] Beijing Neurosurg Inst, Beijing, Peoples R China.
   [Gao, Pei-Yi] Beijing Key Lab Magnet Resonance Imaging & Brain, Beijing, Peoples R China.
C3 Capital Medical University; Capital Medical University; Beijing
   Neurosurgical Institute
RP Gao, PY (通讯作者)，Capital Med Univ, Beijing Tiantan Hosp, Dept Radiol, Tiantan West 6, Beijing 100050, Peoples R China.
EM
FU National Natural Science Foundation of China [81361120402]
FX This work was supported by National Natural Science Foundation of China
   (no. 81361120402). The authors declare no conflict of interest.
CR Achenbach TM., 1991, INTEGRATIVE GUIDE 19
   Aron AR, 2004, TRENDS COGN SCI, V8, P170, DOI 10.1016/j.tics.2004.02.010
   Barkley RA, 1997, PSYCHOL BULL, V121, P65, DOI 10.1037/0033-2909.121.1.65
   BARKLEY RA, 1995, ATTENTION MEMORY EXE, P307
   BISWAL B, 1995, MAGNET RESON MED, V34, P537, DOI 10.1002/mrm.1910340409
   Bush G, 2011, BIOL PSYCHIAT, V69, P1160, DOI 10.1016/j.biopsych.2011.01.022
   Castellanos FX, 2012, TRENDS COGN SCI, V16, P17, DOI 10.1016/j.tics.2011.11.007
   Castellanos FX, 2002, JAMA-J AM MED ASSOC, V288, P1740, DOI 10.1001/jama.288.14.1740
   Castellanos FX, 1996, ARCH GEN PSYCHIAT, V53, P607
   EPSTEIN F, 1988, J NEURO-ONCOL, V6, P309
   Fassbender C, 2006, CLIN PSYCHOL REV, V26, P445, DOI 10.1016/j.cpr.2006.01.003
   Fox MD, 2007, NAT REV NEUROSCI, V8, P700, DOI 10.1038/nrn2201
   Friedman NP, 2006, PSYCHOL SCI, V17, P172, DOI 10.1111/j.1467-9280.2006.01681.x
   Frodl T, 2012, ACTA PSYCHIAT SCAND, V125, P114, DOI 10.1111/j.1600-0447.2011.01786.x
   Funahashi S, 2001, NEUROSCI RES, V39, P147, DOI 10.1016/S0168-0102(00)00224-8
   Harnishfeger Katherine Kipp, 1995, P175, DOI 10.1016/B978-012208930-5/50007-6
   Hill DE, 2003, NEUROPSYCHOLOGY, V17, P496, DOI 10.1037/0894-4105.17.3.496
   Houde O, 2010, DEVELOPMENTAL SCI, V13, P876, DOI 10.1111/j.1467-7687.2009.00938.x
   Karatekin C, 2000, PEDIATR NEUROL, V22, P106, DOI 10.1016/S0887-8994(99)00128-9
   Langleben DD, 2002, J NUCL MED, V43, P1624
   Lee JS, 2005, HUM BRAIN MAPP, V24, P157, DOI 10.1002/hbm.20067
   Li X, 2017, J HYPERTENS, V35, P1044, DOI 10.1097/HJH.0000000000001250
   Nigg JT, 2005, BIOL PSYCHIAT, V57, P1424, DOI 10.1016/j.biopsych.2004.11.011
   Otten ML, 2012, BRAIN, V135, P1017, DOI 10.1093/brain/aws041
   Pennington BF, 1996, J CHILD PSYCHOL PSYC, V37, P51, DOI 10.1111/j.1469-7610.1996.tb01380.x
   Potes C, 2014, NEUROIMAGE, V97, P188, DOI 10.1016/j.neuroimage.2014.04.045
   Recinos PF, 2007, PEDIATR NEUROSURG, V43, P192, DOI 10.1159/000098831
   Rubia K, 1999, AM J PSYCHIAT, V156, P891, DOI 10.1176/ajp.156.6.891
   Seidman LJ, 2005, BIOL PSYCHIAT, V57, P1263, DOI 10.1016/j.biopsych.2004.11.019
   Sergeant JA, 2002, BEHAV BRAIN RES, V130, P3, DOI 10.1016/S0166-4328(01)00430-2
   Soelva V, 2013, CHILD NERV SYST, V29, P597, DOI 10.1007/s00381-012-1973-8
   Sonuga-Barke EJS, 2008, CHILD ADOL PSYCH CL, V17, P367, DOI 10.1016/j.chc.2007.11.008
   Su TW, 2013, SCHIZOPHR RES, V148, P50, DOI 10.1016/j.schres.2013.05.005
   Vaidya CJ, 1998, P NATL ACAD SCI USA, V95, P14494, DOI 10.1073/pnas.95.24.14494
   Wang ZQ, 2011, HUM BRAIN MAPP, V32, P1720, DOI 10.1002/hbm.21140
   Zang YF, 2007, BRAIN DEV-JPN, V29, P83, DOI 10.1016/j.braindev.2006.07.002
NR 36
TC 4
Z9 4
U1 0
U2 14
PU LIPPINCOTT WILLIAMS & WILKINS
PI PHILADELPHIA
PA TWO COMMERCE SQ, 2001 MARKET ST, PHILADELPHIA, PA 19103 USA
SN 0363-8715
EI 1532-3145
J9 J COMPUT ASSIST TOMO
JI J. Comput. Assist. Tomogr.
PD JUL-AUG
PY 2019
VL 43
IS 4
BP 547
EP 552
DI 10.1097/RCT.0000000000000881
PG 6
WC Radiology, Nuclear Medicine & Medical Imaging
WE Science Citation Index Expanded (SCI-EXPANDED)
SC Radiology, Nuclear Medicine & Medical Imaging
GA IM0NH
UT WOS:000477684500004
PM 31162235
DA 2023-08-27
ER

PT J
AU GIRALDI, FP
   MIZOBUCHI, M
   HOROWITZ, ZD
   DOWNS, TR
   ALEPPO, G
   KIER, A
   WAGNER, T
   YUN, JS
   KOPCHICK, JJ
   FROHMAN, LA
AF GIRALDI, FP
   MIZOBUCHI, M
   HOROWITZ, ZD
   DOWNS, TR
   ALEPPO, G
   KIER, A
   WAGNER, T
   YUN, JS
   KOPCHICK, JJ
   FROHMAN, LA
TI DEVELOPMENT OF NEUROEPITHELIAL TUMORS OF THE ADRENAL-MEDULLA IN
   TRANSGENIC MICE EXPRESSING A MOUSE HYPOTHALAMIC GROWTH HORMONE-RELEASING
   HORMONE PROMOTER-SIMIAN VIRUS-40 T-ANTIGEN FUSION GENE
SO ENDOCRINOLOGY
LA English
DT Article
ID TISSUE-SPECIFIC EXPRESSION; NEURO-BLASTOMA CELLS; CHOROID-PLEXUS; LINES;
   DIFFERENTIATION; TUMORIGENESIS; NEUROBLASTOMA; PAPILLOMAS; PITUITARY;
   PRECURSOR
AB Expression of the mouse OH-releasing hormone (GRH) gene is restricted to neurons within the hypothalamus and to placenta. In an attempt to generate immortalized mouse hypothalamic neurons expressing GRH, the proximal 872-nucleotide segment of the 5'-flanking region of the hypothalamic mouse GRH gene was cloned by polymerase chain reaction and ligated to a 2.7-kilobase DNA sequence encoding the simian virus-40 (SV40) T-antigen, so that regulation of SV40 T-antigen expression was dependent on sequences within the mGRH 5'-flanking region. This region contains both TATA and CAAT boxes. The mouse GRH/SV40 T-antigen fusion gene was injected into 1-cell mouse embryos, and SV40 T-antigen incorporation in the mouse genome was found in 11 of 77 live births (3 males and 8 females). Although no evidence of hypothalamic tumors was found, all mice that expressed the transgene also developed tumors originating in the adrenal medulla. Gene copy number varied from 1-20 and was inversely proportional to survival, which ranged from 7-16 weeks. Corticosterone levels were normal. The male transgenic mice were fertile, and their progeny expressed the transgene and developed similar tumors. Microscopic examination of the tumors revealed a primitive neuroectodermal neoplasm that exhibited hematogenous and lymph node metastases and contained 100 ng norepinephrine, 2.85 ng epinephrine, and 1.1 ng dopamine/mg tumor tissue. Primary culture of dispersed tumor cells released norepinephrine into the medium (180 pg/ml.24 h). Cell lines from 2 tumors were established and exhibited characteristics similar to those of mixed neuroblastoma or primitive neuroectodermal tumors. In conclusion, the proximal 872 nucleotides of the hypothalamic mouse GRH promoter contain elements directing tissue-specific expression limited to early adrenal neuroectodermal cells. Other GRH DNA sequences appear to be required for restricted expression of mouse GRH within the hypothalamus.
C1 UNIV ILLINOIS,COLL MED,DEPT MED MC 787,CHICAGO,IL 60612.
   UNIV CINCINNATI,COLL MED,DEPT MED,DIV ENDOCRINOL,CINCINNATI,OH 45267.
   UNIV CINCINNATI,COLL MED,DEPT PATHOL,DIV COMPARAT PATHOL,CINCINNATI,OH 45267.
   OHIO UNIV,EDISON ANIM BIOTECHNOL CTR,ATHENS,OH 45701.
   UNIV ILLINOIS,COLL MED,DEPT MED,CHICAGO,IL 60612.
C3 University of Illinois System; University of Illinois Chicago;
   University of Illinois Chicago Hospital; University System of Ohio;
   University of Cincinnati; University System of Ohio; University of
   Cincinnati; University System of Ohio; Ohio University; University of
   Illinois System; University of Illinois Chicago; University of Illinois
   Chicago Hospital
RI Giraldi, Francesca Pecori/F-5081-2011
OI Giraldi, Francesca Pecori/0000-0001-6065-7803
FU NIDDK NIH HHS [DK-30667] Funding Source: Medline
CR BABINET C, 1989, GENOME, V31, P938, DOI 10.1139/g89-165
   BAETSCHER M, 1991, ONCOGENE, V6, P1133
   BEHRINGER RR, 1988, GENE DEV, V2, P453, DOI 10.1101/gad.2.4.453
   BOTTERI FM, 1987, MOL CELL BIOL, V7, P3178, DOI 10.1128/MCB.7.9.3178
   BRAR AK, 1989, ENDOCRINOLOGY, V125, P801, DOI 10.1210/endo-125-2-801
   BRINSTER RL, 1988, P NATL ACAD SCI USA, V85, P836, DOI 10.1073/pnas.85.3.836
   BRUBAKER PL, 1992, J BIOL CHEM, V267, P20728
   CHEN JD, 1991, MOL CELL BIOL, V11, P5968, DOI 10.1128/MCB.11.12.5968
   CHO HJ, 1989, J NEUROSCI RES, V24, P115, DOI 10.1002/jnr.490240116
   CHOI OR, 1986, NATURE, V323, P731, DOI 10.1038/323731a0
   CHOMCZYNSKI P, 1987, ANAL BIOCHEM, V162, P156, DOI 10.1016/0003-2697(87)90021-2
   CICCARONE V, 1989, CANCER RES, V49, P219
   COOPER MJ, 1990, CELL GROWTH DIFFER, V1, P149
   EFRAT S, 1989, CURR TOP MICROBIOL, V144, P89
   FROHMAN MA, 1989, MOL ENDOCRINOL, V3, P1529, DOI 10.1210/mend-3-10-1529
   GUILLEMIN R, 1982, SCIENCE, V218, P585, DOI 10.1126/science.6812220
   HANAHAN D, 1985, NATURE, V315, P115, DOI 10.1038/315115a0
   HJEMDAHL P, 1979, LIFE SCI, V25, P131, DOI 10.1016/0024-3205(79)90384-9
   LEDENT C, 1991, ENDOCRINOLOGY, V129, P1391, DOI 10.1210/endo-129-3-1391
   LING N, 1984, P NATL ACAD SCI-BIOL, V81, P4302, DOI 10.1073/pnas.81.14.4302
   MARKS JR, 1989, J VIROL, V63, P790, DOI 10.1128/JVI.63.2.790-797.1989
   MAYO KE, 1985, NATURE, V314, P464, DOI 10.1038/314464a0
   MAYO KE, 1983, NATURE, V306, P86, DOI 10.1038/306086a0
   MCGRANE MM, 1988, J BIOL CHEM, V263, P11443
   MELLON PL, 1990, NEURON, V5, P1, DOI 10.1016/0896-6273(90)90028-E
   MIZOBUCHI M, 1993, BIOTECHNIQUES, V15, P215
   OBRIEN JM, 1990, ARCH OPHTHALMOL-CHIC, V108, P1145, DOI 10.1001/archopht.1990.01070100101043
   PAHLMAN S, 1984, CELL DIFFER DEV, V14, P135, DOI 10.1016/0045-6039(84)90038-1
   RAY SK, 1974, ANAL LETT, V7, P109
   REYNOLDS RK, 1988, P NATL ACAD SCI USA, V5, P3135
   RINDI G, 1988, VIRCHOWS ARCH A, V412, P255, DOI 10.1007/BF00737150
   RIVIER J, 1982, NATURE, V300, P276, DOI 10.1038/300276a0
   RORKE LB, 1985, CANCER-AM CANCER SOC, V56, P1869, DOI 10.1002/1097-0142(19851001)56:7+<1869::AID-CNCR2820561330>3.0.CO;2-0
   SIDELL N, 1983, EXP CELL RES, V148, P21, DOI 10.1016/0014-4827(83)90184-2
   SKALNIK DG, 1991, MOL CELL BIOL, V11, P4518, DOI 10.1128/MCB.11.9.4518
   SUHR ST, 1989, MOL ENDOCRINOL, V3, P1693, DOI 10.1210/mend-3-11-1693
   WAGNER TE, 1981, P NATL ACAD SCI-BIOL, V78, P6376, DOI 10.1073/pnas.78.10.6376
   WINDLE JJ, 1990, MOL ENDOCRINOL, V4, P597, DOI 10.1210/mend-4-4-597
   YAMADA H, 1989, BIOMED RES, V3, P127
   ZAFAR MS, 1979, J CLIN ENDOCR METAB, V48, P66
NR 40
TC 12
Z9 12
U1 0
U2 0
PU ENDOCRINE SOC
PI BETHESDA
PA 4350 EAST WEST HIGHWAY SUITE 500, BETHESDA, MD 20814-4110
SN 0013-7227
J9 ENDOCRINOLOGY
JI Endocrinology
PD MAR
PY 1994
VL 134
IS 3
BP 1219
EP 1224
DI 10.1210/en.134.3.1219
PG 6
WC Endocrinology & Metabolism
WE Science Citation Index Expanded (SCI-EXPANDED)
SC Endocrinology & Metabolism
GA MZ443
UT WOS:A1994MZ44300032
PM 8119162
OA Bronze
DA 2023-08-27
ER

PT J
AU Cairns, LA
   Crotta, S
   Minuzzo, M
   RicciardiCastagnoli, P
   Pozzi, L
   Ottolenghi, S
AF Cairns, LA
   Crotta, S
   Minuzzo, M
   RicciardiCastagnoli, P
   Pozzi, L
   Ottolenghi, S
TI Immortalization of neuro-endocrine cells from adrenal tumors arising in
   SV40 T-transgenic mice
SO ONCOGENE
LA English
DT Article
DE pheochromocytoma; transgenic mice; adrenal medullary tumor; ret
ID INTENSELY FLUORESCENT CELLS; NERVE GROWTH-FACTOR; RET PROTOONCOGENE;
   CHROMAFFIN CELLS; CREST DERIVATIVES; EXPRESSION; GENE; DIFFERENTIATION;
   RAT; GLUCOCORTICOIDS
AB Pheochromocytomas are adrenal medullary tumors which arise from the transformation of neural crest-derived cells. In the course of studies of mice transgenic for an SV40 T-gene ectopically expressed in the adrenal medulla, we observed the occurrence of large, mainly bilateral tumors in a high proportion of transgenic animals. From these tumors we established immortalized cell lines which grow in vitro at 32 degrees C (the permissive temperature for the tsA58 T-protein encoded by the transgene), but not at 38 degrees C. These cells demonstrate characteristics of both neuronal (160 kd neurofilament) and endocrine (chromogranins) cells. The expression of Mash-1 and ret supports their initial characterization as early bipotential neuro-endocrine progenitors.
C1 CNR,CTR STUDIO FARMACOL CELLULARE & MOL,I-20133 MILAN,ITALY.
   UNIV ROME,SEZ BIOL CELLULARE,DIP BIOPATHOL UMANA,ROME,ITALY.
C3 Consiglio Nazionale delle Ricerche (CNR); CIVIS; Sapienza University
   Rome
RP Cairns, LA (通讯作者)，UNIV MILAN,DIP GENET & BIOL MICROORGANISMI,MILAN,ITALY.
OI Crotta, Stefania/0000-0003-4676-8616
CR Anderson D J, 1993, Curr Opin Neurobiol, V3, P8, DOI 10.1016/0959-4388(93)90028-W
   ANDERSON DJ, 1989, TRENDS GENET, V5, P74
   BABINET C, 1989, GENOME, V31, P938, DOI 10.1139/g89-165
   BIRREN SJ, 1990, NEURON, V4, P189, DOI 10.1016/0896-6273(90)90094-V
   BOTTERI FM, 1987, MOL CELL BIOL, V7, P3178, DOI 10.1128/MCB.7.9.3178
   CAIRNS LA, 1994, EMBO J, V13, P4577, DOI 10.1002/j.1460-2075.1994.tb06779.x
   CHOMCZYNSKI P, 1987, ANAL BIOCHEM, V162, P156, DOI 10.1016/0003-2697(87)90021-2
   CHURCH GM, 1984, P NATL ACAD SCI-BIOL, V81, P1991, DOI 10.1073/pnas.81.7.1991
   CLAUDE P, 1988, NEURON, V1, P783, DOI 10.1016/0896-6273(88)90126-2
   DOUPE AJ, 1985, J NEUROSCI, V5, P2143
   DOUPE AJ, 1985, J NEUROSCI, V5, P2119
   EIDEN LE, 1987, NEUROSCIENCE, V21, P1019, DOI 10.1016/0306-4522(87)90056-X
   ERANKO O, 1972, HISTOCHEM J, V4, P49, DOI 10.1007/BF01005268
   GEORGE KM, 1994, DEVELOPMENT, V120, P2673
   GREENE LA, 1976, P NATL ACAD SCI USA, V73, P2424, DOI 10.1073/pnas.73.7.2424
   GROVES AK, 1995, DEVELOPMENT, V121, P887
   GUILLEMOT F, 1993, MECH DEVELOP, V42, P171, DOI 10.1016/0925-4773(93)90006-J
   GUILLEMOT F, 1993, CELL, V75, P463, DOI 10.1016/0092-8674(93)90381-Y
   IKEDA I, 1990, ONCOGENE, V5, P1291
   IP NY, 1994, NEURON, V13, P443, DOI 10.1016/0896-6273(94)90359-X
   JAT PS, 1991, P NATL ACAD SCI USA, V88, P5096, DOI 10.1073/pnas.88.12.5096
   Jing SQ, 1996, CELL, V85, P1113, DOI 10.1016/S0092-8674(00)81311-2
   JOHNSON JE, 1990, NATURE, V346, P858, DOI 10.1038/346858a0
   LANDIS SC, 1981, TRENDS NEUROSCI, V4, P172, DOI 10.1016/0166-2236(81)90056-4
   MOSMANN T, 1983, J IMMUNOL METHODS, V65, P55, DOI 10.1016/0022-1759(83)90303-4
   ORKIN SH, 1995, J BIOL CHEM, V270, P4955, DOI 10.1074/jbc.270.10.4955
   PACHNIS V, 1993, DEVELOPMENT, V119, P1005
   POSSENTI R, 1989, EMBO J, V8, P2217, DOI 10.1002/j.1460-2075.1989.tb08345.x
   ROBINSON MO, 1994, P NATL ACAD SCI USA, V91, P12798, DOI 10.1073/pnas.91.26.12798
   SANTORO M, 1990, ONCOGENE, V5, P1595
   SHALNIK DG, 1991, MOL CELL BIOL, V11, P4518
   STEMPLE DL, 1988, NEURON, V1, P517, DOI 10.1016/0896-6273(88)90182-1
   TAKAHASHI M, 1987, MOL CELL BIOL, V7, P1378, DOI 10.1128/MCB.7.4.1378
   Treanor JJS, 1996, NATURE, V382, P80, DOI 10.1038/382080a0
   Trupp M, 1996, NATURE, V381, P785, DOI 10.1038/381785a0
   UNSICKER K, 1978, P NATL ACAD SCI USA, V75, P3498, DOI 10.1073/pnas.75.7.3498
   UNSICKER K, 1989, INT J DEV NEUROSCI, V7, P413, DOI 10.1016/0736-5748(89)90002-6
   VANHEYNINGEN V, 1994, NATURE, V367, P319, DOI 10.1038/367319a0
   WHITE LA, 1994, J NEUROSCI, V14, P6744
   YOMOGIDA K, 1994, DEVELOPMENT, V120, P1759
NR 40
TC 12
Z9 12
U1 0
U2 0
PU STOCKTON PRESS
PI BASINGSTOKE
PA HOUNDMILLS, BASINGSTOKE, HAMPSHIRE, ENGLAND RG21 6XS
SN 0950-9232
J9 ONCOGENE
JI Oncogene
PD JUN 26
PY 1997
VL 14
IS 25
BP 3093
EP 3098
DI 10.1038/sj.onc.1201137
PG 6
WC Biochemistry & Molecular Biology; Oncology; Cell Biology; Genetics &
   Heredity
WE Science Citation Index Expanded (SCI-EXPANDED)
SC Biochemistry & Molecular Biology; Oncology; Cell Biology; Genetics &
   Heredity
GA XG100
UT WOS:A1997XG10000012
PM 9223673
OA Bronze
DA 2023-08-27
ER

PT J
AU Wang, SJ
   Hsu, WC
   Young, YH
AF Wang, SJ
   Hsu, WC
   Young, YH
TI Reversible cochleo-vestibular deficits in two cases of jugular foramen
   tumor after surgery
SO EUROPEAN ARCHIVES OF OTO-RHINO-LARYNGOLOGY
LA English
DT Article
DE jugular foramen schwannoma; glomus jugulare tumor; sudden deafness;
   vocal palsy
ID VOCAL FOLD IMMOBILITY; GLOMUS-JUGULARE; PRESERVATION; SCHWANNOMA
AB Primary jugular foramen (JF) tumor, such as glomus jugular tumor or JF schwannoma, may manifest as a lower cranial nerve deficit; in addition, it can be accompanied by deafness or vertigo if it affects the cranial nerve (CN) VIII. Recently, we encountered JF schwannoma 1 and glomus jugulare tumor 1. Both cases invaded the adjacent cerebellopontine angle, leading to cochleo-vestibular deficits prior to the operation. After surgery, recovery of the audiovestibular function, including hearing, auditory brainstem response and caloric response, was anticipated in both patients. Therefore, cochleo-vestibular deficits in JF tumors can be attributed to compression neuropathy, rather than tumor infiltration.
C1 Natl Taiwan Univ Hosp, Dept Otolaryngol, Taipei, Taiwan.
   Natl Taiwan Univ, Coll Med, Taipei 10018, Taiwan.
C3 National Taiwan University; National Taiwan University Hospital;
   National Taiwan University
RP Young, YH (通讯作者)，Natl Taiwan Univ Hosp, Dept Otolaryngol, 1 Chang Te St, Taipei, Taiwan.
EM
OI Hsu, Wei-Chung/0000-0001-8583-8459
CR Al-Mefty O, 2002, J NEUROSURG, V97, P1356, DOI 10.3171/jns.2002.97.6.1356
   Altman JS, 1997, J VOICE, V11, P364, DOI 10.1016/S0892-1997(97)80016-1
   Benninger MS, 1998, LARYNGOSCOPE, V108, P1346, DOI 10.1097/00005537-199809000-00016
   Bertrand R A, 1976, J Otolaryngol, V5, P44
   BLUMENFELD J, 1993, AM J HYPERTENS, V6, P382, DOI 10.1093/ajh/6.5.382
   Chao CK, 2000, OTOLARYNG HEAD NECK, V122, P922, DOI 10.1067/mhn.2000.104522
   Foote RL, 2002, HEAD NECK-J SCI SPEC, V24, P332, DOI 10.1002/hed.10005
   KAYE AH, 1984, J NEUROSURG, V60, P1045, DOI 10.3171/jns.1984.60.5.1045
   Kim CJ, 2001, LARYNGOSCOPE, V111, P2071, DOI 10.1097/00005537-200111000-00038
   MATISHAK MZ, 1987, J NEUROSURG, V66, P604, DOI 10.3171/jns.1987.66.4.0604
   Murofushi T, 2001, ARCH OTOLARYNGOL, V127, P1069, DOI 10.1001/archotol.127.9.1069
   MURPHY TP, 1989, LARYNGOSCOPE, V99, P1244
   PELLET W, 1988, J NEUROSURG, V69, P887, DOI 10.3171/jns.1988.69.6.0887
   PLUTA RM, 1994, J NEUROSURG, V80, P1091, DOI 10.3171/jns.1994.80.6.1091
   Rapana A, 1997, CLIN NEUROL NEUROSUR, V99, P196, DOI 10.1016/S0303-8467(97)00016-4
   Sen C, 2001, NEUROSURGERY, V48, P838, DOI 10.1097/00006123-200104000-00029
   Slaba S, 2001, J Med Liban, V49, P231
   Takahashi M, 1997, EUR ARCH OTO-RHINO-L, V254, P474, DOI 10.1007/BF02439985
   Tekdemir I, 2001, J CLIN NEUROSCI, V8, P351, DOI 10.1054/jocn.2000.0865
   Young YH, 2002, LARYNGOSCOPE, V112, P509, DOI 10.1097/00005537-200203000-00019
NR 20
TC 4
Z9 4
U1 0
U2 2
PU SPRINGER
PI NEW YORK
PA 233 SPRING ST, NEW YORK, NY 10013 USA
SN 0937-4477
EI 1434-4726
J9 EUR ARCH OTO-RHINO-L
JI Eur. Arch. Oto-Rhino-Laryn.
PD MAY
PY 2004
VL 261
IS 5
BP 247
EP 250
DI 10.1007/s00405-003-0666-z
PG 4
WC Otorhinolaryngology
WE Science Citation Index Expanded (SCI-EXPANDED)
SC Otorhinolaryngology
GA 819AF
UT WOS:000221284900004
PM 13680260
DA 2023-08-27
ER

PT J
AU HIRATO, M
   NAKAMURA, M
   INOUE, HK
   OHYE, C
   HIRATO, J
   SHIBAZAKI, T
   ANDOU, Y
AF HIRATO, M
   NAKAMURA, M
   INOUE, HK
   OHYE, C
   HIRATO, J
   SHIBAZAKI, T
   ANDOU, Y
TI GAMMA-KNIFE RADIOSURGERY FOR THE TREATMENT OF BRAIN-STEM TUMORS
SO STEREOTACTIC AND FUNCTIONAL NEUROSURGERY
LA English
DT Article; Proceedings Paper
CT 1994 Meeting of the Leksell-Gamma-Knife-Society
CY MAY 08-11, 1994
CL KYOTO, JAPAN
SP Leksell Gamma Knife Soc
DE GAMMA KNIFE; RADIOSURGERY; BRAIN-STEM TUMOR; HISTOPATHOLOGY; EPENDYMOMA;
   LYMPHOMA; GLIOMA
ID CENTRAL-NERVOUS-SYSTEM; FRACTIONATED STEREOTAXIC RADIOTHERAPY;
   RADIOBIOLOGY; LYMPHOMA; MODEL
AB Ten patients with brainstem tumors (BST) underwent Gamma Knife radiosurgery. In 5 cases, there were definitive histological diagnoses; ependymoma (n = 3) and CNS lymphoma (n = 2). The others were diagnosed as ependymoma (n = 1), CNS lymphoma (n = 2) and glioma (n = 2) on clinical grounds. Of 4 cases with ependymoma, the tumor showed a marked response in 3 cases. Of 4 cases with CNS lymphoma, all responded rapidly and the clinical symptoms improved markedly. However, in the 2 cases with glioma, the tumor remained unchanged or continued to grow Gamma Knife radiosurgery was effective in local growth control of BST without adverse effects. An alternative treatment design will be required in glioma cases.
C1 GUNMA UNIV,SCH MED,DEPT PATHOL,MAEBASHI,GUMMA 371,JAPAN.
   HIDAKA HOSP,TAKASAKI,GUMMA,JAPAN.
C3 Gunma University
RP HIRATO, M (通讯作者)，GUNMA UNIV,SCH MED,DEPT NEUROSURG,3-39-15 SHOWA MACHI,MAEBASHI,GUMMA 371,JAPAN.
CR AHO R, 1993, ACTA NEUROPATHOL, V86, P71, DOI 10.1007/BF00454901
   BLATT DR, 1994, J NEUROSURG, V80, P1046, DOI 10.3171/jns.1994.80.6.1046
   DEANGELIS LM, 1990, NEUROLOGY, V40, P80, DOI 10.1212/WNL.40.1.80
   FLICKINGER JC, 1992, INT J RADIAT ONCOL, V23, P19, DOI 10.1016/0360-3016(92)90539-T
   HOCHBERG FH, 1988, J NEUROSURG, V68, P835, DOI 10.3171/jns.1988.68.6.0835
   KONDZIOLKA D, 1992, NEUROSURGERY, V31, P280, DOI 10.1227/00006123-199208000-00013
   KONDZIOLKA D, 1992, NEUROSURGERY, V31, P271, DOI 10.1227/00006123-199208000-00012
   Kondziolka D, 1993, Neurosurg Clin N Am, V4, P469
   LARSON DA, 1993, INT J RADIAT ONCOL, V25, P557, DOI 10.1016/0360-3016(93)90080-F
   LINSKEY ME, 1993, J NEUROSURG, V78, P645, DOI 10.3171/jns.1993.78.4.0645
   MCKENZIE MR, 1993, CAN J NEUROL SCI, V20, P279
   OBRIEN MS, 1985, NEUROSURGERY, V1, P762
   PLUNKETT SR, 1985, NEUROSURGERY, V1, P1096
   PODGORSAK EB, 1993, INT J RADIAT ONCOL, V27, P1225, DOI 10.1016/0360-3016(93)90547-9
   SOCIE G, 1990, CANCER, V65, P322, DOI 10.1002/1097-0142(19900115)65:2<322::AID-CNCR2820650223>3.0.CO;2-D
NR 15
TC 22
Z9 22
U1 0
U2 0
PU KARGER
PI BASEL
PA ALLSCHWILERSTRASSE 10, CH-4009 BASEL, SWITZERLAND
SN 1011-6125
J9 STEREOT FUNCT NEUROS
JI Stereotact. Funct. Neurosurg.
PY 1995
VL 64
SU 1
BP 32
EP 41
DI 10.1159/000098762
PG 10
WC Neurosciences; Neuroimaging; Surgery
WE Conference Proceedings Citation Index - Science (CPCI-S); Science Citation Index Expanded (SCI-EXPANDED)
SC Neurosciences & Neurology; Surgery
GA TG375
UT WOS:A1995TG37500005
PM 8584838
DA 2023-08-27
ER

PT J
AU Burkey, JM
   Rizer, FM
   Schuring, AG
   Fucci, MJ
   Lippy, WH
AF Burkey, JM
   Rizer, FM
   Schuring, AG
   Fucci, MJ
   Lippy, WH
TI Acoustic reflexes, auditory brainstem response, and MRI in the
   evaluation of acoustic neuromas
SO LARYNGOSCOPE
LA English
DT Article
ID STEM RESPONSE; LATENCY; TESTS
AB Patient records were reviewed to determine whether persons with absent acoustic reflexes have a higher incidence of abnormal auditory brainstem response (ABR) results in the absence of a cerebellopontine angle (CPA) tumor than those with normal acoustic reflexes. Results showed patients with absent reflexes to have borderline or abnormal ABR results in 45.2% of the cases. Patients with normal reflexes had borderline or abnormal ABR results in 14.2% of the cases. Results indicate that magnetic resonance imaging is a more appropriate test for patients with absent reflexes, since ABR was often nondiagnostic for a CPA tumor in this group.
C1 WARREN OTOL GRP,WARREN,OH.
CR CHANDRASEKHAR SS, 1995, AM J OTOL, V16, P63
   COLLET L, 1992, INT J NEUROSCI, V67, P187, DOI 10.3109/00207459208994784
   EGGERMONT JJ, 1980, ANN OTO RHINOL LARYN, V89, P1
   GORDON ML, 1995, AM J OTOL, V16, P136
   HART RG, 1981, NEUROSURGERY, V9, P450, DOI 10.1227/00006123-198110000-00021
   JOSEY AF, 1988, AM J OTOL, V9, P12
   LIGHTFOOT GR, 1992, BRIT J AUDIOL, V26, P217, DOI 10.3109/03005369209076640
   MAIR IWS, 1988, SCAND AUDIOL, V17, P163, DOI 10.3109/01050398809042188
   TELIAN SA, 1989, LARYNGOSCOPE, V99, P10
   THOMSEN J, 1990, AM J OTOL, V11, P12
   TURNER RG, 1984, EAR HEARING, V5, P125, DOI 10.1097/00003446-198405000-00002
   TURNER RG, 1984, EAR HEARING, V5, P187, DOI 10.1097/00003446-198407000-00002
   WILSON DF, 1992, LARYNGOSCOPE, V102, P961
NR 13
TC 16
Z9 16
U1 0
U2 6
PU LARYNGOSCOPE CO
PI ST LOUIS
PA 10 S BROADWAY 14TH FLOOR, ST LOUIS, MO 63102-1741
SN 0023-852X
J9 LARYNGOSCOPE
JI Laryngoscope
PD JUL
PY 1996
VL 106
IS 7
BP 839
EP 841
DI 10.1097/00005537-199607000-00010
PG 3
WC Medicine, Research & Experimental; Otorhinolaryngology
WE Science Citation Index Expanded (SCI-EXPANDED)
SC Research & Experimental Medicine; Otorhinolaryngology
GA UW659
UT WOS:A1996UW65900010
PM 8667979
DA 2023-08-27
ER

PT J
AU Tennekes, H
   Gembardt, C
   Dammann, M
   van Ravenzwaay, B
AF Tennekes, H
   Gembardt, C
   Dammann, M
   van Ravenzwaay, B
TI The stability of historical control data for common neoplasms in
   laboratory rats: adrenal gland (medulla), mammary gland, liver,
   endocrine pancreas, and pituitary gland
SO REGULATORY TOXICOLOGY AND PHARMACOLOGY
LA English
DT Article
DE spontaneous tumors; rat; adrenals; liver; mammary gland; pituitary;
   pancreas
AB Time-related changes in the incidences of spontaneous neoplasms in adrenals (medulla), mamma, liver, pituitary, and (endocrine) pancreas were assessed statistically in Wistar, Sprague-Dawley, and F344 rats employed by BASF, Germany and major European contract research organizations over the last 20 years. Negative trends (7 of 80 cases) were observed for pituitary pars distalis adenomas in Sprague-Dawley males and females, for pancreas islet cell adenomas in BASF Wistar males and females, for benign adrenal pheochromocytomas in Sprague-Dawley males, for malignant pheochromocytomas in F344 males, and for mammary gland fibroadenomas in BASF Wistar females. Positive trends (13 of 80 cases) were observed for benign pheochromocytomas, mammary gland adenocarcinomas, and pancreas islet cell carcinomas in HanWistar females, for malignant pheochromocytomas and islet cell carcinomas in BASF Wistar males, for benign pheochromocytomas and islet cell adenomas in F344 males, for mammary gland fibroadenomas in Sprague-Dawley females, and for benign hepatocellular tumors in HanWistar males and females, and in BASF Wistar and Sprague-Dawley females. In 60 of 80 cases there were no statistically significant trends. These results indicate that the majority of tumor types showed no time trends and that, in each rat strain, certain tumor types are susceptible to slight positive or negative time trends. Accordingly, the validity and use of historical control data should be based on an organ- and strain-specific statistical analysis of tumor incidence over time. (C) 2004 Elsevier Inc. All rights reserved.
C1 BASF AG, Prod Safety, D-67056 Ludwigshafen, Germany.
   Nederland BV, ETS, NL-7201 EA Zutphen, Netherlands.
C3 BASF
RP van Ravenzwaay, B (通讯作者)，BASF AG, Prod Safety, Z 470, D-67056 Ludwigshafen, Germany.
EM
CR Eiben R, 1999, EXP TOXICOL PATHOL, V51, P523, DOI 10.1016/S0940-2993(99)80133-X
   HASEMAN JK, 1989, FUND APPL TOXICOL, V12, P793, DOI 10.1016/0272-0590(89)90011-0
   JONCKHEERE AR, 1954, BIOMETRIKA, V41, P133, DOI 10.2307/2333011
   MOHR U, 1994, IARC SCI PUBLICATION, V122
   MOHR U, 1997, IARC SCI PUBLICATION, V122
   MOHR U, 1993, IARC SCI PUBLICATION, V122
   *REG IND TOX AN DA, 1999, EXP TOXICOL PATHOL, V51, P461
   van Ravenzwaay B, 2002, REGUL TOXICOL PHARM, V36, P86, DOI 10.1006/rtph.2002.1567
NR 8
TC 22
Z9 23
U1 0
U2 4
PU ACADEMIC PRESS INC ELSEVIER SCIENCE
PI SAN DIEGO
PA 525 B ST, STE 1900, SAN DIEGO, CA 92101-4495 USA
SN 0273-2300
EI 1096-0295
J9 REGUL TOXICOL PHARM
JI Regul. Toxicol. Pharmacol.
PD AUG
PY 2004
VL 40
IS 1
BP 18
EP 27
DI 10.1016/j.yrtph.2004.04.003
PG 10
WC Medicine, Legal; Pharmacology & Pharmacy; Toxicology
WE Science Citation Index Expanded (SCI-EXPANDED)
SC Legal Medicine; Pharmacology & Pharmacy; Toxicology
GA 844HM
UT WOS:000223148900003
PM 15265603
DA 2023-08-27
ER

PT J
AU Su, JM
   Kilburn, LB
   Mansur, DB
   Krailo, M
   Buxton, A
   Adekunle, A
   Gajjar, A
   Adamson, PC
   Weigel, B
   Fox, E
   Blaney, SM
   Fouladi, M
AF Su, Jack M.
   Kilburn, Lindsay B.
   Mansur, David B.
   Krailo, Mark
   Buxton, Allen
   Adekunle, Adesina
   Gajjar, Amar
   Adamson, Peter C.
   Weigel, Brenda
   Fox, Elizabeth
   Blaney, Susan M.
   Fouladi, Maryam
TI Phase I/II trial of vorinostat and radiation and maintenance vorinostat
   in children with diffuse intrinsic pontine glioma: A Children's Oncology
   Group report
SO NEURO-ONCOLOGY
LA English
DT Article
DE children; diffuse intrinsic pontine glioma; phase I; II clinical trials;
   suberoylanilide hydroxamic acid; Vorinostat
ID SUBEROYLANILIDE HYDROXAMIC ACID; BRAIN-STEM TUMORS; HISTONE DEACETYLASE
   INHIBITOR; HIGH-DOSE-CHEMOTHERAPY; HYPERFRACTIONATED RADIOTHERAPY;
   IN-VITRO; II TRIAL; THERAPY; CARBOPLATIN; VINCRISTINE
AB Background A phase I/II trial of vorinostat (suberoylanilide hydroxamic acid), an oral histone deacetylase inhibitor, was conducted in children with newly diagnosed diffuse intrinsic pontine glioma (DIPG) through the Children's Oncology Group (COG) to: 1) determine the recommended phase II dose (RP2D) of vorinostat given concurrently with radiation therapy; 2) document the toxicities of continuing vorinostat as maintenance therapy after radiation; and 3) to determine the efficacy of this regimen by comparing the risk of progression or death with a historical model from past COG trials. Methods Vorinostat was given once daily, Monday through Friday, during radiation therapy (54 Gy in 30 fractions), and then continued at 230 mg/m(2) daily for a maximum of twelve 28-day cycles. Results Twelve patients enrolled in the phase I study; the RP2D of vorinostat given concurrently with radiation was 230 mg/m(2)/day, Monday through Friday weekly. The six patients enrolled at the RP2D and an additional 64 patients enrolled in the phase II study contributed to the efficacy assessment. Although vorinostat was well-tolerated, did not interrupt radiation therapy, and was permanently discontinued in only 8.6% of patients due to toxicities, risk for EFS-event was not significantly reduced compared with the target risk derived from historical COG data (P = 0.32; 1-sided). The 1-year EFS was 5.85% (95% CI 1.89-13.1%) and 1-year OS was 39.2% (27.8-50.5%). Conclusions Vorinostat given concurrently with radiation followed by vorinostat monotherapy was well tolerated in children with newly diagnosed DIPG but failed to improve outcome.
C1 [Su, Jack M.; Blaney, Susan M.] Baylor Coll Med, Texas Childrens Canc Ctr, 6701 Fannin St,CC1510, Houston, TX 77030 USA.
   [Kilburn, Lindsay B.] Childrens Natl Med Ctr, Ctr Canc & Blood Disorders, Washington, DC USA.
   [Mansur, David B.] Rainbow Babies & Childrens Hosp, Dept Radiat Oncol, Cleveland, OH USA.
   [Krailo, Mark; Buxton, Allen] Childrens Oncol Grp, Dept Stat, Monrovia, CA USA.
   [Adekunle, Adesina] Texas Childrens Hosp, Dept Pathol, Houston, TX USA.
   [Gajjar, Amar; Fox, Elizabeth] St Jude Childrens Res Hosp, Dept Oncol, Memphis, TN USA.
   [Adamson, Peter C.] Sanofi, Global Head, Oncol Dev, Cambridge, MA USA.
   [Weigel, Brenda] Univ Minnesota, Dept Pediat, Hem Onc BMT, Masonic Canc Ctr, Minneapolis, MN USA.
   [Fouladi, Maryam] Nationwide Childrens Hosp, Neuro Oncol Program, Columbus, OH USA.
C3 Baylor College of Medicine; Texas Children's Cancer Center; Children's
   National Health System; Case Western Reserve University; Case Western
   Reserve University Hospital; University Hospitals of Cleveland; Rainbow
   Babies & Children's Hospital; Children's Oncology Group (COG); Baylor
   College of Medicine; St Jude Children's Research Hospital;
   Sanofi-Aventis; University of Minnesota System; University of Minnesota
   Twin Cities; University System of Ohio; Ohio State University;
   Nationwide Childrens Hospital
RP Su, JM (通讯作者)，Baylor Coll Med, Texas Childrens Canc Ctr, 6701 Fannin St,CC1510, Houston, TX 77030 USA.
EM
RI Fox, Elizabeth/AAD-5341-2020; Adamson, Peter C./C-5210-2009; Gajjar,
   Amar/N-8059-2018
FU National Institute of Health/National Cancer Institute (IROC grant)
   [U24CA180803]; National Cancer Institute, National Clinical Trials
   Network (Operations Center Grant) [U10CA180886]; St. Baldrick's
   Foundation Consortium Grant; National Cancer Institute, National
   Clinical Trials Network (Statistics & Data Center Grant) [U10CA180899]
FX This work was supported by National Institute of Health/National Cancer
   Institute (IROC grant, U24CA180803), National Cancer Institute, National
   Clinical Trials Network (Operations Center Grant, U10CA180886), National
   Cancer Institute, National Clinical Trials Network (Statistics & Data
   Center Grant, U10CA180899) and St. Baldrick's Foundation Consortium
   Grant.
CR Allen J, 1999, CANCER, V86, P1064, DOI 10.1002/(SICI)1097-0142(19990915)86:6<1064::AID-CNCR24>3.0.CO;2-1
   Bernier-Chastagner V, 2005, CANCER-AM CANCER SOC, V104, P2792, DOI 10.1002/cncr.21534
   Bouffet E, 2000, CANCER, V88, P685, DOI 10.1002/(SICI)1097-0142(20000201)88:3<685::AID-CNCR27>3.0.CO;2-K
   Broniscer A, 2005, CANCER-AM CANCER SOC, V103, P133, DOI 10.1002/cncr.20741
   Broniscer A, 2000, J CLIN ONCOL, V18, P1246, DOI 10.1200/JCO.2000.18.6.1246
   Buczkowicz P, 2014, NAT GENET, V46, P451, DOI 10.1038/ng.2936
   Castel D, 2015, ACTA NEUROPATHOL, V130, P815, DOI 10.1007/s00401-015-1478-0
   Chinnaiyan P, 2005, INT J RADIAT ONCOL, V62, P223, DOI 10.1016/j.ijrobp.2004.12.088
   Cohen KJ, 2011, NEURO-ONCOLOGY, V13, P317, DOI 10.1093/neuonc/noq191
   DEMETS DL, 1994, STAT MED, V13, P1341, DOI 10.1002/sim.4780131308
   Doz F, 2002, EUR J CANCER, V38, P815, DOI 10.1016/S0959-8049(02)00029-1
   Dunkel IJ, 1998, J NEURO-ONCOL, V37, P67, DOI 10.1023/A:1005874508975
   Eyupoglu IY, 2005, J NEUROCHEM, V93, P992, DOI 10.1111/j.1471-4159.2005.03098.x
   Fouladi M, 2010, J CLIN ONCOL, V28, P3623, DOI 10.1200/JCO.2009.25.9119
   Galanis E, 2009, J CLIN ONCOL, V27, P2052, DOI 10.1200/JCO.2008.19.0694
   Grasso CS, 2015, NAT MED, V21, P555, DOI 10.1038/nm.3855
   Greenberg ML, 2005, PEDIATR BLOOD CANCER, V45, P644, DOI 10.1002/pbc.20382
   Guntner AS, 2020, ACTA NEUROPATHOL COM, V8, DOI 10.1186/s40478-020-00953-2
   Haas-Kogan DA, 2011, NEURO-ONCOLOGY, V13, P298, DOI 10.1093/neuonc/noq202
   Hanson JE, 2013, PLOS ONE, V8, DOI 10.1371/journal.pone.0069964
   Hargrove D, 2006, LANCET ONCOL, V7, P241, DOI 10.1016/S1470-2045(06)70615-5
   Hockly E, 2003, P NATL ACAD SCI USA, V100, P2041, DOI 10.1073/pnas.0437870100
   Hoffman LM, 2018, J CLIN ONCOL, V36, P1963, DOI 10.1200/JCO.2017.75.9308
   Jakacki RI, 1999, J NEURO-ONCOL, V44, P77, DOI 10.1023/A:1006360222643
   Jennings MT, 2002, J CLIN ONCOL, V20, P3431, DOI 10.1200/JCO.2002.04.109
   Kalbfleisch J. D., 2002, STAT ANAL FAILURE TI
   KAPLAN EL, 1958, J AM STAT ASSOC, V53, P457, DOI 10.2307/2281868
   Kelly WK, 2005, J CLIN ONCOL, V23, P3923, DOI 10.1200/JCO.2005.14.167
   Korones DN, 2008, PEDIATR BLOOD CANCER, V50, P227, DOI 10.1002/pbc.21154
   KRETSCHMAR CS, 1993, CANCER-AM CANCER SOC, V72, P1404, DOI 10.1002/1097-0142(19930815)72:4<1404::AID-CNCR2820720441>3.0.CO;2-G
   Mandell LR, 1999, INT J RADIAT ONCOL, V43, P959, DOI 10.1016/S0360-3016(98)00501-X
   Marcus KJ, 2003, INT J RADIAT ONCOL, V55, P1182, DOI 10.1016/S0360-3016(02)04391-2
   Marks PA, 2007, ONCOGENE, V26, P1351, DOI 10.1038/sj.onc.1210204
   Mielcarek M, 2011, PLOS ONE, V6, DOI 10.1371/journal.pone.0027746
   O'Connor OA, 2006, J CLIN ONCOL, V24, P166, DOI 10.1200/JCO.2005.01.9679
   Olsen EA, 2007, J CLIN ONCOL, V25, P3109, DOI 10.1200/JCO.2006.10.2434
   Packer RJ, 2005, CANCER-AM CANCER SOC, V104, P1968, DOI 10.1002/cncr.21403
   Packer RJ, 1996, CANCER-AM CANCER SOC, V77, P2150, DOI 10.1002/(SICI)1097-0142(19960515)77:10<2150::AID-CNCR28>3.0.CO;2-T
   Pollack IF, 2011, NEURO-ONCOLOGY, V13, P290, DOI 10.1093/neuonc/noq199
   Rasmussen RD, 2016, MOL ONCOL, V10, P751, DOI 10.1016/j.molonc.2015.12.014
   Sandri A, 2006, CHILD NERV SYST, V22, P1127, DOI 10.1007/s00381-006-0083-x
   Schemper M, 1996, CONTROL CLIN TRIALS, V17, P343, DOI 10.1016/0197-2456(96)00075-X
   Skolnik JM, 2008, J CLIN ONCOL, V26, P190, DOI 10.1200/JCO.2007.12.7712
   Spiller SE, 2006, J NEURO-ONCOL, V79, P259, DOI 10.1007/s11060-006-9142-0
   Ugur HC, 2007, J NEURO-ONCOL, V83, P267, DOI 10.1007/s11060-007-9337-z
   Walter AW, 1998, MED PEDIATR ONCOL, V30, P28, DOI 10.1002/(SICI)1096-911X(199801)30:1<28::AID-MPO9>3.0.CO;2-2
   Wolff JEA, 2002, BRIT J CANCER, V87, P945, DOI 10.1038/sj.bjc.6600552
   WOOLSON RF, 1981, BIOMETRICS, V37, P687, DOI 10.2307/2530150
   Wu G, 2012, NAT GENET, V44, P251, DOI 10.1038/ng.1102
   Yin D, 2007, CLIN CANCER RES, V13, P1045, DOI 10.1158/1078-0432.CCR-06-1261
NR 50
TC 12
Z9 12
U1 1
U2 4
PU OXFORD UNIV PRESS INC
PI CARY
PA JOURNALS DEPT, 2001 EVANS RD, CARY, NC 27513 USA
SN 1522-8517
EI 1523-5866
J9 NEURO-ONCOLOGY
JI Neuro-Oncology
PD APR 1
PY 2022
VL 24
IS 4
BP 655
EP 664
DI 10.1093/neuonc/noab188
EA AUG 2021
PG 10
WC Oncology; Clinical Neurology
WE Science Citation Index Expanded (SCI-EXPANDED)
SC Oncology; Neurosciences & Neurology
GA 0E6KI
UT WOS:000776787900018
PM 34347089
OA Green Published
DA 2023-08-27
ER

PT J
AU Li, ZP
   Gao, Z
   Li, SF
   Zhang, YY
   Xing, LZ
   Zhang, LJ
   Ma, GJ
   Zhao, XB
   Shao, MT
AF Li, Zongpeng
   Gao, Zong
   Li, Shufa
   Zhang, Yuanyuan
   Xing, Lizhi
   Zhang, Lanju
   Ma, Guijie
   Zhao, Xinbo
   Shao, Mingtao
TI Microinjection of calcitonin in midbrain periaqueductal gray attenuates
   hyperalgesia in a chronic constriction injury rat model
SO IRANIAN JOURNAL OF BASIC MEDICAL SCIENCES
LA English
DT Article
DE Analgesia; Calcitonin; Calcitonin receptor; Pain; Periaqueductal gray
ID CENTRAL-NERVOUS-SYSTEM; BINDING-SITES; VENTROMEDIAL MEDULLA; PAIN;
   RECEPTORS; GENE; LOCALIZATION; NOCICEPTION; INHIBITION; EXPRESSION
AB Objective(s): As heat, pain is one of the most common clinical symptoms. Generally, calcitonin (CT) is prescribed as an analgesic agent for the treatment of pain, especially for the pain caused by osteoporosis or primary and metastatic bone tumor. However, the detailed mechanism remains unknown.
   Materials and Methods: In this study, chronic constriction injury (CCI) rat model was created, and hot plate test and von frey filaments test were employed to evaluate thermal withdrawal latency (TWL) and mechanical withdrawal threshold (MWT), respectively. Immunohistochemistry staining and western blot analyses were used to assess the distribution and expression of calcitonin receptor (CT-R) in the midbrain periaqueductal gray (PAG), which was a pivotal site in the modulatory system for the endogenous pain.
   Results: The TWL and MWT before the surgery (19.6 +/- 1.19 sec) were significantly longer than that at day 2 (12.5 +/- 1.60 sec), and day 14 (7.75 +/- 0.89 sec) (P<0.01; P<0.01), respectively. The TWL and MWT at day 14 were significantly increased compared to that at day 7 after microinjection of salmon calcitonin (sCT) with different doses. Interestingly, the expression of CT-R was up-regulated in neuropathic pain. Furthermore, the expression of CT-R was significantly up-regulated and algesia was remarkably relieved when CT was microinjected into PAG.
   Conclusion: These results suggested that an increased CT-R might be associated with hyperalgesia in CCI rat, and CT had a potent antinociceptive effect by the up-regulation of CT-R in the PAG. Thus, it might provide a potential approach for the treatment of bone pain.
C1 [Li, Zongpeng] Linyi Peoples Hosp, Linyi 276000, Peoples R China.
   [Gao, Zong] Shandong Univ Tradit Chinese Med, Affiliated Hosp, Neurosurg Dept, Jinan 250011, Peoples R China.
   [Li, Shufa; Zhang, Yuanyuan; Xing, Lizhi; Zhang, Lanju; Ma, Guijie; Zhao, Xinbo] Linyi Peoples Hosp, Dept Endocrinol, Linyi 276000, Peoples R China.
   [Shao, Mingtao] Linyi Peoples Hosp, Dept Surg, Linyi 276000, Peoples R China.
C3 Shandong University of Traditional Chinese Medicine
RP Li, SF (通讯作者)，Linyi Peoples Hosp, Dept Endocrinol, Linyi 276000, Peoples R China.
EM
RI zhang, yuanyuan/GYA-4428-2022
OI li, shufa/0000-0002-0272-549X
FU Eleventh Five Gerontology Research Program of Civil Affairs Ministry of
   China (Beijing China Fund) [[2008]47-1-48]
FX This study was supported by the Eleventh Five Gerontology Research
   Program of Civil Affairs Ministry of China (Beijing China Fund No.
   [2008]47-1-48). Zongpeng Li and Lanju Zhang contributed equal to this
   study.
CR Aboufatima R, 2004, NEUROSCI LETT, V359, P5, DOI 10.1016/j.neulet.2003.09.022
   Backryd E, 2014, PAIN MED, V15, P111, DOI 10.1111/pme.12248
   BASBAUM AI, 1984, ANNU REV NEUROSCI, V7, P309, DOI 10.1146/annurev.ne.07.030184.001521
   BASBAUM AI, 1978, ANN NEUROL, V4, P451, DOI 10.1002/ana.410040511
   BENNETT GJ, 1988, PAIN, V33, P87, DOI 10.1016/0304-3959(88)90209-6
   Blau LA, 2003, ANN PHARMACOTHER, V37, P564, DOI 10.1345/aph.1C350
   COOPER C, 1992, J BONE MINER RES, V7, P221, DOI 10.1002/jbmr.5650070214
   FABBRI A, 1985, BRAIN RES, V343, P205, DOI 10.1016/0006-8993(85)90736-X
   FISCHER JA, 1985, PEPTIDES, V6, P265, DOI 10.1016/0196-9781(85)90384-5
   Fukui M, 2001, EUR J PHARMACOL, V419, P25, DOI 10.1016/S0014-2999(01)00947-5
   HARGREAVES K, 1988, PAIN, V32, P77, DOI 10.1016/0304-3959(88)90026-7
   Hermann DM, 1997, J CHEM NEUROANAT, V13, P1, DOI 10.1016/S0891-0618(97)00019-7
   JENCK F, 1986, BRAIN RES, V399, P24, DOI 10.1016/0006-8993(86)90597-4
   Kaye AD, 2014, PAIN PHYSICIAN, V17, pE225
   Liu XH, 2010, NEUROCHEM RES, V35, P804, DOI 10.1007/s11064-010-0138-y
   Mason P, 2005, J COMP NEUROL, V493, P2, DOI 10.1002/cne.20751
   Meyer PJ, 2009, PSYCHOPHARMACOLOGY, V204, P531, DOI 10.1007/s00213-009-1482-y
   Morgado C, 2010, EUR J PAIN, V14, P693, DOI 10.1016/j.ejpain.2009.11.011
   MORTON CR, 1986, BRAIN RES, V372, P149, DOI 10.1016/0006-8993(86)91468-X
   Murotani T, 2010, BRAIN RES, V1306, P62, DOI 10.1016/j.brainres.2009.09.117
   OLGIATI VR, 1983, BRAIN RES, V265, P209, DOI 10.1016/0006-8993(83)90334-7
   Paxinos G, 2004, J CHEM NEUROANAT, V27, P217, DOI 10.1016/j.jchemneu.2004.03.005
   PECILE A, 1975, EXPERIENTIA, V31, P332, DOI 10.1007/BF01922569
   SEXTON PM, 1991, MOL NEUROBIOL, V5, P251, DOI 10.1007/BF02935550
   Shamsizadeh A, 2014, IRAN J BASIC MED SCI, V17, P209
   Shen N, 2014, CNS NEUROSCI THER, V15
   Shinki T, 1999, P NATL ACAD SCI USA, V96, P8253, DOI 10.1073/pnas.96.14.8253
   Silverman SL, 2002, OSTEOPOROSIS INT, V13, P858, DOI 10.1007/s001980200118
   SZANTO J, 1992, ONCOLOGY, V49, P180
   Takayama B, 2008, BMC MUSCULOSKEL DIS, V9, DOI 10.1186/1471-2474-9-164
   Tamaddonfard E, 2014, IRAN J BASIC MED SCI, V17, P55
   Tavares I, 2007, J ANAT, V211, P261, DOI 10.1111/j.1469-7580.2007.00759.x
   Vanegas H, 2004, BRAIN RES REV, V46, P295, DOI 10.1016/j.brainresrev.2004.07.004
   Wen LL, 2014, MEDIAT INFLAMM, V2014, DOI 10.1155/2014/171839
   Xia H, 2014, BRAIN RES, V1554, P29, DOI 10.1016/j.brainres.2014.01.032
   Xiao Z, 2010, BRAIN RES, V1330, P31, DOI 10.1016/j.brainres.2010.03.030
   YAKSH TL, 1989, PAIN, V37, P111, DOI 10.1016/0304-3959(89)90160-7
   Yoshimura M, 2000, J BONE MINER METAB, V18, P230, DOI 10.1007/s007740070026
   Youssefi MR, 2012, TROP BIOMED, V29, P519
   ZAIDI M, 1991, VITAM HORM, V46, P87
   ZIMMERMANN M, 1983, PAIN, V16, P109, DOI 10.1016/0304-3959(83)90201-4
NR 41
TC 3
Z9 3
U1 0
U2 4
PU MASHHAD UNIV MED SCIENCES
PI MASHHAD
PA VICE-CHANCELLOR FOR RES CTR OFF IJBMS, DANESHGAH ST, PO BOX 9138813944 -
   445, MASHHAD, 00000, IRAN
SN 2008-3866
EI 2008-3874
J9 IRAN J BASIC MED SCI
JI Iran. J. Basic Med. Sci.
PD JAN
PY 2015
VL 18
IS 1
BP 72
EP 79
PG 8
WC Medicine, Research & Experimental; Pharmacology & Pharmacy
WE Science Citation Index Expanded (SCI-EXPANDED)
SC Research & Experimental Medicine; Pharmacology & Pharmacy
GA CM9PJ
UT WOS:000358040900001
PM 25810879
DA 2023-08-27
ER

PT J
AU GUYOT, JP
   HAUSLER, R
   REVERDIN, A
   BERNEY, J
   MONTANDON, PB
AF GUYOT, JP
   HAUSLER, R
   REVERDIN, A
   BERNEY, J
   MONTANDON, PB
TI DIAGNOSIS OF CEREBELLOPONTINE ANGLE TUMORS
SO ORL-JOURNAL FOR OTO-RHINO-LARYNGOLOGY AND ITS RELATED SPECIALTIES
LA English
DT Article
DE CEREBELLOPONTINE ANGLE TUMORS; BRAIN-STEM AUDITORY EVOKED POTENTIALS;
   INTERAURAL TIME DISCRIMINATION; ELECTRONYSTAGMOGRAPHY; CT SCAN; MAGNETIC
   RESONANCE IMAGING
AB Between 1980 and 1990, 9,176 patients suffering from otoneurological disorders were investigated in our clinic. Sixty-six (0.72%) internal auditory canal or other cerebellopontine angle tumors (CPAT) were diagnosed. Brainstem auditory evoked potentials (BAEP), interaural time discrimination, and magnetic resonance imaging (MRI) revealed to be the most sensitive and efficient tests for the detection of CPAT. However, the investigation of otoneurological symptoms cannot be limited to BAEP and MRI on the a priori of a hypothetical CPAT. Electrophysiologic tests such as impedancemetry, and electronystagmographic testing are also needed to elucidate the many causes of otoneurological symptoms.
RP GUYOT, JP (通讯作者)，UNIV GENEVA,HOP CANTONAL,DEPT OTONEUROOPHTHALMOL,24 RUE MICHELI DU CREST,CH-1211 GENEVA 4,SWITZERLAND.
NR 0
TC 14
Z9 14
U1 1
U2 2
PU KARGER
PI BASEL
PA ALLSCHWILERSTRASSE 10, CH-4009 BASEL, SWITZERLAND
SN 0301-1569
J9 ORL J OTO-RHINO-LARY
JI ORL-J. Oto-Rhino-Laryngol. Relat. Spec.
PD MAY-JUN
PY 1992
VL 54
IS 3
BP 139
EP 143
DI 10.1159/000276283
PG 5
WC Otorhinolaryngology
WE Science Citation Index Expanded (SCI-EXPANDED)
SC Otorhinolaryngology
GA HY781
UT WOS:A1992HY78100006
PM 1518661
DA 2023-08-27
ER

PT J
AU van der Louw, EJTM
   Reddingius, RE
   Olieman, JF
   Neuteboom, RF
   Catsman-Berrevoets, CE
AF van der Louw, Elles J. T. M.
   Reddingius, Roel E.
   Olieman, Joanne F.
   Neuteboom, Rinze F.
   Catsman-Berrevoets, Coriene E.
TI Ketogenic diet treatment in recurrent diffuse intrinsic pontine glioma
   in children: A safety and feasibility study
SO PEDIATRIC BLOOD & CANCER
LA English
DT Article
DE brain tumor; coping; metabolic alteration; side effects
AB Background The mean overall survival rate of children with diffuse intrinsic pontine glioma (DIPG) is 9-11 months, with current standard treatment with fractionated radiotherapy and adjuvant chemotherapy. So far, novel therapeutic strategies have not yet resulted in significantly better survival. The main source of energy for glioblastoma cells is glucose. Therefore, metabolic alterations induced by the use of the extremely carbohydrate-restricted ketogenic diet (KD) as adjuvant therapy are subject of interest in cancer research. Procedure This study explores the safety and feasibility of the KD in children with recurrent DIPG and no remaining treatment options. Safety was defined as the number of adverse effects. Feasibility was defined as the number of patients who were able to use the KD for three months. Coping of patients and parents was measured with questionnaires. Results Three of 14 children referred to our hospital between 2010 and 2015 were included. Two patients completed the study, and one died before the end of the study. Hospitalizations were needed for placing a nasogastric tube (n = 1) and epileptic seizures (n = 1). Adverse effects related to the diet were mild and transient. Parents were highly motivated during the study. Conclusion Use of KD is safe and feasible, but the effect on survival has to be proven in a larger cohort of children who start the KD earlier after diagnosis, preferably as adjuvant therapy to fractionated radiotherapy.
C1 [van der Louw, Elles J. T. M.; Olieman, Joanne F.] Erasmus MC, Univ Med Ctr, Dept Dietet, Sophia Childrens Hosp, Wytemaweg 80, NL-3015 CN Rotterdam, Netherlands.
   [Reddingius, Roel E.] Princess Maxima Ctr Paediat Oncol, Utrecht, Netherlands.
   [Neuteboom, Rinze F.; Catsman-Berrevoets, Coriene E.] Erasmus MC, Univ Med Ctr, Dept Paediat Neurol, Sophia Childrens Hosp, Rotterdam, Netherlands.
C3 Erasmus University Rotterdam; Erasmus MC; Erasmus MC - Sophia Children's
   Hospital; Erasmus University Rotterdam; Erasmus MC; Erasmus MC - Sophia
   Children's Hospital
RP van der Louw, EJTM (通讯作者)，Erasmus MC, Univ Med Ctr, Dept Dietet, Sophia Childrens Hosp, Wytemaweg 80, NL-3015 CN Rotterdam, Netherlands.
EM
RI Ollieman, Joanne/AAQ-2237-2021
OI VAN DER LOUW, ELLES/0000-0003-3245-0471; Olieman,
   Joanne/0000-0003-4100-6030
FU Julians Foundation; Ouders van een Overleden Kind (VOOK)
FX This study was supported by the Julians Foundation and the Ouders van
   een Overleden Kind (VOOK).
CR Abdelwahab MG, 2012, PLOS ONE, V7, DOI 10.1371/journal.pone.0036197
   Chassot A, 2012, J NEURO-ONCOL, V106, P399, DOI 10.1007/s11060-011-0681-7
   Doolittle ND, 2007, J NEURO-ONCOL, V81, P81, DOI 10.1007/s11060-006-9209-y
   El-Khouly FE, 2017, FRONT ONCOL, V7, DOI 10.3389/fonc.2017.00254
   Green AL, 2015, CURR ONCOL REP, V17, DOI 10.1007/s11912-014-0436-7
   Hargrave DR, 2006, NEURO-ONCOLOGY, V8, P175, DOI 10.1215/15228517-2005-008
   Hurk van den TAM, 2010, DIETARY TREATMENT GU
   Kiebish MA, 2008, J LIPID RES, V49, P2545, DOI 10.1194/jlr.M800319-JLR200
   Kossoff E, 2018, EPILEPSIA OPEN, P1
   Martuscello RT, 2016, CLIN CANCER RES, V22, P2482, DOI 10.1158/1078-0432.CCR-15-0916
   Maurer GD, 2011, BMC CANCER, V11, DOI 10.1186/1471-2407-11-315
   National Cancer Institute, 2010, COMM TERM CRIT ADV E
   NEBELING LC, 1995, J AM COLL NUTR, V14, P202
   Robison NJ, 2014, PEDIATR BLOOD CANCER, V61, P636, DOI 10.1002/pbc.24794
   Schmidt M, 2011, NUTR METAB, V8, DOI 10.1186/1743-7075-8-54
   Seyfried Thomas N, 2005, Nutr Metab (Lond), V2, P30, DOI 10.1186/1743-7075-2-30
   Seyfried TN, 2015, FRONT CELL DEV BIOL, V3, DOI 10.3389/fcell.2015.00043
   Seyfried TN, 2003, BRIT J CANCER, V89, P1375, DOI 10.1038/sj.bjc.6601269
   Wood E, 2000, DEV MED CHILD NEUROL, V42, P292, DOI 10.1017/S0012162200000529
   Zuccoli G, 2010, NUTR METAB, V7, DOI 10.1186/1743-7075-7-33
NR 20
TC 16
Z9 16
U1 2
U2 26
PU WILEY
PI HOBOKEN
PA 111 RIVER ST, HOBOKEN 07030-5774, NJ USA
SN 1545-5009
EI 1545-5017
J9 PEDIATR BLOOD CANCER
JI Pediatr. Blood Cancer
PD MAR
PY 2019
VL 66
IS 3
AR e27561
DI 10.1002/pbc.27561
PG 7
WC Oncology; Hematology; Pediatrics
WE Science Citation Index Expanded (SCI-EXPANDED)
SC Oncology; Hematology; Pediatrics
GA HI8DN
UT WOS:000456685600014
PM 30484948
OA Green Published, hybrid
DA 2023-08-27
ER

PT J
AU Holzer, P
   Danzer, M
   Schicho, R
   Samberger, C
   Painsipp, E
   Lippe, IT
AF Holzer, P
   Danzer, M
   Schicho, R
   Samberger, C
   Painsipp, E
   Lippe, IT
TI Vagal afferent input from the acid-challenged rat stomach to the
   brainstem: Enhancement by interleukin-1 beta
SO NEUROSCIENCE
LA English
DT Article
DE acid insult; gastric mucosa; expression of c-Fos; nucleus tractus
   solitarii; proinflammatory cytokines; sensitization of vagal afferent
   pathways
ID MESSENGER-RNA EXPRESSION; C-FOS EXPRESSION; INDUCED SICKNESS BEHAVIOR;
   NECROSIS-FACTOR-ALPHA; EPITHELIAL PERMEABILITY; INTRAVENOUS-INJECTION;
   PROSTAGLANDIN E-2; SOLITARY TRACT; RECEPTOR; NUCLEUS
AB Exposure of the gastric mucosa to back-diffusing concentrations of HCI (0.25 M, pH 0.51) stimulates vagal afferent input to the brainstem. Here we have examined whether pretreatment of rats with the proinflammatory cytokines interleukin-1beta and tumor necrosis factor-alpha causes sensitization of vagal afferent pathways to HCI. Rats were pretreated i.p. with interleukin-1beta, tumor necrosis factor-alpha (10 mug/kg) or their vehicle (sterile saline) 24, 48 and 96 h before intragastric administration of HCI (0.25 M, 1 m1/100 g). Activation of neurons in the nucleus tractus solitarii was visualized by c-Fos immunohistochemistry 2 h after the HCI challenge. I.p. administration of interleukin-1beta and tumor necrosis factor-alpha alone induced c-Fos in the brainstem, an effect that was gone after 24 h. At this time, however, the effect of HCI to cause expression of c-Fos in the nucleus tractus solitarii was significantly enhanced by pretreatment with interleukin-1beta and tumor necrosis factor-alpha. The sensitizing effect of i.p.-administered interleukin-1beta was sustained for more than 48 h and prevented by the interleukin-1 receptor antagonist anakinra. Intracisternal administration of interleukin-1beta and tumor necrosis factor-alpha (100 ng) failed to amplify the HCI-evoked expression of c-Fos in the brainstem.
   These results show that peripheral administration of the proinflammatory cytokines interleukin-1beta and tumor necrosis factor-alpha induces prolonged sensitization of vagal afferent pathways to gastric HCI challenge. This effect seems to arise from a peripheral action on vagal afferents and may be of relevance to gastric chemonociception. 2004 Published by Elsevier Ltd on behalf of IBRO.
C1 Med Univ Graz, Dept Expt & Clin Pharmacol, A-8010 Graz, Austria.
C3 Medical University of Graz
RP Holzer, P (通讯作者)，Med Univ Graz, Dept Expt & Clin Pharmacol, Univ Pl 4, A-8010 Graz, Austria.
EM
RI Holzer, Peter/F-7578-2011
OI Holzer, Peter/0000-0002-5754-395X; Schicho, Rudolf/0000-0002-5726-4731
CR ALTSCHULER SM, 1989, J COMP NEUROL, V283, P248, DOI 10.1002/cne.902830207
   BLUTHE RM, 1995, BRAIN RES, V677, P171, DOI 10.1016/0006-8993(95)00194-U
   Bluthe RM, 2000, PHYSIOL BEHAV, V70, P367, DOI 10.1016/S0031-9384(00)00269-9
   BONHAM AC, 1993, AM J PHYSIOL, V264, pH1674, DOI 10.1152/ajpheart.1993.264.5.H1674
   BRADY LS, 1994, J NEUROSCI, V14, P4951
   Bucinskaite V, 1997, NEUROSCI LETT, V229, P33, DOI 10.1016/S0304-3940(97)00406-0
   CLARKE GD, 1978, J PHYSIOL-LONDON, V284, P55, DOI 10.1113/jphysiol.1978.sp012527
   Danzer M, 2004, EUR J NEUROSCI, V19, P85, DOI 10.1111/j.1460-9568.2004.03109.x
   Danzer M, 2004, AM J PHYSIOL-GASTR L, V286, pG403, DOI 10.1152/ajpgi.00308.2003
   Day HEW, 1996, NEUROENDOCRINOLOGY, V63, P207, DOI 10.1159/000126959
   Ek M, 2000, J COMP NEUROL, V428, P5, DOI 10.1002/1096-9861(20001204)428:1<5::AID-CNE2>3.0.CO;2-M
   Ek M, 1998, J NEUROSCI, V18, P9471
   ERICSSON A, 1995, J COMP NEUROL, V361, P681, DOI 10.1002/cne.903610410
   ERICSSON A, 1994, J NEUROSCI, V14, P897
   Fu LW, 1999, J PHYSIOL-LONDON, V521, P249, DOI 10.1111/j.1469-7793.1999.00249.x
   Gislason H, 1996, EUR SURG RES, V28, P278, DOI 10.1159/000129467
   Goehler LE, 2000, AUTON NEUROSCI-BASIC, V85, P49, DOI 10.1016/S1566-0702(00)00219-8
   Hillsley K, 1998, J PHYSIOL-LONDON, V509, P717, DOI 10.1111/j.1469-7793.1998.717bm.x
   Holzer P, 2002, NEUROGASTROENT MOTIL, V14, P459, DOI 10.1046/j.1365-2982.2002.00353.x
   KENT S, 1992, P NATL ACAD SCI USA, V89, P9117, DOI 10.1073/pnas.89.19.9117
   Konsman JP, 2002, TRENDS NEUROSCI, V25, P154, DOI 10.1016/S0166-2236(00)02088-9
   Konsman JP, 1999, NEUROSCIENCE, V89, P535, DOI 10.1016/S0306-4522(98)00368-6
   Kovacs KJ, 1998, NEUROCHEM INT, V33, P287, DOI 10.1016/S0197-0186(98)00023-0
   Kreis ME, 2000, GASTROENTEROLOGY, V118, pA431, DOI 10.1016/S0016-5085(00)83838-X
   Kurosawa M, 1997, J AUTONOM NERV SYST, V62, P72, DOI 10.1016/S0165-1838(96)00111-7
   Lamb K, 2003, GASTROENTEROLOGY, V125, P1410, DOI 10.1016/j.gastro.2003.07.010
   Matysiak-Budnik T, 2001, DIGEST DIS SCI, V46, P1558, DOI 10.1023/A:1010664626431
   Michl T, 2001, PAIN, V92, P19, DOI 10.1016/S0304-3959(00)00467-X
   Montuschi P, 1996, AM J PHYSIOL-GASTR L, V271, pG275, DOI 10.1152/ajpgi.1996.271.2.G275
   Niijima A, 1996, J AUTONOM NERV SYST, V61, P287, DOI 10.1016/S0165-1838(96)00098-7
   Niimi M, 1997, NEUROENDOCRINOLOGY, V66, P47, DOI 10.1159/000127218
   NORGREN R, 1988, J COMP NEUROL, V273, P207, DOI 10.1002/cne.902730206
   Opree A, 2000, J NEUROSCI, V20, P6289, DOI 10.1523/JNEUROSCI.20-16-06289.2000
   Parada CA, 2003, EUR J NEUROSCI, V17, P1847, DOI 10.1046/j.1460-9568.2003.02626.x
   Paton JFR, 2000, J AUTONOM NERV SYST, V80, P117, DOI 10.1016/S0165-1838(00)00077-1
   Paxinos G., 1998, RAT BRAIN STEREOTAXI, V4th
   Quan N, 1998, NEUROSCIENCE, V83, P281, DOI 10.1016/S0306-4522(97)00350-3
   RIVEST S, 1992, BRAIN RES, V587, P13, DOI 10.1016/0006-8993(92)91424-D
   Schuligoi R, 1998, GASTROENTEROLOGY, V115, P649, DOI 10.1016/S0016-5085(98)70144-1
   Taylor CT, 1998, GASTROENTEROLOGY, V114, P657, DOI 10.1016/S0016-5085(98)70579-7
   Traub RJ, 1996, NEUROSCIENCE, V74, P873, DOI 10.1016/0306-4522(96)00173-X
NR 41
TC 14
Z9 15
U1 0
U2 0
PU PERGAMON-ELSEVIER SCIENCE LTD
PI OXFORD
PA THE BOULEVARD, LANGFORD LANE, KIDLINGTON, OXFORD OX5 1GB, ENGLAND
SN 0306-4522
J9 NEUROSCIENCE
JI Neuroscience
PY 2004
VL 129
IS 2
BP 439
EP 445
DI 10.1016/j.neuroscience.2004.07.040
PG 7
WC Neurosciences
WE Science Citation Index Expanded (SCI-EXPANDED)
SC Neurosciences & Neurology
GA 869HE
UT WOS:000224973200017
PM 15501601
DA 2023-08-27
ER

PT J
AU Ben-Hur, H
   Kossoy, G
   Kossoy, N
   Zusman, I
AF Ben-Hur, H
   Kossoy, G
   Kossoy, N
   Zusman, I
TI Response of the immune system of mammary tumor-bearing rats to
   cyclophosphamide and soluble low-molecular-mass tumor-associated
   antigens: The bone marrow and thymus
SO INTERNATIONAL JOURNAL OF MOLECULAR MEDICINE
LA English
DT Article
DE bone marrow; mammary tumors; T cells; thymus; tumor-associated antigens
ID LINEAGE COMMITMENT; CELLS; THYMOCYTES; PROGRESSION; PROTEINS; THERAPY;
   CANCER; MICE; DRUG
AB We showed in a previous study that soluble low-molecular-mass tumor-associated antigens (sTAA) promote the anti-tumor effect of the anticancer drug cyclophosphamide (CPA) on rat mammary carcinogenesis. In this report, we analyzed the possible mechanism underlying this phenomenon. Studies were performed on the bone marrow and thymus from the following groups of rats: i) control rats, ii) rats treated with sTAA, iii) rats treated with CPA, iv) rats treated with CPA and sTAA. The cellular content of the bone marrow and thymus (CD4(+) and CD8(+) lymphocytes) was analyzed morphometrically and immunohistochemically. In the bone marrow, CPA caused significant substitution of cellular components with fatty tissue whereas sTAA repaired this process. We found that CPA affects mainly the process of myelogenesis whereas sTAA protect the production of lymphocytes. In the thymus, CPA alone or in combination with sTAA repaired the inhibition effect of DMBA on synthesis of CD4(+) and CD8(+) thymocytes. sTAA further increased the amount of CD8(+) T lymphocytes in the medulla of the thymus. Data in the literature as well as the findings presented here demonstrate that the tested treatment, including vaccination with sTAA, actively promotes the generation of the host's antitumor immune response.
C1 Hebrew Univ Jerusalem, Koret Sch Vet Med, Expt Oncol Lab, Fac Agr Food & Environm Qual Sci, IL-76100 Rehovot, Israel.
   Assaf Harofeh Med Ctr, Inst Pathol, Expt Med Lab, Tserifin, Israel.
C3 Hebrew University of Jerusalem; Shamir Medical Center (Assaf Harofeh);
   Tel Aviv University
RP Zusman, I (通讯作者)，Hebrew Univ Jerusalem, Koret Sch Vet Med, Expt Oncol Lab, Fac Agr Food & Environm Qual Sci, POB 12, IL-76100 Rehovot, Israel.
CR Barsukov V. S., 1997, Arkhiv Patologii, V59, P48
   Ben-Hur H, 2001, INT J ONCOL, V19, P407
   Braun S, 2001, ONCOLOGIST, V6, P125, DOI 10.1634/theoncologist.6-2-125
   De Veerman M, 1999, J IMMUNOL, V162, P144
   Diel IJ, 2001, SEMIN ONCOL, V28, P236, DOI 10.1053/sonc.2001.23489
   Ehrke MJ, 1998, INT J CANCER, V76, P579, DOI 10.1002/(SICI)1097-0215(19980518)76:4<579::AID-IJC22>3.0.CO;2-1
   Eichhorst ST, 2001, CANCER RES, V61, P243
   Feuerer M, 2001, NAT MED, V7, P452, DOI 10.1038/86523
   GALLO F, 1993, BREAST CANCER RES TR, V27, P221, DOI 10.1007/BF00665692
   King LB, 1999, INT IMMUNOL, V11, P1203, DOI 10.1093/intimm/11.8.1203
   Prockop S, 2000, SEMIN IMMUNOL, V12, P435, DOI 10.1006/smim.2000.0267
   Punt JA, 1996, J EXP MED, V184, P2091, DOI 10.1084/jem.184.6.2091
   Rossmann P, 1997, FOLIA MICROBIOL, V42, P277, DOI 10.1007/BF02819003
   Sandler B, 1999, EXP ONCOL, V21, P55
   Schwartz GN, 1998, BONE MARROW TRANSPL, V22, P457, DOI 10.1038/sj.bmt.1701364
   Seredenin S. B., 1999, Eksperimental'naya i Klinicheskaya Farmakologiya, V62, P51
   Singer A, 1999, SEMIN IMMUNOL, V11, P273, DOI 10.1006/smim.1999.0183
   Whiteside TL, 1999, CANCER IMMUNOL IMMUN, V48, P346, DOI 10.1007/s002620050585
   WHUTELAND JL, 1995, J HISTOCHEM CYTOCHEM, V43, P313
   ZALESKIS G, 1995, ONCOL RES, V7, P307
NR 20
TC 6
Z9 6
U1 0
U2 1
PU PROFESSOR D A SPANDIDOS
PI ATHENS
PA 1, S MERKOURI ST, EDITORIAL OFFICE,, ATHENS 116 35, GREECE
SN 1107-3756
J9 INT J MOL MED
JI Int. J. Mol. Med.
PD OCT
PY 2002
VL 10
IS 4
BP 517
EP 521
PG 5
WC Medicine, Research & Experimental
WE Science Citation Index Expanded (SCI-EXPANDED)
SC Research & Experimental Medicine
GA 596JT
UT WOS:000178162100026
PM 12239604
DA 2023-08-27
ER

PT J
AU Borner, T
   Arnold, M
   Ruud, J
   Breit, SN
   Langhans, W
   Lutz, TA
   Blomqvist, A
   Riediger, T
AF Borner, Tito
   Arnold, Myrtha
   Ruud, Johan
   Breit, Samuel N.
   Langhans, Wolfgang
   Lutz, Thomas A.
   Blomqvist, Anders
   Riediger, Thomas
TI Anorexia-cachexia syndrome in hepatoma tumour-bearing rats requires the
   area postrema but not vagal afferents and is paralleled by increased
   MIC-1/GDF15
SO JOURNAL OF CACHEXIA SARCOPENIA AND MUSCLE
LA English
DT Article
DE Cancer; Food intake; Energy balance; Brainstem; Muscle; AP-lesion;
   Subdiaphragmatic vagal deafferentation
ID MACROPHAGE INHIBITORY CYTOKINE-1; CANCER-INDUCED ANOREXIA; FOOD-INTAKE;
   NECROSIS-FACTOR; BODY-WEIGHT; MIC-1; LIPOPOLYSACCHARIDE; AVERSIONS;
   SERUM; CHOLECYSTOKININ
AB Background The cancer-anorexia-cachexia syndrome (CACS) negatively affects survival and therapy success in cancer patients. Inflammatory mediators and tumour-derived factors are thought to play an important role in the aetiology of CACS. However, the central and peripheral mechanisms contributing to CACS are insufficiently understood. The area postrema (AP) and the nucleus tractus solitarii are two important brainstem centres for the control of eating during acute sickness conditions. Recently, the tumour-derived macrophage inhibitory cytokine-1 (MIC-1) emerged as a possible mediator of cancer anorexia because lesions of these brainstem areas attenuated the anorectic effect of exogenous MIC-1 in mice.
   Methods Using a rat hepatoma tumour model, we examined the roles of the AP and of vagal afferents in the mediation of CACS. Specifically, we investigated whether a lesion of the AP (APX) or subdiaphragmatic vagal deafferentation (SDA) attenuate anorexia, body weight, muscle, and fat loss. Moreover, we analysed MIC-1 levels in this tumour model and their correlation with tumour size and the severity of the anorectic response.
   Results In tumour-bearing sham-operated animals mean daily food intake significantly decreased. The anorectic response was paralleled by a significant loss of body weight and muscle mass. APX rats were protected against anorexia, body weight loss, and muscle atrophy after tumour induction. In contrast, subdiaphragmatic vagal deafferentation did not attenuate cancer-induced anorexia or body weight loss. Tumour-bearing rats had substantially increased MIC-1 levels, which positively correlated with tumour size and cancer progression and negatively correlated with food intake.
   Conclusions These findings demonstrate the importance of the AP in the mediation of cancer-dependent anorexia and body weight loss and support a pathological role of MIC-1 as a tumour-derived factor mediating CACS, possibly via an AP-dependent action.
C1 [Borner, Tito; Lutz, Thomas A.; Riediger, Thomas] Univ Zurich, Inst Vet Physiol, Vetsuisse Fac, Zurich, Switzerland.
   [Borner, Tito; Langhans, Wolfgang; Lutz, Thomas A.; Riediger, Thomas] Univ Zurich, Zurich Ctr Integrat Human Physiol ZIHP, Zurich, Switzerland.
   [Arnold, Myrtha; Langhans, Wolfgang] Swiss Fed Inst Technol, Physiol & Behav Lab, Schwerzenbach, Switzerland.
   [Ruud, Johan; Blomqvist, Anders] Univ Linkoping, Dept Clin & Expt Med, Linkoping, Sweden.
   [Breit, Samuel N.] Univ New South Wales, St Vincents Hosp, St Vincents Ctr Appl Med Res, Sydney, NSW, Australia.
C3 University of Zurich; University of Zurich; Zurich Center Integrative
   Human Physiology (ZIHP); Swiss Federal Institutes of Technology Domain;
   ETH Zurich; Linkoping University; St Vincents Hospital Sydney;
   University of New South Wales Sydney
RP Riediger, T (通讯作者)，Univ Zurich, Inst Vet Physiol, Winterthurerstr 260, CH-8057 Zurich, Switzerland.
EM
RI Ruud, Johan/W-7407-2019; Breit, Samuel N/B-6177-2008
OI Borner, Tito/0000-0001-9438-695X
FU Swiss National Science Foundation [31003A-135541]; Krebsliga Zurich;
   Forschungskredit University of Zurich; Swiss National Science Foundation
   (SNF) [31003A_135541] Funding Source: Swiss National Science Foundation
   (SNF)
FX This study was funded by the Swiss National Science Foundation (#
   31003A-135541), Krebsliga Zurich, Forschungskredit University of Zurich.
CR Arnold M, 2006, J NEUROSCI, V26, P11052, DOI 10.1523/JNEUROSCI.2606-06.2006
   Asarian L, 2013, AM J PHYSIOL-REG I, V305, pR1215, DOI 10.1152/ajpregu.00446.2012
   Bauskin AR, 2006, CANCER RES, V66, P4983, DOI 10.1158/0008-5472.CAN-05-4067
   Bennani-Baiti N, 2011, SUPPORT CARE CANCER, V19, P1451, DOI 10.1007/s00520-010-0972-0
   BERNSTEIN IL, 1985, AM J PHYSIOL, V249, pR296, DOI 10.1152/ajpregu.1985.249.3.R296
   Bernstein IL, 1996, NEUROSCI BIOBEHAV R, V20, P177, DOI 10.1016/0149-7634(95)00046-H
   BERNSTEIN IL, 1991, AM J PHYSIOL, V260, pR906, DOI 10.1152/ajpregu.1991.260.5.R906
   Bluthe RM, 1996, NEUROREPORT, V7, P1485, DOI 10.1097/00001756-199606170-00008
   BLUTHE RM, 1994, CR ACAD SCI III-VIE, V317, P499
   Borner T, 2016, AM J PHYSL REGUL INT
   Braegger FE, 2014, EUR J NEUROSCI, V40, P3055, DOI 10.1111/ejn.12672
   Breit SN, 2011, GROWTH FACTORS, V29, P187, DOI 10.3109/08977194.2011.607137
   Brown DA, 2003, CLIN CANCER RES, V9, P2642
   Burckart K, 2010, CURR OPIN CLIN NUTR, V13, P410, DOI 10.1097/MCO.0b013e328339fdd2
   EDWARDS GL, 1986, AM J PHYSIOL, V251, pR971, DOI 10.1152/ajpregu.1986.251.5.R971
   Evans WJ, 2008, CLIN NUTR, V27, P793, DOI 10.1016/j.clnu.2008.06.013
   Fairlie WD, 1999, J LEUKOCYTE BIOL, V65, P2, DOI 10.1002/jlb.65.1.2
   Ferrari B, 2005, AM J PHYSIOL-REG I, V289, pR1027, DOI 10.1152/ajpregu.00736.2004
   Gordon JN, 2005, QJM-INT J MED, V98, P779, DOI 10.1093/qjmed/hci127
   Grill HJ, 2009, INT J OBESITY, V33, pS11, DOI 10.1038/ijo.2009.10
   Heiden MGV, 2009, SCIENCE, V324, P1029, DOI 10.1126/science.1160809
   Ihnatko R, 2013, BRIT J CANCER, V109, P1867, DOI 10.1038/bjc.2013.525
   Johnen H, 2007, NAT MED, V13, P1333, DOI 10.1038/nm1677
   Koopmann J, 2004, CLIN CANCER RES, V10, P2386, DOI 10.1158/1078-0432.CCR-03-0165
   Langhans W, 1999, NEUROPEPTIDES, V33, P415, DOI 10.1054/npep.1999.0048
   Laviano A, 2005, NAT CLIN PRACT ONCOL, V2, P158, DOI 10.1038/ncponc0112
   Laviano A, 2003, LANCET ONCOL, V4, P686, DOI 10.1016/S1470-2045(03)01247-6
   Lerner L, 2015, J CACHEXIA SARCOPENI, V6, P317, DOI 10.1002/jcsm.12033
   Lu ZH, 2014, PLOS ONE, V9, DOI 10.1371/journal.pone.0088553
   Lutz TA, 1998, PEPTIDES, V19, P309, DOI 10.1016/S0196-9781(97)00292-1
   Maier SF, 1998, ANN NY ACAD SCI, V840, P289, DOI 10.1111/j.1749-6632.1998.tb09569.x
   MARESPERLMAN JA, 1988, CANCER RES, V48, P602
   McCarthy D O, 2000, Biol Res Nurs, V1, P287, DOI 10.1177/109980040000100405
   Moran TH, 1997, AM J PHYSIOL-REG I, V272, pR1245
   NORGREN R, 1994, AM J PHYSIOL-REG I, V267, pR1136, DOI 10.1152/ajpregu.1994.267.4.R1136
   NORGREN R, 1988, J COMP NEUROL, V273, P207, DOI 10.1002/cne.902730206
   Plata-Salaman C, 2001, INT J OBESITY, V25, pS48, DOI 10.1038/sj.ijo.0801911
   Porter MH, 1998, AM J PHYSIOL-REG I, V275, pR384, DOI 10.1152/ajpregu.1998.275.2.R384
   POWLEY TL, 1987, AM J PHYSIOL, V253, pR361, DOI 10.1152/ajpregu.1987.253.2.R361
   Riediger T, 2010, PHARMACOL BIOCHEM BE, V97, P112, DOI 10.1016/j.pbb.2010.04.015
   Ruud J, 2007, J COMP NEUROL, V504, P275, DOI 10.1002/cne.21407
   Schwartz GJ, 1997, AM J PHYSIOL-REG I, V273, pR1193, DOI 10.1152/ajpregu.1997.273.3.R1193
   Skipworth RJE, 2010, BRIT J CANCER, V102, P665, DOI 10.1038/sj.bjc.6605532
   Staff AC, 2010, GYNECOL ONCOL, V118, P237, DOI 10.1016/j.ygyno.2010.05.032
   Talbert E, 2014, J CACHEXIA SARCOPENI, V5, P321, DOI 10.1007/s13539-014-0141-2
   Tisdale MJ, 2009, PHYSIOL REV, V89, P381, DOI 10.1152/physrev.00016.2008
   Tisdale MJ, 2002, NAT REV CANCER, V2, P862, DOI 10.1038/nrc927
   Tisdale MJ, 1997, J NATL CANCER I, V89, P1763, DOI 10.1093/jnci/89.23.1763
   Tisdale MJ, 2000, NUTRITION, V16, P1013, DOI 10.1016/S0899-9007(00)00409-3
   Tsai VWW, 2015, PLOS ONE, V10, DOI 10.1371/journal.pone.0133362
   Tsai VWW, 2014, PLOS ONE, V9, DOI 10.1371/journal.pone.0100370
   VIGIER D, 1979, ARCH ITAL BIOL, V117, P325
   von Haehling S, 2015, J CACHEXIA SARCOPENI, V6, P315, DOI 10.1002/jcsm.12089
   WALLS EK, 1995, AM J PHYSIOL-REG I, V269, pR1279, DOI 10.1152/ajpregu.1995.269.5.R1279
   WEINGARTEN S, 1993, PHYSIOL BEHAV, V54, P961, DOI 10.1016/0031-9384(93)90309-4
   Welsh JB, 2003, P NATL ACAD SCI USA, V100, P3410, DOI 10.1073/pnas.0530278100
   Zimmers TA, 2008, J CANCER RES CLIN, V134, P753, DOI 10.1007/s00432-007-0336-4
NR 57
TC 26
Z9 26
U1 0
U2 3
PU WILEY
PI HOBOKEN
PA 111 RIVER ST, HOBOKEN 07030-5774, NJ USA
SN 2190-6009
J9 J CACHEXIA SARCOPENI
JI J. Caxhexia Sarcopenia Muscle
PD JUN
PY 2017
VL 8
IS 3
BP 417
EP 427
DI 10.1002/jcsm.12169
PG 11
WC Geriatrics & Gerontology; Medicine, General & Internal
WE Science Citation Index Expanded (SCI-EXPANDED)
SC Geriatrics & Gerontology; General & Internal Medicine
GA EY0PG
UT WOS:000403659900006
PM 28025863
OA Green Published, gold
DA 2023-08-27
ER

PT J
AU Feng, RL
   Zhang, T
   Ke, CX
   Tao, YP
   Wang, Y
AF Feng, Runlin
   Zhang, Tao
   Ke, Changxing
   Tao, Yanping
   Wang, Yan
TI Mixed epithelial and stromal tumor of the kidney: A retrospective
   clinicopathological evaluation
SO ANNALS OF DIAGNOSTIC PATHOLOGY
LA English
DT Article
DE Kidney neoplasms; Mixed epithelial and stromal tumor; Clinicopathology;
   Immunohistochemistry
ID CYSTIC NEPHROMA; URINARY SYSTEM
AB Objective: Mixed epithelial and stromal tumor of the kidney (MESTK) is a rare renal tumor for which there is limited clinical case experience. The aim of this study was to investigate the clinicopathological features, imaging manifestations, immunohistochemical phenotype, diagnosis, and differential diagnosis of MESTK.Methods: We systematically evaluated the clinicopathological characteristics of 13 patients diagnosed with MESTK from 2016 to 2022. Results: There were 12 female cases and 1 male case, and their age ranged from 24 to 62 (mean age 47 years). The tumor was located in the left kidney in 11 patients. Seven of these patients were located in the upper pole of the left kidney. The mean diameter of the tumor was 4.6 cm (range 1.3-8.6 cm), and the main body of the tumor was mainly located in the medulla or medulla. In 7 cases, the tumors were clearly outlined, 4 cases had clear but irregular borders, and in 3 cases, the masses broke through the renal capsule but did not invade the surrounding organs. All tumors had varying proportions of cystic and solid components. In most tumors, the epithelial cell component was predominant. Among the epithelial components, we observed a predominance of large cysts. Small cysts and tubular structures were also seen; the least common were papillary structures. The most common types of lining epithelium were flat and cuboidal. The cell cytoplasm was predominantly eosinophilic with hyaline degeneration. The tumor cells were composed of different proportions of sparse hypocellular areas and dense hypercellular areas between these cells. In most cases, the cellular stroma had an ovarian-like appearance. It is characterized histologically by mature adipocytes, thick-walled blood vessels, and chronic inflammatory cells. Calcification and collagenization were seen in a few cases. In all female patients, estrogen and progesterone receptors in the interstitial component were almost always positively expressed. In male patients, all interstitial components showed locally positive expression of androgens.Conclusion: MESTK had unique characteristics with complex and variable lesion shapes. There was a high degree of overlap with cystic kidney cancer, and the rate of missed and misdiagnosis was extremely high. The diagnosis could not be confirmed by preoperative imaging. The final diagnosis depended on pathomorphology.
C1 [Feng, Runlin; Wang, Yan] Kunming Med Univ, Affiliated Hosp 2, Dept Pathol, Kunming 650101, Yunnan, Peoples R China.
   [Zhang, Tao; Ke, Changxing] Kunming Med Univ, Affiliated Hosp 2, Dept Urol, Kunming 650101, Yunnan, Peoples R China.
   [Tao, Yanping] Third Peoples Hosp Kunming, Dept Emergency, Kunming 650000, Yunnan, Peoples R China.
   [Tao, Yanping] Third Peoples Hosp Kunming, Dept Emergency, Kunming, Peoples R China.
   [Wang, Yan] Kunming Med Univ, Affiliated Hosp 2, Dept Pathol, Kunming, Peoples R China.
C3 Kunming Medical University; Kunming Medical University; Kunming Medical
   University
RP Tao, YP (通讯作者)，Third Peoples Hosp Kunming, Dept Emergency, Kunming, Peoples R China.; Wang, Y (通讯作者)，Kunming Med Univ, Affiliated Hosp 2, Dept Pathol, Kunming, Peoples R China.
EM
OI , zhang tao/0000-0002-3848-4632
FU Fund for the in -hospital Project of the Second Affiliated Hospital of
   Kunming Medical University in Yunnan Province [2021yk015]
FX This research was supported by the Fund for the in -hospital Project of
   the Second Affiliated Hospital of Kunming Medical University in Yunnan
   Province (Approval No: 2021yk015) .
CR Adsay NV, 2000, AM J SURG PATHOL, V24, P958, DOI 10.1097/00000478-200007000-00007
   Calio A, 2016, AM J SURG PATHOL, V40, P1538, DOI 10.1097/PAS.0000000000000733
   Chuang CK, 2018, UROL CASE REP, V17, P122, DOI 10.1016/j.eucr.2017.11.025
   Ferdjallah A, 2019, PEDIATR TRANSPLANT, V23, DOI 10.1111/petr.13575
   Huang HW, 2021, TRANSL CANCER RES, V10, P4256, DOI 10.21037/tcr-21-308
   Jiang H, 2022, SAGE OPEN MED CASE R, V10, DOI 10.1177/2050313X211068646
   Kum JBJ, 2011, AM J SURG PATHOL, V35, P1114, DOI 10.1097/PAS.0b013e3182233fb6
   Kuroda N, 2008, APMIS, V116, P1013, DOI 10.1111/j.1600-0463.2008.01063.x
   Li YJ, 2017, AM J SURG PATHOL, V41, P472, DOI 10.1097/PAS.0000000000000816
   Magnelli LL, 2021, ABDOM RADIOL, V46, P696, DOI 10.1007/s00261-020-02687-0
   Mai KT, 2007, PATHOLOGY, V39, P235, DOI 10.1080/00313020701230799
   Michal M, 1998, PATHOL RES PRACT, V194, P445, DOI 10.1016/S0344-0338(98)80038-1
   Minoda R, 2017, MOL CLIN ONCOL, V7, P1005, DOI 10.3892/mco.2017.1444
   Moch H, 2022, EUR UROL, V82, P458, DOI 10.1016/j.eururo.2022.06.016
   Moch H, 2016, EUR UROL, V70, P93, DOI 10.1016/j.eururo.2016.02.029
   Montironi R, 2016, EUR UROL, V70, P120, DOI 10.1016/j.eururo.2016.03.011
   Pitra T, 2022, CANCERS, P14
   Suzuki T, 2013, INT J UROL, V20, P448, DOI 10.1111/j.1442-2042.2012.03155.x
   Tian JL, 2019, SAUDI MED J, V40, P499, DOI 10.15537/smj.2019.5.24177
   Tsukamoto S, 2022, FRONT PHARMACOL, V13, DOI 10.3389/fphar.2022.885457
   Turbiner J, 2007, AM J SURG PATHOL, V31, P489, DOI 10.1097/PAS.0b013e31802bdd56
   Vujanic GM, 2018, NAT REV UROL
   Wang CJ, 2013, WORLD J SURG ONCOL, V11, DOI 10.1186/1477-7819-11-207
   Yang FY, 2017, INT BRAZ J UROL, V43, P563, DOI 10.1590/S1677-5538.IBJU.2015.0300
   Zou L, 2014, INT J CLIN EXP PATHO, V7, P2658
NR 25
TC 0
Z9 0
U1 0
U2 0
PU ELSEVIER SCIENCE INC
PI NEW YORK
PA STE 800, 230 PARK AVE, NEW YORK, NY 10169 USA
SN 1092-9134
EI 1532-8198
J9 ANN DIAGN PATHOL
JI Ann. Diagn. Pathol.
PD APR
PY 2023
VL 63
AR 152088
DI 10.1016/j.anndiagpath.2022.152088
EA DEC 2022
PG 10
WC Pathology
WE Science Citation Index Expanded (SCI-EXPANDED)
SC Pathology
GA E4FS4
UT WOS:000975123300001
PM 36577187
DA 2023-08-27
ER

PT J
AU Chevaleyre, C
   Kereselidze, D
   Caille, F
   Tournier, N
   Olaciregui, NG
   Winkeler, A
   Decleves, X
   Jego, B
   Cisternino, S
   Auvity, S
   Truillet, C
AF Chevaleyre, Celine
   Kereselidze, Dimitri
   Caille, Fabien
   Tournier, Nicolas
   Olaciregui, Nagore G.
   Winkeler, Alexandra
   Decleves, Xavier
   Jego, Benoit
   Cisternino, Salvatore
   Auvity, Sylvain
   Truillet, Charles
TI TSPO PET Imaging as a Potent Non-Invasive Biomarker for Diffuse
   Intrinsic Pontine Glioma in a Patient-Derived Orthotopic Rat Model
SO INTERNATIONAL JOURNAL OF MOLECULAR SCIENCES
LA English
DT Article
DE DIPG biomarker; TSPO-PET imaging; PDOX
ID TRANSLOCATOR PROTEIN; MANAGEMENT; SURVIVAL; TARGET; TUMORS; MRI
AB Diffuse intrinsic pontine gliomas (DIPG), the first cause of cerebral pediatric cancer death, will greatly benefit from specific and non-invasive biomarkers for patient follow-up and monitoring of drug efficacy. Since biopsies are challenging for brain tumors, molecular imaging may be a technique of choice to target and follow tumor evolution. So far, MR remains the imaging technique of reference for DIPG, although it often fails to define the extent of tumors, an essential parameter for therapeutic efficacy assessment. Thanks to its high sensitivity, positron emission tomography (PET) offers a unique way to target specific biomarkers in vivo. We demonstrated in a patient-derived orthotopic xenograft (PDOX) model in the rat that the translocator protein of 18 kDa (TSPO) may be a promising biomarker for monitoring DIPG tumors. We studied the distribution of 18F-DPA-714, a TSPO radioligand, in rats inoculated with HSJD-DIPG-007 cells. The primary DIPG human cell line HSJD-DIPG-007 highly represents this pediatric tumor, displaying the most prevalent DIPG mutations, H3F3A (K27M) and ACVR1 (R206H). Kinetic modeling and parametric imaging using the brain 18F-DPA-714 PET data enabled specific delineation of the DIPG tumor area, which is crucial for radiotherapy dose management.
C1 [Chevaleyre, Celine; Kereselidze, Dimitri; Caille, Fabien; Tournier, Nicolas; Winkeler, Alexandra; Jego, Benoit; Auvity, Sylvain; Truillet, Charles] Univ Paris Saclay, INSERM, CNRS, BioMaps,Serv Hosp Frederic Joliot,CEA, 4 Pl Gen Leclerc, F-91401 Orsay, France.
   [Olaciregui, Nagore G.] St Joan de Deu Hosp, Inst Recerca, Carrer St Quinti 77, Barcelona 08041, Spain.
   [Decleves, Xavier; Cisternino, Salvatore; Auvity, Sylvain] Univ Paris Cite, Inserm UMR S1144, F-75006 Paris, France.
   [Cisternino, Salvatore; Auvity, Sylvain] Hop Univ Necker Enfants Malad, AP HP, 149 Rue Sevres, F-75015 Paris, France.
C3 CEA; Centre National de la Recherche Scientifique (CNRS); Institut
   National de la Sante et de la Recherche Medicale (Inserm); UDICE-French
   Research Universities; Universite Paris Saclay; Autonomous University of
   Barcelona; Hospital Universitari Vall d'Hebron; Vall d'Hebron Institut
   de Recerca (VHIR); Institut National de la Sante et de la Recherche
   Medicale (Inserm); UDICE-French Research Universities; Universite Paris
   Cite; Assistance Publique Hopitaux Paris (APHP); Hopital Universitaire
   Paul-Brousse - APHP; UDICE-French Research Universities; Universite
   Paris Cite; Hopital Universitaire Cochin - APHP; Hopital Universitaire
   Hotel-Dieu - APHP; Hopital Universitaire Necker-Enfants Malades - APHP;
   Hopital Universitaire Saint-Louis - APHP
RP Truillet, C (通讯作者)，Univ Paris Saclay, INSERM, CNRS, BioMaps,Serv Hosp Frederic Joliot,CEA, 4 Pl Gen Leclerc, F-91401 Orsay, France.
EM
RI Truillet, Charles/ACX-1954-2022; DECLEVES, Xavier/S-6978-2016;
   Cisternino, Salvatore/S-8714-2016; TOURNIER, Nicolas/N-9277-2017
OI DECLEVES, Xavier/0000-0002-9526-2294; Cisternino,
   Salvatore/0000-0001-8500-3574; TOURNIER, Nicolas/0000-0002-0755-2030;
   Winkeler, Alexandra/0000-0003-3397-2570
FU CEA "Bottom up" [ANR-11-INBS-0006]
FX This work was performed on a platform member of France Life Imaging
   network (grant ANR-11-INBS-0006). The authors acknowledge the financial
   support from the CEA "Bottom up" Grant REGIME.
CR Alghamri MS, 2021, FRONT PHARMACOL, V12, DOI 10.3389/fphar.2021.680021
   Ammer LM, 2020, CANCERS, V12, DOI 10.3390/cancers12102973
   Aziz-Bose R, 2019, CURR OPIN ONCOL, V31, P522, DOI 10.1097/CCO.0000000000000577
   Banati RB, 2020, SCI REP-UK, V10, DOI 10.1038/s41598-020-67036-z
   Betlazar C, 2021, FRONT CELL DEV BIOL, V9, DOI 10.3389/fcell.2021.715444
   Caretti V, 2011, BRAIN PATHOL, V21, P441, DOI 10.1111/j.1750-3639.2010.00468.x
   Castel D, 2015, ACTA NEUROPATHOL, V130, P815, DOI 10.1007/s00401-015-1478-0
   Chaves C, 2020, PHARMACEUTICS, V12, DOI 10.3390/pharmaceutics12050399
   Cistaro A, 2021, CURR ONCOL, V28, P2481, DOI 10.3390/curroncol28040226
   Cohen KJ, 2017, NEURO-ONCOLOGY, V19, P1025, DOI 10.1093/neuonc/nox021
   Damont A, 2008, J LABELLED COMPD RAD, V51, P286, DOI 10.1002/jlcr.1523
   Duclos V, 2021, INT J MOL SCI, V22, DOI 10.3390/ijms22084159
   Feldman AT, 2014, METHODS MOL BIOL, V1180, P31, DOI 10.1007/978-1-4939-1050-2_3
   Galldiks N, 2019, NEURO-ONCOL ADV, V1, DOI 10.1093/noajnl/vdz010
   Gallitto M, 2019, ADV RADIAT ONCOL, V4, P520, DOI 10.1016/j.adro.2019.03.009
   Goda JS, 2013, PEDIATR NEUROSURG, V49, P274, DOI 10.1159/000366167
   Guilarte Tomas R, 2022, Pharmacol Ther, V234, P108048, DOI 10.1016/j.pharmthera.2021.108048
   Guo N, 2012, MOL IMAGING BIOL, V14, P743, DOI 10.1007/s11307-012-0541-7
   Hankinson TC, 2011, J NEUROSURG-PEDIATR, V8, P97, DOI 10.3171/2011.4.PEDS1180
   Haumann R., 2022, TRANSL MED COMMUN, V7, P8, DOI [10, DOI 10.1186/S41231-022-00115-7]
   Hoffman LM, 2018, J CLIN ONCOL, V36, P1963, DOI 10.1200/JCO.2017.75.9308
   Jain P, 2020, J NUCL MED, V61, P1107, DOI 10.2967/jnumed.119.229443
   Kluiver TA, 2020, FRONT ONCOL, V10, DOI 10.3389/fonc.2020.00092
   Koopman T, 2020, J CEREBR BLOOD F MET, V40, P895, DOI 10.1177/0271678X19851878
   Kossatz S, 2017, CANCER RES, V77, P2112, DOI 10.1158/0008-5472.CAN-16-2850
   Lin GL, 2018, ACTA NEUROPATHOL COM, V6, DOI 10.1186/s40478-018-0553-x
   Logan J, 2000, NUCL MED BIOL, V27, P661, DOI 10.1016/S0969-8051(00)00137-2
   Louis DN, 2021, NEURO-ONCOLOGY, V23, P1231, DOI 10.1093/neuonc/noab106
   Mathew RK, 2018, J KOREAN NEUROSURG S, V61, P343, DOI 10.3340/jkns.2018.0008
   Mohammad F, 2017, NAT MED, V23, P483, DOI 10.1038/nm.4293
   Morana G, 2020, THERANOSTICS, V10, P11881, DOI 10.7150/thno.50598
   Pigeon H, 2019, NEURO-ONCOLOGY, V21, P755, DOI 10.1093/neuonc/noz029
   Pottier G, 2017, J CEREBR BLOOD F MET, V37, P1163, DOI 10.1177/0271678X16685105
   R Core Team, 2012, R R PROJ STAT COMP
   Roncaroli F, 2016, CLIN TRANSL IMAGING, V4, P145, DOI 10.1007/s40336-016-0168-9
   Schiffer WK, 2006, J NEUROSCI METH, V155, P272, DOI 10.1016/j.jneumeth.2006.01.027
   Shoshan-Barmatz V, 2019, INT J MOL SCI, V20, DOI 10.3390/ijms20133348
   Srikanthan Dilakshan, 2021, Chin Neurosurg J, V7, P6, DOI 10.1186/s41016-020-00218-w
   Su ZJ, 2015, J NUCL MED, V56, P512, DOI 10.2967/jnumed.114.151621
   Unterrainer M, 2020, EUR J NUCL MED MOL I, V47, P1368, DOI 10.1007/s00259-019-04491-5
   Van Gool Stefaan W, 2020, Medicines (Basel), V7, DOI 10.3390/medicines7050029
   Warren KE, 2012, FRONT ONCOL, V2, DOI 10.3389/fonc.2012.00205
   Zinnhardt B, 2021, EUR J NUCL MED MOL I, V49, P174, DOI 10.1007/s00259-021-05276-5
   Zukotynski KA, 2017, J NUCL MED, V58, P1264, DOI 10.2967/jnumed.116.185389
NR 44
TC 0
Z9 0
U1 2
U2 4
PU MDPI
PI BASEL
PA ST ALBAN-ANLAGE 66, CH-4052 BASEL, SWITZERLAND
EI 1422-0067
J9 INT J MOL SCI
JI Int. J. Mol. Sci.
PD OCT
PY 2022
VL 23
IS 20
AR 12476
DI 10.3390/ijms232012476
PG 12
WC Biochemistry & Molecular Biology; Chemistry, Multidisciplinary
WE Science Citation Index Expanded (SCI-EXPANDED)
SC Biochemistry & Molecular Biology; Chemistry
GA 5P2IC
UT WOS:000872979700001
PM 36293329
OA Green Published, gold
DA 2023-08-27
ER

PT J
AU Tomita, T
   Frassanito, P
AF Tomita, Tadanori
   Frassanito, Paolo
TI Tumors of the superior medullary velum in infancy and childhood: report
   of 6 cases Clinical article
SO JOURNAL OF NEUROSURGERY-PEDIATRICS
LA English
DT Article
DE atypical teratoid/rhabdoid tumor; cerebellar tumor; occipital
   transtentorial approach; pineal tumor; quadrigeminal plate; superior
   medullary velum; oncology
ID INTERHEMISPHERIC TRANSTENTORIAL APPROACH; PINEAL TUMORS;
   MEDULLOBLASTOMA; CEREBELLUM; CHILDREN; DISEASE; FOSSA
AB Object. The superior medullary velum (SMV) is a thin lamina of white matter located between the superior cerebellar peduncles horizontally and between the midbrain and cerebellum vertically. The SMV has not previously been described as the primary location of a posterior fossa tumor, although it can be secondarily invaded by a tumor from the cerebellum or quadrigeminal plate. This paper aims to define clinical and radiological features of tumors primarily arising from the SMV during childhood.
   Methods. The authors observed 6 infants and children harboring neoplasms of the SMV who were treated at Ann & Robert Lurie Children's Hospital of Chicago (formerly Children's Memorial Hospital) in Chicago, Illinois. Pathological diagnosis of the neoplasms was an atypical teratoid/rhabdoid tumor (ATRT) in 5 patients, and a juvenile pilocytic astrocytoma (JPA) in the remaining child. The tumors were diagnosed during infancy in all patients, with ages ranging from 3 months to 10 months, except for the patient with a JPA (diagnosed at 5 years old). All patients presented with signs and symptoms of increased intracranial pressure due to obstructive hydrocephalus.
   Results. Characteristic MRI features were noted, consistent with a mass in both the fourth ventricle and the cerebellomesencephalic fissure and quadrigeminal cistern, resulting in the circumferential displacement of the neural structures surrounding the SMV. The tumor was removed effectively in gross-total fashion through the occipital transtentorial approach in all patients. This approach offers a wide exposure of the region. However, all infants with ATRT suffered tumor dissemination and died between 4 and 11 months after diagnosis, in spite of radical resection and oncological treatment. The 1 child with JPA is alive and well 30 months after tumor resection.
   Conclusions. To the best of the authors' knowledge, this is the first description in the literature that focuses on tumors originating from the SMV. This entity must be promptly recognized on preoperative radiological studies to carefully plan the subsequent surgical and clinical management. (http://thejns.org/doi/abs/10.3171/2012.9.PEDS12236)
C1 [Tomita, Tadanori] Ann & Robert H Lurie Childrens Hosp Chicago, Div Pediat Neurosurg, Chicago, IL 60611 USA.
   Northwestern Univ, Feinberg Sch Med, Chicago, IL 60611 USA.
C3 Ann & Robert H. Lurie Children's Hospital of Chicago; Northwestern
   University; Feinberg School of Medicine
RP Tomita, T (通讯作者)，Ann & Robert H Lurie Childrens Hosp Chicago, Div Pediat Neurosurg, 225 E Chicago Ave,Box 28, Chicago, IL 60611 USA.
EM
RI FRASSANITO, PAOLO/K-4099-2018
OI FRASSANITO, PAOLO/0000-0003-0071-3126
CR [Anonymous], 1995, GRAYS ANATOMY
   Baryawno N, 2010, CELL CYCLE, V9, P2548, DOI 10.4161/cc.9.13.12170
   Cardenas RJ, 2011, J CLIN NEUROSCI, V18, P128, DOI 10.1016/j.jocn.2010.04.053
   Cho BK, 1998, CHILD NERV SYST, V14, P53, DOI 10.1007/s003810050175
   Crosby E, 1962, CORRELATIVE ANATOMY
   HALLONET MER, 1990, DEVELOPMENT, V108, P19
   Jagani Z, 2010, NAT MED, V16, P1429, DOI 10.1038/nm.2251
   Kurokawa Y, 1999, SURG NEUROL, V51, P421, DOI 10.1016/S0090-3019(98)00123-2
   LANG J, 1991, ACTA NEUROCHIR, V111, P147, DOI 10.1007/BF01400505
   Louvi A, 2003, DEVELOPMENT, V130, P5319, DOI 10.1242/dev.00736
   Millen KJ, 2008, CURR OPIN NEUROBIOL, V18, P12, DOI 10.1016/j.conb.2008.05.010
   MULLER F, 1988, ANAT EMBRYOL, V179, P55, DOI 10.1007/BF00305100
   O'Brien DF, 2006, J NEUROSURG, V105, P219, DOI 10.3171/ped.2006.105.3.219
   Parisi MA, 2003, MOL GENET METAB, V80, P36, DOI 10.1016/j.ymgme.2003.08.010
   RAIMONDI AJ, 1982, CHILD BRAIN, V9, P239
   Ramelli GP, 2011, J CHILD NEUROL, V26, P377, DOI 10.1177/0883073810382141
   Rhoton AL, 2000, NEUROSURGERY, V47, pS69, DOI 10.1097/00006123-200009001-00012
   Rhoton AL, 2000, NEUROSURGERY, V47, pS29, DOI 10.1097/00006123-200009001-00010
   Santi L, 2000, CHILD NERV SYST, V16, P129, DOI 10.1007/s003810050478
   Sato T, 2009, DEVELOPMENT, V136, P3617, DOI 10.1242/dev.041210
   Tanaka R, 2003, OPER TECH NEUROSURG, V6, P215, DOI [10.1016/j.otns.2003.09.001, DOI 10.1016/J.OTNS.2003.09.001]
   Ternier J, 2006, J NEUROSURG, V104, P369, DOI 10.3171/ped.2006.104.6.369
   Tomita T, 2008, PRINCIPLES PRACTICE, P585
   Trasimeni G, 2008, CHILD NERV SYST, V24, P1037, DOI 10.1007/s00381-008-0635-3
   Tubbs RS, 2004, CLIN ANAT, V17, P82, DOI 10.1002/ca.10185
   Venneti S, 2011, AM J SURG PATHOL, V35, P1463, DOI 10.1097/PAS.0b013e318224d2cd
   von Hoff K, 2011, PEDIATR BLOOD CANCER, V57, P978, DOI 10.1002/pbc.23236
   Yamamoto I, 2001, J NEURO-ONCOL, V54, P263, DOI 10.1023/A:1012790213818
NR 28
TC 8
Z9 9
U1 0
U2 2
PU AMER ASSOC NEUROLOGICAL SURGEONS
PI ROLLING MEADOWS
PA 5550 MEADOWBROOK DRIVE, ROLLING MEADOWS, IL 60008 USA
SN 1933-0707
J9 J NEUROSURG-PEDIATR
JI J. Neurosurg.-Pediatr.
PD JAN
PY 2013
VL 11
IS 1
BP 52
EP 59
DI 10.3171/2012.9.PEDS12236
PG 8
WC Clinical Neurology; Pediatrics; Surgery
WE Science Citation Index Expanded (SCI-EXPANDED)
SC Neurosciences & Neurology; Pediatrics; Surgery
GA 062VP
UT WOS:000312951700009
PM 23121115
DA 2023-08-27
ER

PT J
AU Fukuchi, K
   Suzuki, M
   Sato, S
   Iseki, J
AF Fukuchi, Kazuki
   Suzuki, Makoto
   Sato, Shinsuke
   Iseki, Jouji
TI Unusually increased metaiodobenzylguanidine uptake in a gastrointestinal
   stromal tumor of the stomach
SO ANNALS OF NUCLEAR MEDICINE
LA English
DT Article
DE Iodine-123 MIBG; Gastrointestinal stromal tumor; Scintigraphy; Gamma
   camera
ID INTERSTITIAL-CELLS; CAJAL; PHEOCHROMOCYTOMA; CARCINOMA; SCAN; GIST
AB A 70-year-old woman with a submucosal gastric tumor was referred to our hospital for surgical treatment. Upon examination, it was found that she had hypertension, and abdominal computed tomography revealed swelling on both adrenal glands. The patient was examined with gamma camera imaging and iodine-123 metaiodobenzylguanidine (MIBG), because her hypertension was thought to be due to a suspected adrenomedullary tumor. The planar image showed an unexpected abnormal uptake of MIBG in the upper abdomen. On single-photon emission computed tomographic images, the area of abnormal tracer uptake was thought to correspond to the known gastric tumor. The surgical procedure and histological assessments revealed that the gastric tumor was a gastrointestinal stromal tumor (GIST). MIBG can accumulate in GISTs as well as in neuroendocrine tumors of the medulla of the adrenal glands. Although the cause of radiolabeled MIBG uptake in GISTs is uncertain, further studies are necessary to establish the significance of MIBG scintigraphy in GIST imaging.
C1 [Fukuchi, Kazuki] Shizuoka Prefectural Gen Hosp, Dept Radiol, Shizuoka 4208527, Japan.
   [Suzuki, Makoto] Shizuoka Prefectural Gen Hosp, Dept Pathol, Shizuoka 4208527, Japan.
   [Sato, Shinsuke; Iseki, Jouji] Shizuoka Prefectural Gen Hosp, Dept Surg, Shizuoka 4208527, Japan.
C3 Shizuoka General Hospital; Shizuoka General Hospital; Shizuoka General
   Hospital
RP Fukuchi, K (通讯作者)，Shizuoka Prefectural Gen Hosp, Dept Radiol, Kita Ando 4-27-1, Shizuoka 4208527, Japan.
EM
OI Sato, Shinsuke/0000-0002-6644-1066; suzuki, makoto/0009-0007-4761-4877
CR Balan KK, 2010, CLIN NUCL MED, V35, P196, DOI 10.1097/RLU.0b013e3181cc63cb
   BEIERWALTES WH, 1991, J NUCL MED, V32, P1627
   Corless CL, 2004, J CLIN ONCOL, V22, P3813, DOI 10.1200/JCO.2004.05.140
   Debiec-Rychter M, 2002, MODERN PATHOL, V15, P692, DOI 10.1097/01.MP.0000017564.15834.1F
   Frappaz D, 1997, MED PEDIATR ONCOL, V29, P589, DOI 10.1002/(SICI)1096-911X(199712)29:6<589::AID-MPO14>3.0.CO;2-1
   Inoue Y, 2002, BRIT J RADIOL, V75, P548, DOI 10.1259/bjr.75.894.750548
   Jun JY, 2004, BRIT J PHARMACOL, V141, P670, DOI 10.1038/sj.bjp.0705665
   Kindblom LG, 1998, AM J PATHOL, V152, P1259
   Min KW, 2006, J CELL MOL MED, V10, P995, DOI 10.1111/j.1582-4934.2006.tb00541.x
   Rainis T, 2000, J CLIN ENDOCR METAB, V85, P5, DOI 10.1210/jc.85.1.5
   SISSON JC, 1981, NEW ENGL J MED, V305, P12, DOI 10.1056/NEJM198107023050103
   Sone H, 1996, HORM RES, V46, P138, DOI 10.1159/000185010
   TOBES MC, 1985, J NUCL MED, V26, P897
   Ward SM, 2006, J PHYSIOL-LONDON, V576, P675, DOI 10.1113/jphysiol.2006.117390
NR 14
TC 2
Z9 2
U1 0
U2 0
PU SPRINGER
PI NEW YORK
PA 233 SPRING ST, NEW YORK, NY 10013 USA
SN 0914-7187
J9 ANN NUCL MED
JI Ann. Nucl. Med.
PD OCT
PY 2012
VL 26
IS 8
BP 684
EP 687
DI 10.1007/s12149-012-0612-5
PG 4
WC Radiology, Nuclear Medicine & Medical Imaging
WE Science Citation Index Expanded (SCI-EXPANDED)
SC Radiology, Nuclear Medicine & Medical Imaging
GA 024CJ
UT WOS:000310085000012
PM 22644561
DA 2023-08-27
ER

PT J
AU Pitskhelauri, DI
   Konovalov, AN
   Kornienko, VN
   Serova, NK
   Arutiunov, NV
   Kopachev, DN
AF Pitskhelauri, David I.
   Konovalov, Alexander N.
   Kornienko, Valeri N.
   Serova, Natalia K.
   Arutiunov, Nikita V.
   Kopachev, Dmitry N.
TI INTRAOPERATIVE DIRECT THIRD VENTRICULOSTOMY AND AQUEDUCTAL STENTING IN
   DEEP-SEATED MIDLINE BRAIN TUMOR SURGERY
SO NEUROSURGERY
LA English
DT Article
DE Aqueductal stenting; Hydrocephalus; Midline tumors; Ventriculostomy
ID POSTERIOR-FOSSA TUMORS; PINEAL REGION TUMORS; LAMINA TERMINALIS;
   ENDOSCOPIC AQUEDUCTOPLASTY; OBSTRUCTIVE HYDROCEPHALUS; THALAMIC TUMORS;
   MANAGEMENT; FENESTRATION; CHILDREN; FAILURE
AB OBJECTIVE: Surgical resection of deep-seated midline brain tumors does not always resolve obstruction of cerebrospinal fluid pathways, and an additional operation-ventricular shunting-is required. To prevent postoperative obstructive hydrocephalus, we combine tumor removal and internal ventricular shunting in 1 stage.
   METHODS: Between 2000 and 2006, 82 patients with deep-seated midline brain tumors (tumors of the third ventricle, pineal region, thalamus, upper brainstem, and superior half of the fourth ventricle) underwent 84 tumor resections with intraoperative internal ventricular shunting. Two types of intraoperative shunting were performed: direct third ventriculostomy with fenestration of the premammillary membrane of the third ventricle floor and Liliequist's membrane, 53 operations; and aqueductal stenting, 30 operations. In 1 patient, third ventriculostomy and aqueductal stenting were performed simultaneously.
   RESULTS: As most of the tumors had an infiltrative growth pattern, gross total tumor removal was achieved in only 31% of patients in this series. There were no fatal outcomes related to the surgery. Follow-up data were collected in 73 patients (89%) and ranged from 2 to 68 months (median, 16 months). Additional shunting because of inadequate function of stoma or stent was performed in 13 patients at various times after surgery (median, 30 days). The Kaplan-Meier survival analysis demonstrated that at 12 and 24 months the intraoperative direct third ventriculostomy success rates were 67 and 61%, respectively; aqueductal stenting success rates were 93% at both 12 and 24 months.
   CONCLUSION: Intraoperative direct third ventriculostomy and aqueductal stenting under direct visual control were found to be reliable methods of hydrocephalus management in patients with deep-seated midline brain tumors.
C1 [Pitskhelauri, David I.; Konovalov, Alexander N.; Kornienko, Valeri N.; Serova, Natalia K.; Arutiunov, Nikita V.; Kopachev, Dmitry N.] NN Burdenko Inst Neurosurg, Moscow 125047, Russia.
C3 Russian Academy of Medical Sciences; N.N.Burdenko National Medical
   Research Center of Neurosurgery
RP Pitskhelauri, DI (通讯作者)，NN Burdenko Inst Neurosurg, 4 Tverskaya Yamskaya St 16, Moscow 125047, Russia.
EM
RI Serova, Natalia/ABB-5451-2020; Konovalov, Alexander/ABG-6541-2021;
   Pitzkhelauri, David/AAO-6736-2020
CR Akyuz M, 2006, ACTA NEUROCHIR, V148, P725, DOI 10.1007/s00701-006-0738-0
   APUZZO MLJ, 1993, BRAIN SURG COMPLICAT, P486
   BERNSTEIN M, 1984, J NEUROSURG, V61, P649, DOI 10.3171/jns.1984.61.4.0649
   Brockmeyer D, 1998, PEDIATR NEUROSURG, V28, P236, DOI 10.1159/000028657
   Bruce JN, 1993, BRAIN SURG COMPLICAT, P511
   BRUCE JN, 2006, OPERATIVE NEUROSURGI, P786
   Buxton N, 2001, SURG NEUROL, V55, P74, DOI 10.1016/S0090-3019(01)00352-4
   CHANG SM, 1995, NEUROSURGERY, V37, P383, DOI 10.1227/00006123-199509000-00003
   Cinalli G, 1999, J NEUROSURG, V90, P448, DOI 10.3171/jns.1999.90.3.0448
   Cuccia V, 1997, CHILD NERV SYST, V13, P514, DOI 10.1007/s003810050128
   CULLEY DJ, 1994, NEUROSURGERY, V34, P402, DOI 10.1227/00006123-199403000-00003
   DANDY WE, 1922, J HOPKINS HOSP B, V33, P189
   DELATORRE E, 1963, J Neurosurg, V20, P461
   DIAS MS, 1989, PEDIATR NEUROSCI, V15, P283
   ELVIDGE AR, 1966, J NEUROSURG, V24, P11, DOI 10.3171/jns.1966.24.1.0011
   Ersahin Y, 2007, CHILD NERV SYST, V23, P143, DOI 10.1007/s00381-006-0227-z
   Feng HL, 2004, J NEUROSURG, V100, P626, DOI 10.3171/jns.2004.100.4.0626
   Fritsch MJ, 2004, CHILD NERV SYST, V20, P137, DOI 10.1007/s00381-003-0860-8
   Gaab MR, 1998, J NEUROSURG, V88, P496, DOI 10.3171/jns.1998.88.3.0496
   Gangemi M, 1999, MINIM INVAS NEUROSUR, V42, P128, DOI 10.1055/s-2008-1053384
   GEFFEN G, 1980, BRAIN, V103, P773, DOI 10.1093/brain/103.4.773
   Grant JA, 1997, SURG NEUROL, V47, P210, DOI 10.1016/S0090-3019(96)00457-0
   Grunert P, 2003, MINIM INVAS NEUROSUR, V46, P16, DOI 10.1055/s-2003-37957
   HIGASHI K, 1979, SHINKEI GEKA, V11, P1109
   Hopf NJ, 1999, NEUROSURGERY, V44, P795, DOI 10.1097/00006123-199904000-00062
   Komotar RJ, 2002, NEUROSURGERY, V51, P1403, DOI 10.1097/00006123-200212000-00010
   Konovalov AN, 2003, SURG NEUROL, V59, P250, DOI 10.1016/S0090-3019(03)00080-6
   Konovalov AN, 2004, TREATMENT PINEAL REG
   KORSHUNOV AE, 2004, ZH VOPR NEIROKHIR IM, V4, P18
   Lapras C, 1980, Neurochirurgie, V26, P1
   LEE M, 1994, PEDIATR NEUROSURG, V20, P240, DOI 10.1159/000120797
   Leksell L, 1949, ACTA PSYCH NEUROL, V24, P559, DOI 10.1111/j.1600-0447.1949.tb07339.x
   Lutterbach J, 2002, NEUROSURGERY, V51, P44, DOI 10.1097/00006123-200207000-00006
   McGirt MJ, 2002, PEDIATR NEUROSURG, V36, P248, DOI 10.1159/000058428
   Melikian A G, 2002, Zh Vopr Neirokhir Im N N Burdenko, P5
   Mohanty A, 2001, NEUROSURGERY, V49, P531
   MOLLOY PT, 1995, NEUROLOGY, V45, P1897, DOI 10.1212/WNL.45.10.1897
   NAZZARO JM, 1992, J NEUROSURG, V76, P746, DOI 10.3171/jns.1992.76.5.0746
   Norlen G, 1949, ACTA PSYCH NEUROL, V24, P629
   Omay SB, 2006, OPERATIVE NEUROSURGI, P753
   PAPO I, 1982, NEUROSURGERY, V10, P13, DOI 10.1227/00006123-198201000-00002
   PENDL G, 1990, NEUROSURGERY, V26, P641, DOI 10.1227/00006123-199004000-00013
   PIATT JH, 1995, PEDIATR NEUROSURG, V23, P133, DOI 10.1159/000120950
   Piepmeier J, 1993, BRAIN SURG COMPLICAT, P581
   Pitskhelauri D I, 2004, Zh Vopr Neirokhir Im N N Burdenko, P28
   PUDENZ RH, 1991, SURG NEUROL, V35, P200, DOI 10.1016/0090-3019(91)90072-H
   RAIMONDI AJ, 1981, J NEUROSURG, V55, P174, DOI 10.3171/jns.1981.55.2.0174
   Ray Pulak, 2005, Neurosurg Focus, V19, pE8
   Rickert CH, 1998, NEUROSURGERY, V42, P1378, DOI 10.1097/00006123-199806000-00118
   Sainte-Rose C, 2001, J NEUROSURG, V95, P791, DOI 10.3171/jns.2001.95.5.0791
   SAINTEROSE C, 1992, PEDIATR NEUROSURG, V17, P2, DOI 10.1159/000120557
   SCHMID UD, 1986, J NEUROSURG, V65, P649, DOI 10.3171/jns.1986.65.5.0649
   Schroeder HWS, 2004, CHILD NERV SYST, V20, P821, DOI 10.1007/s00381-004-0937-z
   Steiger HJ, 2000, ACTA NEUROCHIR, V142, P1327, DOI 10.1007/s007010070001
   THOMSON AF, 1951, HIST NEUROLOGICAL SU, P136
   Tomasello F, 1999, NEUROSURGERY, V45, P827, DOI 10.1097/00006123-199910000-00018
   TURNBULL IM, 1966, J NEUROSURG, V24, P24, DOI 10.3171/jns.1966.24.1.0024
   VANDERTOP WP, 1992, NEUROSURGERY, V31, P186, DOI 10.1227/00006123-199208000-00003
   Vandertop WP, 2001, J NEUROSURG, V95, P919
   Yamini B, 2004, J NEUROSURG, V100, P437, DOI 10.3171/ped.2004.100.5.0437
NR 60
TC 7
Z9 10
U1 0
U2 1
PU OXFORD UNIV PRESS INC
PI CARY
PA JOURNALS DEPT, 2001 EVANS RD, CARY, NC 27513 USA
SN 0148-396X
EI 1524-4040
J9 NEUROSURGERY
JI Neurosurgery
PD FEB
PY 2009
VL 64
IS 2
BP 256
EP 266
DI 10.1227/01.NEU.0000338260.05545.84
PG 11
WC Clinical Neurology; Surgery
WE Science Citation Index Expanded (SCI-EXPANDED)
SC Neurosciences & Neurology; Surgery
GA 399JY
UT WOS:000262797400033
PM 19190455
DA 2023-08-27
ER

PT J
AU Tomita, Y
   Shimazu, Y
   Somasundaram, A
   Tanaka, Y
   Takata, N
   Ishi, Y
   Gadd, S
   Hashizume, R
   Angione, A
   Pinero, G
   Hambardzumyan, D
   Brat, DJ
   Hoeman, CM
   Becher, OJ
AF Tomita, Yusuke
   Shimazu, Yosuke
   Somasundaram, Agila
   Tanaka, Yoshihiro
   Takata, Nozomu
   Ishi, Yukitomo
   Gadd, Samantha
   Hashizume, Rintaro
   Angione, Angelo
   Pinero, Gonzalo
   Hambardzumyan, Dolores
   Brat, Daniel J.
   Hoeman, Christine M.
   Becher, Oren J.
TI A novel mouse model of diffuse midline glioma initiated in neonatal
   oligodendrocyte progenitor cells highlights cell-of-origin dependent
   effects of H3K27M
SO GLIA
LA English
DT Article
DE diffuse intrinsic pontine glioma; diffuse midline glioma;
   oligodendrocyte progenitor cells
ID TRANSCRIPTION FACTOR OLIG2; CENTRAL-NERVOUS-SYSTEM; PEDIATRIC
   HIGH-GRADE; POSTNATAL-GROWTH; GENE-EXPRESSION; HISTONE H3.3; MUTATION;
   GLIOMAGENESIS; SUBGROUPS; CHILDREN
AB Diffuse midline glioma (DMG) is a type of lethal brain tumor that develops mainly in children. The majority of DMG harbor the K27M mutation in histone H3. Oligodendrocyte progenitor cells (OPCs) in the brainstem are candidate cells-of-origin for DMG, yet there is no genetically engineered mouse model of DMG initiated in OPCs. Here, we used the RCAS/Tv-a avian retroviral system to generate DMG in Olig2-expressing progenitors and Nestin-expressing progenitors in the neonatal mouse brainstem. PDGF-A or PDGF-B overexpression, along with p53 deletion, resulted in gliomas in both models. Exogenous overexpression of H3.3K27M had a significant effect on tumor latency and tumor cell proliferation when compared with H3.3WT in Nestin+ cells but not in Olig2+ cells. Further, the fraction of H3.3K27M-positive cells was significantly lower in DMGs initiated in Olig2+ cells relative to Nestin+ cells, both in PDGF-A and PDGF-B-driven models, suggesting that the requirement for H3.3K27M is reduced when tumorigenesis is initiated in Olig2+ cells. RNA-sequencing analysis revealed that the differentially expressed genes in H3.3K27M tumors were non-overlapping between Olig2;PDGF-B, Olig2;PDGF-A, and Nestin;PDGF-A models. GSEA analysis of PDGFA tumors confirmed that the transcriptomal effects of H3.3K27M are cell-of-origin dependent with H3.3K27M promoting epithelial-to-mesenchymal transition (EMT) and angiogenesis when Olig2 marks the cell-of-origin and inhibiting EMT and angiogenesis when Nestin marks the cell-of-origin. We did observe some overlap with H3.3K27M promoting negative enrichment of TNFA_Signaling_Via_NFKB in both models. Our study suggests that the tumorigenic effects of H3.3K27M are cell-of-origin dependent, with H3.3K27M being more oncogenic in Nestin+ cells than Olig2+ cells.
C1 [Tomita, Yusuke; Shimazu, Yosuke; Ishi, Yukitomo; Hashizume, Rintaro; Hoeman, Christine M.; Becher, Oren J.] Northwestern Univ, Dept Pediat, Feinberg Sch Med, Chicago, IL 60611 USA.
   [Tomita, Yusuke] Hiroshima City Hiroshima Citizens Hosp, Dept Neurosurg & Neuroendovasc Surg, Hiroshima, Japan.
   [Somasundaram, Agila; Hashizume, Rintaro; Becher, Oren J.] Ann & Robert H Lurie Childrens Hosp Chicago, Div Hematol Oncol & Stem Cell Transplant, Chicago, IL 60611 USA.
   [Tanaka, Yoshihiro] Northwestern Univ, Feinberg Sch Med, Dept Prevent Med, Chicago, IL 60611 USA.
   [Tanaka, Yoshihiro] Northwestern Univ, Ctr Arrhythmia Res, Dept Cardiol, Feinberg Sch Med, Chicago, IL 60611 USA.
   [Takata, Nozomu] Northwestern Univ, Feinberg Cardiovasc & Renal Res Inst FCVRRI, Ctr Vasc & Dev Biol, Chicago, IL 60611 USA.
   [Takata, Nozomu] Northwestern Univ, Simpson Querrey Inst BioNanotechnol, Chicago, IL 60611 USA.
   [Gadd, Samantha] Ann & Robert H Lurie Childrens Hosp Chicago, Dept Pathol, Chicago, IL 60611 USA.
   [Hashizume, Rintaro; Becher, Oren J.] Northwestern Univ, Feinberg Sch Med, Dept Biochem & Mol Genet, Chicago, IL 60611 USA.
   [Angione, Angelo; Pinero, Gonzalo; Hambardzumyan, Dolores] Mt Sinai Sch Med, Dept Neurosurg & Oncol Sci, New York, NY USA.
   [Brat, Daniel J.] Northwestern Univ, Dept Pathol, Feinberg Sch Med, Chicago, IL 60611 USA.
   [Becher, Oren J.] Mt Sinai Kravis Childrens Hosp, Jack Martin Div Pediat Hematol Oncol, New York, NY USA.
C3 Northwestern University; Feinberg School of Medicine; Hiroshima City
   Hospital; Ann & Robert H. Lurie Children's Hospital of Chicago;
   Northwestern University; Feinberg School of Medicine; Northwestern
   University; Feinberg School of Medicine; Northwestern University;
   Northwestern University; Ann & Robert H. Lurie Children's Hospital of
   Chicago; Northwestern University; Feinberg School of Medicine; Icahn
   School of Medicine at Mount Sinai; Northwestern University; Feinberg
   School of Medicine
RP Becher, OJ (通讯作者)，Northwestern Univ, Dept Pediat, Feinberg Sch Med, Chicago, IL 60611 USA.
EM
RI Ishi, Yukitomo/ISU-2030-2023; Angione, Angelo/ISV-2923-2023; Becher,
   Oren/GXN-3269-2022
OI Ishi, Yukitomo/0000-0003-4834-2619; Becher, Oren/0000-0003-4016-8492;
   Gadd, Samantha/0000-0001-5033-8585; Tanaka,
   Yoshihiro/0000-0002-6150-1592; Takata, Nozomu/0000-0003-0171-6994;
   Shimazu, Yosuke/0000-0002-7696-7811; Angione, Angelo/0000-0001-7713-5833
FU Foundation for the National Institutes of Health [K02 NS086917, R01
   CA197313, R21 1R21NS114431-01]
FX Foundation for the National Institutes of Health, Grant/Award Numbers:
   K02 NS086917, R01 CA197313, R21 1R21NS114431-01
CR Anderson JL, 2017, NEURO-ONCOLOGY, V19, P1068, DOI 10.1093/neuonc/now299
   Barton KL, 2013, PLOS ONE, V8, DOI 10.1371/journal.pone.0077639
   Becher OJ, 2010, CANCER RES, V70, P2548, DOI 10.1158/0008-5472.CAN-09-2503
   Bechet D, 2014, ACTA NEUROPATHOL, V128, P733, DOI 10.1007/s00401-014-1337-4
   Bender S, 2013, CANCER CELL, V24, P660, DOI 10.1016/j.ccr.2013.10.006
   Bhat KPL, 2013, CANCER CELL, V24, P331, DOI 10.1016/j.ccr.2013.08.001
   Brien GL, 2021, NAT GENET, V53, P1221, DOI 10.1038/s41588-021-00897-w
   Buczkowicz P, 2015, FRONT ONCOL, V5, DOI 10.3389/fonc.2015.00147
   Buczkowicz P, 2014, NAT GENET, V46, P451, DOI 10.1038/ng.2936
   Chan KM, 2013, GENE DEV, V27, P985, DOI 10.1101/gad.217778.113
   Cordero FJ, 2017, MOL CANCER RES, V15, P1243, DOI 10.1158/1541-7786.MCR-16-0389
   Khuong-Quang DA, 2012, ACTA NEUROPATHOL, V124, P439, DOI 10.1007/s00401-012-0998-0
   Encinas JM, 2006, P NATL ACAD SCI USA, V103, P8233, DOI 10.1073/pnas.0601992103
   Filbin MG, 2018, SCIENCE, V360, P331, DOI 10.1126/science.aao4750
   Funato K, 2014, SCIENCE, V346, P1529, DOI 10.1126/science.1253799
   Haag D, 2021, CANCER CELL, V39, P407, DOI 10.1016/j.ccell.2021.01.005
   Hargrove D, 2006, LANCET ONCOL, V7, P241, DOI 10.1016/S1470-2045(06)70615-5
   Hoeman CM, 2019, NAT COMMUN, V10, DOI 10.1038/s41467-019-08823-9
   Larson JD, 2019, CANCER CELL, V35, P140, DOI 10.1016/j.ccell.2018.11.015
   Lewis PW, 2013, SCIENCE, V340, P857, DOI 10.1126/science.1232245
   Lindquist RA, 2016, NAT COMMUN, V7, DOI 10.1038/ncomms11628
   Louis DN, 2016, ACTA NEUROPATHOL, V131, P803, DOI 10.1007/s00401-016-1545-1
   Mackay A, 2017, CANCER CELL, V32, P520, DOI 10.1016/j.ccell.2017.08.017
   Mehta S, 2011, CANCER CELL, V19, P359, DOI 10.1016/j.ccr.2011.01.035
   Mikkelsen TS, 2007, NATURE, V448, P553, DOI 10.1038/nature06008
   Misuraca KL, 2016, NEOPLASIA, V18, P60, DOI 10.1016/j.neo.2015.12.002
   Misuraca KL, 2014, ACTA NEUROPATHOL COM, V2, DOI 10.1186/s40478-014-0134-6
   Mohammad F, 2017, NAT MED, V23, P483, DOI 10.1038/nm.4293
   Monje M, 2011, P NATL ACAD SCI USA, V108, P4453, DOI 10.1073/pnas.1101657108
   Nagaraja S, 2017, CANCER CELL, V31, P635, DOI 10.1016/j.ccell.2017.03.011
   Nordby Y, 2017, SCI REP-UK, V7, DOI 10.1038/srep43378
   Paugh BS, 2011, J CLIN ONCOL, V29, P3999, DOI 10.1200/JCO.2011.35.5677
   Puget S, 2012, PLOS ONE, V7, DOI 10.1371/journal.pone.0030313
   Ross JL, 2021, BRAIN, V144, P53, DOI 10.1093/brain/awaa382
   Sanders LM, 2020, GIGASCIENCE, V9, DOI 10.1093/gigascience/giaa136
   Schuller U, 2008, CANCER CELL, V14, P123, DOI 10.1016/j.ccr.2008.07.005
   Schwartzentruber J, 2012, NATURE, V482, P226, DOI [10.1038/nature10833, 10.1007/s13311-012-0137-6]
   Singh D, 2002, CANCER CELL, V1, P203, DOI 10.1016/S1535-6108(02)00030-2
   Tate MC, 2015, J COMP NEUROL, V523, P449, DOI 10.1002/cne.23690
   Wagner S, 2006, BRIT J CANCER, V95, P991, DOI 10.1038/sj.bjc.6603402
   Walker AS, 2010, NEURAL PLAST, V2010, DOI 10.1155/2010/894374
   WESTERMARK B, 1995, GLIA, V15, P257, DOI 10.1002/glia.440150307
   Wu G, 2012, NAT GENET, V44, P251, DOI 10.1038/ng.1102
NR 43
TC 7
Z9 7
U1 0
U2 6
PU WILEY
PI HOBOKEN
PA 111 RIVER ST, HOBOKEN 07030-5774, NJ USA
SN 0894-1491
EI 1098-1136
J9 GLIA
JI Glia
PD SEP
PY 2022
VL 70
IS 9
BP 1681
EP 1698
DI 10.1002/glia.24189
EA MAY 2022
PG 18
WC Neurosciences
WE Science Citation Index Expanded (SCI-EXPANDED)
SC Neurosciences & Neurology
GA 2T6YM
UT WOS:000791795000001
PM 35524725
OA hybrid, Green Published
DA 2023-08-27
ER

PT J
AU Gupta, N
   Goumnerova, LC
   Manley, P
   Chi, SN
   Neuberg, D
   Puligandla, M
   Fangusaro, J
   Goldman, S
   Tomita, T
   Alden, T
   DiPatri, A
   Rubin, JB
   Gauvain, K
   Limbrick, D
   Leonard, J
   Geyer, JR
   Leary, S
   Browd, S
   Wang, ZH
   Sood, S
   Bendel, A
   Nagib, M
   Gardner, S
   Karajannis, MA
   Harter, D
   Ayyanar, K
   Gump, W
   Bowers, DC
   Weprin, B
   MacDonald, TJ
   Aguilera, D
   Brahma, B
   Robison, NJ
   Kiehna, E
   Krieger, M
   Sandler, E
   Aldana, P
   Khatib, Z
   Ragheb, J
   Bhatia, S
   Mueller, S
   Banerjee, A
   Bredlau, AL
   Gururangan, S
   Fuchs, H
   Cohen, KJ
   Jallo, G
   Dorris, K
   Handler, M
   Comito, M
   Dias, M
   Nazemi, K
   Baird, L
   Murray, J
   Lindeman, N
   Hornick, JL
   Malkin, H
   Sinai, C
   Greenspan, L
   Wright, KD
   Prados, M
   Bandopadhayay, P
   Ligon, KL
   Kieran, MW
AF Gupta, Nalin
   Goumnerova, Liliana C.
   Manley, Peter
   Chi, Susan N.
   Neuberg, Donna
   Puligandla, Maneka
   Fangusaro, Jason
   Goldman, Stewart
   Tomita, Tadanori
   Alden, Tord
   DiPatri, Arthur
   Rubin, Joshua B.
   Gauvain, Karen
   Limbrick, David
   Leonard, Jeffrey
   Geyer, J. Russel
   Leary, Sarah
   Browd, Samuel
   Wang, Zhihong
   Sood, Sandeep
   Bendel, Anne
   Nagib, Mahmoud
   Gardner, Sharon
   Karajannis, Matthias A.
   Harter, David
   Ayyanar, Kanyalakshmi
   Gump, William
   Bowers, Daniel C.
   Weprin, Bradley
   MacDonald, Tobey J.
   Aguilera, Dolly
   Brahma, Barunashish
   Robison, Nathan J.
   Kiehna, Erin
   Krieger, Mark
   Sandler, Eric
   Aldana, Philipp
   Khatib, Ziad
   Ragheb, John
   Bhatia, Sanjiv
   Mueller, Sabine
   Banerjee, Anu
   Bredlau, Amy-Lee
   Gururangan, Sri
   Fuchs, Herbert
   Cohen, Kenneth J.
   Jallo, George
   Dorris, Kathleen
   Handler, Michael
   Comito, Melanie
   Dias, Mark
   Nazemi, Kellie
   Baird, Lissa
   Murray, Jeff
   Lindeman, Neal
   Hornick, Jason L.
   Malkin, Hayley
   Sinai, Claire
   Greenspan, Lianne
   Wright, Karen D.
   Prados, Michael
   Bandopadhayay, Pratiti
   Ligon, Keith L.
   Kieran, Mark W.
TI Prospective feasibility and safety assessment of surgical biopsy for
   patients with newly diagnosed diffuse intrinsic pontine glioma
SO NEURO-ONCOLOGY
LA English
DT Article
DE bevacizumab; DIPG; erlotinib; stereotactic biopsy; temozolomide
ID ACTIVATING ACVR1 MUTATIONS; BRAIN-STEM GLIOMA; CHILDREN
AB Background. Diagnosis of diffuse intrinsic pontine glioma (DIPG) has relied on imaging studies, since the appearance is pathognomonic, and surgical risk was felt to be high and unlikely to affect therapy. The DIPG Biology and Treatment Study (DIPG-BATS) reported here incorporated a surgical biopsy at presentation and stratified subjects to receive FDA-approved agents chosen on the basis of specific biologic targets.
   Methods. Subjects were eligible for the trial if the clinical features and imaging appearance of a newly diagnosed tumor were consistent with a DIPG. Surgical biopsies were performed after enrollment and prior to definitive treatment. All subjects were treated with conventional external beam radiotherapy with bevacizumab, and then stratified to receive bevacizumab with erlotinib or temozolomide, both agents, or neither agent, based on O6-methylguanine-DNA methyltransferase status and epidermal growth factor receptor expression. Whole-genome sequencing and RNA sequencing were performed but not used for treatment assignment.
   Results. Fifty-three patients were enrolled at 23 institutions, and 50 underwent biopsy. The median age was 6.4 years, with 24 male and 29 female subjects. Surgical biopsies were performed with a specified technique and no deaths were attributed to the procedure. Two subjects experienced grade 3 toxicities during the procedure (apnea, n = 1; hypertension, n = 1). One subject experienced a neurologic deficit (left hemiparesis) that did not fully recover. Of the 50 tumors biopsied, 46 provided sufficient tissue to perform the study assays (92%, two-stage exact binomial 90% CI: 83%-97%).
   Conclusions. Surgical biopsy of DIPGs is technically feasible, associated with acceptable risks, and can provide biologic data that can inform treatment decisions.
C1 [Gupta, Nalin; Mueller, Sabine] Univ Calif San Francisco, Benioff Childrens Hosp, San Francisco, CA 94143 USA.
   [Gupta, Nalin; Mueller, Sabine] Univ Calif San Francisco, San Francisco, CA 94143 USA.
   [Goumnerova, Liliana C.; Manley, Peter; Chi, Susan N.; Neuberg, Donna; Malkin, Hayley; Sinai, Claire; Greenspan, Lianne; Wright, Karen D.; Bandopadhayay, Pratiti; Ligon, Keith L.; Kieran, Mark W.] Dana Farber Canc Inst, Boston, MA 02115 USA.
   [Goumnerova, Liliana C.; Manley, Peter; Chi, Susan N.; Wright, Karen D.; Bandopadhayay, Pratiti; Kieran, Mark W.] Boston Childrens Hosp, Boston, MA USA.
   [Fangusaro, Jason; Tomita, Tadanori; Alden, Tord; DiPatri, Arthur] Ann & Robert H Lurie Childrens Hosp Chicago, Chicago, IL 60611 USA.
   [Fangusaro, Jason; Tomita, Tadanori; Alden, Tord; DiPatri, Arthur] Northwestern Univ, Chicago, IL 60611 USA.
   [Rubin, Joshua B.; Gauvain, Karen; Limbrick, David; Leonard, Jeffrey] Washington Univ, Med Ctr, St Louis, MO USA.
   [Rubin, Joshua B.; Gauvain, Karen; Limbrick, David; Leonard, Jeffrey] St Louis Childrens Hosp, St Louis, MO 63178 USA.
   [Geyer, J. Russel; Leary, Sarah] Seattle Childrens Hosp, Seattle, WA USA.
   [Geyer, J. Russel; Leary, Sarah] Univ Washington, Seattle, WA 98195 USA.
   [Wang, Zhihong; Sood, Sandeep] Childrens Hosp Michigan, Detroit, MI 48201 USA.
   [Wang, Zhihong; Sood, Sandeep] Wayne State Univ, Detroit, MI USA.
   [Nagib, Mahmoud] Childrens Hosp & Clin Minnesota, Minneapolis, MN USA.
   [Karajannis, Matthias A.; Harter, David] NYU, New York, NY USA.
   [Ayyanar, Kanyalakshmi; Gump, William] Univ Louisville, Louisville, KY 40292 USA.
   [Ayyanar, Kanyalakshmi; Gump, William] Nortons Childrens Hosp, Louisville, KY USA.
   [Bowers, Daniel C.; Weprin, Bradley] Univ Texas Southwestern Med Ctr Dallas, Dallas, TX 75390 USA.
   [MacDonald, Tobey J.; Aguilera, Dolly; Brahma, Barunashish] Childrens Healthcare Atlanta, Atlanta, GA USA.
   [MacDonald, Tobey J.; Aguilera, Dolly; Brahma, Barunashish] Emory Univ, Atlanta, GA 30322 USA.
   [Robison, Nathan J.; Kiehna, Erin; Krieger, Mark] Childrens Hosp Los Angeles, Los Angeles, CA 90027 USA.
   [Sandler, Eric; Aldana, Philipp] Wolfsons Childrens Hosp, Nemours Childrens Clin, Jacksonville, FL USA.
   [Sandler, Eric; Aldana, Philipp] Univ Florida, Jacksonville, FL USA.
   [Khatib, Ziad; Ragheb, John] Nicklaus Childrens Hosp, Miami, FL USA.
   [Bredlau, Amy-Lee] Med Univ South Carolina, Charleston, SC 29425 USA.
   [Fuchs, Herbert] Preston Robert Tisch Brain Tumor Ctr, Durham, NC USA.
   [Fuchs, Herbert] Duke Univ, Med Ctr, Durham, NC USA.
   [Cohen, Kenneth J.; Jallo, George] Johns Hopkins Univ, Baltimore, MD USA.
   [Dorris, Kathleen; Handler, Michael] Childrens Hosp Colorado, Denver, CO USA.
   [Dorris, Kathleen; Handler, Michael] Univ Colorado, Sch Med, Denver, CO USA.
   [Comito, Melanie; Dias, Mark] Penn State Hlth Childrens Hosp, Hershey, PA USA.
   [Nazemi, Kellie; Baird, Lissa] Oregon Hlth & Sci Univ, Portland, OR 97201 USA.
   [Nazemi, Kellie; Baird, Lissa] Doernbecher Childrens Hosp, Portland, OR USA.
   [Murray, Jeff] Cook Childrens Med Ctr, Ft Worth, TX USA.
   [Bandopadhayay, Pratiti] Broad Inst, Cambridge, MA USA.
   [Lindeman, Neal; Hornick, Jason L.; Ligon, Keith L.] Brigham & Womens Hosp, 75 Francis St, Boston, MA 02115 USA.
C3 University of California System; University of California San Francisco;
   University of California System; University of California San Francisco;
   Harvard University; Dana-Farber Cancer Institute; Harvard University;
   Boston Children's Hospital; Ann & Robert H. Lurie Children's Hospital of
   Chicago; Northwestern University; Washington University (WUSTL); St.
   Louis Children's Hospital; Washington University (WUSTL); Seattle
   Children's Hospital; University of Washington; University of Washington
   Seattle; Children's Hospital of Michigan; Wayne State University;
   Children's Hospitals & Clinics of Minnesota; New York University;
   University of Louisville; University of Texas System; University of
   Texas Southwestern Medical Center Dallas; Children's Healthcare of
   Atlanta (CHOA); Emory University; Children's Hospital Los Angeles; State
   University System of Florida; University of Florida; Medical University
   of South Carolina; Duke University; Johns Hopkins University; Children's
   Hospital Colorado; University of Colorado System; University of Colorado
   Anschutz Medical Campus; University of Colorado Denver; Pennsylvania
   Commonwealth System of Higher Education (PCSHE); Pennsylvania State
   University; Penn State Health; Oregon Health & Science University; Cook
   Children's Medical Center; Harvard University; Massachusetts Institute
   of Technology (MIT); Broad Institute; Harvard University; Brigham &
   Women's Hospital
RP Gupta, N (通讯作者)，Univ Calif San Francisco, Benioff Childrens Hosp, Div Pediat Neurosurg, 550 16th St,4th Floor,Box 0137, San Francisco, CA 94143 USA.
EM
RI Karajannis, Matthias A/H-8845-2019; Sandler, Eric/AAE-8656-2019
OI Karajannis, Matthias A/0000-0002-7151-6528; gardner,
   sharon/0000-0002-8857-5487; Bowers, Daniel/0000-0002-3947-2481; Sood,
   Sandeep/0000-0001-9305-446X; Robison, Nathan/0000-0001-8613-9772
FU Zach Carson Fund; Ellie Kavalieros DIPG Fund; Ryan Harvey Fund; Mikey
   Czech DIPG Foundation; Caroline Cronk Fund; Prayers from Maria;
   Guglietti DIPG Fund; Markoff Art in Giving Foundation; Brock Fleming
   Fund; Stop and Shop Pediatric Brain Tumor Program; DIPG Collaborative;
   Hope for Caroline Foundation; Reflections of Grace Foundation; Cure
   Starts Now Foundation; Soar with Grace Foundation; Abbie's Army Charity
   Trust; Julian Boivin Courage for Cures Foundation; Smiles for Sophie
   Forever; Caroline's Miracle Foundation; Love; Chloe Foundation; Benny's
   World Foundation; Pray Hope Believe Foundation; Jeffrey Thomas Hayden
   Foundation;  [5P30 CA006516]
FX This work was supported by the Zach Carson Fund, Ellie Kavalieros DIPG
   Fund, Ryan Harvey Fund, Mikey Czech DIPG Foundation, Caroline Cronk
   Fund, Prayers from Maria, Guglietti DIPG Fund, Markoff Art in Giving
   Foundation, Brock Fleming Fund, Stop and Shop Pediatric Brain Tumor
   Program, the DIPG Collaborative (which includes the Hope for Caroline
   Foundation, Reflections of Grace Foundation, Cure Starts Now Foundation,
   Soar with Grace Foundation, Abbie's Army Charity Trust, Julian Boivin
   Courage for Cures Foundation, Smiles for Sophie Forever, Caroline's
   Miracle Foundation, Love, Chloe Foundation, Benny's World Foundation,
   Pray Hope Believe Foundation, Jeffrey Thomas Hayden Foundation), and
   5P30 CA006516.
CR Buczkowicz P, 2014, NAT GENET, V46, P451, DOI 10.1038/ng.2936
   Cage TA, 2013, CHILD NERV SYST, V29, P1313, DOI 10.1007/s00381-013-2101-0
   Chan KM, 2013, GENE DEV, V27, P985, DOI 10.1101/gad.217778.113
   Fontebasso AM, 2014, NAT GENET, V46, P462, DOI 10.1038/ng.2950
   Gilbertson RJ, 2003, CLIN CANCER RES, V9, P3620
   Grasso CS, 2015, NAT MED, V21, P555, DOI 10.1038/nm.3855
   Hashizume R, 2014, NAT MED, V20, P1394, DOI 10.1038/nm.3716
   Jansen MHA, 2012, CANCER TREAT REV, V38, P27, DOI 10.1016/j.ctrv.2011.06.007
   Kebudi R, 2013, PEDIATR DRUGS, V15, P351, DOI 10.1007/s40272-013-0033-5
   Paugh BS, 2011, J CLIN ONCOL, V29, P3999, DOI 10.1200/JCO.2011.35.5677
   Paugh BS, 2010, J CLIN ONCOL, V28, P3061, DOI 10.1200/JCO.2009.26.7252
   Puget S, 2015, CHILD NERV SYST, V31, P1773, DOI 10.1007/s00381-015-2832-1
   Robison NJ, 2014, J NEURO-ONCOL, V119, P7, DOI 10.1007/s11060-014-1448-8
   Roujeau T, 2007, J NEUROSURG, V107, P1, DOI 10.3171/PED-07/07/001
   Taylor KR, 2014, NAT GENET, V46, P457, DOI 10.1038/ng.2925
   Wu G, 2014, NAT GENET, V46, P444, DOI 10.1038/ng.2938
   Wu G, 2012, NAT GENET, V44, P251, DOI 10.1038/ng.1102
NR 17
TC 60
Z9 62
U1 1
U2 5
PU OXFORD UNIV PRESS INC
PI CARY
PA JOURNALS DEPT, 2001 EVANS RD, CARY, NC 27513 USA
SN 1522-8517
EI 1523-5866
J9 NEURO-ONCOLOGY
JI Neuro-Oncology
PD NOV
PY 2018
VL 20
IS 11
BP 1547
EP 1555
DI 10.1093/neuonc/noy070
PG 9
WC Oncology; Clinical Neurology
WE Science Citation Index Expanded (SCI-EXPANDED)
SC Oncology; Neurosciences & Neurology
GA GY6AE
UT WOS:000448665500014
PM 29741745
OA Green Published, Bronze
DA 2023-08-27
ER

PT J
AU Kastriti, ME
   Kameneva, P
   Adameyko, I
AF Kastriti, Maria Eleni
   Kameneva, Polina
   Adameyko, Igor
TI Stem cells, evolutionary aspects and pathology of the adrenal medulla: A
   new developmental paradigm
SO MOLECULAR AND CELLULAR ENDOCRINOLOGY
LA English
DT Review
DE Schwann cell precursors; Chromaffin cells; Oxygen-sensing;
   Neuroblastoma; Paraganglioma; Pheochromocytoma
ID VASOACTIVE INTESTINAL POLYPEPTIDE; CHROMAFFIN CELL; NEURAL-CREST;
   CAROTID-BODY; COMPOSITE PHEOCHROMOCYTOMA; ADRENOMEDULLARY FUNCTION;
   POSTNATAL-DEVELOPMENT; ADRENERGIC PHENOTYPE; NEUROPEPTIDE-Y;
   GROWTH-FACTORS
AB The mammalian adrenal gland is composed of two main components; the catecholaminergic neural crest-derived medulla, found in the center of the gland, and the mesoderm-derived cortex producing steroidogenic hormones. The medulla is composed of neuroendocrine chromaffin cells with oxygen-sensing properties and is dependent on tissue interactions with the overlying cortex, both during development and in adulthood. Other relevant organs include the Zuckerkandl organ containing extra-adrenal chromaffin cells, and carotid oxygen-sensing bodies containing glomus cells. Chromaffin and glomus cells reveal a number of important similarities and are derived from the multipotent nerve-associated descendants of the neural crest, or Schwann cell precursors. Abnormalities in complex developmental processes during differentiation of nerve-associated and other progenitors into chromaffin and oxygen-sensing populations may result in different subtypes of paraganglioma, neuroblastoma and pheochromocytoma. Here, we summarize recent findings explaining the development of chromaffin and oxygen-sensing cells, as well as the potential mechanisms driving neuroendocrine tumor initiation.
C1 [Kastriti, Maria Eleni; Kameneva, Polina; Adameyko, Igor] Karolinska Inst, Dept Physiol & Pharmacol, Solna, Sweden.
   [Kastriti, Maria Eleni; Adameyko, Igor] Med Univ Vienna, Ctr Brain Res, Dept Mol Neurosci, Vienna, Austria.
   [Kameneva, Polina] Russian Acad Sci, Natl Sci Ctr Marine Biol, Far Eastern Branch, Vladivostok, Russia.
   [Adameyko, Igor] Med Univ Vienna, Ctr Brain Res, Dept Neuroimmunol, Vienna, Austria.
C3 Karolinska Institutet; Medical University of Vienna; Russian Academy of
   Sciences; National Scientific Center of Marine Biology, Far East Branch
   of the Russian Academy of Sciences; Medical University of Vienna
RP Adameyko, I (通讯作者)，Karolinska Inst, Dept Physiol & Pharmacol, Solna, Sweden.
EM
RI Kameneva, Polina/N-4474-2016
OI Kameneva, Polina/0000-0002-5513-8221; Adameyko, Igor/0000-0001-5471-0356
FU Swedish Research Council grant (Vetenskapsradet, VR); ERC Consolidator
   grant "STEMMING-FROM-NERVE" [N647844]; Paradifference Foundation; Bertil
   Hallsten Research Foundation; Novo Nordisk Foundation (Postdoc
   fellowship in Endocrinology and Metabolism at International Elite
   Environments) [NNF17OC0026874]; Stiftelsen Riksbankens Jubileumsfond
   (Erik Ronnbergs fond stipend)
FX The authors would like to thank Ingrid Lekk and Thibault G. Bouderlique
   for valuable discussions and feedback. I.A. acknowledges funding from a
   Swedish Research Council grant (Vetenskapsradet, VR), ERC Consolidator
   grant "STEMMING-FROM-NERVE" N647844, the Paradifference Foundation and
   the Bertil Hallsten Research Foundation. M.E.K. was supported by the
   Novo Nordisk Foundation (Postdoc fellowship in Endocrinology and
   Metabolism at International Elite Environments, NNF17OC0026874) and
   Stiftelsen Riksbankens Jubileumsfond (Erik Ronnbergs fond stipend).
CR AFEWORK M, 1995, CELL TISSUE RES, V280, P291, DOI 10.1007/s004410050356
   Ambros PF, 2009, BRIT J CANCER, V100, P1471, DOI 10.1038/sj.bjc.6605014
   Rabadan MA, 2013, DEV NEUROBIOL, V73, P815, DOI 10.1002/dneu.22099
   Annese V, 2017, CELL REP, V19, P471, DOI 10.1016/j.celrep.2017.03.065
   ARIASSTELLA J, 1976, HUM PATHOL, V7, P361, DOI 10.1016/S0046-8177(76)80052-4
   AXELROD J, 1974, SCI AM, V230, P59
   Bandiera R, 2013, DEV CELL, V27, P5, DOI 10.1016/j.devcel.2013.09.003
   Basham KJ, 2019, GENE DEV, V33, P209, DOI 10.1101/gad.317412.118
   Boeva V, 2017, NAT GENET, V49, P1408, DOI 10.1038/ng.3921
   BOHN MC, 1981, DEV BIOL, V82, P1, DOI 10.1016/0012-1606(81)90423-1
   Bornstein S.R., 2019, CURR OPIN ENDOCR MET, V8, P9, DOI [10.1016/j.coemr.2019.06.005, DOI 10.1016/J.COEMR.2019.06.005]
   Bornstein SR, 1999, FASEB J, V13, P1185, DOI 10.1096/fasebj.13.10.1185
   Bornstein SR, 2000, P NATL ACAD SCI USA, V97, P14742, DOI 10.1073/pnas.97.26.14742
   Brindley RL, 2017, ACS CHEM NEUROSCI, V8, P943, DOI 10.1021/acschemneuro.7b00026
   Britsch S, 1998, GENE DEV, V12, P1825, DOI 10.1101/gad.12.12.1825
   Brodeur GM, 2018, CELL TISSUE RES, V372, P277, DOI 10.1007/s00441-017-2761-2
   Brodeur GM, 2014, NAT REV CLIN ONCOL, V11, P704, DOI 10.1038/nrclinonc.2014.168
   BROGI MP, 1959, J PHYSIOL-LONDON, V146, P165, DOI 10.1113/jphysiol.1959.sp006186
   Burnichon N, 2011, HUM MOL GENET, V20, P3974, DOI 10.1093/hmg/ddr324
   Chai C, 2003, DEV BIOL, V260, P226, DOI 10.1016/S0012-1606(03)00219-7
   Chan WH, 2016, DEV NEUROBIOL, V76, P137, DOI 10.1002/dneu.22304
   CHIOCCHI.SR, 1966, NATURE, V212, P834, DOI 10.1038/212834a0
   Chung KF, 2011, MOL CELL ENDOCRINOL, V336, P141, DOI 10.1016/j.mce.2010.11.028
   CLAUDE P, 1988, NEURON, V1, P783, DOI 10.1016/0896-6273(88)90126-2
   COLE TJ, 1995, GENE DEV, V9, P1608, DOI 10.1101/gad.9.13.1608
   COUPLAND RE, 1954, J ANAT, V88, P455
   Dagerlind A, 1990, Neuroreport, V1, P137, DOI 10.1097/00001756-199010000-00014
   Dahia PLM, 2005, PLOS GENET, V1, P72, DOI 10.1371/journal.pgen.0010008
   Das B, 2008, STEM CELLS, V26, P1818, DOI 10.1634/stemcells.2007-0724
   de Celis MFR, 2015, STEM CELLS, V33, P2037, DOI 10.1002/stem.2002
   Dyachuk V, 2014, SCIENCE, V345, P82, DOI 10.1126/science.1253281
   Eisenhofer G, 2011, ENDOCR-RELAT CANCER, V18, P97, DOI 10.1677/ERC-10-0211
   Eisenhofer G, 2011, J CLIN ENDOCR METAB, V96, P375, DOI 10.1210/jc.2010-1588
   Engeland WC, 1996, ENDOCR RES, V22, P395, DOI 10.1080/07435809609043723
   EPPLE A, 1985, J MORPHOL, V183, P225, DOI 10.1002/jmor.1051830210
   Ernsberger U, 2005, CELL TISSUE RES, V319, P1, DOI 10.1007/s00441-004-0996-1
   Espinosa-Medina I, 2014, SCIENCE, V345, P87, DOI 10.1126/science.1253286
   Finco I, 2018, ENDOCRINOLOGY, V159, P579, DOI 10.1210/en.2017-03061
   Finotto S, 1999, DEVELOPMENT, V126, P2935
   Fishbein L, 2018, CELL TISSUE RES, V372, P339, DOI 10.1007/s00441-018-2795-0
   Fishbein L, 2017, CANCER CELL, V31, P181, DOI 10.1016/j.ccell.2017.01.001
   Furlan A, 2018, DEV BIOL, V444, pS25, DOI 10.1016/j.ydbio.2018.02.008
   Furlan A, 2017, SCIENCE, V357, P46, DOI 10.1126/science.aal3753
   Garcia-Fernandez M, 2007, PFLUG ARCH EUR J PHY, V454, P93, DOI 10.1007/s00424-006-0186-y
   Gaskell JF, 1912, J PHYSIOL-LONDON, V44, P59
   GONZALEZ C, 1994, PHYSIOL REV, V74, P829, DOI 10.1152/physrev.1994.74.4.829
   Gotlieb N, 2018, ENDOCRINOLOGY, V159, P248, DOI 10.1210/en.2017-00505
   Graham GS, 1916, J MED RES, V34, P241
   GUILLEMOT F, 1993, CELL, V75, P463, DOI 10.1016/0092-8674(93)90381-Y
   Gut P, 2005, DEVELOPMENT, V132, P4611, DOI 10.1242/dev.02052
   HAMPEL R, 1994, EXP CLIN ENDOCRINOL, V102, P90, DOI 10.1055/s-0029-1211268
   Hershkovitz L, 2007, ENDOCRINOLOGY, V148, P976, DOI 10.1210/en.2006-1100
   HIROKAWA N, 1974, Z ANAT ENTWICKLUNGS, V144, P85, DOI 10.1007/BF00518635
   Hockman D, 2017, ELIFE, V6, DOI 10.7554/eLife.21231
   Hockman D, 2018, DEV BIOL, V444, pS308, DOI 10.1016/j.ydbio.2018.05.016
   Holgert H, 1996, J COMP NEUROL, V371, P603
   HOLZWARTH MA, 1980, NEUROENDOCRINOLOGY, V31, P168, DOI 10.1159/000123069
   Howard MJ, 2000, DEVELOPMENT, V127, P4073
   Hsu HJ, 2003, DEVELOPMENT, V130, P2107, DOI 10.1242/dev.00427
   Huber K, 2006, DEV BIOL, V298, P335, DOI 10.1016/j.ydbio.2006.07.010
   Huber K, 2018, CELL TISSUE RES, V372, P163, DOI 10.1007/s00441-018-2831-0
   Huber K, 2009, AUTON NEUROSCI-BASIC, V151, P10, DOI 10.1016/j.autneu.2009.07.020
   Jessen KR, 2005, NAT REV NEUROSCI, V6, P671, DOI 10.1038/nrn1746
   Jessen KR, 1997, J ANAT, V191, P501, DOI 10.1046/j.1469-7580.1997.19140501.x
   Jessen KR, 2019, FRONT MOL NEUROSCI, V12, DOI 10.3389/fnmol.2019.00069
   Jessen KR, 2015, CSH PERSPECT BIOL, V7, DOI 10.1101/cshperspect.a020487
   Kameda Y, 2002, MICROSC RES TECHNIQ, V59, P196, DOI 10.1002/jemt.10194
   Kasemeier-Kulesa JC, 2010, J NEUROSCI, V30, P13078, DOI 10.1523/JNEUROSCI.0892-10.2010
   Kastriti ME, 2019, FRONT MOL NEUROSCI, V12, DOI 10.3389/fnmol.2019.00006
   Kerosuo L, 2018, P NATL ACAD SCI USA, V115, pE7351, DOI 10.1073/pnas.1800039115
   KNUDSON AG, 1972, AM J HUM GENET, V24, P514
   Knudson AG, 1996, J CANCER RES CLIN, V122, P135, DOI 10.1007/BF01366952
   Kohn A., 1902, ERGEBNISSE ANATOMIE, V12, P253
   LAWSON W, 1980, LARYNGOSCOPE, V90, P120, DOI 10.1288/00005537-198001000-00014
   LEDOUARI.N, 1972, CR ACAD SCI D NAT, V275, P583
   LEDOUARI.NM, 1974, DEV BIOL, V41, P162, DOI 10.1016/0012-1606(74)90291-7
   Lee MJ, 1997, MOL CELL NEUROSCI, V8, P336, DOI 10.1006/mcne.1996.0589
   Lee MJ, 2001, DEV DYNAM, V222, P40, DOI 10.1002/dvdy.1165
   Lim KC, 2000, NAT GENET, V25, P209, DOI 10.1038/76080
   Lopez-Barneo J, 2008, EUR RESPIR J, V32, P1386, DOI 10.1183/09031936.00056408
   LORING JF, 1987, DEV BIOL, V121, P220, DOI 10.1016/0012-1606(87)90154-0
   Lumb R, 2018, DEVELOPMENT, V145, DOI 10.1242/dev.162552
   Lumb R, 2014, NEURAL DEV, V9, DOI 10.1186/1749-8104-9-24
   Matthay KK, 2016, NAT REV DIS PRIMERS, V2, DOI 10.1038/nrdp.2016.78
   MAUBERT E, 1990, NEUROSCI LETT, V113, P121, DOI 10.1016/0304-3940(90)90290-P
   MILANO EG, 1986, J MORPHOL, V189, P249, DOI 10.1002/jmor.1051890304
   Milano EG, 1997, GEN COMP ENDOCR, V108, P483, DOI 10.1006/gcen.1997.7005
   MochizukiOda N, 1997, J NEUROCHEM, V69, P377
   Morohashi K, 2011, MOL CELL ENDOCRINOL, V336, P193, DOI 10.1016/j.mce.2010.11.026
   Nagatsu T, 1998, Adv Pharmacol, V42, P1
   OOMORI Y, 1994, CELL TISSUE RES, V275, P201, DOI 10.1007/BF00319418
   OOMORI Y, 1994, CELL TISSUE RES, V278, P249, DOI 10.1007/BF00414167
   PAIEMENT JM, 1975, GEN COMP ENDOCR, V27, P495, DOI 10.1016/0016-6480(75)90070-2
   Pardal R, 2007, CELL, V131, P364, DOI 10.1016/j.cell.2007.07.043
   Pattyn A, 1999, NATURE, V399, P366, DOI 10.1038/20700
   PEARSE AGE, 1973, HISTOCHEMISTRY, V34, P191, DOI 10.1007/BF00303435
   Peifer M, 2015, NATURE, V526, P700, DOI 10.1038/nature14980
   Pihlajoki M, 2015, FRONT ENDOCRINOL, V6, DOI 10.3389/fendo.2015.00027
   Pugh TJ, 2013, NAT GENET, V45, P279, DOI 10.1038/ng.2529
   Rai R, 2012, SOUTH ASIAN J CANCER, V1, P98, DOI 10.4103/2278-330X.103727
   Ramirez M, 2012, ADV EXP MED BIOL, V758, P255, DOI 10.1007/978-94-007-4584-1_35
   Rao RN, 2013, UROL ANNALS, V5, P115, DOI 10.4103/0974-7796.110011
   Reiprich S, 2008, MOL BIOL CELL, V19, P1575, DOI 10.1091/mbc.E07-08-0782
   Reissmann E, 1996, DEVELOPMENT, V122, P2079
   Rychkov GY, 1998, J PHYSIOL-LONDON, V509, P887, DOI 10.1111/j.1469-7793.1998.887bm.x
   Saito D, 2012, SCIENCE, V336, P1578, DOI 10.1126/science.1222369
   Sano H, 2007, PEDIATR DEVEL PATHOL, V10, P229, DOI 10.2350/06-06-0117.1
   Schneider C, 1999, NEURON, V24, P861, DOI 10.1016/S0896-6273(00)81033-8
   Schober A, 2013, J NEUROENDOCRINOL, V25, P34, DOI 10.1111/j.1365-2826.2012.02367.x
   Schwarz Q, 2009, P NATL ACAD SCI USA, V106, P6164, DOI 10.1073/pnas.0811521106
   Scriba LD, 2020, FRONT ENDOCRINOL, V11, DOI 10.3389/fendo.2020.00079
   SEIDLER FJ, 1985, J PHYSIOL-LONDON, V358, P1
   Shah NM, 1996, CELL, V85, P331, DOI 10.1016/S0092-8674(00)81112-5
   Sharma A, 2012, FRONT CELL NEUROSCI, V6, DOI 10.3389/fncel.2012.00028
   Shida Yohei, 2015, BMC Res Notes, V8, P257, DOI 10.1186/s13104-015-1233-6
   Shtukmaster S, 2013, NEURAL DEV, V8, DOI 10.1186/1749-8104-8-12
   Sicard F, 2006, ANN NY ACAD SCI, V1073, P312, DOI 10.1196/annals.1353.036
   Sicard F, 2007, P NATL ACAD SCI USA, V104, P2007, DOI 10.1073/pnas.0610898104
   Smith Sean M, 2006, Dialogues Clin Neurosci, V8, P383
   Steenblock C, 2018, P NATL ACAD SCI USA, V115, P12997, DOI 10.1073/pnas.1814072115
   Stoyek MR, 2017, AUTON NEUROSCI-BASIC, V206, P43, DOI 10.1016/j.autneu.2017.07.004
   Tajima T, 1999, GENE THER, V6, P1898, DOI 10.1038/sj.gt.3301018
   Takahashi T, 1999, CELL, V99, P59, DOI 10.1016/S0092-8674(00)80062-8
   Takahashi Y, 2013, SCIENCE, V341, P860, DOI 10.1126/science.1230717
   Thompson RJ, 1997, J PHYSIOL-LONDON, V498, P503, DOI 10.1113/jphysiol.1997.sp021876
   Tischler AS, 1995, TOXICOL APPL PHARM, V135, P254, DOI 10.1006/taap.1995.1231
   TISCHLER AS, 1989, INT J DEV NEUROSCI, V7, P439, DOI 10.1016/0736-5748(89)90004-X
   TISCHLER AS, 1988, LAB INVEST, V58, P733
   To TT, 2007, MOL ENDOCRINOL, V21, P472, DOI 10.1210/me.2006-0216
   Tsarovina K, 2004, DEVELOPMENT, V131, P4775, DOI 10.1242/dev.01370
   Tsubota Shoma, 2017, Oncoscience, V4, P70, DOI 10.18632/oncoscience.360
   UNSICKER K, 1993, J ANAT, V183, P207
   Valentijn LJ, 2015, NAT GENET, V47, P1411, DOI 10.1038/ng.3438
   van Groningen T, 2017, NAT GENET, V49, P1261, DOI 10.1038/ng.3899
   VERHOFSTAD AAJ, 1993, J ANAT, V183, P315
   Veschi V, 2019, FRONT ENDOCRINOL, V10, DOI 10.3389/fendo.2019.00782
   Vidal V, 2016, GENE DEV, V30, P1389, DOI 10.1101/gad.277756.116
   Vinson GP, 2016, FRONT NEUROSCI-SWITZ, V10, DOI 10.3389/fnins.2016.00238
   VONEULER US, 1949, NATURE, V163, P642, DOI 10.1038/163642a0
   Wang ZY, 2002, MICROSC RES TECHNIQ, V59, P168, DOI 10.1002/jemt.10191
   Wildner H, 2008, DEVELOPMENT, V135, P473, DOI 10.1242/dev.011783
   Wood MA, 2013, DEVELOPMENT, V140, P4522, DOI 10.1242/dev.092775
   WURTMAN RJ, 1968, ENDOCRINOLOGY, V82, P584, DOI 10.1210/endo-82-3-584
   Yates R, 2013, CURR TOP DEV BIOL, V106, P239, DOI 10.1016/B978-0-12-416021-7.00007-9
   Yuan JY, 2000, NATURE, V407, P802, DOI 10.1038/35037739
   Zubair M, 2009, MOL ENDOCRINOL, V23, P1657, DOI 10.1210/me.2009-0055
   Zuckerkandl E., 1901, VERHANDLUNGEN ANAT G, V15, P95
NR 147
TC 14
Z9 14
U1 1
U2 6
PU ELSEVIER IRELAND LTD
PI CLARE
PA ELSEVIER HOUSE, BROOKVALE PLAZA, EAST PARK SHANNON, CO, CLARE, 00000,
   IRELAND
SN 0303-7207
EI 1872-8057
J9 MOL CELL ENDOCRINOL
JI Mol. Cell. Endocrinol.
PD DEC 1
PY 2020
VL 518
AR 110998
DI 10.1016/j.mce.2020.110998
PG 12
WC Cell Biology; Endocrinology & Metabolism
WE Science Citation Index Expanded (SCI-EXPANDED)
SC Cell Biology; Endocrinology & Metabolism
GA OK0ZN
UT WOS:000584381600047
PM 32818585
OA hybrid
DA 2023-08-27
ER

PT J
AU Surowiec, RK
   Ferris, SF
   Apfelbaum, A
   Espinoza, C
   Mehta, RK
   Monchamp, K
   Sirihorachai, VR
   Bedi, K
   Ljungman, M
   Galban, S
AF Surowiec, Rachel K.
   Ferris, Sarah F.
   Apfelbaum, April
   Espinoza, Carlos
   Mehta, Ranjit K.
   Monchamp, Karamoja
   Sirihorachai, Veerin R.
   Bedi, Karan
   Ljungman, Mats
   Galban, Stefanie
TI Transcriptomic Analysis of Diffuse Intrinsic Pontine Glioma (DIPG)
   Identifies a Targetable ALDH-Positive Subset of Highly Tumorigenic
   Cancer Stem-like Cells
SO MOLECULAR CANCER RESEARCH
LA English
DT Article
ID ALDEHYDE DEHYDROGENASE 1A1; 1ST-IN-HUMAN PHASE-I; HIGH-GRADE; PI3K;
   INHIBITOR; TUMOR; RESISTANCE; GROWTH; PATHWAYS; GDC-0084
AB Understanding the cancer stem cell (CSC) landscape in diffuse intrinsic pontine glioma (DIPG) is desperately needed to address treatment resistance and identify novel therapeutic approaches. Patient-derived DIPG cells demonstrated heterogeneous expression of aldehyde dehydrogenase (ALDH) and CD133 by flow cytometry. Transcriptome-level characterization identified elevated mRNA levels of MYC, E2F, DNA damage repair (DDR) genes, glycolytic metabolism, and mTOR signaling in ALDH(+) compared with ALDH(-), supporting a stem-like phenotype and indicating a druggable target. ALDH(+) cells demonstrated increased proliferation, neurosphere formation, and initiated tumors that resulted in decreased survival when orthotopically implanted. Pharmacologic MAPKJPI3K/mTOR targeting downregulated MYC, E2F, and DDR mRNAs and reduced glycolytic metabolism. In vivo PI3K/mTOR targeting inhibited tumor growth in both flank and an ALDH(+) orthotopic tumor model likely by reducing cancer sternness. In summary, we describe existence of ALDH(+) DIPGs with proliferative properties due to increased metabolism, which may be regulated by the microenvironment and likely contributing to drug resistance and tumor recurrence.
C1 [Surowiec, Rachel K.; Ferris, Sarah F.; Apfelbaum, April; Espinoza, Carlos; Mehta, Ranjit K.; Monchamp, Karamoja; Galban, Stefanie] Univ Michigan, Med Sch, Ctr Mol Imaging, Ann Arbor, MI 48109 USA.
   [Surowiec, Rachel K.; Ferris, Sarah F.; Apfelbaum, April; Espinoza, Carlos; Mehta, Ranjit K.; Monchamp, Karamoja; Galban, Stefanie] Univ Michigan, Med Sch, Dept Radiol, Ann Arbor, MI 48109 USA.
   [Apfelbaum, April; Sirihorachai, Veerin R.; Bedi, Karan] Univ Michigan, Med Sch, Canc Biol Grad Program, Ann Arbor, MI 48109 USA.
   [Sirihorachai, Veerin R.; Ljungman, Mats; Galban, Stefanie] Univ Michigan, Med Sch, Rogel Canc Ctr, Ann Arbor, MI 48109 USA.
   [Ljungman, Mats] Univ Michigan, Med Sch, Dept Radiat Oncol, Ann Arbor, MI 48109 USA.
   [Ljungman, Mats] Univ Michigan, Med Sch, Dept Environm Hlth Sci, Ann Arbor, MI 48109 USA.
   [Ljungman, Mats] Univ Michigan, Med Sch, Ctr RNA Biomed, Ann Arbor, MI 48109 USA.
C3 University of Michigan System; University of Michigan; University of
   Michigan System; University of Michigan; University of Michigan System;
   University of Michigan; University of Michigan System; University of
   Michigan; University of Michigan System; University of Michigan;
   University of Michigan System; University of Michigan; University of
   Michigan System; University of Michigan
RP Galban, S (通讯作者)，Univ Michigan, Med Sch, Dept Radiol, Ctr Mol Imaging, 109 Zina Pitcher Pl, Ann Arbor, MI 48109 USA.
EM
RI Mehta, Ranjit K/N-7433-2019; Bedi, Karan/AAA-3331-2022
OI Bedi, Karan/0000-0002-7843-738X
FU ChadTough/Michael Mosier Defeat DIPG foundation; MD Anderson Cancer
   Center support Grant [5 P30 CA016672-40]; Bru-Sequencing [1 R01
   CA213214, UM1 HG009382]; NCI of the NIH [P30CA046592, P30 CA04659229]
FX This work was supported by ChadTough/Michael Mosier Defeat DIPG
   foundation research grant funding (R. Surowiec, S. Ferris, S. Galban).
   The authors wish to thank the functional proteomics RPPA core facility
   supported by MD Anderson Cancer Center support Grant #5 P30 CA016672-40
   and Michelle Paulsen in assisting with Bru-Sequencing under grant
   support from 1 R01 CA213214 and UM1 HG009382 (K. Bedi, M. Ljungman).
   Flow cytometry work was performed at the University of Michigan Flow
   cytometry Core, which is supported by the NCI of the NIH under award
   numbers P30CA046592 and P30 CA04659229. The authors would also like to
   thank Dr. Mohammed Farhoud from Emit Imaging for performing the
   cryocomputed tomography. The authors wish to thank Drs. Uday B.
   Maachani, Mark M. Souweidane, and Sriram Venneti as well as Michelle
   Monje-Deisseroth for providing us with the DIPG cell lines SU-DIPG IV,
   SU-DIPG XIII, HSJD-DIPG 007, and SU-DIPG 29, respectively. We would also
   like to acknowledge Dr. Filbin's group who has made the Sc-RNA
   sequencing data publicly available
   (https://singlecell.broadinstitute.org/single_cell/study/SCP147/single-c
   ell-analysis-in-pediatric-mid line-gliomas-with-histone-h3k27m-mutation;
   ref. 48) and Cara Spencer from the Galban lab for aiding in analyzing
   the scRNA seq data.
CR Ajani JA, 2014, MOL ONCOL, V8, P142, DOI 10.1016/j.molonc.2013.10.007
   Blaj C, 2017, CANCER RES, V77, P1763, DOI 10.1158/0008-5472.CAN-16-2821
   Butler DE, 2017, ONCOTARGET, V8, P56698, DOI 10.18632/oncotarget.18082
   Capello M, 2016, ONCOTARGET, V7, P5598, DOI 10.18632/oncotarget.6798
   Castel D, 2015, ACTA NEUROPATHOL, V130, P815, DOI 10.1007/s00401-015-1478-0
   Chae YC, 2018, BMB REP, V51, P319, DOI 10.5483/BMBRep.2018.51.7.112
   Chappell WH, 2011, ONCOTARGET, V2, P135, DOI 10.18632/oncotarget.240
   Chiou SH, 2010, CANCER RES, V70, P10433, DOI 10.1158/0008-5472.CAN-10-2638
   Choi SA, 2014, EUR J CANCER, V50, P137, DOI 10.1016/j.ejca.2013.09.004
   Clark DW, 2016, ANN TRANSL MED, V4, DOI 10.21037/atm.2016.11.82
   Clement V, 2009, INT J CANCER, V125, P244, DOI 10.1002/ijc.24352
   Croker AK, 2012, BREAST CANCER RES TR, V133, P75, DOI 10.1007/s10549-011-1692-y
   Daina A, 2017, SCI REP-UK, V7, DOI 10.1038/srep42717
   Deng JL, 2019, BMC CANCER, V19, DOI 10.1186/s12885-019-5824-9
   Du FY, 2019, WORLD J STEM CELLS, V11, P398, DOI 10.4252/wjsc.v11.i7.398
   Duchatel R, 2019, NEURO-ONCOLOGY, V21, P68, DOI 10.1093/neuonc/noz036.024
   Emery B, 2015, CSH PERSPECT BIOL, V7, DOI 10.1101/cshperspect.a020461
   Engelman JA, 2008, NAT MED, V14, P1351, DOI 10.1038/nm.1890
   Feo S, 2000, FEBS LETT, V473, P47, DOI 10.1016/S0014-5793(00)01494-0
   Filbin MG, 2018, SCIENCE, V360, P331, DOI 10.1126/science.aao4750
   Galban S, 2017, MOL CANCER THER, V16, P2340, DOI 10.1158/1535-7163.MCT-17-0207
   Gao XF, 2019, NEOPLASMA, V66, P532, DOI 10.4149/neo_2018_180829N656
   Ginestier C, 2007, CELL STEM CELL, V1, P555, DOI 10.1016/j.stem.2007.08.014
   Glumac PM, 2018, CLIN TRANSL MED, V7, DOI 10.1186/s40169-018-0198-1
   Gopal YNV, 2010, CANCER RES, V70, P8736, DOI 10.1158/0008-5472.CAN-10-0902
   Grasso CS, 2015, NAT MED, V21, P555, DOI 10.1038/nm.3855
   Harutyunyan AS, 2019, NAT COMMUN, V10, DOI 10.1038/s41467-019-09140-x
   Heffron TP, 2016, ACS MED CHEM LETT, V7, P351, DOI 10.1021/acsmedchemlett.6b00005
   Kopetz S, 2015, J CLIN ONCOL, V33, P4032, DOI 10.1200/JCO.2015.63.2497
   Kusoglu A, 2019, GENE, V681, P80, DOI 10.1016/j.gene.2018.09.052
   Lapin DH, 2017, FRONT ONCOL, V7, DOI 10.3389/fonc.2017.00057
   Lowe BR, 2019, CANCERS, V11, DOI 10.3390/cancers11050660
   Ma I, 2011, STEM CELL REV REP, V7, P292, DOI 10.1007/s12015-010-9208-4
   Mackay A, 2017, CANCER CELL, V32, P520, DOI 10.1016/j.ccell.2017.08.017
   Mandal PK, 2011, NAT REV MOL CELL BIO, V12, P198, DOI 10.1038/nrm3060
   Mao Z, 2018, INT J MOL SCI, V19, DOI 10.3390/ijms19072043
   Matulonis U, 2015, GYNECOL ONCOL, V136, P246, DOI 10.1016/j.ygyno.2014.12.019
   Meng S, 2019, FOOD CHEM X, V1, DOI 10.1016/j.fochx.2019.100004
   Misuraca KL, 2015, FRONT ONCOL, V5, DOI 10.3389/fonc.2015.00172
   Modarai Shirin R, 2018, Oncotarget, V9, P34658, DOI 10.18632/oncotarget.26157
   Monje M, 2011, P NATL ACAD SCI USA, V108, P4453, DOI 10.1073/pnas.1101657108
   Munster P, 2016, CLIN CANCER RES, V22, P1932, DOI 10.1158/1078-0432.CCR-15-1665
   Murat A, 2008, J CLIN ONCOL, V26, P3015, DOI 10.1200/JCO.2007.15.7164
   Nagaraja S, 2019, MOL CELL, V76, P965, DOI 10.1016/j.molcel.2019.08.030
   Najafi M, 2019, LIFE SCI, V234, DOI 10.1016/j.lfs.2019.116781
   Paulsen MT, 2014, METHODS, V67, P45, DOI 10.1016/j.ymeth.2013.08.015
   Paulsen MT, 2013, P NATL ACAD SCI USA, V110, P2240, DOI 10.1073/pnas.1219192110
   Sanchez-Diaz PC, 2017, PLOS ONE, V12, DOI 10.1371/journal.pone.0176879
   Singh SK, 2003, CANCER RES, V63, P5821
   Singh SK, 2004, NATURE, V432, P396, DOI 10.1038/nature03128
   Smith MCP, 2004, CANCER RES, V64, P8604, DOI 10.1158/0008-5472.CAN-04-1844
   Stacer AC, 2015, MOL IMAGING, V14, P414, DOI 10.2310/7290.2015.00016
   Tomita H, 2016, ONCOTARGET, V7, P11018, DOI 10.18632/oncotarget.6920
   Vansteenkiste JF, 2015, J THORAC ONCOL, V10, P1319, DOI 10.1097/JTO.0000000000000607
   Vasilogiannakopoulou T, 2018, TRENDS PHARMACOL SCI, V39, P605, DOI 10.1016/j.tips.2018.04.001
   Vinci M, 2018, NAT MED, V24, P1204, DOI 10.1038/s41591-018-0086-7
   Wang Qi-En, 2015, World J Biol Chem, V6, P57, DOI 10.4331/wjbc.v6.i3.57
   Wang YN, 2014, CANCER RES, V74, P4922, DOI 10.1158/0008-5472.CAN-14-1022
   Wen PY, 2020, CLIN CANCER RES, V26, P1820, DOI 10.1158/1078-0432.CCR-19-2808
   Wu YLD, 2017, TRANSL ONCOL, V10, P221, DOI 10.1016/j.tranon.2016.12.008
   Xu SL, 2015, AM J CANCER RES, V5, P1471
   Ying HQ, 2012, CELL, V149, P656, DOI 10.1016/j.cell.2012.01.058
   Yu JSL, 2016, DEVELOPMENT, V143, P3050, DOI 10.1242/dev.137075
   Yuen HF, 2012, BIOSCIENCE REP, V32, P413, DOI 10.1042/BSR20120050
   Zogg CK, 2014, J ONCOL, V2014, DOI 10.1155/2014/524101
NR 65
TC 10
Z9 10
U1 1
U2 4
PU AMER ASSOC CANCER RESEARCH
PI PHILADELPHIA
PA 615 CHESTNUT ST, 17TH FLOOR, PHILADELPHIA, PA 19106-4404 USA
SN 1541-7786
EI 1557-3125
J9 MOL CANCER RES
JI Mol. Cancer Res.
PD FEB 1
PY 2021
VL 19
IS 2
BP 223
EP 239
DI 10.1158/1541-7786.MCR-20-0464
PG 17
WC Oncology; Cell Biology
WE Science Citation Index Expanded (SCI-EXPANDED)
SC Oncology; Cell Biology
GA QH3YW
UT WOS:000618213800006
PM 33106374
OA Green Accepted, Bronze, Green Submitted
DA 2023-08-27
ER

PT J
AU Martin-Vaquero, P
   da Costa, RC
   Wolk, KE
   Premanandan, C
   Oglesbee, MJ
AF Martin-Vaquero, Paula
   da Costa, Ronaldo C.
   Wolk, Kendra E.
   Premanandan, Christopher
   Oglesbee, Michael J.
TI MRI FEATURES OF GLIOMATOSIS CEREBRI IN A DOG
SO VETERINARY RADIOLOGY & ULTRASOUND
LA English
DT Article
DE brain tumor; dog; glioma; gliomatosis; MRI
AB The features of gliomatosis cerebri involving the brainstem and cerebellum in a 3-year-old dog are described. In magnetic resonance (MR) images, there was diffuse loss of the cerebellar folia and cerebellar gray and white matter contrast. Multiple illdefined T2-hyperintensities were present in the cerebellar parenchyma. A poorly defined, T2-hyperintense mass effect was present ventral to the pons and rostral medulla. No contrast enhancement was noted. Cerebrospinal fluid (CSF) was normal. Postmortem examination was consistent with gliomatosis cerebri, based on compatible histopathology and immunohistochemical findings. Although rare, gliomatosis cerebri should be included as a differential for diffuse infiltrative central nervous system (CNS) lesions. (C) 2012 Veterinary Radiology & Ultrasound.
C1 [Martin-Vaquero, Paula; da Costa, Ronaldo C.] Ohio State Univ, Coll Vet Med, Dept Vet Clin Sci, Columbus, OH 43210 USA.
   [Wolk, Kendra E.; Premanandan, Christopher; Oglesbee, Michael J.] Ohio State Univ, Coll Vet Med, Dept Biosci, Columbus, OH 43210 USA.
C3 University System of Ohio; Ohio State University; University System of
   Ohio; Ohio State University
RP Martin-Vaquero, P (通讯作者)，Ohio State Univ, Coll Vet Med, Dept Vet Clin Sci, Columbus, OH 43210 USA.
EM
OI CASIMIRO DA COSTA, RONALDO/0000-0002-4034-9800
CR [Anonymous], 2009, VET NEUROANATOMY CLI
   ARTIGAS J, 1985, CLIN NEUROPATHOL, V4, P135
   Braun U, 2005, VET J, V170, P381, DOI 10.1016/j.tvjl.2004.09.010
   delCarpioODonovan R, 1996, RADIOLOGY, V198, P831, DOI 10.1148/radiology.198.3.8628879
   Essig M, 2001, EUR RADIOL, V11, P303, DOI 10.1007/s003300000587
   Freund M, 2001, EUR RADIOL, V11, P309, DOI 10.1007/s003300000653
   Fuller GN, 2007, WHO CLASSIFICATION T, V1, P50
   Galan A, 2010, VET COMP ONCOL, V8, P254, DOI 10.1111/j.1476-5829.2010.00219.x
   Gruber A, 2006, J VET MED A, V53, P435, DOI 10.1111/j.1439-0442.2006.00864.x
   Harrison JF, 2010, J NEUROSURG-PEDIATR, V6, P291, DOI 10.3171/2010.5.PEDS09176
   Ide T, 2010, VET PATHOL, V47, P741, DOI 10.1177/0300985810363486
   Inoue T, 2011, NEUROSURG REV, V34, P197, DOI 10.1007/s10143-010-0306-1
   Koestner A, 1999, HISTOLOGICAL CLASSIF, VV, P17
   Louis DN, 2007, ACTA NEUROPATHOL, V114, P547, DOI 10.1007/s00401-007-0278-6
   Nevin S, 1938, BRAIN, V61, P170, DOI 10.1093/brain/61.2.170
   Ponce P, 1998, EUR J RADIOL, V28, P226, DOI 10.1016/S0720-048X(97)00172-1
   Porter B, 2003, VET PATHOL, V40, P97, DOI 10.1354/vp.40-1-97
   Rodenas S, 2011, VET J, V187, P85, DOI 10.1016/j.tvjl.2009.10.011
   Sant'Ana F. J. F. de, 2011, Brazilian Journal of Veterinary Pathology, V4, P58
   SHIN YM, 1993, AM J ROENTGENOL, V161, P859, DOI 10.2214/ajr.161.4.8372774
   Stierstorfer B, 2002, TIERARZTL PRAX K H, V30, P282
   Taillibert S, 2006, J NEURO-ONCOL, V76, P201, DOI 10.1007/s11060-005-5263-0
   Vates GE, 2003, NEUROSURGERY, V53, P261, DOI 10.1227/01.NEU.0000073527.20655.E6
   Yip M, 2003, RADIOGRAPHICS, V23, P247, DOI 10.1148/rg.231025113
NR 24
TC 7
Z9 7
U1 0
U2 8
PU WILEY
PI HOBOKEN
PA 111 RIVER ST, HOBOKEN 07030-5774, NJ USA
SN 1058-8183
EI 1740-8261
J9 VET RADIOL ULTRASOUN
JI Vet. Radiol. Ultrasound
PD MAR-APR
PY 2012
VL 53
IS 2
BP 189
EP 192
DI 10.1111/j.1740-8261.2011.01895.x
PG 4
WC Veterinary Sciences
WE Science Citation Index Expanded (SCI-EXPANDED)
SC Veterinary Sciences
GA 928WU
UT WOS:000303020100011
PM 22413939
OA Green Accepted
DA 2023-08-27
ER

PT J
AU Braslis, KG
   Jones, A
AF Braslis, KG
   Jones, A
TI Ganglioneuroma of the adrenal medulla
SO AUSTRALIAN AND NEW ZEALAND JOURNAL OF SURGERY
LA English
DT Article
DE adrenal tumour; ganglioneuroma
C1 GEELONG HOSP,DEPT UROL,GEELONG,VIC,AUSTRALIA.
C3 Geelong Hospital
CR BRAVO EL, 1989, UROL CLIN N AM, V16, P481
   BRENNAN MF, 1982, CANCER PRINCIPLES PR, P971
   BRETAN PN, 1989, UROL CLIN N AM, V16, P505
   Felix E L, 1981, Curr Probl Cancer, V6, P1, DOI 10.1016/S0147-0272(81)80011-6
   GREENE JP, 1989, UROL CLIN N AM, V16, P487
   KLOOS RT, 1995, ENDOCR REV, V16, P460, DOI 10.1210/er.16.4.460
   LADENSTEIN R, 1995, EUR J CANCER, V31A, P637, DOI 10.1016/0959-8049(95)00069-U
   Samaan N A, 1990, Curr Opin Oncol, V2, P101
   SILVERMAN ML, 1989, UROL CLIN N AM, V16, P417
   STAREN ED, 1995, SURG CLIN N AM, V75, P499, DOI 10.1016/S0039-6109(16)46636-3
NR 10
TC 2
Z9 2
U1 0
U2 1
PU BLACKWELL SCIENCE
PI CARLTON
PA 54 UNIVERSITY ST, P O BOX 378, CARLTON VICTORIA 3053, AUSTRALIA
SN 0004-8682
J9 AUST NZ J SURG
JI Aust. N. Z. J. Surg.
PD NOV
PY 1997
VL 67
IS 11
BP 816
EP 818
DI 10.1111/j.1445-2197.1997.tb04596.x
PG 3
WC Surgery
WE Science Citation Index Expanded (SCI-EXPANDED)
SC Surgery
GA YH237
UT WOS:A1997YH23700025
PM 9397007
DA 2023-08-27
ER

PT J
AU Cuccia, V
   Monges, J
AF Cuccia, V
   Monges, J
TI Thalamic tumors in children
SO CHILDS NERVOUS SYSTEM
LA English
DT Article
DE thalamic tumors; astrocytomas; surgery; prognosis; children
ID BRAIN-STEM; RADIATION-THERAPY; ASTROCYTOMAS; MIDBRAIN
AB Thalamic tumors (TT) merit individual analysis and must not be confused with tumors that, while involving the entire thalamus have a different origin. We analyzed 26 patients who fulfilled our criteria of having ''strictly'' TT. We examined incidence, clinical features, histology, response to treatment (mainly surgery), recurrence rate, mortality and prognosis. We considered that histology and surgical treatment were the most important items related to prognosis. Low-grade tumors (LGT) had a good prognosis, while anaplastic tumors (AT) had a discouraging one; nevertheless both must be operated on. We believe that total removal of LGT is curative and total removal of AT, even if it is not curative, can extend survival by some months. Radiotherapy and chemotherapy seemed to be of little value in our series of TT.
RP Cuccia, V (通讯作者)，HOSP NACL PEDIAT PROF JUAN P GARRAHAN, UNIT PEDIAT NEUROSURG, COMBATE POZOS 1881, RA-1245 BUENOS AIRES, BUENOS AIRES, ARGENTINA.
CR BERNSTEIN M, 1984, J NEUROSURG, V61, P649, DOI 10.3171/jns.1984.61.4.0649
   BLOOM HJG, 1982, INT J RADIAT ONCOL, V8, P1083, DOI 10.1016/0360-3016(82)90056-6
   CHEEK WR, 1966, J NEUROSURG, V24, P505, DOI 10.3171/jns.1966.24.2.0505
   COHEN ME, 1994, BRAIN TUMORS CHILDRE, P303
   EIFEL PJ, 1987, INT J RADIAT ONCOL, V13, P847, DOI 10.1016/0360-3016(87)90098-8
   FORSYTH PA, 1993, CANCER-AM CANCER SOC, V72, P1335, DOI 10.1002/1097-0142(19930815)72:4<1335::AID-CNCR2820720431>3.0.CO;2-E
   FRANZINI A, 1994, NEUROSURGERY, V35, P817, DOI 10.1227/00006123-199411000-00003
   GREENBERGER JS, 1977, RADIOLOGY, V122, P463, DOI 10.1148/122.2.463
   Greenwood J Jr, 1973, Surg Neurol, V1, P29
   GRIGSBY PW, 1989, INT J RADIAT ONCOL, V16, P649, DOI 10.1016/0360-3016(89)90480-X
   GRIGSBY PW, 1987, CANCER, V60, P2901, DOI 10.1002/1097-0142(19871215)60:12<2901::AID-CNCR2820601210>3.0.CO;2-W
   HIROSE G, 1975, ARCH NEUROL-CHICAGO, V32, P740, DOI 10.1001/archneur.1975.00490530062005
   HOFFMAN HJ, 1993, NEUROSURGERY, V33, P964
   KELLY PJ, 1989, NEUROSURGERY, V25, P185, DOI 10.1227/00006123-198908000-00006
   KROUWER HGJ, 1995, J NEUROSURG, V82, P548, DOI 10.3171/jns.1995.82.4.0548
   LEE F, 1975, J NEUROSURG, V43, P65, DOI 10.3171/jns.1975.43.1.0065
   MCKISSOCK W, 1958, BRAIN, V81, P41, DOI 10.1093/brain/81.1.41
   NISHIO S, 1990, NEUROSURGERY, V27, P224, DOI 10.1227/00006123-199008000-00009
   TOVI D, 1961, J NEUROSURG, V18, P730, DOI 10.3171/jns.1961.18.6.0730
   VILLAREJO F, 1994, CHILD NERV SYST, V10, P111, DOI 10.1007/BF00302774
NR 20
TC 45
Z9 54
U1 0
U2 1
PU SPRINGER
PI NEW YORK
PA 233 SPRING ST, NEW YORK, NY 10013 USA
SN 0256-7040
EI 1433-0350
J9 CHILD NERV SYST
JI Childs Nerv. Syst.
PD OCT
PY 1997
VL 13
IS 10
BP 514
EP 520
DI 10.1007/s003810050128
PG 7
WC Clinical Neurology; Pediatrics; Surgery
WE Science Citation Index Expanded (SCI-EXPANDED)
SC Neurosciences & Neurology; Pediatrics; Surgery
GA YF755
UT WOS:A1997YF75500005
PM 9403198
DA 2023-08-27
ER

PT J
AU Mizumoto, M
   Oshiro, Y
   Tsuboi, K
AF Mizumoto, Masashi
   Oshiro, Yoshiko
   Tsuboi, Koji
TI Proton beam therapy for intracranial and skull base tumors
SO TRANSLATIONAL CANCER RESEARCH
LA English
DT Review
DE Proton beam therapy (PBT); brain tumor; glioma; meningioma; chordoma
ID NEUTRON-CAPTURE THERAPY; CARBON ION RADIOTHERAPY; RELATIVE BIOLOGICAL
   EFFECTIVENESS; LOW-GRADE GLIOMA; RADIATION-THERAPY; ATYPICAL
   MENINGIOMAS; STEREOTACTIC RADIOTHERAPY; GLIOBLASTOMA-MULTIFORME;
   PHASE-II; PROTON/PHOTON IRRADIATION
AB Radiotherapy is widely used for treating malignant and benign intracranial or skull base tumors because of the difficulty in performing wide or complete tumor resection due to the involvement of eloquent areas, vital vessels, or cranial nerves. Similarly, local radiotherapy with conventional methods may sometimes be inefficient for treating brain tumors because the tolerance doses for the optic nerves, eyes, and brainstem are strictly limited. Under the circumstances, particle radiotherapy with proton or carbon ion beams has excellent dose localization to the target volume owing to the sharp Bragg peak ionization in the human body. With this property, it is possible to deliver higher doses to tumors of various shapes while preserving the surrounding critical brain structures. Although the efficacy of proton beam therapy (PBT) has been suggested for skull base chordoma and unresectable meningioma, further clinical evidence is required for optimization and standardization of the therapy. In refractory tumors such as glioblastoma multiforme and anaplastic meningioma, it is necessary to deliver high doses to improve tumor control. Under these circumstances, protons or carbon ions have potential to deliver sufficient doses to the target while keeping the healthy brain intact. In this context, we focus on the current status of proton beam radiotherapy for treating intracranial and skull base tumors.
C1 [Mizumoto, Masashi; Oshiro, Yoshiko; Tsuboi, Koji] Univ Tsukuba, Dept Radiat Oncol, Tsukuba, Ibaraki, Japan.
C3 University of Tsukuba
RP Mizumoto, M (通讯作者)，Tennoudai 1-1-1, Tsukuba, Ibaraki, Japan.
EM
RI Mizumoto, Masashi/HJP-4287-2023
OI Mizumoto, Masashi/0000-0001-7837-1301
FU Grants-in-Aid for Scientific Research [24659644] Funding Source: KAKEN
CR Ares C, 2009, INT J RADIAT ONCOL, V75, P1111, DOI 10.1016/j.ijrobp.2008.12.055
   AUSTINSEYMOUR M, 1990, RADIOTHER ONCOL, V17, P29, DOI 10.1016/0167-8140(90)90046-Y
   BARBARO NM, 1987, NEUROSURGERY, V20, P525, DOI 10.1227/00006123-198704000-00003
   Baxendale S, 1998, SEIZURE-EUR J EPILEP, V7, P15, DOI 10.1016/S1059-1311(98)90003-6
   Beltran Chris, 2012, Int J Radiat Oncol Biol Phys, V82, pe281, DOI 10.1016/j.ijrobp.2011.01.005
   Boehling NS, 2012, INT J RADIAT ONCOL, V82, P643, DOI 10.1016/j.ijrobp.2010.11.027
   Boskos C, 2009, INT J RADIAT ONCOL, V75, P399, DOI 10.1016/j.ijrobp.2008.10.053
   Busse PM, 2003, J NEURO-ONCOL, V62, P111, DOI 10.1023/A:1023249224364
   Calaminus G, 2010, KLIN PADIATR, V222, P175, DOI 10.1055/s-0030-1252018
   CASTRO JR, 1985, INT J RADIAT ONCOL, V11, P1795, DOI 10.1016/0360-3016(85)90034-3
   Catton C, 1996, RADIOTHER ONCOL, V41, P67, DOI 10.1016/S0167-8140(96)01805-1
   Chan AW, 2012, TECHNOL CANCER RES T, V11, P607, DOI 10.7785/tcrt.2012.500267
   Cheung MC, 2000, ARCH NEUROL-CHICAGO, V57, P1347, DOI 10.1001/archneur.57.9.1347
   Compter I, 2012, STRAHLENTHER ONKOL, V188, P887, DOI 10.1007/s00066-012-0155-7
   Condra KS, 1997, INT J RADIAT ONCOL, V39, P427, DOI 10.1016/S0360-3016(97)00317-9
   Di Maio S, 2011, J NEUROSURG, V115, P1094, DOI 10.3171/2011.7.JNS11355
   Drappatz J, 2010, INT J RADIAT ONCOL, V78, P85, DOI 10.1016/j.ijrobp.2009.07.1741
   Dziuk TW, 1998, J NEURO-ONCOL, V37, P177, DOI 10.1023/A:1005853720926
   Fitzek MM, 1999, J NEUROSURG, V91, P251, DOI 10.3171/jns.1999.91.2.0251
   Fitzek MM, 2001, INT J RADIAT ONCOL, V51, P131, DOI 10.1016/S0360-3016(01)01589-9
   Foweraker KL, 2007, CLIN ONCOL-UK, V19, P509, DOI 10.1016/j.clon.2007.04.004
   Fuji H, 2011, SKULL BASE-INTERD AP, V21, P201, DOI 10.1055/s-0031-1275636
   FULLER DB, 1988, INT J RADIAT ONCOL, V15, P331, DOI 10.1016/S0360-3016(98)90012-8
   GLAHOLM J, 1990, INT J RADIAT ONCOL, V18, P755, DOI 10.1016/0360-3016(90)90394-Y
   GOLDSMITH BJ, 1994, J NEUROSURG, V80, P195, DOI 10.3171/jns.1994.80.2.0195
   GREINER R, 1990, RADIOTHER ONCOL, V17, P37, DOI 10.1016/0167-8140(90)90047-Z
   Halasz LM, 2011, INT J RADIAT ONCOL, V81, P1428, DOI 10.1016/j.ijrobp.2010.07.1991
   Hauswald H, 2012, RADIAT ONCOL, V7, DOI 10.1186/1748-717X-7-189
   Henriksson R, 2008, RADIOTHER ONCOL, V88, P183, DOI 10.1016/j.radonc.2006.04.015
   Henzel M, 2006, NEUROSURGERY, V59, P1188, DOI 10.1227/01.NEU.0000245626.93215.F6
   Hug EB, 2002, INT J RADIAT ONCOL, V52, P1017, DOI 10.1016/S0360-3016(01)02725-0
   Hug EB, 2002, STRAHLENTHER ONKOL, V178, P10, DOI 10.1007/s00066-002-0874-2
   Hug EB, 2000, J NEURO-ONCOL, V48, P151, DOI 10.1023/A:1006434124794
   Igaki H, 2004, INT J RADIAT ONCOL, V60, P1120, DOI 10.1016/j.ijrobp.2004.05.064
   Joensuu H, 2003, J NEURO-ONCOL, V62, P123, DOI 10.1007/BF02699939
   Kagawa K, 2002, INT J RADIAT ONCOL, V54, P928, DOI 10.1016/S0360-3016(02)02949-8
   Kano H, 2007, J NEURO-ONCOL, V84, P41, DOI 10.1007/s11060-007-9338-y
   Karim ABMF, 1996, INT J RADIAT ONCOL, V36, P549, DOI 10.1016/S0360-3016(96)00352-5
   Kawabata S, 2009, J RADIAT RES, V50, P51, DOI 10.1269/jrr.08043
   Koike S, 2002, J RADIAT RES, V43, P247, DOI 10.1269/jrr.43.247
   Komotar RJ, 2012, J NEUROSURG, V117, P679, DOI 10.3171/2012.7.JNS112113
   Kosaki K, 2012, RADIAT ONCOL, V7, DOI 10.1186/1748-717X-7-44
   KRISTIANSEN K, 1981, CANCER, V47, P649, DOI 10.1002/1097-0142(19810215)47:4<649::AID-CNCR2820470405>3.0.CO;2-W
   Lebrun C, 2004, NEUROLOGY, V62, P1783, DOI 10.1212/01.WNL.0000125196.88449.89
   Lee CT, 2005, INT J RADIAT ONCOL, V63, P362, DOI 10.1016/j.ijrobp.2005.01.060
   Liu JK, 2002, NEUROSURGERY, V50, P950, DOI 10.1097/00006123-200205000-00006
   MacDonald SM, 2008, INT J RADIAT ONCOL, V71, P979, DOI 10.1016/j.ijrobp.2007.11.065
   MacDonald SM, 2011, INT J RADIAT ONCOL, V79, P121, DOI 10.1016/j.ijrobp.2009.10.069
   Maclean J, 2013, INT J RADIAT ONCOL, V85, pE179, DOI 10.1016/j.ijrobp.2012.10.032
   MAGRINI SM, 1992, ACTA ONCOL, V31, P847, DOI 10.3109/02841869209089717
   Mair R, 2011, J NEUROSURG, V115, P811, DOI 10.3171/2011.5.JNS11112
   Milosevic MF, 1996, INT J RADIAT ONCOL, V34, P817, DOI 10.1016/0360-3016(95)02166-3
   MIRIMANOFF RO, 1985, J NEUROSURG, V62, P18, DOI 10.3171/jns.1985.62.1.0018
   Mizoe JE, 2007, INT J RADIAT ONCOL, V69, P390, DOI 10.1016/j.ijrobp.2007.03.003
   Mizoe JE, 2009, SKULL BASE-INTERD AP, V19, P219, DOI 10.1055/s-0028-1114295
   Mizumoto M, 2010, INT J RADIAT ONCOL, V77, P98, DOI 10.1016/j.ijrobp.2009.04.054
   Munzenrider JE, 1999, STRAHLENTHER ONKOL, V175, P57, DOI 10.1007/BF03038890
   Noel G, 2005, ACTA ONCOL, V44, P700, DOI 10.1080/02841860500326257
   Noel G, 2005, INT J RADIAT ONCOL, V62, P1412, DOI 10.1016/j.ijrobp.2004.12.048
   Paganetti H, 2002, INT J RADIAT ONCOL, V53, P407, DOI 10.1016/S0360-3016(02)02754-2
   Pearlman A W, 1970, Am J Roentgenol Radium Ther Nucl Med, V108, P332
   Pearson BE, 2008, NEUROSURG FOCUS, V24, DOI 10.3171/FOC/2008/24/5/E3
   Pehlivan B, 2012, INT J RADIAT ONCOL, V83, P1432, DOI 10.1016/j.ijrobp.2011.10.042
   Prados MD, 2009, J CLIN ONCOL, V27, P579, DOI 10.1200/JCO.2008.18.9639
   RICH TA, 1985, CANCER, V56, P182, DOI 10.1002/1097-0142(19850701)56:1<182::AID-CNCR2820560131>3.0.CO;2-J
   ROMERO J, 1993, RADIOTHER ONCOL, V29, P27, DOI 10.1016/0167-8140(93)90169-9
   SANDBERGWOLLHEIM M, 1991, CANCER, V68, P22, DOI 10.1002/1097-0142(19910701)68:1<22::AID-CNCR2820680105>3.0.CO;2-2
   Santoni R, 1998, INT J RADIAT ONCOL, V41, P59, DOI 10.1016/S0360-3016(98)00031-5
   Schulz-Ertner D, 2007, INT J RADIAT ONCOL, V68, P449, DOI 10.1016/j.ijrobp.2006.12.059
   Selch MT, 2004, INT J RADIAT ONCOL, V59, P101, DOI 10.1016/j.ijrobp.2003.09.003
   Shaw E, 2002, J CLIN ONCOL, V20, P2267, DOI 10.1200/JCO.2002.09.126
   SHAW EG, 1994, NEUROSURGERY, V34, P577, DOI 10.1227/00006123-199404000-00002
   Slater JD, 2012, INT J RADIAT ONCOL, V83, pE633, DOI 10.1016/j.ijrobp.2012.01.079
   Smith SJ, 2007, BRIT J NEUROSURG, V21, P588, DOI 10.1080/02688690701684246
   St Clair WH, 2004, INT J RADIAT ONCOL, V58, P727, DOI 10.1016/S0360-3016(03)01574-8
   Stuer C, 2006, NEUROL MED-CHIR, V46, P118, DOI 10.2176/nmc.46.118
   Stupp R, 2009, LANCET ONCOL, V10, P459, DOI 10.1016/S1470-2045(09)70025-7
   Suit H, 2002, INT J RADIAT ONCOL, V53, P798, DOI 10.1016/S0360-3016(02)02851-1
   Takahashi S, 2009, ACTA NEUROCHIR, V151, P759, DOI 10.1007/s00701-009-0383-5
   TAYLOR BW, 1988, INT J RADIAT ONCOL, V15, P299, DOI 10.1016/S0360-3016(98)90008-6
   van den Bent MJ, 2005, LANCET, V366, P985, DOI 10.1016/S0140-6736(05)67070-5
   Vernimmen FJ, 2001, INT J RADIAT ONCOL, V49, P99, DOI 10.1016/S0360-3016(00)01457-7
   WALKER MD, 1978, J NEUROSURG, V49, P333, DOI 10.3171/jns.1978.49.3.0333
   WALKER MD, 1980, NEW ENGL J MED, V303, P1323, DOI 10.1056/NEJM198012043032303
   Weber DC, 2004, RADIOTHER ONCOL, V71, P251, DOI 10.1016/j.radonc.2004.02.011
   Wenkel E, 2000, INT J RADIAT ONCOL, V48, P1363, DOI 10.1016/S0360-3016(00)01411-5
   Yasuda M, 2012, NEUROSURG REV, V35, P171, DOI 10.1007/s10143-011-0334-5
NR 87
TC 5
Z9 5
U1 0
U2 0
PU AME PUBL CO
PI SHATIN
PA FLAT-RM C 16F, KINGS WING PLAZA 1, NO 3 KWAN ST, SHATIN, HONG KONG
   00000, PEOPLES R CHINA
SN 2218-676X
EI 2219-6803
J9 TRANSL CANCER RES
JI Transl. Cancer Res.
PD APR
PY 2013
VL 2
IS 2
BP 87
EP 96
DI 10.3978/j.issn.2218-676X.2013.04.08
PG 10
WC Oncology
WE Science Citation Index Expanded (SCI-EXPANDED)
SC Oncology
GA V42WD
UT WOS:000209643000006
DA 2023-08-27
ER

PT S
AU Manoranjan, B
   Garg, N
   Bakhshinyan, D
   Singh, SK
AF Manoranjan, Branavan
   Garg, Neha
   Bakhshinyan, David
   Singh, Sheila K.
BE Ehtesham, M
TI The Role of Stem Cells in Pediatric Central Nervous System Malignancies
SO STEM CELL BIOLOGY IN NEOPLASMS OF THE CENTRAL NERVOUS SYSTEM
SE Advances in Experimental Medicine and Biology
LA English
DT Article; Book Chapter
DE Pediatric brain tumor; Cancer stem cell; Brain tumor-initiating cell;
   Medulloblastoma; Ependymoma; Supratentorial primitive neuroectodermal
   tumor; Low-grade glioma; Pediatric glioblastoma; Diffuse intrinsic
   pontine glioma; Atypical teratoid/rhabdoid tumor; Germ cell tumor;
   Craniopharyngioma
ID BETA-CATENIN STATUS; MOUSE MODEL; BRAIN-TUMORS; CHILDHOOD
   MEDULLOBLASTOMA; DISTINCT SUBGROUPS; NESTIN EXPRESSION; DRIVER
   MUTATIONS; GENETIC PROFILES; HISTONE H3.3; SELF-RENEWAL
AB Representing the leading cause of childhood cancer mortality, pediatric brain tumors are comprised of diverse histological features, genetic perturbations, cellular populations, treatment protocols, and clinical outcomes. In this chapter we discuss recent and emerging data that implicate cancer stem cells (also known as brain tumor-initiating cells) in initiating and maintaining the growth of a number of pediatric brain tumors including: medulloblastoma, supratentorial primitive neuroectodermal tumor, atypical teratoid/rhabdoid tumor, ependymoma, low-grade glioma, glioblastoma, diffuse intrinsic pontine glioma, germ cell tumor, and craniopharyngioma. The development of a stem cell framework for the study and treatment of these tumors will enable future clinical approaches to harness the heterogeneous cellular and genomic landscape of these solid tumors as an avenue for developing targeted patient-oriented therapies, thereby improving the overall survivorship for the most lethal childhood cancer.
C1 [Manoranjan, Branavan; Garg, Neha; Bakhshinyan, David; Singh, Sheila K.] McMaster Univ, McMaster Stem Cell & Canc Res Inst, Hamilton, ON L8S 4K1, Canada.
   [Manoranjan, Branavan; Singh, Sheila K.] McMaster Univ, Michael G DeGroote Sch Med, Hamilton, ON L8N 3Z5, Canada.
   [Manoranjan, Branavan; Bakhshinyan, David; Singh, Sheila K.] McMaster Univ, Fac Hlth Sci, Dept Biochem & Biomed Sci, Hamilton, ON L8N 3Z5, Canada.
   [Singh, Sheila K.] McMaster Univ, Fac Hlth Sci, Dept Surg, Hamilton, ON L8N 3Z5, Canada.
C3 McMaster University; McMaster University; McMaster University; McMaster
   University
RP Singh, SK (通讯作者)，McMaster Univ, McMaster Stem Cell & Canc Res Inst, Hamilton, ON L8S 4K1, Canada.
EM
RI Garg, Neha/AAG-5180-2020; Singh, Sheila K/W-2799-2018
OI Garg, Neha/0000-0003-2227-8292; Singh, Sheila K/0000-0003-1272-5300
CR Abdouh M, 2009, J NEUROSCI, V29, P8884, DOI 10.1523/JNEUROSCI.0968-09.2009
   Alimova I, 2013, NEURO-ONCOLOGY, V15, P149, DOI 10.1093/neuonc/nos285
   Andoniadou CL, 2012, ACTA NEUROPATHOL, V124, P259, DOI 10.1007/s00401-012-0957-9
   [Anonymous], PRIM BRAIN CENTR NER
   Avci Z, 2003, PEDIATR PATHOL MOL M, V22, P443, DOI 10.1080/15227950307706
   Bailey P, 1925, ARCH NEURO PSYCHIATR, V14, P192, DOI 10.1001/archneurpsyc.1925.02200140055002
   Ballester LY, 2013, AM J SURG PATHOL, V37, P1357, DOI 10.1097/PAS.0b013e318294e817
   Bamberg M, 1999, J CLIN ONCOL, V17, P2585, DOI 10.1200/JCO.1999.17.8.2585
   Becher OJ, 2010, CANCER RES, V70, P2548, DOI 10.1158/0008-5472.CAN-09-2503
   Beier D, 2007, CANCER RES, V67, P4010, DOI 10.1158/0008-5472.CAN-06-4180
   Biegel Jaclyn A, 2006, Neurosurg Focus, V20, pE11
   Bjerke L, 2013, CANCER DISCOV, V3, P512, DOI 10.1158/2159-8290.CD-12-0426
   Bruggeman SWM, 2007, CANCER CELL, V12, P328, DOI 10.1016/j.ccr.2007.08.032
   Bunin GR, 1998, J NEUROSURG, V89, P547, DOI 10.3171/jns.1998.89.4.0547
   CALAMINUS G, 1994, NEUROPEDIATRICS, V25, P26, DOI 10.1055/s-2008-1071577
   Chen RH, 2010, CANCER CELL, V17, P362, DOI 10.1016/j.ccr.2009.12.049
   Chiou SH, 2008, PLOS ONE, V3, DOI 10.1371/journal.pone.0002090
   Cho YJ, 2011, J CLIN ONCOL, V29, P1424, DOI 10.1200/JCO.2010.28.5148
   Clarke Michael F, 2006, Cancer Res, V66, P9339, DOI 10.1158/0008-5472.CAN-06-3126
   Clement V, 2009, INT J CANCER, V125, P244, DOI 10.1002/ijc.24352
   Clifford SC, 2006, CELL CYCLE, V5, P2666, DOI 10.4161/cc.5.22.3446
   Cocce MC, 2011, J NEURO-ONCOL, V104, P375, DOI 10.1007/s11060-010-0478-0
   Conover JC, 2000, NAT NEUROSCI, V3, P1091, DOI 10.1038/80606
   Dasen JS, 2003, NATURE, V425, P926, DOI 10.1038/nature02051
   Dolecek TA, 2012, NEURO-ONCOLOGY, V14, pv1, DOI 10.1093/neuonc/nos218
   Khuong-Quang DA, 2012, ACTA NEUROPATHOL, V124, P439, DOI 10.1007/s00401-012-0998-0
   Elliott RE, 2010, J NEUROSURG-PEDIATR, V6, P403, DOI 10.3171/2010.8.PEDS09385
   Ellison DW, 2010, ACTA NEUROPATHOL, V120, P305, DOI 10.1007/s00401-010-0726-6
   Ellison DW, 2005, J CLIN ONCOL, V23, P7951, DOI 10.1200/JCO.2005.01.5479
   Fattet S, 2009, J PATHOL, V218, P86, DOI 10.1002/path.2514
   Fuccillo M, 2006, NAT REV NEUROSCI, V7, P772, DOI 10.1038/nrn1990
   Garre ML, 2007, CURR OPIN PEDIATR, V19, P471, DOI 10.1097/MOP.0b013e3282495a22
   Gaspar N, 2010, CANCER RES, V70, P9243, DOI 10.1158/0008-5472.CAN-10-1250
   Gaston-Massuet C, 2011, P NATL ACAD SCI USA, V108, P11482, DOI 10.1073/pnas.1101553108
   Gibson P, 2010, NATURE, V468, P1095, DOI 10.1038/nature09587
   Glinsky GV, 2005, J CLIN INVEST, V115, P1503, DOI 10.1172/JCI23412
   Goodrich LV, 1997, SCIENCE, V277, P1109, DOI 10.1126/science.277.5329.1109
   Grammel D, 2012, ACTA NEUROPATHOL, V123, P601, DOI 10.1007/s00401-012-0961-0
   Hallahan AR, 2004, CANCER RES, V64, P7794, DOI 10.1158/0008-5472.CAN-04-1813
   Hargrove D, 2006, LANCET ONCOL, V7, P241, DOI 10.1016/S1470-2045(06)70615-5
   Hemmati HD, 2003, P NATL ACAD SCI USA, V100, P15178, DOI 10.1073/pnas.2036535100
   HERRMANN HD, 1994, NEUROSURGERY, V34, P524, DOI 10.1227/00006123-199403000-00021
   Hoei-Hansen CE, 2006, J PATHOL, V209, P25, DOI 10.1002/path.1948
   Holsken A, 2010, ACTA NEUROPATHOL, V119, P631, DOI 10.1007/s00401-010-0642-9
   Ichimura K, 2012, NEURO-ONCOLOGY, V14, P90, DOI 10.1093/neuonc/nos204
   Johnson RA, 2010, NATURE, V466, P632, DOI 10.1038/nature09173
   Jones DTW, 2013, NAT GENET, V45, P927, DOI 10.1038/ng.2682
   Kao CL, 2009, INT J RADIAT ONCOL, V74, P219, DOI 10.1016/j.ijrobp.2008.12.035
   Karavitaki N, 2006, ENDOCR REV, V27, P371, DOI 10.1210/er.2006-0002
   Kawauchi D, 2012, CANCER CELL, V21, P168, DOI 10.1016/j.ccr.2011.12.023
   Khatua S, 2012, INDIAN J PEDIATR, V79, P922, DOI 10.1007/s12098-012-0689-9
   Kilday JP, 2009, MOL CANCER RES, V7, P765, DOI 10.1158/1541-7786.MCR-08-0584
   Kool M, 2012, ACTA NEUROPATHOL, V123, P473, DOI 10.1007/s00401-012-0958-8
   Kool M, 2008, PLOS ONE, V3, DOI 10.1371/journal.pone.0003088
   Kyritsis AP, 2010, J NEURO-ONCOL, V96, P143, DOI 10.1007/s11060-009-9951-z
   Lee DY, 2012, CANCER CELL, V22, P131, DOI 10.1016/j.ccr.2012.05.036
   Leung C, 2004, NATURE, V428, P337, DOI 10.1038/nature02385
   Li P, 2013, NAT NEUROSCI, V16, P1737, DOI 10.1038/nn.3553
   Liu Z, 2014, NEURO-ONCOLOGY, DOI [10.1093/neuonc/not244, DOI 10.1093/NEUONC/N0T244]
   Liu ZG, 2013, NEURO-ONCOLOGY, V15, P1173, DOI 10.1093/neuonc/not065
   Louis DN, 2007, ACTA NEUROPATHOL, V114, P547, DOI 10.1007/s00401-007-0278-6
   Ma HI, 2010, CHILD NERV SYST, V26, P293, DOI 10.1007/s00381-009-1016-2
   Mack SC, 2009, CHILD NERV SYST, V25, P1195, DOI 10.1007/s00381-009-0928-1
   Magee JA, 2012, CANCER CELL, V21, P283, DOI 10.1016/j.ccr.2012.03.003
   Manoranjan B, 2013, STEM CELLS, V31, P1266, DOI 10.1002/stem.1401
   Monje M, 2011, P NATL ACAD SCI USA, V108, P4453, DOI 10.1073/pnas.1101657108
   Muller HL, 2013, PITUITARY, V16, P56, DOI 10.1007/s11102-012-0401-0
   Nicolaides T, 2010, J NEURO-ONCOL, V98, P117, DOI 10.1007/s11060-009-0071-6
   Nielsen EH, 2011, J NEURO-ONCOL, V104, P755, DOI 10.1007/s11060-011-0540-6
   Northcott PA, 2011, J CLIN ONCOL, V29, P1408, DOI 10.1200/JCO.2009.27.4324
   Oliver TG, 2005, DEVELOPMENT, V132, P2425, DOI 10.1242/dev.01793
   Panosyan EH, 2010, PEDIATR BLOOD CANCER, V55, P644, DOI 10.1002/pbc.22627
   PARHAM DM, 1994, AM J SURG PATHOL, V18, P1010, DOI 10.1097/00000478-199410000-00005
   Paugh BS, 2010, J CLIN ONCOL, V28, P3061, DOI 10.1200/JCO.2009.26.7252
   Pei YX, 2012, DEVELOPMENT, V139, P1724, DOI 10.1242/dev.050104
   Pei YX, 2012, CANCER CELL, V21, P155, DOI 10.1016/j.ccr.2011.12.021
   Pfister SM, 2010, ACTA NEUROPATHOL, V120, P553, DOI 10.1007/s00401-010-0751-5
   Picard D, 2012, LANCET ONCOL, V13, P838, DOI 10.1016/S1470-2045(12)70257-7
   Pollack IF, 2001, CANCER RES, V61, P7404
   Pomeroy SL, 2002, NATURE, V415, P436, DOI 10.1038/415436a
   Puget S, 2012, PLOS ONE, V7, DOI 10.1371/journal.pone.0030313
   Raffel C, 1997, CANCER RES, V57, P842
   Read TA, 2009, CANCER CELL, V15, P135, DOI 10.1016/j.ccr.2008.12.016
   Reynolds BA, 1996, DEV BIOL, V175, P1, DOI 10.1006/dbio.1996.0090
   REYNOLDS BA, 1992, SCIENCE, V255, P1707, DOI 10.1126/science.1553558
   Rorke LB, 1996, J NEUROSURG, V85, P56, DOI 10.3171/jns.1996.85.1.0056
   Sakuma Y, 2004, CANCER SCI, V95, P716, DOI 10.1111/j.1349-7006.2004.tb03251.x
   Sakurada K, 2008, NEUROSURG REV, V31, P173, DOI 10.1007/s10143-007-0115-3
   Saratsis AM, 2014, ACTA NEUROPATHOL, V127, P881, DOI 10.1007/s00401-013-1218-2
   Schild SE, 1996, CANCER, V78, P2564, DOI 10.1002/(SICI)1097-0142(19961215)78:12<2564::AID-CNCR16>3.0.CO;2-U
   Schroeder KM, 2014, PEDIATR RES, V75, P205, DOI 10.1038/pr.2013.194
   Schuller U, 2008, CANCER CELL, V14, P123, DOI 10.1016/j.ccr.2008.07.005
   Schwartzentruber J, 2012, NATURE, V482, P226, DOI [10.1038/nature10833, 10.1007/s13311-012-0137-6]
   Scotting PJ, 2006, NEUROPATH APPL NEURO, V32, P569, DOI 10.1111/J.1365-2990.2006.00797.x
   Singh SK, 2003, CANCER RES, V63, P5821
   Singh SK, 2004, NATURE, V432, P396, DOI 10.1038/nature03128
   Sotelo C, 2004, PROG NEUROBIOL, V72, P295, DOI 10.1016/j.pneurobio.2004.03.004
   Sun YR, 2009, PLOS ONE, V4, DOI 10.1371/journal.pone.0005498
   Tan C, 2013, J PATHOL, V229, P4, DOI 10.1002/path.4098
   Taylor MD, 2002, NAT GENET, V31, P306, DOI 10.1038/ng916
   Taylor MD, 2005, CANCER CELL, V8, P323, DOI 10.1016/j.ccr.2005.09.001
   Taylor MD, 2012, ACTA NEUROPATHOL, V123, P465, DOI 10.1007/s00401-011-0922-z
   Thakkar JP, 2013, MED ONCOL, V30, DOI 10.1007/s12032-013-0496-9
   Thirant C, 2011, PLOS ONE, V6, DOI 10.1371/journal.pone.0016375
   THOMAS KR, 1990, NATURE, V346, P847, DOI 10.1038/346847a0
   Thompson MC, 2006, J CLIN ONCOL, V24, P1924, DOI 10.1200/JCO.2005.04.4974
   Uchida N, 2000, P NATL ACAD SCI USA, V97, P14720, DOI 10.1073/pnas.97.26.14720
   Versteege I, 1998, NATURE, V394, P203, DOI 10.1038/28212
   Wang VY, 2001, NAT REV NEUROSCI, V2, P484, DOI 10.1038/35081558
   Wang X, 2012, ONCOGENE, V31, P187, DOI 10.1038/onc.2011.232
   Ward RJ, 2009, CANCER RES, V69, P4682, DOI 10.1158/0008-5472.CAN-09-0342
   Wechsler-Reya RJ, 1999, NEURON, V22, P103, DOI 10.1016/S0896-6273(00)80682-0
   Wiese C, 2004, CELL MOL LIFE SCI, V61, P2510, DOI 10.1007/s00018-004-4144-6
   Wilson BG, 2011, NAT REV CANCER, V11, P481, DOI 10.1038/nrc3068
   Wisoff JH, 2008, J NEUROSURG-PEDIATR, V1, P124, DOI 10.3171/PED/2008/1/2/124
   Witt H, 2011, CANCER CELL, V20, P143, DOI 10.1016/j.ccr.2011.07.007
   Wright JH, 1910, J EXP MED, V12, P556, DOI 10.1084/jem.12.4.556
   Wu G, 2012, NAT GENET, V44, P251, DOI 10.1038/ng.1102
   Yachnis AT, 1998, J NEUROPATH EXP NEUR, V57, P961, DOI 10.1097/00005072-199810000-00008
   Yang MH, 2010, NAT CELL BIOL, V12, P982, DOI 10.1038/ncb2099
   Yang ZJ, 2008, CANCER CELL, V14, P135, DOI 10.1016/j.ccr.2008.07.003
   Zakrzewska M, 2011, CHILD NERV SYST, V27, P79, DOI 10.1007/s00381-010-1260-5
   Zhang JH, 2013, NAT GENET, V45, P602, DOI 10.1038/ng.2611
   Zhu XY, 2007, PHYSIOL REV, V87, P933, DOI 10.1152/physrev.00006.2006
NR 124
TC 5
Z9 5
U1 0
U2 7
PU SPRINGER-VERLAG BERLIN
PI BERLIN
PA HEIDELBERGER PLATZ 3, D-14197 BERLIN, GERMANY
SN 0065-2598
EI 2214-8019
BN 978-3-319-16537-0; 978-3-319-16536-3
J9 ADV EXP MED BIOL
JI Adv.Exp.Med.Biol.
PY 2015
VL 853
BP 49
EP 68
DI 10.1007/978-3-319-16537-0_4
D2 10.1007/978-3-319-16537-0
PG 20
WC Cell & Tissue Engineering; Oncology
WE Book Citation Index– Science (BKCI-S); Science Citation Index Expanded (SCI-EXPANDED)
SC Cell Biology; Oncology
GA BD5TR
UT WOS:000361815500004
PM 25895707
DA 2023-08-27
ER

PT J
AU Charles, T
   Morgan, B
AF Charles, Teo
   Morgan, Broggi
TI Surgical outcome of patients considered to have "inoperable" tumors by
   specialized pediatric neuro-oncological multidisciplinary teams
SO CHILDS NERVOUS SYSTEM
LA English
DT Article
DE Brain tumor; Brainstem glioma; Multidisciplinary team; Neuro-oncology;
   Pediatric neurosurgery
ID CENTRAL-NERVOUS-SYSTEM; BREAST-CANCER; BRAIN-TUMORS; UNITED-STATES;
   CARE; MANAGEMENT; CENTRALIZATION; METASTASES; SURGERY
AB Despite the lack of evidence in literature, it is widely felt that patient outcomes will be improved by adopting a multidisciplinary team (MDT) approach to children with brain tumors. This study focuses on a series of pediatric patients treated surgically despite a MDT recommendation against surgery.
   A retrospective study was conducted on all pediatric brain and spinal cord tumor patients operated in a single center from 1999 to 2009. Of the 256 surgical patients, 47 patients (18%) had been previously seen by a MDT who had recommended against surgery. Details of preoperative treatment, diagnosis and clinical status, postoperative diagnosis, early and late outcomes, progression-free survival and overall survival, and parental satisfaction were reviewed.
   There was a single case of surgical mortality, and 14 patients have since died from their primary disease an average of 21 months after surgery. Of the patients who are alive, only four (12.5%) have permanent neurological sequelae despite nine patients presenting in a terminal status. In ten cases, radical removal of the tumor resulted in a change in histological diagnosis, usually from a presumed diagnosis of malignancy to a more benign variety (n = 6). Not a single parent expressed regret over the decision to undergo surgery.
   In the majority of patients, surgical decision making is congruent with the collective opinion of dedicated pediatric neuro-oncological MDT. However, sometimes the surgeon's opinion may be incongruous with MDT recommendation. This series demonstrates the dramatic and favorable potential long-term outcomes that may be achieved with surgery of so-called inoperable lesions.
C1 [Charles, Teo; Morgan, Broggi] Prince Wales Private Hosp, Ctr Minimally Invas Neurosurg, Randwick, NSW 2031, Australia.
RP Charles, T (通讯作者)，Prince Wales Private Hosp, Ctr Minimally Invas Neurosurg, 29 Barker St, Randwick, NSW 2031, Australia.
EM
RI Broggi, Morgan/K-3772-2018
OI Broggi, Morgan/0000-0001-8818-1317
CR Al-Shamy G, 2009, J NEURO-ONCOL, V92, P275, DOI 10.1007/s11060-009-9839-y
   ALLEN JC, 1985, PEDIATR CLIN N AM, V32, P633
   Barker FG, 2005, NEURO-ONCOLOGY, V7, P49, DOI 10.1215/S1152851704000146
   Dixon JM, 1996, BRIT MED J, V312, P1155
   Eichler AF, 2007, ONCOLOGIST, V12, P884, DOI 10.1634/theoncologist.12-7-884
   Gillis CR, 1996, BRIT MED J, V312, P145, DOI 10.1136/bmj.312.7024.145
   Gurney JG, 2001, CURR OPIN ONCOL, V13, P160, DOI 10.1097/00001622-200105000-00005
   Gurney JG, 1999, NIH PUBLICATION, V99-4649
   Gururangan S, 2004, ONCOLOGY-NY, V18, P1649
   HAMMOND GD, 1978, CANCER, V41, P29, DOI 10.1002/1097-0142(197801)41:1<29::AID-CNCR2820410107>3.0.CO;2-D
   Houssami N, 2006, EUR J CANCER, V42, P2480, DOI 10.1016/j.ejca.2006.05.023
   Kaufman Cary S, 2004, Breast J, V10, P469, DOI 10.1111/j.1075-122X.2004.21000.x
   Laurent JP, 1985, J PEDIAT NEUROSCI, V1, P15
   Lefranc F, 2009, ACTA NEUROCHIR, V151, P109, DOI 10.1007/s00701-009-0185-9
   Levy Adam S, 2005, Curr Probl Pediatr Adolesc Health Care, V35, P230, DOI 10.1016/j.cppeds.2005.04.001
   Morris E, 2006, BRIT J CANCER, V95, P979, DOI 10.1038/sj.bjc.6603372
   Rabinowitz B, 2004, ONCOLOGY-NY, V18, P1263
   Reddy A T, 2001, Curr Neurol Neurosci Rep, V1, P137, DOI 10.1007/s11910-001-0009-7
   Schuz J, 2001, MED PEDIATR ONCOL, V36, P274, DOI 10.1002/1096-911X(20010201)36:2<274::AID-MPO1065>3.3.CO;2-4
   Shuster TD, 2000, SURG CLIN N AM, V80, P505, DOI 10.1016/S0039-6109(05)70199-7
   Smith ER, 2004, NEUROSURGERY, V54, P553, DOI 10.1227/01.NEU.0000108421.69822.67
   Tripathy Debu, 2003, Breast J, V9, P60, DOI 10.1046/j.1524-4741.2003.09118.x
   Watson GA, 2001, SEMIN RADIAT ONCOL, V11, P152, DOI 10.1053/srao.2001.21421
   Whelan JM, 2006, BREAST, V15, P119, DOI 10.1016/j.breast.2005.02.010
NR 24
TC 6
Z9 6
U1 1
U2 4
PU SPRINGER
PI NEW YORK
PA 233 SPRING ST, NEW YORK, NY 10013 USA
SN 0256-7040
J9 CHILD NERV SYST
JI Childs Nerv. Syst.
PD SEP
PY 2010
VL 26
IS 9
BP 1219
EP 1225
DI 10.1007/s00381-010-1199-6
PG 7
WC Clinical Neurology; Pediatrics; Surgery
WE Science Citation Index Expanded (SCI-EXPANDED)
SC Neurosciences & Neurology; Pediatrics; Surgery
GA 637UB
UT WOS:000280843100014
PM 20563727
DA 2023-08-27
ER

PT J
AU Poh, B
   Koso, H
   Momota, H
   Komori, T
   Suzuki, Y
   Yoshida, N
   Ino, Y
   Todo, T
   Watanabe, S
AF Poh, Boonmin
   Koso, Hideto
   Momota, Hiroyuki
   Komori, Takashi
   Suzuki, Yutaka
   Yoshida, Nobuaki
   Ino, Yasushi
   Todo, Tomoki
   Watanabe, Sumiko
TI Foxr2 promotes formation of CNS-embryonal tumors in a Trp53-deficient
   background
SO NEURO-ONCOLOGY
LA English
DT Article
DE CNS-embryonal tumor; CNS NB-Foxr2; Foxr2; medulloblastoma; PNET
ID CENTRAL-NERVOUS-SYSTEM; BRAIN-TUMORS; CLASSIFICATION; DIFFERENTIATION;
   IDENTIFICATION; GROWTH; MYC; P53
AB Background. Embryonal tumors in the central nervous system (CNS) are primary, aggressive, and poorly differentiated pediatric brain tumors. We identified forkhead box R2 (Foxr2) as an oncogene for medulloblastoma through a transposon-based insertional mutagenesis screen. Foxr2 translocation has been identified in a subset of human embryonal tumors of the CNS, designated as CNS neuroblastoma with Foxr2 activation (CNS NB-Foxr2); however, the in vivo functions of Foxr2 remain elusive.
   Methods. We analyzed the effect of Foxr2 overexpression in the mouse brain by generating a transgenic strain that expresses Foxr2 in the entire brain under a transformation related protein 53 (Trp53)-deficient background. We performed histological analysis of tumors and characterized tumor-derived sphere-forming cells. We investigated gene expression profiles of tumor-derived cells.
   Results. Foxr2 and Trp53 loss promoted tumor formation in the olfactory bulb (OB) and brainstem (BS). The tumors showed the common morphological features of small round blue cell tumors, exhibiting divergent, mainly neuronal and glial, patterns of differentiation, which corresponds to the definition of CNS-embryonal tumors. Importantly, all mice developed CNS-embryonal tumors. In the OB, early proliferative lesions consisting of oligodendrocyte transcription factor 2 (Olig2+) cells were observed, indicating that Foxr2 expression expanded Olig2+ cells in the OB. Tumor-derived cells formed spheres in vitro and induced tumors that recapitulated the parental tumor upon transplantation, indicating the presence of tumor-initiating cells. Gene expression profiling revealed that OB and BS tumor cells were enriched for the expression of the genes specific to CNS NB-Foxr2.
   Conclusion. Our data demonstrate that Foxr2 plays a causative role in the formation of CNS-embryonal tumors.
C1 [Poh, Boonmin; Koso, Hideto; Watanabe, Sumiko] Univ Tokyo, Inst Med Sci, Div Mol & Dev Biol, Tokyo, Japan.
   [Momota, Hiroyuki; Ino, Yasushi; Todo, Tomoki] Univ Tokyo, Inst Med Sci, Div Innovat Canc Therapy, Tokyo, Japan.
   [Komori, Takashi] Tokyo Metoropolitan Neurologial Hosp, Dept Pathol, Tokyo, Japan.
   [Suzuki, Yutaka] Univ Tokyo, Grad Sch Frontier Sci, Dept Bioinformat & Syst Biol, Tokyo, Japan.
   [Yoshida, Nobuaki] Univ Tokyo, Inst Med Sci, Lab Dev Genet, Tokyo, Japan.
C3 University of Tokyo; University of Tokyo; University of Tokyo;
   University of Tokyo
RP Koso, H (通讯作者)，Univ Tokyo, Inst Med Sci, Div Mol & Dev Biol, Minato Ku, 4-6-1 Shirokanedai, Tokyo 1088639, Japan.
EM
OI KOSO, HIDETO/0000-0002-8986-7617
FU Japan Society for the Promotion of Science (JSPS) [26430108, 15H01482,
   17H04988]; JSPS [26253073, 16K08731]; Japan Agency for Medical Research
   and Development (AMED) [16cm0106218h0001]; Grants-in-Aid for Scientific
   Research [26253073, 17H04988, 15H01482, 26430108, 16K08731] Funding
   Source: KAKEN
FX Funding for this work was provided by the Japan Society for the
   Promotion of Science (JSPS) Grant-in-Aid for Scientific Research (C)
   (26430108), Grant-in-Aid for Scientific Research on Innovative Areas
   (15H01482), and Grant-in-Aid for Young Scientists (A) (17H04988) to HK.
   JSPS Grant-in-Aid for Scientific Research (C) (16K08731) to HM, and JSPS
   Grant-in-Aid for Scientific Research (A) (26253073) and the Platform
   Project for Supporting in Drug Discovery and Life Science Research
   (Platform for Drug Discovery, Informatics, and Structural Life Science)
   from Japan Agency for Medical Research and Development (AMED)
   (16cm0106218h0001) to TT.
CR Eberhart CG, 2005, BMC CANCER, V5, DOI 10.1186/1471-2407-5-19
   Espinosa JS, 2008, J NEUROSCI, V28, P2301, DOI 10.1523/JNEUROSCI.5157-07.2008
   Fleming AJ, 2012, CURR PROB PEDIATR AD, V42, P80, DOI 10.1016/j.cppeds.2011.12.002
   Gessi M, 2011, AM J SURG PATHOL, V35, P573, DOI 10.1097/PAS.0b013e31820f1ce0
   Hayashi Y, 2014, MOL BRAIN, V7, DOI 10.1186/1756-6606-7-44
   Jacques TS, 2010, EMBO J, V29, P222, DOI 10.1038/emboj.2009.327
   Komori T, 2017, NEUROL MED-CHIR, V57, P301, DOI 10.2176/nmc.ra.2017-0010
   Koso H, 2016, CANCER RES, V76, P2254, DOI 10.1158/0008-5472.CAN-15-2308
   Koso H, 2014, CANCER RES, V74, P2351, DOI 10.1158/0008-5472.CAN-13-1523
   Kuribayashi H, 2016, EXP EYE RES, V152, P34, DOI 10.1016/j.exer.2016.09.005
   Lam EWF, 2013, NAT REV CANCER, V13, P482, DOI 10.1038/nrc3539
   Li X, 2016, CELL REP, V16, P487, DOI 10.1016/j.celrep.2016.06.004
   Louis DN, 2016, ACTA NEUROPATHOL, V131, P803, DOI 10.1007/s00401-016-1545-1
   Marino S, 2000, GENE DEV, V14, P994
   Massimino M, 2016, CRIT REV ONCOL HEMAT, V105, P35, DOI 10.1016/j.critrevonc.2016.05.012
   Modzelewska K, 2016, CELL REP, V17, P1255, DOI 10.1016/j.celrep.2016.09.081
   Momota H, 2008, ONCOGENE, V27, P4392, DOI 10.1038/onc.2008.81
   Ostrom QT, 2015, NEURO-ONCOLOGY, V17, P1, DOI [10.1093/neuonc/nov189, 10.1093/neuonc/now207]
   Perry A, 2016, BRAIN TUMOR PATHOL, V33, P157, DOI 10.1007/s10014-016-0266-4
   Picard D, 2012, LANCET ONCOL, V13, P838, DOI 10.1016/S1470-2045(12)70257-7
   Rahrmann EP, 2013, NAT GENET, V45, P756, DOI 10.1038/ng.2641
   Schwalbe EC, 2013, ACTA NEUROPATHOL, V126, P943, DOI 10.1007/s00401-013-1206-6
   Sturm D, 2016, CELL, V164, P1060, DOI 10.1016/j.cell.2016.01.015
   Sumiyoshi K, 2018, CANCER SCI, V109, P1513, DOI 10.1111/cas.13579
   Taylor MD, 2012, ACTA NEUROPATHOL, V123, P465, DOI 10.1007/s00401-011-0922-z
   Wang X, 2016, TUMOR BIOL, V37, P10459, DOI 10.1007/s13277-016-4923-3
   Xu WB, 2017, BIOMED PHARMACOTHER, V87, P471, DOI 10.1016/j.biopha.2016.12.120
   Zheng HW, 2008, NATURE, V455, P1129, DOI 10.1038/nature07443
NR 28
TC 9
Z9 10
U1 1
U2 4
PU OXFORD UNIV PRESS INC
PI CARY
PA JOURNALS DEPT, 2001 EVANS RD, CARY, NC 27513 USA
SN 1522-8517
EI 1523-5866
J9 NEURO-ONCOLOGY
JI Neuro-Oncology
PD AUG
PY 2019
VL 21
IS 8
BP 993
EP 1004
DI 10.1093/neuonc/noz067
PG 12
WC Oncology; Clinical Neurology
WE Science Citation Index Expanded (SCI-EXPANDED)
SC Oncology; Neurosciences & Neurology
GA JH9EE
UT WOS:000493069500007
PM 30976792
OA Bronze, Green Published
DA 2023-08-27
ER

PT J
AU KOFLER, M
   MOROTA, N
   RUSKIN, K
   EPSTEIN, FJ
   DELETIS, V
AF KOFLER, M
   MOROTA, N
   RUSKIN, K
   EPSTEIN, FJ
   DELETIS, V
TI NEUROPHYSIOLOGICAL AIDS IN BRAIN-STEM TUMOR SURGERY
SO CESKA A SLOVENSKA NEUROLOGIE A NEUROCHIRURGIE
LA English
DT Article
DE BRAIN-STEM; BRAIN-STEM TUMOR; FACIAL NERVE; CRANIAL NERVE;
   INTRAOPERATIVE MONITORING; MAPPING; 4TH-VENTRICLE; RHOMBOID FOSSA
ID 4TH VENTRICLE; STEM; STIMULATION; FLOOR
AB We present a method of intraoperative neurophysiological mapping of the floor of the fourth ventricle to localize the nuclei of the facial, glossopharyngeal, vagal, and hypoglossal nerves. In eight patients undergoing brainstem tumor surgery, the fourth ventricular floor was electrically stimulated, and electromyographic responses were recorded with wire electrodes from the orbicularis oculi and orbicularis oris muscles, the lateral posterior pharyngeal wall, and the intrinsic muscles of the tongue, Neurophysiological localization of the cranial motor nuclei provided the surgeon with the necessary anatomical orientation, especially in cases where normal anatomy was severely distorted by tumor or edema. The presence of a muscle response following tumor resection, however, did not always indicate preserved post-operative function in the target muscle. The preliminary results are encouraging, but at the same time, reveal some limitations of the current technique.
C1 UNIV INNSBRUCK, DEPT NEUROL, A-6020 INNSBRUCK, AUSTRIA.
   NYU, MED CTR, DEPT ANESTHESIOL, NEW YORK, NY USA.
   NYU, MED CTR, DIV PEDIAT NEUROSURG, NEW YORK, NY USA.
C3 University of Innsbruck; New York University; New York University
RI Ruskin, Keith/AAR-8777-2021; Kofler, Markus/P-5585-2019
OI Kofler, Markus/0000-0002-2962-0903
CR AGNEW WF, 1987, NEUROSURGERY, V20, P143
   Carpenter MB., 1983, HUMAN NEUROANATOMY M, V8, P315
   EPSTEIN F, 1986, J NEUROSURG, V64, P11, DOI 10.3171/jns.1986.64.1.0011
   EPSTEIN F, 1987, J NEUROSURG, V67, P483, DOI 10.3171/jns.1987.67.4.0483
   EPSTEIN FJ, 1993, J NEUROSURG, V78, P408, DOI 10.3171/jns.1993.78.3.0408
   EPSTEIN FJ, 1990, NEUROSURG CLIN N AM, P111
   Fahlbusch R, 1991, Zentralbl Neurochir, V52, P25
   KASHIWAGI S, 1990, J NEUROSURG, V72, P27, DOI 10.3171/jns.1990.72.1.0027
   KATSUTA T, 1993, NEUROSURGERY, V32, P861, DOI 10.1227/00006123-199305000-00025
   KONOVALOV AN, 1990, J NEUROSURG, V73, P181, DOI 10.3171/jns.1990.73.2.0181
   KYOSHIMA K, 1993, J NEUROSURG, V78, P987, DOI 10.3171/jns.1993.78.6.0987
   LANG J, 1991, ACTA NEUROCHIR, V113, P84, DOI 10.1007/BF01402120
   POLLACK IF, 1993, J NEUROSURG, V78, P859, DOI 10.3171/jns.1993.78.6.0859
   STRAUSS C, 1993, J NEUROSURG, V79, P393, DOI 10.3171/jns.1993.79.3.0393
NR 14
TC 0
Z9 0
U1 0
U2 1
PU CZECH MEDICAL SOC
PI PRAGUE 2
PA SOKOLSKA 31, PRAGUE 2 120 26, CZECH REPUBLIC
SN 1210-7859
J9 CESK SLOV NEUROL N
JI Cesk. Slov. Neurol. Neurochir.
PY 1995
VL 58
IS 5
BP 228
EP 232
PG 5
WC Neurosciences; Surgery
WE Science Citation Index Expanded (SCI-EXPANDED)
SC Neurosciences & Neurology; Surgery
GA RZ143
UT WOS:A1995RZ14300003
DA 2023-08-27
ER

PT J
AU Skrap, M
   Melatini, A
   Strukelj, S
AF Skrap, M
   Melatini, A
   Strukelj, S
TI The stereotactic volumetric information: Its role in two-step resection
   of brainstem and thalamic giant tumor. Report of three cases and
   technical note.
SO SURGICAL NEUROLOGY
LA English
DT Article
DE stereotactic resection; brain stem tumor
ID ASSISTED RESECTION; LESIONS; ASTROCYTOMAS; MANAGEMENT; BIOPSIES;
   CHILDREN; GLIOMAS; SURGERY
AB BACKGROUND
   A compact intracerebral tumoral lesion is usually considered to be completely resectable. Nevertheless, radical resection of a huge lesion located in a critical area may damage the surrounding compressed brain tissue. In cases with a good prognosis, a two-step removal appears to be a safer strategy.
   METHODS
   In three cases, two with huge brain stem lesions and one with a thalamic lesion, a two-step volumetric stereotactic resection was planned. This strategy allowed us to evaluate the amount of tumor to be removed during the first procedure and to have, during the second operation, an exact definition of the reduced mass with regard to the scar tissue and postoperative adhesions. Furthermore, we avoided significant shifting of the cerebral structures during both procedures.
   RESULTS
   There was a very good final recovery in the cases with brain stem lesions and a minimal deficit in the patient with the thalamic lesion. The patient with a mesencephalic lesion remained comatose for almost 2 days after the first procedure, confirming our fears about too radical a one-step resection.
   CONCLUSIONS
   We think that by using current techniques, it is possible to remove a well circumscribed lesion regardless of its position. This is probably easier with giant lesions where a safe trajectory can be planned. In these cases, with lesions located in very critical areas but with a good prognosis, a two-step resection appears to be a good option. (C) 1999 by Elsevier Science Inc.
C1 Osped S Maria della Misericordia, Div Neurochirurg, Udine, Italy.
   Osped Cattinara, Div Neurochirurg, Trieste, Italy.
C3 Hospital Santa Maria della Misericordia
RP Skrap, M (通讯作者)，Osped S Maria della Misericordia, Div Neurochirurg, Udine, Italy.
RI Skrap, Miran/J-2092-2012; skrap, miran/AAB-1708-2022
OI skrap, miran/0000-0002-6592-6416
CR ALBRIGHT AL, 1993, NEUROSURGERY, V33, P1026
   Bricolo A, 1991, Acta Neurochir Suppl (Wien), V53, P148
   DRAKE JM, 1991, NEUROSURGERY, V29, P27, DOI 10.1227/00006123-199107000-00005
   EPSTEIN F, 1986, J NEUROSURG, V64, P11, DOI 10.3171/jns.1986.64.1.0011
   EPSTEIN F, 1987, J NEUROSURG, V67, P483, DOI 10.3171/jns.1987.67.4.0483
   Fahlbusch R, 1991, Acta Neurochir Suppl (Wien), V53, P37
   FRANZINI A, 1994, NEUROSURGERY, V35, P817, DOI 10.1227/00006123-199411000-00003
   HEFFEZ DS, 1990, NEUROSURGERY, V27, P789, DOI 10.1227/00006123-199011000-00018
   Hitchcock E, 1991, Acta Neurochir Suppl (Wien), V52, P9
   HOFFMAN HJ, 1993, NEUROSURGERY, V33, P964
   HOOD TW, 1989, NEUROSURGERY, V24, P373, DOI 10.1227/00006123-198903000-00011
   Kelly P J, 1991, Acta Neurochir Suppl (Wien), V52, P26
   KELLY PJ, 1987, J NEUROSURG, V66, P865, DOI 10.3171/jns.1987.66.6.0865
   Kelly PJ, 1991, TUMOR STEREOTAXIS
   KRATIMENOS GP, 1992, ACTA NEUROCHIR, V116, P164, DOI 10.1007/BF01540871
   MORITA A, 1993, NEUROSURGERY, V32, P920, DOI 10.1227/00006123-199306000-00006
   Pendl G, 1991, Acta Neurochir Suppl (Wien), V53, P137
   RAJSHEKHAR V, 1995, J NEUROSURG, V82, P976, DOI 10.3171/jns.1995.82.6.0976
   Skrap M, 1996, ACTA NEUROCHIR, V138, P210, DOI 10.1007/BF01411363
NR 19
TC 1
Z9 1
U1 0
U2 0
PU ELSEVIER SCIENCE INC
PI NEW YORK
PA 655 AVENUE OF THE AMERICAS, NEW YORK, NY 10010 USA
SN 0090-3019
J9 SURG NEUROL
JI Surg. Neurol.
PD NOV
PY 1999
VL 52
IS 5
BP 515
EP 523
DI 10.1016/S0090-3019(99)00126-3
PG 9
WC Clinical Neurology; Surgery
WE Science Citation Index Expanded (SCI-EXPANDED)
SC Neurosciences & Neurology; Surgery
GA 263GP
UT WOS:000084117300029
PM 10595773
DA 2023-08-27
ER

PT J
AU Aminpour, S
   Tinling, SP
   Brodie, HA
AF Aminpour, S
   Tinling, SP
   Brodie, HA
TI Role of tumor necrosis factor-alpha in sensorineural hearing loss after
   bacterial meningitis
SO OTOLOGY & NEUROTOLOGY
LA English
DT Article
DE hearing loss; meningitis; disease; tumor necrosis factor-alpha
ID EXPERIMENTAL PNEUMOCOCCAL MENINGITIS; BLOOD-BRAIN-BARRIER;
   CEREBROSPINAL-FLUID; INNER-EAR; ENDOTHELIAL-CELLS; UNITED-STATES;
   TNF-ALPHA; INFLAMMATION; INDUCTION; CHILDREN
AB Hypothesis: Blockade of tumor necrosis factor-alpha with tumor necrosis factor-alpha antibody will reduce the extent of cochlear injury and hearing loss associated with Streptococcus pneumoniae meningitis.
   Background: Inflammatory mediators play a significant role in the morbidity associated with bacterial meningitis, including hearing loss and labyrinthitis ossificans. Previous studies have shown the attenuation of hearing loss by the nonspecific blockade of such pathways.
   Methods: Fifty Mongolian gerbils were divided into four groups. Auditory brainstem response testing was conducted to measure hearing thresholds. Streptococcus pneumoniae meningitis was induced in Groups I and 2. Group 2 was then given a single intraperitoneal injection of tumor necrosis factor-alpha antibody, whereas Group I received phosphate-buffered saline. Uninfected animals in Groups 3 and 4 were implanted with osmotic pumps that delivered a continuous 8-day intrathecal flow of either tumor necrosis factor-alpha (Group 4) or phosphate-buffered saline (Group 3). After 6 weeks, auditory brainstem response testing was repeated. The cochleas were harvested and analyzed histomorphometrically.
   Results: Group 2 animals with Streptococcus pneumoniae meningitis that also received tumor necrosis factor-alpha antibody developed significantly less hearing loss than Group I animals with meningitis alone. The decrease in the average threshold at 4, 8, 16, and 32 kHz was 31, 30, 25, and 28 dB sound pressure level, respectively (p < 0.0092 for each). Furthermore, histomorphometric analysis showed significantly less damage to the organ of Corti, spiral ganglion, spiral ligament, and stria vascularis in Group 2. Conversely, tumor necrosis factor-alpha induced meningitis animals (Group 3) showed increased hearing loss compared with phosphate-buffered saline controls (Group 4), with p < 0.0001 at all frequencies.
   Conclusion: Tumor necrosis factor-alpha plays an important role in cochlear injury after bacterial meningitis. Blockade of tumor necrosis factor-alpha reduces postmeningitic hearing loss and cochlear injury. Induction of meningitis with intrathecal tumor necrosis factor-alpha also resulted in hearing loss and cochlear injury similar to bacterial meningitis.
C1 Univ Calif Davis, Dept Otolaryngol Head & Neck Surg, Sch Med, Davis, CA 95616 USA.
C3 University of California System; University of California Davis
RP Brodie, HA (通讯作者)，Univ Calif Davis, Dept Otolaryngol, Sch Med, 1515 Newton Court, Davis, CA 95616 USA.
EM
CR Adams JC, 2002, OTOL NEUROTOL, V23, P316, DOI 10.1097/00129492-200205000-00015
   BERLOW SJ, 1980, LARYNGOSCOPE, V90, P1445, DOI 10.1288/00005537-198009000-00004
   BEUTLER B, 1986, SCIENCE, V232, P977, DOI 10.1126/science.3754653
   BHATT S, 1989, LARYNGOSCOPE, V77, P1974
   Bogdan I, 1997, J INFECT DIS, V176, P693, DOI 10.1086/514092
   Brodie HA, 1998, OTOLARYNG HEAD NECK, V118, P15, DOI 10.1016/S0194-5998(98)70369-9
   Chandross KJ, 1996, MOL CELL NEUROSCI, V7, P479, DOI 10.1006/mcne.1996.0035
   DODGE PR, 1984, NEW ENGL J MED, V311, P869, DOI 10.1056/NEJM198410043111401
   EISENBERG LS, 1984, OTOLARYNG HEAD NECK, V92, P700, DOI 10.1177/019459988409200619
   FARZIN RA, 1997, ACTA OTOLARYNGOL STO, V117, P329
   FINITZOHIEBER Y, 1981, INT J PEDIATR OTORHI, V3, P275
   FISCHER H, 1985, J BACTERIOL, V163, P46, DOI 10.1128/JB.163.1.46-54.1985
   FRIE K, 1987, EUR J IMMUNOL, V17, P1271
   FRIEDMANN I, 1993, PATHOLOGY EAR, P752
   Gasque P, 1998, J IMMUNOL, V160, P3543
   Ge NN, 2004, OTOLARYNG HEAD NECK, V131, P563, DOI 10.1016/j.otohns.2004.03.046
   HARRIS JP, 1983, OTOLARYNG HEAD NECK, V91, P18, DOI 10.1177/019459988309100105
   HINOJOSA R, 1980, ARCH OTOLARYNGOL, V106, P193
   HUMMELL DS, 1985, INFECT IMMUN, V47, P384, DOI 10.1128/IAI.47.2.384-387.1985
   KEANE WM, 1979, ARCH OTOLARYNGOL, V105, P9
   KEITHLEY EM, 1989, LARYNGOSCOPE, V99, P409, DOI 10.1288/00005537-198904000-00010
   Keithley EM, 1996, LARYNGOSCOPE, V106, P341, DOI 10.1097/00005537-199603000-00019
   KIKUCHI T, 1995, ANAT EMBRYOL, V191, P101, DOI 10.1007/BF00186783
   KIM KS, 1992, LYMPHOKINE CYTOK RES, V11, P293
   Koedel U, 1999, J NEUROIMMUNOL, V96, P190, DOI 10.1016/S0165-5728(99)00033-8
   LARRICK JW, 1990, FASEB J, V4, P3214
   Leib SL, 2001, BRAIN, V124, P1734, DOI 10.1093/brain/124.9.1734
   LIEBERMAN AP, 1989, P NATL ACAD SCI USA, V86, P6348, DOI 10.1073/pnas.86.16.6348
   Lopez-Cortes LF, 2000, CRIT CARE MED, V28, P215, DOI 10.1097/00003246-200001000-00035
   Mabbott NA, 2002, J VIROL, V76, P5131, DOI 10.1128/JVI.76.10.5131-5139.2002
   MCALLISTER CK, 1975, J INFECT DIS, V132, P355, DOI 10.1093/infdis/132.4.355
   MOVAT HZ, 1987, J LAB CLIN MED, V110, P668
   MUSTAFA MM, 1989, J PEDIATR-US, V115, P208, DOI 10.1016/S0022-3476(89)80067-8
   NADOL JB, 1978, LARYNGOSCOPE, V88, P739
   Nietsch HH, 2000, J BIOL CHEM, V275, P20556, DOI 10.1074/jbc.M002535200
   NOVAK MA, 1990, OTOLARYNG HEAD NECK, V103, P351, DOI 10.1177/019459989010300303
   OHGA S, 1994, ARCH DIS CHILD, V70, P123, DOI 10.1136/adc.70.2.123
   PFISTER HW, 1994, CLIN INFECT DIS, V19, P63
   QUAGLIARELLO V, 1992, NEW ENGL J MED, V327, P864, DOI 10.1056/NEJM199209173271208
   ROSENHALL U, 1978, ACTA OTO-LARYNGOL, V85, P243
   SAEZLLORENS X, 1990, J PEDIATR-US, V116, P671, DOI 10.1016/S0022-3476(05)82647-2
   Satoh H, 2002, LARYNGOSCOPE, V112, P1627, DOI 10.1097/00005537-200209000-00019
   SCHAAD UB, 1995, CLIN INFECT DIS, V20, P685, DOI 10.1093/clinids/20.3.685
   SCHLECH WF, 1985, JAMA-J AM MED ASSOC, V253, P1749, DOI 10.1001/jama.253.12.1749
   STUTMAN HR, 1987, CLIN PEDIATR, V26, P431, DOI 10.1177/000992288702600901
   Tang RB, 2001, CHILD NERV SYST, V17, P453, DOI 10.1007/s003810000422
   TRACEY KJ, 1988, J INFECT DIS, V157, P413, DOI 10.1093/infdis/157.3.413
   Tsao N, 2002, FEMS IMMUNOL MED MIC, V32, P133, DOI 10.1016/S0928-8244(01)00290-5
   TSUJIMOTO M, 1986, BIOCHEM BIOPH RES CO, V137, P1094, DOI 10.1016/0006-291X(86)90337-2
   TUOMANEN E, 1985, J INFECT DIS, V151, P535, DOI 10.1093/infdis/151.3.535
   TUOMANEN E, 1985, J INFECT DIS, V151, P859, DOI 10.1093/infdis/151.5.859
   TUOMANEN E, 1993, SCI AM, V268, P80, DOI 10.1038/scientificamerican0293-80
   TUOMANEN E, 1987, J INFECT DIS, V155, P985, DOI 10.1093/infdis/155.5.985
   TUOMANEN E, 1986, MICROB PATHOGENESIS, V1, P15, DOI 10.1016/0882-4010(86)90028-8
   TUOMANEN E, 1987, AM REV RESPIR DIS, V135, P869, DOI 10.1164/arrd.1987.135.4.869
   TUOMANEN E, 1987, PEDIATR INFECT DIS J, V6, P1146
   TUOMANEN E, 1997, ANN NY ACAD SCI, V797, P42
   Tuomanen EI, 1997, PEDIATR RES, V42, P253, DOI 10.1203/00006450-199709000-00001
   van Rijen HVM, 1998, CYTOKINE, V10, P258, DOI 10.1006/cyto.1997.0287
   VERNON M, 1967, LARYNGOSCOPE, V77, P1856, DOI 10.1288/00005537-196710000-00008
   WAAGE A, 1989, J EXP MED, V170, P1859, DOI 10.1084/jem.170.6.1859
   Wang XB, 2003, OTOL NEUROTOL, V24, P52, DOI 10.1097/00129492-200301000-00012
   WENGER JD, 1990, J INFECT DIS, V162, P1316, DOI 10.1093/infdis/162.6.1316
   WOOLF NK, 1986, ACTA OTO-LARYNGOL, V102, P353, DOI 10.3109/00016488609119418
   YEUNG A, IN PRESS OTOLARYNGOL
NR 65
TC 50
Z9 53
U1 0
U2 2
PU LIPPINCOTT WILLIAMS & WILKINS
PI PHILADELPHIA
PA TWO COMMERCE SQ, 2001 MARKET ST, PHILADELPHIA, PA 19103 USA
SN 1531-7129
EI 1537-4505
J9 OTOL NEUROTOL
JI Otol. Neurotol.
PD JUL
PY 2005
VL 26
IS 4
BP 602
EP 609
DI 10.1097/01.mao.0000178121.28365.0d
PG 8
WC Clinical Neurology; Otorhinolaryngology
WE Science Citation Index Expanded (SCI-EXPANDED)
SC Neurosciences & Neurology; Otorhinolaryngology
GA 957ZL
UT WOS:000231412000009
PM 16015154
DA 2023-08-27
ER

PT J
AU Su, L
   Gao, PY
AF Su, Lu
   Gao, Peiyi
TI Volume mismatch indicates tumors in paramedial bithalamic diseases: a
   retrospective study
SO FRONTIERS IN NEUROLOGY
LA English
DT Article
DE paramedial bilateral thalamus; volume mismatch; discriminating; tumors;
   non-tumors
ID RESONANCE-IMAGING FINDINGS; ENCEPHALOPATHY; LESIONS; MRI
AB ObjectiveThis study aimed to investigate the diagnostic performance of volume mismatch sign on discriminating paramedial bithalamic tumors from non-tumors. MethodsIn this study, we recruited patients with tumors or non-tumors of the paramedial bithalamus. We confirmed the diagnosis by pathology, laboratory tests documented in medical records, medical imaging at the baseline, or through at least 1 year of follow-up. Cases with paramedial thalamic lesions on only one side or purely midbrain illnesses were excluded. Additionally, any case without involvement of the medial thalami (i.e., those with one or both-sided anterior, lateral, or posterior thalamic lesions) was excluded. Two neuroradiologists were trained independently to evaluate volume mismatch sign on magnetic resonance T2-weighted images or T2 fluid-attenuated inversion recovery images. A positive volume mismatch sign means that the ratio of the larger-sided lesion volume to the smaller-sided lesion volume is >150%. The volume of each lesion was calculated by multiplying the anteroposterior diameter by the left-right diameter and by the height of the lesion and then dividing by 2. The kappa value was calculated to show the consistency between the two observers. The chi-square test was used to evaluate differences in volume mismatch sign between the bilthalamic midline tumor and non-tumor groups. The positive (PPV) and negative (NPV) predictive values, sensitivity, and specificity were calculated to evaluate the ability of volume mismatch sign to differentiate paramedial bilateral thalamus tumors from non-tumors. A two-tailed P & LE; 0.05 was considered to be statistically significant. The analyses were performed using the statistical software SPSS version 26. ResultsA total of 96 patients were enrolled in this study between March 2012 and October 2022. A high agreement between the two observers on the volume mismatch sign of bilateral paramedian thalamic diseases was found, and the Kappa value was 0.828. A statistically significant difference was observed for the volume mismatch sign between the paramedial bithalamic tumor and the non-tumorous groups (& chi;(2) = 35.465, P < 0.001). The presence of volume mismatch sign in paramedial bithalamic illnesses predicted the presence of tumors with a sensitivity and specificity of 69.2% and 90.9%, respectively, and PPV and NPV were 90.0% and 71.4%. ConclusionVolume mismatch sign may indicate tumors in paramedian bithalamic diseases.
C1 [Su, Lu; Gao, Peiyi] Capital Med Univ, Beijing Tiantan Hosp, Beijing, Peoples R China.
   [Gao, Peiyi] Neuroradiol Ctr Beijing, Neurosurg Inst, Beijing, Peoples R China.
C3 Capital Medical University
RP Gao, PY (通讯作者)，Capital Med Univ, Beijing Tiantan Hosp, Beijing, Peoples R China.; Gao, PY (通讯作者)，Neuroradiol Ctr Beijing, Neurosurg Inst, Beijing, Peoples R China.
EM
CR Adams LC, 2017, SCI REP-UK, V7, DOI 10.1038/s41598-017-15860-1
   Araujo R, 2016, PEDIATR NEUROL, V65, P96, DOI 10.1016/j.pediatrneurol.2016.08.008
   Autti T, 2007, NEURORADIOLOGY, V49, P571, DOI 10.1007/s00234-007-0220-6
   Badar F, 2011, NEUROL INDIA, V59, P108, DOI 10.4103/0028-3886.84345
   Cinalli G, 2018, J NEUROSURG-PEDIATR, V21, P247, DOI 10.3171/2017.7.PEDS16463
   Cocozza S, 2018, INSIGHTS IMAGING, V9, P1077, DOI 10.1007/s13244-018-0664-8
   Dardiotis E, 2009, J CHILD NEUROL, V24, P1001, DOI 10.1177/0883073808331364
   Gururaj A, 2005, J CHILD NEUROL, V20, P57, DOI 10.1177/08830738050200010901
   Hoch MJ, 2019, AM J NEURORADIOL, V40, P1095, DOI 10.3174/ajnr.A6088
   Johnsen SD, 2006, PEDIATR NEUROL, V34, P405, DOI 10.1016/j.pediatrneurol.2005.10.005
   Khanna PC, 2011, AM J ROENTGENOL, V197, P1449, DOI 10.2214/AJR.11.6702
   Kim E, 2007, AM J NEURORADIOL, V28, P1652, DOI 10.3174/ajnr.A0655
   Kumar D, 2014, NEUROL INDIA, V62, P475, DOI 10.4103/0028-3886.141311
   Lazzaro NA, 2010, AM J NEURORADIOL, V31, P1283, DOI 10.3174/ajnr.A2044
   Linn J, 2007, ROFO-FORTSCHR RONTG, V179, P234, DOI 10.1055/s-2007-962857
   Mach JC, 2020, CLIN NEUROL NEUROSUR, V196, DOI 10.1016/j.clineuro.2020.106034
   Marjelund S, 2004, ACTA RADIOL, V45, P88, DOI 10.1080/02841850410003356
   Nishijima H, 2022, NEUROLOGY, V98, P204, DOI 10.1212/WNL.0000000000013091
   Ohsaka M, 2006, PEDIATR NEUROL, V34, P160, DOI 10.1016/j.pediatrneurol.2005.07.007
   Pols T, 2019, NEUROPEDIATRICS, V50, P152, DOI 10.1055/s-0039-1683864
   Poyiadji N, 2020, RADIOLOGY, V296, pE119, DOI 10.1148/radiol.2020201187
   Reis E, 2020, J BELG SOC RADIOL, V104, DOI 10.5334/jbsr.2017
   Roitberg BZ, 2002, ACTA NEUROCHIR, V144, P301, DOI 10.1007/s007010200040
   Sahin H, 2016, SURG RADIOL ANAT, V38, P711, DOI 10.1007/s00276-016-1619-8
   Schmahmann JD, 2003, STROKE, V34, P2264, DOI 10.1161/01.STR.0000087786.38997.9E
   Smith AB, 2009, AM J ROENTGENOL, V192, pW53, DOI 10.2214/AJR.08.1585
   Steenweg ME, 2010, BRAIN, V133, P2971, DOI 10.1093/brain/awq257
   Steinborn M, 2004, PEDIATR RADIOL, V34, P805, DOI 10.1007/s00247-004-1289-2
   U-King-Im JM, 2011, AM J NEURORADIOL, V32, P413, DOI 10.3174/ajnr.A2290
   Van Cauter S, 2020, NEURORADIOLOGY, V62, P1565, DOI 10.1007/s00234-020-02511-y
   Wu XJ, 2015, MEDIAT INFLAMM, V2015, DOI 10.1155/2015/792578
   Zawadzki R, 2017, POL J RADIOL, V82, P742, DOI 10.12659/PJR.903940
   Zuccoli G, 2009, AM J NEURORADIOL, V30, P171, DOI 10.3174/ajnr.A1280
NR 33
TC 0
Z9 0
U1 0
U2 0
PU FRONTIERS MEDIA SA
PI LAUSANNE
PA AVENUE DU TRIBUNAL FEDERAL 34, LAUSANNE, CH-1015, SWITZERLAND
SN 1664-2295
J9 FRONT NEUROL
JI Front. Neurol.
PD JUL 25
PY 2023
VL 14
AR 1154823
DI 10.3389/fneur.2023.1154823
PG 7
WC Clinical Neurology; Neurosciences
WE Science Citation Index Expanded (SCI-EXPANDED)
SC Neurosciences & Neurology
GA O5DL4
UT WOS:001044014300001
PM 37560447
DA 2023-08-27
ER

PT J
AU Bordi, C
AF Bordi, C
TI Multiple endocrine neoplasia (MEN)-associated tumours
SO DIGESTIVE AND LIVER DISEASE
LA English
DT Article; Proceedings Paper
CT Meeting on Somatostatin Receptors
CY OCT, 2001
CL Borgo Monastero, ITALY
DE endocrine tumours; MEN 1 gene; MEN 1 syndrome; MEN 2 syndrome;
   preneoplastic endocrine lesions; pathology; RET gene
ID MEDULLARY-THYROID CARCINOMA; ZOLLINGER-ELLISON SYNDROME; PARATHYROID
   TUMORS; CLINICAL-FEATURES; ALLELIC DELETIONS; CHROMOSOME 11Q13; MEN1
   GENE; TYPE-1; GASTRINOMAS; PROGNOSIS
AB The pathological changes of turnouts associated with the two main MEN syndromes, types I and 2, and their relation with the genetic defects responsible for the individual syndromes are reviewed. MEN I-associated tumours, affecting mainly the pituitary, the parathyroids and the pancreas. are due to inactivation of the MEN 1 oncosuppressor gene located in 11q13. Although at least one peculiar phenotype of MEN I syndrome is known, no genotype-phenotype relation has been disclosed. MEN 2-associated turnouts, affecting mainly the thyroid C cells, the parathyroids and the adrenal medulla, are due to oncogenic point mutations of the RET proto-oncogene located in 10q11.2. The two main phenotypes of the MEN 2 syndrome, known as MEN 2A and MEN 213, are associated with different mutations of the RET oncogene, mostly located on exons 10 or 11 in MEN 2A and in codon 918 of exon 16 in MEN 2B. Independent of the type of MEN disease, the development of tumours follows a substantially similar pattern in all MEN target organs. The initial lesion is a diffuse hyperplastic proliferation of the affected endocrine tissue with bilateral involvement of pair organs, followed by development of multiple micro- and, eventually, macronodular lesions. Such association of endocrine hyperplastic changes and turnouts is suggestive of a MEN condition in patients with apparently sporadic tumours. LOH investigations in MEN I cases indicate that each multiple tumour is the result of an independent genetic event. (C) 2003 Editrice Gastroenterologica Italiana S.r.l. Published by Elsevier Ltd. All rights reserved.
C1 Univ Parma, Anat Pathol Sect, Dept Pathol & Lab Med, I-43100 Parma, Italy.
C3 University of Parma; University Hospital of Parma
RP Bordi, C (通讯作者)，Univ Parma, Anat Pathol Sect, Dept Pathol & Lab Med, I-43100 Parma, Italy.
EM
CR Agarwal SK, 1997, HUM MOL GENET, V6, P1169, DOI 10.1093/hmg/6.7.1169
   ARNOLD A, 1995, J CLIN INVEST, V95, P2047, DOI 10.1172/JCI117890
   BORDI C, 1987, AM J CLIN PATHOL, V88, P153, DOI 10.1093/ajcp/88.2.153
   Bordi C, 1995, AM J SURG PATHOL, V19, P8
   BRANDI ML, 1991, J ENDOCRINOL INVEST, V14, P61, DOI 10.1007/BF03350266
   BRANDI ML, 1987, ENDOCR REV, V8, P391, DOI 10.1210/edrv-8-4-391
   Darling TN, 1997, ARCH DERMATOL, V133, P853, DOI 10.1001/archderm.133.7.853
   Debelenko LV, 1997, GASTROENTEROLOGY, V113, P773, DOI 10.1016/S0016-5085(97)70171-9
   Debelenko LV, 1997, CANCER RES, V57, P2238
   DELELLIS RA, 1986, INT REV EXP PATHOL, V28, P163
   DELELLIS RA, 1995, LAB INVEST, V72, P494
   HOFSTRA RMW, 1994, NATURE, V367, P375, DOI 10.1038/367375a0
   Jackson C E, 1989, Horm Metab Res Suppl, V21, P9
   Komminoth P, 1997, Verh Dtsch Ges Pathol, V81, P125
   LARSSON C, 1988, NATURE, V332, P85, DOI 10.1038/332085a0
   LIPS CJM, 1984, CRC CR REV ONCOL-HEM, V2, P117, DOI 10.1016/S1040-8428(84)80017-7
   Lubensky IA, 1996, CANCER RES, V56, P5272
   MAJEWSKI JT, 1979, SURGERY, V86, P475
   MELVIN WS, 1993, SURGERY, V114, P1183
   MENDELSOHN G, 1991, FUNCTIONAL ENDOCRINE, P814
   Morelli A, 1996, BIOCHEM BIOPH RES CO, V227, P736, DOI 10.1006/bbrc.1996.1578
   Olufemi SE, 1998, HUM MUTAT, V11, P264, DOI 10.1002/(SICI)1098-1004(1998)11:4<264::AID-HUMU2>3.0.CO;2-V
   PILATO FP, 1988, CANCER, V61, P1815, DOI 10.1002/1097-0142(19880501)61:9<1815::AID-CNCR2820610916>3.0.CO;2-L
   PIPELEERSMARICHAL M, 1990, NEW ENGL J MED, V322, P723, DOI 10.1056/NEJM199003153221103
   SAAD MF, 1984, MEDICINE, V63, P319, DOI 10.1097/00005792-198411000-00001
   SAMAAN NA, 1988, J CLIN ENDOCR METAB, V67, P801, DOI 10.1210/jcem-67-4-801
   SKOGSEID B, 1994, ENDOCRIN METAB CLIN, V23, P1
   TISELL LE, 1988, BRIT J SURG, V75, P154, DOI 10.1002/bjs.1800750222
   Trump D, 1996, QJM-INT J MED, V89, P653
   VASEN HFA, 1992, ARCH INTERN MED, V152, P1250, DOI 10.1001/archinte.152.6.1250
NR 30
TC 3
Z9 3
U1 0
U2 0
PU PACINI EDITORE
PI PISA
PA VIA DELLA GHERARDESCA-ZONA INDUSTRIALE OSPEDALETTO, 56121 PISA, ITALY
SN 1590-8658
J9 DIGEST LIVER DIS
JI Dig. Liver Dis.
PD FEB
PY 2004
VL 36
SU 1
BP S31
EP S34
DI 10.1016/j.dld.2003.11.011
PG 4
WC Gastroenterology & Hepatology
WE Conference Proceedings Citation Index - Science (CPCI-S); Science Citation Index Expanded (SCI-EXPANDED)
SC Gastroenterology & Hepatology
GA 806IQ
UT WOS:000220428000006
PM 15077909
DA 2023-08-27
ER

PT J
AU Wang, YL
   Ma, GM
   Han, JZ
   Wang, HM
   Kang, ZL
AF Wang, Yali
   Ma, Guomei
   Han, Jingzhe
   Wang, Hongmei
   Kang, Zhilei
TI Third ventricle tumor with Bruns sign as the first manifestation: a case
   report
SO INTERNATIONAL JOURNAL OF CLINICAL AND EXPERIMENTAL PATHOLOGY
LA English
DT Article
DE Bruns syndrome; third ventricle tumor; MRI
AB This article reported a case of a third ventricle tumor with Bruns syndrome sudden disturbance of consciousness as the first manifestation, to improve the clinician's understanding and awareness of the fatal signs. A 38-year-old healthy man was admitted to our hospital for a sudden onset coma for 2 hours. Head magnetic resonance imaging (MRI) showed midbrain aqueduct occlusion, intraductal abnormal nodule signal, considering space-occupying lesions. On the fourth day of admission, the patient was scheduled to undergo ventriculoscopic resection of the mass, but the patient had a small ventricular foramen, which was difficult to explore the posterior part of the third ventricle, and the possibility of injuring the vein was high. Finally, the third ventriculostomy was performed by ventriculoscope. 6 months later, the tumor grew slowly and the patient had no hydrocephalus.
C1 [Wang, Yali; Ma, Guomei; Han, Jingzhe; Wang, Hongmei] Harrison Int Peace Hosp, Dept Neurol, 180 Renmin Dong Rd,Taocheng Dist, Hengshui 053000, Hebei, Peoples R China.
   [Kang, Zhilei] Harrison Int Peace Hosp, MRI, Hengshui 053000, Hebei, Peoples R China.
RP Han, JZ (通讯作者)，Harrison Int Peace Hosp, Dept Neurol, 180 Renmin Dong Rd,Taocheng Dist, Hengshui 053000, Hebei, Peoples R China.
EM
CR Bruns L., 1902, NEUROL CENTRALBL, V21, P561
   Cuetter AC, 1997, CLIN INFECT DIS, V24, P157, DOI 10.1093/clinids/24.2.157
   Shahani L, 2015, AM J TROP MED HYG, V92, P1261, DOI 10.4269/ajtmh.14-0842
   Torres-Corzo JG, 2006, J NEUROSURG, V104, P746, DOI 10.3171/jns.2006.104.5.746
NR 4
TC 1
Z9 1
U1 1
U2 3
PU E-CENTURY PUBLISHING CORP
PI MADISON
PA 40 WHITE OAKS LN, MADISON, WI 53711 USA
SN 1936-2625
J9 INT J CLIN EXP PATHO
JI Int. J. Clin. Exp. Pathol.
PY 2019
VL 12
IS 12
BP 4358
EP 4360
PG 3
WC Oncology; Pathology
WE Science Citation Index Expanded (SCI-EXPANDED)
SC Oncology; Pathology
GA JZ4CF
UT WOS:000505048200016
PM 31933839
DA 2023-08-27
ER

PT J
AU Gao, X
   Yamazaki, Y
   Pecori, A
   Tezuka, Y
   Ono, Y
   Omata, K
   Morimoto, R
   Nakamura, Y
   Satoh, F
   Sasano, H
AF Gao, Xin
   Yamazaki, Yuto
   Pecori, Alessio
   Tezuka, Yuta
   Ono, Yoshikiyo
   Omata, Kei
   Morimoto, Ryo
   Nakamura, Yasuhiro
   Satoh, Fumitoshi
   Sasano, Hironobu
TI Histopathological Analysis of Tumor Microenvironment and Angiogenesis in
   Pheochromocytoma
SO FRONTIERS IN ENDOCRINOLOGY
LA English
DT Article
DE pheochromocytoma; pathology; immunohistochemistry; tumor
   microenvironment; angiogenesis
ID GLAND SCALED SCORE; MACROPHAGES; PARAGANGLIOMAS; EXPRESSION; MATURATION;
   CARCINOMA; CELLS; PASS; VEGF; MICE
AB Pheochromocytomas (PHEOs) are relatively rare catecholamine-producing tumors derived from adrenal medulla. Tumor microenvironment (TME) including neoangiogenesis has been explored in many human neoplasms but not necessarily in PHEOs. Therefore, in this study, we examined tumor infiltrating lymphocytes (CD4 and CD8), tumor associated macrophages (CD68 and CD163), sustentacular cells (S100p), and angiogenic markers (CD31 and areas of intratumoral hemorrhage) in 39 cases of PHEOs in the quantitative fashion. We then compared the results with pheochromocytoma of the adrenal gland scaled score (PASS), grading system for pheochromocytoma and paraganglioma (GAPP) and the status of intra-tumoral catecholamine-synthesizing enzymes (TH, DDC, and PNMT) as well as their clinicopathological factors. Intratumoral CD8 (p = 0.0256), CD31 (p = 0.0400), and PNMT (p = 0.0498) status was significantly higher in PHEOs with PASS <4 than PASS >= 4. In addition, intratumoral CD8(+) lymphocytes were also significantly more abundant in well-than moderately differentiated PHEO according to GAPP score (p = 0.0108) and inversely correlated with tumor size (p = 0.0257). Intratumoral CD68(+) cells were significantly higher in PHEOs with regular or normal histological patterns than those not (p = 0.0370) and inversely correlated with tumor size (p = 0.0457). The status of CD163 was significantly positively correlated with that of CD8 positive cells (p = 0.0032). The proportion of intratumoral hemorrhage areas was significantly higher in PHEOs with PASS >= 4 (p = 0.0172). DDC immunoreactivity in tumor cells was significantly positively correlated with PASS score (p = 0.0356) and TH status was significantly higher in PHEOs harboring normal histological patterns (p = 0.0236) and cellular monotony (p = 0.0219) than those not. Results of our present study did demonstrate that abundant CD8(+) and CD68(+) cells could represent a histologically low-scored tumor. In particular, PHEOs with increased intratumoral hemorrhage should be considered rather malignant. In addition, abnormal catecholamine-producing status of tumor cells such as deficient PNMT and TH and increased DDC could also represent more aggressive PHEOs.
C1 [Gao, Xin; Yamazaki, Yuto; Pecori, Alessio; Sasano, Hironobu] Tohoku Univ, Grad Sch Med, Dept Pathol, Sendai, Miyagi, Japan.
   [Tezuka, Yuta; Omata, Kei; Satoh, Fumitoshi] Tohoku Univ, Grad Sch Med, Div Clin Hypertens Endocrinol & Metab, Sendai, Miyagi, Japan.
   [Tezuka, Yuta; Ono, Yoshikiyo; Omata, Kei; Morimoto, Ryo; Satoh, Fumitoshi] Tohoku Univ Hosp, Div Nephrol Endocrinol & Vasc Med, Sendai, Miyagi, Japan.
   [Nakamura, Yasuhiro] Tohoku Med & Pharmaceut Univ, Fac Med, Div Pathol, Sendai, Miyagi, Japan.
C3 Tohoku University; Tohoku University; Tohoku University; Tohoku Medical
   & Pharmaceutical University
RP Sasano, H (通讯作者)，Tohoku Univ, Grad Sch Med, Dept Pathol, Sendai, Miyagi, Japan.
EM
RI Tezuka, Yuta/AAU-7861-2021; Gao, Xin/AAF-8792-2021
OI Tezuka, Yuta/0000-0003-1546-6949; 
FU Global Hagi scholarship by Tohoku University; Health Labour Sciences
   Research Grant [H29-Nanji-Ippan-046]
FX XG was supported by Global Hagi scholarship by Tohoku University. This
   work was supported by a Health Labour Sciences Research Grant (No.
   H29-Nanji-Ippan-046).
CR Amorim-Pires D, 2016, CYTOGENET GENOME RES, V150, P227, DOI 10.1159/000457479
   BRAVO EL, 1991, KIDNEY INT, V40, P544, DOI 10.1038/ki.1991.244
   Chan MSM, 2014, INT J BIOL MARKER, V29, pE193, DOI 10.5301/jbm.5000082
   Chan MSM, 2012, EXPERT OPIN THER TAR, V16, pS69, DOI 10.1517/14728222.2011.628938
   Chew V, 2012, J ONCOL, V2012, DOI 10.1155/2012/608406
   Edgar VA, 2003, BIOCHEM PHARMACOL, V65, P15, DOI 10.1016/S0006-2952(02)01457-0
   Farhat NA, 2019, ENDOCR PATHOL, V30, P90, DOI 10.1007/s12022-019-9575-6
   Farhood B, 2019, J CELL PHYSIOL, V234, P8509, DOI 10.1002/jcp.27782
   Farrugia F A, 2017, Endocr Regul, V51, P168, DOI 10.1515/enr-2017-0018
   Farrugia Frederick-Anthony, 2019, Endocr Regul, V53, P191, DOI 10.2478/enr-2019-0020
   Farrugia Frederick-Anthony, 2016, Khirurgiia (Sofiia), V82, P120
   Favier J, 2010, BEST PRACT RES CL EN, V24, P957, DOI 10.1016/j.beem.2010.10.004
   Feng F, 2011, J UROLOGY, V185, P1583, DOI 10.1016/j.juro.2010.12.050
   Fishbein L, 2016, HEMATOL ONCOL CLIN N, V30, P135, DOI 10.1016/j.hoc.2015.09.006
   Gao X, 2019, J STEROID BIOCHEM, V193, DOI 10.1016/j.jsbmb.2019.105434
   Genard G, 2018, CELL DEATH DIS, V9, DOI 10.1038/s41419-018-0757-9
   Hanahan D, 2012, CANCER CELL, V21, P309, DOI 10.1016/j.ccr.2012.02.022
   Inbar S, 2011, PLOS ONE, V6, DOI 10.1371/journal.pone.0019246
   Joyce JA, 2015, SCIENCE, V348, P74, DOI 10.1126/science.aaa6204
   KIMURA N, 1992, VIRCHOWS ARCH A, V421, P25, DOI 10.1007/BF01607135
   Kimura N, 2014, ENDOCR-RELAT CANCER, V21, P405, DOI 10.1530/ERC-13-0494
   Kitawaki Y, 2018, HUM PATHOL, V77, P88, DOI 10.1016/j.humpath.2018.03.016
   Konno-Kumagai T, 2019, DIS ESOPHAGUS, V32, DOI 10.1093/dote/doy063
   Konosu-Fukaya S, 2018, ENDOCR PATHOL, V29, P302, DOI 10.1007/s12022-018-9544-5
   Konosu-Fukaya S, 2015, MOL CELL ENDOCRINOL, V408, P205, DOI 10.1016/j.mce.2014.10.008
   Kulkarni MM, 2016, J CLIN DIAGN RES, V10, pEC1, DOI 10.7860/JCDR/2016/20565.8419
   Lam AKY, 2017, ENDOCR PATHOL, V28, P213, DOI 10.1007/s12022-017-9484-5
   Landwehr LS, 2020, J IMMUNOTHER CANCER, V8, DOI 10.1136/jitc-2019-000469
   Lin ZY, 2019, J CANCER, V10, P211, DOI 10.7150/jca.26444
   Liu Q, 1996, SURGERY, V120, P938, DOI 10.1016/S0039-6060(96)80037-7
   Lloyd RV, 2017, WHO CLASSIFICATION T
   MADDEN KS, 1995, ANNU REV PHARMACOL, V35, P417, DOI 10.1146/annurev.pa.35.040195.002221
   Mantovani A, 2017, NAT REV CLIN ONCOL, V14, P399, DOI 10.1038/nrclinonc.2016.217
   MCCARTY KS, 1985, ARCH PATHOL LAB MED, V109, P716
   Nasi G, 2019, J NEUROIMMUNOL, V332, P233, DOI 10.1016/j.jneuroim.2019.02.007
   Ni YH, 2015, TUMOR BIOL, V36, P5291, DOI 10.1007/s13277-015-3189-5
   Nouri-Shirazi M, 2000, J IMMUNOL, V165, P3797, DOI 10.4049/jimmunol.165.7.3797
   Oka N, 2020, NEUROENDOCRINOLOGY, V110, P393, DOI 10.1159/000502776
   Parayath NN, 2018, NANO LETT, V18, P3571, DOI 10.1021/acs.nanolett.8b00689
   Parise IZS, 2019, CANCERS, V11, DOI 10.3390/cancers11111730
   Pollard PJ, 2006, J CLIN ENDOCR METAB, V91, P4593, DOI 10.1210/jc.2006-0920
   Quail DF, 2013, NAT MED, V19, P1423, DOI 10.1038/nm.3394
   RAHAT MA, 2016, MEDIAT INFLAMM, V2016, DOI DOI 10.1155/2016/5894347
   Rasmuson T, 2001, ACTA ONCOL, V40, P83
   Ruffell B, 2015, CANCER CELL, V27, P462, DOI 10.1016/j.ccell.2015.02.015
   Salmenkivi K, 2003, APMIS, V111, P458, DOI 10.1034/j.1600-0463.2003.1110402.x
   Sarkar C, 2013, J NEUROIMMUNE PHARM, V8, P7, DOI 10.1007/s11481-012-9395-7
   Sasano H, 2005, BIOMED PHARMACOTHER, V59, pS334, DOI 10.1016/S0753-3322(05)80068-X
   TAKAHASHI H, 1987, ACTA NEUROPATHOL, V73, P227, DOI 10.1007/BF00686615
   Taves MD, 2017, HORM BEHAV, V88, P4, DOI 10.1016/j.yhbeh.2016.10.022
   Thompson LDR, 2002, AM J SURG PATHOL, V26, P551, DOI 10.1097/00000478-200205000-00002
   Trenti A, 2018, INT J MOL SCI, V19, DOI 10.3390/ijms19030859
   Trigunaite A, 2015, CELL IMMUNOL, V294, P87, DOI 10.1016/j.cellimm.2015.02.004
   Troiano G, 2019, ORAL ONCOL, V93, P66, DOI 10.1016/j.oraloncology.2019.04.019
   Wang N, 2014, FRONT IMMUNOL, V5, DOI 10.3389/fimmu.2014.00614
   Wu D, 2009, AM J SURG PATHOL, V33, P599, DOI 10.1097/PAS.0b013e318190d12e
   Yamate Jyoji, 2016, Food Saf (Tokyo), V4, P61, DOI 10.14252/foodsafetyfscj.2016012
   Yamazaki Y, 2018, HYPERTENSION, V72, P632, DOI 10.1161/HYPERTENSIONAHA.118.10907
   Yamazaki Y, 2017, J CLIN ENDOCR METAB, V102, P1182, DOI 10.1210/jc.2016-2986
   Yazdani S, 2014, HUM PATHOL, V45, P866, DOI 10.1016/j.humpath.2013.09.024
   Yazdani S, 2013, HUM PATHOL, V44, P1586, DOI 10.1016/j.humpath.2013.01.004
   Zielke A, 2002, SURGERY, V132, P1056, DOI 10.1067/msy.2002.128613
NR 62
TC 11
Z9 11
U1 0
U2 3
PU FRONTIERS MEDIA SA
PI LAUSANNE
PA AVENUE DU TRIBUNAL FEDERAL 34, LAUSANNE, CH-1015, SWITZERLAND
SN 1664-2392
J9 FRONT ENDOCRINOL
JI Front. Endocrinol.
PD NOV 10
PY 2020
VL 11
AR 587779
DI 10.3389/fendo.2020.587779
PG 10
WC Endocrinology & Metabolism
WE Science Citation Index Expanded (SCI-EXPANDED)
SC Endocrinology & Metabolism
GA OV0EY
UT WOS:000591895600001
PM 33244312
OA Green Published, gold
DA 2023-08-27
ER

PT J
AU Fischbein, NJ
   Prados, MD
   Wara, W
   Russo, C
   Edwards, MSB
   Barkovich, AJ
AF Fischbein, NJ
   Prados, MD
   Wara, W
   Russo, C
   Edwards, MSB
   Barkovich, AJ
TI Radiologic classification of brain stem tumors: Correlation of magnetic
   resonance imaging appearance with clinical outcome
SO PEDIATRIC NEUROSURGERY
LA English
DT Article
DE brain stem tumors; magnetic resonance imaging; neoplasm; glioma;
   gadolinium
ID SURGICAL INDICATIONS; PROGNOSTIC FACTORS; GLIOMAS; CHILDREN; CHILDHOOD;
   MANAGEMENT; DIAGNOSIS; SYSTEM
AB Although tumors of the brain stem have traditionally been classified as a single entity, these tumors are increasingly being recognized as a heterogeneous group, with some subgroups having better prognoses for long-term survival. Although several systems for classification of brain stem tumors have been proposed, none have been based on data derived from contrast-enhanced magnetic resonance (MR) imaging. In this review, we present a classification scheme based on our review of the literature and of the MR scans of 64 patients with brain stem tumors. In addition, we assess the contribution of gadolinium to the classification of brain stem tumors and correlate the various tumor subtypes, based on MR appearance, with prognosis.
   Our results suggest that the most important factor in determining prognosis based on MR characteristics is whether the tumor is diffuse or focal. Focal tumors have an excellent prognosis regardless of the site of tumor origin. Diffuse tumors of the mesencephalon and pens have a significantly poorer prognosis than focal tumors (p = 0.0013), with diffuse pontine tumors having the worst prognosis. Differentiation of diffuse and focal medullary tumors was difficult, possibly explaining the lack of significant difference in the survival of patients with diffuse versus focal medullary tumors. The presence or absence of enhancement after the administration of paramagnetic contrast has no significant relation with outcome, overall or within specific tumor subgroups.
RP Fischbein, NJ (通讯作者)，UNIV CALIF SAN FRANCISCO,DEPT RADIOL,505 PARNASSUS AVE,BOX 0628,SAN FRANCISCO,CA 94143, USA.
RI Russo, Carolyn/N-3997-2018; Prados, Michael/N-5220-2019
OI Russo, Carolyn/0000-0002-8034-7260; Fischbein, Nancy/0000-0003-2895-3586
CR ALBRIGHT AL, 1986, J NEUROSURG, V65, P751, DOI 10.3171/jns.1986.65.6.0751
   ALBRIGHT AL, 1993, NEUROSURGERY, V33, P1026
   ATLAS SW, 1988, RADIOLOGY, V168, P803, DOI 10.1148/radiology.168.3.3406410
   BARKOVICH AJ, 1991, PEDIATR NEUROSURG, V16, P73, DOI 10.1159/000120511
   BRACAC GB, 1985, NEURORADIOLOGY, V27, P202
   BYDDER GM, 1988, NEURORADIOLOGY, V30, P367, DOI 10.1007/BF00404099
   COHEN ME, 1986, NEUROLOGY, V36, P602, DOI 10.1212/WNL.36.5.602
   EDWARDS MSB, 1994, J NEUROSURG, V80, P20, DOI 10.3171/jns.1994.80.1.0020
   EPSTEIN F, 1985, CANCER, V56, P1804, DOI 10.1002/1097-0142(19851001)56:7+<1804::AID-CNCR2820561316>3.0.CO;2-5
   EPSTEIN F, 1988, J NEURO-ONCOL, V6, P309
   EPSTEIN F, 1986, J NEUROSURG, V64, P11, DOI 10.3171/jns.1986.64.1.0011
   EPSTEIN F, 1987, J NEUROSURG, V67, P483, DOI 10.3171/jns.1987.67.4.0483
   EPSTEIN FJ, 1993, J NEUROSURG, V78, P408, DOI 10.3171/jns.1993.78.3.0408
   FARWELL JR, 1977, CANCER, V40, P3123, DOI 10.1002/1097-0142(197712)40:6<3123::AID-CNCR2820400656>3.0.CO;2-6
   FREEMAN CR, 1993, INT J RADIAT ONCOL, V27, P197, DOI 10.1016/0360-3016(93)90228-N
   GUINEY MJ, 1993, INT J RADIAT ONCOL, V25, P235, DOI 10.1016/0360-3016(93)90344-U
   HOFFMAN HJ, 1980, NEUROSURGERY, V7, P243, DOI 10.1227/00006123-198009000-00007
   KHATIB ZA, 1994, PEDIATR NEUROSURG, V20, P2, DOI 10.1159/000120759
   LEE BCP, 1985, AM J NEURORADIOL, V6, P159
   MAY PL, 1991, J NEUROSURG, V74, P867, DOI 10.3171/jns.1991.74.6.0867
   MILSTEIN JM, 1989, J NEURO-ONCOL, V7, P367, DOI 10.1007/BF02147094
   MOLLOY PT, 1995, NEUROLOGY, V45, P1897, DOI 10.1212/WNL.45.10.1897
   PACKER R J, 1991, Pediatric Neurosurgery, V17, P37, DOI 10.1159/000120565
   PACKER RJ, 1985, NEUROLOGY, V35, P397, DOI 10.1212/WNL.35.3.397
   PACKER RJ, 1994, CANCER-AM CANCER SOC, V74, P1827, DOI 10.1002/1097-0142(19940915)74:6<1827::AID-CNCR2820740628>3.0.CO;2-Q
   PIERREKAHN A, 1993, J NEUROSURG, V79, P845, DOI 10.3171/jns.1993.79.6.0845
   POLLACK IF, 1994, J NEUROSURG, V80, P681, DOI 10.3171/jns.1994.80.4.0681
   ROBERTSON PL, 1995, PEDIATR NEUROSURG, V22, P65, DOI 10.1159/000120879
   SANFORD RA, 1988, SURG NEUROL, V30, P276, DOI 10.1016/0090-3019(88)90299-6
   SHRIEVE DC, 1992, INT J RADIAT ONCOL, V24, P599, DOI 10.1016/0360-3016(92)90704-L
   STROINK AR, 1987, NEUROSURGERY, V20, P439, DOI 10.1227/00006123-198703000-00014
   STROINK AR, 1986, J NEUROSURG, V65, P745, DOI 10.3171/jns.1986.65.6.0745
   VANDERTOP WP, 1992, NEUROSURGERY, V31, P186, DOI 10.1227/00006123-199208000-00003
   WHITE HH, 1963, NEUROLOGY, V13, P292, DOI 10.1212/WNL.13.4.292
NR 34
TC 123
Z9 125
U1 0
U2 2
PU KARGER
PI BASEL
PA ALLSCHWILERSTRASSE 10, CH-4009 BASEL, SWITZERLAND
SN 1016-2291
J9 PEDIATR NEUROSURG
JI Pediatr. Neurosurg.
PD JAN
PY 1996
VL 24
IS 1
BP 9
EP 23
DI 10.1159/000121010
PG 15
WC Clinical Neurology; Pediatrics; Surgery
WE Science Citation Index Expanded (SCI-EXPANDED)
SC Neurosciences & Neurology; Pediatrics; Surgery
GA UT878
UT WOS:A1996UT87800004
PM 8817611
DA 2023-08-27
ER

PT J
AU Ruge, MI
   Kickingereder, P
   Simon, T
   Treuer, H
   Sturm, V
AF Ruge, Maximilian I.
   Kickingereder, Philipp
   Simon, Thorsten
   Treuer, Harald
   Sturm, Volker
TI Stereotactic iodine-125 brachytherapy for treatment of inoperable focal
   brainstem gliomas of WHO grades I and II: feasibility and long-term
   outcome
SO JOURNAL OF NEURO-ONCOLOGY
LA English
DT Article
DE Brainstem gliomas; Low-grade gliomas; Stereotactic brachytherapy;
   Interstitial radiosurgery; Iodine-125
ID CONFORMAL RADIATION-THERAPY; GAMMA-KNIFE RADIOSURGERY; CHILDHOOD-CANCER
   SURVIVOR; CENTRAL-NERVOUS-SYSTEM; PEDIATRIC-PATIENTS; INTERSTITIAL
   IRRADIATION; PHASE-II; PROGNOSTIC-FACTORS; CEREBRAL GLIOMAS; MALIGNANT
   GLIOMA
AB Microsurgical resection is the most frequently suggested treatment option for accessible focal brainstem gliomas (F-BSG) of World Health Organization (WHO) grades I and II. Because of their location in the highly eloquent brain, however, resection is associated with permanent postoperative morbidity, ranging from 12 to 33 %. Only a few reports have suggested stereotactic brachytherapy (SBT) with implantation of iodine-125 seeds as a local treatment alternative. Between 1993 and 2010, 47 patients were treated with SBT (iodine-125 seeds; cumulative surface dose 50-65 Gy) for inoperable F-BSG, WHO grades I and II, in one of the largest reported patient series. We evaluated procedure-related complications, clinical outcome, and progression-free and overall survival (PFS, OS). Median follow-up was 81.6 months. Procedure-related mortality was zero. Within 30 days of seed implantation six patients (12.8 %) had transient neurological deficits. Two patients (4.3 %) deteriorated permanently. Space-occupying cysts occurred in six patients (12.8 %) after a median of 28.5 months, and required surgical intervention. Nine patients (19.1 %) presented with tumor relapse after a median of 56.6 months (range 7.9-118.0 months). For the remaining 38 patients complete response was observed for 23.4 %, partial response for 29.8 %, and stable disease for 27.7 %. Actuarial PFS was 97.7 +/- A 2.2, 92.8 +/- A 4.0, 81.2 +/- A 6.5, and 62.0 +/- A 10.4 % after 1, 2, 5, and 10 years, respectively. Corresponding OS was 100 +/- A 0.0 % (1 and 2 years), 97.4 +/- A 2.6 % (5 years), and 87.6 +/- A 7.0 % (10 years). SBT is a comparatively safe, minimally invasive, and highly effective local treatment option for patients with inoperable F-BSG WHO grades I and II; it merits further evaluation in prospective randomized trials.
C1 [Ruge, Maximilian I.; Kickingereder, Philipp; Treuer, Harald; Sturm, Volker] Univ Hosp Cologne, Dept Stereotact & Funct Neurosurg, D-50937 Cologne, Germany.
   [Simon, Thorsten] Univ Hosp Cologne, Dept Pediat Hematol & Oncol, Cologne, Germany.
C3 University of Cologne; University of Cologne
RP Ruge, MI (通讯作者)，Univ Hosp Cologne, Dept Stereotact & Funct Neurosurg, Kerpener Str 62,Main Clin Bldg,11th Floor, D-50937 Cologne, Germany.
EM
RI Simon, Thorsten/AAY-3958-2021; Vollmuth (né Kickingereder),
   Philipp/AAK-3267-2021
OI Simon, Thorsten/0000-0002-3425-8451; Vollmuth (né Kickingereder),
   Philipp/0000-0002-6224-0064
CR Bernstein M, 1995, STEREOT FUNCT NEUROS, V65, P167, DOI 10.1159/000098689
   BERSTEIN M, 1981, NEUROSURGERY, V9, P741, DOI 10.1227/00006123-198112000-00022
   Bowers DC, 2006, J CLIN ONCOL, V24, P5277, DOI 10.1200/JCO.2006.07.2884
   CHAMBERLAIN MC, 1995, ARCH NEUROL-CHICAGO, V52, P162, DOI 10.1001/archneur.1995.00540260066018
   Chuba PJ, 1998, CHILD NERV SYST, V14, P570, DOI 10.1007/s003810050274
   Di Pinto M, 2010, INT J RADIAT ONCOL, V77, P1002, DOI 10.1016/j.ijrobp.2009.06.003
   DIRKS PB, 1994, NEUROSURGERY, V34, P68
   DUFFNER PK, 1985, CANCER-AM CANCER SOC, V56, P1841, DOI 10.1002/1097-0142(19851001)56:7+<1841::AID-CNCR2820561325>3.0.CO;2-C
   Farmer JP, 2001, PEDIATR NEUROSURG, V34, P206, DOI 10.1159/000056021
   Fisher BJ, 1998, J NEUROSURG, V88, P969, DOI 10.3171/jns.1998.88.6.0969
   Fisher PG, 1996, CONT NEUROSURG, V18, P1
   Freeman CR, 1998, INT J RADIAT ONCOL, V40, P265, DOI 10.1016/S0360-3016(97)00572-5
   Fuchs I, 2002, ACT NEUR S, V84, P85
   Gnekow AK, 2004, KLIN PADIATR, V216, P331, DOI 10.1055/s-2004-832355
   Gururangan S, 2002, J CLIN ONCOL, V20, P2951, DOI 10.1200/JCO.2002.12.008
   GUTIN PH, 1984, J NEUROSURG, V60, P61, DOI 10.3171/jns.1984.60.1.0061
   HIRATO M, 1995, STEREOT FUNCT NEUROS, V64, P32, DOI 10.1159/000098762
   Hong Semie, 2005, Cancer Res Treat, V37, P109, DOI 10.4143/crt.2005.37.2.109
   Hubbell JH, 1995, 5632 NISTIR, P20899
   Jallo GI, 2004, CHILD NERV SYST, V20, P143, DOI 10.1007/s00381-003-0870-6
   Julow Jeno, 2004, Strahlenther Onkol, V180, P449
   Karnofsky DA., 1949, CLIN EVALUATION CHEM
   Kestle J, 2004, J NEUROSURG, V101, P1, DOI 10.3171/ped.2004.101.2.0001
   Khaw SL, 2007, PEDIATR BLOOD CANCER, V49, P808, DOI 10.1002/pbc.21270
   KIHLSTROM L, 1994, ACT NEUR S, V62, P55
   Korinthenberg R, 2011, INT J RADIAT ONCOL, V79, P1131, DOI 10.1016/j.ijrobp.2009.12.040
   Kreth FW, 2010, ADV TECH STAND NEURO, V35, P180
   Kreth FW, 2010, ONCOLOGY OF CNS TUMORS, SECOND EDITION, P135, DOI 10.1007/978-3-642-02874-8_6
   Kreth FW, 2006, CANCER, V106, P1372, DOI 10.1002/cncr.21750
   KRETH FW, 1995, J NEUROSURG, V82, P418, DOI 10.3171/jns.1995.82.3.0418
   KRETH FW, 1993, NERVENARZT, V64, P633
   Lafay-Cousin L, 2008, CANCER, V112, P892, DOI 10.1002/cncr.23249
   Laperriere N, 2002, RADIOTHER ONCOL, V64, P259, DOI 10.1016/S0167-8140(02)00078-6
   Laperriere NJ, 1998, INT J RADIAT ONCOL, V41, P1005, DOI 10.1016/S0360-3016(98)00159-X
   Lesniak MS, 2003, PEDIATR NEUROSURG, V39, P314, DOI 10.1159/000075260
   Liao CH, 2012, CHILD NERV SYST, V28, P175, DOI 10.1007/s00381-011-1620-9
   MACDONALD DR, 1990, J CLIN ONCOL, V8, P1277, DOI 10.1200/JCO.1990.8.7.1277
   Malkin M G, 1994, Recent Results Cancer Res, V135, P117
   Marcus KJ, 2005, INT J RADIAT ONCOL, V61, P374, DOI 10.1016/j.ijrobp.2004.06.012
   Massimino M, 2002, J CLIN ONCOL, V20, P4209, DOI 10.1200/JCO.2002.08.087
   Merchant TE, 2002, INT J RADIAT ONCOL, V52, P325, DOI 10.1016/S0360-3016(01)01807-7
   Merchant TE, 2009, J CLIN ONCOL, V27, P3691, DOI 10.1200/JCO.2008.21.2738
   MUNDINGER F, 1991, J NEUROSURG, V75, P740, DOI 10.3171/jns.1991.75.5.0740
   Neglia JP, 2006, J NATL CANCER I, V98, P1528, DOI 10.1093/jnci/djj411
   Ostertag C B, 1989, J Neurosurg Sci, V33, P83
   OSTERTAG CB, 1992, ACTA NEUROCHIR, V119, P53, DOI 10.1007/BF01541782
   PACKER RJ, 1993, J CLIN ONCOL, V11, P850, DOI 10.1200/JCO.1993.11.5.850
   Packer RJ, 1997, J NEUROSURG, V86, P747, DOI 10.3171/jns.1997.86.5.0747
   Peraud A, 2007, CHILD NERV SYST, V23, P39, DOI 10.1007/s00381-006-0203-7
   PIERREKAHN A, 1993, J NEUROSURG, V79, P845, DOI 10.3171/jns.1993.79.6.0845
   POLLACK IF, 1993, J NEUROSURG, V78, P859, DOI 10.3171/jns.1993.78.6.0859
   PONS MD, 1992, J NEURO-ONCOL, V14, P151
   RANKIN J, 1957, Scott Med J, V2, P200
   Recinos PF, 2007, PEDIATR NEUROSURG, V43, P192, DOI 10.1159/000098831
   Ronghe M, 2010, PEDIATR BLOOD CANCER, V55, P471, DOI 10.1002/pbc.22557
   Ruge MI, 2011, J CLIN ONCOL, V29, P4151, DOI 10.1200/JCO.2011.37.3381
   Ruge MI, 2011, J NEURO-ONCOL, V105, P109, DOI 10.1007/s11060-011-0571-z
   Ruge MI, 2011, J NEURO-ONCOL, V103, P129, DOI 10.1007/s11060-010-0364-9
   Ruge MI, 2011, NEUROSURGERY, V68, P1209, DOI 10.1227/NEU.0b013e31820b526a
   Sandri A, 2006, CHILD NERV SYST, V22, P1127, DOI 10.1007/s00381-006-0083-x
   Saran FH, 2002, INT J RADIAT ONCOL, V53, P43, DOI 10.1016/S0360-3016(02)02734-7
   Schnell O, 2008, J NEUROL, V255, P1495, DOI 10.1007/s00415-008-0948-x
   Selker RG, 2002, NEUROSURGERY, V51, P343, DOI 10.1097/00006123-200208000-00009
   Sneed PK, 1997, SEMIN SURG ONCOL, V13, P157, DOI 10.1002/(SICI)1098-2388(199705/06)13:3<157::AID-SSU2>3.0.CO;2-6
   STURM V, 1983, ACTA NEUROCHIR, V68, P11, DOI 10.1007/BF01406197
   Suchorska B, 2011, NEURO-ONCOLOGY, V13, P1133, DOI 10.1093/neuonc/nor100
   Teo C, 2008, CHILD NERV SYST, V24, P1307, DOI 10.1007/s00381-008-0647-z
   Treuer H, 2005, RADIOTHER ONCOL, V77, P202, DOI 10.1016/j.radonc.2005.08.006
   VANSWIETEN JC, 1988, STROKE, V19, P604, DOI 10.1161/01.STR.19.5.604
   Viola A, 2006, RADIAT RES, V165, P695, DOI 10.1667/RR3529.1
   Voges J, 1999, CRIT REV NEUROSURG, V9, P223, DOI 10.1007/s003290050137
   VOGES J, 1993, ACTA NEUROCHIR, P108
   Walter AW, 2000, J PEDIAT HEMATOL ONC, V22, P247, DOI 10.1097/00043426-200005000-00010
   WHITE HH, 1963, NEUROLOGY, V13, P292, DOI 10.1212/WNL.13.4.292
   Yen CP, 2007, J NEUROSURG, V106, P8, DOI 10.3171/jns.2007.106.1.8
NR 75
TC 18
Z9 24
U1 0
U2 11
PU SPRINGER
PI NEW YORK
PA 233 SPRING ST, NEW YORK, NY 10013 USA
SN 0167-594X
EI 1573-7373
J9 J NEURO-ONCOL
JI J. Neuro-Oncol.
PD SEP
PY 2012
VL 109
IS 2
BP 273
EP 283
DI 10.1007/s11060-012-0889-1
PG 11
WC Oncology; Clinical Neurology
WE Science Citation Index Expanded (SCI-EXPANDED)
SC Oncology; Neurosciences & Neurology
GA 998OO
UT WOS:000308250200007
PM 22580799
DA 2023-08-27
ER

PT J
AU Muller, K
   Scheithauer, H
   Pietschmann, S
   Hoffmann, M
   Rossler, J
   Graf, N
   Baumert, BG
   Christiansen, H
   Kortmann, RD
   Kramm, CM
   von Bueren, AO
AF Mueller, Klaus
   Scheithauer, Heike
   Pietschmann, Sophie
   Hoffmann, Marion
   Roessler, Jochen
   Graf, Norbert
   Baumert, Brigitta G.
   Christiansen, Hans
   Kortmann, Rolf-Dieter
   Kramm, Christof M.
   von Bueren, Andre O.
TI Reirradiation as part of a salvage treatment approach for progressive
   non-pontine pediatric high-grade gliomas: preliminary experiences from
   the German HIT-HGG study group
SO RADIATION ONCOLOGY
LA English
DT Article
DE Progressive; Relapsed; High-grade gliomas; Pediatric; Reirradiation;
   Children
ID CHEMOTHERAPY; CHILDREN; BRAIN
AB Background and purpose: The aim of the present analysis was to assess the feasibility, toxicity, and the tumor control of reirradiation as a salvage treatment for progressive pediatric non-pontine high-grade gliomas (HGG).
   Patients and methods: The database of the Reference Center for Radiation Oncology of the German HIT (HIT = German acronym for brain tumor) treatment network for childhood brain tumors was screened for children who were reirradiated for progressive non-pontine HGG.
   Results: We identified eight patients (WHO grade III: n = 5; WHO grade IV: n = 3) who underwent reirradiation between April 2006 and July 2012. Median age was 13.5 years at primary diagnosis and 14.8 years at first progression. All patients initially underwent surgery (incomplete resection, n = 7; biopsy, n = 1) followed by radiochemotherapy. Relapses occurred inside (n = 2), at the margin (n = 4), and outside of the preirradiated area (n = 2). In all patients, reirradiation was tolerated well without significant acute toxicity. Temporary clinical improvement and tumor regression on magnetic resonance imaging (MRI) following reirradiation was reported (n = 3). However, all patients finally died by disease progression. Median survival time was 26.2 months from initial diagnosis and 11.4 months after first progression. Median time interval between initial radiotherapy and first reirradiation was 9.0 months. In six patients, all macroscopic tumor deposits were reirradiated. In these patients, median progression-free (overall) survival from the start of reirradiation was 2.4 (4.6) months.
   Conclusion: Our analysis, although based on a limited patient number, suggests that reirradiation of progressive non-pontine HGG is feasible in children. Benefit in terms of quality of life and/or survival needs to be assessed in a prospective and ideally in a randomized manner.
C1 [Mueller, Klaus; Pietschmann, Sophie; Kortmann, Rolf-Dieter] Univ Med Ctr Leipzig, Dept Radiat Oncol, Leipzig, Germany.
   [Scheithauer, Heike] Univ Munich, Dept Radiat Oncol, Munich, Germany.
   [Hoffmann, Marion; Kramm, Christof M.; von Bueren, Andre O.] Univ Med Ctr Goettingen, Dept Pediat & Adolescent Med, Div Pediat Hematol & Oncol, Gottingen, Germany.
   [Roessler, Jochen] Univ Hosp Freiburg, Ctr Pediat & Adolescent Med, Clin Pediat Hematol & Oncol 4, Freiburg, Germany.
   [Graf, Norbert] Univ Saarland, Dept Pediat Hematol & Oncol, Homburg, Germany.
   [Baumert, Brigitta G.] MediClin Robert Janker Clin, Dept Radiat Oncol, Bonn, Germany.
   [Baumert, Brigitta G.] MediClin Robert Janker Clin, Clin Cooperat Unit Neurooncol, Bonn, Germany.
   [Baumert, Brigitta G.] Univ Bonn Med Ctr, Bonn, Germany.
   [Christiansen, Hans] Hannover Med Sch, Dept Radiotherapy, Hannover, Germany.
C3 Leipzig University; University of Munich; University of Gottingen;
   University of Freiburg; Saarland University; University of Bonn;
   Hannover Medical School
RP von Bueren, AO (通讯作者)，Univ Med Ctr Goettingen, Dept Pediat & Adolescent Med, Div Pediat Hematol & Oncol, Gottingen, Germany.
EM
RI Baumert, Brigitta G./AAM-6549-2020; von Bueren, Andre/AAF-2065-2020;
   Graf, Norbert/H-1328-2011
OI Baumert, Brigitta G./0000-0001-8871-4654; von Bueren,
   Andre/0000-0003-4197-6264; Graf, Norbert/0000-0002-2248-323X; Rossler,
   Jochen/0000-0003-4022-4917
FU German Children's Cancer Foundation (Deutsche Kinderkrebsstiftung); Open
   Access Publication Funds of the Goettingen University
FX We thank all the participating centers for their valuable cooperation.
   The HIT-HGG trial office and the reference institutions (Neuropathology,
   Prof. Dr. Torsten Pietsch et al. / Neuro-Radiology: Prof. Dr. Monika
   Warmuth-Metz et al./Radio-Oncology: Prof. Dr. Rolf-Dieter Kortmann et
   al.) are supported by the German Children's Cancer Foundation (Deutsche
   Kinderkrebsstiftung). We thank Michael Josef Kerber for proofreading and
   linguistic revision of the manuscript. We would like to thank Gudrun
   Leyerer for administrative support. We acknowledge the contributions by
   Drs. Torsten Pietsch (central pathologic review), Monika Warmuth-Metz
   (central review of imaging), Uwe Kordes and Annette Sander (provision of
   study patients). We also acknowledge support by the Open Access
   Publication Funds of the Goettingen University.
CR Ciammella P, 2013, RADIAT ONCOL, V8, DOI 10.1186/1748-717X-8-222
   Combs SE, 2013, ACTA ONCOL, V52, P147, DOI 10.3109/0284186X.2012.692882
   Combs SE, 2007, BMC CANCER, V7, DOI 10.1186/1471-2407-7-167
   De Vleeschouwer S, 2008, CLIN CANCER RES, V14, P3098, DOI 10.1158/1078-0432.CCR-07-4875
   Ernst-Stecken A, 2007, J NEURO-ONCOL, V81, P287, DOI 10.1007/s11060-006-9231-0
   Fogh SE, 2010, J CLIN ONCOL, V28, P3048, DOI 10.1200/JCO.2009.25.6941
   Fokas E, 2009, STRAHLENTHER ONKOL, V185, P235, DOI 10.1007/s00066-009-1753-x
   Fontanilla HP, 2012, AM J CLIN ONCOL-CANC, V35, P51, DOI 10.1097/COC.0b013e318201a2b7
   Henke G, 2009, STRAHLENTHER ONKOL, V185, P113, DOI 10.1007/s00066-009-1969-9
   Lawrence YR, 2010, INT J RADIAT ONCOL, V76, pS20, DOI 10.1016/j.ijrobp.2009.02.091
   MacDonald TJ, 2011, NEURO-ONCOLOGY, V13, P1049, DOI 10.1093/neuonc/nor092
   Mayer R, 2008, INT J RADIAT ONCOL, V70, P1350, DOI 10.1016/j.ijrobp.2007.08.015
   Niyazi M, 2011, RADIOTHER ONCOL, V98, P1, DOI 10.1016/j.radonc.2010.11.006
   Scholtyssek F, 2013, RADIAT ONCOL, V8, DOI 10.1186/1748-717X-8-161
   SPOSTO R, 1989, J NEURO-ONCOL, V7, P165, DOI 10.1007/BF00165101
   Weller M, 2013, NEURO-ONCOLOGY, V15, P4, DOI 10.1093/neuonc/nos273
   Zapletalova D, 2012, ONCOLOGY-BASEL, V82, P249, DOI 10.1159/000336483
NR 17
TC 12
Z9 12
U1 1
U2 5
PU BIOMED CENTRAL LTD
PI LONDON
PA 236 GRAYS INN RD, FLOOR 6, LONDON WC1X 8HL, ENGLAND
SN 1748-717X
J9 RADIAT ONCOL
JI Radiat. Oncol.
PD AUG 12
PY 2014
VL 9
AR 177
DI 10.1186/1748-717X-9-177
PG 8
WC Oncology; Radiology, Nuclear Medicine & Medical Imaging
WE Science Citation Index Expanded (SCI-EXPANDED)
SC Oncology; Radiology, Nuclear Medicine & Medical Imaging
GA AO2ZT
UT WOS:000341198300001
PM 25112658
OA Green Accepted, gold, Green Published
DA 2023-08-27
ER

PT J
AU Hirano, M
   Hatzoglou, V
   Karimi, S
   Young, RJ
AF Hirano, Miki
   Hatzoglou, Vaios
   Karimi, Sasan
   Young, Robert J.
TI Hypertrophic olivary degeneration resulting from posterior fossa masses
   and their treatments
SO CLINICAL IMAGING
LA English
DT Article
DE MRI; All oncology; Radiation-therapy tumor; Surgical-therapy tumor
ID OF-THE-LITERATURE; BRAIN-STEM
AB Purpose: Characterize hypertrophic olivary degeneration (HOD) that develops from posterior fossa masses and their treatments.
   Methods: Retrospectively reviewed MR images and clinical data of 10 patients with posterior fossa masses and HOD.
   Results: Eight patients had cerebellar lesions, and two patients had pontine lesions. Lesions consisted of tumors, demyelination, and nonspecific necrosis. MRI showed T2 hyperintense signal in the inferior olive a median 86 days after the diagnosis of a posterior fossa lesion. HOD presented prior to surgery (n=2), after surgery (n=3), after surgery/radiation therapy (n=4), or without treatment (n=1).
   Conclusions: HOD may develop from posterior fossa masses and surgical and/or radiation therapy. (C) 2015 Elsevier Inc. All rights reserved.
C1 [Hirano, Miki; Hatzoglou, Vaios; Karimi, Sasan; Young, Robert J.] Mem Sloan Kettering Canc Ctr, Dept Radiol, New York, NY 10065 USA.
   [Hatzoglou, Vaios; Karimi, Sasan; Young, Robert J.] Mem Sloan Kettering Canc Ctr, Brain Tumor Ctr, New York, NY 10065 USA.
C3 Memorial Sloan Kettering Cancer Center; Memorial Sloan Kettering Cancer
   Center
RP Young, RJ (通讯作者)，Mem Sloan Kettering Canc Ctr, 1275 York Ave, New York, NY 10065 USA.
EM;
  
RI Young, Robert J/AAD-9823-2020
OI Young, Robert J/0000-0001-7003-3017; Hatzoglou,
   Vaios/0000-0003-0252-0694
FU Brain Tumor Center of Memorial Sloan Kettering Cancer Center
FX Mild Hirano was supported by a Medical Student Summer Fellowship from
   the Brain Tumor Center of Memorial Sloan Kettering Cancer Center.
CR GAUTIER JC, 1961, BRAIN, V84, P341, DOI 10.1093/brain/84.3.341
   Goyal M, 2000, AM J NEURORADIOL, V21, P1073
   Hornyak M, 2008, ACTA NEUROCHIR, V150, P149, DOI 10.1007/s00701-007-1470-0
   KITAJIMA M, 1994, RADIOLOGY, V192, P539, DOI 10.1148/radiology.192.2.8029428
   Krings T, 2003, J NEUROL NEUROSUR PS, V74, P797, DOI 10.1136/jnnp.74.6.797
   Litkowski P, 2012, CLIN IMAG, V36, P371, DOI 10.1016/j.clinimag.2011.09.014
   Phatouros CC, 1998, PEDIATR RADIOL, V28, P830, DOI 10.1007/s002470050475
   Sanverdi SE, 2012, BRIT J RADIOL, V85, P511, DOI 10.1259/bjr/60727602
   Shah R, 2010, AM J NEURORADIOL, V31, P1729, DOI 10.3174/ajnr.A1911
   Shinohara Y, 2013, ACTA RADIOL, V54, P462, DOI 10.1258/ar.2012.120537
NR 10
TC 13
Z9 14
U1 0
U2 3
PU ELSEVIER SCIENCE INC
PI NEW YORK
PA 360 PARK AVE SOUTH, NEW YORK, NY 10010-1710 USA
SN 0899-7071
EI 1873-4499
J9 CLIN IMAG
JI Clin. Imaging
PD SEP-OCT
PY 2015
VL 39
IS 5
BP 787
EP 790
DI 10.1016/j.clinimag.2015.05.015
PG 4
WC Radiology, Nuclear Medicine & Medical Imaging
WE Science Citation Index Expanded (SCI-EXPANDED)
SC Radiology, Nuclear Medicine & Medical Imaging
GA CQ7IT
UT WOS:000360776900009
PM 26104225
OA Green Accepted
DA 2023-08-27
ER

PT J
AU Almosnino, G
   Sikora, MJ
   Farrokhi, FR
   Schwartz, SR
   Zeitler, DM
AF Almosnino, Galit
   Sikora, Matt J.
   Farrokhi, Farrokh R.
   Schwartz, Seth R.
   Zeitler, Daniel M.
TI Tumor-Related and Patient-Related Variables Affecting Length of Hospital
   Stay Following Vestibular Schwannoma Microsurgery
SO ANNALS OF OTOLOGY RHINOLOGY AND LARYNGOLOGY
LA English
DT Article
DE vestibular schwannoma; microsurgery; brainstem compression; length of
   stay; discharge
ID ACOUSTIC NEUROMA; NATURAL-HISTORY; UNITED-STATES; MANAGEMENT; TRENDS;
   EPIDEMIOLOGY; EXCISION; SURGERY; COMPLICATIONS; DISPARITIES
AB Objective: Review a single institution's vestibular schwannoma (VS) microsurgery experience to determine (1) correlations between demographics, comorbidities, and/or surgical approach on hospital length of stay (LOS) and discharge disposition and (2) trends in surgical approach over time. Methods: Retrospective case series from a multidisciplinary skull base program at a tertiary care, academic hospital. All adult (>18 years) patients undergoing primary microsurgery for VS between 2008 and 2018 were included. Results: A total of 147 subjects were identified. Surgical approach was split between middle fossa (MF) (16%), retrosigmoid (RS) (35%), and translabyrinthine (TL) (49%) craniotomies. For the 8% of patients had other than routine (OTR) discharge. Mean LOS was significantly longer for patients undergoing RS than either MF or TL. Brainstem compression by the tumor was associated with longer LOS as were diagnoses of chronic obstructive pulmonary disease (COPD) and peripheral vascular disease (PVD). For all discharges, the 40 to 50- and 50 to 60-year-old subgroups had significantly shorter LOS than the 70-years-and-older patients. For the 92% of patients routinely discharged, there was a significantly shorter LOS in the 40 to 50-year-olds compared to the 70-years-and-older patients. There was a significant shift in surgical approach from RS to TL over the study period. Conclusion: Over 90% of VS microsurgery patients were routinely discharged with a median hospital LOS of 3.2 days, both of which are consistent with published data. There is an inverse relationship between age and LOS with patients older than 70 years having significantly longer LOS. Brainstem compression, COPD, PVD, and the RS approach negatively affect LOS.
C1 [Almosnino, Galit] Dartmouth Hitchcock Med Ctr, Dept Otolaryngol, Lebanon, NH 03766 USA.
   [Sikora, Matt J.] Virginia Mason Med Ctr, Neurosci Inst, Seattle, WA 98101 USA.
   [Farrokhi, Farrokh R.] Virginia Mason Med Ctr, Dept Neurosurg, Seattle, WA 98101 USA.
   [Schwartz, Seth R.; Zeitler, Daniel M.] Virginia Mason Med Ctr, Dept Otolaryngol Head & Neck Surg, Listen Life Ctr, Seattle, WA 98101 USA.
C3 Dartmouth College; Virginia Mason Medical Center; Virginia Mason Medical
   Center; Virginia Mason Medical Center
RP Zeitler, DM (通讯作者)，Virginia Mason Med Ctr, Dept Otolaryngol Head & Neck Surg, Mailstop X10-ON,1100 Ninth Ave, Seattle, WA 98004 USA.
EM
OI Zeitler, Daniel/0000-0001-5506-5483
CR Ahmed OH, 2014, OTOLARYNG HEAD NECK, V150, P266, DOI 10.1177/0194599813507234
   Alkins RD, 2021, OTOL NEUROTOL, V42, P1067, DOI 10.1097/MAO.0000000000003107
   Babu R, 2013, J NEUROSURG, V119, P121, DOI 10.3171/2013.1.JNS121370
   Barker FG, 2003, LARYNGOSCOPE, V113, P1332, DOI 10.1097/00005537-200308000-00013
   Bashjawish B, 2019, LARYNGOSCOPE, V129, P1197, DOI 10.1002/lary.27568
   Carlson ML, 2018, J NEUROL SURG PART B, V79, P289, DOI 10.1055/s-0037-1607319
   Carlson ML, 2015, OTOLARYNG HEAD NECK, V153, P440, DOI 10.1177/0194599815590105
   Cole T, 2015, CUREUS J MED SCIENCE, V7, DOI 10.7759/cureus.369
   Doris L, 2005, ARCH OTOLARYNGOL, V131, P241, DOI 10.1001/archotol.131.3.241
   FINK K, ACAD RADIOL
   Gal TJ, 2010, OTOLARYNG HEAD NECK, V142, P677, DOI 10.1016/j.otohns.2010.01.037
   Goshtasbi K, 2020, LARYNGOSCOPE, V130, P482, DOI 10.1002/lary.27950
   Hachesu PR, 2013, HEALTHC INFORM RES, V19, P121
   Hatch JL, 2018, OTOLARYNG HEAD NECK, V159, P102, DOI 10.1177/0194599818765717
   Hein OV, 2006, ANN THORAC SURG, V81, P880, DOI 10.1016/j.athoracsur.2005.09.077
   Kirchmann M, 2017, NEUROSURGERY, V80, P49, DOI 10.1227/NEU.0000000000001414
   Kshettry VR, 2015, J NEURO-ONCOL, V124, P223, DOI 10.1007/s11060-015-1827-9
   Lau T, 2012, J NEUROSURG, V117, P514, DOI 10.3171/2012.5.JNS111858
   Leon J, 2019, J CLIN NEUROSCI, V68, P174, DOI 10.1016/j.jocn.2019.07.002
   Lin EP, 2017, AM J NEURORADIOL, V38, P2034, DOI 10.3174/ajnr.A5213
   Maldonado ST, 2019, OTOLARYNG HEAD NECK, V161, P137, DOI 10.1177/0194599819835495
   McClelland S, 2017, J CLIN NEUROSCI, V42, P143, DOI 10.1016/j.jocn.2017.03.018
   McClelland S, 2011, NEURO-ONCOLOGY, V13, P1252, DOI 10.1093/neuonc/nor118
   Newman JD, 2018, DIABETES CARE, V41, P1268, DOI 10.2337/dc17-2046
   Obaid S, 2018, J AUDIOL OTOL, V22, P236, DOI 10.7874/jao.2018.00164
   Ostrom QT, 2014, NEURO-ONCOLOGY, V16, P1, DOI [10.1093/neuonc/nov189, 10.1093/neuonc/nou327, 10.1093/neuonc/nou223]
   Patel J, 2014, BRIT J NEUROSURG, V28, P20, DOI 10.3109/02688697.2013.815323
   Patel S, 2013, WORLD NEUROSURG, V80, P142, DOI 10.1016/j.wneu.2012.06.029
   Polanczyk CA, 2001, ANN INTERN MED, V134, P637, DOI 10.7326/0003-4819-134-8-200104170-00008
   Raghavan AM, 2020, OTOL NEUROTOL, V41, pE268, DOI 10.1097/MAO.0000000000002510
   Rosenberg SI, 2000, LARYNGOSCOPE, V110, P497, DOI 10.1097/00005537-200004000-00002
   Semaan MT, 2016, LARYNGOSCOPE, V126, pS5, DOI 10.1002/lary.25729
   Sonig A, 2012, NEUROSURG FOCUS, V33, DOI 10.3171/2012.7.FOCUS12193
   Stangerup SE, 2006, OTOL NEUROTOL, V27, P547, DOI 10.1097/00129492-200606000-00018
   Stangerup SE, 2012, OTOLARYNG CLIN N AM, V45, P257, DOI 10.1016/j.otc.2011.12.008
   STEWART TJ, 1975, ARCH OTOLARYNGOL, V101, P91
   TOS M, 1988, ACTA OTO-LARYNGOL, P38
   Visagan R, 2019, J NEUROL SURG PART B, V80, P310, DOI 10.1055/s-0038-1668541
   Vorasubin N, 2018, OTOL NEUROTOL, V39, P1203, DOI 10.1097/MAO.0000000000001960
NR 39
TC 0
Z9 0
U1 2
U2 2
PU SAGE PUBLICATIONS INC
PI THOUSAND OAKS
PA 2455 TELLER RD, THOUSAND OAKS, CA 91320 USA
SN 0003-4894
EI 1943-572X
J9 ANN OTO RHINOL LARYN
JI Ann. Otol. Rhinol. Laryngol.
PD MAY
PY 2022
VL 131
IS 5
BP 535
EP 543
AR 00034894211029103
DI 10.1177/00034894211029103
EA JUL 2021
PG 9
WC Otorhinolaryngology
WE Science Citation Index Expanded (SCI-EXPANDED)
SC Otorhinolaryngology
GA 0O5OR
UT WOS:000679738100001
PM 34210194
DA 2023-08-27
ER

PT J
AU Colletti, V
   Shannon, RV
AF Colletti, V
   Shannon, RV
TI Open set speech perception with auditory brainstem implant?
SO LARYNGOSCOPE
LA English
DT Article; Proceedings Paper
CT Combined Otolaryngological Spring Meeting
CY MAY 15-16, 2005
CL Boca Raton, FL
DE stem implant; speech perception; amplitude modulation
ID ANTEROVENTRAL COCHLEAR NUCLEUS; ELECTRICAL-STIMULATION; NERVE; HEARING
AB Objective. Only a small percentage of auditory brainstem implant (ABI) recipients treated for neurofibromatosis type 2 (NT2) have proved capable of identifying words using only the sound from the ABI. Recently, the ABI was applied to a series of patients with no cochlear nerve or with cochlear disorders that could not benefit from a cochlear implant (i.e., cochlear nerve aplasia or posttraumatic avulsion) or whose benefit was or would be severely compromised. A significant number of these patients have proven capable of understanding speech, including effortless telephone use. In the present study, a series of psychophysical tests were administered to determine the cause of the difference in performance between tumor (T) and nontumor (NT) ABI patients. Study Design. Retrospective case review. Setting. Tertiary referral center. Patients. Twenty patients with ABIs participated in the investigation. Ten were NF2 patients and 10 NT subjects. Patient ages ranged from 24 to 61 years. Eleven were males and nine females. Intervention: Auditory rehabilitation in auditory disconnection caused by cochlea or cochlear nerve disorders. Results: There was a significant correlation between modulation detection thresholds and speech understanding and a significant difference in modulation detection between T and NIT patients. Conclusions. The difference in modulation detection between the two groups suggests a difference in the survival of specific cells in the cochlear nucleus that support modulation. The pattern of results indicates a separate pathway of auditory processing that is specialized for modulated sounds, and that pathway is critical for speech understanding. In NF2 patients, the tumor and surgery may selectively damage this pathway, resulting in poor speech recognition with prosthetic stimulation.
C1 Univ Verona, Dept Otolaryngol, I-37100 Verona, Italy.
   House Ear Res Inst, Los Angeles, CA USA.
C3 University of Verona; House Research Institute
RP Colletti, V (通讯作者)，Policlin G Rossi, P LA Scuro 10, I-37134 Verona, Italy.
EM
CR BRACKMANN DE, 1993, OTOLARYNG HEAD NECK, V108, P624, DOI 10.1177/019459989310800602
   Colletti V, 2000, AM J OTOL, V21, P826
   Colletti V, 2005, OTOLARYNG HEAD NECK, V133, P126, DOI 10.1016/j.otohns.2005.03.022
   Colletti V, 2002, OTOLARYNG HEAD NECK, V127, P84, DOI 10.1067/mhn.2002.126723
   Fu QJ, 2002, NEUROREPORT, V13, P1635, DOI 10.1097/00001756-200209160-00013
   Ghoshal S, 1996, NEUROSCI LETT, V205, P71, DOI 10.1016/0304-3940(96)12386-7
   HILLENBRAND J, 1995, J ACOUST SOC AM, V97, P3099, DOI 10.1121/1.411872
   Laszig R, 1999, CURR OPIN NEUROL, V12, P41, DOI 10.1097/00019052-199902000-00007
   LIBERMAN MC, 1991, J COMP NEUROL, V313, P240, DOI 10.1002/cne.903130205
   NILSSON M, 1994, J ACOUST SOC AM, V95, P1085, DOI 10.1121/1.408469
   RANCK JB, 1975, BRAIN RES, V98, P417, DOI 10.1016/0006-8993(75)90364-9
   SHANNON RV, 1983, HEARING RES, V12, P1, DOI 10.1016/0378-5955(83)90115-6
   SHANNON RV, 1993, OTOLARYNG HEAD NECK, V108, P634, DOI 10.1177/019459989310800603
   SHANNON RV, 1997, IEEE T REHABIL ENG, V5, P1
   ZHAO HB, 1995, BRAIN RES, V700, P295, DOI 10.1016/0006-8993(95)01078-A
NR 15
TC 138
Z9 142
U1 0
U2 14
PU WILEY
PI HOBOKEN
PA 111 RIVER ST, HOBOKEN 07030-5774, NJ USA
SN 0023-852X
EI 1531-4995
J9 LARYNGOSCOPE
JI Laryngoscope
PD NOV
PY 2005
VL 115
IS 11
BP 1974
EP 1978
DI 10.1097/01.mlg.0000178327.42926.ec
PG 5
WC Medicine, Research & Experimental; Otorhinolaryngology
WE Science Citation Index Expanded (SCI-EXPANDED); Conference Proceedings Citation Index - Science (CPCI-S)
SC Research & Experimental Medicine; Otorhinolaryngology
GA 991UM
UT WOS:000233839600013
PM 16319608
DA 2023-08-27
ER

PT J
AU Li, Q
   Zhang, XQ
   Nie, L
   Chen, GS
   Li, H
   Zhang, F
   Zhang, LY
   Hong, L
   Wang, SF
   Wang, H
AF Li, Q.
   Zhang, X-q
   Nie, L.
   Chen, G-s
   Li, H.
   Zhang, F.
   Zhang, L-y
   Hong, L.
   Wang, S-f
   Wang, H.
TI Expression of interferon-gamma in human adrenal gland and kidney tumours
SO BRITISH JOURNAL OF CANCER
LA English
DT Article
DE interferon-gamma; adrenal tumour; clear cell renal cell carcinoma
ID IFN-GAMMA; SIGNAL-TRANSDUCTION; CELLS; INTERLEUKIN-10; PROLIFERATION;
   TRANSCRIPTION; SURVEILLANCE; ESCAPE; GROWTH
AB It is known that interferon-gamma ( IFN-gamma) is produced by activated T and NK lymphoid cells, mononuclear cells, and macrophage and dendritic cells. Our previous studies have shown that IFN- gamma like immunoreactivity also appears in human adrenal cortical tumour and phaeochromocytoma. To investigate whether human tumour cells can produce IFN- gamma, we examined 429 biopsy specimens of 30 kinds of tumour and tumour- surrounding tissues in adrenal glands and in kidneys by using immunohistochemistry and in situ hybridisation. IFN-gamma immunoactivity was shown in 34.3% of the adrenal cortical adenomas, 50% of the adrenal cortical carcinomas, 26.7% of the phaeochromocytomas, 26.7% of the clear cell renal cell carcinomas ( RCCs), 22% of the adrenal cortexes and 40% of medullas adjacent to tumours. The positive samples and expression areas were well overlapped between the IFN- g mRNA and the immunohistochemistry staining. Western blot analysis has further confirmed the immunohistochemistry results by showing a distinct IFN-gamma band corresponding to 17.4 kDa in tissue extracts from adrenal cortical adenoma, phaeochromocytoma and clear cell RCCs. These results indicate that IFN- gamma is produced by some types of tumour cells, suggesting it may play a dual role in the development of these tumours.
C1 Fourth Mil Med Univ, State Key Lab Canc Biol, Xian 710032, Peoples R China.
   Fourth Mil Med Univ, Dept Pathol, Xijing Hosp, Xian 710032, Peoples R China.
   Biomed Ctr, Dept Med Biochem & Microbiol, S-75123 Uppsala, Sweden.
   Fourth Mil Med Univ, Xijing Hosp, Dept Urinary Surg, Xian 710032, Peoples R China.
C3 Air Force Military Medical University; Air Force Military Medical
   University; Air Force Military Medical University
RP Li, Q (通讯作者)，Fourth Mil Med Univ, State Key Lab Canc Biol, Xian 710032, Peoples R China.
EM
CR Buard A, 1998, CANCER RES, V58, P840
   Chia CS, 2002, IMMUNOL LETT, V84, P163, DOI 10.1016/S0165-2478(02)00176-1
   Dovhey SE, 2000, CANCER RES, V60, P5789
   Dunn GP, 2006, NAT REV IMMUNOL, V6, P836, DOI 10.1038/nri1961
   Dunn GP, 2005, NAT IMMUNOL, V6, P722, DOI 10.1038/ni1213
   Dunn GP, 2005, CANCER RES, V65, P3447, DOI 10.1158/0008-5472.CAN-04-4316
   Dunn GP, 2004, IMMUNITY, V21, P137, DOI 10.1016/j.immuni.2004.07.017
   Dunn GP, 2004, ANNU REV IMMUNOL, V22, P329, DOI 10.1146/annurev.immunol.22.012703.104803
   Fukao T, 2000, J IMMUNOL, V164, P64, DOI 10.4049/jimmunol.164.1.64
   Gessani S, 1998, CYTOKINE GROWTH F R, V9, P117, DOI 10.1016/S1359-6101(98)00007-0
   Haque SJ, 1998, SEMIN ONCOL, V25, P14
   Ikeda H, 2002, CYTOKINE GROWTH F R, V13, P95, DOI 10.1016/S1359-6101(01)00038-7
   JACOB CO, 1987, J EXP MED, V166, P798, DOI 10.1084/jem.166.3.798
   Kaplan DH, 1998, P NATL ACAD SCI USA, V95, P7556, DOI 10.1073/pnas.95.13.7556
   KIEFER R, 1991, NEUROSCIENCE, V45, P551, DOI 10.1016/0306-4522(91)90270-X
   Kooy AJW, 1998, J PATHOL, V184, P169
   Lee CK, 2000, J IMMUNOL, V164, P1286, DOI 10.4049/jimmunol.164.3.1286
   Li Q, 1995, Zhonghua Bing Li Xue Za Zhi, V24, P372
   LI Q, 1996, ZHONGHUAN NE FEN MI, V12, P90
   Murphy KM, 2000, ANNU REV IMMUNOL, V18, P451, DOI 10.1146/annurev.immunol.18.1.451
   Nagao M, 2000, HEPATOLOGY, V32, P491, DOI 10.1053/jhep.2000.16470
   NENNESMO I, 1989, BRAIN RES, V504, P306, DOI 10.1016/0006-8993(89)91374-7
   Schweyer S, 2003, BRIT J CANCER, V89, P915, DOI 10.1038/sj.bjc.6601209
   Schweyer S, 2002, J PATHOL, V197, P89, DOI 10.1002/path.1094
   Shankaran V, 2001, NATURE, V410, P1107, DOI 10.1038/35074122
   Street SEA, 2001, BLOOD, V97, P192, DOI 10.1182/blood.V97.1.192
   TRAUGOTT U, 1988, J NEUROL SCI, V84, P257, DOI 10.1016/0022-510X(88)90130-X
   TRIPP CS, 1993, P NATL ACAD SCI USA, V90, P3725, DOI 10.1073/pnas.90.8.3725
   VANDERMEIDE PH, 1986, J GEN VIROL, V67, P1059, DOI 10.1099/0022-1317-67-6-1059
   Wallace WAH, 1999, J PATHOL, V187, P475, DOI 10.1002/(SICI)1096-9896(199903)187:4<475::AID-PATH268>3.0.CO;2-N
   WANDERS A, 1992, J HEART LUNG TRANSPL, V11, P142
   Weber-Nordt RM, 1998, LEUKEMIA LYMPHOMA, V28, P459, DOI 10.3109/10428199809058353
   Wei YP, 2000, J INTERF CYTOK RES, V20, P403, DOI 10.1089/107999000312342
NR 33
TC 4
Z9 4
U1 0
U2 1
PU NATURE PUBLISHING GROUP
PI LONDON
PA MACMILLAN BUILDING, 4 CRINAN ST, LONDON N1 9XW, ENGLAND
SN 0007-0920
EI 1532-1827
J9 BRIT J CANCER
JI Br. J. Cancer
PD JUL 31
PY 2007
VL 97
IS 3
BP 420
EP 425
DI 10.1038/sj.bjc.6603870
PG 6
WC Oncology
WE Science Citation Index Expanded (SCI-EXPANDED)
SC Oncology
GA 195XE
UT WOS:000248444900020
PM 17622250
OA hybrid, Green Published
DA 2023-08-27
ER

PT J
AU Schonfeld, MH
   Ritzel, RM
   Kemmling, A
   Ernst, M
   Fiehler, J
   Gellissen, S
AF Schoenfeld, Michael H.
   Ritzel, Robert M.
   Kemmling, Andre
   Ernst, Marielle
   Fiehler, Jens
   Gellissen, Susanne
TI Improved detectability of acute and subacute brainstem infarctions by
   combining standard axial and thin-sliced sagittal DWI
SO PLOS ONE
LA English
DT Article
ID DIFFUSION-WEIGHTED MR; POSTERIOR-CIRCULATION STROKE; MAGNETIC-RESONANCE;
   ISCHEMIC-STROKE; NIHSS SCORE; SEQUENCES; DIAGNOSIS; PERFUSION; DEFICITS;
   LESION
AB Background and purpose
   Most false negative findings in DWI of ischemic stroke are in patients with minor deficits clinically localized to the brainstem. Our goal was to evaluate the benefit of a thin-sliced sagittal DWI in addition to conventional axial DWI at 1.5T for the detection of brainstem infarctions.
   Methods
   Data of patients with symptoms consistent with acute and subacute brainstem infarction and an MRI examination including standard axial DWI and thin-sliced sagittal DWI were retrospectively analyzed. Patients with the later diagnosis of a TIA, an inflammation or a tumor of the brainstem were excluded from analysis. Diffusion restrictions were identified by two independent raters blinded for the final clinical diagnosis in three separate reading steps: First, only axial DWI, secondly only sagittal DWI, and lastly both DWIs together. Presence and size of DWI-lesions were documented for each plane. Differences between the observers were settled in consensus in a separate joint reading.
   Results
   Of 73 included patients, 46 patients were clinically diagnosed with brainstem infarction. Inter-observer agreement was excellent for the detection of brainstem lesions in axial and sagittal DWI (kappa = 0.94 and 0.97). In 28/46 patients (60.9%) lesions were detected in the axial plane alone, whereas in 6 more patients (73.9%) lesions were detected in the review of both sequences together. All lesions undetectable in the axial plane were smaller than 5 mm in cranio-caudal direction.
   Conclusions
   Thin-sliced sagittal DWI in addition to axial DWI improves the detection rate of brainstem infarction with little additional expenditure of time.
C1 [Schoenfeld, Michael H.; Ernst, Marielle; Fiehler, Jens; Gellissen, Susanne] Univ Med Ctr Hamburg Eppendorf, Dept Diagnost & Intervent Neuroradiol, Hamburg, Germany.
   [Ritzel, Robert M.] German Armed Forces Hosp Hamburg, Dept Radiol, Hamburg, Germany.
   [Kemmling, Andre] Univ Hosp Luebeck, Dept Neuroradiol, Lubeck, Germany.
C3 University of Hamburg; University Medical Center Hamburg-Eppendorf;
   University of Lubeck
RP Schonfeld, MH (通讯作者)，Univ Med Ctr Hamburg Eppendorf, Dept Diagnost & Intervent Neuroradiol, Hamburg, Germany.
EM
RI Ernst, Marielle/IAQ-0190-2023
OI Ernst, Marielle/0000-0003-2870-4109; Schonfeld,
   Michael/0000-0003-2096-3448
CR [Anonymous], 2011, ATLAS KLIN NEURORADI
   Ay H, 1999, NEUROLOGY, V52, P1784, DOI 10.1212/WNL.52.9.1784
   Byeon J, 2015, CLIN IMAG, V39, P765, DOI 10.1016/j.clinimag.2015.06.001
   Chalela JA, 2007, LANCET, V369, P293, DOI 10.1016/S0140-6736(07)60151-2
   Engelter ST, 2004, NEUROLOGY, V62, P574, DOI 10.1212/01.WNL.0000110310.48162.F7
   Etgen T, 2004, EUR NEUROL, V52, P145, DOI 10.1159/000081623
   Felfeli P, 2017, BRAIN BEHAV, V7, DOI 10.1002/brb3.666
   Fitzek S, 2001, EUR J RADIOL, V39, P180, DOI 10.1016/S0720-048X(01)00372-2
   Gerraty RP, 2002, STROKE, V33, P2019, DOI 10.1161/01.STR.0000020841.74704.5B
   Hayashi S, 2009, NOSOTCHU, V31, P34, DOI [10.3995/jstroke.31.34, DOI 10.3995/JSTROKE.31.34]
   Hong GS, 2017, EUR RADIOL, V27, P859, DOI 10.1007/s00330-016-4388-1
   Huo J, 2016, CLIN NEURORADIOL, V26, P423, DOI 10.1007/s00062-015-0381-3
   Kuker W, 2002, J NEUROL, V249, P33, DOI 10.1007/PL00007845
   Lee H, 2014, J STROKE, V16, P124, DOI 10.5853/jos.2014.16.3.124
   Linfante I, 2001, ARCH NEUROL-CHICAGO, V58, P621, DOI 10.1001/archneur.58.4.621
   Lovblad KO, 1998, AM J NEURORADIOL, V19, P1061
   Marx JJ, 2002, J NEUROL NEUROSUR PS, V72, P572, DOI 10.1136/jnnp.72.5.572
   Moritani T., 2009, DIFFUSION WEIGHTED M, V2nd
   Narisawa A, 2001, No To Shinkei, V53, P1021
   Oppenheim C, 2000, AM J NEURORADIOL, V21, P1434
   Pandya A, 2016, PLOS ONE, V11, DOI 10.1371/journal.pone.0148106
   Paul NLM, 2013, LANCET NEUROL, V12, P65, DOI 10.1016/S1474-4422(12)70299-5
   Porter DA, 2009, MAGN RESON MED, V62, P468, DOI 10.1002/mrm.22024
   Seeger A, 2016, CLIN NEURORADIOL, V26, P177, DOI 10.1007/s00062-014-0342-2
   Seo Min-Jung, 2006, J Clin Neurol, V2, P107, DOI 10.3988/jcn.2006.2.2.107
   Sorimachi T, 2008, NEUROL MED-CHIR, V48, P108, DOI 10.2176/nmc.48.108
   Toi H, 2003, NEURORADIOLOGY, V45, P352, DOI 10.1007/s00234-002-0897-5
   Tong DC, 1998, NEUROLOGY, V50, P864, DOI 10.1212/WNL.50.4.864
   Wardlaw J, 2014, HEALTH TECHNOL ASSES, V18, P1
   Wilson CA, 2017, CLIN NEURORADIOL, V27, P31, DOI 10.1007/s00062-015-0409-8
NR 30
TC 14
Z9 16
U1 0
U2 0
PU PUBLIC LIBRARY SCIENCE
PI SAN FRANCISCO
PA 1160 BATTERY STREET, STE 100, SAN FRANCISCO, CA 94111 USA
SN 1932-6203
J9 PLOS ONE
JI PLoS One
PD JUL 3
PY 2018
VL 13
IS 7
AR e0200092
DI 10.1371/journal.pone.0200092
PG 11
WC Multidisciplinary Sciences
WE Science Citation Index Expanded (SCI-EXPANDED)
SC Science & Technology - Other Topics
GA GL5XD
UT WOS:000437246700042
PM 29969485
OA Green Published, gold, Green Submitted
DA 2023-08-27
ER

PT J
AU Herbach, N
   Nagel, L
   Zwick, T
   Hermanns, W
AF Herbach, N.
   Nagel, L.
   Zwick, T.
   Hermanns, W.
TI Multiple Glucagon-Producing Pancreatic Neuroendocrine Tumors in a Horse
   (Equus caballus)
SO VETERINARY PATHOLOGY
LA English
DT Article
DE glucagonoma; pancreatic neuroendocrine tumor; multiple islet-cell
   adenomas; horse
AB Pancreatic neuroendocrine tumors of glucagon-producing cells are extremely rare in domestic animals. In this report, we describe for the first time, to our knowledge, the incidental finding of multiple glucagon-producing neuroendocrine tumors of the pancreas of a horse. The animal was euthanized due to severe local infection after tooth extraction. On postmortem examination, multiple white nodules of up to 4 cm in diameter were observed in the pancreas. Histologically, pancreatic nodules had the appearance of neuroendocrine neoplasms with positive immunoreactivity for glucagon, synaptophysin, chromogranin A, and neuron-specific enolase. Electron microscopy revealed numerous electron-dense granules, similar to those observed in normal pancreatic alpha cells, in the neoplastic cells. In addition, the left adrenal gland showed multiple hyperplastic foci and adenomas in the medulla that were identified as pheochromocytomas. Based on the morphologic appearance and immunohistochemical staining pattern of pancreatic nodules, a diagnosis of multiple glucagon-producing neuroendocrine tumors was made.
C1 [Herbach, N.; Nagel, L.; Hermanns, W.] Univ Munich, Ctr Clin Vet Med, Inst Vet Pathol, D-80539 Munich, Germany.
   [Zwick, T.] Vet Clin Gessertshausen, Gessertshausen, Germany.
C3 University of Munich
RP Herbach, N (通讯作者)，Univ Munich, Ctr Clin Vet Med, Inst Vet Pathol, Vet Str 13, D-80539 Munich, Germany.
EM
OI Herbach, Nadja/0000-0001-5893-0356
CR Charles J, 2007, JUBB KENNEDY PALMERS, V2, P389
   Heitz PU, 2004, PATHOLOGY GENETICS T, P177
   Jabbour SA, 2006, CLIN DERMATOL, V24, P299, DOI 10.1016/j.clindermatol.2006.04.005
   Kelley LC, 1996, VET PATHOL, V33, P398, DOI 10.1177/030098589603300405
   Kiupel M., 2008, HISTOLOGICAL CLASSIF
   Kloppel G, 2007, BEST PRACT RES CL EN, V21, P15, DOI 10.1016/j.beem.2007.01.004
   Kloppel G, 2011, ENDOCR-RELAT CANCER, V18, pS1, DOI 10.1530/ERC-11-0013
   Lurye JC, 2001, VET CLIN N AM-SMALL, V31, P1083, DOI 10.1016/S0195-5616(01)50014-5
   OBRIEN TD, 1987, VET PATHOL, V24, P308, DOI 10.1177/030098588702400404
   Ouyang D, 2011, WORLD J GASTROENTERO, V17, P137, DOI 10.3748/wjg.v17.i2.137
   PRIESTER WA, 1974, J NATL CANCER I, V53, P227, DOI 10.1093/jnci/53.1.227
   Ramos-Vara JA, 2000, J ZOO WILDLIFE MED, V31, P563, DOI 10.1638/1042-7260(2000)031[0563:GIAJPO]2.0.CO;2
NR 12
TC 5
Z9 5
U1 0
U2 5
PU SAGE PUBLICATIONS INC
PI THOUSAND OAKS
PA 2455 TELLER RD, THOUSAND OAKS, CA 91320 USA
SN 0300-9858
EI 1544-2217
J9 VET PATHOL
JI Vet. Pathol.
PD MAY
PY 2014
VL 51
IS 3
BP 607
EP 611
DI 10.1177/0300985813492803
PG 5
WC Pathology; Veterinary Sciences
WE Science Citation Index Expanded (SCI-EXPANDED)
SC Pathology; Veterinary Sciences
GA AJ5CB
UT WOS:000337697400008
PM 23774746
OA Green Published
DA 2023-08-27
ER

PT J
AU NAZZARO, JM
   SHULTS, WT
   NEUWELT, EA
AF NAZZARO, JM
   SHULTS, WT
   NEUWELT, EA
TI NEUROOPHTHALMOLOGICAL FUNCTION OF PATIENTS WITH PINEAL REGION TUMORS
   APPROACHED TRANSTENTORIALLY IN THE SEMISITTING POSITION
SO JOURNAL OF NEUROSURGERY
LA English
DT Article
DE PINEAL TUMOR; PARINAUD SYNDROME; NEUROOPHTHALMOLOGY; OCCIPITAL
   TRANSTENTORIAL APPROACH
ID MICROSURGICAL ANATOMY; MICRO-SURGERY; 3RD VENTRICLE; INFRATENTORIAL;
   LESIONS; CT
AB To optimize orientation and operative exposure for aggressive resection, the authors approached pineal region tumors transtentorially with the patient in a semisitting position. In the current report, 12 consecutive patients were evaluated to document operative ocular morbidity referable to the brain stem as well as visual deficits secondary to occipital lobe retraction. Before craniotomy, ophthalmological findings related to dorsal midbrain dysfunction were evident in four of the 10 patients who had previously undergone ventricular shunting. The other patients developed a partial or complete Parinaud's syndrome in the early postoperative period and some suffered additional deficits such as cranial nerve palsies. These deficits improved to varying degrees in all patients. Although each had full visual fields preoperatively, an absolute or incomplete left homonymous hemianopsia developed in the immediate postoperative period. Such visual field deficits fully resolved over a variable period of time in 10 of the 12 patients. One patient has a permanent left homonymous hemianopsia, while another has a left homonymous paracentral scotoma. Eight patients were able to return to preoperative pursuits. While ocular abnormalities related to dorsal midbrain dysfunction are most probably independent of operative approach, visual field deficits attributable to occipital lobe retraction were common in patients after a occipital transtentorial approach performed in the semisitting position. Reading difficulties associated with ocular motor dysfunction due to dorsal midbrain involvement represent the principal long-term neuro-ophthalmological complaint of patients who have undergone pineal region surgery.
C1 OREGON HLTH SCI UNIV,DEPT SURG,DIV NEUROSURG,MAIL STOP L603,3181 SW SAM JACKSON PK RD,PORTLAND,OR 97201.
   OREGON HLTH SCI UNIV,DEPT NEUROL,PORTLAND,OR 97201.
   OREGON HLTH SCI UNIV,DEPT MOLEC BIOL,PORTLAND,OR 97201.
   OREGON HLTH SCI UNIV,DEPT BIOCHEM,PORTLAND,OR 97201.
   GOOD SAMARITAN HOSP,DEVERS EYE INST,PORTLAND,OR.
C3 Oregon Health & Science University; Oregon Health & Science University;
   Oregon Health & Science University; Oregon Health & Science University;
   Devers Eye Institute
FU PHS HHS [31770] Funding Source: Medline
CR ANDERSON RE, 1984, DIAGNOSIS TREATMENT, P61
   AUSMAN JI, 1988, SURG NEUROL, V29, P298, DOI 10.1016/0090-3019(88)90161-9
   BALOH RW, 1985, NEUROLOGY, V35, P54, DOI 10.1212/WNL.35.1.54
   CHANG T, 1989, AM J ROENTGENOL, V153, P1269, DOI 10.2214/ajr.153.6.1269
   CHAPMAN PH, 1980, CANCER, V46, P1253, DOI 10.1002/1097-0142(19800901)46:5<1253::AID-CNCR2820460528>3.0.CO;2-D
   Clark WK, 1987, SURG 3 VENTRICLE, P591
   Cushing H., 1932, INTRACRANIAL TUMOURS
   Daroff R.B., 1971, CONTROL EYE MOVEMENT, P175
   DAVIDOFF LM, 1967, B NEW YORK ACAD MED, V43, P537
   DAVIS RL, 1987, SURG 3 VENTRICLE, P235
   DEGIROLAMI U, 1973, J NEUROSURG, V39, P455, DOI 10.3171/jns.1973.39.4.0455
   DONAT JF, 1978, ARCH NEUROL-CHICAGO, V35, P736, DOI 10.1001/archneur.1978.00500350040008
   EDWARDS MSB, 1988, J NEUROSURG, V68, P689, DOI 10.3171/jns.1988.68.5.0689
   FUTRELL NN, 1981, AM J ROENTGENOL, V137, P951, DOI 10.2214/ajr.137.5.951
   GANTI SR, 1986, AM J ROENTGENOL, V146, P451
   HERRICK MK, 1984, DIAGNOSIS TREATMENT, P31
   Hoffman HJ, 1983, CONCEPTS PEDIATR NEU, V4, P360
   HORRAX G, 1950, ARCH NEURO PSYCHIATR, V64, P227, DOI 10.1001/archneurpsyc.1950.02310260065004
   JAMIESON KG, 1971, J NEUROSURG, V35, P550, DOI 10.3171/jns.1971.35.5.0550
   KEANE JR, 1975, ARCH NEUROL-CHICAGO, V32, P185, DOI 10.1001/archneur.1975.00490450065009
   KOBAYASHI S, 1983, J NEUROSURG, V58, P141, DOI 10.3171/jns.1983.58.1.0141
   KUNICKI A, 1960, J NEUROSURG, V17, P815, DOI 10.3171/jns.1960.17.5.0815
   KWAN E, 1987, SURG 3 VENTRICLE, P262
   LAPRAS C, 1987, PROG EXP TUMOR RES, V30, P268
   Lapras C., 1984, DIAGNOSIS TREATMENT, P289
   LUO S, 1989, SURG NEUROL, V32, P36, DOI 10.1016/0090-3019(89)90032-3
   McComb J, 1988, CONCEPTS PEDIAT NEUR, V8, P186
   Neuwelt E A, 1985, Clin Neurosurg, V32, P397
   NEUWELT EA, 1979, J NEUROSURG, V51, P597, DOI 10.3171/jns.1979.51.5.0597
   NEUWELT EA, 1985, NEUROSURGERY, V16, P373, DOI 10.1227/00006123-198503000-00015
   NEUWELT EA, 1984, DIAGNOSIS TREATMENT, P208
   NEUWELT EA, 1984, DIAGNOSIS TREATMENT, P273
   Page L K, 1977, Neurosurgery, V1, P36
   Parinaud H., 1886, BRAIN, V9, P330, DOI 10.1093/brain/9.3.330
   Pendl G, 1984, DIAGNOSIS TREATMENT, P139
   PENDL G, 1984, DIAGNOSIS TREATMENT, P155
   POPPEN JL, 1968, J NEUROSURG, V28, P357, DOI 10.3171/jns.1968.28.4.0357
   POPPEN JL, 1966, J NEUROSURG, V25, P706, DOI 10.3171/jns.1966.25.6.0706
   POSNER M, 1946, J NEUROSURG, V3, P15, DOI 10.3171/jns.1946.3.1.0015
   REID WS, 1978, NEUROSURGERY, V3, P1
   RHOTON AL, 1981, NEUROSURGERY, V8, P357, DOI 10.1227/00006123-198103000-00007
   RINGERTZ N, 1954, J NEUROPATH EXP NEUR, V13, P540, DOI 10.1097/00005072-195410000-00004
   Russell WO, 1943, ARCH PATHOL, V35, P869
   Scheithauer B W, 1985, Clin Neurosurg, V32, P351
   SEYBOLD ME, 1971, NEUROLOGY, V21, P232, DOI 10.1212/WNL.21.3.232
   SHULTS WT, 1984, DIAGNOSIS TREATMENT, P108
   Stein B, 1987, SURG 3 VENTRICLE, P570
   STEIN BM, 1971, J NEUROSURG, V35, P197, DOI 10.3171/jns.1971.35.2.0197
   STONE JL, 1990, ACTA NEUROCHIR, V102, P133, DOI 10.1007/BF01405427
   SUZUKI J, 1965, J NEUROSURG, V23, P565, DOI 10.3171/jns.1965.23.6.0565
   TIEN RD, 1990, AM J NEURORADIOL, V11, P557
   VENTUREYRA ECG, 1981, SURG NEUROL, V16, P77, DOI 10.1016/S0090-3019(81)80073-0
   WRAY SH, 1977, PINEAL TUMORS, P21
   YAMAMOTO I, 1980, J NEUROSURG, V53, P205, DOI 10.3171/jns.1980.53.2.0205
   YAMAMOTO I, 1981, NEUROSURGERY, V8, P334, DOI 10.1227/00006123-198103000-00006
NR 55
TC 56
Z9 58
U1 0
U2 0
PU AMER ASSOC NEUROLOGICAL SURGEONS
PI CHARLOTTESVILLE
PA UNIV VIRGINIA, 1224 WEST MAIN ST, STE 450, CHARLOTTESVILLE, VA 22903
SN 0022-3085
J9 J NEUROSURG
JI J. Neurosurg.
PD MAY
PY 1992
VL 76
IS 5
BP 746
EP 751
DI 10.3171/jns.1992.76.5.0746
PG 6
WC Clinical Neurology; Surgery
WE Science Citation Index Expanded (SCI-EXPANDED)
SC Neurosciences & Neurology; Surgery
GA HQ269
UT WOS:A1992HQ26900004
PM 1564536
DA 2023-08-27
ER

PT J
AU Akhavan-Sigari, R
   Ostertag, H
   Rohde, V
   Tezval, H
AF Akhavan-Sigari, Reza
   Ostertag, Helmut
   Rohde, Veit
   Tezval, Hossein
TI Connection between expression of inducible nitric oxide synthase (iNOS)
   in skull base chordoma and lower urinary tract symptoms
SO INTERNATIONAL UROLOGY AND NEPHROLOGY
LA English
DT Article
DE Lower urinary tract symptoms; Skull base chordoma; Brainstem; INOS
ID PONTINE MICTURITION CENTER; GROWTH-FACTOR RECEPTOR; POSTERIOR-FOSSA;
   GASTRIC-CANCER; CONTINENCE; ASSOCIATION; DISTURBANCE; RETENTION; KI-M1P;
   TUMORS
AB To provide first insights into the potential role of iNOS expressed by skull base chordoma, which causes brainstem compression in and around Barrington's nucleus, and its effect on the micturition center.
   Urodynamic testing of 22 symptomatic patients was performed. All women and men with skull base chordoma treated in two hospitals in Germany between 1986 and 2007 were studied. Lower urinary tract symptoms (LUTS) were documented in patients with acute brainstem compression due to local chordoma growth positive for iNOS expression. Brain magnetic resonance (MRI) images of the lesions of the symptomatic patients were performed.
   Of 74 treated patients, 22 (7 women, 15 men) with a median age of 37 years were evaluated with voiding diaries and computer urodynamic investigation. Urodynamic testing of 22 symptomatic patients with positive iNOS expression of skull base chordoma revealed detrusor overactivity in 55 %, low-compliance bladder in 14 %, detrusor sphincter dyssynergia in 45 % and uninhibited sphincter relaxation in 27 %. There was a significant correlation between strong iNOS expression (score 3-6) in skull base chordoma and severe urinary symptoms (p = 0.003, Spearman rho = 0.526).
   The expression of iNOS in skull base chordoma compressing the dorsolateral pons, in and around Barrington's nucleus, may influence the pontine micturition center (PMC) and be responsible for lower urinary tract symptoms. Nitric oxide may possibly act as a neurotransmitter. We assume that the high infiltration of chordoma with monocyte/macrophages enhances the release of nitric oxide, as monocyte/macrophages are the main source of iNOS.
C1 [Akhavan-Sigari, Reza; Rohde, Veit] Univ Gottingen, Dept Neurosurg, Univ Med Ctr Gottingen, D-37075 Gottingen, Germany.
   [Ostertag, Helmut] Klinikum Hannover, Dept Pathol, Nordstadt Med Ctr, D-30167 Hannover, Germany.
   [Tezval, Hossein] Hannover Med Sch, Dept Urol, D-30625 Hannover, Germany.
C3 University of Gottingen; Hannover Medical School
RP Akhavan-Sigari, R (通讯作者)，Univ Gottingen, Dept Neurosurg, Univ Med Ctr Gottingen, Robert Koch Str 40, D-37075 Gottingen, Germany.
EM;
  
RI Akhavan-Sigari, Reza/HQY-3303-2023
OI Akhavan-Sigari, Reza/0000-0001-6118-1704
CR Abrams P, 2003, UROLOGY, V61, P37, DOI 10.1016/S0090-4295(02)02243-4
   Akhavan-Sigari R, 2014, UROLOGY, V83, P756, DOI 10.1016/j.urology.2013.12.018
   Akhavan-Sigari R, 2014, NEUROSURG REV, V37, P79, DOI 10.1007/s10143-013-0495-5
   [Anonymous], 1960, NEUROL MED-CHIR, DOI DOI 10.2176/NMC.2.25
   Baillieux H, 2006, CEREBELLUM, V5, P289, DOI 10.1080/14734220601009606
   BENNETT BC, 1995, J UROLOGY, V153, P2004, DOI 10.1016/S0022-5347(01)67391-9
   BETTS CD, 1992, BRIT J UROL, V70, P100, DOI 10.1111/j.1464-410X.1992.tb15679.x
   Blok BFM, 2002, UROLOGY, V59, P13, DOI 10.1016/S0090-4295(01)01633-8
   Dalmose AL, 2005, BJU INT, V95, P886, DOI 10.1111/j.1464-410X.2005.05421.x
   de Groat WC, 1997, UROLOGY, V50, P36, DOI 10.1016/S0090-4295(97)00587-6
   FEDER M, 1987, EUR NEUROL, V27, P101, DOI 10.1159/000116140
   FEHSEL K, 1993, DIABETES, V42, P496, DOI 10.2337/diabetes.42.3.496
   Forstermann U, 2012, EUR HEART J, V33, P829, DOI 10.1093/eurheartj/ehr304
   Holman E, 1926, ARCH NEURO PSYCHIATR, V15, P371, DOI 10.1001/archneurpsyc.1926.02200210092009
   HSU SM, 1981, J HISTOCHEM CYTOCHEM, V29, P577, DOI 10.1177/29.4.6166661
   Iwasaki H, 2004, UROLOGY, V63, P994, DOI 10.1016/j.urology.2003.11.036
   Lee KB, 2008, NEUROLOGIST, V14, P312, DOI 10.1097/NRL.0b013e3181714a09
   Manente G, 1996, J NEUROL NEUROSUR PS, V61, P528, DOI 10.1136/jnnp.61.5.528-a
   Mattern J, 1996, BRIT J CANCER, V73, P931, DOI 10.1038/bjc.1996.166
   NATHAN CF, 1991, CURR OPIN IMMUNOL, V3, P65, DOI 10.1016/0952-7915(91)90079-G
   Nicotera P, 1995, ADV NEUROIMMUNOL, V5, P411, DOI 10.1016/0960-5428(95)00025-9
   RADZUN HJ, 1991, LAB INVEST, V65, P306
   Rimm DL, 2001, CANCER J, V7, P24
   Sakakibara R, 1996, J NEUROL SCI, V141, P105, DOI 10.1016/0022-510X(96)00079-2
   Sakakibara R, 1998, J NEUROL NEUROSUR PS, V64, P269, DOI 10.1136/jnnp.64.2.269
   Sakakibara R, 1995, J AUTONOM NERV SYST, V50, P362
   Son HJ, 2001, J CLIN GASTROENTEROL, V33, P383, DOI 10.1097/00004836-200111000-00008
   Song ZJ, 2002, WORLD J GASTROENTERO, V8, P591, DOI 10.3748/wjg.v8.i4.591
   Verne GN, 2003, PAIN, V103, P99, DOI 10.1016/S0304-3959(02)00416-5
   Weinberger PM, 2005, ARCH OTOLARYNGOL, V131, P707, DOI 10.1001/archotol.131.8.707
   Zhang H, 2005, NEUROIMAGE, V24, P174, DOI 10.1016/j.neuroimage.2004.08.027
NR 31
TC 0
Z9 0
U1 0
U2 1
PU SPRINGER
PI DORDRECHT
PA VAN GODEWIJCKSTRAAT 30, 3311 GZ DORDRECHT, NETHERLANDS
SN 0301-1623
EI 1573-2584
J9 INT UROL NEPHROL
JI Int. Urol. Nephrol.
PD NOV
PY 2014
VL 46
IS 11
BP 2109
EP 2116
DI 10.1007/s11255-014-0806-y
PG 8
WC Urology & Nephrology
WE Science Citation Index Expanded (SCI-EXPANDED)
SC Urology & Nephrology
GA AS5RS
UT WOS:000344328000008
PM 25113512
DA 2023-08-27
ER

PT J
AU Gajjar, A
   Bowers, DC
   Karajannis, MA
   Leary, S
   Witt, H
   Gottardo, NG
AF Gajjar, Amar
   Bowers, Daniel C.
   Karajannis, Matthias A.
   Leary, Sarah
   Witt, Hendrik
   Gottardo, Nicholas G.
TI Pediatric Brain Tumors: Innovative Genomic Information Is Transforming
   the Diagnostic and Clinical Landscape
SO JOURNAL OF CLINICAL ONCOLOGY
LA English
DT Review
ID CENTRAL-NERVOUS-SYSTEM; PRIMITIVE NEUROECTODERMAL TUMORS; TERATOID
   RHABDOID TUMORS; LOW-GRADE GLIOMAS; CRANIOSPINAL RADIATION-THERAPY;
   INTRINSIC PONTINE GLIOMA; HIGH-DOSE CHEMOTHERAPY; EMBRYONAL TUMOR;
   CHILDHOOD MEDULLOBLASTOMA; PILOCYTIC ASTROCYTOMAS
AB Pediatric neuro-oncology has undergone an exciting and dramatic transformation during the past 5 years. This article summarizes data from collaborative group and institutional trials that have advanced the science of pediatric brain tumors and survival of patients with these tumors. Advanced genomic analysis of the entire spectrum of pediatric brain tumors has heralded an era in which stakeholders in the pediatric neuro-oncology community are being challenged to reconsider their current research and diagnostic and treatment strategies. The incorporation of this new information into the next-generation treatment protocols will unleash new challenges. This review succinctly summarizes the key advances in our understanding of the common pediatric brain tumors (ie, medulloblastoma, low- and high-grade gliomas, diffuse intrinsic pontine glioma, and ependymoma) and some selected rare tumors (ie, atypical teratoid/rhabdoid tumor and CNS primitive neuroectodermal tumor). The potential impact of this new information on future clinical protocols also is discussed. Cutting-edge genomics technologies and the information gained from such studies are facilitating the identification of molecularly defined subgroups within patients with particular pediatric brain tumors. The number of evaluable patients in each subgroup is small, particularly in the subgroups of rare diseases. Therefore, international collaboration will be crucial to draw meaningful conclusions about novel approaches to treating pediatric brain tumors. (C) 2015 by American Society of Clinical Oncology
C1 [Gajjar, Amar] St Jude Childrens Res Hosp, Memphis, TN 38105 USA.
   [Bowers, Daniel C.] Univ Texas SW Med Ctr Dallas, Dallas, TX 75390 USA.
   [Karajannis, Matthias A.] NYU, Perlmutter Canc Ctr, New York, NY USA.
   [Karajannis, Matthias A.] NYU, Langone Med Ctr, New York, NY USA.
   [Leary, Sarah] Univ Washington, Sch Med, Seattle, WA USA.
   [Leary, Sarah] Fred Hutchinson Canc Res Ctr, Seattle, WA 98104 USA.
   [Witt, Hendrik] German Canc Res Ctr, Heidelberg, Germany.
   [Witt, Hendrik] Heidelberg Univ, Heidelberg, Germany.
   [Gottardo, Nicholas G.] Princess Margaret Hosp Children, Perth, WA, Australia.
   [Gottardo, Nicholas G.] Univ Western Australia, Perth, WA 6009, Australia.
C3 St Jude Children's Research Hospital; University of Texas System;
   University of Texas Southwestern Medical Center Dallas; New York
   University; New York University; NYU Langone Medical Center; University
   of Washington; University of Washington Seattle; Fred Hutchinson Cancer
   Center; Helmholtz Association; German Cancer Research Center (DKFZ);
   Ruprecht Karls University Heidelberg; University of Western Australia;
   University of Western Australia
RP Gajjar, A (通讯作者)，St Jude Childrens Res Hosp, Dept Oncol, MS 260,262 Danny Thomas Pl, Memphis, TN 38105 USA.
EM
RI Gajjar, Amar/N-8059-2018; Karajannis, Matthias A/H-8845-2019; mohamady,
   mahin/G-2175-2016
OI Karajannis, Matthias A/0000-0002-7151-6528; Bowers,
   Daniel/0000-0002-3947-2481; Gottardo, Nicholas/0000-0002-1082-6776
FU St Jude Children's Research Hospital [CA21765]; Raine Medical Research
   Foundation's Clinical Research Fellowship Program; Cancer Center
FX Supported by St Jude Children's Research Hospital with Grant No. CA21765
   (A.G.) provided by the Cancer Center Support, the Noyes Brain Tumor
   Foundation, Musicians Against Childhood Cancer, and the American
   Lebanese-Syrian Associated Charities. N.G.G. is supported by The Raine
   Medical Research Foundation's Clinical Research Fellowship Program.
CR Ater JL, 2012, J CLIN ONCOL, V30, P2641, DOI 10.1200/JCO.2011.36.6054
   Athale UH, 2009, J PEDIAT HEMATOL ONC, V31, P651, DOI 10.1097/MPH.0b013e3181b258a9
   Atkinson JM, 2011, CANCER CELL, V20, P384, DOI 10.1016/j.ccr.2011.08.013
   Bandopadhayay P, 2014, PEDIATR BLOOD CANCER, V61, P1173, DOI 10.1002/pbc.24958
   Bandopadhayay P, 2014, CLIN CANCER RES, V20, P912, DOI 10.1158/1078-0432.CCR-13-2281
   Bar EE, 2008, J NEUROPATH EXP NEUR, V67, P878, DOI 10.1097/NEN.0b013e3181845622
   Biegel JA, 1999, CANCER RES, V59, P74
   Bradley KA, 2013, INT J RADIAT ONCOL, V85, pE55, DOI 10.1016/j.ijrobp.2012.09.004
   Buccoliero AM, 2012, FETAL PEDIATR PATHOL, V31, P278, DOI 10.3109/15513815.2012.659383
   Buczkowicz P, 2014, NAT GENET, V46, P451, DOI 10.1038/ng.2936
   Chapman PB, 2011, NEW ENGL J MED, V364, P2507, DOI 10.1056/NEJMoa1103782
   Chi SN, 2009, J CLIN ONCOL, V27, P385, DOI 10.1200/JCO.2008.18.7724
   Chinot OL, 2014, NEW ENGL J MED, V370, P709, DOI 10.1056/NEJMoa1308345
   Chintagumpala M, 2009, NEURO-ONCOLOGY, V11, P33, DOI 10.1215/15228517-2008-079
   Cin H, 2011, ACTA NEUROPATHOL, V121, P763, DOI 10.1007/s00401-011-0817-z
   COHEN BH, 1995, J CLIN ONCOL, V13, P1687, DOI 10.1200/JCO.1995.13.7.1687
   Cohen KJ, 2011, NEURO-ONCOLOGY, V13, P410, DOI 10.1093/neuonc/noq205
   Cohen KJ, 2011, NEURO-ONCOLOGY, V13, P317, DOI 10.1093/neuonc/noq191
   Dawson MA, 2012, CELL, V150, P12, DOI 10.1016/j.cell.2012.06.013
   Dhanak D, 2014, BIOCHEM BIOPH RES CO, V455, P58, DOI 10.1016/j.bbrc.2014.07.006
   Eisenhardt AE, 2011, INT J CANCER, V129, P2297, DOI 10.1002/ijc.25893
   Ellison DW, 2011, J NEGAT RESULTS BIOM, V10, DOI 10.1186/1477-5751-10-7
   Ellison DW, 2011, J CLIN ONCOL, V29, P1400, DOI 10.1200/JCO.2010.30.2810
   Ellison DW, 2005, J CLIN ONCOL, V23, P7951, DOI 10.1200/JCO.2005.01.5479
   Fangusaro JR, 2008, PEDIATR BLOOD CANCER, V50, P715, DOI 10.1002/pbc.21032
   Fisher PG, 2008, PEDIATR BLOOD CANCER, V51, P245, DOI 10.1002/pbc.21563
   Fontebasso AM, 2014, NAT GENET, V46, P462, DOI 10.1038/ng.2950
   Forshew T, 2009, J PATHOL, V218, P172, DOI 10.1002/path.2558
   Franz DN, 2006, ANN NEUROL, V59, P490, DOI 10.1002/ana.20784
   Gajjar A, 1997, J CLIN ONCOL, V15, P2792, DOI 10.1200/JCO.1997.15.8.2792
   Gajjar A, 2006, LANCET ONCOL, V7, P813, DOI 10.1016/S1470-2045(06)70867-1
   Gajjar A, 2013, CLIN CANCER RES, V19, P6305, DOI 10.1158/1078-0432.CCR-13-1425
   Gajjar AJ, 2014, NAT REV CLIN ONCOL, V11, P714, DOI 10.1038/nrclinonc.2014.181
   Gardner SL, 2008, PEDIATR BLOOD CANCER, V51, P235, DOI 10.1002/pbc.21578
   Gatta G, 2014, LANCET ONCOL, V15, P35, DOI 10.1016/S1470-2045(13)70548-5
   Gerber NU, 2014, INT J RADIAT ONCOL, V89, P863, DOI 10.1016/j.ijrobp.2014.04.017
   Geyer JR, 2005, J CLIN ONCOL, V23, P7621, DOI 10.1200/JCO.2005.09.095
   Gibson P, 2010, NATURE, V468, P1095, DOI 10.1038/nature09587
   Gilbert MR, 2014, NEW ENGL J MED, V370, P699, DOI 10.1056/NEJMoa1308573
   Gnekow AK, 2012, NEURO-ONCOLOGY, V14, P1265, DOI 10.1093/neuonc/nos202
   Gottardo NG, 2014, ACTA NEUROPATHOL, V127, P189, DOI 10.1007/s00401-013-1213-7
   Grundy RG, 2010, EUR J CANCER, V46, P120, DOI 10.1016/j.ejca.2009.09.013
   Gururangan S, 2014, NEURO-ONCOLOGY, V16, P310, DOI 10.1093/neuonc/not154
   Hawkins C, 2011, CLIN CANCER RES, V17, P4790, DOI 10.1158/1078-0432.CCR-11-0034
   Hoffman LM, 2014, ACTA NEUROPATHOL, V127, P731, DOI 10.1007/s00401-013-1212-8
   Jacob K, 2009, BRIT J CANCER, V101, P722, DOI 10.1038/sj.bjc.6605179
   Johnson RA, 2010, NATURE, V466, P632, DOI 10.1038/nature09173
   Jones DTW, 2009, ONCOGENE, V28, P2119, DOI 10.1038/onc.2009.73
   Jones DTW, 2012, NATURE, V488, P100, DOI 10.1038/nature11284
   Karajannis MA, 2014, NEURO-ONCOLOGY, V16, P1408, DOI 10.1093/neuonc/nou059
   Kieran MW, 2013, PEDIAT BLOOD CANC SI, V60
   Kim J, 2013, CANCER CELL, V23, P23, DOI 10.1016/j.ccr.2012.11.017
   Kool M, 2014, CANCER CELL, V25, P393, DOI 10.1016/j.ccr.2014.02.004
   Kool M, 2012, MEMO-MAG EUR MED ONC, V5, P228, DOI 10.1007/s12254-012-0037-9
   Kool M, 2012, ACTA NEUROPATHOL, V123, P473, DOI 10.1007/s00401-012-0958-8
   Korshunov A, 2015, ACTA NEUROPATHOL, V129, P669, DOI 10.1007/s00401-015-1405-4
   Korshunov A, 2014, ACTA NEUROPATHOL, V128, P279, DOI 10.1007/s00401-013-1228-0
   Korshunov A, 2012, ACTA NEUROPATHOL, V124, P875, DOI 10.1007/s00401-012-1068-3
   Korshunov A, 2009, ACTA NEUROPATHOL, V118, P401, DOI 10.1007/s00401-009-0550-z
   Krueger DA, 2013, NEUROLOGY, V80, P574, DOI 10.1212/WNL.0b013e3182815428
   Lafay-Cousin L, 2012, EUR J CANCER, V48, P353, DOI 10.1016/j.ejca.2011.09.005
   Li MH, 2009, CANCER CELL, V16, P533, DOI 10.1016/j.ccr.2009.10.025
   Lin A, 2012, J NEUROPATH EXP NEUR, V71, P66, DOI 10.1097/NEN.0b013e31823f2cb0
   Louis DN, 2007, ACTA NEUROPATHOL, V114, P547, DOI 10.1007/s00401-007-0278-6
   Mack SC, 2014, NATURE, V506, P445, DOI 10.1038/nature13108
   McGuire CS, 2009, J NEUROSURG, V110, P725, DOI 10.3171/2008.9.JNS08117
   Merchant TE, 2009, J CLIN ONCOL, V27, P3598, DOI 10.1200/JCO.2008.20.9494
   Morfouace M, 2014, CANCER CELL, V25, P516, DOI 10.1016/j.ccr.2014.02.009
   Muller K, 2013, STRAHLENTHER ONKOL, V189, P647, DOI 10.1007/s00066-013-0357-7
   Narayana A, 2010, NEURO-ONCOLOGY, V12, P985, DOI 10.1093/neuonc/noq033
   Northcott PA, 2014, NATURE, V511, P428, DOI 10.1038/nature13379
   Northcott PA, 2012, NAT REV CANCER, V12, P818, DOI 10.1038/nrc3410
   Northcott PA, 2012, NATURE, V488, P49, DOI 10.1038/nature11327
   Northcott PA, 2010, NEUROSURG FOCUS, V28, DOI 10.3171/2009.10.FOCUS09218
   Northcott PA, 2009, NAT GENET, V41, P465, DOI 10.1038/ng.336
   Packer RJ, 1999, J CLIN ONCOL, V17, P2127, DOI 10.1200/JCO.1999.17.7.2127
   PACKER RJ, 1993, J CLIN ONCOL, V11, P850, DOI 10.1200/JCO.1993.11.5.850
   Packer RJ, 2006, J CLIN ONCOL, V24, P4202, DOI 10.1200/JCO.2006.06.4980
   Pajtler KW, 2015, CANCER CELL, V27, P728, DOI 10.1016/j.ccell.2015.04.002
   Parker M, 2014, NATURE, V506, P451, DOI 10.1038/nature13109
   Perreault S, 2013, J NEURO-ONCOL, V115, P209, DOI 10.1007/s11060-013-1213-4
   Pfister S, 2008, J CLIN INVEST, V118, P1739, DOI 10.1172/JCI33656
   Pfister SM, 2010, ACTA NEUROPATHOL, V120, P553, DOI 10.1007/s00401-010-0751-5
   Picard D, 2012, LANCET ONCOL, V13, P838, DOI 10.1016/S1470-2045(12)70257-7
   Pomeroy SL, 2002, NATURE, V415, P436, DOI 10.1038/415436a
   Pugh TJ, 2012, NATURE, V488, P106, DOI 10.1038/nature11329
   Pui CH, 2011, NAT REV CLIN ONCOL, V8, P540, DOI 10.1038/nrclinonc.2011.95
   Reddy AT, 2000, CANCER, V88, P2189, DOI 10.1002/(SICI)1097-0142(20000501)88:9<2189::AID-CNCR27>3.0.CO;2-G
   Robinson G, 2012, NATURE, V488, P43, DOI 10.1038/nature11213
   Robinson GW, 2014, BMC CANCER, V14, DOI 10.1186/1471-2407-14-258
   Rodriguez EF, 2011, ACTA NEUROPATHOL, V121, P407, DOI 10.1007/s00401-010-0784-9
   RORKE LB, 1995, J NEURO-ONCOL, V24, P21, DOI 10.1007/BF01052653
   Rudin CM, 2009, NEW ENGL J MED, V361, P1173, DOI 10.1056/NEJMoa0902903
   Rutkowski S, 2005, NEW ENGL J MED, V352, P978, DOI 10.1056/NEJMoa042176
   Rutkowski S, 2010, J CLIN ONCOL, V28, P4961, DOI 10.1200/JCO.2010.30.2299
   Schindler G, 2011, ACTA NEUROPATHOL, V121, P397, DOI 10.1007/s00401-011-0802-6
   Schwartzentruber J, 2012, NATURE, V482, P226, DOI [10.1038/nature10833, 10.1007/s13311-012-0137-6]
   Shih DJH, 2014, J CLIN ONCOL, V32, P886, DOI 10.1200/JCO.2013.50.9539
   Sievert AJ, 2013, P NATL ACAD SCI USA, V110, P5957, DOI 10.1073/pnas.1219232110
   Sievert AJ, 2009, BRAIN PATHOL, V19, P449, DOI 10.1111/j.1750-3639.2008.00225.x
   Slavc I, 2014, CANC MED, V3, P91, DOI DOI 10.1002/CAM4.161
   Spence T, 2014, NEURO-ONCOLOGY, V16, P62, DOI 10.1093/neuonc/not162
   Stupp R, 2005, NEW ENGL J MED, V352, P987, DOI 10.1056/NEJMoa043330
   Sturm D, 2014, NAT REV CANCER, V14, P92, DOI 10.1038/nrc3655
   Sturm D, 2012, CANCER CELL, V22, P425, DOI 10.1016/j.ccr.2012.08.024
   Tang YJ, 2014, NAT MED, V20, P732, DOI 10.1038/nm.3613
   Tarbell NJ, 2013, J CLIN ONCOL, V31, P2936, DOI 10.1200/JCO.2012.43.9984
   Taylor KR, 2014, NAT GENET, V46, P457, DOI 10.1038/ng.2925
   Taylor MD, 2012, ACTA NEUROPATHOL, V123, P465, DOI 10.1007/s00401-011-0922-z
   Taylor RE, 2003, J CLIN ONCOL, V21, P1581, DOI 10.1200/JCO.2003.05.116
   Tekautz TM, 2005, J CLIN ONCOL, V23, P1491, DOI 10.1200/JCO.2005.05.187
   von Hoff K, 2011, PEDIATR BLOOD CANCER, V57, P978, DOI 10.1002/pbc.23236
   Wani K, 2012, ACTA NEUROPATHOL, V123, P727, DOI 10.1007/s00401-012-0941-4
   Wisoff JH, 2011, NEUROSURGERY, V68, P1548, DOI 10.1227/NEU.0b013e318214a66e
   Witt H, 2011, CANCER CELL, V20, P143, DOI 10.1016/j.ccr.2011.07.007
   Wolff JEA, 2010, CANCER-AM CANCER SOC, V116, P705, DOI 10.1002/cncr.24730
   Wu G, 2014, NAT GENET, V46, P444, DOI 10.1038/ng.2938
   Yalon M, 2013, PEDIATR BLOOD CANCER, V60, P71, DOI 10.1002/pbc.24142
   Zhukova N, 2013, J CLIN ONCOL, V31, P2927, DOI 10.1200/JCO.2012.48.5052
NR 119
TC 137
Z9 141
U1 0
U2 22
PU LIPPINCOTT WILLIAMS & WILKINS
PI PHILADELPHIA
PA TWO COMMERCE SQ, 2001 MARKET ST, PHILADELPHIA, PA 19103 USA
SN 0732-183X
EI 1527-7755
J9 J CLIN ONCOL
JI J. Clin. Oncol.
PD SEP 20
PY 2015
VL 33
IS 27
BP 2986
EP U83
DI 10.1200/JCO.2014.59.9217
PG 15
WC Oncology
WE Science Citation Index Expanded (SCI-EXPANDED)
SC Oncology
GA CX9IL
UT WOS:000366018800006
PM 26304884
OA Green Published
DA 2023-08-27
ER

PT J
AU Pongtanakul, B
   Sirachainan, N
   Surapolchai, P
   Charoenkwan, P
   Choeyprasert, W
   Komwilaisak, P
   Chainansamit, SO
   Witsanuyothin, N
   Lertrakul, Y
   Rujkijyanont, P
   Kanjanapongkul, S
   Sosothikul, D
   Sanpakit, K
   Chotsampancharoen, T
   Wangruangsathit, SP
   Thokanit, NS
   Tabjareon, S
   Hongeng, S
AF Pongtanakul, Bunchoo
   Sirachainan, Nongnuch
   Surapolchai, Pacharapan
   Charoenkwan, Pimlak
   Choeyprasert, Worawut
   Komwilaisak, Patcharee
   Chainansamit, Su-on
   Witsanuyothin, Nittaya
   Lertrakul, Yujinda
   Rujkijyanont, Piya
   Kanjanapongkul, Somjai
   Sosothikul, Darintr
   Sanpakit, Kleebsabai
   Chotsampancharoen, Thirachit
   Wangruangsathit, Som Porn
   Thokanit, Nintita Sripaiboonkij
   Tabjareon, Sommaphun
   Hongeng, Suradej
TI Pediatric primary central nervous system tumors registry in Thailand
   under National Health Security Office schemes
SO JOURNAL OF NEURO-ONCOLOGY
LA English
DT Article
DE Asia; CNS tumor; Developing country; Pediatrics; Registry; Thailand
ID PRIMARY INTRACRANIAL TUMORS; BRAIN-TUMORS; CHILDHOOD-CANCER;
   UNITED-STATES; SURVIVAL; TAIWAN
AB Background Few epidemiological studies of pediatric central nervous system (CNS) tumors have been performed using data from Southeast Asian national registries. Therefore, we aimed to examine data on CNS tumors from the first national childhood CNS tumor registry in Thailand. Methods Newly diagnosed children with benign and malignant primary CNS tumors from 20 nationwide hospitals were included. Two eras in the Thai registry were studied to compare national protocol effectiveness, including 2003-2005 (before establishment of a pediatric CNS tumor protocol) and 2011-2012 (post-establishment). Results The first study period had 300 patients with an incidence of 7.5/1,000,000 person-years and the second had 168 patients with an incidence of 13.24/1,000,000 person-years. The three most common tumors were gliomas, medulloblastoma/primitive neuroectodermal tumor (PNET), and germ cell tumors. The most common tumor site was the cerebellum, followed by the brainstem and pineal region. Five- and 10-year overall survival (OS) rates were 46.62% (95% confidence interval [CI] 40.85-52.18) and 41.78% (95% CI 36.11-47.34), respectively, for the first period. The second period had a 5-year OS of 64.75% (95% CI 56.70-71.68). OS rates for gliomas, germ cell tumors, medulloblastoma/PNET, and ependymomas were better in the second period than in the first period. Conclusions The incidence of primary childhood CNS tumors in our study is lower compared with other reports. Improvement of OS in the second study period might be because of establishment of the Thai Pediatric Oncology Group, and national protocols for childhood CNS tumors.
C1 [Pongtanakul, Bunchoo; Sanpakit, Kleebsabai; Tabjareon, Sommaphun] Mahidol Univ, Siriraj Hosp, Fac Med, Dept Pediat, Bangkok 10700, Thailand.
   [Sirachainan, Nongnuch; Hongeng, Suradej] Mahidol Univ, Ramathibodi Hosp, Fac Med, Dept Pediat, Bangkok 10400, Thailand.
   [Surapolchai, Pacharapan] Thammasat Univ, Fac Med, Dept Pediat, Pathum Thani 12120, Thailand.
   [Charoenkwan, Pimlak; Choeyprasert, Worawut] Chiang Mai Univ, Fac Med, Dept Pediat, Chiang Mai 50200, Thailand.
   [Komwilaisak, Patcharee] Khon Kaen Univ, Fac Med, Dept Pediat, Khon Kaen 40002, Thailand.
   [Chainansamit, Su-on] Khon Kaen Hosp, Dept Pediat, Khon Kaen 40000, Thailand.
   [Witsanuyothin, Nittaya] Nakhon Ratchasima Hosp, Dept Pediat, Nakhon Ratchasima 30000, Thailand.
   [Lertrakul, Yujinda] Sappasitthiprasong Hosp, Dept Pediat, Ubon Ratchathani 34000, Thailand.
   [Rujkijyanont, Piya] Phramongkutklao Hosp, Dept Pediat, Bangkok 10400, Thailand.
   [Kanjanapongkul, Somjai] Queen Sirikit Natl Inst Child Hlth, Dept Pediat, Bangkok 10400, Thailand.
   [Sosothikul, Darintr] Chulalongkorn Univ, King Chulalongkorn Mem Hosp, Fac Med, Dept Pediat, Bangkok 10330, Thailand.
   [Chotsampancharoen, Thirachit] Prince Songkla Univ, Fac Med, Dept Pediat, Hat Yai 90110, Songkhla, Thailand.
   [Wangruangsathit, Som Porn] Buddhachinaraj Phitsanulok Hosp, Dept Pediat, Phitsanulok 65000, Thailand.
   [Thokanit, Nintita Sripaiboonkij] Mahidol Univ, Ramathibodi Hosp, Fac Med, Ramathibodi Comprehens Canc Ctr, Bangkok 10400, Thailand.
C3 Mahidol University; Mahidol University; Thammasat University; Chiang Mai
   University; Khon Kaen University; Phramongkutklao Hospital;
   Chulalongkorn University; Prince of Songkla University; Mahidol
   University
RP Hongeng, S (通讯作者)，Mahidol Univ, Ramathibodi Hosp, Fac Med, Dept Pediat, Bangkok 10400, Thailand.
EM
RI Charoenkwan, Pimlak/AGL-9582-2022; Sanpakit, Kleebsabai/ISA-0780-2023;
   Charoenkwan, Pimlak/AAP-5905-2021
OI Charoenkwan, Pimlak/0000-0002-9123-9177; Chotsampancharoen,
   Thirachit/0000-0002-0111-6318; Rujkijyanont, Piya/0000-0002-5006-9810;
   Pongtanakul, Bunchoo/0000-0003-4204-0118
FU Ramathibodi Foundation; Children Cancer Fund under The Patronage of HRH
   Princess Soamsawali; My Child Matters Grant Award UICC; National health
   security office of Thailand
FX This study was financial supported by Ramathibodi Foundation, Children
   Cancer Fund under The Patronage of HRH Princess Soamsawali and My Child
   Matters Grant Award UICC awarded to Dr. Suradej Hongeng, and National
   health security office of Thailand. We thank Ellen Knapp, PhD, from
   Edanz Group (https ://www.edanz editing.com/ac) for editing a draft of
   this manuscript.
CR Ahmed N, 2007, ASIAN PAC J CANCER P, V8, P399
   Azad TD, 2015, WORLD NEUROSURG, V84, P1832, DOI 10.1016/j.wneu.2015.07.074
   Ba-Saddik IA, 2013, CANCER EPIDEMIOL, V37, P803, DOI 10.1016/j.canep.2013.10.001
   Elhassan MMA, 2019, CHILD NERV SYST, V35, P437, DOI 10.1007/s00381-018-04032-9
   Ezzat S, 2016, J NEURO-ONCOL, V126, P371, DOI 10.1007/s11060-015-1979-7
   Florencia M, 2015, CANCER EPIDEMIOL, V39, P505, DOI 10.1016/j.canep.2015.04.010
   Gittleman HR, 2015, CANCER-AM CANCER SOC, V121, P102, DOI 10.1002/cncr.29015
   Howlader N, 2016, SEER CANC STAT REV 1
   Kang JM, 2019, CANCER EPIDEM BIOMAR, V28, P409, DOI 10.1158/1055-9965.EPI-18-0634
   Kong ZR, 2018, J CHILD NEUROL, V33, P610, DOI 10.1177/0883073818772470
   Kruchko C, 2019, NEURO-ONCOL PRACT, V6, P330, DOI 10.1093/nop/npz029
   Liu YL, 2015, CANCER EPIDEMIOL, V39, P21, DOI 10.1016/j.canep.2014.11.010
   Louis DN, 2016, ACTA NEUROPATHOL, V131, P803, DOI 10.1007/s00401-016-1545-1
   Louis DN., 2016, WHO CLASSIFICATION T
   Makino K, 2010, CHILD NERV SYST, V26, P1029, DOI 10.1007/s00381-010-1126-x
   Moore MA, 2013, ASIAN PAC J CANCER P, V14, P4461, DOI 10.7314/APJCP.2013.14.8.4461
   Nomura E, 2011, JPN J CLIN ONCOL, V41, P291, DOI 10.1093/jjco/hyq204
   Ostrom Quinn T, 2019, Neuro Oncol, V21, pv1, DOI 10.1093/neuonc/noz150
   Pouchieu C, 2018, BMC CANCER, V18, DOI 10.1186/s12885-018-4545-9
   Riaz Q, 2019, CHILD NERV SYST, V35, P2347, DOI 10.1007/s00381-019-04260-7
   Rivas-Vilela S, 2019, CLIN TRANSL ONCOL, V21, P1177, DOI 10.1007/s12094-019-02043-9
   Steliarova-Foucher E, 2017, LANCET ONCOL, V18, P719, DOI 10.1016/S1470-2045(17)30186-9
   Stiller CA, 2019, BMC CANCER, V19, DOI 10.1186/s12885-019-5344-7
   Suresh SG, 2017, J PEDIATR NEUROSCI, V12, P237, DOI 10.4103/jpn.JPN_31_17
   Villano JL, 2010, NEURO-ONCOLOGY, V12, P257, DOI 10.1093/neuonc/nop029
   Wiangnon S, 2011, ASIAN PAC J CANCER P, V12, P2215
   Wong TT, 2005, CANCER, V104, P2156, DOI 10.1002/cncr.21430
NR 27
TC 5
Z9 6
U1 0
U2 6
PU SPRINGER
PI NEW YORK
PA ONE NEW YORK PLAZA, SUITE 4600, NEW YORK, NY, UNITED STATES
SN 0167-594X
EI 1573-7373
J9 J NEURO-ONCOL
JI J. Neuro-Oncol.
PD AUG
PY 2020
VL 149
IS 1
BP 141
EP 151
DI 10.1007/s11060-020-03582-w
EA JUL 2020
PG 11
WC Oncology; Clinical Neurology
WE Science Citation Index Expanded (SCI-EXPANDED)
SC Oncology; Neurosciences & Neurology
GA NF3CC
UT WOS:000552153800001
PM 32710301
DA 2023-08-27
ER

PT J
AU Guggilam, A
   Cardinale, JP
   Mariappan, N
   Sriramula, S
   Haque, M
   Francis, J
AF Guggilam, Anuradha
   Cardinale, Jeffrey P.
   Mariappan, Nithya
   Sriramula, Srinivas
   Haque, Masudul
   Francis, Joseph
TI Central TNF inhibition results in attenuated neurohumoral excitation in
   heart failure: a role for superoxide and nitric oxide
SO BASIC RESEARCH IN CARDIOLOGY
LA English
DT Article
DE Tumor necrosis factor; Congestive heart failure; Paraventricular
   nucleus; Rostral ventrolateral medullary nucleus; Sympathoexcitation;
   Oxidant stress
ID SYMPATHETIC-NERVE ACTIVITY; NECROSIS-FACTOR-ALPHA; BLOOD-BRAIN-BARRIER;
   ROSTRAL VENTROLATERAL MEDULLA; LEFT-VENTRICULAR DYSFUNCTION;
   II-DEPENDENT STIMULATION; ANGIOTENSIN-II; MYOCARDIAL-INFARCTION;
   OXIDATIVE STRESS; PARAVENTRICULAR NUCLEUS
AB This study examined the effect of central tumor necrosis factor-alpha (TNF) blockade on the imbalance between nitric oxide and superoxide production in the paraventricular nucleus (PVN) and ventrolateral medulla (VLM), key autonomic regulators, and their contribution to enhanced sympathetic drive in mice with congestive heart failure (CHF). We also used a TNF gene knockout (KO) mouse model to study the involvement of TNF in body fluid homeostasis and sympathoexcitation in CHF. After implantation of intracerebroventricular (ICV) cannulae, myocardial infarction (MI) was induced in wild-type (WT) and KO mice by coronary artery ligation. Osmotic mini-pumps were implanted into one set of WT + MI/Sham mice for continuous ICV infusion of Etanercept (ETN), a TNF receptor fusion protein, or vehicle (VEH). Gene expressions of neuronal nitric oxide synthase (NOS) and angiotensin receptor-type 2 were reduced, while those of inducible NOS, Nox2 homologs, superoxide, peroxynitrite and angiotensin receptor-type 1 were elevated in the brainstem and hypothalamus of MI + VEH. Plasma norepinephrine levels and the number of Fos-positive neurons were also increased in the PVN and VLM in MI + VEH. MI + ETN and KO + MI mice exhibited reduced oxidative stress, reduced sympathoexcitation and an improved cardiac function. These changes in WT + MI were associated with increased sodium and fluid retention. These results indicate that elevated TNF in these autonomic regulatory regions of the brain alter the production of superoxide and nitric oxide, contributing to fluid imbalance and sympathoexcitation in CHF.
C1 [Guggilam, Anuradha; Cardinale, Jeffrey P.; Mariappan, Nithya; Sriramula, Srinivas; Haque, Masudul; Francis, Joseph] Louisiana State Univ, Sch Vet Med, Baton Rouge, LA 70803 USA.
C3 Louisiana State University School of Veterinary Medicine; Louisiana
   State University System; Louisiana State University
RP Francis, J (通讯作者)，Louisiana State Univ, Sch Vet Med, 1909 Skip Bertman Dr, Baton Rouge, LA 70803 USA.
EM
RI Guggilam, Anuradha/C-8875-2009; Sriramula, Srinivas/N-1297-2019
OI Sriramula, Srinivas/0000-0001-9937-1589
FU National Heart, Lung, and Blood Institute [1RO1 HL-080544-01]
FX This work was supported by National Heart, Lung, and Blood Institute
   Grant (1RO1 HL-080544-01 to J.F.). The authors acknowledge the expert
   technical assistance of Elizabeth McIlwain, Philip J. Ebenezer, Michael
   Broussard and Sherry Ring.
CR Banks WA, 2005, CURR PHARM DESIGN, V11, P973, DOI 10.2174/1381612053381684
   Bozkurt B, 1998, CIRCULATION, V97, P1382, DOI 10.1161/01.CIR.97.14.1382
   Campese VM, 2005, HYPERTENSION, V46, P533, DOI 10.1161/01.HYP.0000179088.57586.26
   Campese VM, 2002, HYPERTENSION, V39, P519, DOI 10.1161/hy0202.102815
   Canton M, 2006, EUR HEART J, V27, P875, DOI 10.1093/eurheartj/ehi751
   Chan JYH, 2002, HYPERTENSION, V40, P335, DOI 10.1161/01.HYP.0000029241.33421.EB
   Chappell D, 2009, BASIC RES CARDIOL, V104, P78, DOI 10.1007/s00395-008-0749-5
   Chorianopoulos E, 2010, BASIC RES CARDIOL, V105, P301, DOI 10.1007/s00395-009-0046-y
   Dikalov SI, 2002, ARCH BIOCHEM BIOPHYS, V402, P218, DOI 10.1016/S0003-9861(02)00064-4
   Fildes JE, 2009, J AM COLL CARDIOL, V53, P1013, DOI 10.1016/j.jacc.2008.11.046
   Fleegal-DeMotta MA, 2009, J CEREBR BLOOD F MET, V29, P640, DOI 10.1038/jcbfm.2008.158
   Francis J, 2001, AM J PHYSIOL-REG I, V281, pR1734, DOI 10.1152/ajpregu.2001.281.5.R1734
   Galiano M, 2001, EUR J NEUROSCI, V14, P327, DOI 10.1046/j.0953-816x.2001.01647.x
   Gao L, 2005, AM J PHYSIOL-HEART C, V288, pH2271, DOI 10.1152/ajpheart.00949.2004
   Gloor SM, 2001, BRAIN RES REV, V36, P258, DOI 10.1016/S0165-0173(01)00102-3
   Guggilam A, 2008, EUR J HEART FAIL, V10, P625, DOI 10.1016/j.ejheart.2008.05.004
   Guggilam A, 2007, AM J PHYSIOL-HEART C, V293, pH599, DOI 10.1152/ajpheart.00286.2007
   Han Y, 2007, EUR J HEART FAIL, V9, P967, DOI 10.1016/j.ejheart.2007.07.004
   Hill JA, 2000, CIRCULATION, V101, P2863, DOI 10.1161/01.CIR.101.24.2863
   Hu K, 2001, BASIC RES CARDIOL, V96, P258, DOI 10.1007/s003950170056
   Huang CH, 2009, BASIC RES CARDIOL, V104, P228, DOI 10.1007/s00395-008-0765-5
   Johnson AK, 1997, FRONT NEUROENDOCRIN, V18, P292, DOI 10.1006/frne.1997.0153
   Khaleduzzaman M, 2007, J VIROL, V81, P12307, DOI 10.1128/JVI.01002-07
   Kleinbongard P, 2010, PHARMACOL THERAPEUT, V127, P295, DOI 10.1016/j.pharmthera.2010.05.002
   Krown KA, 1996, J CLIN INVEST, V98, P2854, DOI 10.1172/JCI119114
   Lacerda L, 2010, BASIC RES CARDIOL, V105, P751, DOI 10.1007/s00395-010-0113-4
   Li SM, 2010, BASIC RES CARDIOL, V105, P139, DOI 10.1007/s00395-009-0067-6
   Li YF, 2006, AM J PHYSIOL-REG I, V290, pR1035, DOI 10.1152/ajpregu.00338.2004
   Lindley TE, 2004, CIRC RES, V94, P402, DOI 10.1161/01.RES.0000112964.40701.93
   Liu JL, 1999, CIRC RES, V84, P417, DOI 10.1161/01.RES.84.4.417
   Mann DM, 2004, CIRCULATION, V109, pE126
   Mayorov DN, 2004, HYPERTENSION, V44, P101, DOI 10.1161/01.HYP.0000131290.12255.04
   McCann SM, 2000, ANN NY ACAD SCI, V917, P4, DOI 10.1111/j.1749-6632.2000.tb05368.x
   RAIVICH G, 1994, NEUROPATH APPL NEURO, V20, P209
   SAKUMA I, 1992, CIRC RES, V70, P607, DOI 10.1161/01.RES.70.3.607
   SASAKI S, 1990, HYPERTENSION, V15, P274, DOI 10.1161/01.HYP.15.3.274
   SCHILLER NB, 1979, CIRCULATION, V60, P547, DOI 10.1161/01.CIR.60.3.547
   Sheng JG, 1998, ACTA NEUROPATHOL, V95, P229, DOI 10.1007/s004010050792
   SHI P, HYPERTENSION, V56, P297
   Thielmann M, 2002, CIRC RES, V90, P807, DOI 10.1161/01.RES.0000014451.75415.36
   TorreAmione G, 1996, J AM COLL CARDIOL, V27, P1201, DOI 10.1016/0735-1097(95)00589-7
   Tsutamoto T, 1998, J AM COLL CARDIOL, V31, P391, DOI 10.1016/S0735-1097(97)00494-4
   UNGER T, 1988, CIRCULATION, V77, P40
   Vahid-Ansari F, 1998, AM J PHYSIOL-HEART C, V275, pH2140, DOI 10.1152/ajpheart.1998.275.6.H2140
   Veerasingham SJ, 1997, BRAIN RES, V749, P157, DOI 10.1016/S0006-8993(96)01381-9
   Wei SG, 2009, J HYPERTENS, V27, P543, DOI 10.1097/HJH.0b013e3283200442
   Yu Y, 2010, HYPERTENSION, V55, P652, DOI 10.1161/HYPERTENSIONAHA.109.142836
   ZANZINGER J, 1995, AM J PHYSIOL-REG I, V268, pR958, DOI 10.1152/ajpregu.1995.268.4.R958
   Zanzinger J, 2002, AUTON NEUROSCI-BASIC, V98, P24, DOI 10.1016/S1566-0702(02)00025-5
   Zhang CH, 2010, BASIC RES CARDIOL, V105, P453, DOI 10.1007/s00395-010-0083-6
   Zhang M, 2010, NEUROSCIENCE, V171, P852, DOI 10.1016/j.neuroscience.2010.09.029
   Zimmerman MC, 2004, CIRC RES, V95, P210, DOI 10.1161/01.RES.0000135483.12297.e4
   Zimmerman MC, 2002, CIRC RES, V91, P1038, DOI 10.1161/01.RES.0000043501.47934.FA
   Zucker IH, 2004, PROG BIOPHYS MOL BIO, V84, P217, DOI 10.1016/j.pbiomolbio.2003.11.010
   Zucker Irving H, 2002, Heart Fail Monit, V3, P2
NR 55
TC 48
Z9 54
U1 0
U2 14
PU SPRINGER HEIDELBERG
PI HEIDELBERG
PA TIERGARTENSTRASSE 17, D-69121 HEIDELBERG, GERMANY
SN 0300-8428
EI 1435-1803
J9 BASIC RES CARDIOL
JI Basic Res. Cardiol.
PD MAR
PY 2011
VL 106
IS 2
BP 273
EP 286
DI 10.1007/s00395-010-0146-8
PG 14
WC Cardiac & Cardiovascular Systems
WE Science Citation Index Expanded (SCI-EXPANDED)
SC Cardiovascular System & Cardiology
GA 715ZK
UT WOS:000286934300009
PM 21246206
OA Green Accepted
DA 2023-08-27
ER

PT J
AU Bredel, M
   Pollack, IF
   Hamilton, RL
   James, CD
AF Bredel, M
   Pollack, IF
   Hamilton, RL
   James, CD
TI Epidermal growth factor receptor expression and gene amplification in
   high-grade non-brainstem gliomas of childhood
SO CLINICAL CANCER RESEARCH
LA English
DT Article
ID HUMAN GLIOBLASTOMA-MULTIFORME; MALIGNANT GLIOMAS; EGF RECEPTOR;
   PROGNOSTIC RELEVANCE; MONOCLONAL-ANTIBODY; FACTOR-ALPHA; CELL-LINES;
   TUMORS; ASTROCYTOMA; CHILDREN
AB Epidermal growth factor receptor (EGFR) is commonly overexpressed in adult high-grade gliomas, Forty to 50% of such tumors demonstrate amplification of the EGFR gene, often with rearrangement and constitutive activation of the gene product, suggesting that EGFR might play a role in the malignant progression of a subset of these neoplasms. In this regard, several groups have shown that overexpression of EGFR is associated with an adverse outcome in adult gliomas, In contrast to the extensive studies of EGFR status that have been performed in adult high-grade gliomas, little information has been reported about EGFR expression and amplification, as well as their prognostic relevance in highgrade gliomas of childhood, which carry a somewhat more favorable prognosis than their adult counterparts. To address this issue, we examined the expression of EGFR using immunohistochemistry and screened for amplification of the EGFR gene using a competitive PCR in a series of 27 archival pediatric high-grade nonbrainstem gliomas treated consecutively at our institution between 1975 and 1992, Tumors were categorized based on protein expression patterns, and the association between expression status and outcome was examined, Although elevated immunoreactivity for EGFR was observed in 80% of tumors, only two of the cases had gene amplification. No difference in outcome was observed between tumors that exhibited extensive EGFR immunoreactivity and those that did not (P > 0.3), Although EGFR expression did not seem to be of prognostic relevance for the outcome of pediatric patients harboring high-grade non-brainstem gliomas, the consistently high levels of expression of EGFR in these neoplasms suggest that this receptor plays a role in the malignant phenotype of these tumors. Accordingly, treatment approaches targeting EGFR might be of potential therapeutic benefit for high-grade gliomas of childhood.
C1 Childrens Hosp Pittsburgh, Dept Neurosurg, Pittsburgh, PA 15213 USA.
   Univ Pittsburgh, Sch Med, Dept Neurosurg, Canc Inst,Brain Tumor Ctr, Pittsburgh, PA 15213 USA.
   Univ Pittsburgh, Sch Med, Dept Pathol,Div Neuropathol, Canc Inst,Brain Tumor Ctr, Pittsburgh, PA 15213 USA.
   Mayo Clin & Mayo Fdn, Dept Neurosurg, Rochester, MN 55905 USA.
C3 Pennsylvania Commonwealth System of Higher Education (PCSHE); University
   of Pittsburgh; Pennsylvania Commonwealth System of Higher Education
   (PCSHE); University of Pittsburgh; Pennsylvania Commonwealth System of
   Higher Education (PCSHE); University of Pittsburgh; Mayo Clinic
RP Pollack, IF (通讯作者)，Childrens Hosp Pittsburgh, Dept Neurosurg, 3705 5th Ave, Pittsburgh, PA 15213 USA.
RI James, Charles D/E-2721-2012
OI James, Charles D/0000-0002-1027-203X
FU NCI NIH HHS [CA55728] Funding Source: Medline; NINDS NIH HHS [NS37704,
   NS01810] Funding Source: Medline
CR AARONSON SA, 1991, SCIENCE, V254, P1146, DOI 10.1126/science.1659742
   ALLEN JC, 1985, PEDIATR CLIN N AM, V32, P633
   BATRA SK, 1995, CELL GROWTH DIFFER, V6, P1251
   Bredel M, 1997, CLIN CANCER RES, V3, P2157
   Bredel M, 1998, NEUROSURGERY, V43, P124, DOI 10.1097/00006123-199807000-00081
   Campbell JW, 1996, NEUROSURGERY, V38, P258, DOI 10.1097/00006123-199602000-00006
   DIETRICH U, 1991, J NEUROL, V238, P221
   Duffner PK, 1996, J NEURO-ONCOL, V28, P245
   EKSTRAND AJ, 1991, CANCER RES, V51, P2164
   FINLAY JL, 1995, J CLIN ONCOL, V13, P112, DOI 10.1200/JCO.1995.13.1.112
   HE J, 1995, CANCER RES, V55, P4833
   HIESIGER EM, 1993, J NEURO-ONCOL, V16, P93, DOI 10.1007/BF01324695
   HILLS D, 1995, INT J CANCER, V63, P537, DOI 10.1002/ijc.2910630414
   HSU SM, 1981, J HISTOCHEM CYTOCHEM, V29, P577, DOI 10.1177/29.4.6166661
   HUNTER SB, 1995, J NEUROPATH EXP NEUR, V54, P57, DOI 10.1097/00005072-199501000-00007
   JAROS E, 1992, BRIT J CANCER, V66, P373, DOI 10.1038/bjc.1992.273
   KAPLAN EL, 1958, J AM STAT ASSOC, V53, P457, DOI 10.2307/2281868
   KLEIHUES P, 1990, INT HISTOLOGICAL CLA, V21, P11
   KUNWAR S, 1993, J NEUROSURG, V79, P569, DOI 10.3171/jns.1993.79.4.0569
   LANG FF, 1994, J NEUROSURG, V81, P427, DOI 10.3171/jns.1994.81.3.0427
   Laurent JP, 1985, J PEDIAT NEUROSCI, V1, P15
   LIBERMANN TA, 1985, NATURE, V313, P144, DOI 10.1038/313144a0
   Louis DN, 1997, BRAIN PATHOL, V7, P755, DOI 10.1111/j.1750-3639.1997.tb01062.x
   LUNDJOHANSEN M, 1990, CANCER RES, V50, P6039
   MERLINO GT, 1985, MOL CELL BIOL, V5, P1722, DOI 10.1128/MCB.5.7.1722
   Newcomb EW, 1998, BRAIN PATHOL, V8, P655, DOI 10.1111/j.1750-3639.1998.tb00191.x
   NISHIKAWA R, 1994, P NATL ACAD SCI USA, V91, P7727, DOI 10.1073/pnas.91.16.7727
   POLLACK IF, 1994, NEW ENGL J MED, V331, P1500, DOI 10.1056/NEJM199412013312207
   Pollack IF, 1997, CANCER RES, V57, P304
   Pollack IF, 1997, CANCER-AM CANCER SOC, V79, P849, DOI 10.1002/(SICI)1097-0142(19970215)79:4<849::AID-CNCR23>3.3.CO;2-V
   RASHEED BKA, 1994, CANCER RES, V54, P1324
   SCHOBER R, 1995, CLIN NEUROPATHOL, V14, P169
   SCHOBER R, 1991, J HISTOCHEM CYTOCHEM, V39, P741
   SPOSTO R, 1989, J NEURO-ONCOL, V7, P165, DOI 10.1007/BF00165101
   SUGAWA N, 1990, P NATL ACAD SCI USA, V87, P8602, DOI 10.1073/pnas.87.21.8602
   VONDEIMLING A, 1992, J NEUROSURG, V77, P295, DOI 10.3171/jns.1992.77.2.0295
   VONDEIMLING A, 1993, BRAIN PATHOL, V3, P19
   VONDEIMLING A, 1992, CANCER RES, V52, P2987
   Waha A, 1996, J NEUROSURG, V85, P634, DOI 10.3171/jns.1996.85.4.0634
   WERNER MH, 1988, ACTA NEUROPATHOL, V77, P196
   WESTPHAL M, 1988, CANCER LETT, V38, P283, DOI 10.1016/0304-3835(88)90020-1
   WIKSTRAND CJ, 1995, CANCER RES, V55, P3140
   WONG AJ, 1992, P NATL ACAD SCI USA, V89, P2965, DOI 10.1073/pnas.89.7.2965
   WONG AJ, 1987, P NATL ACAD SCI USA, V84, P6899, DOI 10.1073/pnas.84.19.6899
   YOUNG JL, 1975, J PEDIATR-US, V86, P254, DOI 10.1016/S0022-3476(75)80484-7
NR 45
TC 145
Z9 150
U1 0
U2 0
PU AMER ASSOC CANCER RESEARCH
PI BIRMINGHAM
PA PO BOX 11806, BIRMINGHAM, AL 35202 USA
SN 1078-0432
J9 CLIN CANCER RES
JI Clin. Cancer Res.
PD JUL
PY 1999
VL 5
IS 7
BP 1786
EP 1792
PG 7
WC Oncology
WE Science Citation Index Expanded (SCI-EXPANDED)
SC Oncology
GA 217TG
UT WOS:000081514600027
PM 10430083
DA 2023-08-27
ER

PT J
AU Wu, HW
   Wu, CH
   Lin, SC
   Wu, CC
   Chen, HH
   Chen, YW
   Lee, YY
   Chang, FC
AF Wu, Hsin-Wei
   Wu, Chia-Hung
   Lin, Shih-Chieh
   Wu, Chih-Chun
   Chen, Hsin-Hung
   Chen, Yi-Wei
   Lee, Yi-Yen
   Chang, Feng-Chi
TI MRI features of pediatric atypical teratoid rhabdoid tumors and
   medulloblastomas of the posterior fossa
SO CANCER MEDICINE
LA English
DT Article
DE atypical teratoid rhabdoid tumor (AT; RT); embryonal brain tumor;
   magnetic resonance imaging (MRI); medulloblastoma; pediatric brain tumor
ID CENTRAL-NERVOUS-SYSTEM; APPARENT DIFFUSION-COEFFICIENT; BRAIN-TUMORS;
   TERATOID/RHABDOID TUMOR; SURVIVAL OUTCOMES; SIGNAL INTENSITY;
   EPIDEMIOLOGY; SURVEILLANCE; DIFFERENTIATION; CLASSIFICATION
AB Background: Atypical teratoid rhabdoid tumor (AT/RT) occurs at a younger age and is associated with a worse prognosis than medulloblastoma; however, these two tumor entities are mostly indistinguishable on neuroimaging. The aim of our study was to differentiate AT/RT and medulloblastoma based on their clinical and MRI features to enhance treatment planning and outcome prediction.Methods: From 2005-2021, we retrospectively enrolled 16 patients with histopathologically diagnosed AT/RT and 57 patients with medulloblastoma at our institute. We evaluated their clinical data and MRI findings, including lesion signals, intratumoral morphologies, and peritumoral/distal involvement.Results: The age of children with AT/RT was younger than that of children with medulloblastoma (2.8 +/- 4.9 [0-17] vs. 6.5 +/- 4.0 [0-18], p < 0.001), and the overall survival rate was lower (21.4% vs. 66.0%, p = 0.005). Regarding lesion signals on MRI, AT/RT had a lower ADC(min) (cutoff value & LE;544.7 x 10(-6) mm(2)/s, p < 0.001), a lower ADC ratio (cutoff value <= 0.705, p < 0.001), and a higher DWI ratio (cutoff value >=;1.595, p < 0.001) than medulloblastoma. Regarding intratumoral morphology, the "tumor central vein sign " was mostly exclusive to medulloblastoma (24/57, 42.1%; AT/RT 1/16, 6.3%; p = 0.007). Regarding peritumoral invasion on T2WI, AT/RT was more prone to invasion of the brainstem (p < 0.001) and middle cerebellar peduncle (p < 0.001) than medulloblastoma.Conclusions: MRI findings of a lower ADC value, more peritumoral invasion, and absence of the "tumor central vein sign " may be helpful to differentiate AT/RT from medulloblastoma. These distinct MRI findings together with the younger age of AT/RT patients may explain the worse outcomes in AT/RT patients.
C1 [Wu, Hsin-Wei; Wu, Chia-Hung; Wu, Chih-Chun; Chang, Feng-Chi] Taipei Vet Gen Hosp, Dept Radiol, 201 Sec 2,Shih Pai Rd, Taipei 11217, Taiwan.
   [Wu, Hsin-Wei; Wu, Chia-Hung; Lin, Shih-Chieh; Wu, Chih-Chun; Chen, Hsin-Hung; Chen, Yi-Wei; Lee, Yi-Yen; Chang, Feng-Chi] Natl Yang Ming Chiao Tung Univ, Sch Med, Taipei, Taiwan.
   [Wu, Chia-Hung] Natl Yang Ming Chiao Tung Univ, Inst Clin Med, Taipei, Taiwan.
   [Lin, Shih-Chieh] Taipei Vet Gen Hosp, Dept Pathol & Lab Med, Taipei, Taiwan.
   [Chen, Hsin-Hung; Lee, Yi-Yen] Taipei Vet Gen Hosp, Neurol Inst, Dept Neurosurg, Div Pediat Neurosurg, Taipei, Taiwan.
   [Chen, Yi-Wei] Taipei Vet Gen Hosp, Dept Oncol, Taipei, Taiwan.
   [Chen, Yi-Wei] Yuanpei Univ Med Technol, Dept Med Imaging & Radiol Technol, Hsinchu, Taiwan.
C3 Taipei Veterans General Hospital; National Yang Ming Chiao Tung
   University; National Yang Ming Chiao Tung University; Taipei Veterans
   General Hospital; Taipei Veterans General Hospital; Taipei Veterans
   General Hospital
RP Chang, FC (通讯作者)，Taipei Vet Gen Hosp, Dept Radiol, 201 Sec 2,Shih Pai Rd, Taipei 11217, Taiwan.
EM
RI 張, 豐基/IXN-6983-2023
OI Chang, Feng-Chi/0000-0001-7267-6497; Wu, Chia-Hung/0000-0003-1723-4797
FU Ministry of Science and Technology, Taiwan [MOST 109-2314-B-075-036,
   MOST 110-2314-B-075-005, MOST 110-2314-B-075-032, MOST
   111-2314-B-075-025-MY3]; Professor Tsuen CHANG's Scholarship Program
   from Medical Scholarship Foundation In Memory Of Professor Albert
   Ly-Young Shen; Taipei Veterans General Hospital [V110C-037, V111B-032,
   V111C-028, V112B-007, V112C-059, V112D67-002-MY3-1]; Veterans General
   Hospitals and University System of Taiwan Joint Research Program [VGHUST
   109V1-5-2, VGHUST 110-G1-5-2]; Vivian W. Yen Neurological Foundation;
   Yen Tjing Ling Medical Foundation [CI-109-3, CI-111-2, CI-112-2]
FX Ministry of Science and Technology, Taiwan, Grant/Award Number: MOST
   109-2314-B-075-036, MOST 110-2314-B-075-005, MOST 110-2314-B-075-032 and
   MOST 111-2314-B-075-025-MY3; Professor Tsuen CHANG's Scholarship Program
   from Medical Scholarship Foundation In Memory Of Professor Albert
   Ly-Young Shen; Taipei Veterans General Hospital, Grant/Award Number:
   V110C-037, V111B-032, V111C-028, V112B-007, V112C-059 and
   V112D67-002-MY3-1; Veterans General Hospitals and University System of
   Taiwan Joint Research Program, Grant/Award Number: VGHUST 109V1-5-2 and
   VGHUST 110-G1-5-2; Vivian W. Yen Neurological Foundation; Yen Tjing Ling
   Medical Foundation, Grant/Award Number: CI-109-3, CI-111-2 and CI-112-2
CR Abo-bakrkhattabb OM., 2018, EGYPTIAN J HOSP MED, V73, P6090, DOI [10.21608/ejhm.2018.12521, DOI 10.21608/EJHM.2018.12521]
   Baliga S, 2021, PEDIATR BLOOD CANCER, V68, DOI 10.1002/pbc.28395
   Bishop AJ, 2012, INT J RADIAT ONCOL, V82, P341, DOI 10.1016/j.ijrobp.2010.08.020
   BRADLEY WG, 1993, RADIOLOGY, V189, P15, DOI 10.1148/radiology.189.1.8372185
   Buscariollo DL, 2012, CANCER-AM CANCER SOC, V118, P4212, DOI 10.1002/cncr.27373
   Cohen AR, 2022, NEW ENGL J MED, V386, P1922, DOI 10.1056/NEJMra2116344
   Docampo J, 2013, NEURORADIOL J, V26, P493, DOI 10.1177/197140091302600501
   Eran A, 2010, PEDIATR RADIOL, V40, P1254, DOI 10.1007/s00247-009-1429-9
   Fischer-Valuck BW, 2017, CANCER-AM CANCER SOC, V123, P682, DOI 10.1002/cncr.30405
   Fossey M, 2017, J NEURO-ONCOL, V132, P155, DOI 10.1007/s11060-016-2353-0
   Gajjar A, 2021, J CLIN ONCOL, V39, P822, DOI 10.1200/JCO.20.01372
   Gauvain KM, 2001, AM J ROENTGENOL, V177, P449, DOI 10.2214/ajr.177.2.1770449
   Goo HW, 2017, KOREAN J RADIOL, V18, P194, DOI 10.3348/kjr.2017.18.1.194
   HENKELMAN RM, 1991, RADIOLOGY, V179, P199, DOI 10.1148/radiology.179.1.1848714
   Jin B, 2013, PEDIATR RADIOL, V43, P1001, DOI 10.1007/s00247-013-2646-9
   Kline CN, 2017, NEURO-ONCOL PRACT, V4, P138, DOI 10.1093/nop/npx011
   Koral K, 2008, AM J ROENTGENOL, V190, P809, DOI 10.2214/AJR.07.3069
   Kram David E., 2018, Bioengineering-Basel, V5, P78, DOI 10.3390/bioengineering5040078
   Lau CSM, 2015, CANCER MANAG RES, V7, P301, DOI 10.2147/CMAR.S88561
   이정민, 2016, Investigative Magnetic Resonance Imaging, V20, P158, DOI 10.13104/imri.2016.20.3.158
   Louis DN, 2021, NEURO-ONCOLOGY, V23, P1231, DOI 10.1093/neuonc/noab106
   Louis DN, 2016, ACTA NEUROPATHOL, V131, P803, DOI 10.1007/s00401-016-1545-1
   Maaz AUR, 2021, CHILD NERV SYST, V37, P465, DOI 10.1007/s00381-020-04815-z
   Millard NE, 2016, J CHILD NEUROL, V31, P1341, DOI 10.1177/0883073815600866
   Mittal S, 2009, AM J NEURORADIOL, V30, P232, DOI 10.3174/ajnr.A1461
   Mohapatra I, 2010, NEUROPATHOLOGY, V30, P251, DOI 10.1111/j.1440-1789.2009.01075.x
   Packer RJ, 2002, J PEDIAT HEMATOL ONC, V24, P337, DOI 10.1097/00043426-200206000-00004
   Phuttharak Warinthorn, 2021, Asian Pac J Cancer Prev, V22, P1129, DOI 10.31557/APJCP.2021.22.4.1129
   Pierce TT, 2014, NEURORADIOL J, V27, P63, DOI 10.15274/NRJ-2014-10007
   Reddy AT, 2020, J CLIN ONCOL, V38, P1175, DOI 10.1200/JCO.19.01776
   Rumboldt Z, 2006, AM J NEURORADIOL, V27, P1362
   Sari Erkan, 2015, Pediatr Neurosurg, V50, P12, DOI 10.1159/000370033
   Tassanawipas A, 2005, J Orthop Surg (Hong Kong), V13, P228
   Wu CC, 2012, J CHIN MED ASSOC, V75, P581, DOI 10.1016/j.jcma.2012.08.019
   Wu Z, 2009, J MAGN RESON IMAGING, V29, P177, DOI 10.1002/jmri.21617
   Yamashita Y, 2009, NEUROL RES, V31, P940, DOI 10.1179/174313209X382520
   Zeng Q, 2019, FRONT ONCOL, V9, DOI 10.3389/fonc.2019.00424
   Zhang M, 2021, AM J NEURORADIOL, V42, P1702, DOI 10.3174/ajnr.A7200
   Zitouni S, 2017, JPN J RADIOL, V35, P448, DOI 10.1007/s11604-017-0652-9
NR 39
TC 0
Z9 0
U1 0
U2 0
PU WILEY
PI HOBOKEN
PA 111 RIVER ST, HOBOKEN 07030-5774, NJ USA
SN 2045-7634
J9 CANCER MED-US
JI Cancer Med.
PD MAY
PY 2023
VL 12
IS 9
BP 10449
EP 10461
DI 10.1002/cam4.5780
EA MAR 2023
PG 13
WC Oncology
WE Science Citation Index Expanded (SCI-EXPANDED)
SC Oncology
GA H2EW2
UT WOS:000949178900001
PM 36916326
OA gold, Green Published
DA 2023-08-27
ER

PT J
AU Pan, JJ
   Zhang, HT
   Man, FY
   Shen, YG
   Wang, YW
   Zhong, Y
   Ma, L
   Wang, HY
   Ye, HY
AF Pan, Jingjing
   Zhang, Hongtao
   Man, Fengyuan
   Shen, Yanguang
   Wang, Yingwei
   Zhong, Yan
   Ma, Lu
   Wang, Haiyi
   Ye, Huiyi
TI Measurement and scan reproducibility of parameters of intravoxel
   incoherent motion in renal tumor and normal renal parenchyma: a
   preliminary research at 3.0 T MR
SO ABDOMINAL RADIOLOGY
LA English
DT Article
DE Intravoxel incoherent motion, IVIM; Diffusion-weighted imaging, DWI;
   Magnetic resonance imaging, MRI; Renal tumors; Reproducibility
ID APPARENT-DIFFUSION-COEFFICIENT; B-VALUES; INITIAL-EXPERIENCE;
   PROSTATE-CANCER; WEIGHTED MRI; PERFUSION; CARCINOMA; LESIONS;
   DIFFERENTIATION; LIVER
AB To prospectively estimate measurement and scan reproducibility of parameters of intravoxel incoherent motion (IVIM) in renal tumors, normal renal cortex, and medulla.
   Twenty-four consecutive patients (twelve males and twelve females; median age 56.7 years, range 32-71 years) with 25 renal tumors (20 renal cell carcinomas, one urothelium carcinoma, three angiomyolipomas, and one oncocytoma) were examined twice using IVIM1 and IVIM2 with 9 and 16 b values, respectively, at 3.0 T. All the patients were re-scanned in 24-48 h. Regions of interest (ROIs) were placed in solid part of tumor, normal cortex, and medulla to derive IVIM parameters D (true diffusion coefficient), D* (pseudodiffusion coefficient), and f (perfusion fraction of pseudodiffusion). Differences in parameters between two IVIM sets and intra-observer, inter-observer, and scan-rescan differences were assessed using paired t tests. Intra-observer, inter-observer, and scan-rescan reproducibility were assessed by measuring coefficient of variation and Bland-Altman limits of agreements.
   Intra-observer reproducibility of renal tumors, normal renal cortex, and medulla was excellent for apparent diffusion coefficient (ADC; CV: 3.45%-5.34%, BA-LA: -14% to 18%) and D (CV: 3.65% to 6.04%, BA-LA: -18% to 19%), good for f (CV: 11.96%-16.08%, BA-LA: -76.4% to 92.1% except f of medulla with CV of 32.59% and BA-LA of -76.4% to 92.1% in IVIM1), and poor for D* (CV: 25.0% to 75.4%, BA-LA: -111% to 150%). The same order was in inter-observer reproducibility analysis. Scan-rescan reproducibility was the worst of the three parameters. Renal medulla showed worse reproducibility than renal tumors and the normal cortex. The metrics of IVIM2 had better reproducibility than IVIM1.
   Excellent reproducibility evaluation for ADC and D, good for f, and poor for D* derived from IVIM was performed in renal tumors, normal renal cortex, and medulla. D* has limited reliability and scan-rescan reproducibility should be improved.
C1 [Pan, Jingjing; Zhang, Hongtao; Shen, Yanguang; Wang, Yingwei; Zhong, Yan; Ma, Lu; Wang, Haiyi; Ye, Huiyi] Chinese Peoples Liberat Army Gen Hosp, Dept Radiol, 28 Fuxing Rd,Box 100853, Beijing, Peoples R China.
   [Pan, Jingjing; Man, Fengyuan] PLA Rocket Force, Gen Hosp, Dept Radiol, Beijing, Peoples R China.
   [Zhang, Hongtao] PLA, Hosp 307, Dept Radiol, Beijing, Peoples R China.
C3 Chinese People's Liberation Army General Hospital
RP Wang, HY; Ye, HY (通讯作者)，Chinese Peoples Liberat Army Gen Hosp, Dept Radiol, 28 Fuxing Rd,Box 100853, Beijing, Peoples R China.
EM
RI zhang, hongtao/IWV-4469-2023; Wang, Haiyi/ACM-8538-2022
OI Wang, Haiyi/0000-0002-0717-1748
FU National Natural Science Foundation of China [81471641]
FX This study was funded by the National Natural Science Foundation of
   China (Grant Number 81471641). The funders had no role in research
   design, data collection and analysis, decision to publish, or
   preparation of the manuscript.
CR Andreou A, 2013, EUR RADIOL, V23, P428, DOI 10.1007/s00330-012-2604-1
   [Anonymous], 2009, NEOPLASIA, V11, P102, DOI 10.1593/neo.81328
   Bisdas S, 2013, NEURORADIOLOGY, V55, P1189, DOI 10.1007/s00234-013-1229-7
   Chandarana H, 2011, INVEST RADIOL, V46, P285, DOI 10.1097/RLI.0b013e3181ffc485
   Cohen AD, 2015, MAGN RESON MED, V73, P306, DOI 10.1002/mrm.25109
   Ebrahimi B, 2014, INVEST RADIOL, V49, P640, DOI 10.1097/RLI.0000000000000066
   Federau C, 2014, AM J NEURORADIOL, V35, P256, DOI 10.3174/ajnr.A3686
   Ichikawa S, 2013, MAGN RESON IMAGING, V31, P414, DOI 10.1016/j.mri.2012.08.004
   Kakite S, 2015, J MAGN RESON IMAGING, V41, P149, DOI 10.1002/jmri.24538
   Klauss M, 2011, INVEST RADIOL, V46, P57, DOI 10.1097/RLI.0b013e3181fb3bf2
   Koh DM, 2011, AM J ROENTGENOL, V196, P1351, DOI 10.2214/AJR.10.5515
   Lai V, 2013, EUR RADIOL, V23, P2793, DOI 10.1007/s00330-013-2889-8
   LEBIHAN D, 1986, RADIOLOGY, V161, P401, DOI 10.1148/radiology.161.2.3763909
   LEBIHAN D, 1988, RADIOLOGY, V168, P497, DOI 10.1148/radiology.168.2.3393671
   Lemke A, 2011, MAGN RESON IMAGING, V29, P766, DOI 10.1016/j.mri.2011.03.004
   Lemke A, 2009, INVEST RADIOL, V44, P769, DOI 10.1097/RLI.0b013e3181b62271
   Luciani A, 2008, RADIOLOGY, V249, P891, DOI 10.1148/radiol.2493080080
   Notohamiprodjo M, 2008, INVEST RADIOL, V43, P677, DOI 10.1097/RLI.0b013e31817d14e6
   Notohamiprodjo M, 2010, EUR J RADIOL, V76, P337, DOI 10.1016/j.ejrad.2010.05.033
   Pang YX, 2013, MAGN RESON MED, V69, P553, DOI 10.1002/mrm.24277
   Priola AM, 2017, J MAGN RESON IMAGING, V46, P769, DOI 10.1002/jmri.25633
   Rheinheimer S, 2012, EUR J RADIOL, V81, pE310, DOI 10.1016/j.ejrad.2011.10.016
   Shinmoto H, 2012, AM J ROENTGENOL, V199, pW496, DOI 10.2214/AJR.11.8347
   Sigmund EE, 2012, RADIOLOGY, V263, P758, DOI 10.1148/radiol.12111327
   Stieb S, 2016, NEUROIMAGE-CLIN, V11, P780, DOI 10.1016/j.nicl.2016.05.022
   Thoeny HC, 2005, RADIOLOGY, V235, P911, DOI 10.1148/radiol.2353040554
   TURNER R, 1990, RADIOLOGY, V177, P407, DOI 10.1148/radiology.177.2.2217777
   Wang HY, 2010, RADIOLOGY, V257, P135, DOI 10.1148/radiol.10092396
   Wu WC, 2015, EUR RADIOL, V25, P2485, DOI 10.1007/s00330-015-3655-x
   Wurnig MC, 2015, MAGN RESON MED, V74, P1414, DOI 10.1002/mrm.25506
   Yamada I, 1999, RADIOLOGY, V210, P617, DOI 10.1148/radiology.210.3.r99fe17617
   Yu XD, 2012, EUR J RADIOL, V81, P3061, DOI 10.1016/j.ejrad.2012.04.028
   Zhang YD, 2015, EUR RADIOL, V25, P994, DOI 10.1007/s00330-014-3511-4
NR 33
TC 17
Z9 18
U1 0
U2 7
PU SPRINGER
PI NEW YORK
PA 233 SPRING ST, NEW YORK, NY 10013 USA
SN 2366-004X
EI 2366-0058
J9 ABDOM RADIOL
JI Abdom. Radiol.
PD JUL
PY 2018
VL 43
IS 7
BP 1739
EP 1748
DI 10.1007/s00261-017-1361-7
PG 10
WC Radiology, Nuclear Medicine & Medical Imaging
WE Science Citation Index Expanded (SCI-EXPANDED)
SC Radiology, Nuclear Medicine & Medical Imaging
GA GM6EF
UT WOS:000438253000025
PM 29071436
DA 2023-08-27
ER

PT J
AU [Anonymous]
AF [Anonymous]
TI Recommendations for reporting of tumors of the adrenal cortex and
   medulla
SO MODERN PATHOLOGY
LA English
DT Article
ID PEDIATRIC-ONCOLOGY-GROUP; NEUROBLASTIC TUMORS; PROGNOSTIC-SIGNIFICANCE;
   SHIMADA CLASSIFICATION; CORTICAL CARCINOMA; SYSTEM; INDEX
CR Brodeur GM, 1997, J PEDIAT HEMATOL ONC, V19, P93, DOI 10.1097/00043426-199703000-00001
   DEHNER LP, 1988, AM J PEDIAT HEMATOL, V10, P143
   DEHNER LP, 1998, NEUROBLASTOMAS TUMOR, P112
   HOUGH AJ, 1979, AM J CLIN PATHOL, V72, P390
   JOSHI VV, 1994, SEMIN DIAGN PATHOL, V11, P107
   JOSHI VV, 1992, CANCER-AM CANCER SOC, V69, P2183, DOI 10.1002/1097-0142(19920415)69:8<2183::AID-CNCR2820690828>3.0.CO;2-C
   JOSHI VV, 1991, MODERN PATHOL, V4, P139
   JOSHI VV, 1992, CANCER-AM CANCER SOC, V69, P2197, DOI 10.1002/1097-0142(19920415)69:8<2197::AID-CNCR2820690829>3.0.CO;2-X
   JOSHI VV, 1993, CANCER-AM CANCER SOC, V71, P3173, DOI 10.1002/1097-0142(19930515)71:10<3173::AID-CNCR2820711045>3.0.CO;2-P
   Lack EE, 1997, ATLAS TUMOR PATHOLOG, V19
   MACFARLANE D A, 1958, Ann R Coll Surg Engl, V23, P155
   PHILIP T, 1992, AM J PEDIAT HEMATOL, V14, P97
   SHIMADA H, 1984, J NATL CANCER I, V73, P405, DOI 10.1093/jnci/73.2.405
   SULLIVAN M, 1978, J UROLOGY, V120, P660, DOI 10.1016/S0022-5347(17)57317-6
   VONSLOOTEN H, 1985, CANCER, V55, P766
   WEISS LM, 1989, AM J SURG PATHOL, V13, P202, DOI 10.1097/00000478-198903000-00004
   WOOTEN MD, 1993, CANCER, V72, P3145, DOI 10.1002/1097-0142(19931201)72:11<3145::AID-CNCR2820721105>3.0.CO;2-N
NR 17
TC 0
Z9 0
U1 0
U2 0
PU NATURE PUBLISHING GROUP
PI NEW YORK
PA 75 VARICK ST, 9TH FLR, NEW YORK, NY 10013-1917 USA
SN 0893-3952
EI 1530-0285
J9 MODERN PATHOL
JI Mod. Pathol.
PD AUG
PY 1999
VL 12
IS 8
BP 835
EP 839
PG 5
WC Pathology
WE Science Citation Index Expanded (SCI-EXPANDED)
SC Pathology
GA 226BH
UT WOS:000082000100013
DA 2023-08-27
ER

PT J
AU Wong, RH
   Copeland, WR
   Jacob, JT
   Sivakanthan, S
   Van Gompel, JJ
   van Loveren, H
   Link, MJ
   Agazzi, S
AF Wong, Ricky H.
   Copeland, William R.
   Jacob, Jeffrey T.
   Sivakanthan, Sananthan
   Van Gompel, Jamie J.
   van Loveren, Harry
   Link, Michael J.
   Agazzi, Siviero
TI Anterior Extension of Tumor is as Important as Tumor Size to Facial
   Nerve Outcome and Extent of Resection for Vestibular Schwannomas
SO JOURNAL OF NEUROLOGICAL SURGERY PART B-SKULL BASE
LA English
DT Article
DE acoustic neuroma; anterior extension; facial nerve outcome; tumor
   dimension; vestibular schwannoma
ID ACOUSTIC NEUROMA; CLINICAL ARTICLE; SURGERY
AB Objectives We examined vestibular schwannoma tumor dimension and direction of growth to determine whether these correlate with facial nerve outcome as well as extent of resection (EOR).
   Design Retrospective review of prospectively maintained databases.
   Participants 206 patients were a part of this study.
   Main Outcome Measures Tumor dimensions were measured using preoperative magnetic resonance imaging, and a series of ratios were then calculated to further characterize tumor dimension. Regression analyses were performed to investigate correlation with facial nerve outcome and EOR.
   Results Patients with tumor extending >1.5 cm anterior to the internal auditory canal (IAC) (AB measurement) were three times more likely to have postoperative House-Brackman grades of 3 or worse. We also found that an EB/BF ratio (representing elongated growth parallel to the IAC axis) 1.1 was associated with half the risk of poor facial nerve outcome. Tumors with anterior-posterior diameter (AC measurement) >1.9 cm were five times less likely to undergo gross total resection (GTR). Furthermore, an increased degree of tumor extension into the IAC (DE measurement >2.4 cm) or an increased amount of brainstem compression (EB measurement >1.1 cm) were each associated with a nearly 3-fold decrease in the likelihood of GTR.
   Conclusion Our study demonstrates that anterior extent of the tumor is as important as tumor size to facial nerve outcome and degree of resection for vestibular schwannomas.
C1 [Wong, Ricky H.] NorthShore Univ Hlth Syst, Pritzker Sch Med, Dept Neurol Surg, Evanston, IL USA.
   [Copeland, William R.; Jacob, Jeffrey T.; Van Gompel, Jamie J.; Link, Michael J.] Mayo Clin, Dept Neurol Surg, Rochester, MN USA.
   [van Loveren, Harry; Agazzi, Siviero] Univ S Florida, Dept Neurol Surg, 2 Tampa Gen Circle, Tampa, FL 33606 USA.
   [Sivakanthan, Sananthan] Univ New Mexico, Dept Neurol Surg, Albuquerque, NM 87131 USA.
C3 NorthShore University Health System; Mayo Clinic; State University
   System of Florida; University of South Florida; University of New Mexico
RP Agazzi, S (通讯作者)，Univ S Florida, Dept Neurol Surg, 2 Tampa Gen Circle, Tampa, FL 33606 USA.
EM
RI Van Gompel, Jamie/AAW-4459-2020; agazzi, siviero/AAA-5653-2021
OI Van Gompel, Jamie/0000-0001-8087-7870; agazzi,
   siviero/0000-0001-5760-0187
CR Ansari SF, 2012, NEUROSURG FOCUS, V33, DOI 10.3171/2012.6.FOCUS12163
   Bloch O, 2011, J NEURO-ONCOL, V102, P281, DOI 10.1007/s11060-010-0315-5
   Brackmann DE, 2007, OTOLARYNG HEAD NECK, V136, P773, DOI 10.1016/j.otohns.2006.10.009
   Carlson ML, 2012, LARYNGOSCOPE, V122, P378, DOI 10.1002/lary.22411
   Chen ZN, 2014, J NEUROSURG, V120, P1278, DOI 10.3171/2014.2.JNS131497
   Copeland WR, 2013, J NEUROL SURG PART B, V74, P347, DOI 10.1055/s-0033-1347369
   Falcioni M, 2011, J NEUROSURG, V115, P820, DOI 10.3171/2011.5.JNS101597
   Gerganov VM, 2011, J NEUROSURG, V115, P1087, DOI 10.3171/2011.7.JNS11495
   Gerganov VM, 2009, ACTA NEUROCHIR, V151, P581, DOI 10.1007/s00701-009-0288-3
   Grey PL, 1996, CLIN OTOLARYNGOL, V21, P409, DOI 10.1046/j.1365-2273.1996.00816.x
   HARDY DG, 1989, J NEUROSURG, V71, P799, DOI 10.3171/jns.1989.71.6.0799
   HOUSE JW, 1985, OTOLARYNG HEAD NECK, V93, P146, DOI 10.1177/019459988509300202
   Jacob JT, 2016, LARYNGOSCOPE, V126, P1877, DOI 10.1002/lary.25779
   LEE T K Y, 1990, British Journal of Neurosurgery, V4, P5, DOI 10.3109/02688699009000675
   RUTH HR, 1985, OTOLARYNG HEAD NECK, V93, P160, DOI 10.1177/019459988509300206
   Ryzenman JM, 2005, OTOL NEUROTOL, V26, P516, DOI 10.1097/01.mao.0000169786.22707.12
   Sameshima T, 2013, J NEUROL SURG PART B, V74, P39, DOI 10.1055/s-0032-1329625
   Sampath P, 1997, J NEUROSURG, V87, P60, DOI 10.3171/jns.1997.87.1.0060
   Sharma M, 2013, J NEUROL SURG PART B, V74, P317, DOI 10.1055/s-0033-1349060
   STERKERS JM, 1994, OTOLARYNG HEAD NECK, V110, P146, DOI 10.1177/019459989411000202
   Symon L, 1989, Br J Neurosurg, V3, P343, DOI 10.3109/02688698909002814
   Taoka T, 2006, J MAGN RESON IMAGING, V24, P1005, DOI 10.1002/jmri.20725
NR 22
TC 12
Z9 14
U1 0
U2 5
PU THIEME MEDICAL PUBL INC
PI NEW YORK
PA 333 SEVENTH AVE, NEW YORK, NY 10001 USA
SN 2193-6331
EI 2193-634X
J9 J NEUROL SURG PART B
JI Journal of Neurol. Surg. Part B
PD DEC
PY 2017
VL 78
IS 6
BP 473
EP 480
DI 10.1055/s-0037-1604331
PG 8
WC Clinical Neurology; Surgery
WE Science Citation Index Expanded (SCI-EXPANDED)
SC Neurosciences & Neurology; Surgery
GA FM4CW
UT WOS:000414959900006
PM 29134166
OA Green Published
DA 2023-08-27
ER

PT J
AU Bhatnagar, AK
   Gerszten, PC
   Ozhasaglu, C
   Vogel, WJ
   Kalnicki, S
   Welch, WC
   Burton, SA
AF Bhatnagar, AK
   Gerszten, PC
   Ozhasaglu, C
   Vogel, WJ
   Kalnicki, S
   Welch, WC
   Burton, SA
TI CyberKnife frameless radiosurgery for the treatment of extracranial
   benign tumors
SO TECHNOLOGY IN CANCER RESEARCH & TREATMENT
LA English
DT Article
DE CyberKnife; extracranial; radiosurgery; and benign tumors
ID STEREOTACTIC RADIOSURGERY; RADIATION-THERAPY; PHASE-I; CONFORMAL
   RADIOTHERAPY; CLINICAL-EXPERIENCE; PARASPINAL TUMORS; SPINAL LESIONS;
   MENINGIOMAS; METASTASES; PREVENTION
AB Limited data exists for the use of radiosurgery for benign extracranial tumors. The purpose of this study was to evaluate the feasibility, toxicity, and local control of patients with benign extracranial lesions treated with the CyberKnife Frameless Radiosurgery System. From September 2001 thru January 2004, 59 benign tumors in 44 patients were treated using the CyberKnife a frameless image-guided radiosurgery system. Of these tumors, there were 21 neurofibromas, ten schwannomas, eight meningiomas, eight hemangioblastomas, seven paragangliomas, two hemangiopericytomas, one pseudotumor, one ependymoma, and one arteriovenous malformation (AVM). The anatomic locations of these tumors were spinal (25 cervical, four thoracic, 14 lumbar, and two sacral), neck (eight), orbital (three), brainstem (one), and foramen magnum (one). All patients were treated in a single fraction except three lesions were treated in a fractionated manner. The median treatment delivery time per fraction was 59 minutes (range 11-194). Twenty three lesions initially underwent surgical resection. Ten lesions received prior external beam radiation with a median dose 48 Gy (range 40-54 Gy), and one lesion received two prior CyberKnife treatments for a total dose of 32 Gy to the 80% isodose line. The median follow-up was eight months (range 1-25 months). Acute and late toxicity was graded using the National Cancer Institute Common Toxicity Criteria (CTC) scale. Symptomatic response was documented as "improved," "stable," or "progression". The median tumor dose delivered was 16.0 Gy to the 80% isodose line (range 10-31 Gy). The median tumor volume was 4.3 cc (range 0.14-98.6 cc). The median spinal cord volume receiving more than 8 Gy was 0.035 cc (range 0-2.5 cc) and the median maximum spinal cord dose 11.5 Gy (range 0-19.8 Gy). There were no patients that suffered a significant (Grade 3, 4, or 5) acute toxicity. There was no observed late toxicity. 78% of patients experienced an improvement of their pre-treatment symptoms while only one patient experienced symptom progression. Of the 26 patients who underwent follow-up imaging, the local control rate was 96%. This study suggests that CyberKnife Radiosurgery is a safe and efficacious treatment modality for benign tumors, even for those patients with recurrent previously irradiated lesions.
C1 Univ Pittsburgh, Inst Canc, Dept Radiat Oncol, Pittsburgh, PA 15232 USA.
   Univ Pittsburgh, Inst Canc, Dept Neurol Surg, Pittsburgh, PA 15232 USA.
C3 Pennsylvania Commonwealth System of Higher Education (PCSHE); University
   of Pittsburgh; Pennsylvania Commonwealth System of Higher Education
   (PCSHE); University of Pittsburgh
RP Bhatnagar, AK (通讯作者)，Univ Pittsburgh, Inst Canc, Dept Radiat Oncol, 5230 Ctr Ave, Pittsburgh, PA 15232 USA.
EM
CR Adler JR, 1997, STEREOT FUNCT NEUROS, V69, P124, DOI 10.1159/000099863
   Adler JR, 1999, NEUROSURGERY, V44, P1299, DOI 10.1097/00006123-199906000-00079
   Bilsky MH, 2004, NEUROSURGERY, V54, P823, DOI 10.1227/01.NEU.0000114263.01917.1E
   BLOMGREN H, 1995, ACTA ONCOL, V34, P861, DOI 10.3109/02841869509127197
   Chang EL, 2004, INT J RADIAT ONCOL, V59, P1288, DOI 10.1016/j.ijrobp.2004.04.025
   Flickinger JC, 1996, INT J RADIAT ONCOL, V36, P873, DOI 10.1016/S0360-3016(96)00316-1
   FLICKINGER JC, 1991, CANCER, V67, P345, DOI 10.1002/1097-0142(19910115)67:2<345::AID-CNCR2820670205>3.0.CO;2-M
   Fuss M, 2004, ANN SURG ONCOL, V11, P130, DOI 10.1245/ASO.2004.10.907
   Gerszten PC, 2004, NEUROSURGERY, V55, P89, DOI 10.1227/01.NEU.0000126873.00845.A7
   Gerszten PC, 2003, STEREOT FUNCT NEUROS, V81, P84, DOI 10.1159/000075109
   Keilholz L, 1996, INT J RADIAT ONCOL, V36, P891, DOI 10.1016/S0360-3016(96)00421-X
   KLUMPAR DI, 1994, J AM ACAD DERMATOL, V31, P225, DOI 10.1016/S0190-9622(94)70152-0
   Kondziolka D, 2003, NEUROSURGERY, V53, P815, DOI 10.1093/neurosurgery/53.4.815
   KONDZIOLKA D, 1991, J NEUROSURG, V74, P552, DOI 10.3171/jns.1991.74.4.0552
   Koong AC, 2004, INT J RADIAT ONCOL, V58, P1017, DOI 10.1016/j.ijrobp.2003.11.004
   Kuo JS, 2003, NEUROSURGERY, V53, P1235, DOI 10.1227/01.NEU.0000089485.47590.05
   LEKSELL L, 1971, ACTA CHIR SCAND, V137, P311
   Leksell L, 1971, Lakartidningen, V68, P5145
   LEKSELL L, 1971, ACTA CHIR SCAND, V137, P763
   LEKSELL L, 1951, ACTA CHIR SCAND, V102, P316
   Lukacs S, 1978, J Dermatol Surg Oncol, V4, P620
   LUNSFORD LD, 1991, J NEUROSURG, V75, P512, DOI 10.3171/jns.1991.75.4.0512
   Milker-Zabel S, 2003, INT J RADIAT ONCOL, V55, P162, DOI 10.1016/S0360-3016(02)03864-6
   Pirzkall A, 2000, INT J RADIAT ONCOL, V48, P1197, DOI 10.1016/S0360-3016(00)00739-2
   Ryu SI, 2001, NEUROSURGERY, V49, P838, DOI 10.1097/00006123-200110000-00011
   Seegenschmiedt M. H., 1996, Strahlentherapie und Onkologie, V172, P376
   SEEGENSCHMIEDT MH, 1994, INT J RADIAT ONCOL, V30, P63
   Shiu AS, 2003, INT J RADIAT ONCOL, V57, P605, DOI 10.1016/S0360-3016(03)00792-2
   SOLAN MJ, 1985, INT J RADIAT ONCOL, V11, P675, DOI 10.1016/0360-3016(85)90297-4
   Steiner L, 1974, Acta Neurochir (Wien), VSuppl 21, P195
   Takeuchi H, 2003, MINIM INVAS NEUROSUR, V46, P82, DOI 10.1055/s-2003-39342
   Whyte RI, 2003, ANN THORAC SURG, V75, P1097, DOI 10.1016/S0003-4975(02)04681-7
NR 32
TC 30
Z9 32
U1 0
U2 4
PU ADENINE PRESS
PI SCHENECTADY
PA 2066 CENTRAL AVE, SCHENECTADY, NY 12304 USA
SN 1533-0346
J9 TECHNOL CANCER RES T
JI Technol. Cancer Res. Treat.
PD OCT
PY 2005
VL 4
IS 5
BP 571
EP 576
DI 10.1177/153303460500400511
PG 6
WC Oncology
WE Science Citation Index Expanded (SCI-EXPANDED)
SC Oncology
GA 974EA
UT WOS:000232572900012
PM 16173828
DA 2023-08-27
ER

PT J
AU Chung, S
   Moon, JI
   Leung, A
   Aldrich, D
   Lukianov, S
   Kitayama, Y
   Park, S
   Li, Y
   Bolshakov, VY
   Lamonerie, T
   Kim, KS
AF Chung, Sangmi
   Moon, Jung-Il
   Leung, Amanda
   Aldrich, Daniel
   Lukianov, Stefan
   Kitayama, Yui
   Park, Sara
   Li, Yan
   Bolshakov, Vadim Y.
   Lamonerie, Thomas
   Kim, Kwang-Soo
TI ES cell-derived renewable and functional midbrain dopaminergic
   progenitors
SO PROCEEDINGS OF THE NATIONAL ACADEMY OF SCIENCES OF THE UNITED STATES OF
   AMERICA
LA English
DT Article
DE neural precursors; dopaminergic neurons; transplantation
ID FIBROBLAST-GROWTH-FACTOR; EMBRYONIC STEM-CELLS; PARKINSONS-DISEASE;
   ANIMAL-MODEL; NEURONS; PLATE; DIFFERENTIATION; FGF; TRANSPLANTATION;
   GENERATION
AB During early development, midbrain dopaminergic (mDA) neuronal progenitors (NPs) arise from the ventral mesencephalic area by the combined actions of secreted factors and their downstream transcription factors. These mDA NPs proliferate, migrate to their final destinations, and develop into mature mDA neurons in the substantia nigra and the ventral tegmental area. Here, we show that such authentic mDA NPs can be efficiently isolated from differentiated ES cells (ESCs) using a FACS method combining two markers, Otx2 and Corin. Purified Otx2(+) Corin(+) cells coexpressed other mDA NP markers, including FoxA2, Lmx1b, and Glast. Using optimized culture conditions, these mDA NPs continuously proliferated up to 4 wk with almost 1,000-fold expansion without significant changes in their phenotype. Furthermore, upon differentiation, Otx2(+) Corin(+) cells efficiently generated mDA neurons, as evidenced by coexpression of mDA neuronal markers (e. g., TH, Pitx3, Nurr1, and Lmx1b) and physiological functions (e. g., efficient DA secretion and uptake). Notably, these mDA NPs differentiated into a relatively homogenous DA population with few serotonergic neurons. When transplanted into PD model animals, aphakia mice, and 6-OHDA-lesioned rats, mDA NPs differentiated into mDA neurons in vivo and generated well-integrated DA grafts, resulting in significant improvement in motor dysfunctions without tumor formation. Furthermore, grafted Otx2(+) Corin(+) cells exhibited significant migratory function in the host striatum, reaching >3.3 mm length in the entire striatum. We propose that functional and expandable mDA NPs can be efficiently isolated by this unique strategy and will serve as useful tools in regenerative medicine, bioassay, and drug screening.
C1 [Chung, Sangmi; Moon, Jung-Il; Leung, Amanda; Aldrich, Daniel; Lukianov, Stefan; Kitayama, Yui; Park, Sara; Kim, Kwang-Soo] Harvard Univ, McLean Hosp, Sch Med, Mol Neurobiol Lab, Belmont, MA 02478 USA.
   [Chung, Sangmi; Moon, Jung-Il; Leung, Amanda; Aldrich, Daniel; Lukianov, Stefan; Kitayama, Yui; Park, Sara; Li, Yan; Bolshakov, Vadim Y.; Kim, Kwang-Soo] Harvard Univ, McLean Hosp, Sch Med, Dept Psychiat, Belmont, MA 02478 USA.
   [Chung, Sangmi; Moon, Jung-Il; Leung, Amanda; Aldrich, Daniel; Lukianov, Stefan; Kitayama, Yui; Park, Sara; Kim, Kwang-Soo] Harvard Univ, McLean Hosp, Sch Med, Program Neurosci, Belmont, MA 02478 USA.
   [Chung, Sangmi; Moon, Jung-Il; Leung, Amanda; Aldrich, Daniel; Lukianov, Stefan; Kitayama, Yui; Park, Sara; Kim, Kwang-Soo] Harvard Univ, McLean Hosp, Sch Med, Harvard Stem Cell Inst, Belmont, MA 02478 USA.
   [Li, Yan; Bolshakov, Vadim Y.] Harvard Univ, McLean Hosp, Sch Med, Cellular Neurobiol Lab, Belmont, MA 02478 USA.
   [Lamonerie, Thomas] Univ Nice Sophia Antipolis, Unite Mixte Rech, Inst Dev Biol & Canc, Ctr Natl Rech Sci 6543, F-06108 Nice, France.
C3 Harvard University; McLean Hospital; Harvard University; McLean
   Hospital; Harvard University; McLean Hospital; Harvard University;
   McLean Hospital; Harvard University; McLean Hospital; UDICE-French
   Research Universities; Universite Cote d'Azur
RP Chung, S (通讯作者)，Harvard Univ, McLean Hosp, Sch Med, Mol Neurobiol Lab, Belmont, MA 02478 USA.
EM
RI Li, Yan/HNR-8933-2023
OI Li, Yan/0000-0003-3673-181X; Lamonerie, Thomas/0000-0003-2371-2708
FU National Institutes of Health [NS070577, MH087903, MH048866]
FX This study was supported by National Institutes of Health Grants
   NS070577, MH087903, and MH048866.
CR Barker RA, 2010, CELL STEM CELL, V7, P148, DOI 10.1016/j.stem.2010.07.003
   Broccoli V, 1999, NATURE, V401, P164, DOI 10.1038/43670
   Castelo-Branco GA, 2003, P NATL ACAD SCI USA, V100, P12747, DOI 10.1073/pnas.1534900100
   Chellaiah A, 1999, J BIOL CHEM, V274, P34785, DOI 10.1074/jbc.274.49.34785
   Chung S, 2006, J NEUROCHEM, V97, P1467, DOI 10.1111/j.1471-4159.2006.03841.x
   Chung S, 2002, EUR J NEUROSCI, V16, P1829, DOI 10.1046/j.1460-9568.2002.02255.x
   Chung S, 2009, CELL STEM CELL, V5, P646, DOI 10.1016/j.stem.2009.09.015
   Enshell-Seijffers D, 2008, DEVELOPMENT, V135, P217, DOI 10.1242/dev.011031
   Fasano CA, 2010, CELL STEM CELL, V6, P336, DOI 10.1016/j.stem.2010.03.001
   Fossat N, 2007, BMC DEV BIOL, V7, DOI 10.1186/1471-213X-7-122
   Grothe C, 2007, BRAIN RES REV, V54, P80, DOI 10.1016/j.brainresrev.2006.12.001
   Hedlund E, 2008, STEM CELLS, V26, P1526, DOI 10.1634/stemcells.2007-0996
   Hockemeyer D, 2009, NAT BIOTECHNOL, V27, P851, DOI 10.1038/nbt.1562
   Hwang DY, 2005, J NEUROSCI, V25, P2132, DOI 10.1523/JNEUROSCI.3718-04.2005
   Joksimovic M, 2009, NAT NEUROSCI, V12, P125, DOI 10.1038/nn.2243
   Kim JH, 2002, NATURE, V418, P50, DOI 10.1038/nature00900
   Kittappa R, 2007, PLOS BIOL, V5, P2875, DOI 10.1371/journal.pbio.0050325
   Ladewig J, 2008, STEM CELLS, V26, P1705, DOI 10.1634/stemcells.2008-0007
   Lee SH, 2000, NAT BIOTECHNOL, V18, P675, DOI 10.1038/76536
   Louvi A, 2006, NAT REV NEUROSCI, V7, P93, DOI 10.1038/nrn1847
   Muguruma K, 2010, NAT NEUROSCI, V13, P1171, DOI 10.1038/nn.2638
   Ohmachi S, 2003, J NEUROSCI RES, V72, P436, DOI 10.1002/jnr.10592
   Ono Y, 2007, DEVELOPMENT, V134, P3213, DOI 10.1242/dev.02879
   Peljto M, 2010, CELL STEM CELL, V7, P355, DOI 10.1016/j.stem.2010.07.013
   Politis M, 2010, SCI TRANSL MED, V2, DOI 10.1126/scitranslmed.3000976
   Saarimaki-Vire J, 2007, J NEUROSCI, V27, P8581, DOI 10.1523/JNEUROSCI.0192-07.2007
   Sasaki H, 1997, DEVELOPMENT, V124, P1313
   Shi YH, 2008, CRIT REV ONCOL HEMAT, V65, P43, DOI 10.1016/j.critrevonc.2007.06.004
   Smidt MP, 2007, NAT REV NEUROSCI, V8, P21, DOI 10.1038/nrn2039
   Tang MZ, 2009, DEVELOPMENT, V136, P2027, DOI 10.1242/dev.034330
   Timmer M, 2007, J NEUROSCI, V27, P459, DOI 10.1523/JNEUROSCI.4493-06.2007
   Tropepe V, 1999, DEV BIOL, V208, P166, DOI 10.1006/dbio.1998.9192
   Tsuji O, 2010, P NATL ACAD SCI USA, V107, P12704, DOI 10.1073/pnas.0910106107
   Wichterle H, 2002, CELL, V110, P385, DOI 10.1016/S0092-8674(02)00835-8
   Ye WL, 1998, CELL, V93, P755, DOI 10.1016/S0092-8674(00)81437-3
NR 35
TC 72
Z9 76
U1 1
U2 15
PU NATL ACAD SCIENCES
PI WASHINGTON
PA 2101 CONSTITUTION AVE NW, WASHINGTON, DC 20418 USA
SN 0027-8424
J9 P NATL ACAD SCI USA
JI Proc. Natl. Acad. Sci. U. S. A.
PD JUN 7
PY 2011
VL 108
IS 23
BP 9703
EP 9708
DI 10.1073/pnas.1016443108
PG 6
WC Multidisciplinary Sciences
WE Science Citation Index Expanded (SCI-EXPANDED)
SC Science & Technology - Other Topics
GA 773WE
UT WOS:000291341400075
PM 21606375
OA Green Published, Bronze
DA 2023-08-27
ER

PT J
AU Gomes, ND
   Rezende, CD
   Fontes, SP
   Hovell, AMC
   Landgraf, RG
   Matheus, ME
   Pinto, AD
   Fernandes, PD
AF Gomes, Niele de Matos
   Rezende, Claudia de Moraes
   Fontes, Silvia Paredes
   Hovell, Ana Maria Celestino
   Landgraf, Richardt Gama
   Matheus, Maria Eline
   Pinto, Angelo da Cunha
   Fernandes, Patricia Dias
TI Antineoplasic activity of Copaifera multijuga oil and fractions against
   ascitic and solid Ehrlich tumor
SO JOURNAL OF ETHNOPHARMACOLOGY
LA English
DT Article
DE Copaifera multijuga; Ehrlich ascitic tumor; cancer; copaiba
ID IN-VITRO; ANTITUMOR-ACTIVITY; NITRATE; ENZYME; CELLS
AB The aim of this study was to investigate the effect of chronic treatment with C. multijuga oil on Ehrlich tumor evolution. C multijuga was fractionated in a KOH impregnated silica gel column chromatography to give three distinct fractions, i.e., hexanic, chloroformic, and methanolic, mainly composed by hydrocarbon sesquiterpenes, oxygenated sesquiterpenes and acidic diterpenes, respectively. Results demonstrated that the C multijuga oil, the hexanic, and chloroformic fractions did not develop toxic effects. The oil, hexanic and chloroformic fractions (doses varying between 100 and 200 mg/kg) showed antineoplasic properties against Ehrlich ascitic tumor (EAT) and solid tumor during 10 consecutive days of treatment inhibiting ascitic tumor cell number, reverting medulla and blood cell counts to values similar to control group, and inhibiting the increase on several inflammatory mediators (total protein, PGE(2), nitric oxide, and TNF) on ascitic fluid. The treatment also inhibited the increase in paw volume on tumor-inoculated mice. In conclusion, C. multijugo as well as its fractions demonstrated antineoplasic effect even after oral administration confirming its use by traditional medicine. (C) 2008 Elsevier Ireland Ltd. All rights reserved.
C1 [Gomes, Niele de Matos; Matheus, Maria Eline; Fernandes, Patricia Dias] Univ Fed Rio de Janeiro, Programa Desenvolvimento Farm, ICB, BR-21944970 Rio De Janeiro, Brazil.
   [Rezende, Claudia de Moraes; Fontes, Silvia Paredes; Hovell, Ana Maria Celestino; Pinto, Angelo da Cunha] Univ Fed Rio de Janeiro, Inst Quim, Ctr Tecnol, BR-21941 Rio De Janeiro, Brazil.
   [Landgraf, Richardt Gama] Univ Sao Paulo, ICB, Dept Imunol, BR-05508 Sao Paulo, Brazil.
C3 Universidade Federal do Rio de Janeiro; Universidade Federal do Rio de
   Janeiro; Universidade de Sao Paulo
RP Fernandes, PD (通讯作者)，Univ Fed Rio de Janeiro, Programa Desenvolvimento Farm, ICB, Caixa Postal 68016, BR-21944970 Rio De Janeiro, Brazil.
EM
RI Rezende, Claudia/H-6138-2013; Landgraf, Richardt/F-7146-2012; Fernandes,
   Patricia Dias/C-4489-2013; Fernandes, Patricia Dias/W-1565-2019;
   Fernandes, Patricia/B-4117-2010
OI Rezende, Claudia/0000-0003-2710-5702; Landgraf,
   Richardt/0000-0001-7774-7880; Fernandes, Patricia
   Dias/0000-0001-6870-6745; Fernandes, Patricia Dias/0000-0001-6870-6745; 
FU CAPES
FX This work was supported by grants from CAPES (fellowship to NMG and
   SPF), FAPERJ, and CNPq.
CR BARTHOLOMEW B, 1984, FOOD CHEM TOXICOL, V22, P541, DOI 10.1016/0278-6915(84)90224-2
   BASILE AC, 1988, J ETHNOPHARMACOL, V22, P101, DOI 10.1016/0378-8741(88)90235-8
   Bergami-Santos PC, 2004, LIFE SCI, V75, P245, DOI 10.1016/j.lfs.2004.02.003
   Braga WF, 1998, PHYTOCHEMISTRY, V49, P263, DOI 10.1016/S0031-9422(97)00899-6
   Cavalieri E, 2004, BIOCHEM BIOPH RES CO, V315, P589, DOI 10.1016/j.bbrc.2004.01.088
   CHEN L, 1970, NATURE, V225, P734, DOI 10.1038/225734a0
   FASTAIA J, 1976, J NATL CANCER I, V56, P547, DOI 10.1093/jnci/56.3.547
   FECCHIO D, 1990, INFLAMMATION, V14, P125, DOI 10.1007/BF00914035
   FERREIRA S H, 1979, Journal of Pharmacy and Pharmacology, V31, P648
   FLICK DA, 1984, J IMMUNOL METHODS, V68, P167, DOI 10.1016/0022-1759(84)90147-9
   GREEN LC, 1982, ANAL BIOCHEM, V126, P131, DOI 10.1016/0003-2697(82)90118-X
   Gupta M, 2004, J PHARMACOL SCI, V94, P177, DOI 10.1254/jphs.94.177
   Kleeb S. R., 1997, Life Sciences, V60, P69
   Legault J, 2003, PLANTA MED, V69, P402, DOI 10.1055/s-2003-39695
   Lima SRM, 2003, PHYTOTHER RES, V17, P1048, DOI 10.1002/ptr.1295
   OHSAKI A, 1994, BIOORG MED CHEM LETT, V4, P2889, DOI 10.1016/S0960-894X(01)80834-9
   Paiva LAF, 2004, LIFE SCI, V75, P1979, DOI 10.1016/j.lfs.2004.05.011
   Paiva LAF, 2004, J ETHNOPHARMACOL, V93, P51, DOI 10.1016/j.jep.2004.03.028
   Pinto AC, 2000, J BRAZIL CHEM SOC, V11, P355, DOI 10.1590/S0103-50532000000400005
   PRADELLES P, 1985, ANAL CHEM, V57, P1170, DOI 10.1021/ac00284a003
   Sant'Anna BMP, 2007, J BRAZIL CHEM SOC, V18, P984, DOI 10.1590/S0103-50532007000500016
   Segura JA, 2000, IMMUNOL LETT, V74, P111, DOI 10.1016/S0165-2478(00)00208-X
   Tao L, 2006, CANCER CHEMOTH PHARM, V58, P24, DOI 10.1007/s00280-005-0137-x
   Wang GH, 2002, J NAT PROD, V65, P887, DOI 10.1021/np010586i
NR 24
TC 45
Z9 47
U1 1
U2 15
PU ELSEVIER IRELAND LTD
PI CLARE
PA ELSEVIER HOUSE, BROOKVALE PLAZA, EAST PARK SHANNON, CO, CLARE, 00000,
   IRELAND
SN 0378-8741
EI 1872-7573
J9 J ETHNOPHARMACOL
JI J. Ethnopharmacol.
PD SEP 2
PY 2008
VL 119
IS 1
BP 179
EP 184
DI 10.1016/j.jep.2008.06.033
PG 6
WC Plant Sciences; Chemistry, Medicinal; Integrative & Complementary
   Medicine; Pharmacology & Pharmacy
WE Science Citation Index Expanded (SCI-EXPANDED)
SC Plant Sciences; Pharmacology & Pharmacy; Integrative & Complementary
   Medicine
GA 350EY
UT WOS:000259336600028
PM 18647645
DA 2023-08-27
ER

PT J
AU Shields, LBE
   Bond, C
   Odom, A
   Sun, DA
   Spalding, AC
AF Shields, Lisa B. E.
   Bond, Cindy
   Odom, Aaron
   Sun, David A.
   Spalding, Aaron C.
TI Heterogeneity correction for intensity-modulated frameless SRS in
   pituitary and cavernous sinus tumors: a retrospective study
SO RADIATION ONCOLOGY
LA English
DT Article
DE Radiation; Oncology; Radiosurgery; Dosimetry; Pituitary tumor; Cavernous
   sinus tumor
ID INITIAL CLINICAL-EXPERIENCE; PENCIL BEAM ALGORITHM; ACUROS XB ALGORITHM;
   STEREOTACTIC RADIOSURGERY; EXPERIMENTAL VALIDATION; DOSE CALCULATION;
   CRANIAL NERVES; IMRT; OPTIMIZATION; ACCURACY
AB Background: Frameless immobilization allows for planning and quality assurance of intensity-modulated radiosurgery (IM-SRS) plans. We tested the hypothesis that IM-SRS planning with uniform tissue density corrections results in dose inaccuracy compared to heterogeneity-corrected algorithms.
   Methods: Fifteen patients with tumors of the pituitary or cavernous sinus underwent frameless IM-SRS. Treatment planning CT and MRI scans were obtained and fused to delineate the tumor, optic nerves, chiasm, and brainstem. The plan was developed with static gantry IM-SRS fields using a pencil beam (PB), analytical anisotropic (AAA), and Acuros XB (AXB) algorithms. We evaluated measures of target coverage as well as doses to organs at risk (OAR) for each algorithm. We compared the results of each algorithm in the cases where PTV overlapped OAR (n = 10) to cases without overlapping OAR with PTV (n = 5). Utilizing film dosimetry, we measured the dose distribution for each algorithm through a uniform density target to a rando phantom with non-uniform density of air, tissue, and bone.
   Results: There was no difference in target coverage measured by D-MaxPTV, D-MinPTV, D-95%PTV, or the isodose surface (IDS) covering 95 % of the PTV regardless of algorithm. However, there were differences in dose to OAR. PB predicted higher (p < 0.05) Dmax for the brainstem, chiasm, right optic nerve, and left optic nerve. In cases of PTV overlapping an optic nerve (n = 7), PB was unable to limit dose to 8Gy while achieving PTV coverage (PB 855 cGy vs. AAA 769 cGy, p = 0.05 vs. AXB 658 cGy, p = 0.03). Within the rando phantom, the PB and AAA algorithms over-estimated the dose delivered in the bone-tissue-air interface of the sinus (+ 17 %), while the AXB algorithm closely predicted the actual dose delivered through the inhomogeneous tissue (+/-1 % max, p < 0.05).
   Conclusions: Patients undergoing frameless SRS benefit from heterogeneity corrected dose plans when the lesion lies in areas of widely varying tissue density and near critical normal structures such as the skull base. Film dosimetry confirms that the AXB dose calculation algorithm more accurately predicts actual dose delivered though tissues of varying densities than PB or AAA dose calculation algorithms.
C1 [Shields, Lisa B. E.; Sun, David A.] Norton Healthcare, Norton Neurosci Inst, Louisville, KY 40202 USA.
   [Shields, Lisa B. E.; Bond, Cindy; Odom, Aaron; Sun, David A.; Spalding, Aaron C.] Norton Canc Inst Radiat Ctr, Louisville, KY USA.
   [Shields, Lisa B. E.; Sun, David A.; Spalding, Aaron C.] Norton Healthcare, Brain Tumor Ctr, Louisville, KY 40202 USA.
   [Bond, Cindy; Odom, Aaron] Associates Med Phys LLC, Greenbelt, MD USA.
RP Shields, LBE (通讯作者)，Norton Healthcare, Norton Neurosci Inst, 210 East Gray St,Suite 1102, Louisville, KY 40202 USA.
EM
OI Odom, Aaron/0000-0001-8530-4561
CR Ackerly T, 2011, MED PHYS, V38, P5040, DOI 10.1118/1.3611044
   Breitman K, 2007, J APPL CLIN MED PHYS, V8, P76, DOI 10.1120/jacmp.v8i2.2350
   Dogan N, 2006, PHYS MED BIOL, V51, P4967, DOI 10.1088/0031-9155/51/19/015
   Fogliata A, 2011, RADIAT ONCOL, V6, DOI 10.1186/1748-717X-6-82
   Gil E, 2009, MED PHYS, V36, P4318, DOI 10.1118/1.3244175
   Han T, MED PHYS
   Han T, 2012, MED PHYS, V39, P2193, DOI 10.1118/1.3692180
   Jelen U, 2007, PHYS MED BIOL, V52, P617, DOI 10.1088/0031-9155/52/3/006
   Kamath R, 2005, INT J RADIAT ONCOL, V61, P1467, DOI 10.1016/j.ijrobp.2004.08.021
   Kan MWK, 2013, INT J RADIAT ONCOL, V85, pE73, DOI 10.1016/j.ijrobp.2012.08.031
   Khan FM, 2014, KHANS PHYS RAD THERA, P195
   Lawson JD, 2010, J NEURO-ONCOL, V97, P59, DOI 10.1007/s11060-009-9987-0
   Leber KA, 1998, J NEUROSURG, V88, P43, DOI 10.3171/jns.1998.88.1.0043
   LUTZ W, 1988, INT J RADIAT ONCOL, V14, P373, DOI 10.1016/0360-3016(88)90446-4
   Pacific Northwest X-Ray Inc, 2015, RAND PHANT
   Ryken TC, 2001, INT J RADIAT ONCOL, V51, P1152, DOI 10.1016/S0360-3016(01)01756-4
   Salter BJ, 2009, INT J RADIAT ONCOL, V73, P546, DOI 10.1016/j.ijrobp.2008.09.011
   TISHLER RB, 1993, INT J RADIAT ONCOL, V27, P215, DOI 10.1016/0360-3016(93)90230-S
   Van Esch A, 2006, MED PHYS, V33, P4130, DOI 10.1118/1.2358333
   Wang JZ, 2012, MED DOSIM, V37, P31, DOI 10.1016/j.meddos.2010.12.010
   Yamashita T, 2012, PHYS MED BIOL, V57, P7673, DOI 10.1088/0031-9155/57/22/7673
   Zacarias AS, 2009, J APPL CLIN MED PHYS, V10, P281, DOI 10.1120/jacmp.v10i4.3061
NR 22
TC 3
Z9 3
U1 0
U2 1
PU BIOMED CENTRAL LTD
PI LONDON
PA 236 GRAYS INN RD, FLOOR 6, LONDON WC1X 8HL, ENGLAND
SN 1748-717X
J9 RADIAT ONCOL
JI Radiat. Oncol.
PD SEP 17
PY 2015
VL 10
AR 193
DI 10.1186/s13014-015-0500-y
PG 8
WC Oncology; Radiology, Nuclear Medicine & Medical Imaging
WE Science Citation Index Expanded (SCI-EXPANDED)
SC Oncology; Radiology, Nuclear Medicine & Medical Imaging
GA CR6HF
UT WOS:000361445300007
PM 26377815
OA gold, Green Published
DA 2023-08-27
ER

PT J
AU Jung, SH
   Jung, S
   Moon, KS
   Park, HW
   Kang, SS
AF Jung, Seung-Hoon
   Jung, Shin
   Moon, Kyung-Sub
   Park, Hyun-Woong
   Kang, Sam-Suk
TI Tailored Surgical Approaches for Benign Craniovertebral Junction Tumors
SO JOURNAL OF KOREAN NEUROSURGICAL SOCIETY
LA English
DT Article
DE Benign tumor; Complication; Craniovertebral junction; Neuroradiology;
   Surgical resection
ID FORAMEN MAGNUM; CRANIOCERVICAL JUNCTION; TRANSCONDYLAR APPROACH;
   ANTERIOR PORTION; LATERAL APPROACH; MANAGEMENT; MENINGIOMAS;
   HEMANGIOBLASTOMAS; REMOVAL; LESIONS
AB Objective : We report our surgical experience in the treatment of 16 consecutive patients with benign craniovertebral junction (CVJ) tumor, observed from 2003 to 2008 at our department.
   Methods : We had treated 6 foramen magnum meningiomas, 6 cervicomedullary hemangioblastomas, 1 accessory nerve schwannoma, 1 hypoglossal nerve schwannoma, 1 C2 root schwannoma, and 1 cavernous hemangioma. Clinical results were evaluated by Karnofsky Performance Scale (KPS) and all patients underwent preoperative neuroradiological evaluation with computed tomography (CT) and magnetic resonance image (MRI). Angiography was performed in 15 patients and preoperative embolization was done in 2 patients.
   Results: Five far-lateral, 1 supracondylar and 10 midline suboccipital approaches were performed. Gross total removal was achieved in 15 cases (94%) and subtotal removal in 1 patient (6%). None of the patients required occipitocervical fusion. Radiological follow-up showed no recurrence in cases totally removed. Postoperative decrease of KPS scores was recorded in only 1 patient. The treatment of cervicomedullary solid hemangioblastoma presented particular issues : by preoperative embolization, we removed tumor totally without an excessive bleeding or brainstem injury. In one of foramen magnum meningioma, we carried out subtotal removal due to hard tumor consistency and encasement of neurovascular structures.
   Conclusion : The choice of surgical approaches and the extent of bone resection should be defined according to the location and size of individual tumors. Moreover, we emphasize that preoperative neuroradiological evaluations on presumptive tumor type could be helpful to the surgeon in tailoring the technique and providing the required exposure for different lesions, without unnecessary surgical steps.
C1 Chonnam Natl Univ, Dept Neurosurg, Hwasun Hosp, Kwangju, South Korea.
   Sch Med, Kwangju, South Korea.
C3 Chonnam National University
RP Jung, S (通讯作者)，Chonnam Natl Univ, Dept Neurosurg, Hwasun Hosp, 160 Ilsim Ri, Hwasun Eup 519809, Hwasun, South Korea.
EM
FU Center for Biomedical Human Resources at Chonnam National University
FX This work was supported by the Brain Korea 21 Project, Center for
   Biomedical Human Resources at Chonnam National University.
CR BABU RP, 1994, J NEUROSURG, V81, P49, DOI 10.3171/jns.1994.81.1.0049
   BERTALANFFY H, 1991, NEUROSURGERY, V29, P815, DOI 10.1227/00006123-199112000-00002
   Boulton Melfort R, 2003, Neurosurg Focus, V14, pe10
   Bruneau M, 2008, NEUROSURG REV, V31, P19, DOI 10.1007/s10143-007-0097-1
   Bulsara KR, 2008, J NEUROSURG, V109, P794, DOI 10.3171/JNS/2008/109/11/0794
   Catapano D, 2005, NEUROSURGERY, V56, P1215, DOI 10.1227/01.NEU.0000159646.15026.D6
   CHALJUB G, 1992, AM J ROENTGENOL, V159, P1069, DOI 10.2214/ajr.159.5.1414777
   DEOLIVEIRA E, 1985, SURG NEUROL, V24, P293, DOI 10.1016/0090-3019(85)90042-4
   GEORGE B, 1988, SURG NEUROL, V29, P484, DOI 10.1016/0090-3019(88)90145-0
   Guidetti B, 1988, Adv Tech Stand Neurosurg, V16, P83
   HAKUBA A, 1977, Neurologia Medico-Chirurgica, V17, P63, DOI 10.2176/nmc.17pt1.63
   HAMMON WM, 1972, J NEUROSURG, V37, P339, DOI 10.3171/jns.1972.37.3.0339
   HEROS RC, 1986, J NEUROSURG, V64, P559, DOI 10.3171/jns.1986.64.4.0559
   HO KL, 1981, J NEUROSURG, V55, P132, DOI 10.3171/jns.1981.55.1.0132
   KRATIMENOS GP, 1993, BRIT J NEUROSURG, V7, P129, DOI 10.3109/02688699309103469
   M Samii, 1996, NEUROSURGERY, V39, P1094
   Malis LI, 2002, J NEUROSURG, V97, P1, DOI 10.3171/spi.2002.97.1.0001
   Mathiesen T, 2003, J NEUROSURG, V99, P31, DOI 10.3171/jns.2003.99.1.0031
   Menezes A H, 1994, Clin Neurosurg, V41, P187
   MILLER E, 1987, NEUROSURGERY, V20, P966, DOI 10.1227/00006123-198706000-00026
   Parlato C, 2003, ACTA NEUROCHIR, V145, P31, DOI 10.1007/s00701-002-1024-4
   Russel DS, 1989, PATHOLOGY TUMOURS NE
   Samii M, 1996, NEUROSURGERY, V39, P1086, DOI 10.1097/00006123-199612000-00003
   SEN CN, 1991, ACTA NEUROCHIR, V108, P70, DOI 10.1007/BF01407670
   STEIN BM, 1963, J NEUROSURG, V20, P740, DOI 10.3171/jns.1963.20.9.0740
   Winestone JS, 2007, CHILD NERV SYST, V23, P761, DOI 10.1007/s00381-006-0279-0
NR 26
TC 7
Z9 10
U1 0
U2 2
PU KOREAN NEUROSURGICAL SOC
PI SEOUL
PA #402 POSCO # OFFICE BUILDING 151 SUNHWA-DONG, JUNG-GU, SEOUL, 100-130,
   SOUTH KOREA
SN 2005-3711
EI 1598-7876
J9 J KOREAN NEUROSURG S
JI J. Korean. Neurosurg. Soc.
PD AUG
PY 2010
VL 48
IS 2
BP 139
EP 144
DI 10.3340/jkns.2010.48.2.139
PG 6
WC Clinical Neurology; Surgery
WE Science Citation Index Expanded (SCI-EXPANDED)
SC Neurosciences & Neurology; Surgery
GA 650WX
UT WOS:000281886100009
PM 20856663
OA gold, Green Published
DA 2023-08-27
ER

PT J
AU Kreydiyyeh, SI
   Markossian, S
AF Kreydiyyeh, SI
   Markossian, S
TI Tumor necrosis factor alpha down-regulates the Na+-K+ ATPase and the
   Na+-K(+)2Cl(-) cotransporter in the kidney cortex and medulla
SO CYTOKINE
LA English
DT Article
DE TNF-alpha; PGE(2); Na+-K(+)2Cl(-); Na+-K(+)ATPase; kidney
ID THICK ASCENDING LIMB; CL COTRANSPORTER; BETA-SUBUNIT; CYCLOOXYGENASE-2
   EXPRESSION; MOLECULAR CHARACTERIZATION; NA-K-2CL COTRANSPORTER;
   CYCLIC-AMP; RAT; PHOSPHORYLATION; LOCALIZATION
AB The effect of TNF-alpha on the renal Na+-K+ pump and the Na+-K(+)2Cl(-) cotransporter was investigated in the rat. Animals were injected with the cytokine, and 4 h later, a homogenate from the cortical and medullary tissues was prepared and used to assay the activity of the Na+-K+ ATPase and the protein expression of the pump and symporter. TNF-a reduced the activity and expression of the pump in both cortex and medulla, and its effect disappeared when animals were pre-treated with indomethacin, suggesting that TNF-a acts via PGE(2). Higher levels of PGE(2) were detected by enzyme immunoassay, in kidney tissues isolated from rats treated with PGE(2), thus confirming this hypothesis. The cytokine also down-regulated the Na+-K(+)2Cl(-) cotransporter but this effect was not abrogated by indomethacin. PGE(2), injected into animals, exerted a dose-dependent effect. Low doses did not have any effect on the two transporters in the cortex while high doses inhibited and down-regulated the pump and up-regulated the cotransporter. In the medulla low doses increased the activity and expression of the pump but down-regulated the cotransporter while high doses exerted an exactly opposite effect on the two transporters. It was concluded that the effect of TNF-alpha on the pump is mediated via PGE(2) which is released at relatively high doses. The effect of the cytokine on the cotransporter is, however, independent of PGE(2). (c) 2006 Elsevier Ltd. All rights reserved.
C1 Amer Univ Beirut, Dept Biol, Fac Arts & Sci, Beirut, Lebanon.
C3 American University of Beirut
RP Kreydiyyeh, SI (通讯作者)，Amer Univ Beirut, Dept Biol, Fac Arts & Sci, Beirut, Lebanon.
EM
OI Kreydiyyeh, Sawsan/0000-0002-4149-4081
CR Baert FJ, 1999, GASTROENTEROLOGY, V116, P22, DOI 10.1016/S0016-5085(99)70224-6
   BASTIEN L, 1994, J BIOL CHEM, V269, P11873
   BATSHAKE B, 1995, EUR J BIOCHEM, V231, P809, DOI 10.1111/j.1432-1033.1995.0809d.x
   BENADOR NM, 1995, MICROVASC RES, V50, P154, DOI 10.1006/mvre.1995.1049
   BEUTLER BA, 1985, J IMMUNOL, V135, P3972
   Bhattacharya M, 1998, P NATL ACAD SCI USA, V95, P15792, DOI 10.1073/pnas.95.26.15792
   Breyer MD, 1998, KIDNEY INT, V54, pS88, DOI 10.1046/j.1523-1755.1998.06718.x
   Breyer MD, 2000, CURR OPIN NEPHROL HY, V9, P23, DOI 10.1097/00041552-200001000-00005
   Bryer MD, 2001, ANNU REV PHYSIOL, V63, P579, DOI 10.1146/annurev.physiol.63.1.579
   Carranza ML, 1998, J PHYSIOL-LONDON, V511, P235, DOI 10.1111/j.1469-7793.1998.235bi.x
   Cheng XJ, 1997, AM J PHYSIOL-CELL PH, V273, pC1981, DOI 10.1152/ajpcell.1997.273.6.C1981
   Chibalin AV, 1998, J BIOL CHEM, V273, P8814, DOI 10.1074/jbc.273.15.8814
   Chibalin AV, 1999, J BIOL CHEM, V274, P1920, DOI 10.1074/jbc.274.4.1920
   Cunningham PN, 2002, J IMMUNOL, V168, P5817, DOI 10.4049/jimmunol.168.11.5817
   DECOLLOGNE S, 1993, J CARDIOVASC PHARM, V22, pS96, DOI 10.1097/00005344-199322002-00031
   Ecelbarger CA, 1996, AM J PHYSIOL-RENAL, V271, pF619, DOI 10.1152/ajprenal.1996.271.3.F619
   Ferreri NR, 1999, AM J PHYSIOL-RENAL, V277, pF360, DOI 10.1152/ajprenal.1999.277.3.F360
   GAMBA G, 1994, J BIOL CHEM, V269, P17713
   Geering K, 1996, J CELL BIOL, V133, P1193, DOI 10.1083/jcb.133.6.1193
   Guan YF, 1998, J CLIN INVEST, V102, P194, DOI 10.1172/JCI2872
   Haas M, 2000, ANNU REV PHYSIOL, V62, P515, DOI 10.1146/annurev.physiol.62.1.515
   IGARASHI P, 1995, AM J PHYSIOL-RENAL, V269, pF405, DOI 10.1152/ajprenal.1995.269.3.F405
   KANSRA V, 1995, EUR J PHARM-MOLEC PH, V289, P391, DOI 10.1016/0922-4106(95)90119-1
   Kaplan MR, 1996, KIDNEY INT, V49, P40, DOI 10.1038/ki.1996.6
   Kim GH, 1999, AM J PHYSIOL-RENAL, V276, pF96, DOI 10.1152/ajprenal.1999.276.1.F96
   KIRLEY TL, 1989, J BIOL CHEM, V264, P7185
   LINGREL JB, 1990, PROG NUCLEIC ACID RE, V38, P37
   Lopina OD, 2001, BIOCHEMISTRY-MOSCOW+, V66, P1122, DOI 10.1023/A:1012432913689
   MACICA CM, 1994, KIDNEY INT, V46, P113, DOI 10.1038/ki.1994.250
   MCDONOUGH AA, 1994, AM J PHYSIOL-CELL PH, V267, pC901, DOI 10.1152/ajpcell.1994.267.4.C901
   Morisset S, 1998, J RHEUMATOL, V25, P1146
   Obermuller N, 1996, J CLIN INVEST, V98, P635, DOI 10.1172/JCI118834
   PALFREY HC, 1993, PFLUG ARCH EUR J PHY, V425, P321, DOI 10.1007/BF00374182
   Patrizio M, 2004, GLIA, V48, P241, DOI 10.1002/glia.20074
   PAYNE JA, 1995, CURR OPIN CELL BIOL, V7, P493, DOI 10.1016/0955-0674(95)80005-0
   REGAN JW, 1994, MOL PHARMACOL, V46, P213
   Saklatvala J, 1999, BIOCHEM SOC SYMP, P63
   SATOH T, 1992, J CLIN INVEST, V89, P1496, DOI 10.1172/JCI115740
   SATOH T, 1993, AM J PHYSIOL, V265, pF399, DOI 10.1152/ajprenal.1993.265.3.F399
   Stewart WC, 1998, GLIA, V23, P349, DOI 10.1002/(SICI)1098-1136(199808)23:4<349::AID-GLIA7>3.0.CO;2-Y
   SUGIMOTO Y, 1994, AM J PHYSIOL, V266, pF823, DOI 10.1152/ajprenal.1994.266.5.F823
   TAKEMOTO F, 1992, PFLUG ARCH EUR J PHY, V421, P302, DOI 10.1007/BF00374216
   TAUSSKY HH, 1953, J BIOL CHEM, V202, P675
   VANDEVENTER SJH, 1990, BLOOD, V76, P2520
   VANLANSCHOT JJB, 1991, SURG GYNECOL OBSTET, V172, P293
   Vasilets LA, 1997, ANN NY ACAD SCI, V834, P585, DOI 10.1111/j.1749-6632.1997.tb52326.x
   WINTERS CJ, 1991, SEMIN NEPHROL, V11, P236
   Yang TX, 1996, AM J PHYSIOL-RENAL, V271, pF931, DOI 10.1152/ajprenal.1996.271.4.F931
NR 48
TC 18
Z9 18
U1 0
U2 2
PU ACADEMIC PRESS LTD- ELSEVIER SCIENCE LTD
PI LONDON
PA 24-28 OVAL RD, LONDON NW1 7DX, ENGLAND
SN 1043-4666
EI 1096-0023
J9 CYTOKINE
JI Cytokine
PD FEB 7
PY 2006
VL 33
IS 3
BP 138
EP 144
DI 10.1016/j.cyto.2005.12.007
PG 7
WC Biochemistry & Molecular Biology; Cell Biology; Immunology
WE Science Citation Index Expanded (SCI-EXPANDED)
SC Biochemistry & Molecular Biology; Cell Biology; Immunology
GA 027VP
UT WOS:000236444200003
PM 16488623
DA 2023-08-27
ER

PT J
AU Kushlinskii, NE
   Britvin, TA
   Polyakova, GA
   Abbasova, SG
   Baronin, AA
   Bogatyrev, OP
   Kalinin, AP
   Lipkin, VM
AF Kushlinskii, NE
   Britvin, TA
   Polyakova, GA
   Abbasova, SG
   Baronin, AA
   Bogatyrev, OP
   Kalinin, AP
   Lipkin, VM
TI Soluble fas antigen (sFAS) in the serum from patients with adrenal
   tumors
SO BULLETIN OF EXPERIMENTAL BIOLOGY AND MEDICINE
LA English
DT Article
DE apoptosis; soluble Fas antigen; adrenal tumors; diffuse-and-nodular
   adrenal cortex hyperplasia
ID APOPTOSIS; LIGAND; MOLECULE; DEATH
AB Serum content of soluble Fas antigen was measured by enzyme immunoassay in 60 healthy donors, 31 patients with adrenal tumors, and 16 patients with diffuse-and-nodular hyperplasia of the adrenal cortex accompanying primary aldosteronism and Cushing's disease. sFas was more often detected in the serum from patients with tumors of the adrenal cortex and medulla and diffuse-and-nodular hyperplasia of the adrenal cortex and its content varied in a wider range in patients compared to healthy donors. No correlations were found between the incidence of sFas, its content, sex, and age of healthy donors and patients. The highest content of sFas was found in patients with pheochromocytoma and primary aldosteronism. sFas probably plays a role in the pathogenesis of adrenal tumors and hyperplasia.
C1 Russian Acad Med Sci, NN Blokhin Oncol Res Ctr, Moscow 115478, Russia.
   MF Vladimirskii Reg Clin Res Inst, Moscow, Russia.
   Russian Acad Sci, MM Shemyakin Bioorgan Chem Inst, Moscow, Russia.
