## Supplemental Material 1 for "Bibliometric analysis of emerging trends and research foci in brainstem tumor field over thirty years (1992-2023)": savedrecs (4).txt

FN Clarivate Analytics Web of Science
VR 1.0
PT J
AU Grigoryan, YA
   Onopchenko, CV
AF Grigoryan, YA
   Onopchenko, CV
TI Persistent trigeminal neuralgia after removal of contralateral posterior
   cranial fossa tumor. Report of two cases.
SO SURGICAL NEUROLOGY
LA English
DT Article
DE trigeminal neuralgia; false localizing sign; contralateral mass;
   rhizotomy; facial pain
ID FALSE LOCALIZING SIGN; INTRACRANIAL TUMORS; FACIAL-PAIN; MASS
AB BACKGROUND Contralateral trigeminal neuralgia as a false localizing sign in patients with posterior cranial fossa tumors is rare. Persistent contralateral trigeminal neuralgia after removal of the posterior fossa expanding lesion with microsurgical exploration of the affected trigeminal nerve root has been described in only a few reports. Displacement of the brainstem and the trigeminal nerve root, arachnoid adhesions, and vascular compression of the nerve root entry zone have been reported as causes of persistent contralateral trigeminal neuralgia.
   METHODS One patient developed transformation of the contralateral constant burning facial pain into trigeminal neuralgia after removal of a posterior fossa meningioma. A typical right-sided tic douloureux in our second patient did not disappear after removal of a left acoustic neurinoma. CT scan revealed brainstem displacement to the side of trigeminal neuralgia. Microsurgical exploration in both cases demonstrated the squeezed and distorted trigeminal nerve root and displaced brain stem with no vascular involvement. Both patients underwent partial trigeminal rhizotomy for pain control.
   RESULTS Complete disappearance of the trigeminal neuralgia was evident in both cases with postoperative facial sensory loss. The postoperative course in the first case was uneventful; the second patient died from purulent meningoencephalitis.
   CONCLUSION Persistent contralateral trigeminal neuralgia after removal of a posterior fossa tumor is caused by distortion of the fifth nerve root by the displaced brainstem. Partial trigeminal rhizotomy can be performed for alleviation of facial neuralgic pain in cases without neurovascular compression. (C) 1999 by Elsevier Science Inc.
C1 Russian Med Acad Postgrad Educ, Botkin Hosp, Dept Neurosurg, Moscow 125101, Russia.
C3 Ministry of Health of the Russian Federation
RP Grigoryan, YA (通讯作者)，Russian Med Acad Postgrad Educ, Botkin Hosp, Dept Neurosurg, 2nd Botkinski Pr 5, Moscow 125101, Russia.
OI Grigoryan, Yuri/0000-0003-3157-6187
CR Barker FG, 1996, J NEUROSURG, V84, P818, DOI 10.3171/jns.1996.84.5.0818
   BULLITT E, 1986, J NEUROSURG, V64, P865, DOI 10.3171/jns.1986.64.6.0865
   Cappabianca P, 1997, J NEUROSURG, V86, P171
   CHENG TMW, 1993, NEUROLOGY, V43, P2298, DOI 10.1212/WNL.43.11.2298
   FLORENSA R, 1987, NEUROSURGERY, V20, P1
   HADDAD FS, 1990, NEUROSURGERY, V26, P1033, DOI 10.1227/00006123-199006000-00019
   HAMBY WB, 1947, J NEUROSURG, V4, P179, DOI 10.3171/jns.1947.4.2.0179
   HOFFMAN RA, 1979, NEW YORK STATE J MED, V79, P1436
   JANNETTA PJ, 1990, TRIGEMINAL NEURALGIA, P201
   KOENIG M, 1984, NEUROSURGERY, V14, P335, DOI 10.1227/00006123-198403000-00014
   LEVIN P, 1968, J MT SINAI HOSP, V35, P343
   Matsuura N, 1996, J NEUROSURG, V84, P1067, DOI 10.3171/jns.1996.84.6.1067
   MAURICEWILLIAMS RS, 1975, J NEUROL NEUROSUR PS, V38, P1232, DOI 10.1136/jnnp.38.12.1232
   MICHELUCCI R, 1989, J NEUROL NEUROSUR PS, V52, P1202, DOI 10.1136/jnnp.52.10.1202
   OCONNELL JEA, 1978, BRAIN, V101, P119, DOI 10.1093/brain/101.1.119
   PAILLAS JE, 1969, REV NEUROL-FRANCE, V121, P452
   Parker HL, 1937, J NEUROL PSYCHOPATHO, V17, P256
   SNOW RB, 1987, NEUROSURGERY, V21, P84, DOI 10.1227/00006123-198707000-00018
   Stechison MT, 1996, NEUROSURGERY, V38, P76, DOI 10.1097/00006123-199601000-00018
   YANG DY, 1989, CHUNG HUA I HSUEH TS, V44, P135
NR 20
TC 14
Z9 15
U1 0
U2 1
PU ELSEVIER SCIENCE INC
PI NEW YORK
PA 655 AVENUE OF THE AMERICAS, NEW YORK, NY 10010 USA
SN 0090-3019
J9 SURG NEUROL
JI Surg. Neurol.
PD JUL
PY 1999
VL 52
IS 1
BP 56
EP 60
DI 10.1016/S0090-3019(99)00050-6
PG 5
WC Clinical Neurology; Surgery
WE Science Citation Index Expanded (SCI-EXPANDED)
SC Neurosciences & Neurology; Surgery
GA 209FD
UT WOS:000081036800031
PM 10390175
DA 2023-08-27
ER

PT J
AU Totsune, K
   Takahashi, K
   Murakami, O
   Satoh, F
   Sone, M
   Ohneda, M
   Miura, Y
   Mouri, T
AF Totsune, K
   Takahashi, K
   Murakami, O
   Satoh, F
   Sone, M
   Ohneda, M
   Miura, Y
   Mouri, T
TI Immunoreactive brain natriuretic peptide in human adrenal glands and
   adrenal tumors
SO EUROPEAN JOURNAL OF ENDOCRINOLOGY
LA English
DT Article
ID SEQUENCE DETERMINATION; CARDIAC ATRIUM; RAT; MEDULLA; TISSUES; CELLS;
   PHEOCHROMOCYTOMAS; STEROIDOGENESIS; IDENTIFICATION; LOCALIZATION
AB The presence of brain natriuretic peptide (BNP) ih tissues of human adrenal glands and adrenal tumors was investigated by radioimmunoassay. Immunoreactive BNP concentrations were 0.203+/-0.061 pmol/g wet tissue (mean+/-SEM) in normal parts of adrenal glands (cortex and medulla, N=8), 0.205+/-0.037 pmol/g wet tissue in pheochromocytomas (N=8), 0.230+/-0.062 pmol/g wet tissue in aldosteronomas (N=11) and 0.180+/-0.054 pmol/g wet tissue in adrenocortical adenomas with Cushings syndrome (N=4). Sephadex G-50 superfine column chromatography and reverse-phase high-performance liquid chromatography showed that most (>70%) of the immunoreactive BNP in the normal part of adrenal glands was eluted in the position of human BNP-32. Sephadex G-50 superfine column chromatography of immunoreactive BNP in the pheochromocytoma and aldosteronoma showed four peaks: one in the position of gamma-BNP, one in the position of BNP-32, one between gamma-BNP and BNP-32 and one in the smaller molecular weight region, The present study has shown that immunoreactive BNP is present both in normal human adrenal glands and in adrenal tumors, Multiple molecular forms of BNP were found to be present in the tumor tissues of pheochromocytoma and aldosteronoma.
C1 TOHOKU UNIV, SCH MED, DEPT APPL PHYSIOL & MOL BIOL, AOBA KU, SENDAI, MIYAGI 98077, JAPAN.
   TOHOKU UNIV, SCH MED, DEPT INTERNAL MED 2, SENDAI, MIYAGI 98077, JAPAN.
   TOHOKU UNIV, SCH MED, STUDENT HLTH CTR, SENDAI, MIYAGI 98077, JAPAN.
   TOHOKU UNIV, SCH MED, DEPT PATHOL 2, SENDAI, MIYAGI 98077, JAPAN.
C3 Tohoku University; Tohoku University; Tohoku University; Tohoku
   University
RI Takahashi, Kazuhiro/A-4887-2010
OI Takahashi, Kazuhiro/0000-0001-8943-1875
CR ADRIAN TE, 1983, LANCET, V2, P540
   DELEAN A, 1984, ENDOCRINOLOGY, V115, P1636
   DELEAN A, 1985, LIFE SCI, V36, P2375, DOI 10.1016/0024-3205(85)90341-8
   GERBES AL, 1994, J CLIN ENDOCR METAB, V78, P1307, DOI 10.1210/jc.78.6.1307
   HASHIGUCHI T, 1989, CLIN ENDOCRINOL, V31, P623, DOI 10.1111/j.1365-2265.1989.tb01287.x
   INAGAKI S, 1986, REGUL PEPTIDES, V15, P249, DOI 10.1016/0167-0115(86)90066-2
   KAMBAYASHI Y, 1990, FEBS LETT, V259, P341, DOI 10.1016/0014-5793(90)80043-I
   KAMBAYASHI Y, 1989, BIOCHEM BIOPH RES CO, V163, P233, DOI 10.1016/0006-291X(89)92126-8
   KOJIMA M, 1989, BIOCHEM BIOPH RES CO, V159, P1420, DOI 10.1016/0006-291X(89)92268-7
   KONRAD EM, 1992, REGUL PEPTIDES, V39, P177, DOI 10.1016/0167-0115(92)90539-7
   KUDO T, 1984, NATURE, V312, P756, DOI 10.1038/312756a0
   LEE YJ, 1994, J CLIN ENDOCR METAB, V79, P1476, DOI 10.1210/jc.79.5.1476
   MCKENZIE JC, 1985, J HISTOCHEM CYTOCHEM, V33, P828, DOI 10.1177/33.8.3160763
   MINAMINO N, 1988, BIOCHEM BIOPH RES CO, V157, P402, DOI 10.1016/S0006-291X(88)80061-5
   MOREL G, 1988, ENDOCRINOLOGY, V123, P149, DOI 10.1210/endo-123-1-149
   MOURI T, 1989, PEPTIDES, V10, P201, DOI 10.1016/0196-9781(89)90097-1
   MUKOYAMA M, 1990, LANCET, V335, P801, DOI 10.1016/0140-6736(90)90925-U
   NARUSE M, 1994, HYPERTENSION, V23, pI231, DOI 10.1161/01.HYP.23.1_Suppl.I231
   NAWATA H, 1991, J STEROID BIOCHEM, V40, P367, DOI 10.1016/0960-0760(91)90204-I
   NGUYEN TT, 1988, FEBS LETT, V231, P393, DOI 10.1016/0014-5793(88)80857-3
   OGAWA Y, 1990, ENDOCRINOLOGY, V126, P2225, DOI 10.1210/endo-126-4-2225
   OHASHI M, 1991, Fukuoka Acta Medica, V82, P131
   SCAFF Z, 1987, J EXP PATHOL, V3, P229
   SONE M, 1991, J CARDIOVASC PHARM, V17, pS427, DOI 10.1097/00005344-199100177-00122
   SUDOH T, 1988, NATURE, V332, P78, DOI 10.1038/332078a0
   SUDOH T, 1989, BIOCHEM BIOPH RES CO, V159, P1427, DOI 10.1016/0006-291X(89)92269-9
   TAKAHASHI K, 1992, PEPTIDES, V13, P121, DOI 10.1016/0196-9781(92)90149-W
   TATEYAMA H, 1990, BIOCHEM BIOPH RES CO, V166, P1080, DOI 10.1016/0006-291X(90)90977-U
   TOTSUNE K, 1994, HYPERTENSION, V24, P758, DOI 10.1161/01.HYP.24.6.758
   TOTSUNE K, 1994, PEPTIDES, V15, P287, DOI 10.1016/0196-9781(94)90014-0
   TOTSUNE K, 1996, IN PRESS REGUL PEPT
   WILCOX JN, 1991, MOL CELL BIOL, V11, P3454, DOI 10.1128/MCB.11.7.3454
   YANAGIHARA N, 1991, N-S ARCH PHARMACOL, V343, P289
NR 33
TC 13
Z9 14
U1 0
U2 1
PU BIOSCIENTIFICA LTD
PI BRISTOL
PA EURO HOUSE, 22 APEX COURT WOODLANDS, BRADLEY STOKE, BRISTOL BS32 4JT,
   ENGLAND
SN 0804-4643
EI 1479-683X
J9 EUR J ENDOCRINOL
JI Eur. J. Endocrinol.
PD SEP
PY 1996
VL 135
IS 3
BP 352
EP 356
DI 10.1530/eje.0.1350352
PG 5
WC Endocrinology & Metabolism
WE Science Citation Index Expanded (SCI-EXPANDED)
SC Endocrinology & Metabolism
GA VN031
UT WOS:A1996VN03100016
PM 8890728
DA 2023-08-27
ER

PT J
AU Chang, SW
   Wu, A
   Gore, P
   Beres, E
   Porter, RW
   Preul, MC
   Spetzler, RF
   Bambakidis, NC
AF Chang, Steve W.
   Wu, Anhua
   Gore, Pankaj
   Beres, Elisa
   Porter, Randall W.
   Preul, Mark C.
   Spetzler, Robert F.
   Bambakidis, Nicholas C.
TI QUANTITATIVE COMPARISON OF KAWASE'S APPROACH VERSUS THE RETROSIGMOID
   APPROACH: IMPLICATIONS FOR TUMORS INVOLVING BOTH MIDDLE AND POSTERIOR
   FOSSAE
SO NEUROSURGERY
LA English
DT Article
DE Cranial base approaches; Cranial base tumors; Kawase; Meningiomas;
   Retrosigmoid; Schwannomas; Skull base approaches; Skull base tumors
ID TRANSLABYRINTHINE-TRANSTENTORIAL APPROACH; INTRADURAL SUPRAMEATAL
   APPROACH; PETROCLIVAL MENINGIOMAS; INFRATENTORIAL APPROACH;
   TRANSPETROSAL APPROACH; SURGICAL TECHNIQUE; REGION; RESECTION;
   SUPRATENTORIAL; EXPERIENCE
AB OBJECTIVE: Few quantitative data are available to describe Kawase's exposure of the posterior fossa. We used a cadaveric model to compare Kawase's and the retrosigmoid approach to the petroclival region.
   METHODS: Eighteen cadaveric specimens were dissected and analyzed (6 retrosigmoid, 6 Kawase's, and 6 retrosigmoid intradural suprameatal approaches). Clival and brainstem working areas and surgical freedom were measured.
   RESULTS: The retrosigmoid approach provided a significantly larger clival and brainstem working area than Kawase's approach. Surgical freedom at the trigeminal root entry zone, origin of the anterior inferior cerebellar artery, and Dorello's canal was equivalent across approaches. Kawase's approach provided the most surgical freedom at the trigeminal porus. However, the addition of a suprameatal extension significantly improved the surgical freedom provided by the retrosigmoid approach.
   CONCLUSION: The retrosigmoid approach is a powerful approach to lesions of the cerebellopontine angle and ventral brainstem. Lesions involving the trigeminal porus and Meckel's cave can be approached through Kawase's approach or a suprameatal extension of the retrosigmoid approach. Kawase's approach is best suited for accessing middle fossa lesions with smaller petroclival components located above the internal auditory canal.
C1 [Chang, Steve W.; Wu, Anhua; Gore, Pankaj; Beres, Elisa; Porter, Randall W.; Preul, Mark C.; Spetzler, Robert F.; Bambakidis, Nicholas C.] St Josephs Hosp, Barrow Neurol Inst, Div Neurol Surg, Phoenix, AZ 85013 USA.
   [Preul, Mark C.] St Josephs Hosp, Barrow Neurol Inst, Div Neurosurg Res, Phoenix, AZ 85013 USA.
C3 Barrow Neurological Institute; St. Joseph's Hospital and Medical Center;
   Barrow Neurological Institute; St. Joseph's Hospital and Medical Center
RP Bambakidis, NC (通讯作者)，St Josephs Hosp, Barrow Neurol Inst, Div Neurol Surg, 350 W Thomas Rd, Phoenix, AZ 85013 USA.
EM
RI Chang, Steve Fuhwa/HIR-3815-2022
CR Al-Mefty O, 1991, MENINGIOMAS, P517
   ALMEFTY O, 1988, NEUROSURGERY, V22, P510, DOI 10.1227/00006123-198803000-00010
   Aziz KMA, 2000, NEUROSURGERY, V47, P139, DOI 10.1097/00006123-200007000-00030
   BALDWIN HZ, 1994, J NEUROSURG, V81, P60, DOI 10.3171/jns.1994.81.1.0060
   Bambakidis NC, 2007, NEUROSURGERY, V61, P202, DOI [10.1227/01.neu.0000303218.61230.39, 10.1227/01.NIEU.0000280119.36270.BA]
   Chanda A, 2006, NEUROSURGERY, V59, P1, DOI 10.1227/01.NEU.0000220673.79877.30
   David CA, 1999, BARROW Q, V15, P4
   DAY JD, 1994, NEUROSURGERY, V34, P1009, DOI 10.1227/00006123-199406000-00009
   Fukushima T., 1996, MANUAL SKULL BASE DI
   HAKUBA A, 1988, SURG NEUROL, V30, P108, DOI 10.1016/0090-3019(88)90095-X
   HOUSE WF, 1976, ARCH OTOLARYNGOL, V102, P334
   HOUSE WF, 1978, OTOLARYNGOLOGY, V86
   Hsu FPK, 2004, J NEUROSURG, V100, P695, DOI 10.3171/jns.2004.100.4.0695
   Ichimura S, 2008, ACTA NEUROCHIR, V150, P637, DOI 10.1007/s00701-008-1586-x
   Jittapiromsak P, 2009, NEUROSURGERY, V64, pS35, DOI [10.1227/01.NEU.0000334048.96772.A7, 10.1227/01.NFU.0000334048.96722.A7]
   KANZAKI J, 1977, ARCH OTO-RHINO-LARYN, V217, P119, DOI 10.1007/BF00453899
   KAWASE T, 1994, ACTA NEUROCHIR, V129, P113, DOI 10.1007/BF01406489
   KAWASE T, 1991, NEUROSURGERY, V28, P869, DOI 10.1227/00006123-199106000-00014
   KAWASE T, 1985, J NEUROSURG, V63, P857, DOI 10.3171/jns.1985.63.6.0857
   KING TT, 1970, P ROY SOC MED, V63, P780, DOI 10.1177/003591577006300819
   LITTLE AS, 2008, NEUROSURGERY S2, V62, pNS324
   Little KM, 2005, NEUROSURGERY, V56, P546, DOI 10.1227/01.NEU.0000153906.12640.62
   MALIS LI, 1991, CLIN NEUR, V37, P528
   MORRISON AW, 1973, J NEUROSURG, V38, P382, DOI 10.3171/jns.1973.38.3.0382
   Quinones-Hinojosa A, 2006, NEUROSURGERY S2, V58
   Safavi-Abbasi S, 2007, J NEUROSURG, V107, P797, DOI 10.3171/JNS-07/10/0797
   SAMII M, 1989, NEUROSURGERY, V24, P12, DOI 10.1227/00006123-198901000-00003
   Samii M, 1999, J CLIN NEUROSCI, V6, P27, DOI 10.1016/S0967-5868(99)90598-2
   SAMII M, 1988, ACTA NEUROCHIR, V95, P6, DOI 10.1007/BF01793075
   Samii M, 2000, J NEUROSURG, V92, P235, DOI 10.3171/jns.2000.92.2.0235
   SEKHAR LN, 1990, NEUROSURGERY, V27, P764, DOI 10.1227/00006123-199011000-00015
   Sekhar LN, 1996, J NEURO-ONCOL, V29, P249, DOI 10.1007/BF00165655
   Seoane E, 1999, NEUROSURGERY, V44, P553, DOI 10.1097/00006123-199903000-00065
   Siwanuwatn R, 2006, J NEUROSURG, V104, P137, DOI 10.3171/jns.2006.104.1.137
   SPETZLER RF, 1992, J NEUROSURG, V76, P588, DOI 10.3171/jns.1992.76.4.0588
   Suhardja Agustinus, 2003, Neurosurg Focus, V14, pe9
   Van Havenbergh T, 2003, NEUROSURGERY, V52, P55, DOI 10.1097/00006123-200301000-00006
NR 37
TC 46
Z9 49
U1 0
U2 9
PU OXFORD UNIV PRESS INC
PI CARY
PA JOURNALS DEPT, 2001 EVANS RD, CARY, NC 27513 USA
SN 0148-396X
EI 1524-4040
J9 NEUROSURGERY
JI Neurosurgery
PD MAR
PY 2009
VL 64
IS 3
SU S
BP S44
EP S52
DI 10.1227/01.NEU.0000334410.24984.DD
PG 9
WC Clinical Neurology; Surgery
WE Science Citation Index Expanded (SCI-EXPANDED)
SC Neurosciences & Neurology; Surgery
GA 411MN
UT WOS:000263653800006
PM 19240572
DA 2023-08-27
ER

PT J
AU Giagnacovo, M
   Antonelli, M
   Biassoni, V
   Schiavello, E
   Warmuth-Metz, M
   Buttarelli, FR
   Modena, P
   Massimino, M
AF Giagnacovo, Marzia
   Antonelli, Manila
   Biassoni, Veronica
   Schiavello, Elisabetta
   Warmuth-Metz, Monika
   Buttarelli, Francesca R.
   Modena, Piergiorgio
   Massimino, Maura
TI Retrospective analysis on the consistency of MRI features with
   histological and molecular markers in diffuse intrinsic pontine glioma
   (DIPG)
SO CHILDS NERVOUS SYSTEM
LA English
DT Article
DE DIPG; Glioma
ID HISTONE H3.3; MUTATIONS; REIRRADIATION; SUBGROUPS; CHILDREN
AB Purpose The diagnosis of diffuse intrinsic pontine glioma (DIPG) is based largely on a combination of clinical and radiological findings due to the difficulty of obtaining a biopsy. An accurate evaluation of magnetic resonance imaging (MRI) scans is consequently essential. Recent analyses on the genomic landscape of DIPG revealed recurrent mutations in the H3F3A and HIST1H3B histone genes. We reviewed cases with available tumor tissue from institutional DIPG series to ascertain the consistency between their histo-molecular findings and clinical-radiological features. Methods We conducted a radiological and pathological central review of 22 cases enrolled in institutional DIPG trials. We performed immunohistochemical analyses to detect H3F3A/HIST1H3B K27M mutations, histone trimethylation, and EZH2 expression. Mutational analysis was performed for ACVR1, H3F3A, and HIST1H3B genes. Results Patients' median age at diagnosis was 8 years, and their median overall survival was 11 months. Nineteen/22 cases (86%) showed evidence of K27M mutation on immunohistochemistry and/or mutation analysis. Histone trimethylation expression was low or lacking in these mutated cases. Sequence analysis revealed 13 cases with H3F3A and 1 case with HIST1H3B K27M mutation. There was no significant difference in EZH2 expression between the K27M mutant and wild-type DIPGs. Upon external, blinded MRI re-evaluation one lesion not consistent with DIPG showed no evidence of K27M mutation and retained histone trimethylation expression. Conclusion In conclusion, our study demonstrates a high frequency of histone K27M mutations in DIPG when MRI features are carefully assessed, thus confirming the consistency of imaging with biological markers in our institutional series of DIPG.
C1 [Giagnacovo, Marzia; Modena, Piergiorgio] ASST Lariana Gen Hosp, Genet Unit, Como, Italy.
   [Antonelli, Manila; Buttarelli, Francesca R.] Sapienza Univ, Dept Radiol Oncol & Anatomopathol Sci, Rome, Italy.
   [Biassoni, Veronica; Schiavello, Elisabetta; Massimino, Maura] Fdn IRCCS Ist Nazl Tumori, Pediat Oncol Unit, Milan, Italy.
   [Warmuth-Metz, Monika] Wurzburg Univ Hosp, Reference Ctr Neuroradiol, Wurzburg, Germany.
C3 Sapienza University Rome; Fondazione IRCCS Istituto Nazionale Tumori
   Milan; University of Wurzburg
RP Modena, P (通讯作者)，ASST Lariana Gen Hosp, Genet Unit, Como, Italy.; Massimino, M (通讯作者)，Fdn IRCCS Ist Nazl Tumori, Pediat Oncol Unit, Milan, Italy.
EM
RI Schiavello, Elisabetta ES/K-8091-2016; massimino, maura/K-7310-2016
OI Schiavello, Elisabetta ES/0000-0002-8049-7445; massimino,
   maura/0000-0002-5506-2001
CR Castel D, 2015, ACTA NEUROPATHOL, V130, P815, DOI 10.1007/s00401-015-1478-0
   Deb G, 2014, MOL CANCER RES, V12, P639, DOI 10.1158/1541-7786.MCR-13-0546
   Donaldson SS, 2006, J CLIN ONCOL, V24, P1266, DOI 10.1200/JCO.2005.04.6599
   Fleischhack G, 2019, J NEURO-ONCOL, V143, P107, DOI 10.1007/s11060-019-03140-z
   Gianno F, 2018, GLIOMA, V1, P117, DOI [DOI 10.4103/GLIOMA.GLIOMA_27_18, 10.4103/glioma.glioma_27_18]
   Hargrove D, 2006, LANCET ONCOL, V7, P241, DOI 10.1016/S1470-2045(06)70615-5
   Hoffman LM, 2018, J CLIN ONCOL, V36, P1963, DOI 10.1200/JCO.2017.75.9308
   Janssens GO, 2017, EUR J CANCER, V73, P38, DOI 10.1016/j.ejca.2016.12.007
   Lober RM, 2014, J NEURO-ONCOL, V117, P175, DOI 10.1007/s11060-014-1375-8
   Louis DN, 2016, ACTA NEUROPATHOL, V131, P803, DOI 10.1007/s00401-016-1545-1
   Massimino M, 2008, J NEURO-ONCOL, V87, P355, DOI 10.1007/s11060-008-9525-5
   Massimino M, 2014, J NEURO-ONCOL, V118, P305, DOI 10.1007/s11060-014-1428-z
   Schwartzentruber J, 2012, NATURE, V482, P226, DOI [10.1038/nature10833, 10.1007/s13311-012-0137-6]
   Shanmugam M. K, 2017, ONCOTARGET, V9, P11414
   Taylor KR, 2014, NAT GENET, V46, P457, DOI 10.1038/ng.2925
   van Zanten SEMV, 2017, J NEURO-ONCOL, V132, P255, DOI 10.1007/s11060-016-2363-y
   Wu G, 2012, NAT GENET, V44, P251, DOI 10.1038/ng.1102
   Yuen BTK, 2013, CANCER CELL, V24, P567, DOI 10.1016/j.ccr.2013.09.015
NR 18
TC 10
Z9 12
U1 0
U2 5
PU SPRINGER
PI NEW YORK
PA ONE NEW YORK PLAZA, SUITE 4600, NEW YORK, NY, UNITED STATES
SN 0256-7040
EI 1433-0350
J9 CHILD NERV SYST
JI Childs Nerv. Syst.
PD APR
PY 2020
VL 36
IS 4
BP 697
EP 704
DI 10.1007/s00381-019-04463-y
PG 8
WC Clinical Neurology; Pediatrics; Surgery
WE Science Citation Index Expanded (SCI-EXPANDED)
SC Neurosciences & Neurology; Pediatrics; Surgery
GA KU4FE
UT WOS:000519664600006
PM 31848724
DA 2023-08-27
ER

PT J
AU Simmons, JH
   Shoemaker, AH
   Roth, CL
AF Simmons, Jill H.
   Shoemaker, Ashley H.
   Roth, Christian L.
TI Treatment with Glucagon-Like Peptide-1 Agonist Exendin-4 in a Patient
   with Hypothalamic Obesity Secondary to Intracranial Tumor
SO HORMONE RESEARCH IN PAEDIATRICS
LA English
DT Article
DE Hypothalamic lesion; Exenatide; Glucagon-like peptide-1; Body weight;
   Food intake; Germ cell tumor
ID GASTRIC BYPASS-SURGERY; EXENATIDE EXENDIN-4; GLYCEMIC CONTROL; CHILDHOOD
   CRANIOPHARYNGIOMA; TREATED PATIENTS; RISK-FACTORS; WEIGHT-LOSS;
   CHILDREN; THERAPY; GLP-1
AB Background/Aims: Patients with hypothalamic tumors frequently experience severe obesity, and its treatment with diet, exercise, and/or pharmacologic treatment has had limited effect. Glucagon-like peptide-1 agonist exenatide (exendin-4), used for treatment of type 2 diabetes, causes persistent weight loss via signaling in the brainstem. Methods: We report the case of a 17-year-old patient with obesity resulting from a hypothalamic germ cell tumor. He was treated by chemoradiotherapy and exenatide at a dose of 5 mu g subcutaneously twice daily. Results: Exenatide resulted in a 29-kg weight loss (BMI reduction from 37.1 to 29.1) after 2.5 years of treatment; significant weight gain occurred shortly after exenatide was discontinued. Conclusion: Exenatide resulted in considerable reduction of body weight in a patient with severe hypothalamic obesity. This novel observation requires follow-up clinical studies for establishing the effects of exenatide in patients with disrupted hypothalamic energy regulatory pathways. Copyright (C) 2012 S. Karger AG, Basel
C1 [Roth, Christian L.] Seattle Childrens Res Inst, Div Endocrinol, Seattle, WA 98101 USA.
   [Simmons, Jill H.; Shoemaker, Ashley H.] Vanderbilt Univ, Med Ctr, Dept Pediat, Div Endocrinol, Nashville, TN 37232 USA.
C3 Seattle Children's Hospital; Vanderbilt University
RP Roth, CL (通讯作者)，Seattle Childrens Res Inst, Div Endocrinol, 1900 9th Ave, Seattle, WA 98101 USA.
EM
OI Shoemaker, Ashley/0000-0003-1628-3677; Simmons, Jill/0000-0002-4189-6718
CR Ahmet A, 2006, J PEDIATR ENDOCR MET, V19, P121
   Bailey Clifford J, 2005, Curr Diab Rep, V5, P353, DOI 10.1007/s11892-005-0093-1
   Blonde L, 2006, DIABETES OBES METAB, V8, P436, DOI 10.1111/j.1463-1326.2006.00602.x
   Bloom S, 2005, CLIN MED, V5, P147, DOI 10.7861/clinmedicine.5-2-147
   Bojanowska E, 2005, MED SCI MONITOR, V11, pRA271
   BRAUNER R, 1987, ARCH FR PEDIATR, V44, P765
   Bulow B, 1998, J CLIN ENDOCR METAB, V83, P3897, DOI 10.1210/jc.83.11.3897
   Buse JB, 2004, DIABETES CARE, V27, P2628, DOI 10.2337/diacare.27.11.2628
   Danielsson P, 2007, J CLIN ENDOCR METAB, V92, P4101, DOI 10.1210/jc.2007-0826
   DeFronzo RA, 2005, DIABETES CARE, V28, P1092, DOI 10.2337/diacare.28.5.1092
   Drucker DJ, 2006, LANCET, V368, P1696, DOI 10.1016/S0140-6736(06)69705-5
   Dushay J, 2012, DIABETES CARE, V35, P4, DOI 10.2337/dc11-0931
   Falken Y, 2011, J CLIN ENDOCR METAB, V96, P2227, DOI 10.1210/jc.2010-2876
   Gardner PA, 2008, J NEUROSURG, V109, P6, DOI 10.3171/JNS/2008/109/7/0006
   Gedulin BR, 2005, ENDOCRINOLOGY, V146, P2069, DOI 10.1210/en.2004-1349
   Gutzwiller JP, 2004, PHYSIOL BEHAV, V82, P17, DOI 10.1016/j.physbeh.2004.04.019
   Hamilton JK, 2011, INT J PEDIATR ENDOCR, DOI 10.1155/2011/417949
   Hayes MR, 2010, PHYSIOL BEHAV, V100, P503, DOI 10.1016/j.physbeh.2010.02.029
   Inge TH, 2007, NAT CLIN PRACT ENDOC, V3, P606, DOI 10.1038/ncpendmet0579
   Kelly AS, 2012, OBESITY, V20, P364, DOI 10.1038/oby.2011.337
   Kendall DM, 2005, DIABETES CARE, V28, P1083, DOI 10.2337/diacare.28.5.1083
   Lustig RH, 2003, J CLIN ENDOCR METAB, V88, P2586, DOI 10.1210/jc.2002-030003
   Mason PW, 2002, ARCH PEDIAT ADOL MED, V156, P887, DOI 10.1001/archpedi.156.9.887
   Muller HL, 2008, HORM RES, V69, P193, DOI 10.1159/000113019
   Muller HL, 2007, KLIN PADIATR, V219, P323, DOI 10.1055/s-2007-985848
   Muller HL, 2004, J CLIN ENDOCR METAB, V89, P3298, DOI 10.1210/jc.2003-031751
   Muller HL, 2001, KLIN PADIATR, V213, P244, DOI 10.1055/s-2001-16855
   O'Gorman CS, 2010, J CLIN ENDOCR METAB, V95, P2211, DOI 10.1210/jc.2009-2003
   Rosenstock J, 2010, DIABETES CARE, V33, P1173, DOI 10.2337/dc09-1203
   Roth CL, 2011, OBESITY, V19, P36, DOI 10.1038/oby.2010.80
   Schroeder JW, 2011, PEDIATR RADIOL, V41, P287, DOI 10.1007/s00247-010-1968-0
   Schultes B, 2009, EUR J ENDOCRINOL, V161, P201, DOI 10.1530/EJE-09-0079
   SORVA R, 1988, ACTA PAEDIATR SCAND, V77, P587, DOI 10.1111/j.1651-2227.1988.tb10705.x
   Umeda LM, 2011, OBES SURG, V21, P896, DOI 10.1007/s11695-011-0412-3
   Vilsboll T, 2012, BMJ-BRIT MED J, V344, DOI 10.1136/bmj.d7771
   Vinchon M, 2009, CHILD NERV SYST, V25, P347, DOI 10.1007/s00381-008-0754-x
NR 36
TC 17
Z9 18
U1 0
U2 2
PU KARGER
PI BASEL
PA ALLSCHWILERSTRASSE 10, CH-4009 BASEL, SWITZERLAND
SN 1663-2818
EI 1663-2826
J9 HORM RES PAEDIAT
JI Horm. Res. Paediatr.
PY 2012
VL 78
IS 1
BP 54
EP 58
DI 10.1159/000339469
PG 5
WC Endocrinology & Metabolism; Pediatrics
WE Science Citation Index Expanded (SCI-EXPANDED)
SC Endocrinology & Metabolism; Pediatrics
GA 997AA
UT WOS:000308135000008
PM 22831918
DA 2023-08-27
ER

PT J
AU Doherty, GH
   Oldreive, C
   Harvey, J
AF Doherty, G. H.
   Oldreive, C.
   Harvey, J.
TI Neuroprotective actions of leptin on central and peripheral neurons in
   vitro
SO NEUROSCIENCE
LA English
DT Article
DE apoptosis; dopaminergic; midbrain; obesity; Ob-R; trigeminal
ID HIPPOCAMPAL SYNAPTIC PLASTICITY; CENTRAL-NERVOUS-SYSTEM; SENSORY
   NEURONS; CELL-DEATH; PARKINSONS-DISEASE; HYPOTHALAMIC NEURONS; OXIDATIVE
   STRESS; GROWTH-FACTOR; BK CHANNELS; RAT-BRAIN
AB Neuronal cell death and its regulation have been extensively studied as an essential process of both neurodevelopment and neurodegenerative conditions. However it is not clear how circulating hormones influence such processes. Therefore we aimed to determine whether the anti-obesity hormone leptin could promote the survival of murine central and peripheral neurons in vitro. Thus we established primary neuronal cultures of dopaminergic midbrain neurons and trigeminal sensory neurons and induced cell death via either toxic insult or growth factor withdrawal. We demonstrate that leptin promotes the survival of developing peripheral and central neurons via activation of mitogen-activated protein kinase (MAPK) and phosphoinositide 3-kinase (PI3kinase)/Akt/nuclear factor kappa B (NF-kappa B) -dependent signaling cascades. Specifically, leptin protects dopaminergic midbrain neurons from the apoptotic stimuli, tumor necrosis factor alpha (TNF-alpha) and 6-hydroxydopamine (6-OHDA). In addition, it promotes the survival of postnatal, but not embryonic, trigeminal sensory neurons following neurotrophin withdrawal. Our data reveal a novel neuroprotective role for leptin in the peripheral nervous system while expanding on the known anti-apoptotic role of leptin in the CNS. These findings have important implications for our understanding of neuronal viability. (C) 2008 IBRO. Published by Elsevier Ltd. All rights reserved.
C1 [Doherty, G. H.; Oldreive, C.] Univ St Andrews, Sch Biol, St Andrews KY16 9TS, Fife, Scotland.
   [Harvey, J.] Univ Dundee, Ninewells Hosp & Med Sch, Inst Neurosci, Div Pathol & Neurosci, Dundee DD1 9SY, Scotland.
C3 University of St Andrews; University of Dundee
RP Doherty, GH (通讯作者)，Univ St Andrews, Sch Biol, Bute Bldg,W Burn Lane, St Andrews KY16 9TS, Fife, Scotland.
EM
OI Doherty, Gayle/0000-0003-3494-5857; Harvey, Jenni/0000-0002-9858-001X
FU Alzheimer&apos;s Society [93] Funding Source: researchfish; Wellcome
   Trust [075821] Funding Source: Medline; Alzheimer&apos;s Society [93]
   Funding Source: Medline
CR ABBOTT RA, 1992, EUR J CLIN NUTR, V46, P879
   Ahima RS, 1999, ENDOCRINOLOGY, V140, P2755, DOI 10.1210/en.140.6.2755
   ALESSI DR, 1995, J BIOL CHEM, V270, P27489, DOI 10.1074/jbc.270.46.27489
   Alonzi T, 2001, MOL CELL NEUROSCI, V18, P270, DOI 10.1006/mcne.2001.1018
   Bartlett SE, 1997, BRAIN RES, V761, P257, DOI 10.1016/S0006-8993(97)00329-6
   Bornstein SR, 2000, DIABETES, V49, P532, DOI 10.2337/diabetes.49.4.532
   Bouret SG, 2004, SCIENCE, V304, P108, DOI 10.1126/science.1095004
   Brown R, 2005, MOL BRAIN RES, V139, P357, DOI 10.1016/j.molbrainres.2005.05.009
   Buss Robert R., 2004, Anatomical Science International, V79, P191, DOI 10.1111/j.1447-073x.2004.00088.x
   Buyse M, 2001, EUR J NEUROSCI, V14, P64, DOI 10.1046/j.0953-816x.2001.01628.x
   Caro JF, 1996, LANCET, V348, P159, DOI 10.1016/S0140-6736(96)03173-X
   Chen HL, 2004, AM J EPIDEMIOL, V159, P547, DOI 10.1093/aje/kwh059
   Chen SC, 1999, J RECEPT SIGNAL TR R, V19, P245, DOI 10.3109/10799899909036649
   Cui H, 2006, FASEB J, V20, P2654, DOI 10.1096/fj.06-5989fje
   de la Rosa EJ, 2000, TRENDS NEUROSCI, V23, P454, DOI 10.1016/S0166-2236(00)01628-3
   Dicou E, 2001, NEUROREPORT, V12, P3947, DOI 10.1097/00001756-200112210-00019
   Doherty GH, 2007, NEUROSCI RES, V57, P296, DOI 10.1016/j.neures.2006.10.019
   Durakoglugil M, 2005, J NEUROCHEM, V95, P396, DOI 10.1111/j.1471-4159.2005.03375.x
   Elmquist JK, 1998, NAT NEUROSCI, V1, P445, DOI 10.1038/2164
   Guo ZH, 2008, J BIOL CHEM, V283, P1754, DOI 10.1074/jbc.M703753200
   Hakansson ML, 1998, J NEUROSCI, V18, P559
   Hamanoue M, 1999, MOL CELL NEUROSCI, V14, P28, DOI 10.1006/mcne.1999.0770
   Hanrott K, 2006, J BIOL CHEM, V281, P5373, DOI 10.1074/jbc.M511560200
   Harvey J, 2003, NEUROPHARMACOLOGY, V44, P845, DOI 10.1016/S0028-3908(03)00076-5
   Harvey J, 2006, PROG LIPID RES, V45, P369, DOI 10.1016/j.plipres.2006.03.001
   Hebert G, 2003, NEUROSCI LETT, V349, P191, DOI 10.1016/S0304-3940(03)00832-2
   Horton AR, 1998, EUR J NEUROSCI, V10, P673, DOI 10.1046/j.1460-9568.1998.00079.x
   Hosoi T, 2002, ENDOCRINOLOGY, V143, P3498, DOI 10.1210/en.2002-220077
   Li XL, 2002, NEUROSCIENCE, V113, P607, DOI 10.1016/S0306-4522(02)00162-8
   Lu JN, 2006, NEUROSCI LETT, V407, P240, DOI 10.1016/j.neulet.2006.08.053
   Matsuda J, 1999, ENDOCRINOLOGY, V140, P5233, DOI 10.1210/en.140.11.5233
   Mattioli B, 2005, J IMMUNOL, V174, P6820, DOI 10.4049/jimmunol.174.11.6820
   McGuire SO, 2001, EXP NEUROL, V169, P219, DOI 10.1006/exnr.2001.7688
   Middleton G, 2000, J CELL BIOL, V148, P325, DOI 10.1083/jcb.148.2.325
   Middleton G, 2001, DEVELOPMENT, V128, P447
   NAGATSU T, 2000, J NEURAL TRANSM-SUPP, V107, P277
   O'Malley D, 2005, FASEB J, V19, P1917, DOI 10.1096/fj.05-4166fje
   Oldreive CE, 2008, NEUROCHEM INT, V52, P1394, DOI 10.1016/j.neuint.2008.02.010
   Orike N, 2001, J CELL BIOL, V154, P995, DOI 10.1083/jcb.200101068
   QUI LQ, 1994, DEV BIOL, V163, P516, DOI 10.1006/dbio.1994.1167
   Reichardt LF, 2004, CELL, V118, P141, DOI 10.1016/j.cell.2004.07.008
   Rouet-Benzineb R, 2004, J BIOL CHEM, V279, P16495, DOI 10.1074/jbc.M312999200
   Salinas M, 2003, J BIOL CHEM, V278, P13898, DOI 10.1074/jbc.M209164200
   Saxena NK, 2004, FASEB J, V18, P1612, DOI 10.1096/fj.04-1847fje
   Shanley LJ, 2002, J PHYSIOL-LONDON, V545, P933, DOI 10.1113/jphysiol.2002.029488
   Shanley LJ, 2002, NAT NEUROSCI, V5, P299, DOI 10.1038/nn824
   Shanley LJ, 2001, J NEUROSCI, V21, part. no., DOI 10.1523/JNEUROSCI.21-24-j0001.2001
   Spanswick D, 1997, NATURE, V390, P521, DOI 10.1038/37379
   Spiegelman BM, 2001, CELL, V104, P531, DOI 10.1016/S0092-8674(01)00240-9
   Sriram K, 2002, FASEB J, V16, P1474, DOI 10.1096/fj.02-0216fje
   Tartaglia LA, 1995, CELL, V83, P1263, DOI 10.1016/0092-8674(95)90151-5
   Ur E, 2002, NEUROENDOCRINOLOGY, V75, P264, DOI 10.1159/000054718
   Wayner MJ, 2004, PEPTIDES, V25, P991, DOI 10.1016/j.peptides.2004.03.018
   Weng Z, 2007, J BIOL CHEM, V282, P34479, DOI 10.1074/jbc.M705426200
   Williams HM, 2008, NEUROSCI RES, V60, P380, DOI 10.1016/j.neures.2007.12.007
   Yang YF, 2005, P NATL ACAD SCI USA, V102, P13670, DOI 10.1073/pnas.0504610102
NR 56
TC 44
Z9 46
U1 0
U2 10
PU PERGAMON-ELSEVIER SCIENCE LTD
PI OXFORD
PA THE BOULEVARD, LANGFORD LANE, KIDLINGTON, OXFORD OX5 1GB, ENGLAND
SN 0306-4522
EI 1873-7544
J9 NEUROSCIENCE
JI Neuroscience
PD JUL 17
PY 2008
VL 154
IS 4
BP 1297
EP 1307
DI 10.1016/j.neuroscience.2008.04.052
PG 11
WC Neurosciences
WE Science Citation Index Expanded (SCI-EXPANDED)
SC Neurosciences & Neurology
GA 326RZ
UT WOS:000257677900013
PM 18550292
DA 2023-08-27
ER

PT J
AU Gibson, EG
   Campagne, O
   Selvo, NS
   Gajjar, A
   Stewart, CF
AF Gibson, Elizabeth G.
   Campagne, Olivia
   Selvo, Nicholas S.
   Gajjar, Amar
   Stewart, Clinton F.
TI Population pharmacokinetic analysis of crizotinib in children with
   progressive/recurrent high-grade and diffuse intrinsic pontine gliomas
SO CANCER CHEMOTHERAPY AND PHARMACOLOGY
LA English
DT Article
DE Crizotinib; Pharmacokinetic modeling; Dasatinib; Pediatric; High-grade
   glioma
ID DASATINIB; TUMORS
AB Purpose Crizotinib, a potent oral tyrosine kinase inhibitor, was evaluated in combination with dasatinib in a phase 1 trial (NCT01644773) in children with progressive or recurrent high-grade and diffuse intrinsic pontine gliomas (HGG and DIPG). This study aimed to characterize the pharmacokinetics of crizotinib in this population and identify significant covariates. Methods Patients (N = 36, age range 2.9-21.3 years) were treated orally once or twice-daily with 100-215 mg/m(2) crizotinib and 50-65 mg/m(2) dasatinib. Pharmacokinetic studies were performed for crizotinib alone after the first dose and at steady state, and for the drug combination at steady state. Crizotinib plasma concentrations were measured using a validated LC-MS/MS method. Population modeling was performed (Monolix) and the impact of factors including patient demographics and co-medications were investigated on crizotinib pharmacokinetics. Results Crizotinib concentrations were described with a linear two-compartment model and absorption lag time. Concomitant dasatinib and overweight/obese status significantly influenced crizotinib pharmacokinetics, resulting in clinically relevant impact (> 20%) on drug exposure. Crizotinib mean apparent clearance (CL/F) was 66.7 L/h/m(2) after single-dose and decreased to 26.5 L/h/m(2) at steady state when given alone, but not when combined with dasatinib (mean 60.8 L/h/m(2)). Overweight/obese patients exhibited lower crizotinib CL/F and apparent volume V-1/F (mean 46.2 L/h/m(2) and 73.3 L/m(2)) compared to other patients (mean 75.5 L/h/m(2) and 119.3 L/m(2), p < 0.001). Conclusion A potential pharmacokinetic interaction was observed between crizotinib and dasatinib in children with HGG and DIPG. Further, crizotinib exposure was significantly higher in overweight/obese patients, who may require a dosing adjustment.
C1 [Gibson, Elizabeth G.; Campagne, Olivia; Selvo, Nicholas S.; Stewart, Clinton F.] St Jude Childrens Res Hosp, Dept Pharm & Pharmaceut Sci, 262 Danny Thomas Pl, Memphis, TN 38105 USA.
   [Gibson, Elizabeth G.] Bristol Myers Squibb Co, Princeton, NJ USA.
   [Gajjar, Amar] St Jude Childrens Res Hosp, Dept Oncol, Memphis, TN 38105 USA.
C3 St Jude Children's Research Hospital; Bristol-Myers Squibb; St Jude
   Children's Research Hospital
RP Stewart, CF (通讯作者)，St Jude Childrens Res Hosp, Dept Pharm & Pharmaceut Sci, 262 Danny Thomas Pl, Memphis, TN 38105 USA.
EM
RI Gajjar, Amar/N-8059-2018
OI Gibson, Elizabeth/0000-0001-6032-2528; Campagne,
   Olivia/0000-0001-6348-8910
FU Cancer Center Support (CORE) Grant [CA21765]; American Lebanese Syrian
   Associated Charities (ALSAC) at St. Jude Children's Research Hospital
FX This work was supported by a Cancer Center Support (CORE) Grant
   (CA21765) and the American Lebanese Syrian Associated Charities (ALSAC)
   at St. Jude Children's Research Hospital. The content is solely the
   responsibility of the authors and does not necessarily represent the
   official views of the National Institutes of Health.
CR Andrae J, 2008, GENE DEV, V22, P1276, DOI 10.1101/gad.1653708
   [Anonymous], 2013, WEIGHT FOR AGE CHART
   Appleman LJ, 2011, J CLIN ONCOL, V29, P4837, DOI 10.1200/JCO.2011.37.7929
   Balis FM, 2017, CANCER CHEMOTH PHARM, V79, P181, DOI 10.1007/s00280-016-3220-6
   Beal SL, 2001, J PHARMACOKINET PHAR, V28, P481, DOI 10.1023/A:1012299115260
   Bergstrand M, 2011, AAPS J, V13, P143, DOI 10.1208/s12248-011-9255-z
   Broniscer A, 2018, PEDIATR BLOOD CANCER, V65, DOI 10.1002/pbc.27035
   Broniscer A, 2013, CLIN CANCER RES, V19, P3050, DOI 10.1158/1078-0432.CCR-13-0306
   Chamdine Omar, 2014, CNS Oncol, V3, P433, DOI 10.2217/cns.14.43
   Demetri GD, 2009, CLIN CANCER RES, V15, P6232, DOI 10.1158/1078-0432.CCR-09-0224
   El-Khoueiry AB, 2018, CANCER CHEMOTH PHARM, V81, P659, DOI 10.1007/s00280-018-3517-8
   Fangusaro J, 2012, FRONT ONCOL, V2, DOI 10.3389/fonc.2012.00105
   Greengard E, 2020, CANCER CHEMOTH PHARM, V86, P829, DOI 10.1007/s00280-020-04171-4
   Hirota T, 2019, CLIN PHARMACOKINET, V58, P403, DOI 10.1007/s40262-018-0689-7
   Thai HT, 2014, J PHARMACOKINET PHAR, V41, P15, DOI 10.1007/s10928-013-9343-z
   Knibbe CAJ, 2015, ANNU REV PHARMACOL, V55, P149, DOI 10.1146/annurev-pharmtox-010814-124354
   Korshunov A, 2015, ACTA NEUROPATHOL, V129, P669, DOI 10.1007/s00401-015-1405-4
   Leveque D, 2020, CLIN PHARMACOKINET, V59, P849, DOI 10.1007/s40262-020-00872-4
   Millisor VE, 2017, PEDIATR NEPHROL, V32, P1575, DOI 10.1007/s00467-017-3693-5
   Mould DR, 2012, CPT-PHARMACOMET SYST, V1, DOI 10.1038/psp.2012.4
   Ostman A, 2007, ADV CANCER RES, V97, P247, DOI 10.1016/S0065-230X(06)97011-0
   Papa E, 2017, CELL DEATH DIS, V8, DOI 10.1038/s41419-017-0051-2
   Paugh BS, 2011, J CLIN ONCOL, V29, P3999, DOI 10.1200/JCO.2011.35.5677
   Roberts MS, 2014, J CHROMATOGR B, V960, P151, DOI 10.1016/j.jchromb.2014.04.035
   Vanan MI, 2015, FRONT ONCOL, V5, DOI 10.3389/fonc.2015.00237
   Wang EJ, 2016, CLIN CANCER RES, V22, P5722, DOI 10.1158/1078-0432.CCR-16-0536
   Xiong Y, 2017, PEDIATR CLIN N AM, V64, P1417, DOI 10.1016/j.pcl.2017.08.011
NR 27
TC 1
Z9 2
U1 0
U2 1
PU SPRINGER
PI NEW YORK
PA ONE NEW YORK PLAZA, SUITE 4600, NEW YORK, NY, UNITED STATES
SN 0344-5704
EI 1432-0843
J9 CANCER CHEMOTH PHARM
JI Cancer Chemother. Pharmacol.
PD DEC
PY 2021
VL 88
IS 6
BP 1009
EP 1020
DI 10.1007/s00280-021-04357-4
EA SEP 2021
PG 12
WC Oncology; Pharmacology & Pharmacy
WE Science Citation Index Expanded (SCI-EXPANDED)
SC Oncology; Pharmacology & Pharmacy
GA WL0ZH
UT WOS:000701385100001
PM 34586478
OA Green Accepted
DA 2023-08-27
ER

PT J
AU Zeng, Z
   Zhou, ZY
   Zhan, N
   Yuan, JP
   Ye, BX
   Gu, LJ
   Wang, J
   Jian, ZH
   Xiong, XX
AF Zeng, Zhi
   Zhou, Ziying
   Zhan, Na
   Yuan, Jingping
   Ye, Baixin
   Gu, Lijuan
   Wang, Jun
   Jian, Zhihong
   Xiong, Xiaoxing
TI USP10 Expression in Normal Adrenal Gland and Various Adrenal Tumors
SO ENDOCRINE PATHOLOGY
LA English
DT Article
DE USP10; Adrenal gland; Adrenocortical adenoma; Adrenocortical carcinoma;
   Pheochromocytoma
ID UBIQUITIN-SPECIFIC PROTEASE; AIRWAY EPITHELIAL-CELLS; ANDROGEN RECEPTOR;
   ADRENOCORTICAL TUMORS; DEUBIQUITINATION; ACTIVATION; STABILIZATION
AB Ubiquitin-specific protease 10 (USP10), a novel deubiquitinating enzyme, is associated with androgen receptor transcriptional activity and pathological processes of tumor. However, information between USP10 and the adrenal gland is limited. In particular, the role of USP10 in adrenal tumors has not been elucidated yet. This study aims to investigate the expression of USP10 in the human normal adrenal gland and various adrenal tumors. Tissue samples were obtained from 30 adrenocortical adenomas, nine adrenocortical adenocarcinomas, and 20 pheochromocytomas following laparoscopic surgery. Twenty normal adrenal glands were obtained from kidney surgical resection conducted due to renal cell carcinomas. USP10 expression was investigated on protein levels using immunohistochemistry and on mRNA levels using bioinformatics analysis in the Gene Expression Omnibus (GEO) Datasets. In the 20 cases of normal adrenal glands analyzed, USP10 protein was constantly expressed in situ in the cortex of the adrenal glands, but in the medulla of the gland, only the sustentacular cells were detected positive. In adrenal tumors, detectable levels of USP10 protein were found in 100 % (30/30) adrenocortical adenomas, 88.89 % (8/9) adrenocortical carcinomas, and 10 % (2/20) pheochromocytomas. Bioinformatics analysis did not show a significant difference in USP10 messenger RNA (mRNA) expression between adrenal tumors and normal adrenal gland tissues. A positive USP10 immunoreaction can be useful in distinguishing adrenal cortical tumors from pheochromocytoma.
C1 [Zeng, Zhi; Zhou, Ziying; Zhan, Na; Yuan, Jingping] Wuhan Univ, Renmin Hosp, Dept Pathol, Wuhan 430060, Hubei, Peoples R China.
   [Ye, Baixin] Wuhan Univ, Renmin Hosp, Dept Hematol, Wuhan 430060, Hubei, Peoples R China.
   [Gu, Lijuan; Xiong, Xiaoxing] Wuhan Univ, Renmin Hosp, Cent Lab, Wuhan 430060, Hubei, Peoples R China.
   [Wang, Jun; Jian, Zhihong; Xiong, Xiaoxing] Wuhan Univ, Renmin Hosp, Dept Neurosurg, Wuhan 430060, Hubei, Peoples R China.
C3 Wuhan University; Wuhan University; Wuhan University; Wuhan University
RP Xiong, XX (通讯作者)，Wuhan Univ, Renmin Hosp, Cent Lab, Wuhan 430060, Hubei, Peoples R China.
EM
OI Zeng, Zhi/0000-0003-4131-050X; Xiong, Xiaoxing/0000-0001-6983-8547
FU National Natural Science Foundation of China [81571147, 81301019]; Wu
   Han University [2042015kf0076]
FX This work was supported by the National Natural Science Foundation of
   China (Grant Nos. 81571147 and 81301019) and Independent Scientific
   Research Subject for Young Teachers of Wu Han University (Grant No.
   2042015kf0076).
CR Aubert S, 2002, AM J SURG PATHOL, V26, P1612, DOI 10.1097/00000478-200212000-00009
   Bentvelsen FM, 1996, ENDOCRINOLOGY, V137, P2659, DOI 10.1210/en.137.7.2659
   Bomberger JM, 2010, CHANNELS, V4, P150, DOI 10.4161/chan.4.3.11223
   Bomberger JM, 2009, J BIOL CHEM, V284, P18778, DOI 10.1074/jbc.M109.001685
   Cheng LL, 2012, CLIN CANCER RES, V18, P4201, DOI 10.1158/1078-0432.CCR-12-0036
   Deng SS, 2007, BREAST CANCER RES TR, V104, P21, DOI 10.1007/s10549-006-9393-7
   Dezso Z, 2008, BMC BIOL, V6, DOI 10.1186/1741-7007-6-49
   Draker R, 2011, NUCLEIC ACIDS RES, V39, P3529, DOI 10.1093/nar/gkq1352
   Faus H, 2005, MOL CELL ENDOCRINOL, V245, P138, DOI 10.1016/j.mce.2005.11.011
   Ge XJ, 2005, GENOMICS, V86, P127, DOI 10.1016/j.ygeno.2005.04.008
   Giordano TJ, 2009, CLIN CANCER RES, V15, P668, DOI 10.1158/1078-0432.CCR-08-1067
   Giubellino A, 2014, PLOS ONE, V9, DOI 10.1371/journal.pone.0090458
   Grunda JM, 2006, J NEURO-ONCOL, V80, P261, DOI 10.1007/s11060-006-9191-4
   Lin ZH, 2013, CELL REP, V5, P1639, DOI 10.1016/j.celrep.2013.11.029
   Liu H, 2014, TUMOR BIOL, V35, P12157, DOI 10.1007/s13277-014-2521-9
   Liu JL, 2011, CELL, V147, P223, DOI 10.1016/j.cell.2011.08.037
   Niu JX, 2013, EMBO J, V32, P3206, DOI 10.1038/emboj.2013.247
   Pan LN, 2014, BIOCHEM BIOPH RES CO, V449, P289, DOI 10.1016/j.bbrc.2014.05.037
   Soncini C, 2001, ONCOGENE, V20, P3869, DOI 10.1038/sj.onc.1204553
   Takahashi M, 2013, BLOOD, V122, P715, DOI 10.1182/blood-2013-03-493718
   Yuan J, 2010, CELL, V140, P384, DOI 10.1016/j.cell.2009.12.032
   Zeng Z, 2014, TUMOR BIOL, V35, P3845, DOI 10.1007/s13277-013-1509-1
NR 22
TC 4
Z9 6
U1 2
U2 11
PU HUMANA PRESS INC
PI TOTOWA
PA 999 RIVERVIEW DRIVE SUITE 208, TOTOWA, NJ 07512 USA
SN 1046-3976
EI 1559-0097
J9 ENDOCR PATHOL
JI Endocr. Pathol.
PD DEC
PY 2015
VL 26
IS 4
BP 302
EP 308
DI 10.1007/s12022-015-9406-3
PG 7
WC Endocrinology & Metabolism; Pathology
WE Science Citation Index Expanded (SCI-EXPANDED)
SC Endocrinology & Metabolism; Pathology
GA CX0XR
UT WOS:000365421300004
PM 26555087
DA 2023-08-27
ER

PT J
AU Bondan, EF
   Martins, MDM
   Dossa, PD
   Viebig, LB
   Cardoso, CV
   Martins, JL
   Bernardi, MM
AF Bondan, Eduardo Fernandes
   Monteiro Martins, Maria de Fatima
   Dossa, Pietro Domingues
   Viebig, Ligia Bocamino
   Cardoso, Carolina Vieira
   Martins Junior, Joao Lopes
   Bernardi, Maria Martha
TI Propentofylline reduces glial scar development following gliotoxic
   damage in the rat brainstem
SO ARQUIVOS DE NEURO-PSIQUIATRIA
LA English
DT Article
DE astrocytes; ethidium; gliosis; gliotoxin; xanthine
ID TUMOR-NECROSIS-FACTOR; AUTOIMMUNE ENCEPHALOMYELITIS; CYTOKINE
   PRODUCTION; SPINAL-CORD; REMYELINATION; DEMYELINATION; REGENERATION;
   ACTIVATION; RELEASE; ALPHA
AB Propentofylline is a xanthine derivative that depresses activation of glial cells, whose responses contribute to neural tissue damage during inflammation. Ethidium bromide injection into the central nervous system induces local oligodendroglial and astrocytic loss, resulting in primary demyelination, neuroinflammation and blood-brain barrier disruption. Surviving astrocytes present a vigorous reaction around the injury site with increased immunoreactivity to glial fibrillary acidic protein (GFAP). Objective: This study aimed to evaluate the effect of propentofylline administration on astrocytic response following gliotoxic injury. Method: Wistar rats were injected with ethidium bromide into the cisterna pontis and treated or not with propentofylline (12.5mg/kg/day, intraperitoneal) during the experimental period. Brainstem sections were collected from 15 to 31 days after gliotoxic injection and processed for GFAP immunohistochemistry. Results and Conclusion: Results demonstrate that propentofylline decreased astrocytic activation until the 21st day, suggesting that this drug may have a role in reducing glial scar development following injury.
C1 [Bondan, Eduardo Fernandes; Monteiro Martins, Maria de Fatima; Dossa, Pietro Domingues; Viebig, Ligia Bocamino; Cardoso, Carolina Vieira; Martins Junior, Joao Lopes; Bernardi, Maria Martha] Univ Paulista, Dept Patol Ambiental & Expt, Sao Paulo, SP, Brazil.
   [Bondan, Eduardo Fernandes; Monteiro Martins, Maria de Fatima] Univ Cruzeiro Sul, Dept Vet Med, Sao Paulo, SP, Brazil.
C3 Universidade Paulista; Universidade Cruzeiro do Sul
RP Bondan, EF (通讯作者)，Rua Caconde 125-51, BR-01425011 Sao Paulo, SP, Brazil.
EM
RI Bernardi, Maria/T-6813-2019; Bernardi, M. M./H-8418-2012
OI Bernardi, M. M./0000-0002-6860-9416; Martins, Maria de
   Fatima/0000-0002-3970-0659
CR Balasingam V, 1996, J NEUROSCI, V16, P2945
   Bondan E. F., 2000, Journal of Submicroscopic Cytology and Pathology, V32, P603
   Bondan EF, 2015, ARCH ENDOCRIN METAB, V59, P47, DOI 10.1590/2359-3997000000009
   Bondan EF, 2014, J NEUROIMMUNOL, V275, P139, DOI 10.1016/j.jneuroim.2014.08.372
   Bondan EF, 2014, MICROSC RES TECHNIQ, V77, P23, DOI 10.1002/jemt.22308
   Bondan EF, 2003, ARQ NEURO-PSIQUIAT, V61, P642, DOI 10.1590/S0004-282X2003000400022
   Cregg JM, 2014, EXP NEUROL, V253, P197, DOI 10.1016/j.expneurol.2013.12.024
   Fitch MT, 2001, GLIAL CELL DEV, P263
   Gwak YS, 2008, PAIN, V138, P410, DOI 10.1016/j.pain.2008.01.021
   Hayakawa T, 1998, J BIOL CHEM, V273, P20300, DOI 10.1074/jbc.273.32.20300
   Herrera F, 2010, J BIOL CHEM, V285, P38915, DOI 10.1074/jbc.M110.170274
   Jung S, 1997, J AUTOIMMUN, V10, P519, DOI 10.1006/jaut.1997.0159
   KAMMER GM, 1988, IMMUNOL TODAY, V9, P222, DOI 10.1016/0167-5699(88)91220-0
   Kuypers NJ, 2013, EXP NEUROL, V247, P615, DOI 10.1016/j.expneurol.2013.02.014
   Li LX, 2011, METAB BRAIN DIS, V26, P37, DOI 10.1007/s11011-011-9235-0
   LUNDBLAD R, 1995, SHOCK, V3, P210, DOI 10.1097/00024382-199503000-00009
   NEUNER P, 1994, IMMUNOLOGY, V83, P262
   Paintlia AS, 2009, GLIA, V57, P182, DOI 10.1002/glia.20745
   Pekny M, 2004, J PATHOL, V204, P428, DOI 10.1002/path.1645
   Plaschke K, 2001, BRIT J PHARMACOL, V133, P107, DOI 10.1038/sj.bjp.0704061
   Schubert P, 1997, ANN NY ACAD SCI, V826, P337, DOI 10.1111/j.1749-6632.1997.tb48484.x
   Schubert P, 2000, ANN NY ACAD SCI, V903, P24, DOI 10.1111/j.1749-6632.2000.tb06346.x
   Si QS, 1998, BRAIN RES, V812, P97, DOI 10.1016/S0006-8993(98)00954-8
   Sofroniew MV, 2010, ACTA NEUROPATHOL, V119, P7, DOI 10.1007/s00401-009-0619-8
   Sofroniew MV, 2009, TRENDS NEUROSCI, V32, P638, DOI 10.1016/j.tins.2009.08.002
   SOMMER N, 1995, NAT MED, V1, P244, DOI 10.1038/nm0395-244
   Sweitzer Sarah, 2011, Handb Exp Pharmacol, P235, DOI 10.1007/978-3-642-13443-2_8
   Tawfik VL, 2008, NEUROSCIENCE, V152, P1086, DOI 10.1016/j.neuroscience.2008.01.065
   Titus DJ, 2015, CURR PHARM DESIGN, V21, P332
   Yoshikawa M, 1999, MULT SCLER J, V5, P126, DOI 10.1191/135245899678847194
NR 30
TC 2
Z9 3
U1 0
U2 0
PU ASSOC ARQUIVOS NEURO- PSIQUIATRIA
PI SAO PAULO SP
PA PR AMADEU AMARAL 47/33, 01327-010 SAO PAULO SP, BRAZIL
SN 0004-282X
EI 1678-4227
J9 ARQ NEURO-PSIQUIAT
JI Arq. Neuro-Psiquiatr.
PD SEP
PY 2016
VL 74
IS 9
BP 730
EP 736
DI 10.1590/0004-282X20160108
PG 7
WC Neurosciences; Psychiatry
WE Science Citation Index Expanded (SCI-EXPANDED)
SC Neurosciences & Neurology; Psychiatry
GA DY2ZV
UT WOS:000384961500009
PM 27706422
OA gold, Green Published, Green Submitted
DA 2023-08-27
ER

PT J
AU Zhang, WH
   Li, X
   Zhang, JY
   Luft, A
   Hanley, DF
   van Zijl, P
   Miller, MI
   Younes, L
   Mori, S
AF Zhang, Weihong
   Li, Xin
   Zhang, Jiangyang
   Luft, Andreas
   Hanley, Daniel F.
   van Zijl, Peter
   Miller, Michael I.
   Younes, Laurent
   Mori, Susumu
TI Landmark-referenced voxel-based analysis of diffusion tensor images of
   the brainstem white matter tracts Application in patients with middle
   cerebral artery stroke
SO NEUROIMAGE
LA English
DT Article
DE Diffusion tensor imaging; White matter tract; Normalization;
   Registration; Stroke; Landmark
ID WALLERIAN DEGENERATION; AXONAL PROJECTIONS; PYRAMIDAL TRACT;
   MULTIPLE-SCLEROSIS; FIBER TRACTOGRAPHY; TRACKING; MRI; REGISTRATION;
   MORPHOMETRY; ARCHITECTURE
AB Although DTI can provide detailed information about white matter anatomy, it is not yet straightforward enough to quantify the anatomical information it visualizes. In this study, we developed and tested a new tool to perform brain normalization and voxel-based analysis of DTI data. For the normalization part, manually placed landmarks ensured that the visualized white matter tracts were well-registered among the populations. A standard landmark set in ICBM-152 space and an interface to remap them to subject data were integrated in the procedure. After landmark placement, highly elastic non-linear Large Deformation Diffeomorphic Metric Mapping (LDDMM) was driven by the landmarks to normalize the brainstem anatomy of normal subjects. The approach was then applied to delineate brainstem tract abnormalities in patients with left chronic middle cerebral artery (MCA) stroke. The voxel-based comparison between control and patient groups identified abnormalities in the ipsilesional corticospinal tract and contralesional cerebellar peduncles. We believe that this tool is useful for regional brain normalization of patients with severe anatomical alterations, such as stroke, brain tumor, and lobectomy, for whom standard automated normalization tools may not work properly. (C) 2008 Elsevier Inc. All rights reserved.
C1 [Zhang, Weihong; Zhang, Jiangyang; van Zijl, Peter; Mori, Susumu] Johns Hopkins Univ, Sch Med, Dept Radiol, Baltimore, MD 21205 USA.
   [Luft, Andreas; Hanley, Daniel F.] Johns Hopkins Univ, Div Brain Injury Outcomes, Dept Neurol, Baltimore, MD 21205 USA.
   [Zhang, Weihong; Li, Xin; Zhang, Jiangyang; van Zijl, Peter; Mori, Susumu] Kennedy Krieger Inst, FM Kirby Ctr Funct Brain Imaging, Baltimore, MD 21205 USA.
   [Zhang, Weihong] Chinese Acad Med Sci, Beijing 100730, Peoples R China.
   [Zhang, Weihong] Beijing Union Med Coll Hosp, Peking Union Med Coll, Dept Radiol, Beijing 100730, Peoples R China.
   [Luft, Andreas] Univ Tubingen, Dept Gen Neurol, D-72074 Tubingen, Germany.
   [Luft, Andreas] Univ Tubingen, Hertie Inst Clin Brain Res, D-72074 Tubingen, Germany.
   [Miller, Michael I.; Younes, Laurent] Johns Hopkins Univ, Sch Med, Dept Biomed Engn, Baltimore, MD 21205 USA.
C3 Johns Hopkins University; Johns Hopkins University; Kennedy Krieger
   Institute; Chinese Academy of Medical Sciences - Peking Union Medical
   College; Chinese Academy of Medical Sciences - Peking Union Medical
   College; Peking Union Medical College; Peking Union Medical College
   Hospital; Eberhard Karls University of Tubingen; Eberhard Karls
   University of Tubingen; Eberhard Karls University Hospital; Johns
   Hopkins University
RP Mori, S (通讯作者)，Johns Hopkins Univ, Sch Med, Dept Radiol & Radiol Sci, 217 Traylor Bldg,720 Rutland Ave, Baltimore, MD 21205 USA.
EM
RI Van Zijl, Peter C/ABF-3732-2020; Zhang, Wei/HOF-7252-2023; Younes, E.
   Laurent/A-3349-2010; Miller, Michael I./A-3213-2010
OI Li, Xin/0000-0002-7414-5734; Younes, Laurent/0000-0003-2017-9565; Zhang,
   Jiangyang/0000-0003-0740-2662; Zhang, Jiangyang/0000-0003-4146-8163;
   Hanley, Daniel/0000-0002-9536-4258
FU NIH [U24RR021382, P41 RR015241, RO1AG20012]; Veterans Affairs
   Rehabilitation Research and Development Maryland Exercise and Robotics
   Center of Excellence (MERCE); Rehabilitation Research & Development
   Exercise and Robotics Center of Excellence [B3390K]; NIA [P60AG 12583];
   University of Maryland Claude D. Pepper Older Americans Independence
   Center; Department of Veterans Affairs and Baltimore Veterans Affairs
   Medical Center Geriatrics Research, Education and Clinical Center
   (GRECC); NINDS [1RO1 NS 24282-08]; National Center for Research
   Resources (NCRR)
FX This research was supported by NIH grants U24RR021382, P41 RR015241, and
   RO1AG20012. We wish to acknowledge Richard Macko MD from the Veterans
   Affairs Rehabilitation Research and Development Maryland Exercise and
   Robotics Center of Excellence (MERCE), the Rehabilitation Research &
   Development Exercise and Robotics Center of Excellence (B3390K), and
   Stroke (REAP), as well as the NIA (P60AG 12583) University of Maryland
   Claude D. Pepper Older Americans Independence Center; the Department of
   Veterans Affairs and Baltimore Veterans Affairs Medical Center
   Geriatrics Research, Education and Clinical Center (GRECC), and NINDS
   1RO1 NS 24282-08. One of the supporting agencies, the National Center
   for Research Resources (NCRR), is a component of the National Institutes
   of Health (NIH). The content of this paper is solely the responsibility
   of the authors and does not necessarily represent the of. cial viewof
   NCRR or NIH. Dr. Peter C. M. van Zijl is a paid lecturer for Philips
   Medical Systems. This arrangement has been approved by the Johns Hopkins
   University in accordance with its con. ict of interest policies.
CR Alexander DC, 2001, IEEE T MED IMAGING, V20, P1131, DOI 10.1109/42.963816
   Allassonniere S, 2005, LECT NOTES COMPUT SC, V3757, P365, DOI 10.1007/11585978_24
   Ashburner J, 2000, NEUROIMAGE, V11, P805, DOI 10.1006/nimg.2000.0582
   Basser PJ, 2000, MAGNET RESON MED, V44, P625, DOI 10.1002/1522-2594(200010)44:4<625::AID-MRM17>3.0.CO;2-O
   BASSER PJ, 1994, BIOPHYS J, V66, P259, DOI 10.1016/S0006-3495(94)80775-1
   Cao Y, 2005, IEEE T MED IMAGING, V24, P1216, DOI 10.1109/TMI.2005.853923
   Catani M, 2002, NEUROIMAGE, V17, P77, DOI 10.1006/nimg.2002.1136
   Chiang MC, 2008, IEEE T MED IMAGING, V27, P442, DOI 10.1109/TMI.2007.907326
   Conturo TE, 1999, P NATL ACAD SCI USA, V96, P10422, DOI 10.1073/pnas.96.18.10422
   Genovese CR, 2002, NEUROIMAGE, V15, P870, DOI 10.1006/nimg.2001.1037
   Glenn OA, 2003, J MAGN RESON IMAGING, V18, P641, DOI 10.1002/jmri.10420
   Jiang HY, 2006, COMPUT METH PROG BIO, V81, P106, DOI 10.1016/j.cmpb.2005.08.004
   Jones DK, 1999, MAGNET RESON MED, V42, P515, DOI 10.1002/(SICI)1522-2594(199909)42:3<515::AID-MRM14>3.0.CO;2-Q
   Khong PL, 2004, CEREBROVASC DIS, V18, P240, DOI 10.1159/000079961
   Kirwan CB, 2007, HUM BRAIN MAPP, V28, P959, DOI 10.1002/hbm.20331
   Luft AR, 2005, NEUROIMAGE, V26, P184, DOI 10.1016/j.neuroimage.2005.01.027
   Makris N, 1997, ANN NEUROL, V42, P951, DOI 10.1002/ana.410420617
   Miller MI, 2002, ANNU REV BIOMED ENG, V4, P375, DOI 10.1146/annurev.bioeng.4.092101.125733
   Moller M, 2007, J NEUROL NEUROSUR PS, V78, P587, DOI 10.1136/jnnp.2006.100248
   Mori S, 1999, ANN NEUROL, V45, P265, DOI 10.1002/1531-8249(199902)45:2<265::AID-ANA21>3.0.CO;2-3
   Mori S., 2005, MRI ATLAS HUMAN WHIT
   Mukherjee P, 2005, NEUROIMAG CLIN N AM, V15, P655, DOI 10.1016/j.nic.2005.08.010
   Pagani E, 2005, NEUROIMAGE, V26, P258, DOI 10.1016/j.neuroimage.2005.01.008
   Pajevic S, 1999, MAGNET RESON MED, V42, P526, DOI 10.1002/(SICI)1522-2594(199909)42:3<526::AID-MRM15>3.0.CO;2-J
   Parker GJM, 2002, NEUROIMAGE, V15, P797, DOI 10.1006/nimg.2001.0994
   Partridge SC, 2004, NEUROIMAGE, V22, P1302, DOI 10.1016/j.neuroimage.2004.02.038
   Pierpaoli C, 1996, MAGNET RESON MED, V36, P893, DOI 10.1002/mrm.1910360612
   Pierpaoli C, 2001, NEUROIMAGE, V13, P1174, DOI 10.1006/nimg.2001.0765
   Poupon C, 2000, NEUROIMAGE, V12, P184, DOI 10.1006/nimg.2000.0607
   Pruessmann KP, 1999, MAGNET RESON MED, V42, P952, DOI 10.1002/(SICI)1522-2594(199911)42:5<952::AID-MRM16>3.0.CO;2-S
   Smith SM, 2006, NEUROIMAGE, V31, P1487, DOI 10.1016/j.neuroimage.2006.02.024
   Stieltjes B, 2001, NEUROIMAGE, V14, P723, DOI 10.1006/nimg.2001.0861
   Thomalla G, 2004, NEUROIMAGE, V22, P1767, DOI 10.1016/j.neuroimage.2004.03.041
   Verma R, 2005, P NATL ACAD SCI USA, V102, P6978, DOI 10.1073/pnas.0407828102
   Virta A, 1999, MAGN RESON IMAGING, V17, P1121, DOI 10.1016/S0730-725X(99)00048-X
   Wakana S, 2004, RADIOLOGY, V230, P77, DOI 10.1148/radiol.2301021640
   Werring DJ, 2000, J NEUROL NEUROSUR PS, V69, P269, DOI 10.1136/jnnp.69.2.269
   Wilson M, 2003, J NEUROL NEUROSUR PS, V74, P203, DOI 10.1136/jnnp.74.2.203
   Woods RP, 1998, J COMPUT ASSIST TOMO, V22, P139, DOI 10.1097/00004728-199801000-00027
   Wright IC, 1995, NEUROIMAGE, V2, P244, DOI 10.1006/nimg.1995.1032
   XU D, 2002, IEEE INT S BIOM IM M
   Xu DR, 2003, MAGN RESON MED, V50, P175, DOI 10.1002/mrm.10489
   Xue R, 1999, MAGNET RESON MED, V42, P1123, DOI 10.1002/(SICI)1522-2594(199912)42:6<1123::AID-MRM17>3.0.CO;2-H
   Yushkevich PA, 2008, NEUROIMAGE, V41, P448, DOI 10.1016/j.neuroimage.2008.01.013
   Zhang H, 2006, MED IMAGE ANAL, V10, P764, DOI 10.1016/j.media.2006.06.004
   Zhang JY, 2003, NEUROIMAGE, V20, P1639, DOI 10.1016/S1053-8119(03)00410-5
NR 46
TC 20
Z9 20
U1 0
U2 8
PU ACADEMIC PRESS INC ELSEVIER SCIENCE
PI SAN DIEGO
PA 525 B ST, STE 1900, SAN DIEGO, CA 92101-4495 USA
SN 1053-8119
EI 1095-9572
J9 NEUROIMAGE
JI Neuroimage
PD FEB 1
PY 2009
VL 44
IS 3
BP 906
EP 913
DI 10.1016/j.neuroimage.2008.09.013
PG 8
WC Neurosciences; Neuroimaging; Radiology, Nuclear Medicine & Medical
   Imaging
WE Science Citation Index Expanded (SCI-EXPANDED)
SC Neurosciences & Neurology; Radiology, Nuclear Medicine & Medical Imaging
GA 392KL
UT WOS:000262301500030
PM 18852056
OA Green Accepted
DA 2023-08-27
ER

PT J
AU U-King-Im, JM
   Taylor, MD
   Raybaud, C
AF U-King-Im, Jean Marie
   Taylor, Michael D.
   Raybaud, Charles
TI Posterior fossa ependymomas: new radiological classification with
   surgical correlation
SO CHILDS NERVOUS SYSTEM
LA English
DT Article
DE Ependymomas; Paediatric; Magnetic resonance; Tumour
ID INTRACRANIAL EPENDYMOMAS; PROGNOSTIC-FACTORS; STEM-CELLS; CHILDHOOD;
   CHILDREN; SURVIVAL; MANAGEMENT; TUMOR; LONG
AB The key determinant of long-term outcome in infratentorial ependymomas remains the extent of surgical resection. We describe a new radiological classification system which is validated against surgical findings and correlated with risk of post-operative residual tumour.
   Twenty-five consecutive patients (12 females, mean age 4.9 years, range 0.5-17 years) with infratentorial ependymomas were studied. Lesions were classified on pre-operative MRI according to the pattern of extension, brainstem displacement and involvement of the obex, as lateral-type or midfloor-type tumours. Twenty-one operative records were reviewed with respect to the microanatomical tumour origin by a paediatric neurosurgeon, blinded to MRI findings. Follow-up imaging studies were evaluated for residual tumour.
   There were 15 cases of midfloor-type tumour (anterior displacement of brainstem, infiltration of obex) and 10 cases of lateral-type tumour (lateral displacement of brainstem, obex free of tumour). Extension into prepontine or cerebellopontine cisterns was more common in lateral-type tumours. Agreement between the radiological classification and tumour origin, as defined by operative records, was seen in 18 out of 20 cases. Risk of residual tumour in lateral-type tumours was more than twice that of midfloor-type tumours (80% vs. 33%, p = 0.04). Risk of tumour residual was also significantly higher when vessel encasement or prepontine extension was observed.
   Infratentorial ependymomas can be pre-operatively classified as lateral-type or midfloor-type tumours. This correlates well with operative findings. Lateral-type tumours have significantly increased risk of residual tumour compared to midfloor- type tumours and this may influence intensity of imaging surveillance.
C1 [U-King-Im, Jean Marie; Raybaud, Charles] Hosp Sick Children, Dept Diagnost Imaging, Toronto, ON M5G 1X8, Canada.
   [U-King-Im, Jean Marie; Taylor, Michael D.; Raybaud, Charles] Univ Toronto, Toronto, ON M5G 1X8, Canada.
   [Taylor, Michael D.] Hosp Sick Children, Div Neurosurg, Toronto, ON M5G 1X8, Canada.
C3 University of Toronto; Hospital for Sick Children (SickKids); University
   of Toronto; University of Toronto; Hospital for Sick Children (SickKids)
RP Raybaud, C (通讯作者)，Hosp Sick Children, Dept Diagnost Imaging, 555 Univ Ave, Toronto, ON M5G 1X8, Canada.
EM
OI Taylor, Michael/0000-0001-7009-3466
CR Boop FA, 2009, CHILD NERV SYST, V25, P1163, DOI 10.1007/s00381-009-0945-0
   COSGROVE GR, 1985, SURG NEUROL, V24, P433, DOI 10.1016/0090-3019(85)90304-0
   Dirven CMF, 1997, CHILD NERV SYST, V13, P17, DOI 10.1007/s003810050033
   ERNESTUS RI, 1989, NEUROSURG REV, V12, P157, DOI 10.1007/BF01741491
   FERRANTE L, 1994, ACTA NEUROCHIR, V131, P67, DOI 10.1007/BF01401455
   Figarella-Branger D, 2000, J NEUROSURG, V93, P605, DOI 10.3171/jns.2000.93.4.0605
   GOLDWEIN JW, 1990, CANCER-AM CANCER SOC, V66, P557, DOI 10.1002/1097-0142(19900801)66:3<557::AID-CNCR2820660325>3.0.CO;2-8
   GOLDWEIN JW, 1990, INT J RADIAT ONCOL, V19, P1497, DOI 10.1016/0360-3016(90)90362-N
   Good CD, 2001, J NEUROSURG, V94, P27, DOI 10.3171/jns.2001.94.1.0027
   HEALEY EA, 1991, NEUROSURGERY, V28, P666, DOI 10.1227/00006123-199105000-00005
   IKEZAKI K, 1993, NEUROSURGERY, V32, P38, DOI 10.1227/00006123-199301000-00006
   JAYAWICKREME DP, 1995, CHILD NERV SYST, V11, P409, DOI 10.1007/BF00717407
   Kasliwal MK, 2007, J NEURO-ONCOL, V83, P31, DOI 10.1007/s11060-007-9330-6
   Mack SC, 2009, CHILD NERV SYST, V25, P1195, DOI 10.1007/s00381-009-0928-1
   Massimino M, 2009, FUTURE ONCOL, V5, P207, DOI 10.2217/14796694.5.2.207
   McLaughlin MP, 1998, INT J RADIAT ONCOL, V40, P845, DOI 10.1016/S0360-3016(97)00893-6
   Merchant TE, 2009, LANCET ONCOL, V10, P258, DOI 10.1016/S1470-2045(08)70342-5
   NAZAR GB, 1990, J NEUROSURG, V72, P408, DOI 10.3171/jns.1990.72.3.0408
   Perilongo G, 1997, MED PEDIATR ONCOL, V29, P79
   POLLACK IF, 1995, NEUROSURGERY, V37, P655, DOI 10.1227/00006123-199510000-00008
   Poppleton H, 2007, BRIT J CANCER, V96, P6, DOI 10.1038/sj.bjc.6603519
   Sala F, 1998, PEDIATR NEUROSURG, V28, P135, DOI 10.1159/000028637
   Sanford RA, 2009, CHILD NERV SYST, V25, P1229, DOI 10.1007/s00381-009-0886-7
   Smyth MD, 2000, PEDIATR NEUROSURG, V33, P138, DOI 10.1159/000028995
   SUTTON LN, 1991, PEDIATR NEUROSURG, V16, P57, DOI 10.1159/000120509
   Tamburrini G, 2009, CHILD NERV SYST, V25, P1303, DOI 10.1007/s00381-009-0874-y
   Taylor MD, 2005, CANCER CELL, V8, P323, DOI 10.1016/j.ccr.2005.09.001
   UNDJIAN S, 1990, CHILD NERV SYST, V6, P131, DOI 10.1007/BF00308488
   Yuh EL, 2009, CHILD NERV SYST, V25, P1203, DOI 10.1007/s00381-009-0878-7
NR 29
TC 13
Z9 13
U1 0
U2 1
PU SPRINGER
PI NEW YORK
PA 233 SPRING ST, NEW YORK, NY 10013 USA
SN 0256-7040
EI 1433-0350
J9 CHILD NERV SYST
JI Childs Nerv. Syst.
PD DEC
PY 2010
VL 26
IS 12
BP 1765
EP 1772
DI 10.1007/s00381-010-1251-6
PG 8
WC Clinical Neurology; Pediatrics; Surgery
WE Science Citation Index Expanded (SCI-EXPANDED)
SC Neurosciences & Neurology; Pediatrics; Surgery
GA 680XL
UT WOS:000284270700020
PM 20680298
DA 2023-08-27
ER

PT J
AU Jahns, H
   McElroy, MC
AF Jahns, Hanne
   McElroy, Maire C.
TI Bovine intracranial neoplasia: A retrospective case series
SO VETERINARY PATHOLOGY
LA English
DT Article
DE brain; cattle; immunohistochemistry; isocitrate dehydrogenase gene;
   neoplasia; neuropathology; OLIG2
ID SQUAMOUS-CELL CARCINOMA; CHOROID-PLEXUS CARCINOMA; CEREBELLAR
   MEDULLOBLASTOMA; SPONGIFORM ENCEPHALOPATHY; NEUROPATHOLOGICAL FINDINGS;
   NERVOUS-SYSTEM; TUMORS; CANINE; CATTLE; MENINGIOMAS
AB This case series describes the clinical and pathological findings of intracranial neoplasms in cattle, a rare entity. Data and archived tissues from 24 intracranial tumors were reviewed and investigated by immunohistochemistry for S100, glial fibrillary acidic protein, synaptophysin, pancytokeratin, vimentin, neuron-specific enolase, oligodendrocyte transcription factor 2, and isocitrate dehydrogenase 1. Ages of affected cattle ranged from 6 months to 14 years (5.7 +/- 3.6 years; mean +/- SD). Predominant clinical signs were altered mental state, central vestibular dysfunction, and cerebellar incoordination. Twelve gliomas, all high grade, were the most common tumors observed: oligodendrogliomas (n = 6), astrocytomas (n = 4), and undefined gliomas (n = 2). The oligodendrogliomas were located in the brainstem and extended into the ventricles, whereas all astrocytomas were located in the forebrain. Isocitrate dehydrogenase 1 gene mutation as described in humans was not detected. The 5 meningiomas exhibited microcystic, chordoid, atypical, papillary, and anaplastic subtypes. Metastatic carcinomas (n = 4) were the only secondary tumor type present, and these were located at the level of the medulla with infiltration of cranial nerves and in one case leptomeningeal carcinomatosis. In addition, 2 medulloblastomas and 1 choroid plexus carcinoma were diagnosed. Immunohistochemistry for vimentin and pancytokeratin was particularly useful to distinguish meningiomas and choroid plexus carcinoma (positive for vimentin only) from mestastatic carcinomas (positive for cytokeratin only) as all showed a papillary growth pattern. Overall, the morphological features were comparable with other species and the human and canine classifications could be applied.
C1 [Jahns, Hanne] Univ Coll Dublin, Dublin, Ireland.
   [McElroy, Maire C.] Dept Agr Food & Marine, Celbridge, Ireland.
C3 University College Dublin
RP Jahns, H (通讯作者)，Univ Coll Dublin, UCD, Sch Vet Med, Vet Sci Ctr R010, Dublin D04W 6F6, Ireland.
EM
OI Jahns, Hanne/0000-0001-6944-154X
CR Agerholm JS, 2002, APMIS, V110, P54, DOI 10.1034/j.1600-0463.2002.100107.x
   Akiyama N, 2020, J VET MED SCI, V82, P1436, DOI 10.1292/jvms.20-0295
   Barnhart KF, 2002, VET PATHOL, V39, P311, DOI 10.1354/vp.39-3-311
   Barrett TF, 2018, NEUROSURG FOCUS, V44, DOI 10.3171/2018.2.FOCUS17761
   BASTIANELLO SS, 1982, ONDERSTEPOORT J VET, V49, P195
   Belluco S, 2019, VET PATHOL, V56, P342, DOI 10.1177/0300985818824952
   Bianchi E, 2015, J VET INTERN MED, V29, P1117, DOI 10.1111/jvim.13568
   Bouchart C, 2020, CANCER MED-US, V9, P1069, DOI 10.1002/cam4.2592
   Brooks AN, 2012, J VET DIAGN INVEST, V24, P587, DOI 10.1177/1040638712443358
   Cantile C, 2002, J COMP PATHOL, V126, P183, DOI 10.1053/jcpa.2001.0544
   Capper D, 2009, ACTA NEUROPATHOL, V118, P599, DOI 10.1007/s00401-009-0595-z
   D'Angelo A, 2015, J VET INTERN MED, V29, P967, DOI 10.1111/jvim.12592
   Dalton MF, 2019, FRONT VET SCI, V6, DOI 10.3389/fvets.2019.00224
   Carvalho FKD, 2014, CIENC RURAL, V44, P678, DOI 10.1590/S0103-84782014000400017
   DUKES TW, 1982, CAN VET J, V23, P28
   Eroksuz Y, 1999, AUST VET J, V77, P400, DOI 10.1111/j.1751-0813.1999.tb10317.x
   FANKHAUSER R, 1974, B WORLD HEALTH ORGAN, V50, P53
   FENNER WR, 1990, SEMIN VET MED SURG, V5, P253
   Fernandez F, 2016, VET J, V209, P125, DOI 10.1016/j.tvjl.2015.10.039
   GORDON DE, 1968, CANCER RES, V28, P2423
   Hasselblatt M, 2006, AM J SURG PATHOL, V30, P66, DOI 10.1097/01.pas.0000176430.88702.e0
   HAYES HM, 1975, INT J CANCER, V15, P39, DOI 10.1002/ijc.2910150106
   Hoenerhoff MJ, 2006, J COMP PATHOL, V135, P146, DOI 10.1016/j.jcpa.2006.04.002
   Ide T, 2010, VET PATHOL, V47, P741, DOI 10.1177/0300985810363486
   Iulini B, 2012, RES VET SCI, V93, P872, DOI 10.1016/j.rvsc.2011.10.008
   JEFFREY M, 1992, VET REC, V131, P332, DOI 10.1136/vr.131.15.332
   Johnson GC, 2014, VET PATHOL, V51, P146, DOI 10.1177/0300985813509387
   JOLLY RD, 1969, PATHOL VET, V6, P463, DOI 10.1177/030098586900600510
   JOSEPHSON GKA, 1990, CAN VET J, V31, P700
   Kauer RV, 2020, VET PATHOL, V57, P253, DOI 10.1177/0300985819887534
   Kishimoto TE, 2020, J VET MED SCI, V82, P77, DOI 10.1292/jvms.19-0486
   Kleinschmidt S, 2009, J COMP PATHOL, V140, P72, DOI 10.1016/j.jcpa.2008.10.001
   Koehler JW, 2018, J NEUROPATH EXP NEUR, V77, P2018, DOI 10.1093/jnen/nly085
   Koestner A., 1999, HISTOLOGICAL CLASSIF
   Kosty J, 2017, CELL CYCLE, V16, P1654, DOI 10.1080/15384101.2017.1361062
   Kwon SM, 2019, WORLD NEUROSURG, V125, pE1, DOI 10.1016/j.wneu.2018.11.252
   Ligon KL, 2004, J NEUROPATH EXP NEUR, V63, P499, DOI 10.1093/jnen/63.5.499
   Louis DN, 2016, ACTA NEUROPATHOL, V131, P803, DOI 10.1007/s00401-016-1545-1
   Lucena RB, 2011, J COMP PATHOL, V145, P20, DOI 10.1016/j.jcpa.2010.11.002
   Mayhew I, 1989, LARGE ANIMAL NEUROLO
   MCGILL IS, 1993, J COMP PATHOL, V108, P241, DOI 10.1016/S0021-9975(08)80288-5
   Merickel JL, 2021, VET PATHOL, V58, P945, DOI 10.1177/03009858211025795
   Miller AD, 2019, VET PATHOL, V56, P860, DOI 10.1177/0300985819859872
   Misdorp W, 2002, VET QUART, V24, P1
   Mizukami C, 2021, J COMP PATHOL, V182, P32, DOI 10.1016/j.jcpa.2020.11.005
   MONLUX AW, 1956, AM J VET RES, V17, P646
   Motta L, 2012, VET J, V192, P153, DOI 10.1016/j.tvjl.2011.10.008
   Ohfuji S., 2013, Comparative Clinical Pathology, V22, P789, DOI 10.1007/s00580-013-1724-6
   OMI K, 1994, J COMP PATHOL, V111, P1, DOI 10.1016/S0021-9975(05)80106-9
   Ortloff A, 2017, J COMP PATHOL, V156, P25, DOI 10.1016/j.jcpa.2016.10.007
   Pace LW, 1997, J VET DIAGN INVEST, V9, P106, DOI 10.1177/104063879700900124
   Pant I., 2014, ASTROCYTE, V1, P124
   PLUMMER P. J. G., 1956, CANADIAN JOUR COMP MED AND VET SCI, V20, P239
   Ramos-Vara JA, 2010, VET PATHOL, V47, P725, DOI 10.1177/0300985810364528
   Sammin D, 1996, IRISH VET J, V49, P664
   SHORTRIDGE E H, 1971, New Zealand Veterinary Journal, V19, P5, DOI 10.1080/00480169.1971.33920
   Snyder JM, 2008, J VET INTERN MED, V22, P172, DOI 10.1111/j.1939-1676.2007.0002.x
   Song RB, 2013, J VET INTERN MED, V27, P1143, DOI 10.1111/jvim.12136
   SUMMERS BA, 1979, VET PATHOL, V16, P132, DOI 10.1177/030098587901600118
   SWEENEY RW, 1986, J AM VET MED ASSOC, V189, P555
   Troxel MT, 2003, J VET INTERN MED, V17, P850, DOI 10.1892/0891-6640(2003)017&lt;0850:FINRRO&gt;2.3.CO;2
   Tyagi I, 2013, BRAIN TUMOR PATHOL, V30, P40, DOI 10.1007/s10014-012-0096-y
   Vandevelde M., 2012, VET NEUROPATHOLOGY E
   Yamada M, 1998, J COMP PATHOL, V119, P75, DOI 10.1016/S0021-9975(98)80073-X
   ZEMAN DH, 1986, CORNELL VET, V76, P236
NR 65
TC 1
Z9 1
U1 0
U2 3
PU SAGE PUBLICATIONS INC
PI THOUSAND OAKS
PA 2455 TELLER RD, THOUSAND OAKS, CA 91320 USA
SN 0300-9858
EI 1544-2217
J9 VET PATHOL
JI Vet. Pathol.
PD SEP
PY 2022
VL 59
IS 5
BP 824
EP 835
AR 03009858221100433
DI 10.1177/03009858221100433
EA MAY 2022
PG 12
WC Pathology; Veterinary Sciences
WE Science Citation Index Expanded (SCI-EXPANDED)
SC Pathology; Veterinary Sciences
GA 3O2LL
UT WOS:000806289800001
PM 35638647
OA Green Published, hybrid
DA 2023-08-27
ER

PT J
AU Kiss, NB
   Muth, A
   Andreasson, A
   Juhlin, CC
   Geli, J
   Backdahl, M
   Hoog, A
   Wangberg, B
   Nilsson, O
   Ahlman, H
   Larsson, C
AF Kiss, Nimrod B.
   Muth, Andreas
   Andreasson, Adam
   Juhlin, C. Christofer
   Geli, Janos
   Backdahl, Martin
   Hoog, Anders
   Wangberg, Bo
   Nilsson, Ola
   Ahlman, Hakan
   Larsson, Catharina
TI Acquired hypermethylation of the P16(INK4A) promoter in abdominal
   paraganglioma: relation to adverse tumor phenotype and predisposing
   mutation
SO ENDOCRINE-RELATED CANCER
LA English
DT Article
DE Molecular genetics; Gene regulation; Metastasis; Adrenal medulla;
   Endocrine therapy
ID MALIGNANT PHEOCHROMOCYTOMA; SPORADIC PHEOCHROMOCYTOMAS;
   SUCCINATE-DEHYDROGENASE; PROLYL HYDROXYLASE; GENE-MUTATIONS;
   METHYLATION; EXPRESSION; CANCER; CELLS; SDHB
AB Recurrent alterations in promoter methylation of tumor suppressor genes (TSGs) and LINE1 (L1RE1) repeat elements were previously reported in pheochromocytoma and abdominal paraganglioma. This study was undertaken to explore CpG methylation abnormalities in an extended tumor panel and assess possible relationships between metastatic disease and mutation status. CpG methylation was quantified by bisulfite pyrosequencing for selected TSG promoters and LINE1 repeats. Methylation indices above normal reference were observed for DCR2 (TNFRSF10D), CDH1, P16 (CDKN2A), RARB, and RASSF1A. Z-scores for overall TSG, and individual TSG methylation levels, but not LINE1, were significantly correlated with metastatic disease, paraganglioma, disease predisposition, or outcome. Most strikingly, P16 hypermethylation was strongly associated with SDHB mutation as opposed to RET/MEN2, VHL/VHL, or NF1-related disease. Parallel analyses of constitutional, tumor, and metastasis DNA implicate an order of events where constitutional SDHB mutations are followed by TSG hypermethylation and 1p loss in primary tumors, later transferred to metastatic tissue. In the combined material, P16 hypermethylation was prevalent in SDHB-mutated samples and was associated with short disease-related survival. The findings verify the previously reported importance of P16 and other TSG hypermethylation in an independent tumor series. Furthermore, a constitutional SDHB mutation is proposed to predispose for an epigenetic tumor phenotype occurring before the emanation of clinically recognized malignancy.
C1 [Kiss, Nimrod B.; Andreasson, Adam; Geli, Janos; Backdahl, Martin; Larsson, Catharina] Karolinska Inst, Karolinska Univ Hosp, Dept Mol Med & Surg, SE-17176 Stockholm, Sweden.
   [Kiss, Nimrod B.; Andreasson, Adam; Juhlin, C. Christofer; Larsson, Catharina] Karolinska Univ Hosp, Canc Ctr Karolinska, Stockholm, Sweden.
   [Muth, Andreas; Wangberg, Bo; Ahlman, Hakan] Sahlgrens Univ Hosp, Dept Surg, Gothenburg, Sweden.
   [Juhlin, C. Christofer; Hoog, Anders] Karolinska Inst, Dept Oncol Pathol, Stockholm, Sweden.
   [Nilsson, Ola] Sahlgrens Univ Hosp, Dept Pathol, Gothenburg, Sweden.
C3 Karolinska Institutet; Karolinska University Hospital; Karolinska
   Institutet; Karolinska University Hospital; Sahlgrenska University
   Hospital; Karolinska Institutet; Sahlgrenska University Hospital
RP Kiss, NB (通讯作者)，Karolinska Inst, Karolinska Univ Hosp, Dept Mol Med & Surg, CCK R8 04, SE-17176 Stockholm, Sweden.
EM
RI Juhlin, Carl Christofer/C-1125-2016; Stenman, Adam/C-6131-2015; Muth,
   Andreas/AAV-5144-2021
OI Juhlin, Carl Christofer/0000-0002-5945-9081; Stenman,
   Adam/0000-0003-3248-7629; Muth, Andreas/0000-0003-0857-1521
FU Swedish Cancer Society; Swedish Research Council; Goran Gustafsson
   Foundation for Research in Natural Sciences and Medicine; Gustav V
   Jubilee Foundation; Karolinska Institutet; Stockholm County Council
FX The study was financially supported by grants from Swedish Cancer
   Society, Swedish Research Council, Goran Gustafsson Foundation for
   Research in Natural Sciences and Medicine, Gustav V Jubilee Foundation,
   Karolinska Institutet, and Stockholm County Council.
CR Agger K, 2008, CURR OPIN GENET DEV, V18, P159, DOI 10.1016/j.gde.2007.12.003
   Baysal BE, 2000, SCIENCE, V287, P848, DOI 10.1126/science.287.5454.848
   Benn DE, 2003, ONCOGENE, V22, P1358, DOI 10.1038/sj.onc.1206300
   Boedeker C C, 2011, Laryngorhinootologie, V90 Suppl 1, pS56, DOI 10.1055/s-0030-1270447
   Burnichon N, 2010, HUM MOL GENET, V19, P3011, DOI 10.1093/hmg/ddq206
   Cascon A, 2004, J MED GENET, V41, DOI 10.1136/jmg.2003.012658
   Castellano M, 2006, ANN NY ACAD SCI, V1073, P156, DOI 10.1196/annals.1353.016
   Cervera AM, 2008, CANCER RES, V68, P4058, DOI 10.1158/0008-5472.CAN-07-5580
   Cervera AM, 2009, MOL CANCER, V8, DOI 10.1186/1476-4598-8-89
   Comino-Mendez I, 2011, NAT GENET, V43, P663, DOI 10.1038/ng.861
   DeLellis RA, 2004, WHO CLASSIFICATION T, P136
   Elder EE, 2005, J SURG ONCOL, V89, P193, DOI 10.1002/jso.20177
   Elder EE, 2003, EUR J SURG ONCOL, V29, P278, DOI 10.1053/ejso.2002.1413
   Geli J, 2008, CLIN CANCER RES, V14, P2551, DOI 10.1158/1078-0432.CCR-07-1867
   Geli J, 2007, ENDOCR-RELAT CANCER, V14, P125, DOI 10.1677/ERC-06-0031
   Hao HX, 2009, SCIENCE, V325, P1139, DOI 10.1126/science.1175689
   Khorram-Manesh A, 2005, J INTERN MED, V258, P55, DOI 10.1111/j.1365-2796.2005.01504.x
   Kiss NB, 2008, ENDOCR-RELAT CANCER, V15, P609, DOI 10.1677/ERC-07-0285
   Kiss NB, 2012, BMC MED GENET, V13, DOI 10.1186/1471-2350-13-83
   Lee S, 2005, CANCER CELL, V8, P155, DOI 10.1016/j.ccr.2005.06.015
   Lin YC, 2007, ONCOGENE, V26, P7017, DOI 10.1038/sj.onc.1210507
   Margetts CDE, 2005, ENDOCR-RELAT CANCER, V12, P161, DOI 10.1677/erc.1.00865
   Muscarella P, 2008, GENE EXPRESSION, V14, P207, DOI 10.3727/105221608786883825
   Muth A, 2012, WORLD J SURG, V36, P1389, DOI 10.1007/s00268-012-1430-6
   Neumann HPH, 2002, NEW ENGL J MED, V346, P1459, DOI 10.1056/NEJMoa020152
   Pollard PJ, 2005, HUM MOL GENET, V14, P2231, DOI 10.1093/hmg/ddi227
   Qin YJ, 2010, NAT GENET, V42, P229, DOI 10.1038/ng.533
   Ricketts CJ, 2010, HUM MUTAT, V31, P41, DOI 10.1002/humu.21136
   Sandgren J, 2010, EXP MOL MED, V42, P484, DOI 10.3858/emm.2010.42.7.050
   Sandgren J, 2010, ENDOCR-RELAT CANCER, V17, P561, DOI 10.1677/ERC-09-0310
   Selak MA, 2005, CANCER CELL, V7, P77, DOI 10.1016/j.ccr.2004.11.022
   Smith EH, 2007, HUM MOL GENET, V16, P3136, DOI 10.1093/hmg/ddm275
   Tischler AS, 2008, ARCH PATHOL LAB MED, V132, P1272, DOI 10.1043/1543-2165(2008)132[1272:PAEPU]2.0.CO;2
   Toyota M, 1999, P NATL ACAD SCI USA, V96, P8681, DOI 10.1073/pnas.96.15.8681
   van Nederveen FH, 2009, LANCET ONCOL, V10, P764, DOI 10.1016/S1470-2045(09)70164-0
   Wangberg B, 2006, ANN NY ACAD SCI, V1073, P512, DOI 10.1196/annals.1353.054
   Yao L, 2010, JAMA-J AM MED ASSOC, V304, P2611, DOI 10.1001/jama.2010.1830
   You MJ, 2002, P NATL ACAD SCI USA, V99, P1455, DOI 10.1073/pnas.022632099
NR 38
TC 13
Z9 13
U1 0
U2 1
PU BIOSCIENTIFICA LTD
PI BRISTOL
PA EURO HOUSE, 22 APEX COURT WOODLANDS, BRADLEY STOKE, BRISTOL BS32 4JT,
   ENGLAND
SN 1351-0088
EI 1479-6821
J9 ENDOCR-RELAT CANCER
JI Endocr.-Relat. Cancer
PD FEB
PY 2013
VL 20
IS 1
BP 65
EP 78
DI 10.1530/ERC-12-0267
PG 14
WC Oncology; Endocrinology & Metabolism
WE Science Citation Index Expanded (SCI-EXPANDED)
SC Oncology; Endocrinology & Metabolism
GA 121RZ
UT WOS:000317263200008
PM 23154831
OA Green Published
DA 2023-08-27
ER

PT J
AU Eckardt, L
   Prange-Barczynska, M
   Hodson, EJ
   Fielding, JW
   Cheng, XT
   Lima, JDCC
   Kurlekar, S
   Douglas, G
   Ratcliffe, PJ
   Bishop, T
AF Eckardt, Luise
   Prange-Barczynska, Maria
   Hodson, Emma J.
   Fielding, James W.
   Cheng, Xiaotong
   Lima, Joanna D. C. C.
   Kurlekar, Samvid
   Douglas, Gillian
   Ratcliffe, Peter J.
   Bishop, Tammie
TI Developmental role of PHD2 in the pathogenesis of pseudohypoxic
   pheochromocytoma
SO ENDOCRINE-RELATED CANCER
LA English
DT Article
DE PHD; HIF; hypoxia; adrenal medulla; pheochromocytoma
ID HETEROZYGOUS DEFICIENCY; TYROSINE-HYDROXYLASE; ADRENAL-MEDULLA; HIF2A
   MUTATIONS; CRE RECOMBINASE; PARAGANGLIOMA; HYPOXIA; HYPERPLASIA;
   HIF1-ALPHA; EXPRESSION
AB Despite a general role for the HIF hydroxylase system in cellular oxygen sensing and tumour hypoxia, cancer-associated mutations of genes in this pathway, including PHD2, PHD1, EPAS1 (encoding HIF-2 alpha) are highly tissue-restricted, being observed in pseudohypoxic pheochromocytoma and paraganglioma (PPGL) but rarely, if ever, in other tumours. In an effort to understand that paradox and gain insights into the pathogenesis of pseudohypoxic PPGL, we constructed mice in which the principal HIF prolyl hydroxylase, Phd2, is inactivated in the adrenal medulla using TH-restricted Cre recombinase. Investigation of these animals revealed a gene expression pattern closely mimicking that of pseudohypoxic PPGL. Spatially resolved analyses demonstrated a binary distribution of two contrasting patterns of gene expression among adrenal medullary cells. Phd2 inactivation resulted in a marked shift in this distribution towards a Pnmt(-)/Hif-2 alpha(+)/Rgs5(+) population. This was associated with morphological abnormalities of adrenal development, including ectopic TH+ cells within the adrenal cortex and external to the adrenal gland. These changes were ablated by combined inactivation of Phd2 with Hif-2 alpha, but not Hif-1 alpha. However, they could not be reproduced by inactivation of Phd2 in adult life, suggesting that they arise from dysregulation of this pathway during adrenal development. Together with the clinical observation that pseudohypoxic PPGL manifests remarkably high heritability, our findings suggest that this type of tumour likely arises from dysregulation of a tissue-restricted action of the PHD2/HIF-2 alpha pathway affecting adrenal development in early life and provides a model for the study of the relevant processes.
C1 [Eckardt, Luise; Prange-Barczynska, Maria; Fielding, James W.; Cheng, Xiaotong; Lima, Joanna D. C. C.; Kurlekar, Samvid; Ratcliffe, Peter J.; Bishop, Tammie] Univ Oxford, Target Discovery Inst, Oxford, England.
   [Eckardt, Luise] Heidelberg Univ, Inst Physiol & Pathophysiol, Heidelberg, Germany.
   [Prange-Barczynska, Maria; Fielding, James W.; Cheng, Xiaotong; Ratcliffe, Peter J.] Univ Oxford, Ludwig Inst Canc Res, Oxford, England.
   [Hodson, Emma J.; Ratcliffe, Peter J.] Francis Crick Inst, London, England.
   [Hodson, Emma J.] Univ Cambridge, Dept Expt Med & Immunotherapeut, Cambridge, England.
   [Douglas, Gillian] Univ Oxford, John Radcliffe Hosp, BHF Ctr Res Excellence, Radcliffe Dept Med,Div Cardiovasc Med, Oxford, England.
C3 University of Oxford; Ruprecht Karls University Heidelberg; Ludwig
   Institute for Cancer Research; University of Oxford; Francis Crick
   Institute; University of Cambridge; University of Oxford
RP Ratcliffe, PJ; Bishop, T (通讯作者)，Univ Oxford, Target Discovery Inst, Oxford, England.; Ratcliffe, PJ (通讯作者)，Univ Oxford, Ludwig Inst Canc Res, Oxford, England.; Ratcliffe, PJ (通讯作者)，Francis Crick Inst, London, England.
EM
RI Bishop, Tammie/HPI-1663-2023
OI Bishop, Tammie/0000-0003-3308-9139; Carola Correia Lima, Joanna
   Darck/0000-0001-6804-8666; Prange-Barczynska, Maria/0000-0002-5378-5994;
   Eckardt, Luise/0000-0003-2390-0012; Kurlekar, Samvid/0000-0003-1633-1827
FU Oxford Branch of the Ludwig Institute for Cancer Research; Wellcome
   Trust [FC001501, 106241/Z/14/Z]; Paradifference Foundation; Francis
   Crick Institute from Cancer Research UK [FC001501]; UK Medical Research
   Council [FC001501]; Boehringer Ingelheim Fonds; FAPESP fellowship
   [2018/20083-1]; Christopher Welch Scholarship; Clarendon Fund
FX Funding for the work was received from the Oxford Branch of the Ludwig
   Institute for Cancer Research, the Wellcome Trust (106241/Z/14/Z) and
   the Paradifference Foundation. This work was also supported by the
   Francis Crick Institute, which receives its core funding from Cancer
   Research UK (FC001501), the UK Medical Research Council (FC001501), and
   the Wellcome Trust (FC001501). L E was sponsored by an MD fellowship
   from Boehringer Ingelheim Fonds; J D C C L by a FAPESP fellowship
   (2018/20083-1); S K by a Christopher Welch Scholarship and the Clarendon
   Fund.
CR Al Khazal F, 2021, FASEB J, V35, DOI 10.1096/fj.202002100R
   Toledo RA, 2017, ENDOCR-RELAT CANCER, V24, pC9, DOI 10.1530/ERC-16-0479
   Bankhead P, 2017, SCI REP-UK, V7, DOI 10.1038/s41598-017-17204-5
   Bayley JP, 2009, PLOS ONE, V4, DOI 10.1371/journal.pone.0007987
   Bishop T, 2008, MOL CELL BIOL, V28, P3386, DOI 10.1128/MCB.02041-07
   Bishop T, 2014, HYPOXIA, V2, P197, DOI 10.2147/HP.S47598
   Bishop T, 2013, J PHYSIOL-LONDON, V591, P3565, DOI 10.1113/jphysiol.2012.247254
   Buffet A, 2020, BEST PRACT RES CL EN, V34, DOI 10.1016/j.beem.2020.101416
   Burnichon N, 2011, HUM MOL GENET, V20, P3974, DOI 10.1093/hmg/ddr324
   Chan WH, 2019, SCI REP-UK, V9, DOI 10.1038/s41598-019-40501-0
   Cheng XT, 2020, J CLIN INVEST, V130, P2237, DOI 10.1172/JCI133194
   Choueiri TK, 2020, NAT MED, V26, P1519, DOI 10.1038/s41591-020-1093-z
   Coupland Douglas., 2017, HISTORY
   COUPLAND RE, 1966, NATURE, V209, P590, DOI 10.1038/209590a0
   Cramer T, 2003, CELL, V112, P645, DOI 10.1016/S0092-8674(03)00154-5
   Crona J, 2017, ENDOCR REV, V38, P489, DOI 10.1210/er.2017-00062
   Dahia PLM, 2005, PLOS GENET, V1, P72, DOI 10.1371/journal.pgen.0010008
   Diaz-Castro B, 2012, MOL CELL BIOL, V32, P3347, DOI 10.1128/MCB.00128-12
   Eisenhofer G, 2001, J CLIN ENDOCR METAB, V86, P1999, DOI 10.1210/jc.86.5.1999
   Favier J, 2012, NEW ENGL J MED, V367, P2161, DOI 10.1056/NEJMc1211953
   Favier J, 2009, PLOS ONE, V4, DOI 10.1371/journal.pone.0007094
   Fielding JW, 2018, J PHYSIOL-LONDON, V596, P4393, DOI 10.1113/JP275996
   Fishbein L, 2018, CELL TISSUE RES, V372, P339, DOI 10.1007/s00441-018-2795-0
   Fishbein L, 2017, CANCER CELL, V31, P181, DOI 10.1016/j.ccell.2017.01.001
   Furlan A, 2017, SCIENCE, V357, P46, DOI 10.1126/science.aal3753
   Gao L, 2019, RESP PHYSIOL NEUROBI, V265, P100, DOI 10.1016/j.resp.2018.08.007
   Gruber M, 2007, P NATL ACAD SCI USA, V104, P2301, DOI 10.1073/pnas.0608382104
   Hanemaaijer ES, 2021, P NATL ACAD SCI USA, V118, DOI 10.1073/pnas.2022350118
   Hodson EJ, 2016, J PHYSIOL-LONDON, V594, P1179, DOI 10.1113/JP271050
   Jin Y, 2009, J BIOL CHEM, V284, P23436, DOI 10.1074/jbc.M109.032664
   Kaelin WG, 2008, NAT REV CANCER, V8, P865, DOI 10.1038/nrc2502
   Ladroue C, 2008, NEW ENGL J MED, V359, P2685, DOI 10.1056/NEJMoa0806277
   Lee S, 2005, CANCER CELL, V8, P155, DOI 10.1016/j.ccr.2005.06.015
   Lepoutre-Lussey C, 2016, MOL CELL ENDOCRINOL, V421, P40, DOI 10.1016/j.mce.2015.06.027
   Lindeberg J, 2004, GENESIS, V40, P67, DOI 10.1002/gene.20065
   Lorenzo FR, 2013, J MOL MED, V91, P507, DOI 10.1007/s00109-012-0967-z
   Macias D, 2018, ELIFE, V7, DOI [10.7554/elife.34681, 10.7554/eLife.34681]
   Macias D, 2014, EMBO MOL MED, V6, P1577, DOI 10.15252/emmm.201404153
   Mazzone M, 2009, CELL, V136, P839, DOI 10.1016/j.cell.2009.01.020
   Nagendran M, 2018, ELIFE, V7, DOI 10.7554/eLife.30510
   Opotowsky AR, 2015, J CLIN ENDOCR METAB, V100, P1325, DOI 10.1210/jc.2014-3863
   Park WJ, 2015, ENDOCR-RELAT CANCER, V22, P623, DOI 10.1530/ERC-15-0058
   Piruat JI, 2004, MOL CELL BIOL, V24, P10933, DOI 10.1128/MCB.24.24.10933-10940.2004
   Rotolo T, 2008, PLOS ONE, V3, DOI 10.1371/journal.pone.0004099
   Semenza GL, 2019, ANNU REV PHARMACOL, V59, P379, DOI 10.1146/annurev-pharmtox-010818-021637
   Shen C, 2011, CANCER DISCOV, V1, P222, DOI 10.1158/2159-8290.CD-11-0098
   Smith-Hicks CL, 2000, EMBO J, V19, P612, DOI 10.1093/emboj/19.4.612
   Tai TC, 2009, J NEUROCHEM, V109, P513, DOI 10.1111/j.1471-4159.2009.05978.x
   Thompson RJ, 1997, J PHYSIOL-LONDON, V498, P503, DOI 10.1113/jphysiol.1997.sp021876
   Tischler AS, 2008, ARCH PATHOL LAB MED, V132, P1272, DOI 10.1043/1543-2165(2008)132[1272:PAEPU]2.0.CO;2
   Toledo RA, 2013, ENDOCR-RELAT CANCER, V20, P349, DOI 10.1530/ERC-13-0101
   Vaidya A, 2018, NEW ENGL J MED, V378, P1259, DOI 10.1056/NEJMc1716652
   VERHOFSTAD AAJ, 1979, CELL TISSUE RES, V200, P1
   Vollmer RR, 1996, CLIN EXP HYPERTENS, V18, P731, DOI 10.3109/10641969609081778
   Vooijs M, 2001, EMBO REP, V2, P292, DOI 10.1093/embo-reports/kve064
   Waldmann J, 2010, ENDOCR-RELAT CANCER, V17, P743, DOI 10.1677/ERC-09-0118
   Welander J, 2014, ENDOCR-RELAT CANCER, V21, P495, DOI 10.1530/ERC-13-0384
   Yang CZ, 2015, J MOL MED, V93, P93, DOI 10.1007/s00109-014-1205-7
   Zhou T, 2016, J PHYSIOL-LONDON, V594, P4225, DOI 10.1113/JP271936
   Zhuang ZP, 2012, NEW ENGL J MED, V367, P922, DOI 10.1056/NEJMoa1205119
NR 60
TC 5
Z9 5
U1 0
U2 11
PU BIOSCIENTIFICA LTD
PI BRISTOL
PA STARLING HOUSE, 1600 BRISTOL PARKWAY N, BRISTOL, ENGLAND
SN 1351-0088
EI 1479-6821
J9 ENDOCR-RELAT CANCER
JI Endocr.-Relat. Cancer
PD DEC 1
PY 2021
VL 28
IS 12
BP 757
EP 772
DI 10.1530/ERC-21-0211
PG 16
WC Oncology; Endocrinology & Metabolism
WE Science Citation Index Expanded (SCI-EXPANDED)
SC Oncology; Endocrinology & Metabolism
GA YT3PF
UT WOS:000751275100003
PM 34658364
OA Green Published, hybrid
DA 2023-08-27
ER

PT J
AU Acharya, S
   Liu, JF
   Tatevossian, RG
   Chiang, J
   Qaddoumi, I
   Gajjar, A
   Walker, D
   Harreld, JH
   Merchant, TE
   Ellison, DW
AF Acharya, Sahaja
   Liu, Jo-Fen
   Tatevossian, Ruth G.
   Chiang, Jason
   Qaddoumi, Ibrahim
   Gajjar, Amar
   Walker, David
   Harreld, Julie H.
   Merchant, Thomas E.
   Ellison, David W.
TI Risk stratification in pediatric low-grade glioma and glioneuronal tumor
   treated with radiation therapy: an integrated clinicopathologic and
   molecular analysis
SO NEURO-ONCOLOGY
LA English
DT Article
DE pediatric low-grade glioma; radiation; risk stratification; survival
ID BRAF V600E STATUS; CHILDHOOD; CHILDREN; ENDOCRINE; ONCOLOGY; SURVIVAL;
   MUTATION; OUTCOMES
AB Background. Management of unresectable pediatric low-grade glioma and glioneuronal tumor (LGG/LGGNT) is controversial.There are no validated prognostic features to guide use of radiation therapy (RT). Our study aimed to identify negative prognostic features in patients treated with RT using clinicopathologic and molecular data and validate these findings in an external dataset.
   Methods. Children with non-metastatic, biopsy-proven unresectable LGG/LGGNT treated with RT at a single institution between 1997 and 2017 were identified. Recursive partitioning analysis (RPA) was used to stratify patients into low- and high-risk prognostic groups based on overall survival (OS). CNS9702 data were used for validation.
   Results. One hundred and fifty patients met inclusion criteria. Median follow-up was 11.4 years. RPA yielded low-and high-risk groups with 10-year OS of 95.6% versus 76.4% (95% CI: 88.7%-98.4% vs 59.3%-87.1%, P= 0.003), respectively.These risk groups were validated using CNS9702 dataset (n = 48) (4-year OS: low-risk vs high-risk: 100% vs 64%, P < 0.001). High-risk tumors included diffuse astrocytoma or location within thalamus/midbrain. Low-risk tumors included pilocytic astrocytoma/ganglioglioma located outside of the thalamus/midbrain. In the subgroup with known BRAF status (n = 49), risk stratification remained prognostic independently of BRAF alteration (V600E or fusion). Within the high-risk group, delayed RT, defined as RT after at least one line of chemotherapy, was associated with a further decrement in overall survival (P= 0.021).
   Conclusion. A high-risk subgroup of patients, defined by diffuse astrocytoma histology or midbrain/thalamus tumor location, have suboptimal long-term survival and might benefit from timely use of RT.These results require validation.
C1 [Acharya, Sahaja; Merchant, Thomas E.] St Jude Childrens Res Hosp, Dept Radiat Oncol, 262 DannyThomas Pl,MS 210, Memphis, TN 38105 USA.
   [Liu, Jo-Fen; Walker, David] Univ Nottingham, Childrens Brain Tumor Res Ctr, Nottingham, England.
   [Qaddoumi, Ibrahim; Gajjar, Amar] St Jude Childrens Res Hosp, Div Neurooncol, Memphis, TN 38105 USA.
   [Harreld, Julie H.] St Jude Childrens Res Hosp, Dept Diagnost Imaging, Memphis, TN 38105 USA.
   [Tatevossian, Ruth G.; Chiang, Jason; Ellison, David W.] St Jude Childrens Res Hosp, Dept Pathol, Memphis, TN 38105 USA.
C3 St Jude Children's Research Hospital; University of Nottingham; St Jude
   Children's Research Hospital; St Jude Children's Research Hospital; St
   Jude Children's Research Hospital
RP Acharya, S (通讯作者)，St Jude Childrens Res Hosp, Dept Radiat Oncol, 262 DannyThomas Pl,MS 210, Memphis, TN 38105 USA.
EM
RI Chiang, Jason/N-2766-2018; Qaddoumi, Ibrahim/N-8101-2018; Ellison,
   David/N-4806-2018; Gajjar, Amar/N-8059-2018; Tatevossian,
   Ruth/G-6377-2016
OI Chiang, Jason/0000-0003-3623-8996; Tatevossian, Ruth/0000-0002-1653-3026
FU American Lebanese Syrian Associated Charities (ALSAC); National Cancer
   Institute Support Grant [P30CA021765]
FX All authors affiliated with St Jude Children's Research Hospital receive
   research support from the American Lebanese Syrian Associated Charities
   (ALSAC) and the National Cancer Institute Support Grant (P30CA021765).
CR Acharya S, 2019, J NEURO-ONCOL, V144, P603, DOI 10.1007/s11060-019-03264-2
   Acharya S, 2019, NEURO-ONCOLOGY, V21, P1175, DOI 10.1093/neuonc/noz068
   Acharya S, 2015, NEURO-ONCOLOGY, V17, P741, DOI 10.1093/neuonc/nou311
   Ater JL, 2012, J CLIN ONCOL, V30, P2641, DOI 10.1200/JCO.2011.36.6054
   Benton CB, 2018, AM J HEMATOL, V93, P1245, DOI 10.1002/ajh.25234
   Bitterman DS, 2019, J CLIN ONCOL, V37, P3335, DOI 10.1200/JCO.19.01270
   Capper D, 2012, ACTA NEUROPATHOL, V123, P223, DOI 10.1007/s00401-011-0887-y
   Cherlow JM, 2019, INT J RADIAT ONCOL, V103, P861, DOI 10.1016/j.ijrobp.2018.11.004
   CIAMPI A, 1988, J CLIN EPIDEMIOL, V41, P737, DOI 10.1016/0895-4356(88)90160-6
   Eisenhauer EA, 2009, EUR J CANCER, V45, P228, DOI 10.1016/j.ejca.2008.10.026
   Fangusaro J, 2019, LANCET ONCOL, V20, P1011, DOI 10.1016/S1470-2045(19)30277-3
   Fisher PG, 2008, PEDIATR BLOOD CANCER, V51, P245, DOI 10.1002/pbc.21563
   Friedman DL, 2010, JNCI-J NATL CANCER I, V102, P1083, DOI 10.1093/jnci/djq238
   Fuller C, 2018, NEURO-ONCOLOGY, V20, P109
   Gajjar A, 1997, J CLIN ONCOL, V15, P2792, DOI 10.1200/JCO.1997.15.8.2792
   Garcia MA, 2016, FUTURE ONCOL, V12, P1493, DOI 10.2217/fon-2016-0039
   Gilles FH, 2008, PEDIATR DEVEL PATHOL, V11, P108, DOI 10.2350/07-06-0303.1
   GNEKOW AK, 1995, MED PEDIATR ONCOL, V24, P104, DOI 10.1002/mpo.2950240209
   Gnekow AK, 2012, NEURO-ONCOLOGY, V14, P1265, DOI 10.1093/neuonc/nos202
   Greenberger BA, 2014, INT J RADIAT ONCOL, V89, P1060, DOI 10.1016/j.ijrobp.2014.04.053
   Indelicato DJ, 2019, INT J RADIAT ONCOL, V104, P149, DOI 10.1016/j.ijrobp.2019.01.078
   Jones DTW, 2008, CANCER RES, V68, P8673, DOI 10.1158/0008-5472.CAN-08-2097
   Jones DTW, 2018, J CLIN ONCOL, V36, P96, DOI 10.1200/JCO.2017.75.8987
   Kieran MW, 2018, J CLIN ONCOL, V36, DOI 10.1200/JCO.2018.36.15_suppl.10506
   Lassaletta A, 2017, J CLIN ONCOL, V35, P2934, DOI 10.1200/JCO.2016.71.8726
   Lassaletta A, 2016, J CLIN ONCOL, V34, P3537, DOI 10.1200/JCO.2016.68.1585
   Merchant TE, 2009, J CLIN ONCOL, V27, P3691, DOI 10.1200/JCO.2008.21.2738
   Merchant TE, 2009, J CLIN ONCOL, V27, P3598, DOI 10.1200/JCO.2008.20.9494
   Mistry M, 2015, J CLIN ONCOL, V33, P1015, DOI 10.1200/JCO.2014.58.3922
   Myung JK, 2012, TRANSL ONCOL, V5, P430, DOI 10.1593/tlo.12328
   Paulino AC, 2013, CANCER-AM CANCER SOC, V119, P2654, DOI 10.1002/cncr.28118
   Pignatti F, 2002, J CLIN ONCOL, V20, P2076, DOI 10.1200/JCO.2002.08.121
   POLLACK IF, 1995, J NEUROSURG, V82, P536, DOI 10.3171/jns.1995.82.4.0536
   Qaddoumi I, 2016, ACTA NEUROPATHOL, V131, P833, DOI 10.1007/s00401-016-1539-z
   Salloum R, 2017, ACTA NEUROPATHOL COM, V5, DOI 10.1186/s40478-017-0479-8
   Schindler G, 2011, ACTA NEUROPATHOL, V121, P397, DOI 10.1007/s00401-011-0802-6
   Stokland T, 2010, NEURO-ONCOLOGY, V12, P1257, DOI 10.1093/neuonc/noq092
   Tsang DS, 2017, INT J RADIAT ONCOL, V99, P642, DOI 10.1016/j.ijrobp.2017.07.023
   van den Bent MJ, 2010, ACTA NEUROPATHOL, V120, P297, DOI 10.1007/s00401-010-0725-7
   van Putten W., 2006, CART STATA MODULE PE
   Vatner RE, 2018, J CLIN ONCOL, V36, P2854, DOI 10.1200/JCO.2018.78.1492
NR 41
TC 10
Z9 10
U1 0
U2 9
PU OXFORD UNIV PRESS INC
PI CARY
PA JOURNALS DEPT, 2001 EVANS RD, CARY, NC 27513 USA
SN 1522-8517
EI 1523-5866
J9 NEURO-ONCOLOGY
JI Neuro-Oncology
PD AUG
PY 2020
VL 22
IS 8
BP 1203
EP 1213
DI 10.1093/neuonc/noaa031
PG 11
WC Oncology; Clinical Neurology
WE Science Citation Index Expanded (SCI-EXPANDED)
SC Oncology; Neurosciences & Neurology
GA OW8HH
UT WOS:000593120100017
PM 32052049
OA Green Accepted, Green Published, Bronze
DA 2023-08-27
ER

PT J
AU Rogers, HA
   Estranero, J
   Gudka, K
   Grundy, RG
AF Rogers, Hazel A.
   Estranero, Jasper
   Gudka, Keshni
   Grundy, Richard G.
TI The therapeutic potential of targeting the PI3K pathway in pediatric
   brain tumors
SO ONCOTARGET
LA English
DT Article
DE PI3K pathway; brain tumor; pediatric; therapy; cancer
ID CENTRAL-NERVOUS-SYSTEM; REFRACTORY SOLID TUMORS; PRECLINICAL TESTING
   PROGRAM; INTRINSIC PONTINE GLIOMA; CHILDRENS ONCOLOGY GROUP; LOW-GRADE
   GLIOMA; MEDULLOBLASTOMA CELL-PROLIFERATION; PHOSPHOINOSITIDE 3-KINASE
   PATHWAY; CHRONIC LYMPHOCYTIC-LEUKEMIA; PHASE-I
AB Central nervous system tumors are the most common cancer type in children and the leading cause of cancer related deaths. There is therefore a need to develop novel treatments. Large scale profiling studies have begun to identify alterations that could be targeted therapeutically, including the phosphoinositide 3-kinase (PI3K) signaling pathway, which is one of the most commonly activated pathways in cancer with many inhibitors under clinical development. PI3K signaling has been shown to be aberrantly activated in many pediatric CNS neoplasms. Pre-clinical analysis supports a role for PI3K signaling in the control of tumor growth, survival and migration as well as enhancing the cytotoxic effects of current treatments. Based on this evidence agents targeting PI3K signaling have begun to be tested in clinical trials of pediatric cancer patients. Overall, targeting the PI3K pathway presents as a promising strategy for the treatment of pediatric CNS tumors. In this review we examine the genetic alterations found in the PI3K pathway in pediatric CNS tumors and the pathological role it plays, as well as summarizing the current pre-clinical and clinical data supporting the use of PI3K pathway inhibitors for the treatment of these tumors.
C1 [Rogers, Hazel A.; Estranero, Jasper; Gudka, Keshni; Grundy, Richard G.] Univ Nottingham, Childrens Brain Tumour Res Ctr, Sch Med, Nottingham NG7 2RD, England.
C3 University of Nottingham
RP Rogers, HA (通讯作者)，Univ Nottingham, Childrens Brain Tumour Res Ctr, Sch Med, Nottingham NG7 2RD, England.
EM
OI Rogers, Hazel/0000-0002-1450-5363; Grundy, Richard/0000-0002-2585-2539
FU University of Nottingham
FX There was no specific funding source for the information presented in
   this review article. Authors were supported by funding from the
   University of Nottingham.
CR Adriaensen MEAPM, 2009, EUR J NEUROL, V16, P691, DOI 10.1111/j.1468-1331.2009.02567.x
   Ali K, 2014, NATURE, V510, P407, DOI 10.1038/nature13444
   Arcaro A, 2007, BIOCHEM J, V406, P57, DOI 10.1042/BJ20070309
   Atkinson JM, 2011, CANCER CELL, V20, P384, DOI 10.1016/j.ccr.2011.08.013
   Backer JM, 2008, BIOCHEM J, V410, P1, DOI 10.1042/BJ20071427
   Bagatell R, 2014, PEDIATR BLOOD CANCER, V61, P833, DOI 10.1002/pbc.24874
   Baryawno N, 2010, CANCER RES, V70, P266, DOI 10.1158/0008-5472.CAN-09-0578
   Bax DA, 2009, CLIN CANCER RES, V15, P5753, DOI 10.1158/1078-0432.CCR-08-3210
   Blom T, 2010, J NEURO-ONCOL, V97, P217, DOI 10.1007/s11060-009-0014-2
   Boller D, 2012, ANTICANCER RES, V32, P3015
   BOS JL, 1989, CANCER RES, V49, P4682
   Broderick DK, 2004, CANCER RES, V64, P5048, DOI 10.1158/0008-5472.CAN-04-1170
   Broniscer A, 2004, ONCOLOGIST, V9, P197, DOI 10.1634/theoncologist.9-2-197
   Brugge J, 2007, CANCER CELL, V12, P104, DOI 10.1016/j.ccr.2007.07.014
   Buonamici S, 2010, SCI TRANSL MED, V2, DOI 10.1126/scitranslmed.3001599
   Carol H, 2010, PEDIATR BLOOD CANCER, V55, P1329, DOI 10.1002/pbc.22710
   Chakravarti A, 2004, J CLIN ONCOL, V22, P1926, DOI 10.1200/JCO.2004.07.193
   Chan TSY, 2016, CANCER RES, V76, P2802, DOI 10.1158/0008-5472.CAN-15-2194
   Crawford J, 2013, PEDIATR REV, V34, P63, DOI 10.1542/pir.34-2-63
   Dasgupta T, 2013, NEUROONCOLOGY S1, V15, DOI [10.1093/neuonc/not047, DOI 10.1093/NEUONC/NOT047]
   Dodd KM, 2016, SEMIN CELL DEV BIOL, V52, P3, DOI 10.1016/j.semcdb.2016.01.025
   Ehrhardt M, 2015, ONCOTARGET, V6, P802, DOI 10.18632/oncotarget.2742
   Faivre S, 2006, NAT REV DRUG DISCOV, V5, P671, DOI 10.1038/nrd2062
   Falasca M, 2012, BIOCHEM J, V443, P587, DOI 10.1042/BJ20120008
   Faury D, 2007, J CLIN ONCOL, V25, P1196, DOI 10.1200/JCO.2006.07.8626
   Fouladi M, 2007, J CLIN ONCOL, V25, P4806, DOI 10.1200/JCO.2007.11.4017
   Fouladi M, 2015, CLIN CANCER RES, V21, P1558, DOI 10.1158/1078-0432.CCR-14-0595
   Fouladi M, 2014, PEDIATR BLOOD CANCER, V61, P1246, DOI 10.1002/pbc.25023
   Frappaz D, 2016, EUR J CANCER, V62, P9, DOI 10.1016/j.ejca.2016.03.084
   Fritsch R, 2013, CELL, V153, P1050, DOI 10.1016/j.cell.2013.04.031
   Gallia GL, 2006, MOL CANCER RES, V4, P709, DOI 10.1158/1541-7786.MCR-06-0172
   Geoerger B, 2012, EUR J CANCER, V48, P253, DOI 10.1016/j.ejca.2011.09.021
   Gilbertson RJ, 1997, CANCER RES, V57, P3272
   Gilbertson RJ, 2002, CLIN CANCER RES, V8, P3054
   Gills JJ, 2009, CURR ONCOL REP, V11, P102, DOI 10.1007/s11912-009-0016-4
   Gore L, 2013, CLIN CANCER RES, V19, P3649, DOI 10.1158/1078-0432.CCR-12-3166
   Grill J, 2012, PEDIATR BLOOD CANCER, V58, P489, DOI 10.1002/pbc.24060
   Guan S, 2011, NEURO-ONCOLOGY, V13, P748, DOI 10.1093/neuonc/nor037
   Guerreiro AS, 2008, CLIN CANCER RES, V14, P6761, DOI 10.1158/1078-0432.CCR-08-0385
   Guerreiro AS, 2011, MOL CANCER RES, V9, P925, DOI 10.1158/1541-7786.MCR-10-0200
   Guo YA, 2013, J PATHOL, V230, P17, DOI 10.1002/path.4176
   Hartmann W, 2006, CLIN CANCER RES, V12, P3019, DOI 10.1158/1078-0432.CCR-05-2187
   Hirsch E, 2014, ONCOGENE, V33, P3083, DOI 10.1038/onc.2013.265
   Hoellenriegel J, 2011, BLOOD, V118, P3603, DOI 10.1182/blood-2011-05-352492
   Hutt-Cabezas M, 2013, NEURO-ONCOLOGY, V15, P1604, DOI 10.1093/neuonc/not132
   Huynh H, 2015, MOL CANCER THER, V14, P1224, DOI 10.1158/1535-7163.MCT-14-0768
   Inoki K, 2002, NAT CELL BIOL, V4, P648, DOI 10.1038/ncb839
   Jentoft M, 2011, INT J CLIN EXP PATHO, V4, P43
   Johnson RA, 2010, NATURE, V466, P632, DOI 10.1038/nature09173
   Jones DTW, 2008, CANCER RES, V68, P8673, DOI 10.1158/0008-5472.CAN-08-2097
   Kagawa N, 2006, BRAIN TUMOR PATHOL, V23, P41, DOI 10.1007/s10014-006-0201-1
   Kang MH, 2014, PEDIATR BLOOD CANCER, V61, P1486, DOI 10.1002/pbc.24989
   Kaul A, 2012, GENE DEV, V26, P2561, DOI 10.1101/gad.200907.112
   Kieran M, 2013, PEDIAT BLOOD CANC, V60, DOI [10.1002/pbc.24719, DOI 10.1002/PBC.24719]
   Kilday JP, 2014, CURR NEUROL NEUROSCI, V14, DOI 10.1007/s11910-014-0441-0
   Klempner SJ, 2013, CANCER DISCOV, V3, P1345, DOI 10.1158/2159-8290.CD-13-0063
   Knobbe CB, 2003, BRAIN PATHOL, V13, P507
   Kool M, 2012, ACTA NEUROPATHOL, V123, P473, DOI 10.1007/s00401-012-0958-8
   Li J, 1997, SCIENCE, V275, P1943, DOI 10.1126/science.275.5308.1943
   Liu PD, 2015, CANCER DISCOV, V5, P1194, DOI 10.1158/2159-8290.CD-15-0460
   Liu PX, 2009, NAT REV DRUG DISCOV, V8, P627, DOI 10.1038/nrd2926
   Liu WD, 2014, DIAGN PATHOL, V9, DOI 10.1186/1746-1596-9-132
   Louis DN, 2016, ACTA NEUROPATHOL, V131, P803, DOI 10.1007/s00401-016-1545-1
   Mack SC, 2014, NATURE, V506, P445, DOI 10.1038/nature13108
   Modena P, 2012, NEURO-ONCOLOGY, V14, P1346, DOI 10.1093/neuonc/nos245
   Mohankumar KM, 2015, NAT GENET, V47, P878, DOI 10.1038/ng.3323
   Morgenstern DA, 2014, PEDIATR BLOOD CANCER, V61, P128, DOI 10.1002/pbc.24656
   Motzer RJ, 2008, LANCET, V372, P449, DOI 10.1016/S0140-6736(08)61039-9
   Mueller S, 2012, NEURO-ONCOLOGY, V14, P1146, DOI 10.1093/neuonc/nos140
   Nikbakht H, 2016, NAT COMMUN, V7, DOI 10.1038/ncomms11185
   Northcott PA, 2012, NAT REV CANCER, V12, P818, DOI 10.1038/nrc3410
   Okkenhaug K, 2016, CANCER DISCOV, V6, P1090, DOI 10.1158/2159-8290.CD-16-0716
   Parsons DW, 2008, SCIENCE, V321, P1807, DOI 10.1126/science.1164382
   Parsons DW, 2011, SCIENCE, V331, P435, DOI 10.1126/science.1198056
   Paugh BS, 2013, CANCER RES, V73, P6219, DOI 10.1158/0008-5472.CAN-13-1491
   Paugh BS, 2010, J CLIN ONCOL, V28, P3061, DOI 10.1200/JCO.2009.26.7252
   Pearson AD, 2016, ONCOTARGET, DOI 10.18632/oncotarget.12450
   Peris-Bonet R, 2006, EUR J CANCER, V42, P2064, DOI 10.1016/j.ejca.2006.05.009
   Pfister S, 2008, J CLIN INVEST, V118, P1739, DOI 10.1172/JCI33656
   Pollack IF, 2006, J NEUROSURG, V105, P418, DOI 10.3171/ped.2006.105.5.418
   Pollack IF, 2010, J NEURO-ONCOL, V99, P155, DOI 10.1007/s11060-010-0297-3
   Reddy AT, 2005, J NEURO-ONCOL, V75, P309, DOI 10.1007/s11060-005-6762-8
   Reynolds CP, 2013, PEDIATR BLOOD CANCER, V60, P791, DOI 10.1002/pbc.24301
   Rodriguez EF, 2011, ACTA NEUROPATHOL, V121, P407, DOI 10.1007/s00401-010-0784-9
   Rogers HA, 2013, CLIN CANCER RES, V19, P6450, DOI 10.1158/1078-0432.CCR-13-0222
   Samad N, 2010, ONCOTARGETS THER, V3, P167, DOI 10.2147/OTT.S8147
   Samuels Y, 2004, SCIENCE, V304, P554, DOI 10.1126/science.1096502
   Shymanets A, 2013, J BIOL CHEM, V288, P31059, DOI 10.1074/jbc.M113.508234
   Singh A, 2013, MOL ONCOL, V7, P497, DOI 10.1016/j.molonc.2013.01.001
   Spence T, 2014, ACTA NEUROPATHOL, V128, P291, DOI 10.1007/s00401-014-1291-1
   Spence T, 2014, NEURO-ONCOLOGY, V16, P62, DOI 10.1093/neuonc/not162
   Spunt SL, 2011, J CLIN ONCOL, V29, P2933, DOI 10.1200/JCO.2010.33.4649
   Stemke-Hale K, 2008, CANCER RES, V68, P6084, DOI 10.1158/0008-5472.CAN-07-6854
   Thorarinsdottir HK, 2008, CLIN CANCER RES, V14, P3386, DOI 10.1158/1078-0432.CCR-07-1616
   Thorpe LM, 2015, NAT REV CANCER, V15, P7, DOI 10.1038/nrc3860
   Vanhaesebroeck B, 1999, EXP CELL RES, V253, P239, DOI 10.1006/excr.1999.4701
   Vilar E, 2011, MOL CANCER THER, V10, P395, DOI 10.1158/1535-7163.MCT-10-0905
   Wheless JW, 2014, J CHILD NEUROL, V29, P1562, DOI 10.1177/0883073813501870
   Wojtalla A, 2012, PLOS ONE, V7, DOI 10.1371/journal.pone.0047109
   Wu G, 2014, NAT GENET, V46, P444, DOI 10.1038/ng.2938
   Yalon M, 2013, PEDIATR BLOOD CANCER, V60, P71, DOI 10.1002/pbc.24142
   Yang QS, 2015, CLIN CANCER RES, V21, P1537, DOI 10.1158/1078-0432.CCR-14-2034
   Yap TA, 2015, CURR OPIN PHARMACOL, V23, P98, DOI 10.1016/j.coph.2015.05.016
   Yuan RR, 2009, J HEMATOL ONCOL, V2, DOI 10.1186/1756-8722-2-45
   Zacharoulis S, 2007, PEDIATR BLOOD CANCER, V49, P34, DOI 10.1002/pbc.20935
   Zarghooni M, 2010, J CLIN ONCOL, V28, P1337, DOI 10.1200/JCO.2009.25.5463
   Zhang JH, 2013, NAT GENET, V45, P602, DOI 10.1038/ng.2611
   Zhao L, 2010, CELL CYCLE, V9, P596, DOI 10.4161/cc.9.3.10599
   Zoncu R, 2011, NAT REV MOL CELL BIO, V12, P21, DOI 10.1038/nrm3025
NR 109
TC 14
Z9 14
U1 0
U2 9
PU IMPACT JOURNALS LLC
PI ORCHARD PARK
PA 6666 E QUAKER ST, STE 1, ORCHARD PARK, NY 14127 USA
EI 1949-2553
J9 ONCOTARGET
JI Oncotarget
PD JAN 10
PY 2017
VL 8
IS 2
BP 2083
EP 2095
DI 10.18632/oncotarget.13781
PG 13
WC Oncology; Cell Biology
WE Science Citation Index Expanded (SCI-EXPANDED)
SC Oncology; Cell Biology
GA EH1EL
UT WOS:000391506300015
PM 27926496
OA Green Published, Green Submitted, gold
DA 2023-08-27
ER

PT J
AU Lyons, JL
   Neagu, MR
   Norton, IH
   Klein, JP
AF Lyons, Jennifer L.
   Neagu, Martha R.
   Norton, Isaiah H.
   Klein, Joshua P.
TI Diffusion tensor imaging in brainstem tuberculoma
SO JOURNAL OF CLINICAL NEUROSCIENCE
LA English
DT Article
DE CNS infection; Diffusion tensor imaging; DTI; Tractography; Tuberculoma
AB Integrity of descending white matter tracts can be evaluated by diffusion tensor imaging. In rim-enhancing intraparenchymal lesions, this technique can assist in the differentiation of demyelinating disease from tumor or abscess. Diffusion tensor imaging characteristics of tuberculoma have not been previously reported to our knowledge. A patient with headaches, dizziness, and mild left-sided weakness underwent MRI with diffusion tensor imaging. A large, rim-enhancing lesion within the pons was discovered, which subsequently was diagnosed as tuberculoma. Tractography maps prepared from diffusion tensor imaging data revealed predominantly displaced descending fiber tracts in the region of the rim-enhancing lesion. A few tracts adjacent to the lesion appeared truncated, and this abnormal finding correlated to the patient's clinical deficit. The tractography characteristics of diffusion tensor imaging in this patient potentially are distinct from those seen with demyelinating lesions, which may show more extensive tract truncation. Together with the consonance of exam findings and tract truncation seen in this patient, tractography may prove useful in the diagnosis of suspected tuberculoma. (C) 2013 Elsevier Ltd. All rights reserved.
C1 [Lyons, Jennifer L.; Neagu, Martha R.; Norton, Isaiah H.; Klein, Joshua P.] Brigham & Womens Hosp, Dept Neurol, Boston, MA 02115 USA.
   [Lyons, Jennifer L.; Neagu, Martha R.; Norton, Isaiah H.; Klein, Joshua P.] Harvard Univ, Sch Med, Boston, MA 02115 USA.
   [Klein, Joshua P.] Brigham & Womens Hosp, Dept Radiol, Boston, MA 02115 USA.
C3 Harvard University; Brigham & Women's Hospital; Harvard University;
   Harvard Medical School; Harvard University; Brigham & Women's Hospital
RP Klein, JP (通讯作者)，Brigham & Womens Hosp, Dept Neurol, 75 Francis St, Boston, MA 02115 USA.
EM
CR Giussani C, 2010, NEUROIMAGE, V52, P217, DOI 10.1016/j.neuroimage.2010.03.079
   Phillips NS, 2005, J NEUROSURG, V103, P557, DOI 10.3171/ped.2005.103.6.0557
   Sadeghi N, 2003, EUR RADIOL, V13, P2049, DOI 10.1007/s00330-002-1737-z
   Sahaiu-Srivastava S, 2008, CLIN NEUROL NEUROSUR, V110, P302, DOI 10.1016/j.clineuro.2007.11.002
NR 4
TC 5
Z9 6
U1 0
U2 2
PU ELSEVIER SCI LTD
PI OXFORD
PA THE BOULEVARD, LANGFORD LANE, KIDLINGTON, OXFORD OX5 1GB, OXON, ENGLAND
SN 0967-5868
EI 1532-2653
J9 J CLIN NEUROSCI
JI J. Clin. Neurosci.
PD NOV
PY 2013
VL 20
IS 11
BP 1598
EP 1599
DI 10.1016/j.jocn.2013.01.003
PG 2
WC Clinical Neurology; Neurosciences
WE Science Citation Index Expanded (SCI-EXPANDED)
SC Neurosciences & Neurology
GA 251CW
UT WOS:000326906400025
PM 23707604
DA 2023-08-27
ER

PT J
AU Csaki, C
   Ferencz, T
   Sipos, G
   Kopper, L
   Schuler, D
   Borsi, JD
AF Csaki, C
   Ferencz, T
   Sipos, G
   Kopper, L
   Schuler, D
   Borsi, JD
TI Diffuse plasmacytosis in a child with brainstem glioma following
   multiagent chemotherapy and intensive growth factor support
SO MEDICAL AND PEDIATRIC ONCOLOGY
LA English
DT Article
DE plasmacytosis; glioma; chemotherapy; growth factor; GM-CSF; G-CSF
ID COLONY-STIMULATING FACTOR; MULTIPLE-MYELOMA; GM-CSF; PROLIFERATION;
   LEUKEMIA; THERAPY
AB The use of granulocyte colony-stimulating factor (G-CSF) or granulocyte-macrophage-colony-stimulating factor (GM-CSF) in order to abrogate chemotherapy-induced neutropenia has become a routine part of many cancer treatment regimens. However, there are still very few data available about possible complications related to repeated or prolonged use of these agents in patients with malignant solid tumors. The authors report a child with brainstem glioma who received repeated cycles of multiagent chemotherapy with G- or GM-CSF support. During this period of 10 months, no clinical side effects were observed that could have been attributed to growth factor administration. However, postmortem histological examination revealed the presence of diffuse plasmacytosis, a rare hematological disorder in childhood. Undifferentiated plasma cells of nonmonoclonal origin could be demonstrated infiltrating bone marrow, lungs, and lymph nodes of the patient. Based on previously published in vitro and in vivo evidence on the interleukin-6 (IL-6)-mediated stimulatory effect of G- and GM-CSF on myeloma cell proliferation, the authors suggest a possible link between extensive growth factor support and the development of plasmacytosis in this patient. (C) 1996 Wiley-Liss, Inc.
C1 SEMMELWEIS UNIV MED, DEPT PEDIAT 2, H-1094 BUDAPEST, HUNGARY.
   SEMMELWEIS UNIV MED, DEPT PATHOL & EXPTL CANC RES 1, H-1094 BUDAPEST, HUNGARY.
C3 Semmelweis University; Semmelweis University
CR BARONI CD, 1977, CANCER, V40, P2329, DOI 10.1002/1097-0142(197711)40:5<2329::AID-CNCR2820400548>3.0.CO;2-M
   BONE RC, 1991, CHEST, V100, P802, DOI 10.1378/chest.100.3.802
   BONILLA MA, 1990, BLOOD, V76, pA133
   BOOGAERTS MA, 1994, EUR J CANCER, V30A, P238, DOI 10.1016/0959-8049(94)90099-X
   BRONCHUD M, 1993, ANTI-CANCER DRUG, V4, P127, DOI 10.1097/00001813-199304000-00002
   CELSING F, 1992, EUR J HAEMATOL, V49, P108
   GLASPY JA, 1988, ANN INTERN MED, V109, P789, DOI 10.7326/0003-4819-109-10-789
   Gonzalez Silva M., 1993, Sangre (Saragossa), V38, P47
   GROOPMAN JE, 1989, NEW ENGL J MED, V321, P1449, DOI 10.1056/NEJM198911233212106
   HERRMANN F, 1989, LEUKEMIA, V3, P335
   HORTEN BC, 1979, CANCER, V43, P258, DOI 10.1002/1097-0142(197901)43:1<258::AID-CNCR2820430137>3.0.CO;2-9
   KANZ L, 1991, AM J CLIN ONCOL-CANC, V14, pS27, DOI 10.1097/00000421-199112001-00006
   KEPES JJ, 1988, CANCER, V62, P391, DOI 10.1002/1097-0142(19880715)62:2<391::AID-CNCR2820620226>3.0.CO;2-7
   KLEIN B, 1992, HEMATOL ONCOL CLIN N, V6, P273, DOI 10.1016/S0889-8588(18)30344-7
   LEUSCHNER I, 1994, MED PEDIATR ONCOL, V23, P241
   LIESCHKE GJ, 1992, NEW ENGL J MED, V327, P28
   MARINA NM, 1994, MED PEDIATR ONCOL, V23, P328, DOI 10.1002/mpo.2950230403
   NORDAN RP, 1986, SCIENCE, V233, P566, DOI 10.1126/science.3726549
   POWLES R, 1990, LANCET, V336, P1417, DOI 10.1016/0140-6736(90)93111-2
   SCHRIBER JR, 1993, DRUG SAFETY, V8, P457, DOI 10.2165/00002018-199308060-00006
   VINK A, 1988, J IMMUNOL, V141, P1996
   ZHANG XG, 1990, BLOOD, V76, P2599
NR 22
TC 1
Z9 1
U1 0
U2 0
PU WILEY-LISS
PI HOBOKEN
PA DIV JOHN WILEY & SONS INC, 111 RIVER ST, HOBOKEN, NJ 07030 USA
SN 0098-1532
J9 MED PEDIATR ONCOL
JI Med. Pediatr. Oncol.
PD MAY
PY 1996
VL 26
IS 5
BP 367
EP 371
DI 10.1002/(SICI)1096-911X(199605)26:5<367::AID-MPO10>3.0.CO;2-A
PG 5
WC Oncology; Pediatrics
WE Science Citation Index Expanded (SCI-EXPANDED)
SC Oncology; Pediatrics
GA UA442
UT WOS:A1996UA44200010
PM 8614371
DA 2023-08-27
ER

PT J
AU Rachinger, J
   Buslei, R
   Prell, J
   Strauss, C
AF Rachinger, Jens
   Buslei, Rolf
   Prell, Julian
   Strauss, Christian
TI Solid haemangioblastomas of the CNS: a review of 17 consecutive cases
SO NEUROSURGICAL REVIEW
LA English
DT Article
DE Solid haemangioblastoma; Von Hippel-Lindau disease; Surgical outcome
ID HIPPEL-LINDAU-DISEASE; CENTRAL-NERVOUS-SYSTEM; GAMMA-KNIFE RADIOSURGERY;
   SPINAL-CORD HEMANGIOBLASTOMAS; BRAIN-STEM HEMANGIOBLASTOMAS;
   TUMOR-SUPPRESSOR GENE; SURGICAL-MANAGEMENT; PREOPERATIVE EMBOLIZATION;
   POSTERIOR-FOSSA; ENDOVASCULAR EMBOLIZATION
AB A retrospective study on solid central nervous system haemangioblastomas was performed to characterize clinical features, treatment strategies and outcome in these rare lesions. Between 1993 and 2006 23, solid haemangioblastomas were surgically removed in 17 patients. Eight lesions were located within pons Varolii and medulla oblongata, six within the cerebellar hemispheres and three in the cerebellopontine angle. Three haemangioblastomas were located supratentorially and three within the spinal cord. All patients except two underwent pre-operative magnetic resonance imaging (MRI). Post-operative digital subtraction angiography and/or MRI was performed in all surviving patients. Except for spinal cord lesions, rather unsystematic clinical symptoms were observed. Twenty-two tumours could be resected completely. Two patients with brainstem lesions died within 10 weeks after surgery from infectious complications. Persistent new neurological deficits occurred in two patients. Three patients underwent radiosurgery prior to or following the surgical procedure. Solid haemangioblastomas represent a surgical challenge due to their arteriovenous malformation-like vascularisation and their frequent location in eloquent areas. Surgery is the therapy of choice. Circumferential dissection with devascularization and en bloc removal yields good functional results. A location within the brainstem carries the most unfavourable prognosis.
C1 [Rachinger, Jens; Prell, Julian; Strauss, Christian] Univ Halle Wittenberg, Dept Neurosurg, D-06097 Halle, Germany.
   [Buslei, Rolf] Univ Erlangen Nurnberg, Dept Neuropathol, Erlangen, Germany.
C3 Martin Luther University Halle Wittenberg; University of Erlangen
   Nuremberg
RP Rachinger, J (通讯作者)，Univ Halle Wittenberg, Dept Neurosurg, Ernst Grube Str 40, D-06097 Halle, Germany.
EM
CR Biondi A, 2005, AM J NEURORADIOL, V26, P936
   Chang SD, 1998, NEUROSURGERY, V43, P28, DOI 10.1097/00006123-199807000-00018
   CHEN F, 1995, HUM MUTAT, V5, P66, DOI 10.1002/humu.1380050109
   Chen F, 1996, J MED GENET, V33, P716, DOI 10.1136/jmg.33.8.716
   CHOU SN, 1975, J NEUROSURG, V42, P23, DOI 10.3171/jns.1975.42.1.0023
   Conway JE, 2001, NEUROSURGERY, V48, P55, DOI 10.1097/00006123-200101000-00009
   Cornelius JF, 2007, J NEUROSURG, V106, P994, DOI 10.3171/jns.2007.106.6.994
   Dow GR, 2002, BRIT J NEUROSURG, V16, P168, DOI 10.1080/02688690220131804
   Eskridge JM, 1996, AM J NEURORADIOL, V17, P525
   FILLINGKATZ MR, 1991, NEUROLOGY, V41, P41, DOI 10.1212/WNL.41.1.41
   GLENN GM, 1992, JAMA-J AM MED ASSOC, V267, P1226, DOI 10.1001/jama.267.9.1226
   HO YS, 1990, SURG NEUROL, V33, P407, DOI 10.1016/0090-3019(90)90154-H
   HOSOE S, 1990, GENOMICS, V8, P634, DOI 10.1016/0888-7543(90)90249-T
   Jawahar A, 2000, ACTA NEUROCHIR, V142, P641, DOI 10.1007/s007010070107
   JULOW J, 1994, ACTA NEUROCHIR, V128, P109, DOI 10.1007/BF01400659
   Krishnan KG, 2006, ZBL NEUROCHIR, V67, P137, DOI 10.1055/s-2006-933360
   LATIF F, 1993, SCIENCE, V260, P1317, DOI 10.1126/science.8493574
   Lonser RR, 2003, J NEUROSURG, V98, P106, DOI 10.3171/jns.2003.98.1.0106
   Machein MR, 2000, J NEURO-ONCOL, V50, P109, DOI 10.1023/A:1006416003964
   Mans DA, 2008, EXP CELL RES, V314, P1229, DOI 10.1016/j.yexcr.2007.12.020
   NAIK RTS, 1995, CHILD NERV SYST, V11, P499, DOI 10.1007/BF00334976
   NEUMANN HPH, 1994, NEPHROL DIAL TRANSPL, V9, P1832
   Niemela M, 1996, J NEUROSURG, V85, P591, DOI 10.3171/jns.1996.85.4.0591
   OKAWARA SH, 1973, J NEUROSURG, V39, P514, DOI 10.3171/jns.1973.39.4.0514
   PALMER JJ, 1973, ACTA NEUROCHIR, V27, P125, DOI 10.1007/BF01401877
   Pan L, 1998, STEREOT FUNCT NEUROS, V70, P179, DOI 10.1159/000056420
   RAWE SE, 1978, SURG NEUROL, V9, P337
   Resche F, 1993, Adv Tech Stand Neurosurg, V20, P197
   Rodesch G, 2008, NEURORADIOLOGY, V50, P145, DOI 10.1007/s00234-007-0308-z
   Russel DS, 1989, PATHOLOGY TUMORS NER
   Ryu Stephen I, 2003, Neurosurg Focus, V15, pE10
   SANFORD RA, 1986, J NEUROSURG, V64, P317, DOI 10.3171/jns.1986.64.2.0317
   Santarpia L, 2007, J CELL MOL MED, V11, P1408, DOI 10.1111/j.1582-4934.2007.00122.x
   SEIZINGER BR, 1988, NATURE, V332, P268, DOI 10.1038/332268a0
   SINGOUNAS EG, 1978, ACTA NEUROCHIR, V44, P107, DOI 10.1007/BF01401634
   Spetzger U, 1996, NEUROSURG REV, V19, P147, DOI 10.1007/BF00512042
   SYMON L, 1993, ACTA NEUROCHIR, V120, P103, DOI 10.1007/BF02112026
   Tago M, 2005, J NEUROSURG, V102, P171, DOI 10.3171/jns.2005.102.s_supplement.0171
   Takeuchi S, 2001, NEUROL MED-CHIR, V41, P246, DOI 10.2176/nmc.41.246
   TAMPIERI D, 1993, NEUROSURGERY, V33, P502
   Van Velthoven V, 2003, NEUROSURGERY, V53, P1306, DOI 10.1227/01.NEU.0000093497.81390.29
   VazquezAnon V, 1997, NEURORADIOLOGY, V39, P86, DOI 10.1007/s002340050372
   Wanebo JE, 2003, J NEUROSURG, V98, P82, DOI 10.3171/jns.2003.98.1.0082
   Wang CC, 2001, SURG NEUROL, V56, P218, DOI 10.1016/S0090-3019(01)00590-0
   Wang EM, 2005, J NEUROSURG, V102, P225, DOI 10.3171/jns.2005.102.s_supplement.0225
   Weil RJ, 2003, J NEUROSURG, V98, P95, DOI 10.3171/jns.2003.98.1.0095
   Yasargil M G, 1976, Surg Neurol, P141
   YOUNG S, 1987, J NEUROL NEUROSUR PS, V50, P155, DOI 10.1136/jnnp.50.2.155
   Zbar B, 1996, HUM MUTAT, V8, P348, DOI 10.1002/(SICI)1098-1004(1996)8:4<348::AID-HUMU8>3.3.CO;2-9
   Zhou LF, 2005, SURG NEUROL, V63, P307, DOI 10.1016/j.surneu.2004.07.038
   Zimmermann M, 1997, ZBL NEUROCHIR, V58, P1
NR 51
TC 48
Z9 61
U1 0
U2 6
PU SPRINGER
PI NEW YORK
PA 233 SPRING ST, NEW YORK, NY 10013 USA
SN 0344-5607
EI 1437-2320
J9 NEUROSURG REV
JI Neurosurg. Rev.
PD JAN
PY 2009
VL 32
IS 1
BP 37
EP 47
DI 10.1007/s10143-008-0166-0
PG 11
WC Clinical Neurology; Surgery
WE Science Citation Index Expanded (SCI-EXPANDED)
SC Neurosciences & Neurology; Surgery
GA 371TH
UT WOS:000260853700010
PM 18810515
DA 2023-08-27
ER

PT J
AU Hopfenmueller, VL
   Perner, B
   Reuter, H
   Bates, TJD
   Grosse, A
   Englert, C
AF Hopfenmueller, Vera L.
   Perner, Birgit
   Reuter, Hanna
   Bates, Thomas J. D.
   Grosse, Andreas
   Englert, Christoph
TI The Wilms Tumor Gene wt1a Contributes to Blood-Cerebrospinal Fluid
   Barrier Function in Zebrafish
SO FRONTIERS IN CELL AND DEVELOPMENTAL BIOLOGY
LA English
DT Article
DE Wilms tumor protein; zebrafish; choroid plexus; CRISPR; cas9; dye
   accumulation assay; fluorescent tracer
ID CHOROID-PLEXUS; BRAIN VENTRICLES; SONIC HEDGEHOG; BETA-CATENIN;
   EXPRESSION; CELLS; IDENTIFICATION; ROLES; DNA
AB The Wilms tumor suppressor gene Wt1 encodes a zinc finger transcription factor, which is highly conserved among vertebrates. It is a key regulator of urogenital development and homeostasis but also plays a role in other organs including the spleen and the heart. More recently additional functions for Wt1 in the mammalian central nervous system have been described. In contrast to mammals, bony fish possess two paralogous Wt1 genes, namely wt1a and wt1b. By performing detailed in situ hybridization analyses during zebrafish development, we discovered new expression domains for wt1a in the dorsal hindbrain, the caudal medulla and the spinal cord. Marker analysis identified wt1a expressing cells of the dorsal hindbrain as ependymal cells of the choroid plexus in the myelencephalic ventricle. The choroid plexus acts as a blood-cerebrospinal fluid barrier and thus is crucial for brain homeostasis. By employing wt1a mutant larvae and a dye accumulation assay with fluorescent tracers we demonstrate that Wt1a is required for proper choroid plexus formation and function. Thus, Wt1a contributes to the barrier properties of the choroid plexus in zebrafish, revealing an unexpected role for Wt1 in the zebrafish brain.
C1 [Hopfenmueller, Vera L.; Perner, Birgit; Reuter, Hanna; Bates, Thomas J. D.; Grosse, Andreas; Englert, Christoph] Fritz Lipmann Inst FLI, Leibniz Inst Aging, Jena, Germany.
   [Englert, Christoph] Friedrich Schiller Univ Jena, Inst Biochem & Biophys, Jena, Germany.
   [Reuter, Hanna] Carl Zeiss Microscopy GmbH, Jena, Germany.
   [Bates, Thomas J. D.] Univ Reading, Reading, Berks, England.
   [Grosse, Andreas] Altona Diagnost GmbH, Hamburg, Germany.
C3 Leibniz Institut fur Alternsforschung - Fritz-Lipmann-Institut (FLI);
   Friedrich Schiller University of Jena; Carl Zeiss AG; University of
   Reading
RP Englert, C (通讯作者)，Fritz Lipmann Inst FLI, Leibniz Inst Aging, Jena, Germany.; Englert, C (通讯作者)，Friedrich Schiller Univ Jena, Inst Biochem & Biophys, Jena, Germany.
EM
OI Hopfenmuller, Vera Luise/0000-0002-9807-6403
FU Federal Government of Germany; State of Thuringia; Thuringian state
   program ProExzellenz [RegenerAging-FSU-I-03/14]
FX The FLI is a member of the Leibniz Association and is financially
   supported by the Federal Government of Germany and the State of
   Thuringia. Acquisition of the lightsheet microsope was supported by the
   Thuringian state program ProExzellenz (RegenerAging-FSU-I-03/14).
CR Alvira-Botero Ximena, 2010, Curr Aging Sci, V3, P219
   ARMSTRONG JF, 1993, MECH DEVELOP, V40, P85, DOI 10.1016/0925-4773(93)90090-K
   Awatramani R, 2003, NAT GENET, V35, P70, DOI 10.1038/ng1228
   Bill BR, 2008, PLOS ONE, V3, DOI 10.1371/journal.pone.0003114
   Bollig F, 2006, DEV DYNAM, V235, P554, DOI 10.1002/dvdy.20645
   Bollig F, 2009, DEVELOPMENT, V136, P2883, DOI 10.1242/dev.031773
   Chau YY, 2011, PLOS GENET, V7, DOI 10.1371/journal.pgen.1002404
   Christensen HL, 2013, FRONT PHYSIOL, V4, DOI 10.3389/fphys.2013.00304
   Currle DS, 2005, DEVELOPMENT, V132, P3549, DOI 10.1242/dev.01915
   D'Gama PP, 2021, CELL REP, V37, DOI 10.1016/j.celrep.2021.109775
   Dani N, 2021, CELL, V184, P3056, DOI 10.1016/j.cell.2021.04.003
   Del Bigio MR, 2010, ACTA NEUROPATHOL, V119, P55, DOI 10.1007/s00401-009-0624-y
   del Pozo A, 2020, ENEURO, V7, DOI 10.1523/ENEURO.0047-20.2020
   Dong LH, 2015, KIDNEY INT, V88, P684, DOI 10.1038/ki.2015.198
   Dong LH, 2015, J AM SOC NEPHROL, V26, P2118, DOI 10.1681/ASN.2014080819
   Falk A, 2002, J NEUROSCI RES, V69, P757, DOI 10.1002/jnr.10410
   Garcia-Lecea M, 2008, PLOS ONE, V3, DOI 10.1371/journal.pone.0003090
   Haque F, 2018, J NEUROSCI, V38, P5666, DOI 10.1523/JNEUROSCI.0328-18.2018
   HAUPTMANN G, 1994, TRENDS GENET, V10, P266, DOI 10.1016/0168-9525(90)90008-T
   Henson HE, 2014, FRONT NEUROSCI-SWITZ, V8, DOI 10.3389/fnins.2014.00364
   Herzer U, 1999, CURR BIOL, V9, P837, DOI 10.1016/S0960-9822(99)80369-8
   Huang X, 2009, DEVELOPMENT, V136, P2535, DOI 10.1242/dev.033795
   Imayoshi I, 2008, DEVELOPMENT, V135, P2531, DOI 10.1242/dev.021535
   Irvin DK, 2004, J NEUROSCI RES, V75, P330, DOI 10.1002/jnr.10843
   Kim MS, 2010, J BIOL CHEM, V285, P14585, DOI 10.1074/jbc.M109.094334
   KIMMEL CB, 1995, DEV DYNAM, V203, P253, DOI 10.1002/aja.1002030302
   KREIDBERG JA, 1993, CELL, V74, P679, DOI 10.1016/0092-8674(93)90515-R
   Liddelow SA, 2015, FRONT NEUROSCI-SWITZ, V9, DOI 10.3389/fnins.2015.00032
   Lowery LA, 2005, DEVELOPMENT, V132, P2057, DOI 10.1242/dev.01791
   Lun MP, 2015, NAT REV NEUROSCI, V16, P445, DOI 10.1038/nrn3921
   Mirzadeh Z, 2017, NAT COMMUN, V8, DOI 10.1038/ncomms13759
   Moore AW, 1999, DEVELOPMENT, V126, P1845
   MUNDLOS S, 1993, DEVELOPMENT, V119, P1329
   Nielsen CM, 2010, DEV BIOL, V340, P430, DOI 10.1016/j.ydbio.2010.01.032
   Ohtoshi Akihira, 2008, Cerebrospinal Fluid Res, V5, P16, DOI 10.1186/1743-8454-5-16
   Pavenstadt H, 2003, PHYSIOL REV, V83, P253, DOI 10.1152/physrev.00020.2002
   Perner B, 2007, DEV BIOL, V309, P87, DOI 10.1016/j.ydbio.2007.06.022
   Pfaffl MW, 2001, NUCLEIC ACIDS RES, V29, DOI 10.1093/nar/29.9.e45
   Sanz-Morejon A, 2019, CELL REP, V28, P1296, DOI 10.1016/j.celrep.2019.06.091
   Sathyanesan M, 2012, TRANSL PSYCHIAT, V2, DOI 10.1038/tp.2012.64
   Schnerwitzki D, 2020, FRONT NEUROSCI-SWITZ, V14, DOI 10.3389/fnins.2020.529487
   Schnerwitzki D, 2018, LIFE SCI ALLIANCE, V1, DOI 10.26508/lsa.201800106
   Schnerwitzki D, 2014, DEV BIOL, V393, P24, DOI 10.1016/j.ydbio.2014.06.026
   Shah PT, 2018, CELL, V173, P1045, DOI 10.1016/j.cell.2018.03.063
   SHARMA PM, 1992, CANCER RES, V52, P6407
   Spector R, 2015, EXP NEUROL, V267, P78, DOI 10.1016/j.expneurol.2015.02.032
   Taher MM, 2019, ONCOL LETT, V18, P5063, DOI 10.3892/ol.2019.10882
   Tong YA, 2015, CANCER CELL, V27, P712, DOI 10.1016/j.ccell.2015.04.005
   van Leeuwen LM, 2018, BIOL OPEN, V7, DOI 10.1242/bio.030494
   Vercellino M, 2008, J NEUROIMMUNOL, V199, P133, DOI 10.1016/j.jneuroim.2008.04.035
   von Gise A, 2011, DEV BIOL, V356, P421, DOI 10.1016/j.ydbio.2011.05.668
   Wang XN, 2013, PLOS GENET, V9, DOI 10.1371/journal.pgen.1003645
   Wang X, 2019, THERIOGENOLOGY, V138, P84, DOI 10.1016/j.theriogenology.2019.07.007
   Wolburg H, 2010, ACTA NEUROPATHOL, V119, P75, DOI 10.1007/s00401-009-0627-8
   Zhang Xuemei, 2018, Aquaculture and Fisheries, V3, P99, DOI 10.1016/j.aaf.2018.04.004
NR 55
TC 0
Z9 0
U1 1
U2 4
PU FRONTIERS MEDIA SA
PI LAUSANNE
PA AVENUE DU TRIBUNAL FEDERAL 34, LAUSANNE, CH-1015, SWITZERLAND
SN 2296-634X
J9 FRONT CELL DEV BIOL
JI Front. Cell. Dev. Biol.
PD JAN 11
PY 2022
VL 9
AR 809962
DI 10.3389/fcell.2021.809962
PG 13
WC Cell Biology; Developmental Biology
WE Science Citation Index Expanded (SCI-EXPANDED)
SC Cell Biology; Developmental Biology
GA YO6CP
UT WOS:000748027400001
PM 35087838
OA Green Submitted, gold, Green Published
DA 2023-08-27
ER

PT J
AU Dinh, CT
   Bracho, O
   Mei, C
   Bas, E
   Fernandez-Valle, C
   Telischi, F
   Liu, XZ
AF Dinh, Christine T.
   Bracho, Olena
   Mei, Christine
   Bas, Esperanza
   Fernandez-Valle, Cristina
   Telischi, Fred
   Liu, Xue-Zhong
TI A Xenograft Model of Vestibular Schwannoma and Hearing Loss
SO OTOLOGY & NEUROTOLOGY
LA English
DT Article
DE Acoustic neuroma; Auditory brainstem response; Bioluminescence; Hearing;
   Neurofibromatosis type II; NF2; Tumor; Vestibular schwannoma
ID NEUROFIBROMATOSIS TYPE-2; STEREOTACTIC RADIOSURGERY; FACIAL-NERVE;
   BEVACIZUMAB; MANAGEMENT; GROWTH; PREDICTORS; INHIBITOR; THERAPY
AB Hypothesis:Microsurgical implantation of mouse merlin-deficient Schwann cells (MD-SC) into the cerebellopontine angle of immunodeficient rats will initiate tumor formation, hearing loss, and vestibular dysfunction.Background:The progress in identifying effective drug therapies for treatment of Neurofibromatosis type II (NF2) is limited by the availability of animal models of VS that develop hearing loss and imbalance.Methods:A microsurgical technique for implanting MD-SCs onto the cochleovestibular nerve of rats was developed. Ten Rowett Nude rats were implanted with either approximate to 10(5) MD-SCs expressing luciferase (N=5) or vehicle (N=5). Rats received bioluminescence imaging, auditory brainstem response testing, and were observed for head tilt every 2 weeks after surgery, for a total of 6 weeks. Tumors were harvested and processed with hematoxylin & eosin staining and immunohistochemistry was performed for S100.Results:Rats implanted with MD-SCs developed significantly higher tumor bioluminescence measurements and hearing threshold shifts at multiple frequencies by the 4th and 6th weeks post-implantation, compared with control rats. Rats implanted with MD-SCs also developed gross tumor. The tumor volume was significantly greater than nerve volumes obtained from rats in the control group. All rats with tumors developed a head tilt, while control rats had no signs of vestibular dysfunction. Tumors demonstrated histological features of schwannoma and express S100.Conclusion:Using this microsurgical technique, this xenograft rat model of VS develops tumors involving the cochleovestibular nerve, shifts in hearing thresholds, and vestibular dysfunction. This animal model can be used to investigate tumor-mediated hearing loss and perform preclinical drug studies for NF2.
C1 [Dinh, Christine T.; Bracho, Olena; Mei, Christine; Bas, Esperanza; Telischi, Fred; Liu, Xue-Zhong] Univ Miami, Miller Sch Med, Dept Otolaryngol, 1120 NW 14th St,Suite 579, Miami, FL 33136 USA.
   [Fernandez-Valle, Cristina] Univ Cent Florida, Burnett Sch Biomed Sci, Coll Med, Orlando, FL 32816 USA.
C3 University of Miami; State University System of Florida; University of
   Central Florida
RP Dinh, CT (通讯作者)，Univ Miami, Miller Sch Med, Dept Otolaryngol, 1120 NW 14th St,Suite 579, Miami, FL 33136 USA.
EM
FU NIDCD [1R01DC010189-06]
FX The cell-line utilized in this study was created in part through funding
   by the NIDCD 1R01DC010189-06 research grant to Dr. Cristina
   Fernandez-Valle
CR Bas E, 2016, HEARING RES, V337, P12, DOI 10.1016/j.heares.2016.02.003
   Baser ME, 2002, AM J HUM GENET, V71, P715, DOI 10.1086/342716
   Blakeley J, 2012, CURR OPIN OTOLARYNGO, V20, P372, DOI 10.1097/MOO.0b013e328357d2ee
   Bonne NX, 2016, J NEURO-ONCOL, V129, P47, DOI 10.1007/s11060-016-2150-9
   Dilwali S, 2015, SCI REP-UK, V5, DOI 10.1038/srep18599
   Evans DG, 2010, AM J MED GENET A, V152A, P327, DOI 10.1002/ajmg.a.33139
   Evans DGR, 2009, ORPHANET J RARE DIS, V4, DOI 10.1186/1750-1172-4-16
   Evans DGR, 2005, OTOL NEUROTOL, V26, P93, DOI 10.1097/00129492-200501000-00016
   Fuse MA, 2017, MOL CANCER THER, V16, P2387, DOI 10.1158/1535-7163.MCT-17-0417
   Gehlhausen JR, 2015, HUM MOL GENET, V24, P1, DOI 10.1093/hmg/ddu414
   Giovannini M, 2014, NEURO-ONCOLOGY, V16, P493, DOI 10.1093/neuonc/not242
   JAASKELAINEN J, 1994, J NEUROSURG, V80, P541, DOI 10.3171/jns.1994.80.3.0541
   Jacob A, 2012, LARYNGOSCOPE, V122, P174, DOI 10.1002/lary.22392
   Jethanamest D, 2015, LARYNGOSCOPE, V125, P2163, DOI 10.1002/lary.25159
   Karajannis MA, 2012, NEURO-ONCOLOGY, V14, P1163, DOI 10.1093/neuonc/nos146
   Kim BS, 2016, NEUROSURG REV, V39, P643, DOI 10.1007/s10143-016-0728-5
   Licciulli S, 2013, J BIOL CHEM, V288, P29105, DOI 10.1074/jbc.M113.510933
   Mautner VF, 2010, ANN ONCOL, V21, P2294, DOI 10.1093/annonc/mdq566
   Meijer OWM, 2008, NEUROSURGERY, V62, pA37, DOI 10.1227/01.neu.0000325935.23852.9d
   Nowak A, 2015, NEUROL NEUROCHIR POL, V49, P295, DOI 10.1016/j.pjnns.2015.06.008
   Otsuka G, 2003, J NEUROSURG, V99, P480, DOI 10.3171/jns.2003.99.3.0480
   Petrilli AM, 2016, ONCOGENE, V35, P537, DOI 10.1038/onc.2015.125
   Petrilli AM, 2014, AM J TRANSL RES, V6, P471
   Plotkin SR, 2012, OTOL NEUROTOL, V33, P1046, DOI 10.1097/MAO.0b013e31825e73f5
   Samii M, 1997, NEUROSURGERY, V40, P696, DOI 10.1097/00006123-199704000-00007
   Slusarz KM, 2014, CANCER CHEMOTH PHARM, V73, P1197, DOI 10.1007/s00280-014-2456-2
   SOBEL RA, 1993, J NEUROPATH EXP NEUR, V52, P106, DOI 10.1097/00005072-199303000-00002
   Subach BR, 1999, J NEUROSURG, V90, P815, DOI 10.3171/jns.1999.90.5.0815
   Sughrue ME, 2011, J NEUROSURG, V114, P381, DOI 10.3171/2010.4.JNS091962
   Sun SB, 2014, J NEUROSURG, V121, P143, DOI 10.3171/2014.8.GKS141503
   van de Langenberg R, 2011, OTOL NEUROTOL, V32, P338, DOI 10.1097/MAO.0b013e3182040d9f
   Workman P, 2010, BRIT J CANCER, V102, P1555, DOI 10.1038/sj.bjc.6605642
NR 32
TC 9
Z9 10
U1 0
U2 7
PU LIPPINCOTT WILLIAMS & WILKINS
PI PHILADELPHIA
PA TWO COMMERCE SQ, 2001 MARKET ST, PHILADELPHIA, PA 19103 USA
SN 1531-7129
EI 1537-4505
J9 OTOL NEUROTOL
JI Otol. Neurotol.
PD JUN
PY 2018
VL 39
IS 5
BP E362
EP E369
DI 10.1097/MAO.0000000000001766
PG 8
WC Clinical Neurology; Otorhinolaryngology
WE Science Citation Index Expanded (SCI-EXPANDED)
SC Neurosciences & Neurology; Otorhinolaryngology
GA HK2WU
UT WOS:000457774400012
PM 29557843
OA Green Accepted
DA 2023-08-27
ER

PT J
AU Kulkarni, AV
   Piscione, J
   Shams, I
   Bouffet, E
AF Kulkarni, Abhaya V.
   Piscione, Janine
   Shams, Iffat
   Bouffet, Eric
TI Long-term quality of life in children treated for posterior fossa brain
   tumors
SO JOURNAL OF NEUROSURGERY-PEDIATRICS
LA English
DT Article
DE astrocytoma; brain tumor; ependymoma; oncology; medulloblastoma;
   pediatric; quality of life
ID HEALTH UTILITIES INDEX; FAMILY ASSESSMENT DEVICE; OCCIPITAL HORN RATIO;
   GENERIC CORE SCALES; COGNITIVE-DEVELOPMENT; VENTRICULAR SIZE;
   SPINA-BIFIDA; RELIABILITY; HYDROCEPHALUS; VALIDITY
AB Object. In the face of increasing survival, quality of life (QOL) has become an important indicator of treatment success in children with posterior fossa brain tumors (PFBTs). The authors' objective was to assess the long-term QOL in survivors of PEBT.
   Methods. The authors conducted a cross-sectional study of children who, between birth and age 18 years at diagnosis, had previously been treated at their institution for a PFBT. At the time of assessment for this study, children were between 5 and 19 years old and had received standard treatment for PFBT ending at least 6 months before the assessment. The QOL was measured with the Pediatric Quality of Life Inventory (PedsQL) generic score scales and the Health Utilities Index Mark 3 (HUI3). Multivariate analyses were used to assess several variables (patient related, treatment related, and socioeconomic) for association with QOL.
   Results. A total of 62 children participated in the study (median age at assessment 11.9 years, interquartile range [IQR] 7.8-14.8, and median age at tumor diagnosis of 4.9 years, IQR 2.5-6.9). Median time since active treatment for their PFBT was 5.2 years (IQR 24-10.1). Tumor types included cerebellar pilocytic astrocytoma (45.2%), medulloblastoma (30.6%), ependymoma (11.3%), and brainstem astrocytoma (11.3%). Adjuvant therapy included chemotherapy (40.3%) or radiotherapy (14.5% focal and 21.0% craniospinal radiotherapy). Permanent treatment for hydrocephalus was required in 38.7% of the patients. Tumors recurred in 11.3%, requiring repeat treatment in these patients. The median HUI3 utility score was 0.91 (IQR 0.71-1.00) and the median PedsQL total score was 78.3 (IQR 64.1-92.4). Only the following variables were significantly associated with decreased QOL in multivariable model testing (all p<0.05): need for permanent hydrocephalus treatment, large ventricle size, decreased family functioning, and lower family income.
   Conclusions. As a group, long-term survivors of pediatric PFBT appear to have QOL indicators that are similar to those of the general population, although a reasonable minority of patients experience poor outcomes. Although several confounding variables likely remain in this retrospective study, important associations with QOL include the presence of hydrocephalus and socioeconomic factors. The study sample size, however, was limited and the presence of other important factors cannot be excluded.
C1 [Kulkarni, Abhaya V.; Shams, Iffat] Univ Toronto, Hosp Sick Children, Div Neurosurg, Toronto, ON M5S 1A1, Canada.
   [Bouffet, Eric] Univ Toronto, Hosp Sick Children, Div Oncol, Toronto, ON M5S 1A1, Canada.
   [Piscione, Janine] Univ Toronto, Hosp Sick Children, Div Rehabil, Toronto, ON M5S 1A1, Canada.
C3 University of Toronto; Hospital for Sick Children (SickKids); University
   of Toronto; Hospital for Sick Children (SickKids); University of
   Toronto; Hospital for Sick Children (SickKids)
RP Kulkarni, AV (通讯作者)，Hosp Sick Children, Div Neurosurg, 555 Univ Ave,Room 1503, Toronto, ON M5G 1X8, Canada.
EM
RI Bouffet, Eric/AAB-8999-2020
FU b.r.a.i.n.child
FX This studied was funded by a grant from b.r.a.i.n.child.
CR Aarsen FK, 2004, NEUROLOGY, V62, P1311, DOI 10.1212/01.WNL.0000120549.77188.36
   Anderson VA, 2000, BRIT J CANCER, V82, P255
   BOYLE MH, 1995, QUAL LIFE RES, V4, P249, DOI 10.1007/BF02260864
   BYLES J, 1988, FAM PROCESS, V27, P97, DOI 10.1111/j.1545-5300.1988.00097.x
   Chen E, 2006, AM J PUBLIC HEALTH, V96, P702, DOI 10.2105/AJPH.2004.048124
   ELLENBERG L, 1987, NEUROSURGERY, V21, P638, DOI 10.1227/00006123-198711000-00006
   Feeny D, 2004, SOC SCI MED, V58, P799, DOI 10.1016/S0277-9536(03)00254-5
   FLETCHER JM, 1992, ARCH NEUROL-CHICAGO, V49, P818, DOI 10.1001/archneur.1992.00530320042010
   Fletcher JM, 1996, CHILD NERV SYST, V12, P192, DOI 10.1007/BF00301250
   Fletcher JM, 1996, ARCH NEUROL-CHICAGO, V53, P549, DOI 10.1001/archneur.1996.00550060093022
   Fuemmeler BF, 2002, CLIN PSYCHOL REV, V22, P547, DOI 10.1016/S0272-7358(01)00120-9
   Furlong WJ, 2001, ANN MED, V33, P375, DOI 10.3109/07853890109002092
   Gemke RJBJ, 1996, J CLIN EPIDEMIOL, V49, P327, DOI 10.1016/0895-4356(95)00528-5
   Grill J, 1999, INT J RADIAT ONCOL, V45, P137, DOI 10.1016/S0360-3016(99)00177-7
   Grill J, 2004, PEDIATR BLOOD CANCER, V42, P452, DOI 10.1002/pbc.10469
   Hetherington R, 2006, CHILD NERV SYST, V22, P117, DOI 10.1007/s00381-005-1231-4
   Hungerford A, 2006, EVALUATION REV, V30, P631, DOI 10.1177/0193841X06291532
   Hunt GM, 1999, J NEUROL NEUROSUR PS, V67, P591, DOI 10.1136/jnnp.67.5.591
   Jenkin D, 1998, MED PEDIATR ONCOL, V31, P506, DOI 10.1002/(SICI)1096-911X(199812)31:6<506::AID-MPO7>3.3.CO;2-O
   Kennedy CR, 1999, INT J CANCER, P106
   Kulkarni AV, 2008, J PEDIATR-US, V153, P689, DOI 10.1016/j.jpeds.2008.04.068
   Kulkarni AV, 2007, J NEUROSURG, V107, P358, DOI 10.3171/PED-07/11/358
   Kulkarni AV, 1999, PEDIATR NEUROSURG, V31, P65, DOI 10.1159/000028836
   KUN LE, 1983, J NEUROSURG, V58, P1, DOI 10.3171/jns.1983.58.1.0001
   MILLER IW, 1985, J MARITAL FAM THER, V11, P345, DOI 10.1111/j.1752-0606.1985.tb00028.x
   Mulhern RK, 1999, INT J CANCER, P91
   O'Hayon BB, 1998, PEDIATR NEUROSURG, V29, P245, DOI 10.1159/000028730
   PACKER RJ, 1989, J NEUROSURG, V70, P707, DOI 10.3171/jns.1989.70.5.0707
   PACKER RJ, 1987, PEDIATR NEUROSCI, V13, P169, DOI 10.1159/000120325
   Pollack IF, 2011, J NEUROSURG-PEDIATR, V8, P135, DOI 10.3171/2011.5.PEDS1178
   Poretti A, 2004, DEV MED CHILD NEUROL, V46, P220, DOI 10.1017/S0012162204000374
   Riva-Cambrin J, 2009, J NEUROSURG-PEDIATR, V3, P378, DOI 10.3171/2009.1.PEDS08298
   Saigal S, 1996, JAMA-J AM MED ASSOC, V276, P453, DOI 10.1001/jama.276.6.453
   Trudel JG, 1998, QUAL LIFE RES, V7, P421, DOI 10.1023/A:1008857920624
   Varni JW, 2006, DEV MED CHILD NEUROL, V48, P442, DOI 10.1017/S001216220600096X
   Varni JW, 2006, QUAL LIFE RES, V15, P203, DOI 10.1007/s11136-005-1388-z
   Varni JW, 1999, MED CARE, V37, P126, DOI 10.1097/00005650-199902000-00003
   Varni JW, 2002, CANCER-AM CANCER SOC, V94, P2090, DOI 10.1002/cncr.10428
NR 38
TC 40
Z9 41
U1 0
U2 6
PU AMER ASSOC NEUROLOGICAL SURGEONS
PI ROLLING MEADOWS
PA 5550 MEADOWBROOK DRIVE, ROLLING MEADOWS, IL 60008 USA
SN 1933-0707
EI 1933-0715
J9 J NEUROSURG-PEDIATR
JI J. Neurosurg.-Pediatr.
PD SEP
PY 2013
VL 12
IS 3
BP 235
EP 240
DI 10.3171/2013.6.PEDS12535
PG 6
WC Clinical Neurology; Pediatrics; Surgery
WE Science Citation Index Expanded (SCI-EXPANDED); Social Science Citation Index (SSCI)
SC Neurosciences & Neurology; Pediatrics; Surgery
GA 204IK
UT WOS:000323358800006
PM 23829377
DA 2023-08-27
ER

PT J
AU Hug, EB
   Loredo, LN
   Slater, JD
   DeVries, A
   Grove, RI
   Schaefer, RA
   Rosenberg, AE
   Slater, JM
AF Hug, EB
   Loredo, LN
   Slater, JD
   DeVries, A
   Grove, RI
   Schaefer, RA
   Rosenberg, AE
   Slater, JM
TI Proton radiation therapy for chordomas and chondrosarcomas of the skull
   base
SO JOURNAL OF NEUROSURGERY
LA English
DT Article
DE chordoma; chondrosarcoma; skull base tumor; charged-particle therapy;
   radiation therapy; proton
ID CERVICAL-SPINE; FOLLOW-UP; IRRADIATION; PHOTON; TUMORS
AB Object. Local tumor control, patient survival, and treatment failure outcomes were analyzed to assess treatment efficacy in 58 patients in whom fractionated proton radiation therapy (RT) was administered for skull base chordomas and chondrosarcomas.
   Methods. Between March 1992 and January 1998, a total of 58 patients who could be evaluated were treated for skull base tumors, 33 for chordoma and 25 for chondrosarcoma. Following various surgical procedures, residual tumor was detected in 91% of patients; 59% demonstrated brainstem involvement. Target dosages ranged from 64.8 and 79.2 (mean 70.7) Co Gy equivalent. The range of follow up was 7 to 75 months (mean 33 months).
   In 10 patients (17%) the treatment failed locally, resulting in local control rates of 92% (23 of 25 patients) for chondrosarcomas and 76% (25 of 33 patients) for chordomas. Tumor volume and brainstem involvement influenced control rates. All tumors with volumes of 25 mi or less remained locally controlled, compared with 56% of tumors larger than 25 mi (p = 0.02); 94% of patients without brainstem involvement did not experience recurrence; in patients with brainstem involvement land dose reduction because of brainstem tolerance constraints) the authors achieved a tumor control rate of 53% (p = 0.04). Three patients died of their disease, and one died of intercurrent disease. Actuarial 5-year survival rates were 100% for patients with chondrosarcoma and 79% for patients with chordoma. Grade 3 and 4 late toxicities were observed in four patients (7%) and were symptomatic in three (5%).
   Conclusions. High-dose proton RT offers excellent chances of lasting tumor control and survival, with acceptable risks. In this series all small- and medium-sized tumors with no demonstrable brainstem involvement have been controlled; all such patients are alive. Surgical debulking enhanced delivery of full tumoricidal doses, but even patients with large tumors and disease abutting crucial normal structures benefited.
C1 Loma Linda Univ, Med Ctr, Dept Radiat Med, Loma Linda, CA 92354 USA.
   Massachusetts Gen Hosp, Dept Pathol, Boston, MA 02114 USA.
   Univ Innsbruck, Dept Radiat Therapy, A-6020 Innsbruck, Austria.
C3 Loma Linda University; Harvard University; Massachusetts General
   Hospital; University of Innsbruck
RP Hug, EB (通讯作者)，Loma Linda Univ, Med Ctr, Dept Radiat Med, 11234 Anderson St, Loma Linda, CA 92354 USA.
EM
CR AMENDOLA BE, 1986, RADIOLOGY, V158, P839, DOI 10.1148/radiology.158.3.3945761
   [Anonymous], 1995, INT J RAD ONCOL BIOL, V31, P1049
   AUSTIN JP, 1993, INT J RADIAT ONCOL, V25, P439, DOI 10.1016/0360-3016(93)90065-4
   AUSTINSEYMOUR M, 1989, J NEUROSURG, V70, P13, DOI 10.3171/jns.1989.70.1.0013
   BERSON AM, 1988, INT J RADIAT ONCOL, V15, P559, DOI 10.1016/0360-3016(88)90295-7
   Brown E, 1994, Neuroimaging Clin N Am, V4, P529
   Catton C, 1996, RADIOTHER ONCOL, V41, P67, DOI 10.1016/S0167-8140(96)01805-1
   COX DR, 1972, J R STAT SOC B, V34, P187
   CUMMINGS BJ, 1983, INT J RADIAT ONCOL, V9, P633, DOI 10.1016/0360-3016(83)90228-6
   Dahlin DC, 1986, BONE TUMORS, P119
   Debus J, 1997, INT J RADIAT ONCOL, V39, P967, DOI 10.1016/S0360-3016(97)00364-7
   FAGUNDES MA, 1995, INT J RADIAT ONCOL, V33, P579, DOI 10.1016/0360-3016(95)02014-3
   FORSYTH PA, 1993, J NEUROSURG, V78, P741, DOI 10.3171/jns.1993.78.5.0741
   FULLER DB, 1988, INT J RADIAT ONCOL, V15, P331, DOI 10.1016/S0360-3016(98)90012-8
   GAY E, 1995, NEUROSURGERY, V36, P887, DOI 10.1227/00006123-199505000-00001
   GOITEIN M, 1983, INT J RADIAT ONCOL, V9, P789, DOI 10.1016/0360-3016(83)90003-2
   HEFFELFINGER MJ, 1973, CANCER-AM CANCER SOC, V32, P410, DOI 10.1002/1097-0142(197308)32:2<410::AID-CNCR2820320219>3.0.CO;2-S
   Hess C. F., 1993, Strahlentherapie und Onkologie, V169, P660
   HUG EB, 1995, INT J RADIAT ONCOL, V31, P467, DOI 10.1016/0360-3016(94)00390-7
   ICRU (International Commission on Radiation Units and Measurements), 1993, J ICRU
   KAPLAN EL, 1958, J AM STAT ASSOC, V53, P457, DOI 10.2307/2281868
   MUNZENRIDER JE, 1985, STRAHLENTHER ONKOL, V161, P756
   MUNZENRIDER JE, 1993, HEAD NECK CANCER, V3, P649
   Muthukumar N, 1998, INT J RADIAT ONCOL, V41, P387, DOI 10.1016/S0360-3016(98)00051-0
   Pearlman A W, 1970, Am J Roentgenol Radium Ther Nucl Med, V108, P332
   RICH TA, 1985, CANCER, V56, P182, DOI 10.1002/1097-0142(19850701)56:1<182::AID-CNCR2820560131>3.0.CO;2-J
   Santoni R, 1998, INT J RADIAT ONCOL, V41, P59, DOI 10.1016/S0360-3016(98)00031-5
   SAXTON JP, 1981, INT J RADIAT ONCOL, V7, P913, DOI 10.1016/0360-3016(81)90008-0
   SLATER JM, 1992, INT J RADIAT ONCOL, V22, P383, DOI 10.1016/0360-3016(92)90058-P
   SUIT HD, 1982, J NEUROSURG, V56, P377, DOI 10.3171/jns.1982.56.3.0377
   URANO M, 1984, INT J RADIAT ONCOL, V10, P509, DOI 10.1016/0360-3016(84)90031-2
   Weber A L, 1994, Neuroimaging Clin N Am, V4, P515
NR 32
TC 344
Z9 354
U1 0
U2 6
PU AMER ASSOC NEUROLOGICAL SURGEONS
PI ROLLING MEADOWS
PA 5550 MEADOWBROOK DRIVE, ROLLING MEADOWS, IL 60008 USA
SN 0022-3085
J9 J NEUROSURG
JI J. Neurosurg.
PD SEP
PY 1999
VL 91
IS 3
BP 432
EP 439
DI 10.3171/jns.1999.91.3.0432
PG 8
WC Clinical Neurology; Surgery
WE Science Citation Index Expanded (SCI-EXPANDED)
SC Neurosciences & Neurology; Surgery
GA 232WN
UT WOS:000082393100012
PM 10470818
DA 2023-08-27
ER

PT J
AU Haapaniemi, JJ
   Laurikainen, ET
   Johansson, R
   Rinne, T
   Varpula, M
AF Haapaniemi, JJ
   Laurikainen, ET
   Johansson, R
   Rinne, T
   Varpula, M
TI Audiovestibular findings and location of an acoustic neuroma
SO EUROPEAN ARCHIVES OF OTO-RHINO-LARYNGOLOGY
LA English
DT Article
DE acoustic neuroma; auditory brainstem response electronystagmography;
   topodiagnosis
ID BRAIN-STEM RESPONSE; CEREBELLOPONTINE-ANGLE TUMORS; DIAGNOSIS; HEARING;
   MRI
AB Forty-one patients with unilateral acoustic neuroma (AN) were diagnosed by magnetic resonance imaging (MRI) between 1992 and 1997. All cases were analyzed with respect to tumor location and the results of audiometric examinations, auditory brainstem response (ABR) testing, and electronystagmography (ENG), Tumor location was determined by MRI and cases were divided into intracanalicular and extracanalicular sites. Intracanalicular tumors were significantly smaller than the extracanalicular ones The pure-tone hearing thresholds were better in ears with intracanalicular lesions than in those with extracanalicular ones. Respectively, speech reception thresholds were 33 dB and 45 dB, and speech discrimination scores 79% and 65%. ABR was abnormal in 98% of ANs, but was insufficient for determining tumor location. The ENG pursuit test was more frequently nor mal and the caloric side difference was smaller in ears with intracanalicular than extracanalicular AN. These findings show that the results of pure-tone and speech audiometry and ENG are better in ears with intracanalicular AN, while ABR results are similar regardless of tumor location.
C1 Univ Turku, Cent Hosp, Dept Radiol, FIN-20520 Turku, Finland.
   Univ Turku, Cent Hosp, Dept Otolaryngol, FIN-20520 Turku, Finland.
C3 University of Turku; University of Turku
RP Haapaniemi, JJ (通讯作者)，Ilveksenkatu 10, Piispanristi, Finland.
CR BAGULEY DM, 1989, BRIT J AUDIOL, V23, P158
   BERGENIUS J, 1988, ACTA OTO-LARYNGOL, V106, P361, DOI 10.3109/00016488809122258
   Berrettini S, 1997, J OTOLARYNGOL, V26, P97
   Burkey JM, 1996, LARYNGOSCOPE, V106, P839, DOI 10.1097/00005537-199607000-00010
   CHANDRASEKHAR SS, 1995, AM J OTOL, V16, P63
   CLEMIS JD, 1986, ARCH OTOLARYNGOL, V112, P190
   DORNHOFFER JL, 1994, OTOLARYNG HEAD NECK, V111, P232, DOI 10.1016/S0194-5998(94)70596-8
   GORDON ML, 1995, AM J OTOL, V16, P136
   GUYOT JP, 1992, ORL J OTO-RHINO-LARY, V54, P139, DOI 10.1159/000276283
   HENDRIX RA, 1990, OTOLARYNGOL HEAD NEC, V103, P364
   JOSEY AF, 1988, AM J OTOL, V9, P12
   LEVINE SC, 1991, AM J OTOL, V12, P341
   MOFFAT DA, 1993, J LARYNGOL OTOL, V107, P290, DOI 10.1017/S0022215100122856
   MOFFAT DA, 1989, J LARYNGOL OTOL, V103, P51, DOI 10.1017/S0022215100108023
   MUSIEK FE, 1986, OTOLARYNG HEAD NECK, V94, P568
   Nadol JB, 1996, AM J OTOL, V17, P312
   Naessens B., 1996, Acta Oto-Rhino-Laryngologica Belgica, V50, P99
   PFALTZ CR, 1991, ORL J OTO-RHINO-LARY, V53, P121, DOI 10.1159/000276202
   Schuknecht HF, 1974, PATHOLOGY EAR
   SELESNICK SH, 1992, OTOLARYNG CLIN N AM, V25, P521
   SELESNICK SH, 1993, LARYNGOSCOPE, V103, P431, DOI 10.1002/lary.5541030412
   Shelton C, 1996, OTOLARYNG HEAD NECK, V114, P71, DOI 10.1016/S0194-5998(96)70286-3
   TELIAN SA, 1989, LARYNGOSCOPE, V99, P10
   WELLING DB, 1990, OTOLARYNG HEAD NECK, V103, P364, DOI 10.1177/019459989010300305
   WIESS MH, 1990, OTOLARYNGOL HEAD NEC, V103, P583
   WILSON DF, 1992, LARYNGOSCOPE, V102, P961
NR 26
TC 6
Z9 7
U1 0
U2 0
PU SPRINGER VERLAG
PI NEW YORK
PA 175 FIFTH AVE, NEW YORK, NY 10010 USA
SN 0937-4477
J9 EUR ARCH OTO-RHINO-L
JI Eur. Arch. Oto-Rhino-Laryn.
PD MAY
PY 2000
VL 257
IS 5
BP 237
EP 241
DI 10.1007/s004050050230
PG 5
WC Otorhinolaryngology
WE Science Citation Index Expanded (SCI-EXPANDED)
SC Otorhinolaryngology
GA 329MA
UT WOS:000087909700001
PM 10923934
DA 2023-08-27
ER

PT J
AU Melnyk, T
   Masia, E
   Zagorodko, O
   Conejos-Sanchez, I
   Vicent, MJ
AF Melnyk, Tetiana
   Masia, Esther
   Zagorodko, Oleksandr
   Conejos-Sanchez, Inmaculada
   Vicent, Maria J.
TI Rational design of poly-L-glutamic acid-palbociclib conjugates for
   pediatric glioma treatment
SO JOURNAL OF CONTROLLED RELEASE
LA English
DT Article
DE Poly-L-glutamic acid; Polypeptide-drug conjugates; Palbociclib; Solution
   conformation; Pediatric gliomas; Glioblastoma; Diffuse intrinsic pontine
   glioma
ID GLIOBLASTOMA; RESISTANCE; MECHANISMS
AB Brain tumors represent the second most common cause of pediatric cancer death, with malignant gliomas accounting for similar to 75% of pediatric deaths. Palbociclib, a selective cyclin-dependent kinase 4/6 (CDK4/6) inhibitor, has shown promise in phase I clinical trials of pediatric patients with progressive/refractory brain tumors using the oral administration route; however, pharmacokinetic limitations and toxicity issues remain. We synthesized a family of well-defined linear and star-shaped polyglutamate (PGA)-palbociclib conjugates using redox-sensitive self-immolative linkers to overcome limitations associated with free palbociclib. Exhaustive characterization of this conjugate family provided evidence for a transition towards the formation of more organized conformational structures upon increased drug loading. We evaluated the activity of conjugates in patient-derived glioblastoma and diffuse intrinsic pontine glioma cells, which display differing reducing environments due to differential glutathione expression levels. We discovered that microenvironmental parameters and the identified conformational changes determined palbociclib release kinetics and therapeutic output; furthermore, we identified a star-shaped PGA-palbociclib conjugate with low drug loading as an optimal therapeutic approach in diffuse intrinsic pontine glioma cells.
C1 [Melnyk, Tetiana; Masia, Esther; Zagorodko, Oleksandr; Conejos-Sanchez, Inmaculada; Vicent, Maria J.] Polymer Therapeut Lab, Ave Eduardo Primo Yufera 3, Valencia 46012, Spain.
   [Masia, Esther; Vicent, Maria J.] Prince Felipe Res Ctr, Screening Platform, Ave Eduardo Primo Yufera 3, Valencia 46012, Spain.
C3 Prince Felipe Research Center
RP Conejos-Sanchez, I; Vicent, MJ (通讯作者)，Polymer Therapeut Lab, Ave Eduardo Primo Yufera 3, Valencia 46012, Spain.
EM
RI ; Vicent, Maria J./A-9690-2014
OI Conejos-Sanchez, Inmaculada/0000-0002-4196-0855; Masia Sanchis,
   Esther/0000-0003-2293-0359; Vicent, Maria J./0000-0001-7771-3373
FU Asociacion Espanola contra el Cancer (AECC) [TM PRDVA 172014MELN, ICS
   INVES 211323CONE]; Spanish Ministry of Economy [PID2019-108806RB-I00];
   European Regional Development Fund (PO FEDER Comunitat Valenciana
   2014-2020) Generalitat Valenciana; La Xocolata funding
FX The authors acknowledge AM. Carcaboso at Hospital Sant Joan de Deu,
   Barcelona (Spain) for cell donation and fruitful discussions, E. Solano
   and M. Malfois from BL-11 NCDSWEET beamline at ALBA Synchrotron for
   their support with SAXS experiments, Stuart P. Atkinson for manuscript
   preparation, V. Moreno-Manzano from the Neuronal and Tissue Regeneration
   Laboratory at CIPF for providing the primary neurons, A. Hernandez from
   the Confocal Microscopy service at CIPF, and Alicia Martinez from the
   Cytomics Service at CIPF. We thank the Asociacion Espanola contra el
   Cancer (AECC, TM PRDVA 172014MELN Ph.D. grant; ICS INVES 211323CONE
   Junior AECC grant), Spanish Ministry of Economy (PID2019-108806RB-I00),
   European Regional Development Fund (PO FEDER Comunitat Valenciana
   2014-2020) Generalitat Valenciana, and La Xocolata funding for financial
   support.
CR Alvarez-Fernandez M, 2020, CANCER CELL, V37, P514, DOI 10.1016/j.ccell.2020.03.010
   Arroyo-crespo J.J., 2018, ADV FUNCT MATER, V22, P1
   Barz M, 2013, POLYM CHEM-UK, V4, P2989, DOI 10.1039/c3py00189j
   Blau R, 2018, THERANOSTICS, V8, P3437, DOI 10.7150/thno.23853
   Chipman SD, 2006, INT J NANOMED, V1, P375, DOI 10.2147/nano.2006.1.4.375
   Clark AS, 2016, JAMA ONCOL, V2, P253, DOI 10.1001/jamaoncol.2015.4701
   Duro-Castano A, 2021, SCI ADV, V7, DOI 10.1126/sciadv.abf9180
   Ekladious I, 2019, NAT REV DRUG DISCOV, V18, P273, DOI 10.1038/s41573-018-0005-0
   Hauser P, 2021, CHILDREN-BASEL, V8, DOI 10.3390/children8090739
   Jansen MHA, 2016, MOL CANCER THER, V15, P2166, DOI 10.1158/1535-7163.MCT-15-0558
   Kavand A, 2020, J CONTROL RELEASE, V321, P285, DOI 10.1016/j.jconrel.2020.02.019
   Kelly SM, 2005, BBA-PROTEINS PROTEOM, V1751, P119, DOI 10.1016/j.bbapap.2005.06.005
   Kim Y, 2012, BIOMACROMOLECULES, V13, P814, DOI 10.1021/bm201730p
   Klein ME, 2018, CANCER CELL, V34, P9, DOI 10.1016/j.ccell.2018.03.023
   Knudsen ES, 2017, TRENDS CANCER, V3, P39, DOI 10.1016/j.trecan.2016.11.006
   Louis DN, 2016, ACTA NEUROPATHOL, V131, P803, DOI 10.1007/s00401-016-1545-1
   Cuesta CM, 2021, NANOMED-NANOTECHNOL, V34, DOI 10.1016/j.nano.2021.102376
   Manzanares D, 2020, PHARMACEUTICS, V12, DOI 10.3390/pharmaceutics12040371
   Melnyk T, 2020, ADV DRUG DELIVER REV, V160, P136, DOI 10.1016/j.addr.2020.10.007
   Michaud K, 2010, CANCER RES, V70, P3228, DOI 10.1158/0008-5472.CAN-09-4559
   Nguyen Long V, 2019, Oncotarget, V10, P6317, DOI 10.18632/oncotarget.27238
   Omuro A, 2013, JAMA-J AM MED ASSOC, V310, P1842, DOI 10.1001/jama.2013.280319
   Sangar MLC, 2017, CLIN CANCER RES, V23, P5802, DOI 10.1158/1078-0432.CCR-16-2943
   Schettini F, 2018, FRONT ONCOL, V8, DOI 10.3389/fonc.2018.00608
   Such GK, 2015, ADV MATER, V27, P2278, DOI 10.1002/adma.201405084
   Sun Y, 2019, EBIOMEDICINE, V43, P171, DOI 10.1016/j.ebiom.2019.04.043
   Tarassoli SP, 2017, NANOTECHNOLOGY, V28, DOI 10.1088/1361-6528/28/5/055101
   Taylor JW, 2018, J NEURO-ONCOL, V140, P477, DOI 10.1007/s11060-018-2977-3
   Tsuchikama K, 2018, PROTEIN CELL, V9, P33, DOI 10.1007/s13238-016-0323-0
   Van Mater D., 2021, PEDIATR BLOOD CANCER, V68, P1
   Vinci M, 2018, NAT MED, V24, P1204, DOI 10.1038/s41591-018-0086-7
   Ward E, 2014, CA-CANCER J CLIN, V64, P83, DOI 10.3322/caac.21219
   Warren KE, 2012, FRONT ONCOL, V2, DOI 10.3389/fonc.2012.00205
   Yang HY, 2015, NAT PROTOC, V10, DOI 10.1038/nprot.2015.024
   Zagorodko O, 2017, MACROMOL BIOSCI, V17, DOI 10.1002/mabi.201600316
NR 35
TC 3
Z9 3
U1 2
U2 2
PU ELSEVIER
PI AMSTERDAM
PA RADARWEG 29, 1043 NX AMSTERDAM, NETHERLANDS
SN 0168-3659
EI 1873-4995
J9 J CONTROL RELEASE
JI J. Control. Release
PD MAR
PY 2023
VL 355
BP 385
EP 394
DI 10.1016/j.jconrel.2023.01.079
EA FEB 2023
PG 10
WC Chemistry, Multidisciplinary; Pharmacology & Pharmacy
WE Science Citation Index Expanded (SCI-EXPANDED)
SC Chemistry; Pharmacology & Pharmacy
GA 9P9XB
UT WOS:000944627900001
PM 36746338
DA 2023-08-27
ER

PT J
AU Reuland, P
   Overkamp, D
   Aicher, KP
   Bien, S
   MullerSchauenburg, W
   Feine, U
AF Reuland, P
   Overkamp, D
   Aicher, KP
   Bien, S
   MullerSchauenburg, W
   Feine, U
TI Catecholamine secreting glomus tumor detected by iodine-123-MIBG
   scintigraphy
SO JOURNAL OF NUCLEAR MEDICINE
LA English
DT Article
DE iodine-123-MIGB; endocrine active glomus tumor; pheochromocytoma
ID CONCISE COMMUNICATION; ADRENAL-MEDULLA; NEURO-BLASTOMA;
   PHEOCHROMOCYTOMA; PARAGANGLIOMAS; LOCALIZATION; MANAGEMENT; DIAGNOSIS;
   SURGERY
AB We report the case of a 41-yr-old woman who presented with arterial hypertension and tinnitus in the right ear synchronous with pulse. She had previously undergone surgery for suspected pheochromocytoma without positive therapeutic effect, CT and MRI revealed a homogenous tumor with contrast enhancement in the right hypotympanon and foramen jugulare, and [I-123]metaiodobenzylguanidine (MIBG) scintigraphy demonstrated strong tracer uptake in the same area. Selective venous sampling of catecholamines in the ipsilateral jugular vein confirmed the tumor to have originated from hormone production.
C1 UNIV TUBINGEN,INST PATHOL,DEPT NUCL MED,W-7400 TUBINGEN,GERMANY.
   UNIV TUBINGEN,INST PATHOL,DEPT RADIOL,W-7400 TUBINGEN,GERMANY.
   UNIV TUBINGEN,MED CLIN,TUBINGEN,GERMANY.
   INST DIAGNOST NUCL MED,FREIBURG,GERMANY.
C3 CIVIS; Eberhard Karls University of Tubingen; Eberhard Karls University
   Hospital; CIVIS; Eberhard Karls University of Tubingen; Eberhard Karls
   University Hospital; CIVIS; Eberhard Karls University of Tubingen
CR BOGDASARIAN RS, 1979, OTOLARYNG HEAD NECK, V87, P648, DOI 10.1177/019459987908700520
   BOULOUX PMG, 1992, CLIN ENDOCRINOL, P459
   BRAVO EL, 1984, NEW ENGL J MED, V311, P1298, DOI 10.1056/NEJM198411153112007
   CONE TE, 1958, PEDIATRICS, V21, P994
   DEANGELIS LM, 1987, NEUROLOGY, V37, P129, DOI 10.1212/WNL.37.1.129
   DUKE WW, 1964, ANN INTERN MED, V60, P1040, DOI 10.7326/0003-4819-60-6-1040
   Fisch U., 1977, NEUROLOGICAL SURGERY, V2, P34
   FISCHER M, 1985, CARDIOLOGY, V72, P143, DOI 10.1159/000173961
   HOEFNAGEL CA, 1987, J MED IMAG, V1, P57
   JACKSON CG, 1990, AM J SURG, V159, P389, DOI 10.1016/S0002-9610(05)81279-6
   LANGER M, 1986, HNO, V34, P61
   LECOMPTE PM, 1949, AM J PATHOL, V24, P305
   LEVIT SA, 1969, NEW ENGL J MED, V281, P805, DOI 10.1056/NEJM196910092811502
   LYNN MD, 1984, J NUCL MED, V25, P436
   MANN W, 1984, HNO, V32, P249
   PARKIN JL, 1981, ANN OTO RHINOL LARYN, V90, P60, DOI 10.1177/000348948109000115
   SCHWABER MK, 1984, LARYNGOSCOPE, V94, P1008
   SHAPIRO B, 1985, J NUCL MED, V26, P576
   SHAPIRO B, 1982, P 3 WORLD C NUCL MED, P1265
   Sisson J C, 1986, Am J Physiol Imaging, V1, P96
   SWANSON DP, P 3 INT RAD DOS S, V198, P213
   SZEKELY T, 1984, HNO, V32, P54
   TISCHLER AS, 1977, NEW ENGL J MED, V296, P919
   VONMOLL L, 1987, J NUCL MED, V28, P979
   WEISSMAN AF, 1994, CLIN NUCL MED, V19, P527, DOI 10.1097/00003072-199406000-00013
   WIELAND DM, 1981, J NUCL MED, V22, P358
   Zak F. G, 1982, PARAGANGLIONIC CHEMO
NR 27
TC 7
Z9 7
U1 0
U2 0
PU SOC NUCLEAR MEDICINE INC
PI RESTON
PA 1850 SAMUEL MORSE DR, RESTON, VA 22090-5316
SN 0161-5505
J9 J NUCL MED
JI J. Nucl. Med.
PD MAR
PY 1996
VL 37
IS 3
BP 463
EP 465
PG 3
WC Radiology, Nuclear Medicine & Medical Imaging
WE Science Citation Index Expanded (SCI-EXPANDED)
SC Radiology, Nuclear Medicine & Medical Imaging
GA TZ413
UT WOS:A1996TZ41300019
PM 8772645
DA 2023-08-27
ER

PT J
AU Moller, P
   Bruderlein, S
   Strater, J
   Leithauser, F
   Hasel, C
   Bataille, F
   Moldenhauer, G
   Pawlita, M
   Barth, TFE
AF Moller, P
   Bruderlein, S
   Strater, J
   Leithauser, F
   Hasel, C
   Bataille, F
   Moldenhauer, G
   Pawlita, M
   Barth, TFE
TI MedB-1, a human tumor cell line derived from a primary mediastinal large
   B-cell lymphoma
SO INTERNATIONAL JOURNAL OF CANCER
LA English
DT Article
DE human B-cell lymphoma; primary mediastinal B-cell lymphoma; human B-cell
   lymphoma cell line; immunophenotype; thymic medulla
ID HIGH-GRADE MALIGNANCY; GENE REARRANGEMENTS; EXPRESSION; SCLEROSIS;
   ANTIGENS; AMPLIFICATION; LYMPHOCYTES; IMBALANCES; THYMUS; CLONES
AB Primary mediastinal B-cell lymphoma is a locally highly aggressive but poorly disseminating tumor composed of medium sized or large cells most probably of thymic medullary origin. it has a mature B-cell phenotype, typically lacks immunoglobulin expression and has variable defects in expression of HLA-molecules. We present here a cell line, MedB-1, derived from such a tumor. As is frequently found in mediastinal B-cell lymphomas in situ, MedB-1 is CD10(-), CD19(+), CD21(-). CD22(+), CD23(+) CD25(-), CD37(+), CD38(-), CD39(+), CD40(+), CD54(+), CD95(+), Like the parental tumor, MedB-1 lacks HLA-A,B,C alpha -chains and beta (2)microglobulin and expresses HLA-D molecules at decreased levels. Both parental tumor and MedB-1 cells are clonally related as shown by immunoglobulin heavy chain gene rearrangement analysis, Unlike the parental tumor tissue, the MedB-1 cell line cytoplasmically expresses IgG/kappa in a very small subset of cells under standard culture conditions. MedB-1 does not contain any Epstein-Barr virus DNA, In a tissue adhesion assay MedB-1 cells showed an extensive binding to the medullary region of normal thymus, Altogether, MedB-1 is a suitable tool for functional and molecular analysis of this distinct lymphoma entity. (C) 2001 Wiley-Liss. Inc.
C1 Univ Ulm, Inst Pathol, D-89081 Ulm, Germany.
   Deutsch Krebsforschungszentrum, Tumor Immunol Program, D-6900 Heidelberg, Germany.
   Deutsch Krebsforschungszentrum, Appl Tumor Virol Program, D-6900 Heidelberg, Germany.
C3 Ulm University; Helmholtz Association; German Cancer Research Center
   (DKFZ); Helmholtz Association; German Cancer Research Center (DKFZ)
RP Moller, P (通讯作者)，Univ Ulm, Inst Pathol, Albert Einstein Allee 11, D-89081 Ulm, Germany.
RI Pawlita, Labor/C-9720-2011
OI Pawlita, Michael/0000-0002-4720-8306
CR ADDIS BJ, 1986, HISTOPATHOLOGY, V10, P379, DOI 10.1111/j.1365-2559.1986.tb02491.x
   ALSHARABATI M, 1991, CANCER, V67, P2579, DOI 10.1002/1097-0142(19910515)67:10<2579::AID-CNCR2820671030>3.0.CO;2-H
   Bentz M, 1998, BLOOD, V92, p240A
   BENTZ M, IN PRESS GENES CHROM
   Bishop PC, 1999, J CLIN ONCOL, V17, P2479, DOI 10.1200/JCO.1999.17.8.2479
   BRANDTER LB, 1989, LEUKEMIA, V3, P122
   CARR TF, 1993, J CLIN PATHOL, V46, P513, DOI 10.1136/jcp.46.6.513
   Copie-Bergman R, 1999, BLOOD, V94, P3567, DOI 10.1182/blood.V94.10.3567.422k06_3567_3575
   DAVIS RE, 1990, HUM PATHOL, V21, P1262, DOI 10.1016/S0046-8177(06)80040-7
   EICHELMANN A, 1992, AM J PATHOL, V141, P729
   Harris NL, 1999, J CLIN ONCOL, V17, P3835, DOI 10.1200/JCO.1999.17.12.3835
   HOFMANN WJ, 1988, HUM PATHOL, V19, P1280, DOI 10.1016/S0046-8177(88)80282-X
   ISAACSON PG, 1987, LANCET, V2, P1488
   Joos S, 2000, CANCER RES, V60, P549
   Joos S, 1996, BLOOD, V87, P1571, DOI 10.1182/blood.V87.4.1571.bloodjournal8741571
   KIRN D, 1993, J CLIN ONCOL, V11, P1336, DOI 10.1200/JCO.1993.11.7.1336
   KNAUF WU, 1991, LEUKEMIA LYMPHOMA, V6, P49, DOI 10.3109/10428199109064878
   Kuppers R, 1997, EUR J IMMUNOL, V27, P1398, DOI 10.1002/eji.1830270616
   LAMARRE L, 1989, AM J SURG PATHOL, V13, P730, DOI 10.1097/00000478-198909000-00002
   LAWRENCE JB, 1988, CELL, V52, P51
   MENESTRINA F, 1986, HISTOPATHOLOGY, V10, P589, DOI 10.1111/j.1365-2559.1986.tb02512.x
   MILLER SA, 1988, NUCLEIC ACIDS RES, V16, P1215, DOI 10.1093/nar/16.3.1215
   MOLLER P, 1986, VIRCHOWS ARCH A, V409, P79, DOI 10.1007/BF00705408
   MOLLER P, 1987, INT J CANCER, V40, P32, DOI 10.1002/ijc.2910400107
   MOLLER P, 1987, BLOOD, V69, P1087
   MOLLER P, 1993, BLOOD, V81, P2067
   MOLLER P, 1989, PATHOLOGE, V10, P234
   MOLLER P, 1989, INT J CANCER, V43, P10, DOI 10.1002/ijc.2910430104
   MOMBURG F, 1987, INT J CANCER, V40, P598, DOI 10.1002/ijc.2910400504
   NACHEVA E, 1994, BLOOD, V84, P3422, DOI 10.1182/blood.V84.10.3422.bloodjournal84103422
   NADKARNI JS, 1969, CANCER, V23, P64, DOI 10.1002/1097-0142(196901)23:1<64::AID-CNCR2820230107>3.0.CO;2-M
   Paulli M, 1999, HUM PATHOL, V30, P178, DOI 10.1016/S0046-8177(99)90273-3
   PERRONE T, 1986, AM J SURG PATHOL, V10, P176, DOI 10.1097/00000478-198603000-00005
   POLACK A, 1984, GENE, V27, P279, DOI 10.1016/0378-1119(84)90072-6
   RAMASAMY I, 1992, J CLIN PATHOL, V45, P770, DOI 10.1136/jcp.45.9.770
   SCARPA A, 1987, VIRCHOWS ARCH A, V412, P17, DOI 10.1007/BF00750725
   Scarpa A, 1999, GENE CHROMOSOME CANC, V26, P203, DOI 10.1002/(SICI)1098-2264(199911)26:3<203::AID-GCC3>3.3.CO;2-5
   SCHNABL E, 1990, J EXP MED, V171, P1431, DOI 10.1084/jem.171.5.1431
   STAMPER HB, 1976, J EXP MED, V144, P828, DOI 10.1084/jem.144.3.828
   Tamura A, 1998, INT J CANCER, V78, P100
   TAYLOR KJ, 1990, BLOOD, V75, P2220
   Tsang P, 1996, AM J PATHOL, V148, P2017
NR 42
TC 30
Z9 32
U1 0
U2 0
PU WILEY-LISS
PI NEW YORK
PA DIV JOHN WILEY & SONS INC, 605 THIRD AVE, NEW YORK, NY 10158-0012 USA
SN 0020-7136
J9 INT J CANCER
JI Int. J. Cancer
PD MAY 1
PY 2001
VL 92
IS 3
BP 348
EP 353
DI 10.1002/ijc.1211
PG 6
WC Oncology
WE Science Citation Index Expanded (SCI-EXPANDED)
SC Oncology
GA 417TZ
UT WOS:000167851400008
PM 11291070
DA 2023-08-27
ER

PT J
AU Takami, H
   Graffeo, CS
   Perry, A
   Giannini, C
   Daniels, DJ
AF Takami, Hirokazu
   Graffeo, Christopher S.
   Perry, Avital
   Giannini, Caterina
   Daniels, David J.
TI The Third Eye Sees Double: Cohort Study of Clinical Presentation,
   Histology, Surgical Approaches, and Ophthalmic Outcomes in Pineal Region
   Germ Cell Tumors
SO WORLD NEUROSURGERY
LA English
DT Article
DE Endoscopic third ventriculostomy; Germinoma; Germ cell tumor;
   Hydrocephalus; Neuroophthalmology; Pineal gland
ID VENTRICULOSTOMY; HYDROCEPHALUS; MANAGEMENT
AB BACKGROUND: Intracranial germ cell tumors (GCTs) predominantly occur in the adolescent and young adult population and are most frequently located at the pineal gland. Tumor masses in the pineal region may cause ophthalmic symptoms due to compression to the midbrain, frequently presenting with Parinaud syndrome and hydrocephalus due to aqueductal compression.
   METHODS: We conducted a single-institution cohort study of primary, pineal region GCTs to characterize the clinical presentation, as well as associated ophthalmic and hydrocephalus outcomes.
   RESULTS: Fifty-six primary pineal GCTs were identified. Among the 40 isolated pineal region GCTs, 15 were germinomas while 25 were nongerminomatous GCTs. Among 43 cases of hydrocephalus, endoscopic third ventriculostomy was the primary treatment in 27 cases, which was successful in 23 but failed and required additional treatment for the rest.
   Pineal tumor mass was significantly larger in cases with hydrocephalus compared with those without, and the 20-mm diameter of the tumor was the crucial point for obstructive hydrocephalus.
   Ophthalmic symptoms were commonly observed at presentation, which included diplopia (74.3%), upward-gaze palsy (69.7%), and Argyll Robertson pupil (40%). These symptoms tended to remain, and the existence of these symptoms at presentation predicted the remaining symptoms in the follow-up.
   CONCLUSIONS: Intracranial GCTs presenting with ophthalmic abnormalities appear to be at increased risk of residual posttreatment symptoms, while second-look surgery presents a significant risk factor for the development of new deficits. Hydrocephalus often accompanies pineal region GCTs, and in most cases both cerebrospinal fluid diversion and tissue diagnosis can be successfully achieved via endoscopic third ventriculostomy.
C1 [Takami, Hirokazu; Graffeo, Christopher S.; Perry, Avital; Daniels, David J.] Mayo Clin, Dept Neurol Surg, Rochester, MN 55905 USA.
   [Giannini, Caterina] Mayo Clin, Lab Med & Pathol, Rochester, MN USA.
C3 Mayo Clinic; Mayo Clinic
RP Daniels, DJ (通讯作者)，Mayo Clin, Dept Neurol Surg, Rochester, MN 55905 USA.
EM
RI Giannini, Caterina/AAB-7551-2022
OI Takami, Hirokazu/0000-0001-9742-7462
CR BRUCE JN, 1995, ACTA NEUROCHIR, V134, P130, DOI 10.1007/BF01417679
   Feng HL, 2004, J NEUROSURG, V100, P626, DOI 10.3171/jns.2004.100.4.0626
   Fujimaki T, 2009, J CHILD NEUROL, V24, P1439, DOI 10.1177/0883073809342127
   Grand W, 2016, NEUROSURGERY, V78, P109, DOI 10.1227/NEU.0000000000000994
   Hankinson EV, 2016, J NEUROSURG-PEDIATR, V17, P558, DOI 10.3171/2015.10.PEDS15415
   Hart MG, 2013, J NEUROSURG, V119, P420, DOI 10.3171/2013.3.JNS122137
   HOFFMAN HJ, 1991, J NEUROSURG, V74, P545, DOI 10.3171/jns.1991.74.4.0545
   Isaacs AM, 2016, NEUROSURG FOCUS, V41, DOI 10.3171/2016.6.FOCUS16193
   JENNINGS MT, 1985, J NEUROSURG, V63, P155, DOI 10.3171/jns.1985.63.2.0155
   Jiang T, 2019, J CLIN NEUROSCI, V61, P93, DOI 10.1016/j.jocn.2018.10.136
   Kulkarni AV, 2010, J NEUROSURG-PEDIATR, V6, P310, DOI 10.3171/2010.8.PEDS103
   Kyritsis AP, 2010, J NEURO-ONCOL, V96, P143, DOI 10.1007/s11060-009-9951-z
   LOGOTHETIS CJ, 1982, CANCER, V50, P1629, DOI 10.1002/1097-0142(19821015)50:8<1629::AID-CNCR2820500828>3.0.CO;2-1
   Louis DNOH., 2016, WHO CLASSIFICATION T, V4th ed
   Matsutani M, 1997, J NEUROSURG, V86, P446, DOI 10.3171/jns.1997.86.3.0446
   Ray P., 2005, NEUROSURG FOCUS, V19, P1
   Reddy AT, 2004, NEURO-ONCOLOGY, V6, P127, DOI 10.1215/S1152851703000243
   Takami H, 2020, NEUROSURGERY, V87, P563, DOI 10.1093/neuros/nyaa108
   Takami H, 2021, J NEUROSURG, V134, P437, DOI 10.3171/2019.10.JNS191136
   Takami H, 2019, NEURO-ONCOLOGY, V21, P1565, DOI 10.1093/neuonc/noz139
   Villano JL, 2008, NEURO-ONCOLOGY, V10, P121, DOI 10.1215/15228517-2007-054
   Waqar M, 2016, WORLD NEUROSURG, V94, P386, DOI 10.1016/j.wneu.2016.07.028
NR 22
TC 7
Z9 7
U1 0
U2 0
PU ELSEVIER SCIENCE INC
PI NEW YORK
PA STE 800, 230 PARK AVE, NEW YORK, NY 10169 USA
SN 1878-8750
EI 1878-8769
J9 WORLD NEUROSURG
JI World Neurosurg.
PD JUN
PY 2021
VL 150
BP E482
EP E490
DI 10.1016/j.wneu.2021.03.030
EA JUN 2021
PG 9
WC Clinical Neurology; Surgery
WE Science Citation Index Expanded (SCI-EXPANDED)
SC Neurosciences & Neurology; Surgery
GA SM8GR
UT WOS:000657837500050
PM 33722723
DA 2023-08-27
ER

PT J
AU Yamakami, I
   Yoshinori, H
   Saeki, N
   Wada, M
   Oka, N
AF Yamakami, I.
   Yoshinori, H.
   Saeki, N.
   Wada, M.
   Oka, N.
TI Hearing preservation and intraoperative auditory brainstem response and
   cochlear nerve compound action potential monitoring in the removal of
   small acoustic neurinoma via the retrosigmoid approach
SO JOURNAL OF NEUROLOGY NEUROSURGERY AND PSYCHIATRY
LA English
DT Article
ID VESTIBULAR SCHWANNOMA SURGERY; CEREBELLOPONTINE ANGLE TUMOR; NEUROMA
   SURGERY; OPERATIONS; OUTCOMES
AB Objective: Hearing preservation is the main focus of small acoustic neurinoma (AN) removal. Refinement of intraoperative auditory monitoring may improve postoperative hearing. We have introduced a newly designed intracranial electrode enabling continuous monitoring of the cochlear nerve compound action potential (CNAP). We performed simultaneous monitoring of the auditory brainstem response (ABR) and CNAP during retrosigmoid small AN removal, and clarified the surgical outcome and the usefulness of CNAP monitoring.
   Methods: Twenty-two consecutive patients with a small AN underwent retrosigmoid tumour removal with attempting hearing preservation. ABR and CNAP were simultaneously monitored during tumour removal.
   Results: AN was totally removed in all patients without facial palsy. Preservation rate of useful and serviceable hearing was 82% and 91%, respectively. During microsurgical tumour removal, various surgical equipments and procedures intensified artefacts of ABR, and reliable ABR monitoring with distinct wave V was obtained in 9/22 patients. Unaffected by artefacts, reliable CNAP monitoring was obtained more frequently (in 20/22 patients) than ABR (p = 0.0005). CNAP on completion of tumour removal predicted hearing preservation with no false positive or negative (100% sensitivity and 100% specificity). CNAP changed dynamically and stepwise with surgical manipulations.
   Conclusion: The retrosigmoid approach using auditory monitoring for a small AN can accomplish total tumour removal with an excellent hearing preservation rate. CNAP provides reliable auditory monitoring more frequently than ABR, reflects the intraoperative auditory function almost in real-time, predicts postoperative hearing with excellent sensitivity and specificity, and is more useful for monitoring in the removal of small AN with hearing preservation.
C1 [Yamakami, I.] Chiba Univ, Grad Sch Med, Chuo Ku, Chiba 2608670, Japan.
C3 Chiba University
RP Yamakami, I (通讯作者)，Chiba Univ, Grad Sch Med, Chuo Ku, 1-8-1 Inohana, Chiba 2608670, Japan.
EM
RI Higuchi, Yoshinori/B-3459-2017
OI Higuchi, Yoshinori/0000-0001-5689-3416
CR [Anonymous], 1995, Otolaryngol Head Neck Surg, V113, P179
   Arriaga MA, 1997, LARYNGOSCOPE, V107, P1043, DOI 10.1097/00005537-199708000-00007
   Arts HA, 2006, OTOL NEUROTOL, V27, P234, DOI 10.1097/01.mao.0000185153.54457.16
   Battista RA, 2000, AM J OTOL, V21, P244, DOI 10.1016/S0196-0709(00)80017-4
   Chopra R, 2007, INT J RADIAT ONCOL, V68, P845, DOI 10.1016/j.ijrobp.2007.01.001
   Colletti V, 1996, SKULL BASE SURG, V6, P153, DOI 10.1055/s-2008-1058639
   Colletti V, 1998, AUDIOLOGY, V37, P27
   COLLETTI V, 1994, SKULL BASE SURG, V4, P1, DOI 10.1055/s-2008-1058981
   Colletti V, 1999, OTOLARYNG HEAD NECK, V120, P122, DOI 10.1016/S0194-5998(99)70382-7
   Danner C, 2004, OTOL NEUROTOL, V25, P826, DOI 10.1097/00129492-200409000-00029
   Gjuric M, 2001, OTOL NEUROTOL, V22, P223, DOI 10.1097/00129492-200103000-00019
   Hempel JM, 2006, EUR ARCH OTO-RHINO-L, V263, P714, DOI 10.1007/s00405-006-0054-6
   Jackson LE, 2000, AM J OTOL, V21, P249, DOI 10.1016/S0196-0709(00)80018-6
   KAGA K, 1979, ARCH OTO-RHINO-LARYN, V225, P199, DOI 10.1007/BF00455255
   Koos WT, 1998, J NEUROSURG, V88, P506, DOI 10.3171/jns.1998.88.3.0506
   MANNINEN PH, 1985, ANESTH ANALG, V64, P43
   Matthies C, 1997, NEUROSURGERY, V40, P919, DOI 10.1097/00006123-199705000-00007
   Mazzoni A, 2000, AM J OTOL, V21, P98, DOI 10.1016/S0196-0709(00)80082-4
   Meyer TA, 2006, OTOL NEUROTOL, V27, P380, DOI 10.1097/00129492-200604000-00015
   Mohr G, 2005, J NEUROSURG, V102, P1, DOI 10.3171/jns.2005.102.1.0001
   MOLLER AR, 1982, ARCH OTOLARYNGOL, V108, P77
   MOLLER AR, 1981, EXP NEUROL, V74, P862, DOI 10.1016/0014-4886(81)90259-4
   Moller AR, 1996, AM J OTOL, V17, P452
   NADOL JB, 1992, LARYNGOSCOPE, V102, P1153, DOI 10.1288/00005537-199210000-00010
   Piccirillo E, 2008, AUDIOL NEURO-OTOL, V13, P58, DOI 10.1159/000108623
   Prasad D, 2000, J NEUROSURG, V92, P745, DOI 10.3171/jns.2000.92.5.0745
   Roberson J, 1996, AM J OTOL, V17, P653
   Roberson JB, 1999, LARYNGOSCOPE, V109, P904, DOI 10.1097/00005537-199906000-00012
   Samii M, 2006, J NEUROSURG, V105, P527, DOI 10.3171/jns.2006.105.4.527
   Schmerber S, 2004, ACTA OTO-LARYNGOL, V124, P53, DOI 10.1080/00016480310014840
   SEKIYA T, 1987, NEUROSURGERY, V21, P631, DOI 10.1227/00006123-198711000-00005
   SELTERS WA, 1977, ARCH OTOLARYNGOL, V103, P181
   Slattery WH, 1997, AM J OTOL, V18, P596
   Thomassin JM, 1998, STEREOT FUNCT NEUROS, V70, P74, DOI 10.1159/000056409
   van Eck ATCJ, 2005, J NEUROSURG, V102, P204, DOI 10.3171/jns.2005.102.s_supplement.0204
   Yamakami I, 2000, NEUROL MED-CHIR, V40, P597, DOI 10.2176/nmc.40.597
   Yamakami I, 2004, J NEUROL NEUROSUR PS, V75, P453, DOI 10.1136/jnnp.2003.010827
   Yamakami I, 2003, J CLIN NEUROSCI, V10, P567, DOI 10.1016/S0967-5868(03)00143-7
   Yamakami I, 2002, NEUROL SURG TOKYO, V30, P275
NR 39
TC 45
Z9 52
U1 0
U2 1
PU BMJ PUBLISHING GROUP
PI LONDON
PA BRITISH MED ASSOC HOUSE, TAVISTOCK SQUARE, LONDON WC1H 9JR, ENGLAND
SN 0022-3050
EI 1468-330X
J9 J NEUROL NEUROSUR PS
JI J. Neurol. Neurosurg. Psychiatry
PD FEB
PY 2009
VL 80
IS 2
BP 218
EP 227
DI 10.1136/jnnp.2008.156919
PG 10
WC Clinical Neurology; Psychiatry; Surgery
WE Science Citation Index Expanded (SCI-EXPANDED)
SC Neurosciences & Neurology; Psychiatry; Surgery
GA 396HH
UT WOS:000262582300020
PM 18977821
DA 2023-08-27
ER

PT J
AU Kohzuki, H
   Matsuda, M
   Miki, S
   Shibuya, M
   Ishikawa, E
   Matsumura, A
AF Kohzuki, Hidehiro
   Matsuda, Masahide
   Miki, Shunichiro
   Shibuya, Makoto
   Ishikawa, Eiichi
   Matsumura, Akira
TI Diffusely Infiltrating Cerebellar Anaplastic Astrocytoma Effectively
   Controlled with Bevacizumab: Case Report and Literature Review
SO WORLD NEUROSURGERY
LA English
DT Review
DE Bevacizumab; Cerebellar glioma; Diffuse infiltration
ID GLIOMATOSIS CEREBELLI; CEREBRI
AB BACKGROUND: Gliomas that show extensive diffuse infiltration from the cerebellumto the brainstem without masslike expansion are extremely rare. The efficacy of bevacizumab treatment for diffusely infiltrating gliomas remains uncertain.
   CASE DESCRIPTION: A 75-year-old man presented with a cerebellar anaplastic astrocytoma showing diffuse infiltration to the brainstem without a definite mass. He had experienced rapidly progressive nausea and dysarthria, as well as vertigo and headache for 2 months. Magnetic resonance imaging (MRI) revealed a poorly demarcated T2 high-intensity area in the right cerebellum and brainstem. The tumor in the right cerebellum showed sparse enhancement with gadolinium (Gd). Suboccipital decompressive craniotomy and partial removal of the tumor was emergently performed because of the rapid progression of symptoms and severe tonsillar herniation demonstrated on MRI. The pathologic diagnosis was anaplastic astrocytoma, and genomic analyses revealed no mutation in IDH1, H3F3A, or BRAF. During concomitant chemoradiotherapy with temozolomide, rapid worsening of the neurologic symptoms developed and significant enlargement of the T2 high-intensity area extending to the cerebral peduncle was seen, as well as a new Gd-enhancing lesion in the midbrain. After administration of bevacizumab, the neurologic symptoms gradually improved, the T2 high-intensity area decreased, and the Gd-enhancing lesion disappeared. At follow-up 2 years after the operation, no worsening of neurologic symptoms was seen and the residual T2 high-intensity area remained unchanged on MRI.
   CONCLUSIONS: Bevacizumab treatment may be a salvage treatment option for patients with diffusely infiltrating cerebellar gliomas that exhibits rapid progression during standard treatment.
C1 [Kohzuki, Hidehiro; Matsuda, Masahide; Miki, Shunichiro; Ishikawa, Eiichi; Matsumura, Akira] Univ Tsukuba, Fac Med, Dept Neurosurg, Tsukuba, Ibaraki, Japan.
   [Shibuya, Makoto] Tokyo Med Univ, Hachioji Med Ctr, Cent Clin Lab, Hachioji, Tokyo, Japan.
C3 University of Tsukuba; Tokyo Medical University
RP Matsuda, M (通讯作者)，Univ Tsukuba, Fac Med, Dept Neurosurg, Tsukuba, Ibaraki, Japan.
EM
CR [Anonymous], 2016, WHO CLASSIFICATION T
   Bates DO, 2010, CARDIOVASC RES, V87, P262, DOI 10.1093/cvr/cvq105
   BURGER MC, 2017, INT J MOL SCI, V18
   Chen S, 2013, J NEURO-ONCOL, V112, P267, DOI 10.1007/s11060-013-1058-x
   Chinot OL, 2014, NEW ENGL J MED, V370, P709, DOI 10.1056/NEJMoa1308345
   de Chadarevian JP, 2012, J CHILD NEUROL, V27, P511, DOI 10.1177/0883073811419315
   Inoue T, 2011, NEUROSURG REV, V34, P197, DOI 10.1007/s10143-010-0306-1
   KIM KJ, 1993, NATURE, V362, P841, DOI 10.1038/362841a0
   Matsuda K, 2013, BRAIN TUMOR PATHOL, V30, P180, DOI 10.1007/s10014-012-0121-1
   Matsuda M, 2017, J CLIN NEUROSCI, V40, P115, DOI 10.1016/j.jocn.2017.02.025
   Nakahara A, 2014, NEUROPATHOLOGY, V34, P295, DOI 10.1111/neup.12083
   Rajz GG, 2012, ONCOL LETT, V3, P209, DOI 10.3892/ol.2011.445
   Rorke-Adams LB, 2008, J NEUROSURG-PEDIATR, V2, P346, DOI 10.3171/PED.2008.2.11.346
   Solomon DA, 2016, BRAIN PATHOL, V26, P569, DOI 10.1111/bpa.12336
   Taillibert S, 2006, J NEURO-ONCOL, V76, P201, DOI 10.1007/s11060-005-5263-0
NR 15
TC 1
Z9 1
U1 0
U2 1
PU ELSEVIER SCIENCE INC
PI NEW YORK
PA 360 PARK AVE SOUTH, NEW YORK, NY 10010-1710 USA
SN 1878-8750
EI 1878-8769
J9 WORLD NEUROSURG
JI World Neurosurg.
PD JUL
PY 2018
VL 115
BP 181
EP 185
DI 10.1016/j.wneu.2018.04.110
PG 5
WC Clinical Neurology; Surgery
WE Science Citation Index Expanded (SCI-EXPANDED)
SC Neurosciences & Neurology; Surgery
GA GK8YL
UT WOS:000436519600133
PM 29704688
OA Green Submitted
DA 2023-08-27
ER

PT J
AU Kline, C
   Liu, SJ
   Duriseti, S
   Banerjee, A
   Nicolaides, T
   Raber, S
   Gupta, N
   Haas-Kogan, D
   Braunstein, S
   Mueller, S
AF Kline, Cassie
   Liu, S. John
   Duriseti, Sai
   Banerjee, Anuradha
   Nicolaides, Theodore
   Raber, Shannon
   Gupta, Nalin
   Haas-Kogan, Daphne
   Braunstein, Steve
   Mueller, Sabine
TI Reirradiation and PD-1 inhibition with nivolumab for the treatment of
   recurrent diffuse intrinsic pontine glioma: a single-institution
   experience
SO JOURNAL OF NEURO-ONCOLOGY
LA English
DT Article
DE DIPG; Reirradiation; PD-1 inhibition; Survival
ID IMMUNE CHECKPOINT BLOCKADE; MELANOMA BRAIN METASTASES; LIGAND 1
   EXPRESSION; HYPERFRACTIONATED RADIOTHERAPY; STEM GLIOMAS; RADIATION;
   CHILDREN; SURVIVAL; GLIOBLASTOMA; IPILIMUMAB
AB Background Diffuse intrinsic pontine glioma (DIPG) is a rare, aggressive brain tumor with no known cure. Reirradiation (reRT) at recurrence can prolong survival. The impact of irradiation may be heightened when combined with PD-1 inhibition. We describe our experience using reRT, with or without PD-1 inhibition, in a cohort of patients with recurrent DIPG.
   Methods We performed a retrospective cohort analysis of children who received reRT with or without concomitant PD-1 inhibition for recurrent DIPG at a single institution between 2005 and 2016. We compared progression-free (PFS) and overall survival (OS) between those who received reRT alone or in combination with PD-1 inhibition. We then compared reRT to a cohort of patients who did not receive reRT.
   Results Thirty-one patients were included (8-reRT with nivolumab; 4-reRT alone; 19-no reRT). Patients who received reRT had prolonged OS compared to no reRT (22.9 months-reRT with nivolumab; 20.4 months-reRT alone; 8.3monthsno reRT; p < 0.0001). Patients who received reRT with nivolumab vs. reRT only had slightly prolonged OS from diagnosis and from reRT (22.9 vs. 20.4 months for time from diagnosis; 6.8 vs. 6.0 months for time from reRT). All patients receiving reRT with or without nivolumab tolerated the therapy without acute or late toxicity.
   Conclusions Our experience demonstrates the tolerability of reRT with concurrent PD-1 inhibition for recurrent DIPG and suggests that combination therapy may offer survival benefit. Future prospective studies are needed to confirm the benefits of this combination therapy.
C1 [Kline, Cassie; Banerjee, Anuradha; Raber, Shannon; Gupta, Nalin; Mueller, Sabine] Univ Calif San Francisco, Dept Pediat, San Francisco, CA 94143 USA.
   [Liu, S. John] Univ Calif San Francisco, Sch Med, San Francisco, CA 94143 USA.
   [Duriseti, Sai] Washington Univ, Dept Radiat Oncol, St Louis, MO USA.
   [Kline, Cassie; Mueller, Sabine] Univ Calif San Francisco, Dept Neurol, San Francisco, CA 94143 USA.
   [Banerjee, Anuradha; Nicolaides, Theodore; Gupta, Nalin; Mueller, Sabine] Univ Calif San Francisco, Dept Neurol Surg, San Francisco, CA 94143 USA.
   [Nicolaides, Theodore] NYU, Dept Pediat, New York, NY 10016 USA.
   [Haas-Kogan, Daphne] Dana Farber Canc Inst, Dept Radiat Oncol, Boston, MA 02115 USA.
   [Braunstein, Steve] Univ Calif San Francisco, Dept Radiat Oncol, San Francisco, CA 94143 USA.
C3 University of California System; University of California San Francisco;
   University of California System; University of California San Francisco;
   Washington University (WUSTL); University of California System;
   University of California San Francisco; University of California System;
   University of California San Francisco; New York University; Harvard
   University; Dana-Farber Cancer Institute; University of California
   System; University of California San Francisco
RP Kline, C; Mueller, S (通讯作者)，Univ Calif San Francisco, Dept Pediat, San Francisco, CA 94143 USA.; Kline, C; Mueller, S (通讯作者)，Univ Calif San Francisco, Dept Neurol, San Francisco, CA 94143 USA.; Mueller, S (通讯作者)，Univ Calif San Francisco, Dept Neurol Surg, San Francisco, CA 94143 USA.
EM
OI Nicolaides, Theodore/0000-0002-0637-1086
FU Frank A. Campini Foundation; Cannonball Kids' Cancer [A129749]; National
   Institutes of Health National Center for Advancing Translational
   Sciences [KL2TR001870]; William M. Wood Foundation
FX C.N.K. is supported by Frank A. Campini Foundation, Cannonball Kids'
   Cancer [A129749], and National Institutes of Health National Center for
   Advancing Translational Sciences [KL2TR001870-This content is solely the
   responsibility of the authors and does not necessarily represent the
   official views of the NIH]. The research was supported in part by the
   William M. Wood Foundation (D.H.K).
CR Allen J, 1999, CANCER, V86, P1064, DOI 10.1002/(SICI)1097-0142(19990915)86:6<1064::AID-CNCR24>3.0.CO;2-1
   Berghoff AS, 2015, NEURO-ONCOLOGY, V17, P1064, DOI 10.1093/neuonc/nou307
   Bouffet E, 2016, J CLIN ONCOL, V34, P2206, DOI 10.1200/JCO.2016.66.6552
   Broniscer A, 2004, ONCOLOGIST, V9, P197, DOI 10.1634/theoncologist.9-2-197
   Castel D, 2015, ACTA NEUROPATHOL, V130, P815, DOI 10.1007/s00401-015-1478-0
   Chintagumpala M, 2015, PEDIATR CLIN N AM, V62, P167, DOI 10.1016/j.pcl.2014.09.011
   Cohen K J, 2001, Curr Treat Options Oncol, V2, P529
   Donahue B, 1998, INT J RADIAT ONCOL, V40, P677, DOI 10.1016/S0360-3016(97)00846-8
   Khuong-Quang DA, 2012, ACTA NEUROPATHOL, V124, P439, DOI 10.1007/s00401-012-0998-0
   Fontanilla HP, 2012, AM J CLIN ONCOL-CANC, V35, P51, DOI 10.1097/COC.0b013e318201a2b7
   FREEMAN CR, 1993, INT J RADIAT ONCOL, V27, P197, DOI 10.1016/0360-3016(93)90228-N
   Freese C, 2017, PRACT RADIAT ONCOL, V7, P86, DOI 10.1016/j.prro.2016.11.005
   Fried I, 2018, J NEURO-ONCOL, V136, P189, DOI 10.1007/s11060-017-2643-1
   Garber ST, 2016, NEURO-ONCOLOGY, V18, P1357, DOI 10.1093/neuonc/now132
   Hargrove D, 2006, LANCET ONCOL, V7, P241, DOI 10.1016/S1470-2045(06)70615-5
   Hellmann MD, 2016, ADV IMMUNOL, V130, P251, DOI 10.1016/bs.ai.2015.12.005
   Hwang EOA, 2018, INT SOC PEDIAT NEURO
   Hwang K, 2018, J NEUROONCOL, V54, P307
   Janssens GO, 2017, EUR J CANCER, V73, P38, DOI 10.1016/j.ejca.2016.12.007
   Kaye Erica C, 2014, CNS Oncol, V3, P421, DOI 10.2217/cns.14.47
   Kiess AP, 2015, INT J RADIAT ONCOL, V92, P368, DOI 10.1016/j.ijrobp.2015.01.004
   Knisely JPS, 2012, J NEUROSURG, V117, P227, DOI 10.3171/2012.5.JNS111929
   Lam S, 2015, J PEDIATR NEUROSCI, V10, P199, DOI 10.4103/1817-1745.165656
   Lassaletta A, 2018, PEDIATR BLOOD CANCER, V65, DOI 10.1002/pbc.26988
   Majzner RG, 2017, CANCER-AM CANCER SOC, V123, P3807, DOI 10.1002/cncr.30724
   Mandell LR, 1999, INT J RADIAT ONCOL, V43, P959, DOI 10.1016/S0360-3016(98)00501-X
   Massimino M, 2014, J NEURO-ONCOL, V118, P305, DOI 10.1007/s11060-014-1428-z
   Miyazaki T, 2017, J NEURO-ONCOL, V133, P277, DOI 10.1007/s11060-017-2451-7
   Nazarian J, 2016, ONCOTARGET, V7, P42837, DOI 10.18632/oncotarget.10034
   Okada Hideho, 2015, Lancet Oncol, V16, pe534, DOI 10.1016/S1470-2045(15)00088-1
   Packer Roger J., 1994, Current Opinion in Oncology, V6, P240, DOI 10.1097/00001622-199405000-00004
   Reardon DA, 2016, CANCER IMMUNOL RES, V4, P124, DOI 10.1158/2326-6066.CIR-15-0151
   Schild SE, 1998, J NEURO-ONCOL, V40, P171, DOI 10.1023/A:1006193306286
   Sethi R, 2011, J NEURO-ONCOL, V102, P121, DOI 10.1007/s11060-010-0301-y
   Sharabi AB, 2015, LANCET ONCOL, V16, pE498, DOI 10.1016/S1470-2045(15)00007-8
   Svolos P, 2017, AM J NEURORADIOL, V38, P1235, DOI 10.3174/ajnr.A5159
   Trainer H, 2016, ENDOCRINOL DIAB META, DOI 10.1530/EDM-16-0108
   van den Bent MJ, 2009, J CLIN ONCOL, V27, P2905, DOI 10.1200/JCO.2009.22.4998
   van Zanten SEMV, 2015, EXPERT REV ANTICANC, V15, P157, DOI 10.1586/14737140.2015.974563
   Vandiver JW, 2016, TARGET ONCOL, V11, P553, DOI 10.1007/s11523-016-0426-9
   Vlahovic G, 2015, NEURO-ONCOLOGY, V17, P1043, DOI 10.1093/neuonc/nov071
   Wainwright DA, 2014, CLIN CANCER RES, V20, P5290, DOI 10.1158/1078-0432.CCR-14-0514
   Walston S, 2015, FRONT ONCOL, V5, DOI 10.3389/fonc.2015.00028
   Wolchok JD, 2013, NEW ENGL J MED, V369, P122, DOI 10.1056/NEJMoa1302369
   Wolff JE, 2012, J NEURO-ONCOL, V106, P391, DOI 10.1007/s11060-011-0677-3
   Wong ET, 2015, BRIT J CANCER, V113, P232, DOI 10.1038/bjc.2015.238
   Zeng J, 2013, INT J RADIAT ONCOL, V86, P343, DOI 10.1016/j.ijrobp.2012.12.025
NR 47
TC 31
Z9 32
U1 1
U2 6
PU SPRINGER
PI NEW YORK
PA 233 SPRING ST, NEW YORK, NY 10013 USA
SN 0167-594X
EI 1573-7373
J9 J NEURO-ONCOL
JI J. Neuro-Oncol.
PD DEC
PY 2018
VL 140
IS 3
BP 629
EP 638
DI 10.1007/s11060-018-2991-5
PG 10
WC Oncology; Clinical Neurology
WE Science Citation Index Expanded (SCI-EXPANDED)
SC Oncology; Neurosciences & Neurology
GA HC2MG
UT WOS:000451635500016
PM 30206764
OA Green Published
DA 2023-08-27
ER

PT J
AU Nomikos, IN
   Zizi-Serbetzoglou, A
   Matsakis, G
   Elemenoglou, J
   Vamvakopoulos, NC
AF Nomikos, I. N.
   Zizi-Serbetzoglou, A.
   Matsakis, G.
   Elemenoglou, J.
   Vamvakopoulos, N. C.
TI Association of an oversized adrenal cortical adenoma with expression of
   pheochromocytoma-like neurosecretory features
SO JOURNAL OF BUON
LA English
DT Article
DE adrenal tumor; pheochromocytoma
ID INCIDENTALOMAS; MANAGEMENT; DIAGNOSIS; MARKERS
AB Abnormal stimulation of adrenal function may be either direct, affecting similarly cortical and medullary secretion, or indirect affecting primarily the medulla. Indirect activation of clinically detectable adrenomedullary function may develop as a physical consequence of a non-functional adrenal tumor exerting pressure on the medulla by its size, location and direction of growth. Our case of an oversized and overweight adrenal tumor associated with expression of late-onset pheochromocytoma-like clinical symptoms may be explained by the physical indirect rather than the biological direct activation of adrenomedullary function like hyperplasia or cancer.
C1 [Nomikos, I. N.; Matsakis, G.] Metaxa Canc Hosp, Dept Surg B Unit, Piraeus, Greece.
   [Zizi-Serbetzoglou, A.; Elemenoglou, J.] Tzanion Gen Hosp, Dept Pathol, Piraeus, Greece.
   [Vamvakopoulos, N. C.] Univ Thessalia, Sch Med, Dept Biol & Genet, Larisa, Greece.
C3 University of Thessaly
RP Nomikos, IN (通讯作者)，55 Psarron St, Koridallos 18120, Piraeus, Greece.
EM
CR CAPLAN RH, 1994, ARCH SURG-CHICAGO, V129, P291
   Clarke MR, 1998, HUM PATHOL, V29, P522, DOI 10.1016/S0046-8177(98)90070-3
   Graham D J, 1998, Surg Oncol Clin N Am, V7, P749
   Ivsic T, 2002, ARCH PATHOL LAB MED, V126, P1530
   KOMMINOTH P, 1995, LAB INVEST, V72, P424
   Mansmann G, 2004, ENDOCR REV, V25, P309, DOI 10.1210/er.2002-0031
   Nawar R, 2005, ENDOCR-RELAT CANCER, V12, P585, DOI 10.1677/erc.1.00951
   Nomikos IN, 2006, WOUNDS, V18, P177
   Nomikos LN, 2001, AM J SURG, V182, P278, DOI 10.1016/S0002-9610(01)00701-2
   Sidhu S, 2003, ANZ J SURG, V73, P727, DOI 10.1046/j.1445-2197.2003.02746.x
   Stone JH, 2006, NEW ENGL J MED, V354, P2748, DOI 10.1056/NEJMp058264
   VAMVAKOPOULOS NV, 1995, MEDIAT INFLAMM, V4, P163, DOI 10.1155/S0962935195000275
   Young WF, 2000, ENDOCRIN METAB CLIN, V29, P159, DOI 10.1016/S0889-8529(05)70122-5
NR 13
TC 1
Z9 1
U1 0
U2 0
PU ZERBINIS MEDICAL PUBL
PI ATHENS
PA 42 ANTIFILOU STR, ATHENS, 157 71, GREECE
SN 1107-0625
J9 J BUON
JI J. BUON
PD JUL-SEP
PY 2008
VL 13
IS 3
BP 425
EP 428
PG 4
WC Oncology
WE Science Citation Index Expanded (SCI-EXPANDED)
SC Oncology
GA 355DM
UT WOS:000259689200018
PM 18979561
DA 2023-08-27
ER

PT J
AU FULHAM, MJ
   BROOKS, RA
   HALLETT, M
   DICHIRO, G
AF FULHAM, MJ
   BROOKS, RA
   HALLETT, M
   DICHIRO, G
TI CEREBELLAR DIASCHISIS REVISITED - PONTINE HYPOMETABOLISM AND DENTATE
   SPARING
SO NEUROLOGY
LA English
DT Article
ID POSITRON EMISSION TOMOGRAPHY; CEREBRAL GLUCOSE-UTILIZATION; 2 MAINTAINED
   POSTURES; BLOOD-FLOW; PROMPT MOVEMENTS; CELL OUTPUT; NEURONS;
   METABOLISM; INFARCTION; MONKEY
AB A unilateral supratentorial lesion may cause hypometabolism in the contralateral cerebellar hemisphere (crossed cerebellar diaschisis). We analyzed glucose metabolism, measured by PET-FDG, in the posterior fossa in 67 patients (78 PET studies) with primary unilateral supratentorial brain tumors selected for visually obvious metabolic asymmetry in the cerebellar hemispheres. We found that glucose utilization was 17% lower in the contralateral cerebellar cortex (compared with the ipsilateral one), consistent with the selection criterion, and 19% lower in the ipsilateral pons, wherein lie the first order synapses of the corticopontocerebellar pathway. This finding helps to validate the prevalent view that cerebellar diaschisis is due to interruption of afferent input from the corticopontocerebellar pathway. However, glucose-metabolism in the contralateral dentate nucleus was relatively preserved-only 2% less than the ipsilateral dentate. This "dentate sparing" suggests preservation of afferent input to the largest of the deep cerebellar nuclei from the Purkinje cells in the cortex, despite interruption of the major excitatory input to the Purkinje cells.
C1 NINCDS,NEUROIMAGING BRANCH,BETHESDA,MD 20892.
   NINCDS,MED NEUROL BRANCH,BETHESDA,MD 20892.
C3 National Institutes of Health (NIH) - USA; NIH National Institute of
   Neurological Disorders & Stroke (NINDS); National Institutes of Health
   (NIH) - USA; NIH National Human Genome Research Institute (NHGRI); NIH
   National Institute of Neurological Disorders & Stroke (NINDS)
OI Fulham, Michael/0000-0003-0602-6319
CR AFSHAR F, 1978, STEREOTAXIC ATLAS HU, P211
   ARMSTRONG DM, 1979, J PHYSIOL-LONDON, V289, P425, DOI 10.1113/jphysiol.1979.sp012745
   BARON JC, 1980, ANN NEUROL, V8, P128
   BRODAL A, 1981, NEUROLOGICAL ANATOMY, P294
   BRODAL P, 1978, BRAIN, V101, P251, DOI 10.1093/brain/101.2.251
   BROOKS RA, 1982, J NUCL MED, V23, P538
   BROOKS RA, 1985, METABOLISM HUMAN BRA, P351
   Brooks VB, 1981, HDB PHYSL NERVOUS SY, V2, P877
   BROOKS VB, 1984, NEURAL BASIS MOTOR C, P256
   CREPAX P, 1955, Boll Soc Ital Biol Sper, V31, P1229
   CREPEL F, 1976, J NEUROBIOL, V7, P579, DOI 10.1002/neu.480070610
   DICHIRO G, 1982, NEUROLOGY, V32, P1323, DOI 10.1212/WNL.32.12.1323
   DICHIRO G, 1984, ANN NEUROL S, V15, P138
   ECCLES JC, 1967, CEREBELLUM NEURONAL, P188
   FEENEY DM, 1986, STROKE, V17, P817, DOI 10.1161/01.STR.17.5.817
   FULHAM M, 1989, ANN NEUROL, V26, P172
   HEIDARY H, 1969, ACTA ANAT, V74, P290
   Ito M, 1964, Experientia, V20, P295, DOI 10.1007/BF02151821
   ITO M, 1984, CEREBELLUM NEURAL CO, P13
   KENNEDY C, 1978, ANN NEUROL, V4, P293, DOI 10.1002/ana.410040402
   KENNEDY C, 1991, J CEREBR BLOOD F MET, V11, P885, DOI 10.1038/jcbfm.1991.153
   KUSHNER M, 1984, ANN NEUROL, V15, P425, DOI 10.1002/ana.410150505
   LENZI GL, 1982, J CEREBR BLOOD F MET, V2, P321, DOI 10.1038/jcbfm.1982.33
   LLINAS RR, 1988, SCIENCE, V242, P1654, DOI 10.1126/science.3059497
   MARTIN WRW, 1983, ANN NEUROL, V14, P168
   MENEGHETTI G, 1984, J CEREBR BLOOD F MET, V4, P235, DOI 10.1038/jcbfm.1984.32
   NUDO RJ, 1986, J COMP NEUROL, V245, P553, DOI 10.1002/cne.902450410
   PALKOVITS M, 1977, EXP BRAIN RES, V28, P189
   PANTANO P, 1986, BRAIN, V109, P677, DOI 10.1093/brain/109.4.677
   PATRONAS NJ, 1984, BRAIN RES, V291, P93, DOI 10.1016/0006-8993(84)90654-1
   PERANI D, 1987, J CEREBR BLOOD F MET, V7, P507, DOI 10.1038/jcbfm.1987.95
   PHELPS ME, 1979, ANN NEUROL, V6, P371, DOI 10.1002/ana.410060502
   RAICHLE ME, 1986, HDB PHYSL NERVOUS 5, P643
   ROZENTAL JM, 1990, J NEURO-ONCOL, V8, P153
   SEJNOWSKI TJ, 1980, NATURE, V287, P449, DOI 10.1038/287449a0
   SHIMOYAMA I, 1988, ANN NEUROL, V24, P718, DOI 10.1002/ana.410240605
   SOKOLOFF L, 1977, J NEUROCHEM, V28, P897, DOI 10.1111/j.1471-4159.1977.tb10649.x
   THACH WT, 1970, J NEUROPHYSIOL, V33, P527, DOI 10.1152/jn.1970.33.4.527
   THACH WT, 1970, J NEUROPHYSIOL, V33, P537, DOI 10.1152/jn.1970.33.4.537
   VIEUSSENS R, 1684, NEUROGRAPHIA UNIVERS, P73
   VONMONAKOW C, 1910, NEUE GESICHTSPUNKTE, P1
   WAGNER HJ, 1981, BRAIN RES, V224, P31, DOI 10.1016/0006-8993(81)91114-8
   WEST JR, 1978, BEHAV BIOL, V22, P413, DOI 10.1016/S0091-6773(78)92546-4
NR 43
TC 51
Z9 51
U1 0
U2 0
PU LIPPINCOTT-RAVEN PUBL
PI PHILADELPHIA
PA 227 EAST WASHINGTON SQ, PHILADELPHIA, PA 19106
SN 0028-3878
J9 NEUROLOGY
JI Neurology
PD DEC
PY 1992
VL 42
IS 12
BP 2267
EP 2273
DI 10.1212/WNL.42.12.2267
PG 7
WC Clinical Neurology
WE Science Citation Index Expanded (SCI-EXPANDED)
SC Neurosciences & Neurology
GA KC375
UT WOS:A1992KC37500005
PM 1461377
DA 2023-08-27
ER

PT J
AU Eisenhofer, G
   Goldstein, DS
   Kopin, IJ
   Crout, JR
AF Eisenhofer, G
   Goldstein, DS
   Kopin, IJ
   Crout, JR
TI Pheochromocytoma: Rediscovery as a catecholamine-metabolizing tumor
SO ENDOCRINE PATHOLOGY
LA English
DT Review
DE pheochromocytoma; paraganglioma; chromaffin cells; adrenal glands;
   hypertension; catecholamines; norepinephrine; epinephrine;
   normetanephrine; metanephrine; synthesis; metabolism; secretion
ID BIOCHEMICAL-DIAGNOSIS; PLASMA METANEPHRINES; LIQUID-CHROMATOGRAPHY;
   NOREPINEPHRINE METABOLISM; O-METHYLTRANSFERASE; URINARY;
   NORMETANEPHRINE; INACTIVATION; ACID; CONVERSION
AB Catecholamine-producing tumors are rare neoplasms derived mainly from chromaffin cells of the adrenal medulla (pheochromocytomas)or, in about 10% of cases, from paraganglia (paragangliomas). Diagnosis of these tumors relies heavily on measurements of urinary or plasma catecholamines or catecholamine metabolites. The metabolites are usually thought to be produced after release of catecholamines into the bloodstream. This, however, ignores observations of over 40 yr ago that catecholamines are metabolized within pheochromocytoma tumor cells. Development of improved methods for measurement of catecholamine metabolites, in particular, plasma concentrations of free normetanephrine and metanephrine, has reestablished the importance of intratumoral catecholamine metabolism. In patients with pheochromocytoma, over 90% of the elevations in plasma free normetanephrine and metanephrine result from metabolism of catecholamines within pheochromocytoma tumor cells. This process occurs continuously and independently of variations in catecholamine release. As a consequence, measurements of plasma concentrations and urinary outputs of normetanephrine and metanephrine provide more reliable methods for diagnosis of pheochromocytoma than measurements of the parent amines. Rediscovery of the importance of intratumoral catecholamine metabolism is leading to a reevaluation of the procedures used to diagnose pheochromocytoma. This review provides an update on the diagnosis of pheochromocytoma, with emphasis on identifying and correcting relevant misconceptions about catecholamine metabolism.
C1 Natl Inst Neurol Disorders & Stroke, Clin Neurocardiol Sect, NIH, Bethesda, MD USA.
C3 National Institutes of Health (NIH) - USA; NIH National Institute of
   Neurological Disorders & Stroke (NINDS)
RP Eisenhofer, G (通讯作者)，Natl Inst Neurol Disorders & Stroke, Clin Neurocardiol Sect, NIH, Bethesda, MD USA.
RI Eisenhofer, Graeme/AAU-9829-2021
OI Eisenhofer, Graeme/0000-0002-8601-9903
CR ARMSTRONG MD, 1957, BIOCHIM BIOPHYS ACTA, V25, P422, DOI 10.1016/0006-3002(57)90491-2
   ARMSTRONG MD, 1958, PHARMACOL REV, V11, P394
   ATLAS SA, 2001, MASSRY GLASSOCKS TXB, P1169
   AXELROD J, 1957, SCIENCE, V126, P400, DOI 10.1126/science.126.3270.400
   Blaschko H, 1937, J PHYSIOL-LONDON, V90, P1
   BLOMBERY PA, 1980, ARCH GEN PSYCHIAT, V37, P1095
   BOULOUX PMG, 1998, CLIN ENDOCRINOL, P497
   BRAVO EL, 1979, NEW ENGL J MED, V301, P682, DOI 10.1056/NEJM197909273011302
   BRAVO EL, 1984, NEW ENGL J MED, V311, P1298, DOI 10.1056/NEJM198411153112007
   Bravo EL, 2002, CLEV CLIN J MED, V69, P257, DOI 10.3949/ccjm.69.3.257
   BRAVO EL, 2001, ENDOCRINOL METAB, P337
   BREESE GR, 1969, J PHARMACOL EXP THER, V165, P9
   BROWN MJ, 1984, EUR J CLIN INVEST, V14, P67, DOI 10.1111/j.1365-2362.1984.tb00706.x
   CROUT JR, 1964, J CLIN INVEST, V43, P94, DOI 10.1172/JCI104898
   CROUT JR, 1961, P SOC EXP BIOL MED, V108, P482
   CROUT JR, 1961, AM HEART J, V61, P375, DOI 10.1016/0002-8703(61)90609-3
   CRYER PE, 2001, ENDOCRINOL METAB, P525
   CUCHE JL, 1990, J LAB CLIN MED, V116, P681
   DEQUATTRO V, 1980, CLIN SCI, V59, pS275, DOI 10.1042/cs059275s
   Eisenhofer G, 1999, CLIN EXP PHARMACOL P, V26, pS41
   EISENHOFER G, 1995, CLIN SCI, V88, P533, DOI 10.1042/cs0880533
   Eisenhofer G, 1997, J CLIN ENDOCR METAB, V82, P3864, DOI 10.1210/jc.82.11.3864
   Eisenhofer G, 1996, N-S ARCH PHARMACOL, V354, P287
   Eisenhofer G, 1996, CIRCULATION, V93, P1667, DOI 10.1161/01.CIR.93.9.1667
   EISENHOFER G, 1995, J CLIN ENDOCR METAB, V80, P3009, DOI 10.1210/jc.80.10.3009
   EISENHOFER G, 1987, CLIN SCI, V73, P123, DOI 10.1042/cs0730123
   Eisenhofer G, 2003, J CLIN ENDOCR METAB, V88, P2656, DOI 10.1210/jc.2002-030005
   Eisenhofer G, 2003, J CLIN ENDOCR METAB, V88, P550, DOI 10.1210/jc.2002-021913
   Eisenhofer G, 1999, NEW ENGL J MED, V340, P1872, DOI 10.1056/NEJM199906173402404
   Eisenhofer G, 1998, J CLIN ENDOCR METAB, V83, P2175, DOI 10.1210/jc.83.6.2175
   Eisenhofer G, 2001, Rev Endocr Metab Disord, V2, P297, DOI 10.1023/A:1011572617314
   Eisenhofer G, 2001, CLIN CHEM, V47, P988
   Eisenhofer G, 2001, PHARMACOL THERAPEUT, V91, P35, DOI 10.1016/S0163-7258(01)00144-9
   Eisenhofer G, 1996, J NEUROCHEM, V66, P1565
   EISENHOFER G, 1992, AM J PHYSIOL, V268, pG641
   Ellingson T, 1999, J CHROMATOGR B, V729, P347, DOI 10.1016/S0378-4347(99)00125-5
   ERIKSSON BM, 1987, J CHROMATOGR, V386, P1
   GERLO EAM, 1994, CLIN CHEM, V40, P250
   GITLOW SE, 1960, J CLIN INVEST, V39, P221, DOI 10.1172/JCI104022
   GOLDENBERG M, 1954, AM J MED, V16, P310, DOI 10.1016/0002-9343(54)90349-6
   Goldstein DS, 2003, J PHARMACOL EXP THER, V305, P800, DOI 10.1124/jpet.103.049270
   Goldstein DS, 1999, J CLIN ENDOCR METAB, V84, P2523, DOI 10.1210/jc.84.7.2523
   HERTTING G, 1961, NATURE, V189, P66, DOI 10.1038/189066a0
   IVERSEN LL, 1965, BRIT J PHARM CHEMOTH, V25, P18, DOI 10.1111/j.1476-5381.1965.tb01753.x
   Kawamura M, 2002, AUTON NEUROSCI-BASIC, V96, P131, DOI 10.1016/S1566-0702(01)00385-X
   Kawamura M, 1997, J AUTONOM NERV SYST, V66, P145, DOI 10.1016/S0165-1838(97)00086-6
   KEISER HR, 2001, ENDOCRINOLOGY, P1862
   KOBAYASHI K, 1980, CLIN CHIM ACTA, V107, P163, DOI 10.1016/0009-8981(80)90444-1
   KOCH CA, 2003, IN PRESS ENDOCR PRAC
   KOPIN IJ, 1960, NATURE, V185, P788, DOI 10.1038/185788a0
   KOPIN IJ, 1964, PHARMACOL REV, V16, P179
   KOPIN IJ, 1962, J PHARMACOL EXP THER, V138, P351
   LABROSSE EH, 1958, SCIENCE, V128, P593, DOI 10.1126/science.128.3324.593
   LABROSSE EH, 1970, J CLIN ENDOCR METAB, V30, P580, DOI 10.1210/jcem-30-5-580
   LABROSSE EH, 1976, JNCI-J NATL CANCER I, V57, P633, DOI 10.1093/jnci/57.3.633
   LABROSSE EH, 1961, J CLIN INVEST, V40, P253, DOI 10.1172/JCI104251
   Lagerstedt SA, 2003, CLIN CHEM, V49, pA43
   LANDSBERG L, 2001, HARRISONS PRINCIPLES, P439
   LANDSBERG L, 2001, HARRISONS PRINCIPLES, P2105
   LENDERS JWM, 1995, ANN INTERN MED, V123, P101, DOI 10.7326/0003-4819-123-2-199507150-00004
   LENDERS JWM, 1993, CLIN CHEM, V39, P97
   Lenders JWM, 2002, JAMA-J AM MED ASSOC, V287, P1427, DOI 10.1001/jama.287.11.1427
   Lenz Tomas, 2002, Clin Lab, V48, P5
   LITCHFIELD JW, 1956, LANCET, V271, P1283
   MAAS JW, 1970, J PHARMACOL EXP THER, V174, P381
   Mannelli M, 1999, EUR J ENDOCRINOL, V141, P619, DOI 10.1530/eje.0.1410619
   Mantero F, 2000, J CLIN ENDOCR METAB, V85, P637, DOI 10.1210/jc.85.2.637
   MARDH G, 1985, P NATL ACAD SCI USA, V82, P4979, DOI 10.1073/pnas.82.15.4979
   MARDH G, 1981, J NEUROCHEM, V36, P1181, DOI 10.1111/j.1471-4159.1981.tb01716.x
   MARDH G, 1984, J NEUROCHEM, V42, P43, DOI 10.1111/j.1471-4159.1984.tb09695.x
   MORNEX R, 1991, HORM RES, V36, P220, DOI 10.1159/000182167
   OCONNOR DT, 2000, ADRENAL MEDULLA CARE, P1257
   Pacak K, 2001, J CLIN ENDOCR METAB, V86, P3641, DOI 10.1210/jc.86.8.3641
   PEASTON RT, 1993, J CLIN PATHOL, V46, P734, DOI 10.1136/jcp.46.8.734
   PISANO JJ, 1960, CLIN CHIM ACTA, V5, P406
   Raber W, 2000, ARCH INTERN MED, V160, P2957, DOI 10.1001/archinte.160.19.2957
   Raber W, 2000, ENDOCRINOLOGIST, V10, P65, DOI 10.1097/00019616-200010010-00010
   Richter D, 1940, J PHYSIOL-LONDON, V98, P361, DOI 10.1113/jphysiol.1940.sp003854
   Roden M, 2001, CLIN CHEM, V47, P1061
   ROTH JA, 1992, REV PHYSIOL BIOCH P, V120, P1, DOI 10.1007/BFb0036121
   Rubin GL, 1996, XENOBIOTICA, V26, P1113, DOI 10.3109/00498259609050256
   Sawka AM, 2003, J CLIN ENDOCR METAB, V88, P553, DOI 10.1210/jc.2002-021251
   Shawar L, 1996, J La State Med Soc, V148, P535
   SINCLAIR D, 1991, ANN CLIN BIOCHEM, V28, P417, DOI 10.1177/000456329102800420
   SJOERDSMA A, 1959, J CLIN INVEST, V38, P31, DOI 10.1172/JCI103793
   SJOERDSMA A, 1958, SCIENCE, V127, P876, DOI 10.1126/science.127.3303.876
   STEWART MF, 1993, J CLIN PATHOL, V46, P280, DOI 10.1136/jcp.46.3.280
   TABAKOFF B, 1973, MOL PHARMACOL, V9, P428
   Taylor RL, 2002, CLIN CHEM, V48, P533
   VANHEERDEN JA, 1982, SURGERY, V91, P367
   Walther MM, 1999, J UROLOGY, V162, P659, DOI 10.1097/00005392-199909010-00004
   Young William F. Jr., 1997, Comprehensive Therapy, V23, P319
NR 92
TC 43
Z9 46
U1 2
U2 20
PU HUMANA PRESS INC
PI TOTOWA
PA 999 RIVERVIEW DRIVE SUITE 208, TOTOWA, NJ 07512 USA
SN 1046-3976
EI 1559-0097
J9 ENDOCR PATHOL
JI Endocr. Pathol.
PD FAL
PY 2003
VL 14
IS 3
BP 193
EP 212
DI 10.1385/EP:14:3:193
PG 20
WC Endocrinology & Metabolism; Pathology
WE Science Citation Index Expanded (SCI-EXPANDED)
SC Endocrinology & Metabolism; Pathology
GA 738YD
UT WOS:000186314700002
PM 14586065
DA 2023-08-27
ER

PT J
AU Romagnani, P
   Annunziato, F
   Manetti, R
   Mavilia, C
   Lasagni, L
   Manuelli, C
   Vannelli, GB
   Vanini, V
   Maggi, E
   Pupilli, C
   Romagnani, S
AF Romagnani, P
   Annunziato, F
   Manetti, R
   Mavilia, C
   Lasagni, L
   Manuelli, C
   Vannelli, GB
   Vanini, V
   Maggi, E
   Pupilli, C
   Romagnani, S
TI High CD30 ligand expression by epithelial cells and Hassal's corpuscles
   in the medulla of human thymus
SO BLOOD
LA English
DT Article
ID T-CELLS; RECEPTOR EXPRESSION; CYTOKINE PRODUCTION; NEGATIVE SELECTION;
   UP-REGULATION; ANTIGEN; APOPTOSIS; LYMPHOMA; DEFINES; FAMILY
AB CD30 is a member of tumor necrosis factor (TNF) receptor superfamily that is expressed by activated T cells in the presence of interleukin-4 (IL-4). Although CD30 can mediate a variety of signals, CD30-deficient mice have impaired negative selection of T cells, suggesting that at least in the context of murine thymus, CD30 is a cell death-mediating molecule. The ligand for CD30 (CD30L) is a membrane-associated glycoprotein related to TNF, which is known to be expressed mainly by activated T cells and other leukocytes. However, the nature of CD30L-expressing cells involved in the interaction with CD30+ thymocytes is unclear. We report here that in postnatal human thymus the great majority of CD30+ cells are double positive (CD4+CD8+), activated, IL-4 receptor-expressing T cells which selectively localize in the medullary areas. Moreover, many medullary epithelial cells and Hassal's corpuscles in the same thymus specimens showed unusually high expression of CD30L in comparison with other lymphoid or nonlymphoid tissues. These findings provide additional information on the nature and localization of CD30+ thymocytes and show that epithelial cells are the major holder of CD30L in the thymic medulla. (C) 1998 by The American Society of Hematology.
C1 Univ Florence, Ist Med Interna & Immunoalllergol, I-50134 Florence, Italy.
   Univ Florence, Dept Clin Physiopathol, Endocrinol Unit, I-50134 Florence, Italy.
   Univ Florence, Dept Anat, I-50134 Florence, Italy.
   Apuano Pediat Hosp, Massa Carrara, Italy.
C3 University of Florence; University of Florence; University of Florence
RP Romagnani, S (通讯作者)，Univ Florence, Ist Med Interna & Immunoalllergol, Viale Morgagni 85, I-50134 Florence, Italy.
RI Lasagni, Laura/J-6856-2018; Annunziato, Francesco/C-1155-2013;
   Romagnani, Paola/A-4309-2008; Maggi, Enrico/AAA-8045-2019
OI Lasagni, Laura/0000-0001-6553-9160; Annunziato,
   Francesco/0000-0001-8798-7589; Romagnani, Paola/0000-0002-1774-8088;
   Maggi, Enrico/0000-0002-1824-3583
CR Amakawa R, 1996, CELL, V84, P551, DOI 10.1016/S0092-8674(00)81031-4
   Annunziato F, 1997, EUR J IMMUNOL, V27, P2239, DOI 10.1002/eji.1830270918
   Bowen MA, 1996, J IMMUNOL, V156, P442
   Chilosi M, 1996, EUR J IMMUNOL, V26, P329, DOI 10.1002/eji.1830260209
   DElios MM, 1997, J LEUKOCYTE BIOL, V61, P539, DOI 10.1002/jlb.61.5.539
   DELPRETE G, 1995, FASEB J, V9, P81, DOI 10.1096/fasebj.9.1.7821763
   FERNANDEZ E, 1994, BLOOD, V83, P3245
   GAVRIELI Y, 1992, J CELL BIOL, V119, P493, DOI 10.1083/jcb.119.3.493
   Gruss HJ, 1996, AM J PATHOL, V149, P469
   GRUSS HJ, 1994, BLOOD, V83, P2045
   HAYNES BF, 1984, J EXP MED, V159, P1149, DOI 10.1084/jem.159.4.1149
   MANETTI R, 1994, J EXP MED, V180, P2407, DOI 10.1084/jem.180.6.2407
   MILTENYI S, 1990, CYTOMETRY, V11, P231, DOI 10.1002/cyto.990110203
   MURPHY KM, 1990, SCIENCE, V250, P1720, DOI 10.1126/science.2125367
   Nakamura T, 1997, J IMMUNOL, V158, P1085
   Nakamura T, 1997, J IMMUNOL, V158, P2090
   OHARA J, 1988, P NATL ACAD SCI USA, V85, P8221, DOI 10.1073/pnas.85.21.8221
   Parronchi P, 1997, AM J PATHOL, V150, P823
   PUPILLI C, 1994, AM J PHYSIOL-RENAL, V267, pF679, DOI 10.1152/ajprenal.1994.267.4.F679
   RENZ H, 1991, J IMMUNOL, V146, P3049
   SANGER F, 1977, P NATL ACAD SCI USA, V74, P5463, DOI 10.1073/pnas.74.12.5463
   SCHWAB U, 1982, NATURE, V299, P65, DOI 10.1038/299065a0
   SMITH CA, 1993, CELL, V73, P1349, DOI 10.1016/0092-8674(93)90361-S
   SMITH CA, 1990, SCIENCE, V248, P1019, DOI 10.1126/science.2160731
   SPITS H, 1995, BLOOD, V85, P2654, DOI 10.1182/blood.V85.10.2654.bloodjournal85102654
   STEIN H, 1985, BLOOD, V66, P848
   SURH CD, 1994, NATURE, V372, P100, DOI 10.1038/372100a0
   VANDET NF, 1995, BIOCHEM J, V307, P759, DOI 10.1042/bj3070759
   Wiley SR, 1996, J IMMUNOL, V157, P3635
   Younes A, 1996, BRIT J HAEMATOL, V93, P569, DOI 10.1046/j.1365-2141.1996.d01-1686.x
NR 30
TC 75
Z9 78
U1 0
U2 0
PU W B SAUNDERS CO
PI PHILADELPHIA
PA INDEPENDENCE SQUARE WEST CURTIS CENTER, STE 300, PHILADELPHIA, PA
   19106-3399 USA
SN 0006-4971
J9 BLOOD
JI Blood
PD MAY 1
PY 1998
VL 91
IS 9
BP 3323
EP 3332
DI 10.1182/blood.V91.9.3323.3323_3323_3332
PG 10
WC Hematology
WE Science Citation Index Expanded (SCI-EXPANDED)
SC Hematology
GA ZJ995
UT WOS:000073274500028
PM 9558389
OA Bronze
DA 2023-08-27
ER

PT J
AU James, PE
   Bacic, G
   Grinberg, OY
   Goda, F
   Dunn, JF
   Jackson, SK
   Swartz, HM
AF James, PE
   Bacic, G
   Grinberg, OY
   Goda, F
   Dunn, JF
   Jackson, SK
   Swartz, HM
TI Endotoxin-induced changes in intrarenal PO2, measured by in vivo
   electron paramagnetic resonance oximetry and magnetic resonance imaging
SO FREE RADICAL BIOLOGY AND MEDICINE
LA English
DT Article
DE renal tissue oxygen; renal medulla; free radicals
ID PERFUSED RAT-KIDNEY; BLOOD OXYGENATION; RENAL ISCHEMIA; NITRIC-OXIDE;
   SHOCK; PATHOPHYSIOLOGY; SPECTROSCOPY; FAILURE; SYSTEM; TUMOR
AB Electron Paramagnetic Resonance (EPR) oximetry was used to measure tissue oxygen tension (pO(2)-partial pressure of oxygen) simultaneously in the kidney cortex and outer medulla in vivo in mice. pO(2) in the cortex region was higher compared to that in the outer medulla. An intravenous injection of endotoxin resulted in a sharp drop in pO(2) in the cortex and an increase in the medulla region, resulting in a transient period of equal pO? in both regions. In control kidneys, functional Magnetic Resonance (MR) images showed the cortex region to have high signal intensity (T-2*-weighted images), indicating that this region was well supplied with oxygenated hemoglobin, whereas the outer medulla showed low signal intensity. After administration of endotoxin?rin, we observed an immediate increase in signal intensity in the outer medulla region, reflecting an increased level of oxygenated blood in this region. Pretreatment of mice with N-G-monomethyl-L-arginine prevented both the changes in tissue pO(2) and distribution of oxygenated hemoglobin, suggesting that localized production of nitric oxide has a critical role to play in renal medullary hemodynamics. In combining in vivo EPR with MR images of kidneys, we demonstrate the usefulness of these techniques for monitoring renal pO(2) and changes in the distribution of oxygen.
C1 DARTMOUTH COLL, SCH MED, ELECTRON PARAMAGNET RESONANCE CTR, RADIOL DEPT HB 7252, HANOVER, NH 03755 USA.
   UNIV WALES COLL MED, CARDIFF CF4 4XN, S GLAM, WALES.
   DARTMOUTH COLL, SCH MED, DEPT RADIOL, NUCL MAGNET RESONANCE CTR, HANOVER, NH 03755 USA.
C3 Dartmouth College; Cardiff University; Dartmouth College
OI Bacic, Goran/0000-0003-0235-0170
FU NIGMS NIH HHS [GM-34250] Funding Source: Medline
CR ANTONSSON JB, 1991, ACTA CHIR-EUR J SURG, V157, P3
   ARCHIE JP, 1977, ANN SURG, V186, P171, DOI 10.1097/00000658-197708000-00008
   ATKINS JL, 1991, AM J PHYSIOL, V261, pF849, DOI 10.1152/ajprenal.1991.261.5.F849
   BAUMGARTL H, 1972, MICROVASC RES, V4, P247, DOI 10.1016/0026-2862(72)90036-2
   BREZIS M, 1991, J CLIN INVEST, V88, P390, DOI 10.1172/JCI115316
   BREZIS M, 1994, AM J PHYSIOL-RENAL, V267, pF1063, DOI 10.1152/ajprenal.1994.267.6.F1063
   CARRIERE S, 1966, CIRC RES, V19, P167, DOI 10.1161/01.RES.19.1.167
   CAVANAGH D, 1970, AM J OBSTET GYNECOL, V108, P705, DOI 10.1016/0002-9378(70)90535-1
   COHEN JJ, 1990, KIDNEY INT, V37, P1219, DOI 10.1038/ki.1990.105
   De Navasquez S, 1938, J PATHOL BACTERIOL, V46, P47, DOI 10.1002/path.1700460106
   DINARELLO CA, 1989, ADV IMMUNOL, V44, P153, DOI 10.1016/S0065-2776(08)60642-2
   EPSTEIN FH, 1982, AM J PHYSIOL, V243, pF356, DOI 10.1152/ajprenal.1982.243.4.F356
   FREY L, 1990, EUR SURG RES S1, V22, P66
   GLOCKNER JF, 1992, OXYGEN TRANSPORT TIS, V14, P221
   GREEN LC, 1982, ANAL BIOCHEM, V126, P131, DOI 10.1016/0003-2697(82)90118-X
   GULLICHSEN E, 1991, ACTA CHIR-EUR J SURG, P7
   HANSELL P, 1992, RENAL PHYSIOL BIOCH, V15, P217
   HINSHAW LB, 1961, AM J PHYSIOL, V201, P144, DOI 10.1152/ajplegacy.1961.201.1.144
   HOPPEL BE, 1993, MAGN RESON MED, V30, P715, DOI 10.1002/mrm.1910300609
   JAATTELA M, 1991, LAB INVEST, V64, P724
   JAMES PE, 1995, FREE RADICAL BIO MED, V18, P641, DOI 10.1016/0891-5849(94)00179-N
   LEICHTWEISS HP, 1969, PFLUG ARCH EUR J PHY, V309, P328, DOI 10.1007/BF00587756
   LOHR J, 1991, MAGN RESON IMAGING, V9, P93, DOI 10.1016/0730-725X(91)90102-R
   LUI KJ, 1993, P NATL ACAD SCI USA, V90, P5438
   MAROTTI M, 1987, MAGN RESON MED, V5, P160, DOI 10.1002/mrm.1910050207
   MATTSON DL, 1994, AM J PHYSIOL, V266, pH1918, DOI 10.1152/ajpheart.1994.266.5.H1918
   NAGRANI NK, 1994, 1019 SMRM
   Nilges M.J., 1989, PHYS MEDICA, V5, P195
   OGAWA S, 1990, MAGNET RESON MED, V14, P68, DOI 10.1002/mrm.1910140108
   OHAIR DP, 1989, CIRC SHOCK, V27, P199
   PALLONE TL, 1990, PHYSIOL REV, V70, P885, DOI 10.1152/physrev.1990.70.3.885
   PASSMORE JC, 1973, J TRAUMA, V13, P1066, DOI 10.1097/00005373-197312000-00005
   POHL U, 1990, EXPERIENTIA, V46, P1175, DOI 10.1007/BF01936931
   RAMSAY G, 1989, UPDATE INTENSIVE CAR, P368
   REIDY MA, 1983, LAB INVEST, V48, P25
   SMIRNOV AI, 1993, MAGNET RESON MED, V30, P213, DOI 10.1002/mrm.1910300210
   SWARTZ HM, 1995, OXYGEN TRANSPORT TIS, V16, P119
   TERRIER F, 1989, MAGNET RESON MED, V12, P114, DOI 10.1002/mrm.1910120114
   TUREK P, 1987, CHEM PHYS LETT, V134, P471, DOI 10.1016/0009-2614(87)87175-0
   VERDON CP, 1995, ANAL BIOCHEM, V224, P502, DOI 10.1006/abio.1995.1079
   VEXLER VS, 1993, JMRI-J MAGN RESON IM, V3, P483, DOI 10.1002/jmri.1880030310
   VINCENT JL, 1989, CRIT CARE CLIN, V5, P369
   WANG QZ, 1991, LIFE SCI, V49, pPL55, DOI 10.1016/0024-3205(91)90251-6
   WARDLE EN, 1975, NEPHRON, V14, P321
   WESTENBERGER U, 1990, FREE RADICAL RES COM, V11, P167, DOI 10.3109/10715769009109680
   WILSON DF, 1977, AM J PHYSIOL, V233, pC135, DOI 10.1152/ajpcell.1977.233.5.C135
   XIA YY, 1993, AM J PHYSIOL, V264, pF774, DOI 10.1152/ajprenal.1993.264.5.F774
NR 47
TC 58
Z9 58
U1 0
U2 4
PU ELSEVIER SCIENCE INC
PI NEW YORK
PA 360 PARK AVE SOUTH, NEW YORK, NY 10010-1710 USA
SN 0891-5849
EI 1873-4596
J9 FREE RADICAL BIO MED
JI Free Radic. Biol. Med.
PY 1996
VL 21
IS 1
BP 25
EP 34
DI 10.1016/0891-5849(95)02221-X
PG 10
WC Biochemistry & Molecular Biology; Endocrinology & Metabolism
WE Science Citation Index Expanded (SCI-EXPANDED)
SC Biochemistry & Molecular Biology; Endocrinology & Metabolism
GA UT321
UT WOS:A1996UT32100005
PM 8791090
OA hybrid
DA 2023-08-27
ER

PT J
AU Zaghloul, MS
   Nasr, A
   Tolba, M
   Refaat, A
   Youssef, A
   Mosaab, A
   Enayet, A
   Arafa, O
   Maher, E
   Eldebawy, E
AF Zaghloul, Mohamed S.
   Nasr, Azza
   Tolba, Marwan
   Refaat, Amal
   Youssef, Ayda
   Mosaab, Amal
   Enayet, Abdelrahman
   Arafa, Omar
   Maher, Eslam
   Eldebawy, Eman
TI Hypofractionated Radiation Therapy For Diffuse Intrinsic Pontine Glioma:
   A Noninferiority Randomized Study Including 253 Children
SO INTERNATIONAL JOURNAL OF RADIATION ONCOLOGY BIOLOGY PHYSICS
LA English
DT Article
ID RADIOTHERAPY; SUBGROUPS; MUTATION; TUMORS
AB Purpose: Pediatric diffuse intrinsic pontine glioma is an orphen disease. This study aimed to confirm the noninferiority of hypofractionated (HF) radiation therapy. Identification of the prognostic factors that determine the overall survival (OS) and progression-free survival (PFS) was the secondary objective.
   Methods and Materials: We randomized 253 patients into 3 arms of radiation therapy regimens: HF1, receiving 39 Gy in 13 fractions; HF2, receiving 45 Gy in 15 fractions; and conventional fractionation (CF), receiving 54 Gy in 30 fractions. The OS and PFS were calculated using Kaplan-Meier methods, and the noninferiority was estimated against the CF arm.
   Results: The median OS for the HF1, HF2, and CF were 9.6, 8.2, and 8.7 months, respectively. The 1-, 1.5-, and 2-year OS were 34.6%, 17.9%, and 10.7% for HF1; 26.2%, 13.1%, and 4.8% for HF2; and 25.3%, 12.1%, and 8.4% for CF, respectively (P = .3). The hazard ratio was 0.776 and 1.124 for HF1 and HF2, respectively. Considering the noninferiority margin (Delta) of 15% and a power of 90%, the lower inferiority confidence interval for HF1 was -14.34% and for HF2 it was 11.37% (both below Delta), confirming its noninferiority at 18-months OS. Younger patients (2-5 years of age) had better median OS in the whole cohort (11.6 months), HF1 (13.5), and CF (12.1) but not HF2 (6.2) (P = .003). Furthermore, the OS rates at 1, 1.5, and 2 years for children 2 to 5 years of age in the HF2 arm were lower than those in the HF1 and CF arms. However, similar acute and late side effects were reported in the 3 arms.
   Conclusions: Two hypofractionated radiation therapy proved to be noninferior to conventional fractionation. Young age (2-5 years) is the only prognostic factor determining both OS and PFS. The young age superiority was lost with a higher hypofractionated radiation therapy dose, necessitating more caution in applying 45 Gy in 15 fractions in younger children (2-5 years of age). (C) 2022 Elsevier Inc. All rights reserved.
C1 [Zaghloul, Mohamed S.; Nasr, Azza; Eldebawy, Eman] Cairo Univ, Natl Canc Inst, Radiat Oncol Dept, Giza, Egypt.
   [Zaghloul, Mohamed S.; Nasr, Azza; Refaat, Amal; Youssef, Ayda; Mosaab, Amal; Enayet, Abdelrahman; Arafa, Omar; Maher, Eslam; Eldebawy, Eman] Egypt tMcGill Univ, Childrens Canc Hosp, Montreal, PQ, Canada.
   [Tolba, Marwan] McGill Univ, Montreal, PQ, Canada.
   [Refaat, Amal; Youssef, Ayda] Cairo Univ, Natl Canc Inst, Radiodiag Dept, Cairo, Egypt.
   [Enayet, Abdelrahman] Cairo Univ, Fac Med, Neurosurg Dept, Cairo, Egypt.
   [Arafa, Omar] Cairo Univ, Natl Canc Inst, Pediat Oncol Dept, Giza, Egypt.
C3 Egyptian Knowledge Bank (EKB); Cairo University; National Cancer
   Institute - Egypt; McGill University; Egyptian Knowledge Bank (EKB);
   Cairo University; National Cancer Institute - Egypt; Egyptian Knowledge
   Bank (EKB); Cairo University; Egyptian Knowledge Bank (EKB); Cairo
   University; National Cancer Institute - Egypt
RP Zaghloul, MS (通讯作者)，Cairo Univ, Natl Canc Inst, Radiat Oncol Dept, Giza, Egypt.; Zaghloul, MS (通讯作者)，Egypt tMcGill Univ, Childrens Canc Hosp, Montreal, PQ, Canada.
EM
RI TOLBA, MARWAN/GLN-7804-2022
OI TOLBA, MARWAN/0000-0003-2812-7348; Maher, Eslam/0000-0001-5428-1555
CR [Anonymous], 2007, ACTA NEUROPATHOL, DOI DOI 10.1007/s00401-007-0243-4
   Castel D, 2015, ACTA NEUROPATHOL, V130, P815, DOI 10.1007/s00401-015-1478-0
   Cavalheiro S, 2015, CHILD NERV SYST, V31, P1815, DOI 10.1007/s00381-015-2799-y
   Cooney T, 2017, NEURO-ONCOLOGY, V19, P1279, DOI 10.1093/neuonc/nox107
   COX JD, 1995, INT J RADIAT ONCOL, V31, P1341, DOI 10.1016/0360-3016(95)00060-C
   Khuong-Quang DA, 2012, ACTA NEUROPATHOL, V124, P439, DOI 10.1007/s00401-012-0998-0
   Freeman CR, 1998, INT J RADIAT ONCOL, V40, P265, DOI 10.1016/S0360-3016(97)00572-5
   Freitag G, 2005, BIOMETRICAL J, V47, P88, DOI 10.1002/bimj.200410083
   Grasso CS, 2015, NAT MED, V21, P555, DOI 10.1038/nm.3855
   HaasKogan DA, 1996, INT J RADIAT ONCOL, V36, P95, DOI 10.1016/S0360-3016(96)00244-1
   Indelicato DJ, 2014, ACTA ONCOL, V53, P1298, DOI 10.3109/0284186X.2014.957414
   Izzuddeen Y, 2020, J NEURO-ONCOL, V146, P91, DOI 10.1007/s11060-019-03340-7
   Jansen MH, 2015, NEURO-ONCOLOGY, V17, P160, DOI 10.1093/neuonc/nou104
   Janssens GORJ, 2009, INT J RADIAT ONCOL, V73, P722, DOI 10.1016/j.ijrobp.2008.05.030
   LANGMOEN IA, 1991, CHILD NERV SYST, V7, P13, DOI 10.1007/BF00263825
   Lewis PW, 2013, SCIENCE, V340, P857, DOI 10.1126/science.1232245
   Machin D, 2000, STAT CONFIDENCE, P93
   Negretti L, 2011, J NEURO-ONCOL, V104, P773, DOI 10.1007/s11060-011-0542-4
   Park J, 2020, MEDICINE, V99, DOI 10.1097/MD.0000000000022721
   Rashed WM, 2019, CANCER METAST REV, V38, P759, DOI 10.1007/s10555-019-09824-2
   Robison NJ, 2014, J NEURO-ONCOL, V119, P7, DOI 10.1007/s11060-014-1448-8
   Vitanza NA, 2019, CURR TREAT OPTION NE, V21, DOI 10.1007/s11940-019-0577-y
   Wu G, 2014, NAT GENET, V46, P444, DOI 10.1038/ng.2938
   Zaghloul MS, 2014, RADIOTHER ONCOL, V111, P35, DOI 10.1016/j.radonc.2014.01.013
   Zaghloul MS, 2015, CHILD NERV SYST, V31, P1221, DOI 10.1007/s00381-015-2678-6
   Zaghloul MS, 2016, PEDIAT BLOOD TER CAN, V36, P948
   Zaghloul MS, 2019, CLIN ONCOL-UK, V22, P850
NR 27
TC 4
Z9 4
U1 0
U2 2
PU ELSEVIER SCIENCE INC
PI NEW YORK
PA STE 800, 230 PARK AVE, NEW YORK, NY 10169 USA
SN 0360-3016
EI 1879-355X
J9 INT J RADIAT ONCOL
JI Int. J. Radiat. Oncol. Biol. Phys.
PD JUN 1
PY 2022
VL 113
IS 2
BP 360
EP 368
DI 10.1016/j.ijrobp.2022.01.054
EA MAY 2022
PG 9
WC Oncology; Radiology, Nuclear Medicine & Medical Imaging
WE Science Citation Index Expanded (SCI-EXPANDED)
SC Oncology; Radiology, Nuclear Medicine & Medical Imaging
GA 1Y4TE
UT WOS:000808134300020
PM 35150788
DA 2023-08-27
ER

PT J
AU Baliga, S
   Yock, TI
AF Baliga, Sujith
   Yock, Torunn I.
TI Proton beam therapy in pediatric oncology
SO CURRENT OPINION IN PEDIATRICS
LA English
DT Review
DE brainstem toxicity; endocrinopathy; protons; quality of life
ID MODULATED RADIATION-THERAPY; QUALITY-OF-LIFE; LOW-GRADE GLIOMAS;
   PHASE-II TRIAL; ADJUVANT CHEMOTHERAPY; BRAIN-STEM; CRANIOSPINAL
   IRRADIATION; CHILDHOOD-CANCER; ADULT SURVIVORS; RADIOTHERAPY
AB Purpose of review
   The advent of proton beam therapy (PBT) has initiated a paradigm shift in the field of pediatric radiation oncology, with increasing promise to alleviate both short-term and long-term toxicities. Given the dramatic rise in proton therapy centers in the United States, a discussion of the quality of evidence supporting its use in pediatric cancers is warranted.
   Recent findings
   Proton radiotherapy appears to decrease the incidence and severity of late effects with the strongest evidence in pediatric brain tumor cohorts that shows benefits in neurocognitive, hearing, and endocrine outcomes. However, emerging data has shown that more conservative brainstem dose limits with protons compared with photons are required to limit brainstem toxicity; these modified recommendations have been incorporated into national cooperative group studies. Decreased toxicity in tumors outside of the CNS for PBT have also been reported in sarcomas, Hodgkin disease and neuroblastoma. Similarly, QoL outcomes are improved in brain tumor and other cohorts of patients treated with PBT.
   Summary
   The collective findings demonstrate improved understanding and refinement of PBT in pediatric cancers. Data on QOL, toxicity and disease outcomes with PBT should continue to be collected and reported in order to understand the full extent of the risks and benefits associated with PBT.
C1 [Baliga, Sujith; Yock, Torunn I.] Harvard Med Sch, Massachusetts Gen Hosp, Dept Radiat Oncol, Boston, MA 02115 USA.
C3 Harvard University; Harvard Medical School; Massachusetts General
   Hospital
RP Yock, TI (通讯作者)，Harvard Med Sch, Massachusetts Gen Hosp, Dept Radiat Oncol, Serv Pediat, 55 Fruit St, Boston, MA 02114 USA.; Yock, TI (通讯作者)，Harvard Med Sch, Massachusetts Gen Hosp, 55 Fruit St, Boston, MA 02114 USA.
EM
CR [Anonymous], PEDIAT BLOOD CANC
   [Anonymous], 2017, SEER CANC STAT REV
   [Anonymous], AM SOC CLIN ONCOL ED
   [Anonymous], 2014, INT J PART THER
   [Anonymous], PEDIAT BLOOD CANC
   [Anonymous], J NATL CANC I
   [Anonymous], PEDIAT BLOOD CANC
   [Anonymous], J CLIN ONCOL
   [Anonymous], PEDIAT BLOOD CANC
   [Anonymous], PROTON CRANIOSPINAL
   Armstrong GT, 2013, JNCI-J NATL CANCER I, V105, P899, DOI 10.1093/jnci/djt089
   Armstrong GT, 2010, EUR J PAEDIATR NEURO, V14, P298, DOI 10.1016/j.ejpn.2009.12.006
   Bhakta N, 2017, LANCET, V390, P2569, DOI 10.1016/S0140-6736(17)31610-0
   Blomstrand M, 2012, NEURO-ONCOLOGY, V14, P882, DOI 10.1093/neuonc/nos120
   Brodin NP, 2014, NEURO-ONCOLOGY, V16, P594, DOI 10.1093/neuonc/not225
   Cambruzzi E, 2018, CHILD NERV SYST, V34, P809, DOI 10.1007/s00381-018-3765-2
   Casey DL, 2018, INT J RADIAT ONCOL, V100, P1204, DOI 10.1016/j.ijrobp.2018.01.008
   CASTLEBERRY RP, 1991, J CLIN ONCOL, V9, P789, DOI 10.1200/JCO.1991.9.5.789
   Ciernik IF, 2011, CANCER-AM CANCER SOC, V117, P4522, DOI 10.1002/cncr.26037
   Demizu Y, 2017, CANCER SCI, V108, P972, DOI 10.1111/cas.13192
   Eaton BR, 2016, NEURO-ONCOLOGY, V18, P881, DOI 10.1093/neuonc/nov302
   Edelstein K, 2011, NEURO-ONCOLOGY, V13, P536, DOI 10.1093/neuonc/nor015
   Frange P, 2009, J NEURO-ONCOL, V95, P271, DOI 10.1007/s11060-009-9927-z
   Fuji H, 2013, RADIAT ONCOL, V8, DOI 10.1186/1748-717X-8-255
   Gatcombe HG, 2009, INT J RADIAT ONCOL, V74, P1549, DOI 10.1016/j.ijrobp.2008.10.069
   Gentile MS, 2018, INT J RADIAT ONCOL, V100, P719, DOI 10.1016/j.ijrobp.2017.11.026
   Giantsoudi D, 2017, INT J RADIAT ONCOL, V98, P37, DOI 10.1016/j.ijrobp.2017.01.226
   Gnekow AK, 2012, NEURO-ONCOLOGY, V14, P1265, DOI 10.1093/neuonc/nos202
   Green DM, 1999, J CLIN ONCOL, V17, P3207, DOI 10.1200/JCO.1999.17.10.3207
   Greenberger BA, 2014, INT J RADIAT ONCOL, V89, P1060, DOI 10.1016/j.ijrobp.2014.04.053
   Harrabi S, 2016, STRAHLENTHER ONKOL, V192, P759, DOI 10.1007/s00066-016-1005-9
   Hattangadi JA, 2012, INT J RADIAT ONCOL, V83, P1015, DOI 10.1016/j.ijrobp.2011.08.035
   Hawkins DS, 2018, J CLIN ONCOL, V36, P2770, DOI 10.1200/JCO.2018.77.9694
   Hess CB, 2018, FRONT ONCOL, V8, DOI 10.3389/fonc.2018.00165
   Hoppe BS, 2017, ANN ONCOL, V28, P2179, DOI 10.1093/annonc/mdx287
   Indelicato DJ, 2014, ACTA ONCOL, V53, P1298, DOI 10.3109/0284186X.2014.957414
   Kahalley LS, 2016, J CLIN ONCOL, V34, P1043, DOI 10.1200/JCO.2015.62.1383
   Kuhlthau KA, 2012, J CLIN ONCOL, V30, P2079, DOI 10.1200/JCO.2011.37.0577
   Ladra MM, 2014, J CLIN ONCOL, V32, P3762, DOI 10.1200/JCO.2014.56.1548
   Leiser D, 2016, RADIOTHER ONCOL, V120, P163, DOI 10.1016/j.radonc.2016.05.013
   Lockney NA, 2016, PEDIATR BLOOD CANCER, V63, P1608, DOI 10.1002/pbc.26061
   Mabbott DJ, 2008, NEUROPSYCHOLOGY, V22, P159, DOI 10.1037/0894-4105.22.2.159
   MacEwan I, 2017, ADV RADIAT ONCOL, V2, P220, DOI 10.1016/j.adro.2017.03.001
   Merchant TE, 2009, J CLIN ONCOL, V27, P3691, DOI 10.1200/JCO.2008.21.2738
   Merchant TE, 2009, J CLIN ONCOL, V27, P3598, DOI 10.1200/JCO.2008.20.9494
   Miralbell R, 1997, INT J RADIAT ONCOL, V38, P477, DOI 10.1016/S0360-3016(97)00004-7
   Moxon-Emre I, 2014, J CLIN ONCOL, V32, P1760, DOI 10.1200/JCO.2013.52.3290
   Nanda R, 2017, INT J RADIAT ONCOL, V99, P494, DOI 10.1016/j.ijrobp.2017.04.001
   Odei B, 2017, INT J RADIAT ONCOL, V97, P60, DOI 10.1016/j.ijrobp.2016.09.011
   Oh KS, 2011, INT J RADIAT ONCOL, V81, pE481, DOI 10.1016/j.ijrobp.2011.01.019
   Oshiro Y, 2013, RADIAT ONCOL, V8, DOI 10.1186/1748-717X-8-142
   Ostrom QT, 2014, NEURO-ONCOLOGY, V16, P1, DOI [10.1093/neuonc/nov189, 10.1093/neuonc/nou327, 10.1093/neuonc/nou223]
   Packer RJ, 1999, J CLIN ONCOL, V17, P2127, DOI 10.1200/JCO.1999.17.7.2127
   Packer RJ, 2003, J CLIN ONCOL, V21, P3255, DOI 10.1200/JCO.2003.01.202
   Packer RJ, 2008, ARCH NEUROL-CHICAGO, V65, P1419, DOI 10.1001/archneur.65.11.1419
   Packer RJ, 2006, J CLIN ONCOL, V24, P4202, DOI 10.1200/JCO.2006.06.4980
   Palmer SL, 2008, DEV DISABIL RES REV, V14, P203, DOI 10.1002/ddrr.32
   Palmer SL, 2001, J CLIN ONCOL, V19, P2302, DOI 10.1200/JCO.2001.19.8.2302
   Paulino AC, 2004, INT J RADIAT ONCOL, V60, P265, DOI 10.1016/j.ijrobp.2004.02.001
   Paulino AC, 2018, RADIOTHER ONCOL, V128, P128, DOI 10.1016/j.radonc.2018.01.002
   Paulino AC, 2010, INT J RADIAT ONCOL, V78, P1445, DOI 10.1016/j.ijrobp.2009.09.031
   Pinnix CC, 2015, INT J RADIAT ONCOL, V92, P175, DOI 10.1016/j.ijrobp.2015.02.010
   POLLACK IF, 1995, J NEUROSURG, V82, P536, DOI 10.3171/jns.1995.82.4.0536
   Raney RB, 1999, MED PEDIATR ONCOL, V33, P362, DOI 10.1002/(SICI)1096-911X(199910)33:4<362::AID-MPO4>3.0.CO;2-I
   Ris MD, 2001, J CLIN ONCOL, V19, P3470, DOI 10.1200/JCO.2001.19.15.3470
   Rombi B, 2012, INT J RADIAT ONCOL, V82, P1142, DOI 10.1016/j.ijrobp.2011.03.038
   ROSEN EM, 1984, J CLIN ONCOL, V2, P719, DOI 10.1200/JCO.1984.2.7.719
   Siegel RL, 2017, CA-CANCER J CLIN, V67, P7, DOI [10.3322/caac.20073, 10.3322/caac.20006, 10.3322/caac.21387]
   St Clair WH, 2004, INT J RADIAT ONCOL, V58, P727, DOI 10.1016/S0360-3016(03)01574-8
   Vatner RE, 2018, J CLIN ONCOL, V36, P2854, DOI 10.1200/JCO.2018.78.1492
   Weber DC, 2017, PEDIATR BLOOD CANCER, V64, DOI 10.1002/pbc.26688
   Williams NL, 2018, AM J CLIN ONCOL-CANC, V41, P307, DOI 10.1097/COC.0000000000000267
   Wisoff JH, 2011, NEUROSURGERY, V68, P1548, DOI 10.1227/NEU.0b013e318214a66e
   Wray J, 2016, PEDIATR BLOOD CANCER, V63, P1522, DOI 10.1002/pbc.26044
   Yock T, 2005, INT J RADIAT ONCOL, V63, P1161, DOI 10.1016/j.ijrobp.2005.03.052
   Yock TI, 2016, LANCET ONCOL, V17, P287, DOI 10.1016/S1470-2045(15)00167-9
   Yock TI, 2014, RADIOTHER ONCOL, V113, P89, DOI 10.1016/j.radonc.2014.08.017
   Yock TI, 2014, ACTA ONCOL, V53, P1279, DOI 10.3109/0284186X.2014.957415
   Youland RS, 2013, J PEDIAT HEMATOL ONC, V35, P197, DOI 10.1097/MPH.0b013e3182678bf8
NR 79
TC 23
Z9 23
U1 1
U2 6
PU LIPPINCOTT WILLIAMS & WILKINS
PI PHILADELPHIA
PA TWO COMMERCE SQ, 2001 MARKET ST, PHILADELPHIA, PA 19103 USA
SN 1040-8703
EI 1531-698X
J9 CURR OPIN PEDIATR
JI CURR. OPIN. PEDIATR.
PD FEB
PY 2019
VL 31
IS 1
BP 28
EP 34
DI 10.1097/MOP.0000000000000724
PG 7
WC Pediatrics
WE Science Citation Index Expanded (SCI-EXPANDED)
SC Pediatrics
GA HX5LI
UT WOS:000467442300005
PM 30531222
DA 2023-08-27
ER

PT J
AU Sainte-Rose, C
   Cinalli, G
   Roux, FE
   Maixner, W
   Chumas, PD
   Mansour, M
   Carpentier, A
   Bourgeois, M
   Zerah, M
   Pierre-Kahn, A
   Renier, D
AF Sainte-Rose, C
   Cinalli, G
   Roux, FE
   Maixner, W
   Chumas, PD
   Mansour, M
   Carpentier, A
   Bourgeois, M
   Zerah, M
   Pierre-Kahn, A
   Renier, D
TI Management of hydrocephalus in pediatric patients with posterior fossa
   tumors: the role of endoscopic third ventriculostomy
SO JOURNAL OF NEUROSURGERY
LA English
DT Article
DE posterior fossa tumor; hydrocephalus; third ventriculostomy; endoscopy;
   children
ID 3RD VENTRICULOSTOMY; OBSTRUCTIVE HYDROCEPHALUS; CHILDREN; DRAINAGE;
   SECONDARY; MEDULLOBLASTOMA; SURGERY; SHUNT
AB Object. The authors undertook a study to evaluate the effectiveness of endoscopic third ventriculostomy in the management of hydrocephalus before and after surgical intervention for posterior fossa tumors in children.
   Methods. Between October 1, 1993, and December 31, 1997, a total of 206 consecutive children with posterior fossa tumors underwent surgery at Hopital Necker-Enfants Malades in Paris. Excluded were 10 patients in whom shunts had been placed at the referring hospital. The medical records and neuroimaging studies of the remaining 196 patients were reviewed and categorized into three groups: Group A, 67 patients with hydrocephalus present on admission in whom endoscopic third ventriculostomy was performed prior to tumor removal; Group B, 82 patients with hydrocephalus who did not undergo preliminary third ventriculostomy but instead received conventional treatment; and Group C, 47 patients in whom no ventricular dilation was present on admission.
   There were no significant differences between patients in Group A or B with respect to the following variables: age at presentation, evidence of metastatic disease, extent of tumor resection, or follow-up duration. In patients in Group A, however, more severe hydrocephalus was demonstrated (p < 0.01); the patients in Group C were in this respect different from those in the other two groups.
   Ultimately, there were only four patients (6%) in Group A compared with 22 patients (26.8%) in Group B (p 0.001) in whom progressive hydrocephalus required treatment following removal of the posterior fossa tumor. Sixteen patients (20%) in Group B underwent insertion of a ventriculoperitoneal shunt, which is similar to the incidence reported in the literature and significantly different from that demonstrated in Group A (p < 0.016). The other six patients (7.3%) were treated by endoscopic third ventriculostomy after tumor resection. In Group C, two patients (4.3%) with postoperative hydrocephalus underwent endoscopic third ventriculostomy.
   In three patients who required placement of CSF shunts several episodes of shunt malfunction occurred that were ultimately managed by endoscopic third ventriculostomy and definitive removal of the shunt. There were no deaths; however, there were four cases of transient morbidity associated with third ventriculostomy.
   Conclusions. Third ventriculostomy is feasible even in the presence of posterior fossa tumors (including brainstem tumors). When performed prior to posterior fossa surgery, it significantly reduces the incidence of postoperative hydrocephalus. The procedure provides a valid alternative to placement of a permanent shunt in cases in which hydrocephalus develops following posterior fossa surgery, and it may negate the need for the shunt in cases in which the shunt malfunctions. Furthermore, in patients in whom CSF has caused spread of the tumor at presentation, third ventriculostomy allows chemotherapy to be undertaken prior to tumor excision by controlling hydrocephalus.
   Although the authors acknowledge that the routine application of third ventriculostomy in selected patients results in a proportion of patients undergoing an "unnecessary" procedure, they believe that because patients' postoperative courses are less complicated and because the incidence of morbidity is low and the success rate is high in those patients with severe hydrocephalus that further investigation of this protocol is warranted.
C1 Hop Necker Enfants Malad, Serv Neurochirurg, F-75743 Paris 15, France.
   Osped Santobono, Div Neurochirurg Pediat, Naples, Italy.
   Royal Childrens Hosp, Dept Neurosurg, Melbourne, Vic, Australia.
   Gen Infirm Leeds, Dept Neurosurg, Leeds, W Yorkshire, England.
C3 UDICE-French Research Universities; Universite Paris Cite; Assistance
   Publique Hopitaux Paris (APHP); Hopital Universitaire Cochin - APHP;
   Hopital Universitaire Necker-Enfants Malades - APHP; Royal Children's
   Hospital Melbourne; Leeds General Infirmary
RP Sainte-Rose, C (通讯作者)，Hop Necker Enfants Malad, Serv Neurochirurg, 149 Rue Sevres, F-75743 Paris 15, France.
RI cinalli, giuseppe/AAC-3889-2022; ZERAH, Michel/E-6473-2012
OI cinalli, giuseppe/0000-0002-7797-8483; 
CR ABRAHAM J, 1963, J NEUROSURG, V20, P252, DOI 10.3171/jns.1963.20.3.0252
   ALBRIGHT L, 1977, J NEUROSURG, V46, P52, DOI 10.3171/jns.1977.46.1.0052
   ALLEN JC, 1982, J NEUROSURG, V57, P446, DOI 10.3171/jns.1982.57.4.0446
   Cinalli G, 1998, NEUROSURGERY, V43, P1323, DOI 10.1097/00006123-199812000-00030
   Cinalli G, 1999, J NEUROSURG, V90, P448, DOI 10.3171/jns.1999.90.3.0448
   CULLEY DJ, 1994, NEUROSURGERY, V34, P402, DOI 10.1227/00006123-199403000-00003
   DIAS MS, 1989, PEDIATR NEUROSCI, V15, P283
   EPSTEIN F, 1978, NEUROSURGERY, V3, P348, DOI 10.1227/00006123-197811000-00003
   FORREST DM, 1968, J NEUROSURG, V29, P506, DOI 10.3171/jns.1968.29.5.0506
   HEKMATPANAH J, 1967, J NEUROSURG, V26, P609, DOI 10.3171/jns.1967.26.6.0609
   Hopf NJ, 1999, NEUROSURGERY, V44, P795, DOI 10.1097/00006123-199904000-00062
   JONES RFC, 1990, NEUROSURGERY, V26, P86, DOI 10.1227/00006123-199001000-00012
   KELLY PJ, 1986, NEUROSURGERY, V18, P791, DOI 10.1227/00006123-198606000-00021
   KEUCHER TR, 1979, J NEUROSURG, V50, P179, DOI 10.3171/jns.1979.50.2.0179
   LEE M, 1994, PEDIATR NEUROSURG, V20, P240, DOI 10.1159/000120797
   McLaurin R L, 1983, Clin Neurosurg, V30, P286
   MUSZYNSKI CA, 1994, PEDIATR NEUROSURG, V21, P227, DOI 10.1159/000120840
   PAPO I, 1982, NEUROSURGERY, V10, P13, DOI 10.1227/00006123-198201000-00002
   RAIMONDI AJ, 1981, J NEUROSURG, V55, P174, DOI 10.3171/jns.1981.55.2.0174
   RAPPAPORT ZH, 1989, ACTA NEUROCHIR, V96, P118, DOI 10.1007/BF01456169
   Sainte-Rose C, 1992, NEUROENDOSCOPY, P47
   SCHMID UD, 1986, J NEUROSURG, V65, P649, DOI 10.3171/jns.1986.65.5.0649
   SHALIT MN, 1979, ACTA NEUROCHIR, V47, P161, DOI 10.1007/BF01406401
   STEIN BM, 1972, J NEUROSURG, V36, P763, DOI 10.3171/jns.1972.36.6.0763
NR 24
TC 162
Z9 169
U1 0
U2 2
PU AMER ASSOC NEUROLOGICAL SURGEONS
PI CHARLOTTESVILLE
PA UNIV VIRGINIA, 1224 WEST MAIN ST, STE 450, CHARLOTTESVILLE, VA 22903 USA
SN 0022-3085
J9 J NEUROSURG
JI J. Neurosurg.
PD NOV
PY 2001
VL 95
IS 5
BP 791
EP 797
DI 10.3171/jns.2001.95.5.0791
PG 7
WC Clinical Neurology; Surgery
WE Science Citation Index Expanded (SCI-EXPANDED)
SC Neurosciences & Neurology; Surgery
GA 487WC
UT WOS:000171901200011
PM 11702869
DA 2023-08-27
ER

PT J
AU Desai, K
   Nadkarni, T
   Bhayani, R
   Goel, A
AF Desai, K
   Nadkarni, T
   Bhayani, R
   Goel, A
TI Cerebellopontine angle epidermoid tumor presenting with 'tic convulsif'
   and tinnitus - Case report
SO NEUROLOGIA MEDICO-CHIRURGICA
LA English
DT Article
DE epidermoid; hemifacial spasm; trigeminal neuralgia; tinnitus;
   microvascular decompression
ID HEMIFACIAL SPASM; TRIGEMINAL NEURALGIA; MICROVASCULAR DECOMPRESSION;
   MICROSURGICAL TREATMENT; VASCULAR COMPRESSION; SURGICAL-TREATMENT; NERVE
AB A 22-year-old female presented with a cerebellopontine angle epidermoid tumor manifesting as a rare combination of hemifacial spasm, trigeminal neuralgia, and tinnitus. Magnetic resonance imaging demonstrated the tumor distorting the brainstem and the fourth ventricle. The tumor was almost completely resected and the seventh-eighth cranial nerve complex was decompressed by mobilizing the anterior inferior cerebellar artery loop. No arterial loop was related to the trigeminal nerve. The patient was completely relieved of the "tic convulsif" and tinnitus after the surgery. The inflammatory nature of epidermoid tumor may be involved in the etiology of the syndrome. Microvascular decompression may be needed in addition to tumor removal in such cases.
C1 King Edward Mem Hosp, Dept Neurosurg, Seth GS Med Coll, Mumbai, India.
C3 Seth Gordhandas Sunderdas Medical College & King Edward Memorial
   Hospital
RP Desai, K (通讯作者)，King Edward Mem Hosp, Dept Neurosurg, Seth GS Med Coll, Mumbai, India.
EM
OI Nadkarni, Trimurti/0000-0002-6387-3912
CR AUGER RG, 1986, MAYO CLIN PROC, V61, P640, DOI 10.1016/S0025-6196(12)62028-X
   BURCHIEL KJ, 1988, J NEUROSURG, V69, P35, DOI 10.3171/jns.1988.69.1.0035
   Cushing H, 1920, AM J MED SCI, V160, P157, DOI 10.1097/00000441-192008000-00001
   FRAIOLI B, 1989, NEUROSURGERY, V25, P630, DOI 10.1227/00006123-198910000-00018
   Fukuda H, 2001, ACTA NEUROCHIR, V143, P203, DOI 10.1007/s007010170129
   Fukuda Masafumi, 1997, Neurologia Medico-Chirurgica, V37, P771, DOI 10.2176/nmc.37.771
   GOYA T, 1983, Neurologia Medico-Chirurgica, V23, P651, DOI 10.2176/nmc.23.651
   HAINES SJ, 1979, J NEUROSURG, V50, P257, DOI 10.3171/jns.1979.50.2.0257
   Hasegawa Koichi, 1995, Neurological Surgery, V23, P315
   Janetta PJ, 1993, APS J, V2, P217, DOI 10.1016/S1058-9139(05)80246-3
   JANNETTA PJ, 1977, J NEUROSURG, V47, P321, DOI 10.3171/jns.1977.47.3.0321
   JANNETTA PJ, 1980, ANN SURG, V192, P518, DOI 10.1097/00000658-198010000-00010
   KONDO A, 1980, Neurologia Medico-Chirurgica, V20, P739
   Kuroki A, 1999, NEUROL MED-CHIR, V39, P447, DOI 10.2176/nmc.39.447
   Lin P J, 1991, Gaoxiong Yi Xue Ke Xue Za Zhi, V7, P329
   Matsuka Y, 2000, J OROFAC PAIN, V14, P147
   MATSUMOTO K, 1991, Neurologia Medico-Chirurgica, V31, P410, DOI 10.2176/nmc.31.410
   MIYAZAKI S, 1987, Neurological Surgery, V15, P683
   Miyazaki S, 1983, No To Shinkei, V35, P951
   NAGATA S, 1992, SURG NEUROL, V38, P204, DOI 10.1016/0090-3019(92)90170-R
   OTSUKA S-I, 1989, Archiv fuer Japanische Chirurgie, V58, P245
   Pyen JS, 2001, YONSEI MED J, V42, P255, DOI 10.3349/ymj.2001.42.2.255
   RAPPAPORT ZH, 1985, NEUROCHIRURGIA, V28, P211
   REVILLA AG, 1947, J NEUROSURG, V4, P233, DOI 10.3171/jns.1947.4.3.0233
   SAMII M, 1995, ACTA NEUROCHIR, V134, P148, DOI 10.1007/BF01417682
   Samii M, 1996, J NEUROSURG, V84, P14, DOI 10.3171/jns.1996.84.1.0014
   SINGH AK, 1994, NEUROL RES, V16, P321
   Takano S, 1998, NEUROL MED-CHIR, V38, P100, DOI 10.2176/nmc.38.100
   WAKABAYASHI T, 1983, SURG NEUROL, V19, P244, DOI 10.1016/S0090-3019(83)80008-1
   WILKINS RH, 1991, SURG NEUROL, V36, P251, DOI 10.1016/0090-3019(91)90087-P
   Yasuoka S, 1983, No To Shinkei, V35, P1097
NR 31
TC 17
Z9 17
U1 0
U2 2
PU JAPAN NEUROSURGICAL SOC
PI TOKYO
PA 5-25-16 HONGO, BUNKYO-KU, TOKYO, 113-0033, JAPAN
SN 0470-8105
EI 1349-8029
J9 NEUROL MED-CHIR
JI Neurol. Med.-Chir.
PD APR
PY 2002
VL 42
IS 4
BP 162
EP 165
DI 10.2176/nmc.42.162
PG 4
WC Clinical Neurology; Surgery
WE Science Citation Index Expanded (SCI-EXPANDED)
SC Neurosciences & Neurology; Surgery
GA 541EF
UT WOS:000174971700005
PM 12013668
OA gold
DA 2023-08-27
ER

PT J
AU De Clercq, E
   Grotzer, M
   Landolt, MA
   von Helversen, B
   Flury, M
   Rossler, J
   Kurzo, A
   Streuli, J
AF De Clercq, Eva
   Grotzer, Michael
   Landolt, Markus A.
   von Helversen, Bettina
   Flury, Maria
   Roessler, Jochen
   Kurzo, Andrea
   Streuli, Juerg
TI No wrong decisions in an all-wrong situation. A qualitative study on the
   lived experiences of families of children with diffuse intrinsic pontine
   glioma
SO PEDIATRIC BLOOD & CANCER
LA English
DT Article
DE coping strategies; death; decision-making; DIPG; lived experiences;
   regret
ID PALLIATIVE CARE; CANCER; PARENTS; PERSPECTIVES; LIFE; VIEWS
AB Background Diffuse intrinsic pontine glioma (DIPG) is a rare, but lethal pediatric brain tumor with a median survival of less than 1 year. Existing treatment may prolong life and control symptoms, but may cause toxicity and side effects. In order to improve child- and family-centered care, we aimed to better understand the treatment decision-making experiences of parents, as studies on this topic are currently lacking. Procedure The data for this study came from 24 semistructured interviews with parents whose children were diagnosed with DIPG in two children's hospitals in Switzerland and died between 2000 and 2016. Analysis of the dataset was done using reflexive thematic analysis. Results For most parents, the decision for or against treatment was relatively straightforward given the fatality of the tumor and the absence of treatment protocols. Most of them had no regrets about their decision for or against treatment. The most distressing factor for them was observing their child's gradual loss of independence and informing them about the inescapability of death. To counter this powerlessness, many parents opted for complementary or alternative medicine in order to "do something." Many parents reported psychological problems in the aftermath of their child's death and coping strategies between mothers and fathers often differed. Conclusion The challenges of DIPG are unique and explain why parental and shared decision-making is different in DIPG compared to other cancer diagnoses. Considering that treatment decisions shape parents' grief trajectory, clinicians should reassure parents by framing treatment decisions in terms of family's deeply held values and goals.
C1 [De Clercq, Eva; Streuli, Juerg] Univ Zurich, Inst Bioeth & Hist Med, Winterthurer Str 30, CH-8006 Zurich, Switzerland.
   [Grotzer, Michael; Landolt, Markus A.; Flury, Maria; Streuli, Juerg] Univ Zurich, Univ Childrens Hosp Zurich, Zurich, Switzerland.
   [Landolt, Markus A.] Univ Zurich, Dept Psychol, Zurich, Switzerland.
   [von Helversen, Bettina] Univ Bremen, Dept Psychol, Bremen, Germany.
   [Roessler, Jochen; Kurzo, Andrea] Inselspital Univ, Canc Ctr Inselspital UCI, Bern, Switzerland.
C3 University of Zurich; University Children's Hospital Zurich; University
   of Zurich; University of Zurich; University of Bremen
RP De Clercq, E (通讯作者)，Univ Zurich, Inst Bioeth & Hist Med, Winterthurer Str 30, CH-8006 Zurich, Switzerland.
EM
RI von Helversen, Bettina/AFP-9715-2022
OI von Helversen, Bettina/0000-0003-2004-6922; De Clercq,
   Eva/0000-0002-7835-9553; Landolt, Markus/0000-0003-0760-5558
FU Starr International Foundation; Universitat Zurich
FX The research was funded by the Starr International Foundation. The
   authors sincerely thank the parents who participated in this research
   and generously shared their experiences with them.r Open access funding
   provided by Universitat Zurich.
CR Aein F, 2014, IRAN RED CRESCENT ME, V16, DOI 10.5812/ircmj.8197
   Benedikt MH, 2019, SWISS MED WKLY, V149, DOI 10.4414/smw.2019.20091
   Bergstraesser E, 2015, DEATH STUD, V39, P128, DOI 10.1080/07481187.2014.920434
   Bishop FL, 2010, PEDIATRICS, V125, P768, DOI 10.1542/peds.2009-1775
   Bjork M, 2011, EUR J CANCER CARE, V20, P163, DOI 10.1111/j.1365-2354.2009.01159.x
   Bjork Maria, 2005, J Pediatr Oncol Nurs, V22, P265, DOI 10.1177/1043454205279303
   Bjork Maria, 2009, J Pediatr Nurs, V24, P423, DOI 10.1016/j.pedn.2008.01.082
   Braun V., 2006, QUAL RES PSYCHOL, V3, P77, DOI [https://doi.org/10.1191/1478088706qp063oa, DOI 10.1191/1478088706QP063OA, 10.1191/1478088706qp063oa]
   Braun V, 2019, QUAL RES SPORT EXERC, V11, P589, DOI 10.1080/2159676X.2019.1628806
   Bruce A, 2014, J HOLIST NURS, V32, P35, DOI 10.1177/0898010113498823
   Chiang J, 2020, ACTA NEUROPATHOL COM, V8, DOI 10.1186/s40478-020-00930-9
   Coyne I, 2014, EUR J ONCOL NURS, V18, P273, DOI 10.1016/j.ejon.2014.01.006
   De Clercq E, 2017, EUR J CANCER CARE, V26, DOI 10.1111/ecc.12651
   De Clercq E, 2019, BMC PALLIAT CARE, V18, DOI 10.1186/s12904-019-0438-1
   Gwak HS, 2017, CRIT REV ONCOL HEMAT, V120, P111, DOI 10.1016/j.critrevonc.2017.10.013
   Holmberg C, 2012, BMC COMPLEM ALTERN M, V12, DOI 10.1186/1472-6882-12-218
   Jalmsell L, 2016, ACTA PAEDIATR, V105, P1094, DOI 10.1111/apa.13496
   Jonas D, 2022, J PAIN SYMPTOM MANAG, V63, pE111, DOI 10.1016/j.jpainsymman.2021.07.029
   Lassaletta A, 2018, PEDIATR BLOOD CANCER, V65, DOI 10.1002/pbc.26988
   Lazow MA, 2022, NEURO-ONCOLOGY, V24, P821, DOI 10.1093/neuonc/noab245
   Leemann T, 2020, BMC PALLIAT CARE, V19, DOI 10.1186/s12904-020-00621-1
   Lichtenthal WG, 2020, PALLIATIVE MED, V34, P367, DOI 10.1177/0269216319900301
   Lin JL, 2020, ACAD PEDIATR, V20, P1101, DOI 10.1016/j.acap.2020.06.008
   Lou HL, 2015, CANCER NURS, V38, P298, DOI 10.1097/NCC.0000000000000178
   Macauley R, 2019, PAEDIATR RESPIR REV, V29, P9, DOI 10.1016/j.prrv.2018.05.004
   Mack JW, 2008, J CLIN ONCOL, V26, P4759, DOI 10.1200/JCO.2007.15.6059
   Madrigal VN, 2018, PEDIATRICS, V142, pS170, DOI 10.1542/peds.2018-0516H
   Magi T, 2015, PLOS ONE, V10, DOI 10.1371/journal.pone.0145787
   Pohlkamp L, 2019, PSYCHO-ONCOLOGY, V28, P1530, DOI 10.1002/pon.5112
   Rashed WM, 2019, CANCER METAST REV, V38, P759, DOI 10.1007/s10555-019-09824-2
   Renjilian CB, 2013, PEDIATRICS, V131, pE566, DOI 10.1542/peds.2012-1957
   Rensen N, 2019, PEDIATR BLOOD CANCER, V66, DOI 10.1002/pbc.27728
   Robinson JE, 2019, CHILDREN-BASEL, V6, DOI 10.3390/children6100113
   Rocque BG, 2019, J NEUROSURG-PEDIATR, V23, P40, DOI 10.3171/2018.7.PEDS18297
   Rost M, 2020, J PEDIATR ONCOL NURS, V37, P35, DOI 10.1177/1043454219871082
   Ruch C., 2013, PALLIATIVE CH, V2, P45
   Schuelke MR, 2019, J IMMUNOTHER CANCER, V7, DOI 10.1186/s40425-019-0673-2
   Shortman RI, 2013, CHILD CARE HLTH DEV, V39, P743, DOI 10.1111/cch.12005
   Sim CW, 2020, PEDIATR BLOOD CANCER, V67, DOI 10.1002/pbc.28265
   Snaman J, 2020, SUPPORT CARE CANCER, V28, P4131, DOI 10.1007/s00520-019-05249-3
   Soleman J, 2021, CHILD NERV SYST, V37, P1449, DOI 10.1007/s00381-021-05148-1
   Souweidane MM, 2018, LANCET ONCOL, V19, P1040, DOI 10.1016/S1470-2045(18)30322-X
   Streuli JC, 2021, EUR J PEDIATR, V180, P759, DOI 10.1007/s00431-020-03756-8
   Stub T, 2020, BMC COMPLEMENT MED, V20, DOI 10.1186/s12906-020-02882-y
   Valdez-Martinez E, 2014, BMC PEDIATR, V14, DOI 10.1186/1471-2431-14-124
   VERBI Software, 2019, MAXQDA 2020
   Weaver MS, 2015, PEDIATR BLOOD CANCER, V62, pS829, DOI 10.1002/pbc.25695
   Whitney SN, 2006, J CLIN ONCOL, V24, P160, DOI 10.1200/JCO.2005.01.8390
   Williams J, 2013, UNITED NATIONS CONVENTION ON THE RIGHTS OF THE CHILD IN WALES, P1
   Williams JR, 2020, NEUROSURG FOCUS, V48, DOI 10.3171/2019.9.FOCUS19745
   Wolfe J, 2000, JAMA-J AM MED ASSOC, V284, P2469, DOI 10.1001/jama.284.19.2469
   Woodgate RL, 2006, EUR J CANCER CARE, V15, P8, DOI 10.1111/j.1365-2354.2005.00614.x
   Young K, 2021, PSYCHO-ONCOLOGY, V30, P1643, DOI 10.1002/pon.5745
   Zelcer S, 2010, ARCH PEDIAT ADOL MED, V164, P225, DOI 10.1001/archpediatrics.2009.284
NR 54
TC 0
Z9 0
U1 1
U2 2
PU WILEY
PI HOBOKEN
PA 111 RIVER ST, HOBOKEN 07030-5774, NJ USA
SN 1545-5009
EI 1545-5017
J9 PEDIATR BLOOD CANCER
JI Pediatr. Blood Cancer
PD SEP
PY 2022
VL 69
IS 9
AR e29792
DI 10.1002/pbc.29792
EA JUN 2022
PG 12
WC Oncology; Hematology; Pediatrics
WE Science Citation Index Expanded (SCI-EXPANDED)
SC Oncology; Hematology; Pediatrics
GA 3D1YF
UT WOS:000804729700001
PM 35652529
OA Green Published
DA 2023-08-27
ER

PT J
AU DEMPSEY, PK
   KONDZIOLKA, D
   LUNSFORD, LD
AF DEMPSEY, PK
   KONDZIOLKA, D
   LUNSFORD, LD
TI STEREOTAXIC DIAGNOSIS AND TREATMENT OF PINEAL REGION TUMORS AND VASCULAR
   MALFORMATIONS
SO ACTA NEUROCHIRURGICA
LA English
DT Article
DE PINEAL REGION; RADIOSURGERY; STEREOTAXIC BIOPSY; VASCULAR MALFORMATION
ID INTRACRANIAL ARTERIOVENOUS-MALFORMATIONS; GAMMA-KNIFE;
   LINEAR-ACCELERATOR; TUMORS; RADIOSURGERY; MANAGEMENT; EXPERIENCE;
   BIOPSY; RADIATION; SURGERY
AB Diagnosis and treatment of tumours and vascular malformations in the region of the pineal gland continue to challenge the neurosurgeon's skill. Due to vital vascular and brain structures in the region, microsurgical removal of such masses is often impossible.
   During the past nine years, we have managed 47 patients with pineal region mass lesions using stereotactic techniques for diagnosis and treatment. In order to determine further therapeutic options, 15 patients underwent stereotactic biopsy of pineal region tumours. In all patients, the histologic diagnosis obtained served to direct further therapy. Thirty-two patients were treated with stereotactic radiosurgery for pineal region tumours or vascular malformations. During the follow-up period, one patient underwent delayed microsurgical resection of a midbrain angiographically occult vascular malformation. No other patient required microsurgical intervention after a stereotactic procedure. In all 47 patients, no significant morbidity or mortality occurred after stereotactic biopsy or radiosurgery.
   Empiric treatment of pineal region tumours with fractionated radiation therapy is no longer warranted. Image-guided stereotactic technology provides a safe method to accurately diagnose and effectively treat selected pineal region masses. After definitive histologic diagnosis is established, proper treatment may be instituted. Options for treatment include stereotactic radiosurgery for selected tumours and vascular malformations, microsurgical resection of benign tumours or fractionated external beam radiation therapy for malignant germ cell and glial tumours. Stereotactic surgery should be the first option in the diagnosis and therapy of pineal region masses.
C1 UNIV PITTSBURGH, PRESBYTERIAN UNIV HOSP,SCH MED,DEPT NEUROL SURG, ROOM F-948, PITTSBURGH, PA 15213 USA.
C3 Pennsylvania Commonwealth System of Higher Education (PCSHE); University
   of Pittsburgh; UPMC Presbyterian
RI Lunsford, L. Dade/AHI-7993-2022
CR ABAY EO, 1981, J NEUROSURG, V55, P889, DOI 10.3171/jns.1981.55.6.0889
   APUZZO MLZ, 1987, NEUROSURGERY STATE A, P287
   BACKLUND EO, 1974, ACTA RADIOL THER PHY, V13, P368, DOI 10.3109/02841867409134490
   Bruce J N, 1990, Neurosurg Clin N Am, V1, P123
   CAMINS MB, 1978, ACTA NEUROCHIR, V40, P131, DOI 10.1007/BF01773121
   CASENTINI L, 1990, SURG NEUROL, V34, P79, DOI 10.1016/0090-3019(90)90101-T
   CHANDRASOMA PT, 1989, NEUROSURGERY, V24, P160, DOI 10.1227/00006123-198902000-00002
   CHAPMAN PH, 1980, CANCER, V46, P1253, DOI 10.1002/1097-0142(19800901)46:5<1253::AID-CNCR2820460528>3.0.CO;2-D
   COFFEY RJ, 1985, NEUROSURGERY, V17, P12, DOI 10.1227/00006123-198507000-00003
   COLOMBO F, 1989, NEUROSURGERY, V24, P833, DOI 10.1227/00006123-198906000-00008
   CONWAY LW, 1973, J NEUROSURG, V38, P453, DOI 10.3171/jns.1973.38.4.0453
   CUMMINS FM, 1960, NEUROLOGY, V10, P1031, DOI 10.1212/WNL.10.11.1031
   CUSHING H, 1932, INTRACRANIAL TUMORS, P62
   Dandy WE, 1936, ARCH SURG-CHICAGO, V33, P19, DOI 10.1001/archsurg.1936.01190010022002
   DEMAKAS JJ, 1982, SURG NEUROL, V17, P435, DOI 10.1016/S0090-3019(82)80014-1
   DONAT JF, 1978, ARCH NEUROL-CHICAGO, V35, P736, DOI 10.1001/archneur.1978.00500350040008
   EDWARDS MSB, 1988, J NEUROSURG, V68, P689, DOI 10.3171/jns.1988.68.5.0689
   FRIEDMAN WA, 1989, NEUROSURGERY, V25, P180, DOI 10.1227/00006123-198908000-00005
   HITCHON PW, 1983, NEUROSURGERY, V13, P248, DOI 10.1227/00006123-198309000-00006
   HORRAX G, 1950, ARCH NEURO PSYCHIATR, V64, P227, DOI 10.1001/archneurpsyc.1950.02310260065004
   JAMIESON KG, 1971, J NEUROSURG, V35, P550, DOI 10.3171/jns.1971.35.5.0550
   JENKIN RDT, 1978, J NEUROSURG, V48, P99, DOI 10.3171/jns.1978.48.1.0099
   JOOMA R, 1983, J NEUROSURG, V58, P654, DOI 10.3171/jns.1983.58.5.0654
   KONDZIOLKA D, 1990, NEUROSURGERY, V27, P892, DOI 10.1227/00006123-199012000-00006
   KONDZIOLKA D, 1991, J NEUROSURG, V74, P552, DOI 10.3171/jns.1991.74.4.0552
   LINQUIST C, 1988, MODERN STEREOTACTIC, P491
   LINSKEY ME, 1990, NEUROSURGERY, V26, P736, DOI 10.1227/00006123-199005000-00002
   LOEFFLER JS, 1989, INT J RADIAT ONCOL, V17, P673, DOI 10.1016/0360-3016(89)90123-5
   LUNSFORD LD, 1990, ARCH NEUROL-CHICAGO, V47, P169, DOI 10.1001/archneur.1990.00530020071018
   LUNSFORD LD, 1989, NEUROSURGERY, V24, P151
   LUNSFORD LD, 1991, STEREOT FUNCT NEUROS, V54, P375
   MARSH WR, 1984, CLIN NEUROSURG, V32, P384
   MOSER RP, 1984, DIAGNOSIS TREATMENT, P236
   NAZZARO JM, 1990, J NEUROSURG, V73, P331, DOI 10.3171/jns.1990.73.3.0331
   NEUWELT ER, 1984, CLIN NEUROSURG, V32, P397
   Nor┬u┬n G., 1988, MODERN STEREOTACTIC, P481, DOI 10.1007/978-1-4613-1081-5_38
   OBRADOR S, 1976, ACTA NEUROCHIR, V34, P159, DOI 10.1007/BF01405870
   OGILVY CS, 1988, J NEUROSURG, V69, P350, DOI 10.3171/jns.1988.69.3.0350
   ONO M, 1984, J NEUROSURG, V60, P365, DOI 10.3171/jns.1984.60.2.0365
   OSTERTAG CB, 1988, MODERN STEREOTACTIC, P129
   PECKER J, 1979, SURG NEUROL, V12, P341
   PENDL G, 1984, DIAGNOSIS TREATMENT, P155
   PERTUISET B, 1976, ACTA NEUROCHIR, V34, P151, DOI 10.1007/BF01405868
   POPPEN JL, 1968, J NEUROSURG, V28, P357, DOI 10.3171/jns.1968.28.4.0357
   RAND RW, 1953, J NEUROSURG, V10, P1, DOI 10.3171/jns.1953.10.1.0001
   REID WS, 1978, NEUROSURGERY, V3, P1
   RINGERTZ N, 1954, J NEUROPATH EXP NEUR, V13, P540, DOI 10.1097/00005072-195410000-00004
   ROSENFELD JV, 1990, J NEUROSURG, V73, P287, DOI 10.3171/jns.1990.73.2.0287
   Russell DS, 1989, PATHOLOGY TUMOURS NE, P664
   SANO K, 1987, SURGERY 3RD VENTRICL, P663
   STEIN BM, 1971, J NEUROSURG, V35, P197, DOI 10.3171/jns.1971.35.2.0197
   STEIN BM, 1984, CLIN NEUROSURG, V31, P445
   STEIN BM, 1988, OPERATIVE NEUROSURGI, P401
   STEIN BM, 1984, DIAGNOSIS TREATMENT, P254
   STEINBERG GK, 1990, NEW ENGL J MED, V323, P96, DOI 10.1056/NEJM199007123230205
   WOOD JH, 1981, SURG NEUROL, V16, P192, DOI 10.1016/0090-3019(81)90008-2
   WU A, 1990, INT J RADIAT ONCOL, V18, P941, DOI 10.1016/0360-3016(90)90421-F
   Yasargil M., 1988, MICRONEUROSURGERY B, VIIIB, P323
   Yasargil M G, 1976, Surg Neurol, P195
   ZEE CS, 1991, J COMPUT ASSIST TOMO, V15, P56, DOI 10.1097/00004728-199101000-00007
NR 60
TC 57
Z9 59
U1 0
U2 2
PU SPRINGER WIEN
PI WIEN
PA SACHSENPLATZ 4-6, PO BOX 89, A-1201 WIEN, AUSTRIA
SN 0001-6268
EI 0942-0940
J9 ACTA NEUROCHIR
JI Acta Neurochir.
PY 1992
VL 116
IS 1
BP 14
EP 22
DI 10.1007/BF01541248
PG 9
WC Clinical Neurology; Surgery
WE Science Citation Index Expanded (SCI-EXPANDED)
SC Neurosciences & Neurology; Surgery
GA HX204
UT WOS:A1992HX20400003
PM 1615765
DA 2023-08-27
ER

PT J
AU McNamara, C
   Mankad, K
   Thust, S
   Dixon, L
   Limback-Stanic, C
   D'Arco, F
   Jacques, TS
   Lobel, U
AF McNamara, Cillian
   Mankad, Kshitij
   Thust, Stefanie
   Dixon, Luke
   Limback-Stanic, Clara
   D'Arco, Felice
   Jacques, Thomas S.
   Lobel, Ulrike
TI 2021 WHO classification of tumours of the central nervous system: a
   review for the neuroradiologist
SO NEURORADIOLOGY
LA English
DT Review
DE World Health Organization; Brain tumour; Central nervous system;
   Classification; Diagnosis
ID VACUOLATING NEURONAL TUMOR; TERT PROMOTER MUTATIONS; ATYPICAL
   TERATOID/RHABDOID TUMORS; CRIBRIFORM NEUROEPITHELIAL TUMOR; INTRINSIC
   PONTINE GLIOMA; DIFFUSE MIDLINE GLIOMAS; LOWER-GRADE GLIOMAS;
   ANGIOCENTRIC GLIOMA; MOLECULAR SUBGROUPS; CLINICOPATHOLOGICAL FEATURES
AB The fifth edition of the World Health Organization Classification of Tumours of the Central Nervous System (WHO CNS5) published in 2021 builds on the 2016 edition and incorporates output from the Consortium to Inform Molecular and Practical Approaches to CNS Tumour Taxonomy (cIMPACT-NOW). WHO CNS5 introduces fundamental changes to brain tumour classification through the introduction of new tumour families and types, especially in the paediatric population, and a revision of diagnostic criteria for some of the existing neoplasms. Neuroradiologists are central to brain tumour diagnostics, and it is therefore essential that they become familiar with the key updates. This review aims to summarise the most relevant updates for the neuroradiologist and, where available, discuss the known radiophenotypes of various new tumour types to allow for increased accuracy of language and diagnosis. Of particular importance, WHO CNS5 places greater emphasis on organising tumours by molecular type to reflect biology, as well as to allow for better planning of treatment. The principal updates in adult tumours concern the molecular definition of glioblastoma, restructuring of diffuse gliomas, and the introduction of several new tumour types. The updates to the paediatric classification are protean, ranging from the introduction of new types to establishing separate tumour families for paediatric-type gliomas. This review summarises the most significant revisions and captures the rationale and radiological implications for the major updates.
C1 [McNamara, Cillian; Mankad, Kshitij; D'Arco, Felice; Lobel, Ulrike] Great Ormond St Hosp Children NHS Fdn Trust, Dept Radiol, London, England.
   [McNamara, Cillian; Thust, Stefanie] Natl Hosp Neurol & Neurosurg, Dept Neuroradiol, Queen Sq, London, England.
   [Dixon, Luke] Imperial Coll Healthcare NHS Trust, Dept Neuroradiol, London, England.
   [Limback-Stanic, Clara] Dept Cellular Pathol, Imperial Coll Healthcare NHS Trust, London, England.
   [Jacques, Thomas S.] Great Ormond St Hosp Children NHS Fdn Trust, DBC Programme, UCL GOS Inst Child Hlth, London, England.
   [Jacques, Thomas S.] Great Ormond St Hosp Children NHS Fdn Trust, Dept Histopathol, London, England.
C3 University of London; University College London; Great Ormond Street
   Hospital for Children NHS Foundation Trust; University of London;
   University College London; UCL Medical School; University College London
   Hospitals NHS Foundation Trust; Imperial College London; Imperial
   College London; University of London; University College London; Great
   Ormond Street Hospital for Children NHS Foundation Trust; University of
   London; University College London; Great Ormond Street Hospital for
   Children NHS Foundation Trust
RP McNamara, C (通讯作者)，Great Ormond St Hosp Children NHS Fdn Trust, Dept Radiol, London, England.
EM
RI Dixon, Luke/IZE-9083-2023; Jacques, Thomas/C-2209-2008; McNamara,
   Cillian/AAS-5109-2021
OI Jacques, Thomas/0000-0002-7833-2158; McNamara,
   Cillian/0000-0001-8164-7794; Thust, Stefanie/0000-0001-5136-6000
FU Brain Tumour Charity; Children with Cancer UK; Great Ormond Street
   Hospital Children's Charity; Olivia Hodson Cancer Fund; Cancer Research
   UK; National Institute of Health Research; UCL/UCLH NIHR Biomedical
   Research Centre
FX No funding was received for this review. TJ is grateful for funding from
   the Brain Tumour Charity, Children with Cancer UK, Great Ormond Street
   Hospital Children's Charity, Olivia Hodson Cancer Fund, Cancer Research
   UK, and the National Institute of Health Research. All research at Great
   Ormond Street Hospital NHS Foundation Trust and UCL Great Ormond Street
   Institute of Child Health is made possible by the NIHR Great Ormond
   Street Hospital Biomedical Research Centre. ST receives funding from the
   UCL/UCLH NIHR Biomedical Research Centre. The views expressed are those
   of the author(s) and not necessarily those of the NHS, the NIHR or the
   Department of Health.
CR Abdallah A, 2020, WORLD NEUROSURG, V136, pE245, DOI 10.1016/j.wneu.2019.12.128
   Razek AAKA, 2020, J COMPUT ASSIST TOMO, V44, P356, DOI 10.1097/RCT.0000000000001010
   Aboian MS, 2019, AM J NEURORADIOL, V40, P1804, DOI 10.3174/ajnr.A6302
   Al-Battashi Abeer, 2019, Sultan Qaboos Univ Med J, V19, pe153, DOI 10.18295/squmj.2019.19.02.012
   Alsufayan R, 2017, NEURORADIOLOGY, V59, P873, DOI 10.1007/s00234-017-1884-1
   Ampie L, 2016, J CLIN NEUROSCI, V28, P117, DOI 10.1016/j.jocn.2015.11.015
   Andreiuolo F, 2019, BRAIN PATHOL, V29, P205, DOI 10.1111/bpa.12659
   Bag AK, 2021, NEURORADIOLOGY, V63, P1185, DOI 10.1007/s00234-021-02691-1
   Bandopadhayay P, 2016, NAT GENET, V48, P273, DOI 10.1038/ng.3500
   Bandopadhayay P, 2016, J NEURO-ONCOL, V126, P165, DOI 10.1007/s11060-015-1955-2
   Buffa GB, 2020, J NEURORADIOLOGY, V47, P216, DOI 10.1016/j.neurad.2019.05.010
   Bender K, 2021, J NEURO-ONCOL, V153, P109, DOI 10.1007/s11060-021-03749-z
   Bender S, 2016, NAT MED, V22, P1314, DOI 10.1038/nm.4204
   Berzero G, 2021, NEURO-ONCOLOGY, V23, P955, DOI 10.1093/neuonc/noaa258
   Bianchi F, 2018, CHILD NERV SYST, V34, P817, DOI 10.1007/s00381-018-3764-3
   Blumcke I, 2017, NEW ENGL J MED, V377, P1648, DOI 10.1056/NEJMoa1703784
   Blumcke I, 2014, ACTA NEUROPATHOL, V128, P39, DOI 10.1007/s00401-014-1288-9
   Bodi I, 2014, ACTA NEUROPATHOL COM, V2, DOI 10.1186/2051-5960-2-7
   Brat DJ, 2020, ACTA NEUROPATHOL, V139, P603, DOI 10.1007/s00401-020-02127-9
   Brat DJ, 2018, ACTA NEUROPATHOL, V136, P805, DOI 10.1007/s00401-018-1913-0
   Brat DJ, 2000, BRAIN PATHOL, V10, P342
   Broniscer A, 2018, BRAIN PATHOL, V28, P112, DOI 10.1111/bpa.12484
   Bruford EA, 2021, LEUKEMIA, V35, P3040, DOI 10.1038/s41375-021-01436-6
   Capper D, 2018, NATURE, V555, P469, DOI 10.1038/nature26000
   Chan E, 2017, ACTA NEUROPATHOL, V134, P671, DOI 10.1007/s00401-017-1759-x
   Chen H, 2019, EUR J RADIOL, V114, P152, DOI 10.1016/j.ejrad.2019.03.006
   Chen WW, 2020, BRAIN TUMOR PATHOL, V37, P145, DOI 10.1007/s10014-020-00372-0
   Chen YQ, 2020, BMC NEUROL, V20, DOI 10.1186/s12883-020-01679-3
   Chiang J, 2019, ACTA NEUROPATHOL, V138, P1091, DOI 10.1007/s00401-019-02081-1
   Chiang JCH, 2019, NEURO-ONCOLOGY, V21, P800, DOI 10.1093/neuonc/noz037
   Chiang JCH, 2017, ACTA NEUROPATHOL, V134, P159, DOI 10.1007/s00401-017-1728-4
   Clarke M, 2020, CANCER DISCOV, V10, P942, DOI 10.1158/2159-8290.CD-19-1030
   Covington DB, 2009, PEDIATR NEUROSURG, V45, P429, DOI 10.1159/000277616
   De Lima L, 2020, CHILD NERV SYST, V36, P1589, DOI 10.1007/s00381-020-04692-6
   den Dunnen JT, 2016, HUM MUTAT, V37, P564, DOI 10.1002/humu.22981
   Deng MY, 2020, NEUROPATH APPL NEURO, V46, P422, DOI 10.1111/nan.12590
   Eckel-Passow JE, 2015, NEW ENGL J MED, V372, P2499, DOI 10.1056/NEJMoa1407279
   Ellison DW, 2020, BRAIN PATHOL, V30, P863, DOI 10.1111/bpa.12866
   Ellison DW, 2019, ACTA NEUROPATHOL, V137, P683, DOI 10.1007/s00401-019-01987-0
   Ellison DW, 2011, ACTA NEUROPATHOL, V121, P381, DOI 10.1007/s00401-011-0800-8
   Ellison DW, 2005, J CLIN ONCOL, V23, P7951, DOI 10.1200/JCO.2005.01.5479
   Ferris SP, 2020, BRAIN PATHOL, V30, P46, DOI 10.1111/bpa.12747
   Fiechter M, 2016, CLIN NEUROL NEUROSUR, V147, P98, DOI 10.1016/j.clineuro.2016.06.005
   Fruehwald-Pallamar J, 2011, NEURORADIOLOGY, V53, P387, DOI 10.1007/s00234-010-0829-8
   Fukushima S, 2015, BRAIN TUMOR PATHOL, V32, P131, DOI 10.1007/s10014-014-0198-9
   Gamboa Nicholas T, 2018, Surg Neurol Int, V9, P144, DOI 10.4103/sni.sni_116_18
   Gareton A, 2020, ACTA NEUROPATHOL, V139, P287, DOI 10.1007/s00401-019-02088-8
   Ghasemi DR, 2019, ACTA NEUROPATHOL, V138, P1075, DOI 10.1007/s00401-019-02056-2
   Gilbert AR, 2018, PEDIATR NEUROSURG, V53, P59, DOI 10.1159/000481513
   Goncalves FG, 2021, FRONT NEUROL, V12, DOI 10.3389/fneur.2021.733323
   Goutagny S, 2014, BRAIN PATHOL, V24, P184, DOI 10.1111/bpa.12110
   Guyot A, 2019, J NEURO-ONCOL, V145, P449, DOI 10.1007/s11060-019-03333-6
   Hasselblatt M, 2018, J NEUROPATH EXP NEUR, V77, P422, DOI 10.1093/jnen/nly012
   Hasselblatt M, 2009, J NEUROPATH EXP NEUR, V68, P1249, DOI 10.1097/NEN.0b013e3181c06a51
   Hill RM, 2020, LANCET CHILD ADOLESC, V4, P865, DOI 10.1016/S2352-4642(20)30246-7
   Ho B, 2020, NEURO-ONCOLOGY, V22, P613, DOI 10.1093/neuonc/noz235
   Holsken A, 2016, ACTA NEUROPATHOL COM, V4, DOI 10.1186/s40478-016-0287-6
   Holsten T, 2021, J NEUROPATH EXP NEUR, V80, P52, DOI 10.1093/jnen/nlaa128
   Huse JT, 2017, ACTA NEUROPATHOL, V133, P417, DOI 10.1007/s00401-016-1639-9
   Huse JT, 2013, BRAIN PATHOL, V23, P515, DOI 10.1111/bpa.12035
   Isler C, 2018, BRIT J NEUROSURG, V32, P436, DOI 10.1080/02688697.2018.1476671
   Iv M, 2019, AM J NEURORADIOL, V40, P154, DOI 10.3174/ajnr.A5899
   Jenkins RB, 2006, CANCER RES, V66, P9852, DOI 10.1158/0008-5472.CAN-06-1796
   Johann PD, 2017, BRAIN PATHOL, V27, P411, DOI 10.1111/bpa.12413
   Johann PD, 2016, CANCER CELL, V29, P379, DOI 10.1016/j.ccell.2016.02.001
   Johnson DR, 2019, NEURORADIOLOGY, V61, P1327, DOI 10.1007/s00234-019-02269-y
   Junger ST, 2020, ACTA NEUROPATHOL, V140, P405, DOI 10.1007/s00401-020-02169-z
   Juratli TA, 2019, J NEURO-ONCOL, V141, P327, DOI 10.1007/s11060-018-03034-6
   Juratli TA, 2017, ONCOTARGET, V8, P109228, DOI 10.18632/oncotarget.22650
   Karlowee V, 2017, CHILD NERV SYST, V33, P1411, DOI 10.1007/s00381-017-3405-2
   Kathrani N, 2022, NEURORADIOLOGY, V64, P1519, DOI 10.1007/s00234-021-02857-x
   Khalid L, 2012, AM J NEURORADIOL, V33, P852, DOI 10.3174/ajnr.A2895
   Khanna V, 2017, J NEURO-ONCOL, V135, P433, DOI 10.1007/s11060-017-2594-6
   Kirkman MA, 2018, WORLD NEUROSURG, V116, P279, DOI 10.1016/j.wneu.2018.05.158
   Kool M, 2012, ACTA NEUROPATHOL, V123, P473, DOI 10.1007/s00401-012-0958-8
   Koral K, 2012, CLIN IMAG, V36, P61, DOI 10.1016/j.clinimag.2011.04.016
   Korshunov A, 2017, ACTA NEUROPATHOL, V134, P507, DOI 10.1007/s00401-017-1710-1
   Korshunov A, 2016, ACTA NEUROPATHOL, V131, P137, DOI 10.1007/s00401-015-1493-1
   Lakhani DA, 2020, AM J NEURORADIOL, V41, P2155, DOI 10.3174/ajnr.A6737
   Lambo S, 2019, NATURE, V576, P274, DOI 10.1038/s41586-019-1815-x
   Latysheva A, 2019, NEURORADIOLOGY, V61, P545, DOI 10.1007/s00234-019-02173-5
   Leach JL, 2020, NEURO-ONCOLOGY, V22, P1647, DOI 10.1093/neuonc/noaa140
   Lecler A, 2018, AM J NEURORADIOL, V39, pE32, DOI 10.3174/ajnr.A5425
   Lee JK, 2018, CASE REP NEUROL MED, V2018, DOI 10.1155/2018/1391943
   Lehman NL, 2019, ACTA NEUROPATHOL COM, V7, DOI 10.1186/s40478-019-0689-3
   Lellouch-Tubiana A, 2005, BRAIN PATHOL, V15, P281
   Liu APY, 2021, ACTA NEUROPATHOL, V141, P771, DOI 10.1007/s00401-021-02284-5
   Liu TT, 2016, GENES-BASEL, V7, DOI 10.3390/genes7070038
   Lobel U, 2011, AM J NEURORADIOL, V32, P315, DOI 10.3174/ajnr.A2277
   Louis DN, 2021, NEURO-ONCOLOGY, V23, P1231, DOI 10.1093/neuonc/noab106
   Louis DN, 2020, BRAIN PATHOL, V30, P844, DOI 10.1111/bpa.12832
   Louis DN, 2020, ARCH PATHOL LAB MED, V144, P196, DOI 10.5858/arpa.2018-0565-OA
   Louis DN, 2014, BRAIN PATHOL, V24, P429, DOI 10.1111/bpa.12171
   Lucas CHG, 2020, BRAIN PATHOL, V30, P479, DOI 10.1111/bpa.12797
   Mackay A, 2017, CANCER CELL, V32, P520, DOI 10.1016/j.ccell.2017.08.017
   Marburger T, 2011, ARCH PATHOL LAB MED, V135, P1037, DOI 10.5858/2010-0668-OAR
   Massimino M, 2021, NEURO-ONCOLOGY, V23, P848, DOI 10.1093/neuonc/noaa257
   Maynard J, 2020, RADIOLOGY, V296, P111, DOI 10.1148/radiol.2020191832
   Mete O, 2013, AM J SURG PATHOL, V37, P1694, DOI 10.1097/PAS.0b013e31829723e7
   Meyronet D, 2017, NEURO-ONCOLOGY, V19, P1127, DOI 10.1093/neuonc/now274
   Mhatre R, 2019, BRAIN TUMOR PATHOL, V36, P112, DOI 10.1007/s10014-019-00346-x
   Monday G, 2020, ACTA NEUROPATHOL, V139, P1071, DOI 10.1007/s00401-020-02155-5
   Muller HL, 2019, NAT REV DIS PRIMERS, V5, DOI 10.1038/s41572-019-0125-9
   Mynarek M, 2020, J CLIN ONCOL, V38, P2028, DOI 10.1200/JCO.19.03057
   Nagib MG, 1996, PEDIATR NEUROSURG, V24, P299, DOI 10.1159/000121059
   Naitoh Y, 2002, AM J NEURORADIOL, V23, P1564
   Narvaez ED, 2021, BJR CASE REP, V7, DOI 10.1259/bjrcr.20200139
   Ng A, 2019, BMJ CASE REP, V12, DOI 10.1136/bcr-2018-228248
   Ni HC, 2015, NEUROPATH APPL NEURO, V41, P333, DOI 10.1111/nan.12158
   Nowak J, 2014, AM J NEURORADIOL, V35, P1996, DOI 10.3174/ajnr.A4002
   Nowak J, 2018, NEURO-ONCOLOGY, V20, P1672, DOI 10.1093/neuonc/noy111
   Nowak J, 2019, CLIN NEURORADIOL, V29, P595, DOI 10.1007/s00062-018-0704-2
   Nunes RH, 2017, AM J NEURORADIOL, V38, P1899, DOI 10.3174/ajnr.A5281
   Olsen TK, 2015, NEURO-ONCOLOGY, V17, P1365, DOI 10.1093/neuonc/nov039
   Onishi S, 2018, BRAIN TUMOR PATHOL, V35, P41, DOI 10.1007/s10014-017-0301-0
   Ostrom QT, 2014, NEURO-ONCOLOGY, V16, P1, DOI [10.1093/neuonc/nov189, 10.1093/neuonc/nou327, 10.1093/neuonc/nou223]
   Pajtler KW, 2018, ACTA NEUROPATHOL, V136, P211, DOI 10.1007/s00401-018-1877-0
   Pajtler KW, 2015, CANCER CELL, V27, P728, DOI 10.1016/j.ccell.2015.04.002
   Park JY, 2012, NEUROPATHOLOGY, V32, P570, DOI 10.1111/j.1440-1789.2011.01293.x
   Patay Z, 2015, AM J NEURORADIOL, V36, P2386, DOI 10.3174/ajnr.A4495
   Patel SH, 2017, CLIN CANCER RES, V23, P6078, DOI 10.1158/1078-0432.CCR-17-0560
   Perreault S, 2014, AM J NEURORADIOL, V35, P1263, DOI 10.3174/ajnr.A3990
   Perry A, 2002, BRAIN PATHOL, V12, P183
   Pfister S, 2009, ACTA NEUROPATHOL, V117, P457, DOI 10.1007/s00401-008-0467-y
   Pfister SM, 2022, CANCER DISCOV, V12, P331, DOI 10.1158/2159-8290.CD-21-1094
   Phoenix TN, 2016, CANCER CELL, V29, P508, DOI 10.1016/j.ccell.2016.03.002
   Picart T., 2021, NEUROONCOL ADV, V3
   Pickles JC, 2021, NEUROPATH APPL NEURO, V47, P464, DOI 10.1111/nan.12680
   Pickles JC, 2020, J PATHOL, V250, P510, DOI 10.1002/path.5397
   Pickles JC, 2020, LANCET CHILD ADOLESC, V4, P121, DOI 10.1016/S2352-4642(19)30342-6
   Pietsch T, 2014, ACTA NEUROPATHOL, V127, P609, DOI 10.1007/s00401-014-1264-4
   Pratt D, 2020, ACTA NEUROPATHOL COM, V8, DOI 10.1186/s40478-020-00905-w
   Priesterbach-Ackley LP, 2020, NEUROPATH APPL NEURO, V46, P478, DOI 10.1111/nan.12610
   Raffeld M, 2020, ACTA NEUROPATHOL COM, V8, DOI 10.1186/s40478-020-00973-y
   Ramaswamy V, 2016, ACTA NEUROPATHOL, V131, P821, DOI 10.1007/s00401-016-1569-6
   Ramkissoon LA, 2013, P NATL ACAD SCI USA, V110, P8188, DOI 10.1073/pnas.1300252110
   Reinhardt A, 2019, ACTA NEUROPATHOL COM, V7, DOI 10.1186/s40478-019-0801-8
   Reinhardt A, 2018, ACTA NEUROPATHOL, V136, P273, DOI 10.1007/s00401-018-1837-8
   Reuss DE, 2015, ACTA NEUROPATHOL, V130, P407, DOI 10.1007/s00401-015-1454-8
   Rindi G, 2018, MODERN PATHOL, V31, P1770, DOI 10.1038/s41379-018-0110-y
   Robinson GW, 2020, J CLIN ONCOL, V38, P2010, DOI 10.1200/JCO.20.00593
   Rock K, 2012, BRIT J RADIOL, V85, pE729, DOI 10.1259/bjr/83796755
   Rodriguez FJ, 2012, ACTA NEUROPATHOL, V124, P627, DOI 10.1007/s00401-012-1037-x
   Roux A, 2020, NEURO-ONCOLOGY, V22, P993, DOI 10.1093/neuonc/noaa022
   Roux A, 2020, NEURO-ONCOLOGY, V22, P1190, DOI 10.1093/neuonc/noaa024
   Ryall S, 2020, CANCER CELL, V37, P569, DOI 10.1016/j.ccell.2020.03.011
   Ryall S, 2020, ACTA NEUROPATHOL COM, V8, DOI 10.1186/s40478-020-00902-z
   Sahm F, 2016, JNCI-J NATL CANCER I, V108, DOI 10.1093/jnci/djv377
   Schramm J, 2004, NEUROSURGERY, V55, P340, DOI 10.1227/01.NEU.0000129546.38675.1B
   Shakur SF, 2009, J NEUROSURG-PEDIATR, V3, P197, DOI 10.3171/2008.11.PEDS0858
   Shirahata M, 2018, ACTA NEUROPATHOL, V136, P153, DOI 10.1007/s00401-018-1849-4
   Sievers P, 2021, NEURO-ONCOLOGY, V23, P34, DOI 10.1093/neuonc/noaa251
   Slegers RJ, 2020, ACTA NEUROPATHOL COM, V8, DOI 10.1186/s40478-020-00904-x
   Smits M, 2017, RADIOLOGY, V284, P316, DOI 10.1148/radiol.2017151930
   Smits M, 2016, BRIT J RADIOL, V89, DOI 10.1259/bjr.20150857
   Solomon DA, 2016, BRAIN PATHOL, V26, P569, DOI 10.1111/bpa.12336
   Stichel D, 2018, ACTA NEUROPATHOL, V136, P793, DOI 10.1007/s00401-018-1905-0
   Stucklin ASG, 2019, NAT COMMUN, V10, DOI 10.1038/s41467-019-12187-5
   Sturm D, 2016, CELL, V164, P1060, DOI 10.1016/j.cell.2016.01.015
   Suchorska B, 2019, EUR J CANCER, V107, P15, DOI 10.1016/j.ejca.2018.10.019
   Suzuki H, 2015, NAT GENET, V47, P458, DOI 10.1038/ng.3273
   Swanson AA, 2019, J NEUROPATH EXP NEUR, V78, P791, DOI 10.1093/jnen/nlz064
   Tatevossian RG, 2010, ACTA NEUROPATHOL, V120, P731, DOI 10.1007/s00401-010-0763-1
   Tauziede-Espariat A, 2020, ACTA NEUROPATHOL COM, V8, DOI 10.1186/s40478-020-00974-x
   Tauziede-Espariat A, 2019, ACTA NEUROPATHOL COM, V7, DOI 10.1186/s40478-019-0738-y
   Taylor MD, 2012, ACTA NEUROPATHOL, V123, P465, DOI 10.1007/s00401-011-0922-z
   Tesileanu CMS, 2020, NEURO-ONCOLOGY, V22, P515, DOI 10.1093/neuonc/noz200
   Thomas C, 2020, ACTA NEUROPATHOL, V139, P277, DOI 10.1007/s00401-019-02094-w
   U-King-Im JM, 2010, CHILD NERV SYST, V26, P1765, DOI 10.1007/s00381-010-1251-6
   Upadhyaya SA, 2019, NEURO-ONCOLOGY, V21, P1319, DOI 10.1093/neuonc/noz069
   Valera ET, 2020, PEDIATR BLOOD CANCER, V67, DOI 10.1002/pbc.28015
   van den Bent MJ, 2011, LANCET ONCOL, V12, P583, DOI 10.1016/S1470-2045(11)70057-2
   Vettermann FJ, 2018, CLIN NUCL MED, V43, P895, DOI 10.1097/RLU.0000000000002300
   Wang M, 2005, J NEUROPATH EXP NEUR, V64, P875, DOI 10.1097/01.jnen.0000182981.02355.10
   Weaver KJ, 2017, J NEUROSURG-PEDIATR, V20, P347, DOI 10.3171/2017.5.PEDS16402
   Wefers AK, 2020, ACTA NEUROPATHOL, V139, P193, DOI 10.1007/s00401-019-02078-w
   Whitehead MT, 2015, ANTICANCER RES, V35, P6267
   WHO Classification of Tumours Editorial Board, 2021, CENTRAL NERVOUS SYST, V6
   Wijnenga MMJ, 2018, NEURO-ONCOLOGY, V20, P103, DOI 10.1093/neuonc/nox176
   Witt H, 2018, NEURO-ONCOLOGY, V20, P1616, DOI 10.1093/neuonc/noy118
   Witt H, 2011, CANCER CELL, V20, P143, DOI 10.1016/j.ccr.2011.07.007
   Yamaguchi M, 2016, PATHOL INT, V66, P34, DOI 10.1111/pin.12366
   Yeom KW, 2013, AM J ROENTGENOL, V200, P895, DOI 10.2214/AJR.12.9249
   Yonezawa U, 2020, WORLD NEUROSURG, V140, pE320, DOI 10.1016/j.wneu.2020.05.063
   Yoshida Y, 2018, BRAIN PATHOL, V28, P710, DOI 10.1111/bpa.12585
   Yoshimoto K, 2017, BRAIN TUMOR PATHOL, V34, P103, DOI 10.1007/s10014-017-0287-7
   Zaytseva M, 2021, CANCERS, V13, DOI 10.3390/cancers13194954
   Zetterling M, 2017, PLOS ONE, V12, DOI 10.1371/journal.pone.0188419
   Zhang C, 2020, ACTA NEUROPATHOL COM, V8, DOI 10.1186/s40478-020-01038-w
   Zhang JH, 2013, NAT GENET, V45, P602, DOI 10.1038/ng.2611
   Zhao K, 2021, ACAD RADIOL, V28, pE189, DOI 10.1016/j.acra.2020.03.008
NR 191
TC 11
Z9 11
U1 2
U2 8
PU SPRINGER
PI NEW YORK
PA ONE NEW YORK PLAZA, SUITE 4600, NEW YORK, NY, UNITED STATES
SN 0028-3940
EI 1432-1920
J9 NEURORADIOLOGY
JI Neuroradiology
PD OCT
PY 2022
VL 64
IS 10
BP 1919
EP 1950
DI 10.1007/s00234-022-03008-6
EA JUL 2022
PG 32
WC Clinical Neurology; Neuroimaging; Radiology, Nuclear Medicine & Medical
   Imaging
WE Science Citation Index Expanded (SCI-EXPANDED)
SC Neurosciences & Neurology; Radiology, Nuclear Medicine & Medical Imaging
GA 4N2UY
UT WOS:000828980800001
PM 35869291
DA 2023-08-27
ER

PT J
AU Fukuda, H
   Takahashi, J
   Watanabe, K
   Hayashi, H
   Morizane, A
   Koyanagi, M
   Sasai, Y
   Hashimoto, N
AF Fukuda, Hitosih
   Takahashi, Jun
   Watanabe, Khchi
   Hayashi, Hidekt
   Morizane, Asuka
   Koyanagi, Masaomi
   Sasai, Yoshiki
   Hashimoto, Nobuo
TI Fluorescence-activated cell sorting-based purification of embryonic stem
   cell-derived neural precursors averts tumor formation after
   transplantation
SO STEM CELLS
LA English
DT Article
DE fluorescence-activated cell sorting; embryonic stem cell; Sox1;
   teratoma; transplantation; dopaminergic neuron
ID MIDBRAIN DOPAMINERGIC-NEURONS; IN-VITRO; ES CELLS; EXPRESSION; GENES;
   BLASTOCYSTS; PROGENITORS; GENERATION; INDUCTION; DEVELOP
AB The differentiation of dopaminergic (DA) neurons from mouse embryonic stem cells (ESCs) can be efficiently induced, making these neurons a potential source for transplantation as a treatment for Parkinson's disease, a condition characterized by the gradual loss of midbrain DA neurons. One of the major persistent obstacles to the successful implementation of therapeutic ESC transplantation is the propensity of ESC-derived grafts to form tumors in vivo. To address this problem, we used fluorescence-activated cell sorting to purify mouse ESC-derived neural pre-cursors expressing the neural precursor marker Sox-1. ESC-derived, Sox1(+) cells began to express neuronal cell markers and differentiated into DA neurons upon transplantation into mouse brains but did not generate tumors in this site. In contrast, Sox1(-) cells that expressed ESC markers frequently formed tumors in vivo. These results indicate that Sox1-based cell sorting of neural precursors prevents graft-derived tumor formation after transplantation, providing a promising strategy for cell transplantation therapy of neurodegenerative disorders.
C1 Kyoto Univ, Grad Sch Med, Dept Neurosurg, Sakyo Ku, Kyoto 6068507, Japan.
   RIKEN, Ctr Dev Biol, Organogenesis & Neurogenesis Grp, Kobe, Hyogo, Japan.
C3 Kyoto University; RIKEN
RP Takahashi, J (通讯作者)，Kyoto Univ, Grad Sch Med, Dept Neurosurg, Sakyo Ku, 54 Shogoinkawahara Cho, Kyoto 6068507, Japan.
EM
RI Watanabe, Kiichi/ABA-4854-2020; Koyanagi, Masaomi/AAN-1663-2020
OI Koyanagi, Masaomi/0000-0002-1661-2082; Morizane,
   Asuka/0000-0001-7411-1828
CR ABERCROMBIE M, 1946, ANAT REC, V94, P239, DOI 10.1002/ar.1090940210
   Aubert J, 2003, P NATL ACAD SCI USA, V100, P11836, DOI 10.1073/pnas.1734197100
   Barberi T, 2003, NAT BIOTECHNOL, V21, P1200, DOI 10.1038/nbt870
   Ben-Hur T, 2004, STEM CELLS, V22, P1246, DOI 10.1634/stemcells.2004-0094
   BJORKLUND A, 1983, ACTA PHYSIOL SCAND, P1
   Bjorklund LM, 2002, P NATL ACAD SCI USA, V99, P2344, DOI 10.1073/pnas.022438099
   Brundin P, 1988, Brain Res, V467, P233
   D'Amour KA, 2003, P NATL ACAD SCI USA, V100, P11866, DOI 10.1073/pnas.1834200100
   Erdo F, 2003, J CEREBR BLOOD F MET, V23, P780, DOI 10.1097/01.WCB.0000071886.63724.FB
   Harkany T, 2004, J NEUROCHEM, V88, P1229, DOI 10.1046/j.1471-4159.2003.02243.x
   Kawasaki H, 2000, NEURON, V28, P31, DOI 10.1016/S0896-6273(00)00083-0
   Kim JH, 2002, NATURE, V418, P50, DOI 10.1038/nature00900
   Lee SH, 2000, NAT BIOTECHNOL, V18, P675, DOI 10.1038/76536
   LENDAHL U, 1990, CELL, V60, P585, DOI 10.1016/0092-8674(90)90662-X
   Lenka N, 2002, J CELL SCI, V115, P1471
   Levy G, 2002, TRANSPLANTATION, V73, P953, DOI 10.1097/00007890-200203270-00022
   Mitsui K, 2003, CELL, V113, P631, DOI 10.1016/S0092-8674(03)00393-3
   Morizane A, 2002, J NEUROSCI RES, V69, P934, DOI 10.1002/jnr.10363
   MOUNTFORD P, 1994, P NATL ACAD SCI USA, V91, P4303, DOI 10.1073/pnas.91.10.4303
   Nishimura F, 2003, STEM CELLS, V21, P171, DOI 10.1634/stemcells.21-2-171
   Nishino H, 2000, EXP NEUROL, V164, P209, DOI 10.1006/exnr.2000.7426
   NIWA H, 1991, GENE, V108, P193, DOI 10.1016/0378-1119(91)90434-D
   Paxinos, 2008, MOUSE BRAIN STEREOTA
   Perrier AL, 2004, P NATL ACAD SCI USA, V101, P12543, DOI 10.1073/pnas.0404700101
   Pevny LH, 1998, DEVELOPMENT, V125, P1967
   Reubinoff BE, 2001, NAT BIOTECHNOL, V19, P1134, DOI 10.1038/nbt1201-1134
   Schierle GS, 1999, NAT MED, V5, P97, DOI 10.1038/4785
   Simon HH, 2001, J NEUROSCI, V21, P3126, DOI 10.1523/JNEUROSCI.21-09-03126.2001
   Suemori H, 2001, DEV DYNAM, V222, P273, DOI 10.1002/dvdy.1191
   Takahashi K, 2003, NATURE, V423, P541, DOI 10.1038/nature01646
   Thomson JA, 1998, SCIENCE, V282, P1145, DOI 10.1126/science.282.5391.1145
   Wood HB, 1999, MECH DEVELOP, V86, P197, DOI 10.1016/S0925-4773(99)00116-1
   Yeom YI, 1996, DEVELOPMENT, V122, P881
   Ying QL, 2003, NAT BIOTECHNOL, V21, P183, DOI 10.1038/nbt780
   Zhang SC, 2001, NAT BIOTECHNOL, V19, P1129, DOI 10.1038/nbt1201-1129
NR 35
TC 128
Z9 142
U1 0
U2 9
PU WILEY
PI HOBOKEN
PA 111 RIVER ST, HOBOKEN 07030-5774, NJ USA
SN 1066-5099
EI 1549-4918
J9 STEM CELLS
JI Stem Cells
PD MAR
PY 2006
VL 24
IS 3
BP 763
EP 771
DI 10.1634/stemcells.2005-0137
PG 9
WC Cell & Tissue Engineering; Biotechnology & Applied Microbiology;
   Oncology; Cell Biology; Hematology
WE Science Citation Index Expanded (SCI-EXPANDED)
SC Cell Biology; Biotechnology & Applied Microbiology; Oncology; Hematology
GA 085WX
UT WOS:000240636200032
PM 16223855
OA Bronze
DA 2023-08-27
ER

PT J
AU Mayr, L
   Gojo, J
   Peyrl, A
   Azizi, AA
   Stepien, NM
   Pletschko, T
   Czech, T
   Dorfer, C
   Lambo, S
   Dieckmann, K
   Haberler, C
   Kool, M
   Slavc, I
AF Mayr, Lisa
   Gojo, Johannes
   Peyrl, Andreas
   Azizi, Amedeo A.
   Stepien, Natalia M.
   Pletschko, Thomas
   Czech, Thomas
   Dorfer, Christian
   Lambo, Sander
   Dieckmann, Karin
   Haberler, Christine
   Kool, Marcel
   Slavc, Irene
TI Potential Importance of Early Focal Radiotherapy Following Gross Total
   Resection for Long-Term Survival in Children With Embryonal Tumors With
   Multilayered Rosettes
SO FRONTIERS IN ONCOLOGY
LA English
DT Article
DE embryonal tumor with multilayered rosette; embryonal tumor with abundant
   neuropil and true rosette; radiotherapy; focal radiotherapy; intrathecal
   therapy; embryonal brain tumors
ID CENTRAL-NERVOUS-SYSTEM; INTRATHECAL LIPOSOMAL CYTARABINE; ABUNDANT
   NEUROPIL; TRUE ROSETTES; HIT 2000; MEDULLOEPITHELIOMA; MEDULLOBLASTOMA;
   CLASSIFICATION; CHEMOTHERAPY; THERAPY
AB Embryonal tumor with multilayered rosettes (ETMR) is a rare, aggressive embryonal central nervous system tumor characterized by LIN28A expression and alterations in the C19MC locus. ETMRs predominantly occur in young children, have a dismal prognosis, and no definitive treatment guidelines have been established. We report on nine consecutive patients and review the role of initiation/timing of radiotherapy on survival. Between 2006 and 2018, nine patients were diagnosed with ETMR. Diagnosis was confirmed histopathologically, immunohistochemically and molecularly. Median age was 25 months (5-38). Location was supratentorial in five, pineal in three, and brainstem in one. Seven patients had a gross total resection, one a partial resection and one a biopsy at initial diagnosis. Chemotherapy augmented with intrathecal therapy started a median of 10 days (7-20) after surgery. Only two patients who after gross total resection received radiotherapy very early on (six weeks after diagnosis) are alive and in complete remission 56 and 50 months after diagnosis. All remaining patients for whom radiotherapy was deferred until the end of chemotherapy recurred, albeit none with leptomeningeal disease. A literature research identified 228 patients with ETMR. Including our patients only 26 (11%) of 237 patients survived >36 months with no evidence of disease at last follow-up. All but two long-term (>36 months) survivors received radiotherapy, ten of whom early on following gross total resection (GTR). GTR followed by early focal radiotherapy and intrathecal therapy to prevent leptomeningeal disease are potentially important to improve survival of ETMR in the absence of effective targeted therapies.
C1 [Mayr, Lisa; Gojo, Johannes; Peyrl, Andreas; Azizi, Amedeo A.; Stepien, Natalia M.; Pletschko, Thomas; Slavc, Irene] Med Univ Vienna, Dept Pediat & Adolescent Med, Vienna, Austria.
   [Mayr, Lisa; Gojo, Johannes; Peyrl, Andreas; Azizi, Amedeo A.; Stepien, Natalia M.; Pletschko, Thomas; Slavc, Irene] Med Univ Vienna, Comprehens Ctr Pediat, Vienna, Austria.
   [Czech, Thomas; Dorfer, Christian] Med Univ Vienna, Dept Neurosurg, Vienna, Austria.
   [Lambo, Sander; Kool, Marcel] Hopp Childrens Canc Ctr Heidelberg KiTZ, Div Pediat Neurooncol, Heidelberg, Germany.
   [Lambo, Sander; Kool, Marcel] German Canc Res Ctr, Div Pediat Neurooncol, Heidelberg, Germany.
   [Lambo, Sander; Kool, Marcel] German Canc Consortium DKTK, Heidelberg, Germany.
   [Dieckmann, Karin] Med Univ Vienna, Dept Radiotherapy, Vienna, Austria.
   [Haberler, Christine] Med Univ Vienna, Dept Neurol, Div Neuropathol & Neurochem, Vienna, Austria.
   [Kool, Marcel] Princess Maxima Ctr Pediat Oncol, Res Dept, Utrecht, Netherlands.
C3 Medical University of Vienna; Medical University of Vienna; Medical
   University of Vienna; Helmholtz Association; German Cancer Research
   Center (DKFZ); Helmholtz Association; German Cancer Research Center
   (DKFZ); Medical University of Vienna; Medical University of Vienna
RP Slavc, I (通讯作者)，Med Univ Vienna, Dept Pediat & Adolescent Med, Vienna, Austria.; Slavc, I (通讯作者)，Med Univ Vienna, Comprehens Ctr Pediat, Vienna, Austria.
EM
RI Dorfer, Christian/ACX-8758-2022; Slavc, Irene/IQV-7519-2023
OI Dorfer, Christian/0000-0002-1843-7732; Gojo,
   Johannes/0000-0002-8113-3416; czech, thomas/0000-0001-8112-2795
FU Austrian Science Fund within the TRANSCAN-2 project "BRCAddict" [I
   4164]; Solving Kid's Cancer foundation; Bibi Fund for Childhood Cancer
   Research
FX This study was supported by the Austrian Science Fund (I 4164 to J.G.)
   within the TRANSCAN-2 project "BRCAddict". SL and MK are supported by
   the Solving Kid's Cancer foundation and the Bibi Fund for Childhood
   Cancer Research.
CR Alexiou GA, 2013, J CHILD NEUROL, V28, P1709, DOI 10.1177/0883073812471434
   [Anonymous], 2010, BEERY VMI SUPPLEMENT
   [Anonymous], 2013, VERHALTENSINVENTAR B
   Cairo MS, 2012, J CLIN ONCOL, V30, P387, DOI 10.1200/JCO.2010.33.3369
   Capper D, 2018, NATURE, V555, P469, DOI 10.1038/nature26000
   Dhall G, 2008, PEDIATR BLOOD CANCER, V50, P1169, DOI 10.1002/pbc.21525
   Ding DL, 2014, J NEUROSURG-PEDIATR, V13, P658, DOI 10.3171/2014.3.PEDS13405
   Drechsler R., 2013, VERHALTENSINVENTAR B
   Dufour C, 2014, PEDIATR BLOOD CANCER, V61, P1398, DOI 10.1002/pbc.25009
   Eaton BR, 2020, NEURO-ONCOLOGY, V22, P1379, DOI 10.1093/neuonc/noaa042
   Gerber NU, 2014, INT J RADIAT ONCOL, V89, P863, DOI 10.1016/j.ijrobp.2014.04.017
   Gessi M, 2009, AM J SURG PATHOL, V33, P211, DOI 10.1097/PAS.0b013e318186235b
   Glei<ss>ner U, 2011, FRAGEBOGEN ERFASSUNG
   Goodman R, 2000, BRIT J PSYCHIAT, V177, P534, DOI 10.1192/bjp.177.6.534
   Grob A., 2018, INTELLIGENCE DEV SCA
   Helmstaedter C, 2001, VERBALER LERN MERKFA
   Horwitz M, 2016, CHILD NERV SYST, V32, P299, DOI 10.1007/s00381-015-2920-2
   Jaramillo S, 2019, CLIN TRANSL RAD ONCO, V15, P31, DOI 10.1016/j.ctro.2018.11.002
   Kastner-Koller U., 2012, WIENER ENTWICKLUNGST
   Korshunov A, 2014, ACTA NEUROPATHOL, V128, P279, DOI 10.1007/s00401-013-1228-0
   Kram David E., 2018, Bioengineering-Basel, V5, P78, DOI 10.3390/bioengineering5040078
   Lambo S, 2019, NATURE, V576, P274, DOI 10.1038/s41586-019-1815-x
   Louis DN, 2016, ACTA NEUROPATHOL, V131, P803, DOI 10.1007/s00401-016-1545-1
   Manjila S, 2011, NEUROSURG FOCUS, V30, DOI 10.3171/2010.10.FOCUS10226
   Matsumoto M, 2007, NEUROL MED-CHIR, V47, P428, DOI 10.2176/nmc.47.428
   Mozes P, 2016, J NEURO-ONCOL, V126, P99, DOI 10.1007/s11060-015-1938-3
   Muller K, 2011, STRAHLENTHER ONKOL, V187, P757, DOI 10.1007/s00066-011-2256-0
   Pajtler KW, 2016, J NEURO-ONCOL, V128, P463, DOI 10.1007/s11060-016-2133-x
   Paulus W, 2010, ACTA NEUROPATHOL, V120, P269, DOI 10.1007/s00401-010-0710-1
   Petermann F., 2018, WECHSLER PRESCHOOL P
   Peyrl A, 2014, CLIN PHARMACOKINET, V53, P165, DOI 10.1007/s40262-013-0106-1
   Peyrl A, 2009, CLIN PHARMACOKINET, V48, P265, DOI 10.2165/00003088-200948040-00004
   Pfister S, 2009, ACTA NEUROPATHOL, V117, P457, DOI 10.1007/s00401-008-0467-y
   Potter SLP, 2012, PEDIATR BLOOD CANCER, V58, P362, DOI 10.1002/pbc.23317
   Ravens-Sieberer U, 1998, QUAL LIFE RES, V7, P399, DOI 10.1023/A:1008853819715
   Schmidt C, 2017, NEURO-ONCOLOGY, V19, P1607, DOI 10.1093/neuonc/nox093
   Schuhfried GmbH, 2007, MOT LEIST SER MLS
   Sin-Chan P, 2019, CANCER CELL, V36, P51, DOI 10.1016/j.ccell.2019.06.002
   Slavc I, 2003, J NEURO-ONCOL, V64, P239, DOI 10.1023/A:1025633704071
   Slavc I, 2014, CANCER MED-US, V3, P91, DOI 10.1002/cam4.161
   Spence T, 2014, ACTA NEUROPATHOL, V128, P291, DOI 10.1007/s00401-014-1291-1
   Sturm D, 2016, CELL, V164, P1060, DOI 10.1016/j.cell.2016.01.015
   von Bueren AO, 2011, NEURO-ONCOLOGY, V13, P669, DOI 10.1093/neuonc/nor025
   Wang B, 2018, J NEUROIMAGING, V28, P483, DOI 10.1111/jon.12524
   Wechsler, 2017, WISC V WECHSLER INTE
   Wen PY, 2010, J CLIN ONCOL, V28, P1963, DOI 10.1200/JCO.2009.26.3541
   Woehrer A, 2011, ACTA NEUROPATHOL, V122, P787, DOI 10.1007/s00401-011-0903-2
   Zimmermann P, 2017, TESTBATTERIE AUFMERK
NR 48
TC 9
Z9 9
U1 0
U2 1
PU FRONTIERS MEDIA SA
PI LAUSANNE
PA AVENUE DU TRIBUNAL FEDERAL 34, LAUSANNE, CH-1015, SWITZERLAND
SN 2234-943X
J9 FRONT ONCOL
JI Front. Oncol.
PD DEC 17
PY 2020
VL 10
AR 584681
DI 10.3389/fonc.2020.584681
PG 10
WC Oncology
WE Science Citation Index Expanded (SCI-EXPANDED)
SC Oncology
GA PM2EF
UT WOS:000603618700001
PM 33392079
OA gold, Green Published
DA 2023-08-27
ER

PT J
AU Ohira, S
   Ikawa, T
   Kanayama, N
   Inui, S
   Ueda, Y
   Miyazaki, M
   Nishio, T
   Koizumi, M
   Konishi, K
AF Ohira, Shingo
   Ikawa, Toshiki
   Kanayama, Naoyuki
   Inui, Shoki
   Ueda, Yoshihiro
   Miyazaki, Masayoshi
   Nishio, Teiji
   Koizumi, Masahiko
   Konishi, Koji
TI Dose reduction of hippocampus using HyperArc planning in postoperative
   radiotherapy for primary brain tumors
SO MEDICAL DOSIMETRY
LA English
DT Article
DE Primary brain tumor; HyperArc; Hippocampus
ID RADIATION-THERAPY; CONFORMAL RADIOTHERAPY; GRADE GLIOMA; NECK-CANCER;
   TOXICITY; BENIGN; HEAD
AB To compare dosimetric parameters for the hippocampus, organs at risk (OARs), and targets of volumetric modulated arc therapy (VMAT), noncoplanar VMAT (NC-VMAT), and HyperArc (HA) plans in patients un-dergoing postoperative radiotherapy for primary brain tumors. For 20 patients, HA plans were generated to deliver 40.05 to 60 Gy for the planning target volume (PTV). In addition, doses for the hippocam-pus and OARs were minimized. The VMAT and NC-VMAT plans were retrospectively generated using the same optimization parameters as those in the HA plans. For the hippocampus, the equivalent dose to be administered in 2 Gy fractions (EQD2) was calculated assuming alpha/beta = 2. Dosimetric parameters for the PTV, hippocampus, and OARs in the VMAT, NC-VMAT, and HA plans were compared. For PTV, the HA plans provided significantly lower Dmax and D 1% than the VMAT and NC-VMAT plans ( p < 0.05), whereas the D 99% and Dmin were significantly higher ( p < 0.05). For the contralateral hippocampus, the dosimetric parameters in the HA plans (8.1 +/- 9.6, 6.5 +/- 7.2, 5.6 +/- 5.8, and 4.8 +/- 4.7 Gy for D 20% , D 40% , D 60% and D 80% , respectively) were significantly smaller ( p < 0.05) than those in the VMAT and NC-VMAT plans. Except for the optic chiasm, the Dmax in the HA plans (brainstem, lens, optic nerves, and retinas) was the smallest ( p < 0.05). In addition, the doses in the HA plans for the brain and skin were the small-est ( p < 0.05) among the 3 plans. HA planning, instead of coplanar and noncoplanar VMAT, significantly reduces the dosage to which the contralateral hippocampus as well as other OARs are exposed without compromising on target coverage.(c) 2022 American Association of Medical Dosimetrists. Published by Elsevier Inc. All rights reserved.
C1 [Ohira, Shingo; Ikawa, Toshiki; Kanayama, Naoyuki; Inui, Shoki; Ueda, Yoshihiro; Miyazaki, Masayoshi; Konishi, Koji] Osaka Int Canc Inst, Dept Radiat Oncol, Osaka, Japan.
   [Ohira, Shingo; Nishio, Teiji; Koizumi, Masahiko] Osaka Univ, Dept Med Phys & Engn, Grad Sch Med, Suita, Japan.
   [Ohira, Shingo] Osaka Int Canc Inst, Dept Radiat Oncol, 3-1-69 Otemae,Chuo Ku, Osaka, Osaka 5378567, Japan.
C3 Osaka University
RP Ohira, S (通讯作者)，Osaka Int Canc Inst, Dept Radiat Oncol, 3-1-69 Otemae,Chuo Ku, Osaka, Osaka 5378567, Japan.
EM
OI IKAWA, TOSHIKI/0000-0001-9097-0227; Ohira, Shingo/0000-0002-6170-1471;
   Kanayama, Naoyuki/0000-0003-0016-2213
FU JSPS KAKENHI [21K07742]
FX This study was supported by JSPS KAKENHI Grant (Grant -in-Aid for
   Scientific Research (C) 21K07742) .
CR Borges C, 2012, MED PHYS, V39, P415, DOI 10.1118/1.3671935
   Boychak A, 2016, AM J CLIN ONCOL-CANC, V39, P160, DOI 10.1097/COC.0000000000000034
   Buckner JC, 2016, NEW ENGL J MED, V374, P1344, DOI 10.1056/NEJMoa1500925
   Bush NAO, 2016, J ONCOL PRACT, V12, P1235, DOI 10.1200/JOP.2016.018622
   Chan JL, 2002, J CLIN ONCOL, V20, P1635, DOI 10.1200/JCO.20.6.1635
   Chang EL, 2009, LANCET ONCOL, V10, P1037, DOI 10.1016/S1470-2045(09)70263-3
   Cheung Eva Yi Wah, 2021, BJR Open, V3, P20210009, DOI 10.1259/bjro.20210009
   De Maria L, 2021, FRONT ONCOL, V11, DOI 10.3389/fonc.2021.652646
   de Robles P, 2015, NEURO-ONCOLOGY, V17, P776, DOI 10.1093/neuonc/nou283
   DEANGELIS LM, 1989, NEUROLOGY, V39, P789, DOI 10.1212/WNL.39.6.789
   Feuvret L, 2006, INT J RADIAT ONCOL, V64, P333, DOI 10.1016/j.ijrobp.2005.09.028
   Ghia A, 2007, INT J RADIAT ONCOL, V68, P971, DOI 10.1016/j.ijrobp.2007.02.016
   Gondi V, 2012, INT J RADIAT ONCOL, V83, pE487, DOI [10.1016/j.ijrobp.2011.10.021, 10.1016/j.ijrobp.2012.11.031]
   Guy JB, 2016, MED DOSIM, V41, P9, DOI 10.1016/j.meddos.2015.06.002
   Hofmaier J, 2016, RADIAT ONCOL, V11, DOI 10.1186/s13014-016-0695-6
   Inui S, 2020, RADIAT ONCOL, V15, DOI 10.1186/s13014-020-01614-3
   Jalali R, 2010, INT J RADIAT ONCOL, V77, P974, DOI 10.1016/j.ijrobp.2009.06.025
   Kazda T, 2014, RADIAT ONCOL, V9, DOI 10.1186/1748-717X-9-139
   Knutson NC, 2020, ADV RADIAT ONCOL, V5, P482, DOI 10.1016/j.adro.2020.01.003
   Liu HF, 2022, MED DOSIM, V47, P258, DOI 10.1016/j.meddos.2022.04.003
   Mayo C, 2010, INT J RADIAT ONCOL, V76, pS28, DOI 10.1016/j.ijrobp.2009.07.1753
   Mayo C, 2010, INT J RADIAT ONCOL, V76, pS36, DOI 10.1016/j.ijrobp.2009.08.078
   Mohile NA, 2022, J CLIN ONCOL, V40, P403, DOI 10.1200/JCO.21.02036
   Mollerberg Marie-Louise, 2021, Tech Innov Patient Support Radiat Oncol, V19, P11, DOI 10.1016/j.tipsro.2021.05.001
   Munch S, 2016, STRAHLENTHER ONKOL, V192, P722, DOI 10.1007/s00066-016-1020-x
   Nevelsky A, 2013, J APPL CLIN MED PHYS, V14, P113, DOI 10.1120/jacmp.v14i3.4205
   O'Shea A, 2016, FRONT AGING NEUROSCI, V8, DOI 10.3389/fnagi.2016.00298
   Obara T, 2020, FRONT ONCOL, V10, DOI 10.3389/fonc.2020.574679
   Ohira S, 2019, PHYS MEDICA, V60, P83, DOI 10.1016/j.ejmp.2019.03.022
   Ohira S, 2018, RADIOTHER ONCOL, V129, P105, DOI 10.1016/j.radonc.2018.01.012
   Ohira S, 2018, RADIAT ONCOL, V13, DOI 10.1186/s13014-017-0948-z
   Pasciuti K, 2017, MED DOSIM, V42, P1, DOI 10.1016/j.meddos.2016.09.003
   Phillips GS, 2020, JAMA DERMATOL, V156, P963, DOI 10.1001/jamadermatol.2020.2127
   Popple RA, 2021, PRACT RADIAT ONCOL, V11, pE163, DOI 10.1016/j.prro.2020.10.013
   Porock D, 2002, EUR J CANCER CARE, V11, P33, DOI 10.1046/j.1365-2354.2002.00287.x
   de Dios NR, 2021, J CLIN ONCOL, V39, P3118, DOI 10.1200/JCO.21.00639
   Sprowls CJ, 2021, MED DOSIM, V46, P264, DOI 10.1016/j.meddos.2021.02.007
   Stupp R, 2014, ANN ONCOL, V25, P93, DOI 10.1093/annonc/mdu050
   Van den Bosch L, 2021, RADIOTHER ONCOL, V157, P147, DOI 10.1016/j.radonc.2021.01.024
   Woods KE, 2022, CANCERS, V14, DOI 10.3390/cancers14040939
NR 40
TC 0
Z9 0
U1 2
U2 2
PU ELSEVIER SCIENCE INC
PI NEW YORK
PA STE 800, 230 PARK AVE, NEW YORK, NY 10169 USA
SN 0958-3947
EI 1873-4022
J9 MED DOSIM
JI Med. Dosim.
PY 2023
VL 48
IS 2
BP 67
EP 72
DI 10.1016/j.meddos.2022.12.001
EA APR 2023
PG 6
WC Oncology; Radiology, Nuclear Medicine & Medical Imaging
WE Science Citation Index Expanded (SCI-EXPANDED)
SC Oncology; Radiology, Nuclear Medicine & Medical Imaging
GA G9QX6
UT WOS:000992426600001
PM 36653285
OA Bronze
DA 2023-08-27
ER

PT J
AU Plowman, PN
   Saunders, CAB
   Maisey, M
AF Plowman, PN
   Saunders, CAB
   Maisey, M
TI On the usefulness of brain PET scanning to the paediatric
   neuro-oncologist
SO BRITISH JOURNAL OF NEUROSURGERY
LA English
DT Article
DE PET scanning; paediatric brain tumours; recurrent tumour
ID TUMORS; FDG
AB Positron emission tomography (PET) scanning adds a functional dimension to brain scanning; levels of metabolic activity are imaged and this information may complement the more "anatomic" imaging of CT and MRI. In a series of 10 young patients the usefulness of PET scanning technique was investigated. The major areas of usefulness were the distinction of posttreatment sequelae from active tumour (both postsurgical MRI changes from tumour and postradiation MRI changes from tumour) and the localization of persisting tumour amenable to radiosurgical treatment. The technique was beneficial in assessing continuing activity in pineal tumours (residual pineal teratoma mass and residual pineocytoma mass) and in assessing activity in brainstem/cerebellar peduncle gliomas (three cases). In one unusual case of widespread leptomeningeal melanoma, the PET scan under-read the situation.
C1 Hosp Sick Children, Dept Haematol & Oncol, London WC1, England.
   United Med & Dent Sch Guys & St Thomas Hosp, Clin PET Ctr, London SE1 9RT, England.
C3 University of London; University College London; Great Ormond Street
   Hospital for Children NHS Foundation Trust; University of London; King's
   College London
RP Plowman, PN (通讯作者)，St Bartholomews Hosp, Dept Radiotherapy, London EC14 7BE, England.
OI Saunders, Catherine AB/0000-0001-6928-7378
CR BRUGGERS CS, 1993, MED PEDIATR ONCOL, V21, P301, DOI 10.1002/mpo.2950210414
   DICHIRO G, 1982, NEUROLOGY, V32, P1328
   ERICSON K, 1993, DEV NUC MED, V23, P215
   FEENEY DM, 1976, STROKE, V82, P822
   HAWKINS RA, 1991, J NUCL MED, V32, P1555
   HOFFMAN JM, 1992, J COMPUT ASSIST TOMO, V16, P62, DOI 10.1097/00004728-199201000-00011
   HOLTHOFF VA, 1993, CANCER, V72, P1394, DOI 10.1002/1097-0142(19930815)72:4<1394::AID-CNCR2820720440>3.0.CO;2-D
   KHAYAT D, 1993, J CLIN ONCOL, V11, P2173, DOI 10.1200/JCO.1993.11.11.2173
   LACIC M, UNPUB 18FDG 11C METH
   OTUAMA LA, 1990, PEDIATR NEUROL, V6, P163, DOI 10.1016/0887-8994(90)90057-8
   PATRONAS NJ, 1982, RADIOLOGY, V144, P885, DOI 10.1148/radiology.144.4.6981123
   PLOWMAN PN, 1984, INT J RADIAT ONCOL, V10, P843, DOI 10.1016/0360-3016(84)90386-9
   PLOWMAN PN, 1997, CLIN ONCOL, V10, P577
   VALK PE, 1988, J NEUROSURG, V69, P830, DOI 10.3171/jns.1988.69.6.0830
NR 14
TC 16
Z9 17
U1 0
U2 0
PU INFORMA HEALTHCARE
PI LONDON
PA TELEPHONE HOUSE, 69-77 PAUL STREET, LONDON EC2A 4LQ, ENGLAND
SN 0268-8697
J9 BRIT J NEUROSURG
JI Br. J. Neurosurg.
PD DEC
PY 1997
VL 11
IS 6
BP 525
EP 532
PG 8
WC Clinical Neurology; Surgery
WE Science Citation Index Expanded (SCI-EXPANDED)
SC Neurosciences & Neurology; Surgery
GA YQ367
UT WOS:000071379400006
PM 11013624
DA 2023-08-27
ER

PT J
AU JACKS, T
   SHIH, TS
   SCHMITT, EM
   BRONSON, RT
   BERNARDS, A
   WEINBERG, RA
AF JACKS, T
   SHIH, TS
   SCHMITT, EM
   BRONSON, RT
   BERNARDS, A
   WEINBERG, RA
TI TUMOR PREDISPOSITION IN MICE HETEROZYGOUS FOR A TARGETED MUTATION IN NF1
SO NATURE GENETICS
LA English
DT Article
ID NEUROFIBROMATOSIS TYPE-1 GENE; TRANSGENIC MOUSE MODEL; NEURAL CREST;
   CELL-LINES; PROTEIN PRODUCT; CDNA SEQUENCE; MESSENGER-RNA; STEM-CELLS;
   IDENTIFICATION; GAP
AB Human neurofibromatosis type 1 is a dominant disease caused by the inheritance of a mutant allele of the NF1 gene. In order to study NF1 function, we have constructed a mouse strain carrying a germline mutation in the murine homologue. Heterozygous animals do not exhibit the classical symptoms of the human disease, but are highly predisposed to the formation of various tumour types, notably phaeochomocytoma, a tumour of the neural crest-derived adrenal medulla, and myeloid leukaemia, both of which occur with increased frequency in human NF1 patients. The wild-type Nf1 allele is lost in approximately half of the tumours from heterozygous animals. In addition, homozygosity for the Nf1 mutation leads to abnormal cardiac development and midgestational embryonic lethality.
C1 MIT, DEPT BIOL, CAMBRIDGE, MA 02139 USA.
   TUFTS UNIV, SCH MED, DEPT PATHOL, BOSTON, MA 02111 USA.
   TUFTS UNIV, SCH VET MED, DEPT PATHOL, BOSTON, MA 02111 USA.
   MASSACHUSETTS GEN HOSP E, MOLEC NEUROGENET UNIT, BOSTON, MA 02129 USA.
   MASSACHUSETTS GEN HOSP E, CTR CANC, BOSTON, MA 02129 USA.
   MIT, WHITEHEAD INST BIOMED RES, CAMBRIDGE, MA 02142 USA.
   MIT, DEPT BIOL, CAMBRIDGE, MA 02142 USA.
C3 Massachusetts Institute of Technology (MIT); Tufts University; Tufts
   University; Massachusetts Institute of Technology (MIT); Whitehead
   Institute; Massachusetts Institute of Technology (MIT)
RP JACKS, T (通讯作者)，MIT, CTR CANC RES, CAMBRIDGE, MA 02139 USA.
CR AINSWORTH PJ, 1993, HUM GENET, V91, P151
   ANDERSEN LB, 1993, NAT GENET, V3, P118, DOI 10.1038/ng0293-118
   [Anonymous], 1988, ARCH NEUROL-CHICAGO, V45, P575, DOI 10.1097/00005537-199606000-00007
   [Anonymous], 1982, NEURAL CREST
   Ausubel F.M, 2002, SHORT PROTOCOLS MOL, V5th
   BALLESTER R, 1990, CELL, V63, P851, DOI 10.1016/0092-8674(90)90151-4
   BASU TN, 1992, NATURE, V356, P713, DOI 10.1038/356713a0
   BERNARDS A, 1993, HUM MOL GENET, V2, P645, DOI 10.1093/hmg/2.6.645
   BOLANDE RP, 1974, HUM PATHOL, V5, P409, DOI 10.1016/S0046-8177(74)80021-3
   BRANNAN CI, 1994, GENE DEV, V8, P1019, DOI 10.1101/gad.8.9.1019
   Bronson R.T., 1990, GENETICS AGING, P280
   CAWTHON RM, 1990, CELL, V62, P193, DOI 10.1016/0092-8674(90)90253-B
   CHETTY R, 1993, AM J SURG PATHOL, V17, P837, DOI 10.1097/00000478-199308000-00009
   CLARKE AR, 1992, NATURE, V359, P328, DOI 10.1038/359328a0
   DASTON MM, 1992, NEURON, V8, P415, DOI 10.1016/0896-6273(92)90270-N
   DASTON MM, 1992, DEV DYNAM, V195, P216, DOI 10.1002/aja.1001950307
   DECLUE JE, 1992, CELL, V69, P265, DOI 10.1016/0092-8674(92)90407-4
   DECLUE JE, 1991, P NATL ACAD SCI USA, V88, P9914, DOI 10.1073/pnas.88.22.9914
   EASTON DF, 1993, AM J HUM GENET, V53, P305
   EPSTEIN DJ, 1991, CELL, V67, P767, DOI 10.1016/0092-8674(91)90071-6
   ESTIVILL X, 1991, HUM GENET, V88, P185
   FRANZ T, 1989, ANAT EMBRYOL, V180, P457, DOI 10.1007/BF00305120
   GOSSLER A, 1986, P NATL ACAD SCI USA, V83, P9065, DOI 10.1073/pnas.83.23.9065
   GREEN JE, 1992, AM J PATHOL, V140, P1401
   GUTMANN DH, 1993, HUM MOL GENET, V2, P989
   HINRICHS SH, 1987, SCIENCE, V237, P1340, DOI 10.1126/science.2888191
   HOOPER M, 1987, NATURE, V326, P292, DOI 10.1038/326292a0
   Hope D G, 1981, Adv Neurol, V29, P33
   HUYNH DP, 1994, DEV BIOL, V161, P538, DOI 10.1006/dbio.1994.1052
   JACKS T, 1992, NATURE, V359, P295, DOI 10.1038/359295a0
   JOHNSON MR, 1993, P NATL ACAD SCI USA, V90, P5539, DOI 10.1073/pnas.90.12.5539
   KIRBY ML, 1990, CIRCULATION, V82, P332, DOI 10.1161/01.CIR.82.2.332
   KIRBY ML, 1983, SCIENCE, V220, P1059, DOI 10.1126/science.6844926
   KNUDSON AG, 1971, P NATL ACAD SCI USA, V68, P820, DOI 10.1073/pnas.68.4.820
   KUEHN MR, 1987, NATURE, V326, P295, DOI 10.1038/326295a0
   LAIRD PW, 1991, NUCLEIC ACIDS RES, V19, P4293, DOI 10.1093/nar/19.15.4293
   LEE EYHP, 1992, NATURE, V359, P288, DOI 10.1038/359288a0
   LEGIUS E, 1993, NAT GENET, V3, P122, DOI 10.1038/ng0293-122
   LI Y, 1992, CELL, V69, P275, DOI 10.1016/0092-8674(92)90408-5
   MANSOUR SL, 1988, NATURE, V336, P348, DOI 10.1038/336348a0
   MARTIN GA, 1990, CELL, V63, P843, DOI 10.1016/0092-8674(90)90150-D
   RICCARDI VM, 1981, NEW ENGL J MED, V305, P1617, DOI 10.1056/NEJM198112313052704
   Riccardi VM, 1992, NEUROFIBROMATOSIS PH
   Robertson E, 1987, TERATOCARCINOMAS EMB
   SEIZINGER BR, 1993, NAT GENET, V3, P97, DOI 10.1038/ng0293-97
   SHACKLEFORD GM, 1987, CELL, V50, P89, DOI 10.1016/0092-8674(87)90665-9
   SHANNON KM, 1994, NEW ENGL J MED, V330, P597, DOI 10.1056/NEJM199403033300903
   TASSABEHJI M, 1993, AM J HUM GENET, V53, P90
   THE I, 1993, NAT GENET, V3, P62, DOI 10.1038/ng0193-62
   THOMAS KR, 1987, CELL, V51, P503, DOI 10.1016/0092-8674(87)90646-5
   VISKOCHIL D, 1990, CELL, V62, P187, DOI 10.1016/0092-8674(90)90252-A
   WALLACE MR, 1990, SCIENCE, V249, P181, DOI 10.1126/science.2134734
   XIE WQ, 1991, BIOTECHNIQUES, V11, P324
   XU GF, 1990, CELL, V63, P835, DOI 10.1016/0092-8674(90)90149-9
   XU GF, 1990, CELL, V62, P599, DOI 10.1016/0092-8674(90)90024-9
   XU W, 1992, GENE CHROMOSOME CANC, V4, P337, DOI 10.1002/gcc.2870040411
NR 56
TC 613
Z9 622
U1 1
U2 16
PU NATURE PUBLISHING GROUP
PI NEW YORK
PA 75 VARICK ST, 9TH FLR, NEW YORK, NY 10013-1917 USA
SN 1061-4036
EI 1546-1718
J9 NAT GENET
JI Nature Genet.
PD JUL
PY 1994
VL 7
IS 3
BP 353
EP 361
DI 10.1038/ng0794-353
PG 9
WC Genetics & Heredity
WE Science Citation Index Expanded (SCI-EXPANDED)
SC Genetics & Heredity
GA NU306
UT WOS:A1994NU30600008
PM 7920653
DA 2023-08-27
ER

PT J
AU MIYAGI, Y
   MORIOKA, T
   GRAFTON, ST
   OTSUKA, M
   FUKUI, M
AF MIYAGI, Y
   MORIOKA, T
   GRAFTON, ST
   OTSUKA, M
   FUKUI, M
TI STRIATAL GLUCOSE-METABOLISM AND [18F]FLUORODOPA UPTAKE IN A PATIENT WITH
   TUMOR-INDUCED HEMIPARKINSONISM
SO NEUROSURGERY
LA English
DT Article
DE DOPAMINE; GLUCOSE METABOLISM; MENINGIOMA; PARKINSONISM; POSITRON
   EMISSION TOMOGRAPHY; STRIATUM
ID F-18 DOPA UPTAKE; PARKINSONS-DISEASE; BRAIN-TUMOR; TOMOGRAPHY
AB WE STUDIED A patient with a falx meningioma in the right supplementary motor area and a left-sided hemiparkinsonism that resolved after the tumor was removed. Because there was no evidence of distortion of the basal ganglia and midbrain by the tumor on neuroradiological examination, the possible mechanism of parkinsonism is an impairment of the basal ganglia output to the supplementary motor area. Positron emission tomography scans with 2-[18F]fluoro-2-deoxy-D-glucose and 6-L-[18F]fluorodopa were performed to measure regional cerebral glucose metabolism and striatal dopamine metabolism, respectively. Regional cerebral glucose metabolism was decreased in the striatum of the side of the lesion, although dopamine metabolism was normal. These data suggest that the tumor may have impaired synaptic function of the striatum as a whole, giving rise to contralateral hemiparkinsonism without an impairment of the presynaptic dopaminergic nerve terminals in the striatum.
C1 KYUSHU UNIV 60,FAC MED,INST NEUROL,DEPT NEUROSURG,3-1-1 MAIDASHI,FUKUOKA 812,JAPAN.
   KYUSHU UNIV 60,FAC MED,DEPT RADIOL,FUKUOKA 812,JAPAN.
C3 Kyushu University; Kyushu University
OI Miyagi, Yasushi/0000-0002-1241-0838
CR Brazin M E, 1985, Pediatr Neurol, V1, P306, DOI 10.1016/0887-8994(85)90034-7
   BROOKS DJ, 1990, ANN NEUROL, V28, P547, DOI 10.1002/ana.410280412
   DEYEBENES JG, 1982, ANN NEUROL, V11, P313
   GARNETT ES, 1983, NATURE, V305, P137, DOI 10.1038/305137a0
   GHERARDI R, 1985, ACTA NEUROPATHOL, V65, P338, DOI 10.1007/BF00687019
   HUANG SC, 1991, J CEREBR BLOOD F MET, V11, P898, DOI 10.1038/jcbfm.1991.155
   JURGENS U, 1984, BRAIN RES, V300, P63, DOI 10.1016/0006-8993(84)91341-6
   LEENDERS KL, 1986, J NEUROL NEUROSUR PS, V49, P853, DOI 10.1136/jnnp.49.8.853
   LEENDERS KL, 1986, NEUROLOGY, V36, P1074, DOI 10.1212/WNL.36.8.1074
   MARTIN WRW, 1986, ADV NEUROL, V45, P95
   NAVARRO JA, 1986, J NEUROL NEUROSUR PS, V49, P1457, DOI 10.1136/jnnp.49.12.1457
   NICHOLSON AN, 1964, J NEUROSURG, V21, P104, DOI 10.3171/jns.1964.21.2.0104
   OLIVER L, 1959, LANCET, V2, P817
   OTSUKA M, 1991, J NEUROL NEUROSUR PS, V54, P898, DOI 10.1136/jnnp.54.10.898
   OTSUKA M, 1992, Journal of Nuclear Medicine, V33, P945
   Parker HL, 1923, J NERV MENT DIS, V58, P1, DOI 10.1097/00005053-192307000-00001
   PEARCE J, 1977, POSTGRAD MED J, V53, P726, DOI 10.1136/pgmj.53.626.726
   PLAYFORD ED, 1991, J CEREB BLOOD FLO S2, V11, pS1
   POLYZOIDIS KS, 1985, SURG NEUROL, V23, P59, DOI 10.1016/0090-3019(85)90161-2
   SANDYK R, 1983, J NEUROSURG, V58, P298, DOI 10.3171/jns.1983.58.2.0298
   SCHELL GR, 1984, J NEUROSCI, V4, P539
   SCIARRA D, 1953, AMA ARCH NEUROL PSY, V69, P450, DOI 10.1001/archneurpsyc.1953.02320280038004
   STRAUBE A, 1988, J NEUROL NEUROSUR PS, V51, P730, DOI 10.1136/jnnp.51.5.730
   WAGNER HN, 1983, SCIENCE, V221, P1264, DOI 10.1126/science.6604315
NR 24
TC 22
Z9 22
U1 1
U2 1
PU WILLIAMS & WILKINS
PI BALTIMORE
PA 351 WEST CAMDEN ST, BALTIMORE, MD 21201-2436
SN 0148-396X
J9 NEUROSURGERY
JI Neurosurgery
PD MAY
PY 1993
VL 32
IS 5
BP 838
EP 841
DI 10.1227/00006123-199305000-00019
PG 4
WC Clinical Neurology; Surgery
WE Science Citation Index Expanded (SCI-EXPANDED)
SC Neurosciences & Neurology; Surgery
GA LA619
UT WOS:A1993LA61900019
PM 8492861
DA 2023-08-27
ER

PT J
AU Sanchez-Pernaute, R
   Lee, H
   Patterson, M
   Reske-Nielsen, C
   Yoshizaki, T
   Sonntag, KC
   Studer, L
   Isacson, O
AF Sanchez-Pernaute, Rosario
   Lee, Hyojin
   Patterson, Michaela
   Reske-Nielsen, Casper
   Yoshizaki, Takahito
   Sonntag, Kai C.
   Studer, Lorenz
   Isacson, Ole
TI Parthenogenetic dopamine neurons from primate embryonic stem cells
   restore function in experimental Parkinsons disease
SO BRAIN
LA English
DT Article
DE stem cells; transplantation; midbrain; Parkinson's disease;
   parthenogenesis
ID SUBSTANTIA-NIGRA; NUCLEAR TRANSFER; RAT MODEL; DIRECTED DIFFERENTIATION;
   INDUCED DYSKINESIA; NEURAL PRECURSORS; ANIMAL-MODEL; ES CELLS;
   TRANSPLANTATION; MICE
AB The identity and functional potential of dopamine neurons derived in vitro from embryonic stem cells are critical for the development of a stem cell-based replacement therapy for Parkinsons disease. Using a parthenogenetic primate embryonic stem cell line, we have generated dopamine neurons that display persistent expression of midbrain regional and cell-specific transcription factors, which establish their proper identity and allow for their survival. We show here that transplantation of parthenogenetic dopamine neurons restores motor function in hemi-parkinsonian, 6-hydroxy-dopamine-lesioned rats. Exposure to Wnt5a and fibroblast growth factors (FGF) 20 and 2 at the final stage of in vitro differentiation enhanced the survival of dopamine neurons and, correspondingly, the extent of motor recovery of transplanted animals. Importantly for future development of clinical applications, dopamine neurons were post-mitotic at the time of transplantation and there was no tumour formation. These data provide proof for the concept that parthenogenetic stem cells are a suitable source of functional neurons for therapeutic applications.
C1 [Sanchez-Pernaute, Rosario; Yoshizaki, Takahito; Sonntag, Kai C.; Isacson, Ole] Harvard Univ, Udall Parkinsons Dis Res Ctr Excellence, McLean Hosp, Belmont, MA 02478 USA.
   [Sanchez-Pernaute, Rosario; Patterson, Michaela; Reske-Nielsen, Casper; Yoshizaki, Takahito; Sonntag, Kai C.; Isacson, Ole] Neuroregenerat Labs, McLean Hosp, Belmont, MA 02478 USA.
   [Lee, Hyojin; Studer, Lorenz] Mem Sloan Kettering Canc Ctr, Div Neurosurg & Dev Biol, Lab Stem Cell & Tumor Biol, New York, NY 10021 USA.
C3 Harvard University; McLean Hospital; National Institutes of Health (NIH)
   - USA; NIH National Library of Medicine (NLM); Harvard University;
   McLean Hospital; Memorial Sloan Kettering Cancer Center
RP Sanchez-Pernaute, R (通讯作者)，Fdn Inbiomed, Neuronal Stem Cells Lab, Paseo Mikeletegi 61, San Sebastian 20009, Spain.
EM
RI Sanchez-Pernaute, Rosario/ABD-8661-2021; Studer, Lorenz/ABH-1491-2021;
   Sanchez-Pernaute, Rosario/AAZ-9566-2021; Sonntag,
   Kai-Christian/AAU-6921-2021
OI Studer, Lorenz/0000-0003-0741-7987; Sanchez-Pernaute,
   Rosario/0000-0003-1144-9025; Isacson, Ole/0000-0001-8091-8302; Sonntag,
   Kai-Christian/0000-0001-7979-8171
FU NINDS NIH HHS [P50 NS039793, R01 NS052671, R01 NS-052671, P50 NS-39793]
   Funding Source: Medline
CR ABERCROMBIE M, 1946, ANAT REC, V94, P239, DOI 10.1002/ar.1090940210
   Alberi L, 2004, DEVELOPMENT, V131, P3229, DOI 10.1242/dev.01128
   Barberi T, 2003, NAT BIOTECHNOL, V21, P1200, DOI 10.1038/nbt870
   Ben-Hur T, 2004, STEM CELLS, V22, P1246, DOI 10.1634/stemcells.2004-0094
   Bjorklund LM, 2002, P NATL ACAD SCI USA, V99, P2344, DOI 10.1073/pnas.022438099
   Brederlau A, 2006, STEM CELLS, V24, P1433, DOI 10.1634/stemcells.2005-0393
   Castelo-Branco GA, 2006, MOL CELL NEUROSCI, V31, P251, DOI 10.1016/j.mcn.2005.09.014
   Castelo-Branco GA, 2003, P NATL ACAD SCI USA, V100, P12747, DOI 10.1073/pnas.1534900100
   Christophersen NS, 2007, NAT MED, V13, P118, DOI 10.1038/nm0207-118a
   Cibelli JB, 2002, SCIENCE, V295, P819, DOI 10.1126/science.1065637
   Correia AS, 2007, FRONT NEUROANAT, V1, DOI [10.3389/neuro.05/004.2007, 10.3389/neuro.05.004.2007]
   Dean W, 1998, DEVELOPMENT, V125, P2273
   DUNNETT SB, 1988, EXP BRAIN RES, V71, P411
   Elkabetz Y, 2008, GENE DEV, V22, P152, DOI 10.1101/gad.1616208
   Ferrari D, 2006, EUR J NEUROSCI, V24, P1885, DOI 10.1111/j.1460-9568.2006.05093.x
   Hedlund E, 2008, STEM CELLS, V26, P1526, DOI 10.1634/stemcells.2007-0996
   Hikichi T, 2007, STEM CELLS, V25, P46, DOI 10.1634/stemcells.2006-0439
   Holm PC, 2006, J NEUROCHEM, V99, P343, DOI 10.1111/j.1471-4159.2006.04039.x
   Humpherys D, 2001, SCIENCE, V293, P95, DOI 10.1126/science.1061402
   Kim JH, 2002, NATURE, V418, P50, DOI 10.1038/nature00900
   Kim K, 2007, CELL STEM CELL, V1, P346, DOI 10.1016/j.stem.2007.07.001
   Kim K, 2007, SCIENCE, V315, P482, DOI 10.1126/science.1133542
   Kittappa R, 2007, PLOS BIOL, V5, P2875, DOI 10.1371/journal.pbio.0050325
   Ko JY, 2007, J NEUROCHEM, V103, P1417, DOI 10.1111/j.1471-4159.2007.04898.x
   Kono T, 2004, NATURE, V428, P860, DOI 10.1038/nature02402
   Kuwagata M, 2007, NEUROTOXICOLOGY, V28, P780, DOI 10.1016/j.neuro.2006.07.017
   Lee G, 2007, NAT BIOTECHNOL, V25, P1468, DOI 10.1038/nbt1365
   LEIGHTON PA, 1995, NATURE, V375, P34, DOI 10.1038/375034a0
   LEVITT P, 1982, DEV BRAIN RES, V4, P35, DOI 10.1016/0165-3806(82)90095-5
   Livak KJ, 2001, METHODS, V25, P402, DOI 10.1006/meth.2001.1262
   Lucifero D, 2002, GENOMICS, V79, P530, DOI 10.1006/geno.2002.6732
   MARSHALL JF, 1977, SCIENCE, V198, P62, DOI 10.1126/science.897683
   McKay R, 2000, NATURE, V406, P361, DOI 10.1038/35019186
   Mela F, 2007, J NEUROCHEM, V101, P483, DOI 10.1111/j.1471-4159.2007.04456.x
   Mendez I, 2005, BRAIN, V128, P1498, DOI 10.1093/brain/awh510
   Mitalipov S, 2007, STEM CELLS, V25, P581, DOI 10.1634/stemcells.2006-0120
   Murase S, 2006, J NEUROSCI, V26, P9750, DOI 10.1523/JNEUROSCI.2745-06.2006
   Nash JE, 2005, FASEB J, V19, P583, DOI 10.1096/fj.04-1854fje
   NIKKHAH G, 1993, NEUROSCIENCE, V56, P33, DOI 10.1016/0306-4522(93)90559-X
   Ohmachi S, 2000, BIOCHEM BIOPH RES CO, V277, P355, DOI 10.1006/bbrc.2000.3675
   Parish CL, 2008, J CLIN INVEST, V118, P149, DOI 10.1172/JCI32273
   Perrier AL, 2004, P NATL ACAD SCI USA, V101, P12543, DOI 10.1073/pnas.0404700101
   Revazova ES, 2007, CLONING STEM CELLS, V9, P432, DOI 10.1089/clo.2007.0033
   Rodriguez-Gomez JA, 2007, STEM CELLS, V25, P918, DOI 10.1634/stemcells.2006-0386
   Roy NS, 2006, NAT MED, V12, P1259, DOI 10.1038/nm1495
   Sanchez-Pernaute R, 2005, STEM CELLS, V23, P914, DOI 10.1634/stemcells.2004-0172
   Sanchez-Pernaute R, 2001, J NEUROSCI RES, V65, P284, DOI 10.1002/jnr.1152
   Sanchez-Pernaute R, 2004, J NEUROINFLAMM, V1, DOI 10.1186/1742-2094-1-6
   Schwarz SC, 2006, PARKINSONISM RELAT D, V12, P302, DOI 10.1016/j.parkreldis.2006.01.003
   Sgado P, 2006, P NATL ACAD SCI USA, V103, P15242, DOI 10.1073/pnas.0602116103
   Skottman H, 2006, FEBS LETT, V580, P2875, DOI 10.1016/j.febslet.2006.03.083
   Sonntag KC, 2007, STEM CELLS, V25, P411, DOI 10.1634/stemcells.2006-0380
   Sonntag KC, 2005, MOL CELL NEUROSCI, V28, P417, DOI 10.1016/j.mcn.2004.06.003
   Tabar V, 2005, NAT BIOTECHNOL, V23, P601, DOI 10.1038/nbt1088
   Takagi Y, 2005, J CLIN INVEST, V115, P102, DOI 10.1172/JCI200521137
   Timmer M, 2007, J NEUROSCI, V27, P459, DOI 10.1523/JNEUROSCI.4493-06.2007
   Wilkinson LS, 2007, NAT REV NEUROSCI, V8, P832, DOI 10.1038/nrn2235
   Wouterlood FG, 2003, J NEUROSCI METH, V128, P129, DOI 10.1016/S0165-0270(03)00171-7
   Wu H, 2007, P NATL ACAD SCI USA, V104, P13821, DOI 10.1073/pnas.0706199104
   Yan YP, 2005, STEM CELLS, V23, P781, DOI 10.1634/stemcells.2004-0365
   Yao WD, 2004, NEURON, V41, P625, DOI 10.1016/S0896-6273(04)00048-0
NR 61
TC 66
Z9 72
U1 1
U2 25
PU OXFORD UNIV PRESS
PI OXFORD
PA GREAT CLARENDON ST, OXFORD OX2 6DP, ENGLAND
SN 0006-8950
EI 1460-2156
J9 BRAIN
JI Brain
PD AUG
PY 2008
VL 131
BP 2127
EP 2139
DI 10.1093/brain/awn144
PN 8
PG 13
WC Clinical Neurology; Neurosciences
WE Science Citation Index Expanded (SCI-EXPANDED)
SC Neurosciences & Neurology
GA 335XW
UT WOS:000258329900019
PM 18669499
OA Bronze, Green Published
DA 2023-08-27
ER

PT J
AU Wolff, JEA
   Kramm, C
   Kortmann, RD
   Pietsch, T
   Rutkowski, S
   Jorch, N
   Gnekow, A
   Driever, PH
AF Wolff, Johannes E. A.
   Kramm, Christof
   Kortmann, Rolf-Dieter
   Pietsch, Torsten
   Rutkowski, Stefan
   Jorch, Norbert
   Gnekow, Astrid
   Driever, Pablo Hernaiz
TI Valproic acid was well tolerated in heavily pretreated pediatric
   patients with high-grade glioma
SO JOURNAL OF NEURO-ONCOLOGY
LA English
DT Article
DE Valproic acid; Glioblastoma; Anaplastic astrocytomas; Pontine glioma;
   Child; Adolescent
ID HISTONE DEACETYLASE INHIBITOR; SUBEROYLANILIDE HYDROXAMIC ACID; ACTIVITY
   IN-VITRO; PHASE-I; MALIGNANT GLIOMA; HIT-GBM; SIMULTANEOUS
   RADIOCHEMOTHERAPY; GLIOBLASTOMA-MULTIFORME; FETAL-HEMOGLOBIN;
   TOPOISOMERASE-II
AB Valproic acid (VPA) inhibits histone deacetylase and has been reported to induce apoptosis in glioma. We report 44 heavily pretreated pediatric patients with high-grade glioma or diffuse intrinsic pontine glioma who received VPA as oral continues maintenance treatment with individual dose adaptation. The tumor status when starting the drug was: no measurable disease in 12, measurable but stable disease in 12, and measurable progressive disease in 22 patients. Average trough blood levels of VPA were 99 mg/l. The most frequent complaint was somnolence (three patients), but no severe toxicity was reported. One relapse patient responded, early progression of disease was observed in three frontline patients and in six relapsed patients. Median overall survival duration for all patients was 1.33 years, with large differences between first-line (5-year overall survival, 44%) and relapse therapy (5-year overall survival, 14%). This shows that valproate is safe in this patient population. The moderate tumor efficacy encourages studying the drug further as an element of multi-agent protocols.
C1 [Wolff, Johannes E. A.] Univ Texas MD Anderson Canc Ctr, Childrens Canc Hosp, Dept Pediat, Unit 87, Houston, TX 77030 USA.
   [Wolff, Johannes E. A.] Univ Texas MD Anderson Canc Ctr, Dept Biostat, Houston, TX 77030 USA.
   [Wolff, Johannes E. A.] Univ Regensburg, Regensburg, Germany.
   [Kramm, Christof] Univ Halle Wittenberg, Dept Pediat Oncol, Halle, Germany.
   [Kortmann, Rolf-Dieter] Univ Leipzig, Dept Radiat Oncol, Leipzig, Germany.
   [Pietsch, Torsten] Univ Bonn, Dept Neuropathol, D-5300 Bonn, Germany.
   [Rutkowski, Stefan] Univ Wurzburg, Childrens Univ Hosp, Wurzburg, Germany.
   [Jorch, Norbert] Krankenanstalten Gilead, Dept Pediat, Bielefeld, Germany.
   [Gnekow, Astrid] Klin Kinder & Jugendl, Dept Pediat, Augsburg, Germany.
   [Driever, Pablo Hernaiz] Charite Univ Med Berlin, Dept Pediat Oncol & Hematol, D-13353 Berlin, Germany.
C3 University of Texas System; UTMD Anderson Cancer Center; University of
   Texas System; UTMD Anderson Cancer Center; University of Regensburg;
   Martin Luther University Halle Wittenberg; Leipzig University;
   University of Bonn; University of Hamburg; University Medical Center
   Hamburg-Eppendorf; University of Wurzburg; Free University of Berlin;
   Humboldt University of Berlin; Charite Universitatsmedizin Berlin
RP Wolff, JEA (通讯作者)，Univ Texas MD Anderson Canc Ctr, Childrens Canc Hosp, Dept Pediat, Unit 87, 1515 Holcombe Blvd, Houston, TX 77030 USA.
EM
OI Wolff, Johannes/0000-0003-0278-1309; Hernaiz Driever,
   Pablo/0000-0003-3135-3872
FU Deutsche Kinderkrebsstiftung; Germany and Core [CA 16672]
FX Supported by Deutsche Kinderkrebsstiftung, Germany and Core Grant CA
   16672. The original HIT- GBM (R) Database has been used before as a
   source for analyses with different focuses and different subsets of data
   [ 39, 42 - 44, 53, 54]. This analysis only uses the data generated
   prospectively for including patients formally enrolled on the treatment
   study and the registry or at least the registry until 2003 with follow
   up updated in 2006. The other HIT- GBM publications have been based on
   overlapping but not identical patient populations.
CR Anthony M, 1977, Clin Exp Neurol, V14, P208
   Atmaca A, 2007, BRIT J CANCER, V97, P177, DOI 10.1038/sj.bjc.6603851
   Benesch M, 2005, J NEURO-ONCOL, V72, P179, DOI 10.1007/s11060-004-3546-5
   Blaheta RA, 2005, MED RES REV, V25, P383, DOI 10.1002/med.20027
   Blumenschein GR, 2008, INVEST NEW DRUG, V26, P81, DOI 10.1007/s10637-007-9075-2
   Bohan TP, 2001, NEUROLOGY, V56, P1405, DOI 10.1212/WNL.56.10.1405
   Bouffet E, 1997, CANCER CHEMOTH PHARM, V39, P376, DOI 10.1007/s002800050586
   Broniscer A, 2004, ONCOLOGIST, V9, P197, DOI 10.1634/theoncologist.9-2-197
   BROWN NA, 1980, LANCET, V1, P660
   Camphausen K, 2005, INT J CANCER, V114, P380, DOI 10.1002/ijc.20774
   Carlisi D, 2008, INT J ONCOL, V32, P177
   Das CM, 2007, J NEURO-ONCOL, V85, P159, DOI 10.1007/s11060-007-9402-7
   DOMINGODOMENECH J, 2008, BREAST CANC IN PRESS
   Donaldson SS, 2006, J CLIN ONCOL, V24, P1266, DOI 10.1200/JCO.2005.04.6599
   Dong G, 2008, J PHARMACOL EXP THER, V325, P978, DOI 10.1124/jpet.108.137398
   Driever PH, 2004, PEDIATR HEMAT ONCOL, V21, P743, DOI 10.1080/08880010490514985
   Driever PH, 1999, KLIN PADIATR, V211, P323, DOI 10.1055/s-2008-1043809
   Estlin EJ, 1998, BRIT J CANCER, V78, P652, DOI 10.1038/bjc.1998.555
   Fournel M, 2008, MOL CANCER THER, V7, P759, DOI 10.1158/1535-7163.MCT-07-2026
   Hassig CA, 2008, MOL CANCER THER, V7, P1054, DOI 10.1158/1535-7163.MCT-07-2347
   Huang LL, 2000, ONCOGENE, V19, P5712, DOI 10.1038/sj.onc.1203963
   ISOJARVI JIT, 1993, NEW ENGL J MED, V329, P1383, DOI 10.1056/NEJM199311043291904
   Johnson J, 2008, TRANSPL P, V40, P459, DOI 10.1016/j.transproceed.2008.01.039
   Kieslich M, 2003, PEDIATR HEMAT ONCOL, V20, P15, DOI 10.1080/08880010390158496
   Knupfer MM, 1998, ANTICANCER RES, V18, P3585
   Koenig SA, 2006, EPILEPSIA, V47, P2027, DOI 10.1111/j.1528-1167.2006.00846.x
   Kramm CM, 2006, ANTICANCER RES, V26, P3773
   LANCE J W, 1975, Proceedings of the Australian Association of Neurologists, V12, P55
   Limdi NA, 2007, EPILEPSIA, V48, P478, DOI 10.1111/j.1528-1167.2007.00989.x
   Liu L, 2001, MED PEDIATR ONCOL, V37, P390, DOI 10.1002/mpo.1215
   Marchion DC, 2005, MOL CANCER THER, V4, P1993, DOI 10.1158/1535-7163.MCT-05-0194
   MASOUDI A, 2008, ANTICANCER IN PRESS
   Petrella A, 2008, EUR J CANCER, V44, P740, DOI 10.1016/j.ejca.2008.01.023
   Phiel CJ, 2001, J BIOL CHEM, V276, P36734, DOI 10.1074/jbc.M101287200
   POST RM, 1983, EPILEPSIA, V24, pS97, DOI 10.1111/j.1528-1157.1983.tb04652.x
   REGAN CM, 1985, BRAIN RES, V347, P394, DOI 10.1016/0006-8993(85)90207-0
   Selby R, 1997, BLOOD, V90, P891, DOI 10.1182/blood.V90.2.891.891_891_892
   Spiller HA, 2000, J TOXICOL-CLIN TOXIC, V38, P755, DOI 10.1081/CLT-100102388
   Spiller SE, 2008, J NEURO-ONCOL, V87, P133, DOI 10.1007/s11060-007-9505-1
   SPOSTO R, 1989, J NEURO-ONCOL, V7, P165, DOI 10.1007/BF00165101
   Stupp R, 2005, NEW ENGL J MED, V352, P987, DOI 10.1056/NEJMoa043330
   Wagner S, 2005, ANTICANCER RES, V25, P2583
   Wagner S, 2006, APMIS, V114, P731, DOI 10.1111/j.1600-0463.2006.apm_516.x
   Wagner S, 2006, J NEURO-ONCOL, V79, P281, DOI 10.1007/s11060-006-9133-1
   Witt O, 2004, PEDIATR BLOOD CANCER, V43, P181, DOI 10.1002/pbc.20083
   Wolff JEA, 2002, ANTICANCER RES, V22, P3569
   Wolff JEA, 2002, BRIT J CANCER, V87, P945, DOI 10.1038/sj.bjc.6600552
   Wolff JEA, 1996, KLIN PADIATR, V208, P193, DOI 10.1055/s-2008-1046473
   Wolff JEA, 1996, KLIN PADIATR, V208, P190, DOI 10.1055/s-2008-1046472
   Wolff JEA, 2000, CANCER, V89, P2131, DOI 10.1002/1097-0142(20001115)89:10<2131::AID-CNCR14>3.0.CO;2-J
   Wolff JEA, 2008, J NEURO-ONCOL, V87, P155, DOI 10.1007/s11060-007-9495-z
   Wolff JEA, 2006, J NEURO-ONCOL, V79, P315, DOI 10.1007/s11060-006-9147-8
   Yi X, 2008, BIOCHEM PHARMACOL, V75, P1697, DOI 10.1016/j.bcp.2007.10.035
   Zaskodova D, 2006, ANN NY ACAD SCI, V1091, P385, DOI 10.1196/annals.1378.082
NR 54
TC 43
Z9 44
U1 0
U2 5
PU SPRINGER
PI NEW YORK
PA 233 SPRING ST, NEW YORK, NY 10013 USA
SN 0167-594X
J9 J NEURO-ONCOL
JI J. Neuro-Oncol.
PD DEC
PY 2008
VL 90
IS 3
BP 309
EP 314
DI 10.1007/s11060-008-9662-x
PG 6
WC Oncology; Clinical Neurology
WE Science Citation Index Expanded (SCI-EXPANDED)
SC Oncology; Neurosciences & Neurology
GA 370NG
UT WOS:000260768300010
PM 18679579
DA 2023-08-27
ER

PT J
AU Link, MJ
   Cohen, PL
   Breneman, JC
   Tew, JM
AF Link, MJ
   Cohen, PL
   Breneman, JC
   Tew, JM
TI Malignant squamous degeneration of a cerebellopontine angle epidermoid
   tumor - Case report
SO JOURNAL OF NEUROSURGERY
LA English
DT Article
DE cerebellopontine angle; epidermoid cyst; squamous cell carcinoma;
   trigeminal neuralgia
ID CELL CARCINOMA; STEREOTAXIC RADIOSURGERY; POSTERIOR-FOSSA; BRAIN-STEM;
   CYST; MR; METASTASES; THERAPY; CT
AB The authors present the case of a woman with a cerebellopontine angle (CPA) epidermoid cyst that degenerated into a squamous cell carcinoma. Malignant degeneration of an epidermoid cyst is an extremely rare occurrence. Malignant transformation must be considered in the differential diagnosis when new contrast enhancement on imaging studies and progressive neurological deficit are seen in a patient harboring an epidermoid cyst.
   The patient initially presented with a 10-year history of left trigeminal neuralgia, subacute left-sided hearing loss, and with facial weakness of 3 weeks' duration. Initial magnetic resonance (MR) imaging revealed a left CPA mass, consistent with an epidermoid. There was faint contrast enhancement where the tumor was in contact with the lateral brainstem.
   A subtotal resection was performed. Histopathological findings were consistent with an epidermoid tumor. One year after initial presentation, the patient's neurological deficit had increased, and follow-up MR imaging demonstrated a large contrast-enhancing tumor filling the left CPA and compressing the brainstem. At repeated surgery a squamous cell carcinoma arising from the previous epidermoid was found. The patient was subsequently treated with external-beam radiotherapy and stereotactic radiosurgery. Her tumor stabilized. Three years and 8 months after the patient's initial presentation, a new area of tumor developed at the torcular Herophili. The patient died shortly thereafter.
   Malignant squamous degeneration is a rare cause of enhancement on MR images, as is progressive neurological deficit in a patient with an epidermoid. The combination of subtotal resection, external-beam radiotherapy, and stereotactic radiosurgery may be useful for local tumor control but the long-term prognosis is guarded.
C1 Mayo Clin & Mayo Fdn, Dept Neurol Surg, Rochester, MN 55905 USA.
   Univ Cincinnati, Dept Neurosurg, Cincinnati, OH USA.
   Univ Cincinnati, Inst Neurosci, Cincinnati, OH USA.
   Mayfield Clin, Cincinnati, OH USA.
C3 Mayo Clinic; University System of Ohio; University of Cincinnati;
   University System of Ohio; University of Cincinnati
RP Link, MJ (通讯作者)，Mayo Clin & Mayo Fdn, Dept Neurol Surg, 200 1st St SW, Rochester, MN 55905 USA.
CR ABRAMSON RC, 1989, NEUROSURGERY, V24, P574, DOI 10.1227/00006123-198904000-00014
   ACCIARRI N, 1993, BRIT J NEUROSURG, V7, P565, DOI 10.3109/02688699308995081
   ALVORD EC, 1977, ANN NEUROL, V2, P367, DOI 10.1002/ana.410020504
   Barasch A, 1998, MT SINAI J MED, V65, P370
   BAUMANN CHH, 1956, J NEUROSURG, V13, P455, DOI 10.3171/jns.1956.13.5.0455
   BERGER MS, 1985, J NEUROSURG, V62, P214, DOI 10.3171/jns.1985.62.2.0214
   BONDESON L, 1984, ACTA CYTOL, V28, P487
   Bourhis Jean, 1997, Current Opinion in Oncology, V9, P262, DOI 10.1097/00001622-199709030-00008
   Breneman JC, 1997, CANCER, V79, P551, DOI 10.1002/(SICI)1097-0142(19970201)79:3<551::AID-CNCR18>3.0.CO;2-2
   BUATTI JM, 1995, INT J RADIAT ONCOL, V32, P1161, DOI 10.1016/0360-3016(94)00461-S
   CHANDLER WF, 1975, J NEUROSURG, V43, P614, DOI 10.3171/jns.1975.43.5.0614
   DAVIDSON SI, 1960, J NEUROL NEUROSUR PS, V23, P176, DOI 10.1136/jnnp.23.2.176
   DAVIS KR, 1976, RADIOLOGY, V119, P347, DOI 10.1148/119.2.347
   DUNN RC, 1981, J NEUROSURG, V55, P654, DOI 10.3171/jns.1981.55.4.0654
   EEKHOF JLA, 1985, SURG NEUROL, V23, P189, DOI 10.1016/0090-3019(85)90344-1
   ELIASH A, 1983, CHILD BRAIN, V10, P414
   Ernst P., 1912, VERH DTSCH PATHOL GE, V15, P226
   FIUME D, 1988, SURG NEUROL, V29, P178, DOI 10.1016/0090-3019(88)90002-X
   FOX H, 1965, J NEUROL NEUROSUR PS, V28, P276, DOI 10.1136/jnnp.28.3.276
   GARCIA CA, 1981, J NEUROSURG, V54, P824, DOI 10.3171/jns.1981.54.6.0824
   Giannotta S L, 1976, Surg Neurol, V5, P164
   GOLDMAN SA, 1987, J NEUROSURG, V66, P618, DOI 10.3171/jns.1987.66.4.0618
   GUIDETTI B, 1977, J NEUROSURG, V47, P12, DOI 10.3171/jns.1977.47.1.0012
   GUTIN PH, 1980, J NEUROSURG, V52, P574, DOI 10.3171/jns.1980.52.4.0574
   KEVILLE FJ, 1959, J NEUROSURG, V16, P564, DOI 10.3171/jns.1959.16.5.0564
   Kubokura T, 1986, Neurol Med Chir (Tokyo), V26, P706, DOI 10.2176/nmc.26.706
   LANDERS JW, 1960, ARCH PATHOL, V70, P419
   LEAL O, 1978, J NEUROSURG, V48, P811, DOI 10.3171/jns.1978.48.5.0811
   LEWIS AJ, 1983, J NEUROSURG, V59, P538, DOI 10.3171/jns.1983.59.3.0538
   MACCARTY CS, 1959, SURG GYNECOL OBSTET, V108, P191
   MATSUNO A, 1987, Neurological Surgery, V15, P851
   Mendenhall WM, 1998, CANCER INVEST, V16, P594, DOI 10.3109/07357909809032890
   MORAN CC, 1995, J COMPUT ASSIST TOMO, V19, P628, DOI 10.1097/00004728-199507000-00024
   MORI Y, 1995, NEUROL SURG TOKYO, V23, P905
   NISHIO S, 1995, NEUROSURGERY, V37, P329, DOI 10.1227/00006123-199508000-00021
   Nishiura I, 1989, Neurol Med Chir (Tokyo), V29, P600, DOI 10.2176/nmc.29.600
   NOSAKA Y, 1979, J NEUROSURG, V50, P830, DOI 10.3171/jns.1979.50.6.0830
   Ogata N, 1996, SURG NEUROL, V46, P477, DOI 10.1016/S0090-3019(96)00222-4
   PARIKH S, 1995, J NEURO-ONCOL, V24, P293, DOI 10.1007/BF01052846
   SCHWARTZ JF, 1978, NEUROLOGY, V28, P124, DOI 10.1212/WNL.28.2.124
   STEFFEY DJ, 1988, J COMPUT ASSIST TOMO, V12, P438, DOI 10.1097/00004728-198805010-00016
   Takado M, 1982, Rinsho Shinkeigaku, V22, P579
   TAMPIERI D, 1989, AM J NEURORADIOL, V10, P351
   Tew JM, 1994, ATLAS OPERATIVE MICR, V1
   TOGLIA JU, 1965, J NEUROSURG, V23, P384, DOI 10.3171/jns.1965.23.4.0384
   TOGNETTI F, 1991, British Journal of Neurosurgery, V5, P303, DOI 10.3109/02688699109005191
   ULRICH J, 1964, J NEUROSURG, V21, P1051, DOI 10.3171/jns.1964.21.12.1051
   WONG SW, 1976, CANCER, V37, P1525, DOI 10.1002/1097-0142(197603)37:3<1525::AID-CNCR2820370341>3.0.CO;2-W
   YANAI Y, 1985, NEUROSURGERY, V16, P252, DOI 10.1227/00006123-198502000-00026
   YUH WTC, 1988, AM J NEURORADIOL, V9, P794
   ZAPOL WM, 1977, NEW ENGL J MED, V296, P1279
   Zhao HY, 1998, STEREOT FUNCT NEUROS, V70, P161, DOI 10.1159/000056418
NR 52
TC 60
Z9 63
U1 1
U2 2
PU AMER ASSOC NEUROLOGICAL SURGEONS
PI CHARLOTTESVILLE
PA UNIV VIRGINIA, 1224 WEST MAIN ST, STE 450, CHARLOTTESVILLE, VA 22903 USA
SN 0022-3085
J9 J NEUROSURG
JI J. Neurosurg.
PD NOV
PY 2002
VL 97
IS 5
BP 1237
EP 1243
DI 10.3171/jns.2002.97.5.1237
PG 7
WC Clinical Neurology; Surgery
WE Science Citation Index Expanded (SCI-EXPANDED)
SC Neurosciences & Neurology; Surgery
GA 613BC
UT WOS:000179109800040
PM 12450053
DA 2023-08-27
ER

PT J
AU Ohira, S
   Ueda, Y
   Isono, M
   Masaoka, A
   Hashimoto, M
   Miyazaki, M
   Takashina, M
   Koizumi, M
   Teshima, T
AF Ohira, Shingo
   Ueda, Yoshihiro
   Isono, Masaru
   Masaoka, Akira
   Hashimoto, Misaki
   Miyazaki, Masayoshi
   Takashina, Masaaki
   Koizumi, Masahiko
   Teshima, Teruki
TI Can clinically relevant dose errors in patient anatomy be detected by
   gamma passing rate or modulation complexity score in
   volumetric-modulated arc therapy for intracranial tumors?
SO JOURNAL OF RADIATION RESEARCH
LA English
DT Article
DE gamma passing rate; modulation complexity score; VMAT; QA; intracranial
   tumors
ID PROSTATE-CANCER; IMRT QA; QUALITY; RADIOTHERAPY; SYSTEM; BEAMS; HEAD
AB We investigated whether methods conventionally used to evaluate patient-specific QA in volumetric-modulated arc therapy (VMAT) for intracranial tumors detect clinically relevant dosimetric errors. VMAT plans with coplanar arcs were designed for 37 intracranial tumors. Dosimetric accuracy was validated by using a 3D array detector. Dose deviations between the measured and planned doses were evaluated by gamma analysis. In addition, modulation complexity score for VMAT (MCSv) for each plan was calculated. Three-dimensional dose distributions in patient anatomy were reconstructed using 3DVH software, and clinical deviations in dosimetric parameters between the 3DVH doses and planned doses were calculated. The gamma passing rate (GPR)/MCSv and the clinical dose deviation were evaluated using Pearson's correlation coefficient. Significant correlation (P < 0.05) between the clinical dose deviation and GPR was observed with both the 3%/3 mm and 2%/2 mm criteria in clinical target volume (D-99), brain (D-2), brainstem (D-2) and chiasm (D-2), albeit that the correlations were not 'strong' (0.38 < vertical bar r vertical bar < 0.54). The maximum dose deviations of brainstem were up to 4.9 Gy and 2.9 Gy for D-max and D-%, respectively in the case of high GPR (98.2(%) with 3%/3 mm criteria). Regarding MCSv, none of the evaluated organs showed a significant correlation with clinical dose deviation, and correlations were 'weak' or absent (0.01 < vertical bar r vertical bar < 0.21). The use of high GPR and MCSv values does not always detect dosimetric errors in a patient. Therefore, in-depth analysis with the DVH for patient-specific QA is considered to be preferable for guaranteeing safe dose delivery.
C1 [Ohira, Shingo; Ueda, Yoshihiro; Isono, Masaru; Masaoka, Akira; Miyazaki, Masayoshi; Teshima, Teruki] Osaka Med Ctr Canc & Cardiovasc Dis, Dept Radiat Oncol, Higashinari Ku, Nakamichi 1-3-3, Osaka 5378511, Japan.
   [Ohira, Shingo; Takashina, Masaaki; Koizumi, Masahiko] Osaka Univ, Dept Med Phys & Engn, Grad Sch Med, Suita, Osaka, Japan.
   [Ueda, Yoshihiro] Osaka Univ, Dept Radiat Oncol, Grad Sch Med, Suita, Osaka, Japan.
   [Hashimoto, Misaki] Yao Municipal Hosp, Dept Radiat Oncol, Yao, Japan.
C3 Osaka Medical Center for Cancer & Cardiovascular Diseases; Osaka
   University; Osaka University
RP Teshima, T (通讯作者)，Osaka Med Ctr Canc & Cardiovasc Dis, Dept Radiat Oncol, Higashinari Ku, Nakamichi 1-3-3, Osaka 5378511, Japan.
EM
OI Ohira, Shingo/0000-0002-6170-1471
FU Japan Society for the Promotion of Science (JSPS) [84409, 23003];
   National Cancer Center Research and Development Fund [84409, 26-A-28];
   JSPS KAKENHI [84409, 15H04913]; Health and Labour Sciences Research
   [84409]; Osaka Cancer Society [84409];  [H26-Cancer Policy-General-014]
FX This work was supported by the Japan Society for the Promotion of
   Science (JSPS; No. 84409) core-to-core program (No. 23003), the National
   Cancer Center Research and Development Fund (No. 84409) (26-A-28), JSPS
   KAKENHI (No. 84409) Grant No. 15H04913, Health and Labour Sciences
   Research (No. 84409) Grants (H26-Cancer Policy-General-014) and the
   Osaka Cancer Society (No. 84409).
CR [Anonymous], 2007, J APPL CLIN MED PHYS
   [Anonymous], 2010, PHYS RAD THERAPY
   Basran PS, 2008, MED PHYS, V35, P2300, DOI 10.1118/1.2919075
   Beriwal S, 2007, INT J RADIAT ONCOL, V68, P166, DOI 10.1016/j.ijrobp.2006.12.023
   Clemente-Gutierrez F, 2015, J APPL CLIN MED PHYS, V16, P198, DOI 10.1120/jacmp.v16i2.5190
   Ghandour S, 2015, J APPL CLIN MED PHYS, V16, P258, DOI 10.1120/jacmp.v16i3.5410
   Hoogeman MS, 2008, INT J RADIAT ONCOL, V70, P609, DOI 10.1016/j.ijrobp.2007.08.066
   Kathirvel M, 2013, RADIAT ONCOL, V8, DOI 10.1186/1748-717X-8-140
   Korevaar EW, 2011, RADIOTHER ONCOL, V100, P446, DOI 10.1016/j.radonc.2011.09.007
   Li GJ, 2013, PHYS MEDICA, V29, P295, DOI 10.1016/j.ejmp.2012.04.005
   Liang Y, 2013, J APPL CLIN MED PHYS, V14, P15, DOI 10.1120/jacmp.v14i2.3852
   Macgarry CK, 2016, BRIT J RADIOL, V89
   Masi L, 2013, MED PHYS, V40, DOI 10.1118/1.4810969
   Matuszak MM, 2010, INT J RADIAT ONCOL, V77, P608, DOI 10.1016/j.ijrobp.2009.08.032
   McNiven AL, 2010, MED PHYS, V37, P505, DOI 10.1118/1.3276775
   Nelms BE, 2013, MED PHYS, V40, DOI 10.1118/1.4826166
   Nelms BE, 2012, MED PHYS, V39, P4228, DOI 10.1118/1.4729709
   Nelms BE, 2011, MED PHYS, V38, P1037, DOI 10.1118/1.3544657
   Ning ZH, 2015, STRAHLENTHER ONKOL, V191, P34, DOI 10.1007/s00066-014-0760-8
   Olch AJ, 2012, MED PHYS, V39, P81, DOI 10.1118/1.3666771
   Palma D, 2008, INT J RADIAT ONCOL, V72, P996, DOI 10.1016/j.ijrobp.2008.02.047
   Quan EZM, 2012, INT J RADIAT ONCOL, V83, P1169, DOI 10.1016/j.ijrobp.2011.09.015
   Rustgi SN, 1995, MED PHYS, V22, P2117, DOI 10.1118/1.597655
   Song JH, 2015, J APPL CLIN MED PHYS, V16, P263, DOI 10.1120/jacmp.v16i6.5696
   SUN NUCLEAR corporation, 2012, 1220011 SUN NUCLEAR
   Tyler MK, 2016, J APPL CLIN MED PHYS, V17, P5781
   Verbakel WFAR, 2009, INT J RADIAT ONCOL, V74, P252, DOI 10.1016/j.ijrobp.2008.12.033
   Westermark M, 2000, PHYS MED BIOL, V45, P685, DOI 10.1088/0031-9155/45/3/308
   Wu C, 2012, MED PHYS, V39, P3051, DOI 10.1118/1.4711755
NR 29
TC 6
Z9 8
U1 0
U2 4
PU OXFORD UNIV PRESS
PI OXFORD
PA GREAT CLARENDON ST, OXFORD OX2 6DP, ENGLAND
SN 0449-3060
EI 1349-9157
J9 J RADIAT RES
JI J. Radiat. Res.
PD SEP
PY 2017
VL 58
IS 5
BP 685
EP 692
DI 10.1093/jrr/rrx006
PG 8
WC Biology; Oncology; Radiology, Nuclear Medicine & Medical Imaging
WE Science Citation Index Expanded (SCI-EXPANDED)
SC Life Sciences & Biomedicine - Other Topics; Oncology; Radiology, Nuclear
   Medicine & Medical Imaging
GA FK4CW
UT WOS:000413437700013
PM 28339918
OA Green Submitted, Green Published, gold
DA 2023-08-27
ER

PT J
AU LAMBERT, PR
   RUTH, RA
   THOMAS, JF
AF LAMBERT, PR
   RUTH, RA
   THOMAS, JF
TI PROMONTORY ELECTRICAL-STIMULATION IN POSTOPERATIVE ACOUSTIC TUMOR
   PATIENTS
SO LARYNGOSCOPE
LA English
DT Article
ID COCHLEAR IMPLANTATION; HEARING PRESERVATION; AUDITORY POTENTIALS;
   MANAGEMENT; SURGERY; LABYRINTHECTOMY; NEUROMA; EAR
AB Complete deafness can follow acoustic tumor surgery and results from labyrinthine injury, auditory nerve trauma, and/or vascular compromise. A central auditory prosthesis is one potential rehabilitative strategy in such patients. Anatomical studies suggest that some spiral ganglion cells may survive after vascular occlusion, and we have demonstrated responses to electrical stimulation in patients after labyrinthectomy. It was thus hypothesized that patients deafened after a hearing conservation attempt, but maintaining an intact auditory nerve, could utilize an intracochlear implant. To investigate this possibility, promontory electrical stimulation was performed on three patients who had tumors less than 2 cm and who had serviceable preoperative hearing, but no responses postoperatively. Behavioral responses and electrically evoked auditory brainstem and middle latency responses were obtained from two patients, one of whom was 6 years postsurgery. These data indicate that a cochlear implant may be possible after acoustic tumor surgery.
RP LAMBERT, PR (通讯作者)，UNIV VIRGINIA,HLTH SCI CTR,DEPT OTOLARYNGOL HEAD & NECK SURG,BOX 430,CHARLOTTESVILLE,VA 22908, USA.
CR BELAL A, 1983, J LARYNGOL OTOL, V97, P1
   BERNSTEIN JM, 1966, ARCHIV OTOLARYNGOL, V83, P422
   CHEN DA, 1988, LARYNGOSCOPE, V98, P1170
   EDGERTON BJ, 1982, ANN OTO RHINOL LARYN, V91, P117
   GAME CJA, 1987, ANN OTO RHINOL LARYN, V96, P94, DOI 10.1177/00034894870960S150
   GARDNER G, 1988, ANN OTO RHINOL LARYN, V97, P55, DOI 10.1177/000348948809700110
   GERKEN GM, 1984, HEARING RES, V13, P249, DOI 10.1016/0378-5955(84)90078-9
   GLASSCOCK ME, 1986, LARYNGOSCOPE, V96, P1088
   GRANTZ BJ, 1986, ANN OTO RHINOL LARYN, V95, P454
   HUGHES GB, 1982, LARYNGOSCOPE, V92, P1351
   KILENY PR, 1987, ARCH OTOLARYNGOL, V113, P1072
   KONISHI T, 1961, J ACOUST SOC AM, V33, P349, DOI 10.1121/1.1908659
   KVETON JF, 1989, LARYNGOSCOPE, V99, P610
   LAMBERT PR, 1990, ARCH OTOLARYNGOL, V116, P197
   MAZZONI A, 1970, ARCHIV OTOLARYNGOL, V91, P128
   MIYAMOTO RT, 1991, ANN OTO RHINOL LARYN, V100, P38, DOI 10.1177/000348949110000107
   NEDZELSKI JM, 1984, J OTOLARYNGOL, V13, P355
   OTTO SR, 1990, ANN OTO RHINOL LARYN, V99, P789, DOI 10.1177/000348949009901006
   PERLMAN H B, 1959, Laryngoscope, V69, P591
   PERLMAN H B, 1957, Ann Otol Rhinol Laryngol, V66, P537
   SEBIYA T, 1985, J NEUROSURG, V63, P598
   SHELTON C, 1989, LARYNGOSCOPE, V99, P405, DOI 10.1288/00005537-198904000-00009
   SHORE SE, 1990, ANN OTO RHINOL LARYN, V99, P571, DOI 10.1177/000348949009900715
   SMITH L, 1983, ANN OTO RHINOL LARYN, V92, P19, DOI 10.1177/000348948309200105
NR 24
TC 15
Z9 16
U1 0
U2 0
PU LIPPINCOTT-RAVEN PUBL
PI PHILADELPHIA
PA 227 EAST WASHINGTON SQ, PHILADELPHIA, PA 19106
SN 0023-852X
J9 LARYNGOSCOPE
JI Laryngoscope
PD JUL
PY 1992
VL 102
IS 7
BP 814
EP 819
DI 10.1288/00005537-199207000-00011
PG 6
WC Medicine, Research & Experimental; Otorhinolaryngology
WE Science Citation Index Expanded (SCI-EXPANDED)
SC Research & Experimental Medicine; Otorhinolaryngology
GA JK415
UT WOS:A1992JK41500011
PM 1614252
DA 2023-08-27
ER

PT J
AU De Jonghe, S
   Johnson, MD
   Mamidi, RNVS
   Vinken, P
   Feyen, B
   Lammens, G
   Proctor, J
AF De Jonghe, Sandra
   Johnson, Mark D.
   Mamidi, Rao N. V. S.
   Vinken, Petra
   Feyen, Bianca
   Lammens, Godelieve
   Proctor, Jim
TI Renal tubular and adrenal medullary tumors in the 2-year rat study with
   canagliflozin confirmed to be secondary to carbohydrate (glucose)
   malabsorption in the 15-month mechanistic rat study
SO CHEMICO-BIOLOGICAL INTERACTIONS
LA English
DT Article
DE Canagliflozin; SGLT2 inhibitor; SGLT1 inhibitor; Kidney tumors;
   Pheochromocytomas; Adrenal medullary hyperplasia; Carbohydrate
   malabsorption
ID RISK-ASSESSMENT; RELEVANCE
AB During preclinical development of canagliflozin, an SGLT2 inhibitor, treatment-related pheochromocytomas, renal tubular tumors (RTT), and testicular Leydig cell tumors were reported in the 2-year rat toxicology study.
   In a previous 6-month rat mechanistic study, feeding a glucose free diet prevented canagliflozin effects on carbohydrate malabsorption as well as the increase in cell proliferation in adrenal medulla and kidneys, implicating carbohydrate malabsorption as the mechanism for tumor formation.
   In this chronic study male Sprague-Dawley rats were dosed orally with canagliflozin at high dose-levels (65 or 100 mg/kg/day) for 15 months and received either a standard diet or a glucose-free diet. Canagliflozin-dosed rats on standard diet showed presence of basophilic renal tubular tumors (6/90) and an increased incidence of adrenal medullary hyperplasia (35/90), which was fully prevented by feeding a glucose-free diet (no RTT's; adrenal medullary hyperplasia in <= 5/90).
   These data further confirm that kidney and adrenal medullary tumors in the 2-year rat study were secondary to carbohydrate (glucose) malabsorption and were not due to a direct effect of canagliflozin on these target tissues. (C) 2017 Elsevier B.V. All rights reserved.
C1 [De Jonghe, Sandra; Vinken, Petra; Feyen, Bianca; Lammens, Godelieve] Janssen Res & Dev, Turnhoutseweg 30, B-2340 Beerse, Belgium.
   [Johnson, Mark D.; Mamidi, Rao N. V. S.; Proctor, Jim] Janssen Res & Dev LLC, 1000 Route 202 South, Raritan, NJ 08869 USA.
C3 Johnson & Johnson; Janssen Pharmaceuticals; Johnson & Johnson; Johnson &
   Johnson USA; Janssen Biotech Inc
RP De Jonghe, S (通讯作者)，Janssen Res & Dev, Turnhoutseweg 30, B-2340 Beerse, Belgium.
EM
FU Janssen Research and Development, LLC; Janssen Research & Development, a
   Division of Janssen Pharmaceutica NV
FX This work was supported by Janssen Research and Development, LLC and
   Janssen Research & Development, a Division of Janssen Pharmaceutica NV.
   The authors are all employees and, as such, designed the studies,
   participated in data collection and analysis, made the decision to
   publish, and prepared the manuscript.
CR [Anonymous], 2012, PREC PACK INS
   [Anonymous], 1987, MULTIPLE COMP PROCED
   Barone S, 2009, J BIOL CHEM, V284, P5056, DOI 10.1074/jbc.M808128200
   De Jonghe S, 2014, CHEM-BIOL INTERACT, V224, P1, DOI 10.1016/j.cbi.2014.09.018
   DUNN OJ, 1964, TECHNOMETRICS, V6, P241, DOI 10.2307/1266041
   Fisher RA, 1935, J R STAT SOC, V98, P39, DOI 10.2307/2342435
   Gorboulev V, 2012, DIABETES, V61, P187, DOI 10.2337/db11-1029
   Greim H, 2009, CRIT REV TOXICOL, V39, P695, DOI 10.1080/10408440903190861
   Nakayama T, 2010, AM J PHYSIOL-RENAL, V298, pF712, DOI 10.1152/ajprenal.00433.2009
   Rao M.V.S., 2014, CHEM-BIOL INTERACT, V224, P109
   Siegel S., 1988, NONPARAMETRIC STAT B
   Ways K, 2015, TOXICOL PATHOL, V43, P48, DOI 10.1177/0192623314557179
   Wood CE, 2015, TOXICOL PATHOL, V43, P760, DOI 10.1177/0192623315576005
NR 13
TC 3
Z9 3
U1 0
U2 1
PU ELSEVIER IRELAND LTD
PI CLARE
PA ELSEVIER HOUSE, BROOKVALE PLAZA, EAST PARK SHANNON, CO, CLARE, 00000,
   IRELAND
SN 0009-2797
EI 1872-7786
J9 CHEM-BIOL INTERACT
JI Chem.-Biol. Interact.
PD NOV 1
PY 2017
VL 277
BP 85
EP 90
DI 10.1016/j.cbi.2017.09.008
PG 6
WC Biochemistry & Molecular Biology; Pharmacology & Pharmacy; Toxicology
WE Science Citation Index Expanded (SCI-EXPANDED)
SC Biochemistry & Molecular Biology; Pharmacology & Pharmacy; Toxicology
GA FN7RM
UT WOS:000416216100010
PM 28916336
DA 2023-08-27
ER

PT J
AU Guillamo, JS
   Doz, F
   Delattre, JY
AF Guillamo, JS
   Doz, F
   Delattre, JY
TI Brain stem gliomas
SO CURRENT OPINION IN NEUROLOGY
LA English
DT Article
ID CHILDRENS-CANCER-GROUP; PEDIATRIC-ONCOLOGY-GROUP; HYPERFRACTIONATED
   RADIOTHERAPY; RADIATION-THERAPY; TECTAL TUMORS; NEUROFIBROMATOSIS
   TYPE-1; PROGNOSTIC FACTORS; PONTINE GLIOMAS; NATURAL-HISTORY; PHASE-I/II
AB Brainstem gliomas are now regarded as a heterogeneous group of tumors that can be distinguished by age of onset, clinical and radiological presentation and biological behavior. This paper will focus on each subtype of tumor, in children and in adults, and on recent advances in diagnostic criteria and therapeutic options. Curr Opin Neurol 14:711-715. (C) 2001 Lippincott Williams Wilkins.
C1 Hop La Pitie Salpetriere, Serv Neurol Mazann, Mazann Div Neurol, F-75651 Paris 13, France.
   Inst Curie, Dept Pediat Oncol, Paris, France.
   Hop Henri Mondor, Dept Neurol, F-94010 Creteil, France.
C3 Assistance Publique Hopitaux Paris (APHP); Hopital Universitaire
   Charles-Foix - APHP; Hopital Universitaire Pitie-Salpetriere - APHP;
   UDICE-French Research Universities; Sorbonne Universite; Hopital
   Universitaire Armand-Trousseau - APHP; UDICE-French Research
   Universities; Universite PSL; UNICANCER; Institut Curie; Assistance
   Publique Hopitaux Paris (APHP); Universite
   Paris-Est-Creteil-Val-de-Marne (UPEC); Hopital Universitaire
   Henri-Mondor - APHP
RP Delattre, JY (通讯作者)，Hop La Pitie Salpetriere, Serv Neurol Mazann, Mazann Div Neurol, F-75651 Paris 13, France.
EM
RI Guillamo, Jean-Sebastien/AAC-9525-2021; Doz, Francois P/K-8616-2012
OI Guillamo, Jean-Sebastien/0000-0003-0165-8333; Doz,
   Francois/0000-0001-9286-4831
CR ALBRIGHT AL, 1986, J NEUROSURG, V65, P751, DOI 10.3171/jns.1986.65.6.0751
   ALBRIGHT AL, 1993, NEUROSURGERY, V33, P1026
   Allen J, 1999, CANCER, V86, P1064, DOI 10.1002/(SICI)1097-0142(19990915)86:6<1064::AID-CNCR24>3.0.CO;2-1
   BARKOVICH AJ, 1991, PEDIATR NEUROSURG, V16, P73, DOI DOI 10.1159/000120511]
   Bouffet E, 2000, CANCER, V88, P685, DOI 10.1002/(SICI)1097-0142(20000201)88:3<685::AID-CNCR27>3.0.CO;2-K
   Bowers DC, 2000, PEDIATR NEUROSURG, V32, P24, DOI 10.1159/000028893
   Bricolo A, 1991, Acta Neurochir Suppl (Wien), V53, P148
   Broniscer A, 2000, J CLIN ONCOL, V18, P1246, DOI 10.1200/JCO.2000.18.6.1246
   Constantini S, 1996, J NEURO-ONCOL, V28, P193
   Del Brutto OH, 1999, NEUROLOGY, V52, P210, DOI 10.1212/WNL.52.1.210
   Donahue B, 1998, INT J RADIAT ONCOL, V40, P677, DOI 10.1016/S0360-3016(97)00846-8
   EPSTEIN FJ, 1993, J NEUROSURG, V78, P408, DOI 10.3171/jns.1993.78.3.0408
   Fischbein NJ, 1996, PEDIATR NEUROSURG, V24, P9, DOI 10.1159/000121010
   Fisher PG, 2000, CANCER, V89, P1569, DOI 10.1002/1097-0142(20001001)89:7<1569::AID-CNCR22>3.0.CO;2-0
   FREEMAN CR, 1986, INT J RADIAT ONCOL, V12, P1823, DOI 10.1016/0360-3016(86)90325-1
   Freeman CR, 1999, CHILD NERV SYST, V15, P545, DOI 10.1007/s003810050542
   Freeman CR, 2000, INT J RADIAT ONCOL, V47, P561, DOI 10.1016/S0360-3016(00)00471-5
   FREEMAN CR, 1993, INT J RADIAT ONCOL, V27, P197, DOI 10.1016/0360-3016(93)90228-N
   Freeman CR, 1998, INT J RADIAT ONCOL, V40, P265, DOI 10.1016/S0360-3016(97)00572-5
   GUILLAMO JS, 2001, IN PRESS BRAIN
   HOFFMAN HJ, 1980, NEUROSURGERY, V7, P243, DOI 10.1227/00006123-198009000-00007
   Kaplan AM, 1996, PEDIATR NEUROSURG, V24, P185, DOI 10.1159/000121036
   Knauer-Fischer S, 1999, PEDIATR NEUROL, V20, P57, DOI 10.1016/S0887-8994(98)00094-0
   Landolfi JC, 1998, NEUROLOGY, V51, P1136, DOI 10.1212/WNL.51.4.1136
   LINSTADT DE, 1991, INT J RADIAT ONCOL, V20, P757, DOI 10.1016/0360-3016(91)90019-Z
   Mandell LR, 1999, INT J RADIAT ONCOL, V43, P959, DOI 10.1016/S0360-3016(98)00501-X
   MANTRAVADI RVP, 1982, CANCER, V49, P1294, DOI 10.1002/1097-0142(19820315)49:6<1294::AID-CNCR2820490636>3.0.CO;2-V
   MOLLOY PT, 1995, NEUROLOGY, V45, P1897, DOI 10.1212/WNL.45.10.1897
   NELSON MD, 1994, RADIOLOGY, V191, P279, DOI 10.1148/radiology.191.1.8134588
   Oka K, 1999, J NEUROSURG, V91, P964, DOI 10.3171/jns.1999.91.6.0964
   PACKER RJ, 1985, NEUROLOGY, V35, P397, DOI 10.1212/WNL.35.3.397
   PACKER RJ, 1983, ANN NEUROL, V14, P177, DOI 10.1002/ana.410140204
   PACKER RJ, 1994, CANCER-AM CANCER SOC, V74, P1827, DOI 10.1002/1097-0142(19940915)74:6<1827::AID-CNCR2820740628>3.0.CO;2-Q
   POLLACK IF, 1994, J NEUROSURG, V80, P681, DOI 10.3171/jns.1994.80.4.0681
   POLLACK IF, 1993, J NEUROSURG, V78, P859, DOI 10.3171/jns.1993.78.6.0859
   Pollack IF, 1996, NEUROLOGY, V46, P1652, DOI 10.1212/WNL.46.6.1652
   Schulz-Ertner D, 2000, RADIOTHER ONCOL, V57, P215, DOI 10.1016/S0167-8140(00)00230-9
   Selvapandian S, 1999, ACTA NEUROCHIR, V141, P721, DOI 10.1007/s007010050367
   SEVICK RJ, 1992, AM J ROENTGENOL, V159, P171, DOI 10.2214/ajr.159.1.1609692
   SHRIEVE DC, 1992, INT J RADIAT ONCOL, V24, P599, DOI 10.1016/0360-3016(92)90704-L
   SQUIRES LA, 1994, NEUROLOGY, V44, P953, DOI 10.1212/WNL.44.5.953
   Walker DA, 1999, ARCH DIS CHILD, V80, P558, DOI 10.1136/adc.80.6.558
   Weaver KD, 1999, J NEUROSURG, V91, P1037, DOI 10.3171/jns.1999.91.6.1037
   Zagzag D, 2000, PEDIATRICS, V106, P1045, DOI 10.1542/peds.106.5.1045
NR 44
TC 68
Z9 76
U1 0
U2 2
PU LIPPINCOTT WILLIAMS & WILKINS
PI PHILADELPHIA
PA TWO COMMERCE SQ, 2001 MARKET ST, PHILADELPHIA, PA 19103 USA
SN 1350-7540
EI 1473-6551
J9 CURR OPIN NEUROL
JI Curr. Opin. Neurol.
PD DEC
PY 2001
VL 14
IS 6
BP 711
EP 715
DI 10.1097/00019052-200112000-00006
PG 5
WC Clinical Neurology; Neurosciences
WE Science Citation Index Expanded (SCI-EXPANDED)
SC Neurosciences & Neurology
GA 510NG
UT WOS:000173213700006
PM 11723378
DA 2023-08-27
ER

PT J
AU KANE, AG
   ROBLES, HA
   SMIRNIOTOPOULOS, JG
   HEIRONIMUS, JD
   FISH, MH
AF KANE, AG
   ROBLES, HA
   SMIRNIOTOPOULOS, JG
   HEIRONIMUS, JD
   FISH, MH
TI RADIOLOGIC-PATHOLOGICAL CORRELATION - DIFFUSE PONTINE ASTROCYTOMA
SO AMERICAN JOURNAL OF NEURORADIOLOGY
LA English
DT Article
DE BRAIN STEM, NEOPLASMS; RADIOLOGIC-PATHOLOGICAL CORRELATIONS
ID BRAIN-STEM GLIOMAS; TUMORS; DIAGNOSIS; CHILDREN; CHILDHOOD; LESIONS;
   BIOPSY
C1 BROOKE ARMY MED CTR,DEPT NUCL MED,FT SAM HOUSTON,TX 78234.
   BROOKE ARMY MED CTR,DEPT PATHOL,FT SAM HOUSTON,TX 78234.
   WALTER REED ARMY MED CTR,DEPT RADIOL,WASHINGTON,DC 20307.
   ARMED FORCES INST PATHOL,DEPT RADIOL PATHOL,WASHINGTON,DC 20306.
C3 San Antonio Military Medical Center; United States Department of
   Defense; United States Army; San Antonio Military Medical Center; United
   States Department of Defense; United States Army; United States
   Department of Defense; United States Army; Walter Reed National Military
   Medical Center; United States Department of Defense
RP KANE, AG (通讯作者)，BROOKE ARMY MED CTR,DEPT RADIOL,FT SAM HOUSTON,TX 78234, USA.
RI Smirniotopoulos, James/D-3726-2011
CR ABERNATHEY CD, 1989, J NEUROSURG, V70, P195, DOI 10.3171/jns.1989.70.2.0195
   ALBRIGHT AL, 1986, J NEUROSURG, V65, P751, DOI 10.3171/jns.1986.65.6.0751
   BALL WS, 1992, AM J NEURORADIOL, V13, P761
   BARHOVICH AJ, 1990, PEDIATRIC NEUROIMAGI, P164
   Bradley WG, 1992, MAGNETIC RESONANCE I, P721
   Buckley RC, 1930, ARCH PATHOL, V9, P779
   BYRNE JV, 1989, NEURORADIOLOGY, V31, P129, DOI 10.1007/BF00698839
   FARWELL JR, 1977, CANCER, V40, P3123, DOI 10.1002/1097-0142(197712)40:6<3123::AID-CNCR2820400656>3.0.CO;2-6
   FLORES LE, 1986, AM J DIS CHILD, V140, P684, DOI 10.1001/archpedi.1986.02140210082031
   FULTON DS, 1981, J NEUROSURG, V54, P721, DOI 10.3171/jns.1981.54.6.0721
   HUEFTLE MG, 1985, J COMPUT ASSIST TOMO, V9, P263, DOI 10.1097/00004728-198503000-00006
   KAPLAN WD, 1987, J NUCL MED, V28, P47
   LASSITER KR, 1971, J NEUROSURG, V34, P719, DOI 10.3171/jns.1971.34.6.0719
   LASSMAN LP, 1967, LANCET, V1, P913
   LEE BCP, 1985, AM J NEURORADIOL, V6, P159
   MARTIN RA, 1991, NEUROLOGY, V41, P1675, DOI 10.1212/WNL.41.10.1675
   MOUNTZ JM, 1988, J NEUROSURG, V68, P705, DOI 10.3171/jns.1988.68.5.0705
   NAIDICH TP, 1984, SEMIN ROENTGENOL, V19, P100, DOI 10.1016/0037-198X(84)90030-0
   PANITCH HS, 1970, AM J DIS CHILD, V119, P465, DOI 10.1001/archpedi.1970.02100050467001
   PILCHER C, 1934, ARCH NEUROL PSYCHIAT, P1210
   REIGEL DH, 1979, CHILD BRAIN, V5, P329
   SEGALL HD, 1982, RADIOL CLIN N AM, V20, P237
   STROINK AR, 1987, NEUROSURGERY, V20, P439, DOI 10.1227/00006123-198703000-00014
   STROINK AR, 1986, J NEUROSURG, V65, P745, DOI 10.3171/jns.1986.65.6.0745
NR 24
TC 5
Z9 5
U1 0
U2 0
PU AMER SOC NEURORADIOLOGY
PI OAK BROOK
PA 2210 MIDWEST RD, OAK BROOK, IL 60521
SN 0195-6108
J9 AM J NEURORADIOL
JI Am. J. Neuroradiol.
PD JUL-AUG
PY 1993
VL 14
IS 4
BP 941
EP 945
PG 5
WC Clinical Neurology; Neuroimaging; Radiology, Nuclear Medicine & Medical
   Imaging
WE Science Citation Index Expanded (SCI-EXPANDED)
SC Neurosciences & Neurology; Radiology, Nuclear Medicine & Medical Imaging
GA LM070
UT WOS:A1993LM07000026
PM 8352167
DA 2023-08-27
ER

PT J
AU LEONETTI, JP
   REICHMAN, OH
   ALMEFTY, O
   LI, J
   SMITH, PG
AF LEONETTI, JP
   REICHMAN, OH
   ALMEFTY, O
   LI, J
   SMITH, PG
TI NEUROTOLOGIC CONSIDERATIONS IN THE TREATMENT OF ADVANCED CLIVAL TUMORS
SO OTOLARYNGOLOGY-HEAD AND NECK SURGERY
LA English
DT Article; Proceedings Paper
CT 1991 ANNUAL MEETING OF THE AMERICAN ACADEMY OF OTOLARYNGOLOGY - HEAD AND
   NECK SURGERY
CY SEP 22-26, 1991
CL KANSAS CITY, MO
SP AMER ACAD OTOLARYNGOL, HEAD & NECK SURG
ID SKULL BASE; MENINGIOMAS; NERVE; BONE
AB Head and neck manifestations of advanced clival tumors result from subtle, yet progressive mass effect or direct involvement of multiple cranial nerves. Misinterpretation of clinical or radiographic findings in these patients may result in inappropriate treatment planning, increased patient morbidity, and probable tumor recurrence. Our combined experience in managing 21 patients with advanced clival tumors has shown that preoperative loss of vision, diplopia, or facial hypesthesia suggests superior parasellar disease. Facial twitching or neurotologic symptoms result from posterior central tumor growth, while inferior extension of disease leads to basal cranial nerve deficits with associated speech, voice, and swallowing dysfunctions. The purpose of this article is to correlate the complex anatomy of the clivus, brainstem, and cranial base and the various neurotologic findings associated with neoplasms in this region. Lateral skull base surgical procedures, based on preoperative clinicoradiographic assessment, will be detailed with particular emphasis on preservation of critical neurovascular structures.
C1 ST JOHNS MERCY MED CTR,MIDWEST OTOL GRP,ST LOUIS,MO 63141.
C3 Saint Johns Mercy Medical Center
RP LEONETTI, JP (通讯作者)，LOYOLA UNIV,MED CTR,DEPT OTOLARYNGOL HEAD & NECK SURG,2160 S 1ST AVE,MAYWOOD,IL 60153, USA.
CR ALMEFTY O, 1988, NEUROSURGERY, V22, P510, DOI 10.1227/00006123-198803000-00010
   ALMEFTY O, 1987, NEUROSURGERY, V21, P474, DOI 10.1227/00006123-198710000-00006
   CASTELLANO F, 1953, Acta Radiol Suppl, V104, P1
   DICHIRO GD, 1964, J NEUROSURG, V21, P447, DOI 10.3171/jns.1964.21.6.0447
   FISCH U, 1982, ANN OTO RHINOL LARYN, V91, P474, DOI 10.1177/000348948209100502
   GREITZ T, 1963, ACTA RADIOL DIAGN, V1, P284, DOI 10.1177/028418516300100222
   HOLLINSHEAD WH, 1974, TXB ANATOMY, P794
   HOUSE WF, 1976, ARCH OTOLARYNGOL, V102, P334
   HUANG VP, 1968, AM J ROENTGENOL, V104, P35
   LEONETTI JP, 1989, OTOLARYNG HEAD NECK, V101, P74, DOI 10.1177/019459988910100112
   LEONETTI JP, 1990, OTOLARYNG HEAD NECK, V103, P46, DOI 10.1177/019459989010300107
   MAYBERG MR, 1986, J NEUROSURG, V65, P160, DOI 10.3171/jns.1986.65.2.0160
   MOLLER AR, 1983, J NEUROSURG, V59, P493, DOI 10.3171/jns.1983.59.3.0493
   OOT RF, 1988, AM J ROENTGENOL, V151, P567, DOI 10.2214/ajr.151.3.567
   PARKINSON D, 1965, J NEUROSURG, V23, P474, DOI 10.3171/jns.1965.23.5.0474
   POTTER GD, 1974, RADIOL CLIN N AM, V12, P483
   RHOTON AL, 1976, J NEUROSURG, V24, P54
   SALAMON GM, 1971, J NEUROSURG, V35, P734
   SAVARA BS, 1979, ANGLE ORTHOD, V49, P141
   SEKHAR LN, 1990, NEUROSURGERY, V27, P764, DOI 10.1227/00006123-199011000-00015
   Yasargil MG, 1980, ADV TECH STAND NEURO, V7, P3, DOI [10.1007/978-3-7091-7051-9_1, DOI 10.1007/978-3-7091-7051-9_1]
NR 21
TC 2
Z9 2
U1 0
U2 0
PU MOSBY-YEAR BOOK INC
PI ST LOUIS
PA 11830 WESTLINE INDUSTRIAL DR, ST LOUIS, MO 63146-3318
SN 0194-5998
J9 OTOLARYNG HEAD NECK
JI Otolaryngol. Head Neck Surg.
PD JUL
PY 1992
VL 107
IS 1
BP 49
EP 56
DI 10.1177/019459989210700108
PG 8
WC Otorhinolaryngology; Surgery
WE Conference Proceedings Citation Index - Science (CPCI-S); Science Citation Index Expanded (SCI-EXPANDED)
SC Otorhinolaryngology; Surgery
GA JD621
UT WOS:A1992JD62100008
PM 1528602
DA 2023-08-27
ER

PT J
AU Kaga, K
   Iwasaki, S
   Tamura, A
   Suzuki, J
   Haebara, H
AF Kaga, K
   Iwasaki, S
   Tamura, A
   Suzuki, J
   Haebara, H
TI Temporal bone pathology of acoustic neuroma correlating with presence of
   electrocochleography and absence of auditory brainstem response
SO JOURNAL OF LARYNGOLOGY AND OTOLOGY
LA English
DT Article
DE schwannoma, vestibular; audiometry, evoked response; temporal bone,
   pathology
ID AUDIOMETRY
AB The temporal bone pathology of a 74-year-old female affected by vestibular schwannoma was compared with findings of auditory brainstem response and electrocochleography. At age 71, she complained of hearing loss in the left ear in which pure tone audiometry revealed threshold elevation in the middle-and high-frequency range. Temporal bone CT scanning revealed a medium-sized cerebellopontine angle tumour in the left ear. ABR showed no response in the left ear, but the electrocochleography showed clear compound action potentials. Three years later, at age 74, she died of metastatic lung cancer and sepsis. The left temporal bone pathology consisted primarily of a large vestibular schwannoma occupying the internal auditory meatus. The organ of Corti was well preserved in each turn. In the modiolus, the numbers of spiral ganglion cells and cochlear nerve fibres in each turn were decreased. These histological findings suggest that clear compound action potentials were recorded from the distal portion of the cochlear nerve in spite of the presence of the vestibular schwannoma, but ABR could not be detected because of the blockade of the proximal portion of the cochlear nerve by the vestibular schwannoma.
C1 TEIKYO UNIV,SCH MED,DEPT PATHOL 2,TOKYO 173,JAPAN.
C3 Teikyo University
RP Kaga, K (通讯作者)，UNIV TOKYO,FAC MED,DEPT OTOLARYNGOL,BUNKYO KU,HONGO 7-3-1,TOKYO 113,JAPAN.
CR Antoni NRE, 1920, RUCKENMARKSTUMOREN N, P413
   BENITEZ JT, 1967, ARCHIV OTOLARYNGOL, V86, P25
   CHANDRASEKHAR SS, 1995, AM J OTOL, V16, P63
   DAUMAN R, 1988, CLIN OTOLARYNGOL, V13, P107, DOI 10.1111/j.1365-2273.1988.tb00750.x
   DEMOURA LFP, 1967, ARCHIV OTOLARYNGOL, V85, P125
   EGGERMONT JJ, 1980, ANN OTO RHINOL LARYN, V89, P1
   HALL JW, 1992, HDB AUDITORY EVOKED, P385
   IGARASHI M, 1984, ARCH OTOLARYNGOL, V99, P379
   PORTMANN M, 1989, ARCH OTO-RHINO-LARYN, V246, P286, DOI 10.1007/BF00463577
   SCHUKNECHT HF, 1966, PATHOLOGY EAR, V80, P1
   SELTERS WA, 1977, ARCH OTOLARYNGOL, V103, P181
   SHANON E, 1981, LARYNGOSCOPE, V91, P254
   SILVERSTEIN H, 1966, ARCHIV OTOLARYNGOL, V84, P395
   SUGA F, 1976, ANN OTO RHINOL LARYN, V85, P343, DOI 10.1177/000348947608500305
   YLIKOSKI J, 1978, ARCH OTOLARYNGOL, V104, P679
NR 15
TC 15
Z9 15
U1 0
U2 1
PU HEADLEY BROTHERS LTD
PI ASHFORD
PA INVICTA PRESS, ASHFORD, KENT, ENGLAND TN24 8HH
SN 0022-2151
J9 J LARYNGOL OTOL
JI J. Laryngol. Otol.
PD OCT
PY 1997
VL 111
IS 10
BP 967
EP 972
DI 10.1017/S002221510013909X
PG 6
WC Otorhinolaryngology
WE Science Citation Index Expanded (SCI-EXPANDED)
SC Otorhinolaryngology
GA YE386
UT WOS:A1997YE38600016
PM 9425489
DA 2023-08-27
ER

PT J
AU Giussani, C
   Guida, L
   Biassoni, V
   Schiavello, E
   Carrabba, G
   Trezza, A
   Sganzerla, E
   Massimino, M
AF Giussani, Carlo
   Guida, Lelio
   Biassoni, Veronica
   Schiavello, Elisabetta
   Carrabba, Giorgio
   Trezza, Andrea
   Sganzerla, Erik
   Massimino, Maura
TI Retrospective analysis of the clinical and radiological features of 94
   consecutive DIPGs patients to investigate the factors determining the
   development of hydrocephalus and its impact on clinical status and
   survival
SO CHILDS NERVOUS SYSTEM
LA English
DT Article
DE Diffuse intrinsic pontine glioma; Hydrocephalus; Endoscopic third
   ventriculostomy; Ventriculo peritoneal shunt
ID INTRINSIC PONTINE GLIOMA; BRAIN-STEM GLIOMA; DIFFUSE; REGISTRY; CHILDREN
AB Purpose There is no consensus in the literature about the impact of hydrocephalus on clinical course and overall survival of diffuse intrinsic pontine gliomas (DIPG) patients as well as about its specific treatment. Authors reviewed a series of DIPG patients to investigate factors related to the onset of hydrocephalus, its treatment, and its impact on clinical course and prognosis. Methods A retrospective observational study was performed enrolling pediatric patients affected by DIPG from 2008 to 2018. Clinical and radiological charts were reviewed to find patients' demographic, pathologic and radiologic features in hydrocephalic and non-hydrocephalic patients. In the hydrocephalus cohort, treatment strategy and its effectiveness and complications were analyzed. Results Ninety-four pediatric patients were enrolled in the study. Patients who developed hydrocephalus showed significantly lesser maximum axial tumor areas than patients without hydrocephalus (respectively 6.5 cm(2) vs 16.45 cm(2), p < 0.005). Hydrocephalus developed in 33 patients (35%) with an onset interval of 5.24 +/- 1.21 months (range 3.2-7.3). The majority of hydrocephalic patients (28 cases, 90%) were treated by ventriculoperitoneal shunt, the remaining 3 patients being treated by endoscopic third ventriculostomy. Mean overall survival was 16.6 months +/- 20 months without significative difference between the groups. Conclusion The onset of hydrocephalus occurs in the first moths of the disease story and found a negative correlation with tumor maximal axial diameter. Early treatment of hydrocephalus presents a very low complications rate with satisfying clinical outcome, as it allows the patients to continue the neurooncological therapies being a part of the treatment armamentarium instead of a palliative solution.
C1 [Giussani, Carlo; Guida, Lelio; Trezza, Andrea; Sganzerla, Erik] Univ Milano Bicocca, Osped San Gerardo, Sch Med, Neurosurg, Monza, Italy.
   [Biassoni, Veronica; Schiavello, Elisabetta; Massimino, Maura] Fdn IRCCS Ist Nazl Tumori, Pediat Oncol, Milan, Italy.
   [Carrabba, Giorgio] Univ Milan, Fdn IRCCS Ca Gran, Sch Med, Neurosurg, Milan, Italy.
C3 San Gerardo Hospital; University of Milano-Bicocca; Fondazione IRCCS
   Istituto Nazionale Tumori Milan; University of Milan
RP Giussani, C (通讯作者)，Univ Milano Bicocca, Osped San Gerardo, Sch Med, Neurosurg, Monza, Italy.
EM
RI Carrabba, Giorgio/AAC-3746-2022; Schiavello, Elisabetta ES/K-8091-2016;
   massimino, maura/K-7310-2016; GIUSSANI, CARLO GIORGIO/AAC-2941-2022
OI Schiavello, Elisabetta ES/0000-0002-8049-7445; massimino,
   maura/0000-0002-5506-2001; GIUSSANI, CARLO GIORGIO/0000-0002-9033-2927;
   Carrabba, Giorgio/0000-0002-0170-0653
CR Amano T, 2002, CHILD NERV SYST, V18, P599, DOI 10.1007/s00381-002-0637-5
   [Anonymous], 1999, PEDIAT NEUROSURGERY
   Cohen BH, 2006, NEURO-ONCOLOGY, V8, P284, DOI [10.1215/15228517-2006-009, DOI 10.1215/15228517-2006-009]
   Fischbein NJ, 1996, PEDIATR NEUROSURG, V24, P9, DOI 10.1159/000121010
   Freeman CR, 1998, INT J RADIAT ONCOL, V40, P265, DOI 10.1016/S0360-3016(97)00572-5
   Guida L, 2019, WORLD NEUROSURG, V124, P29, DOI 10.1016/j.wneu.2018.12.096
   Hargrove D, 2006, LANCET ONCOL, V7, P241, DOI 10.1016/S1470-2045(06)70615-5
   Jansen MHA, 2012, CANCER TREAT REV, V38, P27, DOI 10.1016/j.ctrv.2011.06.007
   Jansen MH, 2015, NEURO-ONCOLOGY, V17, P160, DOI 10.1093/neuonc/nou104
   Khatua S, 2014, CHILD NERV SYST, V30, P925, DOI 10.1007/s00381-013-2350-y
   Khatua S, 2011, CHILD NERV SYST, V27, P1391, DOI 10.1007/s00381-011-1468-z
   Lobel U, 2016, NEURORADIOLOGY, V58, P1027, DOI 10.1007/s00234-016-1724-8
   Poussaint TY, 2016, NEURO-ONCOLOGY, V18, P725, DOI 10.1093/neuonc/nov256
   Roujeau T, 2011, CHILD NERV SYST, V27, P1735, DOI 10.1007/s00381-011-1538-2
   Steffen-Smith EA, 2014, BIOMED RES INT, V2014, DOI 10.1155/2014/647356
   van Zanten SEMV, 2017, J NEURO-ONCOL, V134, P231, DOI 10.1007/s11060-017-2514-9
   van Zanten SEMV, 2017, J NEURO-ONCOL, V132, P255, DOI 10.1007/s11060-016-2363-y
NR 17
TC 5
Z9 5
U1 0
U2 3
PU SPRINGER
PI NEW YORK
PA ONE NEW YORK PLAZA, SUITE 4600, NEW YORK, NY, UNITED STATES
SN 0256-7040
EI 1433-0350
J9 CHILD NERV SYST
JI Childs Nerv. Syst.
PD NOV
PY 2020
VL 36
IS 11
BP 2701
EP 2705
DI 10.1007/s00381-020-04589-4
EA MAR 2020
PG 5
WC Clinical Neurology; Pediatrics; Surgery
WE Science Citation Index Expanded (SCI-EXPANDED)
SC Neurosciences & Neurology; Pediatrics; Surgery
GA OG4CD
UT WOS:000521997800002
PM 32222799
DA 2023-08-27
ER

PT S
AU HENRY, JP
   GASNIER, B
   DESNOS, C
   SCHERMAN, D
   KREJCI, E
   MASSOULIE, J
AF HENRY, JP
   GASNIER, B
   DESNOS, C
   SCHERMAN, D
   KREJCI, E
   MASSOULIE, J
BE Wiedenmann, B
   Kvols, LK
   Arnold, R
   Riecken, EO
TI THE CATECHOLAMINE TRANSPORTER OF ADRENAL-MEDULLA CHROMAFFIN GRANULES
SO MOLECULAR AND CELL BIOLOGICAL ASPECTS OF GASTROENTEROPANCREATIC
   NEUROENDOCRINE TUMOR DISEASE
SE Annals of the New York Academy of Sciences
LA English
DT Article; Proceedings Paper
CT Conference on Molecular and Cell Biological Aspects of
   Gastroenteropancreatic Neuroendocrine Tumor Disease
CY NOV 03-07, 1993
CL FREE UNIV BERLIN, BERLIN, GERMANY
SP Free Univ Berlin
HO FREE UNIV BERLIN
ID VESICULAR MONOAMINE TRANSPORTER; AMINE TRANSPORTER; STORAGE-VESICLES
C1 INST BIOL PHYSICOCHIM, NEUROBIOL PHYSICOCHIM SERV, CNRS, UA 1112, F-75005 PARIS, FRANCE.
   ECOLE NORMALE SUPER, NEUROBIOL LAB, CNRS, UA 1857, F-75005 PARIS, FRANCE.
C3 Centre National de la Recherche Scientifique (CNRS); Centre National de
   la Recherche Scientifique (CNRS)
RI Gasnier, Bruno/AAD-9124-2022
OI Gasnier, Bruno/0000-0003-4458-6296; Scherman, Daniel/0000-0003-1207-1298
CR DARCHEN F, 1989, BIOCHEMISTRY-US, V28, P1692, DOI 10.1021/bi00430a040
   DARCHEN F, 1988, BIOCHEM PHARMACOL, V37, P4381, DOI 10.1016/0006-2952(88)90621-1
   DASILVA JN, 1993, SYNAPSE, V14, P128, DOI 10.1002/syn.890140205
   DESNOS C, 1992, J NEUROCHEM, V59, P2105
   ERICKSON JD, 1992, P NATL ACAD SCI USA, V89, P10993, DOI 10.1073/pnas.89.22.10993
   GASNIER B, 1986, MOL PHARMACOL, V29, P275
   HENRY JP, 1987, ANN NY ACAD SCI, V493, P194, DOI 10.1111/j.1749-6632.1987.tb27201.x
   HENRY JP, 1989, BIOCHEM PHARMACOL, V38, P2395, DOI 10.1016/0006-2952(89)90082-8
   ISAMBERT MF, 1989, BIOCHEMISTRY-US, V28, P2265, DOI 10.1021/bi00431a044
   ISAMBERT MF, 1992, BIOCHEMISTRY-US, V31, P1980, DOI 10.1021/bi00122a012
   JOHNSON RG, 1988, PHYSIOL REV, V68, P232, DOI 10.1152/physrev.1988.68.1.232
   KANNER BI, 1987, CRIT REV BIOCHEM MOL, V22, P1, DOI 10.3109/10409238709082546
   KREJCI E, 1993, FEBS LETT, V335, P27, DOI 10.1016/0014-5793(93)80432-T
   LEVY C, 1990, J NEUROCHEM, V54, P880, DOI 10.1111/j.1471-4159.1990.tb02333.x
   LIU YJ, 1992, CELL, V70, P539, DOI 10.1016/0092-8674(92)90425-C
   NJUS D, 1986, BIOCHIM BIOPHYS ACTA, V853, P237, DOI 10.1016/0304-4173(87)90003-6
   SCHERMAN D, 1989, ANN NEUROL, V26, P551, DOI 10.1002/ana.410260409
   SIETZEN M, 1987, NEUROSCIENCE, V22, P131, DOI 10.1016/0306-4522(87)90203-X
NR 18
TC 7
Z9 7
U1 0
U2 1
PU NEW YORK ACAD SCIENCES
PI NEW YORK
PA 2 EAST 63RD ST, NEW YORK, NY 10021 USA
SN 0077-8923
BN 0-89766-898-7; 0-89766-897-9
J9 ANN NY ACAD SCI
JI Ann.NY Acad.Sci.
PY 1994
VL 733
BP 185
EP 192
DI 10.1111/j.1749-6632.1994.tb17268.x
PG 8
WC Biochemistry & Molecular Biology; Cell Biology; Multidisciplinary
   Sciences
WE Conference Proceedings Citation Index - Science (CPCI-S); Science Citation Index Expanded (SCI-EXPANDED)
SC Biochemistry & Molecular Biology; Cell Biology; Science & Technology -
   Other Topics
GA BD10P
UT WOS:A1994BD10P00018
PM 7978866
DA 2023-08-27
ER

PT J
AU Satoh, F
   Takahashi, K
   Murakami, O
   Totsune, K
   Sone, M
   Ohneda, M
   Abe, K
   Miura, Y
   Mouri, T
AF Satoh, F
   Takahashi, K
   Murakami, O
   Totsune, K
   Sone, M
   Ohneda, M
   Abe, K
   Miura, Y
   Mouri, T
TI Immunoreactive adrenomedullin in human adrenal glands and adrenal tumors
SO CANCER DETECTION AND PREVENTION
LA English
DT Article
DE adrenomedullin; adrenal; pheochromocytoma; radioimmunoassay;
   chromatography; neuropeptide
ID NEUROPEPTIDE-Y; PHEOCHROMOCYTOMA; PLASMA; GANGLIONEUROBLASTOMA;
   NEUROBLASTOMA; TISSUE; CELLS; LUNG
AB Adrenomedullin is a potent vasodilator peptide that was isolated from human pheochromocytoma. We developed a sensitive and specific radioimmunoassay for adrenomedullin and studied the presence of adrenomedullin in human adrenal glands and adrenal tumors, including pheochromocytoma. High concentrations of immunoreactive adrenomedullin were found in normal parts of adrenal glands (cortex and medulla) (12.6 +/- 1.0 pmol/g wet wt, N = 7, mean +/- SEM). High concentrations of immunoreactive adrenomedullin were also present in the tumor tissues of pheochromocytoma (4.5 +/- 1.5 pmol/g wet wt, N = 11). Immunoreactive adrenomedullin was detected in some adrenocortical tumors, but these concentrations were much lower than those in the normal adrenal glands and pheochromocytomas. Reverse phase high-performance liquid chromatography of the normal adrenal gland and pheochromocytoma showed a peak eluting in the position of synthetic adrenomedullin 1-52. The present study has shown the presence of high concentrations of immunoreactive adrenomedullin in the normal adrenal glands and pheochromocytomas.
C1 TOHOKU UNIV, SCH MED, DEPT INTERNAL MED 2, SENDAI, MIYAGI 98077, JAPAN.
C3 Tohoku University
RI Takahashi, Kazuhiro/A-4887-2010
OI Takahashi, Kazuhiro/0000-0001-8943-1875
CR HUNTER WM, 1962, NATURE, V194, P495, DOI 10.1038/194495a0
   ICHIKI Y, 1994, FEBS LETT, V338, P6, DOI 10.1016/0014-5793(94)80106-1
   KITAMURA K, 1993, BIOCHEM BIOPH RES CO, V192, P553, DOI 10.1006/bbrc.1993.1451
   KITAMURA K, 1994, FEBS LETT, V341, P288, DOI 10.1016/0014-5793(94)80474-5
   MARTINEZ A, 1995, ENDOCRINOLOGY, V136, P4099, DOI 10.1210/en.136.9.4099
   Miller MJ, 1996, J BIOL CHEM, V271, P23345
   MOURI T, 1989, PEPTIDES, V10, P201, DOI 10.1016/0196-9781(89)90097-1
   MOURI T, 1992, CLIN SCI, V83, P205, DOI 10.1042/cs0830205
   NISHIKIMI T, 1994, HYPERTENSION, V24, P600, DOI 10.1161/01.HYP.24.5.600
   SAID SI, 1975, NEW ENGL J MED, V293, P155, DOI 10.1056/NEJM197507242930401
   SATOH F, 1995, J CLIN ENDOCR METAB, V80, P1750, DOI 10.1210/jc.80.5.1750
   Satoh F, 1996, NEUROSCI LETT, V203, P207, DOI 10.1016/0304-3940(95)12294-X
   SUDA T, 1984, J CLIN ENDOCR METAB, V58, P919, DOI 10.1210/jcem-58-5-919
   SUGO S, 1994, BIOCHEM BIOPH RES CO, V201, P1160, DOI 10.1006/bbrc.1994.1827
   SUGO S, 1994, BIOCHEM BIOPH RES CO, V203, P719, DOI 10.1006/bbrc.1994.2241
   TAKAHASHI K, 1990, ENDOCRINOL JAPON, V37, P53
   TAKAHASHI K, 1989, ENDOCRINOL JAPON, V36, P627
   TAKAHASHI K, 1993, PEPTIDES, V14, P365, DOI 10.1016/0196-9781(93)90054-K
   TAKAHASHI K, 1997, IN PRESS J NEUROCHEM
NR 19
TC 10
Z9 12
U1 0
U2 1
PU ELSEVIER SCI LTD
PI OXFORD
PA THE BOULEVARD, LANGFORD LANE, KIDLINGTON, OXFORD OX5 1GB, OXON, ENGLAND
SN 0361-090X
J9 CANCER DETECT PREV
JI Cancer Detect. Prev.
PY 1997
VL 21
IS 1
BP 51
EP 54
PG 4
WC Oncology
WE Science Citation Index Expanded (SCI-EXPANDED)
SC Oncology
GA WJ373
UT WOS:A1997WJ37300009
PM 9043763
DA 2023-08-27
ER

PT J
AU Cornelius, JF
   Schipper, J
   Tortorar, A
   Krause-Molle, Z
   Smuga, M
   Petridis, AK
   Steiger, HJ
AF Cornelius, Jan Frederick
   Schipper, Joerg
   Tortorar, Angelo
   Krause-Molle, Zarela
   Smuga, Maria
   Petridis, Athanasios K.
   Steiger, Hans-Jakob
TI Continuous and Dynamic Facial Nerve Mapping During Surgery of
   Cerebellopontine Angle Tumors: Clinical Pilot Series
SO WORLD NEUROSURGERY
LA English
DT Article
DE Acoustic neuroma; Cerebellopontine angle tumor; Facial nerve monitoring;
   Intraoperative; Skull base; Vestibular schwannoma
ID ACOUSTIC NEUROMA SURGERY; MOTOR EVOKED-POTENTIALS; VESTIBULAR SCHWANNOMA
   SURGERY; ELECTRICAL-STIMULATION; PRESERVATION; ELECTROMYOGRAPHY;
   RESECTION; REMOVAL; ARTICLE; TRACT
AB OBJECTIVE: To evaluate a new surgical tool combining suction and monopolar neurostimulation (stimulation sucker) for cerebellopontine angle (CPA) tumors. The usefulness for continuous (time) and dynamic (space) facial nerve mapping was studied.
   METHODS: Patients operated on with the stimulation sucker for a CPA tumor between April 2016 and May 2017 in a tertiary care center were identified. Clinical charts were retrospectively evaluated. The minimum follow-up time of patients was 12 months.
   RESULTS: The study population included 17 patients with tumor (6 women and 11 men). The age range was from 2 to 77 years (mean age, 45.7 years; standard error of the mean, 22.7 years). Most CPA tumors were large (Koos grade 4) vestibular schwannomas (n = 10); other pathologies included petrous meningioma (n = 1), metastasis (n = 1), medulloblastoma (n = 2), ependymoma (n = 2), and pilocytic astrocytoma (n = 1). One patient with trigeminal neuralgia served as the normal control subject. No complications because of the stimulation sucker were encountered. Clinical and radiologic outcomes compared favorably with institutional experience and literature. The stimulation sucker allowed for continuous (time) mapping of the facial nerve at the site of resection (space). This real-time feedback allowed early identification of the facial nerve. Handling and ergonomy were excellent and workflow improved. The shortcomings compared with a bayonet-shaped bipolar probe were decreased visibility of neurovascular structures and lower spatial discrimination. The new device did not simplify delicate at the brainstem and on the nerve. We think it should be considered as a complementary tool in the surgeon's armamentarium.
   CONCLUSIONS: Until now, nerve damage as assessed by neuromonitoring (e. g., facial nerve electromyography, motor evoked potential) served as surrogate for nerve function. This concept should be challenged. The studied stimulation sucker detected the facial nerve earlier than conventional techniques, preventing harm by surgical trauma. A larger, prospective study is warranted to better define its role in CPA surgery.
C1 [Cornelius, Jan Frederick; Tortorar, Angelo; Krause-Molle, Zarela; Smuga, Maria; Petridis, Athanasios K.; Steiger, Hans-Jakob] Heinrich Heine Univ, Dept Neurosurg, Med Fac, Dusseldorf, Germany.
   [Schipper, Joerg] Heinrich Heine Univ, Dept ENT Surg, Med Fac, Dusseldorf, Germany.
C3 Heinrich Heine University Dusseldorf; Heinrich Heine University
   Dusseldorf
RP Cornelius, JF (通讯作者)，Heinrich Heine Univ, Dept Neurosurg, Med Fac, Dusseldorf, Germany.
EM
RI Cornelius, Jan/AFR-1507-2022; Steiger, Hans-Jakob/AAN-8307-2021
OI Cornelius, Jan/0000-0002-7663-4750; Steiger,
   Hans-Jakob/0000-0002-8249-2998
CR Amano M, 2011, ACTA NEUROCHIR, V153, P1059, DOI 10.1007/s00701-010-0937-6
   Bendet E, 1999, AM J OTOL, V20, P793
   Boublata L, 2017, WORLD NEUROSURG, V103, P231, DOI 10.1016/j.wneu.2017.02.053
   Chen LH, 2009, CLIN NEUROL NEUROSUR, V111, P47, DOI 10.1016/j.clineuro.2008.07.012
   Deli G, 2011, PARKINSONISM RELAT D, V17, P50, DOI 10.1016/j.parkreldis.2010.10.012
   Fenton JE, 1999, CLIN OTOLARYNGOL, V24, P483, DOI 10.1046/j.1365-2273.1999.00287.x
   Kombos T, 2000, NEUROL MED-CHIR, V40, P501, DOI 10.2176/nmc.40.501
   LALWANI AK, 1994, OTOLARYNG HEAD NECK, V111, P561, DOI 10.1016/S0194-5998(94)70522-4
   Liu BY, 2007, CHINESE MED J-PEKING, V120, P323, DOI 10.1097/00029330-200702020-00013
   Liu SW, 2015, CLIN NEUROL NEUROSUR, V133, P83, DOI 10.1016/j.clineuro.2015.03.016
   Macdonald David B, 2006, J Clin Monit Comput, V20, P347, DOI 10.1007/s10877-006-9033-0
   MacDonald DB, 2003, SPINE, V28, P194, DOI 10.1097/00007632-200301150-00018
   Matthies C, 2011, CLIN NEUROL NEUROSUR, V113, P872, DOI 10.1016/j.clineuro.2011.06.011
   Neuloh G, 2004, J NEUROSURG, V100, P389, DOI 10.3171/jns.2004.100.3.0389
   Neuloh G, 2009, J NEUROL NEUROSUR PS, V80, P417, DOI 10.1136/jnnp.2008.157792
   Neuloh G, 2007, J NEUROSURG, V106, P582, DOI 10.3171/jns.2007.106.4.582
   Prell J, 2017, HNO, V65, P404, DOI 10.1007/s00106-017-0340-1
   Prell J, 2014, CLIN NEUROPHYSIOL, V125, P415, DOI 10.1016/j.clinph.2013.07.015
   Prell J, 2010, NEUROSURGERY, V66, P1064, DOI 10.1227/01.NEU.0000369605.79765.3E
   Raabe A, 2014, J NEUROSURG, V120, P1015, DOI 10.3171/2014.1.JNS13909
   Raftopoulos C, 2005, ACTA NEUROCHIR, V147, P697, DOI 10.1007/s00701-005-0544-0
   Romstock J, 2000, J NEUROSURG, V93, P586, DOI 10.3171/jns.2000.93.4.0586
   Samii M, 2010, J NEUROSURG, V112, P860, DOI 10.3171/2009.7.JNS0989
   Schlake HP, 1999, NEUROL RES, V21, P11, DOI 10.1080/01616412.1999.11740885
   SILVERSTEIN H, 1986, OTOLARYNG HEAD NECK, V94, P409, DOI 10.1177/019459988609400335
   SILVERSTEIN H, 1993, AM J OTOL, V14, P524
   SILVERSTEIN H, 1991, SKULL BASE SURG, V1, P127, DOI 10.1055/s-2008-1056993
   Slotty PJ, 2017, J NEUROSURG, V126, P281, DOI 10.3171/2015.11.JNS15991
   YINGLING CD, 1992, OTOLARYNG CLIN N AM, V25, P413
NR 29
TC 6
Z9 8
U1 0
U2 10
PU ELSEVIER SCIENCE INC
PI NEW YORK
PA 360 PARK AVE SOUTH, NEW YORK, NY 10010-1710 USA
SN 1878-8750
EI 1878-8769
J9 WORLD NEUROSURG
JI World Neurosurg.
PD NOV
PY 2018
VL 119
BP E855
EP E863
DI 10.1016/j.wneu.2018.07.286
PG 9
WC Clinical Neurology; Surgery
WE Science Citation Index Expanded (SCI-EXPANDED)
SC Neurosciences & Neurology; Surgery
GA GX7IO
UT WOS:000447941300104
PM 30099190
DA 2023-08-27
ER

PT J
AU Vitanza, NA
   Biery, MC
   Myers, C
   Ferguson, E
   Zheng, Y
   Girard, EJ
   Przystal, JM
   Park, G
   Noll, A
   Pakiam, F
   Winter, CA
   Morris, SM
   Sarthy, J
   Cole, BL
   Leary, SES
   Crane, C
   Lieberman, NAP
   Mueller, S
   Nazarian, J
   Gottardo, R
   Brusniak, MY
   Mhyre, AJ
   Olson, JM
AF Vitanza, Nicholas A.
   Biery, Matt C.
   Myers, Carrie
   Ferguson, Eric
   Zheng, Ye
   Girard, Emily J.
   Przystal, Justyna M.
   Park, Giulia
   Noll, Alyssa
   Pakiam, Fiona
   Winter, Conrad A.
   Morris, Shelli M.
   Sarthy, Jay
   Cole, Bonnie L.
   Leary, Sarah E. S.
   Crane, Courtney
   Lieberman, Nicole A. P.
   Mueller, Sabine
   Nazarian, Javad
   Gottardo, Raphael
   Brusniak, Mi-Youn
   Mhyre, Andrew J.
   Olson, James M.
TI Optimal therapeutic targeting by HDAC inhibition in biopsy-derived
   treatment-naive diffuse midline glioma models
SO NEURO-ONCOLOGY
LA English
DT Article
DE diffuse intrinsic pontine glioma (DIPG); diffuse midline glioma; H3
   K27M-mutant (DMG); histone deacetylase inhibitor (HDACi); quisinostat;
   romidepsin
ID HISTONE DEACETYLASE INHIBITOR; INTRINSIC PONTINE GLIOMA; BRAIN-STEM
   GLIOMAS; CANCER; CLASSIFICATION; METASTASIS; ROMIDEPSIN; MIGRATION;
   DIAGNOSIS; SYSTEM
AB Background. Diffuse midline gliomas (DMGs), including diffuse intrinsic pontine gliomas (DIPGs), have a dismal prognosis, with less than 2% surviving 5 years postdiagnosis. The majority of DIPGs and all DMGs harbor mutations altering the epigenetic regulatory histone tail (H3 K27M). Investigations addressing DMG epigenetics have identified a few promising drugs, including the HDAC inhibitor (HDACi) panobinostat. Here, we use clinically relevant DMG models to identify and validate other effective HDACi and their biomarkers of response.
   Methods. HDAC inhibitors were tested across biopsy-derived treatment-naive in vitro and in vivo DMG models with biologically relevant radiation resistance. RNA sequencing was performed to define and compare drug efficacy and to map predictive biomarkers of response.
   Results. Quisinostat and romidepsin showed efficacy with low nanomolar half-maximal inhibitory concentration (IC50) values (similar to 50 and similar to 5 nM, respectively). Comparative transcriptome analyses across quisinostat, romidepsin, and panobinostat showed a greater degree of shared biological effects between quisinostat and panobinostat, and less overlap with romidepsin. However, some transcriptional changes were consistent across all 3 drugs at similar biologically effective doses, such as overexpression of troponinT1 slow skeletal type (TNNT1) and downregulation of collagen type 20 alpha 1 chain (COL20A1), identifying these as potential vulnerabilities or on-target biomarkers in DMG. Quisinostat and romidepsin significantly (P < 0.0001) inhibited in vivo tumor growth.
   Conclusions. Our data highlight the utility of treatment-naive biopsy-derived models; establishes quisinostat and romidepsin as effective in vivo; illuminates potential mechanisms and/or biomarkers of DMG cell lethality due to HDAC inhibition; and emphasizes the need for brain tumor-penetrant versions of potentially efficacious agents.
C1 [Vitanza, Nicholas A.; Biery, Matt C.; Myers, Carrie; Ferguson, Eric; Girard, Emily J.; Park, Giulia; Noll, Alyssa; Pakiam, Fiona; Winter, Conrad A.; Morris, Shelli M.; Sarthy, Jay; Leary, Sarah E. S.; Brusniak, Mi-Youn; Mhyre, Andrew J.; Olson, James M.] Fred Hutchinson Canc Res Ctr, 1124 Columbia St, Seattle, WA 98104 USA.
   [Vitanza, Nicholas A.; Leary, Sarah E. S.; Olson, James M.] Univ Washington, Seattle Childrens Hosp, Dept Pediat, Div Pediat Hematol Oncol, Seattle, WA 98195 USA.
   [Zheng, Ye; Gottardo, Raphael] Fred Hutchinson Canc Res Ctr, Vaccine & Infect Dis Div, 1124 Columbia St, Seattle, WA 98104 USA.
   [Przystal, Justyna M.; Mueller, Sabine; Nazarian, Javad] Univ Childrens Hosp Zurich, Zurich, Switzerland.
   [Noll, Alyssa] Univ Washington, Mol & Cellular Biol Grad Program, Seattle, WA 98195 USA.
   [Noll, Alyssa] Univ Washington, Med Scientist Training Program, Seattle, WA 98195 USA.
   [Cole, Bonnie L.] Seattle Childrens Hosp, Dept Labs, Seattle, WA 98105 USA.
   [Cole, Bonnie L.; Lieberman, Nicole A. P.] Univ Washington, Sch Med, Dept Lab Med & Pathol, Seattle, WA USA.
   [Crane, Courtney] Seattle Childrens Res Inst, Ben Towne Ctr Childhood Canc Res, Seattle, WA USA.
   [Mueller, Sabine] Univ Calif San Francisco, San Francisco, CA 94143 USA.
   [Nazarian, Javad] Childrens Natl Med Ctr, Dept Genet Med Res, Washington, DC 20010 USA.
   [Gottardo, Raphael] Fred Hutchinson Canc Res Ctr, Publ Hlth Sci Div, 1124 Columbia St, Seattle, WA 98104 USA.
   [Gottardo, Raphael] Univ Washington, Dept Stat, Seattle, WA 98195 USA.
C3 Fred Hutchinson Cancer Center; Seattle Children's Hospital; University
   of Washington; University of Washington Seattle; Fred Hutchinson Cancer
   Center; University Children's Hospital Zurich; University of Washington;
   University of Washington Seattle; University of Washington; University
   of Washington Seattle; Seattle Children's Hospital; University of
   Washington; University of Washington Seattle; Seattle Children's
   Hospital; University of California System; University of California San
   Francisco; Children's National Health System; Fred Hutchinson Cancer
   Center; University of Washington; University of Washington Seattle
RP Vitanza, NA (通讯作者)，Seattle Childrens Hosp, M-S MB 8-501,4800 Sand Point Way, Seattle, WA 98105 USA.
EM
RI Gottardo, Raphael/HCH-3169-2022
OI Gottardo, Raphael/0000-0002-3867-0232; Lieberman,
   Nicole/0000-0001-9334-8150; Vitanza, Nicholas/0000-0002-3966-4985; Noll,
   Alyssa/0000-0001-8823-313X; Winter, Conrad/0000-0002-5358-5335; Biery,
   Matthew/0000-0003-3315-982X
FU National Cancer Institute [R01 CA114567]; Seattle Run of Hope; Pediatric
   Brain Tumor Research Fund Guild of Seattle Children's Hospital; McKenna
   Claire Foundation; Unravel Pediatric Cancer; Team Cozzi Foundation; Love
   for Lucy; Julianna Sayler Foundation; Grousemont Foundation; Avery
   Huffman Defeat DIPG Foundation; Michael Mosier Defeat DIPG Foundation;
   ChadTough Foundation; Kellen Joyce Heart of a Warrior Research Fund
FX We are grateful for generous funding from the National Cancer Institute
   (R01 CA114567; JMO), the Seattle Run of Hope (NAV, JMO), the Pediatric
   Brain Tumor Research Fund Guild of Seattle Children's Hospital (NAV,
   JMO), the McKenna Claire Foundation (NAV), Unravel Pediatric Cancer
   (NAV, JMO), Team Cozzi Foundation (NAV), Love for Lucy (NAV), the
   Julianna Sayler Foundation (NAV), the Grousemont Foundation (NAV), Avery
   Huffman Defeat DIPG Foundation (NAV), Michael Mosier Defeat DIPG
   Foundation (NAV), the ChadTough Foundation (NAV), and the Kellen Joyce
   Heart of a Warrior Research Fund (MCB).
CR ALBRIGHT AL, 1993, NEUROSURGERY, V33, P1026
   Anastas JN, 2019, CANCER CELL, V36, P528, DOI 10.1016/j.ccell.2019.09.005
   Aslan JE, 2013, AM J PHYSIOL-CELL PH, V305, pC1230, DOI 10.1152/ajpcell.00053.2013
   Baik FM, 2016, JAMA OTOLARYNGOL, V142, P330, DOI 10.1001/jamaoto.2015.3617
   BARKOVICH AJ, 1991, PEDIATR NEUROSURG, V16, P73, DOI 10.1159/000120511
   Bishton MJ, 2011, BLOOD, V117, P3658, DOI 10.1182/blood-2010-11-318055
   Brabetz S, 2018, NAT MED, V24, P1752, DOI 10.1038/s41591-018-0207-3
   Chen Y, 2020, J GENE MED, V22, DOI 10.1002/jgm.3152
   Cooney T, 2017, NEURO-ONCOLOGY, V19, P1279, DOI 10.1093/neuonc/nox107
   Di Pietro C, 2019, EXP NEUROL, V312, P33, DOI 10.1016/j.expneurol.2018.11.004
   DZIEGIELEWSKA KM, 1991, J PHYSIOL-LONDON, V439, P215, DOI 10.1113/jphysiol.1991.sp018664
   Grasso CS, 2015, NAT MED, V21, P555, DOI 10.1038/nm.3855
   Gupta N, 2018, NEURO-ONCOLOGY, V20, P1547, DOI 10.1093/neuonc/noy070
   Halvorson KG, 2015, PLOS ONE, V10, DOI 10.1371/journal.pone.0118926
   Herz HM, 2014, SCIENCE, V345, P1065, DOI 10.1126/science.1255104
   Iancu-Rubin C, 2012, EXP HEMATOL, V40, P564, DOI 10.1016/j.exphem.2012.02.004
   Ibrahim N, 2016, CANCER MED-US, V5, P3041, DOI 10.1002/cam4.862
   Ishihara E, 2019, NEUROL RES, V41, P1043, DOI 10.1080/01616412.2019.1672390
   Cuenca-Zamora EJ, 2019, INT J MOL SCI, V20, DOI 10.3390/ijms20143484
   Larsson P, 2020, SCI REP-UK, V10, DOI 10.1038/s41598-020-62848-5
   Leng N, 2013, BIOINFORMATICS, V29, P1035, DOI 10.1093/bioinformatics/btt087
   Li CL, 2018, J CELL MOL MED, V22, P6190, DOI 10.1111/jcmm.13906
   Liang AJ, 2017, CANCER CELL INT, V17, DOI 10.1186/s12935-017-0458-y
   Lin GL, 2019, SCI TRANSL MED, V11, DOI 10.1126/scitranslmed.aaw0064
   Louis DN, 2016, ACTA NEUROPATHOL, V131, P803, DOI 10.1007/s00401-016-1545-1
   Marks PA, 2009, J CELL BIOCHEM, V107, P600, DOI 10.1002/jcb.22185
   Maruyama D, 2017, INT J HEMATOL, V106, P655, DOI 10.1007/s12185-017-2286-1
   Nagaraja S, 2017, CANCER CELL, V31, P635, DOI 10.1016/j.ccell.2017.03.011
   Ostrom QT, 2014, NEURO-ONCOLOGY, V16, P1, DOI [10.1093/neuonc/nov189, 10.1093/neuonc/nou327, 10.1093/neuonc/nou223]
   Otterson GA, 2010, J THORAC ONCOL, V5, P1644, DOI 10.1097/JTO.0b013e3181ec1713
   Pritchard CC, 2014, J MOL DIAGN, V16, P56, DOI 10.1016/j.jmoldx.2013.08.004
   Remke M, 2011, J CLIN ONCOL, V29, P3852, DOI 10.1200/JCO.2011.36.2798
   Rose M, 2018, MOL CARCINOGEN, V57, P167, DOI 10.1002/mc.22742
   Sadoul K, 2012, BLOOD, V120, P4215, DOI 10.1182/blood-2012-05-428011
   Sasaki K, 2017, CLIN EXP METASTAS, V34, P229, DOI 10.1007/s10585-017-9840-3
   Schwartzentruber J, 2012, NATURE, V482, P226, DOI [10.1038/nature10833, 10.1007/s13311-012-0137-6]
   Seto E, 2014, CSH PERSPECT BIOL, V6, DOI 10.1101/cshperspect.a018713
   Shi YW, 2018, LIFE SCI, V208, P161, DOI 10.1016/j.lfs.2018.07.034
   Singleton WGB, 2018, J NEUROSURG-PEDIATR, V22, P288, DOI 10.3171/2018.2.PEDS17663
   Srivatsan SR, 2020, SCIENCE, V367, P45, DOI 10.1126/science.aax6234
   Stroud MR, 2011, CURR PHARM DESIGN, V17, P4362
   Valenzuela-Fernandez A, 2008, TRENDS CELL BIOL, V18, P291, DOI 10.1016/j.tcb.2008.04.003
   Venugopal B, 2013, CLIN CANCER RES, V19, P4262, DOI 10.1158/1078-0432.CCR-13-0312
   Vitanza NA, 2019, CURR TREAT OPTION NE, V21, DOI 10.1007/s11940-019-0577-y
   Wang HL, 2020, MOL BIOL REP, V47, P433, DOI 10.1007/s11033-019-05146-2
   Wang XM, 2015, CELL BIOCHEM BIOPHYS, V73, P389, DOI 10.1007/s12013-015-0634-4
   Wood PJ, 2018, CANCER CHEMOTH PHARM, V82, P493, DOI 10.1007/s00280-018-3634-4
   Wu G, 2012, NAT GENET, V44, P251, DOI 10.1038/ng.1102
   Zaja F, 2018, LEUKEMIA LYMPHOMA, P1
   Zhao B, 2013, BMC NEUROL, V13, DOI 10.1186/1471-2377-13-210
NR 50
TC 26
Z9 26
U1 0
U2 11
PU OXFORD UNIV PRESS INC
PI CARY
PA JOURNALS DEPT, 2001 EVANS RD, CARY, NC 27513 USA
SN 1522-8517
EI 1523-5866
J9 NEURO-ONCOLOGY
JI Neuro-Oncology
PD MAR
PY 2021
VL 23
IS 3
BP 376
EP 386
DI 10.1093/neuonc/noaa249
PG 11
WC Oncology; Clinical Neurology
WE Science Citation Index Expanded (SCI-EXPANDED)
SC Oncology; Neurosciences & Neurology
GA SA0QM
UT WOS:000649004500007
PM 33130903
OA Green Published, Green Accepted, Bronze
DA 2023-08-27
ER

PT J
AU Bouchal, SM
   Naylor, RM
   Daniels, DJ
AF Bouchal, Samantha M.
   Naylor, Ryan M.
   Daniels, David J.
TI Weathering the storm: a single-institution experience with paroxysmal
   sympathetic hyperactivity after brain tumor resection in pediatric
   patients
SO JOURNAL OF NEUROSURGERY-PEDIATRICS
LA English
DT Article
DE paroxysmal sympathetic hyperactivity; sympathetic storm; autonomic
   dysfunction; brain tumor; pediatric; neurosurgery
ID INJURY; DYSAUTONOMIA
AB OBJECTIVE Paroxysmal sympathetic hyperactivity (PSH) is a complication of severe traumatic or hypoxic brain injury characterized by transient episodes of tachycardia, tachypnea, hypertension, hyperthermia, diaphoresis, and/or dystonic posturing. Posttraumatic "sympathetic storms" are associated with poor outcomes. PSH rarely occurs after brain tumor resection in pediatric patients; only 4 cases have been published since 1929. Thus, the authors sought to report their experience with postcraniotomy PSH in pediatric brain tumor patients.
   METHODS A retrospective study of patients younger than 18 years of age who underwent craniotomy for brain tumor resection at a single center by a single surgeon over a 7-year period was performed. A clinical diagnosis of postoperative PSH was recorded. Recorded outcomes included the interval between surgery and initiation of cytotoxic therapy, need for long-term CSF diversion, length of hospital stay, and survival.
   RESULTS Of the 150 patients who were included for analysis, 4 patients were diagnosed with postoperative PSH for an overall occurrence of 2.7%. PSH patients were younger than non-PSH patients (1.8 +/- 0.4 years vs 9.2 +/- 5.3 years, p = 0.010) and tended to have intraventricular tumors close to the thalamus, basal ganglia, and/or brainstem. PSH patients experienced longer hospital admissions (44.3 +/- 23.4 days vs 6.8 +/- 9.4 days, p = 0.001), a shorter interval between surgery and initiation of cytotoxic cancer-directed therapy (14.3 +/- 8.0 days vs 90.7 days +/- 232.9 days, p = 0.011), and increased need for long-term CSF diversion compared with non-PSH patients (75% vs 25%, p = 0.005). At the last follow-up, 50% of PSH patients had died compared with 13% of non-PSH patients (p = 0.094).
   CONCLUSIONS PSH is a rare postoperative complication that may affect young children with periventricular tumors and is associated with poorer clinical outcomes. Increasing awareness of this condition is vital to improving patient outcomes.
C1 [Bouchal, Samantha M.] Mayo Clin, Alix Sch Med, Rochester, MN USA.
   [Naylor, Ryan M.; Daniels, David J.] Mayo Clin, Dept Neurol Surg, Rochester, MN USA.
   [Daniels, David J.] Mayo Clin, Rochester, MN 55905 USA.
C3 Mayo Clinic; Mayo Clinic; Mayo Clinic
RP Daniels, DJ (通讯作者)，Mayo Clin, Rochester, MN 55905 USA.
EM
FU National Institute of General Medical Sciences [T32 GM65841]; Mayo
   Clinic Medical Scientist Training Program
FX Acknowledgments S.M.B. was supported by an institutional training grant
   from the National Institute of General Medical Sciences (grant no. T32
   GM65841) and the Mayo Clinic Medical Scientist Training Program.
CR Alofisan TO, 2019, PEDIATR CRIT CARE ME, V20, P252, DOI 10.1097/PCC.0000000000001811
   Baguley IJ, 2008, NEUROCRIT CARE, V8, P293, DOI 10.1007/s12028-007-9021-3
   Baguley IJ, 2014, J NEUROTRAUM, V31, P1515, DOI 10.1089/neu.2013.3301
   Branstetter JW, 2020, J PEDIATR INTENSIVE, V9, P64, DOI 10.1055/s-0039-1698758
   Choi HA, 2013, CURR NEUROL NEUROSCI, V13, DOI 10.1007/s11910-013-0370-3
   ENGEL GL, 1945, ARCH NEURO PSYCHIATR, V54, P37, DOI 10.1001/archneurpsyc.1945.02300070047004
   Feng Y, 2015, PEDIATR NEUROSURG, V50, P301, DOI 10.1159/000439282
   Fernandez-Ortega JF, 2012, J NEUROTRAUM, V29, P1364, DOI 10.1089/neu.2011.2033
   Goh KYC, 1999, PEDIATR NEUROL, V21, P742, DOI 10.1016/S0887-8994(99)00069-7
   Hendricks HT, 2007, CLIN REHABIL, V21, P545, DOI 10.1177/0269215507075260
   Ichimiya Y, 2017, BRAIN DEV-JPN, V39, P621, DOI 10.1016/j.braindev.2017.03.023
   Krishnan Y, 2020, INDIAN J MED PAEDIAT, V41, P254, DOI 10.4103/ijmpo.ijmpo_93_18
   Meyfroidt G, 2017, LANCET NEUROL, V16, P721, DOI 10.1016/S1474-4422(17)30259-4
   Penfield W, 1929, ARCH NEURO PSYCHIATR, V22, P358, DOI 10.1001/archneurpsyc.1929.02220020174010
