## Supplemental Material 1 for "Bibliometric analysis of emerging trends and research foci in brainstem tumor field over thirty years (1992-2023)": savedrecs (5).txt

FN Clarivate Analytics Web of Science
VR 1.0
PT J
AU Cansever, L
   Kocaturk, CI
   Cinar, HU
   Bedirhan, MA
AF Cansever, L.
   Kocaturk, C. I.
   Cinar, H. U.
   Bedirhan, M. A.
TI Benign Posterior Mediastinal Neurogenic Tumors: Results of a Comparative
   Study into Video-Assisted Thoracic Surgery and Thoracotomy (13 Years'
   Experience)
SO THORACIC AND CARDIOVASCULAR SURGEON
LA English
DT Article
DE thoracic surgery; schwannoma; thoracoscopy
ID VIDEOTHORACOSCOPIC EXCISION; THORACOSCOPIC RESECTION
AB Objective: Mediastinal neurogenic tumors originate from the nerve tissues of the thorax and are generally located in the posterior mediastinum. The present study was performed to compare the results of thoracotomy with those of video-assisted thoracic surgery (VATS) for the surgical treatment of posterior mediastinal neurogenic tumors.
   Methods: Twenty patients who underwent surgical resection for posterior neurogenic tumors between January 1996 and January 2009 were examined retrospectively. Thirteen (65%) patients were treated by thoracotomy (group T) and VATS was used in seven (35%) patients (group V).
   Results: The duration of surgery was shorter in group V (83.5 +/- 19 min) than in group T (124.6 +/- 16.6 min; p < 0.0001). Chest drains were withdrawn earlier in group V (after 1 day) than in group T (1.6 +/- 0.5 days; p = 0.005). The hospital stay was shorter for group V (1 day) compared with group T (3 +/- 0.9 days; p < 0.0001) and group V required fewer analgesics than group T (p < 0.0001).
   Conclusion: VATS is the preferred treatment for posterior neurogenic tumors that show no preoperative signs of malignancy and do not involve the medulla spinalis.
C1 [Cansever, L.; Kocaturk, C. I.; Cinar, H. U.; Bedirhan, M. A.] Yedikule Hosp Chest Dis & Thorac Surg, Dept Thorac Surg, TR-34325 Bahcesehir Istanbul, Turkey.
C3 Istanbul Yedikule Chest Diseases & Thoracic Surgery Training & Research
   Hospital
RP Cansever, L (通讯作者)，Yedikule Hosp Chest Dis & Thorac Surg, Dept Thorac Surg, Tahtakale Mahallesi Ispartakule Mevkii Patara Ada, TR-34325 Bahcesehir Istanbul, Turkey.
EM
RI Cansever, Levent/HLX-4971-2023; Cansever, Levent/R-5851-2019; Bedirhan,
   Mehmet Ali/AGJ-8323-2022
OI Cansever, Levent/0000-0003-4928-1790; Cansever,
   Levent/0000-0003-4928-1790; Cinar, Huseyin Ulas/0000-0003-0737-2862
CR Asamura H, 1999, Respirology, V4, P9, DOI 10.1046/j.1440-1843.1999.00154.x
   Cardillo G, 2008, EUR J CARDIO-THORAC, V34, P1210, DOI 10.1016/j.ejcts.2008.09.006
   Gossot D, 2007, EUR J CARDIO-THORAC, V32, P848, DOI 10.1016/j.ejcts.2007.09.003
   ISHIDA T, 1994, SURG ENDOSC-ULTRAS, V8, P402, DOI 10.1007/BF00642441
   LANDRENEAU RJ, 1992, CHEST, V102, P1288, DOI 10.1378/chest.102.4.1288
   LOVE JG, 1952, SURG GYNECOL OBSTET, V94, P161
   Reeder L B, 2000, Semin Thorac Cardiovasc Surg, V12, P261, DOI 10.1053/stcs.2000.16738
   RIBET ME, 1994, ANN THORAC SURG, V58, P1091, DOI 10.1016/0003-4975(94)90464-2
   RIQUET M, 1995, ANN THORAC SURG, V60, P943, DOI 10.1016/0003-4975(95)00487-6
   ROVIARO G, 1994, ANN THORAC SURG, V58, P1679, DOI 10.1016/0003-4975(94)91658-6
   Sakumoto N, 2000, SURG TODAY, V30, P974, DOI 10.1007/s005950070016
   Solaini L, 2008, SURG ENDOSC, V22, P298, DOI 10.1007/s00464-007-9586-0
   Yamaguchi Masafumi, 2004, Ann Thorac Cardiovasc Surg, V10, P148
NR 13
TC 16
Z9 17
U1 0
U2 3
PU GEORG THIEME VERLAG KG
PI STUTTGART
PA RUDIGERSTR 14, D-70469 STUTTGART, GERMANY
SN 0171-6425
J9 THORAC CARDIOV SURG
JI Thorac. Cardiovasc. Surg.
PD DEC
PY 2010
VL 58
IS 8
BP 473
EP 475
DI 10.1055/s-0030-1250376
PG 3
WC Cardiac & Cardiovascular Systems; Respiratory System; Surgery
WE Science Citation Index Expanded (SCI-EXPANDED)
SC Cardiovascular System & Cardiology; Respiratory System; Surgery
GA 685JC
UT WOS:000284624000008
PM 21110270
DA 2023-08-27
ER

PT J
AU ROSENHALL, U
AF ROSENHALL, U
TI PHASE AUDIOMETRY IN NEUROAUDIOLOGICAL PRACTICE
SO ACTA OTO-LARYNGOLOGICA
LA English
DT Article
DE DIRECTIONAL HEARING; CEREBELLOPONTINE ANGLE TUMORS; BRAIN-STEM LESIONS;
   TEMPORAL LOBE LESIONS
AB Six groups of subjects, altogether 107 in number, were studied with phase audiometry. Three control groups were studied (normal-hearing healthy controls, persons with cochlear hearing losses, and patients with neurological disease not affecting the auditory system), and three patient groups were studied (patients with cerebellopontine angle tumours, with brainstem lesions, and with temporal lobe lesions). Two phase audiometers were used. The sensitivity was 85% for patients with cerebellopontine angle tumours, 71% for patients with brainstem lesions, and 69% for patients with temporal lobe lesions. There was good agreement between the phase audiometers, though one of them (BIAB Phase Audiometer) had better results for two of the patient groups. Phase audiometry can be recommended in neuroaudiological practice.
RP ROSENHALL, U (通讯作者)，GOTHENBURG UNIV,SAHLGRENS HOSP,EAR NOSE & THROAT CLIN,DEPT AUDIOL,S-41345 GOTHENBURG,SWEDEN.
CR ALMQVIST U, 1989, SCAND AUDIOL, V18, P155, DOI 10.3109/01050398909070739
   BARS DM, 1985, ARCH OTOLARRYNGOL, V11, P17
   BOSATRA A, 1976, AUDIOLOGY, V15, P141
   CRANFORD J, 1971, SCIENCE, V172, P286, DOI 10.1126/science.172.3980.286
   GROEN JJ, 1969, ACTA OTOLARYNGOL STO, V67, P236
   HAUSLER, 1983, ACTA OTOLARYNGOL S S, V400, P1
   HAUSLER R, 1980, BRAIN RES, V191, P589, DOI 10.1016/0006-8993(80)91312-8
   Hirsch A, 1980, Acta Otolaryngol Suppl, V369, P1
   Liden G, 1983, Adv Otorhinolaryngol, V29, P72
   LIDEN G, 1973, ADV OTORHINOLARYNGOL, P271
   MATZKER J, 1959, Z Laryngol Rhinol Otol, V38, P277
   NILSSON R, 1976, ACTA OTO-LARYNGOL, V81, P291, DOI 10.3109/00016487609119965
   OSENHALL U, 1991, VESTIBULAR IDAGNOSIS, P21
   ROSENHALL U, 1985, SCAND AUDIOL, V14, P187, DOI 10.3109/01050398509045940
   RROSENHAMER H, 1977, SCAND AUDIOL, V6, P205
   RUSSOLO M, 1983, AUDIOLOGY, V22, P50
   THOMSEN J, 1981, ARCH OTOLARYNGOL, V107, P601
NR 17
TC 4
Z9 4
U1 0
U2 0
PU SCANDINAVIAN UNIVERSITY PRESS
PI OSLO
PA PO BOX 2959 TOYEN, JOURNAL DIVISION CUSTOMER SERVICE, N-0608 OSLO,
   NORWAY
SN 0001-6489
J9 ACTA OTO-LARYNGOL
JI Acta Oto-Laryngol.
PD MAY
PY 1992
VL 112
IS 3
BP 429
EP 434
DI 10.3109/00016489209137423
PG 6
WC Otorhinolaryngology
WE Science Citation Index Expanded (SCI-EXPANDED)
SC Otorhinolaryngology
GA JD143
UT WOS:A1992JD14300005
PM 1441984
DA 2023-08-27
ER

PT J
AU Flynn, AA
   Green, AJ
   Pedley, RB
   Boxer, GM
   Boden, R
   Begent, RHJ
AF Flynn, AA
   Green, AJ
   Pedley, RB
   Boxer, GM
   Boden, R
   Begent, RHJ
TI A mouse model for calculating the absorbed beta-particle dose from
   I-131- and Y-90-labeled immunoconjugates, including a method for dealing
   with heterogeneity in kidney and tumor
SO RADIATION RESEARCH
LA English
DT Article
ID RADIOLABELED ANTIBODY DISTRIBUTION; EMITTING RADIONUCLIDES; XENOGRAFT
   MODEL; DOSIMETRY; SIZE; RADIOIMMUNOTHERAPY; CEA; NONUNIFORMITY;
   RADIOTHERAPY; CURABILITY
AB Conventional internal radiation dosimetry methods assume that the beta -particle energy is absorbed uniformly and completely in the source organ and that the radioactivity is distributed uniformly in the source. However, in mice, a considerable proportion of the beta -particle energy can escape the source organ, resulting in large cross-organ doses. Furthermore, the distribution of radioactivity is generally heterogeneous in kidney and tumor. Therefore, a model was developed to account for cross-organ doses and for the effects of heterogeneity in kidney and tumor in mice for two of the most important radionuclides used in therapy,I-131 and Y-90. Most mouse organs were modeled as single-compartment ellipsoids or cylinders, while heterogeneity in kidney and in tumor was addressed by using two compartments to represent the cortex and the medulla and viable and necrotic cells, respectively. The dimensions of these models were taken from previous studies, with the exception of kidney and tumor, which were defined using radioluminography and mosaics of high-power microscopy images. The absorbed fractions in each compartment were calculated using beta -particle point dose kernels, The self-organ dose was significantly higher for I-131 compared to 90Y in all compartments, but a considerable amount of beta -particle energy was shown to escape! the source organ for both radionuclides, with. as much as 85% and 36% escaping the marrow for Y-90 and I-131, respectively. The cortex was found to occupy a greater proportion of the total kidney volume than the medulla, and consequently the self-dose was higher in the cortex, In addition, the thickness of the viable shell in the tumor increased,vith tumor size, as did the self-dose fractions in both necrotic and viable areas. This dosimetry model improves dose estimates in mice and gives a conceptual basis for considering dosimetry in humans. (C) 2001 by Radiation Research Society.
C1 UCL Royal Free & Univ Coll Med Sch, Dept Oncol, CRC, Targeting & Imaging Grp, London NW3 2PF, England.
C3 University of London; University College London; UCL Medical School
RP Flynn, AA (通讯作者)，UCL Royal Free & Univ Coll Med Sch, Dept Oncol, CRC, Targeting & Imaging Grp, Royal Free Campus,Rowland Hill St, London NW3 2PF, England.
EM
CR BARDIES M, 1994, PHYS MED BIOL, V39, P961, DOI 10.1088/0031-9155/39/6/004
   Behr TM, 1997, J NUCL MED, V38, P409
   BERGER MJ, 1971, 7 MIRD SOC NUCL MED
   Casey JL, 1996, BRIT J CANCER, V74, P1397, DOI 10.1038/bjc.1996.555
   CHESTER KA, 1994, LANCET, V343, P455, DOI 10.1016/S0140-6736(94)92695-6
   Chung JK, 1999, ANN NUCL MED, V13, P303, DOI 10.1007/BF03164868
   DURBIN PW, 1992, HEALTH PHYS, V63, P427, DOI 10.1097/00004032-199210000-00007
   Flynn AA, 1999, INT J RADIAT ONCOL, V43, P183, DOI 10.1016/S0360-3016(98)00275-2
   Flynn AA, 1999, PHYS MED BIOL, V44, pN151, DOI 10.1088/0031-9155/44/7/402
   FRAKER PJ, 1978, BIOCHEM BIOPH RES CO, V80, P849, DOI 10.1016/0006-291X(78)91322-0
   HUI TE, 1994, CANCER, V73, P951
   HUMM JL, 1990, J NUCL MED, V31, P75
   LAROCHE S, 1998, MICROSC ANAL, V5, P5
   LOEVINGER R, 1968, SCHEMA ABSORBED DOSE
   Loevinger R., 1988, MIRD PRIMER ABSORBED
   Muthuswamy MS, 1998, J NUCL MED, V39, P1243
   ODONOGHUE JA, 1995, J NUCL MED, V36, P1902
   PEDLEY RB, 1993, BRIT J CANCER, V68, P69, DOI 10.1038/bjc.1993.288
   PEDLEY RB, 1987, EUR J NUCL MED, V13, P197
   ROBERSON PL, 1994, CANCER, V73, P912, DOI 10.1002/1097-0142(19940201)73:3+<912::AID-CNCR2820731325>3.0.CO;2-P
   TOM BH, 1976, IN VITRO CELL DEV B, V12, P180
   WHELDON TE, 1991, RADIOTHER ONCOL, V21, P91, DOI 10.1016/0167-8140(91)90080-Z
   YORKE ED, 1993, MED PHYS, V20, P543, DOI 10.1118/1.597050
NR 23
TC 45
Z9 49
U1 0
U2 3
PU RADIATION RESEARCH SOC
PI LAWRENCE
PA 810 E TENTH STREET, LAWRENCE, KS 66044 USA
SN 0033-7587
J9 RADIAT RES
JI Radiat. Res.
PD JUL
PY 2001
VL 156
IS 1
BP 28
EP 35
DI 10.1667/0033-7587(2001)156[0028:AMMFCT]2.0.CO;2
PG 8
WC Biology; Biophysics; Radiology, Nuclear Medicine & Medical Imaging
WE Science Citation Index Expanded (SCI-EXPANDED)
SC Life Sciences & Biomedicine - Other Topics; Biophysics; Radiology,
   Nuclear Medicine & Medical Imaging
GA 452MG
UT WOS:000169861700005
PM 11418070
DA 2023-08-27
ER

PT J
AU Suzuki, M
   Yamada, C
   Inoue, R
   Kashio, A
   Saito, Y
   Nakanishi, W
AF Suzuki, Mitsuya
   Yamada, Chikako
   Inoue, Rika
   Kashio, Akinori
   Saito, Yuki
   Nakanishi, Wakako
TI Analysis of Vestibular Testing in Patients With Vestibular Schwannoma
   Based on the Nerve of Origin, the Localization, and the Size of the
   Tumor
SO OTOLOGY & NEUROTOLOGY
LA English
DT Article
DE Amplitude; Caloric test; Latency; Vestibular evoked myogenic potential;
   Vestibular schwannoma
ID EVOKED MYOGENIC POTENTIALS; CEREBELLOPONTINE ANGLE TUMORS; ACOUSTIC
   NEUROMAS; PREDICTION; RESPONSES; PRESSURE; SURGERY
AB Objectives: We aimed to analyze the factors influencing caloric response and vestibular evoked myogenic potential (VEMP) in vestibular schwannoma.
   Subjects: The subjects comprised 130 patients with unilateral vestibular schwannoma pathologically diagnosed by surgery.
   Method: Caloric response and the amplitude and latency of VEMP were measured and analyzed based on the nerve of origin, localization, and size of the tumor. The tumors were classified into 3 types based on localization: intracanalicular, intermediate, and medial; and into 4 grades based on size: 9 mm or less, 10 to 19 mm, 20 to 29 turn, and 30 mm or greater.
   Results: Abnormal rates of caloric response and VEMP in patients with tumors arising from the superior vestibular nerve were not significantly different from those in patients with tumors of the inferior vestibular nerve. In the intermediate and medial type-but not in the intracanalicular type-a significant difference in tumor size was observed between patients with normal caloric response and those with canal paresis as also between patients with normal VEMP and those with abnormal VEMP. In patients with tumors that maximally measured 10 to 19 mm or of the intermediate type, the p- and n-wave latencies of VEMP were significantly prolonged compared with those in the normal opposite ear.
   Conclusion: 1) The nerve of origin of tumors cannot be predicted based on caloric response and VEMP. 2) In the intermediate and medial types, caloric response and the VEMP amplitude are significantly diminished in association with an increase in tumor size. 3) Prolonged VEMP latencies seem to be not only caused by tumor compression to the brainstem or vestibular spinal tract but also by tumor compression isolated to inferior vestibular nerve.
C1 [Suzuki, Mitsuya; Kashio, Akinori; Saito, Yuki] Univ Tokyo, Fac Med, Dept Otolaryngol, Bunkyo Ku, Tokyo 1138655, Japan.
   [Yamada, Chikako; Nakanishi, Wakako] Tokyo Metropolitan Police Hosp, Dept Otolaryngol, Tokyo, Japan.
   [Inoue, Rika] Jichi Med Univ Saitama, Med Ctr, Dept Otolaryngol, Saitama, Japan.
C3 University of Tokyo; Jichi Medical University
RP Suzuki, M (通讯作者)，Univ Tokyo, Fac Med, Dept Otolaryngol, Bunkyo Ku, 7-3-1 Hongo, Tokyo 1138655, Japan.
EM
OI Saito, Yuki/0000-0002-4825-7783
CR Badie B, 2001, OTOL NEUROTOL, V22, P696, DOI 10.1097/00129492-200109000-00024
   BERGENIUS J, 1988, ACTA OTO-LARYNGOL, V106, P361, DOI 10.3109/00016488809122258
   Berrettini S, 1996, J NEUROL SCI, V144, P24, DOI 10.1016/S0022-510X(96)00174-8
   Brackmann DE, 2000, AM J OTOL, V21, P417, DOI 10.1016/S0196-0709(00)80054-X
   Cheng BL, 2002, CHAOS, V12, P267, DOI 10.1063/1.1460942
   GREIMAN MC, 1991, AM J OTOLARYNG, V12, P12, DOI 10.1016/0196-0709(91)90068-Q
   Inoue Y, 2002, EUR ARCH OTO-RHINO-L, V259, P73, DOI 10.1007/s00405-001-0426-x
   Iwasaki S, 2005, OTOL NEUROTOL, V26, P1196, DOI 10.1097/01.mao.0000194890.44023.e6
   Matthies C, 1997, NEUROSURGERY, V40, P1
   Murofushi T, 2001, ARCH OTOLARYNGOL, V127, P1069, DOI 10.1001/archotol.127.9.1069
   NEELY JG, 1986, ANN OTO RHINOL LARYN, V95, P460, DOI 10.1177/000348948609500505
   Sartucci F, 2002, BRAIN RES BULL, V59, P59, DOI 10.1016/S0361-9230(02)00842-0
   TALLAN EM, 1993, AM J OTOL, V14, P131
   Tsutsumi T, 2000, AM J OTOL, V21, P712
   VALVASSORI GE, 1988, OTOLARYNG CLIN N AM, V21, P337
   Welgampola MS, 2005, NEUROLOGY, V64, P1682, DOI 10.1212/01.WNL.0000161876.20552.AA
   WILSON DF, 1992, LARYNGOSCOPE, V102, P961
NR 17
TC 41
Z9 42
U1 0
U2 2
PU LIPPINCOTT WILLIAMS & WILKINS
PI PHILADELPHIA
PA 530 WALNUT ST, PHILADELPHIA, PA 19106-3621 USA
SN 1531-7129
J9 OTOL NEUROTOL
JI Otol. Neurotol.
PD OCT
PY 2008
VL 29
IS 7
BP 1027
EP 1031
DI 10.1097/MAO.0b013e3181845854
PG 5
WC Clinical Neurology; Otorhinolaryngology
WE Science Citation Index Expanded (SCI-EXPANDED)
SC Neurosciences & Neurology; Otorhinolaryngology
GA 365ZQ
UT WOS:000260449100024
PM 18698272
DA 2023-08-27
ER

PT J
AU Venneti, S
   Santi, M
   Felicella, MM
   Yarilin, D
   Phillips, JJ
   Sullivan, LM
   Martinez, D
   Perry, A
   Lewis, PW
   Thompson, CB
   Judkins, AR
AF Venneti, Sriram
   Santi, Mariarita
   Felicella, Michelle Madden
   Yarilin, Dmitry
   Phillips, Joanna J.
   Sullivan, Lisa M.
   Martinez, Daniel
   Perry, Arie
   Lewis, Peter W.
   Thompson, Craig B.
   Judkins, Alexander R.
TI A sensitive and specific histopathologic prognostic marker for H3F3A
   K27M mutant pediatric glioblastomas
SO ACTA NEUROPATHOLOGICA
LA English
DT Article
DE Pediatric glioblastoma; H3F3A mutation; K27M mutation; H3K27me3;
   Diagnostic biomarker; Prognosis; Methylation; Epigenetics
ID ACTIVATING ACVR1 MUTATIONS; INTRINSIC PONTINE GLIOMAS; HIGH-GRADE
   ASTROCYTOMAS; GENE-EXPRESSION; HISTONE H3.3; TUMORS; SUBGROUPS;
   TRIMETHYLATION; H3K27ME3; EZH2
AB Pediatric glioblastomas (GBM) are highly aggressive and lethal tumors. Recent sequencing studies have shown that similar to 30 % of pediatric GBM and similar to 80 % of diffuse intrinsic pontine gliomas show K27M mutations in the H3F3A gene, a variant encoding histone H3.3. H3F3A K27M mutations lead to global reduction in H3K27me3. Our goal was to develop biomarkers for the histopathologic detection of these tumors. Therefore, we evaluated the utility of measuring H3K27me3 global reduction as a histopathologic and prognostic biomarker and tested an antibody directed specifically against the H3.3 K27M mutation in 290 samples. The study cohort included 203 pediatric (including 38 pediatric high-grade astrocytomas) and 38 adult brain tumors of various subtypes and grades and 49 non-neoplastic reactive brain tissues. Detection of H3.3 K27M by immunohistochemistry showed 100 % sensitivity and specificity and was superior to global reduction in H3K27me3 as a biomarker in diagnosing H3F3A K27M mutations. Moreover, cases that stained positive for H3.3 K27M showed a significantly poor prognosis compared to corresponding negative tumors. These results suggest that immunohistochemical detection of H3.3 K27M is a sensitive and specific surrogate for the H3F3A K27M mutation and defines a prognostically poor subset of pediatric GBM.
C1 [Venneti, Sriram; Thompson, Craig B.] Mem Sloan Kettering Canc Ctr, Canc Biol & Genet Program, New York, NY 10065 USA.
   [Santi, Mariarita; Sullivan, Lisa M.; Martinez, Daniel] Univ Penn, Childrens Hosp Philadelphia, Perelman Sch Med, Dept Pathol & Lab Med, Philadelphia, PA 19104 USA.
   [Felicella, Michelle Madden] Henry Ford Hlth Syst, Dept Pathol, Detroit, MI USA.
   [Yarilin, Dmitry] Mem Sloan Kettering Canc Ctr, Mol Cytol Core Facil, New York, NY 10065 USA.
   [Phillips, Joanna J.; Perry, Arie] Univ Calif San Francisco, Dept Pathol, San Francisco, CA 94140 USA.
   [Lewis, Peter W.] Univ Wisconsin, Dept Biomol Chem, Madison, WI 53706 USA.
   [Judkins, Alexander R.] Univ So Calif, Keck Sch Med, Childrens Hosp Los Angeles, Dept Pathol & Lab Med, Los Angeles, CA 90027 USA.
C3 Memorial Sloan Kettering Cancer Center; University of Pennsylvania;
   Pennsylvania Medicine; Childrens Hospital of Philadelphia; Henry Ford
   Health System; Henry Ford Hospital; Memorial Sloan Kettering Cancer
   Center; University of California System; University of California San
   Francisco; University of Wisconsin System; University of Wisconsin
   Madison; Children's Hospital Los Angeles; University of Southern
   California
RP Venneti, S (通讯作者)，Mem Sloan Kettering Canc Ctr, Canc Biol & Genet Program, New York, NY 10065 USA.
EM
RI Lewis, Peter W/C-4348-2008
OI Lewis, Peter/0000-0002-9816-7823; Sullivan, Lisa/0000-0003-0726-7149
FU NIH [P30 CA008748]; MSKCC brain tumor center; NCI [K08 CA181475,
   R01CA168802]
FX We thank the MSKCC cytology core (supported by NIH P30 CA008748)
   facility and the Geoffrey Bene Translational core for expert technical
   assistance. We thank C. David Allis and Jason T. Huse for helpful
   comments. This work was supported in part by grants from the MSKCC brain
   tumor center and the NCI to S. V. (K08 CA181475) and the NCI
   (R01CA168802) to C. B. T. We thank Dr. Tullia Lindsten for critical
   reading of the manuscript. C. B. T is co-founder of Agios
   Pharmaceuticals and has financial interest in Agios. The authors of this
   study declare no other potential conflicts of interest.
CR Aihara K, 2014, NEURO-ONCOLOGY, V16, P140, DOI 10.1093/neuonc/not144
   Bender S, 2013, CANCER CELL, V24, P660, DOI 10.1016/j.ccr.2013.10.006
   Buczkowicz P, 2014, ACTA NEUROPATHOL, V128, P573, DOI 10.1007/s00401-014-1319-6
   Buczkowicz P, 2014, NAT GENET, V46, P451, DOI 10.1038/ng.2936
   Chan ES, 2013, PEDIATR DEVEL PATHOL, V16, P272, DOI 10.2350/12-06-1216-OA.1
   Chan KM, 2013, GENE DEV, V27, P985, DOI 10.1101/gad.217778.113
   Khuong-Quang DA, 2012, ACTA NEUROPATHOL, V124, P439, DOI 10.1007/s00401-012-0998-0
   Fontebasso AM, 2014, NAT GENET, V46, P462, DOI 10.1038/ng.2950
   Fontebasso AM, 2013, BRAIN PATHOL, V23, P210, DOI 10.1111/bpa.12023
   Gessi M, 2013, J NEURO-ONCOL, V112, P67, DOI 10.1007/s11060-012-1040-z
   Gielen GH, 2013, AM J CLIN PATHOL, V139, P345, DOI 10.1309/AJCPABOHBC33FVMO
   Hojfeldt JW, 2013, NAT REV DRUG DISCOV, V12, P917, DOI 10.1038/nrd4154
   Jha P, 2014, NEURO-ONCOLOGY, V16, P1607, DOI 10.1093/neuonc/nou113
   Lee RS, 2012, J CLIN INVEST, V122, P2983, DOI 10.1172/JCI64400
   Lewis PW, 2013, SCIENCE, V340, P857, DOI 10.1126/science.1232245
   Pepe MS, 2004, STAT EVALUATION MED
   Schwartzentruber J, 2012, NATURE, V482, P226, DOI [10.1038/nature10833, 10.1007/s13311-012-0137-6]
   Sturm D, 2012, CANCER CELL, V22, P425, DOI 10.1016/j.ccr.2012.08.024
   Sullivan LM, 2012, AM J SURG PATHOL, V36, P73, DOI 10.1097/PAS.0b013e31822cfa7e
   Taylor KR, 2014, NAT GENET, V46, P457, DOI 10.1038/ng.2925
   Venneti S, 2013, BRAIN PATHOL, V23, P558, DOI 10.1111/bpa.12042
   Venneti S, 2013, J NEUROPATH EXP NEUR, V72, P298, DOI 10.1097/NEN.0b013e3182898113
   Venneti S, 2011, AM J SURG PATHOL, V35, P1463, DOI 10.1097/PAS.0b013e318224d2cd
   Wu G, 2014, NAT GENET, V46, P444, DOI 10.1038/ng.2938
   Wu G, 2012, NAT GENET, V44, P251, DOI 10.1038/ng.1102
NR 25
TC 93
Z9 100
U1 0
U2 11
PU SPRINGER
PI NEW YORK
PA 233 SPRING ST, NEW YORK, NY 10013 USA
SN 0001-6322
EI 1432-0533
J9 ACTA NEUROPATHOL
JI Acta Neuropathol.
PD NOV
PY 2014
VL 128
IS 5
BP 743
EP 753
DI 10.1007/s00401-014-1338-3
PG 11
WC Clinical Neurology; Neurosciences; Pathology
WE Science Citation Index Expanded (SCI-EXPANDED)
SC Neurosciences & Neurology; Pathology
GA AS1RL
UT WOS:000344058200011
PM 25200322
OA Green Published, hybrid
DA 2023-08-27
ER

PT J
AU Molina, ES
   van Eck, ATCJ
   Sauerland, C
   Schipmann, S
   Horstmann, G
   Stummer, W
   Brokinkel, B
AF Molina, Eric Suero
   van Eck, Albertus T. C. J.
   Sauerland, Cristina
   Schipmann, Stephanie
   Horstmann, Gerhard
   Stummer, Walter
   Brokinkel, Benjamin
TI Local Tumor Control and Clinical Symptoms After Gamma Knife Radiosurgery
   for Residual and Recurrent Vestibular Schwannomas
SO WORLD NEUROSURGERY
LA English
DT Article
DE Gamma Knife radiosurgery; Microsurgery; Neuroma, acoustic; Schwannoma,
   vestibular
ID ACOUSTIC NEUROMA; FOLLOW-UP; SURGERY; MANAGEMENT; MICROSURGERY;
   RESECTION; EXPERIENCE; OUTCOMES
AB BACKGROUND: The use of Gamma Knife radiosurgery (GKRS) for recurrent or residual vestibular schwarmorna (VS) after microsurgery (MS) has been investigated in several retrospective studies. The purpose of this study vvas to identify potential risk factors for both neurologic deterioration and tumor progression after GKRS for previously operated VSs in a prospective setting.
   METHODS: Patients who underwent GKRS for previously operated and his topathologically confirmed VS between 1998 and 2015 vvere prospectively followed-up. Risk factors for therapy side effects and predictors for tumor control were investigated in uni- and multivariate analyses.
   RESULTS: A total of 160 individuals with a median age of 55 years were included. Median tumor, volume prior to GKRS was 1.40 cm(3) (range, 0.06-35.80 cm(3)). After a median follow-up of 36 months, hearing and facial nerve function were serviceable (modified Gardner-Robertson and HouseBrackmann grades I and II) in 7 (5%) and 82 (55%) patients, respectively. Deterioration to a nonserviceable facial nerve function after GKRS was found in 3% (3/89) and tended to increase with rising tumor volume (odds ratio, 1.65 per cm(3); 95% confidence interval, 1.00-2.71; P = 0.051). Median tumor volume prior to GKRS was higher in patients with radiologic (P = 0.020) or clinical tumor progression (P < 0.001). Critical tumor volume prior to GKRS to predict clinical and radiologic tumor progression was 1.30 cm(3) (P< 0.001) and 3.30 cm(3) (P = 0.019), respectively. However, in multivariate analyses, none of the analyzed variables were found to independently predict tumor progression.
   CONCLUSIONS: Intended submaximal resection followed by GKRS is as viable treatment for VS. Because tumor remnant size after MS is an important predictor for recurrence after adjuvant GKRS, both brainstem and cerebellar decompression and maximal safely achievable resection should remain major goals of microsurgery.
C1 [Molina, Eric Suero; Schipmann, Stephanie; Stummer, Walter; Brokinkel, Benjamin] Univ Hosp Munster, Dept Neurosurg, Munster, Germany.
   [van Eck, Albertus T. C. J.; Horstmann, Gerhard] Gamma Knife Ctr Krefeld, Krefeld, Germany.
   [Sauerland, Cristina] Univ Munster, Inst Biostat & Clin Res, Munster, Germany.
C3 University of Munster; University of Munster
RP Brokinkel, B (通讯作者)，Univ Hosp Munster, Dept Neurosurg, Munster, Germany.
EM
RI Stummer, Walter/AAF-9043-2019; Sauerland, Maria Cristina/A-4154-2014;
   Stummer, Walter/AAA-3319-2020; Suero-Molina, Eric/ABC-7355-2021
OI Suero-Molina, Eric/0000-0001-5652-2105; Brokinkel,
   Benjamin/0000-0003-3462-3479
CR Bloch DC, 2004, OTOLARYNG HEAD NECK, V130, P104, DOI 10.1016/S0194-5998(03)01598-5
   Bloch O, 2011, J NEURO-ONCOL, V102, P281, DOI 10.1007/s11060-010-0315-5
   Brokinkel B, 2014, J CLIN NEUROSCI, V21, P2077, DOI 10.1016/j.jocn.2014.03.037
   Conley GS, 2010, CURR OPIN OTOLARYNGO, V18, P351, DOI 10.1097/MOO.0b013e32833c71a2
   El-Kashlan HK, 2000, AM J OTOL, V21, P389, DOI 10.1016/S0196-0709(00)80049-6
   Falcioni M, 2011, J NEUROSURG, V115, P820, DOI 10.3171/2011.5.JNS101597
   Flickinger JC, 2004, INT J RADIAT ONCOL, V60, P225, DOI 10.1016/j.ijrobp.2004.02.019
   Freeman SRM, 2007, OTOL NEUROTOL, V28, P1076, DOI 10.1097/MAO.0b013e318159e76a
   Fu VX, 2018, J NEUROSURG, V129, P137, DOI 10.3171/2017.3.JNS162033
   Gerganov VM, 2012, WORLD NEUROSURG, V77, P627, DOI 10.1016/j.wneu.2011.10.008
   Godefroy WP, 2009, OTOL NEUROTOL, V30, P629, DOI 10.1097/MAO.0b013e3181a8651f
   Hasegawa T, 2013, J NEUROSURG, V118, P557, DOI 10.3171/2012.10.JNS12523
   Hong BJ, 2014, OTOL NEUROTOL, V35, P171, DOI 10.1097/MAO.0000000000000174
   HOUSE JW, 1985, OTOLARYNG HEAD NECK, V93, P146, DOI 10.1177/019459988509300202
   KAMEYAMA S, 1994, ACTA NEUROCHIR, V129, P127, DOI 10.1007/BF01406491
   Klijn S, 2016, J NEUROSURG, V124, P1619, DOI 10.3171/2015.4.JNS142415
   Kondziolka D, 1998, NEW ENGL J MED, V339, P1426, DOI 10.1056/NEJM199811123392003
   Lee CC, 2014, J NEUROSURG, V121, P150, DOI 10.3171/2014.8.GKS141312
   Liscak R, 2009, ACTA NEUROCHIR, V151, P317, DOI 10.1007/s00701-009-0254-0
   Lobato-Polo J, 2009, NEUROSURGERY, V65, P294, DOI 10.1227/01.NEU.0000345944.14065.35
   Lunsford LD, 2005, J NEUROSURG, V102, P195, DOI 10.3171/jns.2005.102.s_supplement.0195
   Magliulo G, 2000, J OTOLARYNGOL, V29, P344
   Matthies C, 1997, NEUROSURGERY, V40, P469, DOI 10.1097/00006123-199703000-00009
   Matthies C, 1997, NEUROSURGERY, V40, P1
   Matthies C, 1997, NEUROSURGERY, V40, P481
   Milligan BD, 2012, J NEUROSURG, V116, P598, DOI 10.3171/2011.11.JNS11811
   Nagano O, 2008, J NEUROSURG, V109, P811, DOI 10.3171/JNS/2008/109/11/0811
   Nakamura H, 2000, AM J NEURORADIOL, V21, P1540
   Nonaka Y, 2013, NEUROSURGERY, V72, P103, DOI 10.1227/NEU.0b013e3182752b05
   Pan HC, 2012, J NEUROSURG, V117, P69, DOI 10.3171/2012.6.GKS12697
   Petit JH, 2001, NEUROSURGERY, V49, P1299, DOI 10.1097/00006123-200112000-00003
   Pollock BE, 1998, J NEUROSURG, V89, P944, DOI 10.3171/jns.1998.89.6.0944
   Pollock BE, 1998, J NEUROSURG, V89, P949, DOI 10.3171/jns.1998.89.6.0949
   Pollock BE, 2013, J NEUROSURG S, V119, P949
   Pollock Bruce E, 2008, Prog Neurol Surg, V21, P163, DOI 10.1159/000156904
   Prasad D, 2000, J NEUROSURG, V92, P745, DOI 10.3171/jns.2000.92.5.0745
   Rinaldi V, 2012, J NEUROL SURG PART B, V73, P21, DOI 10.1055/s-0032-1304559
   Roberson JB, 1996, AM J OTOL, V17, P307
   Roche Pierre-Hugues, 2008, Prog Neurol Surg, V21, P152, DOI 10.1159/000156902
   Samii M, 1997, NEUROSURGERY, V40, P11, DOI 10.1097/00006123-199701000-00002
   Samii M, 1997, NEUROSURGERY, V40, P29
   Samii M, 2016, J NEUROSURG, V125, P1198, DOI 10.3171/2015.8.JNS151350
   Timmer FCA, 2011, LARYNGOSCOPE, V121, P1359, DOI 10.1002/lary.21763
   Unger F, 2002, ACTA NEUROCHIR, V144, P671, DOI 10.1007/s00701-002-0950-5
   van de Langenberg R, 2009, NEURORADIOLOGY, V51, P517, DOI 10.1007/s00234-009-0529-4
   van Eck ATCJ, 2005, J NEUROSURG, V102, P204, DOI 10.3171/jns.2005.102.s_supplement.0204
   Yang SY, 2008, J NEUROL NEUROSUR PS, V79, P431, DOI 10.1136/jnnp.2007.119602
NR 47
TC 8
Z9 8
U1 0
U2 4
PU ELSEVIER SCIENCE INC
PI NEW YORK
PA 360 PARK AVE SOUTH, NEW YORK, NY 10010-1710 USA
SN 1878-8750
EI 1878-8769
J9 WORLD NEUROSURG
JI World Neurosurg.
PD FEB
PY 2019
VL 122
BP E1240
EP E1246
DI 10.1016/j.wneu.2018.11.022
PG 7
WC Clinical Neurology; Surgery
WE Science Citation Index Expanded (SCI-EXPANDED)
SC Neurosciences & Neurology; Surgery
GA HJ6UM
UT WOS:000457328100150
PM 30447443
DA 2023-08-27
ER

PT J
AU Sensu, S
   Keser, SH
   Gurbuz, Y
   Barisik, NO
   Gul, AE
AF Sensu, Sibel
   Keser, Sevinc Hallac
   Gurbuz, Yesim
   Barisik, Nagehan Ozdemir
   Gul, Aylin Ege
TI Clinicopathological Evaluation of 15 Ectopic Adrenal Tissues
SO ARCHIVES OF IRANIAN MEDICINE
LA English
DT Article
DE Clear cell tumors; Ectopic adrenal; Ovarian tumor; Testicular neoplasms
ID ADRENOCORTICAL TISSUE; CYSTADENOMA; PATIENT; TUMOR; CORD
AB Background: Ectopic adrenal tissue is a rare condition associated with embryological development defects seen in many different areas in the abdomen and pelvis. Here, we aimed to discuss the clinicopathological features of ectopic adrenal tissues diagnosed in our clinic, in light of the literature.
   Methods: We included cases of ectopic adrenal tissues incidentally detected in the specimens from patients undergoing operation for various diagnoses during 2012- March 2020. The cases were examined according to gender, age, location and accompanying pathological diagnoses. During this period, 15 cases of ectopic adrenal tissues (6 paratubal, 3 paraovarian, 2 paratesticular, 1 spermatic cord, 1 paraaortic, 1 liver capsule, 1 omentum) were detected accompanied by two endometrial carcinomas, two serous cystadenomas, one seminoma, one mixed germ cell tumor, one bilateral ovarian serous carcinoma and hepatic high-grade colon adenocarcinoma metastasis.
   Results: In this report, the fifth ectopic adrenal tissue accompanying a malignant testicular tumor, the fifth and sixth ectopic adrenal tissues occurring with ovarian serous cystadenoma, the first case observed with serous cystadenocarcinoma and the first case detected with hepatic adenocarcinoma metastasis are presented. Our cases are mostly women and adult men.
   Conclusion: Ectopic adrenal tissues can lead to hormonal problems and also to adrenal cortex and medulla neoplasms. Microscopically, they may be confused with clear cell gynecological and germ cell tumors. If the ectopic focus is misdiagnosed as an implant, a benign entity may be incorrectly reported as malignant. Also, it is important to prevent mis-staging in malignancies. For precise diagnosis, an immunopanel such as inhibin, melan A, and calretinin can be performed.
C1 [Sensu, Sibel; Gurbuz, Yesim] Istinye Univ, Med Fac, Dept Pathol, Istanbul, Turkey.
   [Keser, Sevinc Hallac; Barisik, Nagehan Ozdemir; Gul, Aylin Ege] Univ Hlth Sci, Dr Lutfi Kirdar Kartal Training & Res Hosp, Dept Pathol, Istanbul, Turkey.
C3 Istinye University; Istanbul Kartal Dr Lutfi Kirdar Training & Research
   Hospital; University of Health Sciences Turkey
RP Sensu, S (通讯作者)，Istinye Univ, Med Fac, Dept Pathol, Istanbul, Turkey.
EM
RI sensu, sibel/AGU-9782-2022
OI sensu, sibel/0000-0002-4607-780X
CR Dotto RS, 2019, UROL CASE REP, V27, DOI 10.1016/j.eucr.2019.100907
   Floyd MS, 2014, CUAJ-CAN UROL ASSOC, V8, pE176, DOI 10.5489/cuaj.1584
   Khandakar B, 2015, J CLIN DIAGN RES, V9, pED13, DOI 10.7860/JCDR/2015/15411.6638
   MARES AJ, 1980, J PEDIATR SURG, V15, P289, DOI 10.1016/S0022-3468(80)80139-4
   Mari M, 2004, UROL INT, V73, P191, DOI 10.1159/000079705
   Mazza E, 1995, J ENDOCRINOL INVEST, V18, P809, DOI 10.1007/BF03349816
   Ohsugi H, 2019, ENDOCRINOL DIAB META, DOI 10.1530/EDM-19-0073
   Penezic L, 2015, EUR UROL SUPPL, V14, pE1297, DOI [10.1016/S1569-9056(15)30334-1, DOI 10.1016/S1569-9056(15)30334-1]
   Resorlu B, 2007, ANKARA U TIP FAK MEC, V60, P93, DOI [10.1501/Tipfak_0000000230, DOI 10.1501/TIPFAK_0000000230]
   Sahin C, 2017, INT J SURG CASE REP, V33, P89, DOI 10.1016/j.ijscr.2017.02.045
   Sangeeta M, 2014, INT J BASIC APPL MED, V4, P42
   Senescende L, 2016, HERNIA, V20, P879, DOI 10.1007/s10029-016-1535-1
   Sisto JM, 2018, GYNECOL ENDOCRINOL, V34, P644, DOI 10.1080/09513590.2018.1441399
   Sugiyama T, 2015, PATHOL INT, V65, P95, DOI 10.1111/pin.12234
   Sullivan JG, 2005, BJU INT, V95, P407, DOI 10.1111/j.1464-410X.2005.05310.x
   Usta U, 2006, BALK MED J, V23, P95
   Yasar L, 2012, J GYNECOL SURG, V28, P220, DOI 10.1089/gyn.2011.0055
   Ye HH, 2009, MODERN PATHOL, V22, P175, DOI 10.1038/modpathol.2008.162
   Zengin T, 2013, CYSTOSCOPE, V1, P55
   김태형, 2008, Obstetrics & Gynecology Science, V51, P1562
NR 20
TC 1
Z9 1
U1 0
U2 3
PU ACAD MEDICAL SCIENCES I R IRAN
PI TEHRAN
PA PO BOX 19395-5655, TEHRAN, 00000, IRAN
SN 1029-2977
EI 1735-3947
J9 ARCH IRAN MED
JI Arch. Iran. Med.
PD APR
PY 2021
VL 24
IS 4
BP 301
EP 305
DI 10.34172/aim.2021.42
PG 5
WC Medicine, General & Internal
WE Science Citation Index Expanded (SCI-EXPANDED)
SC General & Internal Medicine
GA TK5CM
UT WOS:000674176500005
PM 34196190
OA hybrid
DA 2023-08-27
ER

PT J
AU Kadri, PAS
   Arnautovic, KI
   Ibn Essayed, W
   Al-Mefty, O
AF Kadri, Paulo A. S.
   Arnautovic, Kenan, I
   Ibn Essayed, Walid
   Al-Mefty, Ossama
TI Resection of Clival Chordoma Through the Anterior Clivectomy:
   2-Dimensional Operative Video
SO OPERATIVE NEUROSURGERY
LA English
DT Article
DE Anterior maxillotomy; Brainstem; Chordoma; Clivectomy; Microsurgery;
   Neuroendoscopy; Trans-sphenoidal surgery
AB Clival chordomas are rare malignant behaving tumors that grow, locally invade, metastasize, and seed, and they have a high recurrence rate.(,) The longest disease control is achieved by radical resection followed by high doses of radiation therapy, commonly proton beam. To achieve radical tumor removal, multiple surgical procedures through different approaches might be required. Since the chordoma's origin is, and remains, extradural, an extradural approach is preferred, and can lead to intradural extension. Anterior approach is frequently utilized to remove the midline-located tumor and the eroded clivus. Several midline approaches were utilized, including the transbasal, transfacial, transcervical, open door, and Lefort's maxillotomies; however, the same tumor removal can be achieved with a simple extension of the trans-sphenoidal approach, by resecting the anterior maxillary wall, of the contralateral to the lesion preponderant side. This approach coupled with the use of neuronavigation on mobile head and endoscopic-assisted technique allowed to achieve a wide and direct exposure, with the ability to resect extra- and intradural tumors.(,) Lately, the endonasal endoscopic technique became popular as an alternative; however, we found a great advantage in the ability to combine the stereoscopic microsurgical technique with the endoscopic dissection, in addition to avoiding the extensive nasal dissection and its complications. We present a case of a 63-yr old woman with an upper clivus chordoma compressing the brainstem who underwent a gross total resection by endoscopic-assisted microscopic techniques through an anterior clivectomy approach. Patient consented to the procedure and publication of her images.
C1 [Kadri, Paulo A. S.] Univ Fed Mato Grosso do Sul, Dept Neurosurg, Campo Grande, MS, Brazil.
   [Arnautovic, Kenan, I] Univ Tennessee, Semmes Murphey Neurol & Spine Inst, Hlth Sci Ctr, Memphis, TN USA.
   [Arnautovic, Kenan, I] Univ Tennessee, Dept Neurosurg, Hlth Sci Ctr, Memphis, TN USA.
   [Ibn Essayed, Walid; Al-Mefty, Ossama] Harvard Med Sch, Dept Neurosurg, Brigham & Womens Hosp, Boston, MA 02115 USA.
C3 Universidade Federal de Mato Grosso do Sul; University of Tennessee
   System; University of Tennessee Health Science Center; University of
   Tennessee System; University of Tennessee Health Science Center; Harvard
   University; Brigham & Women's Hospital; Harvard Medical School
RP Al-Mefty, O (通讯作者)，Harvard Med Sch, Skull Base Surg, Dept Neurosurg, Brigham & Womens Hosp, 75 Francis St, Boston, MA 02115 USA.
EM
CR Al-Mefty O, 2008, J NEUROSURG, V109, P783, DOI 10.3171/JNS/2008/109/11/0783
   AlMefty O, 1997, J NEUROSURG, V86, P182, DOI 10.3171/jns.1997.86.2.0182
   Arnautovic KI, 2001, J NEUROSURG, V95, P798, DOI 10.3171/jns.2001.95.5.0798
   Colli B O, 2001, Neurosurg Focus, V10, pE1
   Pamir MN, 2017, CHORDOMAS TECHNOLOGI
NR 5
TC 0
Z9 0
U1 0
U2 0
PU OXFORD UNIV PRESS INC
PI CARY
PA JOURNALS DEPT, 2001 EVANS RD, CARY, NC 27513 USA
SN 2332-4252
EI 2332-4260
J9 OPER NEUROSURG
JI Oper. Neurosurg.
PD DEC
PY 2021
VL 21
IS 6
BP E516
EP E517
DI 10.1093/ons/opab334
EA SEP 2021
PG 2
WC Clinical Neurology; Surgery
WE Science Citation Index Expanded (SCI-EXPANDED)
SC Neurosciences & Neurology; Surgery
GA XL8IS
UT WOS:000728385500029
PM 34561705
OA hybrid
DA 2023-08-27
ER

PT J
AU Karsy, M
   Gelbman, M
   Shah, P
   Balumbu, O
   Moy, F
   Arslan, E
AF Karsy, Michael
   Gelbman, Marshall
   Shah, Paarth
   Balumbu, Odessa
   Moy, Fred
   Arslan, Erol
TI Established and emerging variants of glioblastoma multiforme: review of
   morphological and molecular features
SO FOLIA NEUROPATHOLOGICA
LA English
DT Review
DE glioblastoma multiforme; GBM; brain tumours; glioma; astrocytoma;
   variant; WHO
ID PRIMITIVE NEUROECTODERMAL TUMOR; GROWTH-FACTOR-RECEPTOR; GRANULAR-CELL
   ASTROCYTOMAS; INTEGRATED GENOMIC ANALYSIS; HIGH-GRADE GLIOMA;
   GIANT-CELL; MALIGNANT GLIOMAS; MESENCHYMAL DIFFERENTIATION; GEMISTOCYTIC
   ASTROCYTOMAS; GENETIC ALTERATIONS
AB Since the recent publication of the World Health Organization brain tumour classification guidelines in 2007, a significant expansion in the molecular understanding of glioblastoma multiforme (GBM) and its pathological as well as genomic variants has been evident. The purpose of this review article is to evaluate the histopathological, molecular and clinical features surrounding emerging and currently established GBM variants. The tumours discussed include classic glioblastoma multiforme and its four genomic variants, proneural, neural, mesenchymal, classical, as well as gliosarcoma (GS), and giant cell GBM (gcGBM). Furthermore, the emerging variants include fibrillary/epithelial GBM, small cell astrocytoma (SCA), GBM with oligodendroglial component (GBMO), GBM with primitive neuroectodermal features (GBM-PNET), gemistocytic astrocytoma (GA), granular cell astrocytoma (GCA), and paediatric high-grade glioma (HGG) as well as diffuse intrinsic pontine glioma (DIPG). Better understanding of the heterogeneous nature of GBM may provide improved treatment paradigms, prognostic classification, and approaches towards molecularly targeted treatments.
C1 [Karsy, Michael; Balumbu, Odessa; Moy, Fred] New York Med Coll, Dept Neurosurg, Dept Pathol, Valhalla, NY 10595 USA.
   [Shah, Paarth] New York Med Coll, Dept Med, Valhalla, NY 10595 USA.
   [Arslan, Erol] New York Med Coll, Inst Fertil Preservat, Valhalla, NY 10595 USA.
   [Arslan, Erol] Derik State Hosp, Dept Obstet & Gynecol, Mardin, Turkey.
C3 New York Medical College; New York Medical College; New York Medical
   College; Derik State Hospital
RP Karsy, M (通讯作者)，New York Med Coll, Dept Neurosurg, Dept Pathol, Basic Sci Bldg,Room 413, Valhalla, NY 10595 USA.
EM
RI Karsy, Michael/AAH-8583-2020
OI Karsy, Michael/0000-0002-0422-7937; arslan, erol/0000-0002-9111-0744
CR Actor B, 2002, GENE CHROMOSOME CANC, V34, P416, DOI 10.1002/gcc.10087
   Aivazian K, 2012, PATHOLOGY, V44, P270, DOI 10.1097/PAT.0b013e328351bceb
   Alatakis S, 2004, J CLIN NEUROSCI, V11, P650, DOI 10.1016/j.jocn.2003.08.009
   ALBUQUERQUE L, 1992, ACTA NEUROPATHOL, V84, P680
   [Anonymous], 2012, CBTRUS STAT REP PRIM
   [Anonymous], 2007, WHO HISTOLOGICAL CLA
   ASAI A, 1994, J NEURO-ONCOL, V19, P259, DOI 10.1007/BF01053280
   Avninder S, 2006, J NEURO-ONCOL, V78, P123, DOI 10.1007/s11060-005-9077-x
   Bailey P, 1926, CLASSIFICATION TUMOR
   BANERJEE AK, 1989, CANCER, V63, P518, DOI 10.1002/1097-0142(19890201)63:3<518::AID-CNCR2820630320>3.0.CO;2-5
   Barresi V, 2006, ARCH PATHOL LAB MED, V130, P1208
   Barth RF, 2009, J NEURO-ONCOL, V94, P299, DOI 10.1007/s11060-009-9875-7
   Barut F, 2009, TURK NEUROSURG, V19, P417
   Bello L, 2001, NEUROSURGERY, V49, P380, DOI 10.1097/00006123-200108000-00022
   BIERNAT W, 1995, J NEUROPATH EXP NEUR, V54, P651, DOI 10.1097/00005072-199509000-00006
   Birner P, 2011, FOLIA NEUROPATHOL, V49, P88
   Boerman RH, 1996, J NEUROPATH EXP NEUR, V55, P973, DOI 10.1097/00005072-199609000-00004
   Bonavia R, 2011, CANCER RES, V71, P4055, DOI 10.1158/0008-5472.CAN-11-0153
   Brat DJ, 2002, AM J SURG PATHOL, V26, P750, DOI 10.1097/00000478-200206000-00008
   Brennan C, 2009, PLOS ONE, V4, DOI 10.1371/journal.pone.0007752
   BROOKS WH, 1978, ANN NEUROL, V4, P219, DOI 10.1002/ana.410040305
   BURGER PC, 1980, CANCER, V46, P1179, DOI 10.1002/1097-0142(19800901)46:5<1179::AID-CNCR2820460517>3.0.CO;2-0
   Burger PC, 2001, J NEUROPATH EXP NEUR, V60, P1099, DOI 10.1093/jnen/60.11.1099
   Burger PC, 2002, SURG PATHOLOGY NERVO
   CARSTENS PHB, 1995, ANN CLIN LAB SCI, V25, P241
   Castellano-Sanchez AA, 2003, BRAIN PATHOL, V13, P185
   Chakravarti A, 2004, J CLIN ONCOL, V22, P1926, DOI 10.1200/JCO.2004.07.193
   Chen RH, 2010, CANCER CELL, V17, P362, DOI 10.1016/j.ccr.2009.12.049
   CHIMELLI L, 1984, J NEUROPATH EXP NEUR, V43, P634, DOI 10.1097/00005072-198411000-00007
   Chin L, 2008, NATURE, V455, P1061, DOI 10.1038/nature07385
   Chorny JA, 2000, CLIN NEUROPATHOL, V19, P170
   Colman H, 2008, ARCH NEUROL-CHICAGO, V65, P877, DOI 10.1001/archneur.65.7.877
   DAUMASDUPORT C, 1988, CANCER-AM CANCER SOC, V62, P2152, DOI 10.1002/1097-0142(19881115)62:10<2152::AID-CNCR2820621015>3.0.CO;2-T
   DeCarvalho AC, 2010, STEM CELLS, V28, P181, DOI 10.1002/stem.264
   Donev K, 2010, NEUROPATH APPL NEURO, V36, P411, DOI 10.1111/j.1365-2990.2010.01078.x
   du Plessis DG, 2004, CLIN NEUROPATHOL, V23, P141
   Faury D, 2007, J CLIN ONCOL, V25, P1196, DOI 10.1200/JCO.2006.07.8626
   Feigin I, 1959, CANCER, V11, P264
   Figarella-Branger D, 2011, REV NEUROL-FRANCE, V167, P683, DOI 10.1016/j.neurol.2011.07.006
   Frederick L, 2000, CANCER RES, V60, P1383
   Galli R, 2004, CANCER RES, V64, P7011, DOI 10.1158/0008-5472.CAN-04-1364
   Geddes JF, 1996, AM J SURG PATHOL, V20, P55, DOI 10.1097/00000478-199601000-00006
   Geranmayeh F, 2007, NEUROSURGERY, V60, P159, DOI 10.1227/01.NEU.0000249192.30786.67
   Ghods AJ, 2007, STEM CELLS, V25, P1645, DOI 10.1634/stemcells.2006-0624
   Giangaspero F, 1999, SEMIN DIAGN PATHOL, V16, P91
   GRANT JW, 1989, ACTA NEUROPATHOL, V79, P305, DOI 10.1007/BF00294666
   Gravendeel LAM, 2009, CANCER RES, V69, P9065, DOI 10.1158/0008-5472.CAN-09-2307
   Han SJ, 2010, J NEUROSURG, V112, P990, DOI 10.3171/2009.9.JNS09931
   Han SJ, 2010, CANCER-AM CANCER SOC, V116, P1358, DOI 10.1002/cncr.24857
   Han SJ, 2010, J NEURO-ONCOL, V96, P313, DOI 10.1007/s11060-009-9973-6
   Han SJ, 2010, J NEUROSURG, V112, P26, DOI 10.3171/2009.3.JNS081081
   Harada K, 1998, CANCER RES, V58, P4694
   HARRIS CP, 1991, SURG NEUROL, V36, P202, DOI 10.1016/0090-3019(91)90113-N
   Hartmann C, 2009, ACTA NEUROPATHOL, V118, P469, DOI 10.1007/s00401-009-0561-9
   HAYASHI K, 1993, CANCER, V72, P850, DOI 10.1002/1097-0142(19930801)72:3<850::AID-CNCR2820720334>3.0.CO;2-4
   He J, 2001, J NEUROPATH EXP NEUR, V60, P863, DOI 10.1093/jnen/60.9.863
   Heesters MAAM, 1999, J NEURO-ONCOL, V44, P255, DOI 10.1023/A:1006398613605
   Hegi ME, 2005, NEW ENGL J MED, V352, P997, DOI 10.1056/NEJMoa043331
   Hegi ME, 2012, ACTA NEUROPATHOL, V123, P841, DOI 10.1007/s00401-011-0938-4
   HERMANSON M, 1992, CANCER RES, V52, P3213
   HOSHINO T, 1975, J NEUROPATH EXP NEUR, V34, P263, DOI 10.1097/00005072-197505000-00004
   Huse JT, 2010, NAT REV CANCER, V10, P319, DOI 10.1038/nrc2818
   Ishizawa K, 2002, ACTA NEUROPATHOL, V103, P193
   Jansen M, 2010, LANCET NEUROL, V9, P717, DOI 10.1016/S1474-4422(10)70105-8
   Johnson MW, 2010, WORLD NEUROSURG, V73, P108, DOI 10.1016/j.surneu.2009.07.036
   JONES H, 1991, NEUROPATH APPL NEURO, V17, P177, DOI 10.1111/j.1365-2990.1991.tb00713.x
   Joo Mee, 2011, Hum Pathol, DOI 10.1016/j.humpath.2011.08.015
   Jung V, 1999, J NEUROPATH EXP NEUR, V58, P993, DOI 10.1097/00005072-199909000-00009
   Kandemir NO, 2009, TURK NEUROSURG, V19, P260
   Kaneshiro D, 2009, APPL IMMUNOHISTO M M, V17, P512, DOI 10.1097/PAI.0b013e3181a2c6a4
   Karremann M, 2010, J NEURO-ONCOL, V97, P257, DOI 10.1007/s11060-009-0021-3
   Karremann M, 2009, NEURO-ONCOLOGY, V11, P323, DOI 10.1215/15228517-2008-099
   Karsy Michael, 2012, Genes Cancer, V3, P3, DOI 10.1177/1947601912448068
   Karsy M, 2010, ANTICANCER RES, V30, P4915
   KEPES JJ, 1982, CLIN NEUROPATHOL, V1, P139
   KERNOHAN JW, 1949, P STAFF M MAYO CLIN, V24, P71
   Khanna M, 2011, INDIAN J PATHOL MICR, V54, P51, DOI 10.4103/0377-4929.77324
   Kim Y, 2003, J KOREAN MED SCI, V18, P625, DOI 10.3346/jkms.2003.18.4.625
   Klink B, 2011, CELL ONCOL, V34, P261, DOI 10.1007/s13402-011-0034-8
   Koga K, 2001, CANCER RES, V61, P6248
   Kosel S, 2001, NEUROSURGERY, V48, P187
   Kozak KR, 2009, NEURO-ONCOLOGY, V11, P833, DOI 10.1215/15228517-2008-123
   Kraus JA, 2002, NEUROPATH APPL NEURO, V28, P325, DOI 10.1046/j.1365-2990.2002.00413.x
   Kraus JA, 2001, ACTA NEUROPATHOL, V101, P311
   Kros JM, 2000, J NEUROPATH EXP NEUR, V59, P679, DOI 10.1093/jnen/59.8.679
   Kros JM, 1996, ACTA NEUROPATHOL, V91, P99
   KROUWER HGJ, 1991, J NEUROSURG, V74, P399, DOI 10.3171/jns.1991.74.3.0399
   Larysz D, 2011, FOLIA NEUROPATHOL, V49, P28
   Lee D, 2012, J NEURO-ONCOL, V107, P643, DOI 10.1007/s11060-011-0790-3
   Lewis-Tuffin LJ, 2010, PLOS ONE, V5, DOI 10.1371/journal.pone.0013665
   Lusis EA, 2010, NEUROSURGERY, V67, P811, DOI 10.1227/01.NEU.0000375513.12925.5C
   MacDonald TJ, 2011, NEURO-ONCOLOGY, V13, P1049, DOI 10.1093/neuonc/nor092
   MARKESBERY WR, 1973, J NEUROPATH EXP NEUR, V32, P92, DOI 10.1097/00005072-197301000-00006
   Martinez R, 2007, CANCER GENET CYTOGEN, V175, P26, DOI 10.1016/j.cancergencyto.2007.01.006
   Martins DC, 2006, J NEURO-ONCOL, V80, P49, DOI 10.1007/s11060-006-9149-6
   Marucci G, 2011, J NEURO-ONCOL, V104, P621, DOI 10.1007/s11060-010-0523-z
   MCNAB AA, 1991, AUST NZ J OPHTHALMOL, V19, P21, DOI 10.1111/j.1442-9071.1991.tb00317.x
   MEIS JM, 1990, MODERN PATHOL, V3, P19
   MELARAGNO MJ, 1993, HUM PATHOL, V24, P805, DOI 10.1016/0046-8177(93)90020-H
   Metellus P, 2011, ANN SURG ONCOL, V18, P2937, DOI 10.1245/s10434-011-1703-6
   MeyerPuttlitz B, 1997, AM J PATHOL, V151, P853
   Miller CR, 2007, ARCH PATHOL LAB MED, V131, P397
   MORANTZ RA, 1976, J NEUROSURG, V45, P398, DOI 10.3171/jns.1976.45.4.0398
   MORK SJ, 1988, J NEUROPATH EXP NEUR, V47, P101, DOI 10.1097/00005072-198803000-00003
   Motomura K, 2012, CANC SCI
   Mueller W, 2001, BRAIN PATHOL, V11, P39
   MULLER W, 1987, NEUROSURG REV, V10, P213, DOI 10.1007/BF01782050
   Nagaishi M, 2012, BRAIN PATHOL, V22, P670, DOI 10.1111/j.1750-3639.2012.00571.x
   Nagaishi M, 2012, AM J PATHOL, V180, P1816, DOI 10.1016/j.ajpath.2012.01.027
   Nagasaka T, 2007, BRAIN TUMOR PATHOL, V24, P1, DOI 10.1007/s10014-006-0209-6
   Nakamura H, 2011, BRAIN TUMOR PATHOL, V28, P185, DOI 10.1007/s10014-011-0039-z
   Neelima R, 2011, NEUROL INDIA, V59, P918, DOI 10.4103/0028-3886.91386
   Nishikawa Ryo, 2004, Brain Tumor Pathol, V21, P53, DOI 10.1007/BF02484510
   Oh D, 1999, ARCH PATHOL LAB MED, V123, P917
   Ohgaki H, 2011, BRAIN TUMOR PATHOL, V28, P177, DOI 10.1007/s10014-011-0029-1
   Ozolek JA, 2004, MODERN PATHOL, V17, P739, DOI 10.1038/modpathol.3800109
   PALMA L, 1989, ACTA NEUROCHIR, V97, P17, DOI 10.1007/BF01577735
   Parsons DW, 2008, SCIENCE, V321, P1807, DOI 10.1126/science.1164382
   Paugh BS, 2011, J CLIN ONCOL, V29, P3999, DOI 10.1200/JCO.2011.35.5677
   Paugh BS, 2010, J CLIN ONCOL, V28, P3061, DOI 10.1200/JCO.2009.26.7252
   Peraud A, 1997, J NEUROPATH EXP NEUR, V56, P1236, DOI 10.1097/00005072-199711000-00008
   Peraud A, 1999, LAB INVEST, V79, P123
   Perry A, 2004, CANCER, V101, P2318, DOI 10.1002/cncr.20625
   Perry A, 2009, BRAIN PATHOL, V19, P81, DOI 10.1111/j.1750-3639.2008.00167.x
   PERRY JR, 1995, CANCER, V75, P2910, DOI 10.1002/1097-0142(19950615)75:12<2910::AID-CNCR2820751219>3.0.CO;2-A
   Phillips HS, 2006, CANCER CELL, V9, P157, DOI 10.1016/j.ccr.2006.02.019
   Pina-Oviedo S, 2006, ACTA NEUROPATHOL, V111, P388, DOI 10.1007/s00401-006-0050-3
   Pinto LW, 2008, VIRCHOWS ARCH, V452, P481, DOI 10.1007/s00428-007-0562-9
   Pollack IF, 2006, J NEUROSURG, V105, P418, DOI 10.3171/ped.2006.105.5.418
   Pollack IF, 2006, J CLIN ONCOL, V24, P3431, DOI 10.1200/JCO.2006.05.7265
   Pollack IF, 2011, J NEUROSURG-PEDIATR, V8, P135, DOI 10.3171/2011.5.PEDS1178
   Pollack IF, 2010, J NEURO-ONCOL, V99, P155, DOI 10.1007/s11060-010-0297-3
   Pollack IF, 2002, NEW ENGL J MED, V346, P420, DOI 10.1056/NEJMoa012224
   Prayson RA, 2009, ANN DIAGN PATHOL, V13, P36, DOI 10.1016/j.anndiagpath.2007.04.009
   Qaddoumi I, 2009, CANCER-AM CANCER SOC, V115, P5761, DOI 10.1002/cncr.24663
   Quezado M, 2005, CLIN NEUROPATHOL, V24, P163
   Quon H, 2008, COCHRANE DB SYST REV, DOI 10.1002/14651858.CD007104
   Reis RM, 2005, CELL ONCOL, V27, P319
   Reis RM, 2000, AM J PATHOL, V156, P425, DOI 10.1016/S0002-9440(10)64746-3
   Reis RM, 2001, ACTA NEUROPATHOL, V102, P422
   Rickert CH, 2002, ACTA NEUROPATHOL, V103, P309, DOI 10.1007/s00401-002-0516-x
   RINGERTZ N, 1950, ACTA PATHOL MIC SC, V27, P51
   Rodriguez FJ, 2008, CANCER-AM CANCER SOC, V113, P2779, DOI 10.1002/cncr.23899
   RUTKA JT, 1986, CANCER RES, V46, P5893
   Saad A, 2006, AM J CLIN PATHOL, V126, P602, DOI 10.1309/M67J9YF5K43AQ5KC
   Salvati M, 2009, J NEURO-ONCOL, V94, P129, DOI 10.1007/s11060-009-9815-6
   Sanders RP, 2007, PEDIATR BLOOD CANCER, V49, P888, DOI 10.1002/pbc.21272
   Sarkar C, 1997, INDIAN J MED RES, V106, P229
   Schiffman JD, 2010, CANCER RES, V70, P512, DOI 10.1158/0008-5472.CAN-09-1851
   Schittenhelm J, 2010, CLIN NEUROPATHOL, V29, P323
   Schlegel J, 1999, PATHOBIOLOGY, V67, P202, DOI 10.1159/000028073
   Schwartzentruber J, 2012, NATURE, V482, P226, DOI [10.1038/nature10833, 10.1007/s13311-012-0137-6]
   Seker A, 2006, J NEUROSURG, V105, P473, DOI 10.3171/ped.2006.105.6.473
   SHAPIRO JR, 1981, CANCER RES, V41, P2349
   Shi Y, 2008, ARCH PATHOL LAB MED, V132, P1946, DOI 10.1043/1543-2165-132.12.1946
   Shinojima N, 2004, J NEUROSURG, V101, P219, DOI 10.3171/jns.2004.101.2.0219
   Sibenaller Zita A, 2005, Neurosurg Focus, V19, pE1
   Simmons ML, 2001, CANCER RES, V61, P1122
   Singh A, 2012, CASE-BASED NEUROLOGY, P1
   Smith SF, 2006, J NEURO-ONCOL, V80, P75, DOI 10.1007/s11060-006-9158-5
   Song XY, 2011, INT J CLIN EXP PATHO, V4, P651
   Stupp R, 2009, LANCET ONCOL, V10, P459, DOI 10.1016/S1470-2045(09)70025-7
   Takeuchi H, 2012, BRAIN TUMOR PATHOL, V29, P154, DOI 10.1007/s10014-012-0100-6
   Tihan T, 2006, J NEURO-ONCOL, V76, P175, DOI 10.1007/s11060-005-4897-2
   Varlet P, 2004, NEUROSURGERY, V55, P1377, DOI 10.1227/01.NEU.0000143033.36582.40
   Verhaak RGW, 2010, CANCER CELL, V17, P98, DOI 10.1016/j.ccr.2009.12.020
   Vordermark D, 2006, BMC CANCER, V6, DOI 10.1186/1471-2407-6-247
   Wang YZ, 2012, NEURO-ONCOLOGY, V14, P518, DOI 10.1093/neuonc/nor232
   Watanabe K, 1997, LAB INVEST, V76, P277
   Watanabe K, 1998, ACTA NEUROPATHOL, V95, P559, DOI 10.1007/s004010050840
   WIKSTRAND CJ, 1983, CANCER RES, V43, P3327
   Wolanczyk M, 2010, FOLIA NEUROPATHOL, V48, P27
   Zarghooni M, 2010, J CLIN ONCOL, V28, P1337, DOI 10.1200/JCO.2009.25.5463
NR 173
TC 72
Z9 73
U1 0
U2 31
PU TERMEDIA PUBLISHING HOUSE LTD
PI POZNAN
PA KLEEBERGA 2, POZNAN, 61-615, POLAND
SN 1641-4640
EI 1509-572X
J9 FOLIA NEUROPATHOL
JI Folia Neuropathol.
PY 2012
VL 50
IS 4
BP 301
EP 321
DI 10.5114/fn.2012.32361
PG 21
WC Neurosciences; Pathology
WE Science Citation Index Expanded (SCI-EXPANDED)
SC Neurosciences & Neurology; Pathology
GA 064PL
UT WOS:000313087800001
PM 23319187
OA gold, Green Submitted
DA 2023-08-27
ER

PT J
AU Gonzalez-Gomez, P
   Bello, MJ
   Lomas, J
   Arjona, D
   Alonso, ME
   Aminoso, C
   Lopez-Marin, I
   Anselmo, NP
   Sarasa, JL
   Gutierrez, M
   Casartelli, C
   Rey, JA
AF Gonzalez-Gomez, P
   Bello, MJ
   Lomas, J
   Arjona, D
   Alonso, ME
   Aminoso, C
   Lopez-Marin, I
   Anselmo, NP
   Sarasa, JL
   Gutierrez, M
   Casartelli, C
   Rey, JA
TI Aberrant methylation of multiple genes in neuroblastic tumours:
   relationship with MYCN amplification and allelic status at 1p
SO EUROPEAN JOURNAL OF CANCER
LA English
DT Article
DE CpG island; methylation; neuroblastoma; MYCN amplification; 1p deletion;
   caspase 8; THBS1; MGMT; p73; RB1
ID CPG ISLAND METHYLATION; P16 CDKN2A GENE; PROMOTER HYPERMETHYLATION;
   CHILDHOOD NEUROBLASTOMAS; KINASE PROMOTER; P73 GENE; DELETION; CANCER;
   DEATH; SUPPRESSOR
AB Aberrant hypermethylation occurs in tumour cell CpG islands and is an important pathway for the repression of gene transcription in cancers. We investigated aberrant hypermethylation of 11 genes by methylation-specific polymerase chain reaction (PCR), after treatment of the DNA with bisulphite, and correlated the findings with MYCN amplification and allelic status at 1 p in a series of 44 neuroblastic tumours. This tumour series includes five ganglioneuromas (G), one ganglioneuroblastoma (GN) and 38 neuroblastomas (six stage 1 tumours; five stage 2 tumours; six stage 3 cases; 19 stage 4 tumours, and two stage 4S cases). Aberrant methylation of at least one of the 11 genes studied was detected in 95 % (42 of 44) of the cases. The frequencies of aberrant methylation were: 64% for thrombospondin-1 (THBS1); 30% for tissue inhibitor of metalloproteinase 3 (TIMP-3); 27% for O-6-methylguanine-DNA methyltransferase (MGMT); 25% for p73; 18% for RB1; 14% for death-associated protein kinase (DAPK), p14(ARF), p16(INK4a) and caspase 8, and 0% for TP53 and glutathione S-transferase P1 (GSTP1). No aberrant methylation was observed in four control normal tissue samples (brain and adrenal medulla). MYCN amplification was found in 11 cases (all stage 4 neuroblastomas), whereas allelic loss at 1p was identified in 16 samples (13 stage 4 and two stage 3 neuroblastomas, and one ganglioneuroma). All but one case with caspase 8 methylation also displayed MYCN amplification. Our results suggest that promoter hypermethylation is a frequent epigenetic event in the tumorigenesis of neuroblastic tumours, but no specific pattern of hypermethylated genes could be demonstrated. (C) 2003 Elsevier Science Ltd. All rights reserved.
C1 Hosp Univ La Paz, Dept C Expt, Lab Oncogenet Mol, Madrid 28046, Spain.
   Univ Sao Paulo, Fac Med Ribeirao Preto, Dept Genet, BR-14049 Ribeirao Preto, SP, Brazil.
   Fdn Jimenez Diaz, Dept Anat Patol, E-28040 Madrid, Spain.
   Hosp Univ La Paz, Dept Anat Patol, Madrid, Spain.
C3 Hospital Universitario La Paz; Universidade de Sao Paulo; Hospital
   Universitario La Paz
RP Rey, JA (通讯作者)，Hosp Univ La Paz, Dept C Expt, Lab Oncogenet Mol, Paseo Castellana 261, Madrid 28046, Spain.
RI Bello, Maria/IAN-1750-2023; Rey, Juan A/H-5026-2013
CR Bachman KE, 1999, CANCER RES, V59, P798
   Banelli B, 2000, ONCOGENE, V19, P4553, DOI 10.1038/sj.onc.1203807
   Baylin SB, 1998, ADV CANCER RES, V72, P141
   Bello MJ, 2000, CANCER GENET CYTOGEN, V120, P30, DOI 10.1016/S0165-4608(99)00249-6
   Bello MJ, 2000, CANCER GENET CYTOGEN, V116, P142, DOI 10.1016/S0165-4608(99)00122-3
   Boldin MP, 1996, CELL, V85, P803, DOI 10.1016/S0092-8674(00)81265-9
   BRODEUR GM, 1993, J CLIN ONCOL, V11, P1466, DOI 10.1200/JCO.1993.11.8.1466
   Corn PG, 1999, CANCER RES, V59, P3352
   Esteller M, 2001, CANCER RES, V61, P4689
   Esteller M, 2000, CANCER RES, V60, P129
   Esteller M, 2001, CANCER RES, V61, P3225
   FONG CT, 1989, P NATL ACAD SCI USA, V86, P3753, DOI 10.1073/pnas.86.10.3753
   Giordani L, 2002, CANCER GENET CYTOGEN, V135, P42, DOI 10.1016/S0165-4608(01)00640-9
   Gonzalez-Gomes P, 2003, CANCER GENET CYTOGEN, V142, P21, DOI 10.1016/S0165-4608(02)00799-9
   Gonzalez-Gomez P, 2003, ONCOL REP, V10, P1031
   Gonzalez-Gomez P, 2003, BRIT J CANCER, V88, P109, DOI 10.1038/sj.bjc.6600737
   Kong XT, 1997, EUR J CANCER, V33, P1962, DOI 10.1016/S0959-8049(97)00209-8
   MELINO G, 2002, NATURE REVIES CANC, V2, P1
   Nakamura M, 2001, LAB INVEST, V81, P77, DOI 10.1038/labinvest.3780213
   REY JA, 1992, INT J CANCER, V51, P703, DOI 10.1002/ijc.2910510507
   Roberts DD, 1996, FASEB J, V10, P1183, DOI 10.1096/fasebj.10.10.8751720
   Sanchez-Cespedes M, 2000, CANCER RES, V60, P892
   SCHWAB M, 2000, PATHOLOGY GENETICS T, P153
   Sherr CJ, 1996, SCIENCE, V274, P1672, DOI 10.1126/science.274.5293.1672
   Takita J, 1997, CANCER RES, V57, P907
   Takita J, 1998, ONCOGENE, V17, P3137, DOI 10.1038/sj.onc.1202232
   Takita J, 2001, ONCOGENE, V20, P4424, DOI 10.1038/sj.onc.1204521
   Tang XM, 2000, JNCI-J NATL CANCER I, V92, P1511, DOI 10.1093/jnci/92.18.1511
   Teitz T, 2000, NAT MED, V6, P529, DOI 10.1038/75007
   Teitz T, 2001, J MOL MED, V79, P428, DOI 10.1007/s001090100233
   VERSTEEG R, 1995, EUR J CANCER, V31A, P538, DOI 10.1016/0959-8049(95)00037-J
   Wong TS, 2002, CLIN CANCER RES, V8, P433
   Zhang YP, 1998, CELL, V92, P725, DOI 10.1016/S0092-8674(00)81401-4
NR 33
TC 55
Z9 58
U1 0
U2 3
PU PERGAMON-ELSEVIER SCIENCE LTD
PI OXFORD
PA THE BOULEVARD, LANGFORD LANE, KIDLINGTON, OXFORD OX5 1GB, ENGLAND
SN 0959-8049
J9 EUR J CANCER
JI Eur. J. Cancer
PD JUL
PY 2003
VL 39
IS 10
BP 1478
EP 1485
DI 10.1016/S0959-8049(03)00312-5
PG 8
WC Oncology
WE Science Citation Index Expanded (SCI-EXPANDED)
SC Oncology
GA 701ZN
UT WOS:000184199200032
PM 12826052
DA 2023-08-27
ER

PT J
AU Javadpour, M
   Mallucci, C
AF Javadpour, M
   Mallucci, C
TI The role of neuroendoscopy in the management of tectal gliomas
SO CHILDS NERVOUS SYSTEM
LA English
DT Article; Proceedings Paper
CT 2nd World Conference of the International-Study-Group-on-Neuroendoscopy
CY SEP 11-13, 2003
CL Naples, ITALY
SP Int Study Grp Neuroendoscopy
DE hydrocephalus; glioma; tectum; midbrain; neuroendoscopy; aqueductal
   stenosis
ID ENDOSCOPIC 3RD VENTRICULOSTOMY; AQUEDUCTAL STENOSIS; MIDBRAIN GLIOMAS;
   BRAIN-STEM; CHILDREN; TUMORS; PLATE; FAILURE; LESIONS; DEATH
AB Objectives: The objective was to determine the role of neuroendoscopy in the management of tectal plate gliomas. Methods: Clinical and radiological data of children and adults who underwent neuroendoscopy were reviewed. Successful outcome was defined as shunt-freedom and resolution of symptoms and signs of raised intracranial pressure. Results: Between 1 April 1998 and 31 March 2003, 11 patients (6 males and 5 females) underwent endoscopic third ventriculostomy (ETV) for the management of hydrocephalus secondary to a tectal plate glioma. This comprises 4% of 258 patients who have undergone neuroendoscopic procedures in our unit. The median age at the time of surgery was 19 (range 9-59) years. In 4 patients endoscopic biopsy of the tumour was performed at the same time. During a median follow-up period of 31 months (range 2-45), 4 patients underwent a second ETV, of whom 2 subsequently required insertion of ventriculoperitoneal (VP) shunts. Nine patients (82%) remain shunt-independent. Of the 4 tumours that were biopsied, a histological diagnosis of low-grade astrocytoma was made with certainty in only 2 cases. All tumours remained unchanged on follow-up MR imaging. Conclusions: Endoscopic third ventriculostomy can result in excellent control of hydrocephalus in patients with tectal plate gliomas. Tectal gliomas appear to be very slow growing tumours that can be managed conservatively in adults as well as children. Longer follow-up in larger studies is required.
C1 Walton Ctr Neurol & Neurosurg, Dept Neurosurg, Liverpool L9 7LJ, Merseyside, England.
C3 Walton Centre
RP Mallucci, C (通讯作者)，Walton Ctr Neurol & Neurosurg, Dept Neurosurg, Lower Lane, Liverpool L9 7LJ, Merseyside, England.
EM
RI Javadpour, Mohsen/AAX-4657-2020
OI Javadpour, Mohsen/0000-0001-9836-1741; Mallucci,
   Conor/0000-0002-5509-0547
CR ALBRIGHT AL, 1983, CANCER, V52, P2313, DOI 10.1002/1097-0142(19831215)52:12<2313::AID-CNCR2820521226>3.0.CO;2-I
   Bowers DC, 2000, PEDIATR NEUROSURG, V32, P24, DOI 10.1159/000028893
   Bulsara KR, 2003, SURG NEUROL, V59, P58, DOI 10.1016/S0090-3019(02)00942-4
   Cinalli G, 1999, J NEUROSURG, V90, P448, DOI 10.3171/jns.1999.90.3.0448
   EIFEL PJ, 1987, INT J RADIAT ONCOL, V13, P847, DOI 10.1016/0360-3016(87)90098-8
   Grant GA, 1999, PEDIATR NEUROSURG, V31, P170, DOI 10.1159/000028857
   Hader WJ, 2002, J NEUROSURG, V97, P211, DOI 10.3171/jns.2002.97.1.0211
   Javadpour M, 2003, BRIT J NEUROSURG, V17, P266, DOI 10.1080/0268869031000153215
   Kaku Y, 1999, NEUROSURGERY, V44, P338, DOI 10.1097/00006123-199902000-00052
   KERNOHAN GW, 1952, ATLAS TUMOR PATHOL, P18
   KIHLSTROM L, 1994, ACT NEUR S, V62, P55
   MAY PL, 1991, J NEUROSURG, V74, P867, DOI 10.3171/jns.1991.74.6.0867
   Mizoguchi M, 2000, MINIM INVAS NEUROSUR, V43, P53, DOI 10.1055/s-2000-8414
   Oka K, 1999, J NEUROSURG, V91, P964, DOI 10.3171/jns.1999.91.6.0964
   Packer R J, 1992, Neurosurg Clin N Am, V3, P863
   PENDL G, 1990, NEUROSURGERY, V26, P641, DOI 10.1227/00006123-199004000-00013
   POLLACK IF, 1994, J NEUROSURG, V80, P681, DOI 10.3171/jns.1994.80.4.0681
   SQUIRES LA, 1994, NEUROLOGY, V44, P953, DOI 10.1212/WNL.44.5.953
   VANDERTOP WP, 1992, NEUROSURGERY, V31, P186, DOI 10.1227/00006123-199208000-00003
   Wang CC, 2000, SURG NEUROL, V53, P41, DOI 10.1016/S0090-3019(99)00165-2
   Wellons JC, 2002, NEUROSURGERY, V51, P63, DOI 10.1097/00006123-200207000-00010
   Yeh DD, 2002, NEUROSURGERY, V50, P735, DOI 10.1097/00006123-200204000-00009
NR 22
TC 34
Z9 34
U1 0
U2 1
PU SPRINGER
PI NEW YORK
PA 233 SPRING ST, NEW YORK, NY 10013 USA
SN 0256-7040
EI 1433-0350
J9 CHILD NERV SYST
JI Childs Nerv. Syst.
PD NOV
PY 2004
VL 20
IS 11-12
BP 852
EP 857
DI 10.1007/s00381-004-0942-2
PG 6
WC Clinical Neurology; Pediatrics; Surgery
WE Science Citation Index Expanded (SCI-EXPANDED); Conference Proceedings Citation Index - Science (CPCI-S)
SC Neurosciences & Neurology; Pediatrics; Surgery
GA 866DF
UT WOS:000224753300014
PM 15243713
DA 2023-08-27
ER

PT J
AU Fei, XY
   Dong, Y
   An, HD
   Zhang, Q
   Zhang, YC
   Shi, J
AF Fei, Xiaoyan
   Dong, Yun
   An, Hedi
   Zhang, Qi
   Zhang, Yingchun
   Shi, Jun
TI Impact of region of interest size on transcranial sonography based
   computer-aided diagnosis for Parkinson's disease
SO MATHEMATICAL BIOSCIENCES AND ENGINEERING
LA English
DT Article
DE Parkinson's disease; transcranial sonography; region of interest;
   substantia nigra
ID BREAST-TUMOR; CLASSIFICATION
AB Transcranial sonography (TCS) has gained increasing application for diagnosis of Parkinson's disease (PD) in clinical practice in recent years, because most PD patients, even in the early stage of PD, have abnormal hyperechogenicity of the substantia nigra (SN) in brainstem shown in TCS images. Therefore, the region of interest (ROI) for feature extraction should cover the SN region in a computer-aided diagnosis (CAD) system. The ROI size naturally affects the feature representation. However, there currently exist no unified standard for determining the size of ROI. In this work, we quantitatively compare the performance of TCS-based CAD with three sizes of ROIs, namely the entire midbrain (EM) region, the half of midbrain (HoM) region and the SN region. The experimental results on the original extracted features and the features by dimensionality reduction show that ROI covering the EM region achieves the overall best diagnosis performance. The results indicates that the neighboring regions around SN might also have abnormal symptoms, which cannot be clearly observed with naked eyes. It suggests that the large ROI includes more information for feature representation to improve the diagnosis performance of TCS-based CAD for PD.
C1 [Fei, Xiaoyan; Zhang, Qi; Shi, Jun] Shanghai Univ, Key Lab Specialty Fiber Opt & Opt Access Networks, Shanghai Inst Adv Commun & Data Sci, Sch Commun & Informat Engn, Shanghai, Peoples R China.
   [Dong, Yun] Tongji Univ, Shanghai East Hosp, Dept Ultrasonog, Shanghai, Peoples R China.
   [An, Hedi] Tongji Univ, Shanghai East Hosp, Dept Neurol, Shanghai, Peoples R China.
   [Zhang, Yingchun] Soochow Univ, Affiliated Hosp 2, Dept Ultrasound, Suzhou, Peoples R China.
C3 Shanghai University; Tongji University; Tongji University; Soochow
   University - China
RP Shi, J (通讯作者)，Shanghai Univ, Key Lab Specialty Fiber Opt & Opt Access Networks, Shanghai Inst Adv Commun & Data Sci, Sch Commun & Informat Engn, Shanghai, Peoples R China.
EM
RI z, y/HPC-0477-2023; Zhang, Y J/HLG-1022-2023; zhang, ying/HJB-1230-2022;
   Zhang, Yida/T-2546-2018
OI Zhang, Yida/0000-0002-1505-6678
FU National Natural Science Foundation of China [61471231, 81830058,
   81627804]; Shanghai Science and Technology Foundation [18010500600,
   17411953400]; Pre-Research Project of The Second Affiliated Hospital of
   Soochow University [SDFEYGJ1709]
FX This work is supported by the National Natural Science Foundation of
   China (61471231, 81830058, 81627804), Shanghai Science and Technology
   Foundation (18010500600, 17411953400, 18010500600), and the Pre-Research
   Project of The Second Affiliated Hospital of Soochow University
   (SDFEYGJ1709).
CR Berg D, 2007, PARKINSONISM RELAT D, V13, pS429, DOI 10.1016/S1353-8020(08)70043-9
   Burke RE, 2013, EXP NEUROL, V246, P72, DOI 10.1016/j.expneurol.2012.01.011
   Chen L, 2012, LECT NOTES COMPUT SC, V7512, P272, DOI 10.1007/978-3-642-33454-2_34
   Ding Chris, 2005, Journal of Bioinformatics and Computational Biology, V3, P185, DOI 10.1142/S0219720005001004
   Frosini D, 2017, CURR OPIN NEUROL, V30, P416, DOI 10.1097/WCO.0000000000000463
   Gong BM, 2018, NEUROCOMPUTING, V320, P141, DOI 10.1016/j.neucom.2018.09.025
   Guo LH, 2018, CLIN HEMORHEOL MICRO, V69, P343, DOI 10.3233/CH-170275
   Huang QH, 2020, IEEE T KNOWL DATA EN, V32, P728, DOI 10.1109/TKDE.2019.2891622
   Huang QH, 2018, BIOMED RES INT-UK, V2018, DOI 10.1155/2018/5137904
   Ma YY, 2013, INT STAT REV, V81, P134, DOI 10.1111/j.1751-5823.2012.00182.x
   Moon JH, 2018, ACTA RADIOL, V59, P657, DOI 10.1177/0284185117732097
   Mwangi B, 2014, NEUROINFORMATICS, V12, P229, DOI 10.1007/s12021-013-9204-3
   Pauly O, 2012, LECT NOTES COMPUT SC, V7512, P443, DOI 10.1007/978-3-642-33454-2_55
   Plate A, 2012, ULTRASOUND MED BIOL, V38, P2041, DOI 10.1016/j.ultrasmedbio.2012.07.017
   Sakalauskas A., 2018, J ULTRASOUND MED
   Sakalauskas A, 2016, ULTRASOUND MED BIOL, V42, P322, DOI 10.1016/j.ultrasmedbio.2015.09.009
   Shi J, 2019, IEEE T BIO-MED ENG, V66, P2362, DOI 10.1109/TBME.2018.2889398
   Shi J, 2015, NEUROCOMPUTING, V168, P930, DOI 10.1016/j.neucom.2015.05.032
   Shi J, 2015, NEUROCOMPUTING, V149, P1415, DOI 10.1016/j.neucom.2014.08.054
   Skerl K, 2015, CLIN RADIOL, V70, P1421, DOI 10.1016/j.crad.2015.08.010
   Weingarten CP, 2015, NEUROSCI BIOBEHAV R, V59, P16, DOI 10.1016/j.neubiorev.2015.09.007
   Zhang Q, 2017, ULTRASOUND MED BIOL, V43, P1058, DOI 10.1016/j.ultrasmedbio.2016.12.016
   Zhou SC, 2013, BIOMED SIGNAL PROCES, V8, P688, DOI 10.1016/j.bspc.2013.06.011
NR 23
TC 1
Z9 3
U1 3
U2 14
PU AMER INST MATHEMATICAL SCIENCES-AIMS
PI SPRINGFIELD
PA PO BOX 2604, SPRINGFIELD, MO 65801-2604 USA
SN 1547-1063
EI 1551-0018
J9 MATH BIOSCI ENG
JI Math. Biosci. Eng.
PY 2019
VL 16
IS 5
BP 5640
EP 5651
DI 10.3934/mbe.2019280
PG 12
WC Mathematical & Computational Biology
WE Science Citation Index Expanded (SCI-EXPANDED)
SC Mathematical & Computational Biology
GA IF1YA
UT WOS:000472872700097
PM 31499729
OA gold
DA 2023-08-27
ER

PT J
AU Kramm, CM
   Wagner, S
   Van Gool, S
   Schmid, H
   Strater, R
   Gnekow, A
   Rutkowski, S
   Wolff, JEA
AF Kramm, Christof M.
   Wagner, Sabine
   Van Gool, Stefan
   Schmid, Hansjoerg
   Straeter, Ronald
   Gnekow, Astrid
   Rutkowski, Stefan
   Wolff, Johannes E. A.
TI Improved survival after gross total resection of malignant gliomas in
   pediatric patients from the HIT-GBM studies
SO ANTICANCER RESEARCH
LA English
DT Article
DE neurosurgery; tumor resection; glioblastoma; anaplastic astrocytoma;
   malignant glioma; survival
ID HIGH-GRADE GLIOMA; GLIOBLASTOMA-MULTIFORME; SURGICAL RESECTION;
   RANDOMIZED-TRIAL; CHILDREN; EXTENT; CHILDHOOD; PROGNOSIS; ASTROCYTOMAS;
   CHEMOTHERAPY
AB The present study was performed to investigate the prognostic impact of tumor resection on survival in children and adolescents with malignant gliomas. From the HIT-GBM data base of the Gesellschaft fur Paediatrische Onkologie und Haematologie (GPOH), 85 pediatric patients with malignant non-pontine gliomas were analyzed. Histological diagnosis and extent of tumor resection had been confirmed by central neuropathological review and post-surgical imaging. The extent of tumor resection represented the most prominent prognostic factor for overall (OS) and event-free survival (EFS) in univariate and Cox regression analyses. Four-year survival after gross total tumor resection was 48.0 +/- 12.0% (OS) and 14.1 +/- 8.9% (EFS), after non-total resection 13.2 +/- 6.1% and 2.9 +/- 2.8%, respectively. From several clinical parameters, only histological grading displayed a similar statistical significance in Cox regression analysis. In conclusion, gross total tumor resection improves survival in pediatric patients with malignant gliomas and should always be attempted when possible.
C1 Univ Texas, MD Anderson Canc Ctr, Dept Pediat, Sect Ped Neurooncol, Houston, TX 77030 USA.
   Univ Calgary, Alberta Childrens Prov Gen Hosp, Calgary, AB, Canada.
   Univ Wurzburg, Wurzburg, Germany.
   Klinikum Augsburg, Childrens Hosp, Augsburg, Germany.
   Univ Munster, Dept Pediat Hematol & Oncol, D-4400 Munster, Germany.
   Univ Hannover, Sch Med, Dept Pediat Hematol & Oncol, Hannover, Germany.
   Katholieke Univ Leuven, Dept Pediat Hematol & Oncol, Louvain, Belgium.
   Univ Regensburg, Dept Pediat Hematol & Oncol, St Hedwig Hosp, D-8400 Regensburg, Germany.
   Univ Dusseldorf, Childrens Hosp, Dept Pediat Oncol Hematol & Immunol, D-4000 Dusseldorf, Germany.
C3 University of Texas System; UTMD Anderson Cancer Center; Alberta
   Childrens Hospital; University of Calgary; University of Wurzburg;
   Klinikum Augsburg; University of Munster; Leibniz University Hannover;
   KU Leuven; University of Regensburg; Heinrich Heine University
   Dusseldorf
RP Wolff, JEA (通讯作者)，Univ Texas, MD Anderson Canc Ctr, Dept Pediat, Sect Ped Neurooncol, 1515 Holcombe Blvd,Unit 87, Houston, TX 77030 USA.
EM
OI Wolff, Johannes/0000-0003-0278-1309
CR Albright AL, 2000, NEUROSURGERY, V47, P879, DOI 10.1097/00006123-200010000-00018
   Benesch M, 2005, J NEURO-ONCOL, V72, P179, DOI 10.1007/s11060-004-3546-5
   Campbell JW, 1996, NEUROSURGERY, V38, P258, DOI 10.1097/00006123-199602000-00006
   DEVAUX BC, 1993, J NEUROSURG, V78, P767, DOI 10.3171/jns.1993.78.5.0767
   Finlay JL, 1999, CHILD NERV SYST, V15, P786, DOI 10.1007/s003810050471
   Heideman RL, 1997, CANCER-AM CANCER SOC, V80, P497
   Keles GE, 1999, SURG NEUROL, V52, P371, DOI 10.1016/S0090-3019(99)00103-2
   Kortmann RD, 2000, INT J RADIAT ONCOL, V46, P269, DOI 10.1016/S0360-3016(99)00369-7
   Kowalczuk A, 1997, NEUROSURGERY, V41, P1028, DOI 10.1097/00006123-199711000-00004
   KRETH FW, 1993, J NEUROSURG, V78, P762, DOI 10.3171/jns.1993.78.5.0762
   Lacroix M, 2001, J NEUROSURG, V95, P190, DOI 10.3171/jns.2001.95.2.0190
   Pollack IF, 2002, NEW ENGL J MED, V346, P420, DOI 10.1056/NEJMoa012224
   Rickert CH, 2001, AM J PATHOL, V158, P1525, DOI 10.1016/S0002-9440(10)64103-X
   SPOSTO R, 1989, J NEURO-ONCOL, V7, P165, DOI 10.1007/BF00165101
   Wagner S, 2004, CANCER, V100, P1750, DOI 10.1002/cncr.20168
   Wisoff JH, 1998, J NEUROSURG, V89, P52, DOI 10.3171/jns.1998.89.1.0052
   Wolff JEA, 2002, ANTICANCER RES, V22, P3569
   Wolff JEA, 2002, CANCER, V94, P264, DOI 10.1002/cncr.10114
   Wolff JEA, 2000, CANCER, V89, P2131, DOI 10.1002/1097-0142(20001115)89:10<2131::AID-CNCR14>3.0.CO;2-J
NR 19
TC 60
Z9 63
U1 0
U2 2
PU INT INST ANTICANCER RESEARCH
PI ATHENS
PA EDITORIAL OFFICE 1ST KM KAPANDRITIOU-KALAMOU RD KAPANDRITI, PO BOX 22,
   ATHENS 19014, GREECE
SN 0250-7005
J9 ANTICANCER RES
JI Anticancer Res.
PD SEP-OCT
PY 2006
VL 26
IS 5B
BP 3773
EP 3779
PG 7
WC Oncology
WE Science Citation Index Expanded (SCI-EXPANDED)
SC Oncology
GA 100ZA
UT WOS:000241707500025
PM 17094400
DA 2023-08-27
ER

PT J
AU Li, SL
   Wang, C
   Chen, JY
   Lan, YJ
   Zhang, WCB
   Kang, Z
   Zheng, Y
   Zhang, R
   Yu, JY
   Li, WB
AF Li, Shenglan
   Wang, Can
   Chen, Jinyi
   Lan, Yanjie
   Zhang, Weichunbai
   Kang, Zhuang
   Zheng, Yi
   Zhang, Rong
   Yu, Jianyu
   Li, Wenbin
TI Signaling pathways in brain tumors and therapeutic interventions
SO SIGNAL TRANSDUCTION AND TARGETED THERAPY
LA English
DT Review
ID CENTRAL-NERVOUS-SYSTEM; GERM-CELL TUMORS; PHASE-II TRIAL; TERT PROMOTER
   MUTATIONS; PRIMARY CNS LYMPHOMA; GLIOBLASTOMA SUBSET ANALYSIS;
   INTEGRATED GENOMIC ANALYSIS; INTRINSIC PONTINE GLIOMAS; HEPATOCYTE
   GROWTH-FACTOR; HIGH-GRADE MENINGIOMA
AB Brain tumors, although rare, contribute to distinct mortality and morbidity at all ages. Although there are few therapeutic options for brain tumors, enhanced biological understanding and unexampled innovations in targeted therapies and immunotherapies have considerably improved patients' prognoses. Nonetheless, the reduced response rates and unavoidable drug resistance of currently available treatment approaches have become a barrier to further improvement in brain tumor (glioma, meningioma, CNS germ cell tumors, and CNS lymphoma) treatment. Previous literature data revealed that several different signaling pathways are dysregulated in brain tumor. Importantly, a better understanding of targeting signaling pathways that influences malignant behavior of brain tumor cells might open the way for the development of novel targeted therapies. Thus, there is an urgent need for a more comprehensive understanding of the pathogenesis of these brain tumors, which might result in greater progress in therapeutic approaches. This paper began with a brief description of the epidemiology, incidence, risk factors, as well as survival of brain tumors. Next, the major signaling pathways underlying these brain tumors' pathogenesis and current progress in therapies, including clinical trials, targeted therapies, immunotherapies, and system therapies, have been systemically reviewed and discussed. Finally, future perspective and challenges of development of novel therapeutic strategies in brain tumor were emphasized.
C1 [Li, Shenglan; Wang, Can; Chen, Jinyi; Lan, Yanjie; Zhang, Weichunbai; Kang, Zhuang; Zheng, Yi; Zhang, Rong; Yu, Jianyu; Li, Wenbin] Capital Med Univ, Beijing Tiantan Hosp, Canc Ctr, Dept Neuro Oncol, Beijing, Peoples R China.
C3 Capital Medical University
RP Li, WB (通讯作者)，Capital Med Univ, Beijing Tiantan Hosp, Canc Ctr, Dept Neuro Oncol, Beijing, Peoples R China.
EM
FU National Natural Science Foundation of China; National Science and
   Technology Major Project of China; Clinical Major Specialty Projects of
   Beijing; Beijing Advanced Innovation Center for Big Data-based Precision
   Medicine;  [81972338];  [2016ZX09101017];  [2021YFF0901404]
FX AcknowledgementsThis study was sponsored by The National Natural Science
   Foundation of China, grant number 81972338. This work was also supported
   by the National Science and Technology Major Project of China
   (No.2016ZX09101017) and Clinical Major Specialty Projects of Beijing.
   This research was also supported by Beijing Advanced Innovation Center
   for Big Data-based Precision Medicine (No.2021YFF0901404).
CR Aas PA, 2003, NATURE, V421, P859, DOI 10.1038/nature01363
   Abedalthagafi M, 2016, NEURO-ONCOLOGY, V18, P649, DOI 10.1093/neuonc/nov316
   Abramson JS, 2017, NEW ENGL J MED, V377, P783, DOI 10.1056/NEJMc1704610
   Ahmed N, 2017, JAMA ONCOL, V3, P1094, DOI 10.1001/jamaoncol.2017.0184
   Akhavan D, 2010, NEURO-ONCOLOGY, V12, P882, DOI 10.1093/neuonc/noq052
   Al-Nedawi K, 2008, NAT CELL BIOL, V10, P619, DOI 10.1038/ncb1725
   Alapetite C, 2010, NEURO-ONCOLOGY, V12, P1318, DOI 10.1093/neuonc/noq093
   Aldape K, 2007, ARCH PATHOL LAB MED, V131, P242
   Alharbi M, 2020, JCO PRECIS ONCOL, V4, P796, DOI 10.1200/PO.20.00017
   Amaria RN, 2018, NAT MED, V24, P1649, DOI 10.1038/s41591-018-0197-1
   Amen AM, 2021, P NATL ACAD SCI USA, V118, DOI 10.1073/pnas.2008772118
   Anastas JN, 2019, CANCER CELL, V36, P528, DOI 10.1016/j.ccell.2019.09.005
   [Anonymous], 2020, CORRECTION LANCET ON, V21, P796
   [Anonymous], 2020, LANCET ONCOL, V21, pe462
   Asby DJ, 2018, CANCER MANAG RES, V10, P3483, DOI 10.2147/CMAR.S167095
   Bady P, 2018, ACTA NEUROPATHOL, V135, P601, DOI 10.1007/s00401-018-1810-6
   Bagley SJ, 2018, NEURO-ONCOLOGY, V20, P1429, DOI 10.1093/neuonc/noy032
   Bai M., 2022, BIOMED RES INT, V2022
   Banerjee A, 2017, NEURO-ONCOLOGY, V19, P1135, DOI 10.1093/neuonc/now282
   Bardella C, 2016, CANCER CELL, V30, P578, DOI 10.1016/j.ccell.2016.08.017
   Bartee E, 2017, EXP HEMATOL ONCOL, V6, DOI 10.1186/s40164-017-0075-4
   Batchelor T, 2006, J CLIN ONCOL, V24, P1281, DOI 10.1200/JCO.2005.04.8819
   Batchelor TT, 2016, HEMATOL-AM SOC HEMAT, P379, DOI 10.1182/asheducation-2016.1.379
   Baumgarten P, 2014, NEUROPATH APPL NEURO, V40, P205, DOI 10.1111/nan.12088
   Bhimani AD, 2020, PITUITARY, V23, P381, DOI 10.1007/s11102-020-01042-2
   Bi WL, 2022, NEURO-ONCOLOGY, V24, P101, DOI 10.1093/neuonc/noab118
   Biau J, 2018, NEUROCHIRURGIE, V64, P29, DOI 10.1016/j.neuchi.2014.10.112
   Birzu C, 2020, CURR OPIN ONCOL, V32, P613, DOI 10.1097/CCO.0000000000000687
   Bogdahn U, 2011, NEURO-ONCOLOGY, V13, P132, DOI 10.1093/neuonc/noq142
   Borrero LJH, 2021, BBA-REV CANCER, V1876, DOI 10.1016/j.bbcan.2021.188556
   Boussiotis VA, 2016, NEW ENGL J MED, V375, P1767, DOI 10.1056/NEJMra1514296
   Braganza MZ, 2012, NEURO-ONCOLOGY, V14, P1316, DOI 10.1093/neuonc/nos208
   Brandes AA, 2016, NEURO-ONCOLOGY, V18, P1146, DOI 10.1093/neuonc/now009
   Brastianos PK, 2013, NAT GENET, V45, P285, DOI 10.1038/ng.2526
   Brat DJ, 2020, ACTA NEUROPATHOL, V139, P603, DOI 10.1007/s00401-020-02127-9
   Brat DJ, 2018, ACTA NEUROPATHOL, V136, P805, DOI 10.1007/s00401-018-1913-0
   Brat DJ, 2015, NEW ENGL J MED, V372, P2481, DOI 10.1056/NEJMoa1402121
   Brennan C, 2009, PLOS ONE, V4, DOI 10.1371/journal.pone.0007752
   Brennan CW, 2013, CELL, V155, P462, DOI 10.1016/j.cell.2013.09.034
   Brigliadori G, 2016, J NEURO-ONCOL, V128, P333, DOI 10.1007/s11060-016-2116-y
   Bromberg JEC, 2019, LANCET ONCOL, V20, P216, DOI 10.1016/S1470-2045(18)30747-2
   Brown CE, 2016, NEW ENGL J MED, V375, P2561, DOI 10.1056/NEJMoa1610497
   Brown CE, 2015, CLIN CANCER RES, V21, P4062, DOI 10.1158/1078-0432.CCR-15-0428
   Brown NF, 2020, BMC CANCER, V20, DOI 10.1186/s12885-020-6624-y
   Butler M, 2020, TRENDS CANCER, V6, P380, DOI 10.1016/j.trecan.2020.02.010
   Byeon S., 2020, TARGET ONCOL, V15, P185
   Cairncross JG, 2014, J CLIN ONCOL, V32, P783, DOI 10.1200/JCO.2013.49.3726
   Calaminus G, 2004, KLIN PADIATR, V216, P141, DOI 10.1055/s-2004-822626
   Calaminus G, 2017, NEURO-ONCOLOGY, V19, P1661, DOI 10.1093/neuonc/nox122
   Calimeriy T, 2021, ESMO OPEN, V6, DOI 10.1016/j.esmoop.2021.100213
   Cao JY, 2020, AGING-US, V12, P1656, DOI 10.18632/aging.102706
   Carbonneau M, 2016, NAT COMMUN, V7, DOI 10.1038/ncomms12700
   Casamayor-Palleja M, 2001, BLOOD, V97, P3992, DOI 10.1182/blood.V97.12.3992
   Castel D, 2015, ACTA NEUROPATHOL, V130, P815, DOI 10.1007/s00401-015-1478-0
   Chamberlain MC, 2008, CANCER-AM CANCER SOC, V113, P2146, DOI 10.1002/cncr.23803
   Chamberlain MC, 2007, NEUROLOGY, V69, P969, DOI 10.1212/01.wnl.0000271382.62776.b7
   Chamberlain Marc C, 2013, CNS Oncol, V2, P227, DOI 10.2217/cns.13.17
   Chamberlain MC, 2012, J NEURO-ONCOL, V107, P315, DOI 10.1007/s11060-011-0741-z
   Chamberlain MC, 2004, NEUROLOGY, V62, P1210, DOI 10.1212/01.WNL.0000118300.82017.F4
   Chang AL, 2016, CANCER RES, V76, P5671, DOI 10.1158/0008-5472.CAN-16-0144
   Chapuy B, 2016, BLOOD, V127, P869, DOI 10.1182/blood-2015-10-673236
   Chen C, 2021, ACTA PHARMACOL SIN, V42, P108, DOI 10.1038/s41401-020-0418-2
   Chen R, 2015, NEUROSURG FOCUS, V38, DOI 10.3171/2015.1.FOCUS14745
   Chen XY, 2018, NAT COMMUN, V9, DOI 10.1038/s41467-018-05373-4
   Chen YW, 2012, CHILD NERV SYST, V28, P557, DOI 10.1007/s00381-012-1683-2
   Chen ZH, 2017, CANCER RES, V77, P2266, DOI 10.1158/0008-5472.CAN-16-2310
   Cheng FL, 2019, J EXP CLIN CANC RES, V38, DOI 10.1186/s13046-019-1269-x
   Cher L, 1996, NEUROLOGY, V46, P1757, DOI 10.1212/WNL.46.6.1757
   Chinot OL, 2014, NEW ENGL J MED, V370, P709, DOI 10.1056/NEJMoa1308345
   Chongsathidkiet P, 2018, NAT MED, V24, P1459, DOI 10.1038/s41591-018-0135-2
   Clark VE, 2013, SCIENCE, V339, P1077, DOI 10.1126/science.1233009
   Clarke VE, 2016, NAT GENET, V48, P1253, DOI 10.1038/ng.3651
   Cloughesy T, 2017, J CLIN ONCOL, V35, P343, DOI 10.1200/JCO.2015.64.7685
   Cloughesy TF, 2018, NEURO-ONCOLOGY, V20, P259, DOI 10.1093/neuonc/nox151
   Cordova C, 2020, CURR ONCOL REP, V22, DOI 10.1007/s11912-020-00937-4
   Creed JH, 2020, EUR J CANCER, V129, P123, DOI 10.1016/j.ejca.2020.01.012
   Dal Porto JM, 2004, MOL IMMUNOL, V41, P599, DOI 10.1016/j.molimm.2004.04.008
   Dasanu CA, 2019, EXPERT OPIN BIOL TH, V19, P99, DOI 10.1080/14712598.2019.1559292
   Davies H, 2002, NATURE, V417, P949, DOI 10.1038/nature00766
   Davis F, 2011, RADIAT RES, V175, P790, DOI 10.1667/RR2186.1
   Davis RE, 2010, NATURE, V463, P88, DOI 10.1038/nature08638
   DEAN M, 1985, NATURE, V318, P385, DOI 10.1038/318385a0
   DEPREZ RHL, 1994, AM J HUM GENET, V54, P1022
   DEPREZ RHL, 1995, J NEUROPATH EXP NEUR, V54, P224, DOI 10.1097/00005072-199503000-00009
   Desjardins A, 2018, NEW ENGL J MED, V379, P150, DOI 10.1056/NEJMoa1716435
   Di Stefano AL, 2015, CLIN CANCER RES, V21, P3307, DOI 10.1158/1078-0432.CCR-14-2199
   DiNardo CD, 2018, NEW ENGL J MED, V378, P2386, DOI 10.1056/NEJMoa1716984
   DiNardo CD, 2021, J CLIN ONCOL, V39, P57, DOI 10.1200/JCO.20.01632
   Dubinski D, 2016, NEURO-ONCOLOGY, V18, P807, DOI 10.1093/neuonc/nov280
   Dubrow R, 2010, CANCER EPIDEM BIOMAR, V19, P1709, DOI 10.1158/1055-9965.EPI-10-0225
   Duffau H, 2015, NEURO-ONCOLOGY, V17, P332, DOI 10.1093/neuonc/nou153
   Duncan CG, 2012, GENOME RES, V22, P2339, DOI 10.1101/gr.132738.111
   Ellison DW, 2020, BRAIN PATHOL, V30, P863, DOI 10.1111/bpa.12866
   Ellison DW, 2019, ACTA NEUROPATHOL, V137, P683, DOI 10.1007/s00401-019-01987-0
   Elsamadicy AA, 2017, EXPERT OPIN BIOL TH, V17, P507, DOI 10.1080/14712598.2017.1299705
   Erdem-Eraslan L, 2016, CANCER RES, V76, P525, DOI 10.1158/0008-5472.CAN-15-0776
   Ermoian RP, 2002, CLIN CANCER RES, V8, P1100
   Esteller M, 1999, CANCER RES, V59, P793
   Estrada CC, 2019, J AM SOC NEPHROL, V30, P187, DOI 10.1681/ASN.2018080853
   Falnes PO, 2002, NATURE, V419, P178, DOI 10.1038/nature01048
   Fan QW, 2013, CANCER CELL, V24, P438, DOI 10.1016/j.ccr.2013.09.004
   Fan X, 2010, STEM CELLS, V28, P5, DOI 10.1002/stem.254
   Farrera-Sal M, 2021, CLIN CANCER RES, V27, P2979, DOI 10.1158/1078-0432.CCR-20-1565
   Fecci PE, 2006, CANCER RES, V66, P3294, DOI 10.1158/0008-5472.CAN-05-3773
   Ferguson SD, 2018, J NEUROPATH EXP NEUR, V77, P437, DOI 10.1093/jnen/nly022
   Ferguson SD, 2015, J NEURO-ONCOL, V123, P385, DOI 10.1007/s11060-015-1731-3
   Ferrara N, 2005, BIOCHEM BIOPH RES CO, V333, P328, DOI 10.1016/j.bbrc.2005.05.132
   Ferreri AJM, 2016, LANCET HAEMATOL, V3, pE217, DOI 10.1016/S2352-3026(16)00036-3
   Filbin MG, 2013, NAT MED, V19, P1518, DOI 10.1038/nm.3328
   Fisher JL, 2007, NEUROL CLIN, V25, P867, DOI 10.1016/j.ncl.2007.07.002
   Frappaz D, 2022, NEURO-ONCOLOGY, V24, P516, DOI 10.1093/neuonc/noab252
   Frenel J. S., 370MO FOLAGLI PHASE, P31
   Friedman GK, 2021, NEW ENGL J MED, V384, P1613, DOI 10.1056/NEJMoa2024947
   Frigault MJ, 2019, BLOOD, V134, P860, DOI 10.1182/blood.2019001694
   Fu XD, 2015, CELL METAB, V22, P508, DOI 10.1016/j.cmet.2015.06.009
   Fueyo J, 2000, ONCOGENE, V19, P2, DOI 10.1038/sj.onc.1203251
   Fukushima S, 2014, ACTA NEUROPATHOL, V127, P911, DOI 10.1007/s00401-014-1247-5
   Furnari FB, 2015, NAT REV CANCER, V15, P302, DOI 10.1038/nrc3918
   Gagne LM, 2017, TRENDS CELL BIOL, V27, P738, DOI 10.1016/j.tcb.2017.06.002
   Gan HK, 2012, CANCER RES, V72, P2924, DOI 10.1158/0008-5472.CAN-11-3898
   Gao ML, 2021, CANCER RES, V81, P3580, DOI 10.1158/0008-5472.CAN-20-2773
   Gelerstein E, 2017, J CLIN NEUROSCI, V37, P51, DOI 10.1016/j.jocn.2016.11.011
   Ghesquieres H, 2019, ANN ONCOL, V30, P621, DOI 10.1093/annonc/mdz032
   Gilbert CA, 2010, CANCER RES, V70, P6870, DOI 10.1158/0008-5472.CAN-10-1378
   Gilbert MR, 2017, J NEURO-ONCOL, V131, P193, DOI 10.1007/s11060-016-2288-5
   Gilbert MR, 2014, NEW ENGL J MED, V370, P699, DOI 10.1056/NEJMoa1308573
   Giles AJ, 2018, J IMMUNOTHER CANCER, V6, DOI 10.1186/s40425-018-0371-5
   Gittleman H, 2019, J NEURO-ONCOL, V143, P251, DOI 10.1007/s11060-019-03173-4
   Gluck WL, 2020, INVEST NEW DRUG, V38, P831, DOI 10.1007/s10637-019-00840-1
   Goff SL, 2019, J IMMUNOTHER, V42, P126, DOI 10.1097/CJI.0000000000000260
   Gohar Maha K, 2018, Egypt J Immunol, V25, P19
   Goldbrunner R, 2021, NEURO-ONCOLOGY, V23, P1821, DOI 10.1093/neuonc/noab150
   Goldbrunner R, 2016, LANCET ONCOL, V17, pE383, DOI 10.1016/S1470-2045(16)30321-7
   Gonzalez-Aguilar A, 2012, CLIN CANCER RES, V18, P5203, DOI 10.1158/1078-0432.CCR-12-0845
   Gousias K, 2016, J NEUROSURG, V125, P551, DOI 10.3171/2015.9.JNS15754
   Goutagny S, 2014, BRAIN PATHOL, V24, P184, DOI 10.1111/bpa.12110
   Graillon T, 2020, CLIN CANCER RES, V26, P552, DOI 10.1158/1078-0432.CCR-19-2109
   Grasso CS, 2015, NAT MED, V21, P555, DOI 10.1038/nm.3855
   Gritsch S, 2022, CANCER-AM CANCER SOC, V128, P47, DOI 10.1002/cncr.33918
   Grommes C, 2019, BLOOD, V133, P436, DOI 10.1182/blood-2018-09-875732
   Grommes C, 2019, NEURO-ONCOLOGY, V21, P296, DOI 10.1093/neuonc/noy192
   Grommes C, 2017, CANCER DISCOV, V7, P1018, DOI 10.1158/2159-8290.CD-17-0613
   Grunberg SM, 2006, CANCER INVEST, V24, P727, DOI 10.1080/07357900601062339
   Gupta S, 2018, NEUROSURG FOCUS, V44, DOI 10.3171/2018.1.FOCUS17754
   Gurney JG, 1996, EPIDEMIOLOGY, V7, P485, DOI 10.1097/00001648-199609000-00005
   Gutmann DH, 2019, NEURON, V104, P442, DOI 10.1016/j.neuron.2019.08.028
   Haase D, 2010, NEURO-ONCOLOGY, V12, P844, DOI 10.1093/neuonc/noq004
   Haase S, 2018, EXPERT OPIN THER TAR, V22, P599, DOI 10.1080/14728222.2018.1487953
   Hai L, 2018, CELL DEATH DIS, V9, DOI 10.1038/s41419-017-0119-z
   Hambardzumyan D, 2016, NAT NEUROSCI, V19, P20, DOI 10.1038/nn.4185
   Hara T, 2021, CANCER CELL, V39, P779, DOI 10.1016/j.ccell.2021.05.002
   Hargrave DR, 2019, CLIN CANCER RES, V25, P7303, DOI 10.1158/1078-0432.CCR-19-2177
   Harter PN, 2017, CHIN CLIN ONCOL, V6, DOI 10.21037/cco.2017.05.02
   Harutyunyan AS, 2019, NAT COMMUN, V10, DOI 10.1038/s41467-019-09140-x
   Hegi ME, 2005, NEW ENGL J MED, V352, P997, DOI 10.1056/NEJMoa043331
   Hegi ME, 2008, J CLIN ONCOL, V26, P4189, DOI 10.1200/JCO.2007.11.5964
   Hellmann MD, 2019, NEW ENGL J MED, V381, P2020, DOI 10.1056/NEJMoa1910231
   Heynckes S, 2019, BMC CANCER, V19, DOI 10.1186/s12885-019-5308-y
   Heynckes S, 2017, ONCOTARGET, V8, P74170, DOI 10.18632/oncotarget.18819
   Hodges TR, 2017, NEURO-ONCOLOGY, V19, P1047, DOI 10.1093/neuonc/nox026
   Hoeman C, 2018, FRONT ONCOL, V8, DOI 10.3389/fonc.2018.00191
   Holick CN, 2007, AM J CLIN NUTR, V85, P877, DOI 10.1093/ajcn/85.3.877
   Horn S, 2013, SCIENCE, V339, P959, DOI 10.1126/science.1230062
   Hu XM, 2022, FRONT ONCOL, V12, DOI 10.3389/fonc.2022.912166
   Hu YW, 2012, INT J RADIAT ONCOL, V84, P639, DOI 10.1016/j.ijrobp.2011.12.052
   Huang PH, 2009, SCI SIGNAL, V2, DOI 10.1126/scisignal.287re6
   Hui EF, 2017, SCIENCE, V355, P1428, DOI 10.1126/science.aaf1292
   Huntoon K, 2020, FRONT ONCOL, V10, DOI 10.3389/fonc.2020.579599
   Ichimura K, 2016, ACTA NEUROPATHOL, V131, P889, DOI 10.1007/s00401-016-1557-x
   Iorgulescu JB, 2021, CLIN CANCER RES, V27, P276, DOI 10.1158/1078-0432.CCR-20-2291
   Jakola AS, 2017, ANN ONCOL, V28, P1942, DOI 10.1093/annonc/mdx230
   James MF, 2009, MOL CELL BIOL, V29, P4250, DOI 10.1128/MCB.01581-08
   Ji YL, 2015, J CLIN ONCOL, V33, P4093, DOI 10.1200/JCO.2015.61.6490
   Johnson LA, 2015, SCI TRANSL MED, V7, DOI 10.1126/scitranslmed.aaa4963
   Juratli TA, 2018, ACTA NEUROPATHOL, V136, P779, DOI 10.1007/s00401-018-1899-7
   Kaley T, 2018, J CLIN ONCOL, V36, P3477, DOI 10.1200/JCO.2018.78.9990
   Kaley T, 2014, NEURO-ONCOLOGY, V16, P829, DOI 10.1093/neuonc/not330
   Kaley TJ, 2020, ANN CLIN TRANSL NEUR, V7, P429, DOI 10.1002/acn3.51009
   Kaley TJ, 2015, NEURO-ONCOLOGY, V17, P116, DOI 10.1093/neuonc/nou148
   Katz LM, 2018, ACTA NEUROPATHOL, V135, P955, DOI 10.1007/s00401-018-1844-9
   Kellie SJ, 2004, PEDIATR BLOOD CANCER, V43, P126, DOI 10.1002/pbc.20026
   Killela PJ, 2014, ONCOTARGET, V5, P1515, DOI 10.18632/oncotarget.1765
   Kim KH, 2013, NEURO-ONCOLOGY, V15, P161, DOI 10.1093/neuonc/nos299
   Kita D, 2007, ACTA NEUROPATHOL, V113, P295, DOI 10.1007/s00401-006-0186-1
   Klekamp J, 1999, SURG NEUROL, V52, P552, DOI 10.1016/S0090-3019(99)00153-6
   Koelsche C, 2013, ACTA NEUROPATHOL, V126, P907, DOI 10.1007/s00401-013-1195-5
   Korshunov A, 2018, BRAIN PATHOL, V28, P656, DOI 10.1111/bpa.12566
   Koschmann C, 2016, ONCOTARGET, V7, P65696, DOI 10.18632/oncotarget.11602
   KRAUTH J, 1990, EUR ARCH PSY CLIN N, V239, P391, DOI 10.1007/BF01734548
   Kreth S, 2011, PLOS ONE, V6, DOI 10.1371/journal.pone.0017156
   Kros J, 2001, J PATHOL, V194, P367, DOI 10.1002/path.909
   Kuan AS, 2019, NEURO-ONCOLOGY, V21, P944, DOI 10.1093/neuonc/noz013
   KUPPNER MC, 1988, INT J CANCER, V42, P562, DOI 10.1002/ijc.2910420416
   Kurucu N, 2020, J CANCER RES THER, V16, P1459, DOI 10.4103/jcrt.JCRT_314_18
   Lacourt A, 2013, BMC PUBLIC HEALTH, V13, DOI 10.1186/1471-2458-13-340
   Lamouille S, 2014, NAT REV MOL CELL BIO, V15, P178, DOI 10.1038/nrm3758
   Lang FF, 2018, J CLIN ONCOL, V36, P1419, DOI 10.1200/JCO.2017.75.8219
   Lapin DH, 2017, FRONT ONCOL, V7, DOI 10.3389/fonc.2017.00057
   Lapointe S, 2018, LANCET, V392, P432, DOI 10.1016/S0140-6736(18)30990-5
   Larson JD, 2019, CANCER CELL, V35, P140, DOI 10.1016/j.ccell.2018.11.015
   Lassman AB, 2019, NEURO-ONCOLOGY, V21, P106, DOI 10.1093/neuonc/noy091
   Leavitt E, 2019, CURR TREAT OPTION ON, V20, DOI 10.1007/s11864-019-0683-9
   Levine AJ, 1997, CELL, V88, P323, DOI 10.1016/S0092-8674(00)81871-1
   Li J, 2011, J SURG ONCOL, V103, P813, DOI 10.1002/jso.21851
   Li Xiang-Rong, 2016, J Registry Manag, V43, P187
   Li YH, 2015, NAT CELL BIOL, V17, P1327, DOI 10.1038/ncb3240
   LIBERMANN TA, 1984, CANCER RES, V44, P753
   LIBERMANN TA, 1985, NATURE, V313, P144, DOI 10.1038/313144a0
   Limam S, 2020, J NEUROVIROL, V26, P347, DOI 10.1007/s13365-020-00833-4
   Lin CJ, 2016, TOXICOL APPL PHARM, V304, P59, DOI 10.1016/j.taap.2016.05.018
   Lin K, 2022, SCIENCE, V377, P502, DOI 10.1126/science.abn7570
   Liu HL, 2021, CELL REP, V36, DOI 10.1016/j.celrep.2021.109718
   Liu SL, 2018, INVEST NEW DRUG, V36, P961, DOI 10.1007/s10637-018-0575-z
   Liu XJ, 2019, J EXP CLIN CANC RES, V38, DOI 10.1186/s13046-019-1235-7
   Lorzadeh S, 2021, BBA-MOL CELL RES, V1868, DOI 10.1016/j.bbamcr.2020.118926
   Lou E, 2012, J NEURO-ONCOL, V109, P63, DOI 10.1007/s11060-012-0861-0
   Louis DN, 2021, NEURO-ONCOLOGY, V23, P1231, DOI 10.1093/neuonc/noab106
   Louis DN, 2020, BRAIN PATHOL, V30, P844, DOI 10.1111/bpa.12832
   Louis DN, 2019, BRAIN PATHOL, V29, P469, DOI 10.1111/bpa.12732
   Louis DN, 2018, ACTA NEUROPATHOL, V135, P639, DOI 10.1007/s00401-018-1826-y
   Louis DN, 2018, ACTA NEUROPATHOL, V135, P481, DOI 10.1007/s00401-018-1808-0
   Louis DN, 2017, BRAIN PATHOL, V27, P851, DOI 10.1111/bpa.12457
   Louis DN, 2017, ACTA NEUROPATHOL, V133, P1, DOI 10.1007/s00401-016-1646-x
   Louis DN, 2016, ACTA NEUROPATHOL, V131, P803, DOI 10.1007/s00401-016-1545-1
   Lu C, 2012, NATURE, V483, P474, DOI 10.1038/nature10860
   Magnes T, 2021, ESMO OPEN, V6, DOI 10.1016/j.esmoop.2020.100012
   Malumbres M, 2005, TRENDS BIOCHEM SCI, V30, P630, DOI 10.1016/j.tibs.2005.09.005
   Mantovani A, 2004, TRENDS IMMUNOL, V25, P677, DOI 10.1016/j.it.2004.09.015
   Mantovani A, 2002, TRENDS IMMUNOL, V23, P549, DOI 10.1016/S1471-4906(02)02302-5
   Sepulveda-Sanchez JM, 2017, NEURO-ONCOLOGY, V19, P1522, DOI 10.1093/neuonc/nox105
   Marenco-Hillembrand L, 2020, J NEURO-ONCOL, V147, P297, DOI 10.1007/s11060-020-03451-6
   Marin BM, 2021, NEURO-ONCOLOGY, V23, P2042, DOI 10.1093/neuonc/noab133
   Martikainen M, 2019, CANCERS, V11, DOI 10.3390/cancers11020186
   Martin V., 2018, ACTR 39
   Massague J, 2008, CELL, V134, P215, DOI 10.1016/j.cell.2008.07.001
   Mathews JD, 2013, BMJ-BRIT MED J, V346, DOI 10.1136/bmj.f2360
   Maude SL, 2018, NEW ENGL J MED, V378, P439, DOI 10.1056/NEJMoa1709866
   Mawrin C, 2010, J NEURO-ONCOL, V99, P379, DOI 10.1007/s11060-010-0342-2
   McCann KJ, 2009, BLOOD, V113, P4677, DOI 10.1182/blood-2008-09-179366
   Mehta S, 2018, CELL MOL LIFE SCI, V75, P385, DOI 10.1007/s00018-017-2608-8
   Mellinghoff IK, 2020, J CLIN ONCOL, V38, DOI 10.1200/JCO.19.03327
   MERRIAM B, 1989, PRACTITIONER, V233, P649
   Miles X, 2021, FRONT ONCOL, V11, DOI 10.3389/fonc.2021.703442
   Miller KD, 2021, CA-CANCER J CLIN, V71, P381, DOI 10.3322/caac.21693
   Miller TW, 2019, J NEURO-ONCOL, V144, P563, DOI 10.1007/s11060-019-03258-0
   Mirian C, 2020, J NEUROL NEUROSUR PS, V91, P378, DOI 10.1136/jnnp-2019-322257
   Miska J, 2019, CELL REP, V27, P226, DOI 10.1016/j.celrep.2019.03.029
   Mizowaki T, 2015, J NEURO-ONCOL, V124, P165, DOI 10.1007/s11060-015-1843-9
   Montesinos-Rongen M, 2011, ACTA NEUROPATHOL, V122, P791, DOI 10.1007/s00401-011-0891-2
   Mottini C, 2021, SEMIN CANCER BIOL, V68, P59, DOI 10.1016/j.semcancer.2019.09.023
   Na K, 2019, J NEURO-ONCOL, V142, P445, DOI 10.1007/s11060-019-03114-1
   Nagaishi M, 2014, BRAIN TUMOR PATHOL, V31, P257, DOI 10.1007/s10014-013-0168-7
   Nair S, 2021, JCI INSIGHT, V6, DOI 10.1172/jci.insight.144619
   Nair SK, 2014, CLIN CANCER RES, V20, P2684, DOI 10.1158/1078-0432.CCR-13-3268
   Narita Y, 2021, NEURO-ONCOLOGY, V23, P122, DOI 10.1093/neuonc/noaa145
   Nassiri F, 2021, NEURO-ONCOLOGY, V23, P1282, DOI 10.1093/neuonc/noab036
   Nassiri F, 2019, NEURO-ONCOLOGY, V21, P901, DOI 10.1093/neuonc/noz061
   Nayak L, 2017, BLOOD, V129, P3071, DOI 10.1182/blood-2017-01-764209
   Nayak L, 2012, J NEURO-ONCOL, V109, P187, DOI 10.1007/s11060-012-0886-4
   Nduom EK, 2016, NEURO-ONCOLOGY, V18, P195, DOI 10.1093/neuonc/nov172
   Neglia JP, 2006, J NATL CANCER I, V98, P1528, DOI 10.1093/jnci/djj411
   Neyns B, 2009, ANN ONCOL, V20, P1596, DOI 10.1093/annonc/mdp032
   Ngo VN, 2011, NATURE, V470, P115, DOI 10.1038/nature09671
   Nie E, 2021, NEURO-ONCOLOGY, V23, P435, DOI 10.1093/neuonc/noaa198
   Norden AD, 2015, NEUROLOGY, V84, P280, DOI 10.1212/WNL.0000000000001153
   Norden AD, 2010, J NEURO-ONCOL, V96, P211, DOI 10.1007/s11060-009-9948-7
   Nowak-Choi K, 2021, J NEURO-ONCOL, V152, P145, DOI 10.1007/s11060-020-03688-1
   O'Rourke DM, 2017, SCI TRANSL MED, V9, DOI 10.1126/scitranslmed.aaa0984
   Ohashi R, 2014, ONCOL REP, V32, P1163, DOI 10.3892/or.2014.3283
   Okamura R, 2018, JCO PRECIS ONCOL, V2, DOI 10.1200/PO.18.00183
   Olar A, 2017, ACTA NEUROPATHOL, V133, P431, DOI 10.1007/s00401-017-1678-x
   Oldrini B, 2020, NAT COMMUN, V11, DOI 10.1038/s41467-020-17717-0
   Omuro A, 2018, NEURO-ONCOLOGY, V20, P674, DOI 10.1093/neuonc/nox208
   Omuro A, 2015, BLOOD, V125, P1403, DOI 10.1182/blood-2014-10-604561
   Ostrom Q. T., 2020, NEUROONCOL S1
   Ostrom QT, 2021, CURR NEUROL NEUROSCI, V21, DOI 10.1007/s11910-021-01152-9
   Ostrom QT, 2019, NEURO-ONCOLOGY, V21, P1357, DOI 10.1093/neuonc/noz123
   Ostrom QT, 2015, NEURO-ONCOLOGY, V17, P1, DOI [10.1093/neuonc/nov189, 10.1093/neuonc/now207]
   Otvos B, 2016, STEM CELLS, V34, P2026, DOI 10.1002/stem.2393
   Paldor I, 2016, J CLIN NEUROSCI, V31, P37, DOI 10.1016/j.jocn.2016.03.014
   Paramasivam N, 2019, ACTA NEUROPATHOL, V138, P295, DOI 10.1007/s00401-019-02008-w
   Park J, 2015, RADIAT ONCOL, V10, DOI 10.1186/s13014-015-0441-5
   Parsons DW, 2008, SCIENCE, V321, P1807, DOI 10.1126/science.1164382
   Pathania M, 2017, CANCER CELL, V32, P684, DOI 10.1016/j.ccell.2017.09.014
   Pearce MS, 2012, LANCET, V380, P499, DOI 10.1016/S0140-6736(12)60815-0
   Peereboom DM, 2019, JCI INSIGHT, V4, DOI 10.1172/jci.insight.130748
   Petrilli AM, 2016, ONCOGENE, V35, P537, DOI 10.1038/onc.2015.125
   Pierscianek D, 2013, BRAIN PATHOL, V23, P13, DOI 10.1111/j.1750-3639.2012.00609.x
   Piperi C, 2019, CELLS-BASEL, V8, DOI 10.3390/cells8111398
   Platten M, 2021, NATURE, V592, P463, DOI 10.1038/s41586-021-03363-z
   Ponzoni M, 2014, ANN ONCOL, V25, P316, DOI 10.1093/annonc/mdt385
   Prager BC, 2020, CANCER DISCOV, V10, P1722, DOI 10.1158/2159-8290.CD-20-0160
   Pratt D, 2022, ACTA NEUROPATHOL, V143, P403, DOI 10.1007/s00401-022-02404-9
   Preusser M, 2022, NEURO-ONCOLOGY, V24, P755, DOI 10.1093/neuonc/noab243
   Preusser M, 2018, NAT REV NEUROL, V14, P106, DOI 10.1038/nrneurol.2017.168
   Przybylowski CJ, 2021, J NEUROSURG, V135, P515, DOI 10.3171/2020.6.JNS20374
   Ragel BT, 2010, J NEURO-ONCOL, V99, P315, DOI 10.1007/s11060-010-0381-8
   Rahman MA, 2019, BRIT J CANCER, V121, P545, DOI 10.1038/s41416-019-0551-1
   Raychaudhuri B, 2011, NEURO-ONCOLOGY, V13, P591, DOI 10.1093/neuonc/nor042
   Reap EA, 2018, CANCER RES, V78, P256, DOI 10.1158/0008-5472.CAN-17-0469
   Reardon DA, 2020, JAMA ONCOL, V6, P1003, DOI 10.1001/jamaoncol.2020.1024
   Reardon DA, 2020, CLIN CANCER RES, V26, P1586, DOI 10.1158/1078-0432.CCR-18-1140
   Reuss DE, 2015, ACTA NEUROPATHOL, V129, P133, DOI 10.1007/s00401-014-1370-3
   Reuss DE, 2013, ACTA NEUROPATHOL, V125, P351, DOI 10.1007/s00401-013-1093-x
   Riemenschneider MJ, 2006, LANCET NEUROL, V5, P1045, DOI 10.1016/S1474-4422(06)70625-1
   Rizvi NA, 2015, LANCET ONCOL, V16, P257, DOI 10.1016/S1470-2045(15)70054-9
   Robert C, 2019, NEW ENGL J MED, V381, P626, DOI 10.1056/NEJMoa1904059
   Roboz GJ, 2020, BLOOD, V135, P463, DOI 10.1182/blood.2019002140
   Rodriguez PC, 2008, IMMUNOL REV, V222, P180, DOI 10.1111/j.1600-065X.2008.00608.x
   Rollison DEM, 2005, INT J CANCER, V113, P769, DOI 10.1002/ijc.20641
   Ronellenfitsch MW, 2018, ACTA NEUROPATHOL COM, V6, DOI 10.1186/s40478-018-0583-4
   Rubenstein JL, 2018, BLOOD ADV, V2, P1595, DOI 10.1182/bloodadvances.2017014845
   Rubenstein JL, 2013, BLOOD, V121, P745, DOI 10.1182/blood-2012-07-440974
   Rubenstein JL, 2006, BLOOD, V107, P3716, DOI 10.1182/blood-2005-03-0897
   Ruda R, 2019, J NEURO-ONCOL, V145, P115, DOI 10.1007/s11060-019-03277-x
   Ruder AM, 2012, AM J IND MED, V55, P747, DOI 10.1002/ajim.22085
   RUTTLEDGE MH, 1994, NAT GENET, V6, P180, DOI 10.1038/ng0294-180
   Sadetzki S, 2005, RADIAT RES, V163, P424, DOI 10.1667/RR3329
   Sahm F, 2017, LANCET ONCOL, V18, P682, DOI 10.1016/S1470-2045(17)30155-9
   Sahm F, 2016, JNCI-J NATL CANCER I, V108, DOI 10.1093/jnci/djv377
   Sampson JH, 2010, J CLIN ONCOL, V28, P4722, DOI 10.1200/JCO.2010.28.6963
   Sandmann T, 2015, J CLIN ONCOL, V33, P2735, DOI 10.1200/JCO.2015.61.5005
   Santoni G, 2022, INT J MOL SCI, V23, DOI 10.3390/ijms23020688
   Scerrati A, 2020, EXPERT REV ANTICANC, V20, P197, DOI 10.1080/14737140.2020.1736567
   Schindler G, 2011, ACTA NEUROPATHOL, V121, P397, DOI 10.1007/s00401-011-0802-6
   Schreck KC, 2019, CANCERS, V11, DOI 10.3390/cancers11091262
   Schulte SL, 2016, ONCOTARGET, V7, P55026, DOI 10.18632/oncotarget.10392
   Schuster J, 2015, NEURO-ONCOLOGY, V17, P854, DOI 10.1093/neuonc/nou348
   Schuster SJ, 2019, NEW ENGL J MED, V380, P45, DOI 10.1056/NEJMoa1804980
   Schwartzentruber J, 2012, NATURE, V482, P226, DOI [10.1038/nature10833, 10.1007/s13311-012-0137-6]
   Sellers WR, 1997, J CLIN ONCOL, V15, P3301, DOI 10.1200/JCO.1997.15.11.3301
   Shankar GM, 2017, NEURO-ONCOLOGY, V19, P1447, DOI 10.1093/neuonc/nox094
   Shankar GM, 2017, NEURO-ONCOLOGY, V19, P535, DOI 10.1093/neuonc/now235
   Shapiro IM, 2014, SCI TRANSL MED, V6, DOI 10.1126/scitranslmed.3008639
   Sharma M, 2019, J NEURO-ONCOL, V144, P359, DOI 10.1007/s11060-019-03236-6
   Shepherd DJ, 2021, ONCOLOGIST, V26, P919, DOI 10.1002/onco.13835
   Shih KC, 2016, J NEURO-ONCOL, V129, P281, DOI 10.1007/s11060-016-2172-3
   Sievers P, 2020, ACTA NEUROPATHOL, V140, P409, DOI 10.1007/s00401-020-02188-w
   Silveira AB, 2019, ACTA NEUROPATHOL, V137, P637, DOI 10.1007/s00401-019-01975-4
   Simo M, 2014, CANCER CHEMOTH PHARM, V73, P919, DOI 10.1007/s00280-014-2422-z
   SIMPSON D, 1957, J NEUROL NEUROSUR PS, V20, P22, DOI 10.1136/jnnp.20.1.22
   Slot KM, 2018, WORLD NEUROSURG, V111, pE856, DOI 10.1016/j.wneu.2017.12.178
   Solomon JP, 2020, MODERN PATHOL, V33, DOI 10.1038/s41379-019-0324-7
   Song LR, 2020, WORLD NEUROSURG, V136, pE355, DOI 10.1016/j.wneu.2019.12.182
   Sorensen AG, 2012, CANCER RES, V72, P402, DOI 10.1158/0008-5472.CAN-11-2464
   Stock S, 2019, INT J MOL SCI, V20, DOI 10.3390/ijms20246223
   Strand S, 1996, NAT MED, V2, P1361, DOI 10.1038/nm1296-1361
   Stupp R, 2005, NEW ENGL J MED, V352, P987, DOI 10.1056/NEJMoa043330
   Stupp R, 2017, JAMA-J AM MED ASSOC, V318, P2306, DOI 10.1001/jama.2017.18718
   Su R, 2018, CELL, V172, P90, DOI 10.1016/j.cell.2017.11.031
   Sun C, 2018, IMMUNITY, V48, P434, DOI 10.1016/j.immuni.2018.03.014
   Szabo E, 2016, NEURO-ONCOLOGY, V18, P1242, DOI 10.1093/neuonc/now043
   Tabernero J, 2015, J CLIN ONCOL, V33, P3401, DOI 10.1200/JCO.2014.60.7341
   Takahashi M, 2019, CANCER SCI, V110, P2247, DOI 10.1111/cas.14067
   Takami H, 2022, CURR ONCOL REP, V24, P105, DOI 10.1007/s11912-022-01195-2
   Tanaka K, 2011, CANCER DISCOV, V1, P524, DOI 10.1158/2159-8290.CD-11-0124
   Taylor AJ, 2010, J CLIN ONCOL, V28, P5287, DOI 10.1200/JCO.2009.27.0090
   Terry MB, 2009, ANN EPIDEMIOL, V19, P161, DOI 10.1016/j.annepidem.2008.12.010
   Todo T, 2022, NAT COMMUN, V13, DOI 10.1038/s41467-022-31262-y
   Todo T, 2022, NAT MED, V28, P1630, DOI 10.1038/s41591-022-01897-x
   Torre M, 2020, ACTA NEUROPATHOL COM, V8, DOI 10.1186/s40478-020-00980-z
   Touat M, 2020, NATURE, V580, P517, DOI 10.1038/s41586-020-2209-9
   TROFATTER JA, 1993, CELL, V72, P791, DOI 10.1016/0092-8674(93)90406-G
   Tsuda K, 2014, NEUROL MED-CHIR, V54, P907, DOI 10.2176/nmc.oa.2013-0311
   Turajlic S, 2017, LANCET ONCOL, V18, P1009, DOI 10.1016/S1470-2045(17)30516-8
   Turcan S, 2012, NATURE, V483, P479, DOI 10.1038/nature10866
   Turner MC, 2013, CANCER CAUSE CONTROL, V24, P949, DOI 10.1007/s10552-013-0171-7
   Turtle CJ, 2016, SCI TRANSL MED, V8, DOI 10.1126/scitranslmed.aaf8621
   Udaka YT, 2018, NEUROL CLIN, V36, P533, DOI 10.1016/j.ncl.2018.04.009
   Ullrich NJ, 2020, NEURO-ONCOLOGY, V22, P1527, DOI 10.1093/neuonc/noaa071
   Umemura N, 2008, J LEUKOCYTE BIOL, V83, P1136, DOI 10.1189/jlb.0907611
   van den Bent M, 2020, J NEURO-ONCOL, V146, P79, DOI 10.1007/s11060-019-03337-2
   Vaubel RA, 2016, J NEUROPATH EXP NEUR, V75, P44, DOI 10.1093/jnen/nlv006
   Venneti S, 2013, J NEUROPATH EXP NEUR, V72, P298, DOI 10.1097/NEN.0b013e3182898113
   Verhaak RGW, 2010, CANCER CELL, V17, P98, DOI 10.1016/j.ccr.2009.12.020
   Villano JL, 2011, BRIT J CANCER, V105, P1414, DOI 10.1038/bjc.2011.357
   Voldborg BR, 1997, ANN ONCOL, V8, P1197, DOI 10.1023/A:1008209720526
   von Deimling A, 2011, BRAIN PATHOL, V21, P74, DOI 10.1111/j.1750-3639.2010.00454.x
   Wahl M, 2017, CANCER-AM CANCER SOC, V123, P4631, DOI 10.1002/cncr.30909
   Wainwright DA, 2012, CLIN CANCER RES, V18, P6110, DOI 10.1158/1078-0432.CCR-12-2130
   Wakimoto H, 2014, CLIN CANCER RES, V20, P2898, DOI 10.1158/1078-0432.CCR-13-3052
   Walker DG, 2006, J NEUROIMMUNOL, V175, P59, DOI 10.1016/j.jneuroim.2006.03.006
   Wang DW, 2021, FRONT GENET, V12, DOI 10.3389/fgene.2021.709666
   Wang JG, 2016, NAT GENET, V48, P768, DOI 10.1038/ng.3590
   Wang P, 2015, CELL REP, V13, P2353, DOI 10.1016/j.celrep.2015.11.029
   Ward HA, 2018, EUR J CANCER PREV, V27, P379, DOI [10.1097/CEJ.0000000000000331, 10.1097/cej.0000000000000331]
   Watanabe T, 2002, ACTA NEUROPATHOL, V103, P267, DOI 10.1007/s004010100464
   Weber J, 2017, NEW ENGL J MED, V377, P1824, DOI 10.1056/NEJMoa1709030
   WELLENREUTHER R, 1995, AM J PATHOL, V146, P827
   Weller M, 2020, CANCER TREAT REV, V87, DOI 10.1016/j.ctrv.2020.102029
   Weller M, 2017, LANCET ONCOL, V18, P1373, DOI 10.1016/S1470-2045(17)30517-X
   Weller M, 2017, LANCET ONCOL, V18, pE315, DOI 10.1016/S1470-2045(17)30194-8
   Weller M, 2017, JNCI-J NATL CANCER I, V109, DOI 10.1093/jnci/djw320
   Weller M, 2012, NEURO-ONCOLOGY, V14, P1481, DOI 10.1093/neuonc/nos159
   Wen PY, 2019, CLIN CANCER RES, V25, P5799, DOI 10.1158/1078-0432.CCR-19-0261
   Wen PY, 2018, NEURO-ONCOLOGY, V20, P249, DOI 10.1093/neuonc/nox154
   Wen PY, 2011, NEURO-ONCOLOGY, V13, P437, DOI 10.1093/neuonc/noq198
   Wick W, 2019, NEURO-ONCOLOGY, V21, P95, DOI 10.1093/neuonc/noy161
   Wickstrom M, 2015, NAT COMMUN, V6, DOI 10.1038/ncomms9904
   Wiestler B, 2013, ACTA NEUROPATHOL, V126, P443, DOI 10.1007/s00401-013-1156-z
   Williams EA, 2020, ACTA NEUROPATHOL COM, V8, DOI 10.1186/s40478-020-01040-2
   Williams SR, 2019, J NEUROL SURG PART B, V80, P562, DOI 10.1055/s-0038-1676821
   Wolchok JD, 2008, ONCOLOGIST, V13, P2, DOI 10.1634/theoncologist.13-S4-2
   Won WJ, 2019, CELL STRESS, V3, P47, DOI 10.15698/cst2019.02.176
   Woo HY, 2020, BRAIN TUMOR PATHOL, V37, P136, DOI 10.1007/s10014-020-00377-9
   Woroniecka K, 2018, CLIN CANCER RES, V24, P4175, DOI 10.1158/1078-0432.CCR-17-1846
   Wu G, 2014, NAT GENET, V46, P444, DOI 10.1038/ng.2938
   Wu SY, 2020, MOL CANCER, V19, DOI 10.1186/s12943-020-01238-x
   Xie Q, 2012, P NATL ACAD SCI USA, V109, P570, DOI 10.1073/pnas.1119059109
   Xu W, 2011, CANCER CELL, V19, P17, DOI 10.1016/j.ccr.2010.12.014
   Xue NN, 2017, SCI REP-UK, V7, DOI 10.1038/srep39011
   Yalon M, 2013, PEDIATR BLOOD CANCER, V60, P71, DOI 10.1002/pbc.24142
   Yang KY, 2022, MOL CANCER, V21, DOI 10.1186/s12943-022-01513-z
   You H, 2021, CANCER LETT, V509, P121, DOI 10.1016/j.canlet.2021.02.025
   Youngblood MW, 2021, NEURO-ONCOLOGY, V23, P783, DOI 10.1093/neuonc/noaa226
   Youngblood MW, 2020, J NEUROSURG, V133, P1345, DOI 10.3171/2019.8.JNS191266
   Youssef G, 2020, CURR NEUROL NEUROSCI, V20, DOI 10.1007/s11910-020-01040-8
   Yun EJ, 2020, CELL DEATH DIS, V11, DOI 10.1038/s41419-020-02988-8
   ZANG KD, 1967, NATURE, V216, P84, DOI 10.1038/216084a0
   Zang KD, 2001, CYTOGENET CELL GENET, V93, P207, DOI 10.1159/000056986
   Zarghooni M, 2010, J CLIN ONCOL, V28, P1337, DOI 10.1200/JCO.2009.25.5463
   Zhang CC, 2016, JNCI-J NATL CANCER I, V108, DOI 10.1093/jnci/djv375
   Zhang WCB, 2022, FRONT ONCOL, V12, DOI 10.3389/fonc.2022.910476
   Zhang WCB, 2022, FRONT NUTR, V9, DOI 10.3389/fnut.2022.834258
   Zhang XH, 2012, CANCER SCI, V103, P181, DOI 10.1111/j.1349-7006.2011.02154.x
   Zhang YN, 2016, ONCOTARGET, V7, P37370, DOI 10.18632/oncotarget.8276
   Zhao K, 2017, CANCER LETT, V406, P12, DOI 10.1016/j.canlet.2017.07.022
   Zhao SM, 2009, SCIENCE, V324, P261, DOI 10.1126/science.1170944
   Zheng SY, 2019, MOL CANCER THER, V18, P991, DOI 10.1158/1535-7163.MCT-18-1122
   Zhu SM, 2021, J HEMATOL ONCOL, V14, DOI 10.1186/s13045-021-01164-5
   Zhu Z, 2020, CELL STEM CELL, V26, P187, DOI 10.1016/j.stem.2019.11.016
NR 431
TC 3
Z9 3
U1 8
U2 11
PU SPRINGERNATURE
PI LONDON
PA CAMPUS, 4 CRINAN ST, LONDON, N1 9XW, ENGLAND
SN 2095-9907
EI 2059-3635
J9 SIGNAL TRANSDUCT TAR
JI Signal Transduct. Target. Ther.
PD JAN 4
PY 2023
VL 8
IS 1
AR 8
DI 10.1038/s41392-022-01260-z
PG 23
WC Biochemistry & Molecular Biology; Cell Biology
WE Science Citation Index Expanded (SCI-EXPANDED)
SC Biochemistry & Molecular Biology; Cell Biology
GA 7N1MJ
UT WOS:000907109900004
PM 36596785
OA Green Published, gold
DA 2023-08-27
ER

PT J
AU Hong, BJ
   Krauss, JK
   Bremer, M
   Karstens, JH
   Heissler, HE
   Nakamura, M
AF Hong, Bujung
   Krauss, Joachim K.
   Bremer, Michael
   Karstens, Johann H.
   Heissler, Hans E.
   Nakamura, Makoto
TI Vestibular Schwannoma Microsurgery for Recurrent Tumors After Radiation
   Therapy or Previous Surgical Resection
SO OTOLOGY & NEUROTOLOGY
LA English
DT Article
DE Recurrent vestibular schwannoma; Radiation therapy; Surgical resection
ID GAMMA-KNIFE RADIOSURGERY; STEREOTACTIC RADIOSURGERY; ACOUSTIC NEUROMAS;
   MALIGNANT-TRANSFORMATION; LONG-TERM; FOLLOW-UP; MANAGEMENT; SURGERY;
   RADIOTHERAPY; SALVAGE
AB Objective: The purpose of this study is to compare the outcomes in patients who underwent microsurgical resection for recurrent vestibular schwannoma after microsurgical resection and previous radiation therapy.
   Study Design: Retrospective study.
   Setting: University hospital.
   Patients: Fifteen patients, who underwent microsurgical resection for recurrent vestibular schwannoma after previous surgery (group A), and 5 patients, who underwent microsurgical resection after previous radiation therapy (group B) were included.
   Intervention: Surgical resection after radiation therapy or previous surgical resection.
   Main Outcome Measures: Intraoperative findings and postoperative facial nerve function were investigated in groups A and B.
   Results: Mean tumor volumes were 18.4 +/- 2.44 cm(3) in group A and 19.0 +/- 1.53 cm(3) in group B. Total resection was achieved in 10 patients (67%) of group A and in 3 patients (60.0%) of group B. The tumor was more difficult to resect because of severe adhesions to the facial nerve. Anatomic facial nerve preservation could be achieved in 19 patients. Mean follow-up time was 80 months for group A and 28 months for group B. At last follow-up, 7 patients (53.8%) of group A had a good facial nerve function. In 3 patients (75.0%) of group B, the preoperative facial nerve function was preserved postoperatively. Preexistent facial paresis, large tumor with extrameatal growth and brainstem compression correlated with poor postoperative facial nerve function.
   Conclusion: Surgical outcome of recurrent vestibular schwannoma is more unsatisfactory than after primary surgery. It remains to be clarified whether previous surgery may implicate a higher risk for postoperative facial nerve function than previous radiation therapy upon surgery for tumor recurrence.
C1 [Hong, Bujung; Krauss, Joachim K.; Heissler, Hans E.; Nakamura, Makoto] Hannover Med Sch, Dept Neurosurg, D-30625 Hannover, Germany.
   [Bremer, Michael; Karstens, Johann H.] Hannover Med Sch, Dept Radiat Oncol, D-30625 Hannover, Germany.
C3 Hannover Medical School; Hannover Medical School
RP Hong, BJ (通讯作者)，Hannover Med Sch, Dept Neurosurg, Carl Neuberg Str 1, D-30625 Hannover, Germany.
EM
RI Nakamura, Makoto/F-2620-2014
OI Nakamura, Makoto/0000-0001-8597-0193
CR Arlt F, 2011, NEUROL RES, V33, P1032, DOI 10.1179/1743132811Y.0000000027
   Battista RA, 2009, OTOLARYNG CLIN N AM, V42, P635, DOI 10.1016/j.otc.2009.04.009
   Bloch DC, 2004, OTOLARYNG HEAD NECK, V130, P104, DOI 10.1016/S0194-5998(03)01598-5
   Bloch O, 2011, J NEURO-ONCOL, V102, P281, DOI 10.1007/s11060-010-0315-5
   Combs SE, 2010, INT J RADIAT ONCOL, V76, P193, DOI 10.1016/j.ijrobp.2009.01.064
   Delsanti Christine, 2008, Prog Neurol Surg, V21, P93, DOI 10.1159/000156712
   El-Kashlan HK, 2000, AM J OTOL, V21, P389, DOI 10.1016/S0196-0709(00)80049-6
   Freeman SRM, 2007, OTOL NEUROTOL, V28, P1076, DOI 10.1097/MAO.0b013e318159e76a
   Friedman RA, 2005, LARYNGOSCOPE, V115, P1827, DOI 10.1097/01.mlg.0000175063.76945.75
   Friedman RA, 2011, OTOL NEUROTOL, V32, P1322, DOI 10.1097/MAO.0b013e31822e5b76
   Gerganov VM, 2012, J NEUROSURG, V116, P713, DOI 10.3171/2011.12.JNS111682
   Hasegawa T, 2005, NEUROSURGERY, V57, P257, DOI 10.1227/01.NEU.0000166542.00512.84
   Hayhurst C, 2012, NEURO-ONCOLOGY, V14, P87, DOI 10.1093/neuonc/nor171
   HOUSE JW, 1985, OTOLARYNG HEAD NECK, V93, P146, DOI 10.1177/019459988509300202
   Iwai Y, 2007, NEUROSURGERY, V60, P75, DOI 10.1227/01.NEU.0000249251.78794.45
   Kano H, 2010, INT J RADIAT ONCOL, V76, P520, DOI 10.1016/j.ijrobp.2009.01.076
   Lee CC, 2010, J NEURO-ONCOL, V98, P203, DOI 10.1007/s11060-010-0178-9
   Lee DJ, 2003, OTOL NEUROTOL, V24, P650, DOI 10.1097/00129492-200307000-00020
   Lee F, 2002, LARYNGOSCOPE, V112, P948, DOI 10.1097/00005537-200206000-00002
   Limb CJ, 2005, LARYNGOSCOPE, V115, P93, DOI 10.1097/01.mlg.0000150686.93011.49
   Linthicum FH, 2002, ORL J OTO-RHINO-LARY, V64, P138, DOI 10.1159/000057793
   Liscak R, 2009, ACTA NEUROCHIR, V151, P317, DOI 10.1007/s00701-009-0254-0
   Markou K, 2012, AM J OTOLARYNG, V33, P168, DOI 10.1016/j.amjoto.2011.02.006
   Milligan BD, 2012, J NEUROSURG, V116, P598, DOI 10.3171/2011.11.JNS11811
   Murakami K, 2011, ACTA NEUROCHIR, V153, P1201, DOI 10.1007/s00701-011-0972-y
   Murphy ES, 2011, J NEUROSURG, V114, P432, DOI 10.3171/2009.12.JNS091339
   Nagano O, 2008, J NEUROSURG, V109, P811, DOI 10.3171/JNS/2008/109/11/0811
   Nakamura H, 2000, AM J NEURORADIOL, V21, P1540
   Patni AH, 2005, OTOLARYNG HEAD NECK, V132, P11, DOI 10.1016/j.otohns.2004.09.094
   Pollock BE, 1998, J NEUROSURG, V89, P944, DOI 10.3171/jns.1998.89.6.0944
   Pollock BE, 2006, NEUROSURGERY, V58, P241, DOI 10.1227/01.NEU.0000194833.66593.8B
   Pollock Bruce E, 2008, Prog Neurol Surg, V21, P163, DOI 10.1159/000156904
   Ramina R, 2007, SKULL BASE-INTERD AP, V17, P109, DOI 10.1055/s-2006-953510
   Regis J, 2012, CANCER RADIOTHER, V16, pS70, DOI 10.1016/j.canrad.2012.01.006
   Roche PH, 2004, NEUROCHIRURGIE, V50, P383
   Roche PH, 2012, OTOLARYNG CLIN N AM, V45, P367, DOI 10.1016/j.otc.2011.12.007
   Roche Pierre-Hugues, 2008, Prog Neurol Surg, V21, P158, DOI 10.1159/000156903
   Roche Pierre-Hugues, 2008, Prog Neurol Surg, V21, P152, DOI 10.1159/000156902
   Roche Pierre-Hugues, 2008, Prog Neurol Surg, V21, P89, DOI 10.1159/000156711
   Roos DE, 2012, INT J RADIAT ONCOL, V82, P1352, DOI 10.1016/j.ijrobp.2011.04.068
   Samii M, 1997, NEUROSURGERY, V40, P248, DOI 10.1097/00006123-199702000-00005
   Shin M, 2002, LANCET, V360, P309, DOI 10.1016/S0140-6736(02)09521-1
   Shuto T, 2008, ACTA NEUROCHIR, V150, P229, DOI 10.1007/s00701-007-1486-5
   Slattery WH, 2009, OTOLARYNG CLIN N AM, V42, P707, DOI 10.1016/j.otc.2009.04.007
   Sughrue ME, 2011, J NEUROSURG, V114, P1218, DOI 10.3171/2010.11.JNS10257
   Szeifert GT, 2004, NEUROCHIRURGIE, V50, P327
   Timmer FCA, 2011, LARYNGOSCOPE, V121, P1359, DOI 10.1002/lary.21763
   Wanibuchi M, 2009, J NEUROSURG, V111, P845, DOI 10.3171/2008.12.JNS08620
   Wilkinson JS, 2004, J CLIN PATHOL, V57, P109, DOI 10.1136/jcp.57.1.109
   Yomo S, 2009, NEUROSURGERY, V64, P48, DOI 10.1227/01.NEU.0000327692.74477.D5
NR 50
TC 17
Z9 18
U1 2
U2 4
PU LIPPINCOTT WILLIAMS & WILKINS
PI PHILADELPHIA
PA 530 WALNUT ST, PHILADELPHIA, PA 19106-3621 USA
SN 1531-7129
EI 1537-4505
J9 OTOL NEUROTOL
JI Otol. Neurotol.
PD JAN
PY 2014
VL 35
IS 1
BP 171
EP 181
DI 10.1097/MAO.0000000000000174
PG 11
WC Clinical Neurology; Otorhinolaryngology
WE Science Citation Index Expanded (SCI-EXPANDED)
SC Neurosciences & Neurology; Otorhinolaryngology
GA AJ5BB
UT WOS:000337694300038
PM 24335935
DA 2023-08-27
ER

PT J
AU Guclu, B
   Sindou, M
   Meyronet, D
   Streichenberger, N
   Simon, E
   Mertens, P
AF Guclu, Bulent
   Sindou, Marc
   Meyronet, David
   Streichenberger, Nathalie
   Simon, Emile
   Mertens, Patrick
TI Anatomical study of the central myelin portion and transitional zone of
   the vestibulocochlear nerve
SO ACTA NEUROCHIRURGICA
LA English
DT Article
DE Anatomy; Brainstem; Cranial nerves; Transitional zone; Vascular
   compression syndromes; Vestibulocochlear nerve
ID DISABLING POSITIONAL VERTIGO; 8TH CRANIAL NERVE; MICROVASCULAR
   DECOMPRESSION; COMPRESSION SYNDROMES; SELECTION CRITERIA; SEVERE
   TINNITUS; COCHLEAR NERVE; ORIGIN; ROOT; TUMOR
AB The aim of this study was to evaluate gross and microscopic anatomical features of the vestibulocochlear nerve or eighth cranial nerve (CNVIII) from fresh cadavers, especially the nerve's central myelin portion (CMP) and transitional zone (TZ), and to consider any pathological implications.
   Six fresh cadavers were used to examine the CNVIII. Its cisternal length from brainstem to internal auditory meatus was measured. Longitudinal sections were stained to make following measurements: the diameter where the nerve enters the brainstem, the diameter where the TZ begins, the distance to the most distal part of TZ from the brainstem, and the depth of the TZ. The volume of the CMP was calculated as well.
   The cisternal length of ten CNVIIIs measured between 14.2 and 19.2 mm (16.48 +/- 1.78 mm). The thickness where the CNVIII enters the brainstem was between 1.21 and 3.16 mm (2.31 +/- 0.68 mm); the thickness where the TZ begins was between 1.07 and 2.21 mm (1.44 +/- 0.38 mm); the distance of the most distal part of the TZ from the brainstem was between 9.28 and 13.84 mm (11.50 +/- 1.56 mm); the depth of the TZ was between 0.56 and 1.28 mm (0.81 +/- 0.27 mm). The volume of the CMP was between 17.34 and 53.87 mm(3) (33.98 +/- 13.74 mm(3)). The measurements were compared to trigeminal, facial, glossopharyngeal and vagus nerves. CNVIII was the nerve with the longest CMP.
   The measurements showed that the CMP of CNVIII was very long. The implication of this length in the dysfunctional syndromes of this nerve, its propensity to harbor schwannomas, which most frequently arise at the porus of the auditory meatus, and the vulnerability to damages are discussed.
C1 [Guclu, Bulent] Sevket Yilmaz Res & Training Hosp, Minist Hlth, Neurosurg Clin, TR-16310 Yildirim, Bursa, Turkey.
   [Guclu, Bulent; Sindou, Marc; Simon, Emile; Mertens, Patrick] Univ Lyon 1, Hop Neurolog Pierre Wertheimer, Dept Neurosurg, F-69365 Lyon, France.
   [Meyronet, David; Streichenberger, Nathalie] Hosp Civils Lyon, Ctr Pathol & Neuropathol E, F-69003 Lyon, France.
   [Meyronet, David; Streichenberger, Nathalie] Univ Lyon 1, Neurosci Ctr, Inserm U1028, CNRS UMR 5292, F-6900 Lyon, France.
   [Simon, Emile; Mertens, Patrick] Univ Lyon 1, Dept Anat, F-69365 Lyon, France.
C3 Ministry of Health - Turkey; Sevket Yilmaz Training & Research Hospital;
   CHU Lyon; UDICE-French Research Universities; Universite Claude Bernard
   Lyon 1; CHU Lyon; Centre National de la Recherche Scientifique (CNRS);
   CNRS - National Institute for Biology (INSB); UDICE-French Research
   Universities; Universite Claude Bernard Lyon 1; Institut National de la
   Sante et de la Recherche Medicale (Inserm); Universite Jean Monnet;
   UDICE-French Research Universities; Universite Claude Bernard Lyon 1
RP Guclu, B (通讯作者)，Sevket Yilmaz Res & Training Hosp, Minist Hlth, Neurosurg Clin, TR-16310 Yildirim, Bursa, Turkey.
EM
RI Conti, Alfredo/H-3022-2019; Simon, Emile/C-1890-2012; guclu,
   bulent/ITU-7284-2023
OI Conti, Alfredo/0000-0002-9408-2662; Mertens, Patrick/0000-0001-5272-261X
CR BRIDGER MWM, 1980, J LARYNGOL OTOL, V94, P1353, DOI 10.1017/S0022215100090186
   De Ridder D, 2002, NEUROSURGERY, V51, P427, DOI 10.1097/00006123-200208000-00023
   ESCOUROLLE R, 1971, Neurochirurgie, V17, P25
   FONCIN JF, 1979, ANN OTO-LAR CHIR C-F, V96, P11
   Guclu B, 2009, NEUROCHIRURGIE, V55, P92, DOI 10.1016/j.neuchi.2009.02.001
   Guclu B, 2011, ACTA NEUROCHIR, V153, P2365, DOI 10.1007/s00701-011-1168-1
   Guevara N, 2008, EUR ARCH OTO-RHINO-L, V265, P397, DOI 10.1007/s00405-007-0471-1
   Jannetta P J, 1997, Clin Neurosurg, V44, P69
   JANNETTA PJ, 1984, NEW ENGL J MED, V310, P1700, DOI 10.1056/NEJM198406283102604
   JANNETTA PJ, 1975, SURG FORUM, V26, P467
   MOLLER AR, 1984, ANN OTO RHINOL LARYN, V93, P39
   Moller AR, 2006, NEURAL PLASTICITY DI
   MOLLER MB, 1993, ACTA NEUROCHIR, V125, P75, DOI 10.1007/BF01401831
   MOLLER MB, 1993, LARYNGOSCOPE, V103, P421, DOI 10.1002/lary.5541030410
   Murray MR, 1940, AM J PATHOL, V16, P41
   NEELY JG, 1976, LARYNGOSCOPE, V86, P984, DOI 10.1288/00005537-197607000-00012
   Obersteiner H, 1894, ARB NEUROL I WIEN, V1-3, P158
   Pellet W, 2004, NEUROCHIRURGIE, V50, P195
   Pirsig W, 1979, ACOUSTIC TUMORS, V1, P52
   Rubinstein D, 1996, AM J NEURORADIOL, V17, P1099
   Ryu H, 1998, ACTA NEUROCHIR, V140, P1279, DOI 10.1007/s007010050250
   SCHEFTER RP, 1986, ANN OTO RHINOL LARYN, V95, P146, DOI 10.1177/000348948609500207
   Skinner HA, 1929, BRIT J SURG, V16, P440, DOI 10.1002/bjs.1800166308
   Skinner HA, 1931, ARCH NEURO PSYCHIATR, V25, P356, DOI 10.1001/archneurpsyc.1931.02230020144008
   Tarlov IM, 1937, ARCH NEURO PSYCHIATR, V37, P555, DOI 10.1001/archneurpsyc.1937.02260150085005
   Tarlov IM, 1937, ARCH NEURO PSYCHIATR, V37, P1338, DOI 10.1001/archneurpsyc.1937.02260180118008
   Vasama JPI, 1998, NEUROL RES, V20, P242, DOI 10.1080/01616412.1998.11740513
NR 27
TC 12
Z9 12
U1 1
U2 9
PU SPRINGER WIEN
PI WIEN
PA SACHSENPLATZ 4-6, PO BOX 89, A-1201 WIEN, AUSTRIA
SN 0001-6268
J9 ACTA NEUROCHIR
JI Acta Neurochir.
PD DEC
PY 2012
VL 154
IS 12
BP 2277
EP 2283
DI 10.1007/s00701-012-1479-x
PG 7
WC Clinical Neurology; Surgery
WE Science Citation Index Expanded (SCI-EXPANDED)
SC Neurosciences & Neurology; Surgery
GA 040XJ
UT WOS:000311359500021
PM 22914910
DA 2023-08-27
ER

PT J
AU Hwang, EI
   Sayour, EJ
   Flores, CT
   Grant, G
   Wechsler-Reya, R
   Hoang-Minh, LB
   Kieran, MW
   Salcido, J
   Prins, RM
   Figg, JW
   Platten, M
   Candelario, KM
   Hale, PG
   Blatt, JE
   Governale, LS
   Okada, H
   Mitchell, DA
   Pollack, IF
AF Hwang, Eugene, I
   Sayour, Elias J.
   Flores, Catherine T.
   Grant, Gerald
   Wechsler-Reya, Robert
   Hoang-Minh, Lan B.
   Kieran, Mark W.
   Salcido, Joanne
   Prins, Robert M.
   Figg, John W.
   Platten, Michael
   Candelario, Kate M.
   Hale, Paul G.
   Blatt, Jason E.
   Governale, Lance S.
   Okada, Hideho
   Mitchell, Duane A.
   Pollack, Ian F.
TI The current landscape of immunotherapy for pediatric brain tumors
SO NATURE CANCER
LA English
DT Review
ID CENTRAL-NERVOUS-SYSTEM; DENDRITIC CELL VACCINATION; INTRINSIC PONTINE
   GLIOMAS; NEURAL PROGENITOR CELLS; REGULATORY T-CELLS; HIGH-GRADE; MOUSE
   MODEL; IMMUNE-RESPONSES; CHECKPOINT INHIBITION; GLIOBLASTOMA PATIENTS
AB Pediatric central nervous system tumors are the most common solid malignancies in childhood, and aggressive therapy often leads to long-term sequelae in survivors, making these tumors challenging to treat. Immunotherapy has revolutionized prospects for many cancer types in adults, but the intrinsic complexity of treating pediatric patients and the scarcity of clinical studies of children to inform effective approaches have hampered the development of effective immunotherapies in pediatric settings. Here, we review recent advances and ongoing challenges in pediatric brain cancer immunotherapy, as well as considerations for efficient clinical translation of efficacious immunotherapies into pediatric settings.
   Hwang and colleagues discuss recent advances and current challenges in developing immunotherapies for pediatric brain cancer, as well as the clinical implications of ongoing and completed studies.
C1 [Hwang, Eugene, I] Childrens Natl Hosp, Brain Tumor Inst, Div Oncol, Washington, DC 20010 USA.
   [Sayour, Elias J.; Flores, Catherine T.; Hoang-Minh, Lan B.; Figg, John W.; Candelario, Kate M.; Blatt, Jason E.; Governale, Lance S.; Mitchell, Duane A.] Univ Florida, Preston A Wells Jr Ctr Brain Tumor Therapy, Dept Neurosurg, Gainesville, FL USA.
   [Grant, Gerald] Stanford Univ, Lucile Packard Childrens Hosp, Div Pediat Neurosurg, Palo Alto, CA 94304 USA.
   [Wechsler-Reya, Robert] NCI, Tumor Initiat & Maintenance Program, Canc Ctr, Sanford Burnham Prebys Med Discovery Inst, La Jolla, CA USA.
   [Kieran, Mark W.] Day One Biopharmaceut, San Francisco, CA USA.
   [Salcido, Joanne] Brain Tumor Fdn, Asheville, NC USA.
   [Prins, Robert M.] Univ Calif Los Angeles, David Geffen Sch Med, Dept Neurosurg, Los Angeles, CA 90095 USA.
   [Prins, Robert M.] Univ Calif Los Angeles, David Geffen Sch Med, Dept Mol & Med Pharmacol, Los Angeles, CA 90095 USA.
   [Platten, Michael] Heidelberg Univ, Med Fac Mannheim, Dept Neurol, MCTN, Heidelberg, Germany.
   [Platten, Michael] DKFZ, CCU Brain Tumor Immunol, Heidelberg, Germany.
   [Hale, Paul G.] Childrens Brain Trust, Coral Springs, FL USA.
   [Okada, Hideho] Univ Calif San Francisco, Dept Neurosurg, San Francisco, CA USA.
   [Pollack, Ian F.] Univ Pittsburgh, Sch Med, Dept Neurol Surg, Pittsburgh, PA 15261 USA.
C3 Children's National Health System; State University System of Florida;
   University of Florida; Lucile Packard Children's Hospital (LPCH);
   Stanford University; National Institutes of Health (NIH) - USA; NIH
   National Cancer Institute (NCI); Sanford Burnham Prebys Medical
   Discovery Institute; University of California System; University of
   California Los Angeles; University of California Los Angeles Medical
   Center; David Geffen School of Medicine at UCLA; University of
   California System; University of California Los Angeles; University of
   California Los Angeles Medical Center; David Geffen School of Medicine
   at UCLA; Ruprecht Karls University Heidelberg; Helmholtz Association;
   German Cancer Research Center (DKFZ); University of California System;
   University of California San Francisco; Pennsylvania Commonwealth System
   of Higher Education (PCSHE); University of Pittsburgh
RP Hwang, EI (通讯作者)，Childrens Natl Hosp, Brain Tumor Inst, Div Oncol, Washington, DC 20010 USA.
EM
RI Platten, Michael/F-2902-2013
OI Platten, Michael/0000-0002-4746-887X; Blatt, Jason/0000-0001-6892-7724;
   Prins, Robert/0000-0002-6282-6583
FU ReMission Alliance against Brain Tumors
FX This publication was supported in part by the ReMission Alliance against
   Brain Tumors.
CR Acharya UH, 2019, EXPERT REV HEMATOL, V12, P195, DOI 10.1080/17474086.2019.1585238
   Ahluwalia MS, 2019, J CLIN ONCOL, V37, DOI 10.1200/JCO.2019.37.15_suppl.2016
   Ahmed N, 2017, JAMA ONCOL, V3, P1094, DOI 10.1001/jamaoncol.2017.0184
   Akhavan D, 2019, IMMUNOL REV, V290, P60, DOI 10.1111/imr.12773
   Albert ML, 1998, NAT MED, V4, P1321, DOI 10.1038/3315
   Alexandrov LB, 2013, NATURE, V500, P415, DOI 10.1038/nature12477
   Andradas C, 2021, CANCERS, V13, DOI 10.3390/cancers13020330
   Askew K, 2017, CELL REP, V18, P391, DOI 10.1016/j.celrep.2016.12.041
   Aurelian L, 2016, ONCOTARGETS THER, V9, P2627, DOI 10.2147/OTT.S63049
   Badodi S, 2017, CELL REP, V21, P2772, DOI 10.1016/j.celrep.2017.11.021
   Bardella C, 2016, CANCER CELL, V30, P578, DOI 10.1016/j.ccell.2016.08.017
   Beatty GL, 2014, ONCOIMMUNOLOGY, V3, DOI 10.4161/21624011.2014.970027
   Becher OJ, 2010, CANCER RES, V70, P2548, DOI 10.1158/0008-5472.CAN-09-2503
   Bian S, 2018, NAT METHODS, V15, P631, DOI 10.1038/s41592-018-0070-7
   Bloch O, 2013, CLIN CANCER RES, V19, P3165, DOI 10.1158/1078-0432.CCR-12-3314
   Bouffet E, 2016, J CLIN ONCOL, V34, P2206, DOI 10.1200/JCO.2016.66.6552
   Brahmer JR, 2018, J ONCOL PRACT, V14, P247, DOI 10.1200/JOP.18.00005
   Bristol Myers Squibb, 2019, BRIST MYERS SQUIBB A
   Brown CE, 2016, NEW ENGL J MED, V375, P2561, DOI 10.1056/NEJMoa1610497
   Buczkowicz P, 2014, NAT GENET, V46, P451, DOI 10.1038/ng.2936
   Cancer M, 2019, CELL STEM CELL, V25, P855, DOI 10.1016/j.stem.2019.10.005
   Castel D, 2015, ACTA NEUROPATHOL, V130, P815, DOI 10.1007/s00401-015-1478-0
   Ceschin R, 2015, AM J NEURORADIOL, V36, P2170, DOI 10.3174/ajnr.A4428
   Chan TA, 2015, NEW ENGL J MED, V373, P1984, DOI 10.1056/NEJMc1508163
   Chen ZR, 2021, CANCERS, V13, DOI 10.3390/cancers13051114
   Chheda ZS, 2018, J EXP MED, V215, P141, DOI 10.1084/jem.20171046
   Choi BD, 2019, NAT BIOTECHNOL, V37, P1049, DOI 10.1038/s41587-019-0192-1
   Choi BD, 2013, P NATL ACAD SCI USA, V110, P270, DOI 10.1073/pnas.1219817110
   Cloughesy TF, 2019, NAT MED, V25, P477, DOI 10.1038/s41591-018-0337-7
   Cohen KJ, 2011, NEURO-ONCOLOGY, V13, P410, DOI 10.1093/neuonc/noq205
   Cordero FJ, 2017, MOL CANCER RES, V15, P1243, DOI 10.1158/1541-7786.MCR-16-0389
   Davis AA, 2019, J IMMUNOTHER CANCER, V7, DOI 10.1186/s40425-019-0768-9
   de Vries NA, 2010, CLIN CANCER RES, V16, P3431, DOI 10.1158/1078-0432.CCR-09-3414
   Deng JH, 2018, CANCER DISCOV, V8, P216, DOI 10.1158/2159-8290.CD-17-0915
   Desjardins A, 2018, NEW ENGL J MED, V379, P150, DOI 10.1056/NEJMoa1716435
   Dey J, 2012, MOL CELL BIOL, V32, P4104, DOI 10.1128/MCB.00862-12
   Dhar SS, 2018, MOL CELL, V70, P825, DOI 10.1016/j.molcel.2018.04.028
   Khuong-Quang DA, 2012, ACTA NEUROPATHOL, V124, P439, DOI 10.1007/s00401-012-0998-0
   Donson AM, 2009, J IMMUNOL, V183, P7428, DOI 10.4049/jimmunol.0902811
   Doucette T, 2013, CANCER IMMUNOL RES, V1, P112, DOI 10.1158/2326-6066.CIR-13-0028
   Duan SL, 2015, NAT COMMUN, V6, DOI 10.1038/ncomms10068
   Duraiswamy J, 2013, CANCER RES, V73, P3591, DOI 10.1158/0008-5472.CAN-12-4100
   Dutoit V, 2018, ONCOIMMUNOLOGY, V7, DOI 10.1080/2162402X.2017.1391972
   Eder N, 2020, NAT COMMUN, V11, DOI 10.1038/s41467-020-16167-y
   Elshafeey N, 2019, NAT COMMUN, V10, DOI 10.1038/s41467-019-11007-0
   Engelhardt B, 2006, J NEURAL TRANSM, V113, P477, DOI 10.1007/s00702-005-0409-y
   Fenstermaker RA, 2016, CANCER IMMUNOL IMMUN, V65, P1339, DOI 10.1007/s00262-016-1890-x
   Filley AC, 2017, ONCOTARGET, V8, P91779, DOI 10.18632/oncotarget.21586
   Fisher GH, 1999, ONCOGENE, V18, P5253, DOI 10.1038/sj.onc.1203087
   Flores C, 2019, SCI ADV, V5, DOI 10.1126/sciadv.aav9879
   Forget A, 2018, CANCER CELL, V34, P379, DOI 10.1016/j.ccell.2018.08.002
   Franchimont D, 2004, ANN NY ACAD SCI, V1024, P124, DOI 10.1196/annals.1321.009
   Fried I, 2018, J NEURO-ONCOL, V136, P189, DOI 10.1007/s11060-017-2643-1
   Fults D, 2002, NEOPLASIA, V4, P32, DOI 10.1038/sj.neo.7900200
   Funato K, 2014, SCIENCE, V346, P1529, DOI 10.1126/science.1253799
   Gattinoni L, 2005, J EXP MED, V202, P907, DOI 10.1084/jem.20050732
   Geoerger B, 2020, LANCET ONCOL, V21, pE127, DOI 10.1016/S1470-2045(20)30105-4
   Gholamin S, 2017, SCI TRANSL MED, V9, DOI 10.1126/scitranslmed.aaf2968
   Gibson P, 2010, NATURE, V468, P1095, DOI 10.1038/nature09587
   Goff SL, 2019, J IMMUNOTHER, V42, P126, DOI 10.1097/CJI.0000000000000260
   Goodrich LV, 1997, SCIENCE, V277, P1109, DOI 10.1126/science.277.5329.1109
   Grabovska Y, 2020, NAT COMMUN, V11, DOI 10.1038/s41467-020-18070-y
   Grammel D, 2012, ACTA NEUROPATHOL, V123, P601, DOI 10.1007/s00401-012-0961-0
   Gust J, 2017, CANCER DISCOV, V7, P1404, DOI 10.1158/2159-8290.CD-17-0698
   Haanen J B A G, 2018, Ann Oncol, V29, piv264, DOI 10.1093/annonc/mdy162
   Hallahan AR, 2004, CANCER RES, V64, P7794, DOI 10.1158/0008-5472.CAN-04-1813
   Halvorson KG, 2015, PLOS ONE, V10, DOI 10.1371/journal.pone.0118926
   Hamid O, 2013, NEW ENGL J MED, V369, P134, DOI 10.1056/NEJMoa1305133
   Han ZY, 2016, NAT COMMUN, V7, DOI 10.1038/ncomms10421
   Hanahan D, 2011, CELL, V144, P646, DOI 10.1016/j.cell.2011.02.013
   Harutyunyan AS, 2019, NAT COMMUN, V10, DOI 10.1038/s41467-019-09140-x
   Hashizume R, 2012, J NEURO-ONCOL, V110, P305, DOI 10.1007/s11060-012-0973-6
   Hatton BA, 2008, CANCER RES, V68, P1768, DOI 10.1158/0008-5472.CAN-07-5092
   Haydar D, 2021, NEURO-ONCOLOGY, V23, P999, DOI 10.1093/neuonc/noaa278
   He P, 2011, EUR J NUCL MED MOL I, V38, P334, DOI 10.1007/s00259-010-1633-8
   Hede SM, 2009, GLIA, V57, P1143, DOI 10.1002/glia.20837
   Heimberger AB, 2008, CLIN CANCER RES, V14, P5166, DOI 10.1158/1078-0432.CCR-08-0320
   Heiss JD, 2019, J NEUROSURG-PEDIATR, V23, P333, DOI 10.3171/2018.9.PEDS17225
   Hilf N, 2019, NATURE, V565, P240, DOI 10.1038/s41586-018-0810-y
   Holland EC, 2000, TOXICOL PATHOL, V28, P171, DOI 10.1177/019262330002800122
   Hong JJ, 2010, CLIN CANCER RES, V16, P4892, DOI 10.1158/1078-0432.CCR-10-1507
   Huang M, 2019, CELL STEM CELL, V25, P433, DOI 10.1016/j.stem.2019.05.013
   Huq S, 2021, J NEUROSURG-PEDIATR, V27, P482, DOI 10.3171/2020.8.PEDS20561
   Hwang E, 2018, NEURO-ONCOLOGY, V20, P100
   Jenkins NC, 2016, J NEURO-ONCOL, V126, P415, DOI 10.1007/s11060-015-1985-9
   Johnson DB, 2020, LANCET ONCOL, V21, pE398, DOI 10.1016/S1470-2045(20)30107-8
   Johnson DB, 2019, J IMMUNOTHER CANCER, V7, DOI 10.1186/s40425-019-0617-x
   Johnson DB, 2019, CLIN CANCER RES, V25, P1452, DOI 10.1158/1078-0432.CCR-18-3858
   Johnson DB, 2016, NEW ENGL J MED, V375, P1749, DOI 10.1056/NEJMoa1609214
   Ke XY, 2008, FRONT BIOSCI-LANDMRK, V13, P3986, DOI 10.2741/2986
   Kennedy LB, 2019, ONCOL THER, V7, P101, DOI 10.1007/s40487-019-0096-8
   Kodumudi KN, 2012, J IMMUNOL, V189, P5147, DOI 10.4049/jimmunol.1200274
   Koga T, 2020, NAT COMMUN, V11, DOI 10.1038/s41467-020-14312-1
   Koschmann C, 2016, ONCOTARGET, V7, P65696, DOI 10.18632/oncotarget.11602
   Kostine M, 2021, EUR J CANCER, V157, P474, DOI 10.1016/j.ejca.2021.08.036
   Krakowski ML, 1997, EUR J IMMUNOL, V27, P2840, DOI 10.1002/eji.1830271115
   Kreiter S, 2015, NATURE, V520, P692, DOI 10.1038/nature14426
   Krishnadas DK, 2015, J NEURO-ONCOL, V123, P193, DOI 10.1007/s11060-015-1765-6
   Kwon CH, 2008, CANCER RES, V68, P3286, DOI 10.1158/0008-5472.CAN-07-6867
   Lan XY, 2020, ONCOIMMUNOLOGY, V9, DOI 10.1080/2162402X.2020.1776577
   Larson JD, 2019, CANCER CELL, V35, P140, DOI 10.1016/j.ccell.2018.11.015
   Lee C, 2019, NAT COMMUN, V10, DOI 10.1038/s41467-018-08269-5
   Lee DW, 2015, LANCET, V385, P517, DOI 10.1016/S0140-6736(14)61403-3
   Lee Y, 2007, ONCOGENE, V26, P6442, DOI 10.1038/sj.onc.1210467
   Lenting K, 2017, ACTA NEUROPATHOL, V133, P263, DOI 10.1007/s00401-017-1671-4
   Liau LM, 2018, J TRANSL MED, V16, DOI 10.1186/s12967-018-1507-6
   Liau LM, 2005, CLIN CANCER RES, V11, P5515, DOI 10.1158/1078-0432.CCR-05-0464
   Locke FL, 2019, LANCET ONCOL, V20, P31, DOI 10.1016/S1470-2045(18)30864-7
   Lohr J, 2011, CLIN CANCER RES, V17, P4296, DOI 10.1158/1078-0432.CCR-10-2557
   Long AH, 2015, NAT MED, V21, P581, DOI 10.1038/nm.3838
   Louis DN, 2016, ACTA NEUROPATHOL, V131, P803, DOI 10.1007/s00401-016-1545-1
   Mackay A, 2017, CANCER CELL, V32, P520, DOI 10.1016/j.ccell.2017.08.017
   Majzner RG, 2019, CLIN CANCER RES, V25, P2560, DOI 10.1158/1078-0432.CCR-18-0432
   Margol AS, 2015, CLIN CANCER RES, V21, P1457, DOI 10.1158/1078-0432.CCR-14-1144
   Maude SL, 2014, NEW ENGL J MED, V371, P1507, DOI 10.1056/NEJMoa1407222
   Maxwell R, 2018, ONCOIMMUNOLOGY, V7, DOI 10.1080/2162402X.2018.1500108
   Merk DJ, 2018, DEV CELL, V44, P709, DOI 10.1016/j.devcel.2018.02.012
   Mestas J, 2004, J IMMUNOL, V172, P2731, DOI 10.4049/jimmunol.172.5.2731
   Misuraca KL, 2015, FRONT ONCOL, V5, DOI 10.3389/fonc.2015.00172
   Monje M, 2011, P NATL ACAD SCI USA, V108, P4453, DOI 10.1073/pnas.1101657108
   Morgan RA, 2013, J IMMUNOTHER, V36, P133, DOI 10.1097/CJI.0b013e3182829903
   Mount CW, 2018, NAT MED, V24, P572, DOI 10.1038/s41591-018-0006-x
   Mueller S, 2020, J CLIN INVEST, V130, P6325, DOI 10.1172/JCI140378
   Mukherjee J, 2006, CANCER BIOL THER, V5, P156, DOI 10.4161/cbt.5.2.2313
   Ng JMY, 2015, CANCER RES, V75, P4629, DOI 10.1158/0008-5472.CAN-15-0874
   Nguyen AT, 2015, NEUROPATH APPL NEURO, V41, P403, DOI 10.1111/nan.12196
   O'Rourke DM, 2017, SCI TRANSL MED, V9, DOI 10.1126/scitranslmed.aaa0984
   Ochs K, 2017, ONCOIMMUNOLOGY, V6, DOI 10.1080/2162402X.2017.1328340
   Ogawa J, 2018, CELL REP, V23, P1220, DOI 10.1016/j.celrep.2018.03.105
   Okada H, 2004, EXPERT OPIN BIOL TH, V4, P1609, DOI 10.1517/14712598.4.10.1609
   Okada H, 2001, HUM GENE THER, V12, P575
   Okada Hideho, 2015, Lancet Oncol, V16, pe534, DOI 10.1016/S1470-2045(15)00088-1
   Okada H, 2011, J CLIN ONCOL, V29, P330, DOI 10.1200/JCO.2010.30.7744
   Okada H, 2007, J TRANSL MED, V5, DOI 10.1186/1479-5876-5-67
   Okada H, 2009, CRIT REV IMMUNOL, V29, P1
   Ozawa T, 2018, CELL REP, V23, P3787, DOI 10.1016/j.celrep.2018.04.099
   Pajtler KW, 2019, NAT COMMUN, V10, DOI 10.1038/s41467-019-11884-5
   Pajtler KW, 2017, ACTA NEUROPATHOL, V133, P5, DOI 10.1007/s00401-016-1643-0
   Pan EY, 2020, J ONCOL PHARM PRACT, V26, P814, DOI 10.1177/1078155219872786
   Pathania M, 2017, CANCER CELL, V32, P684, DOI 10.1016/j.ccell.2017.09.014
   Patterson JD, 2020, FRONT ONCOL, V10, DOI 10.3389/fonc.2020.01582
   Paugh BS, 2013, CANCER RES, V73, P6219, DOI 10.1158/0008-5472.CAN-13-1491
   Pei YX, 2012, CANCER CELL, V21, P155, DOI 10.1016/j.ccr.2011.12.021
   Pham CD, 2016, CLIN CANCER RES, V22, P582, DOI 10.1158/1078-0432.CCR-15-0713
   Piccioni David E, 2019, CNS Oncol, V8, pCNS34, DOI 10.2217/cns-2018-0015
   Platten M, 2018, SEMIN NEUROL, V38, P62, DOI 10.1055/s-0037-1620274
   Pollack IF, 2016, J NEURO-ONCOL, V130, P517, DOI 10.1007/s11060-016-2245-3
   Pollack IF, 2014, J CLIN ONCOL, V32, P2050, DOI 10.1200/JCO.2013.54.0526
   POLLACK IF, 1994, NEW ENGL J MED, V331, P1500, DOI 10.1056/NEJM199412013312207
   Postow MA, 2018, NEW ENGL J MED, V378, P158, DOI [10.1056/NEJMra1703481, 10.1056/NEJMc1801663]
   Puputti M, 2006, MOL CANCER RES, V4, P927, DOI 10.1158/1541-7786.MCR-06-0085
   Puzanov I, 2017, J IMMUNOTHER CANCER, V5, DOI 10.1186/s40425-017-0300-z
   Regina A, 2015, MOL CANCER THER, V14, P129, DOI 10.1158/1535-7163.MCT-14-0399
   Reilly KM, 2000, NAT GENET, V26, P109, DOI 10.1038/79075
   Robinson G, 2012, NATURE, V488, P43, DOI 10.1038/nature11213
   Roth P, 2021, CANCER TREAT REV, V97, DOI 10.1016/j.ctrv.2021.102189
   Rozeman EA, 2019, NAT MED, V25, P879, DOI 10.1038/s41591-019-0482-7
   RUSSELL WL, 1979, P NATL ACAD SCI USA, V76, P5818, DOI 10.1073/pnas.76.11.5818
   Rutledge WC, 2013, CLIN CANCER RES, V19, P4951, DOI 10.1158/1078-0432.CCR-13-0551
   Schlager C, 2016, NATURE, V530, P349, DOI 10.1038/nature16939
   Schuller U, 2008, CANCER CELL, V14, P123, DOI 10.1016/j.ccr.2008.07.005
   Schuster SJ, 2019, NEW ENGL J MED, V380, P45, DOI 10.1056/NEJMoa1804980
   Schwalbe EC, 2017, LANCET ONCOL, V18, P958, DOI 10.1016/S1470-2045(17)30243-7
   Schwartzentruber J, 2012, NATURE, V482, P226, DOI [10.1038/nature10833, 10.1007/s13311-012-0137-6]
   Shi X, 2016, ONCOGENE, V35, P5746, DOI 10.1038/onc.2016.108
   Shiraishi R, 2021, CANCER SCI, V112, P2948, DOI 10.1111/cas.14990
   Smith KS, 2020, ACTA NEUROPATHOL, V140, P209, DOI 10.1007/s00401-020-02171-5
   Snyder A, 2014, NEW ENGL J MED, V371, P2189, DOI 10.1056/NEJMoa1406498
   Solomon DA, 2016, BRAIN PATHOL, V26, P569, DOI 10.1111/bpa.12336
   Sonabend AM, 2012, J NEURO-ONCOL, V108, P1, DOI 10.1007/s11060-011-0776-1
   Souweidane MM, 2018, LANCET ONCOL, V19, P1040, DOI 10.1016/S1470-2045(18)30322-X
   Stucklin ASG, 2019, NAT COMMUN, V10, DOI 10.1038/s41467-019-12187-5
   Sturm D, 2014, NAT REV CANCER, V14, P92, DOI 10.1038/nrc3655
   Suryadevara CM, 2018, ONCOIMMUNOLOGY, V7, DOI 10.1080/2162402X.2018.1434464
   Terada Y, 2019, CELL REP, V26, P2608, DOI 10.1016/j.celrep.2019.02.009
   Theruvath J, 2020, NAT MED, V26, P712, DOI 10.1038/s41591-020-0821-8
   Thompson JA, 2018, J NATL COMPR CANC NE, V16, P594, DOI 10.6004/jnccn.2018.0047
   Tocagen, 2019, TOC REP RES TOC 5 PH
   Uhrbom L, 1998, CANCER RES, V58, P5275
   Uziel T, 2005, GENE DEV, V19, P2656, DOI 10.1101/gad.1368605
   van der Burg SH, 2016, NAT REV CANCER, V16, P219, DOI 10.1038/nrc.2016.16
   Wainwright DA, 2012, CLIN CANCER RES, V18, P6110, DOI 10.1158/1078-0432.CCR-12-2130
   Walker PR, 2003, BRAIN RES REV, V42, P97, DOI 10.1016/S0165-0173(03)00141-3
   Watanabe K, 1997, CLIN CANCER RES, V3, P523
   Weber JS, 2017, J CLIN ONCOL, V35, P785, DOI 10.1200/JCO.2015.66.1389
   Weller M, 2017, LANCET ONCOL, V18, P1373, DOI 10.1016/S1470-2045(17)30517-X
   Wetmore C, 2001, CANCER RES, V61, P513
   Wheeler CJ, 2008, CANCER RES, V68, P5955, DOI 10.1158/0008-5472.CAN-07-5973
   Xing HL, 2014, BMC PLANT BIOL, V14, DOI 10.1186/s12870-014-0327-y
   Yamanaka R, 2005, CLIN CANCER RES, V11, P4160, DOI 10.1158/1078-0432.CCR-05-0120
   Yang ZJ, 2008, CANCER CELL, V14, P135, DOI 10.1016/j.ccr.2008.07.003
   Yerrabelli RS, 2021, EUR J NUCL MED MOL I, V48, P1166, DOI 10.1007/s00259-020-05050-z
   Yu JS, 2004, CANCER RES, V64, P4973, DOI 10.1158/0008-5472.CAN-03-3505
   Yu K, 2020, NATURE, V578, P166, DOI 10.1038/s41586-020-1952-2
   Yu LH, 2022, J CANCER RES CLIN, V148, P2643, DOI 10.1007/s00432-021-03839-5
   Yu LT, 2010, NEURO-ONCOLOGY, V12, P580, DOI 10.1093/neuonc/nop056
   Zhang LP, 2019, CANCER MED-US, V8, P3553, DOI 10.1002/cam4.2240
   Zhu X, 2017, J NEUROSURG-PEDIATR, V19, P249, DOI 10.3171/2016.8.PEDS16326
   Zhu Y, 2005, CANCER CELL, V8, P119, DOI 10.1016/j.ccr.2005.07.004
   Zook BC, 2000, TOXICOL PATHOL, V28, P193, DOI 10.1177/019262330002800124
NR 200
TC 14
Z9 14
U1 4
U2 16
PU NATURE PORTFOLIO
PI BERLIN
PA HEIDELBERGER PLATZ 3, BERLIN, 14197, GERMANY
EI 2662-1347
J9 NAT CANCER
JI Nat. Cancer
PD JAN
PY 2022
VL 3
IS 1
BP 11
EP 24
DI 10.1038/s43018-021-00319-0
PG 14
WC Oncology
WE Science Citation Index Expanded (SCI-EXPANDED)
SC Oncology
GA YK3JR
UT WOS:000745114100005
PM 35121998
DA 2023-08-27
ER

PT J
AU Santos, MM
   Timoteo, A
   Coiteiro, D
   Pimentel, J
AF Santos, Maria M.
   Timoteo, Angela
   Coiteiro, Domingos
   Pimentel, Jose
TI Schwannoma of the Fourth Ventricle: The Eighth Case Report
SO WORLD NEUROSURGERY
LA English
DT Article
DE Brain tumor; Fourth ventricle tumor; Intra-axial schwannoma
ID CYSTIC INTRAVENTRICULAR SCHWANNOMA; VENTRICLE
AB OBJECTIVE: We report an uncommon case of a surgical resection of a fourth ventricle tumor in an adult that proved to be a schwannoma.
   METHODS: A 53-year-old man presented with a 1.5-year history of gait unsteadiness and vertigo and a few-week history of headache, emesis, and neurogenic dysphagia. A brain magnetic resonance imaging revealed a large, heterogeneously contrast enhancing mass located within the fourth ventricle, compressing the brainstem and causing supratentorial ventricle enlargement. A suboccipital craniotomy and a telovelar approach were performed to resect the tumor. The ventricular system was repermeabilized at the end of the operation.
   RESULTS: A postoperative magnetic resonance imaging confirmed complete tumor removal. There was an initial worsening of the preoperative deficits, which progressively improved. The tumor was classified as a fourth ventricle schwannoma. There has been no evidence of tumor recurrence during the 6 years of follow-up. At present, the patient is ambulatory and reports an intermittent diplopia on conjugated gaze.
   CONCLUSION: This case report intends to reveal the eighth case of a fourth ventricle schwannoma since 1957. Schwannomas of the fourth ventricle are infrequent but should be accounted in the differential diagnosis of spaceoccupying lesions in this location. Gross total resection might be the definite treatment of these tumors if deemed possible.
C1 [Santos, Maria M.; Coiteiro, Domingos] Entidade Publ Empresarial, Ctr Hosp Lisboa Norte, Dept Neurol Surg, Lisbon, Portugal.
   [Pimentel, Jose] Entidade Publ Empresarial, Ctr Hosp Lisboa Norte, Dept Neuropathol, Lisbon, Portugal.
   [Timoteo, Angela] Hosp Fernando Fonseca, Dept Neurol, Amadora, Portugal.
C3 Universidade de Lisboa; Universidade de Lisboa; Hospital Professor
   Doutor Fernando Fonseca, EPE
RP Santos, MM (通讯作者)，Entidade Publ Empresarial, Ctr Hosp Lisboa Norte, Dept Neurol Surg, Lisbon, Portugal.
EM
CR ADELMAN LS, 1972, NEUROLOGY, V22, P726, DOI 10.1212/WNL.22.7.726
   Barbosa MD, 2001, NEUROCIRUGIA, V12, P56
   Benedict WJ, 2008, CHILD NERV SYST, V24, P529, DOI 10.1007/s00381-007-0556-6
   Benedikt M, 1874, VIRCHOWS ARCH PATHOL, V59, P395, DOI DOI 10.1007/BF01968394
   DAVID M, 1965, NEUROCHIRURGIE, V11, P578
   de Vasconcellos LP, 2009, ARQ NEURO-PSIQUIAT, V67, P1100, DOI 10.1590/S0004-282X2009000600027
   Dow GR, 2004, BRIT J NEUROSURG, V18, P561, DOI 10.1080/02688690400012632
   Erdogan E, 2003, MINIM INVAS NEUROSUR, V46, P50, DOI 10.1055/s-2003-37969
   Estrada Mastache J, 2002, REV MED IMSS, V40, P405
   Ghatak N R, 1975, Surg Neurol, V3, P45
   HERREGODTS P, 1991, J NEUROSURG, V74, P816, DOI 10.3171/jns.1991.74.5.0816
   Hodges TR, 2011, CASE REP MED, V2011, P1
   Jaimovich R, 2013, CHILD NERV SYST, V29, P499, DOI 10.1007/s00381-012-1977-4
   JUNG JM, 1995, J NEUROSURG, V82, P121, DOI 10.3171/jns.1995.82.1.0121
   Leveque M, 2007, NEUROCHIRURGIE, V53, P383, DOI 10.1016/j.neuchi.2007.06.005
   MARCHAND L, 1957, Ann Med Psychol (Paris), V115, P108
   MASON TH, 1968, J NEUROSURG, V29, P414, DOI 10.3171/jns.1968.29.4.0414
   Messing-Junger A Martina, 2006, Brain Pathol, V16, P87
   Oertel MF, 2009, CLIN NEUROL NEUROSUR, V111, P768, DOI 10.1016/j.clineuro.2009.06.009
   OST AH, 1990, AM J NEURORADIOL, V11, P1262
   PIMENTEL J, 1988, CHILD NERV SYST, V4, P373, DOI 10.1007/BF00270615
   RAMAMURTHI B, 1958, J NEUROL NEUROSUR PS, V21, P92, DOI 10.1136/jnnp.21.2.92
   REDEKOP G, 1990, J NEUROSURG, V73, P777, DOI 10.3171/jns.1990.73.5.0777
   RIGGS HE, 1957, J NEUROPATH EXP NEUR, V16, P332, DOI 10.1097/00005072-195707000-00004
   Russel D. S., 1977, PATHOLOGY TUMOURS NE
   SHALIT MN, 1982, ACTA NEUROCHIR, V64, P253, DOI 10.1007/BF01406060
   Stohr P, 1922, U Z GESAMTE ANAT, V63, P562
   VANRENSBURG MJ, 1975, J NEUROL NEUROSUR PS, V38, P703
   WEINER HL, 1993, J NEURO-ONCOL, V15, P37, DOI 10.1007/BF01050261
NR 29
TC 5
Z9 5
U1 0
U2 2
PU ELSEVIER SCIENCE INC
PI NEW YORK
PA 360 PARK AVE SOUTH, NEW YORK, NY 10010-1710 USA
SN 1878-8750
EI 1878-8769
J9 WORLD NEUROSURG
JI World Neurosurg.
PD NOV
PY 2015
VL 84
IS 5
AR 1493.e9
DI 10.1016/j.wneu.2015.04.036
PG 5
WC Clinical Neurology; Surgery
WE Science Citation Index Expanded (SCI-EXPANDED)
SC Neurosciences & Neurology; Surgery
GA CV8KM
UT WOS:000364533900051
PM 25920574
DA 2023-08-27
ER

PT J
AU Kaminska, E
   Obolonczyk, L
   Kmiec, P
   Czapiewski, P
   Babinska, A
   Sworczak, K
AF Kaminska, E.
   Obolonczyk, L.
   Kmiec, P.
   Czapiewski, P.
   Babinska, A.
   Sworczak, K.
TI REPORT ON 5 CASES OF ADRENAL GANGLIONEUROMAS
SO ACTA ENDOCRINOLOGICA-BUCHAREST
LA English
DT Article
DE ganglioneuroma; adrenal tumor; adult; catecholamine hypersecretion
ID INCIDENTALOMAS; FEATURES
AB Background. Ganglioneuroma (GN) is a rare, benign, neuroblastic tumor. It may arise from the sympathetic plexus or adrenal medulla. Its most common location is the posterior mediastinum (41.5%), while in approximately 21% of patients the tumor is located in the adrenal gland. Median age of adults at diagnosis is approximately 40 to 50 years of age. Since GN is most commonly asymptomatic, the diagnosis is usually accidental, i.e. when radiological examinations are performed due to other indications. This slow-growing tumor can reach a large size. In a small percentage of patients the tumor secretes catecholamines.
   We present five ganglioneuroma patients, who were hospitalized in our Department from 1993 through 2012. Their mean age was 31 years. In four cases the tumor was located in the adrenal gland and in one - extra-adrenally, which was revealed during surgery. In three out of five presented cases pathological hormone secretion by the tumor was confirmed in laboratory results.
C1 [Kaminska, E.; Obolonczyk, L.; Kmiec, P.; Babinska, A.; Sworczak, K.] Med Univ Gdansk, Dept Endocrinol & Internal Med, PL-80952 Gdansk, Poland.
   [Czapiewski, P.] Med Univ Gdansk, Dept Pathomorphol, PL-80952 Gdansk, Poland.
C3 Fahrenheit Universities; Medical University Gdansk; Fahrenheit
   Universities; Medical University Gdansk
RP Kmiec, P (通讯作者)，Med Univ Gdansk, Dept Endocrinol & Internal Med, 7 Debinki St, PL-80952 Gdansk, Poland.
EM
RI Czapiewski, Piotr/J-7949-2012; Kmieć, Piotr/AAB-6724-2020; Babińska,
   Anna/AAL-6714-2021
OI Babińska, Anna/0000-0003-2028-5878
CR Babinska A, 2012, ARCH MED SCI, V8, P97, DOI 10.5114/aoms.2012.27288
   Babinska Anna, 2006, Wiad Lek, V59, P744
   Barzon L, 2003, EUR J ENDOCRINOL, V149, P273, DOI 10.1530/eje.0.1490273
   Barzon L, 1999, J CLIN ENDOCR METAB, V84, P520, DOI 10.1210/jc.84.2.520
   BOVE KE, 1981, ARCH PATHOL LAB MED, V105, P325
   Erem C, 2014, ACTA ENDOCRINOL-BUCH, V10, P140, DOI 10.4183/aeb.2014.140
   Geoerger B, 2001, CANCER, V91, P1905, DOI 10.1002/1097-0142(20010515)91:10<1905::AID-CNCR1213>3.0.CO;2-4
   Gilshtein H, 2012, CASE REP ONCOL, V5, P487, DOI 10.1159/000342445
   Ichikawa T, 1996, BRIT J RADIOL, V69, P114, DOI 10.1259/0007-1285-69-818-114
   Jain M, 1999, DIAGN CYTOPATHOL, V21, P194, DOI 10.1002/(SICI)1097-0339(199909)21:3<194::AID-DC9>3.0.CO;2-B
   Joshi VV, 2000, PEDIATR DEVEL PATHOL, V3, P184, DOI 10.1007/s100240050024
   Jung HR, 2011, J KOREAN SURG SOC, V80, P297, DOI 10.4174/jkss.2011.80.4.297
   Kanthan R, 2012, WORLD J SURG ONCOL, V10, DOI 10.1186/1477-7819-10-64
   Linos D, 2011, SURGERY, V149, P99, DOI 10.1016/j.surg.2010.03.016
   LUCAS K, 1994, MED PEDIATR ONCOL, V22, P240, DOI 10.1002/mpo.2950220405
   Maweja S, 2007, ACTA CHIR BELG, V107, P670, DOI 10.1080/00015458.2007.11680144
   Sasaki S, 2012, INTERNAL MED, V51, P2365, DOI 10.2169/internalmedicine.51.7726
   Srinivasan R, 2007, Indian J Pathol Microbiol, V50, P32
   Sucandy I, 2011, N AM J MED SCI, V3, P336, DOI 10.4297/najms.2011.3336
   Tarantino RM, 2012, ARQ BRAS ENDOCRINOL, V56, P270, DOI 10.1590/S0004-27302012000400009
   Yang AI, 2013, J NEUROIMAGING, V23, P537, DOI 10.1111/j.1552-6569.2012.00765.x
   Yang Q, 2010, SURGERY, V147, P854, DOI 10.1016/j.surg.2009.11.010
NR 22
TC 1
Z9 1
U1 0
U2 2
PU EDITURA ACAD ROMANE
PI BUCURESTI
PA CALEA 13 SEPTEMBRIE NR 13, SECTOR 5, BUCURESTI 050711, ROMANIA
SN 1841-0987
EI 1843-066X
J9 ACTA ENDOCRINOL-BUCH
JI Acta Endocrinol.
PD JUL-SEP
PY 2014
VL 10
IS 3
BP 477
EP 486
DI 10.4183/aeb.2014.477
PG 10
WC Endocrinology & Metabolism
WE Science Citation Index Expanded (SCI-EXPANDED)
SC Endocrinology & Metabolism
GA AM0KY
UT WOS:000339535700016
DA 2023-08-27
ER

PT J
AU LAHR, G
   MAYERHOFER, A
   BUCHER, S
   BARTHELS, D
   WILLE, W
   GRATZL, M
AF LAHR, G
   MAYERHOFER, A
   BUCHER, S
   BARTHELS, D
   WILLE, W
   GRATZL, M
TI NEURAL CELL-ADHESION MOLECULES IN RAT ENDOCRINE TISSUES AND TUMOR-CELLS
   - DISTRIBUTION AND MOLECULAR ANALYSIS
SO ENDOCRINOLOGY
LA English
DT Article
ID MEMBRANE-SPANNING REGION; COMPLETE CODING SEQUENCE; NERVE GROWTH-FACTOR;
   MOUSE N-CAM; MONOCLONAL-ANTIBODIES; POLYSIALIC ACID; IMMUNOGLOBULIN
   SUPERFAMILY; EXTRACELLULAR DOMAIN; NCAM POLYPEPTIDE; 180-KD COMPONENT
AB The adhesive properties of neural cell adhesion molecules (NCAMs) can be modified by alternative splicing of the primary transcript or posttranslational modifications. In the present study, we describe distinct forms of alternative splicing and posttranslational modification of the extracellular domain of NCAM of various endocrine tissues and derived tumor cells of the rat. Using an antiserum detecting the immunoglobulin-like domains of NCAM as well as a monoclonal antibody recognizing the NCAM-specific polysialic acid (PSA), we observed a similar staining pattern in adrenals, pituitary, and neoplastic endocrine cells. In endocrine tumor cells [pheochromocytoma (PC12), insulinoma (RINA2), and pituitary tumor cells (GH3)], NCAM immunoreactivity was most intense at contact sites between the cells. The immunocytochemical data were substantiated by results of in situ hybridization histochemistry. Specifically, higher levels of NCAM mRNA were detected in the adrenal cortex than in the medulla. In the pituitary, NCAM mRNA was more abundant in the anterior and intermediate lobes than in the neural lobe. The sequence of NCAM mRNAs in endocrine cells was analyzed by polymerase chain reaction and S1 nuclease protection assays. We found that major exons 4-13 of the NCAM mRNA in endocrine tissues and related tumor cell lines were homologous to those in the brain. However, PC12, RINA2, and GH3 tumor cells; normal rat pituitaries; and adrenals contained different amounts of NCAM mRNA with an alternative extra exon, termed VASE (also called pi in mouse) between constitutive exons 7 and 8. In addition, in pituitaries, we detected an alternative exon in splice site a between the constitutive exons 12 and 13, termed a15 with or without an AAG triplett. These sites are thought to be important for the adhesive properties of NCAM. Therefore, these results suggest that modifications of NCAM may be important for adhesive interactions in normal and neoplastic endocrine cells.
C1 UNIV COLOGNE,INST GENET,W-5000 COLOGNE,GERMANY.
C3 University of Cologne
RP LAHR, G (通讯作者)，UNIV ULM,ANAT & ZELLBIOL ABT,ALBERT EINSTEIN ALLEE 11,POSTFACH 4066,W-7900 ULM,GERMANY.
OI Mayerhofer, Artur/0000-0002-9388-4639
CR ALETSEEUFRECHT MC, 1990, FEBS LETT, V267, P295, DOI 10.1016/0014-5793(90)80948-I
   ALETSEEUFRECHT MC, 1990, FEBS LETT, V272, P45, DOI 10.1016/0014-5793(90)80445-O
   [Anonymous], 1989, MOL CLONING LABORATO
   BARBAS JA, 1988, EMBO J, V7, P625, DOI 10.1002/j.1460-2075.1988.tb02856.x
   BARTHELS D, 1992, EUR J NEUROSCI, V4, P327, DOI 10.1111/j.1460-9568.1992.tb00880.x
   BARTHELS D, 1987, EMBO J, V6, P907, DOI 10.1002/j.1460-2075.1987.tb04837.x
   BITTERSUERMANN D, 1987, IMMUNOL RES, V6, P225, DOI 10.1007/BF02935517
   CHEN AS, 1990, MOL CELL BIOL, V10, P3314, DOI 10.1128/MCB.10.7.3314
   DICKSON G, 1987, CELL, V50, P1119, DOI 10.1016/0092-8674(87)90178-4
   DOHERTY P, 1992, NATURE, V356, P791, DOI 10.1038/356791a0
   DOHERTY P, 1990, NEURON, V5, P209, DOI 10.1016/0896-6273(90)90310-C
   EDELMAN GM, 1988, BIOCHEMISTRY-US, V27, P3533, DOI 10.1021/bi00410a001
   EDELMAN GM, 1982, P NATL ACAD SCI-BIOL, V79, P7036, DOI 10.1073/pnas.79.22.7036
   EDELMAN GM, 1985, ANNU REV BIOCHEM, V54, P135, DOI 10.1146/annurev.biochem.54.1.135
   FINNE J, 1983, BIOCHEM BIOPH RES CO, V112, P482, DOI 10.1016/0006-291X(83)91490-0
   FRAWLEY LS, 1991, ENDOCR REV, V12, P337, DOI 10.1210/edrv-12-4-337
   FROSCH M, 1985, P NATL ACAD SCI USA, V82, P1194, DOI 10.1073/pnas.82.4.1194
   GORIDIS C, 1985, EMBO J, V4, P631, DOI 10.1002/j.1460-2075.1985.tb03676.x
   GREENE LA, 1976, P NATL ACAD SCI USA, V73, P2424, DOI 10.1073/pnas.73.7.2424
   HAMSHERE M, 1991, NUCLEIC ACIDS RES, V19, P4709, DOI 10.1093/nar/19.17.4709
   HE HT, 1987, J CELL BIOL, V105, P2489, DOI 10.1083/jcb.105.6.2489
   HE HT, 1986, EMBO J, V5, P2489, DOI 10.1002/j.1460-2075.1986.tb04526.x
   HEMPERLY JJ, 1986, P NATL ACAD SCI USA, V83, P9822, DOI 10.1073/pnas.83.24.9822
   HIRSCH MR, 1990, MOL CELL BIOL, V10, P1959, DOI 10.1128/MCB.10.5.1959
   HOFFMAN S, 1983, P NATL ACAD SCI-BIOL, V80, P5762, DOI 10.1073/pnas.80.18.5762
   HSU SM, 1981, J HISTOCHEM CYTOCHEM, V29, P577, DOI 10.1177/29.4.6166661
   KAPLAN B, 1976, BIOCHEM J, V183, P181
   LAHR G, 1990, NEUROSCIENCE, V39, P605, DOI 10.1016/0306-4522(90)90245-Y
   LANDMESSER L, 1990, NEURON, V4, P655, DOI 10.1016/0896-6273(90)90193-J
   LANGLEY K, 1990, MARKERS NEURAL ENDOC, P133
   LANGLEY OK, 1987, FEBS LETT, V220, P108, DOI 10.1016/0014-5793(87)80885-2
   LANGLEY OK, 1989, J HISTOCHEM CYTOCHEM, V37, P781, DOI 10.1177/37.6.2723399
   MOOS M, 1988, NATURE, V334, P701, DOI 10.1038/334701a0
   MURRAY BA, 1986, J CELL BIOL, V103, P1431, DOI 10.1083/jcb.103.4.1431
   NUSSDORFER GG, 1986, INT REV CYTOL, V98, P319
   NYBROE O, 1986, NEUROCHEM INT, V9, P539, DOI 10.1016/0197-0186(86)90147-6
   OWENS GC, 1987, P NATL ACAD SCI USA, V84, P294, DOI 10.1073/pnas.84.1.294
   POLLERBERG EG, 1985, J CELL BIOL, V101, P1921, DOI 10.1083/jcb.101.5.1921
   POLLERBERG GE, 1987, CELL TISSUE RES, V250, P227, DOI 10.1007/BF00214676
   POLTORAK M, 1990, EXP NEUROL, V110, P52, DOI 10.1016/0014-4886(90)90051-S
   POWELL SK, 1991, NATURE, V353, P76, DOI 10.1038/353076a0
   PREDIGER EA, 1988, P NATL ACAD SCI USA, V85, P9616, DOI 10.1073/pnas.85.24.9616
   PRENTICE HM, 1987, EMBO J, V6, P1859, DOI 10.1002/j.1460-2075.1987.tb02444.x
   REGAN CM, 1991, INT J BIOCHEM, V23, P513
   REYES AA, 1991, MOL CELL BIOL, V11, P1654, DOI 10.1128/MCB.11.3.1654
   ROUGON G, 1986, J BIOL CHEM, V261, P3396
   ROUGON G, 1986, J CELL BIOL, V103, P2429, DOI 10.1083/jcb.103.6.2429
   RUPPERT C, 1986, EMBO J, V5, P1897, DOI 10.1002/j.1460-2075.1986.tb04442.x
   RUTISHAUSER U, 1988, PHYSIOL REV, V68, P819, DOI 10.1152/physrev.1988.68.3.819
   RUTISHAUSER U, 1988, SCIENCE, V240, P53, DOI 10.1126/science.3281256
   SAIKI RK, 1988, SCIENCE, V239, P489
   SANTONI MJ, 1989, EMBO J, V8, P385, DOI 10.1002/j.1460-2075.1989.tb03389.x
   SMALL SJ, 1987, J CELL BIOL, V105, P2335, DOI 10.1083/jcb.105.5.2335
   SMALL SJ, 1990, J CELL BIOL, V111, P2089, DOI 10.1083/jcb.111.5.2089
   THEODOSIS DT, 1991, P NATL ACAD SCI USA, V88, P5494, DOI 10.1073/pnas.88.13.5494
   THOMPSON J, 1989, GENE DEV, V3, P348, DOI 10.1101/gad.3.3.348
   WILLIAMS AF, 1988, ANNU REV IMMUNOL, V6, P381, DOI 10.1146/annurev.iy.06.040188.002121
   YERHOFER A, 1991, ENDOCRINOLOGY, V129, P792
   YOKORO K, 1961, CANCER RES, V21, P178
NR 59
TC 41
Z9 41
U1 0
U2 0
PU ENDOCRINE SOC
PI BETHESDA
PA 4350 EAST WEST HIGHWAY SUITE 500, BETHESDA, MD 20814-4110
SN 0013-7227
J9 ENDOCRINOLOGY
JI Endocrinology
PD MAR
PY 1993
VL 132
IS 3
BP 1207
EP 1217
DI 10.1210/en.132.3.1207
PG 11
WC Endocrinology & Metabolism
WE Science Citation Index Expanded (SCI-EXPANDED)
SC Endocrinology & Metabolism
GA KP290
UT WOS:A1993KP29000038
PM 8440182
OA Green Published
DA 2023-08-27
ER

PT J
AU Cook, RL
   Householder, KT
   Chung, EP
   Prakapenka, AV
   DiPerna, DM
   Sirianni, RW
AF Cook, Rebecca L.
   Householder, Kyle T.
   Chung, Eugene P.
   Prakapenka, Alesia V.
   DiPerna, Danielle M.
   Sirianni, Rachael W.
TI A critical evaluation of drug delivery fromligandmodified nanoparticles:
   Confounding small molecule distribution and efficacy in the central
   nervous system
SO JOURNAL OF CONTROLLED RELEASE
LA English
DT Article
DE Brain; Chemotherapy; Drug delivery; Nanoparticle; Peptide; Spinal cord
ID LOADED PLGA NANOPARTICLES; TARGETING GENE DELIVERY; SPINAL-CORD;
   MOUSE-BRAIN; NANOCARRIERS; PROTEINS; LIGAND; RVG
AB In this work, we sought to test how surface modification of poly(lactic-co-glycolic acid) (PLGA) nanoparticles with peptide ligand alters the brain specific delivery of encapsulated molecules. For biodistribution studies, nanoparticles modified with rabies virus glycoprotein (RVG29) were loaded with small molecule drug surrogates and administered to healthy mice by lateral tail vein injection. Mice were perfused 2 h after injection and major anatomical regions of the CNS were dissected (striatum, midbrain, cerebellum, hippocampus, cortex, olfactory bulb, brainstem, and cervical, thoracic, lumbar and sacral spinal cord). For functional studies, surface modified nanoparticles were loaded with the chemotherapeutic camptothecin (CPT) and administered to mice bearing intracranial GL261-Luc2 gliomas. Outcome measures included tumor growth, as measured by bioluminescent imaging, and median survival time. We observed that small molecule delivery from PLGA nanoparticles varied by as much as 150% for different tissue regions within the CNS. These differences were directly correlated to regional differences in cerebral blood volume. Although the presence of RVG29 enhanced apparent brain delivery for multiple small molecule payloads, we observed minimal evidence for targeting to muscle or spinal cord, which are the known sites for rabies virus entry into the CNS, and enhancements in brain delivery were not prolonged due to an apparent aqueous instability of the RVG29 ligand. Furthermore, we have identified concerning differences in apparent delivery kinetics as measured by different payloads: nanoparticle encapsulated DiR was observed to accumulate in the brain, whereas encapsulated Nile red was rapidly cleared. Although systemically administered CPT loaded nanoparticles slowed the growth of orthotopic brain tumors to prolong survival, the presence of RVG29 did not enhance therapeutic efficacy compared to control nanoparticles. These data are consistent with a model of delivery of hydrophobic small molecules to the brain that does not rely on internalization of polymer nanoparticles in target tissue. We discuss an important risk for discordance between biodistribution, as typically measured by drug surrogate, and therapeutic outcome, as determined by clinically relevant measurement of drug function in a disease model. These results pose critical considerations for the methods used to design and evaluate targeted drug delivery systems in vivo. (C) 2015 Elsevier B. V. All rights reserved.
C1 [Cook, Rebecca L.; Householder, Kyle T.; Chung, Eugene P.; Prakapenka, Alesia V.; DiPerna, Danielle M.; Sirianni, Rachael W.] Barrow Neurol Inst, Barrow Brain Tumor Res Ctr, Phoenix, AZ 85013 USA.
   [Cook, Rebecca L.; Householder, Kyle T.; Sirianni, Rachael W.] Arizona State Univ, Sch Biol & Hlth Syst Engn, Ira A Fulton Sch Engn, Tempe, AZ 85287 USA.
   [Chung, Eugene P.] Arizona State Univ, Sch Life Sci, Tempe, AZ 85287 USA.
   [Prakapenka, Alesia V.; Sirianni, Rachael W.] Arizona State Univ, Sch Life Sci, Interdisciplinary Grad Program Neurosci, Tempe, AZ 85287 USA.
C3 Barrow Neurological Institute; Arizona State University; Arizona State
   University-Tempe; Arizona State University; Arizona State
   University-Tempe; Arizona State University; Arizona State
   University-Tempe
RP Sirianni, RW (通讯作者)，St Josephs Hosp, Ctr Res Neurosci, Room 436,350 W Thomas Rd, Phoenix, AZ 85013 USA.
EM;
  ;
  
RI Cook, Rebecca/HPG-5035-2023
OI Sirianni, Rachael/0000-0002-9196-0239; Householder,
   Kyle/0000-0001-9234-2021; Cook, Rebecca/0000-0001-9009-9937; Prakapenka,
   Alesia/0000-0002-4930-1141
FU Ben and Catherine Ivy Foundation; Barrow Neurological Foundation; U.S.
   Department of Defense [W81XWH-14-1-0311]; National Science Foundation
   [DGE-1311230]; National Institutes of Health [R01-CA149128,
   T32-DA022975]
FX The authors gratefully acknowledge funding from the Ben and Catherine
   Ivy Foundation, the Barrow Neurological Foundation, the U.S. Department
   of Defense (W81XWH-14-1-0311), the National Science Foundation
   (DGE-1311230) and the National Institutes of Health (R01-CA149128,
   T32-DA022975). We also thank Dr. Adrienne C. Scheck at Barrow
   Neurological Institute for providing GL261-Luc2 cells.
CR Abdelwahab MG, 2011, JOVE-J VIS EXP, DOI 10.3791/3403
   Alam MI, 2010, EUR J PHARM SCI, V40, P385, DOI 10.1016/j.ejps.2010.05.003
   Alvarez-Erviti L, 2011, NAT BIOTECHNOL, V29, P341, DOI 10.1038/nbt.1807
   Bartanusz V, 2011, ANN NEUROL, V70, P194, DOI 10.1002/ana.22421
   Bastiat G, 2013, J CONTROL RELEASE, V170, P334, DOI 10.1016/j.jconrel.2013.06.014
   Bertrand N, 2014, ADV DRUG DELIVER REV, V66, P2, DOI 10.1016/j.addr.2013.11.009
   BOWERY NG, 1987, NEUROSCIENCE, V20, P365, DOI 10.1016/0306-4522(87)90098-4
   Calvo P, 2002, EUR J NEUROSCI, V15, P1317, DOI 10.1046/j.1460-9568.2002.01967.x
   Chen HT, 2008, P NATL ACAD SCI USA, V105, P6596, DOI 10.1073/pnas.0707046105
   Chen W, 2011, J DRUG TARGET, V19, P228, DOI 10.3109/1061186X.2010.492523
   Chugh BP, 2009, NEUROIMAGE, V47, P1312, DOI 10.1016/j.neuroimage.2009.03.083
   Chvatal SA, 2008, BIOMATERIALS, V29, P1967, DOI 10.1016/j.biomaterials.2008.01.002
   Dora CL, 2006, J PHARM PHARM SCI, V9, P22
   Fahmy TM, 2005, BIOMATERIALS, V26, P5727, DOI 10.1016/j.biomaterials.2005.02.025
   Fu AL, 2012, PHARM RES-DORDR, V29, P1562, DOI 10.1007/s11095-012-0667-y
   Gaudin A, 2014, NAT NANOTECHNOL, V9, P1054, DOI [10.1038/NNANO.2014.274, 10.1038/nnano.2014.274]
   Gullotti E, 2012, J CONTROL RELEASE, V164, P170, DOI 10.1016/j.jconrel.2012.04.042
   Householder KT, 2015, INT J PHARMACEUT, V479, P374, DOI 10.1016/j.ijpharm.2015.01.002
   Hwang DW, 2011, BIOMATERIALS, V32, P4968, DOI 10.1016/j.biomaterials.2011.03.047
   Jacobs VL, 2011, ASN NEURO, V3, DOI 10.1042/AN20110014
   Kim JY, 2013, BIOMATERIALS, V34, P1170, DOI 10.1016/j.biomaterials.2012.09.047
   Kreuter J, 2014, ADV DRUG DELIVER REV, V71, P2, DOI 10.1016/j.addr.2013.08.008
   Kumar P, 2007, NATURE, V448, P39, DOI 10.1038/nature05901
   Lafon M, 2005, J NEUROVIROL, V11, P82, DOI 10.1080/13550280590900427
   Langevin C, 2002, J BIOL CHEM, V277, P37655, DOI 10.1074/jbc.M201374200
   Li MH, 2014, J NANOSCI NANOTECHNO, V14, P415, DOI 10.1166/jnn.2014.8757
   Liu Y, 2013, PLOS ONE, V8, DOI 10.1371/journal.pone.0062905
   Liu Y, 2012, J CONTROL RELEASE, V163, P203, DOI 10.1016/j.jconrel.2012.09.001
   Liu Y, 2009, BIOMATERIALS, V30, P4195, DOI 10.1016/j.biomaterials.2009.02.051
   McCall RL, 2014, TISSUE BARRIERS, V2, DOI 10.4161/21688362.2014.944449
   McCall Rebecca L, 2013, J Vis Exp, P51015, DOI 10.3791/51015
   McCarron PA, 2008, BIOCONJUGATE CHEM, V19, P1561, DOI 10.1021/bc800057g
   MEIKLE ADS, 1981, STAIN TECHNOL, V56, P235, DOI 10.3109/10520298109067317
   Patel Toral R, 2014, CNS Oncol, V3, P115, DOI 10.2217/cns.14.2
   Prabhakar U, 2013, CANCER RES, V73, P2412, DOI 10.1158/0008-5472.CAN-12-4561
   Saxena T, 2015, ACS NANO, V9, P1492, DOI 10.1021/nn505980z
   Sempf K, 2013, EUR J PHARM BIOPHARM, V85, P53, DOI 10.1016/j.ejpb.2012.11.030
   Sirianni RW, 2014, BIOCONJUGATE CHEM, V25, P2157, DOI 10.1021/bc500315j
   Son S, 2011, J CONTROL RELEASE, V155, P18, DOI 10.1016/j.jconrel.2010.08.011
   Tao YH, 2012, J MATER CHEM, V22, P11808, DOI 10.1039/c2jm31675g
   Tortorella S, 2014, CURR DRUG DELIV, V11, P427, DOI 10.2174/1567201810666140106115436
   Ulukan H, 2002, DRUGS, V62, P2039, DOI 10.2165/00003495-200262140-00004
   van der Meel R, 2013, ADV DRUG DELIVER REV, V65, P1284, DOI 10.1016/j.addr.2013.08.012
   Wang YC, 2008, BIOMATERIALS, V29, P4546, DOI 10.1016/j.biomaterials.2008.07.050
   Xiang LX, 2011, J DRUG TARGET, V19, P632, DOI 10.3109/1061186X.2010.523788
   Xu PS, 2009, MOL PHARMACEUT, V6, P190, DOI 10.1021/mp800137z
   Zadran S, 2013, NEUROMOL MED, V15, P74, DOI 10.1007/s12017-012-8196-8
   Zhou JB, 2013, P NATL ACAD SCI USA, V110, P11751, DOI 10.1073/pnas.1304504110
NR 48
TC 51
Z9 51
U1 2
U2 57
PU ELSEVIER SCIENCE BV
PI AMSTERDAM
PA PO BOX 211, 1000 AE AMSTERDAM, NETHERLANDS
SN 0168-3659
EI 1873-4995
J9 J CONTROL RELEASE
JI J. Control. Release
PD DEC 28
PY 2015
VL 220
BP 89
EP 97
DI 10.1016/j.jconrel.2015.10.013
PN A
PG 9
WC Chemistry, Multidisciplinary; Pharmacology & Pharmacy
WE Science Citation Index Expanded (SCI-EXPANDED)
SC Chemistry; Pharmacology & Pharmacy
GA CY0TN
UT WOS:000366119100011
PM 26471392
OA Green Accepted, hybrid
DA 2023-08-27
ER

PT J
AU Gupta, T
   Sarin, R
   Jalali, R
   Sharma, S
   Kurkure, P
   Goel, A
AF Gupta, Tejpal
   Sarin, Rajiv
   Jalali, Rakesh
   Sharma, Suash
   Kurkure, Purna
   Goel, Atul
TI A pragmatic clinicopathobiological grouping/staging system for gliomas:
   Proposal of the Indian TNM subcommittee on brain tumors
SO NEUROLOGY INDIA
LA English
DT Article
DE Gliomas; prognosis; staging classification
ID LOW-GRADE GLIOMA; PROGNOSTIC-FACTORS; EUROPEAN ORGANIZATION;
   RADIATION-THERAPY; RANDOMIZED-TRIAL; SURVIVAL; RADIOTHERAPY; CHILDREN;
   CHEMOTHERAPY; MANAGEMENT
AB Background: There is no universally accepted staging system for primary brain tumors wherein prognostication is mainly based on complex composite indices . Aim: To develop a simple, pragmatic, and widely applicable grouping/staging system for gliomas, the most common primary brain tumor. Materials and Methods: An expert neurooncology panel with representation from radiation oncology, neurosurgery, pathology, radiology, and medical oncology had several rounds of discussion on issues pertinent to brain tumor staging. The trade off was between the accuracy of prognostic categorization and a pragmatic, widely applicable approach. Results and Recommendations: The Tumor-Node-Metastasis staging was considered irrelevant for gliomas that seldom metastasize to lymphatics or outside the neuraxis. Instead, a 4-point staging/grouping system is proposed, using histological grade as the main prognostic variable and at least one stage migration based on other unfavorable features such as tumor location (brainstem); age ( 5 years for all grades, > 50 years for high-grade, and > 40 years for low-grade gliomas); poor neurological performance status (NPS 2-4); multicentricity and/or gliomatosis; and adverse biological parameters (proliferative index, angiogenesis markers, apoptotic index, cytogenetic abnormalities, and molecular markers). Conclusion: In absence of a grouping/staging system for primary brain tumors, prognostification is mostly based on complex composite indices. The proposed clinicopathobiological grouping/staging system for gliomas is a simple, pragmatic, and user-friendly tool with a potential to fulfill the objectives of staging classification.
C1 [Gupta, Tejpal; Sarin, Rajiv; Jalali, Rakesh] ACTREC TMH, Dept Radiat Oncol, Bombay, Maharashtra, India.
   [Sharma, Suash] Tata Mem Hosp, Dept Pathol, TMH, Bombay, Maharashtra, India.
   [Kurkure, Purna] Tata Mem Hosp, Dept Pediat Oncol, TMH, Bombay, Maharashtra, India.
   [Goel, Atul] King Edward Mem Hosp, Dept Neurosurg, Bombay, Maharashtra, India.
C3 Tata Memorial Centre (TMC); Advance Centre for Treatment, Research &
   Education in Cancer (ACTREC); Tata Memorial Centre (TMC); Tata Memorial
   Hospital; Tata Memorial Centre (TMC); Tata Memorial Hospital; Seth
   Gordhandas Sunderdas Medical College & King Edward Memorial Hospital
RP Sarin, R (通讯作者)，Adv Ctr Treatment Res & Educ Canc, Tata Mem Ctr, Kharghar 410210, Navi Mumbai, India.
EM
OI Sarin, Rajiv/0000-0002-6405-8282
CR ABRAMSON DH, 1982, CA-CANCER J CLIN, V32, P130, DOI 10.3322/canjclin.32.3.130
   [Anonymous], 2007, CANC FACTS FIG 2007
   [Anonymous], GLOBOCAN 2002 CANC I
   Bauman G, 1999, INT J RADIAT ONCOL, V45, P923, DOI 10.1016/S0360-3016(99)00284-9
   Claus EB, 2006, CANCER-AM CANCER SOC, V106, P1358, DOI 10.1002/cncr.21733
   CURRAN WJ, 1993, J NATL CANCER I, V85, P704, DOI 10.1093/jnci/85.9.704
   Duffner P K, 1999, Neuro Oncol, V1, P152, DOI 10.1093/neuonc/1.2.152
   Dufour C, 2006, EUR J CANCER, V42, P2939, DOI 10.1016/j.ejca.2006.06.021
   Fouladi M, 2005, J CLIN ONCOL, V23, P7152, DOI 10.1200/JCO.2005.01.214
   Gospodarowicz MK, 2004, CANCER, V100, P1, DOI 10.1002/cncr.11898
   Grundy RG, 2007, LANCET ONCOL, V8, P696, DOI 10.1016/S1470-2045(07)70208-5
   Gupta T, 2002, LANCET ONCOL, V3, P557, DOI 10.1016/S1470-2045(02)00853-7
   Hermanek P., 1987, TNM CLASSIFICATION M
   Karim ABMF, 1996, INT J RADIAT ONCOL, V36, P549, DOI 10.1016/S0360-3016(96)00352-5
   Karim ABMF, 2002, INT J RADIAT ONCOL, V52, P316, DOI 10.1016/S0360-3016(01)02692-X
   Lamborn KR, 2004, NEURO-ONCOLOGY, V6, P227, DOI 10.1215/S1152851703000620
   Louis DN, 2007, ACTA NEUROPATHOL, V114, P547, DOI 10.1007/s00401-007-0278-6
   *MRC BRAIN TUM WOR, 1990, J NEUROONCOL, V9, P47
   Pignatti F, 2002, J CLIN ONCOL, V20, P2076, DOI 10.1200/JCO.2002.08.121
   Raney RB, 2001, J PEDIAT HEMATOL ONC, V23, P215, DOI 10.1097/00043426-200105000-00008
   Sobin L., 2002, TNM CLASSIFICATION M
   Sobin LH., 1997, TNM CLASSIFICATION M
   Stupp R, 2003, ANN ONCOL, V14, P1695, DOI 10.1093/annonc/mdg501
   Stupp R, 2005, NEW ENGL J MED, V352, P987, DOI 10.1056/NEJMoa043330
   Stupp R, 2006, ONCOLOGIST, V11, P165, DOI 10.1634/theoncologist.11-2-165
   van den Bent MJ, 2005, LANCET, V366, P985, DOI 10.1016/S0140-6736(05)67070-5
NR 26
TC 1
Z9 1
U1 0
U2 3
PU WOLTERS KLUWER MEDKNOW PUBLICATIONS
PI MUMBAI
PA WOLTERS KLUWER INDIA PVT LTD , A-202, 2ND FLR, QUBE, C T S  NO 1498A-2
   VILLAGE MAROL, ANDHERI EAST, MUMBAI, Maharashtra, INDIA
SN 0028-3886
EI 1998-4022
J9 NEUROL INDIA
JI Neurol. India
PD MAY-JUN
PY 2009
VL 57
IS 3
BP 247
EP 251
DI 10.4103/0028-3886.53261
PG 5
WC Neurosciences
WE Science Citation Index Expanded (SCI-EXPANDED)
SC Neurosciences & Neurology
GA 470NK
UT WOS:000267981400004
PM 19587462
OA Green Submitted
DA 2023-08-27
ER

PT J
AU GREEN, JE
   BAIRD, AM
   HINRICHS, SH
   KLINTWORTH, GK
   JAY, G
AF GREEN, JE
   BAIRD, AM
   HINRICHS, SH
   KLINTWORTH, GK
   JAY, G
TI ADRENAL-MEDULLARY TUMORS AND IRIS PROLIFERATION IN A TRANSGENIC MOUSE
   MODEL OF NEUROFIBROMATOSIS
SO AMERICAN JOURNAL OF PATHOLOGY
LA English
DT Article
ID T-CELL LEUKEMIA; VIRUS TYPE-I; TROPICAL SPASTIC PARAPARESIS;
   COLONY-STIMULATING FACTOR; INTERLEUKIN-2 RECEPTOR; VONRECKLINGHAUSEN
   NEUROFIBROMATOSIS; LISCH NODULES; GENE; EXPRESSION; HTLV-1
AB The expression of the human T-cell lymphotropic virus type 1 (HTLV-1) tax gene in transgenic mice has been shown to result in the development of neurofibromas. 1 Further characterization of these transgenic mice bas revealed other significant pathologic similarities between this transgenic mouse model and human neurofibromatosis (NF). Pheochromocytomas of the adrenal medulla and hamartomas of the iris are well-recognized manifestations of human NF. Adrenal medullary tumors have been found in 68% of transgenic animals that were studied. They appear, however, not to be pheochromocytomas, but rather composed of undifferentiated spindle cells. Proliferation of fibroblastlike cells in the iris also occurs in one-half of the transgenic animals surviving more than 6 months. Melanocytes, however, have not been found in the transgenic iris lesion although they are characteristically found in the Lisch nodules of human NF. The similarities between human neurofibromatosis and this transgenic mouse model (in which the overexpression of a single gene results in neoplasia) are discussed This transgenic system may provide further insights into molecular mechanisms involved in the pathogenesis of neurofibromatosis.
C1 UNIV CALIF DAVIS, DAVIS, CA 95616 USA.
   DUKE UNIV, MED CTR, DEPT PATHOL, DURHAM, NC 27710 USA.
   DUKE UNIV, MED CTR, DEPT OPHTHALMOL, DURHAM, NC 27710 USA.
   AMER RED CROSS, JEROME H HOLLAND LAB, VIROL LAB, ROCKVILLE, MD USA.
C3 University of California System; University of California Davis; Duke
   University; Duke University; American Red Cross
RP GREEN, JE (通讯作者)，NCI, FCRDC,MOLEC ONCOL LAB,POB B,BLDG 469, ROOM 237, FREDERICK, MD 21702 USA.
RI Jay, Gregory/C-6346-2013
FU NCI NIH HHS [CA 496624, CA 51779-01] Funding Source: Medline
CR APPLE DJ, 1991, OCULAR PATHOLOGY CLI, P404
   BROWNSTEIN S, 1983, OPHTHALMOLOGY, V90, P1595
   BUREK JD, 1982, MOUSE BIOMEDICAL RES, V3, P432
   CALLENDER GR, 1930, AM J OPHTHALMOL, V13, P121
   CHAN JY, 1986, P NATL ACAD SCI USA, V83, P8669, DOI 10.1073/pnas.83.22.8669
   CROSS SL, 1987, CELL, V49, P47, DOI 10.1016/0092-8674(87)90754-9
   CROWE FW, 1956, CLIN PATHOLOGICAL GE, P146
   DEACON CF, 1986, GEN COMP ENDOCR, V61, P87, DOI 10.1016/0016-6480(86)90253-4
   FUJII M, 1988, P NATL ACAD SCI USA, V85, P8526, DOI 10.1073/pnas.85.22.8526
   GESSAIN A, 1985, LANCET, V2, P407
   GREEN JE, 1991, MOL CELL BIOL, V11, P4635, DOI 10.1128/MCB.11.9.4635
   GREEN JE, 1989, NATURE, V341, P72, DOI 10.1038/341072a0
   GREEN JE, 1989, MOL CELL BIOL, V9, P4731, DOI 10.1128/MCB.9.11.4731
   Greenwald M J, 1984, Pediatr Dermatol, V2, P98, DOI 10.1111/j.1525-1470.1984.tb00454.x
   HEALEY FH, 1958, NEW ENGL J MED, V258, P540, DOI 10.1056/NEJM195803132581106
   HIDAKA M, 1988, EMBO J, V7, P519, DOI 10.1002/j.1460-2075.1988.tb02840.x
   HINRICHS SH, 1987, SCIENCE, V237, P1340, DOI 10.1126/science.2888191
   HINUMA Y, 1981, P NATL ACAD SCI-BIOL, V78, P6476, DOI 10.1073/pnas.78.10.6476
   HSU SM, 1981, J HISTOCHEM CYTOCHEM, V29, P577, DOI 10.1177/29.4.6166661
   HUSON S, 1987, BRIT J OPHTHALMOL, V71, P235, DOI 10.1136/bjo.71.3.235
   INOUE J, 1986, EMBO J, V5, P2883, DOI 10.1002/j.1460-2075.1986.tb04583.x
   INOUE J, 1987, P NATL ACAD SCI USA, V84, P3653, DOI 10.1073/pnas.84.11.3653
   INSLER MS, 1985, AM J OPHTHALMOL, V99, P731, DOI 10.1016/S0002-9394(14)76056-8
   JACOBSON S, 1988, NATURE, V331, P540, DOI 10.1038/331540a0
   KOESTNER A, 1971, AM J PATHOL, V63, P37
   KUSHIDA H, 1974, J ELECTRON MICROSC, V23, P197
   LEWIS RA, 1981, OPHTHALMOLOGY, V88, P348
   LILIENBAUM A, 1990, J VIROL, V64, P256, DOI 10.1128/JVI.64.1.256-263.1990
   MANIATIS T, 1982, MOL CLONING LABORATO, P196
   NAKAMURA T, 1989, AM J PATHOL, V135, P251
   NERENBERG M, 1987, SCIENCE, V237, P1324, DOI 10.1126/science.2888190
   NERENBERG MI, 1989, AM J PATHOL, V135, P1025
   NERENBERG MI, 1991, NEUROLOGY, V41, P1687, DOI 10.1212/WNL.41.10.1687
   PEARSONWEBB MA, 1986, NEW ENGL J MED, V315, P1553
   PERRY HD, 1982, ARCH OPHTHALMOL-CHIC, V100, P1635, DOI 10.1001/archopht.1982.01030040613012
   POIESZ BJ, 1980, P NATL ACAD SCI-BIOL, V77, P7415, DOI 10.1073/pnas.77.12.7415
   RICCARDI VM, 1981, NEW ENGL J MED, V305, P1617, DOI 10.1056/NEJM198112313052704
   ROULEAU GA, 1987, NATURE, V329, P246, DOI 10.1038/329246a0
   SCHMALE MC, 1983, AM J PATHOL, V112, P238
   SHEEHAN DC, 1980, THEORY PRACTICE HIST, P278
   SHIELDS JA, 1983, DIAGNOSIS MANAGEMENT, P656
   SODROSKI J, 1985, SCIENCE, V228, P1430, DOI 10.1126/science.2990028
   SPENCER WH, 1985, OPHTHALMIC PATHOLOGY, V3, P1382
   WALLACE MR, 1990, SCIENCE, V249, P181, DOI 10.1126/science.2134734
   XU GF, 1990, CELL, V62, P599, DOI 10.1016/0092-8674(90)90024-9
   ZEHAVI C, 1986, CLIN GENET, V29, P51
NR 46
TC 24
Z9 25
U1 0
U2 0
PU ELSEVIER SCIENCE INC
PI NEW YORK
PA STE 800, 230 PARK AVE, NEW YORK, NY 10169 USA
SN 0002-9440
EI 1525-2191
J9 AM J PATHOL
JI Am. J. Pathol.
PD JUN
PY 1992
VL 140
IS 6
BP 1401
EP 1410
PG 10
WC Pathology
WE Science Citation Index Expanded (SCI-EXPANDED)
SC Pathology
GA HY135
UT WOS:A1992HY13500014
PM 1605307
DA 2023-08-27
ER

PT J
AU Singh, S
   Das, KK
   Jaiswal, S
   Kumar, R
AF Singh, Suyash
   Das, Kuntal Kanti
   Jaiswal, Sushila
   Kumar, Raj
TI Exophytic Cerebral Hemispheric Low-Grade Glioma: Unusual Growth Pattern
   of Common Central Nervous System Tumor
SO WORLD NEUROSURGERY
LA English
DT Article
DE Cerebral hemisphere; Exophytic growth; Frontal lobe; Low-grade glioma
ID RESECTION; EXTENT
AB BACKGROUND: Exophytic growth (EG), wherein the tumor mass grows beyond the neuraxial boundary formed by pia mater, remains a rare pattern of glioma growth. It has been described in gliomas at several locations like the brainstem, cerebellum, suprasellar area, spinal cord, and insula. However, EG in hemispheric grade 2 astrocytomas, particularly affecting the convexity subarachnoid space, as against the more spacious basal subarachnoid space, is exceedingly rare. To the best of our knowledge, there is only 1 such case reported in the English literature. Due to the extreme rarity, little is known about these tumors, particularly the mechanisms underlying the EG and their possible clinical implications.
   CASE DESCRIPTION: A 32-year-old woman presented with partial motor seizures without any neurologic signs. On magnetic resonance imaging of the head, a nonenhancing, T2-hyperintense, right frontal lobe lesion was noted. The majority of the lesion was occupying the adjacent subdural space while still remaining in continuity with the intra-axial globular component. During surgery, the exophytic nature of the lesion was confirmed. We performed maximal resection of the exophytic portion with subtotal excision of the intra-axial component. She recovered uneventfully after surgery and subsequently received external beam radiotherapy for the residual tumor.
   CONCLUSION: Although extremely rare, cerebral hemispheric grade 2 astrocytomas may have an EG pattern. The exact mechanisms underlying this are not clearly known. More such cases need to be reported so that we can decipher the true nature of these tumors.
C1 [Singh, Suyash; Das, Kuntal Kanti; Kumar, Raj] Sanjay Gandhi Postgrad Inst Med Sci, Dept Neurosurg, Lucknow, Uttar Pradesh, India.
   [Jaiswal, Sushila] Sanjay Gandhi Postgrad Inst Med Sci, Dept Pathol, Lucknow, Uttar Pradesh, India.
C3 Sanjay Gandhi Postgraduate Institute of Medical Sciences; Sanjay Gandhi
   Postgraduate Institute of Medical Sciences
RP Das, KK (通讯作者)，Sanjay Gandhi Postgrad Inst Med Sci, Dept Neurosurg, Lucknow, Uttar Pradesh, India.
EM
RI Das, Kuntal/AAV-6549-2020; Kumar, Rajesh/G-1408-2014
OI Kumar, Rajesh/0000-0002-6019-0702
CR Adeeb N, 2013, CHILD NERV SYST, V29, P17, DOI 10.1007/s00381-012-1910-x
   Das Kuntal Kanti, 2017, Asian J Neurosurg, V12, P224, DOI 10.4103/1793-5482.144151
   Engwer C, 2015, J MATH BIOL, V71, P551, DOI 10.1007/s00285-014-0822-7
   EPSTEIN FJ, 1993, J NEUROSURG, V78, P408, DOI 10.3171/jns.1993.78.3.0408
   Gerganov VM, 2011, J CLIN NEUROSCI, V18, P669, DOI 10.1016/j.jocn.2010.08.017
   Ghodsi M, 2013, PEDIATR NEUROSURG, V49, P195, DOI 10.1159/000362621
   Incekara F, 2016, J NEUROL SURG PART A, V77, P79, DOI 10.1055/s-0035-1551830
   Larjavaara S, 2007, NEURO-ONCOLOGY, V9, P319, DOI 10.1215/15228517-2007-016
   Perry Arie, 2016, Handb Clin Neurol, V134, P71, DOI 10.1016/B978-0-12-802997-8.00005-0
   POLLACK IF, 1993, J NEUROSURG, V78, P859, DOI 10.3171/jns.1993.78.6.0859
   Sanai N, 2008, NEUROSURGERY, V62, P753, DOI 10.1227/01.neu.0000318159.21731.cf
   Singh Gajendra, 2015, Asian J Neurosurg, V10, P195, DOI 10.4103/1793-5482.161173
   Tukaram S.P., 2016, INDIAN J MED RES PHA, V3, P41
   Velasquez C, 2016, WORLD NEUROSURG, V87, P200, DOI 10.1016/j.wneu.2015.12.035
   Yordanova YN, 2017, NEUROCHIRURGIE, V63, P243, DOI 10.1016/j.neuchi.2016.09.006
NR 15
TC 4
Z9 4
U1 0
U2 0
PU ELSEVIER SCIENCE INC
PI NEW YORK
PA 360 PARK AVE SOUTH, NEW YORK, NY 10010-1710 USA
SN 1878-8750
EI 1878-8769
J9 WORLD NEUROSURG
JI World Neurosurg.
PD MAY
PY 2018
VL 113
BP 184
EP 187
DI 10.1016/j.wneu.2018.02.048
PG 4
WC Clinical Neurology; Surgery
WE Science Citation Index Expanded (SCI-EXPANDED)
SC Neurosciences & Neurology; Surgery
GA GG8JD
UT WOS:000432942700124
PM 29454959
DA 2023-08-27
ER

PT J
AU Tannus, M
   Goldman, SM
   Andreoni, C
AF Tannus, Matheus
   Goldman, Suzan M.
   Andreoni, Cassio
TI Practical and Intuitive Surgical Approach Renal Ranking to Predict
   Outcomes in the Management of Renal Tumors: A Novel Score Tool
SO JOURNAL OF ENDOUROLOGY
LA English
DT Article
ID LAPAROSCOPIC PARTIAL NEPHRECTOMY; CELL CARCINOMA; RADICAL NEPHRECTOMY;
   DISEASE; MASSES; SIZE; RISK
AB Background and Purpose: Surgery continues to be the main form of treatment for patients with renal tumors. We create a more practical and intuitive score for renal tumor classification.
   Patients and Methods: Eighty patients underwent surgery for renal tumors and were prospectively enrolled. The tumors were classified using the following variables: (1) tumor size, (2) endophytic or exophytic tumor, (3) longitudinal location of the tumor, (4) the extent of the impairment of the renal parenchyma, (5) relationship with the renal sinus, and (6) anterior or posterior.
   Results: The mean operative time, tumor size, and bleeding increased proportionally to the increased complexity of the tumor measured by scores (P < 0.0001, P < 0.0001, and P = 0.036, respectively). The mean total score was 8.7 points for patients undergoing partial nephrectomy (PN) and 14.4 points for those undergoing radical nephrectomy (RN) (P < 0.0001). Patients with larger tumors, completely endophytic, which exceeded the renal medulla, and were centrally located, underwent RN more often (86.7% - P < 0.0001, 64% - P = 0.01, 77% P < 0.0001, and 78.9% - P < 0.0001, respectively). In univariate analysis, RN was associated with tumors larger than 7 cm (P = 0.001), tumors that exceeded the renal medullary (< 0.001), centrally located tumors (odds ration [OR] = 150, P < 0.001), and tumors of high complexity (P < 0.001). Analysis showed no association between complications and variables in the score. The findings were similar when the tumors were evaluated with the R.E.N.A.L. score system.
   Conclusion: Surgical Approach Renal Ranking is a simple, practical and intuitive classification for renal tumors that can be used in the decision-making process and to predict outcomes in the surgical treatment of patients with renal tumors.
C1 [Tannus, Matheus; Goldman, Suzan M.; Andreoni, Cassio] Univ Fed Sao Paulo, Div Urol, Sao Paulo, Brazil.
C3 Universidade Federal de Sao Paulo (UNIFESP)
RP Tannus, M (通讯作者)，R Alexandrina Ramalho,70 Apt 904, BR-40296230 Salvador, BA, Brazil.
EM
CR Bruner B, 2011, BJU INT, V108, P67, DOI 10.1111/j.1464-410X.2010.09837.x
   Campbell SC, 2009, J UROLOGY, V182, P1271, DOI 10.1016/j.juro.2009.07.004
   Ficarra V, 2009, EUR UROL, V56, P786, DOI 10.1016/j.eururo.2009.07.040
   Finley DS, 2008, J UROLOGY, V180, P492, DOI 10.1016/j.juro.2008.04.019
   Frank I, 2003, J UROLOGY, V170, P2217, DOI 10.1097/01.ju.0000095475.12515.5e
   Heuer R, 2010, EUR UROL, V57, P223, DOI 10.1016/j.eururo.2009.10.023
   Hew MN, 2011, J UROLOGY, V186, P42, DOI 10.1016/j.juro.2011.03.020
   Huang WC, 2006, LANCET ONCOL, V7, P735, DOI 10.1016/S1470-2045(06)70803-8
   Janzen NK, 2003, UROL CLIN N AM, V30, P843, DOI 10.1016/S0094-0143(03)00056-9
   Kontak JA, 2003, UROL CLIN N AM, V30, P467, DOI 10.1016/S0094-0143(03)00020-X
   Kutikov A, 2009, J UROLOGY, V182, P844, DOI 10.1016/j.juro.2009.05.035
   Lane BR, 2007, J UROLOGY, V177, P70, DOI 10.1016/j.juro.2006.08.093
   McKiernan J, 2002, UROLOGY, V59, P816, DOI 10.1016/S0090-4295(02)01501-7
   Meeks JJ, 2008, J UROLOGY, V180, P2375, DOI 10.1016/j.juro.2008.08.018
   Ng CS, 2005, J UROLOGY, V174, P846, DOI 10.1097/01.ju.0000169259.49754.02
   Okhunov Z, 2011, J ENDOUROL, V25, P1921, DOI 10.1089/end.2011.0301
   Patard JJ, 2004, J UROLOGY, V172, P2167, DOI 10.1097/01.ju.0000141137.61330.4d
   ROBSON CJ, 1969, J UROLOGY, V101, P297, DOI 10.1016/S0022-5347(17)62331-0
   Rosevear HM, 2012, BJU INT, V109, P700, DOI 10.1111/j.1464-410X.2011.10452.x
   Simmons MN, 2010, J UROLOGY, V183, P1708, DOI 10.1016/j.juro.2010.01.005
   Thompson RH, 2008, J UROLOGY, V179, P468, DOI 10.1016/j.juro.2007.09.077
   Weizer AZ, 2008, J UROLOGY, V180, P1273, DOI 10.1016/j.juro.2008.06.066
   Wingo MS, 2008, J ENDOUROL, V22, P1261, DOI 10.1089/end.2008.0135
   Zisman A, 2001, J UROLOGY, V166, P54, DOI 10.1016/S0022-5347(05)66075-2
NR 24
TC 13
Z9 14
U1 0
U2 4
PU MARY ANN LIEBERT, INC
PI NEW ROCHELLE
PA 140 HUGUENOT STREET, 3RD FL, NEW ROCHELLE, NY 10801 USA
SN 0892-7790
EI 1557-900X
J9 J ENDOUROL
JI J. Endourol.
PD APR
PY 2014
VL 28
IS 4
BP 487
EP 492
DI 10.1089/end.2013.0148
PG 6
WC Urology & Nephrology
WE Science Citation Index Expanded (SCI-EXPANDED)
SC Urology & Nephrology
GA AI9YM
UT WOS:000337304100016
PM 24437510
DA 2023-08-27
ER

PT J
AU Agamanolis, DP
   Katsetos, CD
   Klonk, CJ
   Bartkowski, HM
   Ganapathy, S
   Staugaitis, SM
   Kuerbitz, SJ
   Patton, DF
   Talaizadeh, A
   Cohen, BH
AF Agamanolis, Dimitri P.
   Katsetos, Christos D.
   Klonk, Christopher J.
   Bartkowski, Henry M.
   Ganapathy, Srinivas
   Staugaitis, Susan M.
   Kuerbitz, Steven J.
   Patton, Donna F.
   Talaizadeh, Ali
   Cohen, Bruce H.
TI An Unusual Form of Superficially Disseminated Glioma in Children: Report
   of 3 Cases
SO JOURNAL OF CHILD NEUROLOGY
LA English
DT Article
DE brain tumors; disseminated gliomas; gliomatosis
ID EXTENSIVE LEPTOMENINGEAL DISSEMINATION; III BETA-TUBULIN; 1P;
   OLIGODENDROGLIOMA; LOCALIZATION; DELETION; TUMORS; 19Q
AB Three children, aged 4, 5, and 9 years, had an insidious onset of ataxia. Magnetic resonance imaging (MRI) showed hydrocephalus and countless foci of high T2 signal coating the cerebellum, basilar cisterns, brainstem, and fourth ventricle. Similar lesions were present in the spinal cord. Symptoms were relatively mild given the massive tumor burden. Biopsies were composed of superficially infiltrating cells with oligodendroglioma-like features (perinuclear halos and cytologic monotony) and microcysts. Classical cytogenetic analysis of 2 cases showed normal karyotypes. Chromosome fluorescence in situ hybridization revealed 1p36 deletion with intact 19q in 2 cases and no abnormality in one. A similar combination of clinical, MRI, and histopathologic findings has been reported previously in 10 other cases. The pathologic findings suggest a glioma with diffuse or multifocal superficial origin and do not correspond to a described entity in the current World Health Organization (WHO) classification of brain tumors.
C1 [Agamanolis, Dimitri P.; Klonk, Christopher J.] Akron Childrens Hosp, Dept Pathol, Akron, OH 44308 USA.
   [Agamanolis, Dimitri P.; Klonk, Christopher J.; Bartkowski, Henry M.; Kuerbitz, Steven J.; Patton, Donna F.; Talaizadeh, Ali; Cohen, Bruce H.] Northeastern Ohio Univ Coll Med & Pharm, Coll Med, Akron, OH USA.
   [Katsetos, Christos D.] Drexel Univ, Dept Pediat, Coll Med, St Christophers Hosp Children, Philadelphia, PA 19104 USA.
   [Katsetos, Christos D.] Drexel Univ, Dept Pathol & Lab Med, Coll Med, St Christophers Hosp Children, Philadelphia, PA 19104 USA.
   [Katsetos, Christos D.] Drexel Univ, Dept Neurol, Coll Med, St Christophers Hosp Children, Philadelphia, PA 19104 USA.
   [Bartkowski, Henry M.] Akron Childrens Hosp, Div Neurosurg, Akron, OH 44308 USA.
   [Ganapathy, Srinivas; Staugaitis, Susan M.] Cleveland Clin, Dept Neurosci & Anat Pathol, Cleveland, OH 44106 USA.
   [Kuerbitz, Steven J.; Patton, Donna F.; Talaizadeh, Ali] Akron Childrens Hosp, Div Hematol Oncol, Akron, OH 44308 USA.
   [Cohen, Bruce H.] Akron Childrens Hosp, Div Pediat Neurol, Akron, OH 44308 USA.
C3 Akron Children's Hospital; Northeast Ohio Medical University (NEOMED);
   Drexel University; Drexel University; Drexel University; Akron
   Children's Hospital; Cleveland Clinic Foundation; Akron Children's
   Hospital; Akron Children's Hospital
RP Agamanolis, DP (通讯作者)，Akron Childrens Hosp, Dept Pathol, 1 Perkins Sq, Akron, OH 44308 USA.
EM
CR Armao DM, 2000, AM J NEURORADIOL, V21, P1122
   Bagchi A, 2007, CELL, V128, P459, DOI 10.1016/j.cell.2006.11.052
   Bourne TD, 2006, J NEUROSURG, V105, P465, DOI 10.3171/ped.2006.105.6.465
   BRODEUR GM, 1977, CANCER-AM CANCER SOC, V40, P2256, DOI 10.1002/1097-0142(197711)40:5<2256::AID-CNCR2820400536>3.0.CO;2-1
   Cairncross JG, 1998, J NATL CANCER I, V90, P1473, DOI 10.1093/jnci/90.19.1473
   Gardiman MP, 2010, BRAIN PATHOL, V20, P361, DOI 10.1111/j.1750-3639.2009.00285.x
   Gilmer-Hill HS, 2000, J NEUROSURG, V92, P109, DOI 10.3171/spi.2000.92.1.0109
   Huang T, 2001, NEURORADIOLOGY, V43, P868, DOI 10.1007/s002340100604
   Ichimura K, 2008, ONCOGENE, V27, P2097, DOI 10.1038/sj.onc.1210848
   Katsetos CD, 2003, J CHILD NEUROL, V18, P851, DOI 10.1177/088307380301801205
   Katsetos CD, 2002, J NEUROPATH EXP NEUR, V61, P307, DOI 10.1093/jnen/61.4.307
   Katsetos CD, 2001, ARCH PATHOL LAB MED, V125, P613
   LEE VMY, 1988, P NATL ACAD SCI USA, V85, P7384, DOI 10.1073/pnas.85.19.7384
   Perilongo G, 2002, CHILD NERV SYST, V18, P505, DOI 10.1007/s00381-002-0626-8
   Perry A, 2003, FRONT BIOSCI-LANDMRK, V8, pA1, DOI 10.2741/896
   Prayson RA, 2002, AM J CLIN PATHOL, V118, P512
   Senaratna S, 2001, J CLIN NEUROSCI, V8, P363, DOI 10.1054/jocn.2000.0802
   SHEA TB, 1993, J NEUROIMMUNOL, V44, P117, DOI 10.1016/0165-5728(93)90274-3
   Smith JS, 1999, ONCOGENE, V18, P4144, DOI 10.1038/sj.onc.1202759
   STERNBERGER LA, 1983, P NATL ACAD SCI-BIOL, V80, P6126, DOI 10.1073/pnas.80.19.6126
   Yamamoto T, 1996, AM J SURG PATHOL, V20, P363, DOI 10.1097/00000478-199603000-00014
   Yip S, 2008, J NEUROPATH EXP NEUR, V67, P1, DOI 10.1097/nen.0b013e31815f65fb
NR 22
TC 22
Z9 22
U1 0
U2 1
PU SAGE PUBLICATIONS INC
PI THOUSAND OAKS
PA 2455 TELLER RD, THOUSAND OAKS, CA 91320 USA
SN 0883-0738
EI 1708-8283
J9 J CHILD NEUROL
JI J. Child Neurol.
PD JUN
PY 2012
VL 27
IS 6
BP 727
EP 733
DI 10.1177/0883073811426500
PG 7
WC Clinical Neurology; Pediatrics
WE Science Citation Index Expanded (SCI-EXPANDED)
SC Neurosciences & Neurology; Pediatrics
GA 943RN
UT WOS:000304143000006
PM 22596013
DA 2023-08-27
ER

PT J
AU Upadhyaya, SA
   Ghazwani, Y
   Wu, SJ
   Broniscer, A
   Boop, FA
   Gajjar, A
   Qaddoumi, I
AF Upadhyaya, Santhosh A.
   Ghazwani, Yahya
   Wu, Shengjie
   Broniscer, Alberto
   Boop, Fredrick A.
   Gajjar, Amar
   Qaddoumi, Ibrahim
TI Mortality in children with low-grade glioma or glioneuronal tumors: A
   single-institution study
SO PEDIATRIC BLOOD & CANCER
LA English
DT Article
DE children; low-grade glioma; progressive disease; suicide; violent deaths
ID TERM-FOLLOW-UP; LONG-TERM; PILOCYTIC ASTROCYTOMAS; GENETIC ALTERATIONS;
   CEREBELLAR ASTROCYTOMAS; ADULT SURVIVORS; BRAF; OUTCOMES; FUSION;
   CHEMOTHERAPY
AB BackgroundWhile pediatric low-grade glioma/glioneuronal tumors (LGG/LGGNTs) are considered slow-growing, indolent tumors with excellent long-term prognosis, mortality due to the disease is not unknown. Few studies have addressed the cause of death in this population.
   MethodsRetrospective review of clinicopathologic and radiologic data for children 21 years or younger with LGG/LGGNT who died at St. Jude Children's Research Hospital between April 1985 and June 2015. Our primary objective was to determine the causes and timing of mortality in affected children.
   ResultsFor the 87 eligible patients, median age at diagnosis was 7.7 years (range, 0.21-21 years), median age at death was 14.26 years (range, 0.58-32 years), and median time to death from diagnosis was 4.02 years (range, 0.21-24 years). Midbrain/thalamus was the most common tumor location (n=34), followed by suprasellar/hypothalamic (n = 18) and cerebrocortical (n = 13). Astrocytoma not otherwise specified (n = 24), pilocytic astrocytoma (n = 23), and fibrillary astrocytoma (n = 11) were the predominant histologic diagnoses. Causes of death included progressive primary disease (PD) (n = 43), progression of PD with histological features of a high-grade glioma at progression or at autopsy (PD-HGG) (n = 15), second cancer (n=3), suicide (n = 4), and vehicular accident (n = 3). Among the 15 patients with PD-HGG, 12 received radiation therapy before histologic confirmation of progression.
   ConclusionsPD and PD-HGG contributed to 66% of the mortality in our patient cohort. Early psychological intervention should be included as part of the multidisciplinary management approach of children with LGG/LGGNT to reduce the risk of suicide in vulnerable subjects.
C1 [Upadhyaya, Santhosh A.; Ghazwani, Yahya; Broniscer, Alberto; Gajjar, Amar; Qaddoumi, Ibrahim] St Jude Childrens Res Hosp, Div Neuro Oncol, Dept Oncol, MS 260,262 Danny Thomas Pl, Memphis, TN 38105 USA.
   [Wu, Shengjie] St Jude Childrens Res Hosp, Dept Biostat, 332 N Lauderdale St, Memphis, TN 38105 USA.
   [Boop, Fredrick A.] St Jude Childrens Res Hosp, Dept Surg, 332 N Lauderdale St, Memphis, TN 38105 USA.
   [Boop, Fredrick A.] LeBonheur Childrens Hosp, Dept Neurosurg, Memphis, TN USA.
C3 St Jude Children's Research Hospital; St Jude Children's Research
   Hospital; St Jude Children's Research Hospital
RP Qaddoumi, I (通讯作者)，St Jude Childrens Res Hosp, Div Neuro Oncol, Dept Oncol, MS 260,262 Danny Thomas Pl, Memphis, TN 38105 USA.
EM
RI Gajjar, Amar/N-8059-2018; Qaddoumi, Ibrahim/N-8101-2018; Upadhyaya,
   Santhosh A/N-3914-2018
OI Upadhyaya, Santhosh A/0000-0001-6101-149X; Qaddoumi,
   Ibrahim/0000-0002-6645-4895; Ghazwani, Yahya/0000-0001-6239-6045
FU National Institutes of Health [CA21765]; ALSAC
FX Grant sponsor: National Institutes of Health; Grant number: CA21765;
   Grant sponsor: ALSAC.
CR Armstrong GT, 2011, NEURO-ONCOLOGY, V13, P223, DOI 10.1093/neuonc/noq178
   Ater JL, 2012, J CLIN ONCOL, V30, P2641, DOI 10.1200/JCO.2011.36.6054
   Bandopadhayay P, 2014, PEDIATR BLOOD CANCER, V61, P1173, DOI 10.1002/pbc.24958
   Becker AP, 2015, J NEUROPATH EXP NEUR, V74, P743, DOI 10.1097/NEN.0000000000000213
   Brinkman TM, 2013, J NEURO-ONCOL, V113, P425, DOI 10.1007/s11060-013-1130-6
   Broadway SJ, 2011, J NEUROSURG-PEDIATR, V7, P589, DOI 10.3171/2011.4.PEDS119
   Broniscer A, 2007, J CLIN ONCOL, V25, P682, DOI 10.1200/JCO.2006.06.8213
   Chalil A, 2016, J CHILD NEUROL, V31, P517, DOI 10.1177/0883073815599259
   Chamdine O, 2016, PEDIATR BLOOD CANCER, V63, P62, DOI 10.1002/pbc.25731
   Cin H, 2011, ACTA NEUROPATHOL, V121, P763, DOI 10.1007/s00401-011-0817-z
   Dahiya S, 2013, ACTA NEUROPATHOL, V125, P901, DOI 10.1007/s00401-013-1120-y
   Gajjar A, 1997, J CLIN ONCOL, V15, P2792, DOI 10.1200/JCO.1997.15.8.2792
   Gnekow AK, 2012, NEURO-ONCOLOGY, V14, P1265, DOI 10.1093/neuonc/nos202
   Gunnes MW, 2017, INT J CANCER, V140, P575, DOI 10.1002/ijc.30474
   Gunny RS, 2005, PEDIATR RADIOL, V35, P1086, DOI 10.1007/s00247-005-1546-z
   Guzman-De-Villoria JA, 2014, CANCER IMAGING, V14, DOI 10.1186/s40644-014-0035-8
   Hawkins C, 2011, CLIN CANCER RES, V17, P4790, DOI 10.1158/1078-0432.CCR-11-0034
   Jones DTW, 2012, CELL MOL LIFE SCI, V69, P1799, DOI 10.1007/s00018-011-0898-9
   Krieger MD, 1997, PEDIATR NEUROSURG, V27, P1, DOI 10.1159/000121218
   Krishnatry R, 2016, CANCER-AM CANCER SOC, V122, P1261, DOI 10.1002/cncr.29907
   Loh JK, 2013, CHILD NERV SYST, V29, P2051, DOI 10.1007/s00381-013-2113-9
   Marton E, 2007, J NEUROL SCI, V252, P144, DOI 10.1016/j.jns.2006.11.008
   MCCORMACK BM, 1992, NEUROSURGERY, V31, P636
   Messing-Junger AM, 2002, CHILD NERV SYST, V18, P445, DOI 10.1007/s00381-002-0644-6
   Mistry M, 2015, J CLIN ONCOL, V33, P1015, DOI 10.1200/JCO.2014.58.3922
   Morana G, 2014, J NUCL MED, V55, P718, DOI 10.2967/jnumed.113.125500
   Ogiwara H, 2012, NEUROSURGERY, V70, P40, DOI 10.1227/NEU.0b013e31822ff0ed
   Oh KS, 2011, INT J RADIAT ONCOL, V81, pE481, DOI 10.1016/j.ijrobp.2011.01.019
   Penman CL, 2015, FRONT ONCOL, V5, DOI 10.3389/fonc.2015.00054
   Perilongo G, 2005, J NEURO-ONCOL, V75, P301, DOI 10.1007/s11060-005-6754-8
   Peters O, 2004, PEDIATR BLOOD CANCER, V43, P250, DOI 10.1002/pbc.20111
   Qaddoumi I, 2016, ACTA NEUROPATHOL, V131, P833, DOI 10.1007/s00401-016-1539-z
   Rakotonjanahary J, 2015, PLOS ONE, V10, DOI 10.1371/journal.pone.0127676
   Rumana CS, 1998, NEUROSURGERY, V42, P1038, DOI 10.1097/00006123-199805000-00049
   Ryall S, 2016, ACTA NEUROPATHOL COM, V4, DOI 10.1186/s40478-016-0353-0
   Shah SS, 2015, CHILD NERV SYST, V31, P653, DOI 10.1007/s00381-015-2669-7
   Sparrow J, 2016, J PEDIATR REHAB MED, V9, P117, DOI 10.3233/PRM-160372
   Stokland T, 2010, NEURO-ONCOLOGY, V12, P1257, DOI 10.1093/neuonc/noq092
   Tabori U, 2006, NEOPLASIA, V8, P136, DOI 10.1593/neo.05715
   Zhang JH, 2013, NAT GENET, V45, P602, DOI 10.1038/ng.2611
NR 40
TC 7
Z9 8
U1 0
U2 5
PU WILEY
PI HOBOKEN
PA 111 RIVER ST, HOBOKEN 07030-5774, NJ USA
SN 1545-5009
EI 1545-5017
J9 PEDIATR BLOOD CANCER
JI Pediatr. Blood Cancer
PD JAN
PY 2018
VL 65
IS 1
AR e26717
DI 10.1002/pbc.26717
PG 9
WC Oncology; Hematology; Pediatrics
WE Science Citation Index Expanded (SCI-EXPANDED)
SC Oncology; Hematology; Pediatrics
GA FN6JE
UT WOS:000416119400011
PM 28708330
OA Green Accepted
DA 2023-08-27
ER

PT J
AU Zhao, Y
   Wang, J
   Li, MH
   Ma, TT
   Zhang, XY
   Xu, X
   Zeng, M
   Peng, YM
AF Zhao, Yan
   Wang, Juan
   Li, Muhan
   Ma, Tingting
   Zhang, Xingyue
   Xu, Xin
   Zeng, Min
   Peng, Yuming
TI The Influence of Trigeminocardiac Reflex on Postoperative Cardiac
   Adverse Events in Patients Undergoing Cerebellopontine Angle Tumor
   Resections: A Case-Control Study
SO WORLD NEUROSURGERY
LA English
DT Article
DE Adverse cardiac events; Cerebellopontine angle; Trigeminocardiac reflex
ID SKULL BASE; SURGERY
AB OBJECTIVE: To assess the potential impact of the trige-minocardiac reflex (TCR) on postoperative adverse cardiac events and to identify predictors of the TCR in cer-ebellopontine angle surgery. -METHODS: Patients undergoing elective cer-ebellopontine angle surgery from October 1, 2015, to September 30, 2020, were recruited consecutively for this retrospective case-control study. The TCR was evaluated by reviewing electronic anesthesia records and defined as a drop in heart rate was >20%. Controls were identified from the same retrospective cohort and matched by age, sex, and similar (+/- 5 days) surgery date in the ratio of 1:2. -RESULTS: Of 2446 patients, 68 (2.78%) experienced TCR episodes. A total of 97 TCR episodes occurred among the 68 patients. In 2 TCR episodes, severe cardiac complications developed after surgery-myocardial injury in one case and cardiac arrest in the other case. The prevalence of adverse cardiovascular events was higher in the TCR group (60.3% vs. 36.0%, P = 0.001) than in the control group. The independent risk factor for the TCR in the multivariate condition logistic regression was tumor compression of the brainstem (odds ratio = 2.36, 95% confidence interval 1.40-3.95; P = 0.001). -CONCLUSIONS: Intraoperative TCR episodes seemed to be associated with postoperative adverse cardiac events in patients undergoing cerebellopontine angle surgery. Moreover, tumor compression of the brainstem might be a risk factor for TCR episodes.
C1 [Zhao, Yan; Wang, Juan; Li, Muhan; Ma, Tingting; Zhang, Xingyue; Zeng, Min; Peng, Yuming] Capital Med Univ, Beijing Tiantan Hosp, Dept Anesthesiol, Beijing, Peoples R China.
   [Xu, Xin] Chinese Acad Med Sci, Canc Hosp, Dept Anesthesiol, Beijing, Peoples R China.
C3 Capital Medical University; Chinese Academy of Medical Sciences - Peking
   Union Medical College
RP Peng, YM (通讯作者)，Capital Med Univ, Beijing Tiantan Hosp, Dept Anesthesiol, Beijing, Peoples R China.
EM
CR Adachi K, 2021, NEUROSURG REV, V44, P1173, DOI 10.1007/s10143-020-01313-3
   Arnold RW, 2021, CLIN OPHTHALMOL, V15, P973, DOI 10.2147/OPTH.S300860
   Chowdhury T, 2022, J NEUROSURG ANESTH, V34, P282, DOI 10.1097/ANA.0000000000000723
   Chowdhury T, 2017, FRONT NEUROL, V8, DOI 10.3389/fneur.2017.00022
   Chowdhury T, 2014, NEUROL RES INT, V2014, DOI 10.1155/2014/974930
   Chowdhury T, 2015, J NEUROSURG ANESTH, V27, P136, DOI 10.1097/ANA.0000000000000065
   Haas CE, 2004, AM J HEALTH-SYST PH, V61, P1661, DOI 10.1093/ajhp/61.16.1661
   Harada Kaori, 2011, Masui, V60, P1169
   Hemmer LB, 2010, J CLIN ANESTH, V22, P205, DOI 10.1016/j.jclinane.2009.03.017
   Koerbel A, 2005, ACTA NEUROCHIR, V147, P727, DOI 10.1007/s00701-005-0535-1
   Li C, 2019, CRIT CARE MED, V47, P478
   Manolis AA, 2021, TRENDS CARDIOVAS MED, V31, P290, DOI 10.1016/j.tcm.2020.04.011
   Meuwly C, 2016, J NEUROSURG ANESTH, V28, P437, DOI 10.1097/ANA.0000000000000240
   Meuwly C, 2017, FRONT NEUROL, V8, DOI 10.3389/fneur.2017.00533
   Meuwly C, 2015, MEDICINE, V94, DOI 10.1097/MD.0000000000000807
   Nalivaiko E, 2003, AUTON NEUROSCI-BASIC, V105, P101, DOI 10.1016/S1566-0702(03)00048-1
   Paton JFR, 2005, BRAIN RES REV, V49, P555, DOI 10.1016/j.brainresrev.2005.02.005
   Paton JFR, 2006, CLIN EXP PHARMACOL P, V33, P1245, DOI 10.1111/j.1440-1681.2006.04518.x
   Reddy KRM, 2006, J NEUROSURG ANESTH, V18, P163
   Sandu N, 2015, FUTUR NEUROL, V10, P7, DOI 10.2217/FNL.14.66
   Schaller B, 2005, CLIN NEUROL NEUROSUR, V107, P468, DOI 10.1016/j.clineuro.2004.12.004
   Schaller B, 1999, J NEUROSURG, V90, P215, DOI 10.3171/jns.1999.90.2.0215
   Schaller B, 2009, J NEUROSURG ANESTH, V21, P187, DOI 10.1097/ANA.0b013e3181a2bf22
   Shakil H, 2019, WORLD NEUROSURG, V122, P424, DOI 10.1016/j.wneu.2018.10.186
   Shibao S, 2017, WORLD NEUROSURG, V106, P939, DOI [10.1016/J.WNEU.2017.07.086, 10.1016/j.wneu.2017.07.086]
   Spiriev T, 2011, J NEUROSURG ANESTH, V23, P379, DOI 10.1097/ANA.0b013e3182312486
   Spiriev T, 2011, J NEUROSURG ANESTH, V23, P271, DOI 10.1097/ANA.0b013e3182204c2c
   Sun ZC, 2022, FRONT SURG, V9, DOI 10.3389/fsurg.2022.989644
NR 28
TC 0
Z9 0
U1 0
U2 0
PU ELSEVIER SCIENCE INC
PI NEW YORK
PA STE 800, 230 PARK AVE, NEW YORK, NY 10169 USA
SN 1878-8750
EI 1878-8769
J9 WORLD NEUROSURG
JI World Neurosurg.
PD APR
PY 2023
VL 172
BP E291
EP E298
DI 10.1016/j.wneu.2023.01.010
EA MAR 2023
PG 8
WC Clinical Neurology; Surgery
WE Science Citation Index Expanded (SCI-EXPANDED)
SC Neurosciences & Neurology; Surgery
GA D9WG1
UT WOS:000972153900001
PM 36623723
OA Bronze
DA 2023-08-27
ER

PT J
AU Hukin, J
   Siffert, J
   Cohen, H
   Velasquez, L
   Zagzag, D
   Allen, J
AF Hukin, J
   Siffert, J
   Cohen, H
   Velasquez, L
   Zagzag, D
   Allen, J
TI Leptomeningeal dissemination at diagnosis of pediatric low-grade
   neuroepithelial tumors
SO NEURO-ONCOLOGY
LA English
DT Article
ID PILOCYTIC ASTROCYTOMAS; GLIOMA; CHILDREN; CHEMOTHERAPY; GROWTH; CELLS
AB The goal of this study was to describe the demographic, histologic, and prognostic features of children with low-grade neuroepithelial tumors (LGN) of the CNS presenting with leptomeningeal metastases (LM) at diagnosis. We identified 528 newly diagnosed LGN children, 13 (3%) of whom had LM at diagnosis. LM was defined by neuroimaging, clinical evidence, and/or biopsy. The charts were reviewed and patients contacted to validate the demographic data, treatment, and clinical status. The distribution of LM patients by primary tumor site was diencephalon, 5; cerebrum, 2; spinal cord, 3; brainstem, 2; and cerebellum, 1. Six of 8 patients with LM had durable objective responses to chemotherapy. The 5-year progression-free survival of patients with LM at diagnosis was 17%, compared to 85% (95% Cl, 80%-91%) for those with localized LGN who had a gross total resection and 51% (95% Cl, 44%-52%) for those with localized LGN who had less aggressive surgery (P < 0.0001). Only 1 of these 13 LM patients died. The 5-year overall survival of the localized LGN group with a gross total resection was 97% (95% Cl, 92%-99.9%), and that of the localized LGN group with less aggressive surgery was 88% (95% CI, 84%-95%) (P = 0.004). The 3% frequency of LM at diagnosis is likely an underestimate since patients with newly diagnosed LGN were not routinely staged. We suggest that staging be considered in the following circumstances: diencephalic primary site, unexplained hydrocephalus, clinical features suggestive of LM, and before adjuvant therapy is initiated. The prognosis for children with LM at diagnosis is favorable, and its identification alters therapeutic strategies.
C1 Womens & Childrens Hosp, Dept Pediat, Div Neurol & Oncol, Vancouver, BC V6H 3V4, Canada.
   CNS Pfizer Inc, New York, NY 10017 USA.
   Beth Israel Deaconess Med Ctr, Inst Neurol & Neurosurg, New York, NY 10128 USA.
   Natl Taiwan Univ Hosp, Taipei 10016, Taiwan.
C3 Pfizer; Harvard University; Beth Israel Deaconess Medical Center;
   National Taiwan University; National Taiwan University Hospital
RP Hukin, J (通讯作者)，Womens & Childrens Hosp, Div Neurol, 4480 Oak St, Vancouver, BC V6H 3V4, Canada.
EM
OI Hukin, Juliette/0000-0002-8727-8456; Allen, Jeffrey/0000-0001-9957-6664
CR Akar Z, 2000, CHILD NERV SYST, V16, P309, DOI 10.1007/s003810050520
   Bauman G, 1999, CAN J NEUROL SCI, V26, P18
   BOURDON MA, 1983, CANCER RES, V43, P2796
   BROOKS PC, 1994, SCIENCE, V264, P569, DOI 10.1126/science.7512751
   Campbell JW, 1996, J NEURO-ONCOL, V28, P223, DOI 10.1007/bf00250201
   CINALLI G, 1995, J NEUROSURG, V83, P480, DOI 10.3171/jns.1995.83.3.0480
   CIVITELLO LA, 1988, NEUROLOGY, V38, P562, DOI 10.1212/WNL.38.4.562
   Claffey KP, 1996, CANCER METAST REV, V15, P165, DOI 10.1007/BF00437469
   DEVRIES C, 1992, SCIENCE, V255, P989, DOI 10.1126/science.1312256
   Di X, 1997, J EXP CLIN CANC RES, V16, P153
   Doireau V, 1999, BRIT J CANCER, V81, P835, DOI 10.1038/sj.bjc.6690772
   EDVARDSEN K, 1993, INT J DEV NEUROSCI, V11, P681, DOI 10.1016/0736-5748(93)90055-I
   GAJJAR A, 1995, J NEUROSURG, V83, P67, DOI 10.3171/jns.1995.83.1.0067
   Giannini C, 1997, BRAIN PATHOL, V7, P785, DOI 10.1111/j.1750-3639.1997.tb01064.x
   Giese A, 1996, INT J CANCER, V67, P275, DOI 10.1002/(SICI)1097-0215(19960717)67:2<275::AID-IJC20>3.0.CO;2-9
   Hukin J, 2002, NEURO-ONCOLOGY, V4, P253, DOI 10.1093/neuonc/4.4.253
   Jaworski DM, 1996, CANCER RES, V56, P2293
   MAMELAK AN, 1994, J NEUROSURG, V81, P24, DOI 10.3171/jns.1994.81.1.0024
   McCowage G, 1996, MED PEDIATR ONCOL, V27, P32, DOI 10.1002/(SICI)1096-911X(199607)27:1<32::AID-MPO7>3.0.CO;2-V
   Packer RJ, 1997, J NEUROSURG, V86, P747, DOI 10.3171/jns.1997.86.5.0747
   Perilongo G, 1997, CANCER, V80, P142, DOI 10.1002/(SICI)1097-0142(19970701)80:1<142::AID-CNCR19>3.0.CO;2-Y
   POLLACK IF, 1995, J NEUROSURG, V82, P536, DOI 10.3171/jns.1995.82.4.0536
   POLLACK IF, 1994, CANCER, V73, P2869, DOI 10.1002/1097-0142(19940601)73:11<2869::AID-CNCR2820731134>3.0.CO;2-I
   Prados MD, 1997, J NEURO-ONCOL, V32, P235, DOI 10.1023/A:1005736104205
   SCOTT EW, 1987, PEDIATR NEUROSCI, V13, P225, DOI 10.1159/000120335
   SHAPIRO K, 1976, CHILD BRAIN, V2, P177
   WOO SY, 1988, J CLIN ONCOL, V6, P1001, DOI 10.1200/JCO.1988.6.6.1001
NR 27
TC 25
Z9 25
U1 0
U2 1
PU OXFORD UNIV PRESS INC
PI CARY
PA JOURNALS DEPT, 2001 EVANS RD, CARY, NC 27513 USA
SN 1522-8517
EI 1523-5866
J9 NEURO-ONCOLOGY
JI Neuro-Oncology
PD JUL
PY 2003
VL 5
IS 3
BP 188
EP 196
DI 10.1215/S1152851702000297
PG 9
WC Oncology; Clinical Neurology
WE Science Citation Index Expanded (SCI-EXPANDED)
SC Oncology; Neurosciences & Neurology
GA 688XN
UT WOS:000183462700006
PM 12816725
OA Green Published, Bronze
DA 2023-08-27
ER

PT J
AU Gao, YF
   Qin, ZG
   Li, DY
   Yu, WD
   Sun, LB
   Liu, NJ
   Zhao, CH
   Zhang, BT
   Hu, Y
   Sun, DJ
   Jin, XY
AF Gao, Yufei
   Qin, Zhigang
   Li, Dongyuan
   Yu, Weidong
   Sun, Libo
   Liu, Naijie
   Zhao, Conghai
   Zhang, Butian
   Hu, Yu
   Sun, Daju
   Jin, Xingyi
TI Intracerebral schwannoma: A case report and literature review
SO ONCOLOGY LETTERS
LA English
DT Review
DE incidence of a disease; intracerebral schwannoma; site of disease; case
   report; literature review
ID TEMPORAL-LOBE EPILEPSY; OF-THE-LITERATURE; MEDULLA-OBLONGATA;
   INTRAPARENCHYMAL SCHWANNOMAS; CYSTIC SCHWANNOMA; CHILD; NERVE; CELLS;
   TUMOR
AB Intracranial schwannoma accounts for between 5 and 8% of intracranial tumors, whereas intracerebral schwannoma, a rare disease, accounts for <1% of intracranial schwannomas. In addition to the present case report, a total of 84 cases reported within China and elsewhere were reviewed and summarized, and the age of the tumor onset, the site of disease, imaging results, clinical presentation, pathological classification and prognosis were analyzed. The present case report described a 12-year-old female with an intracerebral schwannoma in the brainstem, who was followed-up for 5 years using magnetic resonance imaging after a surgical resection without recurrence, and clinical symptoms were reported to have completely resolved. The incidence of intracerebral schwannoma was low among cases, and the correct diagnosis was not able to be made preoperatively, and the majority of cases were diagnosed on the basis of postoperative pathology. The majority of cases analyzed were supratentorial, occurring at an age <40 according to previous literature. In addition, 33% of patients presented with subtentorial schwannoma, occurring at an age >40. The prognosis was classified as good (patient can live independently) for the majority of patients if surgery was able to completely resect the lesion.
C1 [Gao, Yufei; Qin, Zhigang; Li, Dongyuan; Yu, Weidong; Sun, Libo; Liu, Naijie; Zhao, Conghai; Jin, Xingyi] Jilin Univ, China Japan Union Hosp, Dept Neurosurg, 126 Xiantai St, Changchun 130033, Jilin, Peoples R China.
   [Zhang, Butian] Jilin Univ, China Japan Union Hosp, Dept Radiol, Changchun 130033, Jilin, Peoples R China.
   [Hu, Yu; Sun, Daju] Jilin Univ, China Japan Union Hosp, Dept Pathol, Changchun 130033, Jilin, Peoples R China.
C3 Jilin University; Jilin University; Jilin University
RP Jin, XY (通讯作者)，Jilin Univ, China Japan Union Hosp, Dept Neurosurg, 126 Xiantai St, Changchun 130033, Jilin, Peoples R China.
EM
RI Yu, Wei/GZL-3831-2022; li, wei/IUQ-2973-2023; YU, Wei/HTP-9667-2023;
   SUN, Libo/G-7016-2019
OI SUN, Libo/0000-0002-3805-5870
CR AlBatly AA, 2014, PAN AFR MED J, V18, DOI 10.11604/pamj.2014.18.342.5075
   Anton T, 2006, J NEURO-ONCOL, V76, P307, DOI 10.1007/s11060-005-7286-y
   ARYANPUR J, 1988, J NEUROSURG, V69, P446, DOI 10.3171/jns.1988.69.3.0446
   AUER RN, 1982, J NEUROSURG, V56, P154, DOI 10.3171/jns.1982.56.1.0154
   BRUNER JM, 1984, J NEUROPATH EXP NEUR, V43, P296, DOI 10.1097/00005072-198405000-00020
   BRUNI P, 1984, SURG NEUROL, V22, P360, DOI 10.1016/0090-3019(84)90140-X
   CASADEL GP, 1993, J NEUROSURG, V79, P217, DOI 10.3171/jns.1993.79.2.0217
   Celikoglu E, 2007, J CLIN NEUROSCI, V14, P589, DOI 10.1016/j.jocn.2006.04.014
   Chelyshev Iu A, 2000, Usp Fiziol Nauk, V31, P54
   Consales A, 2010, BRIT J NEUROSURG, V24, P306, DOI 10.3109/02688691003624604
   de Andrade GC, 2002, ARQ NEURO-PSIQUIAT, V60, P308, DOI 10.1590/S0004-282X2002000200025
   Deng C, 1985, J NANTONG U, V1, P46
   DIBIASI C, 1994, AM J NEURORADIOL, V15, P1956
   Erongun U, 1996, NEUROSURG REV, V19, P269, DOI 10.1007/BF00314844
   EZURA M, 1992, NEUROSURGERY, V30, P97, DOI 10.1227/00006123-199201000-00017
   FEIGIN I, 1971, J NEUROPATH EXP NEUR, V30, P603, DOI 10.1097/00005072-197110000-00005
   FRIM DM, 1992, PEDIATR NEUROSURG, V18, P190, DOI 10.1159/000120661
   Ghatak N R, 1975, Surg Neurol, V3, P45
   GHOSH S, 1992, BRIT J NEUROSURG, V6, P163, DOI 10.3109/02688699209002921
   GIBSON AAM, 1966, J NEUROSURG, V24, P552, DOI 10.3171/jns.1966.24.2.0552
   GOKAY H, 1984, SURG NEUROL, V22, P69, DOI 10.1016/0090-3019(84)90232-5
   Guha D, 2012, J NEUROSURG, V117, P136, DOI 10.3171/2012.3.JNS111043
   HAHN JF, 1977, SOUTH MED J, V70, P539, DOI 10.1097/00007611-197705000-00010
   Hockley A D, 1975, Surg Neurol, V4, P509
   Ishihara M, 2009, PATHOL INT, V59, P583, DOI 10.1111/j.1440-1827.2009.02410.x
   KASANTIKUL V, 1981, J NEUROL NEUROSUR PS, V44, P1110, DOI 10.1136/jnnp.44.12.1110
   LADOUCEUR D, 1989, CAN J NEUROL SCI, V16, P357, DOI 10.1017/S0317167100029243
   Lee S, 2013, J KOREAN NEUROSURG S, V54, P340, DOI 10.3340/jkns.2013.54.4.340
   Li Q, NERVOUS SYSTEM TUMOR
   Lin JK, 2003, J NEUROSURG, V98, P621, DOI 10.3171/jns.2003.98.3.0621
   Ma L, 2013, J CRANIOFAC SURG, V24, pE541, DOI 10.1097/SCS.0b013e31828601bf
   Maiuri F, 2004, J NEURO-ONCOL, V66, P191, DOI 10.1023/B:NEON.0000013476.26986.7a
   Mirsky R, 2008, J PERIPHER NERV SYST, V13, P122, DOI 10.1111/j.1529-8027.2008.00168.x
   Muzzafar S, 2010, AM J NEURORADIOL, V31, P567, DOI 10.3174/ajnr.A1758
   NELSON E, 1970, BRAIN, V93, P475, DOI 10.1093/brain/93.3.475
   NEW PFJ, 1972, J NEUROSURG, V36, P795, DOI 10.3171/jns.1972.36.6.0795
   Osborn AG, 2007, DIAGNOSTIC IMAGING B
   Paredes I, 2012, CLIN NEUROL NEUROSUR, V114, P42, DOI 10.1016/j.clineuro.2011.07.003
   PIALAT J, 1975, LYON MED, V234, P129
   REDEKOP G, 1990, J NEUROSURG, V73, P777, DOI 10.3171/jns.1990.73.5.0777
   RODRIGUEZSALAZAR A, 1984, CHILD BRAIN, V11, P69
   Russel DS, 1989, PATHOLOGY TUMORS NER
   SARKAR C, 1987, J NEUROSURG, V67, P120, DOI 10.3171/jns.1987.67.1.0120
   Scherer SS, 1997, CURR OPIN NEUROL, V10, P386, DOI 10.1097/00019052-199710000-00006
   SCHWARTZ AM, 1988, SOUTH MED J, V81, P385, DOI 10.1097/00007611-198803000-00022
   SHALIT MN, 1982, ACTA NEUROCHIR, V64, P253, DOI 10.1007/BF01406060
   Sharma MC, 1996, J NEUROL NEUROSUR PS, V60, P200, DOI 10.1136/jnnp.60.2.200
   SHARMA RR, 1993, BRIT J NEUROSURG, V7, P83, DOI 10.3109/02688699308995061
   SHARMA V, 1993, BRIT J NEUROSURG, V7, P427, DOI 10.3109/02688699309103500
   Srinivas R, 2013, CASE REP NEUROL MED, V2013, DOI 10.1155/2013/171494
   STEFANKO SZ, 1986, ACTA NEUROPATHOL, V71, P321, DOI 10.1007/BF00688056
   TRANDINH HD, 1991, NEUROSURGERY, V29, P296, DOI 10.1227/00006123-199108000-00026
   Trotter J, 1993, Perspect Dev Neurobiol, V1, P149
   Tsuiki H, 1997, ACTA NEUROCHIR, V139, P756, DOI 10.1007/BF01420049
   VANRENSBURG MJ, 1975, J NEUROL NEUROSUR PS, V38, P703
   Wang H, 2000, JIANGSU MED J, V26, P869
   Wilson BR, 2016, WORLD NEUROSURG, V92, DOI 10.1016/j.wneu.2016.05.078
   Woodhoo A, 2008, GLIA, V56, P1481, DOI 10.1002/glia.20723
   Xu D, 2007, J RARE UNCOM DIS, V14, P7
   Yi Y, 2008, CASE SCHWANNOMA BRAI, V19, P300
   Zeng E, 2005, CHIN J MINIMA INVAS, V10, P473
   Zhao X, 1994, CHIN J NEUROSURG, V10, P280
   Zhu Q, 2010, ANHUI MED J, V31, P156
   许健, 2002, [中华神经外科杂志, Chinese Journal of Neurosurgery], V18, P86
   钟群, 2004, [中华放射学杂志, Chinese Journal of Radiology], V38, P1119
NR 65
TC 14
Z9 15
U1 2
U2 6
PU SPANDIDOS PUBL LTD
PI ATHENS
PA POB 18179, ATHENS, 116 10, GREECE
SN 1792-1074
EI 1792-1082
J9 ONCOL LETT
JI Oncol. Lett.
PD AUG
PY 2018
VL 16
IS 2
BP 2501
EP 2510
DI 10.3892/ol.2018.8949
PG 10
WC Oncology
WE Science Citation Index Expanded (SCI-EXPANDED)
SC Oncology
GA GT8EJ
UT WOS:000444768100043
PM 30013644
OA gold, Green Published
DA 2023-08-27
ER

PT J
AU Zhang, H
   Verkman, AS
AF Zhang, Hua
   Verkman, A. S.
TI Microfiberoptic Measurement of Extracellular Space Volume in Brain and
   Tumor Slices Based on Fluorescent Dye Partitioning
SO BIOPHYSICAL JOURNAL
LA English
DT Article
ID AQUAPORIN-4 WATER CHANNELS; MACROMOLECULE DIFFUSION; SPINAL-CORD;
   IN-VIVO; MICE; TORTUOSITY; LACKING; FRACTION; HIPPOCAMPUS; TRANSPORT
AB The fractional volume occupied by extracellular space in tissues, termed a, is an important parameter of tissue architecture that affects cellular functions and drug delivery. We report a technically simple fluorescent dye partitioning method to measure alpha in tissue slices based on microfiberoptic detection of dye fluorescence in tissue versus overlying solution. Microfiberoptic tip geometry and dyes were selected for alpha determination from fluorescence intensity ratios, without the need to correct for illumination profile, light scattering/absorption, or dye binding. The method was validated experimentally using cell-embedded gels of specified alpha-values and optical properties. In mouse brain slices, alpha was strongly location-dependent, ranging from 0.16 in thalamus to 0.22 in brainstem, and was sensitive to cell volume changes. Aquaporin-4 water channel gene deletion caused significant extracellular space expansion, with alpha = 0.181 +/- 0.002 in cortex in wild-type mice and 0.211 +/- 0.003 in Aquaporin-4 knockout mice. In slices of LLC1 cell tumors grown in mice to similar to 5 mm diameter, alpha decreased remarkably from similar to 0.45 in superficial tumor to <0.25 in deeper (>100 mu m) tumor. Fluorescent dye partitioning with microfiberoptic detection permits rapid, accurate, and anisotropy-insensitive determination of alpha-values in tissue slices.
C1 [Verkman, A. S.] Univ Calif San Francisco, Dept Med, San Francisco, CA 94143 USA.
   Univ Calif San Francisco, Dept Physiol, San Francisco, CA USA.
C3 University of California System; University of California San Francisco;
   University of California System; University of California San Francisco
RP Verkman, AS (通讯作者)，Univ Calif San Francisco, Dept Med, San Francisco, CA 94143 USA.
EM
FU National Institutes of Health [EB00415, DK35124, EY13574, DK86125,
   DK72517, HL73856]; Guthy-Jackson Charitable Foundation
FX This work was supported by grants EB00415, DK35124, EY13574, DK86125,
   DK72517, and HL73856 from the National Institutes of Health, and a grant
   from the Guthy-Jackson Charitable Foundation.
CR Appelgren L, 1973, Bibl Anat, V12, P504
   Binder DK, 2006, GLIA, V53, P631, DOI 10.1002/glia.20318
   Binder DK, 2004, J NEUROSCI, V24, P8049, DOI 10.1523/JNEUROSCI.2294-04.2004
   BLASBERG RG, 1975, J PHARMACOL EXP THER, V195, P73
   CHOI DW, 1988, NEURON, V1, P623, DOI 10.1016/0896-6273(88)90162-6
   Hrabe J, 2004, BIOPHYS J, V87, P1606, DOI 10.1529/biophysj.103.039495
   Hrabetova S, 2005, HIPPOCAMPUS, V15, P441, DOI 10.1002/hipo.20068
   JACKSON MJ, 1970, BIOCHIM BIOPHYS ACTA, V211, P425, DOI 10.1016/0005-2736(70)90248-8
   JAIN RK, 1987, CANCER RES, V47, P3039
   Jain RK, 1997, ADV DRUG DELIVER REV, V26, P71, DOI 10.1016/S0169-409X(97)00027-6
   JAKOBSEN I, 1995, MAGN RESON IMAGING, V13, P693, DOI 10.1016/0730-725X(95)00019-D
   Jin S, 2008, BIOPHYS J, V95, P1785, DOI 10.1529/biophysj.108.131466
   KARLSSON L, 1980, MICROVASC RES, V19, P71, DOI 10.1016/0026-2862(80)90084-9
   Krol A, 1999, CANCER RES, V59, P4136
   LAW RO, 1982, EXPERIENTIA, V38, P411, DOI 10.1007/BF01952615
   LEVIN VA, 1970, AM J PHYSIOL, V219, P1528, DOI 10.1152/ajplegacy.1970.219.5.1528
   Li J, 2002, INVEST OPHTH VIS SCI, V43, P573
   Li J, 2001, J BIOL CHEM, V276, P31233, DOI 10.1074/jbc.M104368200
   Lu DC, 2008, FASEB J, V22, P3216, DOI 10.1096/fj.07-104836
   Ma TH, 1997, J CLIN INVEST, V100, P957, DOI 10.1172/JCI231
   Magzoub M, 2008, FASEB J, V22, P276, DOI 10.1096/fj.07-9150com
   Magzoub M, 2009, BIOPHYS J, V96, P2382, DOI 10.1016/j.bpj.2008.12.3916
   Mazel T, 1998, NEUROREPORT, V9, P1299
   MCBAIN CJ, 1990, SCIENCE, V249, P674, DOI 10.1126/science.2382142
   Mori K, 1999, SCIENCE, V286, P711, DOI 10.1126/science.286.5440.711
   Nicholson C, 1998, TRENDS NEUROSCI, V21, P207, DOI 10.1016/S0166-2236(98)01261-2
   NICHOLSON C, 1981, J PHYSIOL-LONDON, V321, P225, DOI 10.1113/jphysiol.1981.sp013981
   Prokopova S, 1997, NEUROREPORT, V8, P3527, DOI 10.1097/00001756-199711100-00022
   RICE ME, 1991, J NEUROPHYSIOL, V65, P264, DOI 10.1152/jn.1991.65.2.264
   ROSENBERG LE, 1962, AM J PHYSIOL, V202, P800, DOI 10.1152/ajplegacy.1962.202.4.800
   Simonova Z, 1996, PHYSIOL RES, V45, P11
   Sykova E, 2004, NEUROCHEM INT, V45, P453, DOI 10.1016/j.neuint.2003.11.009
   Sykova E, 2008, PHYSIOL REV, V88, P1277, DOI 10.1152/physrev.00027.2007
   Thiagarajah JR, 2006, NAT METHODS, V3, P275, DOI 10.1038/NMETH863
   Vargova L, 2003, GLIA, V42, P77, DOI 10.1002/glia.10204
   Verkman AS, 2006, BBA-BIOMEMBRANES, V1758, P1085, DOI 10.1016/j.bbamem.2006.02.018
   Yao XM, 2008, J NEUROSCI, V28, P5460, DOI 10.1523/JNEUROSCI.0257-08.2008
   Zador Z, 2008, FASEB J, V22, P870, DOI 10.1096/fj.07-9468com
   Zamecnik J, 2004, NEUROPATH APPL NEURO, V30, P338, DOI 10.1046/j.1365-2990.2003.0541.x
NR 39
TC 36
Z9 37
U1 0
U2 9
PU CELL PRESS
PI CAMBRIDGE
PA 50 HAMPSHIRE ST, FLOOR 5, CAMBRIDGE, MA 02139 USA
SN 0006-3495
EI 1542-0086
J9 BIOPHYS J
JI Biophys. J.
PD AUG 18
PY 2010
VL 99
IS 4
BP 1284
EP 1291
DI 10.1016/j.bpj.2010.06.023
PG 8
WC Biophysics
WE Science Citation Index Expanded (SCI-EXPANDED)
SC Biophysics
GA 641CW
UT WOS:000281103200034
PM 20713014
OA Green Published, hybrid
DA 2023-08-27
ER

PT J
AU Tate, MC
   Lindquist, RA
   Nguyen, T
   Sanai, N
   Barkovich, AJ
   Huang, EJ
   Rowitch, DH
   Alvarez-Buylla, A
AF Tate, Matthew C.
   Lindquist, Robert A.
   Thuhien Nguyen
   Sanai, Nader
   Barkovich, A. James
   Huang, Eric J.
   Rowitch, David H.
   Alvarez-Buylla, Arturo
TI Postnatal Growth of the Human Pons: A Morphometric and
   Immunohistochemical Analysis
SO JOURNAL OF COMPARATIVE NEUROLOGY
LA English
DT Article
DE brainstem; pediatric; development; basis; pontine glioma;
   nif-0000-00217; SciRes_000114; AB_304558; AB_2109815; AB_442102;
   AB_396287; AB_92396; AB_91107; AB_291466; AB_2336877; AB_2336878;
   AB_261856; AB_2304493
ID INTRINSIC PONTINE GLIOMAS; NERVOUS-SYSTEM MYELINATION; HUMAN INFANCY;
   STEM-CELLS; NUCLEI; MEDULLOBLASTOMA; CLASSIFICATION; VIMENTIN; PATTERNS;
   SEQUENCE
AB Despite its critical importance to global brain function, the postnatal development of the human pons remains poorly understood. In the present study, we first performed magnetic resonance imaging (MRI)-based morphometric analyses of the postnatal human pons (0-18 years; n = 6-14/timepoint). Pons volume increased 6-fold from birth to 5 years, followed by continued slower growth throughout childhood. The observed growth was primarily due to expansion of the basis pontis. T2-based MRI analysis suggests that this growth is linked to increased myelination, and histological analysis of myelin basic protein in human postmortem specimens confirmed a dramatic increase in myelination during infancy. Analysis of cellular proliferation revealed many Ki67(+) cells during the first 7 months of life, particularly during the first month, where proliferation was increased in the basis relative to tegmentum. The majority of proliferative cells in the postnatal pons expressed the transcription factor Olig2, suggesting an oligodendrocyte lineage. The proportion of proliferating cells that were Olig2(+) was similar through the first 7 months of life and between basis and tegmentum. The number of Ki67(+) cells declined dramatically from birth to 7 months and further decreased by 3 years, with a small number of Ki67(+) cells observed throughout childhood. In addition, two populations of vimentin/nestin-expressing cells were identified: a dorsal group near the ventricular surface, which persists throughout childhood, and a parenchymal population that diminishes by 7 months and was not evident later in childhood. Together, our data reveal remarkable postnatal growth in the ventral pons, particularly during infancy when cells are most proliferative and myelination increases. J. Comp. Neurol. 523:449-462, 2015. (c) 2014 Wiley Periodicals, Inc.
C1 [Tate, Matthew C.; Lindquist, Robert A.; Thuhien Nguyen; Rowitch, David H.; Alvarez-Buylla, Arturo] Univ Calif San Francisco, Eli & Edythe Broad Ctr Regenerat Med & Stem Cell, San Francisco, CA 94143 USA.
   [Tate, Matthew C.; Thuhien Nguyen; Rowitch, David H.; Alvarez-Buylla, Arturo] Univ Calif San Francisco, Dept Neurol Surg, San Francisco, CA 94143 USA.
   [Lindquist, Robert A.; Huang, Eric J.; Rowitch, David H.; Alvarez-Buylla, Arturo] Univ Calif San Francisco, Neurosci Grad Program, San Francisco, CA 94143 USA.
   [Lindquist, Robert A.] Univ Calif San Francisco, Sch Med, Med Scientist Training Program, San Francisco, CA 94143 USA.
   [Sanai, Nader] Barrow Neurol Inst, Dept Neurol Surg, Phoenix, AZ 85013 USA.
   [Barkovich, A. James] Univ Calif San Francisco, Dept Radiol & Biomed Imaging, Sect Pediat Neuroradiol, San Francisco, CA 94143 USA.
   [Huang, Eric J.] Univ Calif San Francisco, Dept Pathol, San Francisco, CA 94143 USA.
   [Rowitch, David H.] Univ Calif San Francisco, Howard Hughes Med Inst, San Francisco, CA 94143 USA.
   [Rowitch, David H.] Univ Calif San Francisco, Dept Pediat, San Francisco, CA 94143 USA.
C3 University of California System; University of California San Francisco;
   University of California System; University of California San Francisco;
   University of California System; University of California San Francisco;
   University of California System; University of California San Francisco;
   Barrow Neurological Institute; University of California System;
   University of California San Francisco; University of California System;
   University of California San Francisco; Howard Hughes Medical Institute;
   University of California System; University of California San Francisco;
   University of California System; University of California San Francisco
RP Alvarez-Buylla, A (通讯作者)，Univ Calif San Francisco, Dept Neurol Surg, 35 Med Ctr Way RMB 1038,Box 0525, San Francisco, CA 94143 USA.
EM
RI Huang, Eric J./AAH-7997-2019
OI Huang, Eric J./0000-0002-5381-3801; Rowitch, David/0000-0002-0079-0060;
   Alvarez-Buylla, Arturo/0000-0003-4426-8925; Lindquist,
   Robert/0000-0003-4746-0605
FU National Institutes of Health [NS28478]; John G. Bowes Research Fund
   [NRSA 1F32NS067889-01A1]; Pediatric Brain Tumor Foundation; University
   of California [142675]
FX Grant sponsor: National Institutes of Health; Grant number: NS28478;
   Grant sponsor: John G. Bowes Research Fund (to A.A.-B.); Grant number:
   NRSA 1F32NS067889-01A1 (to M.T.); Grant sponsor: Pediatric Brain Tumor
   Foundation (to A.A.-B., D.H.R.), University of California; Grant number:
   MRPI #142675 (to E.J.H.); Grant sponsor: D.H.R. is an HHMI Investigator.
CR Abe S, 2004, PRENATAL DIAG, V24, P352, DOI 10.1002/pd.873
   ADAMS CE, 1980, BRAIN RES BULL, V5, P277, DOI 10.1016/0361-9230(80)90169-0
   Ballester LY, 2013, AM J SURG PATHOL, V37, P1357, DOI 10.1097/PAS.0b013e318294e817
   Barkovich AJ, 2009, BRAIN, V132, P3199, DOI 10.1093/brain/awp247
   Barrow J, 2011, NEURO-ONCOLOGY, V13, P212, DOI 10.1093/neuonc/noq158
   BOHN W, 1992, EXP CELL RES, V201, P1, DOI 10.1016/0014-4827(92)90341-5
   BRODY BA, 1987, J NEUROPATH EXP NEUR, V46, P283, DOI 10.1097/00005072-198705000-00005
   DEBUS E, 1983, DIFFERENTIATION, V25, P193, DOI 10.1111/j.1432-0436.1984.tb01355.x
   Essick CR, 1912, AM J ANAT, V13, P25, DOI 10.1002/aja.1000130103
   Fangusaro J, 2009, J CHILD NEUROL, V24, P1409, DOI 10.1177/0883073809338960
   Fischbein NJ, 1996, PEDIATR NEUROSURG, V24, P9, DOI 10.1159/000121010
   Gibson P, 2010, NATURE, V468, P1095, DOI 10.1038/nature09587
   Grill J, 2012, PEDIATR BLOOD CANCER, V58, P489, DOI 10.1002/pbc.24060
   Hatta Toshihisa, 2007, Congenital Anomalies, V47, P63, DOI 10.1111/j.1741-4520.2007.00145.x
   KINNEY HC, 1988, J NEUROPATH EXP NEUR, V47, P217, DOI 10.1097/00005072-198805000-00003
   Ligon KL, 2006, P NATL ACAD SCI USA, V103, P7853, DOI 10.1073/pnas.0511001103
   Ligon KL, 2004, J NEUROPATH EXP NEUR, V63, P499, DOI 10.1093/jnen/63.5.499
   Messam CA, 2000, EXP NEUROL, V161, P585, DOI 10.1006/exnr.1999.7319
   MIHAILOFF GA, 1984, J COMP NEUROL, V222, P116, DOI 10.1002/cne.902220110
   Monje M, 2011, P NATL ACAD SCI USA, V108, P4453, DOI 10.1073/pnas.1101657108
   NOZAKI H, 1992, DEV BRAIN RES, V65, P13, DOI 10.1016/0165-3806(92)90003-F
   Paugh BS, 2011, J CLIN ONCOL, V29, P3999, DOI 10.1200/JCO.2011.35.5677
   Puget S, 2012, PLOS ONE, V7, DOI 10.1371/journal.pone.0030313
   Quinones-Hinojosa A, 2006, J COMP NEUROL, V494, P415, DOI 10.1002/cne.20798
   Roig B, 2010, BRAIN RES, V1336, P22, DOI 10.1016/j.brainres.2010.03.099
   Saladin K. S., 2012, ANATOMY PHYSL UNITY
   Schuller U, 2008, CANCER CELL, V14, P123, DOI 10.1016/j.ccr.2008.07.005
   STATHOPOULOS E, 1989, J HISTOCHEM CYTOCHEM, V37, P1363, DOI 10.1177/37.9.2671152
   Taylor MD, 2005, CANCER CELL, V8, P323, DOI 10.1016/j.ccr.2005.09.001
   Wagner S, 2006, J NEURO-ONCOL, V79, P281, DOI 10.1007/s11060-006-9133-1
   Wu G, 2012, NAT GENET, V44, P251, DOI 10.1038/ng.1102
   Zarghooni M, 2010, J CLIN ONCOL, V28, P1337, DOI 10.1200/JCO.2009.25.5463
NR 32
TC 28
Z9 28
U1 0
U2 8
PU WILEY-BLACKWELL
PI HOBOKEN
PA 111 RIVER ST, HOBOKEN 07030-5774, NJ USA
SN 0021-9967
EI 1096-9861
J9 J COMP NEUROL
JI J. Comp. Neurol.
PD FEB 15
PY 2015
VL 523
IS 3
BP 449
EP 462
DI 10.1002/cne.23690
PG 14
WC Neurosciences; Zoology
WE Science Citation Index Expanded (SCI-EXPANDED)
SC Neurosciences & Neurology; Zoology
GA AW7WR
UT WOS:000346472800006
PM 25307966
OA Green Accepted, Green Submitted
DA 2023-08-27
ER

PT J
AU Ushio, M
   Murofushi, T
   Iwasaki, S
AF Ushio, Munetaka
   Murofushi, Toshihisa
   Iwasaki, Shinichi
TI Subjective visual horizontal in patients with posterior canal benign
   paroxysmal positional vertigo
SO ACTA OTO-LARYNGOLOGICA
LA English
DT Article
DE benign paroxysmal positional vertigo; subjective visual horizontal;
   ocular tilt reaction; utricle; utricular macula; vestibular;
   cellebello-pontine angle
ID UNILATERAL VESTIBULAR NEURECTOMY; OCULAR TILT REACTION; ORIENTATION;
   MORPHOLOGY; DEVIATION; NEURONS; ROLL; CAT
AB Conclusion. Patients with posterior canal benign paroxysmal positional vertigo (p-BPPV) could have mild excitation in the vestibular system on the affected side. Objective. p-BPPV is considered to be caused by small particles dislodged from the utricular macula. While subjective visual horizontal (SVH) mainly reflects utricular function, there is less documentation on SVH in patients with p-BPPV. The purpose of this study was to clarify whether p-BPPV could affect SVH. Patients and methods. Twenty-eight patients with unilateral idiopathic p-BPPV were enrolled. For comparison, 17 patients with vestibular neuritis and 45 patients with cerebello-pontine angle (CPA) tumor were also examined. For measurement of SVH, we used a device that has a red bar of light-emitting diodes (LEDs) in front of the patient. Subjects were asked to rotate the bar to the position that they felt was horizontal in a completely dark room. Results. Deviation of SVH toward the unaffected side down was seen in 82.1% (23/28) of patients with p-BPPV. Mean +/- SD deviation of SVH from the true horizontal to the unaffected side down was 0.49 +/- 1.2 1 degrees. Conversely, deviation of SVH was toward the affected side down in 88.2% of patients with vestibular neuritis and in 75.6% of patients with CPA tumor.
C1 Univ Tokyo, Fac Med, Dept Otolaryngol, Bunkyo Ku, Tokyo 1138655, Japan.
   Tokyo Postal Serv Agcy Hosp, Dept Otolaryngol, Tokyo, Japan.
C3 University of Tokyo
RP Ushio, M (通讯作者)，Univ Tokyo, Fac Med, Dept Otolaryngol, Bunkyo Ku, 7-3-1 Hongou, Tokyo 1138655, Japan.
EM
RI Ushio, Munetaka/S-3598-2016; murofushi, toshihisa/Q-1159-2019
OI Ushio, Munetaka/0000-0001-9051-634X; 
CR Bohmer A, 1995, J Vestib Res, V5, P35
   BRANDT T, 1987, BRAIN, V110, P649, DOI 10.1093/brain/110.3.649
   BRANDT T, 1993, ANN NEUROL, V33, P528, DOI 10.1002/ana.410330518
   CURTHOYS IS, 1991, ACTA OTO-LARYNGOL, P5
   DAI MJ, 1989, EXP BRAIN RES, V77, P315, DOI 10.1007/BF00274989
   FRIEDMAN.G, 1971, ACTA OTO-LARYNGOL, V71, P289, DOI 10.3109/00016487109125366
   Gall RM, 1999, J OTOLARYNGOL, V28, P162
   GRAF W, 1983, EXP BRAIN RES, V52, P125
   GRAF W, 1986, EXP BRAIN RES, V63, P35
   Halmagyi GM, 1997, ADV OTO-RHINO-LARYNG, V53, P132
   HALMAGYI GM, 1979, ANN NEUROL, V6, P80, DOI 10.1002/ana.410060122
   SCHONE H, 1964, AEROSPACE MED, V35, P764
   Tabak S, 1997, ACTA OTO-LARYNGOL, V117, P1, DOI 10.3109/00016489709117982
   Van Nechel C, 2001, ADV OTO-RHINO-LARYNG, V58, P77
   von Brevern T, 2006, OTOL NEUROTOL, V27, P92, DOI 10.1097/01.mao.0000187238.56583.9b
NR 15
TC 10
Z9 10
U1 0
U2 1
PU TAYLOR & FRANCIS LTD
PI ABINGDON
PA 2-4 PARK SQUARE, MILTON PARK, ABINGDON OR14 4RN, OXON, ENGLAND
SN 0001-6489
EI 1651-2251
J9 ACTA OTO-LARYNGOL
JI Acta Oto-Laryngol.
PY 2007
VL 127
IS 8
BP 836
EP 838
DI 10.1080/00016480601053115
PG 3
WC Otorhinolaryngology
WE Science Citation Index Expanded (SCI-EXPANDED)
SC Otorhinolaryngology
GA 202CR
UT WOS:000248880100009
PM 17762994
DA 2023-08-27
ER

PT J
AU Moser, B
   Janik, S
   Schiefer, AI
   Mullauer, L
   Bekos, C
   Scharrer, A
   Mildner, M
   Renyi-Vamos, F
   Klepetko, W
   Ankersmit, HJ
AF Moser, Bernhard
   Janik, Stefan
   Schiefer, Ana-Iris
   Muellauer, Leonhard
   Bekos, Christine
   Scharrer, Anke
   Mildner, Michael
   Renyi-Vamos, Ferenc
   Klepetko, Walter
   Ankersmit, Hendrik Jan
TI Expression of RAGE and HMGB1 in Thymic Epithelial Tumors, Thymic
   Hyperplasia and Regular Thymic Morphology
SO PLOS ONE
LA English
DT Article
ID GLYCATION END-PRODUCTS; MYASTHENIA-GRAVIS; SOLUBLE RECEPTOR; THERAPEUTIC
   TARGET; DENDRITIC CELLS; T-CELLS; PATHOGENESIS; THYMOMA; CLASSIFICATION;
   BLOCKADE
AB Recently, a role of the receptor for advanced glycation endproducts (RAGE) in myasthenia gravis was described. RAGE and its ligand high mobility group box 1 (HMGB1) play key roles in autoimmunity and cancer. To test whether these molecules are involved in patients with thymic abnormalities we applied immunohistochemical analysis in 33 cases of thymic epithelial tumors, comprising 27 thymomas and 6 thymic carcinomas, and 21 nonneoplastic thymuses. Both molecules were detected in neoplastic epithelial cells: RAGE staining was most intense in WHO type B2 thymomas and thymic carcinomas (p<0.001). HMGB1 nuclear staining was strongest in A and AB, and gradually less in B1 = B2>B3> thymic carcinoma (p<0.001). Conversely, HMGB1 cytoplasmic staining intensities were as follows: A and AB (none), B1 (strong), B2 (moderate), B3 and thymic carcinoma (weak); (p<0.001). Fetal thymic tissue showed a distinct expression of RAGE and HMGB1 in subcapsular cortical epithelial cells which was found in 50% of myasthenic patients. Furthermore RAGE and HMGB1 were expressed in thymocytes, macrophages, Hassall's corpuscles, thymic medulla, and germinal center cells in myasthenic patients. Immunohistochemistry results were complemented by systemic measurements (immunosorbent assay): serum levels of soluble RAGE were significantly reduced in patients with epithelial tumors (p = 0.008); and in invasive tumors (p = 0.008). Whereas RAGE was equally reduced in thymic hyperplasia and epithelial tumors (p = 0.003), HMGB1 was only elevated in malignancies (p = 0.036). Results were most pronounced in thymic carcinomas. Thus, RAGE and HMGB1 are involved in the (patho-) physiology of thymus, as evidenced by differentiated thymic and systemic expression patterns that may act as diagnostic or therapeutic targets in autoimmune disease and cancer.
C1 [Moser, Bernhard; Janik, Stefan; Bekos, Christine; Klepetko, Walter; Ankersmit, Hendrik Jan] Med Univ Vienna, Div Surg, Dept Thorac Surg, Vienna, Austria.
   [Schiefer, Ana-Iris; Muellauer, Leonhard; Scharrer, Anke] Med Univ Vienna, Dept Pathol, Vienna, Austria.
   [Bekos, Christine; Ankersmit, Hendrik Jan] Med Univ Vienna, Christian Doppler Lab Diag & Regenerat Cardiac &, Vienna, Austria.
   [Mildner, Michael] Med Univ Vienna, Dept Dermatol, Vienna, Austria.
   [Renyi-Vamos, Ferenc] Natl Inst Oncol, Dept Gen & Thorac Surg, Budapest, Hungary.
C3 Medical University of Vienna; Medical University of Vienna; Medical
   University of Vienna; Medical University of Vienna; National Institute
   of Oncology Hungary
RP Moser, B (通讯作者)，Med Univ Vienna, Div Surg, Dept Thorac Surg, Vienna, Austria.
EM
RI Schiefer, Ana-Iris/AAX-8123-2020; Müllauer, Leonhard/GSN-9884-2022;
   Moser, Bernhard/ABA-9436-2020
OI Müllauer, Leonhard/0000-0002-3399-078X; Moser,
   Bernhard/0000-0002-6037-5475; Bekos, Christine/0000-0003-4038-6860;
   Ankersmit, Hendrik Jan/0000-0002-8761-3517; Mildner,
   Michael/0000-0002-6892-925X
FU FOLAB - Research Laboratories of the Department of Surgery, Medical
   University Vienna; Christian Doppler Laboratory for the Diagnosis and
   Regeneration of Cardiac and Thoracic Diseases
FX Funding sources: FOLAB - Research Laboratories of the Department of
   Surgery, Medical University Vienna. Christian Doppler Laboratory for the
   Diagnosis and Regeneration of Cardiac and Thoracic Diseases. The funders
   had no role in study design, data collection and analysis, decision to
   publish, or preparation of the manuscript.
CR Bergeron S, 2005, J BIOL CHEM, V280, P31314, DOI 10.1074/jbc.M503063200
   BURNET M, 1957, BRIT MED J, V1, P779, DOI 10.1136/bmj.1.5022.779
   Calder AE, 2011, NEUROIMMUNOMODULAT, V18, P281, DOI 10.1159/000329496
   Castleman B., 1955, ATLAS TUMOR PATHOLOG, V19
   Cataldegirmen G, 2005, J EXP MED, V201, P473, DOI 10.1084/jem.20040934
   Cavalcante P, 2011, MUSCLE NERVE, V44, P467, DOI 10.1002/mus.22103
   Chen G, 2002, CANCER, V95, P420, DOI 10.1002/cncr.10665
   COSGROVE D, 1992, INT IMMUNOL, V4, P707, DOI 10.1093/intimm/4.6.707
   Detterbeck F, 2010, J THORAC ONCOL, V5, P1, DOI 10.1097/JTO.0b013e3181c59224
   Detterbeck FC, 2011, J THORAC ONCOL, V6, pS1710, DOI 10.1097/JTO.0b013e31821e8cff
   Dougan M, 2008, J EXP MED, V205, P267, DOI 10.1084/jem.20080136
   Duwe BV, 2005, CHEST, V128, P2893, DOI 10.1378/chest.128.4.2893
   Engels EA, 2010, J THORAC ONCOL, V5, pS260, DOI 10.1097/JTO.0b013e3181f1f62d
   Fujii Y, 2011, THORAC SURG CLIN, V21, P125, DOI 10.1016/j.thorsurg.2010.08.002
   Gao Ke, 2008, Zhonghua Yixue Yichuanxue Zazhi, V25, P311
   Halayko AJ, 2009, CAN J PHYSIOL PHARM, V87, P743, DOI 10.1139/Y09-054
   Hiwatashi K, 2008, ANN SURG ONCOL, V15, P923, DOI 10.1245/s10434-007-9698-8
   Hofmann MA, 2002, GENES IMMUN, V3, P123, DOI 10.1038/sj.gene.6363861
   Hohlfeld R, 2008, J NEUROIMMUNOL, V201, P21, DOI 10.1016/j.jneuroim.2008.05.020
   Imai K, 2000, LANCET, V356, P1795, DOI 10.1016/S0140-6736(00)03231-1
   Jablonska-Mestanova V, 2013, BRATISL MED J, V114, P402, DOI 10.4149/BLL_2013_086
   Jiao L, 2011, CANCER EPIDEM BIOMAR, V20, P1430, DOI 10.1158/1055-9965.EPI-11-0066
   Jiao L, 2011, CANCER RES, V71, P3582, DOI 10.1158/0008-5472.CAN-10-2573
   Johnson SB, 2001, ONCOLOGIST, V6, P239, DOI 10.1634/theoncologist.6-3-239
   Kalea AZ, 2011, FRONT BIOSCI-LANDMRK, V16, P2756, DOI 10.2741/3884
   Kang R, 2013, CLIN CANCER RES, V19, P4046, DOI 10.1158/1078-0432.CCR-13-0495
   KONDO K, 1990, AM J SURG PATHOL, V14, P1139, DOI 10.1097/00000478-199012000-00006
   Kuniyasu H, 2003, ONCOL REP, V10, P445
   Lara PN, 2000, CANCER TREAT REV, V26, P127
   LE PT, 1991, J EXP MED, V174, P1147, DOI 10.1084/jem.174.5.1147
   Leclerc Estelle, 2010, Cardiovasc Psychiatry Neurol, V2010, P539581, DOI 10.1155/2010/539581
   Li Ronghua, 2010, Lin Chung Er Bi Yan Hou Tou Jing Wai Ke Za Zhi, V24, P823
   Lin LG, 2012, J CANCER RES CLIN, V138, P11, DOI 10.1007/s00432-011-1067-0
   Little AJ, 2013, NUCLEIC ACIDS RES, V41, P3289, DOI 10.1093/nar/gks1461
   Livesey KM, 2012, CANCER RES, V72, P1996, DOI 10.1158/0008-5472.CAN-11-2291
   Ma CY, 2012, SCAND J IMMUNOL, V75, P614, DOI 10.1111/j.1365-3083.2012.02691.x
   Marx A, 2013, AUTOIMMUN REV, V12, P875, DOI 10.1016/j.autrev.2013.03.007
   Miyata T, 1996, J CLIN INVEST, V98, P1088, DOI 10.1172/JCI118889
   Moser B, 2007, AM J TRANSPLANT, V7, P293, DOI 10.1111/j.1600-6143.2006.01617.x
   Moser B, 2007, J IMMUNOL, V179, P8051, DOI 10.4049/jimmunol.179.12.8051
   Moser B, 2014, THORAC CARDIOV SURG, V62, P153, DOI 10.1055/s-0032-1322611
   Moser B, 2012, BIOCHEM BIOPH RES CO, V420, P96, DOI 10.1016/j.bbrc.2012.02.121
   NEEPER M, 1992, J BIOL CHEM, V267, P14998
   Omatsu M, 2012, GEN THORAC CARDIOVAS, V60, P803, DOI 10.1007/s11748-012-0160-x
   PAPATESTAS AE, 1971, AM J MED, V50, P465, DOI 10.1016/0002-9343(71)90336-6
   Papoudou-Bai A, 2012, IN VIVO, V26, P985
   Pisetsky DS, 2008, ARTHRITIS RES THER, V10, DOI 10.1186/ar2440
   Pullerits R, 2005, ARTHRITIS RES THER, V7, pR817, DOI 10.1186/ar1749
   Raica M, 2007, Rom J Morphol Embryol, V48, P101
   Raica M, 2006, ANN ANAT, V188, P345, DOI 10.1016/j.aanat.2006.01.012
   Ramasamy R, 2009, J LEUKOCYTE BIOL, V86, P505, DOI 10.1189/jlb.0409230
   Roithmaier S, 2007, J HEART LUNG TRANSPL, V26, P845, DOI 10.1016/j.healun.2007.05.019
   Rosai J, 1976, ATLAS TUMOR PATHOLOG
   Ruffini E, 2011, J THORAC ONCOL, V6, P614, DOI 10.1097/JTO.0b013e318207cd74
   Sasahira T, 2005, VIRCHOWS ARCH, V446, P411, DOI 10.1007/s00428-005-1210-x
   Savchenko AS, 2006, CELL TISSUE RES, V325, P455, DOI 10.1007/s00441-006-0202-8
   SCHMIDT AM, 1992, J BIOL CHEM, V267, P14987
   Sims GP, 2010, ANNU REV IMMUNOL, V28, P367, DOI 10.1146/annurev.immunol.021908.132603
   Sims GP, 2001, J IMMUNOL, V167, P1935, DOI 10.4049/jimmunol.167.4.1935
   Sparvero LJ, 2009, J TRANSL MED, V7, DOI 10.1186/1479-5876-7-17
   Strobel P, 2005, SURG TODAY, V35, P805, DOI 10.1007/s00595-005-3047-y
   Tafani M, 2011, J NEUROINFLAMM, V8, DOI 10.1186/1742-2094-8-32
   Taguchi A, 2000, NATURE, V405, P354, DOI 10.1038/35012626
   THOMAS L, 1982, YALE J BIOL MED, V55, P329
   Urbonaviciute V, 2011, J INTERN MED, V270, P309, DOI 10.1111/j.1365-2796.2011.02432.x
   Vincent A, 2002, NAT REV IMMUNOL, V2, P797, DOI 10.1038/nri916
   Wang XL, 2012, PLOS ONE, V7, DOI [10.1371/journal.pone.0036663, 10.1371/journal.pone.0040711]
   Watanabe N, 2005, NATURE, V436, P1181, DOI 10.1038/nature03886
   Weksler B, 2012, ANN THORAC SURG, V93, P884, DOI 10.1016/j.athoracsur.2011.05.089
   Xu XC, 2013, INT J IMMUNOPATH PH, V26, P485, DOI 10.1177/039463201302600222
   Yan SS, 2003, NAT MED, V9, P287, DOI 10.1038/nm831
   Zhu XJ, 2013, CELL BIOCHEM BIOPHYS, V66, P843, DOI 10.1007/s12013-013-9515-x
   Zuckerman NS, 2010, EUR J IMMUNOL, V40, P1150, DOI 10.1002/eji.200939914
NR 73
TC 27
Z9 30
U1 0
U2 10
PU PUBLIC LIBRARY SCIENCE
PI SAN FRANCISCO
PA 1160 BATTERY STREET, STE 100, SAN FRANCISCO, CA 94111 USA
SN 1932-6203
J9 PLOS ONE
JI PLoS One
PD APR 4
PY 2014
VL 9
IS 4
AR e94118
DI 10.1371/journal.pone.0094118
PG 14
WC Multidisciplinary Sciences
WE Science Citation Index Expanded (SCI-EXPANDED)
SC Science & Technology - Other Topics
GA AE6MJ
UT WOS:000334107500125
PM 24705787
OA gold, Green Published, Green Submitted
DA 2023-08-27
ER

PT J
AU Brady, S
   Lechan, RM
   Schwaitzberg, SD
   Dayal, Y
   Ziar, J
   Tischler, AS
AF Brady, S
   Lechan, RM
   Schwaitzberg, SD
   Dayal, Y
   Ziar, J
   Tischler, AS
TI Composite pheochromocytoma/ganglioneuroma of the adrenal gland
   associated with multiple endocrine neoplasia 2A - Case report with
   immunohistochemical analysis
SO AMERICAN JOURNAL OF SURGICAL PATHOLOGY
LA English
DT Article
DE composite adrenal medullary tumor; multiple neoplasia; pheochromocytoma;
   ganglioneuroma; tyrosine hydroxylase; phenylethanolamine;
   N-methyltransferase; catecholamine; Ki-67; MIB-1; neuron restrictive
   silencing factor; REST; transdifferentiation
ID VASOACTIVE INTESTINAL PEPTIDE; RET PROTOONCOGENE; ANTIGEN RETRIEVAL;
   NEUROFIBROMATOSIS; MUTATIONS; FEATURES; DISEASE; TYPE-2A; PROTEIN;
   TUMORS
AB We report a case of composite pheochromocytoma/ganglioneuroma arising in a background of diffuse and nodular medullary hyperplasia in the adrenal gland of a 34-year-old man with multiple endocrine neoplasia 2a (MEN 2a). Cells were histologically classified as chromaffin or chromaffin-like (small typical-appearing pheochromocytoma cells), neuron-like (possessing ganglion cell morphology), and intermediate. We speculate that these cell types may represent a spectrum of differentiation of a neoplastic clone, with the intermediate cells representing a transitional stage between chromaffin cells and neurons. All three cell types in the composite tumor and all chromaffin cells in both nodular and nonnodular areas of the remaining medulla were strongly immunoreactive for tyrosine hydroxylase, the rate-limiting enzyme in catecholamine synthesis. In contrast, neuron-like cells (and to a variable extent intermediate cells) displayed selective loss of expression of phenylethanolamine-N-methyltransferase (PNMT), the enzyme that synthesizes epinephrine. Proliferative activity of the composite tumor and both the nodular and nonnodular medulla was studied by staining for the endogenous cell proliferation antigen Ki-67, using monoclonal antibody MIB-1. MIB-1 labeling was highest in Schwann cell areas of the composite tumor, followed by chromaffin-like cells in the composite tumor and in the separate nodules. Labeling was absent in neuron-like cells, consistent with the cells' postulated status as terminally differentiated derivatives of a chromaffin cell precursor, and was highly variable in nonnodular areas of the medulla. The latter observation suggests topographical variation in signals that drive chromaffin cell proliferation in MEN.
C1 TUFTS UNIV,SCH MED,DEPT INTERNAL MED,BOSTON,MA 02111.
   TUFTS UNIV,SCH MED,DEPT SURG,BOSTON,MA 02111.
C3 Tufts University; Tufts University
RP Brady, S (通讯作者)，TUFTS UNIV,SCH MED,DEPT PATHOL,750 WASHINGTON ST,BOSTON,MA 02111, USA.
OI Tischler, Arthur/0000-0002-9726-5739
FU NCI NIH HHS [CA48017, CA27808] Funding Source: Medline
CR ABE H, 1985, ENDOCRINOLOGY, V116, P1383, DOI 10.1210/endo-116-4-1383
   BOLANDE RP, 1974, HUM PATHOL, V5, P409, DOI 10.1016/S0046-8177(74)80021-3
   CHETTY R, 1993, AM J SURG PATHOL, V17, P837, DOI 10.1097/00000478-199308000-00009
   CHONG JHA, 1995, CELL, V80, P949, DOI 10.1016/0092-8674(95)90298-8
   CHOUTET P, 1981, SEM HOP PARIS, V57, P590
   DELELLIS RA, 1995, LAB INVEST, V72, P494
   Donadio F, 1982, Minerva Chir, V37, P551
   DONISKELLER H, 1993, HUM MOL GENET, V2, P851, DOI 10.1093/hmg/2.7.851
   EVINGER M, 1995, 8 INT S CHROM CELL B
   GAGEL RF, 1988, NEW ENGL J MED, V318, P478, DOI 10.1056/NEJM198802253180804
   GUTMANN DH, 1993, NEURON, V10, P335, DOI 10.1016/0896-6273(93)90324-K
   KIMURA N, 1994, ENDOCR PATHOL, V5, P223, DOI 10.1007/BF02921490
   LINNOILA RI, 1990, HUM PATHOL, V21, P1168
   LODA M, 1994, HUM PATHOL, V25, P1050, DOI 10.1016/0046-8177(94)90064-7
   MOORE PJ, 1995, SOUTHERN MED J, V88, P475, DOI 10.1097/00007611-199504000-00020
   MULLIGAN LM, 1993, NATURE, V363, P458, DOI 10.1038/363458a0
   NAKAGAWARA A, 1985, CANCER, V55, P2794, DOI 10.1002/1097-0142(19850615)55:12<2794::AID-CNCR2820551213>3.0.CO;2-L
   PACHNIS V, 1993, DEVELOPMENT, V119, P1005
   SALMI J, 1988, ACTA MED SCAND, V224, P403
   SCHOENHERR CJ, 1995, SCIENCE, V267, P1360, DOI 10.1126/science.7871435
   SHI SR, 1991, J HISTOCHEM CYTOCHEM, V39, P741, DOI 10.1177/39.6.1709656
   TISCHLER AS, 1995, J HISTOCHEM CYTOCHEM, V43, P1, DOI 10.1177/43.1.7822757
   TISCHLER AS, 1976, SCIENCE, V192, P902, DOI 10.1126/science.179139
   TISCHLER AS, 1980, LAB INVEST, V43, P399
   TISCHLER AS, 1987, HUM PATHOL, V18, P909, DOI 10.1016/S0046-8177(87)80268-X
NR 25
TC 47
Z9 50
U1 0
U2 2
PU LIPPINCOTT-RAVEN PUBL
PI PHILADELPHIA
PA 227 EAST WASHINGTON SQ, PHILADELPHIA, PA 19106
SN 0147-5185
J9 AM J SURG PATHOL
JI Am. J. Surg. Pathol.
PD JAN
PY 1997
VL 21
IS 1
BP 102
EP 108
DI 10.1097/00000478-199701000-00011
PG 7
WC Pathology; Surgery
WE Science Citation Index Expanded (SCI-EXPANDED)
SC Pathology; Surgery
GA WB243
UT WOS:A1997WB24300011
PM 8990146
DA 2023-08-27
ER

PT J
AU Pang, Y
   Liu, Y
   Pacak, K
   Yang, CZ
AF Pang, Ying
   Liu, Yang
   Pacak, Karel
   Yang, Chunzhang
TI Pheochromocytomas and Paragangliomas: From Genetic Diversity to Targeted
   Therapies
SO CANCERS
LA English
DT Review
DE pheochromocytoma; paraganglioma; neuroendocrine tumor; targeted therapy;
   therapy resistance
ID HIPPEL-LINDAU DISEASE; SUCCINATE-DEHYDROGENASE; MALIGNANT
   PHEOCHROMOCYTOMAS; SPORADIC PHEOCHROMOCYTOMA; INHIBITOR SUNITINIB; NECK
   PARAGANGLIOMAS; SOMATIC MUTATIONS; ENDOCRINE TUMORS; FACTOR 1-ALPHA; SDH
   MUTATIONS
AB Pheochromocytoma and paraganglioma (PCPGs) are rare neuroendocrine tumors that arise from the chromaffin tissue of adrenal medulla and sympathetic ganglia. Although metastatic PCPGs account for only 10% of clinical cases, morbidity and mortality are high because of the uncontrollable mass effect and catecholamine level generated by these tumors. Despite our expanding knowledge of PCPG genetics, the clinical options to effectively suppress PCPG progression remain limited. Several recent translational studies revealed that PCPGs with different molecular subtypes exhibit distinctive oncogenic pathways and spectrum of therapy resistance. This suggests that therapeutics can be adjusted based on the signature molecular and metabolic pathways of PCPGs. In this review, we summarized the latest findings on PCPG genetics, novel therapeutic targets, and perspectives for future personalized medicine.
C1 [Pang, Ying; Pacak, Karel] NIH, Sect Med Neuroendocrinol, Eunice Kennedy Shriver Natl Inst Child Hlth & Hum, Bethesda, MD 20892 USA.
   [Liu, Yang; Yang, Chunzhang] Natl Canc Inst, Neurooncol Branch, Ctr Canc Res, Bethesda, MD 20892 USA.
C3 National Institutes of Health (NIH) - USA; NIH Eunice Kennedy Shriver
   National Institute of Child Health & Human Development (NICHD); National
   Institutes of Health (NIH) - USA; NIH National Cancer Institute (NCI)
RP Yang, CZ (通讯作者)，Natl Canc Inst, Neurooncol Branch, Ctr Canc Res, Bethesda, MD 20892 USA.
EM
RI Liu, Yang/AAP-7525-2020
OI Yang, Chunzhang/0000-0001-6433-0867
FU Neuro-Oncology Branch, Center for Cancer Research, National Cancer
   Institute; Eunice Kennedy Shriver National Institute of Child Health and
   Human Development, National Institutes of Health, Bethesda MD USA;
   NATIONAL CANCER INSTITUTE [ZIABC011684] Funding Source: NIH RePORTER
FX This study was supported by the Neuro-Oncology Branch, Center for Cancer
   Research, National Cancer Institute and Eunice Kennedy Shriver National
   Institute of Child Health and Human Development, National Institutes of
   Health, Bethesda MD USA 20892.
CR Adler JT, 2008, SURGERY, V144, P956, DOI 10.1016/j.surg.2008.08.027
   Amar L, 2007, J CLIN ENDOCR METAB, V92, P3822, DOI 10.1210/jc.2007-0709
   Astuti D, 2001, AM J HUM GENET, V69, P49, DOI 10.1086/321282
   ATTIE T, 1995, HUM MOL GENET, V4, P1381, DOI 10.1093/hmg/4.8.1381
   Ayala-Ramirez M, 2012, J CLIN ENDOCR METAB, V97, P4040, DOI 10.1210/jc.2012-2356
   Bausch B, 2007, J CLIN ENDOCR METAB, V92, P2784, DOI 10.1210/jc.2006-2833
   Bayley JP, 2014, BMC MED GENET, V15, DOI 10.1186/s12881-014-0111-8
   Baysal BE, 2000, SCIENCE, V287, P848, DOI 10.1126/science.287.5454.848
   Bergsand EK, 2013, SEMIN ONCOL, V40, P4, DOI 10.1053/j.seminoncol.2012.11.013
   BLACKWOOD EM, 1991, SCIENCE, V251, P1211, DOI 10.1126/science.2006410
   Boedeker CC, 2009, J CLIN ENDOCR METAB, V94, P1938, DOI 10.1210/jc.2009-0354
   BOUSSET K, 1993, ONCOGENE, V8, P3211
   Burnichon N, 2012, CLIN CANCER RES, V18, P2828, DOI 10.1158/1078-0432.CCR-12-0160
   Burnichon N, 2011, HUM MOL GENET, V20, P3974, DOI 10.1093/hmg/ddr324
   Burnichon N, 2011, EUR J ENDOCRINOL, V164, P141, DOI 10.1530/EJE-10-0758
   Burnichon N, 2010, HUM MOL GENET, V19, P3011, DOI 10.1093/hmg/ddq206
   Capodimonti S, 2012, J CLIN ONCOL, V30, pE137, DOI 10.1200/JCO.2011.38.6797
   Castro-Vega LJ, 2015, NAT COMMUN, V6, DOI 10.1038/ncomms7044
   Cayo MA, 2009, AM J TRANSL RES, V1, P178
   Chen WF, 2016, NATURE, V539, P112, DOI 10.1038/nature19796
   Chouaib S, 2017, ONCOGENE, V36, P439, DOI 10.1038/onc.2016.225
   Chouchani ET, 2014, NATURE, V515, P431, DOI 10.1038/nature13909
   Comino-Mendez I, 2011, NAT GENET, V43, P663, DOI 10.1038/ng.861
   Correia M J, 2000, Rev Port Cardiol, V19, P11
   Courtney KD, 2018, J CLIN ONCOL, V36, P867, DOI 10.1200/JCO.2017.74.2627
   Crona J, 2013, J CLIN ENDOCR METAB, V98, pE1266, DOI 10.1210/jc.2012-4257
   de Cubas AA, 2015, CLIN CANCER RES, V21, P3020, DOI 10.1158/1078-0432.CCR-14-2804
   Druce MR, 2009, HORM METAB RES, V41, P697, DOI 10.1055/s-0029-1220687
   Erlic Z, 2009, HORM-INT J ENDOCRINO, V8, P29, DOI 10.14310/horm.2002.1219
   Favier J, 2012, ENDOCR PATHOL, V23, P34, DOI 10.1007/s12022-011-9189-0
   Fishbein L, 2017, CANCER CELL, V31, P181, DOI 10.1016/j.ccell.2017.01.001
   Fishbein L, 2012, CANCER GENET-NY, V205, P1, DOI 10.1016/j.cancergen.2012.01.009
   Fitzgerald PA, 2006, ANN NY ACAD SCI, V1073, P465, DOI 10.1196/annals.1353.050
   Friedrich CA, 2001, HUM MOL GENET, V10, P763, DOI 10.1093/hmg/10.7.763
   Ghayee HK, 2013, PLOS ONE, V8, DOI 10.1371/journal.pone.0065624
   Gill AJ, 2010, HUM PATHOL, V41, P805, DOI 10.1016/j.humpath.2009.12.005
   Giubellino A, 2013, ENDOCRINOLOGY, V154, P646, DOI 10.1210/en.2012-1854
   Gottlieb E, 2005, NAT REV CANCER, V5, P857, DOI 10.1038/nrc1737
   Gutmann DH, 2017, NAT REV DIS PRIMERS, V3, DOI 10.1038/nrdp.2017.4
   Guzy RD, 2008, MOL CELL BIOL, V28, P718, DOI 10.1128/MCB.01338-07
   Hadoux J, 2017, HORM CANCER-US, V8, P330, DOI 10.1007/s12672-017-0303-8
   Hadoux J, 2014, INT J CANCER, V135, P2711, DOI 10.1002/ijc.28913
   Hahn NM, 2009, J CLIN ONCOL, V27, P460, DOI 10.1200/JCO.2008.19.9380
   Hao HX, 2009, SCIENCE, V325, P1139, DOI 10.1126/science.1175689
   Hatfield SM, 2016, CURR OPIN PHARMACOL, V29, P90, DOI 10.1016/j.coph.2016.06.009
   Huang H, 2008, CANCER-AM CANCER SOC, V113, P2020, DOI 10.1002/cncr.23812
   Jafri M, 2012, EUR J ENDOCRINOL, V166, P151, DOI 10.1530/EJE-11-0497
   Janeway KA, 2011, P NATL ACAD SCI USA, V108, P314, DOI 10.1073/pnas.1009199108
   Jimenez C, 2009, J CLIN ENDOCR METAB, V94, P386, DOI 10.1210/jc.2008-1972
   Jochmanova I, 2018, TRENDS CANCER, V4, P6, DOI 10.1016/j.trecan.2017.11.001
   Joshua AM, 2009, J CLIN ENDOCR METAB, V94, P5, DOI 10.1210/jc.2008-1836
   Keith B, 2012, NAT REV CANCER, V12, P9, DOI 10.1038/nrc3183
   Killian JK, 2013, CANCER DISCOV, V3, P648, DOI 10.1158/2159-8290.CD-13-0092
   King KS, 2011, J CLIN ONCOL, V29, P4137, DOI 10.1200/JCO.2011.34.6353
   Koch CA, 2002, ONCOGENE, V21, P479, DOI 10.1038/sj.onc.1205133
   Korpershoek E, 2011, J CLIN ENDOCR METAB, V96, pE1472, DOI 10.1210/jc.2011-1043
   Kulke MH, 2006, J CLIN ONCOL, V24, P401, DOI 10.1200/JCO.2005.03.6046
   Kunst HPM, 2011, CLIN CANCER RES, V17, P247, DOI 10.1158/1078-0432.CCR-10-0420
   Kwekkeboom DJ, 2003, EUR J NUCL MED MOL I, V30, P417, DOI 10.1007/s00259-002-1050-8
   Labiano S, 2016, ONCOIMMUNOLOGY, V5, DOI 10.1080/2162402X.2015.1062967
   Letouze E, 2013, CANCER CELL, V23, P739, DOI 10.1016/j.ccr.2013.04.018
   Li ZY, 2016, NEUROCHEM INT, V99, P239, DOI 10.1016/j.neuint.2016.08.004
   Liu Y, 2019, JNCI-J NATL CANCER I, V111, P1033, DOI 10.1093/jnci/djy230
   Lonser RR, 2003, LANCET, V361, P2059, DOI 10.1016/S0140-6736(03)13643-4
   Lu YX, 2017, CANCER RES, V77, P1709, DOI 10.1158/0008-5472.CAN-16-2773
   Luchetti A, 2015, INT J ENDOCRINOL, V2015, DOI 10.1155/2015/138573
   Ma XJM, 2009, NAT REV MOL CELL BIO, V10, P307, DOI 10.1038/nrm2672
   Machens A, 2005, J CLIN ENDOCR METAB, V90, P3999, DOI 10.1210/jc.2005-0064
   Matro J, 2013, HORM METAB RES, V45, P147, DOI 10.1055/s-0032-1331211
   Merlo A, 2013, J CLIN ENDOCR METAB, V98, pE1661, DOI 10.1210/jc.2013-1636
   Nemoto K, 2012, NEUROENDOCRINOL LETT, V33, P260
   Niemann S, 2000, NAT GENET, V26, P268, DOI 10.1038/81551
   Oh DY, 2012, CANCER-AM CANCER SOC, V118, P6162, DOI 10.1002/cncr.27675
   Otte A, 1997, EUR J NUCL MED, V24, P792
   Pang Y, 2018, CLIN CANCER RES, V24, P3423, DOI 10.1158/1078-0432.CCR-17-3406
   Pasini B, 2009, J INTERN MED, V266, P19, DOI 10.1111/j.1365-2796.2009.02111.x
   Percy MJ, 2008, NEW ENGL J MED, V358, P162, DOI 10.1056/NEJMoa073123
   Plaza-Menacho I, 2014, MOL CELL, V53, P738, DOI 10.1016/j.molcel.2014.01.015
   Qi ST, 2012, CANCER SCI, V103, P269, DOI 10.1111/j.1349-7006.2011.02134.x
   Ratner N, 2015, NAT REV CANCER, V15, P290, DOI 10.1038/nrc3911
   Ricketts C, 2008, J NATL CANCER I, V100, P1260, DOI 10.1093/jnci/djn254
   Roe JS, 2006, MOL CELL, V22, P395, DOI 10.1016/j.molcel.2006.04.006
   Safford SD, 2003, SURGERY, V134, P956, DOI 10.1016/S0039-6060(03)00426-4
   Saxena N, 2016, JNCI-J NATL CANCER I, V108, DOI 10.1093/jnci/djv287
   Semenza G.L., 2007, SCI STKE, V407, pcm8, DOI [10.1126/stke.4072007cm8, DOI 10.1126/STKE.4072007CM8]
   Semenza GL, 1996, GENOMICS, V34, P437, DOI 10.1006/geno.1996.0311
   Semenza GL, 2009, BLOOD, V114, P2015, DOI 10.1182/blood-2009-05-189985
   Strosberg J, 2017, NEW ENGL J MED, V376, P125, DOI 10.1056/NEJMoa1607427
   Sulkowski PL, 2018, NAT GENET, V50, P1086, DOI 10.1038/s41588-018-0170-4
   Timmers HJLM, 2007, J CLIN ENDOCR METAB, V92, P779, DOI 10.1210/jc.2006.2315
   Turcan S, 2012, NATURE, V483, P479, DOI 10.1038/nature10866
   Tuthill Mark, 2009, BMJ Case Rep, V2009, DOI 10.1136/bcr.08.2008.0732
   van Hulsteijn LT, 2014, CLIN ENDOCRINOL, V80, P487, DOI 10.1111/cen.12341
   VANDERMEY AGL, 1989, LANCET, V2, P1291
   Vicha A, 2014, ENDOCR-RELAT CANCER, V21, pR261, DOI 10.1530/ERC-13-0398
   Wang HR, 2018, CANCER RES, V78, P1266, DOI 10.1158/0008-5472.CAN-17-1718
   Welander J, 2012, HUM MOL GENET, V21, P5406, DOI 10.1093/hmg/dds402
   Welsh S, 2004, MOL CANCER THER, V3, P233
   Welsh SJ, 2003, MOL CANCER THER, V2, P235
   Yang Chunzhang, 2015, Hum Genome Var, V2, P15053, DOI 10.1038/hgv.2015.53
   Yang CZ, 2012, FASEB J, V26, P4506, DOI 10.1096/fj.12-210146
   Ying P, 2017, ONCOTARGET, V8, P22313, DOI 10.18632/oncotarget.16224
   YOSHIMOTO K, 1992, JPN J CANCER RES, V83, P1057, DOI 10.1111/j.1349-7006.1992.tb02722.x
   Zaknun JJ, 2013, EUR J NUCL MED MOL I, V40, P800, DOI 10.1007/s00259-012-2330-6
   Zhang ZP, 2017, TOXICOL APPL PHARM, V336, P94, DOI 10.1016/j.taap.2017.10.011
   Zhong H, 1999, CANCER RES, V59, P5830
   Zhuang ZP, 2012, NEW ENGL J MED, V367, P922, DOI 10.1056/NEJMoa1205119
NR 107
TC 26
Z9 26
U1 0
U2 2
PU MDPI
PI BASEL
PA ST ALBAN-ANLAGE 66, CH-4052 BASEL, SWITZERLAND
SN 2072-6694
J9 CANCERS
JI Cancers
PD APR
PY 2019
VL 11
IS 4
AR 436
DI 10.3390/cancers11040436
PG 16
WC Oncology
WE Science Citation Index Expanded (SCI-EXPANDED)
SC Oncology
GA HY0BE
UT WOS:000467773400004
PM 30925729
OA Green Published, gold, Green Submitted
DA 2023-08-27
ER

PT J
AU Roche, PH
   Pellet, W
   Fuentes, S
   Thomassin, JM
   Regis, J
AF Roche, PH
   Pellet, W
   Fuentes, S
   Thomassin, JM
   Regis, J
TI Gamma knife radiosurgical management of petroclival meningiomas results
   and indications
SO ACTA NEUROCHIRURGICA
LA English
DT Article
DE cranial nerves; gamma knife; petroclival meningiomas; stereoractic
   radiosurgery
ID STEREOTAXIC RADIOSURGERY; BASE MENINGIOMAS; EXPERIENCE; SURGERY
AB Background. Surgical treatment of petroclival meningiomas remains challenging. In order to refine indications for the use of stereotactic radiosurgery in the treatment of these tumours, we retrospectively evaluated our experience in this field.
   Methods. Thirty-two patients harboring a petroclival meningioma were treated consecutively using a Gamma knife between December 92 and June 1998. Eight patients underwent radiosurgery after one or more attempted surgical removals and 24 had radiosurgery as the primary treatment. The main symptoms before radiosurgery were cranial nerve palsies, including a sixth nerve deficit in 10 patients and a trigeminal nerve disturbance in 9. All patients underwent a conformal multiisocentric treatment (mean isocenter's number 8.8) and the dose delivered at the tumour margin ranged from 10 to 15 Gy (mean dose 13 Gy).
   Findings. The duration of follow-up varied from 24 to 118 months (mean clinical follow-up 56 months, mean radiological follow-up 52.6 months). All 32 patients survived. The turnout volume remained unchanged in 28 patients and decreased slightly in 4. Neurological status worsened permanently in 2 patients with a delayed hemiparesis due to focal pontine infarction. These complications were associated with large meningiomas with vascular involvement and ventral brainstem compression, and occurred at the very early stage of our experience. At last follow-up, preoperative fifth or sixth cranial nerve deficits had improved or recovered in 13 out of 19 patients and any delayed worsening or new cranial nerve deficits were not observed after radiosurgery.
   Conclusions. Stereotactic radiosurgery with a Gamma knife provides effective management of small to middle sized petroclival meningiomas and is an alternative to microsurgery. Careful selection of patients and use of major technical refinements should improve the safety of this treatment.
C1 CHU Timone, Serv Neurochirurg Stereotax & Fonct, F-13005 Marseille, France.
   CHU Timone, Unite Otoneurochirurg, F-13005 Marseille, France.
   CHU Timone, Serv Neurochirurg, F-13005 Marseille, France.
C3 CIVIS; UDICE-French Research Universities; Aix-Marseille Universite;
   Assistance Publique-Hopitaux de Marseille; CIVIS; UDICE-French Research
   Universities; Aix-Marseille Universite; Assistance Publique-Hopitaux de
   Marseille; CIVIS; UDICE-French Research Universities; Aix-Marseille
   Universite; Assistance Publique-Hopitaux de Marseille
RP Regis, J (通讯作者)，CHU Timone, Serv Neurochirurg Stereotax & Fonct, Blvd Jean Moulin, F-13005 Marseille, France.
CR ALMEFTY O, 1988, NEUROSURGERY, V22, P510, DOI 10.1227/00006123-198803000-00010
   BRICOLO AP, 1992, NEUROSURGERY, V31, P813, DOI 10.1227/00006123-199211000-00001
   Couldwell WT, 1996, J NEUROSURG, V84, P20, DOI 10.3171/jns.1996.84.1.0020
   GANZ JC, 1993, STEREOT FUNCT NEUROS, V61, P23, DOI 10.1159/000100656
   KALLIO M, 1992, NEUROSURGERY, V31, P2, DOI 10.1227/00006123-199207000-00002
   KONDZIOLKA D, 1991, J NEUROSURG, V74, P552, DOI 10.3171/jns.1991.74.4.0552
   LARSON DA, 1993, INT J RADIAT ONCOL, V25, P557, DOI 10.1016/0360-3016(93)90080-F
   Mathiesen T, 1996, NEUROSURGERY, V39, P2, DOI 10.1097/00006123-199607000-00002
   Morita A, 1999, J NEUROSURG, V90, P42, DOI 10.3171/jns.1999.90.1.0042
   Regis J, 2002, J NEUROSURG, V97, P1091, DOI 10.3171/jns.2002.97.5.1091
   Roche PH, 2000, J NEUROSURG, V93, P68, DOI 10.3171/jns.2000.93.supplement_3.0068
   SAMII M, 1989, NEUROSURGERY, V24, P12, DOI 10.1227/00006123-198901000-00003
   SEKHAR LN, 1990, NEUROSURGERY, V27, P764, DOI 10.1227/00006123-199011000-00015
   Subach BR, 1998, NEUROSURGERY, V42, P437, DOI 10.1097/00006123-199803000-00001
NR 14
TC 85
Z9 87
U1 0
U2 1
PU SPRINGER-VERLAG WIEN
PI VIENNA
PA SACHSENPLATZ 4-6, PO BOX 89, A-1201 VIENNA, AUSTRIA
SN 0001-6268
J9 ACTA NEUROCHIR
JI Acta Neurochir.
PD OCT
PY 2003
VL 145
IS 10
BP 883
EP 888
DI 10.1007/s00701-003-0123-1
PG 6
WC Clinical Neurology; Surgery
WE Science Citation Index Expanded (SCI-EXPANDED)
SC Neurosciences & Neurology; Surgery
GA 739AW
UT WOS:000186320900006
PM 14577010
DA 2023-08-27
ER

PT J
AU Lancellotti, CLP
   Amary, MFC
   Barbastefano, AM
   Tilbery, CP
AF Lancellotti, CLP
   Amary, MFC
   Barbastefano, AM
   Tilbery, CP
TI ''Gliomatosis cerebri'' simulating an acute diffuse encephalomyelitis
SO ARQUIVOS DE NEURO-PSIQUIATRIA
LA English
DT Article
DE gliomatosis cerebri; neuroradiology; neuropathology; brain; tumour
ID DIAGNOSIS; FEATURES; MR
AB Neuroradiologic, neuropathologic and immunohistochemical features are reported in a young man with a impairment of the central nervous system mimicking an acute diffuse encephalomyelitis. A white male, 17 years old, healthy till 4 months before, when developed a right hemiparesis and after 2 months a bilateral hemiparesis with a progressive impairment of several cranial nerves. Magnetic resonance imaging showed multiple lesions without a mass effect that suggested myelin loss. He remained unconscious for almost one month before dying of pneumonia. The neuropathologic examination showed a heavy brain (1505 g) with herniations and a large right midbrain. There were several soft and pink areas mainly at the right midbrain, left cerebellum and in the white matter of the left cerebral hemisphere. The histopathologic sections showed diffuse blastomatous proliferation without total replacement or destruction of the original tissue. The tumor cells had astrocytic, oligodendrocytic and spongioblastic phenotypes, some of them with a GFAP-positive reactivity. There were focal anaplastic changes. The diagnosis of ''gliomatosis cerebri'' was only possible by the autopsy.
C1 FAC CIENCIAS MED SANTA CASA,DEPT CIENCIAS PATOL,SAO PAULO,BRAZIL.
   FAC CIENCIAS MED SANTA CASA,DISCIPLINA NEUROL,SAO PAULO,BRAZIL.
C3 Faculdade de Ciencias Medicas Santa Casa de Sao Paulo; Faculdade de
   Ciencias Medicas Santa Casa de Sao Paulo
RI Amary, Fernanda/AAI-7339-2020
CR ARTIGAS J, 1985, CLIN NEUROPATHOL, V4, P135
   BALAKRISHNAN V, 1985, PATHOLOGY, V17, P123, DOI 10.3109/00313028509063740
   BALKO MG, 1992, HUM PATHOL, V23, P706
   BURGER PC, 1993, ATLAS TUMOR PATHOL, P72
   CERVOSNAVARRO J, 1987, VIRCHOWS ARCH A, V411, P93, DOI 10.1007/BF00734520
   COUCH JR, 1974, NEUROLOGY, V24, P504, DOI 10.1212/WNL.24.6.504
   DELISLE MB, 1989, ANN PATHOL, V9, P214
   DUNN J, 1957, ARCH PATHOL, V64, P82
   FELSBERG GJ, 1994, AM J NEURORADIOL, V15, P1745
   Galatioto S., 1993, Zentralblatt fuer Pathologie, V139, P261
   HARA A, 1991, J NEUROL, V238, P80, DOI 10.1007/BF00315685
   JENNINGS MT, 1995, J CHILD NEUROL, V10, P37, DOI 10.1177/088307389501000111
   KLEIHUES P, 1993, HISTOLOGICAL TYPING, P21
   KOSLOW SA, 1992, NEURORADIOLOGY, V34, P331, DOI 10.1007/BF00588195
   MILLER RR, 1981, ACTA CYTOL, V25, P37
   MINEURA K, 1991, J NUCL MED, V32, P726
   Nevin S, 1938, BRAIN, V61, P170, DOI 10.1093/brain/61.2.170
   NG HK, 1988, CLIN NEUROPATHOL, V7, P266
   Poser CM, 1994, ANN NEUROL, V36, P180
   POSER CM, 1992, MED PRINCIPLES PRACT, V3, P1
   Priest J R, 1993, J Neurosurg, V78, P158
   ROSS IB, 1991, SURG NEUROL, V36, P431, DOI 10.1016/0090-3019(91)90156-4
   RUSSELL DS, 1989, PATHOL TUMOURS NER, P146
   Scherer HJ, 1940, BRAIN, V63, P1, DOI 10.1093/brain/63.1.1
   SCHOBER R, 1991, ACTA NEUROCHIR, V113, P131, DOI 10.1007/BF01403198
   Sheinker IM., 1943, J NEUROPATH EXP NEUR, V2, P178, DOI [10.1097/00005072-194304000-00007, DOI 10.1097/00005072-194304000-00007]
   SHIN YM, 1993, AM J ROENTGENOL, V161, P859, DOI 10.2214/ajr.161.4.8372774
   TROOST D, 1987, CLIN NEUROL NEUROSUR, V89, P43, DOI 10.1016/S0303-8467(87)80075-6
   WILSON NW, 1987, J NEUROL, V234, P445, DOI 10.1007/BF00314097
   YANAKA K, 1992, AM J NEURORADIOL, V13, P349
   ZULCH KJ, 1986, BRAIN TUMORS THEIR B, P341
NR 31
TC 7
Z9 7
U1 0
U2 0
PU ASSOC ARQUIVOS DE NEURO- PSIQUIATRIA
PI SAO PAULO SP
PA C/O DR OSWALDO LANGE, CAIXA POSTAL 8877, 01065-970 SAO PAULO SP, BRAZIL
SN 0004-282X
J9 ARQ NEURO-PSIQUIAT
JI Arq. Neuro-Psiquiatr.
PD SEP
PY 1997
VL 55
IS 3
BP 488
EP 495
DI 10.1590/S0004-282X1997000300023
PG 8
WC Neurosciences; Psychiatry
WE Science Citation Index Expanded (SCI-EXPANDED)
SC Neurosciences & Neurology; Psychiatry
GA XR522
UT WOS:A1997XR52200023
PM 9629370
OA Green Published
DA 2023-08-27
ER

PT J
AU Lloyd, SK
   Evans, DG
AF Lloyd, S. K.
   Evans, D. G.
TI Neurofibromatosis type 2 service delivery in England
SO NEUROCHIRURGIE
LA English
DT Article
DE Neurofibromatosis type 2; Vestibular schwannoma; Cochlear implantation;
   Auditory brainstem implantation; Bevacizumab
ID TUMOR-PRONE SYNDROMES; AUDITORY BRAIN-STEM; COCHLEAR IMPLANTATION;
   PREDICTORS; MORTALITY; DISEASE; FAMILY
AB Neurofibromatosis type 2 (NF2) is a complex disease characterized by the development of multiple schwannomas, especially vestibular schwannomas, as well as other types of benign tumours including meningioma and spinal ependymoma. Due to its multisystem nature, the management of NF2 requires a multidisciplinary approach. In England, the delivery of care for NF2 patients has been centralized to four-"hub" centres in Manchester, Cambridge, Oxford and London each having associated "spoke" centres. Each centre has a core multidisciplinary team consisting of genetics, otolaryngology, neurosurgery, paediatrics, neurology, audiology, radiology, psychology, physiotherapy, specialist nurses and administrative staff. In addition, the core team has access to plastic surgery, ophthalmology, peripheral nerve surgery and adult and paediatric oncology. There are weekly multidisciplinary clinics each with six to eight patients. Each patient is discussed during a team meeting and the management decisions that are made are then discussed with the patients. All patients are reviewed at least annually and have annual head magnetic resonance imaging (MRI) and three yearly spinal MRI. Annual audiological assessment is performed. Cochlear implantation and auditory brainstem implantation are offered if indicated. Surgery, stereotactic radiosurgery and bevacizumab therapy are available for the management of intracranial and spinal tumours. The integration of the service in England has provided significant benefits to patient care and, in the long term, will provide robust patient outcome data that will provide an evidence base to assist in optimizing management of patients with NF2. (C) 2016 Published by Elsevier Masson SAS.
C1 [Lloyd, S. K.] Univ Manchester, Inst Canc Sci, Manchester M13 9PL, Lancs, England.
   [Lloyd, S. K.; Evans, D. G.] Cent Manchester Univ Hosp NHS Fdn Trust, Manchester Acad Hlth Sci Ctr, Manchester M13 9WL, Lancs, England.
   [Evans, D. G.] Univ Manchester, MAHSC, Manchester Ctr Genom Med, Manchester, Lancs, England.
C3 University of Manchester; University of Manchester; University of
   Manchester
RP Lloyd, SK (通讯作者)，Manchester Royal Infirm, Univ Dept Otolaryngol Head & Neck Surg, Oxford Rd, Manchester M13 9WL, Lancs, England.
EM
RI Evans, D Gareth/AAB-4308-2022
OI Evans, D Gareth/0000-0002-8482-5784
CR Baser ME, 2002, AM J HUM GENET, V71, P715, DOI 10.1086/342716
   Evans DG, 2012, CLIN GENET, V82, P416, DOI 10.1111/j.1399-0004.2011.01816.x
   Evans DG, 2010, AM J MED GENET A, V152A, P327, DOI 10.1002/ajmg.a.33139
   Evans DGR, 2011, ADV OTO-RHINO-LARYNG, V70, P91, DOI 10.1159/000322482
   EVANS DGR, 1993, J LARYNGOL OTOL, V107, P401, DOI 10.1017/S002221510012328X
   Evans DGR, 1998, AM J HUM GENET, V63, P727, DOI 10.1086/512074
   Evans DGR, 2005, BRIT J NEUROSURG, V19, P5, DOI 10.1080/02688690500081206
   Hexter A, 2015, J MED GENET, V52, P699, DOI 10.1136/jmedgenet-2015-103290
   Hornigold RE, 2012, J NEUROL SURG PART B, V73, P104, DOI 10.1055/s-0032-1301396
   Kontorinis G, 2014, EUR ARCH OTORHINOLAR
   Lloyd SKW, 2014, OTOL NEUROTOL, V35, P43, DOI 10.1097/MAO.0000000000000185
   McSorley A, 2015, OTOL NEUROTOL, V36, P873, DOI 10.1097/MAO.0000000000000671
   Moffat DA, 2013, BRIT J NEUROSURG, V27, P446, DOI 10.3109/02688697.2013.771143
   Tysome JR, 2013, OTOL NEUROTOL, V34, P1743, DOI 10.1097/MAO.0b013e3182a1a8b4
NR 14
TC 13
Z9 14
U1 1
U2 6
PU MASSON EDITEUR
PI MOULINEAUX CEDEX 9
PA 21 STREET CAMILLE DESMOULINS, ISSY, 92789 MOULINEAUX CEDEX 9, FRANCE
SN 0028-3770
EI 1773-0619
J9 NEUROCHIRURGIE
JI Neurochirurgie
PD NOV
PY 2018
VL 64
IS 5
BP 375
EP 380
DI 10.1016/j.neuchi.2015.10.006
PG 6
WC Clinical Neurology; Surgery
WE Science Citation Index Expanded (SCI-EXPANDED)
SC Neurosciences & Neurology; Surgery
GA HD6XZ
UT WOS:000452694900008
PM 26826883
DA 2023-08-27
ER

PT J
AU Mureb, MC
   Dastazirgada, Y
   Benjamin, C
   Golfinos, JG
   Kondziolka, D
AF Mureb, Monica C.
   Dastazirgada, Yosef
   Benjamin, Carolina
   Golfinos, John G.
   Kondziolka, Douglas
TI Simultaneous Treatment of Petroclival Meningiomas and the Trigeminal
   Nerve with Gamma Knife Radiosurgery for Tumor-Related Trigeminal
   Neuralgia
SO WORLD NEUROSURGERY
LA English
DT Article
DE Gamma knife radiosurgery; Petroclival meningioma; Stereotactic
   radiosurgery; Trigeminal neuralgia
ID STEREOTACTIC RADIOSURGERY
AB BACKGROUND: Some petroclival meningiomas cause trigeminal nerve compression, leading to disabling trigeminal neuralgia (TN). Tumor resection and nerve decompression can offer pain relief but might not be feasible in all patients. Simultaneous stereotactic radiosurgery (SRS) to the tumor and nerve is another option. SRS is an effective means of treating meningiomas and TN separately, but data on the efficacy and outcomes of their concomitant treatment are limited.
   CASE DESCRIPTION: We report a series of 4 patients who presented with TN secondary to a petroclival mass causing compression of the trigeminal nerve. All patients underwent SRS to both the petroclival mass and trigeminal nerve in a single session. The average margin tumor dose was 12.25 Gy (range, 12-12.5 Gy), and the average maximum trigeminal nerve dose was 80 Gy (range, 75-85 Gy). In all patients, before intervention, the Barrow Neurologic Institute (BNI) pain intensity score was grade IV or V. At last follow-up (average, 29.8 months), all patients were pain-free (BNI I or IIIA). Two patients experienced reduced facial sensation in 1 or all 3 distributions. No brainstem edema was seen.
   CONCLUSIONS: This series highlights the benefits and safety of simultaneous treatment of petroclival tumors and the trigeminal nerve in a single session for patients affected by tumor-related TN.
C1 [Mureb, Monica C.; Dastazirgada, Yosef; Benjamin, Carolina; Golfinos, John G.; Kondziolka, Douglas] NYU, Dept Neurosurg, Sch Med, 550 1St Ave, New York, NY 10016 USA.
C3 New York University
RP Kondziolka, D (通讯作者)，NYU, Dept Neurosurg, Sch Med, 550 1St Ave, New York, NY 10016 USA.
EM
OI Kondziolka, Douglas/0000-0001-8925-7344
CR Flannery TJ, 2010, J NEUROSURG, V112, P957, DOI 10.3171/2009.8.JNS09695
   Flickinger JC, 2001, INT J RADIAT ONCOL, V51, P449, DOI 10.1016/S0360-3016(01)01606-6
   Kano H, 2011, STEREOT FUNCT NEUROS, V89, P17, DOI 10.1159/000321187
   Kim JW, 2017, STEREOT FUNCT NEUROS, V95, P209, DOI 10.1159/000475763
   Kim SK, 2016, J NEUROSURG, V125, P838, DOI 10.3171/2015.7.JNS15451
   LEKSELL L, 1992, RADIOSURGERY, P263
   Ramina R, 2008, ESSENTIALS NEUROSURG, P121
   Squire SE, 2012, STEREOT FUNCT NEUROS, V90, P145, DOI 10.1159/000335873
   Starke R, 2014, J NEURO-ONCOL, V119, P169, DOI 10.1007/s11060-014-1470-x
   Wayhs SY, 2017, CLIN NEUROL NEUROSUR, V152, P68, DOI 10.1016/j.clineuro.2016.11.021
   Young B, 2013, J NEUROSURG, V119, P1166, DOI 10.3171/2013.1.JNS12875
NR 11
TC 7
Z9 7
U1 0
U2 0
PU ELSEVIER SCIENCE INC
PI NEW YORK
PA STE 800, 230 PARK AVE, NEW YORK, NY 10169 USA
SN 1878-8750
EI 1878-8769
J9 WORLD NEUROSURG
JI World Neurosurg.
PD JUL
PY 2020
VL 139
BP 242
EP 244
DI 10.1016/j.wneu.2020.04.082
PG 3
WC Clinical Neurology; Surgery
WE Science Citation Index Expanded (SCI-EXPANDED)
SC Neurosciences & Neurology; Surgery
GA MM7JC
UT WOS:000550328700015
PM 32330619
DA 2023-08-27
ER

PT J
AU COLLETTI, V
   BRICOLO, A
   FIORINO, FG
   BRUNI, L
AF COLLETTI, V
   BRICOLO, A
   FIORINO, FG
   BRUNI, L
TI CHANGES IN DIRECTLY RECORDED COCHLEAR NERVE COMPOUND ACTION-POTENTIALS
   DURING ACOUSTIC TUMOR SURGERY
SO SKULL BASE SURGERY
LA English
DT Article
ID AUDITORY EVOKED-POTENTIALS; HEARING PRESERVATION; NEUROMAS
AB Damage to labyrinthine, neural, and vascular structures may possibly account for failure to achieve hearing preservation after removal of small acoustic neuromas. The pathogenesis of auditory impairment is, however, sometimes unclear. Intraoperative monitoring of auditory evoked potentials enables us to study possible correlations between surgical maneuvers and the function of auditory structures and pathways. To this end, simultaneous recording of brainstem auditory evoked potentials and direct cochlear nerve action potentials (CNAPs) was utilized in 14 patients with small (12 to 18 mm) unilateral acoustic neuromas. The latency of the first negative component (N-1) of the CNAPs proved extremely sensitive in detecting intraoperative auditory damage and was a good predictive index of postoperative auditory outcome. Evaluation of temporal and morphologic CNAP patterns allowed identification of coagulation close to the cochlear nerve, drilling of the internal auditory canal, and removal of the intrameatal portion of the tumor, the most critical steps in hearing preservation. The mechanisms of damage are analyzed and discussed.
C1 UNIV VERONA,DEPT ENT,VERONA,ITALY.
   UNIV VERONA,DEPT NEUROSURG,VERONA,ITALY.
C3 University of Verona; University of Verona
CR ABRAMSON M, 1985, LARYNGOSCOPE, V95, P1318
   COLLETTI V, 1992, 1ST INT C ECOG OAE I
   COLLETTI V, 1990, FRACASTORO, V83, P157
   COLLETTI V, 1993, IN PRESS AM J OTOL
   FIORINO FG, 1992, 1ST INT C ECOG OAE I
   GRUNDY BL, 1982, J NEUROSURG, V57, P674, DOI 10.3171/jns.1982.57.5.0674
   JANNETTA PJ, 1984, ANN SURG, V200, P513, DOI 10.1097/00000658-198410000-00012
   KVETON JF, 1989, OTOLARYNG HEAD NECK, V100, P594, DOI 10.1177/019459988910000613
   MATTHIES C, 1992, 1ST INT SKULL BAS C, P11
   MOLLER AR, 1984, OTOLARYNG HEAD NECK, V92, P434
   Moller AR, 1981, J EXP NEUROL, V74, P862
   OJEMANN RG, 1984, J NEUROSURG, V61, P938, DOI 10.3171/jns.1984.61.5.0938
   RAUDZENS PA, 1982, J NEUROSURG, V57, P341, DOI 10.3171/jns.1982.57.3.0341
   SCHRAMM J, 1988, NEUROSURGERY, V22, P697
   SEKYA T, 1987, NEUROSURGERY, V21, P631
   SHELTON C, 1990, LARYNGOSCOPE, V100, P115
   Silverstein H, 1985, AM J OTOL S, V6, P99
   TATAGIBA M, 1992, J NEUROSURG, V77, P677, DOI 10.3171/jns.1992.77.5.0677
NR 18
TC 36
Z9 36
U1 0
U2 0
PU THIEME MEDICAL PUBL INC
PI NEW YORK
PA 381 PARK AVE SOUTH, NEW YORK, NY 10016
SN 1052-1453
J9 SKULL BASE SURG
JI Skull Base Surg.
PD JAN
PY 1994
VL 4
IS 1
BP 1
EP 9
DI 10.1055/s-2008-1058981
PG 9
WC Clinical Neurology; Otorhinolaryngology; Surgery
WE Science Citation Index Expanded (SCI-EXPANDED)
SC Neurosciences & Neurology; Otorhinolaryngology; Surgery
GA MU892
UT WOS:A1994MU89200001
PM 17170918
OA Green Published
DA 2023-08-27
ER

PT J
AU Kovacs, A
   Toth, L
   Glavak, C
   Liposits, G
   Hadjiev, J
   Antal, G
   Emri, M
   Vandulek, C
   Repa, I
AF Kovacs, Arpad
   Toth, Lilla
   Glavak, Csaba
   Liposits, Gabor
   Hadjiev, Janaki
   Antal, Gergely
   Emri, Miklos
   Vandulek, Csaba
   Repa, Imre
TI Integrating functional MRI information into conventional 3D radiotherapy
   planning of CNS tumors. Is it worth it?
SO JOURNAL OF NEURO-ONCOLOGY
LA English
DT Article
DE Functional MRI; Brain tumour; 3D radiotherapy
ID RISK
AB The purpose of our study was to examine the potential benefits of integrating functional MRI (fMRI) information into the 3D-based planning process for central nervous system (CNS) malignancies. Between 01.01.2008 and 01.12.2009, ten patients with astrocytoma (both low and high-grade histological type) were enrolled in this study. Before the planning process, conventional CT planning, postoperative MR, and individual functional MRI examinations were conducted. For the functional MRI examination four types of conventional stimuli were applied: acoustic, visual, somatosensory, and numeric. To examine the potential benefits of using fMRI-based information, three different types of theoretical planning were applied and compared: 3D conformal plan without fMRI information, 3D conformal plan with fMRI information, and IMRT plan with fMRI information. DVH analysis and the NTCP model were used for plan comparison. When comparing planning methods, distance-related subgroups were generated and studied. By using the additional fMRI information, a significantly higher sparing effect can be achieved on these ORs (both with conventional 3D-based planning and IMRT). In cases when the OR-PTV distance is less than 1 cm, IMRT seems to be a significantly better choice than conventional 3D-based techniques. IMRT also has an additional sparing effect on the optic tract and brainstem, especially for locations close to the midline. Our results demonstrated that using fMRI information in conventional 3D-based treatment planning has the potential benefit of significant dose reduction for the critical organs, with no compromise in PTV coverage even when using conventional 3D planning. fMRI can be widely used in low-grade cases (long life expectancy, lower acute and late toxicity) and also in cases with high-grade astrocytomas or distant metastases (higher dose to PTV with better sparing of risk organs). In cases when the OR-PTV distance is less than 1 cm, IMRT should be the choice of treatment for a higher sparing effect on functional active areas. Longer imaging and clinical follow up are needed to confirm the real sparing effect on these functional areas.
C1 [Kovacs, Arpad; Toth, Lilla; Glavak, Csaba; Liposits, Gabor; Hadjiev, Janaki; Antal, Gergely; Vandulek, Csaba; Repa, Imre] Univ Kaposvar, Dept Diagnost & Oncoradiol, H-7400 Kaposvar, Hungary.
   [Kovacs, Arpad] Univ Pecs, Fac Hlth Sci, Pecs, Hungary.
   [Emri, Miklos] Univ Debrecen, Debrecen, Hungary.
C3 Kaposvar University; University of Pecs; University of Debrecen
RP Kovacs, A (通讯作者)，Univ Kaposvar, Dept Diagnost & Oncoradiol, Guba S St 40, H-7400 Kaposvar, Hungary.
EM
RI Emri, Miklós/AAK-3515-2020; Liposits, Gabor/ABC-9408-2021
OI Emri, Miklós/0000-0001-9884-9202; Liposits, Gabor/0000-0002-8204-3949;
   Kovacs, Arpad/0000-0002-8469-5764
CR ANCA LG, 2010, RADIOTHER ONCOL, V96, P325
   Aoyama H, 2004, INT J RADIAT ONCOL, V58, P1177, DOI 10.1016/j.ijrobp.2003.08.034
   BURMAN C, 1991, INT J RADIAT ONCOL, V21, P123, DOI 10.1016/0360-3016(91)90172-Z
   Chang J, 2008, MED DOSIM, V33, P42, DOI 10.1016/j.meddos.2007.05.002
   Flickinger JC, 2000, INT J RADIAT ONCOL, V46, P1143, DOI 10.1016/S0360-3016(99)00513-1
   Garcia A, 2003, J RADIOTHER PRACT, V3, P63
   Garcia-Alvarez R, 2006, J MAGN RESON IMAGING, V23, P108, DOI 10.1002/jmri.20493
   Gore JC, 2003, J CLIN INVEST, V112, P4, DOI 10.1172/JCI200319010
   Isambert A, 2008, RADIOTHER ONCOL, V87, P93, DOI 10.1016/j.radonc.2007.11.030
   Kovacs A, 2011, PATHOL ONCOL RES, V17, P207, DOI 10.1007/s12253-010-9298-y
   Langen KJ, 2003, NUCL MED BIOL, V30, P501, DOI 10.1016/S0969-8051(03)00023-4
   Liu WC, 2000, MED PHYS, V27, P1563, DOI 10.1118/1.599022
   MARKS JE, 1981, INT J RADIAT ONCOL, V7, P243, DOI 10.1016/0360-3016(81)90443-0
   MOONEN CTW, 1999, FUNCTIONAL MRI
   Ogawa K, 2004, INT J RADIAT ONCOL, V58, P705, DOI 10.1016/j.ijrobp.2003.07.001
   Pantelis E, 2010, INT J RADIAT ONCOL, V78, P257, DOI 10.1016/j.ijrobp.2009.10.064
   Pasciuti K, 2008, RADIOTHER ONCOL, V87, P110, DOI 10.1016/j.radonc.2007.12.019
   Voges J, 1996, INT J RADIAT ONCOL, V36, P1055, DOI 10.1016/S0360-3016(96)00422-1
   Weber WA, 2000, EUR J NUCL MED, V27, P542, DOI 10.1007/s002590050541
   Young RJ, 2010, NEUROIMAG CLIN N AM, V20, P311, DOI 10.1016/j.nic.2010.04.011
NR 20
TC 10
Z9 11
U1 0
U2 6
PU SPRINGER
PI NEW YORK
PA 233 SPRING ST, NEW YORK, NY 10013 USA
SN 0167-594X
EI 1573-7373
J9 J NEURO-ONCOL
JI J. Neuro-Oncol.
PD DEC
PY 2011
VL 105
IS 3
BP 629
EP 637
DI 10.1007/s11060-011-0633-2
PG 9
WC Oncology; Clinical Neurology
WE Science Citation Index Expanded (SCI-EXPANDED)
SC Oncology; Neurosciences & Neurology
GA 858MR
UT WOS:000297803100020
PM 21725803
DA 2023-08-27
ER

PT J
AU Tringali, S
   Dubreuil, C
   Zaouche, S
   Ferber-Viart, C
AF Tringali, Stephane
   Dubreuil, Christian
   Zaouche, Sandra
   Ferber-Viart, Chantal
TI Are Stage IV vestibular schwannomas preoperatively different from other
   stages?
SO OTOLOGY & NEUROTOLOGY
LA English
DT Article
DE auditory brainstem response; large vestibular schwannomas; preoperative
   factors; videonystamography
ID ACOUSTIC NEUROMAS; PRESERVATION; SURGERY
AB Objective: The aim of this study was to focus on the clinical and paraclinical symptoms of patients suffering from Stage IV vestibular schwannomas (VSs).
   Patients: In this prospective study, we included 734 patients who have VS and candidates for operation.
   Main Outcome Measures: Patients were classified as having Stage I, II, III, or IV tumors according to Tos criteria as evaluated by magnetic resonance imaging.
   Preoperative Clinical Evaluation: We recorded the occurrence of complaints (%) and duration (yr) of hearing loss, tinnitus, and balance disorder. Preoperative paraclinical evaluation included pure-tone (PTA) and speech audiometry, auditory brainstem response (ABR) patterns, and vestibular deficit at videonystamography (VNG). Continuous variables were compared between Stage IV and other stages using analysis of variance. Qualitative variables expressed as a percentage of presence were compared between Stage IV and other stages using percentage comparison.
   Results: Quantitative Parameters. Patients with Stage IV VS were significantly younger as compared with patients with other stages. Stage IV hearing loss was greater compared with other stages at 250 and 500 Hz but smaller at 2,000 and 8,000 Hz. We found no difference in the loss of PTA between Stage IV and the other stages. Speech discriminancy score was smaller in Stage IV. The durations of hearing loss, tinnitus, and balance disorders were similar whatever the tumor stage. Auditory brainstem response patterns showed no difference in Wave III latency between Stage IV VS and other stages, whereas Wave V latency and V-I interval were higher in Stage IV. Both ABR threshold and VNG caloric deficit were higher in Stage IV VS compared with other stages. Qualitative Parameters. The percentage of patients with Stage IV was lower than that with Stages II and III. The percentage of men and women was similar in all stages. The occurrence of hearing loss was similar in all stages, whereas that of tinnitus was lower in Stage IV compared with Stages I and II. In contrast, the occurrence of balance disorder was higher in Stage IV compared with all other stages.
   Conclusion: In clinical and paraclinical manifestation, Stage IV VS is different from the other stages. The PTA differences may be attributed to the younger age. Occurrence of clinical symptoms, ABR, and VNG pattern can be explained by the fact that Stage IV develops rapidly in the vestibular, rather than the cochlear nerve and by the fact that larger tumors can be cerebellar compression. This has been confirmed by the higher occurrence of balance disorders in Stage IV and the lower occurrence of tinnitus with similar hearing loss in all stages.
C1 [Tringali, Stephane; Dubreuil, Christian; Zaouche, Sandra] Hosp Civils Lyon, Ctr Hosp Lyon Sud, Serv Otoneurochirurg, F-69495 Pierre Benite, France.
   [Ferber-Viart, Chantal] Ctr Hosp Lyon Sud, F-69310 Pierre Benite, France.
   [Ferber-Viart, Chantal] UCB Lyon 1, Inst Fed Neurosci, UMR Neurosci & Syst Sensoriels, Lyon, France.
C3 CHU Lyon; CHU Lyon; UDICE-French Research Universities; Universite
   Claude Bernard Lyon 1
RP Tringali, S (通讯作者)，Hosp Civils Lyon, Ctr Hosp Lyon Sud, Serv Otoneurochirurg, F-69495 Pierre Benite, France.
EM
RI Tringali, Stephane/AAS-8368-2021
OI Tringali, Stephane/0000-0002-3236-4963
CR Anderson DE, 2005, J NEUROSURG, V102, P643, DOI 10.3171/jns.2005.102.4.0643
   Charabi S, 1996, ACTA OTO-LARYNGOL, V116, P59, DOI 10.3109/00016489609137713
   Ferber-Viart C, 2000, LARYNGOSCOPE, V110, P145, DOI 10.1097/00005537-200001000-00026
   Lanman TH, 1999, J NEUROSURG, V90, P617, DOI 10.3171/jns.1999.90.4.0617
   Matthies C, 1997, NEUROSURGERY, V40, P1
   Nicoucar K, 2006, J NEUROSURG, V105, P205, DOI 10.3171/jns.2006.105.2.205
   Rosenberg SI, 2000, LARYNGOSCOPE, V110, P497, DOI 10.1097/00005537-200004000-00002
   TOS M, 1992, ACOUSTIC NEUROMA, P975
   Yamakami I, 2004, J NEUROL NEUROSUR PS, V75, P453, DOI 10.1136/jnnp.2003.010827
NR 9
TC 10
Z9 10
U1 0
U2 0
PU LIPPINCOTT WILLIAMS & WILKINS
PI PHILADELPHIA
PA 530 WALNUT ST, PHILADELPHIA, PA 19106-3621 USA
SN 1531-7129
J9 OTOL NEUROTOL
JI Otol. Neurotol.
PD JAN
PY 2008
VL 29
IS 1
BP 46
EP 49
DI 10.1097/mao.0b013e31815c2a85
PG 4
WC Clinical Neurology; Otorhinolaryngology
WE Science Citation Index Expanded (SCI-EXPANDED)
SC Neurosciences & Neurology; Otorhinolaryngology
GA 249IT
UT WOS:000252222200010
PM 18046259
DA 2023-08-27
ER

PT J
AU Schmerber, S
   Lavieille, JP
   Dumas, G
   Herve, T
AF Schmerber, S
   Lavieille, JP
   Dumas, G
   Herve, T
TI Intraoperative auditory monitoring in vestibular schwannoma surgery: New
   trends
SO ACTA OTO-LARYNGOLOGICA
LA English
DT Article
DE auditory outcome; brainstem auditory-evoked potential; prognostic
   indicators; retrosigmoid approach
ID HEARING PRESERVATION; PROGNOSTIC-FACTORS; TUMOR SURGERY; RESECTION
AB Objective - To investigate the efficiency of a new method of brainstem auditory-evoked potential (BAEP) monitoring during complete vestibular schwannoma (VS) resection with attempted hearing preservation.
   Material and Methods - Dedicated software providing near real-time recording was developed using a rejection strategy of artifacts based on spectral analysis. A small sample number (maximum 200) is required and results are obtained within 10 s. Fourteen consecutive patients with hearing class A operated on for VS, in an attempt to preserve hearing, participated in the investigation. Postoperatively, 7 patients (50%) had useful hearing (hearing class A, 4/14; hearing class B, 3/14) on the operated side. Seven patients (50%) were reduced to hearing class D.
   Results - Drilling of the internal auditory canal (IAC) and tumor removal at the lateral end of the IAC were identified as the two most critical steps for achieving hearing preservation. Intraoperative BAEP monitoring was sensitive in detecting auditory damage with useful feedback but its effectiveness in preventing irreversible hearing impairment was not demonstrated in this study.
   Conclusion - Combined BAEP and direct auditory nerve monitoring using the same equipment will be performed in the future in an attempt to enhance the chances of preventing irreversible hearing damage, and possibly to improve the hearing outcome significantly. approach.
C1 Grenoble Hosp, Univ Dept Otorhinolaryngol, Grenoble, France.
C3 CHU Grenoble Alpes; Communaute Universite Grenoble Alpes; UDICE-French
   Research Universities; Universite Grenoble Alpes (UGA)
RP Schmerber, S (通讯作者)，Univ Dept Otorhinolaryngol, BP 217, FR-38043 Grenoble 9, France.
EM
RI SCHMERBER, Sebastien/D-1789-2016; SCHMERBER, Sébastien/AAM-6193-2021
OI SCHMERBER, Sebastien/0000-0001-5778-7201; SCHMERBER,
   Sebastien/0000-0003-0189-5574
CR Battista RA, 2000, AM J OTOL, V21, P244, DOI 10.1016/S0196-0709(00)80017-4
   Brackmann DE, 2000, AM J OTOL, V21, P417, DOI 10.1016/S0196-0709(00)80054-X
   COHEN NL, 1993, AM J OTOL, V14, P423
   Colletti V, 1997, OTOLARYNG HEAD NECK, V117, P596, DOI 10.1016/S0194-5998(97)70039-1
   Ferber-Viart C, 2000, LARYNGOSCOPE, V110, P145, DOI 10.1097/00005537-200001000-00026
   GLASSCOCK ME, 1993, J NEUROSURG, V78, P864, DOI 10.3171/jns.1993.78.6.0864
   Harner SG, 1996, AM J OTOL, V17, P150
   Hecht CS, 1997, LARYNGOSCOPE, V107, P1122, DOI 10.1097/00005537-199708000-00021
   Holsinger FC, 2000, AM J OTOL, V21, P695
   HOUSE JW, 1985, OTOLARYNG HEAD NECK, V93, P146, DOI 10.1177/019459988509300202
   Irving RM, 1998, J NEUROSURG, V88, P840, DOI 10.3171/jns.1998.88.5.0840
   Jackson LE, 2000, AM J OTOL, V21, P249, DOI 10.1016/S0196-0709(00)80018-6
   Kaga K, 1997, J LARYNGOL OTOL, V111, P967, DOI 10.1017/S002221510013909X
   KEMINK JL, 1990, LARYNGOSCOPE, V100, P597
   Koos W, 1976, CLIN MICRONEUROSURGE, P91
   KVETON JF, 1990, LARYNGOSCOPE, V100, P1171
   Levo H, 2002, ANN OTO RHINOL LARYN, V111, P392, DOI 10.1177/000348940211100502
   Magnan J, 2002, OTOL NEUROTOL, V23, P141, DOI 10.1097/00129492-200203000-00006
   Matthies C, 1997, NEUROSURGERY, V40, P469, DOI 10.1097/00006123-199703000-00009
   Matthies C, 1997, NEUROSURGERY, V40, P459, DOI 10.1097/00006123-199703000-00007
   MONSELL EM, 1995, OTOLARYNG HEAD NECK, V113, P179
   NADOL JB, 1992, LARYNGOSCOPE, V102, P1153, DOI 10.1288/00005537-199210000-00010
   SHELTON C, 1989, ARCH OTOLARYNGOL, V115, P1213
   Sunderland S, 1981, THE CRANIAL NERVES, P16, DOI [10.1007/978-3-642-67980-3_3, DOI 10.1007/978-3-642-67980-3_3]
   Zappia JJ, 1996, OTOLARYNG HEAD NECK, V115, P98, DOI 10.1016/S0194-5998(96)70144-4
NR 25
TC 11
Z9 21
U1 0
U2 0
PU TAYLOR & FRANCIS LTD
PI ABINGDON
PA 2-4 PARK SQUARE, MILTON PARK, ABINGDON OR14 4RN, OXON, ENGLAND
SN 0001-6489
EI 1651-2251
J9 ACTA OTO-LARYNGOL
JI Acta Oto-Laryngol.
PD JAN
PY 2004
VL 124
IS 1
BP 53
EP 61
DI 10.1080/00016480310014840
PG 9
WC Otorhinolaryngology
WE Science Citation Index Expanded (SCI-EXPANDED)
SC Otorhinolaryngology
GA 763RZ
UT WOS:000188115400012
PM 14977079
DA 2023-08-27
ER

PT J
AU Banks, WA
   Moinuddin, A
   Morley, JE
AF Banks, WA
   Moinuddin, A
   Morley, JE
TI Regional transport of TNF-alpha across the blood-brain barrier in young
   ICR and young and aged SAMP8 mice
SO NEUROBIOLOGY OF AGING
LA English
DT Article
DE blood-brain barrier; tumor necrosis factor-alpha; TNF; aging; SAM mice;
   SAMP8 mice; transport; brain; central nervous system; hypothalamus;
   cortex; striatum; midbrain; cytokine; protein; amyloid; learning;
   memory; cognition
ID TUMOR-NECROSIS-FACTOR; DIFFERENTIAL PERMEABILITY; MOUSE; BETA;
   MICROCIRCULATION; INTERLEUKIN-6; CAPILLARY; PEPTIDES; ALBUMIN; GLUCOSE
AB The blood-brain barrier (BBB) controls the exchange of regulatory substances, including tumor necrosis factor-a! (TNF), between the brain and the blood. Transport across the BBB of some regulatory substances is altered with aging. Here, we measured the blood to brain unidirectional influx rate (Ki) for whole brain and 10 brain regions for radioactively labeled TNF in three groups of mice: young (2 mo old) ICR (the standard outbred albino laboratory mouse also termed CD-1), young SAMP8 (a strain which develops impaired learning and memory with aging that correlates with an age-related increase in brain levels of amyloid beta protein), and aged 117 mo) SAMP8 mice. In ICR mice, the hypothalamus had the fastest (1.73 mul/g-min) and the parietal cortex the slowest (0.189 mul/g-min) rates of uptake, a regional difference of about 9 fold. No differences in transport into whole brain or brain regions occurred between the ICR and young SAMP8, showing a lack of differences between strains. Transport was higher for the occipital cortex, midbrain, and striatum in aged SAMP8 mice. These results show blood-borne TNF enters some regions of the brain much more readily than others and TNF transport is increased into some brain regions of the SAMP8 mice at an age when teaming and memory are impaired. (C) 2001 Elsevier Science Inc. All rights reserved.
C1 Vet Affairs Med Ctr, GRECC, St Louis, MO USA.
   St Louis Univ, Sch Med, Dept Internal Med, Div Geriatr, St Louis, MO 63106 USA.
C3 Geriatric Research Education & Clinical Center; US Department of
   Veterans Affairs; Veterans Health Administration (VHA); Saint Louis
   University
RP Banks, WA (通讯作者)，Vet Affairs Med Ctr, GRECC, St Louis, MO USA.
EM
RI Morley, John Edward/K-1570-2019; Banks, William A/K-1330-2017
OI Banks, William A/0000-0002-0496-3619
FU NIMH NIH HHS [R0-1 MH54979] Funding Source: Medline; NINDS NIH HHS [R0-1
   NS41863] Funding Source: Medline
CR Ando T, 1999, NEUROIMMUNOMODULAT, V6, P319, DOI 10.1159/000026391
   Banks WA, 1996, PEPTIDES, V17, P305, DOI 10.1016/0196-9781(96)00025-3
   Banks WA, 1998, PEPTIDES, V19, P883, DOI 10.1016/S0196-9781(98)00018-7
   Banks WA, 2000, J GERONTOL A-BIOL, V55, pB601, DOI 10.1093/gerona/55.12.B601
   BANKS WA, 1985, NEUROSCI LETT, V61, P171, DOI 10.1016/0304-3940(85)90420-3
   BANKS WA, 1995, PHARMACOL BIOCHEM BE, V51, P139, DOI 10.1016/0091-3057(94)00412-C
   Blanc EM, 1997, J NEUROCHEM, V68, P1870
   BLASBERG RG, 1983, J CEREBR BLOOD F MET, V3, P8, DOI 10.1038/jcbfm.1983.2
   BODNAR RJ, 1989, CANCER RES, V49, P6280
   DANIEL PM, 1978, J PHYSIOL-LONDON, V274, P141, DOI 10.1113/jphysiol.1978.sp012139
   DAVSON H, 1996, PHYSL CSF BLOOD BRAI, P303
   DELATORRE JC, 1993, NEUROL RES, V15, P146
   DELI MA, 1995, J NEUROSCI RES, V41, P717, DOI 10.1002/jnr.490410602
   FABRY Z, 1993, J NEUROIMMUNOL, V47, P23, DOI 10.1016/0165-5728(93)90281-3
   Fiala M, 1998, MOL MED, V4, P480, DOI 10.1007/BF03401753
   Flood JF, 1998, NEUROSCI BIOBEHAV R, V22, P1
   FLOOD JF, 1992, J GERONTOL, V47, pB52, DOI 10.1093/geronj/47.2.B52
   Friedman JM, 1998, NATURE, V395, P763, DOI 10.1038/27376
   FROLICH L, 1991, EUR ARCH PSY CLIN N, V240, P363, DOI 10.1007/BF02279767
   GRAMMAS P, 1995, DEMENTIA, V6, P126, DOI 10.1159/000106934
   GUTIERREZ EG, 1993, J NEUROIMMUNOL, V47, P169, DOI 10.1016/0165-5728(93)90027-V
   HANKES LV, 1991, ADV EXP MED BIOL, V294, P675
   HARIK SI, 1991, ANN NY ACAD SCI, V640, P47
   KALARIA RN, 1992, CEREBROVAS BRAIN MET, V4, P226
   Kety S.S., 1987, ENCY NEUROSCIENCE, V1, P206
   Male D, 1995, NEW CONCEPTS OF A BLOOD-BRAIN BARRIER, P311
   MANESS LM, 1995, BRAIN RES, V700, P83, DOI 10.1016/0006-8993(95)00913-B
   MANESS LM, 1994, LIFE SCI, V55, P1643, DOI 10.1016/0024-3205(94)00331-9
   Mattson MP, 1997, BRAIN RES REV, V23, P47, DOI 10.1016/S0165-0173(96)00014-8
   Mizushima H, 1999, LIFE SCI, V65, P2127, DOI 10.1016/S0024-3205(99)00477-4
   MOINUDDIN A, 2000, IN PRESS NEUROIMMUNO
   Munoz-Fernandez MA, 1998, PROG NEUROBIOL, V56, P307
   Nomura Y, 1996, ANN NY ACAD SCI, V786, P410, DOI 10.1111/j.1749-6632.1996.tb39080.x
   Pan WH, 1997, NEUROSCI BIOBEHAV R, V21, P603, DOI 10.1016/S0149-7634(96)00047-4
   Pan WH, 1996, AM J PHYSIOL-ENDOC M, V271, pE636, DOI 10.1152/ajpendo.1996.271.4.E636
   Pan WH, 1997, EXP NEUROL, V146, P367, DOI 10.1006/exnr.1997.6533
   PATLAK CS, 1983, J CEREBR BLOOD F MET, V3, P1, DOI 10.1038/jcbfm.1983.1
   Rosenberg GA, 1995, BRAIN RES, V703, P151, DOI 10.1016/0006-8993(95)01089-0
   Shah GN, 1997, EXP GERONTOL, V32, P501, DOI 10.1016/S0531-5565(96)00158-1
   Torelli GF, 1999, AM J PHYSIOL-REG I, V277, pR850, DOI 10.1152/ajpregu.1999.277.3.R850
   Uchida D, 1996, BRAIN RES, V736, P280, DOI 10.1016/S0006-8993(96)00716-0
   ZLOKOVIC BV, 1993, BIOCHEM BIOPH RES CO, V197, P1034, DOI 10.1006/bbrc.1993.2582
NR 42
TC 63
Z9 64
U1 0
U2 4
PU ELSEVIER SCIENCE INC
PI NEW YORK
PA STE 800, 230 PARK AVE, NEW YORK, NY 10169 USA
SN 0197-4580
EI 1558-1497
J9 NEUROBIOL AGING
JI Neurobiol. Aging
PD JUL-AUG
PY 2001
VL 22
IS 4
BP 671
EP 676
DI 10.1016/S0197-4580(01)00220-2
PG 6
WC Geriatrics & Gerontology; Neurosciences
WE Science Citation Index Expanded (SCI-EXPANDED)
SC Geriatrics & Gerontology; Neurosciences & Neurology
GA 453JF
UT WOS:000169912200017
PM 11445267
DA 2023-08-27
ER

PT J
AU Chino, K
   Tanyi, JA
   Stea, B
AF Chino, Kazunmi
   Tanyi, James A.
   Stea, Baldassarre
TI STEREOTACTIC RADIOTHERAPY FOR UNILATERAL ORBITAL LYMPHOMA AND ORBITAL
   PSEUDO-TUMORS: A PLANNING STUDY
SO MEDICAL DOSIMETRY
LA English
DT Article
DE Cataracts; Stereotactic IMRT; Orbital lymphoma; Grave's ophthalmopathy
ID GRAVES OPHTHALMOPATHY; TARGET LOCALIZATION; RADIATION-THERAPY; SYSTEM
AB Orbital lymphoma and Grave's ophthalmopathy (GO) are successfully treated with radiation therapy. The lenses are blocked to prevent cataract formation. However, blocking of the lens by traditional methods can be difficult for tumors located anteriorly and extending into the retrobulbar space. We present a series of 3 patients with orbital lymphoma and 1 with GO treated with stereotactic intensity modulated radiation therapy (IMRT) to spare normal structures, including the lens. Three patients with orbital lymphomas and I with unilateral GO were treated with IMRT using a linac with stereotactic capabilities. Avoidance structures included the ipsilateral lens and globe, the contralateral lens and globe, the chiasm, and the brainstem. Two patients with orbital lymphoma were treated with 24 Gy in 12 fractions, and 1 patient was treated with 30.6 Gy in 17 fractions. The patient with GO was treated with 20 Gy in 10 fractions. The dosimetry was analyzed to determine the dose to normal tissues. Patient follow-up varies between 8 and 24 months. The mean minimal dose to the ipsilateral lens was 13.6% of the total dose, the mean maximal dose was 33.5%, and the mean median dose was 19.3%. The mean median dose to the contralateral eye was 1.1 % of the total dose. The mean median dose to the chiasm was 14.9% of the total dose. The mean median dose to the brainstem was 1.9% of the total dose. No cataracts or other complications were noted in the 4 patients treated with this technique so far. IMRT gives a more conformal treatment to the orbital contents while sparing normal tissues such as the ipsilateral lens and adjacent critical structures. This should result in fewer complications such as cataracts. (C) 2009 American Association of Medical Dosimetrists.
C1 [Chino, Kazunmi] Univ Arizona, Hlth Sci Ctr, Dept Radiat Oncol, Tucson, AZ 85724 USA.
   Oregon Hlth & Sci Univ, Dept Radiat Med, Portland, OR 97201 USA.
C3 University of Arizona; University of Arizona Health Sciences; Oregon
   Health & Science University
RP Chino, K (通讯作者)，Univ Arizona, Hlth Sci Ctr, Dept Radiat Oncol, Tucson, AZ 85724 USA.
EM
CR Alonso-Arrizabalaga S, 2007, INT J RADIAT ONCOL, V69, P936, DOI 10.1016/j.ijrobp.2007.06.063
   [Anonymous], 2002, AJCC CANC STAGING HD
   AUSTINSEYMOUR MM, 1985, INT J RADIAT ONCOL, V11, P371, DOI 10.1016/0360-3016(85)90160-9
   BAHN RS, 1993, NEW ENGL J MED, V329, P1468
   Bartalena L, 2000, ENDOCR REV, V21, P168, DOI 10.1210/er.21.2.168
   Beckendorf V, 1999, INT J RADIAT ONCOL, V43, P805, DOI 10.1016/S0360-3016(98)00405-2
   CHRISTENSON J, 2007, EUR SOC THER RAD ONC
   EMAMI B, 1991, INT J RADIAT ONCOL, V21, P109, DOI 10.1016/0360-3016(91)90171-Y
   JEREB B, 1984, INT J RADIAT ONCOL, V10, P1013, DOI 10.1016/0360-3016(84)90172-X
   Jin JY, 2006, MED PHYS, V33, P4557, DOI 10.1118/1.2392605
   PEREZ CA, 2004, PRINCIPLES PRACTICE, P884
   Petric MP, 2005, J APPL CLIN MED PHYS, V6, P63, DOI 10.1120/jacmp.2025.25353
   Verellen D, 2003, RADIOTHER ONCOL, V67, P129, DOI 10.1016/S0167-8140(02)00385-7
   Zhou P, 2005, INT J RADIAT ONCOL, V63, P866, DOI 10.1016/j.ijrobp.2005.03.005
NR 14
TC 5
Z9 5
U1 0
U2 1
PU ELSEVIER SCIENCE INC
PI NEW YORK
PA 360 PARK AVE SOUTH, NEW YORK, NY 10010-1710 USA
SN 0958-3947
J9 MED DOSIM
JI Med. Dosim.
PD SPR
PY 2009
VL 34
IS 1
BP 57
EP 62
DI 10.1016/j.meddos.2007.12.004
PG 6
WC Oncology; Radiology, Nuclear Medicine & Medical Imaging
WE Science Citation Index Expanded (SCI-EXPANDED)
SC Oncology; Radiology, Nuclear Medicine & Medical Imaging
GA 413SK
UT WOS:000263812600010
PM 19181257
DA 2023-08-27
ER

PT J
AU Kamoun, M
   Mnif, MF
   Rekik, N
   Belguith, N
   Charfi, N
   Mnif, L
   Elleuch, M
   Mnif, F
   Kamoun, T
   Mnif, Z
   Kamoun, H
   Sellami-Boudawara, T
   Hachicha, M
   Abid, M
AF Kamoun, Mandi
   Mnif, Mouna Feki
   Rekik, Nabila
   Belguith, Neila
   Charfi, Nadia
   Mnif, Lilia
   Elleuch, Mouna
   Mnif, Fatma
   Kamoun, Thouraya
   Mnif, Zeinab
   Kamoun, Hassen
   Sellami-Boudawara, Tahia
   Hachicha, Mongia
   Abid, Mohamed
TI Ganglioneuroma of adrenal gland in a patient with Turner syndrome
SO ANNALS OF DIAGNOSTIC PATHOLOGY
LA English
DT Article
DE Adrenal ganglioneuroma; Neurogenic tumors; Turner syndrome
ID GROWTH-HORMONE THERAPY; NEUROBLASTOMA; CARCINOMA; GONADOBLASTOMA;
   NEOPLASIA
AB A 15-year-old girl with Turner syndrome was unexpectedly found to have a left suprarenal mass. Extensive investigations showed a clinically and biochemically inapparent mass. Computed tomography disclosed a well-defined solid lesion in the left adrenal measuring 6.5 x 5 cm with minimal contrast enhancement. Laparoscopic adrenalectomy was done. Histologic examination revealed an encapsulated mass originated from the left adrenal medulla. Tumor tissue comprised abundant collagen fibers and spindloid cells admixed with mature ganglion cells. The tumor was diagnosed as left adrenal ganglioneuroma. According to literature, we report the eighth case of ganglioneuroma complicating Turner syndrome. Patients with this syndrome are predisposed to the development of neuroblastoma and related tumors. Reasons for this predisposition might relate to genetic and hormonal factors. Given that these tumors are often limited stage and of good prognosis, we recommend their screening in all patients with Turner syndrome. Published by Elsevier Inc.
C1 [Kamoun, Mandi; Mnif, Mouna Feki; Rekik, Nabila; Charfi, Nadia; Elleuch, Mouna; Mnif, Fatma; Abid, Mohamed] Hedi Chaker Hosp, Dept Endocrinol, Sfax 3029, Tunisia.
   [Belguith, Neila; Kamoun, Hassen] Fac Med, Dept Human Mol Genet, Sfax 3029, Tunisia.
   [Mnif, Lilia; Sellami-Boudawara, Tahia] Habib Bourguiba Hosp, Dept Anat Pathol, Sfax 3029, Tunisia.
   [Kamoun, Thouraya; Hachicha, Mongia] Hedi Chaker Hosp, Dept Pediat, Sfax 3029, Tunisia.
   [Mnif, Zeinab] Hedi Chaker Hosp, Dept Radiol, Sfax 3029, Tunisia.
C3 Universite de Sfax; Hopital Hedi Chaker; Universite de Sfax; Universite
   de Sfax; Hopital Habib Bourguiba; Universite de Sfax; Hopital Hedi
   Chaker; Universite de Sfax; Hopital Hedi Chaker
RP Mnif, MF (通讯作者)，Hedi Chaker Hosp, Dept Endocrinol, Sfax 3029, Tunisia.
EM
RI Aseeri, Mohammed/P-1495-2018
OI kamoun, hassen/0000-0002-1798-3862
CR Bianco B, 2009, INT J GYNECOL PATHOL, V28, P197, DOI 10.1097/PGP.0b013e318186a825
   Blatt J, 1997, J PEDIATR-US, V131, P666, DOI 10.1016/S0022-3476(97)70090-8
   Brant WO, 2006, J UROLOGY, V175, P1858, DOI 10.1016/S0022-5347(05)00932-8
   Cabanas P, 2005, EUR J ENDOCRINOL, V153, P499, DOI 10.1530/eje.1.01988
   Connors MH, 1996, NEUROSURGERY, V39, P1243, DOI 10.1097/00006123-199612000-00037
   Gravholt CH, 2008, LANCET ONCOL, V9, P193, DOI 10.1016/S1470-2045(08)70045-7
   Matsuoka H, 1997, ACTA PAEDIATR JAPON, V39, P628
   Maweja S, 2007, ACTA CHIR BELG, V107, P670, DOI 10.1080/00015458.2007.11680144
   Maweja S, 2007, AM J SURG, V194, P683, DOI 10.1016/j.amjsurg.2007.01.034
   MILLER RW, 1968, AM J DIS CHILD, V115, P253, DOI 10.1001/archpedi.1968.02100010255015
   Morotti RA, 2007, SEMIN LIVER DIS, V27, P427, DOI 10.1055/s-2007-991518
   Park JK, 2008, J OBSTET GYNAECOL RE, V34, P745, DOI 10.1111/j.1447-0756.2008.00918.x
   Sasaki Y, 2000, J PEDIAT HEMATOL ONC, V22, P89, DOI 10.1097/00043426-200001000-00020
   Satge D, 2003, CANCER GENET CYTOGEN, V147, P89, DOI 10.1016/S0165-4608(03)00203-6
   Satge D, 1998, CANCER RES, V58, P448
   Schoemaker MJ, 2008, LANCET ONCOL, V9, P239, DOI 10.1016/S1470-2045(08)70033-0
   Sivakumaran T. A., 1999, Journal of Environmental Pathology Toxicology and Oncology, V18, P339
   Stasik Chad N, 2005, Endocr Pract, V11, P382
   Stochholm K, 2006, J CLIN ENDOCR METAB, V91, P3897, DOI 10.1210/jc.2006-0558
   WERTELECKI W, 1970, CANCER-AM CANCER SOC, V26, P485, DOI 10.1002/1097-0142(197008)26:2<485::AID-CNCR2820260233>3.0.CO;2-3
NR 20
TC 14
Z9 17
U1 0
U2 0
PU ELSEVIER SCIENCE INC
PI NEW YORK
PA 360 PARK AVE SOUTH, NEW YORK, NY 10010-1710 USA
SN 1092-9134
EI 1532-8198
J9 ANN DIAGN PATHOL
JI Ann. Diagn. Pathol.
PD APR
PY 2010
VL 14
IS 2
BP 133
EP 136
DI 10.1016/j.anndiagpath.2009.06.007
PG 4
WC Pathology
WE Science Citation Index Expanded (SCI-EXPANDED)
SC Pathology
GA 574FA
UT WOS:000275972900010
PM 20227019
DA 2023-08-27
ER

PT J
AU Pacholke, HD
   Amdur, RJ
   Louis, DA
   Yang, H
   Mendenhall, WM
AF Pacholke, HD
   Amdur, RJ
   Louis, DA
   Yang, H
   Mendenhall, WM
TI The role of intensity modulated radiation therapy for favorable stage
   tumor of the nasal cavity or ethmoid sinus
SO AMERICAN JOURNAL OF CLINICAL ONCOLOGY-CANCER CLINICAL TRIALS
LA English
DT Article
DE radiation therapy; intensity modulated radiation therapy; nasal cavity;
   paranasal sinus
ID IMRT; RADIOTHERAPY; CARCINOMA
AB Objectives: To compare intensity-modulated radiation therapy (IMRT), 4-field conformal, and the standard 3-field conventional technique of radiotherapy for favorable stage tumors of the nasal cavity or ethmoid sinus.
   Methods and Materials: We compared the 3 techniques in 3 patients with tumors of the nasal cavity or ethmoid sinus that did not involve the eye or optic pathways. We required that each plan deliver the prescription dose (70.2 Gy at 1.8 Gy per fraction) to 95% of the planning target volume. We compared the maximum point dose to critical normal structures (brainstem, optic chiasm, optic nerves, retina, lens).
   Results: IMRT and the 4-field conformal technique were clearly better than the 3-field conventional technique. The 4-field conformal plan was as good as IMRT.
   Conclusions: The conventional 3-field technique is not the best way to treat most patients with tumors between the eyes. A 4-field conformal plan is an excellent alternative to IMRT for some patients with tumors of the nasal cavity or ethmoid sinus. Conformal radiotherapy with a noncoplaner field that exits into the low neck may make it difficult to electively irradiate the neck lymphatics. IMRT may be a better option in this situation.
C1 Univ Florida, Coll Med, Dept Radiat Oncol, Gainesville, FL 32610 USA.
C3 State University System of Florida; University of Florida
RP Mendenhall, WM (通讯作者)，Univ Florida, Coll Med, Dept Radiat Oncol, POB 100385,2000 SW Archer Rd, Gainesville, FL 32610 USA.
EM
CR Adams EJ, 2001, INT J RADIAT ONCOL, V51, P579, DOI 10.1016/S0360-3016(01)01655-8
   Claus F, 2001, INT J RADIAT ONCOL, V51, P318, DOI 10.1016/S0360-3016(01)01627-3
   Huang D, 2003, INT J RADIAT ONCOL, V56, P158, DOI 10.1016/S0360-3016(03)00080-4
   Mock U, 2004, INT J RADIAT ONCOL, V58, P147, DOI 10.1016/S0360-3016(03)01452-4
   Tsien C, 2003, INT J RADIAT ONCOL, V55, P776, DOI 10.1016/S0360-3016(02)04274-8
NR 5
TC 16
Z9 16
U1 0
U2 0
PU LIPPINCOTT WILLIAMS & WILKINS
PI PHILADELPHIA
PA TWO COMMERCE SQ, 2001 MARKET ST, PHILADELPHIA, PA 19103 USA
SN 0277-3732
EI 1537-453X
J9 AM J CLIN ONCOL-CANC
JI Am. J. Clin. Oncol.-Cancer Clin. Trials
PD OCT
PY 2005
VL 28
IS 5
BP 474
EP 478
DI 10.1097/01.coc.0000182600.51019.de
PG 5
WC Oncology
WE Science Citation Index Expanded (SCI-EXPANDED)
SC Oncology
GA 971TP
UT WOS:000232407900011
PM 16199987
DA 2023-08-27
ER

PT J
AU Zhu, QQ
   Wang, ZQ
   Zhu, WR
   Chen, WX
   Wu, JT
AF Zhu, Qing-Qiang
   Wang, Zhong-Qiu
   Zhu, Wen-rong
   Chen, Wen-xin
   Wu, Jing-tao
TI The multislice CT findings of renal carcinoma associated with XP11.2
   translocation/TFE gene fusion and collecting duct carcinoma
SO ACTA RADIOLOGICA
LA English
DT Article
DE Xp11.2-Translocation/TFE gene fusion; renal cell carcinoma; collecting
   duct carcinoma; computed tomography; CT
ID CELL CARCINOMA; FEATURES; DIFFERENTIATION; ONCOCYTOMAS; PATTERNS;
   KIDNEY; TUMORS
AB Background: Renal cell carcinoma associated with Xp11.2 translocation and TFE gene fusion (Xp11.2/TFE RCC), and collecting duct carcinoma (CDC) are uncommon subtypes of renal cell carcinomas.
   Purpose: To investigate the multislice CT (MSCT) characteristics of these two tumor types.
   Material and Methods: Nine patients with Xp11.2/TFE RCC and 10 patients with CDC were studied retrospectively. MSCT was undertaken to investigate differences in tumor characteristics and enhancement patterns.
   Results: All patients had single tumors centered in the renal medulla. Two patients with each tumor type had lymph node involvement and there was a single case of hepatic metastasis (Xp11.2/TFE RCC). The mean tumor diameter of Xp11.2/TFE RCC tumors was significantly larger than for CDC tumors. Two patients with Xp11.2/TFE RCC had cystic components as did eight patients with CDC (P < 0.05). Calcifications were present in six patients, each with CDC. Clear tumor boundaries were visible in two patients with CDC and in nine with Xp11.2/TFE RCC (P < 0.05). The density of Xp11.2/TFE RCC tumors was greater than that of CDC tumors, normal renal cortex, or medulla on unenhanced CT. Enhancement was higher with Xp11.2/TFE RCC than with CDC tumors during all phases. Xp11.2/TFE RCC enhancement was higher than in the renal medulla during cortical and medullary phase but lower than in normal renal medulla during the delayed phase. CDC tumor enhancement was lower than that for normal renal medulla during all enhanced phases.
   Conclusion: Both tumor types originated from the renal medulla. Distinguishing features included density on unenhanced CT, enhancement patterns, and capsule signs. Identifying these differences may aid diagnosis.
C1 [Zhu, Qing-Qiang; Zhu, Wen-rong; Chen, Wen-xin; Wu, Jing-tao] Subei Peoples Hosp, Sch Clin, Coll Med, Yangzhou, Peoples R China.
   [Wang, Zhong-Qiu] Tongji Univ, East Hosp, Sch Med, Dept Radiol, Shanghai 200092, Peoples R China.
C3 Tongji University
RP Wu, JT (通讯作者)，Subei Peoples Hosp, Sch Clin, Coll Med, Yangzhou, Peoples R China.
EM
RI Wang, Zhong-Qiu/ABC-5782-2021; Wu, Jing/GZK-5063-2022
OI Wang, Zhong-Qiu/0000-0002-4204-9430; 
CR ABERCROMBIE JF, 1992, J ROY SOC MED, V85, P625
   Argani P, 2003, AM J SURG PATHOL, V27, P750, DOI 10.1097/00000478-200306000-00005
   Argani P, 2010, AM J SURG PATHOL, V34, P1295, DOI 10.1097/PAS.0b013e3181e8ce5b
   Argani P, 2009, AM J SURG PATHOL, V33, P609, DOI 10.1097/PAS.0b013e31818fbdff
   Armah HB, 2010, ARCH PATHOL LAB MED, V134, P124, DOI 10.1043/2008-0391-RSR.1
   Blitman NM, 2005, AM J ROENTGENOL, V185, P268, DOI 10.2214/ajr.185.1.01850268
   El-Sharkawy MS, 2007, INT UROL NEPHROL, V39, P929, DOI 10.1007/s11255-007-9224-8
   Eren S, 2006, EUR J RADIOL, V59, P93, DOI 10.1016/j.ejrad.2006.01.003
   Farah Raymond, 2005, Ann Diagn Pathol, V9, P46, DOI 10.1053/j.anndiagpath.2004.10.007
   Gakis G, 2011, EUR J RADIOL, V80, P274, DOI 10.1016/j.ejrad.2010.06.049
   Gong Yun, 2003, Arch Pathol Lab Med, V127, pe38
   HONDA H, 1992, UROL RADIOL, V14, P148, DOI 10.1007/BF02926917
   Jayasinghe C, 2010, KLIN PADIATR, V222, P187, DOI 10.1055/s-0030-1252011
   Kato H, 2011, J MAGN RESON IMAGING, V33, P217, DOI 10.1002/jmri.22392
   KENNEDY SM, 1990, HUM PATHOL, V21, P449, DOI 10.1016/0046-8177(90)90209-N
   Kim JK, 2004, HINYOKIKA KIYO, V50, P865
   Lopez-Beltran A, 2006, EUR UROL, V49, P798, DOI 10.1016/j.eururo.2005.11.035
   Low G, 2012, INT J UROL, V19, P90, DOI 10.1111/j.1442-2042.2011.02885.x
   Matz LR, 1997, PATHOLOGY, V29, P354, DOI 10.1080/00313029700169305
   Meyer PN, 2007, AM J CLIN PATHOL, V128, P70, DOI 10.1309/LR5G1VMXPY3G0CUK
   Parikh J, 2009, RARE TUMORS, V1, DOI 10.4081/rt.2009.e53
   Pickhardt PJ, 2001, AM J ROENTGENOL, V176, P627, DOI 10.2214/ajr.176.3.1760627
   Ross H, 2010, PATHOLOGY, V42, P369, DOI 10.3109/00313021003767348
   RUMPELT HJ, 1991, HISTOPATHOLOGY, V18, P115, DOI 10.1111/j.1365-2559.1991.tb01453.x
   Ruppert-Kohlmayr AJ, 2004, AM J ROENTGENOL, V183, P1387, DOI 10.2214/ajr.183.5.1831387
   Sauk SC, 2011, RADIOLOGY, V261, P854, DOI 10.1148/radiol.11101508
   Silverman SG, 2007, RADIOGRAPHICS, V27, P1131, DOI 10.1148/rg.274065147
   Skolarus TA, 2008, J UROLOGY, V179, P439, DOI 10.1016/j.juro.2007.09.076
   Soyer P, 1997, EUR RADIOL, V7, P224, DOI 10.1007/s003300050140
   Tsuda K, 2005, INT J UROL, V12, P795, DOI 10.1111/j.1442-2042.2005.01126.x
   Winarti NW, 2008, INT J SURG PATHOL, V16, P66, DOI 10.1177/1066896907304994
   Yoon SK, 2006, EUR J RADIOL, V57, P453, DOI 10.1016/j.ejrad.2005.09.009
NR 32
TC 16
Z9 18
U1 0
U2 9
PU SAGE PUBLICATIONS LTD
PI LONDON
PA 1 OLIVERS YARD, 55 CITY ROAD, LONDON EC1Y 1SP, ENGLAND
SN 0284-1851
EI 1600-0455
J9 ACTA RADIOL
JI Acta Radiol.
PD APR
PY 2013
VL 54
IS 3
BP 355
EP 362
DI 10.1258/ar.2012.120255
PG 8
WC Radiology, Nuclear Medicine & Medical Imaging
WE Science Citation Index Expanded (SCI-EXPANDED)
SC Radiology, Nuclear Medicine & Medical Imaging
GA 190EN
UT WOS:000322321900018
PM 23446748
DA 2023-08-27
ER

PT J
AU Klonou, A
   Spiliotakopoulou, D
   Themistocleous, MS
   Piperi, C
   Papavassiliou, AG
AF Klonou, Alexia
   Spiliotakopoulou, Danai
   Themistocleous, Marios S.
   Piperi, Christina
   Papavassiliou, Athanasios G.
TI Chromatin remodeling defects in pediatric brain tumors
SO ANNALS OF TRANSLATIONAL MEDICINE
LA English
DT Review
DE Histones; methyltransferases; epigenetic; gliomas; medulloblastoma (MB);
   ependymoma (EPN)
ID INTRINSIC PONTINE GLIOMA; HIGH-GRADE GLIOMAS; HISTONE LYSINE
   METHYLATION; PHASE-I CONSORTIUM; MOLECULAR SUBGROUPS; DISTINCT
   SUBGROUPS; MUTATIONS; MEDULLOBLASTOMA; GENES; EZH2
AB Brain tumors are regarded as the most prevalent solid neoplasms in children and the principal reason of death in this population. Even though surgical resection, radiotherapy and chemotherapy have improved outcome, a significant number of patients die in 6-12 months after diagnosis while those who survive, frequently experience side effects and relapses. Several studies suggest that many types of cancer including pediatric brain tumors are characterized by alterations in epigenetic profiles with deregulated chromatin remodeling and posttranslational covalent histone modifications playing a prominent role. Moreover, interplay of genetic and epigenetic changes has been associated to tumor growth and invasion as well as to modulation of patient's response to current treatment. Therefore, detection of tumor-specific histone changes and elucidation of the underlying gene defects will allow successful tailoring of personalized treatment. The goal of this review is to provide an update of genetic and epigenetic alterations that characterize pediatric brain tumors focusing on histone modifications, aiming at directing future molecular and epigenetic therapeutic targeting.
C1 [Klonou, Alexia; Spiliotakopoulou, Danai; Piperi, Christina; Papavassiliou, Athanasios G.] Univ Athens, Dept Biol Chem, Med Sch, 75 M Asias St, Athens 11527, Greece.
   [Themistocleous, Marios S.] Childrens Hosp Agia Sofia, Dept Neurosurg, Athens, Greece.
C3 CIVIS; National & Kapodistrian University of Athens
RP Papavassiliou, AG (通讯作者)，Univ Athens, Dept Biol Chem, Med Sch, 75 M Asias St, Athens 11527, Greece.
EM
RI Karamouzis, Michalis/AAD-2860-2020; Piperi, Christina/AAF-2009-2020
OI Piperi, Christina/0000-0002-2701-0618
CR Bandopadhayay P, 2014, CLIN CANCER RES, V20, P912, DOI 10.1158/1078-0432.CCR-13-2281
   Bayliss J, 2016, SCI TRANSL MED, V8, DOI 10.1126/scitranslmed.aah6904
   Bender S, 2013, CANCER CELL, V24, P660, DOI 10.1016/j.ccr.2013.10.006
   Bergthold G, 2014, BBA-REV CANCER, V1845, P294, DOI 10.1016/j.bbcan.2014.02.004
   Bernstein BE, 2006, CELL, V125, P315, DOI 10.1016/j.cell.2006.02.041
   Bezecny P, 2014, MED ONCOL, V31, DOI 10.1007/s12032-014-0985-5
   Bjerke L, 2013, CANCER DISCOV, V3, P512, DOI 10.1158/2159-8290.CD-12-0426
   Bunt J, 2013, ACTA NEUROPATHOL, V125, P385, DOI 10.1007/s00401-012-1069-2
   Bunt J, 2012, INT J CANCER, V131, pE21, DOI 10.1002/ijc.26474
   Chamdine Omar, 2014, CNS Oncol, V3, P433, DOI 10.2217/cns.14.43
   Chan KM, 2013, GENE DEV, V27, P985, DOI 10.1101/gad.217778.113
   Ciarapica R, 2014, BMC CANCER, V14, DOI 10.1186/1471-2407-14-139
   Dang Mai, 2017, Continuum (Minneap Minn), V23, P1727, DOI 10.1212/CON.0000000000000545
   Delcuve GP, 2009, J CELL PHYSIOL, V219, P243, DOI 10.1002/jcp.21678
   DeSouza RM, 2014, FRONT ONCOL, V4, DOI 10.3389/fonc.2014.00176
   Khuong-Quang DA, 2012, ACTA NEUROPATHOL, V124, P439, DOI 10.1007/s00401-012-0998-0
   Dubuc AM, 2013, ACTA NEUROPATHOL, V125, P373, DOI 10.1007/s00401-012-1070-9
   Faria CMC, 2011, J NEUROSURG-PEDIATR, V8, P119, DOI 10.3171/2011.5.PEDS1140
   Feinberg AP, 2004, NAT REV CANCER, V4, P143, DOI 10.1038/nrc1279
   Fontebasso AM, 2014, NAT GENET, V46, P462, DOI 10.1038/ng.2950
   Fontebasso AM, 2013, ACTA NEUROPATHOL, V125, P659, DOI 10.1007/s00401-013-1095-8
   Forst DA, 2014, ONCOLOGIST, V19, P403, DOI 10.1634/theoncologist.2013-0345
   Fouladi M, 2010, J CLIN ONCOL, V28, P3623, DOI 10.1200/JCO.2009.25.9119
   Gajjar A, 2015, J CLIN ONCOL, V33, P2986, DOI 10.1200/JCO.2014.59.9217
   Garcia MA, 2016, FUTURE ONCOL, V12, P1493, DOI 10.2217/fon-2016-0039
   Grasso CS, 2015, NAT MED, V21, P827, DOI 10.1038/nm0715-827a
   Haider BA, 2014, PLOS ONE, V9, DOI 10.1371/journal.pone.0089565
   Hashizume R, 2014, NAT MED, V20, P1394, DOI 10.1038/nm.3716
   Hovestadt V, 2014, NATURE, V510, P537, DOI 10.1038/nature13268
   Hummel TR, 2013, PEDIATR BLOOD CANCER, V60, P1452, DOI 10.1002/pbc.24541
   Huse JT, 2015, J CHILD NEUROL, V30, P1838, DOI 10.1177/0883073815579709
   Johnson A, 2017, ONCOLOGIST, V22, P1478, DOI 10.1634/theoncologist.2017-0242
   Johnson KJ, 2014, CANCER EPIDEM BIOMAR, V23, P2716, DOI 10.1158/1055-9965.EPI-14-0207
   Jones C, 2012, NAT REV CLIN ONCOL, V9, P400, DOI 10.1038/nrclinonc.2012.87
   Jones DTW, 2013, BRAIN PATHOL, V23, P193, DOI 10.1111/bpa.12019
   Kaplan Frederick S, 2009, Hum Mutat, V30, P379, DOI 10.1002/humu.20868
   Kieran MW, 2014, NEURO-ONCOLOGY, V16, P1037, DOI 10.1093/neuonc/nou109
   Kleer CG, 2003, P NATL ACAD SCI USA, V100, P11606, DOI 10.1073/pnas.1933744100
   Klonou A, 2017, NEUROMOL MED, V19, P256, DOI 10.1007/s12017-017-8455-9
   Kolb EA, 2010, PEDIATR BLOOD CANCER, V55, P668, DOI 10.1002/pbc.22576
   Kool M, 2012, ACTA NEUROPATHOL, V123, P473, DOI 10.1007/s00401-012-0958-8
   Kool M, 2008, PLOS ONE, V3, DOI 10.1371/journal.pone.0003088
   Kramm CM, 2011, NEURO-ONCOLOGY, V13, P680, DOI 10.1093/neuonc/nor045
   Krueger DA, 2013, NEUROLOGY, V80, P574, DOI 10.1212/WNL.0b013e3182815428
   Ku M., 2008, PLOS GENET, V4, P1
   Le LQ, 2007, ONCOGENE, V26, P4609, DOI 10.1038/sj.onc.1210261
   Lewis PW, 2013, SCIENCE, V340, P857, DOI 10.1126/science.1232245
   Liu KW, 2017, SCI SIGNAL, V10, DOI 10.1126/scisignal.aaf7593
   Liu XY, 2012, ACTA NEUROPATHOL, V124, P615, DOI 10.1007/s00401-012-1031-3
   Mack SC, 2014, NATURE, V506, P445, DOI 10.1038/nature13108
   Mackay A, 2017, CANCER CELL, V32, P520, DOI 10.1016/j.ccell.2017.08.017
   Maury E, 2017, EPIGENETICS-US, V12, P353, DOI 10.1080/15592294.2016.1278095
   Mendenhall EM, 2010, PLOS GENET, V6, DOI 10.1371/journal.pgen.1001244
   Milde T, 2010, CLIN CANCER RES, V16, P3240, DOI 10.1158/1078-0432.CCR-10-0395
   Mohammad F, 2017, NAT MED, V23, P483, DOI 10.1038/nm.4293
   La Madrid AM, 2015, FRONT ONCOL, V5, DOI 10.3389/fonc.2015.00148
   Muscal JA, 2013, PEDIATR BLOOD CANCER, V60, P390, DOI 10.1002/pbc.24271
   Northcott PA, 2017, NATURE, V547, P311, DOI 10.1038/nature22973
   Northcott PA, 2014, NATURE, V511, P428, DOI 10.1038/nature13379
   Northcott PA, 2009, NAT GENET, V41, P465, DOI 10.1038/ng.336
   Olow A, 2016, CLIN CANCER RES, V22, P5312, DOI 10.1158/1078-0432.CCR-15-1101
   Panditharatna E, 2015, CANCER GENET-NY, V208, P367, DOI 10.1016/j.cancergen.2015.04.008
   Parker M, 2014, NATURE, V506, P451, DOI 10.1038/nature13109
   Paugh BS, 2013, CANCER RES, V73, P6219, DOI 10.1158/0008-5472.CAN-13-1491
   Paugh BS, 2010, J CLIN ONCOL, V28, P3061, DOI 10.1200/JCO.2009.26.7252
   Penman CL, 2015, FRONT ONCOL, V5, DOI 10.3389/fonc.2015.00054
   Perez-Ramirez M, 2016, CLIN NEUROL NEUROSUR, V144, P53, DOI 10.1016/j.clineuro.2016.02.041
   Pfister S, 2008, INT J CANCER, V122, P1207, DOI 10.1002/ijc.23283
   Populo H, 2012, INT J MOL SCI, V13, P1886, DOI 10.3390/ijms13021886
   Raabe E, 2013, CLIN CANCER RES, V19, P4553, DOI 10.1158/1078-0432.CCR-13-0662
   Reddington JP, 2013, GENOME BIOL, V14, DOI 10.1186/gb-2013-14-3-r25
   Rizzo D, 2015, BIOMED RES INT, V2015, DOI 10.1155/2015/215135
   Robinson G, 2012, NATURE, V488, P43, DOI 10.1038/nature11213
   Roussel MF, 2018, CEREBELLUM, V17, P28, DOI 10.1007/s12311-017-0899-9
   Saratsis AM, 2014, ACTA NEUROPATHOL, V127, P881, DOI 10.1007/s00401-013-1218-2
   Schuettengruber B, 2011, NAT REV MOL CELL BIO, V12, P799, DOI 10.1038/nrm3230
   Schwalbe EC, 2017, LANCET ONCOL, V18, P958, DOI 10.1016/S1470-2045(17)30243-7
   Schwartzentruber J, 2012, NATURE, V482, P226, DOI [10.1038/nature10833, 10.1007/s13311-012-0137-6]
   Shi JW, 2014, MOL CELL, V54, P728, DOI 10.1016/j.molcel.2014.05.016
   Sievert AJ, 2013, P NATL ACAD SCI USA, V110, P5957, DOI 10.1073/pnas.1219232110
   Sneeringer CJ, 2010, P NATL ACAD SCI USA, V107, P20980, DOI 10.1073/pnas.1012525107
   Sobol-Milejska G, 2017, ADV CLIN EXP MED, V26, P571, DOI 10.17219/acem/62320
   Sparmann A, 2006, NAT REV CANCER, V6, P846, DOI 10.1038/nrc1991
   Staedtke V, 2016, TRENDS CANCER, V2, P338, DOI 10.1016/j.trecan.2016.06.003
   Sturm D, 2012, CANCER CELL, V22, P425, DOI 10.1016/j.ccr.2012.08.024
   Su JM, 2011, CLIN CANCER RES, V17, P589, DOI 10.1158/1078-0432.CCR-10-0738
   Swartling FJ, 2012, CANCER CELL, V21, P601, DOI 10.1016/j.ccr.2012.04.012
   Tang YJ, 2014, NAT MED, V20, P732, DOI 10.1038/nm.3613
   Taylor KR, 2014, NAT GENET, V46, P457, DOI 10.1038/ng.2925
   Taylor MD, 2012, ACTA NEUROPATHOL, V123, P465, DOI 10.1007/s00401-011-0922-z
   Venneti S, 2013, BRAIN PATHOL, V23, P558, DOI 10.1111/bpa.12042
   Veringa SJE, 2013, PLOS ONE, V8, DOI 10.1371/journal.pone.0061512
   Wells Elizabeth M, 2015, Continuum (Minneap Minn), V21, P373, DOI 10.1212/01.CON.0000464176.96311.d1
   Williams Maria J, 2017, Front Oncol, V7, P45, DOI 10.3389/fonc.2017.00045
   Wu G, 2014, NAT GENET, V46, P444, DOI 10.1038/ng.2938
   Wu G, 2012, NAT GENET, V44, P251, DOI 10.1038/ng.1102
NR 96
TC 7
Z9 7
U1 0
U2 5
PU AME PUBL CO
PI SHATIN
PA FLAT-RM C 16F, KINGS WING PLAZA 1, NO 3 KWAN ST, SHATIN, HONG KONG
   00000, PEOPLES R CHINA
SN 2305-5839
EI 2305-5847
J9 ANN TRANSL MED
JI ANN. TRANSL. MED.
PD JUN
PY 2018
VL 6
IS 12
AR 248
DI 10.21037/atm.2018.04.08
PG 12
WC Oncology; Medicine, Research & Experimental
WE Science Citation Index Expanded (SCI-EXPANDED)
SC Oncology; Research & Experimental Medicine
GA GL9CI
UT WOS:000437527200001
PM 30069450
OA hybrid, Green Published
DA 2023-08-27
ER

PT J
AU Kim, H
   Potrebko, P
   Rivera, A
   Liu, HS
   Eldredge-Hindy, HB
   Gunn, V
   Werner-Wasik, M
   Andrews, DW
   Evans, JJ
   Farrell, CJ
   Judy, K
   Shi, WY
AF Kim, Hyun
   Potrebko, Peter
   Rivera, Amanda
   Liu, Haisong
   Eldredge-Hindy, Harriet B.
   Gunn, Vickie
   Werner-Wasik, Maria
   Andrews, David W.
   Evans, James J.
   Farrell, Christopher J.
   Judy, Kevin
   Shi, Wenyin
TI Tumor volume threshold for achieving improved conformity in VMAT and
   Gamma Knife stereotactic radiosurgery for vestibular schwannoma
SO RADIOTHERAPY AND ONCOLOGY
LA English
DT Article
DE Radiosurgery; Vestibular schwannoma; Gamma knife; VMAT
ID ARC; RADIOTHERAPY; RADIATION; INDEX; TOOL
AB Background and purpose: Recent advances in multileaf collimator field shaping technology and inverse planning software have resulted in highly conformal LINAC based stereotactic radiosurgery (SRS) plans with minimal dose to critical structures. This modeling study compares Gamma Knife (GK) and LINAC SRS for vestibular schwannoma (VS).
   Materials and methods: 76 treatment plans from nineteen patients with VS were planned using GK forward planning and volumetric arc therapy (VMAT) inverse planning software. VMAT plans were generated with 1 coplanar, 3 and 5 non-coplanar arcs. Dose to normal structures and beam-on time (dose rate 600 MU/min) were compared using Kruskal-Wallis and Dunn's post hoc test.
   Results: Median tumor volume was 1.2 cm(3) (range 0.1-4.8 cm(3)). A peripheral tumor dose of 12 Gy was prescribed. Tumor coverage was >99.8%. VMAT plans had lower target D2% and mean dose, as well as decreased beam-on time, compared to GK plans (p < 0.0001). Paddick conformity index in VMAT 5 arc plans was superior to that of GK plans for targets >0.5 cm(3) (p = 0.002). Similar dose to cochlea, normal brain tissue and brainstem was observed.
   Conclusion: VMAT should be considered as a safe, alternative modality to GK for VS SRS treatment, especially for tumors larger than 0.5 cm(3). (C) 2015 Elsevier Ireland Ltd. All rights reserved.
C1 [Kim, Hyun; Potrebko, Peter; Rivera, Amanda; Eldredge-Hindy, Harriet B.; Werner-Wasik, Maria; Shi, Wenyin] Thomas Jefferson Univ, Sidney Kimmel Med Coll, Dept Radiat Oncol, Philadelphia, PA 19107 USA.
   [Liu, Haisong; Gunn, Vickie; Andrews, David W.; Evans, James J.; Farrell, Christopher J.; Judy, Kevin] Thomas Jefferson Univ, Sidney Kimmel Med Coll, Dept Neurol Surg, Philadelphia, PA 19107 USA.
C3 Jefferson University; Jefferson University
RP Shi, WY (通讯作者)，Bodine Ctr, Dept Radiat Oncol, Suite G-301,111 S 11th St, Philadelphia, PA 19107 USA.
EM
RI Rivera, Amanda/ISV-6332-2023; Shi, Wenyin/I-1321-2019
OI Shi, Wenyin/0000-0002-6336-3912; Eldredge-Hindy,
   Harriet/0000-0003-3582-968X
CR Abacioglu U, 2014, RADIAT ONCOL, V9, DOI 10.1186/1748-717X-9-119
   Andrews DW, 2004, LANCET, V363, P1665, DOI 10.1016/S0140-6736(04)16250-8
   Brown M, 2011, NEUROSURGERY, V69, P605, DOI 10.1227/NEU.0b013e31821a42f3
   Clark GM, 2010, INT J RADIAT ONCOL, V76, P296, DOI 10.1016/j.ijrobp.2009.05.029
   Deinsberger R, 2005, NEUROSURG REV, V28, P79, DOI 10.1007/s10143-005-0376-7
   Evans DGR, 2006, J MED GENET, V43, P289, DOI 10.1136/jmg.2005.036319
   Fogliata A, 2008, RADIOTHER ONCOL, V89, P254, DOI 10.1016/j.radonc.2008.07.021
   Gerbi Bruce J, 2004, J Appl Clin Med Phys, V5, P80, DOI 10.1120/jacmp.2021.25278
   Gevaert T, 2012, INT J RADIAT ONCOL, V82, P1627, DOI 10.1016/j.ijrobp.2011.01.052
   Griffiths A, 2007, INT J TECHNOL ASSESS, V23, P488, DOI 10.1017/S0266462307070584
   Hasegawa T, 2013, J NEUROSURG, V118, P557, DOI 10.3171/2012.10.JNS12523
   Lagerwaard FJ, 2009, INT J RADIAT ONCOL, V74, P610, DOI 10.1016/j.ijrobp.2008.12.076
   Lasak JM, 2008, OTOL NEUROTOL, V29, P1179, DOI 10.1097/MAO.0b013e31818b6639
   Lee TF, 2008, RADIOTHER ONCOL, V89, P89, DOI 10.1016/j.radonc.2008.05.010
   Loeffler JS, 2003, NEUROSURGERY, V52, P1436, DOI 10.1227/01.NEU.0000064809.59806.E8
   LOEFFLER JS, 2003, NEUROSURGERY, V52, P1440, DOI DOI 10.1227/01.NEU.0000064809.59806.E812762888
   Mayo CS, 2010, INT J RADIAT ONCOL, V78, P1457, DOI 10.1016/j.ijrobp.2009.10.005
   Paddick I, 2000, J NEUROSURG, V93, P219, DOI 10.3171/jns.2000.93.supplement_3.0219
   Paddick I, 2006, J NEUROSURG, V105, P194, DOI 10.3171/sup.2006.105.7.194
   Shiu AS, 1997, INT J RADIAT ONCOL, V37, P679, DOI 10.1016/S0360-3016(96)00507-X
   Wolff HA, 2010, RADIAT ONCOL, V5, DOI 10.1186/1748-717X-5-77
   Yang I, 2009, J CLIN NEUROSCI, V16, P742, DOI 10.1016/j.jocn.2008.09.023
NR 22
TC 9
Z9 9
U1 1
U2 2
PU ELSEVIER IRELAND LTD
PI CLARE
PA ELSEVIER HOUSE, BROOKVALE PLAZA, EAST PARK SHANNON, CO, CLARE, 00000,
   IRELAND
SN 0167-8140
J9 RADIOTHER ONCOL
JI Radiother. Oncol.
PD MAY
PY 2015
VL 115
IS 2
BP 229
EP 234
DI 10.1016/j.radonc.2015.03.031
PG 6
WC Oncology; Radiology, Nuclear Medicine & Medical Imaging
WE Science Citation Index Expanded (SCI-EXPANDED)
SC Oncology; Radiology, Nuclear Medicine & Medical Imaging
GA CM5YN
UT WOS:000357765000013
PM 25998805
DA 2023-08-27
ER

PT J
AU MARX, A
   SCHOMIG, D
   SCHULTZ, A
   GATTENLOHNER, S
   JUNG, A
   KIRCHNER, T
   MELMS, A
   MULLERHERMELINK, HK
AF MARX, A
   SCHOMIG, D
   SCHULTZ, A
   GATTENLOHNER, S
   JUNG, A
   KIRCHNER, T
   MELMS, A
   MULLERHERMELINK, HK
TI DISTRIBUTION OF MOLECULES MEDIATING THYMOCYTE-STROMA-INTERACTIONS IN
   HUMAN THYMUS, THYMITIS AND THYMIC EPITHELIAL TUMORS
SO THYMUS
LA English
DT Article
DE ADHESION; AUTOIMMUNITY; MYASTHENIA GRAVIS; T-CELLS; THYMOMA; THYMUS
ID ACETYLCHOLINE-RECEPTOR EPITOPE; CD4+ LYMPHOCYTES-T; MYASTHENIA-GRAVIS;
   IMMATURE THYMOCYTES; ALPHA-SUBUNIT; CELLS; ADHESION; THYMOMA; B7;
   ACTIVATION
AB Two findings in thymic epithelial tumors are correlated with the occurrence of myasthenia gravis(MG): (I) the expression of an acetylcholine receptor (AChR)-like-epitope in the neoplastic epithelium, and (2) the preservation of thymus-like features in the neoplasms, indicated by the presence of immature thymocytes. On this background it has been proposed that paraneoplastic MG may start with an intratumorous abnormal T cell selection due to aberrantly expressed AChR-epitopes (self-peptides). As appropriate thymocyte-stroma-interactions are prerequisites for thymocyte development in the thymus (and probably in MG-associated thymic tumors, too), we analyzed the expression of CD28/B7(BB1), CD2/LFA3, LFA-1/ICAM-1 and VLA-4/VCAM-1 in human thymus and thymomas by immunohistochemistry. In normal thymuses and thymitis the stromal molecules were expressed at higher levels in the medulla than in the cortex. This was particulrly true for B7(BB1) that was undetectable by immunoperoxidase techniques in the cortex. In contrast, cortical-type thymic epithelial tumors (cortical thymoma and well differentited thymic carcinoma), known to exhibit the highest association with myasthenia, expressed the stromal molecules at almost medullary levels. The findings may be a clue to a functional difference between neoplastic and normal cortical epithelial cells: while we find the former to have the capacity to present soluble antigen to antigen-specific CD4(+) T cells in vitro, normal cortical epithelium failed to do so. This altered microenvironment in thymomas might contribute to autoimmunization by stimulating mature recirculating AChR-specific T cells.
C1 UNIV ERLANGEN NURNBERG,INST PATHOL,W-8520 ERLANGEN,GERMANY.
   UNIV TUBINGEN,DEPT NEUROL,W-7400 TUBINGEN,GERMANY.
C3 University of Erlangen Nuremberg; Eberhard Karls University of Tubingen
RP MARX, A (通讯作者)，UNIV WURZBURG,INST PATHOL,SCHNEIDER STR 2,D-97080 WURZBURG,GERMANY.
CR BAGGI F, 1993, J NEUROIMMUNOL, V46, P57, DOI 10.1016/0165-5728(93)90233-O
   BOYD RL, 1993, IMMUNOL TODAY, V14, P445, DOI 10.1016/0167-5699(93)90248-J
   CARLOW DA, 1992, J IMMUNOL, V148, P1595
   CLARK EA, 1994, NATURE, V367, P425, DOI 10.1038/367425a0
   DALLOUL AH, 1991, BLOOD, V77, P69
   DAMLE NK, 1991, P NATL ACAD SCI USA, V88, P6403, DOI 10.1073/pnas.88.15.6403
   DAMLE NK, 1992, J IMMUNOL, V148, P1985
   DAMLE NK, 1993, EUR J IMMUNOL, V23, P1513, DOI 10.1002/eji.1830230718
   FINE JS, 1992, J IMMUNOL, V147, P2852
   FUJII Y, 1992, J NEUROL, V239, P82, DOI 10.1007/BF00862978
   HARDING FA, 1992, NATURE, V356, P607, DOI 10.1038/356607a0
   Haynes B F, 1990, Semin Immunol, V2, P67
   JENKINS MK, 1993, CURR OPIN IMMUNOL, V5, P361, DOI 10.1016/0952-7915(93)90054-V
   JONES L, 1992, INT IMMUNOL, V5, P503
   KILLEEN N, 1992, EMBO J, V11, P4329, DOI 10.1002/j.1460-2075.1992.tb05532.x
   KIRCHNER T, 1988, AM J PATHOL, V130, P268
   KIRCHNER T, 1992, AM J SURG PATHOL, V16, P153
   Kirchner T., 1989, PROG SURG PATHOL, V10, P167
   LINSLEY PS, 1991, J EXP MED, V173, P721, DOI 10.1084/jem.173.3.721
   MARX A, 1991, LYMPHATIC TISSUES AND IN VIVO IMMUNE RESPONSES, P577
   MARX A, 1989, AM J PATHOL, V134, P865
   MARX A, 1990, LAB INVEST, V62, P279
   MELMS A, 1992, ANN NEUROL, V31, P311, DOI 10.1002/ana.410310314
   MELMS A, 1989, J CLIN INVEST, V83, P785, DOI 10.1172/JCI113958
   MILLER GT, 1993, J EXP MED, V178, P211, DOI 10.1084/jem.178.1.211
   MIZUOCHI T, 1992, J EXP MED, V175, P1601, DOI 10.1084/jem.175.6.1601
   MULLERHERMELINK HK, 1993, ANN NY ACAD SCI, V681, P56, DOI 10.1111/j.1749-6632.1993.tb22869.x
   MULLERHERMELINK HK, 1994, RECENT ADV HISTOPATH, V16, P9
   NICKOLOFF BJ, 1993, AM J PATHOL, V142, P1029
   PAGE DM, 1993, J IMMUNOL, V151, P1868
   PUNT JA, 1994, J EXP MED, V179, P709, DOI 10.1084/jem.179.2.709
   RITTER MA, 1993, IMMUNOL TODAY, V14, P462, DOI 10.1016/0167-5699(93)90250-O
   SHAHINIAN A, 1993, SCIENCE, V261, P609, DOI 10.1126/science.7688139
   SIMPSON JA, 1960, SCOT MED J, V5, P419, DOI DOI 10.1177/003693306000501001
   SOMMER N, 1990, ANN NEUROL, V28, P312, DOI 10.1002/ana.410280303
   TAN RS, 1992, J IMMUNOL, V149, P3217
   TZARTOS SJ, 1992, EUR J BIOCHEM, V207, P915, DOI 10.1111/j.1432-1033.1992.tb17124.x
   UTSUMI K, 1991, P NATL ACAD SCI USA, V88, P5685, DOI 10.1073/pnas.88.13.5685
   VANGOOL SW, 1994, J EXP MED, V179, P715, DOI 10.1084/jem.179.2.715
   VANSEVENTER GA, 1991, J EXP MED, V174, P901, DOI 10.1084/jem.174.4.901
   WILLCOX N, 1987, AM J PATHOL, V127, P447
   YOKOCHI T, 1982, J IMMUNOL, V128, P823
NR 42
TC 17
Z9 17
U1 0
U2 0
PU KLUWER ACADEMIC PUBL
PI DORDRECHT
PA SPUIBOULEVARD 50, PO BOX 17, 3300 AA DORDRECHT, NETHERLANDS
SN 0165-6090
J9 THYMUS
JI Thymus
PY 1994
VL 23
IS 2
BP 83
EP 93
PG 11
WC Immunology
WE Science Citation Index Expanded (SCI-EXPANDED)
SC Immunology
GA QF101
UT WOS:A1994QF10100002
PM 7725387
DA 2023-08-27
ER

PT J
AU DEHAENE, I
   CASSELMAN, J
AF DEHAENE, I
   CASSELMAN, J
TI LEFT SUPERIOR OBLIQUE MYOKYMIA AND RIGHT SUPERIOR OBLIQUE PARALYSIS DUE
   TO A POSTERIOR-FOSSA TUMOR
SO NEURO-OPHTHALMOLOGY
LA English
DT Article
DE TROCHLEAR NERVE PALSY; SUPERIOR OBLIQUE MYOKYMIA
AB A case of right superior oblique paralysis and left superior oblique myokymia is presented, caused by a pilocytic astrocytoma compressing the midbrain tectum.
C1 ALGEMEEN ZIEKENHUIS ST JAN,DEPT MED IMAGING,B-8000 BRUGGE,BELGIUM.
RP DEHAENE, I (通讯作者)，ALGEMEEN ZIEKENHUIS ST JAN,DEPT NEUROL,B-8000 BRUGGE,BELGIUM.
CR MORROW MJ, 1990, NEUROLOGY, V40, P367, DOI 10.1212/WNL.40.2.367
   NEETENS A, 1983, NEURO-OPHTHALMOLOGY, V3, P103, DOI 10.3109/01658108309009725
   THURSTON SE, 1991, NEUROLOGY, V41, P1679, DOI 10.1212/WNL.41.10.1679
NR 3
TC 16
Z9 16
U1 0
U2 0
PU AEOLUS PRESS
PI BUREN
PA PO BOX 740, 4116 ZJ BUREN, NETHERLANDS
SN 0165-8107
J9 NEURO-OPHTHALMOLOGY
JI Neuro-Ophthalmol.
PD FEB
PY 1993
VL 13
IS 1
BP 13
EP 16
DI 10.3109/01658109309036997
PG 4
WC Clinical Neurology; Ophthalmology
WE Science Citation Index Expanded (SCI-EXPANDED)
SC Neurosciences & Neurology; Ophthalmology
GA KT019
UT WOS:A1993KT01900003
DA 2023-08-27
ER

PT J
AU Ahmed, M
   Schmidt, M
   Sohaib, A
   Kong, C
   Burke, K
   Richardson, C
   Usher, M
   Brennan, S
   Riddell, A
   Davies, M
   Newbold, K
   Harrington, KJ
   Nutting, CM
AF Ahmed, Merina
   Schmidt, Maria
   Sohaib, Aslam
   Kong, Christine
   Burke, Kevin
   Richardson, Cheryl
   Usher, Marianne
   Brennan, Sinead
   Riddell, Angela
   Davies, Mark
   Newbold, Kate
   Harrington, Kevin J.
   Nutting, Christopher M.
TI The value of magnetic resonance imaging in target volume delineation of
   base of tongue tumours - A study using flexible surface coils
SO RADIOTHERAPY AND ONCOLOGY
LA English
DT Article
DE MRI; Volume delineation; Oropharynx
ID INTENSITY-MODULATED RADIOTHERAPY; COMPLETE DISTORTION CORRECTION;
   SQUAMOUS-CELL CARCINOMA; DOSE DISTRIBUTION; MR-IMAGES; CT; HEAD;
   LARYNGEAL; IMPACT; CANCER
AB Introduction: Magnetic resonance imaging (MRI) provides superior diagnostic accuracy over computed tomography (CC) in oropharyngeal tumours. Precise delineation of the gross tumour volume (GTV) is mandatory in radiotherapy planning when a GTV boost is required. CT volume definition in this regard is poor. We studied the feasibility of using flexible surface (flex-L) coils to obtain MR images for MR-CT fusion to assess the benefit of MRI over CT alone in planning base of tongue tumours.
   Methods: Eight patients underwent CT and MRI radiotherapy planning scans with an immobilisation device. Distortion-corrected T1-weighted post-contrast MR scans were fused to contrast-enhanced planning CT scans. GTV, clinical target and planning target volumes (CTV, PTV) and organs at risk (OAR) were delineated on CT, then on MRI with blinding to the CT images. The volumetric and spatial differences between MRI and CT volumes for GTV, CTV, PTV and OAR were compared. MR image distortions due to field inhomogeneity and non-linear gradients were corrected and the need for such correction was evaluated.
   Results: The mean primary GTV was larger on MRI (22.2 vs. 9.5 cm(3), p = 0.05) than CT. The mean primary and nodal GTV (i.e. BUT and macroscopic nodes) was significantly larger on MRI (27.2 vs. 14.4 cm(3), p = 0.05). The volume overlap index (VOI) between MRI and CT for the primary was 0.34 suggesting that MRI depicts parts of the primary tumour not detected by CT. There was no significant difference in volume delineation between MR and CT for CTV, PTV, nodal CTV and nodal PTV. MRI volumes for brainstem and spinal cord were significantly smaller due to improved organ definition (p = 0.002). Susceptibility and gradient-related distortions were not found to be clinically significant.
   Conclusion: MRI improves the definition of tongue base tumours and neurological structures. The use of MRI is recommended for GTV dose-escalation techniques to provide precise depiction of GTV and improved sparing of spinal cord and brainstem. (C) 2009 Elsevier Ireland Ltd. All rights reserved. Radiotherapy and Oncology 94 (2010) 161-167
C1 [Ahmed, Merina; Kong, Christine; Burke, Kevin; Davies, Mark; Newbold, Kate; Harrington, Kevin J.; Nutting, Christopher M.] Royal Marsden NHS Fdn Trust, Dept Radiotherapy, London SW3 6JJ, England.
   [Schmidt, Maria; Richardson, Cheryl; Usher, Marianne] Royal Marsden NHS Fdn Trust, Canc Res UK Clin Magnet Resonance Grp, Surrey, England.
   [Sohaib, Aslam; Riddell, Angela] Royal Marsden NHS Fdn Trust, Dept Radiol, London, England.
   [Brennan, Sinead] St James Hosp, Dept Radio2therapy, Dublin, Ireland.
   [Harrington, Kevin J.; Nutting, Christopher M.] Inst Canc Res, London SW3 6JB, England.
C3 Royal Marsden NHS Foundation Trust; Royal Marsden NHS Foundation Trust;
   Royal Marsden NHS Foundation Trust; Trinity College Dublin; Royal
   Marsden NHS Foundation Trust; University of London; Institute of Cancer
   Research - UK
RP Ahmed, M (通讯作者)，Royal Marsden NHS Fdn Trust, Dept Radiotherapy, Fulham Rd, London SW3 6JJ, England.
EM
RI Newbold, Kate/AAA-9176-2022
OI Brennan, sinead/0000-0001-8427-6554; Schmidt, Maria/0000-0001-8993-5301;
   Harrington, Kevin/0000-0002-6014-348X
CR BATTISTA JJ, 1980, INT J RADIAT ONCOL, V6, P99, DOI 10.1016/0360-3016(80)90211-4
   Bhide S, 2007, RADIOTHER ONCOL, V82, pS74, DOI 10.1016/S0167-8140(07)80203-9
   Chung NN, 2004, HEAD NECK-J SCI SPEC, V26, P241, DOI 10.1002/hed.10378
   de Bondt BJ, 2009, HEAD NECK-J SCI SPEC, V31, P928, DOI 10.1002/hed.21060
   Doran SJ, 2005, PHYS MED BIOL, V50, P1343, DOI 10.1088/0031-9155/50/7/001
   Eisbruch A, 1998, INT J RADIAT ONCOL, V41, P559, DOI 10.1016/S0360-3016(98)00082-0
   Geets X, 2005, RADIOTHER ONCOL, V77, P25, DOI 10.1016/j.radonc.2005.04.010
   Geets X, 2006, RADIOTHER ONCOL, V78, P291, DOI 10.1016/j.radonc.2006.01.006
   Geets X, 2007, RADIOTHER ONCOL, V85, P105, DOI 10.1016/j.radonc.2007.05.010
   Heesters M. A. A. M., 1993, Strahlentherapie und Onkologie, V169, P729
   HILL DLG, 1994, RADIOLOGY, V191, P447, DOI 10.1148/radiology.191.2.8153319
   Humphreys M, 2005, RADIOTHER ONCOL, V77, P39, DOI 10.1016/j.radonc.2005.06.039
   Kinoshita T, 2004, CLIN IMAG, V28, P93, DOI 10.1016/S0899-7071(03)00120-7
   Lam P, 2004, AM J ROENTGENOL, V182, P803, DOI 10.2214/ajr.182.3.1820803
   Lenz M, 2000, EUR J RADIOL, V33, P203, DOI 10.1016/S0720-048X(99)00143-6
   NUTTING CM, 2009, J CLIN ONCOL, V17, pS18
   Oldham M, 2001, MED PHYS, V28, P1436, DOI 10.1118/1.1380430
   Reinsberg SA, 2005, PHYS MED BIOL, V50, P2651, DOI 10.1088/0031-9155/50/11/014
   *ROYAL COLL RAD, 2006, RCR061
   Schenck JF, 1996, MED PHYS, V23, P815, DOI 10.1118/1.597854
   Sigal R, 1996, RADIOGRAPHICS, V16, P787, DOI 10.1148/radiographics.16.4.8835972
   Urbano MTG, 2007, CLIN ONCOL-UK, V19, P604, DOI 10.1016/j.clon.2007.07.001
   VANHERK M, 1994, MED PHYS, V21, P1163, DOI 10.1118/1.597344
   Zou KH, 2004, ACAD RADIOL, V11, P178, DOI 10.1016/S1076-6332(03)00671-8
NR 24
TC 51
Z9 55
U1 0
U2 8
PU ELSEVIER IRELAND LTD
PI CLARE
PA ELSEVIER HOUSE, BROOKVALE PLAZA, EAST PARK SHANNON, CO, CLARE, 00000,
   IRELAND
SN 0167-8140
J9 RADIOTHER ONCOL
JI Radiother. Oncol.
PD FEB
PY 2010
VL 94
IS 2
BP 161
EP 167
DI 10.1016/j.radonc.2009.12.021
PG 7
WC Oncology; Radiology, Nuclear Medicine & Medical Imaging
WE Science Citation Index Expanded (SCI-EXPANDED)
SC Oncology; Radiology, Nuclear Medicine & Medical Imaging
GA 578BA
UT WOS:000276267100006
PM 20096947
DA 2023-08-27
ER

PT J
AU McCormick, WE
   Lee, JH
AF McCormick, WE
   Lee, JH
TI Pseudobulbar palsy caused by a large petroclival meningioma: Report of
   two cases
SO SKULL BASE-AN INTERDISCIPLINARY APPROACH
LA English
DT Article
DE meningioma; pathological laughter; pseudobulbar palsy; tumor
ID PATHOLOGICAL LAUGHTER
AB Two patients sought treatment for symptoms of bulbar motor dysfunction and the marked emotional lability that characterizes pseudobulbar palsy (PBP). Magnetic resonance (MR) imaging showed large petroclival masses with severe compression and displacement of the brainstem. A suboccipital craniectomy with a transpetrosal, transtentorial approach to the tumor was performed in both patients. One patient required a second-stage surgery to resect an extension of the tumor into the cavernous sinus. In both patients pathological evaluation confirmed the diagnosis of syncytial meningioma. After surgery, PBP resolved in both patients. Large posterior fossa tumors should be included in the differential diagnosis of patients with a clinical picture of bulbar motor dysfunction coupled with emotional lability.
C1 Cleveland Clin Fdn, Dept Neurosurg S80, Sect Skull Base Surg, Cleveland, OH 44195 USA.
C3 Cleveland Clinic Foundation
RP Lee, JH (通讯作者)，Cleveland Clin Fdn, Dept Neurosurg S80, Sect Skull Base Surg, 9500 Euclid Ave, Cleveland, OH 44195 USA.
CR ACHARI AN, 1976, JAMA-J AM MED ASSOC, V235, P1469
   Adams RD, 1997, PRINCIPLES NEUROLOGY, P508
   Bhatjiwale MG, 2000, NEUROSURGERY, V47, P469, DOI 10.1097/00006123-200008000-00044
   CAIRNS H, 1950, Folia Psychiatr Neurol Neurochir Neerl, V53, P193
   CANTU RC, 1966, J NEUROSURG, V24, P1024, DOI 10.3171/jns.1966.24.6.1024
   Hattori S, 1998, PATHOL RES PRACT, V194, P363, DOI 10.1016/S0344-0338(98)80061-7
   IRONSIDE R, 1956, BRAIN, V79, P589, DOI 10.1093/brain/79.4.589
   LEPINE R, 1877, REV MENSUELLE MED CH, V1, P909
   MAGNUS A, 1837, MULLER ARCH, V258, P256
   MATSUOKA S, 1993, J NEUROSURG, V79, P428, DOI 10.3171/jns.1993.79.3.0428
   Matsuoka S, 1977, Neurol Med Chir (Tokyo), V17, P195, DOI 10.2176/nmc.17pt2.195
   MITSUMOTO H, 1996, NEUROLOGY CLIN PRACT, V1, P261
   Nadkarni TD, 1999, BRIT J NEUROSURG, V13, P212, DOI 10.1080/02688699944023
   Osumi Y, 1976, Rinsho Shinkeigaku, V16, P715
   POECK K, 1985, HDB CLIN NEUROLOGY, V45
   Shafqat S, 1998, NEUROLOGY, V50, P1918, DOI 10.1212/WNL.50.6.1918
   STERN WE, 1957, J NEUROSURG, V14, P129
   STEVENSON GC, 1966, J NEUROSURG, V24, P544, DOI 10.3171/jns.1966.24.2.0544
   TSUZAKA K, 1993, CLIN NEUROL, V33, P194
   Wisoff J H, 1984, Neurosurgery, V15, P707
NR 20
TC 7
Z9 7
U1 0
U2 1
PU THIEME MEDICAL PUBL INC
PI NEW YORK
PA 333 SEVENTH AVE, NEW YORK, NY 10001 USA
SN 1531-5010
J9 SKULL BASE-INTERD AP
JI Skull Base-Interdiscip. Appr.
PD MAY
PY 2002
VL 12
IS 2
BP 67
EP 71
AR UNSP 1531-5010,p;2002,12,02,067,072,ftx,en;sbs00266x
DI 10.1055/s-2002-31568-1
PG 5
WC Clinical Neurology; Otorhinolaryngology; Surgery
WE Science Citation Index Expanded (SCI-EXPANDED)
SC Neurosciences & Neurology; Otorhinolaryngology; Surgery
GA 555BZ
UT WOS:000175774400003
PM 17167648
DA 2023-08-27
ER

PT J
AU Folling, I
   Wennerstrom, AB
   Eide, TJ
   Nilsen, HL
AF Folling, Ivar
   Wennerstrom, Anna B.
   Eide, Tor J.
   Nilsen, Hilde Loge
TI Phaeochromocytomas overexpress insulin transcript and produce insulin
SO ENDOCRINE CONNECTIONS
LA English
DT Article
DE phaeochromocytoma; insulin; INS-IGF2; ectopic production; ectopic
   transcription; tumour-induced hypoglycaemia; tumour-induced
   hyperglycaemia; immunohistochemistry
ID GROWTH-FACTOR-I; RECEPTOR EXPRESSION; HYPOGLYCEMIA; CELL; TUMORS; OVARY
AB Introduction: Phaeochromocytomas are tumours originating in the medulla of the adrenal gland. They produce catecholamines, and some tumours also produce ectopic hormones. Two types of glucose imbalances occur in phaeochromocytoma patients, hyperglycaemia and hypoglycaemic attacks. Therefore, we tested whether insulin transcript (INS), insulin, and a hybrid read-through transcript between exons from insulin and insulin-like growth factor 2 (INS-IGF2) were expressed in phaeochromocytomas.
   Methods: We measured the expression of insulin using immunohistochemistry. The expression of INS-IGF2 was determined by qRT-PCR in formalin-fixed and paraffin-embedded tissue from 20 phaeochromocytomas. The expression of INS and INS-IGF2 transcripts was also analysed in 182 phaeochromocytomas and paragangliomas using publicly available datasets in The Cancer Genome Atlas (TCGA) Database.
   Results: Of 20 phaeochromocytomas, 16 stained positive for insulin. The distribution of positive cells was mostly scattered, with some focal expression indicating clonal expansion. Nineteen tumours expressed high levels of INS and INS-IGF2 transcripts. The expression of the two transcripts corresponded closely. In the TCGA dataset, phaeochromocytoma expresses higher levels of INS and INS-IGF2 transcripts compared to the normal non-tumour adrenal glands. Thus, the expression of INS and INS-IGF2 seems to be a general phenomenon in phaeochromocytoma.
   Conclusion: Most phaeochromocytomas contain cells that overexpress INS and INS-IGF2 transcripts. Most tumours also display heterogeneous expression of polypeptides immunoreactive to monoclonal anti-insulin antibodies. Clinically this may relate to both hyperglycaemia and hypoglycaemic attacks seen in patients with phaeochromocytoma as well as autocrine tumour growth.
C1 [Folling, Ivar] Akershus Univ Hosp, Dept Endocrinol, Lorenskog, Norway.
   [Folling, Ivar; Nilsen, Hilde Loge] Univ Oslo, Inst Clin Med, Oslo, Norway.
   [Folling, Ivar; Wennerstrom, Anna B.; Nilsen, Hilde Loge] Univ Oslo, Dept Clin Mol Biol, Lorenskog, Norway.
   [Folling, Ivar; Wennerstrom, Anna B.; Nilsen, Hilde Loge] Akershus Univ Hosp, Lorenskog, Norway.
   [Eide, Tor J.] Oslo Univ Hosp, Dept Pathol, Div Lab Med, Oslo, Norway.
C3 University of Oslo; University of Oslo; University of Oslo; University
   of Oslo; University of Oslo
RP Folling, I (通讯作者)，Akershus Univ Hosp, Dept Endocrinol, Lorenskog, Norway.; Folling, I (通讯作者)，Univ Oslo, Inst Clin Med, Oslo, Norway.; Folling, I (通讯作者)，Univ Oslo, Dept Clin Mol Biol, Lorenskog, Norway.; Folling, I (通讯作者)，Akershus Univ Hosp, Lorenskog, Norway.
EM
OI Wennerstrom, Anna Berit/0000-0001-8143-9944
FU South-Eastern Regional Infrastructure for Clinical and Translational
   Research (SERIT) [276984]
FX The study was supported by South-Eastern Regional Infrastructure for
   Clinical and Translational Research (SERIT) grant number 276984.
CR Abdulhadi Basma, 2021, AACE Clin Case Rep, V7, P80, DOI 10.1016/j.aace.2020.11.002
   ASHTON MA, 1995, HISTOPATHOLOGY, V27, P463, DOI 10.1111/j.1365-2559.1995.tb00311.x
   Aunis D, 1998, INT REV CYTOL, V181, P213, DOI 10.1016/S0074-7696(08)60419-2
   Battocchio M, 2015, ENDOCRINE, V49, P611, DOI 10.1007/s12020-015-0605-y
   Bedinger DH, 2015, MOL CELL ENDOCRINOL, V415, P143, DOI 10.1016/j.mce.2015.08.013
   Bramswig NC, 2012, CELL MOL LIFE SCI, V69, P2109, DOI 10.1007/s00018-011-0915-z
   Castro-Vega LJ, 2016, ONCOGENE, V35, P1080, DOI 10.1038/onc.2015.172
   Fernandez MC, 2013, CLIN ENDOCRINOL, V79, P623, DOI 10.1111/cen.12205
   Crona J, 2015, CLIN CANCER RES, V21, P4451, DOI 10.1158/1078-0432.CCR-14-2854
   Dekkers T, 2014, J CLIN ENDOCR METAB, V99, pE1341, DOI 10.1210/jc.2013-4255
   Dimitriadis GK, 2017, ENDOCR-RELAT CANCER, V24, pR173, DOI 10.1530/ERC-17-0036
   Erlic Z, 2019, EXP CLIN ENDOCR DIAB, V127, P129, DOI 10.1055/a-0649-0960
   Folling I, 2015, END ABSTR, V37, DOI [10.1530/endoabs.37.EP1156, DOI 10.1530/ENDOABS.37.EP1156]
   Fottner C, 2006, J MOL ENDOCRINOL, V36, P279, DOI 10.1677/jme.1.01975
   Frankton S, 2009, HORM-INT J ENDOCRINO, V8, P65, DOI 10.14310/horm.2002.1224
   Gao SL, 2019, TECHNOL CANCER RES T, V18, DOI 10.1177/1533033818823029
   GELATO MC, 1990, J CLIN ENDOCR METAB, V71, P1168, DOI 10.1210/jcem-71-5-1168
   Gill AJ, 2018, HISTOPATHOLOGY, V72, P106, DOI 10.1111/his.13277
   Ilias I, 2005, J CLIN ENDOCR METAB, V90, P4955, DOI 10.1210/jc.2004-2527
   Johannessen LE, 2016, ONCOL REP, V36, P2653, DOI 10.3892/or.2016.5132
   Kaltsas G, 2010, ENDOCR-RELAT CANCER, V17, pR173, DOI 10.1677/ERC-10-0024
   KAMIO T, 1991, AM J PATHOL, V138, P83
   Kanatsuna N, 2013, J BIOL CHEM, V288, P29013, DOI 10.1074/jbc.M113.478222
   Korner M, 2007, J NUCL MED, V48, P736, DOI 10.2967/jnumed.106.038679
   Ku HT, 2007, DIABETES, V56, P921, DOI 10.2337/db06-0468
   Kuroda A, 2009, PLOS ONE, V4, DOI 10.1371/journal.pone.0006953
   LEACH SD, 1990, WORLD J SURG, V14, P335, DOI 10.1007/BF01658520
   Liu YS, 2016, CELL, V165, P535, DOI 10.1016/j.cell.2016.03.014
   Minn AH, 2004, LANCET, V363, P363, DOI 10.1016/S0140-6736(04)15438-X
   Monk D, 2006, HUM MOL GENET, V15, P1259, DOI 10.1093/hmg/ddl041
   MORGELLO S, 1988, CANCER, V61, P800, DOI 10.1002/1097-0142(19880215)61:4<800::AID-CNCR2820610426>3.0.CO;2-3
   Morken NH, 2007, ACTA OBSTET GYN SCAN, V86, P500, DOI 10.1080/00016340600613477
   Nielsen HM, 2015, ENDOCR-RELAT CANCER, V22, P953, DOI 10.1530/ERC-15-0086
   Ogilvie RF, 1937, Q J MED, V6, P287
   Ramkumar S, 2014, BMC ENDOCR DISORD, V14, DOI 10.1186/1472-6823-14-36
   Saber-Ayad M, 2018, ESMO OPEN, V3, pA199, DOI [10.1136/esmoopen-2018-EACR25.474, DOI 10.1136/ESMOOPEN-2018-EACR25.474]
   Saisho Y, 2013, DIABETES CARE, V36, P111, DOI 10.2337/dc12-0421
   Seckl MJ, 1999, NEW ENGL J MED, V341, P733, DOI 10.1056/NEJM199909023411004
   SHAMES JM, 1968, AM J MED, V44, P632, DOI 10.1016/0002-9343(68)90065-X
   STAGNO PA, 1987, ARCH PATHOL LAB MED, V111, P440
   Tang ZF, 2017, NUCLEIC ACIDS RES, V45, pW98, DOI 10.1093/nar/gkx247
   Thonangi Rajesh Patnaik, 2014, Indian J Endocrinol Metab, V18, P234, DOI 10.4103/2230-8210.129120
   Thorsoe KS, 2010, BIOCHEMISTRY-US, V49, P6234, DOI 10.1021/bi1000118
   Uysal M, 2007, HORM RES, V67, P292, DOI 10.1159/000099291
NR 44
TC 0
Z9 0
U1 0
U2 2
PU BIOSCIENTIFICA LTD
PI BRISTOL
PA STARLING HOUSE, 1600 BRISTOL PARKWAY N, BRISTOL, ENGLAND
SN 2049-3614
J9 ENDOCR CONNECT
JI Endocr. Connect.
PD AUG
PY 2021
VL 10
IS 8
BP 815
EP 824
DI 10.1530/EC-21-0269
PG 10
WC Endocrinology & Metabolism
WE Science Citation Index Expanded (SCI-EXPANDED)
SC Endocrinology & Metabolism
GA TQ0BX
UT WOS:000677956500001
PM 34170845
OA Green Published, gold
DA 2023-08-27
ER

PT J
AU Ladzinski, P
   Majchrzak, H
   Maliszewski, M
   Majchrzak, K
   Kaspera, W
   Tymowski, M
   Adamczyk, P
AF Ladzinski, Piotr
   Majchrzak, Henryk
   Maliszewski, Mariusz
   Majchrzak, Krzysztof
   Kaspera, Wojciech
   Tymowski, Michal
   Adamczyk, Piotr
TI Direct and remote outcome after treatment of tumours involving the
   cavernous sinus and its surroundings
SO NEUROLOGIA I NEUROCHIRURGIA POLSKA
LA English
DT Article
DE cavernous sinus; internal carotid artery; microsurgery; meningioma;
   hypophyseal adenoma; neurinoma
ID OF-THE-LITERATURE; PITUITARY-ADENOMAS; STEREOTACTIC RADIOSURGERY;
   SURGICAL-TREATMENT; MALIGNANT-LYMPHOMA; OCULOMOTOR NERVE; TROCHLEAR
   NERVE; CRANIAL NERVES; CAROTID-ARTERY; MENINGIOMAS
AB Background and purpose: The aim of this study was to present results obtained by the authors after surgical treatment of tumours involving the cavernous sinus (CS) and its surroundings.
   Material and methods: Thirty-eight patients (23 females and 15 males) with tumours of the CS and its surroundings were included in the study The neurological condition of patients and their ability to perform activities of daily living were evaluated prior to treatment, postoperatively and after completion of therapy The following parameters were measured: approximate volume of the operated lesions; their relation to larger vessels, cranial nerves and the brainstem; consistency and vascularization.
   Results: The medical history of the disease ranged from 1.5 to 48 months, mean 12 months. In 89.5% of cases impaired Ocular motility on the side of the lesion was the symptom indicative of the condition. The second most common group of symptoms (29%) included impairments of trigeminal nerve function. The approximate volume of all resected tumours ranged from 0.6 to 60 mL (mean 12.2 mL). In 50% of cases the resected tumours were diagnosed as meningiomas. The remaining entities included: perithelioma, hypophyseal adenoma, adenoid cystic carcinoma, neuroendocrine carcinoma, trigeminal neurinoma, chordoma, cavernous angioma and lymphoma. In 63% resections were complete. Functional capacity of patients improved in 34% of cases, remained unchanged in 53%, deteriorated in 10.5%, and one patient died.
   Conclusions: Surgical intervention involving the CS and its surroundings enables resection of neoplastic tumours of this anatomical region with satisfactory clinical results and low mortality. Most patients suffer from temporary paresis or paralysis of the oculomotor, trochlear and abducent nerves. Reduction of completeness of resections of CS tumours decreases the number of postoperative neurological deficits without a significant increase in the rate of lesion regrowth.
C1 [Ladzinski, Piotr; Majchrzak, Henryk; Maliszewski, Mariusz; Majchrzak, Krzysztof; Kaspera, Wojciech; Tymowski, Michal; Adamczyk, Piotr] Silesian Med Univ Katowice, Dept Neurosurg, St Barbara Reg, Multidisciplinary Hosp 5, PL-41200 Sosnowiec, Poland.
C3 Medical University Silesia
RP Ladzinski, P (通讯作者)，Silesian Med Univ Katowice, Dept Neurosurg, St Barbara Reg, Multidisciplinary Hosp 5, Pl Medyko, PL-41200 Sosnowiec, Poland.
EM
RI Kaspera, Wojciech/AAF-2715-2019
OI Kaspera, Wojciech/0000-0002-7139-3409; Ladzinski,
   Piotr/0000-0001-8957-0162
CR AHMADI J, 1986, AM J ROENTGENOL, V146, P257, DOI 10.2214/ajr.146.2.257
   al-Mefty O, 1991, Acta Neurochir Suppl (Wien), V53, P117
   ALMEFTY O, 1988, SURG NEUROL, V30, P370, DOI 10.1016/0090-3019(88)90200-5
   ANAND VK, 1991, LARYNGOSCOPE, V101, P557
   [Anonymous], CAVERNOUS SINUS COMP
   Aziz KMA, 1999, NEUROSURGERY, V44, P859, DOI 10.1097/00006123-199904000-00096
   Bristot R, 1997, SURG NEUROL, V48, P160, DOI 10.1016/S0090-3019(97)00033-5
   Chen JCT, 2001, NEUROSURGERY, V48, P1022, DOI 10.1097/00006123-200105000-00011
   DeJesus O, 1996, NEUROSURGERY, V39, P915, DOI 10.1097/00006123-199611000-00005
   DELPASSAND ES, 1988, NEUROSURGERY, V23, P501, DOI 10.1227/00006123-198810000-00018
   DEMONTE F, 1994, J NEUROSURG, V81, P245, DOI 10.3171/jns.1994.81.2.0245
   DEMONTE F, 2000, CAVERNOUS SINUS COMP, P329
   DEVRIES EJ, 1988, LARYNGOSCOPE, V98, P960
   Dolenc VV, 1997, NEUROSURGERY, V41, P542, DOI 10.1097/00006123-199709000-00007
   Dolenc VV, 1997, OPERATIVE SKULL BASE, P207
   DUMA CM, 1993, NEUROSURGERY, V32, P699, DOI 10.1227/00006123-199305000-00001
   EISENBERG M, 2000, CAVERNOUS SINUS, P307
   Eisenberg MB, 2000, CAVERNOUS SINUS
   ELKALLINY M, 1992, J NEUROSURG, V77, P508, DOI 10.3171/jns.1992.77.4.0508
   GRIMSON BS, 1984, J NEUROSURG, V61, P191, DOI 10.3171/jns.1984.61.1.0191
   HIRSCH WL, 1993, AM J ROENTGENOL, V160, P1083, DOI 10.2214/ajr.160.5.8470581
   INOUE T, 1990, NEUROSURGERY, V27, P986, DOI 10.1227/00006123-199012000-00021
   KLEINPETER G, 1990, ACTA NEUROCHIR, V103, P87, DOI 10.1007/BF01407511
   Klink DF, 2000, NEUROSURGERY, V47, P24, DOI 10.1097/00006123-200007000-00005
   KNOSP E, 1993, NEUROSURGERY, V33, P610, DOI 10.1227/00006123-199310000-00008
   Knosp E, 1996, NEUROSURGERY, V38, P434, DOI 10.1097/00006123-199603000-00003
   KOTAPKA MJ, 1994, J NEUROSURG, V81, P252, DOI 10.3171/jns.1994.81.2.0252
   KRAJEWSKI R, 1992, NEUROSURGERY, V30, P591
   KRISHT AF, 2000, CAVERNOUS SINUS COMP, P297
   LAM B, 2000, CAVERNOUS SINUS, P111
   LANZINO G, 1993, SURG NEUROL, V40, P359, DOI 10.1016/0090-3019(93)90214-L
   LARSON JJ, 1995, J NEUROSURG, V83, P596, DOI 10.3171/jns.1995.83.4.0596
   Lee JYK, 2002, J NEUROSURG, V97, P65, DOI 10.3171/jns.2002.97.1.0065
   LINSEY M, 2000, CAVERNOUS SINUS, P55
   LINSKEY ME, 1992, NEUROSURGERY, V30, P101, DOI 10.1227/00006123-199201000-00018
   Mariniello G, 1999, J NEUROSURG, V91, P1045, DOI 10.3171/jns.1999.91.6.1045
   Maruyama K, 2004, NEUROSURGERY, V55, P1068, DOI 10.1227/01.NEU.0000140839.47922.5A
   MEYER FB, 1990, J NEUROSURG, V73, P187, DOI 10.3171/jns.1990.73.2.0187
   Nakatomi H, 1996, SURG NEUROL, V46, P272, DOI 10.1016/0090-3019(96)00159-0
   OSullivan MG, 1997, NEUROSURGERY, V40, P238, DOI 10.1097/00006123-199702000-00003
   SCHUBIGER O, 1980, SURG NEUROL, V13, P313
   Sekhar L N, 1996, Acta Neurochir Suppl, V65, P58
   Sekhar L N, 1991, Acta Neurochir Suppl (Wien), V53, P101
   SEKHAR LN, 1989, NEUROSURGERY, V24, P18, DOI 10.1227/00006123-198901000-00004
   SEKHAR LN, 1992, J NEUROSURG, V76, P935, DOI 10.3171/jns.1992.76.6.0935
   SEKIYA T, 1993, NEUROSURGERY, V32, P227, DOI 10.1227/00006123-199302000-00012
   Sen C, 1997, J NEUROSURG, V87, P535, DOI 10.3171/jns.1997.87.4.0535
   SEPEHRNIA A, 1990, NEUROSURGERY, V27, P151, DOI 10.1227/00006123-199007000-00026
   Shin M, 2001, J NEUROSURG, V95, P435, DOI 10.3171/jns.2001.95.3.0435
   Shin M, 2000, J NEUROSURG, V93, P2, DOI 10.3171/jns.2000.93.supplement_3.0002
   SLONIEWSKI P, 1998, CHIRURG PODSTAWY CZA, P41
   Thapar K, 1996, NEUROSURGERY, V38, P99, DOI 10.1097/00006123-199601000-00024
   van Overbeeke JJ, 1998, J NEUROSURG, V88, P336, DOI 10.3171/jns.1998.88.2.0336
   Weisz DJ, 2000, CAVERNOUS SINUS, P123
   Yokoyama S, 2001, NEUROSURGERY, V49, P857, DOI 10.1097/00006123-200110000-00014
   Zielinski Grzegorz, 2003, Neurol Neurochir Pol, V37, P1239
   Zielinski Grzegorz, 2002, Neurol Neurochir Pol, V36, P69
NR 57
TC 2
Z9 2
U1 0
U2 1
PU ELSEVIER URBAN & PARTNER SP Z O O
PI WROCLAW
PA UL MIGDALOWA 4, LOK 59, WROCLAW, 02-796, POLAND
SN 0028-3843
J9 NEUROL NEUROCHIR POL
JI Neurol. Neurochir. Pol.
PY 2008
VL 42
IS 5
BP 402
EP 415
PG 14
WC Clinical Neurology
WE Science Citation Index Expanded (SCI-EXPANDED)
SC Neurosciences & Neurology
GA 408BE
UT WOS:000263407300003
PM 19105109
DA 2023-08-27
ER

PT J
AU Portela-Gomes, GM
   Grimelius, L
   Stridsberg, M
AF Portela-Gomes, Guida M.
   Grimelius, Lars
   Stridsberg, Mats
TI Immunohistochemical and Biochemical Studies with Region-Specific
   Antibodies to Chromogranins A and B and Secretogranins II and III in
   Neuroendocrine Tumors
SO CELLULAR AND MOLECULAR NEUROBIOLOGY
LA English
DT Article; Proceedings Paper
CT 15th International Symposium on Chromaffin Cell Biology
CY NOV 12-16, 2009
CL Merida, MEXICO
DE Chromogranin A; Chromogranin B; Secretogranin II; Secretogranin III;
   Neuroendocrine tumor; Immunohistochemistry; Plasma concentration
   measurements
ID HUMAN GASTROINTESTINAL-TRACT; SYNAPTIC VESICLE PROTEIN-2;
   ADRENAL-MEDULLA; CHROMAFFIN GRANULES; ISLET CELLS; PANCREAS; PEPTIDE;
   POLYPEPTIDE; SECRETION; NEOPLASMS
AB This short review deals with our investigations in neuroendocrine tumors (NETs) with antibodies against defined epitopes of chromogranins (Cgs) A and B and secretogranins (Sgs) II and III. The immunohistochemical expression of different epitopes of the granin family of proteins varies in NE cells in normal human endocrine and non-endocrine organs and in NETs, suggesting post-translational processing. In most NETs one or more epitopes of the granins were lacking, but variations in the expression pattern occurred both in benign and malignant NETs. A few epitopes displayed patterns that may be valuable in differentiating between benign and malignant NET types, e.g., well-differentiated NET types expressed more CgA epitopes than the poorly differentiated ones and C-terminal secretoneurin visualized a cell type related to malignancy in pheochromocytomas. Plasma concentrations of different epitopes of CgA and CgB varied. In patients suffering from carcinoid tumors or endocrine pancreatic tumors the highest concentrations were found with epitopes from the mid-portion of CgA. For CgB the highest plasma concentrations were recorded for the epitope 439-451. Measurements of SgII showed that patients with endocrine pancreatic tumors had higher concentrations than patients with carcinoid tumors or pheochromocytomas. SgIII was not detectable in patients with NETs.
C1 [Portela-Gomes, Guida M.] Univ Lisbon, Dept Gastroenterol, P-1699 Lisbon, Portugal.
   [Grimelius, Lars] Univ Hosp, Dept Genet & Pathol, Uppsala, Sweden.
   [Stridsberg, Mats] Univ Hosp, Dept Med Sci, Uppsala, Sweden.
C3 Universidade de Lisboa; Uppsala University; Uppsala University Hospital;
   Uppsala University; Uppsala University Hospital
RP Portela-Gomes, GM (通讯作者)，Rua Domingos Sequeira 128, P-2765525 S Pedro Do Estoril, Estoril, Portugal.
EM
CR BANKS P, 1965, BIOCHEM J, V97, pC40, DOI 10.1042/bj0970040C
   BORDI C, 1991, CANCER-AM CANCER SOC, V67, P663, DOI 10.1002/1097-0142(19910201)67:3<663::AID-CNCR2820670323>3.0.CO;2-L
   Cunningham RT, 1999, J PATHOL, V187, P321
   FAHRENKAMP AG, 1995, VIRCHOWS ARCH, V426, P361
   Feldman SA, 2003, ENDOCR PATHOL, V14, P3, DOI 10.1385/EP:14:1:3
   FISCHERCOLBRIE R, 1987, ANN NY ACAD SCI, V493, P120, DOI 10.1111/j.1749-6632.1987.tb27189.x
   FISCHERCOLBRIE R, 1995, PROG NEUROBIOL, V46, P49, DOI 10.1016/0301-0082(94)00060-U
   GALINDO E, 1991, P NATL ACAD SCI USA, V88, P1426, DOI 10.1073/pnas.88.4.1426
   Guillemot J, 2006, INT J CANCER, V118, P2003, DOI 10.1002/ijc.21571
   Helle KB, 2007, CELL MOL LIFE SCI, V64, P2863, DOI 10.1007/s00018-007-7254-0
   Helle KB, 2004, BIOL REV, V79, P769, DOI 10.1017/S146479310400644X
   HELLE KB, 1966, MOL PHARMACOL, V2, P298
   HSI KL, 1982, FEBS LETT, V147, P261, DOI 10.1016/0014-5793(82)81055-7
   Ischia R, 1997, J BIOL CHEM, V272, P11657
   Jin L, 2003, ENDOCR PATHOL, V14, P37, DOI 10.1385/EP:14:1:37
   Kruger PG, 2003, REGUL PEPTIDES, V114, P29, DOI 10.1016/S0167-0115(03)00069-7
   LEE RWH, 1983, J BIOL CHEM, V258, P1326
   MARTENS GJM, 1988, FEBS LETT, V234, P160, DOI 10.1016/0014-5793(88)81324-3
   Oberg K, 2004, ACTA ONCOL, V43, P617, DOI 10.1080/02841860410018575
   OCONNOR DT, 1986, NEW ENGL J MED, V314, P1145, DOI 10.1056/NEJM198605013141803
   OTTIGER HP, 1990, J NEUROSCI, V10, P3135
   Portela-Gomes G, 2005, VIRCHOWS ARCH, V446, P604, DOI 10.1007/s00428-005-1222-6
   Portela-Gomes GM, 2000, AM J PATHOL, V157, P1299, DOI 10.1016/S0002-9440(10)64645-7
   Portela-Gomes GM, 2004, APMIS, V112, P663, DOI 10.1111/j.1600-0463.2004.t01-1-apm1121003.x
   Portela-Gomes GM, 2002, J HISTOCHEM CYTOCHEM, V50, P1487, DOI 10.1177/002215540205001108
   Portela-Gomes GM, 2002, J HISTOCHEM CYTOCHEM, V50, P1023, DOI 10.1177/002215540205000804
   Portela-Gomes GM, 2000, ADV EXP MED BIOL, V482, P193
   Portela-Gomes GM, 2001, J HISTOCHEM CYTOCHEM, V49, P483, DOI 10.1177/002215540104900408
   Portela-Gomes GM, 2001, AM J SURG PATHOL, V25, P1261, DOI 10.1097/00000478-200110000-00006
   Portela-Gomes GM, 2010, REGUL PEPTIDES, V165, P12, DOI 10.1016/j.regpep.2010.02.011
   Portela-Gomes GM, 2010, REGUL PEPTIDES, V165, P30, DOI 10.1016/j.regpep.2010.06.002
   PortelaGomes GM, 1997, J HISTOCHEM CYTOCHEM, V45, P815, DOI 10.1177/002215549704500606
   ROSA P, 1981, MOL CELL ENDOCRINOL, V24, P181, DOI 10.1016/0303-7207(81)90058-7
   Solcia E, 2000, HISTOLOGICAL TYPING, P56
   Sonnleitner-Wittauer U.HackerGW, 1996, CELL VISION, V3, P27
   SRIKANTA S, 1986, DIABETES, V35, P300, DOI 10.2337/diabetes.35.3.300
   Stridsberg M, 2008, J ANAT, V212, P229, DOI 10.1111/j.1469-7580.2008.00857.x
   Stridsberg M, 2005, REGUL PEPTIDES, V125, P193, DOI 10.1016/j.regpep.2004.08.027
   Stridsberg M, 2004, REGUL PEPTIDES, V117, P219, DOI 10.1016/j.regpep.2003.10.023
   Stridsberg M, 2000, ADV EXP MED BIOL, V482, P319
   STRIDSBERG M, 1995, J ENDOCRINOL, V144, P49, DOI 10.1677/joe.0.1440049
   Stridsberg M, 2008, REGUL PEPTIDES, V148, P95, DOI 10.1016/j.regpep.2008.03.007
   Stridsberg M, 2007, REGUL PEPTIDES, V139, P80, DOI 10.1016/j.regpep.2006.10.008
   Strub JM, 1996, J BIOL CHEM, V271, P28533, DOI 10.1074/jbc.271.45.28533
   Tartaglia A, 2006, VIRCHOWS ARCH, V448, P399, DOI 10.1007/s00428-005-0113-1
   Thompson LDR, 2002, AM J SURG PATHOL, V26, P551, DOI 10.1097/00000478-200205000-00002
   TWARDZIK DR, 1985, P NATL ACAD SCI USA, V82, P5300, DOI 10.1073/pnas.82.16.5300
   WATKINSON A, 1991, BIOCHEM J, V276, P471, DOI 10.1042/bj2760471
   WEILER R, 1988, AM J SURG PATHOL, V12, P877, DOI 10.1097/00000478-198811000-00008
   Winkler H, 1998, Adv Pharmacol, V42, P257
NR 50
TC 8
Z9 8
U1 0
U2 4
PU SPRINGER/PLENUM PUBLISHERS
PI NEW YORK
PA 233 SPRING ST, NEW YORK, NY 10013 USA
SN 0272-4340
EI 1573-6830
J9 CELL MOL NEUROBIOL
JI Cell. Mol. Neurobiol.
PD NOV
PY 2010
VL 30
IS 8
SI SI
BP 1147
EP 1153
DI 10.1007/s10571-010-9585-x
PG 7
WC Cell Biology; Neurosciences
WE Science Citation Index Expanded (SCI-EXPANDED); Conference Proceedings Citation Index - Science (CPCI-S)
SC Cell Biology; Neurosciences & Neurology
GA 703AX
UT WOS:000285947200003
PM 21046454
DA 2023-08-27
ER

PT J
AU Han, MS
   Jung, S
   Kim, IY
   Moon, KS
   Jung, TY
   Jang, WY
AF Han, Moon-Soo
   Jung, Shin
   Kim, In-Young
   Moon, Kyung-Sub
   Jung, Tae-Young
   Jang, Woo-Youl
TI Hemifacial Spasm Caused by Small Epidermoid Tumor, Misinterpreted as
   Delayed Secondary Hemifacial Spasm Caused by Vestibular Schwannoma
   Treated with Gamma Knife Surgery
SO WORLD NEUROSURGERY
LA English
DT Article
DE Epidermoid cyst; Hemifacial spasm; Secondary hemifacial spasm;
   Vestibular schwannoma
ID CEREBELLOPONTINE ANGLE; NERVE; CYST
AB BACKGROUND: Hemifacial spasm (HFS), one of the most common hyperactive cranial rhizopathies, is a disorder characterized by spontaneous, intermittent, and repetitive contraction of unilateral facial muscle. The most common cause of HFS is a mechanical compression of the facial nerve at the root exit zone (REZ) by blood vessels located in the brainstem. The incidence of cerebellopontine angle (CPA) tumor-induced HFS ranges from 0.3% - 2.5% of all patients with HFS, and the incidence of epidermoid cyst among such tumors is extremely rare (0.2% to 0.25%). We present a patient manifesting HFS induced by a small epidermoid cyst who underwent Gamma Knife surgery (GKS) for ipsilateral vestibular schwannoma (VS) more than 10 years ago.
   CLINICAL DESCRIPTION: A 59-year-old female was admitted to our hospital with HFS. Ten years before admission, the patient underwent GKS for ipsilateral VS. Enhanced brain magnetic resonance imaging demonstrated that the volume of VS was not increased. Also, no definitive mechanical compression of the facial nerve REZ by blood vessels was detected. We concluded that HFS was secondary to the VS treatment by GKS and decided to operate to remove the tumor and identify REZ. A standardized lateral suboccipital retrosigmoid approach was performed. The cochlear nerve was encased by a tumor mass, and the posterior inferior cerebellar artery (PICA) was displaced by the mass. During tumor excision, we ensured that the VS and displaced PICA did not directly compress the REZ. Interestingly, an unexpected lesion was found with a yellowish viscous mass located between the sixth nerve and ventral side of the seventh nerve REZ. After the tumor removal, a small piece of Teflon was inserted between the PICA and REZ of the facial nerve. Postoperatively, the patient experienced no HFS or facial nerve dysfunction. Histopathologic examination of the lesion revealed an epidermoid cyst.
   CONCLUSIONS: We should consider various possibilities for the cause of HFS and review brain imaging meticulously if patients were treated for CPA tumor with GKS a long time ago and the delayed HFS occurred without changes in tumor volume. HFS induced by epidermoid cyst is rare. Therefore it is critical for the neurosurgeon to ensure the absence of any abnormal contact with the REZ of the facial nerve during surgery for HFS induced by CPA tumors.
C1 [Han, Moon-Soo; Jung, Shin; Kim, In-Young; Moon, Kyung-Sub; Jung, Tae-Young; Jang, Woo-Youl] Chonnam Natl Univ, Hwasun Hosp & Med Sch, Brain Tumor Clin, Dept Neurosurg,Res Inst Med Sci, Hwasun, Jeollanam Do, South Korea.
   [Han, Moon-Soo; Jung, Shin; Kim, In-Young; Moon, Kyung-Sub; Jung, Tae-Young; Jang, Woo-Youl] Chonnam Natl Univ, Hwasun Hosp & Med Sch, Gamma Knife Ctr, Dept Neurosurg,Res Inst Med Sci, Hwasun, Jeollanam Do, South Korea.
C3 Chonnam National University; Chonnam National University
RP Jung, S (通讯作者)，Chonnam Natl Univ, Hwasun Hosp & Med Sch, Brain Tumor Clin, Dept Neurosurg,Res Inst Med Sci, Hwasun, Jeollanam Do, South Korea.
EM
CR Berrettini S, 1996, J NEUROL SCI, V144, P24, DOI 10.1016/S0022-510X(96)00174-8
   Cancelli I, 2005, NEUROL SCI, V26, P46, DOI 10.1007/s10072-005-0382-0
   Choi SK, 2009, J KOREAN NEUROSURG S, V45, P196, DOI 10.3340/jkns.2009.45.3.196
   Chung SS, 2000, ACTA NEUROCHIR, V142, P901, DOI 10.1007/s007010070076
   GORMLEY WB, 1994, ACTA NEUROCHIR, V128, P115, DOI 10.1007/BF01400660
   Han HY, 2010, NEUROSURG REV, V33, P335, DOI 10.1007/s10143-010-0250-0
   Harada A, 2002, J NEUROSURG, V97, P482, DOI 10.3171/jns.2002.97.2.0482
   HIGASHI S, 1992, SURG NEUROL, V37, P289, DOI 10.1016/0090-3019(92)90155-G
   Iwai Y, 2001, NEUROL MED-CHIR, V41, P87, DOI 10.2176/nmc.41.87
   JANNETTA PJ, 1977, J NEUROSURG, V47, P321, DOI 10.3171/jns.1977.47.3.0321
   Kobata H, 2002, NEUROSURGERY, V50, P276, DOI 10.1097/00006123-200202000-00008
   Lee SH, 2010, ACTA NEUROCHIR, V152, P1901, DOI 10.1007/s00701-010-0796-1
   Miyazaki S, 1983, No To Shinkei, V35, P951
   NAGATA S, 1992, SURG NEUROL, V38, P204, DOI 10.1016/0090-3019(92)90170-R
   Sandberg DI, 1999, PEDIATR NEUROL, V21, P754, DOI 10.1016/S0887-8994(99)00077-6
   SPRIK C, 1988, OPHTHALMOLOGY, V95, P1042
   Zhang X, 2019, WORLD NEUROSURG, V125, pE10, DOI 10.1016/j.wneu.2018.12.035
NR 17
TC 2
Z9 2
U1 0
U2 1
PU ELSEVIER SCIENCE INC
PI NEW YORK
PA STE 800, 230 PARK AVE, NEW YORK, NY 10169 USA
SN 1878-8750
EI 1878-8769
J9 WORLD NEUROSURG
JI World Neurosurg.
PD OCT
PY 2019
VL 130
BP 410
EP 414
DI 10.1016/j.wneu.2019.07.119
PG 5
WC Clinical Neurology; Surgery
WE Science Citation Index Expanded (SCI-EXPANDED)
SC Neurosciences & Neurology; Surgery
GA JB0MH
UT WOS:000488248200214
PM 31349082
DA 2023-08-27
ER

PT J
AU Zhou, XL
   Spittau, B
AF Zhou, Xiaolai
   Spittau, Bjoern
TI Lipopolysaccharide-Induced Microglia Activation Promotes the Survival of
   Midbrain Dopaminergic Neurons In Vitro
SO NEUROTOXICITY RESEARCH
LA English
DT Article
DE mDA neuron; LPS; Microglia; Astrocytes; TGF beta
ID PARKINSONS-DISEASE; TGF-BETA; GENE-EXPRESSION; INDUCED
   NEURODEGENERATION; INDUCED NEUROTOXICITY; NEUROTROPHIC FACTOR;
   NITRIC-OXIDE; BRAIN-INJURY; RAT MODEL; NEUROPROTECTION
AB Microglia are the resident immune cells of the central nervous system (CNS) and respond to a variety of endogenous and exogenous stimuli in order to restore cell and tissue homeostasis. Lipopolysaccharide (LPS) is one of these exogenous stimuli, constitutes a major component of the outer membrane of Gram-negative bacteria, and binds to the microglial pattern recognition receptor Toll-like receptor 4 (TLR4). LPS-induced microglia activation is believed to promote neurodegeneration by release of neurotoxic factors such as interleukin-1 beta, tumor necrosis factor alpha, or nitric oxide. In the present study, we investigated whether the physical presence of microglia is required to promote neurotoxicity and whether microglia-derived factors are essential. Interestingly, we observed that dopaminergic (mDA) neuron survival was only affected in mixed neuron-glia cultures containing microglia but not in neuron-enriched cultures. Moreover, we clearly demonstrate that microglia-conditioned medium (MCM) after LPS treatment increased mDA neuron survival, process numbers as well as process length. The observed protective effects of MCM was rather caused by microglia-derived factors and only partially dependent on the increase in reactive astrocytes. These results indicate that LPS-induced microglia activation does not necessarily have detrimental effects on mDA neurons and further support the hypothesis that activated microglia support neuron survival by release of neurotrophic and neuroprotective factors.
C1 [Zhou, Xiaolai] Mayo Clin Jacksonville, Dept Neurosci, 4500 San Pablo Rd, Jacksonville, FL 32224 USA.
   [Spittau, Bjoern] Univ Rostock, Inst Anat, Rostock, Germany.
   [Zhou, Xiaolai; Spittau, Bjoern] Univ Freiburg, Inst Anat & Cell Biol, Dept Mol Embryol, Fac Med, Freiburg, Germany.
C3 Mayo Clinic; University of Rostock; University of Freiburg
RP Spittau, B (通讯作者)，Univ Rostock, Inst Anat, Rostock, Germany.; Spittau, B (通讯作者)，Univ Freiburg, Inst Anat & Cell Biol, Dept Mol Embryol, Fac Med, Freiburg, Germany.
EM
RI Spittau, Björn/AGZ-8396-2022; Zhou, Xiaolai/AAF-7227-2020
OI Spittau, Bjorn/0000-0001-5423-0571
FU DFG [SP 1555/2-1] Funding Source: Medline
CR ABE M, 1994, ANAL BIOCHEM, V216, P276, DOI 10.1006/abio.1994.1042
   ADAMS JC, 1981, J HISTOCHEM CYTOCHEM, V29, P775, DOI 10.1177/29.6.7252134
   Barcia C, 2012, SCI REP-UK, V2, DOI 10.1038/srep00809
   Block ML, 2007, NAT REV NEUROSCI, V8, P57, DOI 10.1038/nrn2038
   Bolin LM, 2002, J NEUROCHEM, V83, P167, DOI 10.1046/j.1471-4159.2002.01131.x
   Chen HB, 2016, SCI REP-UK, V6, DOI 10.1038/srep30459
   Chen X, 2017, J NEUROIMMUNE PHARM, V12, P433, DOI 10.1007/s11481-017-9732-y
   'Episcopo Francesca L, 2013, Curr Aging Sci, V6, P45
   Funk GD, 2015, COMP BIOCHEM PHYS A, V186, P83, DOI 10.1016/j.cbpa.2015.01.010
   Gayle DA, 2002, DEV BRAIN RES, V133, P27, DOI 10.1016/S0165-3806(01)00315-7
   Gibbons HM, 2006, BRAIN RES, V1084, P1, DOI 10.1016/j.brainres.2006.02.032
   GREEN LC, 1982, ANAL BIOCHEM, V126, P131, DOI 10.1016/0003-2697(82)90118-X
   Guha M, 2001, CELL SIGNAL, V13, P85, DOI 10.1016/S0898-6568(00)00149-2
   Herrera AJ, 2000, NEUROBIOL DIS, V7, P429, DOI 10.1006/nbdi.2000.0289
   Hirsch EC, 2009, LANCET NEUROL, V8, P382, DOI 10.1016/S1474-4422(09)70062-6
   Huhner L, 2017, FRONT MOL NEUROSCI, V10, DOI 10.3389/fnmol.2017.00062
   Iravani MM, 2012, NEUROSCI LETT, V510, P138, DOI 10.1016/j.neulet.2012.01.022
   Jellinger KA, 2001, ADV NEUROL, V86, P55
   Johnston LC, 2008, MOL THER, V16, P1392, DOI 10.1038/mt.2008.113
   Kim WG, 2000, J NEUROSCI, V20, P6309, DOI 10.1523/JNEUROSCI.20-16-06309.2000
   Kosloski LM, 2013, J NEUROIMMUNOL, V265, P1, DOI 10.1016/j.jneuroim.2013.10.009
   KRIEGLSTEIN K, 1995, EMBO J, V14, P736, DOI 10.1002/j.1460-2075.1995.tb07052.x
   Krieglstein K, 2002, J PHYSIOLOGY-PARIS, V96, P25, DOI 10.1016/S0928-4257(01)00077-8
   LAWSON LJ, 1990, NEUROSCIENCE, V39, P151, DOI 10.1016/0306-4522(90)90229-W
   Le WD, 2016, FRONT MOL NEUROSCI, V9, DOI 10.3389/fnmol.2016.00089
   Li L, 2007, BRAIN RES, V1159, P8, DOI 10.1016/j.brainres.2007.04.066
   Liu W, 2011, LIFE SCI, V89, P141, DOI 10.1016/j.lfs.2011.05.011
   Liu YX, 2008, NEUROTOXICOLOGY, V29, P864, DOI 10.1016/j.neuro.2008.02.014
   Lu YC, 2008, CYTOKINE, V42, P145, DOI 10.1016/j.cyto.2008.01.006
   Machado V, 2016, INT J MOL SCI, V17, DOI 10.3390/ijms17020151
   Machado V, 2016, NEUROBIOL DIS, V88, P1, DOI 10.1016/j.nbd.2015.12.016
   Mccoy MK, 2006, J NEUROSCI, V26, P9365, DOI 10.1523/JNEUROSCI.1504-06.2006
   Polazzi E, 2009, J NEUROCHEM, V110, P545, DOI 10.1111/j.1471-4159.2009.06117.x
   Prinz M, 2014, NAT REV NEUROSCI, V15, P300, DOI 10.1038/nrn3722
   Qian L, 2008, J IMMUNOL, V181, P660, DOI 10.4049/jimmunol.181.1.660
   Qin LY, 2007, GLIA, V55, P453, DOI 10.1002/glia.20467
   Qin LY, 2004, J BIOL CHEM, V279, P1415, DOI 10.1074/jbc.M307657200
   Rickert Uta, 2014, J Neurodegener Dis, V2014, P369468, DOI 10.1155/2014/369468
   Roussa E, 2009, ADV EXP MED BIOL, V651, P81
   Sharaf A, 2013, CELL TISSUE RES, V351, P373, DOI 10.1007/s00441-012-1537-y
   Song SJ, 2016, RESTOR NEUROL NEUROS, V34, P415, DOI 10.3233/RNN-150607
   Spittau B, 2015, GLIA, V63, P142, DOI 10.1002/glia.22740
   Spittau B, 2013, GLIA, V61, P287, DOI 10.1002/glia.22435
   Spittau B, 2012, NEUROMOL MED, V14, P317, DOI 10.1007/s12017-012-8189-7
   Tanaka T, 2008, NEUROSCI LETT, V431, P262, DOI 10.1016/j.neulet.2007.11.051
   Tang SC, 2008, EXP NEUROL, V213, P114, DOI 10.1016/j.expneurol.2008.05.014
   Tang SC, 2007, P NATL ACAD SCI USA, V104, P13798, DOI 10.1073/pnas.0702553104
   Unsicker K, 2002, ADV EXP MED BIOL, V513, P353
   Watanabe H, 2000, NEUROSCI LETT, V289, P53, DOI 10.1016/S0304-3940(00)01252-0
   Wolf SA, 2017, ANNU REV PHYSIOL, V79, P619, DOI 10.1146/annurev-physiol-022516-034406
   Zhou XL, 2015, J NEUROCHEM, V134, P125, DOI 10.1111/jnc.13111
   Zhu CH, 2016, J EXP MED, V213, P1047, DOI 10.1084/jem.20151000
   Zhu Y, 2016, INT J MOL SCI, V17, DOI 10.3390/ijms17010025
   Zlotnik A, 2014, J NEUROIMMUNOL, V270, P22, DOI 10.1016/j.jneuroim.2014.03.006
NR 54
TC 7
Z9 7
U1 0
U2 10
PU SPRINGER
PI NEW YORK
PA 233 SPRING ST, NEW YORK, NY 10013 USA
SN 1029-8428
EI 1476-3524
J9 NEUROTOX RES
JI Neurotox. Res.
PD MAY
PY 2018
VL 33
IS 4
BP 856
EP 867
DI 10.1007/s12640-017-9842-6
PG 12
WC Neurosciences
WE Science Citation Index Expanded (SCI-EXPANDED)
SC Neurosciences & Neurology
GA GD5HB
UT WOS:000430536900016
PM 29188488
DA 2023-08-27
ER

PT J
AU RamachandranNair, R
   Ochi, A
   Akiyama, T
   Buckley, DJ
   Soman, TB
   Weiss, SK
   Otsubo, H
AF RamachandranNair, R
   Ochi, A
   Akiyama, T
   Buckley, DJ
   Soman, TB
   Weiss, SK
   Otsubo, H
TI Partial seizures triggering infantile spasms in the presence of a basal
   ganglia glioma
SO EPILEPTIC DISORDERS
LA English
DT Article
DE infantile spasms; partial seizures; basal ganglia glioma; tumor
ID WEST SYNDROME; INTRACRANIAL TUMORS
AB Infantile spasms associated with brain tumors have been reported. A focal cortical lesion can induce infantile spasms by triggering the brainstem and basal ganglia in this vulnerable age group. We report the case of a female infant with a low-grade glioma in the right basal ganglia, spreading to the cortical area. She presented at the age of five months with left hemiparesis and partial seizures. She developed infantile spasms at the age of 12 months. This is the first video clip report of partial seizures triggering symmetrical spasms in series, secondary to a basal ganglia glioma extending to the cortex.
C1 Univ Toronto, Hosp Sick Children, Div Neurol, Toronto, ON M5G 1X8, Canada.
C3 University of Toronto; Hospital for Sick Children (SickKids)
RP Otsubo, H (通讯作者)，Univ Toronto, Hosp Sick Children, Div Neurol, 555 Univ Ave, Toronto, ON M5G 1X8, Canada.
EM
CR Aktan G, 1997, J CHILD NEUROL, V12, P152, DOI 10.1177/088307389701200216
   ASANUMA H, 1995, PEDIATR NEUROL, V12, P361, DOI 10.1016/0887-8994(95)00057-M
   ASKENASI A, 1991, J CHILD NEUROL, V6, P180
   BRANCH CE, 1979, ANN NEUROL, V5, P302, DOI 10.1002/ana.410050315
   Chugani HT, 2002, ADV EXP MED BIOL, V497, P111
   DIROCCO C, 1991, CHILD NERV SYST, V7, P150, DOI 10.1007/BF00776711
   DONAT JF, 1991, J CHILD NEUROL, V6, P246, DOI 10.1177/088307389100600308
   Frost JD, 2005, J CLIN NEUROPHYSIOL, V22, P25, DOI 10.1097/01.WNP.0000149893.12678.44
   GABRIEL YH, 1980, ANN NEUROL, V7, P287, DOI 10.1002/ana.410070316
   GASTAUT H, 1978, DEV MED CHILD NEUROL, V20, P21
   KUROKAWA T, 1980, PEDIATRICS, V65, P81
   MIMAKI T, 1983, ANN NEUROL, V14, P695, DOI 10.1002/ana.410140621
   OHTAHARA S, 1993, EPILEPSIA, V34, P716, DOI 10.1111/j.1528-1157.1993.tb00451.x
   Ohtsuka Y, 1996, EPILEPSIA, V37, P1060, DOI 10.1111/j.1528-1157.1996.tb01025.x
   Otsubo H, 1999, CLIN NEUROPHYSIOL, V110, P374, DOI 10.1016/S1388-2457(98)00024-8
   RUGGIERI V, 1989, PEDIATR NEUROL, V5, P327, DOI 10.1016/0887-8994(89)90029-5
   SHIELDS WD, 1992, EPILEPSIA, V33, pS26
   Vigevano F, 2001, BRAIN DEV-JPN, V23, P467, DOI 10.1016/S0387-7604(01)00284-4
   Wyllie E, 1996, EPILEPSIA, V37, P625, DOI 10.1111/j.1528-1157.1996.tb00626.x
   YAMAMOTO N, 1988, EPILEPSIA, V29, P34, DOI 10.1111/j.1528-1157.1988.tb05095.x
NR 20
TC 11
Z9 11
U1 0
U2 1
PU JOHN LIBBEY EUROTEXT LTD
PI MONTROUGE
PA 127 AVE DE LA REPUBLIQUE, 92120 MONTROUGE, FRANCE
SN 1294-9361
EI 1950-6945
J9 EPILEPTIC DISORD
JI Epileptic Disord.
PD DEC
PY 2005
VL 7
IS 4
BP 378
EP 382
PG 5
WC Clinical Neurology
WE Science Citation Index Expanded (SCI-EXPANDED)
SC Neurosciences & Neurology
GA 004JP
UT WOS:000234749400013
PM 16338683
DA 2023-08-27
ER

PT J
AU Nakamura, M
   Samii, M
AF Nakamura, M
   Samii, M
TI Surgical management of a meningioma in the retrosellar region
SO ACTA NEUROCHIRURGICA
LA English
DT Article
DE retrosellar region; clivus; meningioma; brain tumour
ID PETROCLIVAL MENINGIOMAS; BASILAR ARTERY
AB Lesions ventral to the brainstem in the retrosellar and interpeduncular region are major challenges to the surgeon because of their location at great depth in the centre of the cranial base. We report the operative management of a patient with a meningioma in an unusual location, extending from the upper clivus, retrosellar and interpeduncular region into the suprasellar area up to the level of the foramen of Monro.
   A 41-year-old man presented with a 3-month history of progressive visual disturbance, episodic headache and signs and symptoms of endocrinological disturbance. Magnetic resonance imaging (MRI) studies showed a homogeneously enhancing tumour ventral to the brainstem with large cranio-caudal extension from the upper clivus to the suprasellar area.
   Operative removal was planned in two stages. First, through a right lateral suboccipital retrosigmoid craniectomy the caudal portion of the tumour at the upper clivus and prepontine region was removed. Second, the residual suprasellar part of the tumour was removed totally through a fronto-lateral craniotomy on the right side one week later.
   When a tumour is very large or involves different areas of the skull base, it is necessary to decide between removal in one stage, which requires a complex and time consuming skull base approach, or in multiple stages. Our case demonstrates how a retrosellar meningioma extending to the suprasellar region can be totally removed using two simple skull base approaches without the risk of compromise to venous drainage and without the need for extensive bone removal as described in other skull base approaches.
C1 Nordstadt Hosp, Dept Neurosurg, D-30167 Hannover, Germany.
   Sch Med, Dept Neurosurg, Hannover, Germany.
   Int Neurosci Inst, Hannover, Germany.
RP Nakamura, M (通讯作者)，Nordstadt Hosp, Dept Neurosurg, Haltenhoffstr 41, D-30167 Hannover, Germany.
RI Nakamura, Makoto/F-2620-2014
OI Nakamura, Makoto/0000-0001-8597-0193
CR al-Mefty O, 1991, Acta Neurochir Suppl (Wien), V53, P166
   Dolenc V V, 1987, Br J Neurosurg, V1, P251, DOI 10.3109/02688698709035309
   DRAKE CG, 1961, J NEUROSURG, V18, P230, DOI 10.3171/jns.1961.18.2.0230
   FUJITSU K, 1985, J NEUROSURG, V62, P340, DOI 10.3171/jns.1985.62.3.0340
   HAKUBA A, 1988, SURG NEUROL, V30, P108, DOI 10.1016/0090-3019(88)90095-X
   HAKUBA A, 1986, SURG NEUROL, V26, P271, DOI 10.1016/0090-3019(86)90161-8
   HITSELBERGER WE, 1966, ARCHIV OTOLARYNGOL, V84, P267
   Javed T, 1991, Acta Neurochir Suppl (Wien), V53, P171
   KAWASE T, 1991, NEUROSURGERY, V28, P869, DOI 10.1227/00006123-199106000-00014
   MacDonald JD, 1998, NEUROSURGERY, V43, P84, DOI 10.1097/00006123-199807000-00054
   Malis LI, 1985, NEUROSURGERY, V1, P1011
   Samii M, 1996, J NEUROSURG, V84, P375, DOI 10.3171/jns.1996.84.3.0375
   SAMII M, 1992, ACTA NEUROCHIR, V118, P27, DOI 10.1007/BF01400723
   Samii M, 1999, J CLIN NEUROSCI, V6, P27, DOI 10.1016/S0967-5868(99)90598-2
   SAMII M, 1988, ACTA NEUROCHIR, V95, P6, DOI 10.1007/BF01793075
   Samii M, 2000, J NEUROSURG, V92, P235, DOI 10.3171/jns.2000.92.2.0235
   Sano K, 1980, Neurol Res, V2, P361
   SEKHAR LN, 1986, TUMORS SKULL BASE EX, P141
   SHIOKAWA Y, 1989, NEUROSURGERY, V25, P793, DOI 10.1227/00006123-198911000-00016
   Symon L., 1982, ADV TECHNICAL STANDA, V9, P69
   Yasargil G., 1980, ADV TECHNICAL STANDA, V7, P1
   Yasargil M G, 1976, Surg Neurol, V6, P83
NR 22
TC 17
Z9 17
U1 0
U2 0
PU SPRINGER WIEN
PI WIEN
PA SACHSENPLATZ 4-6, PO BOX 89, A-1201 WIEN, AUSTRIA
SN 0001-6268
EI 0942-0940
J9 ACTA NEUROCHIR
JI Acta Neurochir.
PD MAR
PY 2003
VL 145
IS 3
BP 215
EP 220
DI 10.1007/s00701-002-1053-z
PG 6
WC Clinical Neurology; Surgery
WE Science Citation Index Expanded (SCI-EXPANDED)
SC Neurosciences & Neurology; Surgery
GA 658GZ
UT WOS:000181714400014
PM 12632118
DA 2023-08-27
ER

PT J
AU Kumar, R
   Kalra, SK
AF Kumar, Raj
   Kalra, Samir K.
TI Pediatric brain stem lesions: introduction of a scoring system for
   clinical evaluation and their treatment analysis
SO CHILDS NERVOUS SYSTEM
LA English
DT Article
DE brainstem lesions; outcome analysis; Karnofsky's performance score;
   Kumar's and Samir's score
ID PROGNOSTIC-FACTORS; GLIOMAS; CHILDREN; TUMORS; CHILDHOOD; MANAGEMENT;
   PATHOLOGY; SURVIVAL; FEATURES; QUALITY
AB Introduction Brainstem lesions in pediatric age group include mainly gliomas. As these are not usually amenable to complete surgical resection, multimodality treatments are used. To assess treatment responses, Karnofsky's Performance Score (KPS) has been the gold standard; however, in pediatric patients, comprehensive neurological examination is more valuable.
   Objective Our purpose was to formulate neurological examination based scoring system and compare its effectiveness with KPS.
   Materials and methods A retrospective analysis of 37 pediatric patients with brainstem lesions was performed. All these patients were assessed both pre- and post-treatment and in every follow up using standard KPS and our proposed scoring system, Kumar's and Samir's Score (K&SS). K&SS was devised choosing the common clinical features and those affecting outcome the most. Twenty-two features were hence selected, and points were allotted to each according to their functional importance, with range from 22 to 100.
   Results and discussion Twenty-seven had gliomas, and in nonglioma group, seven had brainstem tuberculosis (BSTB). Nineteen patients with BSG were less than 6 years of age. Most gliomas were pilocytic astrocytomas (n=21). Outcome analysis of patients in both groups using KPS showed deterioration or no change in the scores in majority whereas using K&SS, most patients showed improvement.
   Conclusion Gliomas form majority of pediatric brainstem lesions, with high occurrence of BSTB in nongliomatous group. Our proposed scoring system, based on comprehensive neurological examination, is more sensitive than KPS in treatment analysis of brainstem lesions.
C1 [Kumar, Raj; Kalra, Samir K.] Sanjay Gandhi Postgrad Inst Med Sci, Dept Neurosurg, Lucknow 226014, Uttar Pradesh, India.
C3 Sanjay Gandhi Postgraduate Institute of Medical Sciences
RP Kumar, R (通讯作者)，Sanjay Gandhi Postgrad Inst Med Sci, Dept Neurosurg, Lucknow 226014, Uttar Pradesh, India.
EM
CR ALBRIGHT AL, 1986, J NEUROSURG, V65, P751, DOI 10.3171/jns.1986.65.6.0751
   Benesch M, 2001, PEDIATR NEUROSURG, V35, P173, DOI 10.1159/000050418
   EPSTEIN F, 1986, J NEUROSURG, V64, P11, DOI 10.3171/jns.1986.64.1.0011
   Epstein F, 1996, PEDIATR NEUROSURG, V24, P24, DOI 10.1159/000121011
   Fisher PG, 2000, CANCER, V89, P1569, DOI 10.1002/1097-0142(20001001)89:7<1569::AID-CNCR22>3.0.CO;2-0
   Freeman CR, 1998, INT J RADIAT ONCOL, V40, P265, DOI 10.1016/S0360-3016(97)00572-5
   Galanis E, 2000, J NEUROSURG, V93, P201, DOI 10.3171/jns.2000.93.2.0201
   Gilbert M, 2000, SEMIN ONCOL, V27, pS20
   HOFFMAN HJ, 1997, IN PRESS CLIN NEUROS, V44, P549
   Kumar R, 2000, BRIT J NEUROSURG, V14, P356, DOI 10.1080/026886900417397
   KUN LE, 1983, J NEUROSURG, V58, P1, DOI 10.3171/jns.1983.58.1.0001
   LASSITER KR, 1971, J NEUROSURG, V34, P719, DOI 10.3171/jns.1971.34.6.0719
   Laws ER, 2003, J NEUROSURG, V99, P467, DOI 10.3171/jns.2003.99.3.0467
   Lesniak MS, 2003, PEDIATR NEUROSURG, V39, P314, DOI 10.1159/000075260
   MARIA BL, 1993, J CHILD NEUROL, V8, P112, DOI 10.1177/088307389300800203
   MILSTEIN JM, 1985, CANCER, V56, P1834, DOI 10.1002/1097-0142(19851001)56:7+<1834::AID-CNCR2820561323>3.0.CO;2-A
   MULHERN RK, 1994, PEDIATR NEUROSURG, V20, P226, DOI 10.1159/000120795
   PACKER RJ, 1994, CANCER-AM CANCER SOC, V74, P1827, DOI 10.1002/1097-0142(19940915)74:6<1827::AID-CNCR2820740628>3.0.CO;2-Q
   PACKER RJ, 1992, PEDIATR NEUROSURG, V17, P1404
   PANITCH HS, 1970, AM J DIS CHILD, V119, P465, DOI 10.1001/archpedi.1970.02100050467001
   POLLACK IF, 1994, NEW ENGL J MED, V331, P1500, DOI 10.1056/NEJM199412013312207
   POLLACK IF, 1993, J NEUROSURG, V78, P859, DOI 10.3171/jns.1993.78.6.0859
   ROBERTSON PL, 1995, PEDIATR NEUROSURG, V22, P65, DOI 10.1159/000120879
   ROBERTSON PL, 1994, NEUROLOGY, V44, P1798, DOI 10.1212/WNL.44.10.1798
   Samii M, 2001, J NEUROSURG, V95, P825, DOI 10.3171/jns.2001.95.5.0825
   Selvapandian S, 1999, ACTA NEUROCHIR, V141, P721, DOI 10.1007/s007010050367
   Shuper A, 1998, CHILD NERV SYST, V14, P578, DOI 10.1007/s003810050275
   STROINK AR, 1986, J NEUROSURG, V65, P745, DOI 10.3171/jns.1986.65.6.0745
   Tortosa A, 2003, CANCER, V97, P1063, DOI 10.1002/cncr.11120
   Wang CC, 2000, SURG NEUROL, V53, P41, DOI 10.1016/S0090-3019(99)00165-2
   Zimmerman RA, 1996, PEDIATR NEUROSURG, V25, P83, DOI 10.1159/000121102
NR 31
TC 3
Z9 3
U1 0
U2 0
PU SPRINGER
PI NEW YORK
PA 233 SPRING ST, NEW YORK, NY 10013 USA
SN 0256-7040
EI 1433-0350
J9 CHILD NERV SYST
JI Childs Nerv. Syst.
PD APR
PY 2008
VL 24
IS 4
BP 467
EP 475
DI 10.1007/s00381-007-0516-1
PG 9
WC Clinical Neurology; Pediatrics; Surgery
WE Science Citation Index Expanded (SCI-EXPANDED)
SC Neurosciences & Neurology; Pediatrics; Surgery
GA 267RA
UT WOS:000253525700009
PM 17978821
DA 2023-08-27
ER

PT J
AU Jacob, JT
   Pollock, BE
   Carlson, ML
   Driscoll, CLW
   Link, MJ
AF Jacob, Jeffrey T.
   Pollock, Bruce E.
   Carlson, Matthew L.
   Driscoll, Colin L. W.
   Link, Michael J.
TI Stereotactic Radiosurgery in the Management of Vestibular Schwannoma and
   Glomus Jugulare Indications, Techniques, and Results
SO OTOLARYNGOLOGIC CLINICS OF NORTH AMERICA
LA English
DT Article
DE Stereotactic radiosurgery; Vestibular schwannoma; Glomus jugulare; Skull
   base tumors
AB Gamma Knife stereotactic radiosurgery (GKS) has become an important management strategy for an increasing number of patients with skull base tumors. For select patients with lateral skull base disorders, given the proximity to sensitive critical structures such as the brainstem, cranial nerves, and cochlea, this technology has emerged as a first-line treatment to achieve the paramount goals of long-term tumor control and maintenance of existing neurologic function. This article reviews the indications, technique, and results of GKS for the treatment of vestibular schwannoma and glomus jugulare tumors, and highlights our experience in treating these tumors at the Mayo Clinic.
C1 [Jacob, Jeffrey T.; Pollock, Bruce E.; Driscoll, Colin L. W.; Link, Michael J.] Mayo Clin, Dept Neurol Surg, Sch Med, Rochester, MN 55905 USA.
   [Pollock, Bruce E.] Mayo Clin, Dept Radiat Oncol, Sch Med, Rochester, MN 55905 USA.
   [Carlson, Matthew L.; Driscoll, Colin L. W.; Link, Michael J.] Mayo Clin, Dept Otolaryngol Head & Neck Surg, Sch Med, Rochester, MN 55905 USA.
C3 Mayo Clinic; Mayo Clinic; Mayo Clinic
RP Link, MJ (通讯作者)，Mayo Clin, Dept Neurol Surg, 200 First St Southwest, Rochester, MN 55905 USA.
EM
NR 0
TC 21
Z9 21
U1 1
U2 5
PU W B SAUNDERS CO-ELSEVIER INC
PI PHILADELPHIA
PA 1600 JOHN F KENNEDY BOULEVARD, STE 1800, PHILADELPHIA, PA 19103-2899 USA
SN 0030-6665
EI 1557-8259
J9 OTOLARYNG CLIN N AM
JI Otolaryngol. Clin. N. Am.
PD JUN
PY 2015
VL 48
IS 3
BP 515
EP +
DI 10.1016/j.otc.2015.02.010
PG 13
WC Otorhinolaryngology
WE Science Citation Index Expanded (SCI-EXPANDED)
SC Otorhinolaryngology
GA CK9EW
UT WOS:000356544400010
PM 25873444
DA 2023-08-27
ER

PT J
AU Wang, SS
   Bandopadhayay, P
   Jenkins, MR
AF Wang, Stacie Shiqi
   Bandopadhayay, Pratiti
   Jenkins, Misty Rayna
TI Towards Immunotherapy for Pediatric Brain Tumors
SO TRENDS IN IMMUNOLOGY
LA English
DT Review
ID CAR-T-CELLS; INTRINSIC PONTINE GLIOMA; VARIANT III EGFRVIII; HIGH-GRADE;
   B-CELL; CANCER-IMMUNOTHERAPY; ANTIGEN PEPTIDES; OPEN-LABEL; PHASE-III;
   GLIOBLASTOMA
AB Pediatric brain tumors are the leading cause of childhood cancer-related death. Immunotherapy is a powerful new approach for treating some refractory cancers; applying this ` fourth pillar' of cancer treatment to pediatric brain tumors is an exciting but challenging prospect. This review offers new perspectives on moving towards successful immunotherapy for pediatric brain tumors, focusing on pediatric high-grade glioma (HGG), a subgroup with universally poor outcomes. We cover chimeric antigen receptor T cell (CAR-T) therapy, vaccine therapy, and checkpoint inhibition in this context, and focus on the need for intimately understanding the growing brain and its immune system. We highlight the challenges associated with the application of immunotherapy in pediatric neuro-oncology, as well as the tissue-specific challenges to be overcome, to achieve improved outcomes.
C1 [Wang, Stacie Shiqi; Jenkins, Misty Rayna] Walter & Eliza Hall Inst Med Res, Div Immunol, Parkville, Vic 3052, Australia.
   [Wang, Stacie Shiqi] Murdoch Childrens Res Inst, Parkville, Vic 3052, Australia.
   [Bandopadhayay, Pratiti] Dana Farber Boston Childrens Canc & Blood Disorde, Boston, MA 02115 USA.
   [Bandopadhayay, Pratiti] Harvard Med Sch, Dept Pediat, Boston, MA 02115 USA.
   [Bandopadhayay, Pratiti] Broad Inst MIT & Harvard, Cambridge, MA 02142 USA.
   [Jenkins, Misty Rayna] Univ Melbourne, Dept Med Biol, Parkville, Vic 3052, Australia.
   [Jenkins, Misty Rayna] La Trobe Univ, La Trobe Inst Mol Sci, Bundoora, Vic 3083, Australia.
C3 Walter & Eliza Hall Institute; Murdoch Children's Research Institute;
   Harvard University; Boston Children's Hospital; Dana-Farber Cancer
   Institute; Harvard University; Harvard Medical School; Harvard
   University; Massachusetts Institute of Technology (MIT); Broad
   Institute; University of Melbourne; La Trobe University
RP Jenkins, MR (通讯作者)，Walter & Eliza Hall Inst Med Res, Div Immunol, Parkville, Vic 3052, Australia.; Jenkins, MR (通讯作者)，Univ Melbourne, Dept Med Biol, Parkville, Vic 3052, Australia.; Jenkins, MR (通讯作者)，La Trobe Univ, La Trobe Inst Mol Sci, Bundoora, Vic 3083, Australia.
EM
RI ; Jenkins, Misty Rayna/I-5338-2015
OI Wang, Stacie Shiqi/0000-0001-5476-9323; Jenkins, Misty
   Rayna/0000-0001-6564-2715
CR Ahmed N, 2017, JAMA ONCOL, V3, P1094, DOI 10.1001/jamaoncol.2017.0184
   [Anonymous], FDA APPR NEW TREATM
   [Anonymous], J CLIN ONCOL
   Askew K, 2017, CELL REP, V18, P391, DOI 10.1016/j.celrep.2016.12.041
