## Supplemental Material 1 for "Bibliometric analysis of emerging trends and research foci in brainstem tumor field over thirty years (1992-2023)": savedrecs (6).txt

FN Clarivate Analytics Web of Science
VR 1.0
PT J
AU Komiyama, Y
   Nishimura, N
   Munakata, M
   Okuda, K
   Nishino, N
   Kosaka, C
   Masuda, M
   Mori, T
   Matsuda, T
   Takahashi, H
AF Komiyama, Y
   Nishimura, N
   Munakata, M
   Okuda, K
   Nishino, N
   Kosaka, C
   Masuda, M
   Mori, T
   Matsuda, T
   Takahashi, H
TI Increases in plasma ouabainlike immunoreactivity during surgical
   extirpation of pheochromocytoma
SO HYPERTENSION RESEARCH-CLINICAL AND EXPERIMENTAL
LA English
DT Article
DE ouabain; adrenalectomy; catecholamines; pheochromocytoma
ID BOVINE ADRENOCORTICAL-CELLS; HYPOTHALAMUS; DIGOXIN; RATS; PUMP
AB The ouabainlike factor (OLF) is thought to be an important modulator of salt and water metabolism. Plasma OLF could be derived from the central nervous system and/or the adrenal gland. Since the adrenal medulla is of neural origin, the cytology of pheochromocytoma of adrenomedullary origin resembles that of neuronal cells. Ouabainlike immunoreactivity (OLI) is, in fact, present in the adrenal medulla as shown by immunohistochemistry. The plasma levels of catecholamines and OLI were significantly elevated during surgical extirpation of pheochromocytoma in this case. To clarify the origin of circulating OLI in a patient with pheochromocytoma, the relationship between plasma OLI and catecholamines during adrenalectomy was investigated, Plasma catecholamine levels exceeded the normal reference interval, and plasma OLI was positively correlated with the patient's plasma level of norepinephrine. The peak level during operation was about 10 times higher than the baseline level. Both levels reached a maximum when the tumor was mechanically pressed, and then gradually decreased thereafter, The level of OLI in the tumor was higher than that of the normal adrenal cortex. When OLI in the tumor was characterized by reversed-phase high-performance liquid chromatography, the retention time of OLI corresponded with that of authentic ouabain. These results suggest that the circulating OLI in this patients was derived mainly from the pheochromocytoma of adrenomedullary origin.
C1 Kansai Med Univ, Dept Clin Sci & Lab Med, Moriguchi, Osaka 5708506, Japan.
   Kansai Med Univ, Dept Surg 2, Moriguchi, Osaka 5708506, Japan.
   Kansai Med Univ, Dept Urol, Moriguchi, Osaka 5708506, Japan.
C3 Kansai Medical University; Kansai Medical University; Kansai Medical
   University
RP Takahashi, H (通讯作者)，Kansai Med Univ, Dept Clin Sci & Lab Med, 10-15 Fumizono, Moriguchi, Osaka 5708506, Japan.
CR [Anonymous], PATHOPHYSIOLOGY
   DEWARDENER H, 1961, CLIN SCI, V21, P249
   DOOLITTLE MH, 1994, CLIN CHEM, V40, P487
   DORIS PA, 1994, HYPERTENSION, V23, P632, DOI 10.1161/01.HYP.23.5.632
   Doris PA, 1996, ENDOCRINOLOGY, V137, P533, DOI 10.1210/en.137.2.533
   HUANG BS, 1994, HYPERTENSION, V23, P1087, DOI 10.1161/01.HYP.23.6.1087
   Komiyama Y, 1997, CLIN EXP HYPERTENS, V19, P363, DOI 10.3109/10641969709080824
   Komiyama Y, 1996, J HYPERTENS, V14, P447
   LAREDO J, 1995, BIOCHEM BIOPH RES CO, V212, P487, DOI 10.1006/bbrc.1995.1996
   LAREDO J, 1994, ENDOCRINOLOGY, V135, P794, DOI 10.1210/en.135.2.794
   LEENEN FHH, 1993, AM J PHYSIOL, V265, pH421, DOI 10.1152/ajpheart.1993.265.1.H421
   Lim Dong-Yoon, 1993, Korean Journal of Internal Medicine (English Edition), V8, P103
   LUDENS JH, 1991, HYPERTENSION, V17, P923, DOI 10.1161/01.HYP.17.6.923
   NARUSE K, 1994, HYPERTENSION S1, V23, P102
   Perrin A, 1997, MOL CELL ENDOCRINOL, V126, P7, DOI 10.1016/S0303-7207(96)03964-0
   PIDGEON GB, 1994, CLIN SCI, V86, P391, DOI 10.1042/cs0860391
   Schneider R, 1998, J BIOL CHEM, V273, P784, DOI 10.1074/jbc.273.2.784
   TAKAHASHI H, 1994, SODIUM PUMP, P775
   Takahashi H, 1994, PATHOPHYSIOLOGY, V1, P25
   TAMURA M, 1994, J BIOL CHEM, V269, P11972
   TOROK TL, 1990, EXP PHYSIOL, V75, P573, DOI 10.1113/expphysiol.1990.sp003433
   YAMADA H, 1992, BRAIN RES, V584, P237, DOI 10.1016/0006-8993(92)90900-T
   YAMADA H, 1992, NEUROSCI LETT, V141, P143, DOI 10.1016/0304-3940(92)90881-7
NR 23
TC 24
Z9 25
U1 1
U2 1
PU JAPANESE SOC HYPERTENSION CENT ACADEMIC SOC, PUBL OFFICE
PI TOYONAKA
PA OSAKA, 14TH FL, SENRI LIFE SCI CENTER BLDG, 4-2 SHINSENRI- HIGASHI-MACHI
   1 CHOME, TOYONAKA, 565-0082, JAPAN
SN 0916-9636
J9 HYPERTENS RES-CLIN E
JI Hypertens. Res.-Clin. Exp.
PD JUL
PY 1999
VL 22
IS 2
BP 135
EP 139
DI 10.1291/hypres.22.135
PG 5
WC Peripheral Vascular Disease
WE Science Citation Index Expanded (SCI-EXPANDED)
SC Cardiovascular System & Cardiology
GA 219YG
UT WOS:000081635700010
PM 10487330
OA Bronze
DA 2023-08-27
ER

PT J
AU James, PE
   Grinberg, OY
   Goda, F
   Panz, T
   OHara, JA
   Swartz, HM
AF James, PE
   Grinberg, OY
   Goda, F
   Panz, T
   OHara, JA
   Swartz, HM
TI Gloxy: An oxygen-sensitive coal for accurate measurement of low oxygen
   tensions in biological systems
SO MAGNETIC RESONANCE IN MEDICINE
LA English
DT Article
DE EPR; oxygen
ID EPR OXIMETRY; BREAST-CANCER; INDIA INK; PROBE; RADIOTHERAPY; RESONANCE;
   FUSINITE; VITRO; VIVO
AB This paper describes the characteristics of a new oxygen sensitive, paramagnetic material that has some significant advantages for measurements of tissue pO(2) by in vivo EPR, This paramagnetic component of Welsh coal, termed ''gloxy'' was found to have valuable EPR features that allow accurate measurement of low oxygen tensions in vivo; these include large oxygen-dependent changes in linewidth, a high number of paramagnetic spin centers (resulting in high signal amplitude), and stability in tissue allowing repeated pO(2) measurements to be made in vivo with high precision, Renal pO(2) was measured deep in the medulla region of isolated perfused kidneys and found to be lower than that in the cortex (1.7 +/- 0.05 and 7.1 +/- 0.3 mm Hg, respectively), The quality of the EPR signal obtained from the renal outer medulla and also from tumors in mice was such that the pO(2) measurements were obtained with a precision of +/-3% of the measured pO(2) (Kidney: 1.7 +/- 0.05 mmHg; Tumor: 1.37 a 0.04 mmHg). In vitro tests on the viability of cells and in vivo studies using Glory demonstrate the stability and inertness of this oxygen-sensitive material.
C1 DARTMOUTH COLL SCH MED,DEPT RADIOL,EPR CTR,HANOVER,NH 03755.
C3 Dartmouth College
RI Panz, Tomasz/AAN-4799-2021
FU NCRR NIH HHS [RR01811, RR11802] Funding Source: Medline; NIGMS NIH HHS
   [GM 51630] Funding Source: Medline
CR BRAME JSS, 1956, FUEL SOLID LIQUID GA
   BREZIS M, 1991, J CLIN INVEST, V88, P390, DOI 10.1172/JCI115316
   BRIGGS H, 1930, COLLIERY ENG     MAY
   Bruley D. F., 1992, OXYGEN TRANSPORT TIS, P229
   CANNON GJ, 1992, J CELL SCI, V101, P907
   EPSTEIN E, 1989, J DERMATOL SURG ONC, V15, P273, DOI 10.1111/j.1524-4725.1989.tb03160.x
   FENNERTY MB, 1992, AM J GASTROENTEROL, V87, P79
   GODA F, 1995, MAGN RESON MED, V33, P237, DOI 10.1002/mrm.1910330214
   GODA F, 1995, CANCER RES, V55, P2249
   HARRIS JR, 1985, NEW ENGL J MED, V313, P1365, DOI 10.1056/NEJM198511213132128
   HUANG X, 1994, CHEM RES TOXICOL, V7, P451, DOI 10.1021/tx00039a025
   HUANG X, 1993, CHEM RES TOXICOL, V6, P453
   JAMES PE, 1995, FREE RADICAL BIO MED, V18, P641, DOI 10.1016/0891-5849(94)00179-N
   James PE, 1996, FREE RADICAL BIO MED, V21, P25, DOI 10.1016/0891-5849(95)02221-X
   JAMES PE, 1996, OXYGEN TRANSPORT TIS, V23
   LIU KJ, 1995, BRAIN RES, V685, P91, DOI 10.1016/0006-8993(95)00413-K
   LIU KJ, 1993, P NATL ACAD SCI USA, V90, P5438, DOI 10.1073/pnas.90.12.5438
   MATSUHASHI M, 1995, J BACTERIOL, V177, P688, DOI 10.1128/jb.177.3.688-693.1995
   MCCORMICK B, 1987, INT J RADIAT ONCOL, V13, P1667, DOI 10.1016/0360-3016(87)90163-5
   NILGES MJ, 1989, PHYS MED, V5, P195
   PARR SW, 1906, ILLINOIS STATE GEOLO, V3
   SMIRNOVA TI, 1994, J PHYS CHEM-US, V98, P2464, DOI 10.1021/j100060a039
   SWARTZ HM, 1991, MAGNET RESON MED, V20, P333, DOI 10.1002/mrm.1910200217
   TANAKA K, 1984, SOLID STATE COMMUN, V52, P343, DOI 10.1016/0038-1098(84)90841-X
   VAHIDI N, 1994, MAGN RESON MED, V31, P139, DOI 10.1002/mrm.1910310207
   WALKER S J, 1988, Radiography Today, V54, P27
   WHETTON JT, APPL ELECT RESISTIVI
   ZWEIER JL, 1994, J MAGN RESON SER B, V105, P52, DOI 10.1006/jmrb.1994.1099
NR 28
TC 43
Z9 44
U1 0
U2 2
PU WILLIAMS & WILKINS
PI BALTIMORE
PA 351 WEST CAMDEN ST, BALTIMORE, MD 21201-2436
SN 0740-3194
J9 MAGNET RESON MED
JI Magn.Reson.Med.
PD JUL
PY 1997
VL 38
IS 1
BP 48
EP 58
DI 10.1002/mrm.1910380109
PG 11
WC Radiology, Nuclear Medicine & Medical Imaging
WE Science Citation Index Expanded (SCI-EXPANDED)
SC Radiology, Nuclear Medicine & Medical Imaging
GA XH272
UT WOS:A1997XH27200008
PM 9211379
DA 2023-08-27
ER

PT J
AU Parekh, K
   LeBlang, S
   Nazarian, J
   Mueller, S
   Zacharoulis, S
   Hynynen, K
   Powlovich, L
AF Parekh, Kavya
   LeBlang, Suzanne
   Nazarian, Javad
   Mueller, Sabine
   Zacharoulis, Stergios
   Hynynen, Kullervo
   Powlovich, Lauren
TI Past, present and future of Focused Ultrasound as an adjunct or
   complement to DIPG/DMG therapy: A consensus of the 2021 FUSF DIPG
   meeting
SO NEOPLASIA
LA English
DT Article
ID BLOOD-BRAIN-BARRIER; DRUG-DELIVERY; DOXORUBICIN; DISRUPTION; TUMORS;
   MODEL; FLUORESCENCE; MANAGEMENT; KINETICS; SYSTEM
AB Diffuse Intrinsic Pontine Glioma (DIPG), now known as Diffuse Midline Glioma (DMG) is a devastating pediatric brain tumor with limited treatment options and a very poor prognosis. Despite more than 250 clinical trials aimed to treat children diagnosed with DMG, no curative therapies currently exist for this patient population. A major obstacle has been the intact blood brain barrier (BBB) which prevents most therapeutics from crossing into the tumor bed. Focused Ultrasound (FUS) is an emerging, noninvasive medical technology which has been shown in both preclinical and clinical research to disrupt the blood brain barrier safely and temporarily. FUS blood brain barrier opening has been studied in combination with chemotherapies in preclinical DMG models, and this technology is now being investigated in clinical trials for the treatment of pediatric brain tumors. Focused ultrasound has additional mechanisms of action, including sonodynamic therapy and radiation sensitization, that hold promise as future DMG therapies as well. This paper, largely based off the proceedings from a workshop held by the Focused Ultrasound Foundation in October of 2021, summarizes the current state of the field of focused ultrasound for DIPG/DMG, including preclinical, technical, and clinical summaries in addition to recommended next steps for continued advancement of the game changing technology of Focused Ultrasound.
C1 [Parekh, Kavya] Univ Virginia, Charlottesville, VA USA.
   [Parekh, Kavya; LeBlang, Suzanne; Powlovich, Lauren] Focused Ultrasound Fdn, Charlottesville, VA 22903 USA.
   [Nazarian, Javad] Childrens Natl Hosp, Washington, DC USA.
   [Mueller, Sabine] Univ Calif San Francisco, San Francisco, CA USA.
   [Zacharoulis, Stergios] Bristol Meyers Squibb, New York, NY USA.
   [Zacharoulis, Stergios] Columbia Univ, Med Ctr, Dept Pediat Oncol, New York, NY USA.
   [Hynynen, Kullervo] Sunnybrook Res Inst, Toronto, ON, Canada.
C3 University of Virginia; Children's National Health System; University of
   California System; University of California San Francisco; Columbia
   University; University of Toronto; Sunnybrook Health Science Center;
   Sunnybrook Research Institute
RP Powlovich, L (通讯作者)，Focused Ultrasound Fdn, Charlottesville, VA 22903 USA.
EM
RI Hynynen, Kullervo/AGR-1889-2022
CR Alkins R, 2013, CANCER RES, V73, P1892, DOI 10.1158/0008-5472.CAN-12-2609
   Alli S, 2018, J CONTROL RELEASE, V281, P29, DOI 10.1016/j.jconrel.2018.05.005
   Alonso A, 2015, FRONT NEUROL NEUROSC, V36, P106, DOI 10.1159/000366242
   [Anonymous], VIEW NEUROVASCULAR U
   Bruzek AK, 2020, CLIN CANCER RES, V26, P6266, DOI 10.1158/1078-0432.CCR-20-2066
   Bunevicius A, 2022, J NEURO-ONCOL, V156, P1, DOI 10.1007/s11060-021-03807-6
   Burgess A, 2014, RADIOLOGY, V273, P736, DOI 10.1148/radiol.14140245
   Carvalho D, 2019, COMMUN BIOL, V2, DOI 10.1038/s42003-019-0420-8
   Chen KT, 2021, SCI ADV, V7, DOI 10.1126/sciadv.abd0772
   Cordero FJ, 2017, MOL CANCER RES, V15, P1243, DOI 10.1158/1541-7786.MCR-16-0389
   Czarnota GJ, 2012, P NATL ACAD SCI USA, V109, pE2033, DOI 10.1073/pnas.1200053109
   Deligne C, 2020, FLUIDS BARRIERS CNS, V17, DOI 10.1186/s12987-020-00198-0
   Englander ZK, 2021, SCI REP-UK, V11, DOI 10.1038/s41598-021-85180-y
   Focused Ultrasound Foundation, 2021, CISC VIS NETW IND GL
   Golbourn BJ, 2022, NAT CANCER, V3, P899, DOI 10.1038/s43018-022-00407-9
   Haumann R., 2022, TRANSL MED COMMUN, V7, P8, DOI [10, DOI 10.1186/S41231-022-00115-7]
   Idbaih A, 2019, CLIN CANCER RES, V25, P3793, DOI 10.1158/1078-0432.CCR-18-3643
   Ishida J, 2021, J CONTROL RELEASE, V330, P1034, DOI 10.1016/j.jconrel.2020.11.010
   Joiner JB, 2020, J IMMUNOL, V205, P2327, DOI 10.4049/jimmunol.1901430
   Kast RE, 2020, BRAIN SCI, V10, DOI 10.3390/brainsci10010051
   Kebudi R, 2013, PEDIATR DRUGS, V15, P351, DOI 10.1007/s40272-013-0033-5
   Kobus T, 2016, J CONTROL RELEASE, V238, P281, DOI 10.1016/j.jconrel.2016.08.001
   LANGMOEN IA, 1991, CHILD NERV SYST, V7, P13, DOI 10.1007/BF00263825
   Lee Y, 2011, SEMIN CUTAN MED SURG, V30, P199, DOI 10.1016/j.sder.2011.08.001
   Lin GL, 2018, ACTA NEUROPATHOL COM, V6, DOI 10.1186/s40478-018-0553-x
   Liu HL, 2014, THERANOSTICS, V4, P432, DOI 10.7150/thno.8074
   Liu HL, 2010, RADIOLOGY, V255, P415, DOI 10.1148/radiol.10090699
   Mainprize T, 2019, SCI REP-UK, V9, DOI 10.1038/s41598-018-36340-0
   McDannold N, 2020, SCI REP-UK, V10, DOI 10.1038/s41598-020-65617-6
   McDannold N, 2019, THERANOSTICS, V9, P6284, DOI 10.7150/thno.35892
   Meng Y, 2021, SCI TRANSL MED, V13, DOI 10.1126/scitranslmed.abj4011
   Moriuchi S, 2011, J NEUROSURG, V115, P278, DOI 10.3171/2011.4.JNS102137
   Papachristodoulou A, 2019, J CONTROL RELEASE, V295, P130, DOI 10.1016/j.jconrel.2018.12.009
   Park EJ, 2012, J CONTROL RELEASE, V163, P277, DOI 10.1016/j.jconrel.2012.09.007
   Park J, 2012, J CONTROL RELEASE, V162, P134, DOI 10.1016/j.jconrel.2012.06.012
   Pi ZK, 2019, ANN BIOMED ENG, V47, P549, DOI 10.1007/s10439-018-02141-9
   Pouliopoulos A. N., SCI REP-UK
   Pouliopoulos AN, 2020, ULTRASOUND MED BIOL, V46, P73, DOI 10.1016/j.ultrasmedbio.2019.09.010
   Raspagliesi L, 2021, FRONT ONCOL, V11, DOI 10.3389/fonc.2021.679989
   Roberts JW, 2022, J NEURO-ONCOL, V157, P237, DOI 10.1007/s11060-022-03974-0
   Schwake M, 2014, ACTA NEUROCHIR, V156, P1077, DOI 10.1007/s00701-014-2096-7
   Silveira A.B., NAT GENET
   Stummer W, 2014, ACTA NEUROCHIR, V156, P2315, DOI 10.1007/s00701-014-2234-2
   Timbie KF, 2015, J CONTROL RELEASE, V219, P61, DOI 10.1016/j.jconrel.2015.08.059
   Tran WT, 2012, BRIT J CANCER, V107, P469, DOI 10.1038/bjc.2012.279
   Treat LH, 2007, INT J CANCER, V121, P901, DOI 10.1002/ijc.22732
   Treat LH, 2012, ULTRASOUND MED BIOL, V38, P1716, DOI 10.1016/j.ultrasmedbio.2012.04.015
   Vitanza NA, 2019, CURR TREAT OPTION NE, V21, DOI 10.1007/s11940-019-0577-y
   Wang ST, 2020, BRIT J RADIOL, V93, DOI 10.1259/bjr.20190214
   Wang YX, 2015, P NATL ACAD SCI USA, V112, P9704, DOI 10.1073/pnas.1511694112
   Wei HJ, 2021, INT J RADIAT ONCOL, V110, P539, DOI 10.1016/j.ijrobp.2020.12.019
   White PJ, 2020, ULTRASOUND MED BIOL, V46, P1998, DOI 10.1016/j.ultrasmedbio.2020.04.013
   Wu SK, 2019, SCI REP-UK, V9, DOI 10.1038/s41598-019-46832-2
   Ye DZ, 2021, PHARMACEUTICS, V13, DOI 10.3390/pharmaceutics13020190
   Ye DZ, 2018, J CONTROL RELEASE, V286, P145, DOI 10.1016/j.jconrel.2018.07.020
   Zhang D.Y., CLIN CANCER RES
NR 56
TC 1
Z9 1
U1 3
U2 4
PU ELSEVIER SCIENCE INC
PI NEW YORK
PA STE 800, 230 PARK AVE, NEW YORK, NY 10169 USA
SN 1476-5586
J9 NEOPLASIA
JI Neoplasia
PD MAR
PY 2023
VL 37
AR 100876
DI 10.1016/j.neo.2023.100876
EA JAN 2023
PG 9
WC Oncology
WE Science Citation Index Expanded (SCI-EXPANDED)
SC Oncology
GA 9H2VA
UT WOS:000938693600001
PM 36709715
OA Green Published, gold
DA 2023-08-27
ER

PT J
AU Toguslu, G
   Erdi, MF
   Arac, D
   Keskin, F
   Kilinc, I
   Cuce, G
AF Toguslu, Gokhan
   Erdi, Mehmet Fatih
   Arac, Densel
   Keskin, Fatih
   Kilinc, Ibrahim
   Cuce, Gokhan
TI Ameliorating the effects of Adalimumab on rabbits with experimental
   cerebral vasospasm after subarachnoid hemorrhage
SO ULUSAL TRAVMA VE ACIL CERRAHI DERGISI-TURKISH JOURNAL OF TRAUMA &
   EMERGENCY SURGERY
LA English
DT Article
DE Adalimumab; cerebral vasospasm; cytokine; inflammation; neuroprotection;
   rabbit; subarachnoid hemorrhage
ID NECROSIS-FACTOR-ALPHA; TNF-ALPHA; INDUCED APOPTOSIS; INJURY;
   PATHOPHYSIOLOGY; ASSOCIATION; TRAUMA
AB BACKGROUND: Adalimumab (ADA), which is a new-generation recombinant human monoclonal antibody for tumor necrosis factor a (TNF alpha), has strong anti-inflammatory effects. The role of enhanced inflammation is well established for the development and progression of cerebral vasospasm. Investigated in the present study is the probable ameliorating and neuroprotective effects of ADA in rabbits using a cerebral vasospasm model with biochemical and histopathological methods.
   METHODS: Thirty male New-Zealand white rabbits were randomly divided into control, subarachnoid hemorrhage (SAH) only and SAH plus ADA treatment groups. SAH was established as a single cisterna magna autologous arterial blood injection. ADA treatment was started just after intracisternal blood injection and continued for 72 hours once a day. The animals were sacrificed 72 hours after the induction of SAH, serum and brainstem tissue obtained for investigations.
   RESULTS: Brainstem tissue and plasma levels of tumor necrosis factor-alpha and Interleukin-1 beta, brainstem tissue Matrix metalloproteinase-9 levels increased after SAH and partly decreased after treatment. Plasma levels of brain-derived neurotrophic factor decreased after SAH and partly restored after treatment. ADA treatment significantly increased the mean cross-sectional area of the vasospastic basilar arteries, reduced the basilar artery wall thickness and also ameliorates enhanced endothelial apoptosis.
   CONCLUSION: Findings obtained in this study suggest that ADA is an effective neuroprotective agent for ameliorating cerebral vasospasm in experimental rabbit vasospasm.
C1 [Toguslu, Gokhan] Kadirli State Hosp, Dept Neurosurg, Osmaniye, Turkey.
   [Erdi, Mehmet Fatih; Arac, Densel; Keskin, Fatih] Necmettin Erbakan Univ, Meram Fac Med, Dept Neurosurg, Konya, Turkey.
   [Kilinc, Ibrahim] Necmettin Erbakan Univ, Meram Fac Med, Dept Biochem, Konya, Turkey.
   [Cuce, Gokhan] Necmettin Erbakan Univ, Meram Fac Med, Dept Histol & Embryol, Konya, Turkey.
C3 Osmaniye Kadirli State Hospital; Necmettin Erbakan University; Necmettin
   Erbakan University; Necmettin Erbakan University
RP Arac, D (通讯作者)，Necmettin Erbakan Univ, Meram Fac Med, Dept Neurosurg, Konya, Turkey.
EM
RI Keskin, Fatih/AAF-4142-2021; KILINC, Ibrahim/B-4565-2013; Erdi,
   Fatih/HJG-4686-2022
OI KILINC, Ibrahim/0000-0002-7729-7557; Erdi, Fatih/0000-0003-3621-6658
CR Achrol AS, 2006, STROKE, V37, P231, DOI 10.1161/01.STR.0000195133.98378.4b
   Bavbek M, 1998, STROKE, V29, P1930, DOI 10.1161/01.STR.29.9.1930
   Bowman G, 2004, NEUROSURGERY, V54, P719, DOI 10.1227/01.NEU.0000108981.73153.6E
   Bowman G, 2006, NEUROCRIT CARE, V5, P222, DOI 10.1385/NCC:5:3:222
   Bramlett HM, 2004, J CEREBR BLOOD F MET, V24, P133, DOI 10.1097/01.WCB.0000111614.19196.04
   Buommino E, 2012, ARCH DERMATOL RES, V304, P481, DOI 10.1007/s00403-012-1251-3
   Celik H, 2016, TURK NEUROSURG, V26, P622, DOI 10.5137/1019-5149.JTN.13210-14.1
   Chen SD, 2017, INT J MOL SCI, V18, DOI 10.3390/ijms18030545
   Dang BQ, 2017, CURR DRUG TARGETS, V18, P1358, DOI 10.2174/1389450118666170427151657
   Decourt B, 2017, CURR ALZHEIMER RES, V14, P412, DOI 10.2174/1567205013666160930110551
   Dumont AS, 2003, NEUROSURGERY, V53, P123, DOI 10.1227/01.NEU.0000068863.37133.9E
   Erdi MF, 2011, TURK NEUROSURG, V21, P527, DOI [10.5137/1019-5149.JTN .4431-11.1, 10.5137/1019-5149.JTN.4431-11.1]
   Fassbender K, 2001, J NEUROL NEUROSUR PS, V70, P534, DOI 10.1136/jnnp.70.4.534
   Kolias AG, 2009, J NEUROSCI RES, V87, P1, DOI 10.1002/jnr.21823
   Lin CC, 2008, TOXICOL APPL PHARM, V229, P386, DOI 10.1016/j.taap.2008.01.032
   Mansoor SR, 2018, J MOL NEUROSCI, V65, P246, DOI 10.1007/s12031-018-1092-4
   Petrache I, 2000, AM J PHYSIOL-LUNG C, V278, pL312, DOI 10.1152/ajplung.2000.278.2.L312
   Polat E, 2016, ULUS TRAVMA ACIL CER, V22, P134, DOI 10.5505/tjtes.2015.54358
   SELMAJ K, 1991, ANN NEUROL, V30, P694, DOI 10.1002/ana.410300510
   Sen O, 2006, NEUROL RES, V28, P859, DOI 10.1179/016164106X115099
   SHARIEF MK, 1991, NEW ENGL J MED, V325, P467, DOI 10.1056/NEJM199108153250704
   Siasios I, 2013, NEUROL RES INT, V2013, DOI 10.1155/2013/571328
   Silva Y, 2005, STROKE, V36, P86, DOI 10.1161/01.STR.0000149615.51204.0b
   Singh S, 2018, NEUROSCIENCE, V376, P40, DOI 10.1016/j.neuroscience.2017.12.021
   Takizawa T, 2001, NEUROL RES, V23, P724, DOI 10.1179/016164101101199243
   Tsilimbaris M, 2009, GRAEF ARCH CLIN EXP, V247, P1119, DOI 10.1007/s00417-009-1065-y
   VanAntwerp DJ, 1996, SCIENCE, V274, P787, DOI 10.1126/science.274.5288.787
   Yang G, 2016, NEUROL SCI, V37, P1253, DOI 10.1007/s10072-016-2576-z
NR 28
TC 4
Z9 4
U1 0
U2 2
PU TURKISH ASSOC TRAUMA EMERGENCY SURGERY
PI ISTANBUL
PA KOPRULU MEHMET PASA SOC, DENIZ ABDAL MAH, DADASOGLU AP, 25-1 SEHREMINI,
   ISTANBUL, 00000, TURKEY
SN 1306-696X
J9 ULUS TRAVMA ACIL CER
JI Ulus. Travma Acil Cerrahi Derg.
PD NOV
PY 2020
VL 26
IS 6
BP 847
EP 852
DI 10.14744/tjtes.2019.52504
PG 6
WC Emergency Medicine
WE Science Citation Index Expanded (SCI-EXPANDED)
SC Emergency Medicine
GA OH6NP
UT WOS:000582710000003
PM 33107964
OA hybrid
DA 2023-08-27
ER

PT J
AU Strauss, C
   Neu, M
   Bischoff, B
   Romstock, J
AF Strauss, C
   Neu, M
   Bischoff, B
   Romstock, J
TI Clinical and neurophysiological observations after superior petrosal
   vein obstruction during surgery of the cerebellopontine angle: Case
   report
SO NEUROSURGERY
LA English
DT Article
DE cerebellopontine angle; cochlear nerve; neurophysiological monitoring;
   superior petrosal vein obstruction
ID SCHWANNOMAS ACOUSTIC NEUROMAS; MICROVASCULAR DECOMPRESSION; MANAGEMENT;
   HEARING; NERVE
AB IMPORTANCE: Preservation of venous drainage during surgery of the cerebellopontine angle has received little attention.
   CLINICAL PRESENTATION: We describe changes in brainstem auditory evoked potentials after temporary obstruction of the superior petrosal vein during surgical resection of a small meningioma at the petrous apex via a standard suboccipital-lateral approach. Temporary clipping of the petrosal vein resulted in deterioration of the brainstem auditory evoked potentials. The tumor was removed with preservation of the superior petrosal vein.
   CONCLUSION: A transient postoperative cochlear nerve deficit emphasizes the importance of venous drainage and its preservation during surgery for small lesions of the cerebellopontine angle that do not distort normal anatomic structures.
C1 Univ Erlangen Nurnberg, Dept Neurosurg, D-91054 Erlangen, Germany.
C3 University of Erlangen Nuremberg
RP Strauss, C (通讯作者)，Univ Erlangen Nurnberg, Dept Neurosurg, Schwabachanlage 6, D-91054 Erlangen, Germany.
CR Bebin J, 1968, Henry Ford Hosp Med J, V16, P61
   Fujimaki T, 1999, J NEUROSURG, V90, P1148
   LUDERS H, 1988, J CLIN NEUROPHYSIOL, V5, P261
   MARTIN RG, 1980, NEUROSURGERY, V6, P483, DOI 10.1227/00006123-198005000-00001
   MATSUSHIMA T, 1989, NEUROSURGERY, V24, P890, DOI 10.1227/00006123-198906000-00016
   Matthies C, 1997, NEUROSURGERY, V40, P459, DOI 10.1097/00006123-199703000-00007
   MAZZONI A, 1969, ANN OTO RHINOL LARYN, V78, P797, DOI 10.1177/000348946907800413
   McLaughlin MR, 1999, J NEUROSURG, V90, P1, DOI 10.3171/jns.1999.90.1.0001
   NAGER GT, 1954, ANN OTO RHINOL LARYN, V63, P51, DOI 10.1177/000348945406300104
   Neu M, 1999, CLIN NEUROPHYSIOL, V110, P1935, DOI 10.1016/S1388-2457(99)00148-0
   RHOTON A L JR, 1984, Neurological Research, V6, P3
   Samii M, 1997, NEUROSURGERY, V40, P11, DOI 10.1097/00006123-199701000-00002
   Samii M, 1997, NEUROSURGERY, V40, P248, DOI 10.1097/00006123-199702000-00005
   SEYER H, 1994, ACTA NEUROCHIR, V131, P157, DOI 10.1007/BF01401467
   STRAUSS C, 1991, NEUROSURGERY, V28, P559, DOI 10.1227/00006123-199104000-00012
   Sweet WH, 1995, OPERATIVE NEUROSURGI, P1543
   TSUKAMOTO H, 1993, SURG NEUROL, V40, P31, DOI 10.1016/0090-3019(93)90166-X
   WATANABE E, 1989, ACTA NEUROCHIR, V98, P118, DOI 10.1007/BF01407337
NR 18
TC 17
Z9 19
U1 0
U2 0
PU LIPPINCOTT WILLIAMS & WILKINS
PI PHILADELPHIA
PA 530 WALNUT ST, PHILADELPHIA, PA 19106-3621 USA
SN 0148-396X
J9 NEUROSURGERY
JI Neurosurgery
PD MAY
PY 2001
VL 48
IS 5
BP 1157
EP 1159
DI 10.1097/00006123-200105000-00043
PG 3
WC Clinical Neurology; Surgery
WE Science Citation Index Expanded (SCI-EXPANDED)
SC Neurosciences & Neurology; Surgery
GA 424GB
UT WOS:000168223000104
PM 11334285
DA 2023-08-27
ER

PT J
AU Banik, S
   Lahiri, T
AF Banik, S
   Lahiri, T
TI Increase in brain serotonin level and concomitant reduction in food
   intake and body weight of mice bearing chemically - Induced fibrosarcoma
SO BIOMEDICAL RESEARCH-TOKYO
LA English
DT Article
ID PLATELET SEROTONIN; DOPAMINE; GROWTH; TUMOR; NOREPINEPHRINE;
   NEUROENDOCRINE; DEPRESSION; REGIONS; STRESS; ACID
AB The investigation was carried out to elucidate whether growth of malignant tumor is associated with alterations in central neurotransmitter level of the host. The concentrations of serotonin (5-HT) and its metabolite 5-hydroxyindoleacetic acid (5-HIAA) were measured by HPLC with electrochemical detection in discrete areas of the brain of Swiss mice bearing benzo(a) pyrene - induced primary fibrosarcoma. The level of 5-HT was markedly elevated in raphe nucleus of the brain from 0.86+/-0.15 mug/g tissue in controls to 2.2+/-0.65 mug/g and 3.7+/-0.86 mug/g tissue in early (tumor volume < 1.5 cm(3)) and advanced stage of the disease (tumor volume > 3 cm(3)) respectively. 5-HT values also increased appreciably in midbrain and hypothalamus. Tn contrast, platelet 5-HT level declined significantly (p < 0.05) in tumor bearing animals. Like 5-HT, 5-HIAA level also elevated in discrete areas of the brain. The rise in 5-HT concentration in brain was paralleled by progressive decline in food intake and body weight. In summary, the results demonstrate marked rise in brain 5-HT level in mice bearing chemically-induced fibrosarcoma and this may account for tumor-associated anorexia and cachexia.
C1 Chittaranjan Natl Canc Inst, Dept Neuroendocrinol, Kolkata 700026, W Bengal, India.
RP Lahiri, T (通讯作者)，Chittaranjan Natl Canc Inst, Dept Neuroendocrinol, 37 SP Mukherjee Rd, Kolkata 700026, W Bengal, India.
EM
CR Baile Walter F., 1996, Current Opinion in Oncology, V8, P182
   BAKER DG, 1977, CANCER RES, V37, P3939
   BANIK S, 1994, NEOPLASMA, V41, P9
   Banik S., 1994, Indian Journal of Experimental Biology, V32, P106
   BANIK S, 1994, MED SCI RES, V22, P145
   BEVERLEY PCL, 1993, INTRO CELLULAR MOL B, P406
   Bortolozzi AA, 1998, BIOGENIC AMINES, V14, P667
   BROWN RE, 1994, INTRO NEUROENDOCRINO, P88
   CHANCE WT, 1988, LIFE SCI, V43, P67, DOI 10.1016/0024-3205(88)90238-X
   DASGUPTA PS, 1992, NEOPLASMA, V39, P163
   DASGUPTA PS, 1991, INDIAN J EXP BIOL, V29, P86
   DONOHOE TP, 1984, LIFE SCI, V34, P203, DOI 10.1016/0024-3205(84)90592-7
   GLOWINSKI J, 1966, J NEUROCHEM, V13, P655, DOI 10.1111/j.1471-4159.1966.tb09873.x
   Jaeckle Kurt A., 1996, Current Opinion in Oncology, V8, P204
   KALISKI MJ, 1991, ENCY HUMAN BIOL, P829
   KAPLAN RD, 1982, LIFE SCI, V31, P583, DOI 10.1016/0024-3205(82)90488-X
   KIM C, 1987, J CHROMATOGR, V386, P25
   LALA P, 1971, METHOD CANCER RES, P4
   LECHIN F, 1990, PSYCHONEUROENDOCRINO, V15, P435, DOI 10.1016/0306-4530(90)90067-J
   MAINWARING WIP, 1993, INTRO CELLULAR MOL B, P357
   MANNUCCI PM, 1989, EUR J CANCER CLIN ON, V25, P1413, DOI 10.1016/0277-5379(89)90098-9
   Mendelson J, 1994, HARRISONS PRINCIPLE, V2, P1814
   MULLER EE, 1995, ENDOCRINOLOGY, P151
   NICHOLS M, 1983, LIFE SCI, V32, P1819, DOI 10.1016/0024-3205(83)90059-0
   PRADA MD, 1988, EXPERIENTIA, V44, P115
   RAY MR, 1994, BIOGENIC AMINES, V10, P555
   REICHLIN S, 1995, ENDOCRINOLOGY, P2964
   REINHARD JF, 1977, LIFE SCI, V21, P1741, DOI 10.1016/0024-3205(77)90153-9
   ROSZMAN TL, 1988, PROG ALLERGY, V43, P140
   SETH PK, 1995, CURR SCI INDIA, V68, P204
   SHERWOOD IM, 1995, ENDOCRINOLOGY, P2754
   TAKADA A, 1995, BIOGENIC AMINES, V11, P363
   WEIGENT DA, 1990, ANN NY ACAD SCI, V579, P17
NR 33
TC 4
Z9 4
U1 0
U2 1
PU BIOMEDICAL RESEARCH PRESS LTD
PI SAPPORO
PA HOKKAIDO UNIV, LAB HISTOLOGY & CYTOLOGY, C/O TOSHIHIKO IWANAGA, KITA
   15-NISHI 7, SAPPORO, 060-8638, JAPAN
SN 0388-6107
EI 1880-313X
J9 BIOMED RES-TOKYO
JI Biomed. Res.
PD OCT
PY 2000
VL 21
IS 5
BP 255
EP 261
PG 7
WC Medicine, Research & Experimental
WE Science Citation Index Expanded (SCI-EXPANDED)
SC Research & Experimental Medicine
GA 413JK
UT WOS:000167609100003
OA Bronze
DA 2023-08-27
ER

PT J
AU Zhang, DD
   Zhang, CY
   Zhang, YX
   Cui, HP
   Chen, J
   Ma, WZ
   Jia, H
AF Zhang, Dan-Dan
   Zhang, Cheng-Yun
   Zhang, Yu-Xin
   Cui, Hai-Peng
   Chen, Jiao
   Ma, Wen-Zhi
   Jia, Hua
TI G-CSF reduces loss of dopaminergic neurons by inhibiting TNF-alpha and
   IL-1 beta in mouse model of Parkinson's disease
SO INTERNATIONAL JOURNAL OF NEUROSCIENCE
LA English
DT Article
DE Parkinson's disease (PD); 1-methyl-4-phenyl-1; 2; 3;
   6-tetrahydropyridine (MPTP); granulocyte-colony stimulating
   factor(G-CSF); tyrosine hydroxylase (TH); inflammatory factor
ID COLONY-STIMULATING FACTOR; MICROGLIAL ACTIVATION; FACTOR PROTECTS;
   CELL-DEATH; MPTP MODEL; INFLAMMATION; BRAIN; NEUROINFLAMMATION;
   NEURODEGENERATION; MODULATION
AB Purpose of the study granulocyte-colony stimulating factor (G-CSF) is a hematopoietic growth factor existing in neutrophils, glial cells and neurons. Increasing researches discovered that G-CSF improved cell survival in neurodegenerative diseases by its anti-inflammatory effect. However, the effect of G-CSF in suppressing inflammation in Parkinson's disease (PD) remains unclear. Thus, the purpose of this study is to explored the anti-inflammatory effect of G-CSF in mouse model of PD. Materials and methods G-CSF was administrated in the PD model induced by MPTP. Subsequently, the protein of tyrosine hydroxylase (TH), ionized calcium-binding adaptor molecule 1 (Iba-1) and the inflammatory cytokines including tumor necrosis factor-alpha (TNF-alpha) and interleukin-1 beta (IL-1 beta) in the midbrain were examined. In addition, the phosphorylated mitogen-activated protein kinases (MAPK) including c-Jun NH2-terminal kinase (JNK), extracellular signal-regulated kinase (ERK) and p38 MAPK in the midbrain were investigated. Results Compared with the MPTP group, the protein of TH in the midbrain was increased, while the Iba-1 and the inflammatory factors were decreased. In addition, the expression of phosphorylated JNK (p-JNK) in the midbrain of the MPTP + G-CSF group was decreased, while the phosphorylated ERK (p-ERK) levels were elevated. Conclusions These findings emphasize that G-CSF inhibited the degradation of DA neurons. The protective effect is associated with the reduction of the inflammatory factors caused by the inhibition of the microglial activation. Moreover, G-CSF may decrease the inflammatory factors through the decrease of P-JNK and the increase of P-ERK.
C1 [Zhang, Dan-Dan; Chen, Jiao; Ma, Wen-Zhi; Jia, Hua] Ningxia Med Univ, Sch Basic Med Sci, Dept Human Anat & Histoembryol, 1160 Shengli St, Yinchuan 750004, Ningxia, Peoples R China.
   [Zhang, Dan-Dan] Chengde Med Univ, Sch Basic Med Sci, Dept Physiol, Chengde, Peoples R China.
   [Zhang, Cheng-Yun] Chengde Med Univ, Sch Basic Med Sci, Dept Human Anat, Chengde, Peoples R China.
   [Zhang, Yu-Xin] North China Univ Sci & Technol, Sch Basic Med Sci, Dept Human Anat, Tangshan, Peoples R China.
   [Cui, Hai-Peng] Chengde Med Univ, Sch Basic Med Sci, Dept Pathophysiol, Chengde, Peoples R China.
   [Ma, Wen-Zhi] Ningxia Med Univ, Minist Educ, Key Lab Reprod & Genet Ningxia Hui Autonomous Reg, Key Lab Fertil Preservat & Maintenance, Yinchuan, Ningxia, Peoples R China.
   [Ma, Wen-Zhi; Jia, Hua] Penn State Univ, Ctr Reprod Biol & Hlth, Sch Agr Sci, University Pk, PA USA.
C3 Ningxia Medical University; Chengde Medical University; Chengde Medical
   University; North China University of Science & Technology; Chengde
   Medical University; Ningxia Medical University; Pennsylvania
   Commonwealth System of Higher Education (PCSHE); Pennsylvania State
   University; Pennsylvania State University - University Park
RP Ma, WZ; Jia, H (通讯作者)，Ningxia Med Univ, Sch Basic Med Sci, Dept Human Anat & Histoembryol, 1160 Shengli St, Yinchuan 750004, Ningxia, Peoples R China.
EM
FU Natural Science Foundation of Ningxia Province [2018AAC03067]; National
   Natural Science Foundation of China [81260193]; Ningxia High School
   First-class Disciplines (West China First-class Disciplines Basic
   Medical Sciences at Ningxia Medical University) [NXYLXK2017B07]
FX The work was supported by the Natural Science Foundation of Ningxia
   Province under Grant [2018AAC03067]; the National Natural Science
   Foundation of China under Grant [81260193]; and Ningxia High School
   First-class Disciplines (West China First-class Disciplines Basic
   Medical Sciences at Ningxia Medical University) under Grant
   [NXYLXK2017B07].
CR Block ML, 2005, PROG NEUROBIOL, V76, P77, DOI 10.1016/j.pneurobio.2005.06.004
   Bohush A, 2018, INT J MOL SCI, V19, DOI 10.3390/ijms19102973
   Cao XQ, 2006, J NEUROCHEM, V99, P861, DOI 10.1111/j.1471-4159.2006.04125.x
   Chamoli M, 2018, J NEURAL TRANSM, V125, P1651, DOI 10.1007/s00702-018-1887-z
   Chen X, 2018, FRONT IMMUNOL, V9, DOI 10.3389/fimmu.2018.02122
   Cui WY, 2010, J NEUROIMMUNE PHARM, V5, P479, DOI 10.1007/s11481-010-9210-2
   Dauer W, 2003, NEURON, V39, P889, DOI 10.1016/S0896-6273(03)00568-3
   Donat CK, 2017, FRONT AGING NEUROSCI, V9, DOI 10.3389/fnagi.2017.00208
   Garcia-Dominguez I, 2018, FRONT CELL NEUROSCI, V12, DOI 10.3389/fncel.2018.00398
   Gibson CL, 2005, J CEREBR BLOOD F MET, V25, P431, DOI 10.1038/sj.jcbfm.9600033
   Hashemzaei M, 2016, DRUG CHEM TOXICOL, V39, P375, DOI 10.3109/01480545.2015.1123267
   Hirsch EC, 1998, ANN NEUROL, V44, pS115, DOI 10.1002/ana.410440717
   Hirsch EC, 2003, ANN NY ACAD SCI, V991, P214
   Huang HY, 2007, NEUROBIOL AGING, V28, P1258, DOI 10.1016/j.neurobiolaging.2006.05.037
   Jiang H, 2010, J INT MED RES, V38, P1305, DOI 10.1177/147323001003800412
   Karpenko MN, 2018, CELL IMMUNOL, V327, P77, DOI 10.1016/j.cellimm.2018.02.011
   Kempuraj D, 2016, J Neurol Neurosurg Spine, V1
   Khan AU, 2019, INT J NEUROSCI, V129, P55, DOI 10.1080/00207454.2018.1486837
   Kim EK, 2015, ARCH TOXICOL, V89, P867, DOI 10.1007/s00204-015-1472-2
   Kim SO, 2006, CELL MICROBIOL, V8, P1958, DOI 10.1111/j.1462-5822.2006.00763.x
   Lee ST, 2008, BRAIN RES, V1194, P130, DOI 10.1016/j.brainres.2007.11.058
   Lofrumento DD, 2011, NEUROIMMUNOMODULAT, V18, P79, DOI 10.1159/000320027
   Luchtman DW, 2009, PHYSIOL BEHAV, V98, P130, DOI 10.1016/j.physbeh.2009.04.021
   Lull ME, 2010, NEUROTHERAPEUTICS, V7, P354, DOI 10.1016/j.nurt.2010.05.014
   Maiolo SA, 2018, PROG NEUROBIOL
   MCGEER PL, 1988, NEUROLOGY, V38, P1285, DOI 10.1212/WNL.38.8.1285
   Menzie-Suderam JM, 2018, BRAIN RES, V1682, P1, DOI 10.1016/j.brainres.2017.12.022
   Meredith GE, 2011, J PARKINSON DIS, V1, P19, DOI 10.3233/JPD-2011-11023
   MOGI M, 1994, NEUROSCI LETT, V180, P147, DOI 10.1016/0304-3940(94)90508-8
   MOGI M, 1994, NEUROSCI LETT, V165, P208, DOI 10.1016/0304-3940(94)90746-3
   Munoz-Manchado AB, 2016, J NEUROCHEM, V136, P373, DOI 10.1111/jnc.13409
   Ouchi Y, 2005, ANN NEUROL, V57, P168, DOI 10.1002/ana.20338
   Pain S, 2013, EXP TOXICOL PATHOL, V65, P689, DOI 10.1016/j.etp.2012.09.001
   Pollari E, 2011, J NEUROINFLAMM, V8, DOI 10.1186/1742-2094-8-74
   Rai SN, 2020, J CHEM NEUROANAT, V104, DOI 10.1016/j.jchemneu.2020.101752
   Ren B, 2015, J NEUROL SCI, V348, P142, DOI 10.1016/j.jns.2014.11.026
   Schabitz WR, 2003, STROKE, V34, P745, DOI 10.1161/01.STR.0000057814.70180.17
   Schneider A, 2005, J CLIN INVEST, V115, P2083, DOI 10.1172/JCI23559
   Su JJ, 2015, PLOS ONE, V10, DOI 10.1371/journal.pone.0120707
   Tejera D, 2019, METHODS MOL BIOL, V2034, P57, DOI 10.1007/978-1-4939-9658-2_5
   Urdzikova L, 2006, J NEUROTRAUM, V23, P1379, DOI 10.1089/neu.2006.23.1379
   Varadi C, 2020, BIOLOGY-BASEL, V9, DOI 10.3390/biology9050103
   Vila M, 2001, P NATL ACAD SCI USA, V98, P2837, DOI 10.1073/pnas.051633998
   Wu DC, 2002, J NEUROSCI, V22, P1763, DOI 10.1523/JNEUROSCI.22-05-01763.2002
   Xu L, 2016, MOL NEUROBIOL, V53, P6709, DOI 10.1007/s12035-015-9593-4
   Zhao LR, 2007, EXP NEUROL, V204, P569, DOI 10.1016/j.expneurol.2006.12.001
NR 46
TC 0
Z9 0
U1 1
U2 4
PU TAYLOR & FRANCIS LTD
PI ABINGDON
PA 2-4 PARK SQUARE, MILTON PARK, ABINGDON OR14 4RN, OXON, ENGLAND
SN 0020-7454
EI 1563-5279
J9 INT J NEUROSCI
JI Int. J. Neurosci.
PD MAR 4
PY 2023
VL 133
IS 3
BP 278
EP 289
DI 10.1080/00207454.2021.1910259
EA MAR 2021
PG 12
WC Neurosciences
WE Science Citation Index Expanded (SCI-EXPANDED)
SC Neurosciences & Neurology
GA G5BY9
UT WOS:000658579200001
PM 33781148
DA 2023-08-27
ER

PT J
AU Tan, B
   Chen, J
   Qin, SY
   Liao, CY
   Zhang, Y
   Wang, D
   Li, SQ
   Zhang, ZJ
   Zhang, P
   Xu, FG
AF Tan, Bei
   Chen, Jie
   Qin, Siyuan
   Liao, Chuyao
   Zhang, Ying
   Wang, Di
   Li, Siqi
   Zhang, Zunjian
   Zhang, Pei
   Xu, Fengguo
TI Tryptophan Pathway-Targeted Metabolomics Study on the Mechanism and
   Intervention of Cisplatin-Induced Acute Kidney Injury in Rats
SO CHEMICAL RESEARCH IN TOXICOLOGY
LA English
DT Article
ID INDOXYL SULFATE; INDUCED NEPHROTOXICITY; 3-INDOXYL SULFATE; INDUCED
   APOPTOSIS; CANCER CELLS; CHEMOTHERAPY; TIME; QUANTIFICATION; INHIBITION;
   ACTIVATION
AB Cisplatin is a chemotherapeutic agent widely employed in the treatment of various solid tumors. However, its use is often restricted by acute kidney injury (AKI) which is the dose-limiting adverse effect of cisplatin. While numerous studies aiming to alleviate the AKI have been conducted, there are no effective remedies in clinical practice. In this paper, a targeted metabolomics study was performed to reveal the potential relationship between tryptophan metabolism and cisplatin-induced AKI. A chemical derivatization integrated liquid chromatography coupled tandem mass spectrometry (LC-MS/MS) approach was utilized to quantify 29 metabolites in the tryptophan pathway in rat kidney medulla and cortex after cisplatin administration. Results showed that tryptophan metabolism was remarkably disturbed both in the medulla and cortex after cisplatin administration. We also found that the tryptophan pathway in the medulla was more sensitive to cisplatin exposure compared with the cortex. Among these metabolites, indoxyl sulfate was focused for further study because it accumulated most significantly in the kidney cortex and medulla in a dose-dependent manner. A function verification study proved that chlormethiazole, a widely used CYP2E1 inhibitor, could reduce the production of indoxyl sulfate in the liver and attenuate cisplatin-induced AKI in rats. In conclusion, our study depicted the tryptophan pathway in cisplatin-induced AKI for the first time and demonstrated tryptophan metabolism is closely associated with the renal toxicity caused by cisplatin, which can be of great use for the discovery of renal toxicity attenuating remedies.
C1 [Tan, Bei; Chen, Jie; Qin, Siyuan; Liao, Chuyao; Zhang, Ying; Wang, Di; Li, Siqi; Zhang, Zunjian; Zhang, Pei; Xu, Fengguo] China Pharmaceut Univ, Key Lab Drug Qual Control & Pharmacovigilance, Minist Educ, State Key Lab Nat Med, Nanjing 210009, Peoples R China.
C3 China Pharmaceutical University
RP Zhang, P; Xu, FG (通讯作者)，China Pharmaceut Univ, Key Lab Drug Qual Control & Pharmacovigilance, Minist Educ, State Key Lab Nat Med, Nanjing 210009, Peoples R China.
EM
OI Chen, Jie/0000-0002-6360-1765
FU NSFC [82073812, 81773682, 81773861]; Jiangsu Provincial National Science
   Foundation for Distinguished Young Scholars [BK20180027]; National
   Science and Technology Major Project [2017ZX09101001]; Fundamental
   Research Funds for the Central Universities [2632021PY03]; Double
   First-Class University project; Program for Jiangsu province Innovative
   Research Team; Priority Academic Program Development of Jiangsu Higher
   Education Institutions (PAPD)
FX This work was supported by the NSFC (nos. 82073812, 81773682, and
   81773861), Jiangsu Provincial National Science Foundation for
   Distinguished Young Scholars (no. BK20180027), National Science and
   Technology Major Project (2017ZX09101001), the Fundamental Research
   Funds for the Central Universities (2632021PY03), Double First-Class
   University project, the Program for Jiangsu province Innovative Research
   Team, and a project funded by the Priority Academic Program Development
   of Jiangsu Higher Education Institutions (PAPD).
CR Agus A, 2018, CELL HOST MICROBE, V23, P716, DOI 10.1016/j.chom.2018.05.003
   Arany I, 2003, SEMIN NEPHROL, V23, P460, DOI 10.1016/S0270-9295(03)00089-5
   Banoglu E, 2002, EUR J DRUG METAB PH, V27, P135, DOI 10.1007/BF03190428
   Chaudhary UB, 2003, DRUGS, V63, P1565, DOI 10.2165/00003495-200363150-00004
   CHEVREAU N, 1995, NUTR CANCER, V23, P205, DOI 10.1080/01635589509514375
   Choi BY, 2017, BMC COMPLEM ALTERN M, V17, DOI 10.1186/s12906-017-1776-2
   Dasari S, 2014, EUR J PHARMACOL, V740, P364, DOI 10.1016/j.ejphar.2014.07.025
   Devlin AS, 2016, CELL HOST MICROBE, V20, P709, DOI 10.1016/j.chom.2016.10.021
   Duarte IF, 2013, MOL PHARMACEUT, V10, P4242, DOI 10.1021/mp400335k
   Gao H, 2019, CHEM-BIOL INTERACT, V297, P130, DOI 10.1016/j.cbi.2018.11.008
   Gillam EMJ, 2000, BIOCHEMISTRY-US, V39, P13817, DOI 10.1021/bi001229u
   GREGG RW, 1992, J CLIN ONCOL, V10, P795, DOI 10.1200/JCO.1992.10.5.795
   dos Santos NAG, 2012, ARCH TOXICOL, V86, P1233, DOI 10.1007/s00204-012-0821-7
   Guo HM, 2017, J CHROMATOGR A, V1504, P83, DOI 10.1016/j.chroma.2017.05.008
   Huang YZ, 2019, ANAL CHIM ACTA, V1063, P99, DOI 10.1016/j.aca.2019.02.001
   Ismaili N, 2011, J HEMATOL ONCOL, V4, DOI 10.1186/1756-8722-4-35
   Jiang RQ, 2017, ANAL CHEM, V89, P12223, DOI 10.1021/acs.analchem.7b03020
   Jiang YL, 2017, ONCOL REP, V38, P3668, DOI 10.3892/or.2017.6035
   Kaluzna-Czaplinska J, 2019, CRIT REV FOOD SCI, V59, P72, DOI 10.1080/10408398.2017.1357534
   Kim HS, 2017, TOXICOL LETT, V280, pS238, DOI 10.1016/j.toxlet.2017.07.659
   Kusumoto M, 2011, CLIN EXP NEPHROL, V15, P820, DOI 10.1007/s10157-011-0524-z
   Li HR, 2017, BIOMED PHARMACOTHER, V86, P324, DOI 10.1016/j.biopha.2016.12.036
   Li YB, 2014, RSC ADV, V4, P8260, DOI 10.1039/c3ra46920d
   Lucas D, 2013, NAT MED, V19, P695, DOI 10.1038/nm.3155
   Manohar S, 2018, J NEPHROL, V31, P15, DOI 10.1007/s40620-017-0392-z
   NIWA T, 1994, J LAB CLIN MED, V124, P96
   Pabla N, 2008, KIDNEY INT, V73, P994, DOI 10.1038/sj.ki.5002786
   Park JS, 2019, KOREAN J INTERN MED, V34, P146, DOI 10.3904/kjim.2016.298
   Perazella MA, 2012, CLIN J AM SOC NEPHRO, V7, P1713, DOI 10.2215/CJN.02780312
   Qu XY, 2020, TOXICOLOGY, V431, DOI 10.1016/j.tox.2020.152366
   RIES F, 1986, AM J KIDNEY DIS, V8, P368, DOI 10.1016/S0272-6386(86)80112-3
   Roager HM, 2018, NAT COMMUN, V9, DOI 10.1038/s41467-018-05470-4
   Rothhammer V, 2016, NAT MED, V22, P586, DOI 10.1038/nm.4106
   Schulman G, 2016, BMC NEPHROL, V17, DOI 10.1186/s12882-016-0357-9
   Shimada Y, 2013, PLOS ONE, V8, DOI 10.1371/journal.pone.0080604
   Vanholder R, 2014, J AM SOC NEPHROL, V25, P1897, DOI 10.1681/ASN.2013101062
   Volarevic V, 2019, J BIOMED SCI, V26, DOI 10.1186/s12929-019-0518-9
   Wang D, 2005, NAT REV DRUG DISCOV, V4, P307, DOI 10.1038/nrd1691
   Wang GJ, 2019, DRUG CHEM TOXICOL, V42, P600, DOI 10.1080/01480545.2018.1455204
   Wang YY, 2020, GUT MICROBES, V12, P1, DOI 10.1080/19490976.2020.1823800
   Wang Z, 2014, KIDNEY INT, V86, P338, DOI 10.1038/ki.2014.65
   Xu L, 2019, CHEM RES TOXICOL, V32, P2411, DOI 10.1021/acs.chemrestox.9b00180
   Zarogoulidis K, 2013, J THORAC DIS, V5, pS389, DOI 10.3978/j.issn.2072-1439.2013.07.10
   Zgoda-Pols JR, 2011, TOXICOL APPL PHARM, V255, P48, DOI 10.1016/j.taap.2011.05.015
   Zhang P., 2017, SCI REP-UK, V7, P44504
   Zhang P, 2016, CHEM RES TOXICOL, V29, P776, DOI 10.1021/acs.chemrestox.5b00519
   Zhang X, 2018, EUR J PHARMACOL, V834, P288, DOI 10.1016/j.ejphar.2018.07.044
NR 47
TC 7
Z9 8
U1 3
U2 20
PU AMER CHEMICAL SOC
PI WASHINGTON
PA 1155 16TH ST, NW, WASHINGTON, DC 20036 USA
SN 0893-228X
EI 1520-5010
J9 CHEM RES TOXICOL
JI Chem. Res. Toxicol.
PD JUL 19
PY 2021
VL 34
IS 7
BP 1759
EP 1768
DI 10.1021/acs.chemrestox.1c00110
EA JUN 2021
PG 10
WC Chemistry, Medicinal; Chemistry, Multidisciplinary; Toxicology
WE Science Citation Index Expanded (SCI-EXPANDED)
SC Pharmacology & Pharmacy; Chemistry; Toxicology
GA TM3XL
UT WOS:000675485500010
PM 34110802
DA 2023-08-27
ER

PT J
AU Ishola, IO
   Akinyede, AA
   Adeluwa, TP
   Micah, C
AF Ishola, Ismail O.
   Akinyede, A. A.
   Adeluwa, T. P.
   Micah, C.
TI Novel action of vinpocetine in the prevention of paraquat-induced
   parkinsonism in mice: involvement of oxidative stress and
   neuroinflammation
SO METABOLIC BRAIN DISEASE
LA English
DT Article
DE Neuroinflammation; Bar test; Memory impairment; Tumour necrotic
   factor-alpha; Catalepsy; Malodialdehyde
ID HERBICIDE PARAQUAT; DOPAMINERGIC-NEURONS; DISEASE; RATS; MECHANISM;
   CATALEPSY; TOXICITY; EXPOSURE; MODEL
AB Parkinson's disease (PD) is a multifactorial chronic progressive neurodegenerative disease caused by age, genetic and environmental factors such as paraquat (PQT). PQT (a quartenary nitrogen herbicide) is implicated in some form of idiopathic PD. This study sought to investigate the protective effect of vinpocetine on paraquat-induced Parkinsonism in mice. Forty-eight male albino mice were randomly divided into 6 groups and treated orally as follows for 21 days; Group 1: vehicle normal (10 ml/kg), group 2: vehicle control (10 ml/kg); groups 3-5: vinpocetine (5, 10 or 20 mg/kg); group 6: vinpocetine (20 mg/kg). Animals in groups 2-5 were given PQT (10 mg/kg, i.p.) every 3 days for 3 weeks. The effect of treatments on spontaneous motor activity (open field test), muscle coordination (rotarod tests), cataleptic behaviour (bar test), and working memory (Y-maze test) were assayed. After the behavioural assay on day 21, the midbrain was isolated for estimation of oxidative stress and TNF-alpha. Intraperitoneal injection of paraquat significantly induced motor deficits, muscle incoordination, catalepsy and working memory impairment which was ameliorated by the pretreatment of mice with vinpocetine. In addition, paraquat injection caused marked increase in nitroso-oxidative stress markers with concomitant deficits in antioxidant enzymes activities (GSH and SOD) as well as induction of tumour necrotic factor-alpha (TNF-alpha) in the mid-brain which were attenuated by the pretreatment of mice with vinpocetine. Findings from this study showed that vinpocetine prevented paraquat-induced motor deficits, memory impairment, oxidative stress and neuroinflammation through enhancement of antioxidant defense system and inhibition of neuroinflammatory cytokine. Thus, could be a potential drug in the management of Parkinsonism.
C1 [Ishola, Ismail O.; Akinyede, A. A.; Adeluwa, T. P.; Micah, C.] Univ Lagos, Dept Pharmacol Therapeut & Toxicol, Fac Basic Med Sci, Coll Med, PMB 12003, Lagos, Nigeria.
C3 University of Lagos
RP Ishola, IO (通讯作者)，Univ Lagos, Dept Pharmacol Therapeut & Toxicol, Fac Basic Med Sci, Coll Med, PMB 12003, Lagos, Nigeria.
EM
RI Ishola, Ismaila/T-5394-2019; Iishola, I/AAQ-8007-2020
OI Ishola, Ismaila/0000-0001-9475-6754; 
CR Adedeji HA, 2014, PROG NEURO-PSYCHOPH, V48, P245, DOI 10.1016/j.pnpbp.2013.10.014
   Berry C, 2010, CELL DEATH DIFFER, V17, P1115, DOI 10.1038/cdd.2009.217
   Blanco-Ayala T, 2014, FREE RADICAL RES, V48, P623, DOI 10.3109/10715762.2014.899694
   Braak H, 2003, NEUROBIOL AGING, V24, P197, DOI 10.1016/S0197-4580(02)00065-9
   Brooks AI, 1999, BRAIN RES, V823, P1, DOI 10.1016/S0006-8993(98)01192-5
   Brown RE, 1999, BEHAV GENET, V29, P263, DOI 10.1023/A:1021694307672
   Buege J A, 1978, Methods Enzymol, V52, P302
   Cai YJ, 2013, BIOCHEM BIOPH RES CO, V434, P439, DOI 10.1016/j.bbrc.2013.03.092
   Cai YJ, 2012, J PHARMACOL EXP THER, V343, P479, DOI 10.1124/jpet.112.195446
   Chen Q, 2010, NEUROTOXICOLOGY, V31, P310, DOI 10.1016/j.neuro.2010.02.006
   Colnat-Coulbois S, 2005, J NEUROL NEUROSUR PS, V76, P780, DOI 10.1136/jnnp.2004.047829
   Corti O, 2005, CR BIOL, V328, P131, DOI 10.1016/j.crvi.2004.10.009
   Costello S, 2009, AM J EPIDEMIOL, V169, P919, DOI 10.1093/aje/kwp006
   Day BJ, 1999, P NATL ACAD SCI USA, V96, P12760, DOI 10.1073/pnas.96.22.12760
   Fahim MA, 2013, PHYSIOL RES, V62, P313, DOI 10.33549/physiolres.932386
   HOFFMAN DC, 1995, PSYCHOPHARMACOLOGY, V120, P128, DOI 10.1007/BF02246184
   Ishola IO, 2013, J ETHNOPHARMACOL, V146, P440, DOI 10.1016/j.jep.2012.12.015
   Ishola IO, 2017, METAB BRAIN DIS, V32, P235, DOI 10.1007/s11011-016-9902-2
   Jeon KI, 2010, P NATL ACAD SCI USA, V107, P9795, DOI 10.1073/pnas.0914414107
   Kuter K, 2007, BRAIN RES, V1155, P196, DOI 10.1016/j.brainres.2007.04.018
   LOWRY OH, 1951, J BIOL CHEM, V193, P265
   Manning-Bog AB, 2002, J BIOL CHEM, V277, P1641, DOI 10.1074/jbc.C100560200
   McCormack AL, 2002, NEUROBIOL DIS, V10, P119, DOI 10.1006/nbdi.2002.0507
   Miranda-Contreras L, 2005, DEV BRAIN RES, V160, P19, DOI 10.1016/j.devbrainres.2005.08.001
   Mitra S, 2011, J NEUROINFLAMM, V8, DOI 10.1186/1742-2094-8-163
   Molnar Peter, 1994, Neurobiology (Budapest), V2, P255
   Ogunrin AO, 2014, ANN MED HEALTH SCI R, V4, P654, DOI 10.4103/2141-9248.139368
   Peng J, 2004, J BIOL CHEM, V279, P32626, DOI 10.1074/jbc.M404596200
   SEDLAK J, 1968, ANAL BIOCHEM, V25, P192, DOI 10.1016/0003-2697(68)90092-4
   Steece-Collier K, 2002, P NATL ACAD SCI USA, V99, P13972, DOI 10.1073/pnas.242594999
   Suganya SN, 2017, METAB BRAIN DIS, V32, P471, DOI 10.1007/s11011-016-9929-4
   Sun Yong-Hong, 2011, Zhonghua Lao Dong Wei Sheng Zhi Ye Bing Za Zhi, V29, P437
   Tweedie D, 2007, CURR ALZHEIMER RES, V4, P378, DOI 10.2174/156720507781788873
   Vaccari C, 2017, SYST REV-LONDON, V6, DOI 10.1186/s13643-017-0491-x
   Zaitone SA, 2012, INDIAN J PHARMACOL, V44, P774, DOI 10.4103/0253-7613.103300
   Zhang YS, 2018, EUR J PHARMACOL, V819, P30, DOI 10.1016/j.ejphar.2017.11.041
NR 36
TC 23
Z9 23
U1 1
U2 7
PU SPRINGER/PLENUM PUBLISHERS
PI NEW YORK
PA 233 SPRING ST, NEW YORK, NY 10013 USA
SN 0885-7490
EI 1573-7365
J9 METAB BRAIN DIS
JI Metab. Brain Dis.
PD OCT
PY 2018
VL 33
IS 5
BP 1493
EP 1500
DI 10.1007/s11011-018-0256-9
PG 8
WC Endocrinology & Metabolism; Neurosciences
WE Science Citation Index Expanded (SCI-EXPANDED)
SC Endocrinology & Metabolism; Neurosciences & Neurology
GA GU3DD
UT WOS:000445154200012
PM 29855979
DA 2023-08-27
ER

PT S
AU Tischler, AS
   Kimura, N
   Mcnicol, AM
AF Tischler, Arthur S.
   Kimura, Noriko
   Mcnicol, Anne Marie
BE Pacak, K
   Eisenhofer, G
TI Pathology of pheochromocytoma and extra-adrenal paraganglioma
SO PHEOCHROMOCYTOMA
SE Annals of the New York Academy of Sciences
LA English
DT Article; Proceedings Paper
CT 1st International Symposium on Pheochromocytoma
CY OCT 20-23, 2005
CL Bethesda, MD
SP NIH
DE pheochromocytoma; paraganglioma; pathology; histology; malignancy;
   reporting
ID SPORADIC PHEOCHROMOCYTOMAS; SURGICAL-PATHOLOGY; ANTIGEN RETRIEVAL;
   ADRENAL-MEDULLA; EPINEPHRINE; BENIGN; RECOMMENDATIONS; NOREPINEPHRINE;
   CHROMOGRANIN; ASSOCIATION
AB The 2004 WHO classification of endocrine tumors defines pheochromocytoma as a tumor arising from chromaffin cells in the adrenal medulla. Closely related tumors in extra-adrenal sympathetic and parasympathetic paraganglia are classified as extra-adrenal paragangliomas. A pheochromocytoma is an intra-adrenal sympathetic paraganglioma. While arbitrary, this nomenclature serves to emphasize important distinctive properties of intra-adrenal tumors that must be taken into account in clinical practice and research. Those include an often adrenergic phenotype, a relatively low rate of malignancy, and a predilection to occur in particular hereditary syndromes. Current roles of pathology are limited to distinguishing primary or metastatic pheochromocytomas/paragangliomas from other endocrine or nonendocrine tumors, and flagging tumors that show features suggestive of malignant potential or syndromic disease. Future roles may involve more definitive assessment of malignancy, genotype-phenotype correlation, and identification of targets for therapy. Pathology practice currently rests mostly on interpretation of conventional histological sections stained with hematoxylin and eosin, with variable ancillary application of immunohistochemical staining. Malignancy is currently defined by the presence of metastases, not local invasion. Local invasion alone is a poor predictor of metastases, and the absence of apparent invasion does not preclude development of metastases. The two types of aggressive behavior might therefore have different biological underpinnings, and those will be resolved most effectively if consistent terminology is employed. In order to be optimally informative, pathology reports must employ consistent nomenclature and incorporate standard elements. Templates or checklists for minimal standard reporting are recommended by several pathology associations, but identification of some recommended and optional elements is currently subjective or inconsistent.
C1 Tufts Univ, New England Med Ctr, Dept Pathol, Sch Med, Boston, MA 02111 USA.
   Tohoku Rosai Hosp, Dept Pathol, Sendai, Miyagi 9818563, Japan.
   Univ Glasgow, Dept Pathol, Glasgow G4 0SF, Lanark, Scotland.
C3 Tufts Medical Center; Tufts University; University of Glasgow
RP Tischler, AS (通讯作者)，Tufts Univ, New England Med Ctr, Dept Pathol, Sch Med, Box 802,750 Washington St, Boston, MA 02111 USA.
EM
RI McNicol, Anne M/C-8075-2011
OI Tischler, Arthur/0000-0002-9726-5739
CR August C, 2004, MODERN PATHOL, V17, P1119, DOI 10.1038/modpathol.3800160
   Bryant J, 2003, J NATL CANCER I, V95, P1196, DOI 10.1093/jnci/djg024
   Dahia PLM, 2005, PLOS GENET, V1, P72, DOI 10.1371/journal.pgen.0010008
   Dannenberg H, 2003, ENDOCR PATHOL, V14, P329, DOI 10.1385/EP:14:4:329
   DeLellis RA, 2004, TUMOURS ENDOCRINE OR
   Dymock BW, 2005, J MED CHEM, V48, P4212, DOI 10.1021/jm050355z
   Eisenhofer G, 2004, ENDOCR-RELAT CANCER, V11, P423, DOI 10.1677/erc.1.00829
   Eisenhofer G, 2004, ENDOCR-RELAT CANCER, V11, P897, DOI 10.1677/erc.1.00838
   KARSNER HT, 1950, ATLAS TUMOR PATHOLOG
   Kimura N, 2005, ENDOCR PATHOL, V16, P23, DOI 10.1385/EP:16:1:023
   Koch CA, 2002, ENDOCR PATHOL, V13, P17, DOI 10.1385/EP:13:1:17
   Lack EE, 1999, VIRCHOWS ARCH, V435, P87, DOI 10.1007/s004280050403
   Lack EE, 2003, HUM PATHOL, V34, P112, DOI 10.1053/hupa.2003.13
   Lenders JWM, 2005, LANCET, V366, P665, DOI 10.1016/S0140-6736(05)67139-5
   LINNOILA RI, 1990, HUM PATHOL, V21, P1168
   Liu Tong-Hua, 2004, Zhonghua Bing Li Xue Za Zhi, V33, P198
   LLOYD RV, 1986, AM J PATHOL, V125, P45
   Portela-Gomes GM, 2004, APMIS, V112, P663, DOI 10.1111/j.1600-0463.2004.t01-1-apm1121003.x
   Srivastava A, 2004, ENDOCR PATHOL, V15, P175, DOI 10.1385/EP:15:2:175
   Thompson LDR, 2002, AM J SURG PATHOL, V26, P551, DOI 10.1097/00000478-200205000-00002
   Tischler AS, 1998, CELL TISSUE RES, V293, P563, DOI 10.1007/s004410051148
NR 21
TC 36
Z9 38
U1 0
U2 3
PU WILEY-BLACKWELL
PI MALDEN
PA COMMERCE PLACE, 350 MAIN STREET, MALDEN 02148, MA USA
SN 0077-8923
BN 1-57331-597-4
J9 ANN NY ACAD SCI
JI Ann.NY Acad.Sci.
PY 2006
VL 1073
BP 557
EP 570
DI 10.1196/annals.1353.059
PG 14
WC Oncology; Endocrinology & Metabolism; Multidisciplinary Sciences
WE Conference Proceedings Citation Index - Science (CPCI-S); Science Citation Index Expanded (SCI-EXPANDED)
SC Oncology; Endocrinology & Metabolism; Science & Technology - Other
   Topics
GA BFD27
UT WOS:000241140400058
PM 17102124
DA 2023-08-27
ER

PT J
AU Bektasoglu, HK
   Cipe, G
   Yardimci, E
   Arici, DS
   Hasbahceci, M
   Karatepe, O
   Muslumanoglu, M
AF Bektasoglu, Huseyin Kazim
   Cipe, Gokhan
   Yardimci, Erkan
   Arici, Dilek Sema
   Hasbahceci, Mustafa
   Karatepe, Oguzhan
   Muslumanoglu, Mahmut
TI Metachronous metastatic paraganglioma in jejunum as a rare entity: A
   case report
SO ONCOLOGY LETTERS
LA English
DT Article
DE paraganglioma; pheochromocytoma; jejunum; bowel obstruction
ID MALIGNANT PHEOCHROMOCYTOMA; LAPAROSCOPY; TUMORS
AB Pheochromocytomas and paragangliomas are neuroendocrine tumors that arise from chromaffin cells of adrenal medulla and extra-adrenal paraganglia, respectively. The recurrence of these neuroendocrine tumors as a jejunal mass causing obstruction in the small intestine is an exceptional entity. The present study reports the case of a,70-year-old male who presented to the Emergency Department of Bezmialem Vakif University Hospital with abdominal pain and vomiting. The patient possessed a history of left nephrectomy due to malignant pheochromocytoma that had invaded into the left kidney eight months prior to presentation. Bowel obstruction was diagnosed and the patient underwent a laparoscopic procedure. Partial resection of the jejunum was performed and immunohistochemical studies revealed the lesion to be malignant paraganglioma. The majority of paragangliomas are chemo- and radioresistant. Surgical excision remains the primary treatment. Metachronous paraganglioma arising from the small intestine is an extremely rare entity and may be a relevant consideration in patients presenting with bowel obstruction.
C1 [Bektasoglu, Huseyin Kazim; Cipe, Gokhan; Yardimci, Erkan; Hasbahceci, Mustafa; Karatepe, Oguzhan; Muslumanoglu, Mahmut] Bezmialem Vakif Univ, Fac Med, Dept Gen Surg, TR-34093 Istanbul, Turkey.
   [Arici, Dilek Sema] Bezmialem Vakif Univ, Fac Med, Dept Pathol, TR-34093 Istanbul, Turkey.
C3 Bezmialem Vakif University; Bezmialem Vakif University
RP Bektasoglu, HK (通讯作者)，Bezmialem Vakif Univ, Fac Med, Dept Gen Surg, Adnan Menderes Bulvari, TR-34093 Istanbul, Turkey.
EM
RI hasbahceci, mustafa/ABG-1763-2020; Yardimci, Erkan/AAR-1013-2020
OI Yardimci, Erkan/0000-0003-0908-4274
CR Adjalle R, 2009, HORM METAB RES, V41, P687, DOI 10.1055/s-0029-1231025
   Al-Mulhim AS, 2008, J LAPAROENDOSC ADV S, V18, P599, DOI 10.1089/lap.2007.0199
   Amar L, 2005, J CLIN ENDOCR METAB, V90, P2110, DOI 10.1210/jc.2004-1398
   Fishbein L, 2012, CANCER GENET-NY, V205, P1, DOI 10.1016/j.cancergen.2012.01.009
   Ilias I, 2008, NUCL MED BIOL, V35, pS27, DOI 10.1016/j.nucmedbio.2008.04.007
   Jimenez C, 2013, CURR ONCOL REP, V15, P356, DOI 10.1007/s11912-013-0320-x
   Karamanakos SN, 2010, SURG LAPARO ENDO PER, V20, P119, DOI 10.1097/SLE.0b013e3181d87178
   Liauw Jennifer J Y, 2005, Asian J Surg, V28, P185, DOI 10.1016/S1015-9584(09)60339-0
   Matro J, 2013, HORM METAB RES, V45, P147, DOI 10.1055/s-0032-1331211
   Moslemi MK, 2012, INT J SURG CASE REP, V3, P537, DOI 10.1016/j.ijscr.2012.07.007
   Young WE, 2006, ANN NY ACAD SCI, V1073, P21, DOI 10.1196/annals.1353.002
NR 11
TC 1
Z9 1
U1 0
U2 0
PU SPANDIDOS PUBL LTD
PI ATHENS
PA POB 18179, ATHENS, 116 10, GREECE
SN 1792-1074
EI 1792-1082
J9 ONCOL LETT
JI Oncol. Lett.
PD MAR
PY 2015
VL 9
IS 3
BP 1278
EP 1280
DI 10.3892/ol.2015.2860
PG 3
WC Oncology
WE Science Citation Index Expanded (SCI-EXPANDED)
SC Oncology
GA CD2OE
UT WOS:000350918100047
PM 25663897
OA Green Published, Green Submitted, gold
DA 2023-08-27
ER

PT J
AU Taha, H
   Schirmer, M
AF Taha, H.
   Schirmer, M.
TI Clear Cell Meningeoma of the Brainstem in a 3.5-year-old Child: Case
   Report and Review of the Literature
SO CENTRAL EUROPEAN NEUROSURGERY
LA English
DT Article
ID 1ST 2 DECADES; RADIATION-INDUCED MENINGIOMAS; INTRACRANIAL MENINGIOMAS;
   NEUROFIBROMATOSIS; TUMORS; LIFE; NF2
C1 [Taha, H.] Klinikum Solingen, Neurochirurg Klin, Dept Neurosurg, D-42653 Solingen, Germany.
RP Taha, H (通讯作者)，Klinikum Solingen, Neurochirurg Klin, Dept Neurosurg, Gotenstr 1, D-42653 Solingen, Germany.
EM
CR ABDULRAUF SI, 2003, TXB NEUROLOGICAL SUR, V2, P1303
   Al-Mefty O, 2004, J NEUROSURG, V100, P1002, DOI 10.3171/jns.2004.100.6.1002
   Amirjamshidi A, 2000, CHILD NERV SYST, V16, P406, DOI 10.1007/s003819900205
   Baumgartner JE, 1996, J NEURO-ONCOL, V29, P223, DOI 10.1007/BF00165652
   Begnami MD, 2007, INT J SURG PATHOL, V15, P110, DOI 10.1177/1066896906299128
   Carra S, 2001, PEDIATR NEUROSURG, V34, P264, DOI 10.1159/000056035
   CHAN RC, 1984, J NEUROSURG, V60, P52, DOI 10.3171/jns.1984.60.1.0052
   DARLING JR, 1994, LEADERSHIP ORG DEV J, V15, P3, DOI DOI 10.1108/01437739410073047
   DAVIDSON GS, 1989, CANCER, V63, P1205, DOI 10.1002/1097-0142(19890315)63:6<1205::AID-CNCR2820630627>3.0.CO;2-U
   DEEN HG, 1982, J NEUROSURG, V56, P317, DOI 10.3171/jns.1982.56.3.0317
   Demirtas E, 2000, PATHOL RES PRACT, V196, P151, DOI 10.1016/S0344-0338(00)80095-3
   Di Rocco C, 1999, CRIT REV NEUROSURG, V9, P180, DOI 10.1007/s003290050129
   Erdincler P, 1998, SURG NEUROL, V49, P136, DOI 10.1016/S0090-3019(97)00343-1
   Ferrante L, 1989, Br J Neurosurg, V3, P189, DOI 10.3109/02688698909002794
   GERMANO IM, 1994, J NEUROSURG, V80, P447, DOI 10.3171/jns.1994.80.3.0447
   GHIM TT, 1993, CANCER, V71, P4091, DOI 10.1002/1097-0142(19930615)71:12<4091::AID-CNCR2820711247>3.0.CO;2-G
   GLASIER CM, 1993, AM J NEURORADIOL, V14, P237
   GOLDSMITH BJ, 1994, J NEUROSURG, V80, P195, DOI 10.3171/jns.1994.80.2.0195
   Kantar M, 2004, CHILD NERV SYST, V20, P46, DOI 10.1007/s00381-003-0798-x
   Mallucci CL, 1996, CHILD NERV SYST, V12, P582
   McCarthy BJ, 1998, J NEUROSURG, V88, P831, DOI 10.3171/jns.1998.88.5.0831
   MIRIMANOFF RO, 1985, J NEUROSURG, V62, P18, DOI 10.3171/jns.1985.62.1.0018
   MOLLESTON MC, 1994, PEDIATR NEUROSURG, V21, P195, DOI 10.1159/000120835
   MOSS SD, 1988, NEUROSURGERY, V22, P758
   NUMAGUCHI Y, 1978, Neurologia Medico-Chirurgica, V18, P119, DOI 10.2176/nmc.18pt1.119
   Nunes F, 2003, J CHILD NEUROL, V18, P718, DOI 10.1177/08830738030180101301
   PERILONGO G, 1992, PEDIATR NEUROSURG, V18, P16, DOI 10.1159/000120637
   Perry A, 2001, J NEUROPATH EXP NEUR, V60, P994, DOI 10.1093/jnen/60.10.994
   Ruggieri M, 2005, NEUROPEDIATRICS, V36, P21, DOI 10.1055/s-2005-837581
   Rushing EJ, 2005, J NEUROSURG, V103, P489, DOI 10.3171/ped.2005.103.6.0489
   Sadetzki S, 2002, J NEUROSURG, V97, P1078, DOI 10.3171/jns.2002.97.5.1078
   SANO K, 1981, CHILD BRAIN, V8, P98
   Sharif S, 1998, BRIT J NEUROSURG, V12, P369, DOI 10.1080/02688699844925
   Sheikh BY, 1996, SURG NEUROL, V45, P328, DOI 10.1016/0090-3019(95)00451-3
   Stettner GM, 2007, NEUROPEDIATRICS, V38, P29, DOI 10.1055/s-2007-980204
   Teo JGC, 1998, PEDIATR NEUROSURG, V28, P27, DOI 10.1159/000028614
   Tufan K, 2005, PEDIATR NEUROSURG, V41, P1, DOI 10.1159/000084858
   Turgut M, 1997, BRIT J NEUROSURG, V11, P501, DOI 10.1080/02688699745655
   ZORLUDEMIR S, 1995, AM J SURG PATHOL, V19, P493, DOI 10.1097/00000478-199505000-00001
   Zwerdling T, 2002, J PEDIAT HEMATOL ONC, V24, P199, DOI 10.1097/00043426-200203000-00008
NR 40
TC 3
Z9 3
U1 2
U2 2
PU GEORG THIEME VERLAG KG
PI STUTTGART
PA RUDIGERSTR 14, D-70469 STUTTGART, GERMANY
SN 1868-4904
EI 1868-4912
J9 CENT EUR NEUROSURG
JI Cent. Eur. Neurosurg.
PD AUG
PY 2010
VL 71
IS 3
BP 148
EP 151
DI 10.1055/s-0029-1220939
PG 4
WC Clinical Neurology; Surgery
WE Science Citation Index Expanded (SCI-EXPANDED)
SC Neurosciences & Neurology; Surgery
GA 642YS
UT WOS:000281258400010
PM 20393894
DA 2023-08-27
ER

PT J
AU Tongren, JE
   Yang, CF
   Collins, WE
   Sullivan, JS
   Lal, AA
   Xiao, LH
AF Tongren, JE
   Yang, CF
   Collins, WE
   Sullivan, JS
   Lal, AA
   Xiao, LH
TI Expression of proinflammatory cytokines in four regions of the brain in
   Macaque mulatta (rhesus) monkeys infected with Plasmodium coatneyi
SO AMERICAN JOURNAL OF TROPICAL MEDICINE AND HYGIENE
LA English
DT Article
ID TUMOR NECROSIS FACTOR; CEREBRAL MALARIA; PARASITIZED ERYTHROCYTES;
   SEQUESTRATION; CEREBELLUM; PATHOLOGY; PROFILES
AB We have characterized brain cytokine expression profiles in the Plasmodium coatneyi/rhesus (Macaque mulatta) malaria model. Eight rhesus monkeys were included in the study; four were infected with P. coatneyi, and four were used as uninfected controls. All inoculated animals became infected. Eleven days after parasite inoculation, the rhesus monkeys were killed and tissue samples from 4 regions of the brain (cortex and white matter of the cerebrum, cerebellum, and midbrain) were collected for quantitation of mRNA expression of cytokines, adhesion molecules, and inducible nitric oxide synthetase (iNOS) by reverse transcriptase-polymerase chain reaction (RT-PCR). The expression levels of tumor necrosis actor-alpha (TNF-alpha), gamma interferon (IFN-gamma), interleukin-l-beta (IL-1 beta), intercellular adhesion molecule-1 (ICAM-1) and inducible nitric oxide synethetase (iNOS) were highest in the cerebellum of infected animals, correlating well with pathologic observations of sequestration of parasitized erythrocytes in this region of the brain. Infected animals also had higher TNF-alpha expression levels in the cortex and IL-1 beta expression levels in the cortex, white matter, and midbrain. Thus, the expression of pro-inflammatory and T helper-1 (TH-1) cytokines, adhesion molecules, and iNOS appears to predominate in the cerebellum of infected rhesus monkeys.
C1 Ctr Dis Control & Prevent, Div Parasit Dis, Natl Ctr Infect Dis, US Dept HHS, Atlanta, GA 30341 USA.
C3 Centers for Disease Control & Prevention - USA
RP Xiao, LH (通讯作者)，Ctr Dis Control & Prevent, Div Parasit Dis, Natl Ctr Infect Dis, US Dept HHS, Mail Stop F-122,4770 Buford Highway, Atlanta, GA 30341 USA.
RI Xiao, Lihua/B-1704-2013; Yang, Chunfu/G-6890-2013
OI Xiao, Lihua/0000-0001-8532-2727; 
CR AIKAWA M, 1988, AM J TROP MED HYG, V39, P3, DOI 10.4269/ajtmh.1988.39.3
   AIKAWA M, 1992, AM J TROP MED HYG, V46, P391, DOI 10.4269/ajtmh.1992.46.391
   BERENDT AR, 1994, PARASITOL TODAY, V10, P412, DOI 10.1016/0169-4758(94)90238-0
   CLARK IA, 1994, PARASITOL TODAY, V10, P410, DOI 10.1016/0169-4758(94)90237-2
   CURFS JHAJ, 1993, EXP PARASITOL, V77, P212, DOI 10.1006/expr.1993.1078
   DEKOSSODO S, 1993, J IMMUNOL, V151, P4811
   DENIS M, 1994, J LEUKOCYTE BIOL, V55, P682, DOI 10.1002/jlb.55.5.682
   ELING WMC, 1994, BIOTHERAPY, V7, P211, DOI 10.1007/BF01878487
   Gerritsen ME, 1996, CARDIOVASC RES, V32, P720, DOI 10.1016/S0008-6363(96)00072-7
   GRAU GE, 1989, NEW ENGL J MED, V320, P1586, DOI 10.1056/NEJM198906153202404
   HARPAZ R, 1992, J CLIN INVEST, V90, P515, DOI 10.1172/JCI115889
   HOLLOWAY B, 1993, NUCLEIC ACIDS RES, V21, P3905, DOI 10.1093/nar/21.16.3905
   Jennings VM, 1997, INFECT IMMUN, V65, P4883, DOI 10.1128/IAI.65.11.4883-4887.1997
   KERN P, 1989, AM J MED, V87, P139, DOI 10.1016/S0002-9343(89)80688-6
   KWIATKOWSKI D, 1990, LANCET, V336, P1201, DOI 10.1016/0140-6736(90)92827-5
   NAGATAKE T, 1992, AM J TROP MED HYG, V47, P259, DOI 10.4269/ajtmh.1992.47.259
   NUSSENZWEIG RS, 1994, SCIENCE, V265, P1381, DOI 10.1126/science.8073276
   PORTA J, 1993, CLIN NEUROPATHOL, V12, P142
   Rudin W, 1997, AM J PATHOL, V150, P257
   SEIN KK, 1993, AM J TROP MED HYG, V49, P513, DOI 10.4269/ajtmh.1993.49.513
   SEIN KK, 1993, AM J TROP MED HYG, V48, P504, DOI 10.4269/ajtmh.1993.48.504
   Smith CD, 1996, AM J TROP MED HYG, V55, P379, DOI 10.4269/ajtmh.1996.55.379
   Sornasse T, 1996, J EXP MED, V184, P473, DOI 10.1084/jem.184.2.473
   Xiao LH, 1996, J IMMUNOL METHODS, V199, P139, DOI 10.1016/S0022-1759(96)00174-3
   Yang CF, 1999, AM J TROP MED HYG, V61, P226, DOI 10.4269/ajtmh.1999.61.226
NR 25
TC 19
Z9 21
U1 0
U2 1
PU AMER SOC TROP MED & HYGIENE
PI MCLEAN
PA 8000 WESTPARK DR, STE 130, MCLEAN, VA 22101 USA
SN 0002-9637
EI 1476-1645
J9 AM J TROP MED HYG
JI Am. J. Trop. Med. Hyg.
PD APR
PY 2000
VL 62
IS 4
BP 530
EP 534
DI 10.4269/ajtmh.2000.62.530
PG 5
WC Public, Environmental & Occupational Health; Tropical Medicine
WE Science Citation Index Expanded (SCI-EXPANDED)
SC Public, Environmental & Occupational Health; Tropical Medicine
GA 401TX
UT WOS:000166945300020
PM 11220773
OA Bronze
DA 2023-08-27
ER

PT J
AU Ng, WH
   Wan, GQ
   Peng, ZN
   Too, HP
AF Ng, Wai Hoe
   Wan, Guo Qiang
   Peng, Zhong Ni
   Too, Heng Phon
TI Glial cell-line derived neurotrophic factor (GDNF) family of ligands
   confer chemoresistance in a ligand-specific fashion in malignant gliomas
SO JOURNAL OF CLINICAL NEUROSCIENCE
LA English
DT Article
DE GDNF; Glioma; Astrocytoma; Brain tumor; Chemotherapy
ID EXPRESSION; RECEPTOR; GFR-ALPHA-1; ACTIVATION; INVASION; ISOFORMS
AB Glial cell-line derived neurotrophic factor (GDNF) is a neurotrophic factor known to promote neuronal survival of dopaminergic neurons in the embryonic midbrain as well as contribute to carcinogenesis in many cancers. Its ubiquitous presence in the central nervous system suggests a role in the mitogenesis of high-grade astrocytoma. GDNF is overexpressed in glioblastoma cell lines and human gliomas. GFR alpha 1b is the predominant spliced receptor isoform in human gliomas and RET9 is the predominant co-receptor. Significantly there is differential overexpression of the GFR alpha 1b spliced isoform compared to the GFR alpha 1a spliced variant. Pre-treatment of glioblastoma cell lines with GDNF but not the alternative ligand neurturin, promoted mitogenic behaviour and conferred chemoresistance to 1,3-bis(2-chloroethyl)-1-nitrosourea (BCNU). Signaling mapping of BCNU and GDNF suggest that the ability of GDNF to promote Akt activity and inhibit JNK activity may contribute to the increased cellular survival after BCNU chemotherapy. (C) 2008 Elsevier Ltd. All rights reserved.
C1 [Ng, Wai Hoe; Wan, Guo Qiang; Peng, Zhong Ni; Too, Heng Phon] Natl Univ Singapore, Fac Med, Dept Biochem, Singapore, Singapore.
C3 National University of Singapore
RP Ng, WH (通讯作者)，Natl Univ Singapore, Fac Med, Dept Biochem, Singapore, Singapore.
EM
RI Wan, Guoqiang/GRR-5628-2022
OI Wan, Guoqiang/0000-0003-3841-9301
FU Singapore Millenium Foundation (SMF)
FX We would like to acknowledge the support of the Singapore Millenium
   Foundation (SMF) for this research.
CR Airaksinen MS, 1999, MOL CELL NEUROSCI, V13, P313, DOI 10.1006/mcne.1999.0754
   Airaksinen MS, 2002, NAT REV NEUROSCI, V3, P383, DOI 10.1038/nrn812
   Charlet-Berguerand N, 2004, CELL SIGNAL, V16, P1425, DOI 10.1016/j.cellsig.2004.05.006
   Dey BK, 1998, NEUROREPORT, V9, P37, DOI 10.1097/00001756-199801050-00008
   Hamel W, 2000, ACTA NEUROCHIR, V142, P113, DOI 10.1007/s007010050015
   Hansford LA, 2005, NEUROSCI LETT, V389, P77, DOI 10.1016/j.neulet.2005.07.034
   Ito Y, 2005, SURGERY, V138, P788, DOI 10.1016/j.surg.2005.07.007
   Iwahashi N, 2002, CANCER, V94, P167, DOI 10.1002/cncr.10169
   Johnson JM, 2003, SCIENCE, V302, P2141, DOI 10.1126/science.1090100
   LANTOS PL, 1986, GREENFIELDS NEUROPAT
   LIN LFH, 1993, SCIENCE, V260, P1130, DOI 10.1126/science.8493557
   Lindahl M, 2000, MOL CELL NEUROSCI, V15, P522, DOI 10.1006/mcne.2000.0845
   Meng XJ, 2000, SCIENCE, V287, P1489, DOI 10.1126/science.287.5457.1489
   Mueller MM, 2003, ACTA NEUROCHIR, V145, P999, DOI 10.1007/s00701-003-0132-0
   Popsueva A, 2003, J CELL BIOL, V161, P119, DOI 10.1083/jcb.200212174
   Powers JF, 2001, NEUROSCIENCE, V108, P341, DOI 10.1016/S0306-4522(01)00410-9
   Sanicola M, 1997, P NATL ACAD SCI USA, V94, P6238, DOI 10.1073/pnas.94.12.6238
   Shefelbine SE, 1998, HUM GENET, V102, P474, DOI 10.1007/s004390050724
   Song H, 2006, NEUROSCI RES, V56, P29, DOI 10.1016/j.neures.2006.04.019
   Wiesenhofer B, 2000, ANTISENSE NUCLEIC A, V10, P311, DOI 10.1089/oli.1.2000.10.311
   Wiesenhofer B, 2000, ACTA NEUROPATHOL, V99, P131, DOI 10.1007/PL00007416
   Wong YW, 1998, NEUROREPORT, V9, P3767, DOI 10.1097/00001756-199812010-00002
   Yoong LF, 2005, MOL BRAIN RES, V139, P1, DOI 10.1016/j.molbrainres.2005.05.016
   Zulch KJ., 1986, BRAIN TUMORS THEIR B
NR 24
TC 29
Z9 32
U1 0
U2 2
PU ELSEVIER SCI LTD
PI OXFORD
PA THE BOULEVARD, LANGFORD LANE, KIDLINGTON, OXFORD OX5 1GB, OXON, ENGLAND
SN 0967-5868
EI 1532-2653
J9 J CLIN NEUROSCI
JI J. Clin. Neurosci.
PD MAR
PY 2009
VL 16
IS 3
BP 427
EP 436
DI 10.1016/j.jocn.2008.06.002
PG 10
WC Clinical Neurology; Neurosciences
WE Science Citation Index Expanded (SCI-EXPANDED)
SC Neurosciences & Neurology
GA 412ZI
UT WOS:000263762300015
PM 19138852
DA 2023-08-27
ER

PT J
AU Stieb, S
   Malla, M
   Graydon, S
   Riesterer, O
   Klock, S
   Studer, G
   Tanadini-Lang, S
AF Stieb, Sonja
   Malla, Michelle
   Graydon, Shaun
   Riesterer, Oliver
   Kloeck, Stephan
   Studer, Gabriela
   Tanadini-Lang, Stephanie
TI Dosimetric influence of pitch in patient positioning for radiotherapy of
   long treatment volumes; the usefulness of six degree of freedom couch
SO BRITISH JOURNAL OF RADIOLOGY
LA English
DT Article
ID ROTATIONAL SETUP ERRORS; MODULATED RADIATION-THERAPY; HEAD;
   IMMOBILIZATION; UNCERTAINTIES; CONSEQUENCES; CANCER; SYSTEM; ROLL
AB Objective: Pitch, the rotation around the transversal axis of the patient during radiotherapy has little impact on the dose distribution of small spherical treatment volumes; however it might affect treatment of long volumes requiring a correction with a six degree of freedom couch.
   Methods: We included 10 patients each with nasopharyngeal carcinoma (NPC) and esophageal cancer, treated with volumetric modulated arc therapy, Pitch was simulated by tilting the planning CT in ventral and dorsal direction by +/- 1.5 degrees and +/- 3 degrees. Verification plans were calculated on the tilted datasets and were compared to the original plan and the dose constraints of the organs at risk (OAR).
   Results: The deviation in dose to the planning target volume is increasing with the degree of pitch with mean changes of up to 2% for NPC and 1% for esophageal cancer. The most affected OAR in NPC patients are brainstem (max. dose +6.0%) and spinal cord (max. dose +10.0%) when tilted by 3 degrees dorsally and lenses (max. dose +3.3%), oral mucosa (mean dose +2.6%) and parotid glands (mean dose +4.3%) when tilted by 3 degrees ventrally. For esophageal cancer patients, there was no significant change in dose to any OAR. Whereas for esophageal cancer, all tilted treatment plans were still clinically acceptable regarding OAR, 5 NPC plans would no longer be acceptable with a pitch of 1.5 degrees ventral (N = 1), 3 degrees ventral (N = 2) and 3 degrees dorsal (N = 2).
   Conclusion: Planning target volume coverage in both tumor entities was only slightly affected, but pitch errors could be relevant for OAR in NPC patients.
   Advances in knowledge: A correction with a six degree of freedom couch is recommended for NPC patients with a pitch mismatch of more than 1.5 degrees to avoid exceeded doses to the OAR.
C1 [Stieb, Sonja; Malla, Michelle; Graydon, Shaun; Riesterer, Oliver; Kloeck, Stephan; Studer, Gabriela; Tanadini-Lang, Stephanie] Univ Hosp Zurich, Dept Radiat Oncol, Zurich, Switzerland.
   [Stieb, Sonja; Malla, Michelle; Graydon, Shaun; Riesterer, Oliver; Kloeck, Stephan; Studer, Gabriela; Tanadini-Lang, Stephanie] Univ Zurich, Zurich, Switzerland.
   [Stieb, Sonja] Univ Hosp Zurich, Inst Diagnost & Intervent Radiol, Zurich, Switzerland.
   [Studer, Gabriela] Cantonal Hosp Lucerne, Inst Radiat Oncol, Luzern, Switzerland.
C3 University of Zurich; University Zurich Hospital; University of Zurich;
   University of Zurich; University Zurich Hospital; Lucerne Cantonal
   Hospital
RP Stieb, S (通讯作者)，Univ Hosp Zurich, Dept Radiat Oncol, Zurich, Switzerland.; Stieb, S (通讯作者)，Univ Zurich, Zurich, Switzerland.; Stieb, S (通讯作者)，Univ Hosp Zurich, Inst Diagnost & Intervent Radiol, Zurich, Switzerland.
EM
RI Klöck, Stephan/GLR-9861-2022; Studer, Gabriela Monika/A-3013-2017
OI Klöck, Stephan/0000-0002-4105-3219; Riesterer,
   Oliver/0000-0002-9508-0546; Tanadini-Lang, Stephanie/0000-0002-4387-1522
FU KFSP Tumor Oxygenation of the University of Zurich
FX This work was supported by the KFSP Tumor Oxygenation of the University
   of Zurich
CR Ahn PH, 2009, INT J RADIAT ONCOL, V73, P626, DOI 10.1016/j.ijrobp.2008.10.007
   Chen YJ, 2007, INT J RADIAT ONCOL, V68, P1537, DOI 10.1016/j.ijrobp.2007.04.023
   Fu WH, 2006, PHYS MED BIOL, V51, P5321, DOI 10.1088/0031-9155/51/20/016
   Fu WH, 2013, MED DOSIM, V38, P125, DOI 10.1016/j.meddos.2012.09.003
   Guckenberger M, 2006, INT J RADIAT ONCOL, V65, P934, DOI 10.1016/j.ijrobp.2006.02.019
   Guckenberger M, 2012, RADIAT ONCOL, V7, DOI 10.1186/1748-717X-7-63
   Hong TS, 2005, INT J RADIAT ONCOL, V61, P779, DOI 10.1016/j.ijrobp.2004.07.696
   Kaiser A, 2006, INT J RADIAT ONCOL, V66, P949, DOI 10.1016/j.ijrobp.2006.05.055
   Kim GY, 2008, MED DOSIM, V33, P93, DOI 10.1016/j.meddos.2007.05.004
   Lang S, 2015, PRACT RADIAT ONCOL, V5, pE689, DOI 10.1016/j.prro.2015.03.007
   Lee J, 2015, BRIT J RADIOL, V88, DOI 10.1259/bjr.20140862
   Lin CG, 2017, J MED RADIAT SCI, V64, P106, DOI 10.1002/jmrs.189
   Mancosu P, 2015, BRIT J RADIOL, V88, DOI 10.1259/bjr.20150468
   Oh YK, 2014, J APPL CLIN MED PHYS, V15, P85, DOI 10.1120/jacmp.v15i2.4418
   Peng JL, 2011, J APPL CLIN MED PHYS, V12, P61, DOI 10.1120/jacmp.v12i3.3422
   Sejpal SV, 2009, INT J RADIAT ONCOL, V75, P40, DOI 10.1016/j.ijrobp.2008.08.042
   Velec M, 2010, INT J RADIAT ONCOL, V76, P949, DOI 10.1016/j.ijrobp.2009.07.004
   Wang WD, 2014, BIOMED RES INT, V2014, DOI 10.1155/2014/814948
   Zhang LF, 2006, INT J RADIAT ONCOL, V64, P1559, DOI 10.1016/j.ijrobp.2005.12.023
NR 19
TC 2
Z9 2
U1 0
U2 2
PU BRITISH INST RADIOLOGY
PI LONDON
PA 36 PORTLAND PLACE, LONDON W1N 4AT, ENGLAND
SN 0007-1285
EI 1748-880X
J9 BRIT J RADIOL
JI Br. J. Radiol.
PY 2018
VL 91
IS 1091
AR 20170704
DI 10.1259/bjr.20170704
PG 8
WC Radiology, Nuclear Medicine & Medical Imaging
WE Science Citation Index Expanded (SCI-EXPANDED)
SC Radiology, Nuclear Medicine & Medical Imaging
GA GX6WC
UT WOS:000447903200004
PM 30004794
OA Green Published
DA 2023-08-27
ER

PT J
AU PARK, SH
   CHI, JG
   CHO, BK
   WANG, KC
AF PARK, SH
   CHI, JG
   CHO, BK
   WANG, KC
TI SPINAL-CORD GANGLIOGLIOMA IN CHILDHOOD
SO PATHOLOGY RESEARCH AND PRACTICE
LA English
DT Article
DE GANGLIOGLIOMA; SPINAL CORD; SYRINGOMYELIA; ULTRASTRUCTURAL STUDY
ID TUMORS; CHILDREN
AB Two childhood cases of spinal cord ganglioglioma are described. Both cases showed a remarkable similarity in presentation, radiologically and morphologically. The first case was a 6 1/2-year-old boy who bad ganglioglioma involving the entire length of the spinal cord. He has been suffering from a long span of scoliosis and gait disturbance since he was 1 year. The tumor contained multiple syrinx. The second case was a 3-year-old girl with ganglioglioma involving a large portion of the spinal cord from lower medulla to T3. This case also contained syringobulbia and syringomyelia in the multiple levels. She suffered from progressive motor weakness of all extremities. Owing to a good demarcation from normal spinal cord, the tumors could easily be removed by the microsurgical dissection through second stage operation in both cases. Histopatbologically and ultrastructurally, the tumors were benign gangliogliomas. The symptoms improved a lot after surgery. Based on our two cases and literature review we confirmed that the spinal cord gangliogliomas are slowly growing tumors occurring in childhood or young adulthood. They often involve usually extensive level or entire length of the spinal cord and may be accompanied by scoliosis and gait disturbance when the tumor presents since infancy or early childhood as in our cases. They also often exhibit a cystic change of the tumor associated with large or small syringomyelia. Nearly total removal is possible due to a good demarcation from the normal spinal cord. Therefore, early diagnosis and surgical management are fundamental for the avoidance of unnecessary delay of surgery and a good result.
C1 SEOUL NATL UNIV,CHILDRENS HOSP,DEPT PATHOL,28 YONGON DONG,CHONGNO GU,SEOUL 110744,SOUTH KOREA.
   SEOUL NATL UNIV,CHILDRENS HOSP,DEPT NEUROSURG,SEOUL 151,SOUTH KOREA.
   SEOUL NATL UNIV,COLL MED,SEOUL 151,SOUTH KOREA.
C3 Seoul National University (SNU); Seoul National University Hospital;
   Seoul National University (SNU); Seoul National University Hospital;
   Seoul National University (SNU)
RI Geun, Je/G-4989-2011; Park, Sung-Hye/B-5325-2011; Wang,
   Kyu-Chang/J-5545-2012
OI Park, Sung-Hye/0000-0002-8681-1597; Chi, Je-Geun/0000-0002-9950-2072
CR ALBRIGHT L, 1980, CHILD BRAIN, V6, P274
   BELL WO, 1988, J NEUROSURG, V69, P295, DOI 10.3171/jns.1988.69.2.0295
   BEVILACQUA G, 1979, ACTA NEUROPATHOL, V48, P239, DOI 10.1007/BF00690528
   COURVILLE CB, 1930, ARCH NEUROL PSYCHIAT, V24, P431
   GARRIDO E, 1978, CHILD BRAIN, V4, P339
   HARDISON HH, 1987, CHILD NERV SYST, V3, P89, DOI 10.1007/BF00271131
   HERRMANN HD, 1988, NEUROSURGERY, V22, P518, DOI 10.1227/00006123-198803000-00011
   JOHANNSSON JH, 1981, J NEUROSURG, V54, P58, DOI 10.3171/jns.1981.54.1.0058
   KALYANARAMAN UP, 1982, HUM PATHOL, V13, P952, DOI 10.1016/S0046-8177(82)80062-2
   KALYANRAMAN UP, 1987, NEUROSURGERY, V20, P428, DOI 10.1227/00006123-198703000-00012
   Kernohan JW, 1932, BRAIN, V55, P287, DOI 10.1093/brain/55.3.287
   KITANO M, 1987, ACTA PATHOL JAPON, V37, P1009
   Lichtenstein BW, 1937, ARCH NEURO PSYCHIATR, V37, P1356, DOI 10.1001/archneurpsyc.1937.02260180136009
   PICK L, 1911, Z GESAMTE NEUROL PSY, V6, P397
   RICHMOND JG, 1966, AM J CLIN PATHOL, V45, P692
   RODEWALD L, 1987, American Journal of Medical Genetics, V26, P7, DOI 10.1002/ajmg.1320260103
   RUSSELL DS, 1962, J NEUROPATH EXP NEUR, V21, P185, DOI 10.1097/00005072-196204000-00002
   Russell DS, 1989, PATHOLOGY TUMORS NER, V5, P289
   WALD U, 1985, J NEUROSURG, V62, P14
   ZIMMERMAN H, 1971, PATHOLOGY NERVOUS SY, P1947
   ZUELCH KJ, 1986, BRAIN TUMORS THEIR B, P292
   1943, 7 MED RES COUNC WAR
NR 22
TC 22
Z9 22
U1 0
U2 1
PU GUSTAV FISCHER VERLAG
PI JENA
PA VILLENGANG 2, D-07745 JENA, GERMANY
SN 0344-0338
J9 PATHOL RES PRACT
JI Pathol. Res. Pract.
PD MAR
PY 1993
VL 189
IS 2
BP 189
EP 196
DI 10.1016/S0344-0338(11)80091-9
PG 8
WC Pathology
WE Science Citation Index Expanded (SCI-EXPANDED)
SC Pathology
GA KY676
UT WOS:A1993KY67600011
PM 8321747
DA 2023-08-27
ER

PT J
AU Yan, J
   Studer, L
   McKay, RDG
AF Yan, J
   Studer, L
   McKay, RDG
TI Ascorbic acid increases the yield of dopaminergic neurons derived from
   basic fibroblast growth factor expanded mesencephalic precursors
SO JOURNAL OF NEUROCHEMISTRY
LA English
DT Article
DE midbrain; progenitor cell; stem cells; tyrosine-hydroxylase
ID TYROSINE-HYDROXYLASE; STEM-CELLS; CULTURE; DIFFERENTIATION; INDUCTION;
   PHENOTYPE; TISSUE; MICE
AB CNS precursors derived from E12 rat mesencephalon proliferate in the presence of basic fibroblast growth factor and differentiate in vitro into functional dopaminergic neurons, which upon transplantation alleviate behavioral symptoms in a rat model of Parkinson's disease. Here we show that the efficiency of dopaminergic differentiation decreases in the mesencephalic precursors that were proliferated or passaged for extended periods in vitro. Ascorbic acid treatment restored dopaminergic differentiation in these precursors and led to a greater than 10-fold increase in dopamine neuron yield compared with untreated cultures. The effect of ascorbic acid was stereospecific and could not be mimicked by any other antioxidants. The expression of sodium-dependent vitamin C transporter, a recently identified stereospecific ascorbic acid transporter, was maintained in mesencephalic precursors for extended in vitro periods. Pre-treatment of in vitro expanded mesencephalic precursors with ascorbic acid might facilitate the large-scale generation of dopaminergic neurons for clinical transplantation.
C1 Mem Sloan Kettering Canc Ctr, Lab Stem Cell & Tumor Biol, New York, NY 10021 USA.
   NINDS, Mol Biol Lab, NIH, Bethesda, MD 20892 USA.
   Johns Hopkins Univ, Sch Med, Dept Pathol, Div Neuropathol, Baltimore, MD 21205 USA.
C3 Memorial Sloan Kettering Cancer Center; National Institutes of Health
   (NIH) - USA; NIH National Institute of Neurological Disorders & Stroke
   (NINDS); Johns Hopkins University
RP Studer, L (通讯作者)，Mem Sloan Kettering Canc Ctr, Lab Stem Cell & Tumor Biol, 1275 York Ave,Box 256, New York, NY 10021 USA.
RI Studer, Lorenz/ABH-1491-2021
OI Studer, Lorenz/0000-0003-0741-7987
CR Brundin P, 1998, NAT NEUROSCI, V1, P537, DOI 10.1038/2773
   Daadi MM, 1999, J NEUROSCI, V19, P4484
   Helbock HJ, 1999, METHOD ENZYMOL, V300, P156
   Jaiswal N, 1997, J CELL BIOCHEM, V64, P295, DOI 10.1002/(SICI)1097-4644(199702)64:2<295::AID-JCB12>3.3.CO;2-6
   KALIR HH, 1991, J NEUROCHEM, V57, P458, DOI 10.1111/j.1471-4159.1991.tb03773.x
   Max SR, 1996, DEV BRAIN RES, V91, P140, DOI 10.1016/0165-3806(95)00163-8
   NAKASHIMA Y, 1970, J VITAMINOL, V16, P276, DOI 10.5925/jnsv1954.16.276
   Pennathur S, 1999, J BIOL CHEM, V274, P34621, DOI 10.1074/jbc.274.49.34621
   REBEC GV, 1994, PROG NEUROBIOL, V43, P537, DOI 10.1016/0301-0082(94)90052-3
   Rice ME, 2000, TRENDS NEUROSCI, V23, P209, DOI 10.1016/S0166-2236(99)01543-X
   RONCHETTI IP, 1996, ASCORBIC ACID BIOCH
   ROSENBERG PA, 1988, J NEUROSCI, V8, P2887, DOI 10.1523/jneurosci.08-08-02887.1988
   Studer L, 1996, BRAIN RES BULL, V41, P143, DOI 10.1016/S0361-9230(96)00114-1
   Studer L, 1998, NAT NEUROSCI, V1, P537, DOI 10.1038/2774
   Studer L, 1998, NAT NEUROSCI, V1, P290, DOI 10.1038/1105
   Studer L, 2000, J NEUROSCI, V20, P7377
   Tsukaguchi H, 1999, NATURE, V399, P70, DOI 10.1038/19986
   Wagner J, 1999, NAT BIOTECHNOL, V17, P653, DOI 10.1038/10862
   Xiao GZ, 1997, MOL ENDOCRINOL, V11, P1103, DOI 10.1210/me.11.8.1103
   Ye WL, 1998, CELL, V93, P755, DOI 10.1016/S0092-8674(00)81437-3
   Zetterstrom RH, 1997, SCIENCE, V276, P248, DOI 10.1126/science.276.5310.248
   Zhou JW, 1996, EUR J NEUROSCI, V8, P2328, DOI 10.1111/j.1460-9568.1996.tb01196.x
NR 22
TC 130
Z9 151
U1 0
U2 4
PU LIPPINCOTT WILLIAMS & WILKINS
PI PHILADELPHIA
PA 530 WALNUT ST, PHILADELPHIA, PA 19106-3621 USA
SN 0022-3042
J9 J NEUROCHEM
JI J. Neurochem.
PD JAN
PY 2001
VL 76
IS 1
BP 307
EP 311
DI 10.1046/j.1471-4159.2001.00073.x
PG 5
WC Biochemistry & Molecular Biology; Neurosciences
WE Science Citation Index Expanded (SCI-EXPANDED)
SC Biochemistry & Molecular Biology; Neurosciences & Neurology
GA 388AE
UT WOS:000166154600033
PM 11146004
OA Bronze
DA 2023-08-27
ER

PT J
AU Qiao, YH
   Zhang, LZ
   Xu, KL
   Zeng, LY
   Meng, LJ
   Wang, J
   Meng, H
AF Qiao Yuehua
   Zhang Longzhen
   Xu Kailin
   Zeng Lingyu
   Meng Lingjian
   Wang Jun
   Meng Hong
TI Inflammatory Lesions of Cochlea in Murine Cytomegalovirus-Infected Mice
   with Hearing Loss
SO CELL BIOCHEMISTRY AND BIOPHYSICS
LA English
DT Article
DE Murine cytomegalovirus; Cochlear inflammation; Hearing loss; Tumor
   necrosis factor (TNF)-alpha; Interleukin (IL)-6
ID CMV INFECTION; VIRAL LOAD; CHILDREN
AB We sought to determine the distribution of tumor necrosis factor (TNF)-alpha, interleukin (IL)-6 and lymphocytes in the cochlea of mice infected with murine cytomegalovirus (MCMV). For this purpose, 16 newborn mice were divided equally into model and control (uninfected) groups. In model group, 10 mu l of MCMV was injected into the brain of each mouse whereas in control group, 10 mu l of physiological saline was injected. Fourteen days after the injection of MCMV, the auditory brainstem response audiometry was performed. Later, the mice were killed and the acoustic capsule samples were collected for polymerase chain reaction analysis, histopathological and immunohistochemical studies. Compared with control mice, the incubation period was longer (F = 13.797; P = 0.003) and the amplitude was lower (F = 5.095; P = 0.043) in model group mice. It indicated that the intracerebral injection of MCMV caused hearing loss in mice. Histopathological examination of cochleas revealed increased levels of lymphocytic infiltration in the membrana vestibularis. Higher levels of TNF-alpha and IL-6 in scala tympani were detected by immunohistochemical staining. Taken together, the hearing loss in mice could be related with the inflammatory changes occurring in cochlea after inception of MCMV infection.
C1 [Qiao Yuehua] Xuzhou Med Coll, Dept Otolaryngol, Xuzhou 221002, Jiangsu, Peoples R China.
   [Zhang Longzhen] Xuzhou Med Coll, Canc Res Ctr, Xuzhou 221002, Jiangsu, Peoples R China.
   [Xu Kailin; Zeng Lingyu] Xuzhou Med Coll, Transplant Immune Lab, Xuzhou 221002, Jiangsu, Peoples R China.
   [Meng Lingjian; Wang Jun] Xuzhou Med Coll, Affiliated Hosp, Dept Paediat, Xuzhou 221002, Jiangsu, Peoples R China.
   [Meng Hong] Shandong Prov Acad Med Sci, Basic Med Inst, Microbiol Lab, Jinan, Peoples R China.
C3 Xuzhou Medical University; Xuzhou Medical University; Xuzhou Medical
   University; Xuzhou Medical University; Shandong First Medical University
   & Shandong Academy of Medical Sciences
RP Xu, KL (通讯作者)，Xuzhou Med Coll, Transplant Immune Lab, Xuzhou 221002, Jiangsu, Peoples R China.
EM
FU Xuzhou Technology Development Fund, China [XM07C071]
FX We thank the Xuzhou Technology Development Fund, China for financial
   support (Grant # XM07C071). We are also indebted to the Microbiological
   Laboratory, Basic Medicine Institute, Shandong Province Academy of
   Medical Sciences, China for kindly providing 3T3 cells and MCMV Smith
   strain.
CR Biheng C., 2007, J PRACTICAL MED, V23, P336
   Dumitriu IE, 2005, TRENDS IMMUNOL, V26, P381, DOI 10.1016/j.it.2005.04.009
   Fowler KB, 2006, J CLIN VIROL, V35, P226, DOI 10.1016/j.jcv.2005.09.016
   Grosse SD, 2008, J CLIN VIROL, V41, P57, DOI 10.1016/j.jcv.2007.09.004
   Jiangying S., 2004, CHINESE J DIAGNOSTIC, V10, P359
   Lanari M, 2006, PEDIATRICS, V117, pE76, DOI 10.1542/peds.2005-0629
   Liangzhen W., 2005, CHINESE J PRACTICAL, V21, P331
   Mo Wei, 2005, Zhejiang Da Xue Xue Bao Yi Xue Ban, V34, P358
   Noyola DE, 2000, PEDIATR INFECT DIS J, V19, P505, DOI 10.1097/00006454-200006000-00003
   Pass Robert F, 2005, Herpes, V12, P50
   Ross SA, 2009, PEDIATR INFECT DIS J, V28, P588, DOI 10.1097/INF.0b013e3181979a27
   Tang-Feldman YJ, 2006, J VIROL METHODS, V131, P122, DOI 10.1016/j.jviromet.2005.07.013
   White DR, 2006, ARCH OTOLARYNGOL, V132, P608, DOI 10.1001/archotol.132.6.608
   Yan F., 2007, CHINESE J BIRTH HLTH, V15, P27
   Zhijian C., 2005, SHANDONG MED J, V45, P3
NR 15
TC 8
Z9 13
U1 0
U2 4
PU HUMANA PRESS INC
PI TOTOWA
PA 999 RIVERVIEW DRIVE SUITE 208, TOTOWA, NJ 07512 USA
SN 1085-9195
EI 1559-0283
J9 CELL BIOCHEM BIOPHYS
JI Cell Biochem. Biophys.
PD MAR
PY 2012
VL 62
IS 2
BP 281
EP 287
DI 10.1007/s12013-011-9292-3
PG 7
WC Biochemistry & Molecular Biology; Biophysics; Cell Biology
WE Science Citation Index Expanded (SCI-EXPANDED)
SC Biochemistry & Molecular Biology; Biophysics; Cell Biology
GA 899XI
UT WOS:000300849600003
PM 22001951
DA 2023-08-27
ER

PT J
AU 't Hart, E
   Bianco, J
   Bruin, MAC
   Derieppe, M
   Besse, HC
   Berkhout, K
   Kie, LACJ
   Su, Y
   Hoving, EW
   Huitema, ADR
   Ries, MG
   van Vuurden, DG
AF 't Hart, E.
   Bianco, J.
   Bruin, M. A. C.
   Derieppe, M.
   Besse, H. C.
   Berkhout, K.
   Kie, L. A. Chin Joe
   Su, Y.
   Hoving, E. W.
   Huitema, A. D. R.
   Ries, M. G.
   van Vuurden, D. G.
TI Radiosensitisation by olaparib through focused ultrasound delivery in a
   diffuse midline glioma model
SO JOURNAL OF CONTROLLED RELEASE
LA English
DT Article
DE Radiosensitisation; PARP1; Blood-brain barrier; Focused ultrasound
ID BRAIN-BARRIER DISRUPTION; RADIATION-THERAPY; RIBOSE POLYMERASE; PONTINE
   GLIOMAS; DRUG-DELIVERY; BREAST-CANCER; RADIOTHERAPY; COMBINATION;
   HEALTH; GROWTH
AB Background and purpose: Diffuse midline glioma H3K27-altered (DMG) is an aggressive, inoperable, predomi-nantly paediatric brain tumour. Treatment strategies are limited, resulting in a median survival of only 11 months. Currently, radiotherapy (RT), often combined with temozolomide, is considered the standard of care but remains palliative, highlighting the urgency for new therapies. Radiosensitisation by olaparib, an inhibitor of PARP1 and subsequently PAR-synthesis, is a promising treatment option. We assessed whether PARP1 inhibition enhances radiosensitivity in vitro and in vivo following focused ultrasound mediated blood-brain barrier opening (FUS-BBBO).Methods: Effects of PARP1 inhibition were evaluated in vitro using viability, clonogenic, and neurosphere assays. In vivo olaparib extravasation and pharmacokinetic profiling following FUS-BBBO was measured by LC-MS/MS. Survival benefit of FUS-BBBO combined with olaparib and RT was assessed using a patient-derived xenograft (PDX) DMG mouse model.Results: Treatment with olaparib in combination with radiation delayed tumour cell proliferation in vitro through the reduction of PAR. Prolonged exposure of low olaparib concentration was more efficient in delaying cell growth than short exposure of high concentration. FUS-BBBO increased olaparib bioavailability in the pons by 5.36-fold without observable adverse effects. A Cmax of 54.09 mu M in blood and 1.39 mu M in the pontine region was achieved following administration of 100 mg/kg olaparib. Although RT combined with FUS-BBBO mediated olaparib extravasation delayed local tumour growth, survival benefits were not observed in an in vivo DMG PDX model.Conclusions: Olaparib effectively radiosensitises DMG cells in vitro and reduces primary tumour growth in vivo when combined with RT. Further studies are needed to investigate the therapeutic benefit of olaparib in suitable preclinical PDX models.
C1 ['t Hart, E.; Bianco, J.; Derieppe, M.; Berkhout, K.; Kie, L. A. Chin Joe; Su, Y.; Hoving, E. W.; Huitema, A. D. R.; van Vuurden, D. G.] Princess Maxima Ctr Pediat Oncol, Heidelberglaan 25, NL-3584 CS Utrecht, Netherlands.
   [Bruin, M. A. C.; Huitema, A. D. R.] Antoni van Leeuwenhoek Hosp, Netherlands Canc Inst, Dept Pharm & Pharmacol, Plesmanlaan 121, NL-1066 CX Amsterdam, Netherlands.
   [Besse, H. C.; Ries, M. G.] Univ Med Ctr Utrecht, Ctr Imaging Sci, Heidelberglaan 100, NL-3584 CX Utrecht, Netherlands.
   [Huitema, A. D. R.] Univ Utrecht, Univ Med Ctr Utrecht, Dept Clin Pharm, Heidelberglaan 100, NL-3584 CX Utrecht, Netherlands.
C3 Netherlands Cancer Institute; Utrecht University; Utrecht University
   Medical Center; Utrecht University; Utrecht University Medical Center
RP van Vuurden, DG (通讯作者)，Princess Maxima Ctr Pediat Oncol, Heidelberglaan 25, NL-3584 CS Utrecht, Netherlands.
EM
RI Bianco, John/F-3448-2012
OI Bianco, John/0000-0001-5717-5125
FU KWF Young Investigator Award [KWF 10911]
FX This project was supported by KWF Young Investigator Award (KWF 10911,
   Dr. D.G. van Vuurden). The authors thank Dr. Angel Montero Carcaboso
   (Sant Joan de Deu Barcelona Hospital, Barcelona, Spain) for kindly
   providing the patient derived DMG cell lines. We also thank Dr. Wissam
   Beaino for hosting the experiments at the Radionuclide Center
   (Department of Radiology and Nuclear Medicine, Amsterdam University
   Medical Center, Amsterdam, Netherlands) , Luc Lucas for his technical
   expertise in LC-MS/MS (Bioanalytical Laboratory of the Pharmacy,
   Netherlands Cancer Institute, Amsterdam, Netherlands) and Prof. Jan
   Molenaar for providing plasmids (Princes Maxima Center for Pediatric
   Oncology, Utrecht, Netherlands) . Graphical abstract and Fig. 1 created
   with BioRender.com
CR Alli S, 2018, J CONTROL RELEASE, V281, P29, DOI 10.1016/j.jconrel.2018.05.005
   Ashworth A, 2008, J CLIN ONCOL, V26, P3785, DOI 10.1200/JCO.2008.16.0812
   ATAC MS, 1979, CLIN NEUROL NEUROSUR, V81, P281, DOI 10.1016/0303-8467(79)90032-5
   Bairoch Amos, 2018, J Biomol Tech, V29, P25, DOI 10.7171/jbt.18-2902-002
   Baxter P.A., NEUROONCOLOGY
   Burgess A, 2015, EXPERT REV NEUROTHER, V15, P477, DOI 10.1586/14737175.2015.1028369
   Chabot P, 2017, J NEURO-ONCOL, V131, P105, DOI 10.1007/s11060-016-2275-x
   Chaudhuri AR, 2017, NAT REV MOL CELL BIO, V18, P610, DOI 10.1038/nrm.2017.53
   Cho H, 2016, SCI REP-UK, V6, DOI 10.1038/srep31201
   Chornenkyy Y., MOL CANC THER
   Committee for Medicinal Products for Human Use (CHMP), 2014, CHMP ASS REP LYNP
   de Haan R, 2019, BMC CANCER, V19, DOI 10.1186/s12885-019-6121-3
   EMA, GUIDELINE BIOANALYTI
   Englander ZK, 2021, SCI REP-UK, V11, DOI 10.1038/s41598-021-85180-y
   FERRIS AM, 1989, ANN NY ACAD SCI, V561, P113, DOI 10.1111/j.1749-6632.1989.tb20975.x
   Franken NAP, 2006, NAT PROTOC, V1, P2315, DOI 10.1038/nprot.2006.339
   Gandhi K, 2022, PHARMACEUTICS, V14, DOI 10.3390/pharmaceutics14040833
   Greis C, 2004, EUR RADIOL, V14, pP11, DOI 10.1007/s10406-004-0076-3
   Hargrove D, 2006, LANCET ONCOL, V7, P241, DOI 10.1016/S1470-2045(06)70615-5
   Haumann R., 2022, TRANSL MED COMMUN, V7, P8, DOI [10, DOI 10.1186/S41231-022-00115-7]
   Haumann R, 2020, JOVE-J VIS EXP, DOI 10.3791/61269
   Hawkins BT, 2005, PHARMACOL REV, V57, P173, DOI 10.1124/pr.57.2.4
   Hoffman LM, 2018, J CLIN ONCOL, V36, P1963, DOI 10.1200/JCO.2017.75.9308
   Ishida J, 2021, J CONTROL RELEASE, V330, P1034, DOI 10.1016/j.jconrel.2020.11.010
   Jannetti S.A., J NUCL MED
   Jansen MH, 2015, NEURO-ONCOLOGY, V17, P160, DOI 10.1093/neuonc/nou104
   Jansen MHA, 2016, MOL CANCER THER, V15, P2166, DOI 10.1158/1535-7163.MCT-15-0558
   Kilkenny C, 2012, OSTEOARTHR CARTILAGE, V20, P256, DOI 10.1016/j.joca.2012.02.010
   LASSMAN LP, 1967, LANCET, V1, P913
   Lesueur P, 2017, ONCOTARGET, V8, P69105, DOI 10.18632/oncotarget.19079
   Loap P, 2021, INT J RADIAT ONCOL, V109, P436, DOI 10.1016/j.ijrobp.2020.09.032
   Louis DN, 2016, ACTA NEUROPATHOL, V131, P803, DOI 10.1007/s00401-016-1545-1
   Kholosy WM, 2021, J PERS MED, V11, DOI 10.3390/jpm11090869
   Mackay A, 2017, CANCER CELL, V32, P520, DOI 10.1016/j.ccell.2017.08.017
   Mehta MP, 2012, INT J RADIAT ONCOL, V84, pS269, DOI 10.1016/j.ijrobp.2012.07.702
   Menezes MCS, 2022, CANCERS, V14, DOI 10.3390/cancers14174332
   Michmerhuizen AR, 2019, MOL CANCER THER, V18, P2063, DOI 10.1158/1535-7163.MCT-19-0520
   Niepel Mario, 2017, Curr Protoc Chem Biol, V9, P55, DOI 10.1002/cpch.21
   Nijenhuis CM, 2013, J CHROMATOGR B, V940, P121, DOI 10.1016/j.jchromb.2013.09.020
   Ostrom QT, 2014, NEURO-ONCOLOGY, V16, P1, DOI [10.1093/neuonc/nov189, 10.1093/neuonc/nou327, 10.1093/neuonc/nou223]
   Pardridge WM, 2007, DRUG DISCOV TODAY, V12, P54, DOI 10.1016/j.drudis.2006.10.013
   Park J, 2012, J CONTROL RELEASE, V162, P134, DOI 10.1016/j.jconrel.2012.06.012
   Pedersen H, 2020, CANCERS, V12, DOI 10.3390/cancers12102813
   Qi L, 2020, CANCER LETT, V493, P197, DOI 10.1016/j.canlet.2020.08.035
   Robson M, 2017, NEW ENGL J MED, V377, P523, DOI 10.1056/NEJMoa1706450
   Rudolph J, 2022, P NATL ACAD SCI USA, V119, DOI 10.1073/pnas.2121979119
   Sa-Pereira I, 2012, MOL NEUROBIOL, V45, P327, DOI 10.1007/s12035-012-8244-2
   Schneider CA, 2012, NAT METHODS, V9, P671, DOI 10.1038/nmeth.2089
   Senra JM, 2011, MOL CANCER THER, V10, P1949, DOI 10.1158/1535-7163.MCT-11-0278
   Sheikov N, 2004, ULTRASOUND MED BIOL, V30, P979, DOI 10.1016/j.ultrasmedbio.2004.04.010
   Sheikov N, 2008, ULTRASOUND MED BIOL, V34, P1093, DOI 10.1016/j.ultrasmedbio.2007.12.015
   Sheikov N, 2006, ULTRASOUND MED BIOL, V32, P1399, DOI 10.1016/j.ultrasmedbio.2006.05.015
   Sim HW, 2021, NEURO-ONCOLOGY, V23, P1736, DOI 10.1093/neuonc/noab111
   Spina R, 2018, JOVE-J VIS EXP, DOI 10.3791/56176
   Stupp R, 2009, LANCET ONCOL, V10, P459, DOI 10.1016/S1470-2045(09)70025-7
   Su GS, 2020, BIOMED CHROMATOGR, V34, DOI 10.1002/bmc.4791
   Sun Kaiming, 2018, Oncotarget, V9, P37080, DOI 10.18632/oncotarget.26354
   t Hart E., BIOMEDICINES
   Todd N, 2018, NEUROIMAGE, V178, P414, DOI 10.1016/j.neuroimage.2018.05.063
   U.F. and D.A. (FDA), GUID IND BIOAN METH
   van Vuurden DG, 2011, ONCOTARGET, V2, P984, DOI 10.18632/oncotarget.362
   Waissi W, 2021, CANCERS, V13, DOI 10.3390/cancers13030527
   Warren KE, 2018, FRONT ONCOL, V8, DOI 10.3389/fonc.2018.00239
   Xiong Y, 2020, NEOPLASIA, V22, P431, DOI 10.1016/j.neo.2020.06.009
   Ye DZ, 2018, J CONTROL RELEASE, V286, P145, DOI 10.1016/j.jconrel.2018.07.020
   Zarghooni M, 2010, J CLIN ONCOL, V28, P1337, DOI 10.1200/JCO.2009.25.5463
NR 66
TC 0
Z9 0
U1 1
U2 1
PU ELSEVIER
PI AMSTERDAM
PA RADARWEG 29, 1043 NX AMSTERDAM, NETHERLANDS
SN 0168-3659
EI 1873-4995
J9 J CONTROL RELEASE
JI J. Control. Release
PD MAY
PY 2023
VL 357
BP 287
EP 298
DI 10.1016/j.jconrel.2023.03.058
EA APR 2023
PG 12
WC Chemistry, Multidisciplinary; Pharmacology & Pharmacy
WE Science Citation Index Expanded (SCI-EXPANDED)
SC Chemistry; Pharmacology & Pharmacy
GA E5YU2
UT WOS:000976304200001
PM 37019285
OA hybrid
DA 2023-08-27
ER

PT J
AU Quester, R
   Schroder, R
AF Quester, R
   Schroder, R
TI Topographic anatomy of the cochlear nuclear region at the floor of the
   fourth ventricle in humans
SO JOURNAL OF NEUROSURGERY
LA English
DT Article
DE anatomical study; cochlear nucleus region; foramen of Luschka; fourth
   ventricle; interindividual and intraindividual variability lateral
   recess
ID AUDITORY BRAIN-STEM; 4TH VENTRICLE; LANDMARKS; HEARING; COMPLEX
AB Object. The development of appropriate methods to stimulate the dorsal and ventral cochlear nucleus by means of an auditory brainstem implant in patients with acquired bilateral anacusis requires a detailed topoanatomical knowledge both of the location and extension of the nuclear surface in the fourth ventricle and lateral recess and of its variability. The goal of this study was to provide that information. Anatomically, it is possible to use a midline surgical approach to the fourth ventricle rather than the translabyrinthine and suboccipital routes of access used hitherto. This is especial ly useful if severe scarring, which occurs as a result of tumor removal in the cerebellopontine angle, make the orientation and placement of an auditory brainstem implant via a lateral surgical approach difficult. There have been only a few studies, involving single cases and small series of patients, in which the focus was the exact extension of the cochlear nuclei, whose microsurgically relevant position in relation to the surface structures is not known in detail.
   Methods. Landmarks that are important for the placement of an auditory brainstem implant through the fourth ventricle were examined and measured in a large series of 28 formalin-fixed human brainstems. In all cases, these examinations were supplemented by addition of a histological section series. For the first time values of unfixed fresh brainstem tissue were determined. Anatomical features are discussed with regard to their possible neurosurgical relevance, taking into account inter- and intraindividual variability.
   Conclusions. The midline approach would provide an opportunity to stimulate the whole area of the dorsal as well as the ventral cochlear nucleus with an auditory brainstem implant.
C1 Univ Cologne, Abt Stereotaxie & Funkt Neurochirurg, Neurochirurg Klin, D-50931 Cologne, Germany.
   Univ Cologne, Dept Neuropathol, D-50931 Cologne, Germany.
C3 University of Cologne; University of Cologne
RP Quester, R (通讯作者)，Univ Cologne, Abt Stereotaxie & Funkt Neurochirurg, Neurochirurg Klin, Joseph Stelzmann Str 9, D-50931 Cologne, Germany.
CR Bogucki J, 1997, ACTA NEUROCHIR, V139, P1014, DOI 10.1007/BF01411553
   BRACKMANN DE, 1993, OTOLARYNG HEAD NECK, V108, P624, DOI 10.1177/019459989310800602
   DIAMOND IT, 1973, ARCH OTOLARYNGOL, V98, P397
   DUBLIN WB, 1982, OTOLARYNG HEAD NECK, V90, P744, DOI 10.1177/019459988209000613
   FUSE G, 1913, ARB HIRNANAT I ZURIC, V7, P1
   JACOB U, 1991, ANAT ANZEIGER, V173, P93
   KUROKI A, 1995, J NEUROSURG, V82, P933, DOI 10.3171/jns.1995.82.6.0933
   Lagerlof H, 1934, J PSYCHOL NEUROL, V46, P178
   LANG J, 1991, ACTA NEUROCHIR, V113, P84, DOI 10.1007/BF01402120
   LANG J, 1981, CRANIAL NERVES, P363
   LANG J, 1979, PRAKTISCHE ANATOM 1B, V1
   LANG J, 1979, PRAKTISCHE ANATO 1A, V1
   LAST RJ, 1953, BRIT J SURG, V40, P525, DOI 10.1002/bjs.18004016403
   LASZIG R, 1995, J LARYNGOL OTOL, V109, P385, DOI 10.1017/S0022215100130257
   Laszig R, 1994, Ann Otolaryngol Chir Cervicofac, V111, P315
   LUETJE CM, 1992, LARYNGOSCOPE, V102, P23
   MATSUSHIMA T, 1982, NEUROSURGERY, V11, P631, DOI 10.1227/00006123-198211000-00008
   MATSUSHIMA T, 1996, NEUROSURGERY, V1, P1155
   MCELVEEN JT, 1987, OTOLARYNG HEAD NECK, V96, P135, DOI 10.1177/019459988709600204
   MCELVEEN JT, 1985, AM J OTOL S, V6, P88
   MONSELL EM, 1987, AM J OTOL, V8, P450
   MOORE JK, 1987, HEARING RES, V29, P1, DOI 10.1016/0378-5955(87)90202-4
   MOORE JK, 1979, AM J ANAT, V154, P393, DOI 10.1002/aja.1001540306
   Olszewski J., 1954, CYTOARCHITECTURE HUM
   Quester R, 1997, J NEUROSCI METH, V75, P81, DOI 10.1016/S0165-0270(97)00050-2
   Ramon y Cajal S, 1909, HISTOLOGIE SYSTEME N, P774
   RHOTON AL, 1996, NEUROSURGERY, V1, P1063
   Rhoton AL, 1993, SURG CRANIAL NERVES, P1
   ROBERTSON EG, 1941, ENCEPHALOGRAPHY
   SELDON HL, 1991, BRAIN RES, V551, P185, DOI 10.1016/0006-8993(91)90932-L
   STENZEL CR, 1992, THESIS U COLOGNE
   TERR LI, 1985, HEARING RES, V17, P51, DOI 10.1016/0378-5955(85)90130-3
   TERR LI, 1987, LARYNGOSCOPE, V97, P1009
   TERR LI, 1990, OTOLARYNG HEAD NECK, V102, P717, DOI 10.1177/019459989010200615
   TERR LI, 1985, ARCH OTOLARYNGOL, V111, P495
   TREFF W M, 1960, Psychiatr Neurol Neurochir, V63, P116
NR 36
TC 18
Z9 20
U1 0
U2 0
PU AMER ASSOC NEUROLOGICAL SURGEONS
PI CHARLOTTESVILLE
PA UNIV VIRGINIA, 1224 WEST MAIN ST, STE 450, CHARLOTTESVILLE, VA 22903 USA
SN 0022-3085
J9 J NEUROSURG
JI J. Neurosurg.
PD SEP
PY 1999
VL 91
IS 3
BP 466
EP 476
DI 10.3171/jns.1999.91.3.0466
PG 11
WC Clinical Neurology; Surgery
WE Science Citation Index Expanded (SCI-EXPANDED)
SC Neurosciences & Neurology; Surgery
GA 232WN
UT WOS:000082393100017
PM 10470823
DA 2023-08-27
ER

PT J
AU Mori, Y
   Hashizume, C
   Shibamoto, Y
   Kobayashi, T
   Nakazawa, H
   Hagiwara, M
   Tsugawa, T
AF Mori, Yoshimasa
   Hashizume, Chisa
   Shibamoto, Yuta
   Kobayashi, Tatsuya
   Nakazawa, Hisato
   Hagiwara, Masahiro
   Tsugawa, Takahiko
TI STEREOTACTIC RADIOTHERAPY FOR SPINAL INTRADURAL METASTASES DEVELOPING
   WITHIN OR ADJACENT TO THE PREVIOUS IRRADIATION FIELD - REPORT OF THREE
   CASES
SO NAGOYA JOURNAL OF MEDICAL SCIENCE
LA English
DT Article
DE IMRT; Intradural; Metastasis; Spine; Stereotactic radiotherapy
ID RADIATION TOLERANCE; CLINICAL-EXPERIENCE; CORD METASTASIS; RADIOSURGERY;
   REIRRADIATION; LESIONS
AB Results of stereotactic radiotherapy (SRT) for spinal intradural metastases developing inside or adjacent to the previous external-beam radiation therapy (EBRT) field are shown in 3 cases. One case of spinal intramedullary metastasis and two cases of intradural extramedullary metastases were treated using a Novalis shaped-beam SRT. Case 1 developed an intramedullary metastatic tumor in the C1 spinal medulla inside the previous whole brain EBRT field and another lesion adjacent to the field in the C2 spinal medulla. Case 2 developed intradural extramedullary metastasis around C6-8 inside the previous EBRT field for the primary lung adenocarcinoma. Case 3 developed multiple spinal intradural extramedullary metastatic deposits after surgical resection and following whole brain EBRT for brain metastasis. We delivered 24 to 36 Gy in 5 to 12 fractions. The treated tumors were stable or decreased in size until the patients' death from the primary cancer (10, 22, and 5 months). Neurological symptoms were stable or improved in all 3 patients. Palliative SRT using Novalis is expected to be safe and effective even if the patient develops spinal intradural metastases within or adjacent to the previous irradiation field.
C1 [Mori, Yoshimasa; Shibamoto, Yuta] Nagoya City Univ, Dept Radiol & Radiat Oncol, Grad Sch Med Sci, Nagoya, Aichi 4678601, Japan.
   [Hashizume, Chisa; Kobayashi, Tatsuya; Nakazawa, Hisato; Hagiwara, Masahiro; Tsugawa, Takahiko] Nagoya Kyoritsu Hosp, Nagoya Radiosurg Ctr, Nagoya, Aichi, Japan.
C3 Nagoya City University
RP Mori, Y (通讯作者)，Nagoya City Univ, Dept Radiol & Radiat Oncol, Grad Sch Med Sci, Mizuho Ku, 1 Kawasumi,Mizuho Cho, Nagoya, Aichi 4678601, Japan.
EM
CR Ang K K, 1994, Oncology (Williston Park), V8, P71
   Benzil DL, 2004, J NEUROSURG, V101, P413, DOI 10.3171/sup.2004.101.supplement3.0413
   De Salles AAF, 2004, J NEUROSURG, V101, P435, DOI 10.3171/sup.2004.101.supplement3.0435
   Endo S, 2008, NEUROL SURG TOKYO, V36, P345
   Gerszten PC, 2004, NEUROSURGERY, V55, P89, DOI 10.1227/01.NEU.0000126873.00845.A7
   GERSZTEN PC, SPINE, V32, P193
   Hashii H, 2011, J RADIAT RES, V52, P641, DOI 10.1269/jrr.10187
   Milano MT, 2011, CANCER TREAT REV, V37, P567, DOI 10.1016/j.ctrv.2011.04.004
   Mizumoto M, 2011, INT J RADIAT ONCOL, V79, P208, DOI 10.1016/j.ijrobp.2009.10.056
   Ryu S, 2000, AM J CLIN ONCOL-CANC, V23, P29, DOI 10.1097/00000421-200002000-00007
   RYU S, 2007, CANCER, V109, P626
   Ryu S, 2008, J PAIN SYMPTOM MANAG, V35, P292, DOI 10.1016/j.jpainsymman.2007.04.020
   Ryu SI, 2001, NEUROSURGERY, V49, P838, DOI 10.1097/00006123-200110000-00011
   Sahgal A, 2012, INT J RADIAT ONCOL, V82, P107, DOI 10.1016/j.ijrobp.2010.08.021
   Sahgal A, 2010, INT J RADIAT ONCOL, V77, P548, DOI 10.1016/j.ijrobp.2009.05.023
   SCHULTHEISS TE, 1990, INT J RADIAT ONCOL, V19, P219, DOI 10.1016/0360-3016(90)90157-F
   Shin DA, 2009, NEUROSURG FOCUS, V27, DOI 10.3171/2009.9.FOCUS09194
   WARA WM, 1975, CANCER, V35, P1558, DOI 10.1002/1097-0142(197506)35:6<1558::AID-CNCR2820350613>3.0.CO;2-7
   WONG CS, 1993, RADIOTHER ONCOL, V26, P132
NR 19
TC 5
Z9 6
U1 0
U2 0
PU NAGOYA UNIV, SCH MED
PI NAGOYA
PA EDITORIAL OFF, NAGOYA UNIV MED LIB, SCH MED, 65 TSURUMAI-CHO SHOWA- KU,
   NAGOYA, 466-8550, JAPAN
SN 2186-3326
EI 0027-7622
J9 NAGOYA J MED SCI
JI Nagoya J. Med. Sci.
PD AUG
PY 2013
VL 75
IS 3-4
BP 263
EP 271
PG 9
WC Medicine, Research & Experimental
WE Science Citation Index Expanded (SCI-EXPANDED)
SC Research & Experimental Medicine
GA 206WQ
UT WOS:000323556700012
PM 24640183
DA 2023-08-27
ER

PT J
AU Su, CH
   Young, YH
AF Su, Chia-Hung
   Young, Yi-Ho
TI Disorders Affecting the Fourth Ventricle: Etiology and Clinical
   Correlates
SO OTOLOGY & NEUROTOLOGY
LA English
DT Article
DE Ataxia; Fourth ventricle; Nystagmus; Ocular and cervical
   vestibular-evoked myogenic potential; Vomiting
ID EVOKED MYOGENIC POTENTIALS; VISUAL-VESTIBULAR INTERACTION; 4TH
   VENTRICLE; EPIDERMOID TUMORS; REBOUND NYSTAGMUS; SUPPRESSION; FLOCCULUS;
   SURGERY; LESIONS; REFLEX
AB Objective: The fourth ventricle encompasses many vital structures including the brainstem as its floor and the cerebellum as its lateral wall and roof. Therefore, lesions affecting the fourth ventricle may present as cerebellar or brainstem manifestations. Herein, we presented our experience in the diagnosis of disorders affecting the fourth ventricle during the past 15 years.
   Study Design: Retrospective study.
   Setting: University hospital.
   Subjects: From September 1995 to August 2010, 24,838 patients with vertigo/dizziness visited the clinic. Of them, 13 patients (0.5%) had tumor/stroke affecting the fourth ventricle.
   Main Outcome Measures: All patients underwent a battery of audiovestibular function test.
   Results: All patients had vertigo (100%) and then ataxia (92%) and nausea/vomiting (85%). Eight patients (68%) displayed persistent nystagmus. Most (> 75%) patients showed central signs in electronystagmographic recordings and abnormal caloric coupled with visual suppression test. Three patients underwent both ocular and cervical vestibular-evoked myogenic potential tests, and abnormal results were noted in all 3 patients indicating a brainstem involvement. However, magnetic resonance imaging failed to demonstrate brainstem lesion in these 3 patients. Final diagnoses consisted of the following: cavernoma, n (-) 3; metastatic cancer, n = 3; astrocytoma, n = 2; meningioma, n = 2; epidermoid cyst, n = 1; ependymoma, n = 1; and lymphoma, n = 1. The prognosis was poor as 10 patients (77%) died within 2 years.
   Conclusion: When a vertiginous patient displayed ataxia, persistent vomiting, and persistent nystagmus, lesion affecting the fourth ventricle should be kept in mind because its prognosis is poor. Abnormal results in electronystagmography and in ocular and cervical vestibular-evoked myogenic potentials may serve as a supplementary to magnetic resonance imaging to delineate the involvement of the lesion.
C1 [Young, Yi-Ho] Natl Taiwan Univ Hosp, Dept Otolaryngol, Taipei, Taiwan.
   [Su, Chia-Hung] Fu Jen Catholic Univ, Catholic Cardinal Tien Hosp, Dept Otolaryngol, Taipei, Taiwan.
C3 National Taiwan University; National Taiwan University Hospital; Fu Jen
   Catholic University
RP Young, YH (通讯作者)，Natl Taiwan Univ Hosp, Dept Otolaryngol, 1 Chang Te St, Taipei, Taiwan.
EM
RI Su, Chia-Hung/C-2807-2008
FU National Science Council, Taipei, Taiwan [NSC 99-2314-B002-049-MY3]
FX This study was supported Grant NSC 99-2314-B002-049-MY3 from the
   National Science Council, Taipei, Taiwan.
CR BALOH RW, 1979, NEUROLOGY, V29, P116, DOI 10.1212/WNL.29.1.116
   BARLOON TJ, 1989, MAGN RESON IMAGING, V7, P635, DOI 10.1016/0730-725X(89)90532-8
   Campbell JW, 1996, J NEURO-ONCOL, V28, P223, DOI 10.1007/bf00250201
   Chang CH, 2007, CLIN NEUROPHYSIOL, V118, P1105, DOI 10.1016/j.clinph.2007.01.020
   Chen CC, 2006, LARYNGOSCOPE, V116, P1678, DOI 10.1097/01.mlg.0000233507.13293.d9
   Craig WM, 1938, J AMER MED ASSOC, V111, P2370, DOI 10.1001/jama.1938.02790520026006
   Curthoys IS, 2010, CLIN NEUROPHYSIOL, V121, P132, DOI 10.1016/j.clinph.2009.09.027
   FIUME D, 1988, SURG NEUROL, V29, P178, DOI 10.1016/0090-3019(88)90002-X
   Gianna CC, 2000, J VESTIBUL RES-EQUIL, V10, P227
   HOOD JD, 1973, BRAIN, V96, P507, DOI 10.1093/brain/96.3.507
   KATSARKAS A, 1981, LARYNGOSCOPE, V91, P71
   Lin CM, 2010, OTOL NEUROTOL, V31, P157, DOI 10.1097/MAO.0b013e3181c2a0e1
   Lin CY, 1999, LARYNGOSCOPE, V109, P1803, DOI 10.1097/00005537-199911000-00015
   MATSUSHIMA T, 1982, NEUROSURGERY, V11, P631, DOI 10.1227/00006123-198211000-00008
   NEMOTO Y, 1989, J COMPUT ASSIST TOMO, V13, P769, DOI 10.1097/00004728-198909000-00004
   Ohue S, 2010, NEUROSURG REV, V33, P315, DOI 10.1007/s10143-010-0256-7
   Price CJ, 2008, NEUROSCIENTIST, V14, P182, DOI 10.1177/1073858407311100
   ROSARIO M, 1981, NEUROSURGERY, V9, P9, DOI 10.1227/00006123-198107000-00003
   SHARPE JA, 1981, NEUROLOGY, V31, P427
   Su CH, 2011, EUR ARCH OTO-RHINO-L, V268, P923, DOI 10.1007/s00405-010-1463-0
   TAKEMORI S, 1974, BRAIN RES, V72, P213, DOI 10.1016/0006-8993(74)90860-9
   Talacchi A, 1998, NEUROSURGERY, V42, P242, DOI 10.1097/00006123-199802000-00020
   Tancredi A, 2003, ACTA NEUROCHIR, V145, P905, DOI 10.1007/s00701-003-0008-3
   Tseng CL, 2010, OTOL NEUROTOL, V31, P959, DOI 10.1097/MAO.0b013e3181e8fb1a
   Tseng CL, 2010, EUR ARCH OTO-RHINO-L, V267, P191, DOI 10.1007/s00405-009-1025-5
   VIBERT N, 1993, EXP BRAIN RES, V97, P263, DOI 10.1007/BF00228695
   Wang SJ, 2010, EAR HEARING, V31, P283, DOI 10.1097/AUD.0b013e3181bdbac0
   Yasargil MG, 2008, NEUROSURGERY, V62, P1029, DOI [10.1227/01.NEU.0000316427.57165.01, 10.1227/01.neu.0000333768.12951.9a]
   Young YH, 2001, LARYNGOSCOPE, V111, P1490, DOI 10.1097/00005537-200108000-00029
   ZEE DS, 1981, J NEUROPHYSIOL, V46, P878, DOI 10.1152/jn.1981.46.4.878
NR 30
TC 11
Z9 12
U1 0
U2 0
PU LIPPINCOTT WILLIAMS & WILKINS
PI PHILADELPHIA
PA 530 WALNUT ST, PHILADELPHIA, PA 19106-3621 USA
SN 1531-7129
J9 OTOL NEUROTOL
JI Otol. Neurotol.
PD OCT
PY 2011
VL 32
IS 8
BP 1329
EP 1335
DI 10.1097/MAO.0b013e31822e5ba7
PG 7
WC Clinical Neurology; Otorhinolaryngology
WE Science Citation Index Expanded (SCI-EXPANDED)
SC Neurosciences & Neurology; Otorhinolaryngology
GA 821BD
UT WOS:000294948900028
PM 21897323
DA 2023-08-27
ER

PT J
AU Msaouel, P
   Tannir, NM
   Walker, CL
AF Msaouel, Pavlos
   Tannir, Nizar M.
   Walker, Cheryl Lyn
TI A Model Linking Sickle Cell Hemoglobinopathies and SMARCB1 Loss in Renal
   Medullary Carcinoma
SO CLINICAL CANCER RESEARCH
LA English
DT Article
ID DOUBLE-STRAND BREAKS; HIGH NACL; CHROMOSOMAL TRANSLOCATIONS; DNA BREAKS;
   DISEASE; REPAIR; TRAIT; NEPHROPATHY; MECHANISMS; DELETIONS
AB Renal medullary carcinoma (RMC) is a highly aggressive malignancy that predominantly afflicts young adults and adolescents with sickle hemoglobinopathies. It is characterized by complete loss of expression of the chromatin remodeler and tumor suppressor SMARCB1. Despite therapy, the outcomes of patients with RMC remain very poor, highlighting the need to understand the etiology of this cancer, and develop new diagnostic, preventive, and therapeutic strategies. A key knowledge gap in RMC biology is why sickle hemoglobinopathies predispose to the development of this cancer. We propose a model wherein the extreme conditions of hypoxia and hypertonicity of the renal medulla, combined with regional ischemia induced by red blood cell sickling, activate DNA repair mechanisms to drive deletions and translocations in SMARCB1, which is localized in a fragile region of chromosome 22. This mechanism would explain the linkage between RMC and sickle hemoglobinopathies, as well as the age dependence and predilection of RMC toward the right kidney.
   Significance: This perspective proposes an integrated and testable model of renal medullary carcinoma pathogenesis. Insights provided by this model can additionally inform other malignancies arising from the renal medulla and/or associated with loss of the SMARCB1 tumor suppressor gene. (C) 2018 AACR.
C1 [Msaouel, Pavlos; Tannir, Nizar M.] Univ Texas MD Anderson Canc Ctr, Dept Genitourinary Med Oncol, Houston, TX 77030 USA.
   [Msaouel, Pavlos; Walker, Cheryl Lyn] Baylor Coll Med, Dept Mol & Cell Biol, Ctr Precis Environm Hlth, Houston, TX 77030 USA.
C3 University of Texas System; UTMD Anderson Cancer Center; Baylor College
   of Medicine
RP Walker, CL (通讯作者)，Baylor Coll Med, Dept Cellular & Mol Biol, Ctr Precis Environm Hlth, Houston, TX 77030 USA.; Walker, CL (通讯作者)，Baylor Coll Med, Dept Med, Ctr Precis Environm Hlth, Houston, TX 77030 USA.; Tannir, NM (通讯作者)，Univ Texas MD Anderson Canc Ctr, Dept Genitourinary Med Oncol, Unit 1374, 1155 Pressler St, Houston, TX 77030 USA.
EM
FU Conquer Cancer Foundation Young Investigator Award;  [NIHT32 CA009666]
FX The authors thank Sarah E. Townsend for critical reading of the
   manuscript and Cora Connor for helpful suggestions on the content. P.
   Msaouel is supported for this research by the NIHT32 CA009666 and a
   Conquer Cancer Foundation Young Investigator Award.
CR ABEL MS, 1948, JAMA-J AM MED ASSOC, V136, P624, DOI 10.1001/jama.1948.72890260003008a
   Alvarez O, 2015, PEDIATR BLOOD CANCER, V62, P1694, DOI 10.1002/pbc.25592
   Anazoeze M, 2016, AFR HEALTH SCI, V16, P490, DOI 10.4314/ahs.v16i2.17
   Beckermann KE, 2017, J ONCOL PRACT, V13, P414, DOI 10.1200/JOP.2017.020909
   Bemanian Shahrooz, 2005, BMC Nephrol, V6, P10
   BREZIS M, 1995, NEW ENGL J MED, V332, P647, DOI 10.1056/NEJM199503093321006
   Bunn HF., 1986, HEMOGLOBIN MOL GENET
   Burnside RD, 2015, CYTOGENET GENOME RES, V146, P89, DOI 10.1159/000438708
   Calderaro J, 2016, EUR UROL, V69, P1055, DOI 10.1016/j.eururo.2015.09.027
   Carlo MI, 2016, J CLIN ONCOL, V34, DOI 10.1200/JCO.2016.34.15_suppl.4556
   DAVIS CJ, 1995, AM J SURG PATHOL, V19, P1, DOI 10.1097/00000478-199501000-00001
   Debelenko LV, 2011, MODERN PATHOL, V24, P430, DOI 10.1038/modpathol.2010.213
   Dmitrieva NI, 2004, CELL CYCLE, V3, P561
   Dmitrieva NI, 2004, P NATL ACAD SCI USA, V101, P2317, DOI 10.1073/pnas.0308463100
   Dmitrieva NI, 2003, AM J PHYSIOL-RENAL, V285, pF266, DOI 10.1152/ajprenal.00060.2003
   Ghezraoui H, 2014, MOL CELL, V55, P829, DOI 10.1016/j.molcel.2014.08.002
   Gladwin MT, 2003, CIRCULATION, V107, P271, DOI 10.1161/01.CIR.0000044943.12533.A8
   Hastings PJ, 2009, NAT REV GENET, V10, P551, DOI 10.1038/nrg2593
   Hodges C, 2016, CSH PERSPECT MED, V6, DOI 10.1101/cshperspect.a026930
   Hollmann TJ, 2011, AM J SURG PATHOL, V35, pE47, DOI 10.1097/PAS.0b013e31822b325b
   Huang LE, 2007, J MOL MED, V85, P139, DOI 10.1007/s00109-006-0133-6
   Iliakis G, 2015, MUTAT RES-GEN TOX EN, V793, P166, DOI 10.1016/j.mrgentox.2015.07.001
   Jackson EM, 2009, CLIN CANCER RES, V15, P1923, DOI 10.1158/1078-0432.CCR-08-2091
   Kang ZJ, 2016, CHIN J CANCER, V35, DOI 10.1186/s40880-016-0108-0
   Kato T, 2012, CURR OPIN GENET DEV, V22, P221, DOI 10.1016/j.gde.2012.02.004
   Mani RS, 2010, NAT REV GENET, V11, P819, DOI 10.1038/nrg2883
   Marino-Enriquez A, 2011, GENE CHROMOSOME CANC, V50, P146, DOI 10.1002/gcc.20839
   McDonald-McGinn DM, 2015, NAT REV DIS PRIMERS, V1, DOI 10.1038/nrdp.2015.71
   MERKLIN R J, 1958, J Int Coll Surg, V29, P41
   MOSTOFI FK, 1957, ARCH PATHOL, V63, P336
   Nahm AM, 2000, NEPHROL DIAL TRANSPL, V15, P549, DOI 10.1093/ndt/15.4.549
   Nickoloff JA, 2017, JNCI-J NATL CANCER I, V109, DOI 10.1093/jnci/djx059
   PERILLIE PE, 1963, J CLIN INVEST, V42, P570, DOI 10.1172/JCI104746
   PFITZNER J, 1976, ANAESTHESIA, V31, P273, DOI 10.1111/j.1365-2044.1976.tb11804.x
   PLATT OS, 1994, NEW ENGL J MED, V330, P1639, DOI 10.1056/NEJM199406093302303
   Redon CE, 2011, P NATL ACAD SCI USA, V108, P20281, DOI 10.1073/pnas.1117713109
   Sands JM, 2009, SEMIN NEPHROL, V29, P178, DOI 10.1016/j.semnephrol.2009.03.008
   Seo AN, 2017, ADV ANAT PATHOL, V24, P65, DOI 10.1097/PAP.0000000000000138
   Shah AY, 2017, BJU INT, V120, P782, DOI 10.1111/bju.13705
   Sirohi D, 2017, HUM PATHOL, V67, P134, DOI 10.1016/j.humpath.2017.07.006
   Sishc BJ, 2017, CANCERS, V9, DOI 10.3390/cancers9070081
   Smith NE, 2014, AM J SURG PATHOL, V38, P858, DOI 10.1097/PAS.0000000000000179
   Statius van Eps L W, 1970, Clin Chim Acta, V27, P501
   Sullivan LM, 2013, MODERN PATHOL, V26, P385, DOI 10.1038/modpathol.2012.175
   Tsaras G, 2009, AM J MED, V122, P507, DOI 10.1016/j.amjmed.2008.12.020
   Wang ML, 2006, NUCLEIC ACIDS RES, V34, P6170, DOI 10.1093/nar/gkl840
NR 46
TC 38
Z9 38
U1 0
U2 4
PU AMER ASSOC CANCER RESEARCH
PI PHILADELPHIA
PA 615 CHESTNUT ST, 17TH FLOOR, PHILADELPHIA, PA 19106-4404 USA
SN 1078-0432
EI 1557-3265
J9 CLIN CANCER RES
JI Clin. Cancer Res.
PD MAY 1
PY 2018
VL 24
IS 9
BP 2044
EP 2049
DI 10.1158/1078-0432.CCR-17-3296
PG 6
WC Oncology
WE Science Citation Index Expanded (SCI-EXPANDED)
SC Oncology
GA GE5YN
UT WOS:000431300500004
PM 29440190
OA Bronze
DA 2023-08-27
ER

PT J
AU Bowers, DC
   Georgiades, C
   Aronson, LJ
   Carson, BS
   Weingart, JD
   Wharam, MD
   Melhem, ER
   Burger, PC
   Cohen, KJ
AF Bowers, DC
   Georgiades, C
   Aronson, LJ
   Carson, BS
   Weingart, JD
   Wharam, MD
   Melhem, ER
   Burger, PC
   Cohen, KJ
TI Tectal gliomas: Natural history of an indolent lesion in pediatric
   patients
SO PEDIATRIC NEUROSURGERY
LA English
DT Article
DE tectal glioma; low-grade glioma; tumor progression
ID BRAIN-STEM GLIOMAS; LOW-GRADE GLIOMAS; AQUEDUCTAL STENOSIS; MIDBRAIN
   TUMORS; CHILDREN; MANAGEMENT; RADIOTHERAPY; THERAPY
AB The mesencephalic tectal glioma is a distinctive form of brain stem glioma with an unusually benign clinical course. Periaqueductal location, lack of contrast enhancement, and long periods of stability are classic features. The clinical management of these lesions, especially at the time of radiographic enlargement varies widely in the published literature. It is unclear whether these progressive lesions need to be treated. Accordingly, clinical and radiologic features of 7 patients were reviewed, with attention to the clinical course of the disease after radiologic enlargement. The age at diagnosis ranged from 3.3 to 16.6 years. Six of 7 had MRI tumor enlargement beginning 0.3-5.7 years after initial diagnosis. One of these 6 patients had radiographic progression coupled with a new clinical symptom which was treated with stereotactic radiation therapy. The remaining 5 patients with MRI progression and normal neurological exams were not treated and remain free of new neurologic deficits 1.8-6.9 years after the first radiographic tumor enlargement. The results suggest that pediatric tectal gliomas are a very low-grade lesion. Conservative management in the absence of new clinical symptoms could be argued, reserving radiotherapy or chemotherapy for clinical progression. Copyright (C) 2000 S. Karger AG. Basel.
C1 Johns Hopkins Univ, Sch Med, Johns Hopkins Oncol Ctr, Baltimore, MD 21205 USA.
   Johns Hopkins Univ, Sch Med, Dept Pediat, Baltimore, MD 21205 USA.
   Johns Hopkins Univ, Sch Med, Dept Radiol, Baltimore, MD 21205 USA.
   Johns Hopkins Univ, Sch Med, Dept Pathol, Baltimore, MD 21205 USA.
   Johns Hopkins Univ, Sch Med, Dept Neurosurg, Baltimore, MD 21205 USA.
C3 Johns Hopkins University; Johns Hopkins Medicine; Johns Hopkins
   University; Johns Hopkins University; Johns Hopkins University; Johns
   Hopkins University
RP Cohen, KJ (通讯作者)，Johns Hopkins Univ Hosp, CMSC-800,600 N Wolfe St, Baltimore, MD 21287 USA.
EM
RI Melhem, Elias R/E-5205-2013
OI Bowers, Daniel/0000-0002-3947-2481
CR BARKOVICH AJ, 1989, AM J NEURORADIOL, V10, P471
   BOYDSTON WR, 1992, PEDIATR NEUROSURG, V17, P234, DOI 10.1159/000120603
   EDWARDS MSB, 1989, J NEUROSURG, V70, P691, DOI 10.3171/jns.1989.70.5.0691
   FREEMAN CR, 1993, INT J RADIAT ONCOL, V27, P197, DOI 10.1016/0360-3016(93)90228-N
   Hamilton MG, 1996, CAN J NEUROL SCI, V23, P204, DOI 10.1017/S031716710003852X
   HOFFMAN HJ, 1980, NEUROSURGERY, V7, P243, DOI 10.1227/00006123-198009000-00007
   KHATIB ZA, 1994, PEDIATR NEUROSURG, V20, P2, DOI 10.1159/000120759
   KIHLSTROM L, 1994, ACT NEUR S, V62, P55
   Levin V A, 1996, Curr Opin Oncol, V8, P175
   MARIA BL, 1993, J CHILD NEUROL, V8, P112, DOI 10.1177/088307389300800203
   MAY PL, 1991, J NEUROSURG, V74, P867, DOI 10.3171/jns.1991.74.6.0867
   PACKER RJ, 1993, J CLIN ONCOL, V11, P850, DOI 10.1200/JCO.1993.11.5.850
   POLLACK IF, 1994, J NEUROSURG, V80, P681, DOI 10.3171/jns.1994.80.4.0681
   POLLACK IF, 1993, J NEUROSURG, V78, P859, DOI 10.3171/jns.1993.78.6.0859
   ROBERTSON PL, 1995, PEDIATR NEUROSURG, V22, P65, DOI 10.1159/000120879
   SHAW EG, 1989, INT J RADIAT ONCOL, V16, P663, DOI 10.1016/0360-3016(89)90482-3
   SQUIRES LA, 1994, NEUROLOGY, V44, P953, DOI 10.1212/WNL.44.5.953
   STROINK AR, 1986, J NEUROSURG, V65, P745, DOI 10.3171/jns.1986.65.6.0745
   VANDERTOP WP, 1992, NEUROSURGERY, V31, P186, DOI 10.1227/00006123-199208000-00003
   ZIMMERMAN RA, 1992, PEDIATR NEUROSURG, V18, P58, DOI 10.1159/000120644
NR 20
TC 73
Z9 75
U1 0
U2 0
PU KARGER
PI BASEL
PA ALLSCHWILERSTRASSE 10, CH-4009 BASEL, SWITZERLAND
SN 1016-2291
EI 1423-0305
J9 PEDIATR NEUROSURG
JI Pediatr. Neurosurg.
PD JAN
PY 2000
VL 32
IS 1
BP 24
EP 29
DI 10.1159/000028893
PG 6
WC Clinical Neurology; Pediatrics; Surgery
WE Science Citation Index Expanded (SCI-EXPANDED)
SC Neurosciences & Neurology; Pediatrics; Surgery
GA 305VZ
UT WOS:000086562400005
PM 10765135
DA 2023-08-27
ER

PT J
AU Kanzaki, J
   Ogawa, K
   Inoue, Y
   Shiobara, R
AF Kanzaki, J
   Ogawa, K
   Inoue, Y
   Shiobara, R
TI Hearing preservation surgery in acoustic neuroma patients with normal
   hearing
SO SKULL BASE SURGERY
LA English
DT Article
ID TUMORS; RECOVERY
AB Acoustic neuroma (AN) patients who had normal hearing accounted for 7% of all our AN cases. It is important to identify AN patients who have normal hearing but are suffering from tinnitus, dizziness, or vertigo, or a combination of these symptoms (without hearing loss), in order to make an early diagnosis: Patients who experience sudden hearing loss and recover completely should also be thoroughly examined in order to rule out AN. In this article, we describe an investigation of the clinical features and results of hearing preservation surgery in AN patients with normal hearing. The overall hearing-preservation:: rate was 57%. The hearing preservation rate did ndt depend on the auditory brainstem response (ABR) pattern, tumor size, or the origin of the tumor. We think that patients must be properly informed concerning all aspects of the surgery, including the possibility that hearing and/or facial nerve function cannot be preserved.
C1 Keio Univ, Sch Med, Dept Otolaryngol, Shinjuku Ku, Tokyo 160, Japan.
   Keio Univ, Sch Med, Dept Neurosurg, Shinjuku Ku, Tokyo 160, Japan.
C3 Keio University; Keio University
RP Kanzaki, J (通讯作者)，Keio Univ, Sch Med, Dept Otolaryngol, Shinjuku Ku, 35 Shinanomachi, Tokyo 160, Japan.
CR BECK HJ, 1986, OTOLARYNG HEAD NECK, V94, P96, DOI 10.1177/019459988609400116
   BERENHOLZ LP, 1992, ANN OTO RHINOL LARYN, V101, P827, DOI 10.1177/000348949210101005
   BERG HM, 1986, OTOLARYNG HEAD NECK, V94, P15, DOI 10.1177/019459988609400103
   HART RG, 1983, NEUROLOGY, V33, P211, DOI 10.1212/WNL.33.2.211
   KANZAKI J, 1991, ACTA OTO-LARYNGOL, P125
   KANZAKI J, 1994, SKULL BASE SURG, V4, P76, DOI 10.1055/s-2008-1058974
   KANZAKI J, 1991, ACTA OTO-LARYNGOL, P120
   KANZAKI J, 1991, ACTA OTO-LARYNGOL, P6
   KANZAKI J, 1986, ORL J OTO-RHINO-LARY, V48, P193, DOI 10.1159/000275868
   KANZAKI J, 1976, OTOLARYNGOLOGY-TOKYO, V48, P193
   MOFFAT DA, 1994, J LARYNGOL OTOL, V108, P116, DOI 10.1017/S0022215100126052
   MONSELL EM, 1995, OTOLARYNG HEAD NECK, V113, P179
   OGAWA K, 1991, ACTA OTO-LARYNGOL, P138
   OGAWA K, 1991, ACTA OTO-LARYNGOL, P144
   PENSAK ML, 1985, LARYNGOSCOPE, V95, P1188
   ROLAND PS, 1987, SOUTHERN MED J, V80, P166, DOI 10.1097/00007611-198702000-00006
   Saleh EA, 1996, AM J OTOL, V17, P127
   SAUNDERS JE, 1995, OTOLARYNG HEAD NECK, V113, P23, DOI 10.1016/S0194-5998(95)70140-0
   YANAGIHARA N, 1993, LARYNGOSCOPE, V103, P308
NR 19
TC 13
Z9 13
U1 0
U2 0
PU THIEME MEDICAL PUBL INC
PI NEW YORK
PA 333 SEVENTH AVE, NEW YORK, NY 10001 USA
SN 1052-1453
J9 SKULL BASE SURG
JI Skull Base Surg.
PD JAN
PY 1997
VL 7
IS 3
BP 109
EP 113
DI 10.1055/s-2008-1058601
PG 5
WC Clinical Neurology; Otorhinolaryngology; Surgery
WE Science Citation Index Expanded (SCI-EXPANDED)
SC Neurosciences & Neurology; Otorhinolaryngology; Surgery
GA 179XA
UT WOS:000079354300001
PM 17171019
OA Green Published
DA 2023-08-27
ER

PT J
AU Ganz, J
   Reda, W
   Abdelkarim, K
AF Ganz, J. C.
   Reda, W. A.
   Abdelkarim, K.
TI Adverse radiation effects after Gamma Knife Surgery in relation to dose
   and volume
SO ACTA NEUROCHIRURGICA
LA English
DT Article
DE Radiosurgery; Gamma Knife; Target volume; Dosimetry; Radiosensitivity;
   Adverse radiation effects; Radionecrosis; Post-radiation oedema; Dose;
   Meningioma; Vestibular schwannoma; Acoustic neuroma; Arteriovenous
   malformation
ID SKULL BASE MENINGIOMAS; INTRACRANIAL ARTERIOVENOUS-MALFORMATIONS;
   ACOUSTIC NEUROMA RADIOSURGERY; CAVERNOUS SINUS MENINGIOMAS; THERAPY
   ONCOLOGY GROUP; PRIMARY BRAIN-TUMORS; STEREOTACTIC RADIOSURGERY;
   VESTIBULAR SCHWANNOMAS; NATURAL-HISTORY; FOLLOW-UP
AB The relationship between target volume and adverse radiation effects (AREs) at low prescription doses requires elucidation. The development of AREs in three series of patients treated in the Gamma Knife is analysed in relation to prescription dose and target volume.
   There were three groups. In group 1, there were of 275 patients with meningiomas; in group 2, 132 patients with vestibular schwannomas; and in group 3, 107 patients with arteriovenous malformations (AVMs). The minimum follow-up for each group was more than 24 months. All patients were followed up at six monthly intervals. The patients with tumours received a prescription dose of 12 Gy, which was varied to protect normal structures but not in relation to tumour volume per se. The desired AVM prescription dose was 25 Gy, but this was also reduced to protect normal structures and to keep the total dose within certain pre-defined limits. All AREs refer to intra-parenchymal increased perilesional T2 signal on MR irrespective of clinical correlation.
   There was no relationship between tumour volume and the development of ARE in the tumour groups. There was a highly significant relationship between target volume and the development of ARE for the AVMs with their much higher dose. Radiation-induced clinical trigeminal and facial nerve deficits with both vestibular schwannomas and meningiomas were always associated with an increased T2 signal in the neighbouring brainstem parenchyma.
   The relationship between target volume and the risk of adverse radiation effects may not apply with lower prescription doses. Individual radiosensitivity may explain why a minority suffer AREs unrelated to target volume. It is possible that radiation-induced brainstem parenchymal damage with concomitant cranial nerve deficits may be commoner after radiosurgery than is usually thought. If tumour control with lower doses is adequate, radiosurgery could be safely considered for larger targets associated with a high risk from microsurgery.
C1 [Ganz, J. C.] Haukeland Hosp, Dept Neurosurg, N-5021 Bergen, Norway.
   [Ganz, J. C.; Reda, W. A.; Abdelkarim, K.] Nasser Inst Shobra, Gamma Knife Ctr Cairo, Cairo, Egypt.
C3 University of Bergen; Haukeland University Hospital
RP Ganz, J (通讯作者)，Haukeland Hosp, Dept Neurosurg, Jonas Liesvei 65, N-5021 Bergen, Norway.
EM
RI Reda, Wael A/E-7447-2010; Karim, Khaled Abdel/C-8895-2015
OI Reda, Wael A/0000-0003-4723-5096; Abdel Karim,
   Khaled/0000-0003-3629-0217
CR Abad J M, 1983, J Neurosurg Sci, V27, P203
   Chang JH, 2003, J NEUROL NEUROSUR PS, V74, P226, DOI 10.1136/jnnp.74.2.226
   Chin LS, 2001, J NEUROSURG, V94, P899, DOI 10.3171/jns.2001.94.6.0899
   Chopra R, 2007, INT J RADIAT ONCOL, V68, P845, DOI 10.1016/j.ijrobp.2007.01.001
   DiBiase SJ, 2004, INT J RADIAT ONCOL, V60, P1515, DOI 10.1016/j.ijrobp.2004.05.073
   DUMA CM, 1993, NEUROSURGERY, V32, P699, DOI 10.1227/00006123-199305000-00001
   Eustacchio S, 2002, ACT NEUR S, V84, P71
   Feigl GC, 2005, J NEUROSURG, V102, P189, DOI 10.3171/jns.2005.102.s_supplement.0189
   Flickinger JC, 2000, INT J RADIAT ONCOL, V46, P1143, DOI 10.1016/S0360-3016(99)00513-1
   FLICKINGER JC, 1992, RADIOSURGERY, P15
   Flickinger JC, 2004, INT J RADIAT ONCOL, V60, P225, DOI 10.1016/j.ijrobp.2004.02.019
   Flickinger JC, 1998, INT J RADIAT ONCOL, V40, P273, DOI 10.1016/S0360-3016(97)00718-9
   FLICKINGER JC, 1990, INT J RADIAT ONCOL, V19, P143, DOI 10.1016/0360-3016(90)90146-B
   Flickinger JC, 2003, INT J RADIAT ONCOL, V56, P801, DOI 10.1016/S0360-3016(03)00126-3
   FLICKINGER JC, 1991, ACTA ONCOL, V30, P363, DOI 10.3109/02841869109092387
   Flickinger JC, 2001, J NEUROSURG, V94, P1, DOI 10.3171/jns.2001.94.1.0001
   FLICKINGER JC, 1991, STEREOT FUNCT NEUROS, V57, P99, DOI 10.1159/000099560
   Foote KD, 2001, J NEUROSURG, V95, P440, DOI 10.3171/jns.2001.95.3.0440
   FULTS D, 1984, NEUROSURGERY, V15, P658, DOI 10.1227/00006123-198411000-00003
   Ganz JC, 1996, STEREOT FUNCT NEUROS, V66, P129, DOI 10.1159/000099778
   GANZ JC, 1993, STEREOT FUNCT NEUROS, V61, P23, DOI 10.1159/000100656
   Ganz JC, 2005, J NEUROSURG, V102, P4, DOI 10.3171/jns.2005.102.s_supplement.0004
   GRAVES VB, 1990, INVEST RADIOL, V25, P952, DOI 10.1097/00004424-199008000-00014
   GUIDETTI B, 1980, J NEUROSURG, V53, P149, DOI 10.3171/jns.1980.53.2.0149
   GUO WY, 1993, ACTA RADIOL, V34, P1
   Harris AE, 2003, SURG NEUROL, V60, P298, DOI 10.1016/S0090-3019(03)00320-3
   HEROS RC, 1987, NEUROLOGY, V37, P279, DOI 10.1212/WNL.37.2.279
   Hudgins WR, 1996, STEREOT FUNCT NEUROS, V66, P121, DOI 10.1159/000099777
   Ide M, 2004, NEUROL MED-CHIR, V44, P321, DOI 10.2176/nmc.44.321
   Inoue HK, 2005, J NEUROSURG, V102, P111, DOI 10.3171/jns.2005.102.s_supplement.0111
   Iwai Y, 1999, SURG NEUROL, V52, P40, DOI 10.1016/S0090-3019(99)00037-3
   Iwai Y, 2000, NEUROL SURG TOKYO, V28, P411
   Iwai Y, 2004, J CLIN NEUROSCI, V11, P715, DOI 10.1016/j.jocn.2003.10.027
   Iwai Y, 2003, NEUROSURGERY, V53, P282, DOI 10.1227/01.NEU.0000073416.22608.B3
   Iwai Y, 2003, NEUROSURGERY, V52, P517, DOI 10.1227/01.NEU.0000047814.18819.9F
   Iwai Y, 2001, J CLIN NEUROSCI, V8, P528, DOI 10.1054/jocn.2000.0890
   Kim CH, 2004, ACTA NEUROCHIR, V146, P741, DOI 10.1007/s00701-004-0226-3
   Kim DG, 2005, J NEUROSURG, V102, P255, DOI 10.3171/jns.2005.102.s_supplement.0255
   Kondziolka D, 1992, Neurosurg Clin N Am, V3, P219
   Kondziolka D, 1998, NEUROSURGERY, V43, P405, DOI 10.1097/00006123-199809000-00001
   Kondziolka D, 1999, J NEUROSURG, V91, P44, DOI 10.3171/jns.1999.91.1.0044
   KONDZIOLKA D, 1998, STEREOTACTIC RADIOSU, P104
   Korytko T, 2006, INT J RADIAT ONCOL, V64, P419, DOI 10.1016/j.ijrobp.2005.07.980
   Kreil W, 2005, J NEUROL NEUROSUR PS, V76, P1425, DOI 10.1136/jnnp.2004.049213
   Kwon J, 1998, STEREOT FUNCT NEUROS, V70, P57, DOI 10.1159/000056407
   Lee JYK, 2002, J NEUROSURG, V97, P65, DOI 10.3171/jns.2002.97.1.0065
   Levegrun S, 2004, STRAHLENTHER ONKOL, V180, P758, DOI 10.1007/s00066-004-1266-6
   Levegrun S, 2004, INT J RADIAT ONCOL, V59, P796, DOI 10.1016/j.ijrobp.2003.11.033
   Linskey M E, 1992, Neurosurg Clin N Am, V3, P191
   Linskey ME, 2005, J NEUROSURG, V102, P59, DOI 10.3171/jns.2005.102.s_supplement.0059
   LINSKEY ME, 1993, INT J RADIAT ONCOL, V25, P227, DOI 10.1016/0360-3016(93)90343-T
   Liscak R, 1995, Cas Lek Cesk, V134, P534
   Liscak R, 2005, J NEUROSURG, V102, P207, DOI 10.3171/jns.2005.102.s_supplement.0207
   Liscak R, 1999, ACTA NEUROCHIR, V141, P473, DOI 10.1007/s007010050327
   LUESSENHOP AJ, 1984, J NEUROSURG, V60, P14, DOI 10.3171/jns.1984.60.1.0014
   Lunsford LD, 2005, J NEUROSURG, V102, P195, DOI 10.3171/jns.2005.102.s_supplement.0195
   Maity A, 2004, PEDIATR NEUROSURG, V40, P207, DOI 10.1159/000082293
   Majhail NS, 2001, STEREOT FUNCT NEUROS, V76, P36, DOI 10.1159/000056493
   Metellus P, 2005, NEUROSURGERY, V57, P873, DOI 10.1227/01.NEU.0000179924.76551.cd
   Michelsen W J, 1979, Clin Neurosurg, V26, P307
   Miyawaki L, 1999, INT J RADIAT ONCOL, V44, P1089, DOI 10.1016/S0360-3016(99)00102-9
   Muthukumar N, 1998, ACTA NEUROCHIR, V140, P315, DOI 10.1007/s007010050104
   Muthukumar N, 1999, SURG NEUROL, V51, P268, DOI 10.1016/S0090-3019(98)00099-8
   Nakamura S, 1996, STEREOT FUNCT NEUROS, V66, P142, DOI 10.1159/000099804
   Nakaya K, 1999, STEREOT FUNCT NEUROS, V72, P67, DOI 10.1159/000056441
   Nicolato A, 1996, STEREOT FUNCT NEUROS, V66, P112, DOI 10.1159/000099776
   Niranjan A, 1999, NEUROSURGERY, V45, P753, DOI 10.1097/00006123-199910000-00003
   Noel G, 2005, INT J RADIAT ONCOL, V63, P1555, DOI 10.1016/j.ijrobp.2005.04.037
   Noren G, 1998, STEREOT FUNCT NEUROS, V70, P65, DOI 10.1159/000056408
   Nussbaum E S, 1995, Clin Neurosurg, V42, P348
   ONDRA SL, 1991, J NEUROSURG, V75, P338
   Pan DHC, 2000, J NEUROSURG, V93, P113, DOI 10.3171/jns.2000.93.supplement_3.0113
   Pan DHC, 1998, STEREOT FUNCT NEUROS, V70, P19, DOI 10.1159/000056403
   Pendl G, 1997, MINIM INVAS NEUROSUR, V40, P87, DOI 10.1055/s-2008-1053423
   PENDL G, 1995, STEREOT FUNCT NEUROS, V64, P11, DOI 10.1159/000098760
   Petit JH, 2001, NEUROSURGERY, V49, P1299, DOI 10.1097/00006123-200112000-00003
   Pollock BE, 2005, INT J RADIAT ONCOL, V62, P1427, DOI 10.1016/j.ijrobp.2004.12.067
   Pollock BE, 2003, INT J RADIAT ONCOL, V55, P1000, DOI 10.1016/S0360-3016(02)04356-0
   Pollock Bruce E, 2003, Neurosurg Focus, V14, pe4
   Regis J, 2002, NEUROCHIRURGIE, V48, P471
   Regis J, 2006, J NEUROSURG, V104, P913, DOI 10.3171/jns.2006.104.6.913
   Roche PH, 2000, J NEUROSURG, V93, P68, DOI 10.3171/jns.2000.93.supplement_3.0068
   Roche PH, 2003, ACTA NEUROCHIR, V145, P883, DOI 10.1007/s00701-003-0123-1
   SHAW E, 1993, INT J RADIAT ONCOL, V27, P1231, DOI 10.1016/0360-3016(93)90548-A
   Shaw E, 1996, INT J RADIAT ONCOL, V34, P647, DOI 10.1016/0360-3016(95)02106-X
   Shaw E, 2000, INT J RADIAT ONCOL, V47, P291, DOI 10.1016/S0360-3016(99)00507-6
   Shin M, 2001, J NEUROSURG, V95, P435, DOI 10.3171/jns.2001.95.3.0435
   Singh VP, 2000, J NEUROSURG, V93, P57, DOI 10.3171/jns.2000.93.supplement_3.0057
   Stafford SL, 2001, NEUROSURGERY, V49, P1029, DOI 10.1097/00006123-200111000-00001
   STEINER L, 1992, RADIOSURGERY BASELIN, P229
   Subach BR, 1998, NEUROSURGERY, V42, P437, DOI 10.1097/00006123-199803000-00001
   Tanaka Takayuki, 1996, Neurologia Medico-Chirurgica, V36, P7, DOI 10.2176/nmc.36.7
   Varlotto JM, 2003, INT J RADIAT ONCOL, V57, P452, DOI 10.1016/S0360-3016(03)00568-6
   Vermeulen S, 1998, STEREOT FUNCT NEUROS, V70, P80, DOI 10.1159/000056410
   VERMEULEN S, 1999, STEREOTACT FUNCT N S, V72, P160
   Voges J, 1996, INT J RADIAT ONCOL, V36, P1055, DOI 10.1016/S0360-3016(96)00422-1
   WILKINS RH, 1985, NEUROSURGERY, V16, P421, DOI 10.1227/00006123-198503000-00026
   YAMAMOTO Y, 1995, J NEUROSURG, V83, P832, DOI 10.3171/jns.1995.83.5.0832
NR 98
TC 47
Z9 48
U1 0
U2 3
PU SPRINGER WIEN
PI WIEN
PA SACHSENPLATZ 4-6, PO BOX 89, A-1201 WIEN, AUSTRIA
SN 0001-6268
EI 0942-0940
J9 ACTA NEUROCHIR
JI Acta Neurochir.
PD JAN
PY 2009
VL 151
IS 1
BP 9
EP 19
DI 10.1007/s00701-008-0174-4
PG 11
WC Clinical Neurology; Surgery
WE Science Citation Index Expanded (SCI-EXPANDED)
SC Neurosciences & Neurology; Surgery
GA 399EY
UT WOS:000262784400002
PM 19129961
DA 2023-08-27
ER

PT J
AU Dinh, CT
   Chen, S
   Nourbakhsh, A
   Padgett, K
   Johnson, P
   Goncalves, S
   Bracho, O
   Bas, E
   Bohorquez, J
   Monje, PV
   Fernandez-Valle, C
   Elsayyad, N
   Liu, XZ
   Welford, SM
   Telischi, F
AF Dinh, Christine T.
   Chen, Si
   Nourbakhsh, Aida
   Padgett, Kyle
   Johnson, Perry
   Goncalves, Stefania
   Bracho, Olena
   Bas, Esperanza
   Bohorquez, Jorge
   Monje, Paula V.
   Fernandez-Valle, Cristina
   Elsayyad, Nagy
   Liu, Xuezhong
   Welford, Scott M.
   Telischi, Fred
TI Single Fraction and Hypofractionated Radiation Cause Cochlear Damage,
   Hearing Loss, and Reduced Viability of Merlin-Deficient Schwann Cells
SO CANCERS
LA English
DT Article
DE vestibular schwannoma; radiation; apoptosis; auditory hair cell;
   hearing; hearing loss; hypofractionation; cell death; Schwann
ID GAMMA-KNIFE RADIOSURGERY; EVIDENCE-BASED GUIDELINES; PRETREATMENT
   GROWTH-RATE; TERM-FOLLOW-UP; VESTIBULAR SCHWANNOMAS; STEREOTACTIC
   RADIOSURGERY; TUMOR-CONTROL; NEUROFIBROMATOSIS TYPE-2; PRESERVATION;
   OUTCOMES
AB Background: Vestibular schwannomas (VS) are benign intracranial tumors caused by loss of function of the merlin tumor suppressor. We tested three hypotheses related to radiation, hearing loss (HL), and VS cell survival: (1) radiation causes HL by injuring auditory hair cells (AHC), (2) fractionation reduces radiation-induced HL, and (3) single fraction and equivalent appropriately dosed multi-fractions are equally effective at controlling VS growth. We investigated the effects of single fraction and hypofractionated radiation on hearing thresholds in rats, cell death pathways in rat cochleae, and viability of human merlin-deficient Schwann cells (MD-SC).Methods: Adult rats received cochlear irradiation with single fraction (0 to 18 Gray [Gy]) or hypofractionated radiation. Auditory brainstem response (ABR) testing was performed for 24 weeks. AHC viabilities were determined using immunohistochemistry. Neonatal rat cochleae were harvested after irradiation, and gene- and cell-based assays were conducted. MD-SCs were irradiated, and viability assays and immunofluorescence for DNA damage and cell cycle markers were performed.Results: Radiation caused dose-dependent and progressive HL in rats and AHC losses by promoting expression of apoptosis-associated genes and proteins. When compared to 12 Gy single fraction, hypofractionation caused smaller ABR threshold and pure tone average shifts and was more effective at reducing MD-SC viability.Conclusions: Investigations into the mechanisms of radiation ototoxicity and VS radiobiology will help determine optimal radiation regimens and identify potential therapies to mitigate radiation-induced HL and improve VS tumor control.
C1 [Dinh, Christine T.; Nourbakhsh, Aida; Goncalves, Stefania; Bracho, Olena; Liu, Xuezhong; Telischi, Fred] Univ Miami, Miller Sch Med, Dept Otolaryngol, Miami, FL 33136 USA.
   [Dinh, Christine T.; Padgett, Kyle; Liu, Xuezhong; Welford, Scott M.; Telischi, Fred] Univ Miami, Sylvester Comprehens Canc Ctr, Miller Sch Med, Miami, FL 33136 USA.
   [Chen, Si] Univ Florida, Coll Med, Dept Otolaryngol, Gainesville, FL 32610 USA.
   [Padgett, Kyle; Welford, Scott M.] Univ Miami, Miller Sch Med, Dept Radiat Oncol, Miami, FL 33136 USA.
   [Padgett, Kyle] Univ Miami, Miller Sch Med, Dept Radiol, Miami, FL 33136 USA.
   [Johnson, Perry] Univ Florida, Coll Med, Dept Radiat Oncol, Gainesville, FL 32610 USA.
   [Bas, Esperanza] Sylvester Comprehens Canc Ctr, Dept Res Pharm, Miami, FL 33136 USA.
   [Bohorquez, Jorge; Telischi, Fred] Univ Miami, Dept Biomed Engn, Miami, FL 33146 USA.
   [Monje, Paula V.] Univ Kentucky, Coll Med, Dept Neurosurg, Lexington, KY 40536 USA.
   [Fernandez-Valle, Cristina] Univ Cent Florida, Coll Med, Burnett Sch Biomed Sci, Orlando, FL 32827 USA.
   [Elsayyad, Nagy] Allina Hlth Canc Inst Radiat Oncol, St Paul, MN 55102 USA.
   [Liu, Xuezhong] Univ Miami, Dept Human Genet, Miller Sch Med, Miami, FL 33136 USA.
C3 University of Miami; University of Miami; State University System of
   Florida; University of Florida; University of Miami; University of
   Miami; State University System of Florida; University of Florida;
   University of Miami; University of Kentucky; State University System of
   Florida; University of Central Florida; University of Miami
RP Dinh, CT (通讯作者)，Univ Miami, Miller Sch Med, Dept Otolaryngol, Miami, FL 33136 USA.; Dinh, CT (通讯作者)，Univ Miami, Sylvester Comprehens Canc Ctr, Miller Sch Med, Miami, FL 33136 USA.
EM
FU American Neurotology Society Research Grant; North American Skull Base
   Society Research Grant; Alpha Omega Alpha Postgraduate Research
   Fellowship Award; NIH/NIDCD [K08DC017508]; Children's Tumor Foundation;
   Department of Defense [33997]; NIH/NCI Sylvester Comprehensive Cancer
   Center K-supplement grant; Miami Project to Cure Paralysis; Indiana
   State Department of Health [43547, T32DC015995]; Department of
   Neurosurgery from University of Kentucky;  [W81-15-1-0446]
FX This research was funded by the American Neurotology Society Research
   Grant to C.T.D., the North American Skull Base Society Research Grant to
   C.T.D. and F.T., the Alpha Omega Alpha Postgraduate Research Fellowship
   Award to A.N., the American Neurotology Society Research Grant to A.N.,
   the NIH/NIDCD K08DC017508 to C.T.D., NIH/NIDCD T32DC015995 to X.Z.L.,
   NIH/NCI Sylvester Comprehensive Cancer Center K-supplement grant to
   C.T.D., and awards from the Children's Tumor Foundation and Department
   of Defense W81-15-1-0446 to C.F.V. The APC was funded by the NIH/NCI
   Sylvester Comprehensive Cancer Center K-supplement grant. P.V.M.
   received funding from The Miami Project to Cure Paralysis, the Indiana
   State Department of Health (grants 33997 and 43547), and the Department
   of Neurosurgery from University of Kentucky.
CR Bas E, 2016, HEARING RES, V337, P12, DOI 10.1016/j.heares.2016.02.003
   Cannan WJ, 2016, J CELL PHYSIOL, V231, P3, DOI 10.1002/jcp.25048
   Carlson ML, 2018, NEUROSURGERY, V82, pE35, DOI 10.1093/neuros/nyx511
   Carlson ML, 2013, J NEUROSURG, V118, P579, DOI 10.3171/2012.9.JNS12919
   Chen ZP, 2018, INT J CLIN ONCOL, V23, P608, DOI 10.1007/s10147-018-1267-6
   Chopra R, 2007, INT J RADIAT ONCOL, V68, P845, DOI 10.1016/j.ijrobp.2007.01.001
   Coughlin AR, 2018, OTOL NEUROTOL, V39, P273, DOI 10.1097/MAO.0000000000001672
   Dilwali S, 2015, SCI REP-UK, V5, DOI 10.1038/srep18599
   Dinh CT, 2020, OTOL NEUROTOL, V41, pE529, DOI 10.1097/MAO.0000000000002613
   Dinh CT, 2018, OTOL NEUROTOL, V39, pE362, DOI 10.1097/MAO.0000000000001766
   Dinh CT, 2017, OTOL NEUROTOL, V38, P180, DOI 10.1097/MAO.0000000000001289
   Evans DGR, 2005, OTOL NEUROTOL, V26, P93, DOI 10.1097/00129492-200501000-00016
   Frischer JM, 2019, J NEUROSURG, V130, P388, DOI 10.3171/2017.8.JNS171281
   Gehlhausen JR, 2015, HUM MOL GENET, V24, P1, DOI 10.1093/hmg/ddu414
   Germano IM, 2018, NEUROSURGERY, V82, pE49, DOI 10.1093/neuros/nyx515
   Ghaderi N, 2022, INT J MOL SCI, V23, DOI 10.3390/ijms23031316
   Govindaraj R, 2022, ADV RADIAT ONCOL, V7, DOI 10.1016/j.adro.2022.101059
   Hasegawa T, 2011, J NEUROSURG, V115, P1078, DOI 10.3171/2011.7.JNS11749
   Horiba A, 2016, NEUROL MED-CHIR, V56, P186, DOI 10.2176/nmc.oa.2015-0212
   Kim YH, 2013, INT J RADIAT ONCOL, V85, P61, DOI 10.1016/j.ijrobp.2012.03.036
   Kirkpatrick JP, 2017, NEURO-ONCOLOGY, V19, P38, DOI 10.1093/neuonc/now301
   Klijn S, 2016, J NEUROSURG, V124, P1619, DOI 10.3171/2015.4.JNS142415
   Kranzinger M, 2014, STRAHLENTHER ONKOL, V190, P798, DOI 10.1007/s00066-014-0630-4
   Kruyt IJ, 2018, J NEUROSURG, V128, P49, DOI 10.3171/2016.9.JNS161463
   Laine H, 2007, J NEUROSCI, V27, P1434, DOI 10.1523/JNEUROSCI.4956-06.2007
   Langenhuizen PPJH, 2019, J NEUROSURG, V131, P1405, DOI 10.3171/2018.6.JNS18516
   Lo A, 2018, INT J RADIAT ONCOL, V100, P443, DOI 10.1016/j.ijrobp.2017.09.024
   Marston AP, 2017, J NEUROSURG, V127, P380, DOI 10.3171/2016.5.JNS153013
   Massager N, 2007, J NEUROSURG, V107, P733, DOI 10.3171/JNS-07/10/0733
   Monje PV, 2018, MOL NEUROBIOL, V55, P6637, DOI 10.1007/s12035-017-0837-3
   National Research Council, 2011, GUIDE CARE USE LAB A, V8th, DOI [DOI 10.17226/12910, 10.17226/12910]
   Nisenbaum E, 2021, OTO OPEN, V5, DOI 10.1177/2473974X211059111
   Niu NN, 2014, INT J RADIAT ONCOL, V89, P113, DOI 10.1016/j.ijrobp.2014.01.038
   Pawlik TM, 2004, INT J RADIAT ONCOL, V59, P928, DOI 10.1016/j.ijrobp.2004.03.005
   Peng KW, 2020, SCI REP-UK, V10, DOI 10.1038/s41598-020-74128-3
   Petrilli AM, 2014, AM J TRANSL RES, V6, P471
   Pollock BE, 2004, STEREOT FUNCT NEUROS, V82, P98, DOI 10.1159/000077659
   Savardekar AR, 2022, WORLD NEUROSURG, V160, P102, DOI 10.1016/j.wneu.2021.11.083
   Schumacher AJ, 2017, J NEUROL SURG PART B, V78, P2, DOI 10.1055/s-0036-1584231
   Sharma MS, 2010, J NEURO-ONCOL, V98, P265, DOI 10.1007/s11060-010-0181-1
   Diaz LS, 2020, J NEURO-ONCOL, V147, P351, DOI 10.1007/s11060-020-03423-w
   Soltys SG, 2021, INT J RADIAT ONCOL, V110, P100, DOI 10.1016/j.ijrobp.2020.11.019
   Sun SB, 2014, J NEUROSURG, V121, P143, DOI 10.3171/2014.8.GKS141503
   Teo M, 2016, WORLD NEUROSURG, V93, P398, DOI 10.1016/j.wneu.2016.06.080
   Thielhelm TP, 2021, CANCERS, V13, DOI 10.3390/cancers13184575
   Nguyen T, 2018, CLIN NEUROL NEUROSUR, V166, P116, DOI 10.1016/j.clineuro.2018.01.005
   Tsao MN, 2017, J RADIOSURGERY SBRT, V5, P5
   Watanabe S, 2016, J NEUROSURG, V125, P64, DOI 10.3171/2016.7.GKS161494
   Yang I, 2010, J NEUROSURG, V112, P851, DOI 10.3171/2009.8.JNS0985
   Yang I, 2009, J NEURO-ONCOL, V93, P41, DOI 10.1007/s11060-009-9842-3
NR 50
TC 0
Z9 0
U1 0
U2 0
PU MDPI
PI BASEL
PA ST ALBAN-ANLAGE 66, CH-4052 BASEL, SWITZERLAND
EI 2072-6694
J9 CANCERS
JI Cancers
PD MAY 18
PY 2023
VL 15
IS 10
AR 2818
DI 10.3390/cancers15102818
PG 23
WC Oncology
WE Science Citation Index Expanded (SCI-EXPANDED)
SC Oncology
GA H6IZ5
UT WOS:000996990800001
PM 37345155
OA Green Published, gold
DA 2023-08-27
ER

PT J
AU Nikniaz, Z
   Ostadrahimi, A
   Mahdavi, R
   Ebrahimi, AA
   Nikniaz, L
AF Nikniaz, Zeinab
   Ostadrahimi, Alireza
   Mahdavi, Reza
   Ebrahimi, Ali Asghar
   Nikniaz, Leila
TI Effects of Elaeagnus angustifolia L. supplementation on serum levels of
   inflammatory cytokines and matrix metalloproteinases in females with
   knee osteoarthritis
SO COMPLEMENTARY THERAPIES IN MEDICINE
LA English
DT Article
DE Cytokines; E. angistifolia L.; Inflammation; Osteoarthritis
ID EXTRACT
AB Objective: In an attempt to investigate new strategies aimed at reducing inflammation in osteoarthritis, the anti-inflammatory effect of Elaeagnus angustifolia L. as a complementary treatment was evaluated in females with knee osteoarthritis.
   Method: In this clinical trial, 90 females with mild to moderate osteoarthritis were assigned to two intervention and one placebo groups. In addition to the conventional therapy, the patients in intervention groups received 15 g/day of E. angustifolia L. medulla and whole fruit powders respectively for 8 weeks. The levels of tumor necrosis factor-alpha (TNF-alpha), interleukine-1 beta (IL-1 beta), interleukine-10 (IL-10), matrix metalloproteinase-1 (MMP-1) and -13 (MMP-13) were measured with human ELISA kits. Paired t-test and ANOVA were used for statistical analysis.
   Results: The statistically significant decrease was observed in the mean levels of serum TNF-alpha in the medulla (0.004) and whole fruit (0.001) groups after 8 weeks of supplementation. In contrast to the placebo group, there was a significant rise in the mean levels of serum IL-10 in medulla (p-value=0.01) and whole fruit groups (p-value=0.009) at the end of study. The interventions resulted in significant decrease in the mean levels of serum MMP-1 in the medulla (0.001) and whole fruit (0.002) groups. After the interventions, no significant changes were observed in the serum IL-1 beta and MMP-13 levels.
   Conclusion: Daily supplementation with E. angustifolia L. in both forms of medulla and whole fruit powders appeared to be effective for decreasing inflammatory cytokines (TNF-alpha and MMP-1) and enhancing anti-inflammatory cytokines (IL-10). (C) 2014 Elsevier Ltd. All rights reserved.
C1 [Nikniaz, Zeinab] Tabriz Univ Med Sci, Student Res Comm, Tabriz, Iran.
   [Ostadrahimi, Alireza] Tabriz Univ Med Sci, Nutr Res Ctr, Tabriz, Iran.
   [Mahdavi, Reza] Tabriz Univ Med Sci, Drug Appl Res Ctr, Tabriz, Iran.
   [Ebrahimi, Ali Asghar] Tabriz Univ Med Sci, Imam Reza Hosp, Connect Tissue Dis Res Ctr, Tabriz, Iran.
   [Nikniaz, Leila] Tabriz Univ Med Sci, Tabriz Hlth Serv Management Res Ctr, Tabriz, Iran.
C3 Tabriz University of Medical Science; Tabriz University of Medical
   Science; Tabriz University of Medical Science; Tabriz University of
   Medical Science; Tabriz University of Medical Science
RP Ostadrahimi, A (通讯作者)，Tabriz Univ Med Sci, Nutr Res Ctr, Tabriz, Iran.
EM
RI nikniaz, zeinab/L-5912-2017; Nikniaz, Leila/M-5747-2017; Mahdavi,
   Reza/C-4773-2017
OI nikniaz, zeinab/0000-0001-6522-1048; Nikniaz, Leila/0000-0003-2951-936X;
   Mahdavi, Reza/0000-0002-5444-2395
FU Nutrition Research Center at Tabriz University of Medical Sciences
   [D/29]
FX The authors wish to thank the Nutrition Research Center at Tabriz
   University of Medical Sciences (NO. D/29) for financial support. We are
   also grateful to all patients for their participation in the study. The
   results of this article are derived from PhD thesis of Zeinab Nikniaz
   (NO of D/29).
CR Ahmadiani A, 2000, J ETHNOPHARMACOL, V72, P287, DOI 10.1016/S0378-8741(00)00222-1
   Ahmed S, 2004, J PHARMACOL EXP THER, V308, P767, DOI 10.1124/jpet.103.059220
   [Anonymous], 1990, MED PLANTS
   Ayaz FA, 2001, J FOOD COMPOS ANAL, V14, P505, DOI 10.1006/jfca.2001.1004
   Bensen WG, 1999, MAYO CLIN PROC, V74, P1095, DOI 10.4065/74.11.1095
   Bucur L, 2009, FARMACIA, V57, P736
   Cauwe B, 2007, CRIT REV BIOCHEM MOL, V42, P113, DOI 10.1080/10409230701340019
   Farahbakhsh S, 2011, BASIC CLIN NEUROSCI, V2, P31
   Fernandes JC, 2002, BIORHEOLOGY, V39, P237
   Fernandez-Moreno M, 2012, FRONT PHYSIOL, V3, DOI 10.3389/fphys.2012.00129
   Hara ES, 2013, BIOCHIMIE, V95, P374, DOI 10.1016/j.biochi.2012.10.016
   Hedbom E, 2002, CELL MOL LIFE SCI, V59, P45, DOI 10.1007/s00018-002-8404-z
   Hulejova H, 2007, CYTOKINE, V38, P151, DOI 10.1016/j.cyto.2007.06.001
   Manicourt DH, 2006, ARTHRITIS RHEUM, V54, P3205, DOI 10.1002/art.22075
   Murakami M, 2000, J BIOL CHEM, V275, P32783, DOI 10.1074/jbc.M003505200
   Nikolaeva A. G., 1970, CHEM NAT COMPD, V6, P659, DOI DOI 10.1007/BF00563479
   Noreen Y, 1998, PLANTA MED, V64, P520, DOI 10.1055/s-2006-957506
   Salaffi F, 2005, CLIN EXP RHEUMATOL, V23, P829
   Schmid B, 2001, PHYTOTHER RES, V15, P344, DOI 10.1002/ptr.981
   Silverstein FE, 2000, JAMA-J AM MED ASSOC, V284, P1247, DOI 10.1001/jama.284.10.1247
   Smith WL, 1996, J BIOL CHEM, V271, P33157, DOI 10.1074/jbc.271.52.33157
   Sondergaard BC, 2006, OSTEOARTHR CARTILAGE, V14, P738, DOI 10.1016/j.joca.2006.01.016
   Wang L, 2013, ADV MATER RES-SWITZ, V750-752, P16, DOI 10.4028/www.scientific.net/AMR.750-752.16
   WELTON AF, 1986, PLANT FLAVONOIDS BIO, P231
   Westacott CI, 1996, SEMIN ARTHRITIS RHEU, V25, P254, DOI 10.1016/S0049-0172(96)80036-9
   Yamazaki Y, 2011, CHEM PHARM BULL, V59, P388, DOI 10.1248/cpb.59.388
NR 26
TC 35
Z9 37
U1 0
U2 20
PU CHURCHILL LIVINGSTONE
PI EDINBURGH
PA JOURNAL PRODUCTION DEPT, ROBERT STEVENSON HOUSE, 1-3 BAXTERS PLACE,
   LEITH WALK, EDINBURGH EH1 3AF, MIDLOTHIAN, SCOTLAND
SN 0965-2299
EI 1873-6963
J9 COMPLEMENT THER MED
JI Complement. Ther. Med.
PD OCT
PY 2014
VL 22
IS 5
BP 864
EP 869
DI 10.1016/j.ctim.2014.07.004
PG 6
WC Integrative & Complementary Medicine
WE Science Citation Index Expanded (SCI-EXPANDED)
SC Integrative & Complementary Medicine
GA AS3UO
UT WOS:000344203700005
PM 25440377
DA 2023-08-27
ER

PT J
AU Fang, D
   Gan, HY
   Cheng, L
   Lee, JH
   Zhou, H
   Sarkaria, JN
   Daniels, DJ
   Zhang, ZG
AF Fang, Dong
   Gan, Haiyun
   Cheng, Liang
   Lee, Jeong-Heon
   Zhou, Hui
   Sarkaria, Jann N.
   Daniels, David J.
   Zhang, Zhiguo
TI H3.3K27M mutant proteins reprogram epigenome by sequestering the PRC2
   complex to poised enhancers
SO ELIFE
LA English
DT Article
ID HISTONE H3.3; DNA HYPOMETHYLATION; PEDIATRIC GLIOMA; ACTIVE GENES;
   CHROMATIN; MUTATIONS; POLYCOMB; INHIBITION; METHYLATION; EXPRESSION
AB Expression of histone H3.3K27M mutant proteins in human diffuse intrinsic pontine glioma (DIPG) results in a global reduction of tri-methylation of H3K27 (H3K27me3), and paradoxically, H3K27me3 peaks remain at hundreds of genomic loci, a dichotomous change that lacks mechanistic insights. Here, we show that the PRC2 complex is sequestered at poised enhancers, but not at active promoters with high levels of H3.3K27M proteins, thereby contributing to the global reduction of H3K27me3. Moreover, the levels of H3.3K27M proteins are low at the retained H3K27me3 peaks and consequently having minimal effects on the PRC2 activity at these loci. H3K27me3-mediated silencing at specific tumor suppressor genes, including Wilms Tumor 1, promotes proliferation of DIPG cells. These results support a model in which the PRC2 complex is redistributed to poised enhancers in H3.3K27M mutant cells and contributes to tumorigenesis in part by locally enhancing H3K27me3, and hence silencing of tumor suppressor genes.
C1 [Fang, Dong; Gan, Haiyun; Zhou, Hui; Zhang, Zhiguo] Columbia Univ, Dept Pediat, Inst Canc Genet, Irving Canc Res Ctr, New York, NY 10027 USA.
   [Fang, Dong; Gan, Haiyun; Zhou, Hui; Zhang, Zhiguo] Columbia Univ, Dept Genet & Dev, Inst Canc Genet, Irving Canc Res Ctr, New York, NY 10027 USA.
   [Cheng, Liang; Lee, Jeong-Heon] Mayo Clin, Dept Biochem & Mol Biol, Rochester, MN USA.
   [Sarkaria, Jann N.] Mayo Clin, Dept Radiat Oncol, Rochester, MN USA.
   [Daniels, David J.] Mayo Clin, Dept Neurosurg, Rochester, MN USA.
   [Fang, Dong] Zhejiang Univ, Innovat Ctr Cell Signaling Network, Life Sci Inst, Hangzhou, Zhejiang, Peoples R China.
C3 Columbia University; Columbia University; Mayo Clinic; Mayo Clinic; Mayo
   Clinic; Zhejiang University
RP Zhang, ZG (通讯作者)，Columbia Univ, Dept Pediat, Inst Canc Genet, Irving Canc Res Ctr, New York, NY 10027 USA.; Zhang, ZG (通讯作者)，Columbia Univ, Dept Genet & Dev, Inst Canc Genet, Irving Canc Res Ctr, New York, NY 10027 USA.
EM
RI gan, haiyun/A-8121-2018
OI Zhang, Zhiguo/0000-0002-9451-2685
FU National Institutes of Health [CA204297]
FX National Institutes of Health CA204297 Zhiguo Zhang; The funders had no
   role in study design, data collection and interpretation, or the
   decision to submit the work for publication.
CR Ahmad K, 2002, MOL CELL, V9, P1191, DOI 10.1016/S1097-2765(02)00542-7
   Ahmad K, 2002, P NATL ACAD SCI USA, V99, P16477, DOI 10.1073/pnas.172403699
   Banaszynski LA, 2013, CELL, V155, P107, DOI 10.1016/j.cell.2013.08.061
   Bardeesy N, 1998, NUCLEIC ACIDS RES, V26, P1784, DOI 10.1093/nar/26.7.1784
   Baylin SB, 2011, NAT REV CANCER, V11, P726, DOI 10.1038/nrc3130
   Bender S, 2013, CANCER CELL, V24, P660, DOI 10.1016/j.ccr.2013.10.006
   Bernstein BE, 2006, CELL, V125, P315, DOI 10.1016/j.cell.2006.02.041
   Campos EI, 2009, ANNU REV GENET, V43, P559, DOI 10.1146/annurev.genet.032608.103928
   Cao R, 2002, SCIENCE, V298, P1039, DOI 10.1126/science.1076997
   Castel D, 2015, ACTA NEUROPATHOL, V130, P815, DOI 10.1007/s00401-015-1478-0
   Chan KM, 2013, CELL CYCLE, V12, P2546, DOI 10.4161/cc.25625
   Chan KM, 2013, GENE DEV, V27, P985, DOI 10.1101/gad.217778.113
   Chen P, 2013, GENE DEV, V27, P2109, DOI 10.1101/gad.222174.113
   Chow CM, 2005, EMBO REP, V6, P354, DOI 10.1038/sj.embor.7400366
   Cordero FJ, 2017, MOL CANCER RES, V15, P1243, DOI 10.1158/1541-7786.MCR-16-0389
   Creyghton MP, 2010, P NATL ACAD SCI USA, V107, P21931, DOI 10.1073/pnas.1016071107
   Deaton AM, 2016, ELIFE, V5, DOI 10.7554/eLife.15316
   Dellaretti M, 2012, J NEUROSURG, V117, P810, DOI 10.3171/2012.7.JNS111992
   Eden A, 2003, SCIENCE, V300, P455, DOI 10.1126/science.1083557
   Ehrlich M, 2009, EPIGENOMICS-UK, V1, P239, DOI 10.2217/EPI.09.33
   Elsasser SJ, 2015, NATURE, V522, P240, DOI 10.1038/nature14345
   Esteller M, 2007, HUM MOL GENET, V16, pR50, DOI 10.1093/hmg/ddm018
   Fang D, 2016, SCIENCE, V352, P1344, DOI 10.1126/science.aae0065
   Feng JX, 2012, NAT PROTOC, V7, P1728, DOI 10.1038/nprot.2012.101
   Fontebasso AM, 2014, NAT GENET, V46, P462, DOI 10.1038/ng.2950
   Funato K, 2014, SCIENCE, V346, P1529, DOI 10.1126/science.1253799
   GESSLER M, 1990, NATURE, V343, P774, DOI 10.1038/343774a0
   Goldberg AD, 2010, CELL, V140, P678, DOI 10.1016/j.cell.2010.01.003
   Han JH, 2013, CELL, V155, P817, DOI 10.1016/j.cell.2013.10.014
   Hashizume R, 2014, NAT MED, V20, P1394, DOI 10.1038/nm.3716
   Herz HM, 2014, SCIENCE, V345, P1065, DOI 10.1126/science.1255104
   Iwami K, 2013, NEURO-ONCOLOGY, V15, P747, DOI 10.1093/neuonc/not007
   Izumoto S, 2008, J NEUROSURG, V108, P963, DOI 10.3171/JNS/2008/108/5/0963
   Jayaram H, 2016, P NATL ACAD SCI USA, V113, P6182, DOI 10.1073/pnas.1605523113
   Jiao LY, 2015, SCIENCE, V350, DOI 10.1126/science.aac4383
   Jones C, 2014, NAT REV CANCER, V14, P651, DOI 10.1038/nrc3811
   Justin N, 2016, NAT COMMUN, V7, DOI 10.1038/ncomms11316
   Langmead B, 2012, NAT METHODS, V9, P357, DOI [10.1038/NMETH.1923, 10.1038/nmeth.1923]
   Lewis PW, 2013, SCIENCE, V340, P857, DOI 10.1126/science.1232245
   Li H, 2009, BIOINFORMATICS, V25, P1754, DOI 10.1093/bioinformatics/btp324
   Lu C, 2016, SCIENCE, V352, P844, DOI 10.1126/science.aac7272
   Maiti S, 2000, CANCER RES, V60, P6288
   Margueron R, 2011, NATURE, V469, P343, DOI 10.1038/nature09784
   Maze I, 2014, NAT REV GENET, V15, P259, DOI 10.1038/nrg3673
   McKittrick E, 2004, P NATL ACAD SCI USA, V101, P1525, DOI 10.1073/pnas.0308092100
   Mohammad F, 2017, NAT MED, V23, P483, DOI 10.1038/nm.4293
   Monje M, 2011, P NATL ACAD SCI USA, V108, P4453, DOI 10.1073/pnas.1101657108
   Ng RK, 2008, NAT CELL BIOL, V10, P102, DOI 10.1038/ncb1674
   Nikbakht H, 2016, NAT COMMUN, V7, DOI 10.1038/ncomms11185
   Orlando DA, 2014, CELL REP, V9, P1163, DOI 10.1016/j.celrep.2014.10.018
   Piunti A, 2017, NAT MED, V23, P493, DOI 10.1038/nm.4296
   Quinlan AR, 2010, BIOINFORMATICS, V26, P841, DOI 10.1093/bioinformatics/btq033
   Rada-Iglesias A, 2011, NATURE, V470, P279, DOI 10.1038/nature09692
   Rai TS, 2014, GENE DEV, V28, P2712, DOI 10.1101/gad.247528.114
   Rauscher J, 2014, J CLIN PATHOL, V67, P556, DOI 10.1136/jclinpath-2013-202114
   Schubeler D, 2004, GENE DEV, V18, P1263, DOI 10.1101/gad.1198204
   Schwartzentruber J, 2012, NATURE, V482, P226, DOI [10.1038/nature10833, 10.1007/s13311-012-0137-6]
   Shan CM, 2016, ELIFE, V5, DOI 10.7554/eLife.17903
   Simon JA, 2013, MOL CELL, V49, P808, DOI 10.1016/j.molcel.2013.02.013
   Solomon DA, 2016, BRAIN PATHOL, V26, P569, DOI 10.1111/bpa.12336
   Stroud H, 2012, P NATL ACAD SCI USA, V109, P5370, DOI 10.1073/pnas.1203145109
   Struhl K, 2014, ELIFE, V3, DOI 10.7554/eLife.02475
   Sturm D, 2012, CANCER CELL, V22, P425, DOI 10.1016/j.ccr.2012.08.024
   Szenker E, 2011, CELL RES, V21, P421, DOI 10.1038/cr.2011.14
   Tagami H, 2004, CELL, V116, P51, DOI 10.1016/S0092-8674(03)01064-X
   Tatavosian R, 2018, NAT COMMUN, V9, DOI 10.1038/s41467-018-04455-7
   Wu G, 2014, NAT GENET, V46, P444, DOI 10.1038/ng.2938
   Wu G, 2012, NAT GENET, V44, P251, DOI 10.1038/ng.1102
   Yeung JT, 2013, J NEURO-ONCOL, V111, P103, DOI 10.1007/s11060-012-0998-x
   Zhang R, 2016, C ELECT INSUL DIEL P, P1007, DOI 10.1109/CEIDP.2016.7785631
NR 70
TC 56
Z9 57
U1 1
U2 7
PU ELIFE SCIENCES PUBLICATIONS LTD
PI CAMBRIDGE
PA SHERATON HOUSE, CASTLE PARK, CAMBRIDGE, CB3 0AX, ENGLAND
SN 2050-084X
J9 ELIFE
JI eLife
PD JUN 22
PY 2018
VL 7
AR e36696
DI 10.7554/eLife.36696
PG 23
WC Biology
WE Science Citation Index Expanded (SCI-EXPANDED)
SC Life Sciences & Biomedicine - Other Topics
GA GL7RS
UT WOS:000437400800001
PM 29932419
OA Green Published, Green Submitted, gold
DA 2023-08-27
ER

PT J
AU LYONS, MK
   KELLY, PJ
AF LYONS, MK
   KELLY, PJ
TI COMPUTER-ASSISTED STEREOTAXIC BIOPSY AND VOLUMETRIC RESECTION OF
   THALAMIC PILOCYTIC ASTROCYTOMAS - REPORT OF 23 CASES
SO STEREOTACTIC AND FUNCTIONAL NEUROSURGERY
LA English
DT Article; Proceedings Paper
CT MEETING OF THE AMERICAN SOC FOR STEREOTACTIC AND FUNCTIONAL NEUROSURGERY
CY JUN 16-19, 1991
CL PITTSBURGH, PA
SP AMER SOC STEREOTACT & FUNCT NEUROSURG
DE STEREOTAXIC SURGERY; RESECTION; VOLUMETRIC; THALAMUS; PILOCYTIC;
   ASTROCYTOMA
ID NEOPLASMS
AB Twenty-three cases of pathologically verified thalamic pilocytic astrocytomas diagnosed at computer-assisted stereotactic biopsy and/or volumetric resection at the Mayo Clinic between January 1985 and October 1990 were reviewed. Computer-assisted stereotactic volumetric resection was performed in 19 patients. Postoperative imaging demonstrated no residual contrast-enhancing tumor in 14 patients and a small amount of contrast-enhancing tumor (< 5% of the original tumor volume) in 5 patients. Biopsy only was performed in 4 patients: 2 with primarily cystic tumors successfully treated with stereotactic instillation of P-32, and 2 patients with stable (nonprogressive) deficits. There was 1 operative death; a patient with tumor extending into the midbrain became comatose and died 10 days after surgery. The remaining 22 patients are alive and well. Computer-assisted stereotactic volumetric resection of thalamic pilocytic astrocytomas can be performed with low morbidity and a favorable long-term prognosis.
C1 MAYO CLIN & MAYO FDN,DEPT NEUROL SURG,ROCHESTER,MN 55905.
   MAYO CLIN & MAYO GRAD SCH MED,ROCHESTER,MN 55901.
C3 Mayo Clinic; Mayo Clinic
RI Kelly, Patrick Joseph/E-8056-2017
OI Kelly, Patrick Joseph/0000-0002-8540-7488
CR BERNSTEIN M, 1984, J NEUROSURG, V61, P649, DOI 10.3171/jns.1984.61.4.0649
   KELLY PJ, 1986, J NEUROSURG, V64, P427, DOI 10.3171/jns.1986.64.3.0427
   KELLY PJ, 1989, NEUROSURGERY, V25, P185, DOI 10.1227/00006123-198908000-00006
   KELLY PJ, 1987, MAYO CLIN PROC, V62, P450, DOI 10.1016/S0025-6196(12)65470-6
   KELLY PJ, 1984, SURG NEUROL, V21, P465, DOI 10.1016/0090-3019(84)90452-X
   MCGIRR SJ, 1987, NEUROSURGERY, V20, P447, DOI 10.1227/00006123-198703000-00016
NR 6
TC 32
Z9 32
U1 0
U2 1
PU KARGER
PI BASEL
PA ALLSCHWILERSTRASSE 10, CH-4009 BASEL, SWITZERLAND
SN 1011-6125
J9 STEREOT FUNCT NEUROS
JI Stereotact. Funct. Neurosurg.
PY 1992
VL 59
IS 1-4
BP 100
EP 104
DI 10.1159/000098924
PG 5
WC Neurosciences; Neuroimaging; Surgery
WE Conference Proceedings Citation Index - Science (CPCI-S); Science Citation Index Expanded (SCI-EXPANDED)
SC Neurosciences & Neurology; Surgery
GA JQ015
UT WOS:A1992JQ01500015
PM 1295026
DA 2023-08-27
ER

PT J
AU Blomstergren, A
   Rydelius, A
   Abul-Kasim, K
   Latt, J
   Sundgren, PC
   Bengzon, J
AF Blomstergren, Adam
   Rydelius, Anna
   Abul-Kasim, Kasim
   Latt, Jimmy
   Sundgren, Pia C.
   Bengzon, Johan
TI Evaluation of reproducibility in MRI quantitative volumetric assessment
   and its role in the prediction of overall survival and progression-free
   survival in glioblastoma
SO ACTA RADIOLOGICA
LA English
DT Article
DE Magnetic resonance imaging (MRI); brain; brainstem; adults; 3D computer
   applications; primary neoplasms; surgery
ID RECURRENT GLIOBLASTOMA; MALIGNANT GLIOMA; TUMOR VOLUME; RESECTION;
   EXTENT; RADIOTHERAPY; CLASSIFICATION; MULTIFORME; DIAGNOSIS; IMPACT
AB Background Residual tumor volume (RTV) and extent of resection (EOR) have previously been shown to affect survival in glioblastoma (GB) patients. Quantitative radiological assessment (QRA) of these factors could potentially affect clinical decision-making in the postoperative period. Purpose The first aim was to evaluate the reproducibility of different volume estimation methods of RTV and EOR by comparing QRA with subjective visual estimation and with objective volume estimations. The second aim was to clarify whether QRA of RTV and EOR would provide accuracy in predicting progression-free survival (PFS) and overall survival (OS) in GB patients. Material and Methods Seventy GB patients were studied retrospectively. Reproducibility of QRA was compared to conventional visual analysis. Intra-rater agreement between two repeated measurements of 25 patients was calculated. QRA for RTV and EOR was made for the entire study population. Survival analysis was performed by multivariate cox-regression analysis. Results QRA of RTV and EOR gave superior intra-rater agreement compared to subjective evaluation. Multivariate survival analysis showed prognostic significance on 18 months PFS (hazard ratio [HR] = 0.44, P = 0.003) and OS (HR = 0.42, P = 0.012) at RTV < 1.6 mL and with EOR > 96% on PFS (HR = 2.152, P = 0.005) but not on OS (HR = 1.92, P = 0.053). Conclusion QRA of tumor volumes is more robust compared to standard evaluation methods. Since EOR and RTV are correlated to the prognosis in GB, quantitative analysis of tumor volumes could aid decision-making and patient management postoperatively.
C1 [Blomstergren, Adam; Bengzon, Johan] Lund Univ, Dept Clin Sci, Div Neurosurg, SE-22184 Lund, Sweden.
   [Blomstergren, Adam; Bengzon, Johan] Reg Skane, SE-22184 Lund, Sweden.
   [Rydelius, Anna] Lund Univ, Dept Clin Sci, Div Neurol, Lund, Sweden.
   [Abul-Kasim, Kasim] Lund Univ, Ctr Imaging & Funct, Sect Neuroradiol, SUS Malmo, Malmo, Sweden.
   [Latt, Jimmy; Sundgren, Pia C.] Lund Univ, Ctr Imaging & Funct, SUS, Lund, Sweden.
   [Sundgren, Pia C.] Lund Univ, Dept Clin Sci, Div Radiol, Lund, Sweden.
   [Bengzon, Johan] Lund Univ, Stem Cell Ctr, BMC B10, Lund, Sweden.
C3 Lund University; Lund University; Lund University; Lund University; Lund
   University; Lund University
RP Blomstergren, A (通讯作者)，Lund Univ, Dept Clin Sci, Div Neurosurg, SE-22184 Lund, Sweden.; Blomstergren, A (通讯作者)，Reg Skane, SE-22184 Lund, Sweden.
EM
OI Sundgren, Pia/0000-0001-9237-1236; Rydelius, Anna/0000-0002-6742-1825
FU Medical Faculty at Lund University, Region Skane; Swedish Cancer Society
   [CAN 2013/321, CAN 2016/365]; Swedish Research Council
   [K2011-52X-21737-01-3]; Regional Research Funds; Crafoord Fesearch
   Foundation [20120612]
FX The author(s) disclosed receipt of the following financial support for
   the research, authorship, and/or publication of this article: This work
   was supported by a donation from Viveca Jeppsson, ALF grants from the
   Medical Faculty at Lund University, Region Skane, the Swedish Cancer
   Society (CAN 2013/321, CAN 2016/365), the Swedish Research Council
   (K2011-52X-21737-01-3), Regional Research Funds, and the Crafoord
   Fesearch Foundation (no. 20120612).
CR ALBERT FK, 1945, NEUROSURGERY, V34, P45
   BLAND JM, 1986, LANCET, V1, P307, DOI 10.1016/s0140-6736(86)90837-8
   Bloch O, 2012, J NEUROSURG, V117, P1032, DOI 10.3171/2012.9.JNS12504
   Chaichana KL, 2014, J NEURO-ONCOL, V120, P625, DOI 10.1007/s11060-014-1597-9
   Chaichana KL, 2014, NEURO-ONCOLOGY, V16, P113, DOI 10.1093/neuonc/not137
   Chaudhry NS, 2013, CANCER INVEST, V31, P287, DOI 10.3109/07357907.2013.789899
   D'Amico RS, 2017, WORLD NEUROSURG, V103, P538, DOI 10.1016/j.wneu.2017.04.041
   Dea N, 2012, CAN J NEUROL SCI, V39, P632, DOI 10.1017/S0317167100015377
   Grabowski MM, 2014, J NEUROSURG, V121, P1115, DOI 10.3171/2014.7.JNS132449
   Kanaly CW, 2011, PLOS ONE, V6, DOI 10.1371/journal.pone.0016031
   KARNOFSKY DA, 1948, CANCER-AM CANCER SOC, V1, P634, DOI 10.1002/1097-0142(194811)1:4<634::AID-CNCR2820010410>3.0.CO;2-L
   Lacroix M, 2001, J NEUROSURG, V95, P190, DOI 10.3171/jns.2001.95.2.0190
   Laperriere N, 2002, RADIOTHER ONCOL, V64, P259, DOI 10.1016/S0167-8140(02)00078-6
   Louis DN, 2007, ACTA NEUROPATHOL, V114, P547, DOI 10.1007/s00401-007-0278-6
   Louis DN, 2016, ACTA NEUROPATHOL, V131, P803, DOI 10.1007/s00401-016-1545-1
   McGirt MJ, 2009, J NEUROSURG, V110, P156, DOI 10.3171/2008.4.17536
   McHugh ML, 2012, BIOCHEM MEDICA, V22, P276, DOI 10.11613/bm.2012.031
   Meier R, 2017, J NEUROSURG, V127, P798, DOI 10.3171/2016.9.JNS16146
   Meier R, 2016, SCI REP-UK, V6, DOI 10.1038/srep23376
   Orringer D, 2012, J NEUROSURG, V117, P851, DOI 10.3171/2012.8.JNS12234
   Porz N, 2016, PLOS ONE, V11, DOI 10.1371/journal.pone.0165302
   Sanai N, 2011, J NEUROSURG, V115, P3, DOI 10.3171/2011.2.JNS10998
   Sawaya R, 1998, NEUROSURGERY, V42, P1044, DOI 10.1097/00006123-199805000-00054
   Stupp R, 2014, ANN ONCOL, V25, P93, DOI 10.1093/annonc/mdu050
   Stupp R, 2009, LANCET ONCOL, V10, P459, DOI 10.1016/S1470-2045(09)70025-7
   Suchorska B, 2016, NEURO-ONCOLOGY, V18, P549, DOI 10.1093/neuonc/nov326
   Upadhyay N, 2011, BRIT J RADIOL, V84, pS107, DOI 10.1259/bjr/65711810
   WALKER MD, 1980, NEW ENGL J MED, V303, P1323, DOI 10.1056/NEJM198012043032303
   Watts C, 2014, CLIN ONCOL-UK, V26, P385, DOI 10.1016/j.clon.2014.04.001
   Weller M, 2014, LANCET ONCOL, V15, pE395, DOI 10.1016/S1470-2045(14)70011-7
   Wen PY, 2008, NEW ENGL J MED, V359, P492, DOI 10.1056/NEJMra0708126
NR 31
TC 9
Z9 9
U1 1
U2 10
PU SAGE PUBLICATIONS LTD
PI LONDON
PA 1 OLIVERS YARD, 55 CITY ROAD, LONDON EC1Y 1SP, ENGLAND
SN 0284-1851
EI 1600-0455
J9 ACTA RADIOL
JI Acta Radiol.
PD APR
PY 2019
VL 60
IS 4
BP 516
EP 525
DI 10.1177/0284185118786060
PG 10
WC Radiology, Nuclear Medicine & Medical Imaging
WE Science Citation Index Expanded (SCI-EXPANDED)
SC Radiology, Nuclear Medicine & Medical Imaging
GA HQ0NQ
UT WOS:000462092300014
PM 29966430
DA 2023-08-27
ER

PT J
AU Karadeniz-Bilgili, MY
   Semelka, RC
   Hyslop, WB
   Pamuklar, E
   Rivero, H
   Firat, Z
   Woosley, JT
AF Karadeniz-Bilgili, MY
   Semelka, RC
   Hyslop, WB
   Pamuklar, E
   Rivero, H
   Firat, Z
   Woosley, JT
TI MRI findings of primary small-cell carcinoma of kidney
SO MAGNETIC RESONANCE IMAGING
LA English
DT Article
DE MRI; small cell carcinoma; kidney
ID URINARY-TRACT; THERAPY
AB We report the MRI findings of primary small-cell carcinoma of the kidney (PSCCK) in a 59-year-old female. This tumor appeared as a 16-cm mass that arose from the right kidney. This lesion had diminished signal on T1-weighted images and heterogeneous mixed signal oil T2-weighted images. The tumor primarily involved the renal medulla with persistent thin renal cortex. Despite the tumors' large size, no substantial central necrosis was present. The predominant medullary location and the lack of central necrosis in this large tumor were features unusual for renal cell carcinoma and should raise the suspicion of another malignancy, the differential diagnosis of which should contain extrapulmonary sinall-cell carcinoma of the kidney. (c) 2005 Elsevier Inc. All rights reserved.
C1 Univ N Carolina, Dept Radiol, Chapel Hill, NC 27599 USA.
   Univ N Carolina, Dept Pathol & Lab Med, Chapel Hill, NC 27599 USA.
C3 University of North Carolina; University of North Carolina Chapel Hill;
   University of North Carolina; University of North Carolina Chapel Hill
RP Semelka, RC (通讯作者)，Univ N Carolina, Dept Radiol, CB 7510, Chapel Hill, NC 27599 USA.
EM
RI bilgili, mirace yasemin karadeniz/ABB-3286-2021
OI bilgili, mirace yasemin karadeniz/0000-0002-2368-5239; Pamuklar,
   Ertan/0000-0002-7066-6786
CR Akkaya BK, 2003, UROL ONCOL-SEMIN ORI, V21, P11, DOI 10.1016/S1078-1439(03)00021-8
   CAPELLA C, 1984, AM J SURG PATHOL, V8, P855, DOI 10.1097/00000478-198411000-00006
   Chuang Cheng-Keng, 2003, Chang Gung Med J, V26, P26
   Fujisawa Toshio, 2003, Nihon Kokyuki Gakkai Zasshi, V41, P440
   Gonzalez-Lois C, 2001, ARCH PATHOL LAB MED, V125, P796
   Majhail NS, 2003, CANCER, V97, P1436, DOI 10.1002/cncr.11199
   Matsumoto Minori, 2004, Hinyokika Kiyo, V50, P111
   REMICK SC, 1992, MED PEDIATR ONCOL, V20, P89, DOI 10.1002/mpo.2950200202
   Semelka RC, 1996, AM J ROENTGENOL, V166, P823, DOI 10.2214/ajr.166.4.8610558
   SEMELKA RC, 2002, ABDOMINAL PELVIC MRI, P793
   WEEKS SM, 1995, ABDOM IMAGING, V20, P365, DOI 10.1007/BF00203373
   YU DS, 1990, BRIT J UROL, V66, P590, DOI 10.1111/j.1464-410X.1990.tb07188.x
NR 12
TC 12
Z9 17
U1 0
U2 1
PU ELSEVIER SCIENCE INC
PI NEW YORK
PA 360 PARK AVE SOUTH, NEW YORK, NY 10010-1710 USA
SN 0730-725X
EI 1873-5894
J9 MAGN RESON IMAGING
JI Magn. Reson. Imaging
PD APR
PY 2005
VL 23
IS 3
BP 515
EP 517
DI 10.1016/j.mri.2004.12.008
PG 3
WC Radiology, Nuclear Medicine & Medical Imaging
WE Science Citation Index Expanded (SCI-EXPANDED)
SC Radiology, Nuclear Medicine & Medical Imaging
GA 924HD
UT WOS:000228968800010
PM 15862654
DA 2023-08-27
ER

PT J
AU Rosenstock, T
   Giampiccolo, D
   Schneider, H
   Runge, SJ
   Bahrend, I
   Vajkoczy, P
   Picht, T
AF Rosenstock, Tizian
   Giampiccolo, Davide
   Schneider, Heike
   Runge, Sophia Jutta
   Baehrend, Ina
   Vajkoczy, Peter
   Picht, Thomas
TI Specific DTI seeding and diffusivity-analysis improve the quality and
   prognostic value of TMS-based deterministic DTI of the pyramidal tract
SO NEUROIMAGE-CLINICAL
LA English
DT Article
DE Navigated transcranial magnetic stimulation (nTMS); Brain tumor surgery;
   Glioma; Motor outcome; Diffusion tensor imaging; Fractional anisotropy;
   Apparent diffusion coefficient
ID TRANSCRANIAL MAGNETIC STIMULATION; CORTICOSPINAL TRACT; BRAIN-TUMORS;
   FIBER TRACKING; NAVIGATED TMS; MOTOR CORTEX; WHITE-MATTER; TRACTOGRAPHY;
   RESECTION; SURGERY
AB Object: Navigated transcranial magnetic stimulation (nTMS) combined with diffusion tensor imaging (DTI) is used preoperatively in patients with eloquent-located brain lesions and allows analyzing non-invasively the spatial relationship between the tumor and functional areas (e.g. the motor cortex and the corticospinal tract [CST]). In this study, we examined the diffusion parameters FA (fractional anisotropy) and ADC (apparent diffusion coefficient) within the CST in different locations and analyzed their interrater reliability and usefulness for predicting the patients' motor outcome with a precise approach of specific region of interest (ROI) seeding based on the color-coded FA-map.
   Methods: Prospectively collected data of 30 patients undergoing bihemispheric nTMS mapping followed by nTMS-based DTI fiber tracking prior to surgery of motor eloquent high-grade gliomas were analyzed by 2 experienced and 1 unexperienced examiner. The following data were scrutinized for both hemispheres after tractography based on nTMS-motor positive cortical seeds and a 2nd region of interest in one layer of the caudal pons defined by the color-coded FA-map: the pre-and postoperative motor status (day of discharge und 3 months), the closest distance between the tracts and the tumor (TTD), the fractional anisotropy (FA) and the apparent diffusion coefficient (ADC). The latter as an average within the CST as well as specific values in different locations (peritumoral, mesencephal, pontine).
   Results: Lower average FA-values within the affected CST as well as higher average ADC-values are significantly associated with deteriorated postoperative motor function (p = 0.006 and p = 0.026 respectively). Segmental analysis within the CST revealed that the diffusion parameters are especially disturbed on a peritumoral level and that the degree of their impairment correlates with motor deficits (FA p = 0.065, ADC p = 0.007). No significant segmental variation was seen in the healthy hemisphere. The interrater reliability showed perfect agreement for almost all analyzed parameters.
   Conclusions: Adding diffusion weighted imaging derived information on the structural integrity of the nTMS-based tractography results improves the predictive power for postoperative motor outcome. Utilizing a second subcortical ROI which is specifically seeded based on the color-coded FA map increases the tracking quality of the CST independently of the examiner's experience. Further prospective studies are needed to validate the nTMS-based prediction of the patient's outcome.
C1 [Rosenstock, Tizian; Schneider, Heike; Runge, Sophia Jutta; Baehrend, Ina; Vajkoczy, Peter; Picht, Thomas] Charite, Dept Neurosurg, Charitepl 1, D-10117 Berlin, Germany.
   [Giampiccolo, Davide] Univ Hosp, Inst Neurosurg, Piazzale Stefani 1, I-37100 Verona, Italy.
C3 Free University of Berlin; Humboldt University of Berlin; Charite
   Universitatsmedizin Berlin; University of Verona; Azienda Ospedaliera
   Universitaria Integrata Verona
RP Rosenstock, T (通讯作者)，Charite, Dept Neurosurg, Charitepl 1, D-10117 Berlin, Germany.
EM
OI Picht, Thomas/0000-0002-8630-1466; Rosenstock,
   Tizian/0000-0002-0620-9118; Giampiccolo, Davide/0000-0003-3849-4168
CR Almenawer SA, 2015, NEURO-ONCOLOGY, V17, P868, DOI 10.1093/neuonc/nou349
   [Anonymous], J NEUROSURG
   Basser P. J., 1994, Biophysical Journal, V66, P259, DOI 10.1016/S0006-3495(94)80775-1
   Bobek-Billewicz B, 2011, FOLIA NEUROPATHOL, V49, P262
   Bucci M, 2013, NEUROIMAGE-CLIN, V3, P361, DOI 10.1016/j.nicl.2013.08.008
   Chen LZ, 2016, NEUROSCI BIOBEHAV R, V68, P838, DOI 10.1016/j.neubiorev.2016.07.022
   Conti A, 2014, OPER NEUROSURG, V10, P542, DOI 10.1227/NEU.0000000000000502
   Davidson T, 2013, PLOS ONE, V8, DOI 10.1371/journal.pone.0070286
   Duffau H, 2007, NEUROPHYSIOL CLIN, V37, P373, DOI 10.1016/j.neucli.2007.09.003
   Duffau H., 2005, J NEUROL NEUROSURG
   Forster MT, 2015, NEUROSURGERY, V76, P766, DOI 10.1227/NEU.0000000000000715
   Forster MT, 2014, OPER NEUROSURG, V10, P51, DOI 10.1227/NEU.0000000000000075
   Forster MT, 2011, NEUROSURGERY, V68, P1317, DOI 10.1227/NEU.0b013e31820b528c
   Frey D, 2012, NEUROIMAGE, V62, P1600, DOI 10.1016/j.neuroimage.2012.05.059
   Frey D, 2014, NEURO-ONCOLOGY, V16, P1365, DOI 10.1093/neuonc/nou110
   Giordano M, 2015, CLIN NEUROL NEUROSUR, V136, P1, DOI 10.1016/j.clineuro.2015.05.004
   Hervey-Jumper SL, 2016, J NEURO-ONCOL, V130, P269, DOI 10.1007/s11060-016-2110-4
   Kombos T, 2009, J CLIN NEUROPHYSIOL, V26, P422, DOI 10.1097/WNP.0b013e3181c2c0dc
   Kombos T, 2009, NEUROSURG FOCUS, V27, DOI 10.3171/2009.8.FOCUS09140
   Krieg SM, 2014, NEURO-ONCOLOGY, V16, P1274, DOI 10.1093/neuonc/nou007
   Krieg SM, 2012, J NEUROSURG, V116, P994, DOI 10.3171/2011.12.JNS111524
   Krieg SM, 2012, ACTA NEUROCHIR, V154, P555, DOI 10.1007/s00701-011-1255-3
   Kumar P, 2016, NEURORADIOLOGY, V58, P1043, DOI 10.1007/s00234-016-1718-6
   Kumar P, 2016, J STROKE, V18, P50, DOI 10.5853/jos.2015.01186
   Lehericy S, 2000, J NEUROSURG, V92, P589, DOI 10.3171/jns.2000.92.4.0589
   Liu WL, 2016, PARKINSONISM RELAT D, V32, P94, DOI 10.1016/j.parkreldis.2016.09.005
   Louis DN, 2007, ACTA NEUROPATHOL, V114, P547, DOI 10.1007/s00401-007-0278-6
   Lu S, 2003, AM J NEURORADIOL, V24, P937
   Lucas CW, 2017, NEUROIMAGE-CLIN, V13, P297, DOI 10.1016/j.nicl.2016.11.022
   Mandelli ML, 2014, J NEUROSURG, V121, P349, DOI 10.3171/2014.4.JNS131160
   McGirt MJ, 2009, NEUROSURGERY, V65, P463, DOI 10.1227/01.NEU.0000349763.42238.E9
   Mori S, 2002, NMR BIOMED, V15, P468, DOI 10.1002/nbm.781
   Negwer C, 2017, BRAIN IMAGING BEHAV, V11, P899, DOI 10.1007/s11682-016-9563-0
   Negwer C, 2017, J NEUROSURG, V126, P1006, DOI 10.3171/2016.2.JNS152382
   Perez MA, 2009, TOP STROKE REHABIL, V16, P254, DOI 10.1310/tsr1604-254
   Picht T, 2016, J NEURO-ONCOL, V126, P535, DOI 10.1007/s11060-015-1993-9
   Picht T, 2011, NEUROSURGERY, V69, P581, DOI 10.1227/NEU.0b013e3182181b89
   Picht T, 2009, NEUROSURGERY, V65, P93, DOI 10.1227/01.NEU.0000348009.22750.59
   Rutten GJ, 2010, NEUROSURG FOCUS, V28, DOI 10.3171/2009.12.FOCUS09251
   Sanai N, 2008, NEUROSURGERY, V62, P753, DOI 10.1227/01.neu.0000318159.21731.cf
   SCHAG CC, 1984, J CLIN ONCOL, V2, P187, DOI 10.1200/JCO.1984.2.3.187
   Schonberg T, 2006, NEUROIMAGE, V30, P1100, DOI 10.1016/j.neuroimage.2005.11.015
   Smits M, 2007, AM J NEURORADIOL, V28, P1354, DOI 10.3174/ajnr.A0538
   Sollmann N, 2017, BRAIN TOPOGR, V30, P98, DOI 10.1007/s10548-016-0536-9
   Stadlbauer A, 2007, AM J NEURORADIOL, V28, P462
   Tarapore PE, 2016, CLIN NEUROPHYSIOL, V127, P1895, DOI 10.1016/j.clinph.2015.11.042
   Tarapore PE, 2012, J NEUROSURG, V117, P354, DOI 10.3171/2012.5.JNS112124
   Weiss C, 2015, NEUROIMAGE-CLIN, V7, P424, DOI 10.1016/j.nicl.2015.01.006
   Zdunczyk A, 2013, ACTA NEUROCHIR, V155, P1309, DOI 10.1007/s00701-013-1665-5
   Zolal A, 2013, J NEURORADIOLOGY, V40, P71, DOI 10.1016/j.neurad.2012.05.006
NR 50
TC 33
Z9 35
U1 0
U2 5
PU ELSEVIER SCI LTD
PI OXFORD
PA THE BOULEVARD, LANGFORD LANE, KIDLINGTON, OXFORD OX5 1GB, OXON, ENGLAND
SN 2213-1582
J9 NEUROIMAGE-CLIN
JI NeuroImage-Clin.
PY 2017
VL 16
BP 276
EP 285
DI 10.1016/j.nicl.2017.08.010
PG 10
WC Neuroimaging
WE Science Citation Index Expanded (SCI-EXPANDED)
SC Neurosciences & Neurology
GA FK1IU
UT WOS:000413235100030
PM 28840099
OA Green Published, gold
DA 2023-08-27
ER

PT J
AU Hu, YS
   Lee, CC
   Wu, CA
   Lin, CJ
   Yang, HC
   Guo, WY
   Liu, KD
   Chung, WY
   Shiau, CY
   Wu, HM
AF Hu, Yong-Sin
   Lee, Cheng-Chia
   Wu, Chia-An
   Lin, Chung-Jung
   Yang, Huai-Che
   Guo, Wan-Yuo
   Liu, Kang-Du
   Chung, Wen-Yuh
   Shiau, Cheng-Ying
   Wu, Hsiu-Mei
TI Magnetic resonance imaging signal characteristics associated with
   prognosis of skull base chordoma after gamma knife radiosurgery
SO JOURNAL OF NEURO-ONCOLOGY
LA English
DT Article
DE Diffusion-weighted imaging; Magnetic resonance imaging; Chordoma; Skull
   base; Stereotactic radiosurgery; Gamma Knife
ID CLINICAL-FEATURES; OUTCOMES
AB Objective:To investigate the association between the magnetic resonance imaging (MRI) signal characteristics of skull base chordoma and radiosurgical outcomes. Methods:Twenty-four patients with skull base chordomas treated with Gamma Knife radiosurgery (GKRS) after previous surgical resection were retrospectively (2001-2021) examined. Pre-GKRS MRIs were analyzed for R-T2 (tumor-to-brainstem signal intensity ratio on T2-weighted imaging), R-CE (tumor-to-brainstem signal intensity ratio on contrast-enhanced T1-weighted imaging), and mean apparent diffusion coefficient (ADC). Correlations of the parameters with patient survival and local tumor progression were made by using Cox regression and Kaplan-Meier analyses. Results:During a median follow-up of 46 months after GKRS, 9 patients died with significantly more local tumor progression events (median number: 2 vs 0, P = .012) than did 15 alive patients. On multivariable analysis, higher mean ADC was associated with longer patient survival (P = .016) after GKRS. The actuarial 5-year overall survival rates were 88.9% versus 54.7% for chordomas with an ADC of >= 1270 x 10(-6) mm(2)/s versus < 1270 x 10(-6) mm(2)/s. R-T2 < 1.5 (P = .038) and R-CE > 1.57 (P = .022) were associated with a lower probability of local tumor control. Conclusion:Lower mean ADC values are associated with shorter patient survival in skull base chordomas after GKRS. Diffusion-weighted imaging may help in GKRS planning and outcome prediction for these patients.
C1 [Hu, Yong-Sin] Minist Hlth & Welf, Taipei Hosp, Dept Radiol, New Taipei, Taiwan.
   [Hu, Yong-Sin; Lee, Cheng-Chia; Wu, Chia-An; Lin, Chung-Jung; Yang, Huai-Che; Guo, Wan-Yuo; Liu, Kang-Du; Chung, Wen-Yuh; Shiau, Cheng-Ying; Wu, Hsiu-Mei] Natl Yang Ming Chiao Tung Univ, Sch Med, Taipei, Taiwan.
   [Hu, Yong-Sin; Wu, Chia-An; Lin, Chung-Jung; Guo, Wan-Yuo; Wu, Hsiu-Mei] Taipei Vet Gen Hosp, Dept Radiol, 201 Shipai Rd,Sec 2, Taipei 112, Taiwan.
   [Lee, Cheng-Chia; Yang, Huai-Che; Liu, Kang-Du; Chung, Wen-Yuh] Taipei Vet Gen Hosp, Neurol Inst, Dept Neurosurg, Taipei, Taiwan.
   [Wu, Chia-An] Taipei Med Univ, Shuang Ho Hosp, Dept Radiol, Taipei, Taiwan.
   [Chung, Wen-Yuh] Kaohsiung Vet Gen Hosp, Dept Neurosurg, Kaohsiung, Taiwan.
   [Shiau, Cheng-Ying] Taipei Vet Gen Hosp, Canc Ctr, Taipei, Taiwan.
C3 National Yang Ming Chiao Tung University; Taipei Veterans General
   Hospital; Taipei Veterans General Hospital; Taipei Medical University;
   Shuang Ho Hospital; Kaohsiung Veterans General Hospital; Taipei Veterans
   General Hospital
RP Wu, HM (通讯作者)，Natl Yang Ming Chiao Tung Univ, Sch Med, Taipei, Taiwan.; Wu, HM (通讯作者)，Taipei Vet Gen Hosp, Dept Radiol, 201 Shipai Rd,Sec 2, Taipei 112, Taiwan.
EM
RI Hu, Yong-Sin/I-8972-2017
OI Hu, Yong-Sin/0000-0002-4786-0791
FU Taipei Veterans General Hospital; Taiwan's Ministry of Science and
   Technology [V110C-056];  [MOST-109-2628-B-010-014-MY2]
FX This work was supported by Taipei Veterans General Hospital (Grant
   Number: V110C-056) and Taiwan's Ministry of Science and Technology
   (Grant Number: MOST-109-2628-B-010-014-MY2).
CR Catton C, 1996, RADIOTHER ONCOL, V41, P67, DOI 10.1016/S0167-8140(96)01805-1
   Erdem E, 2003, RADIOGRAPHICS, V23, P995, DOI 10.1148/rg.234025176
   Ginat DT, 2012, AM J ROENTGENOL, V198, pW597, DOI 10.2214/AJR.11.7424
   Guler E, 2017, J NEUROL SURG PART B, V78, P256, DOI 10.1055/s-0036-1597824
   Holtzman AL, 2021, INT J PART THER, V8, P179, DOI 10.14338/IJPT-20-00066.1
   Kano H, 2011, NEUROSURGERY, V68, P379, DOI 10.1227/NEU.0b013e3181ffa12c
   Lee CC, 2018, J NEURO-ONCOL, V140, P547, DOI 10.1007/s11060-018-2980-8
   Malayeri AA, 2011, RADIOGRAPHICS, V31, P1773, DOI 10.1148/rg.316115515
   Oh HC, 2021, SCI REP-UK, V11, DOI 10.1038/s41598-020-79894-8
   Pikis S, 2022, J NEUROSURG, V137, P977, DOI 10.3171/2021.12.JNS212416
   Raza SM, 2018, OPER NEUROSURG, V15, P131, DOI 10.1093/ons/opx201
   Santegoeds Rene G C, 2018, Curr Radiol Rep, V6, P16, DOI 10.1007/s40134-018-0275-7
   Sasaki T, 2018, AM J NEURORADIOL, V39, P1316, DOI 10.3174/ajnr.A5664
   Shinya Y, 2022, INT J RADIAT ONCOL, V113, P582, DOI 10.1016/j.ijrobp.2022.02.024
   Snell JW, 2006, J NEUROSURG, V104, P157, DOI 10.3171/jns.2006.104.1.157
   Takagi M, 2018, RADIAT ONCOL, V13, DOI 10.1186/s13014-018-1173-0
   Tian K, 2017, AM J NEURORADIOL, V38, P1206, DOI 10.3174/ajnr.A5152
   Tian KB, 2016, WORLD NEUROSURG, V92, P407, DOI 10.1016/j.wneu.2016.05.035
   U.S. Department of Health and Human Service . National Institutes of Health National Cancer Institute, 2017, COMMON TERMINOLOGY C
   Walcott BP, 2012, LANCET ONCOL, V13, pE69, DOI 10.1016/S1470-2045(11)70337-0
   Wang K, 2020, WORLD NEUROSURG, V136, pE310, DOI 10.1016/j.wneu.2019.12.164
   Wang L, 2017, J NEUROSURG, V127, P1257, DOI 10.3171/2016.9.JNS16559
   Yeom KW, 2013, AM J NEURORADIOL, V34, P1056, DOI 10.3174/ajnr.A3333
   Zhou JP, 2018, WORLD NEUROSURG, V117, P46, DOI 10.1016/j.wneu.2018.05.209
NR 24
TC 0
Z9 0
U1 1
U2 2
PU SPRINGER
PI NEW YORK
PA ONE NEW YORK PLAZA, SUITE 4600, NEW YORK, NY, UNITED STATES
SN 0167-594X
EI 1573-7373
J9 J NEURO-ONCOL
JI J. Neuro-Oncol.
PD JAN
PY 2023
VL 161
IS 1
BP 45
EP 56
DI 10.1007/s11060-022-04199-x
EA DEC 2022
PG 12
WC Oncology; Clinical Neurology
WE Science Citation Index Expanded (SCI-EXPANDED)
SC Oncology; Neurosciences & Neurology
GA 8B5NK
UT WOS:000903606700001
PM 36565364
DA 2023-08-27
ER

PT J
AU Burkhardt, K
   Heuberger, F
   Delavelle, J
AF Burkhardt, K.
   Heuberger, F.
   Delavelle, J.
TI Pilocytic astrocytoma in the elderly
SO CLINICAL NEUROPATHOLOGY
LA English
DT Article
DE pilocytic astrocytoma; tumor; related parkinsonism; neuroradiological
   pitfalls; old age
ID ADULT; PARKINSONISM; FEATURES; CHILDREN
AB Pilocytic astrocytoma (WHO Grade I) is a low-grade glioma with a favorable prognosis most commonly diagnosed in patients aged below 20. It is the most common glioma in children, and cases discovered in elderly patients are rare. We report the highly unusual case of an 85-year-old man whose neurological signs included Parkinsonism, and in whom post mortem examination revealed a pilocytic astrocytoma of the brainstem. We also discuss the clinical, neuroradiological and neuropathological differential diagnosis.
C1 Univ Hosp Geneva, Dept Surg Pathol, CH-1211 Geneva, Switzerland.
   Univ Hosp Geneva, Hop de Loex, Geneva, Switzerland.
   Univ Hosp Geneva, Dept Neuroradiol, Geneva, Switzerland.
C3 University of Geneva; University of Geneva; University of Geneva
RP Burkhardt, K (通讯作者)，Univ Hosp Geneva, Dept Surg Pathol, Rue Micheli-du-Crest 24, CH-1211 Geneva, Switzerland.
EM
OI Burkhardt, Karim/0000-0002-0802-3499
CR Bell D, 2004, BRIT J NEUROSURG, V18, P613, DOI 10.1080/02688690400022896
   Burkhard C, 2003, J NEUROSURG, V98, P1170, DOI 10.3171/jns.2003.98.6.1170
   COAKLEY KJ, 1995, MAYO CLIN PROC, V70, P747, DOI 10.4065/70.8.747
   Giannini C, 1999, J NEUROPATH EXP NEUR, V58, P46, DOI 10.1097/00005072-199901000-00006
   Kestle J, 2004, J NEUROSURG, V101, P1, DOI 10.3171/ped.2004.101.2.0001
   KLEIHUES P, 2000, WHO CLASSIFICATION T, P45
   Komotar Ricardo J, 2005, Neurosurg Focus, V18, pE7
   LONES MA, 1991, ACTA NEUROPATHOL, V81, P688, DOI 10.1007/BF00296382
   Lyons MK, 2007, CLIN NEUROL NEUROSUR, V109, P76, DOI 10.1016/j.clineuro.2006.03.002
   Papavero L, 2005, BRAIN PATHOL, V15, P91
   Pohle T, 1999, MOVEMENT DISORD, V14, P842, DOI 10.1002/1531-8257(199909)14:5<842::AID-MDS1020>3.0.CO;2-O
   Rektor I, 2006, J NEUROL SCI, V248, P185, DOI 10.1016/j.jns.2006.05.026
   Scarabino T, 2005, J Neurosurg Sci, V49, P73
   Tihan T, 1999, J NEUROPATH EXP NEUR, V58, P1061, DOI 10.1097/00005072-199910000-00004
   van der Wal EPJ, 2003, PEDIATR RADIOL, V33, P207, DOI 10.1007/s00247-002-0828-y
NR 15
TC 5
Z9 6
U1 0
U2 0
PU DUSTRI-VERLAG DR KARL FEISTLE
PI DEISENHOFEN-MUENCHEN
PA BAHNHOFSTRASSE 9 POSTFACH 49, D-82032 DEISENHOFEN-MUENCHEN, GERMANY
SN 0722-5091
J9 CLIN NEUROPATHOL
JI Clin. Neuropathol.
PD NOV-DEC
PY 2007
VL 26
IS 6
BP 306
EP 310
PG 5
WC Clinical Neurology; Pathology
WE Science Citation Index Expanded (SCI-EXPANDED)
SC Neurosciences & Neurology; Pathology
GA 239IX
UT WOS:000251513100007
PM 18232598
DA 2023-08-27
ER

PT J
AU Li, HY
   Ren, B
   Yu, SL
   Gao, HW
   Sun, PL
AF Li, Huiyang
   Ren, Bo
   Yu, Shili
   Gao, Hongwen
   Sun, Ping-Li
TI The clinicopathological significance of thymic epithelial markers
   expression in thymoma and thymic carcinoma
SO BMC CANCER
LA English
DT Article
DE Immunohistochemical staining; Thymic epithelial markers; Cortex;
   Medulla; Differentiation; Thymoma; Thymic squamous cell carcinoma
ID PROTEASOME SUBUNIT BETA-5T; CD40 EXPRESSION; AIRE EXPRESSION;
   DIFFERENTIATION; CLASSIFICATION; NEOPLASMS; REPRODUCIBILITY; SELECTION;
   CANCER; CELLS
AB BackgroundThe classification of thymomas is based on the morphology of epithelial tumor cells and the proportion of lymphocytes. Type A thymomas are composed of the spindle or oval tumor epithelial cells. Tumor cells of B thymomas are epithelioid-shaped with increasing atypia. Type AB thymomas have the features of epithelial tumor cells of A and B thymomas. The diagnosis can be difficult because of the complex morphology. Some novel thymic epithelial markers have been reported in several preclinical studies, but they have not been applied to clinical practice. Here, we investigated the expression of 3 cortical and 3 medullary markers, which are thymoproteasome-specific subunit beta 5t (beta 5t), thymus-specific serine protease 16 (PRSS16), cathepsin V, autoimmune regulator (AIRE), CD40 and claudin-4.MethodsImmunohistochemistry was used to analyze 53 cases of thymomas and thymic squamous cell carcinomas (TSCC), aiming to explore the expression of cortical and medullary epithelial markers and their correlation with histological classification, Masaoka-Koga stage, and prognosis.ResultsOur results found that for cortical epithelial markers the expression of beta 5t, PRSS16, and cathepsin V was higher in type AB and B thymomas than in micronodular thymoma with lymphoid stroma (MNT), and we observed a dramatic increase of beta 5t and PRSS16 expression in type AB compared to type A thymomas. In medullary epithelial markers, the expression of AIRE was higher in type A than in B3 thymomas. CD40 and beta 5t expression were associated with the Masaoka-Koga stage. High cathepsin V expression was related to a good prognosis and a longer progression-free survival.ConclusionThis is the first comprehensive analysis of the role of thymic cortical and medullary epithelial markers as biomarkers for differential diagnosis and prognosis in thymic tumors. Thymic medullary epithelial immunophenotype was found to exhibit in type A, MNT, and TSCC. Type B thymomas primarily exhibited a cortical epithelial immunophenotype. Type AB thymomas showed cortical, medullary, or mixed corticomedullary epithelial immunophenotype. Our results demonstrated that thymic cortical and medullary epithelial markers including beta 5t, PRSS16, cathepsin V, and AIRE could be used as ancillary markers in the diagnosis and prognosis of thymic epithelial tumors.
C1 [Li, Huiyang; Ren, Bo; Yu, Shili; Gao, Hongwen; Sun, Ping-Li] Jilin Univ, Hosp 2, Dept Pathol, 218 Ziqiang Rd, Changchun 130041, Jilin, Peoples R China.
C3 Jilin University
RP Sun, PL (通讯作者)，Jilin Univ, Hosp 2, Dept Pathol, 218 Ziqiang Rd, Changchun 130041, Jilin, Peoples R China.
EM
RI ren, bo/IST-0814-2023
CR Alkaaki A, 2022, EUR J CARDIO-THORAC, V62, DOI 10.1093/ejcts/ezac274
   Angirekula M, 2022, CANCERS, V14, DOI 10.3390/cancers14092299
   [Anonymous], 2022, REF PERM NCCN CLIN P
   Bardou P, 2014, BMC BIOINFORMATICS, V15, DOI 10.1186/1471-2105-15-293
   BERNATZ PE, 1961, J THORAC CARDIOV SUR, V42, P424
   GALY AHM, 1992, J IMMUNOL, V149, P775
   Gardner JM, 2009, CURR OPIN IMMUNOL, V21, P582, DOI 10.1016/j.coi.2009.08.007
   Gommeaux J, 2009, EUR J IMMUNOL, V39, P956, DOI 10.1002/eji.200839175
   Ichimiya S, 2006, J HISTOCHEM CYTOCHEM, V54, P1277, DOI 10.1369/jhc.6A7028.2006
   Illei PB, 2020, SEMIN DIAGN PATHOL, V37, P166, DOI 10.1053/j.semdp.2020.04.006
   Jing JS, 2018, INT J CLIN EXP PATHO, V11, P4035
   Kawano H, 2015, J IMMUNOL, V195, P5149, DOI 10.4049/jimmunol.1501000
   Kiuchi S, 2017, HUM PATHOL, V60, P66, DOI 10.1016/j.humpath.2016.09.027
   Kojika M, 2009, MODERN PATHOL, V22, P1341, DOI 10.1038/modpathol.2009.105
   Kondo K, 2004, ANN THORAC SURG, V77, P1183, DOI 10.1016/j.athoracsur.2003.07.042
   Lee GD, 2016, J THORAC DIS, V8, P901, DOI 10.21037/jtd.2016.03.53
   Li R, 2009, INT J MOL SCI, V10, P3900, DOI 10.3390/ijms10093900
   Liu PP, 2022, J CLIN PATHOL, V75, P702, DOI 10.1136/jclinpath-2021-207819
   Liu YM, 2014, IMMUNOL LETT, V161, P50, DOI 10.1016/j.imlet.2014.04.008
   MARINO M, 1985, VIRCHOWS ARCH A, V407, P119, DOI 10.1007/BF00737071
   Matsumura Y, 2016, ANTICANCER RES, V36, P4467, DOI 10.21873/anticanres.10991
   Miki Y, 2014, VIRCHOWS ARCH, V464, P725, DOI 10.1007/s00428-014-1587-5
   Mneimneh WS, 2015, MODERN PATHOL, V28, P1415, DOI 10.1038/modpathol.2015.104
   Okumura M, 2008, GEN THORAC CARDIOVAS, V56, P10, DOI 10.1007/s11748-007-0177-8
   Roden AC, 2015, AM J SURG PATHOL, V39, P427, DOI 10.1097/PAS.0000000000000391
   Roden AC, 2015, HUM PATHOL, V46, P17, DOI 10.1016/j.humpath.2014.10.001
   Slobodova Z, 2011, NEOPLASMA, V58, P189, DOI 10.4149/neo_2011_03_189
   Strobel P, 2008, ANN NY ACAD SCI, V1132, P143, DOI 10.1196/annals.1405.018
   Strobel P, 2014, HISTOPATHOLOGY, V64, P557, DOI 10.1111/his.12279
   Suzuki E, 2008, AUTOIMMUNITY, V41, P377, DOI 10.1080/08916930801987573
   Tomaru U, 2013, ARCH IMMUNOL THER EX, V61, P357, DOI 10.1007/s00005-013-0234-1
   Tomaru U, 2012, J CLIN PATHOL, V65, P858, DOI 10.1136/jclinpath-2012-200820
   Toss M, 2020, J CLIN PATHOL, V73, P76, DOI 10.1136/jclinpath-2019-205939
   Tseng YC, 2017, PLOS ONE, V12, DOI 10.1371/journal.pone.0179527
   Valavanis Christos, 2021, J BUON, V26, P1198
   Wang XP, 2012, FRONT IMMUNOL, V3, DOI 10.3389/fimmu.2012.00019
   WHO Classification of Tumours Editorial Board, 2021, WHO CLASS TUM, V5
   Wolf JL, 2020, HISTOPATHOLOGY, V77, P734, DOI 10.1111/his.14167
   Yamada Y, 2022, VIRCHOWS ARCH, V480, P843, DOI 10.1007/s00428-021-03229-9
   Yamada Y, 2014, J CLIN PATHOL, V67, DOI 10.1136/jclinpath-2013-201930
   Yamada Y, 2011, AM J SURG PATHOL, V35, P1296, DOI 10.1097/PAS.0b013e3182237f5d
   Zeng HL, 2020, BMC CANCER, V20, DOI 10.1186/s12885-020-07332-z
   Zucali PA, 2013, LUNG CANCER, V79, P236, DOI 10.1016/j.lungcan.2012.11.015
NR 43
TC 0
Z9 0
U1 2
U2 2
PU BMC
PI LONDON
PA CAMPUS, 4 CRINAN ST, LONDON N1 9XW, ENGLAND
EI 1471-2407
J9 BMC CANCER
JI BMC Cancer
PD FEB 16
PY 2023
VL 23
IS 1
AR 161
DI 10.1186/s12885-023-10619-6
PG 10
WC Oncology
WE Science Citation Index Expanded (SCI-EXPANDED)
SC Oncology
GA 9L0VV
UT WOS:000941276600005
PM 36797681
OA Green Published, gold
DA 2023-08-27
ER

PT J
AU SUZUKI, T
   WATANABE, K
   SUGINO, T
   TANIGAWA, T
   SATOH, S
AF SUZUKI, T
   WATANABE, K
   SUGINO, T
   TANIGAWA, T
   SATOH, S
TI IMMUNOCYTOCHEMICAL DEMONSTRATION OF IGF-II IMMUNOREACTIVITY IN HUMAN
   PHEOCHROMOCYTOMA AND EXTRAADRENAL ABDOMINAL PARAGANGLIOMA
SO JOURNAL OF PATHOLOGY
LA English
DT Article
DE IGF-II; PHEOCHROMOCYTOMA; PARAGANGLIOMA; IMMUNOCYTOCHEMISTRY
ID GROWTH FACTOR-II; NONSUPPRESSIBLE INSULIN-LIKE; STRUCTURAL HOMOLOGY;
   WILMS TUMOR; HUMAN-SERUM; HUMAN-BRAIN; EXPRESSION; DIFFERENTIATION;
   TRANSCRIPTS; CELLS
AB Insulin-like growth factor (IGF)-II immunoreactivity was demonstrated in 14 adrenal glands. six retroperitoneal paraganglia, 18 phaeochromocytomas. and six extra-adrenal abdominal paragangliomas through the use of a monoclonal antibody against rat IGF-II which cross-reacts with human IGF-II. A subpopulation of parenchymal cells in normal adrenal medulla and in retroperitoneal paraganglia was positively immunolabelled. Eighteen cases with phaeochromocytoma including two malignant and four multiple endocrine neoplasias all contained numerous IGF-II-immunoreactive tumour cells. All six extra-adrenal abdominal paragangliomas contained numerous immunoreactive tumour cells. Almost all the other normal human adult tissues examined failed to immunolabel. These results indicate that IGF-II-like immunoreactivity is widely expressed in normal and neoplastic human paraganglionic tissues, although its biological significance in these tissues has not been determined.
C1 FUKUSHIMA MED SCH,DEPT PATHOL,FUKUSHIMA 960,JAPAN.
   NIIGATA UNIV,SCH MED,DEPT UROL,NIIGATA 95021,JAPAN.
C3 Fukushima Medical University; Niigata University
RI Tanigawa, Tetsuya/J-6131-2019
OI Tanigawa, Tetsuya/0000-0002-9305-1637
CR CARLSSONSKWIRUT C, 1986, FEBS LETT, V201, P46, DOI 10.1016/0014-5793(86)80568-3
   CONGOTE LF, 1985, BIOCHEM BIOPH RES CO, V126, P653, DOI 10.1016/0006-291X(85)90234-7
   EWTON DZ, 1984, J CELL PHYSIOL, V120, P263, DOI 10.1002/jcp.1041200302
   FROESCH ER, 1985, ANNU REV PHYSIOL, V47, P443
   GRAHAM RC, 1966, J HISTOCHEM CYTOCHEM, V14, P291, DOI 10.1177/14.4.291
   HASELBACHER G, 1982, ENDOCRINOLOGY, V110, P1822, DOI 10.1210/endo-110-5-1822
   HASELBACHER GK, 1987, P NATL ACAD SCI USA, V84, P1104, DOI 10.1073/pnas.84.4.1104
   HASELBACHER GK, 1985, P NATL ACAD SCI USA, V82, P2153, DOI 10.1073/pnas.82.7.2153
   HERVONEN A, 1987, AM J ANAT, V153, P563
   MOSES AC, 1980, EUR J BIOCHEM, V103, P387, DOI 10.1111/j.1432-1033.1980.tb04325.x
   RECHLER MM, 1978, EUR J BIOCHEM, V82, P5, DOI 10.1111/j.1432-1033.1978.tb11991.x
   RECIOPINTO E, 1984, BRAIN RES, V302, P323, DOI 10.1016/0006-8993(84)90246-4
   REEVE AE, 1985, NATURE, V317, P258, DOI 10.1038/317258a0
   RINDERKNECHT E, 1978, J BIOL CHEM, V253, P2769
   RINDERKNECHT E, 1976, P NATL ACAD SCI USA, V73, P4379, DOI 10.1073/pnas.73.12.4379
   SCHMID C, 1983, FEBS LETT, V161, P117, DOI 10.1016/0014-5793(83)80742-X
   SCOTT J, 1985, NATURE, V317, P260, DOI 10.1038/317260a0
   SU TS, 1989, CANCER RES, V49, P1773
   SUZUKI T, 1989, VIRCHOWS ARCH A, V414, P515, DOI 10.1007/BF00781709
   TANAKA H, 1989, ENDOCRINOLOGY, V124, P870, DOI 10.1210/endo-124-2-870
   ZAPF J, 1978, EUR J BIOCHEM, V87, P285, DOI 10.1111/j.1432-1033.1978.tb12377.x
NR 21
TC 11
Z9 11
U1 0
U2 0
PU JOHN WILEY & SONS LTD
PI W SUSSEX
PA BAFFINS LANE CHICHESTER, W SUSSEX, ENGLAND PO19 1UD
SN 0022-3417
J9 J PATHOL
JI J. Pathol.
PD JUN
PY 1992
VL 167
IS 2
BP 199
EP 203
DI 10.1002/path.1711670207
PG 5
WC Oncology; Pathology
WE Science Citation Index Expanded (SCI-EXPANDED)
SC Oncology; Pathology
GA JB260
UT WOS:A1992JB26000006
PM 1635001
DA 2023-08-27
ER

PT J
AU Wang, J
   Huang, TYT
   Hou, Y
   Bartom, E
   Lu, XY
   Shilatifard, A
   Yue, F
   Saratsis, A
AF Wang, Juan
   Huang, Tina Yi-Ting
   Hou, Ye
   Bartom, Elizabeth
   Lu, Xinyan
   Shilatifard, Ali
   Yue, Feng
   Saratsis, Amanda
TI Epigenomic landscape and 3D genome structure in pediatric high-grade
   glioma
SO SCIENCE ADVANCES
LA English
DT Article
ID HIJACKING; ENHANCERS; OLIG2
AB Pediatric high-grade gliomas (pHGGs), including glioblastoma multiforme (GBM) and diffuse intrinsic pontine glioma (DIPG), are morbid brain tumors. Even with treatment survival is poor, making pHGG the number one cause of cancer death in children. Up to 80% of DIPGs harbor a somatic missense mutation in genes encoding histone H3. To investigate whether H3K27M is associated with distinct chromatin structure that alters transcription regulation, we generated the first high-resolution Hi-C maps of pHGG cell lines and tumor tissue. By integrating transcriptome (RNA-seq), enhancer landscape (ChIP-seq), genome structure (Hi-C), and chromatin accessibility (ATAC-seq) datasets from H3K27M and wild-type specimens, we identified tumor-specific enhancers and regulatory networks for known oncogenes. We identified genomic structural variations that lead to potential enhancer hijacking and gene coamplification, including A2M, JAG2, and FLRT1. Together, our results imply three-dimensional genome alterations may play a critical role in the pHGG epigenetic landscape and contribute to tumorigenesis.
C1 [Wang, Juan; Hou, Ye; Bartom, Elizabeth; Shilatifard, Ali; Yue, Feng; Saratsis, Amanda] Northwestern Univ, Dept Biochem & Mol Genet, Feinberg Sch Med, Chicago, IL 60611 USA.
   [Huang, Tina Yi-Ting; Saratsis, Amanda] Northwestern Univ, Dept Neurol Surg, Feinberg Sch Med, Chicago, IL 60611 USA.
   [Lu, Xinyan] Northwestern Univ, Dept Pathol, Feinberg Sch Med, Chicago, IL 60611 USA.
   [Yue, Feng; Saratsis, Amanda] Northwestern Univ, Robert H Lurie Comprehens Canc Ctr, Chicago, IL 60611 USA.
   [Saratsis, Amanda] Ann & Robert H Lurie Childrens Hosp Chicago, Dept Surg, Div Pediat Neurosurg, Chicago, IL 60611 USA.
C3 Northwestern University; Feinberg School of Medicine; Northwestern
   University; Feinberg School of Medicine; Northwestern University;
   Feinberg School of Medicine; Ann & Robert H. Lurie Children's Hospital
   of Chicago; Northwestern University; Robert H. Lurie Comprehensive
   Cancer Center; Ann & Robert H. Lurie Children's Hospital of Chicago
RP Yue, F; Saratsis, A (通讯作者)，Northwestern Univ, Dept Biochem & Mol Genet, Feinberg Sch Med, Chicago, IL 60611 USA.; Saratsis, A (通讯作者)，Northwestern Univ, Dept Neurol Surg, Feinberg Sch Med, Chicago, IL 60611 USA.; Yue, F; Saratsis, A (通讯作者)，Northwestern Univ, Robert H Lurie Comprehens Canc Ctr, Chicago, IL 60611 USA.; Saratsis, A (通讯作者)，Ann & Robert H Lurie Childrens Hosp Chicago, Dept Surg, Div Pediat Neurosurg, Chicago, IL 60611 USA.
EM
RI ; Bartom, Elizabeth/S-4475-2017
OI Hou, Ye/0000-0001-9079-573X; Saratsis, Amanda/0000-0002-4913-2771; Yue,
   Feng/0000-0002-7954-5462; Bartom, Elizabeth/0000-0002-5618-2582; Wang,
   Juan/0000-0003-0933-3744
FU NIH's Natural Institute of Neurological Disease and Stroke
   [K08NS097264]; NIH [R35GM124820, R01HG009906, R50CA221848]
FX Research reported in this publication was supported by the NIH's Natural
   Institute of Neurological Disease and Stroke, grant number K08NS097264
   (to A.S.). F.Y. is supported by NIH grants R35GM124820 and R01HG009906.
   E.B. is supported by the NIH grant R50CA221848. The content is solely
   the responsibility of the authors and does not necessarily represent the
   official view of the NIH.
CR Anderson JL, 2017, NEURO-ONCOLOGY, V19, P1068, DOI 10.1093/neuonc/now299
   Arabzade A, 2020, NEUROPATH APPL NEURO, V46, P48, DOI 10.1111/nan.12612
   Asby DJ, 2018, CANCER MANAG RES, V10, P3483, DOI 10.2147/CMAR.S167095
   Bai LC, 2017, CANCER RES, V77, P2476, DOI 10.1158/0008-5472.CAN-16-2622
   Barrow J, 2011, NEURO-ONCOLOGY, V13, P212, DOI 10.1093/neuonc/noq158
   Buchanan JA, 2008, GENET MED, V10, P639, DOI [10.1097/GIM.0b013e318183f848, 10.1097GIM.0b013e318183f848]
   Carro MS, 2010, NATURE, V463, P318, DOI 10.1038/nature08712
   Cheng ZX, 2013, CLIN CANCER RES, V19, P1748, DOI 10.1158/1078-0432.CCR-12-3066
   Daubon T, 2019, NAT COMMUN, V10, DOI 10.1038/s41467-019-08480-y
   Dixon JR, 2018, NAT GENET, V50, P1388, DOI 10.1038/s41588-018-0195-8
   Filbin MG, 2018, SCIENCE, V360, P331, DOI 10.1126/science.aao4750
   Flavahan WA, 2016, NATURE, V529, P110, DOI 10.1038/nature16490
   Fortin J, 2020, CANCER CELL, V37, P308, DOI 10.1016/j.ccell.2020.02.002
   Funato K, 2014, SCIENCE, V346, P1529, DOI 10.1126/science.1253799
   Haddad N, 2017, CHROMOSOME RES, V25, P35, DOI 10.1007/s10577-016-9548-2
   Huang JL, 2015, GENOME BIOL, V16, DOI 10.1186/s13059-015-0740-z
   Jones C, 2017, NEURO-ONCOLOGY, V19, P153, DOI 10.1093/neuonc/now101
   Jung E, 2019, NAT NEUROSCI, V22, P1951, DOI 10.1038/s41593-019-0540-y
   Kosty J, 2017, CELL CYCLE, V16, P1654, DOI 10.1080/15384101.2017.1361062
   Labreche K, 2015, NAT COMMUN, V6, DOI 10.1038/ncomms8207
   Lu J, 2015, CHEM BIOL, V22, P755, DOI 10.1016/j.chembiol.2015.05.009
   Lulla RR, 2016, SCI ADV, V2, DOI 10.1126/sciadv.1501354
   Mack SC, 2019, J EXP MED, V216, P1071, DOI 10.1084/jem.20190196
   Mackay A, 2017, CANCER CELL, V32, P520, DOI 10.1016/j.ccell.2017.08.017
   Mohammad F, 2017, NAT MED, V23, P483, DOI 10.1038/nm.4293
   Morgan SL, 2017, NAT COMMUN, V8, DOI 10.1038/ncomms15993
   Morton AR, 2019, CELL, V179, P1330, DOI 10.1016/j.cell.2019.10.039
   Nagaraja S, 2019, MOL CELL, V76, P965, DOI 10.1016/j.molcel.2019.08.030
   Northcott PA, 2014, NATURE, V511, P428, DOI 10.1038/nature13379
   Piunti A, 2017, NAT MED, V23, P493, DOI 10.1038/nm.4296
   Plass C, 2013, NAT REV GENET, V14, P765, DOI 10.1038/nrg3554
   Qiu JG, 2017, DRUG DISCOV TODAY, V22, P148, DOI 10.1016/j.drudis.2016.09.017
   Rowley MJ, 2017, MOL CELL, V67, P837, DOI 10.1016/j.molcel.2017.07.022
   Salameh TJ, 2020, NAT COMMUN, V11, DOI 10.1038/s41467-020-17239-9
   Spielmann M, 2018, NAT REV GENET, V19, P453, DOI 10.1038/s41576-018-0007-0
   Stafford JM, 2018, SCI ADV, V4, DOI 10.1126/sciadv.aau5935
   Suva ML, 2014, CELL, V157, P580, DOI 10.1016/j.cell.2014.02.030
   Towner RA, 2013, NEURO-ONCOLOGY, V15, P1625, DOI 10.1093/neuonc/not124
   Wang S, 2020, GENOME BIOL, V21, DOI 10.1186/s13059-020-01986-5
   Warren KE, 2012, NEURO-ONCOLOGY, V14, P326, DOI 10.1093/neuonc/nor190
   Wen NY, 2019, INT J ONCOL, V55, P879, DOI 10.3892/ijo.2019.4863
   Winter GE, 2015, SCIENCE, V348, P1376, DOI 10.1126/science.aab1433
   Xu L, 2018, P NATL ACAD SCI USA, V115, pE5086, DOI 10.1073/pnas.1712363115
   Zhang SC, 2017, CANCER CELL, V31, P591, DOI 10.1016/j.ccell.2017.02.013
NR 44
TC 27
Z9 27
U1 1
U2 16
PU AMER ASSOC ADVANCEMENT SCIENCE
PI WASHINGTON
PA 1200 NEW YORK AVE, NW, WASHINGTON, DC 20005 USA
SN 2375-2548
J9 SCI ADV
JI Sci. Adv.
PD JUN
PY 2021
VL 7
IS 23
AR eabg4126
DI 10.1126/sciadv.abg4126
PG 16
WC Multidisciplinary Sciences
WE Science Citation Index Expanded (SCI-EXPANDED)
SC Science & Technology - Other Topics
GA SO1RQ
UT WOS:000658757100034
PM 34078608
OA Green Published
DA 2023-08-27
ER

PT J
AU Kerr, G
   Sheldon, H
   Chaikuad, A
   Alfano, I
   von Delft, F
   Bullock, AN
   Harris, AL
AF Kerr, Georgina
   Sheldon, Helen
   Chaikuad, Apirat
   Alfano, Ivan
   von Delft, Frank
   Bullock, Alex N.
   Harris, Adrian L.
TI A small molecule targeting ALK1 prevents Notch cooperativity and
   inhibits functional angiogenesis
SO ANGIOGENESIS
LA English
DT Article
DE ALK1; ACVRL1; BMP9; Notch; Angiogenesis; Hypersprouting
ID FIBRODYSPLASIA OSSIFICANS PROGRESSIVA; ACTIVATING ACVR1 MUTATIONS;
   INTRINSIC PONTINE GLIOMA; RECEPTOR-LIKE KINASE-1; TUMOR-GROWTH; DLL4;
   BMP; INTEGRATION; REFINEMENT; THERAPY
AB Activin receptor-like kinase 1 (ALK1, encoded by the gene ACVRL1) is a type I BMP/TGF-beta receptor that mediates signalling in endothelial cells via phosphorylation of SMAD1/5/8. During angiogenesis, sprouting endothelial cells specialise into tip cells and stalk cells. ALK1 synergises with Notch in stalk cells to induce expression of the Notch targets HEY1 and HEY2 and thereby represses tip cell formation and angiogenic sprouting. The ALK1-Fc soluble protein fusion has entered clinic trials as a therapeutic strategy to sequester the high-affinity extracellular ligand BMP9. Here, we determined the crystal structure of the ALK1 intracellular kinase domain and explored the effects of a small molecule kinase inhibitor K02288 on angiogenesis. K02288 inhibited BMP9-induced phosphorylation of SMAD1/5/8 in human umbilical vein endothelial cells to reduce both the SMAD and the Notch-dependent transcriptional responses. In endothelial sprouting assays, K02288 treatment induced a hypersprouting phenotype reminiscent of Notch inhibition. Furthermore, K02288 caused dysfunctional vessel formation in a chick chorioallantoic membrane assay of angiogenesis. Such activity may be advantageous for small molecule inhibitors currently in preclinical development for specific BMP gain of function conditions, including diffuse intrinsic pontine glioma and fibrodysplasia ossificans progressiva, as well as more generally for other applications in tumour biology.
C1 [Kerr, Georgina; Chaikuad, Apirat; Alfano, Ivan; von Delft, Frank; Bullock, Alex N.] Univ Oxford, Struct Genom Consortium, Oxford OX3 7DQ, England.
   [Sheldon, Helen; Harris, Adrian L.] Univ Oxford, John Radcliffe Hosp, Weatherall Inst Mol Med, Oxford OX3 9DS, England.
C3 University of Oxford; University of Oxford
RP Bullock, AN (通讯作者)，Univ Oxford, Struct Genom Consortium, Old Rd Campus Res Bldg,Roosevelt Dr, Oxford OX3 7DQ, England.
EM
RI Harris, Adrian L/ABA-3343-2020
OI Harris, Adrian L/0000-0003-1376-8409; Chaikuad,
   Apirat/0000-0003-1120-2209; Sheldon, Helen/0000-0002-0945-5496; von
   Delft, Frank/0000-0003-0378-0017
FU Cancer Research United Kingdom (CR-UK) through the Oxford Cancer
   Research Centre Development Fund [C38302/A12278]; AbbVie; Bayer;
   Boehringer Ingelheim; Genome Canada through Ontario Genomics Institute
   Grant [OGI-055]; GlaxoSmithKline; Janssen; Lilly Canada; Novartis
   Research Foundation; Ontario Ministry of Economic Development and
   Innovation; Pfizer; Takeda; Wellcome Trust Grant [092809/Z/10/Z];
   Wellcome Trust [092809/Z/10/Z] Funding Source: Wellcome Trust; Cancer
   Research UK [11359, 16466, 18974] Funding Source: researchfish; National
   Institute for Health Research [NF-SI-0611-10163] Funding Source:
   researchfish
FX The authors would like to thank Diamond Light Source for beamtime
   (proposal mx442), and the staff of beamline I24 for assistance with
   crystal testing and data collection. We thank Greg Cuny (University of
   Houston), Paul Yu (Harvard University) and Ji-Liang Li (University of
   Oxford) for assistance with reagents. This work was supported by Cancer
   Research United Kingdom (CR-UK), including grant number C38302/A12278,
   through the Oxford Cancer Research Centre Development Fund. The SGC is a
   registered charity (No. 1097737) that receives funds from AbbVie, Bayer,
   Boehringer Ingelheim, Genome Canada through Ontario Genomics Institute
   Grant OGI-055, GlaxoSmithKline, Janssen, Lilly Canada, the Novartis
   Research Foundation, the Ontario Ministry of Economic Development and
   Innovation, Pfizer, Takeda, and Wellcome Trust Grant 092809/Z/10/Z.
CR Buczkowicz P, 2014, NAT GENET, V46, P451, DOI 10.1038/ng.2936
   Chaikuad A, 2012, J BIOL CHEM, V287, P36990, DOI 10.1074/jbc.M112.365932
   Cunha SI, 2010, J EXP MED, V207, P85, DOI 10.1084/jem.20091309
   Davis IW, 2007, NUCLEIC ACIDS RES, V35, pW375, DOI 10.1093/nar/gkm216
   Emsley P, 2010, ACTA CRYSTALLOGR D, V66, P486, DOI 10.1107/S0907444910007493
   Evans P, 2006, ACTA CRYSTALLOGR D, V62, P72, DOI 10.1107/S0907444905036693
   Fontebasso AM, 2014, NAT GENET, V46, P462, DOI 10.1038/ng.2950
   Hawinkels LJAC, 2013, EXPERT OPIN INV DRUG, V22, P1371, DOI 10.1517/13543784.2013.837884
   Hellstrom M, 2007, NATURE, V445, P776, DOI 10.1038/nature05571
   Johnson DW, 1996, NAT GENET, V13, P189, DOI 10.1038/ng0696-189
   Kabsch W, 2010, ACTA CRYSTALLOGR D, V66, P133, DOI 10.1107/S0907444909047374
   Larrivee B, 2012, DEV CELL, V22, P489, DOI 10.1016/j.devcel.2012.02.005
   Li JL, 2011, CANCER RES, V71, P6073, DOI 10.1158/0008-5472.CAN-11-1704
   Li JL, 2010, FUTURE ONCOL, V6, P1099, DOI [10.2217/fon.10.62, 10.2217/FON.10.62]
   Mccoy AJ, 2007, J APPL CRYSTALLOGR, V40, P658, DOI 10.1107/S0021889807021206
   Mohedas AH, 2013, ACS CHEM BIOL, V8, P1291, DOI 10.1021/cb300655w
   Moya IM, 2012, DEV CELL, V22, P501, DOI 10.1016/j.devcel.2012.01.007
   Murshudov GN, 1997, ACTA CRYSTALLOGR D, V53, P240, DOI 10.1107/S0907444996012255
   Nakatsu Martin N, 2007, J Vis Exp, P186, DOI 10.3791/186
   Noguera-Troise I, 2006, NATURE, V444, P1032, DOI 10.1038/nature05355
   Oda T, 1997, NAT GENET, V16, P235, DOI 10.1038/ng0797-235
   Ridgway J, 2006, NATURE, V444, P1083, DOI 10.1038/nature05313
   Sanvitale CE, 2013, PLOS ONE, V8, DOI 10.1371/journal.pone.0062721
   Shore EM, 2006, NAT GENET, V38, P525, DOI 10.1038/ng1783
   Siekmann AF, 2007, NATURE, V445, P781, DOI 10.1038/nature05577
   Taylor KR, 2014, NAT GENET, V46, P457, DOI 10.1038/ng.2925
   Upton PD, 2009, J BIOL CHEM, V284, P15794, DOI 10.1074/jbc.M109.002881
   Wu G, 2014, NAT GENET, V46, P444, DOI 10.1038/ng.2938
NR 28
TC 47
Z9 49
U1 0
U2 8
PU SPRINGER
PI DORDRECHT
PA VAN GODEWIJCKSTRAAT 30, 3311 GZ DORDRECHT, NETHERLANDS
SN 0969-6970
EI 1573-7209
J9 ANGIOGENESIS
JI Angiogenesis
PD APR
PY 2015
VL 18
IS 2
BP 209
EP 217
DI 10.1007/s10456-014-9457-y
PG 9
WC Peripheral Vascular Disease
WE Science Citation Index Expanded (SCI-EXPANDED)
SC Cardiovascular System & Cardiology
GA CE0DT
UT WOS:000351475400010
PM 25557927
OA Green Submitted, Green Published, hybrid
DA 2023-08-27
ER

PT J
AU DeBoer, MD
   Zhu, XX
   Levasseur, P
   Meguid, MM
   Suzuki, S
   Inui, A
   Taylor, JE
   Halem, HA
   Dong, JZ
   Datta, R
   Culler, MD
   Marks, DL
AF DeBoer, Mark D.
   Zhu, Xin Xia
   Levasseur, Peter
   Meguid, Michael M.
   Suzuki, Susumu
   Inui, Akio
   Taylor, John E.
   Halem, Heather A.
   Dong, Jesse Z.
   Datta, Rakesh
   Culler, Michael D.
   Marks, Daniel L.
TI Ghrelin treatment causes increased food intake and retention of lean
   body mass in a rat model of cancer cachexia
SO ENDOCRINOLOGY
LA English
DT Article
ID TUMOR-NECROSIS-FACTOR; PLASMA GHRELIN; GASTRIC BYPASS; APPETITE;
   ANOREXIA; EXPRESSION; RECEPTORS; CYTOKINES; SUPPLEMENTATION;
   INFLAMMATION
AB Cancer cachexia is a debilitating syndrome of anorexia and loss of lean body mass that accompanies many malignancies. Ghrelin is an orexigenic hormone with a short half-life that has been shown to improve food intake and weight gain in human and animal subjects with cancer cachexia. We used a rat model of cancer cachexia and administered human ghrelin and a synthetic ghrelin analog BIM-28131 via continuous infusion using sc osmotic minipumps. Tumor-implanted rats receiving human ghrelin or BIM-28131 exhibited a significant increase in food consumption and weight gain vs. saline-treated animals. We used dual-energy x-ray absorptiometry scans to show that the increased weight was due to maintenance of lean mass vs. a loss of lean mass in saline-treated animals. Also, BIM-28131 significantly limited the loss of fat mass normally observed in tumor-implanted rats. We further performed real-time PCR analysis of the hypothalami and brainstems and found that ghrelin-treated animals exhibited a significant increase in expression of orexigenic peptides agouti-related peptide and neuropeptide Y in the hypothalamus and a significant decrease in the expression of IL-1 receptor-I transcript in the hypothalamus and brainstem. We conclude that ghrelin and a synthetic ghrelin receptor agonist improve weight gain and lean body mass retention via effects involving orexigenic neuropeptides and antiinflammatory changes.
C1 Oregon Hlth & Sci Univ, Ctr Study Weight Regulat, Portland, OR 97239 USA.
   SUNY Upstate Med Univ, Dept Surg, Syracuse, NY 13210 USA.
   SUNY Upstate Med Univ, Surg Metab & Nutr Lab, Syracuse, NY 13210 USA.
   Kagoshima Univ, Grad Sch Med & Dent Sci, Dept Behav Med, Kagoshima 8908520, Japan.
   IPSEN, Milford, MA 01757 USA.
C3 Oregon Health & Science University; State University of New York (SUNY)
   System; State University of New York (SUNY) Upstate Medical Center;
   State University of New York (SUNY) System; State University of New York
   (SUNY) Upstate Medical Center; Kagoshima University; Ipsen
RP Marks, DL (通讯作者)，707 SW Gaines Rd,CDRC-P, Portland, OR 97239 USA.
EM
RI Marks, Daniel/C-3712-2009
OI Culler, Michael D./0000-0001-5168-8739
FU NIDDK NIH HHS [R01 DK 70333-01, F32 DK 072820-01A1, 1K08 DK 062207-01]
   Funding Source: Medline; PHS HHS [DK/NCI 43796/70239] Funding Source:
   Medline
CR Abad LW, 2002, CYTOKINE, V18, P286, DOI 10.1006/cyto.2002.0893
   Akamizu T, 2004, EUR J ENDOCRINOL, V150, P447, DOI 10.1530/eje.0.1500447
   Andre R, 2005, J NEUROCHEM, V95, P324, DOI 10.1111/j.1471-4159.2005.03364.x
   BONFIELD TL, 1995, AM J RESP CRIT CARE, V152, P2111, DOI 10.1164/ajrccm.152.6.8520783
   Cone RD, 2005, NAT NEUROSCI, V8, P571, DOI 10.1038/nn1455
   Cummings DE, 2002, NEW ENGL J MED, V346, P1623, DOI 10.1056/NEJMoa012908
   DATTA R, 2005, P 3 INT CACH C ROM I
   Deans C, 2005, CURR OPIN CLIN NUTR, V8, P265, DOI 10.1097/01.mco.0000165004.93707.88
   DeBoer MD, 2006, TRENDS ENDOCRIN MET, V17, P199, DOI 10.1016/j.tem.2006.05.005
   Docagne F, 2005, EUR J NEUROSCI, V21, P1205, DOI 10.1111/j.1460-9568.2005.03965.x
   Eriksson C, 2000, CYTOKINE, V12, P423, DOI 10.1006/cyto.1999.0582
   Fouladiun M, 2005, CANCER-AM CANCER SOC, V103, P2189, DOI 10.1002/cncr.21013
   GABELICA Z, 1995, P 8 SEM BRAS CAT AN, V1, P1
   Garcia JM, 2005, J CLIN ENDOCR METAB, V90, P2920, DOI 10.1210/jc.2004-1788
   Granado M, 2005, AM J PHYSIOL-ENDOC M, V288, pE486, DOI 10.1152/ajpendo.00196.2004
   Guijarro A, 2006, CURR OPIN CLIN NUTR, V9, P423, DOI 10.1097/01.mco.0000232903.04910.7b
   HALEM HA, 2004, 86 ANN M END SOC NEW, P34
   HALEM HA, 2005, 87 ANN M END SOC SAN, P407
   Hanada T, 2004, METABOLISM, V53, P84, DOI 10.1016/j.metabol.2003.06.004
   Hanada T, 2003, BIOCHEM BIOPH RES CO, V301, P275, DOI 10.1016/S0006-291X(02)03028-0
   Huang Q, 2005, BRIT J NUTR, V93, P853, DOI 10.1079/BJN20051412
   Inui A, 2004, FASEB J, V18, P439, DOI 10.1096/fj.03-0641rev
   Inui A, 2003, CURR OPIN CLIN NUTR, V6, P395, DOI 10.1097/01.mco.0000078989.18774.74
   Inui A, 2002, DIABETES OBES METAB, V4, P431, DOI 10.1046/j.1463-1326.2002.00228.x
   Jatoi A, 2002, J CLIN ONCOL, V20, P567, DOI 10.1200/JCO.20.2.567
   Kojima M, 1999, NATURE, V402, P656, DOI 10.1038/45230
   LEVINE B, 1990, NEW ENGL J MED, V323, P236, DOI 10.1056/NEJM199007263230405
   Marks DL, 2001, CANCER RES, V61, P1432
   May PE, 2002, AM J SURG, V183, P471, DOI 10.1016/S0002-9610(02)00823-1
   MCCARTHY HD, 1995, AM J PHYSIOL-ENDOC M, V269, pE852, DOI 10.1152/ajpendo.1995.269.5.E852
   Nagaya N, 2001, CIRCULATION, V104, P1430, DOI 10.1161/hc3601.095575
   Nakazato M, 2001, NATURE, V409, P194, DOI 10.1038/35051587
   Neary NM, 2004, J CLIN ENDOCR METAB, V89, P2832, DOI 10.1210/jc.2003-031768
   NIXON DW, 1980, AM J MED, V68, P683, DOI 10.1016/0002-9343(80)90254-5
   Otto B, 2004, EUR J ENDOCRINOL, V151, P113, DOI 10.1530/eje.0.1510113
   Plata-Salaman C, 2001, INT J OBESITY, V25, pS48, DOI 10.1038/sj.ijo.0801911
   PLATASALAMAN CR, 1988, BRAIN RES, V448, P106, DOI 10.1016/0006-8993(88)91106-7
   Qureshi AR, 2002, J AM SOC NEPHROL, V13, pS28
   Ramos EJB, 2004, J AM COLL SURGEONS, V199, P716, DOI 10.1016/j.jamcollsurg.2004.07.014
   Ramos EJB, 2004, CURR OPIN CLIN NUTR, V7, P427, DOI 10.1097/01.mco.0000134363.53782.cb
   ROUBENOFF R, 1994, J CLIN INVEST, V93, P2379, DOI 10.1172/JCI117244
   Sato T, 2001, AM J PHYSIOL-REG I, V281, pR1907, DOI 10.1152/ajpregu.2001.281.6.R1907
   Shimizu Y, 2003, CLIN CANCER RES, V9, P774
   Simons JPFHA, 1998, CANCER, V82, P553, DOI 10.1002/(SICI)1097-0142(19980201)82:3<553::AID-CNCR18>3.0.CO;2-0
   Sun YX, 2003, MOL CELL BIOL, V23, P7973, DOI 10.1128/MCB.23.22.7973-7981.2003
   Sun YX, 2004, P NATL ACAD SCI USA, V101, P4679, DOI 10.1073/pnas.0305930101
   Tisdale MJ, 2002, NAT REV CANCER, V2, P862, DOI 10.1038/nrc927
   Tisdale MJ, 1997, J NATL CANCER I, V89, P1763, DOI 10.1093/jnci/89.23.1763
   TorreAmione G, 1996, CIRCULATION, V93, P704, DOI 10.1161/01.CIR.93.4.704
   Vigano A, 2004, CANCER, V101, P1090, DOI 10.1002/cncr.20472
   Wigmore SJ, 1997, CLIN SCI, V92, P215, DOI 10.1042/cs0920215
   Wisse BE, 2001, ENDOCRINOLOGY, V142, P3292, DOI 10.1210/en.142.8.3292
   Wolf I, 2006, CANCER-AM CANCER SOC, V106, P966, DOI 10.1002/cncr.21690
   Wren AM, 2001, J CLIN ENDOCR METAB, V86, P5992, DOI 10.1210/jc.86.12.5992
   Wren AM, 2001, DIABETES, V50, P2540, DOI 10.2337/diabetes.50.11.2540
NR 55
TC 139
Z9 145
U1 0
U2 14
PU ENDOCRINE SOC
PI WASHINGTON
PA 2055 L ST NW, SUITE 600, WASHINGTON, DC 20036 USA
SN 0013-7227
EI 1945-7170
J9 ENDOCRINOLOGY
JI Endocrinology
PD JUN
PY 2007
VL 148
IS 6
BP 3004
EP 3012
DI 10.1210/en.2007-0016
PG 9
WC Endocrinology & Metabolism
WE Science Citation Index Expanded (SCI-EXPANDED)
SC Endocrinology & Metabolism
GA 169DH
UT WOS:000246572400052
PM 17347304
OA Bronze
DA 2023-08-27
ER

PT J
AU HARPER, CM
   HARNER, SG
   SLAVIT, DH
   LITCHY, WJ
   DAUBE, JR
   BEATTY, CW
   EBERSOLD, MJ
AF HARPER, CM
   HARNER, SG
   SLAVIT, DH
   LITCHY, WJ
   DAUBE, JR
   BEATTY, CW
   EBERSOLD, MJ
TI EFFECT OF BAEP MONITORING ON HEARING PRESERVATION DURING ACOUSTIC
   NEUROMA RESECTION
SO NEUROLOGY
LA English
DT Article
ID AUDITORY EVOKED-POTENTIALS; MICROVASCULAR DECOMPRESSION; HEMIFACIAL
   SPASM; REMOVAL; NERVE
AB We measured the effect of brainstem auditory evoked potential (BAEP) monitoring on hearing preservation. in acoustic neuroma resection in 90 consecutive patients with monitoring compared with 90 historical controls matched for tumor size and preoperative hearing status. In small tumors (<2 cm), BAEP monitoring was associated with a higher rate of hearing preservation and a greater chance that the hearing preserved was clinically useful. Changes in the BAEP intraoperatively showed a good correlation with postoperative hearing status.
C1 MAYO CLIN & MAYO FDN,DEPT OTOLARYNGOL,ROCHESTER,MN 55905.
   MAYO CLIN & MAYO FDN,DEPT NEUROSURG,ROCHESTER,MN 55905.
C3 Mayo Clinic; Mayo Clinic
RP HARPER, CM (通讯作者)，MAYO CLIN & MAYO FDN,DEPT NEUROL,200 1ST ST SW,ROCHESTER,MN 55905, USA.
CR ABRAMSON M, 1985, LARYNGOSCOPE, V95, P1318
   AUGER RG, 1986, MAYO CLIN PROC, V61, P640, DOI 10.1016/S0025-6196(12)62028-X
   HARNER SG, 1988, LARYNGOSCOPE, V98, P209
   HARNER SG, 1985, J NEUROSURG, V63, P168
   MOLLER AR, 1988, ELECTROEN CLIN NEURO, V70, P248, DOI 10.1016/0013-4694(88)90085-5
   MOLLER AR, 1989, NEUROSURGERY, V24, P257
   MOLLER MB, 1985, J NEUROSURG, V63, P17, DOI 10.3171/jns.1985.63.1.0017
   NADOL JB, 1987, LARYNGOSCOPE, V98, P118
   OJEMANN RG, 1984, J NEUROSURG, V61, P938, DOI 10.3171/jns.1984.61.5.0938
   RADTKE RA, 1989, NEUROLOGY, V39, P187, DOI 10.1212/WNL.39.2.187
   SEKIYA T, 1987, J NEUROSURG, V67, P244, DOI 10.3171/jns.1987.67.2.0244
   SLAVIT DH, 1991, ARCH OTOLARYNGOL, V117, P1153
   TATOR CH, 1985, J NEUROSURG, V63, P168, DOI 10.3171/jns.1985.63.2.0168
   WATANABE E, 1989, ACTA NEUROCHIR, V98, P118, DOI 10.1007/BF01407337
NR 14
TC 38
Z9 38
U1 0
U2 3
PU LIPPINCOTT-RAVEN PUBL
PI PHILADELPHIA
PA 227 EAST WASHINGTON SQ, PHILADELPHIA, PA 19106
SN 0028-3878
J9 NEUROLOGY
JI Neurology
PD AUG
PY 1992
VL 42
IS 8
BP 1551
EP 1553
DI 10.1212/WNL.42.8.1551
PG 3
WC Clinical Neurology
WE Science Citation Index Expanded (SCI-EXPANDED)
SC Neurosciences & Neurology
GA JH221
UT WOS:A1992JH22100021
PM 1641152
DA 2023-08-27
ER

PT J
AU Kleinschmidt-DeMasters, BK
   Levy, JMM
AF Kleinschmidt-DeMasters, Bette K.
   Levy, Jean M. Mulcahy
TI H3 K27M-mutant gliomas in adults vs. children share similar histological
   features and adverse prognosis
SO CLINICAL NEUROPATHOLOGY
LA English
DT Article
DE pilocytic astrocytoma; ganglioglioma; midline; pons; elderly
ID H3F3A K27M MUTATION; MARKER; RARE
AB Background: H3 K27M mutation was originally described in pediatric diffuse intrinsic pontine gliomas (DIPGs), but has been recently recognized to occur also in adult midline diffuse gliomas, as well as midline tumors with other morphologies, including gangliogliomas (GGs), anaplastic GGs, pilocytic astrocytomas (PAs), and posterior fossa ependymomas. In a few patients with H3 K27M-mutant tumors with these alternate morphologies, longer survival has been reported, making grading difficult for the neuropathologist. Few series compare tumors in adult vs. pediatric cohorts; we report our 4-year experience. Materials and methods: Text Word database searches using "H3 K27M" in reports generated between January 2013 and November 10, 2017 were used to identify patients. Clinical and histological features as well as survival were evaluated for each case. Results: 28 H3 K27M-mutant tumors were identified, with equal numbers of adults (13) vs. children (15). For adults, mean and median age was 52 years (range = 27 - 81 years), 2 decades older than a recently-published adult series. Tumors involved thalamic (adult = 7; pediatric = 7), spinal cord (adult = 4: pediatric = 2), pons (adult = 1; pediatric = 6), and hypothalamic (n = 1) sites. Other morphologies at presentation included pure GG (n = 3, pediatric) and PA (n = 1, adult). One adult and 1 pediatric patient each presented with leptomeningeal dissemination or developed leptomeningeal dissemination within 1 year after diagnosis. with transformation from PA or GG histology to glioblastoma. Mean survival was 9.3 (adults) vs. 8.9 (pediatric) months. Patients with tumors of other morphologies (GG, PA) did not enjoy extended survival. Conclusion: H3 K27M-mutant tumors can affect patients at advanced ages. may show leptomeningeal dissemination at time of presentation. and "pure" GG or PA morphology is not rare. Regardless of patient age or tumor morphology, patients fare equally poorly.
C1 [Kleinschmidt-DeMasters, Bette K.] Univ Colorado, Sch Med, Dept Pathol, Boulder, CO 80309 USA.
   [Kleinschmidt-DeMasters, Bette K.] Univ Colorado, Sch Med, Dept Neurol, Aurora, CO 80309 USA.
   [Kleinschmidt-DeMasters, Bette K.] Univ Colorado, Sch Med, Dept Neurosurg, Aurora, CO 80309 USA.
   [Levy, Jean M. Mulcahy] Univ Colorado, Sch Med, Dept Pharmacol, Aurora, CO 80309 USA.
   [Levy, Jean M. Mulcahy] Univ Colorado, Sch Med, Dept Pediat, Aurora, CO 80309 USA.
   [Levy, Jean M. Mulcahy] Childrens Hosp Colorado, Morgan Adams Fdn Pediat, Brain Tumor Res Program, Aurora, CO USA.
C3 University of Colorado System; University of Colorado Anschutz Medical
   Campus; University of Colorado Boulder; University of Colorado System;
   University of Colorado Anschutz Medical Campus; University of Colorado
   System; University of Colorado Anschutz Medical Campus; University of
   Colorado System; University of Colorado Anschutz Medical Campus;
   University of Colorado System; University of Colorado Anschutz Medical
   Campus; Children's Hospital Colorado
RP Kleinschmidt-DeMasters, BK (通讯作者)，Univ Colorado, Sch Med, 12605 E 16th Ave,MS F768,Room 3017, Aurora, CO 80045 USA.
EM
FU NIH/NCI [K08CA193982]; Morgan Adams Foundation
FX Support provided by: NIH/NCI (K08CA193982), and the Morgan Adams
   Foundation (JML).
CR Buczkowicz P, 2014, ACTA NEUROPATHOL, V128, P573, DOI 10.1007/s00401-014-1319-6
   Buczkowicz P, 2014, NAT GENET, V46, P451, DOI 10.1038/ng.2936
   Bugiani M, 2017, ONCOTARGET, V8, P60447, DOI 10.18632/oncotarget.19726
   Caretti V, 2014, ACTA NEUROPATHOL, V128, P605, DOI 10.1007/s00401-014-1307-x
   Gao Y, 2017, NEUROPATHOLOGY
   Gessi M, 2016, ACTA NEUROPATHOL, V132, P635, DOI 10.1007/s00401-016-1608-3
   Gielen GH, 2013, AM J CLIN PATHOL, V139, P345, DOI 10.1309/AJCPABOHBC33FVMO
   Hamano E, 2017, J NEUROSURG, V126, P1, DOI 10.3171/2015.10.JNS151022
   Haque F, 2017, ACTA NEUROPATHOL COM, V5, DOI 10.1186/s40478-017-0449-1
   Hawkins C, 2016, WHO CLASSIFICATION T, P57
   Hochart A, 2015, ANN CLIN TRANSL NEUR, V2, P439, DOI 10.1002/acn3.184
   Joyon N, 2017, NEUROPATH APPL NEURO, V43, P271, DOI 10.1111/nan.12329
   Karremann M, 2017, NEUROONCOL
   Kleinschmidt-DeMasters BK, 2017, BRAIN PATHOL, V27, P846, DOI 10.1111/bpa.12455
   Lopez G, 2017, ACTA NEUROPATHOL COM, V5, DOI 10.1186/s40478-017-0440-x
   Lopez GY, 2017, ACTA NEUROPATHOL, V134, P961, DOI 10.1007/s00401-017-1780-0
   Meyronet D, 2017, NEURO-ONCOLOGY, V19, P1127, DOI 10.1093/neuonc/now274
   Nakata S, 2017, BRAIN TUMOR PATHOL, V34, P113, DOI 10.1007/s10014-017-0288-6
   Orillac C, 2016, ACTA NEUROPATHOL COM, V4, DOI 10.1186/s40478-016-0361-0
   Pages M, 2018, BRAIN PATHOL, V28, P103, DOI 10.1111/bpa.12473
   Ryall S, 2017, CHILD NERV SYST, V33, P1047, DOI 10.1007/s00381-017-3481-3
   Shows J, 2016, BRAIN PATHOL, V26, P120, DOI 10.1111/bpa.12327
   Solomon DA, 2016, BRAIN PATHOL, V26, P569, DOI 10.1111/bpa.12336
   Tinkle CL, 2017, PEDIAT BLOOD CANC, V64, P64
   Venneti S, 2014, ACTA NEUROPATHOL, V128, P743, DOI 10.1007/s00401-014-1338-3
   Zanello M, 2016, J NEUROPATH EXP NEUR, V75, P971, DOI 10.1093/jnen/nlw074
NR 26
TC 70
Z9 76
U1 0
U2 3
PU DUSTRI-VERLAG DR KARL FEISTLE
PI DEISENHOFEN-MUENCHEN
PA BAHNHOFSTRASSE 9 POSTFACH 49, D-82032 DEISENHOFEN-MUENCHEN, GERMANY
SN 0722-5091
J9 CLIN NEUROPATHOL
JI Clin. Neuropathol.
PD MAR-APR
PY 2018
VL 37
IS 2
BP 53
EP 63
DI 10.5414/NP301085
PG 11
WC Clinical Neurology; Pathology
WE Science Citation Index Expanded (SCI-EXPANDED)
SC Neurosciences & Neurology; Pathology
GA HK6UF
UT WOS:000458119400003
PM 29393845
OA Green Submitted, Green Published
DA 2023-08-27
ER

PT J
AU Gruber, A
   Leschnik, M
   Kneissl, S
   Schmidt, P
AF Gruber, A.
   Leschnik, M.
   Kneissl, S.
   Schmidt, P.
TI Gliomatosis cerebri in a dog
SO JOURNAL OF VETERINARY MEDICINE SERIES A-PHYSIOLOGY PATHOLOGY CLINICAL
   MEDICINE
LA English
DT Article
ID BRAIN-TUMORS
AB Gliomatosis cerebri is a rare tumourous disease of the central nervous system consisting of glial cells characterized by diffuse widespread infiltration and preservation of preformed brain structures. This report describes a case of gliomatosis cerebri in a 9-year-old, male, flat-coated retriever dog with central nervous symptoms. Magnetic resonance imaging of the brain demonstrated ill-defined hyperintense zones of paramedian white matter within the right cerebral hemisphere, brainstem and cerebellum. Histological examination showed a proliferation of tumour cells in the brain structures mentioned. Glial fibrillary acidic protein-immunohistochemistry revealed a lot of fibres positively stained. Based on these findings, the alteration was diagnosed as gliomatosis cerebri.
C1 Univ Vet Med Vienna, Inst Pathol & Forens Vet Med, Dept Pathobiol, A-1210 Vienna, Austria.
   Univ Vet Med Vienna, Dept Small Anim & Horses, Clin Internal Med & Infect Dis, A-1210 Vienna, Austria.
   Univ Vet Med Vienna, Clin Radiol, Dept Diagnost Imaging, Infect Dis & Lab Med, A-1210 Vienna, Austria.
C3 University of Veterinary Medicine Vienna; University of Veterinary
   Medicine Vienna; University of Veterinary Medicine Vienna
RP Gruber, A (通讯作者)，Univ Vet Med Vienna, Inst Pathol & Forens Vet Med, Dept Pathobiol, Vet Pl 1, A-1210 Vienna, Austria.
EM
RI Kneissl, Sibylle Maria/ABD-8091-2021; Kneissl, Sibylle/HJG-6480-2022
OI Kneissl, Sibylle Maria/0000-0001-9588-4715; 
CR Braun U, 2005, VET J, V170, P381, DOI 10.1016/j.tvjl.2004.09.010
   delCarpioODonovan R, 1996, RADIOLOGY, V198, P831, DOI 10.1148/radiology.198.3.8628879
   Hulette CM, 1996, MODERN PATHOL, V9, P316
   HULETTE CM, 1992, AM J SURG PATHOL, V16, P493, DOI 10.1097/00000478-199205000-00008
   IBAYASHI N, 1989, LAB INVEST, V61, P310
   KANDLER R H, 1991, British Journal of Neurosurgery, V5, P187, DOI 10.3109/02688699108998466
   Kim DG, 1998, ACTA NEUROCHIR, V140, P755, DOI 10.1007/s007010050176
   KLEIHUES P, 1993, BRAIN PATHOL, V3, P255, DOI 10.1111/j.1750-3639.1993.tb00752.x
   Koestner A, 1999, WHO INT HISTOLOGICAL, VV, P17
   Lantos PL, 2000, WHO CLASSIFICATION T, P92
   MANNOJI H, 1986, ACTA NEUROPATHOL, V71, P341, DOI 10.1007/BF00688060
   Marihart B, 2002, WIEN TIERARZTL MONAT, V89, P29
   PINSKER MO, 2004, MANUAL HIRNTUMOREN P, P84
   Porter B, 2003, VET PATHOL, V40, P97, DOI 10.1354/vp.40-1-97
   Stierstorfer B, 2002, TIERARZTL PRAX K H, V30, P282
   SUMMERS BA, 2001, DEP VET PATHOLOGY WE
   Summers BA., 1995, VET NEUROPATHOLOGY, P351
   Tancredi A, 2000, ACTA NEUROCHIR, V142, P469, DOI 10.1007/s007010050459
   VANDEVELDE M, 1985, ACTA NEUROPATHOL, V66, P111, DOI 10.1007/BF00688685
   Vates GE, 2003, NEUROSURGERY, V53, P261, DOI 10.1227/01.NEU.0000073527.20655.E6
   WILLARD MD, 1982, CORNELL VET, V72, P211
NR 21
TC 13
Z9 13
U1 0
U2 3
PU BLACKWELL PUBLISHING
PI OXFORD
PA 9600 GARSINGTON RD, OXFORD OX4 2DQ, OXON, ENGLAND
SN 0931-184X
J9 J VET MED A
JI J. Vet. Med. Ser. A-Physiol. Pathol. Clin. Med.
PD OCT
PY 2006
VL 53
IS 8
BP 435
EP 438
DI 10.1111/j.1439-0442.2006.00864.x
PG 4
WC Veterinary Sciences
WE Science Citation Index Expanded (SCI-EXPANDED)
SC Veterinary Sciences
GA 095BB
UT WOS:000241282100010
PM 16970635
DA 2023-08-27
ER

PT J
AU Badaloni, ME
   Johnson, AR
   Arganaraz, R
   Mantese, B
AF Badaloni, Maria Eugenia
   Johnson, Agustin Ruiz
   Arganaraz, Romina
   Mantese, Beatriz
TI Intratumoral catheter placement in pediatric patients with diffuse
   midline gliomas
SO CHILDS NERVOUS SYSTEM
LA English
DT Review
DE Diffuse midline glioma; Ommaya; Histone; Symptoms; Radiotherapy
ID RESERVOIRS; TUMORS
AB IntroductionDiffuse midline brainstem gliomas have a poor prognosis and are generally not amenable to surgical resection. Occasionally, palliative surgical procedures can be performed to improve the quality of life of these patients. We describe three patients with solid-cystic brainstem gliomas in whom an Ommaya reservoir catheter was placed to reduce mass effect.ObjectivesTo describe the characteristics, indications for, and operative technique of Ommaya reservoir catheter placement in patients with solid-cystic diffuse midline glioma.Materials and methodsA review was conducted of the medical records of pediatric patients with solid-cystic diffuse midline glioma H3 K27-altered, treated with an Ommaya reservoir at Hospital J.P. Garrahan between 2014 and 2021 together with a search of the literature.ResultsThree cases of stereotaxic Ommaya placement in solid-cystic diffuse midline gliomas, H3 K27M-altered were identified. After the procedure, clinical improvement and reduction of the size of the tumor cyst size was achieved. No associated complications were seen. At the time of the study, one patient died, and the remaining two patients continued in follow-up at our hospital.ConclusionWe believe that the placement of an intratumoral Ommaya reservoir catheter may be considered a therapeutic option to improve symptoms and quality of life of selected patients with solid-cystic diffuse midline glioma.
C1 [Badaloni, Maria Eugenia; Johnson, Agustin Ruiz; Arganaraz, Romina; Mantese, Beatriz] Hosp Pediat JP Garrahan, Buenos Aires, DF, Argentina.
C3 Hospital de Pediatria Doctor Juan Garrahan; University of Buenos Aires
RP Badaloni, ME (通讯作者)，Hosp Pediat JP Garrahan, Buenos Aires, DF, Argentina.
EM
CR Bannykh S, 2017, J NEUROSURG, P1
   Chiarelli P, 2017, PEDIATR NEUROSURG, P1
   Cooney TM, 2020, J NEURO-ONCOL, V150, P27, DOI 10.1007/s11060-020-03553-1
   Dhawan S, 2019, WORLD NEUROSURG, V127, P607, DOI 10.1016/j.wneu.2019.04.016
   Frio F, 2019, WORLD NEUROSURG, V132, DOI 10.1016/j.wneu.2019.07.217
   Hill CS, 2021, CHILD NERV SYST, V37, P1917, DOI 10.1007/s00381-021-05060-8
   Louis DN, 2021, NEURO-ONCOLOGY, V23, P1231, DOI 10.1093/neuonc/noab106
   McCrea J, 2015, PRINCIPLES PRACTICE, P553
   Oshima A, 2017, J CLIN NEUROSCI, V39, P199, DOI 10.1016/j.jocn.2016.12.042
   PACKER RJ, 1994, CURR OPIN NEUROL, V7, P484, DOI 10.1097/00019052-199412000-00003
   Puget S, 2015, CHILD NERV SYST, V31, P1773, DOI 10.1007/s00381-015-2832-1
   RAMASWAMY V, 2017, J NEUROSURG-PEDIATR, P1
   Srikandarajah N, 2014, BRIT J NEUROSURG, V28, P475, DOI 10.3109/02688697.2013.854314
   Yang XT, 2016, WORLD NEUROSURG, V89, P93, DOI 10.1016/j.wneu.2015.12.040
   Zhang YJ, 2021, ADV EXP MED BIOL, V1283, P1, DOI 10.1007/978-981-15-8104-5_1
NR 15
TC 0
Z9 0
U1 0
U2 0
PU SPRINGER
PI NEW YORK
PA ONE NEW YORK PLAZA, SUITE 4600, NEW YORK, NY, UNITED STATES
SN 0256-7040
EI 1433-0350
J9 CHILD NERV SYST
JI Childs Nerv. Syst.
PD AUG
PY 2023
VL 39
IS 8
BP 2065
EP 2070
DI 10.1007/s00381-023-05950-z
EA APR 2023
PG 6
WC Clinical Neurology; Pediatrics; Surgery
WE Science Citation Index Expanded (SCI-EXPANDED)
SC Neurosciences & Neurology; Pediatrics; Surgery
GA O0QU9
UT WOS:000971871400001
PM 37072561
DA 2023-08-27
ER

PT J
AU Wu, GY
   Zhang, RY
   Mao, HM
   Chen, YH
   Liu, GQ
   Zhang, J
AF Wu, Guangyu
   Zhang, Ruiyun
   Mao, Haiming
   Chen, Yonghui
   Liu, Guiqin
   Zhang, Jin
TI The value of blood oxygen level dependent (BOLD) imaging in evaluating
   post-operative renal function outcomes after laparoscopic partial
   nephrectomy
SO EUROPEAN RADIOLOGY
LA English
DT Article
DE Magnetic resonance imaging; Glomerular filtration rate; Renal cell
   carcinoma; Nephrectomy; Renal circulation
ID GLOMERULAR-FILTRATION; KIDNEY INJURY; MRI; DIFFUSION; ISCHEMIA; HYPOXIA;
   VOLUME
AB ObjectivesTo assess the ability of renal blood oxygen level dependent (BOLD) MRI metrics to predict post-operative renal function.MethodsWe studied 152 patients who underwent laparoscopic partial nephrectomy (LPN) and renal MRI examination including BOLD. Short-term and long-term renal function was evaluated using the glomerular filtration rate (GFR) derived from renal scintigraphy. Renal function decline was assessed as the absolute decline (AD), percentage decline (PD) and optimal renal function preservation (OP). T2* values were analysed in the renal cortex and medulla ipsilateral and contralateral to the tumour. Clinical characteristics and imaging metrics were evaluated using univariate and multivariate linear regression analyses. Risk factors obtained using BOLD metrics (determined by multivariate regression) were then combined and compared with RENAL scores to predict OP.ResultsIncreasing warm ischaemia time (WIT), resected and ischaemic volume (RAIV), larger tumour size, higher RENAL score and lower preoperative GFR were short-term risk factors for AD, while increasing WIT and lower preoperative GFR were significant for long-term outcomes. Increasing WIT, RAIV, lower T2* value in the cortex and higher T2* value in the medulla on the ipsilateral side were short-term risk factors for PD, while all of the above factors (except WIT and RAIV) were significant for long-term outcomes. The performance of the combination of T2* values in the cortex and medulla on the ipsilateral side to tumour in predicting OP was better than RENAL score (AUC 0.762 vs 0.634, p = 0.013).ConclusionsRenal BOLD-MRI metrics could provide useful information to the clinician in predicting post-operative renal function outcomes.Key Points center dot Renal fMRI metrics may be useful for prediction of renal functional outcomes and merit further study.center dot Renal fMRI metrics may reflect degree of baseline disease and ability to tolerate warm ischaemia.center dot Combination of T2* values was better than RENAL score for predicting OP.
C1 [Wu, Guangyu; Mao, Haiming; Liu, Guiqin] Shanghai Jiao Tong Univ, Sch Med, Renji Hosp, Dept Radiol, 1630 Dongfang Rd, Shanghai 200120, Peoples R China.
   [Zhang, Ruiyun; Chen, Yonghui; Zhang, Jin] Shanghai Jiao Tong Univ, Renji Hosp, Sch Med, Dept Urinary Surg, 1630 Dongfang Rd, Shanghai 200120, Peoples R China.
C3 Shanghai Jiao Tong University; Shanghai Jiao Tong University
RP Liu, GQ (通讯作者)，Shanghai Jiao Tong Univ, Sch Med, Renji Hosp, Dept Radiol, 1630 Dongfang Rd, Shanghai 200120, Peoples R China.; Zhang, J (通讯作者)，Shanghai Jiao Tong Univ, Renji Hosp, Sch Med, Dept Urinary Surg, 1630 Dongfang Rd, Shanghai 200120, Peoples R China.
EM
FU National Natural Science Foundation of China [81601487/81371622]
FX This study has received funding from the National Natural Science
   Foundation of China (81601487/81371622).
CR Busing KA, 2008, RADIOLOGY, V246, P553, DOI 10.1148/radiol.2462062166
   Chawla LS, 2012, KIDNEY INT, V82, P516, DOI 10.1038/ki.2012.208
   De Santo NG, 1999, NEPHRON, V81, P136, DOI 10.1159/000045268
   dos Santos EA, 2007, INVEST RADIOL, V42, P157, DOI 10.1097/01.rli.0000252492.96709.36
   Evans RG, 2008, CLIN EXP PHARMACOL P, V35, P1405, DOI 10.1111/j.1440-1681.2008.05063.x
   Evans RG, 2013, CLIN EXP PHARMACOL P, V40, P106, DOI 10.1111/1440-1681.12031
   Evans RG, 2011, AM J PHYSIOL-REG I, V300, pR931, DOI 10.1152/ajpregu.00552.2010
   Ficarra V, 2009, EUR UROL, V56, P786, DOI 10.1016/j.eururo.2009.07.040
   Friederich-Persson M, 2013, HYPERTENSION, V62, P914, DOI 10.1161/HYPERTENSIONAHA.113.01425
   Gill IS, 2011, EUR UROL, V59, P128, DOI 10.1016/j.eururo.2010.10.002
   Ginzburg S, 2015, UROLOGY, V86, P300, DOI 10.1016/j.urology.2015.04.043
   Hansell P, 2013, CLIN EXP PHARMACOL P, V40, P123, DOI 10.1111/1440-1681.12034
   Huang YQ, 2015, NMR BIOMED, V28, P154, DOI 10.1002/nbm.3235
   Hung AJ, 2013, J UROLOGY, V189, P36, DOI 10.1016/j.juro.2012.09.042
   Klatte T, 2015, EUR UROL, V68, P980, DOI 10.1016/j.eururo.2015.04.010
   Kopp RP, 2015, UROLOGY, V86, P312, DOI 10.1016/j.urology.2015.02.067
   Kutikov A, 2009, J UROLOGY, V182, P844, DOI 10.1016/j.juro.2009.05.035
   Kwon T, 2015, ANN SURG ONCOL, V22, pS1594, DOI 10.1245/s10434-015-4500-9
   Lee VS, 2003, RADIOLOGY, V227, P289, DOI 10.1148/radiol.2271020383
   Legrand M, 2008, MOL MED, V14, P502, DOI 10.2119/2008-00006.Legrand
   Ljungberg B, 2007, EUR UROL, V51, P1502, DOI 10.1016/j.eururo.2007.03.035
   Ljungberg B, 2015, EUR UROL, V67, P913, DOI 10.1016/j.eururo.2015.01.005
   Prasad PV, 2006, NEPHRON CLIN PRACT, V103, pC58, DOI 10.1159/000090610
   Ries M, 2003, J MAGN RESON IMAGING, V17, P104, DOI 10.1002/jmri.10224
   Seeliger E, 2012, EUR HEART J, V33, P2007, DOI 10.1093/eurheartj/ehr494
   Shin TY, 2015, J UROLOGY, V193, P423, DOI 10.1016/j.juro.2014.07.084
   SHROUT PE, 1979, PSYCHOL BULL, V86, P420, DOI 10.1037/0033-2909.86.2.420
   Simhan J, 2011, EUR UROL, V60, P724, DOI 10.1016/j.eururo.2011.05.030
   Sourbron SP, 2008, INVEST RADIOL, V43, P40, DOI 10.1097/RLI.0b013e31815597c5
   Spaliviero M, 2016, EUR UROL, V69, P72, DOI 10.1016/j.eururo.2015.08.008
   Volpe A, 2015, EUR UROL, V68, P61, DOI 10.1016/j.eururo.2015.01.025
   Wu GY, 2017, EUR RADIOL, V27, P3574, DOI 10.1007/s00330-016-4700-0
   Xu XQ, 2010, EUR RADIOL, V20, P978, DOI 10.1007/s00330-009-1619-8
   Zargar H, 2015, BJU INT, V116, P407, DOI 10.1111/bju.12933
   Zhang JL, 2014, AM J PHYSIOL-RENAL, V306, pF579, DOI 10.1152/ajprenal.00575.2013
   Zhu LD, 2016, MEDICINE, V95, DOI 10.1097/MD.0000000000003043
NR 36
TC 1
Z9 2
U1 0
U2 6
PU SPRINGER
PI NEW YORK
PA 233 SPRING ST, NEW YORK, NY 10013 USA
SN 0938-7994
EI 1432-1084
J9 EUR RADIOL
JI Eur. Radiol.
PD DEC
PY 2018
VL 28
IS 12
BP 5035
EP 5043
DI 10.1007/s00330-018-5525-9
PG 9
WC Radiology, Nuclear Medicine & Medical Imaging
WE Science Citation Index Expanded (SCI-EXPANDED)
SC Radiology, Nuclear Medicine & Medical Imaging
GA HB8PT
UT WOS:000451353500014
PM 29948077
DA 2023-08-27
ER

PT J
AU Sakura, M
   Masuda, H
   Saito, K
   Koga, F
   Kawakami, S
   Kihara, K
AF Sakura, Mizuaki
   Masuda, Hitoshi
   Saito, Kazutaka
   Koga, Fumitaka
   Kawakami, Satoru
   Kihara, Kazunori
TI Collecting duct carcinoma with acquired cystic disease of the kidney in
   a long-term hemodialysis patient
SO INTERNATIONAL JOURNAL OF UROLOGY
LA English
DT Article
DE acquired kidney cystic disease; collecting duct carcinoma; dialysis
   patient
ID RENAL-CELL CARCINOMA
AB We report a very rare case of collecting or Bellini duct carcinoma (CDC) found in a 60-year-old male who had received hemodialysis therapy for 21 years. Screening with ultrasonography revealed a solid tumor originating from the cyst wall in the right kidney with acquired cystic disease of the kidney. Subsequent computed tomography (CT) and angiography could not detect another renal tumor. Right radical nephrectomy was performed. The tumor detected preoperatively was composed of papillary renal cell tumor (RCC) and multiple clear cell carcinoma, pathologically. In addition to the tumors, CDC was revealed in the central medulla with the involvement of regional lymph nodes. Three months later, left nephrectomy was performed because left RCC was suspected during CT. The histological diagnosis was multiple clear cell carcinomas. Peritonitis carcinomatosa appeared and the patient died 13 months later.
C1 [Sakura, Mizuaki; Masuda, Hitoshi; Saito, Kazutaka; Koga, Fumitaka; Kawakami, Satoru; Kihara, Kazunori] Tokyo Med & Dent Univ, Grad Sch, Dept Urol & Reprod Med, Tokyo 1138519, Japan.
C3 Tokyo Medical & Dental University (TMDU)
RP Sakura, M (通讯作者)，Tokyo Med & Dent Univ, Grad Sch, Dept Urol & Reprod Med, 1-5-45 Yushima, Tokyo 1138519, Japan.
EM
RI KOGA, Fumitaka/AAM-8739-2021; Koga, Fumitaka/AAO-2779-2021; ,
   Fumitaka/AAO-2957-2021; Koga, Fumitaka/AAO-4655-2021
OI KOGA, Fumitaka/0000-0001-7888-0085; , Fumitaka/0000-0001-7888-0085;
   Saito, Kazutaka/0000-0002-7261-0188
CR Aita K, 2003, PATHOL INT, V53, P463, DOI 10.1046/j.1440-1827.2003.01502.x
   Cheuk W, 2002, HUM PATHOL, V33, P761, DOI 10.1053/hupa.2002.125370
   EBLE JN, 2004, PATHOLOGY GENETICS T, P33
   Ishikawa I, 2005, J JPN SOC DIAL THER, V38, P1689
   Ito Fumio, 1993, Acta Urologica Japonica, V39, P735
   Karakiewicz PI, 2007, EUR UROL, V52, P1140, DOI 10.1016/j.eururo.2007.01.070
   Kihara K, 2004, INT J UROL, V11, P714, DOI 10.1111/j.1442-2042.2004.00895.x
   Tickoo SK, 2006, AM J SURG PATHOL, V30, P141, DOI 10.1097/01.pas.0000185382.80844.b1
   Tokuda N, 2006, J UROLOGY, V176, P40, DOI 10.1016/S0022-5347(06)00502-7
NR 9
TC 4
Z9 4
U1 0
U2 0
PU WILEY
PI HOBOKEN
PA 111 RIVER ST, HOBOKEN 07030-5774, NJ USA
SN 0919-8172
EI 1442-2042
J9 INT J UROL
JI Int. J. Urol.
PD JAN
PY 2008
VL 15
IS 1
BP 93
EP 95
DI 10.1111/j.1442-2042.2007.01924.x
PG 3
WC Urology & Nephrology
WE Science Citation Index Expanded (SCI-EXPANDED)
SC Urology & Nephrology
GA 247UV
UT WOS:000252109000018
PM 18184183
DA 2023-08-27
ER

PT J
AU Guo, LM
   Qiu, YM
   Ge, JW
   Zhou, DX
AF Guo, Liemei
   Qiu, Yongming
   Ge, Jianwei
   Zhou, Dongxue
TI Glioblastoma Multiforme with Subcutaneous Metastases, Case Report and
   Literature Review
SO JOURNAL OF KOREAN NEUROSURGICAL SOCIETY
LA English
DT Review
DE Glioblastoma multiforme; Glioblastoma metastasis; Subcutaneous
   metastasis; Extracranial metastasis
ID CERVICAL LYMPH-NODE; CUTANEOUS METASTASIS; DIAGNOSIS
AB Glioblastoma multiforme (GBM) is the most common primary brain tumor and the most malignant astrocytoma in adults, with rare extra-cranial metastases, especially for subcutaneous metastases. It could be easily misdiagnosed as primary subcutaneous tumor. In this report, we describe a patient with pontine GBM who developed a subcutaneous swelling at the ipsilateral posterior cervical region 8 months after operation, and the pathological and immunocytochemical examination carry the same characteristics as the primary intracranial GBM cells, which defined it as subcutaneous metastasis. GBM with subcutaneous metastasis is extremely rare, and knowledge of a prior intracranial GBM, pathological examinations and immunocytochemical tests with markers typically expressed by GBM are of vital importance for the diagnosis of GBM metastasis. Surgical resection of subcutaneous swelling, followed by chemotherapy and radiotherapy, could be the best strategy of treatment for the patients with GBM subcutaneous metastasis.
C1 [Guo, Liemei; Qiu, Yongming; Ge, Jianwei] Shanghai Jiao Tong Univ, Sch Med, Renji Hosp, Dept Neurosurg, Shanghai 200127, Peoples R China.
   [Zhou, Dongxue] Shanghai Jiao Tong Univ, Sch Med, Renji Hosp, Gamma Knife Treatment Ctr, Shanghai 200127, Peoples R China.
C3 Shanghai Jiao Tong University; Shanghai Jiao Tong University
RP Qiu, YM (通讯作者)，Shanghai Jiao Tong Univ, Sch Med, Renji Hosp, Dept Neurosurg, 1630 Dongfang Rd, Shanghai 200127, Peoples R China.
EM
CR Allan RS, 2004, J NEUROL NEUROSUR PS, V75, P559
   Ates LE, 2003, NEUROPATHOLOGY, V23, P146, DOI 10.1046/j.1440-1789.2003.00490.x
   Bouillot-Eimer S, 2005, CLIN NEUROPATHOL, V24, P247
   CAMPORA RG, 1993, ACTA CYTOL, V37, P938
   Figueroa P, 2002, J AM ACAD DERMATOL, V46, P297, DOI 10.1067/mjd.2002.104966
   Jain N, 2005, BRIT J NEUROSURG, V19, P65, DOI 10.1080/02688690500081423
   Li Z, 2010, J NEURO-ONCOL, V97, P451, DOI 10.1007/s11060-009-0044-9
   Mentrikoski M, 2008, AM J DERMATOPATH, V30, P381, DOI 10.1097/DAD.0b013e31817532c4
   Miliaras G, 2009, CENT EUR NEUROSURG, V70, P39, DOI 10.1055/s-2008-1080941
   Moon KS, 2004, J KOREAN MED SCI, V19, P911, DOI 10.3346/jkms.2004.19.6.911
   Saad AG, 2007, J PEDIAT HEMATOL ONC, V29, P190, DOI 10.1097/MPH.0b013e31803350a7
   Santos Angelina V, 2003, Acta Med Port, V16, P209
   STEINBOK P, 1985, J NEUROSURG, V62, P918, DOI 10.3171/jns.1985.62.6.0918
   Stupp R, 2005, NEW ENGL J MED, V352, P987, DOI 10.1056/NEJMoa043330
NR 14
TC 11
Z9 11
U1 0
U2 6
PU KOREAN NEUROSURGICAL SOC
PI SEOUL
PA BLDG B, #402, 27, CHILPAE-RO, JUNG-GU, SEOUL, 04511, SOUTH KOREA
SN 2005-3711
EI 1598-7876
J9 J KOREAN NEUROSURG S
JI J. Korean. Neurosurg. Soc.
PD NOV
PY 2012
VL 52
IS 5
BP 484
EP 487
DI 10.3340/jkns.2012.52.5.484
PG 4
WC Clinical Neurology; Surgery
WE Science Citation Index Expanded (SCI-EXPANDED)
SC Neurosciences & Neurology; Surgery
GA 065FU
UT WOS:000313134700010
PM 23323171
OA Green Published, Green Submitted, gold
DA 2023-08-27
ER

PT J
AU SOUSSI, T
   OTTO, SR
AF SOUSSI, T
   OTTO, SR
TI EFFECTS OF ELECTRICAL BRAIN-STEM STIMULATION ON TINNITUS
SO ACTA OTO-LARYNGOLOGICA
LA English
DT Article
DE TINNITUS; ELECTRICAL STIMULATION; BILATERAL ACOUSTIC NEUROMA;
   TRANSLABYRINTHINE APPROACH; AUDITORY BRAIN-STEM IMPLANT
AB The primary aim of the auditory brainstem implant (ABI), an investigational device, is to restore auditory sensations in neurofibromatosis-2 patients deafened by bilateral acoustic neuroma surgery. An electrode is placed on the dorsal cochlear nucleus after the tumor is removed through the translabyrinthine approach. This procedure was performed on 18 patients: 13 were completely deafferentated after bilateral tumor removal and had received the ABI during surgery on the second-sided tumor; 5 were implanted at first-sided tumor removal. Three of these 5 patients had usable hearing remaining on the side of the second acoustic neuroma and were stimulated only during laboratory sessions. All patients had tinnitus. We used a questionnaire to assess the effects of brainstem stimulation on this symptom. Of the 18 patients, one early patient deceased, and one patient with no follow-up was not included in this study. Six patients who were unable to use the ABI because of temporary hardware problems or side effects from electrical stimulation were also excluded. Thus, we studied 10 patients. Of 7 patients who used their implant daily, 6 reported noticeable tinnitus reduction; the ABI had no effect in the remaining case. Of 3 recently implanted patients who used this implant only during laboratory testing, one patient reported complete suppression of tinnitus, one described worse tinnitus, and one reported no effect.
C1 UNIV SO CALIF,SCH MED,HOUSE EAR INST,LOS ANGELES,CA 90033.
C3 House Research Institute; University of Southern California
OI soussi, thierry/0000-0001-8184-3293
CR [Anonymous], 1855, ELECTRISATION LOCALI
   ARAN JM, 1981, CIBA F S, V85
   BERLINER KI, 1992, AM J OTOL, V13, P13
   CHOUARD CH, 1981, ACTA OTO-LARYNGOL, V91, P415, DOI 10.3109/00016488109138522
   Eisenberg L S, 1987, J Rehabil Res Dev, V24, P9, DOI 10.1682/JRRD.1987.07.0009
   Grapengiesser CJC, 1801, VERSUCHE GALVANISMUS
   HOUSE JW, 1981, TINNITUS CIBA F S, V85
   POLITZER A, 1883, DISEASES EAR
   Riccardi VM, 1986, NEUROFIBROMATOSIS PH
   SHANNON RV, 1990, HEARING RES, V47, P159, DOI 10.1016/0378-5955(90)90173-M
   THEDINGER B, 1985, ANN OTO RHINOL LARYN, V94, P10, DOI 10.1177/000348948509400102
   YUEN TGH, 1981, NEUROSURGERY, V9, P292, DOI 10.1227/00006123-198109000-00013
NR 12
TC 57
Z9 59
U1 0
U2 2
PU SCANDINAVIAN UNIVERSITY PRESS
PI OSLO
PA PO BOX 2959 TOYEN, JOURNAL DIVISION CUSTOMER SERVICE, N-0608 OSLO,
   NORWAY
SN 0001-6489
J9 ACTA OTO-LARYNGOL
JI Acta Oto-Laryngol.
PD MAR
PY 1994
VL 114
IS 2
BP 135
EP 140
DI 10.3109/00016489409126031
PG 6
WC Otorhinolaryngology
WE Science Citation Index Expanded (SCI-EXPANDED)
SC Otorhinolaryngology
GA ND135
UT WOS:A1994ND13500004
PM 8203193
DA 2023-08-27
ER

PT J
AU Ishikawa, M
   Kusaka, G
   Takashima, K
   Kamochi, H
   Shinoda, S
AF Ishikawa, Mami
   Kusaka, Gen
   Takashima, Kouichi
   Kamochi, Haruna
   Shinoda, Soji
TI Intraoperative monitoring during surgery for hypoglossal schwannoma
SO JOURNAL OF CLINICAL NEUROSCIENCE
LA English
DT Article
DE Electromyography; Evoked potential; Hypoglossal nerve; Operative
   monitoring; Schwannoma
ID SKULL BASE SURGERY; TRANSCONDYLAR APPROACH; NERVE; NEURINOMA; TUBERCLE
AB A 54-year-old man presented with an intracranial schwannoma of the hypoglossal nerve between the medulla and the left hypoglossal canal. The condylar fossa approach was used with intra-operative electromyography (EMG) monitoring of the lower cranial nerves. The tumor was then removed carefully without decreasing the tongue EMG responses. EMG monitoring enabled us to remove the tumor while maintaining the function of the hypoglossal nerve. Tongue EMG was easily recorded by stimulating the hypoglossal nerve fibers, which was useful in identifying the hypoglossal nerve and evaluating its function. This suggests that tongue EMG is a useful monitoring tool to enhance neurological outcome following removal of tumors in this region. (C) 2009 Elsevier Ltd. All rights reserved.
C1 [Ishikawa, Mami; Kusaka, Gen; Takashima, Kouichi; Kamochi, Haruna; Shinoda, Soji] Jichi Med Univ, Saitama Med Ctr, Dept Neurosurg, Omiya Ku, Saitama 3308503, Japan.
C3 Jichi Medical University
RP Ishikawa, M (通讯作者)，Jichi Med Univ, Saitama Med Ctr, Dept Neurosurg, Omiya Ku, 847 Amanumacho, Saitama 3308503, Japan.
EM
CR BERTALANFFY H, 1991, NEUROSURGERY, V29, P815, DOI 10.1227/00006123-199112000-00002
   Gibo H, 1997, SURG NEUROL, V48, P85, DOI 10.1016/S0090-3019(96)00475-2
   Gilsbach JM, 1998, SURG NEUROL, V50, P563, DOI 10.1016/S0090-3019(97)00378-9
   Kikkawa Y, 2007, MINIM INVAS NEUROSUR, V50, P296, DOI 10.1055/s-2007-984385
   Matsushima T, 2001, J NEUROSURG, V95, P268, DOI 10.3171/jns.2001.95.2.0268
   Mikuni N, 2004, J NEUROSURG, V101, P536, DOI 10.3171/jns.2004.101.3.0536
   Piccirilli M, 2007, ACTA NEUROCHIR, V149, P949, DOI 10.1007/s00701-007-1154-9
   Schlake HP, 2001, CLIN NEUROL NEUROSUR, V103, P72, DOI 10.1016/S0303-8467(01)00115-9
   Topsakal C, 2008, NEUROSURG REV, V31, P45, DOI 10.1007/s10143-007-0105-5
   WEI HT, 1997, J NEUROSURG, V87, P555
NR 10
TC 13
Z9 13
U1 0
U2 2
PU ELSEVIER SCI LTD
PI OXFORD
PA THE BOULEVARD, LANGFORD LANE, KIDLINGTON, OXFORD OX5 1GB, OXON, ENGLAND
SN 0967-5868
J9 J CLIN NEUROSCI
JI J. Clin. Neurosci.
PD AUG
PY 2010
VL 17
IS 8
BP 1053
EP 1056
DI 10.1016/j.jocn.2009.09.040
PG 4
WC Clinical Neurology; Neurosciences
WE Science Citation Index Expanded (SCI-EXPANDED)
SC Neurosciences & Neurology
GA 623GK
UT WOS:000279726700024
PM 20488709
DA 2023-08-27
ER

PT J
AU Hermann, GE
   Rogers, RC
   Bresnahan, JC
   Beattie, MS
AF Hermann, GE
   Rogers, RC
   Bresnahan, JC
   Beattie, MS
TI Tumor necrosis factor-alpha induces cFOS and strongly potentiates
   glutamate-mediated cell death in the rat spinal cord
SO NEUROBIOLOGY OF DISEASE
LA English
DT Article
DE spinal cord injury; cytokines; glutamate; cFOS; excitotoxic cell death;
   microglia
ID TNF-ALPHA; MICROGLIAL ACTIVATION; FUNCTIONAL DEFICITS; INJURY; RECEPTOR;
   BRAIN; EXPRESSION; NEUROTOXICITY; CYTOKINES; MEDULLA
AB Excitotoxic cell death due to glutamate release is important in the secondary injury following CNS trauma or ischemia. Proinflammatory cytokines also play a role. Both glutamate and tumor necrosis factor-alpha (TNFalpha) are released immediately after spinal cord injury. Neurophysiological studies show that TNFalpha can potentiate the effects of glutamatergic afferent input to produce hyperactivation of brain-stem sensory neurons. Therefore, we hypothesized that TNFalpha might act cooperatively with glutamate to affect cell death in the spinal cord as well. Nanoinjections of either TNFalpha (60 pg) or kainate (KA; 32 ng) alone into the thoracic gray resulted in almost no tissue damage or cell death 90 min after injection. However, the combination of TNFalpha plus KA at these same doses produced a large area of tissue necrosis and neuronal cell death, an effect which was blocked by the AMPA receptor antagonist CNQX (17 ng). These results suggest that secondary injury may involve potentiation of AMPA receptor-mediated excitatory cell death by TNFalpha. (C) 2001 Academic Press.
C1 Ohio State Univ, Coll Med & Publ Hlth, Dept Neurosci, Labs Auton Neurosci & Neural Repair, Columbus, OH 43210 USA.
C3 University System of Ohio; Ohio State University
RP Hermann, GE (通讯作者)，Ohio State Univ, Coll Med & Publ Hlth, Dept Neurosci, Labs Auton Neurosci & Neural Repair, 333 W 10th Ave, Columbus, OH 43210 USA.
EM
RI Hermann, Gerlinda E/N-1967-2017; Rogers, Richard/N-1988-2017; Beattie,
   Michael S/A-8953-2009
OI Hermann, Gerlinda E/0000-0003-0405-2820; 
FU NIDDK NIH HHS [DK52142] Funding Source: Medline; NINDS NIH HHS [NS38079]
   Funding Source: Medline
CR Barger SW, 1997, NATURE, V388, P878, DOI 10.1038/42257
   Beattie MS, 2000, CONT NEUROS, P1
   Beattie MS, 1997, EXP NEUROL, V148, P453, DOI 10.1006/exnr.1997.6695
   Bethea JR, 1999, J NEUROTRAUM, V16, P851, DOI 10.1089/neu.1999.16.851
   Bethea JR, 1998, J NEUROSCI, V18, P3251
   BISHOP GA, 1990, NEUROSCIENCE, V39, P251, DOI 10.1016/0306-4522(90)90238-Y
   Brewer KL, 1999, EXP NEUROL, V159, P484, DOI 10.1006/exnr.1999.7173
   BUTCHER LL, 1978, EUR J PHARMACOL, V50, P287, DOI 10.1016/0014-2999(78)90365-5
   CHAO CC, 1994, DEV NEUROSCI-BASEL, V16, P172, DOI 10.1159/000112104
   CHOI DW, 1994, PROG BRAIN RES, V100, P47
   Crowe MJ, 1997, NAT MED, V3, P73, DOI 10.1038/nm0197-73
   Emch GS, 2000, AM J PHYSIOL-GASTR L, V279, pG582, DOI 10.1152/ajpgi.2000.279.3.G582
   FADEN AI, 1988, ANN NEUROL, V23, P623, DOI 10.1002/ana.410230618
   GELBARD HA, 1993, DEV NEUROSCI-BASEL, V15, P417, DOI 10.1159/000111367
   Hall SM, 2000, J NEUROIMMUNOL, V106, P130, DOI 10.1016/S0165-5728(00)00213-7
   Hayashi M, 2000, J NEUROTRAUM, V17, P203, DOI 10.1089/neu.2000.17.203
   HERMANN G, 1995, NEUROIMMUNOMODULAT, V2, P74, DOI 10.1159/000096874
   Hsieh JH, 1998, J AUTONOM NERV SYST, V73, P7, DOI 10.1016/S0165-1838(98)00117-9
   Kimelberg HK, 2000, NEUROSCIENTIST, V6, P14, DOI 10.1177/107385840000600110
   KWAK S, 1995, BRAIN RES, V702, P61, DOI 10.1016/0006-8993(95)01000-6
   Lee YL, 2000, NEUROCHEM INT, V36, P417, DOI 10.1016/S0197-0186(99)00133-3
   Liu D, 1999, NEUROSCIENCE, V93, P1383, DOI 10.1016/S0306-4522(99)00278-X
   Liu SL, 1997, BRAIN RES, V756, P160, DOI 10.1016/S0006-8993(97)00137-6
   McDonald JW, 1998, NEUROREPORT, V9, P2757, DOI 10.1097/00001756-199808240-00014
   McDonald JW, 1998, NAT MED, V4, P291, DOI 10.1038/nm0398-291
   MOHAMMED JR, 1995, BRAIN RES, V695, P100, DOI 10.1016/0006-8993(95)00505-K
   Nadeau S, 2000, J NEUROSCI, V20, P3456, DOI 10.1523/JNEUROSCI.20-09-03456.2000
   Nakamura Y, 1999, NEUROSCI RES, V35, P95, DOI 10.1016/S0168-0102(99)00071-1
   Noda M, 2000, J NEUROSCI, V20, P251, DOI 10.1523/JNEUROSCI.20-01-00251.2000
   Pan WH, 1999, J NEUROSCI, V19, P3649
   PANTER SS, 1990, ANN NEUROL, V27, P96, DOI 10.1002/ana.410270115
   PIANI D, 1991, NEUROSCI LETT, V133, P159, DOI 10.1016/0304-3940(91)90559-C
   Raivich G, 1999, BRAIN RES REV, V30, P77, DOI 10.1016/S0165-0173(99)00007-7
   RINAMAN L, 1993, J COMP NEUROL, V338, P475, DOI 10.1002/cne.903380402
   Rosen CJ, 1999, J CLIN DENSITOM, V2, P3, DOI 10.1385/JCD:2:1:3
   Rothwell NJ, 1999, J PHYSIOL-LONDON, V514, P3, DOI 10.1111/j.1469-7793.1999.003af.x
   Schnell L, 1999, J NEUROPATH EXP NEUR, V58, P245, DOI 10.1097/00005072-199903000-00004
   Shuman SL, 1997, J NEUROSCI RES, V50, P798, DOI 10.1002/(SICI)1097-4547(19971201)50:5<798::AID-JNR16>3.3.CO;2-#
   Sivarao DV, 1999, EUR J PHARMACOL, V368, P173, DOI 10.1016/S0014-2999(99)00015-1
   Wang CX, 1996, J NEUROIMMUNOL, V69, P151
   Wrathall JR, 1996, EXP NEUROL, V137, P119, DOI 10.1006/exnr.1996.0012
   Wrathall JR, 1997, EXP NEUROL, V145, P565, DOI 10.1006/exnr.1997.6506
   WRATHALL JR, 1994, J NEUROSCI, V14, P6598
   Xu J, 1998, MOL BRAIN RES, V59, P135, DOI 10.1016/S0169-328X(98)00142-9
   YAKOVLEV AG, 1994, MOL CHEM NEUROPATHOL, V23, P179, DOI 10.1007/BF02815410
   Yang H, 2000, NEUROSCIENCE, V95, P773
   Yezierski RP, 1998, PAIN, V75, P141, DOI 10.1016/S0304-3959(97)00216-9
NR 47
TC 159
Z9 170
U1 1
U2 4
PU ACADEMIC PRESS INC ELSEVIER SCIENCE
PI SAN DIEGO
PA 525 B ST, STE 1900, SAN DIEGO, CA 92101-4495 USA
SN 0969-9961
EI 1095-953X
J9 NEUROBIOL DIS
JI Neurobiol. Dis.
PD AUG
PY 2001
VL 8
IS 4
BP 590
EP 599
DI 10.1006/nbdi.2001.0414
PG 10
WC Neurosciences
WE Science Citation Index Expanded (SCI-EXPANDED)
SC Neurosciences & Neurology
GA 463EF
UT WOS:000170463300005
PM 11493024
DA 2023-08-27
ER

PT J
AU Eom, KS
   Kim, DW
   Choi, SS
   Choi, KH
   Kim, TY
AF Eom, Ki Seong
   Kim, Doe Won
   Choi, See Sung
   Choi, Keum Ho
   Kim, Tae Young
TI Preoperative embolization of a cerebellar haemangioblastoma using Onyx:
   case report and literature review
SO NEUROLOGIA I NEUROCHIRURGIA POLSKA
LA English
DT Review
DE haemangioblastoma; embolization; Onyx
ID BRAIN
AB Haemangioblastoma is a slow-growing, highly vascular tumour and typically occurs in the cerebellum but can also occur in the brainstem and spinal cord. Because of their hypervascularity and location, cerebellar haemangioblastomas can be difficult to remove. The purpose of preoperative embolization of haemangioblastomas is to decrease the intraoperative blood loss and to facilitate excision. However, the safety and efficacy of this procedure remain controversial. Here, we report the case of a man with cerebellar haemangioblastoma who underwent preoperative embolization with Onyx. The tumour was completely removed with minimal tumour bleeding. There was no complication related to embolization.
C1 [Eom, Ki Seong; Kim, Doe Won; Choi, See Sung; Choi, Keum Ho; Kim, Tae Young] Wonkwang Univ, Coll Med, Iksan 570711, South Korea.
C3 Wonkwang University
RP Kim, TY (通讯作者)，Wonkwang Univ, Coll Med, 344-2 Shinyong Dong, Iksan 570711, South Korea.
EM
FU Wonkwang University
FX This paper was supported by Wonkwang University in 2011.
CR Cornelius JF, 2007, J NEUROSURG, V106, P994, DOI 10.3171/jns.2007.106.6.994
   Elhammady MS, 2010, J NEUROSURG, V112, P1039, DOI 10.3171/2009.7.JNS09351
   Gore P, 2008, NEUROSURGERY, V62, P1204, DOI 10.1227/01.neu.0000333292.74986.ac
   Montano N, 2008, J NEUROSURG, V108, P1063, DOI 10.3171/JNS/2008/108/5/1063
   REHMAN T, BMJ CASE REPORTS
   STANDARD SC, 1994, SURG NEUROL, V41, P405, DOI 10.1016/0090-3019(94)90035-3
   TAMPIERI D, 1993, NEUROSURGERY, V33, P502
   VATES GE, 2005, TXB NEUROONCOLOGY, P294
NR 8
TC 7
Z9 7
U1 0
U2 2
PU TERMEDIA PUBLISHING HOUSE LTD
PI POZNAN
PA WENEDOW ST 9-1, POZNAN, 61-614, POLAND
SN 0028-3843
J9 NEUROL NEUROCHIR POL
JI Neurol. Neurochir. Pol.
PD MAY-JUN
PY 2011
VL 45
IS 3
BP 292
EP 296
PG 5
WC Clinical Neurology
WE Science Citation Index Expanded (SCI-EXPANDED)
SC Neurosciences & Neurology
GA 816GP
UT WOS:000294587000010
PM 21866486
DA 2023-08-27
ER

PT J
AU YOUNG, WF
AF YOUNG, WF
TI PHEOCHROMOCYTOMA - 1926-1993
SO TRENDS IN ENDOCRINOLOGY AND METABOLISM
LA English
DT Review
ID MALIGNANT PHEOCHROMOCYTOMA
AB Pheochromocytoma is a tumor frequently sought and rarely found. It is associated with spectacular cardiovascular disturbances and, when correctly diagnosed and properly treated, it is curable, when undiagnosed or improperly treated, it can be fatal. Catecholamine-producing tumors that arise from chromaffin cells of the adrenal medulla and sympathetic ganglia are termed pheochromocytomas and paragangliomas, respectively. However, the term pheochromocytoma has become the generic name for all catecholamine-producing tumors. The biochemical diagnosis is straightforward. The localization of pheochromocytoma has been greatly facilitated by advances in computerized imaging and meta-iodobenzylguanidine scanning. Treatment with preoperative alpha- and beta-adrenergic blockade followed by surgical excision of the pheochromocytoma are associated with very low morbidity and mortality.
C1 MAYO CLIN & MAYO FDN,SCH MED,DIV INTERNAL MED & ENDOCRINOL,ROCHESTER,MN 55905.
   MAYO CLIN & MAYO FDN,SCH MED,DIV METAB,ROCHESTER,MN 55905.
   MAYO CLIN & MAYO FDN,SCH MED,DIV INTERNAL MED,ROCHESTER,MN 55905.
C3 Mayo Clinic; Mayo Clinic; Mayo Clinic
RP YOUNG, WF (通讯作者)，MAYO CLIN & MAYO FDN,SCH MED,DIV HYPERTENS,ROCHESTER,MN 55905, USA.
CR ATUK NO, 1979, MEDICINE, V58, P209, DOI 10.1097/00005792-197905000-00001
   AVERBUCH SD, 1988, ANN INTERN MED, V109, P267, DOI 10.7326/0003-4819-109-4-267
   COPE C, 1991, AUST NZ J SURG, V61, P695, DOI 10.1111/j.1445-2197.1991.tb00323.x
   FRUSTACI A, 1993, CHEST, V99, P383
   GAGNER M, 1992, NEW ENGL J MED, V327, P1033
   GRAHAM JB, 1951, SURG GYNECOL OBSTET, V92, pS105
   GROSSMAN E, 1991, HYPERTENSION, V17, P733, DOI 10.1161/01.HYP.17.6.733
   HARPER MA, 1989, BRIT J OBSTET GYNAEC, V96, P594, DOI 10.1111/j.1471-0528.1989.tb03262.x
   ITO Y, 1992, WORLD J SURG, V16, P759, DOI 10.1007/BF02067379
   JACKSON JA, 1993, ENDOCRINOLOGIST, V3, P5
   KALFF V, 1982, ARCH INTERN MED, V142, P2092, DOI 10.1001/archinte.142.12.2092
   KREMPF M, 1991, J CLIN ENDOCR METAB, V72, P455, DOI 10.1210/jcem-72-2-455
   LAMBERTS SWJ, 1990, NEW ENGL J MED, V323, P1246, DOI 10.1056/NEJM199011013231805
   Manger W, 1977, PHEOCHROMOCYTOMA
   Mayo CH, 1927, J AMER MED ASSOC, V89, P1047, DOI 10.1001/jama.1927.02690130035013
   Nadler JL, 1991, ENDOCRINOLOGIST, V1, P5
   NATIV O, 1992, CANCER, V69, P2683, DOI 10.1002/1097-0142(19920601)69:11<2683::AID-CNCR2820691110>3.0.CO;2-M
   Saegesser F, 1984, Rev Med Suisse Romande, V104, P403
   Shapiro B, 1991, J Nucl Biol Med, V35, P269
   SHAPIRO B, 1990, ENDOCRINE HYPERTENSI, P235
   SHEPS SG, 1988, ENDOCRIN METAB CLIN, V17, P397, DOI 10.1016/S0889-8529(18)30426-2
   SHULKIN BL, 1992, J NUCL MED, V33, P1125
   SJOBERG RJ, 1992, ARCH INTERN MED, V152, P1193, DOI 10.1001/archinte.152.6.1193
   VANGILS APG, 1991, RADIOGRAPHICS, V11, P37, DOI 10.1148/radiographics.11.1.1671719
   VANHEERDEN JA, 1990, WORLD J SURG, V14, P325, DOI 10.1007/BF01658516
   VANHEERDEN JA, 1982, AM J SURG, V144, P277, DOI 10.1016/0002-9610(82)90525-6
   WELBOURN RB, 1987, BRIT J SURG, V74, P594, DOI 10.1002/bjs.1800740717
   WHALEN RK, 1992, J UROLOGY, V147, P1, DOI 10.1016/S0022-5347(17)37119-7
NR 28
TC 31
Z9 32
U1 0
U2 0
PU ELSEVIER SCIENCE INC
PI NEW YORK
PA 655 AVENUE OF THE AMERICAS, NEW YORK, NY 10010
SN 1043-2760
J9 TRENDS ENDOCRIN MET
JI Trends Endocrinol. Metab.
PD MAY-JUN
PY 1993
VL 4
IS 4
BP 122
EP 127
DI 10.1016/1043-2760(93)90035-D
PG 6
WC Endocrinology & Metabolism
WE Science Citation Index Expanded (SCI-EXPANDED)
SC Endocrinology & Metabolism
GA LK548
UT WOS:A1993LK54800002
PM 18407145
DA 2023-08-27
ER

PT J
AU Zhang, L
   Peterson, TE
   Lu, VM
   Parney, IF
   Daniels, DJ
AF Zhang, Liang
   Peterson, Timothy E.
   Lu, Victor M.
   Parney, Ian F.
   Daniels, David J.
TI Antitumor activity of novel pyrazole-based small molecular inhibitors of
   the STAT3 pathway in patient derived high grade glioma cells
SO PLOS ONE
LA English
DT Article
ID SIGNALING PATHWAYS; GROWTH-FACTOR; STEM-CELLS; CANCER; GLIOBLASTOMA;
   ACTIVATION; INTERLEUKIN-6; APOPTOSIS; PHOSPHORYLATION; RECEPTOR
AB Abnormal activation of signal transducer and activator of transcription 3 ( STAT3) transcription factor has been observed in many human cancers with roles in tumor initiation, progression, drug resistance, angiogenesis and immunosuppression. STAT3 is constitutively activated in a variety of cancers including adult high grade gliomas ( aHGGs) such as glioblastoma ( GBM), and pediatric high grade gliomas ( pHGG). Inhibiting STAT3 is a promising target-specific chemotherapeutic strategy for tumors with aberrant STAT3 signaling. Here we investigated the antitumor effects of novel pyrazole-based STAT3 pathway inhibitors named MNS1 ( Mayo Neurosurgery 1) in both pediatric and adult HGG tumor cells. MNS1 compounds selectively decreased cell viability and proliferation in patient-derived HGG cells with minimal toxicity on normal human astrocytes. These inhibitors selectively blocked IL-6-induced STAT3 phosphorylation and nuclear localization of pSTAT3 without affecting other signaling molecules including Akt, STAT1, JAK2 or ERK1/2 phosphorylation. Functional analysis showed that MNS1 compounds induced apoptosis and decrease tumor migration. The anti-tumor effects extended into a murine pHGG ( diffuse intrinsic pontine glioma) patient derived xenograft, and systemic toxicity was not evident during dose escalation in mice. These results support further development of STAT3 inhibitors for both pediatric and adult HGG.
C1 [Zhang, Liang; Peterson, Timothy E.; Lu, Victor M.; Parney, Ian F.; Daniels, David J.] Mayo Clin, Dept Neurosurg, Rochester, MN 55902 USA.
C3 Mayo Clinic
RP Daniels, DJ (通讯作者)，Mayo Clin, Dept Neurosurg, Rochester, MN 55902 USA.
EM
FU NIH [T32NS07494]; NINDS [K08NS092891]
FX DJD received support via NIH Training Grant T32NS07494 and a K08NS092891
   award through NINDS. The funders had no role in study design, data
   collection and analysis, decision to publish, or preparation of the
   manuscript.
CR Ashizawa T, 2013, INT J ONCOL, V43, P219, DOI 10.3892/ijo.2013.1916
   Ball S, 2011, PLOS ONE, V6, DOI 10.1371/journal.pone.0018820
   Baumann H, 1996, P NATL ACAD SCI USA, V93, P8374, DOI 10.1073/pnas.93.16.8374
   Bowman T, 2000, ONCOGENE, V19, P2474, DOI 10.1038/sj.onc.1203527
   Brantley EC, 2008, MOL CANCER RES, V6, P675, DOI 10.1158/1541-7786.MCR-07-2180
   Bromberg J, 2000, ONCOGENE, V19, P2468, DOI 10.1038/sj.onc.1203476
   Center MDAC, 2013, JAK2 INH WP1066 TREA
   Corcoran RB, 2011, CANCER RES, V71, P5020, DOI 10.1158/0008-5472.CAN-11-0908
   Eason DD, 2001, J IMMUNOL, V166, P1041, DOI 10.4049/jimmunol.166.2.1041
   Ferrajoli A, 2007, CANCER RES, V67, P11291, DOI 10.1158/0008-5472.CAN-07-0593
   Garcia R, 1997, CELL GROWTH DIFFER, V8, P1267
   Griesinger AM, 2015, CANCER IMMUNOL RES, V3, P1165, DOI 10.1158/2326-6066.CIR-15-0061
   Haftchenary S, 2013, ACS MED CHEM LETT, V4, P1102, DOI 10.1021/ml4003138
   Hashizume R, 2012, J NEURO-ONCOL, V110, P305, DOI 10.1007/s11060-012-0973-6
   Heinrich PC, 1998, BIOCHEM J, V334, P297, DOI 10.1042/bj3340297
   Hussain SF, 2007, CANCER RES, V67, P9630, DOI 10.1158/0008-5472.CAN-07-1243
   Iwamaru A, 2007, ONCOGENE, V26, P2435, DOI 10.1038/sj.onc.1210031
   Kelly JJP, 2009, STEM CELLS, V27, P1722, DOI 10.1002/stem.98
   Kohsaka S, 2012, MOL CANCER THER, V11, P1289, DOI 10.1158/1535-7163.MCT-11-0801
   Kumar R, 2017, PLOS ONE, V12, DOI 10.1371/journal.pone.0179012
   Lin GS, 2014, MED ONCOL, V31, DOI 10.1007/s12032-014-0924-5
   Lin L, 2010, NEOPLASIA, V12, P39, DOI 10.1593/neo.91196
   Liu QL, 2010, J NEURO-ONCOL, V100, P165, DOI 10.1007/s11060-010-0158-0
   Liu Y, 2010, CELL CYCLE, V9, P3423, DOI 10.4161/cc.9.17.12946
   Marotta LLC, 2011, J CLIN INVEST, V121, P2723, DOI 10.1172/JCI44745
   McFarland BC, 2011, MOL CANCER THER, V10, P2384, DOI 10.1158/1535-7163.MCT-11-0480
   Meel MH, 2018, CELL MOL LIFE SCI, V75, P871, DOI 10.1007/s00018-017-2714-7
   Nagasaki T, 2014, BRIT J CANCER, V110, P469, DOI 10.1038/bjc.2013.748
   Nakajima K, 1996, EMBO J, V15, P3651, DOI 10.1002/j.1460-2075.1996.tb00734.x
   Oh DY, 2015, CANCER RES TREAT, V47, P607, DOI 10.4143/crt.2014.249
   Rahaman SO, 2002, ONCOGENE, V21, P8404, DOI 10.1038/sj.onc.1206047
   Sai K, 2012, J NEURO-ONCOL, V107, P487, DOI 10.1007/s11060-011-0786-z
   Saidi A, 2009, INT J CANCER, V125, P1054, DOI 10.1002/ijc.24380
   Schust J, 2004, ANAL BIOCHEM, V330, P114, DOI 10.1016/j.ab.2004.03.024
   Schust J, 2006, CHEM BIOL, V13, P1235, DOI 10.1016/j.chembiol.2006.09.018
   Siddiquee K, 2007, P NATL ACAD SCI USA, V104, P7391, DOI 10.1073/pnas.0609757104
   Song H, 2005, P NATL ACAD SCI USA, V102, P4700, DOI 10.1073/pnas.0409894102
   Song LX, 2003, ONCOGENE, V22, P4150, DOI 10.1038/sj.onc.1206479
   Stechishin OD, 2013, NEURO-ONCOLOGY, V15, P198, DOI 10.1093/neuonc/nos302
   Stupp R, 2005, NEW ENGL J MED, V352, P987, DOI 10.1056/NEJMoa043330
   Swiatek-Machado K., 2012, CANC BIOL THERAPY, V13
   Wan SS, 2014, GASTROENTEROLOGY, V147, P1393, DOI 10.1053/j.gastro.2014.08.039
   Wang HM, 2004, LAB INVEST, V84, P941, DOI 10.1038/labinvest.3700123
   Wang H, 2009, STEM CELLS, V27, P2393, DOI 10.1002/stem.188
   Weissenberger J, 2004, ONCOGENE, V23, P3308, DOI 10.1038/sj.onc.1207455
   Weissenberger J, 2010, CLIN CANCER RES, V16, P5781, DOI 10.1158/1078-0432.CCR-10-0446
   Welby JP, 2019, FRONT ONCOL, V9, DOI 10.3389/fonc.2019.00092
   Wong ALA, 2017, EXPERT OPIN INV DRUG, V26, P883, DOI 10.1080/13543784.2017.1351941
   YU CL, 1995, SCIENCE, V269, P81, DOI 10.1126/science.7541555
   Yu DD, 2013, BIOORGAN MED CHEM, V21, P4266, DOI 10.1016/j.bmc.2013.04.069
   Yu H, 2004, NAT REV CANCER, V4, P97, DOI 10.1038/nrc1275
   Yu H, 2007, NAT REV IMMUNOL, V7, P41, DOI 10.1038/nri1995
   Zhang XL, 2013, CANCER RES, V73, P1922, DOI 10.1158/0008-5472.CAN-12-3175
   ZHONG Z, 1994, SCIENCE, V264, P95, DOI 10.1126/science.8140422
NR 54
TC 11
Z9 11
U1 1
U2 4
PU PUBLIC LIBRARY SCIENCE
PI SAN FRANCISCO
PA 1160 BATTERY STREET, STE 100, SAN FRANCISCO, CA 94111 USA
SN 1932-6203
J9 PLOS ONE
JI PLoS One
PD JUL 30
PY 2019
VL 14
IS 7
AR e0220569
DI 10.1371/journal.pone.0220569
PG 18
WC Multidisciplinary Sciences
WE Science Citation Index Expanded (SCI-EXPANDED)
SC Science & Technology - Other Topics
GA IW4WU
UT WOS:000484981300043
PM 31361777
OA Green Published, Green Submitted, gold
DA 2023-08-27
ER

PT J
AU Plouin, PF
   Gimenez-Roqueplo, AP
AF Plouin, Pierre-Francois
   Gimenez-Roqueplo, Anne-Paule
TI Pheochromocytomas and secreting paragangliomas
SO ORPHANET JOURNAL OF RARE DISEASES
LA English
DT Review
ID HEREDITARY PARAGANGLIOMA; BIOCHEMICAL-DIAGNOSIS; GENE MUTATION; SDHD
   GENE; LOCALIZATION; RECURRENCE; FEATURES; TUMORS
AB Catecholamine-producing tumors may arise in the adrenal medulla ( pheochromocytomas) or in extraadrenal chromaffin cells ( secreting paragangliomas). Their prevalence is about 0.1% in patients with hypertension and 4% in patients with a fortuitously discovered adrenal mass. An increase in the production of catecholamines causes symptoms ( mainly headaches, palpitations and excess sweating) and signs ( mainly hypertension, weight loss and diabetes) reflecting the effects of epinephrine and norepinephrine on alpha- and beta- adrenergic receptors. Catecholamine- producing tumors mimic paroxysmal conditions with hypertension and/or cardiac rhythm disorders, including panic attacks, in which sympathetic activation linked to anxiety reproduces the same signs and symptoms. These tumors may be sporadic or part of any of several genetic diseases: familial pheochromocytoma-paraganglioma syndromes, multiple endocrine neoplasia type 2, neurofibromatosis 1 and von Hippel-Lindau disease. Familial cases are diagnosed earlier and are more frequently bilateral and recurring than sporadic cases. The most specific and sensitive diagnostic test for the tumor is the determination of plasma or urinary metanephrines. The tumor can be located by computed tomography, magnetic resonance imaging and metaiodobenzylguanidine scintigraphy. Treatment requires resection of the tumor, generally by laparoscopic surgery. About 10% of tumors are malignant either at first operation or during followup, malignancy being diagnosed by the presence of lymph node, visceral or bone metastases. Recurrences and malignancy are more frequent in cases with large or extraadrenal tumors. Patients, especially those with familial or extraadrenal tumors, should be followed-up indefinitely.
C1 [Plouin, Pierre-Francois] Univ Paris 05, F-75270 Paris 06, France.
   [Plouin, Pierre-Francois] Univ Paris 05, Assistance Publ Hop Paris, Hop Europeen Groeges, Dept Genet, F-75270 Paris 06, France.
C3 UDICE-French Research Universities; Universite Paris Cite; Assistance
   Publique Hopitaux Paris (APHP); UDICE-French Research Universities;
   Aix-Marseille Universite; Assistance Publique-Hopitaux de Marseille;
   Universite Paris Cite
EM;
  
RI GIMENEZ-ROQUEPLO, Anne-Paule/T-6752-2018
OI GIMENEZ-ROQUEPLO, Anne-Paule/0000-0002-4816-670X
CR Amar L, 2005, J CLIN ONCOL, V23, P8812, DOI 10.1200/JCO.2005.03.1484
   Amar L, 2005, J CLIN ENDOCR METAB, V90, P2110, DOI 10.1210/jc.2004-1398
   Baysal BE, 2003, TRENDS ENDOCRIN MET, V14, P453, DOI 10.1016/j.tem.2003.08.004
   Benn DE, 2006, J CLIN ENDOCR METAB, V91, P827, DOI 10.1210/jc.2005-1862
   Bravo EL, 2003, ENDOCR REV, V24, P539, DOI 10.1210/er.2002-0013
   Eisenhofer G, 2004, ENDOCR-RELAT CANCER, V11, P423, DOI 10.1677/erc.1.00829
   Eisenhofer G, 2003, J CLIN ENDOCR METAB, V88, P2656, DOI 10.1210/jc.2002-030005
   Favier J, 2002, AM J PATHOL, V161, P1235, DOI 10.1016/S0002-9440(10)64400-8
   Gimenez-Roqueplo AP, 2003, CANCER RES, V63, P5615
   Gimenez-Roqueplo AP, 2002, J CLIN ENDOCR METAB, V87, P4771, DOI 10.1210/jc.2002-020525
   Gimenez-Roqueplo AP, 2001, AM J HUM GENET, V69, P1186, DOI 10.1086/324413
   Heron E, 1996, ANN INTERN MED, V125, P300, DOI 10.7326/0003-4819-125-4-199608150-00008
   HSIAO RJ, 1991, MEDICINE, V70, P33, DOI 10.1097/00005792-199101000-00003
   Ichihara M, 2004, CANCER LETT, V204, P197, DOI 10.1016/S0304-3835(03)00456-7
   Ilias I, 2004, J CLIN ENDOCR METAB, V89, P479, DOI 10.1210/jc.2003-031091
   Kim WY, 2004, J CLIN ONCOL, V22, P4991, DOI 10.1200/JCO.2004.05.061
   Kudva YC, 2003, J CLIN ENDOCR METAB, V88, P4533, DOI 10.1210/jc.2003-030720
   Lenders JWM, 2002, JAMA-J AM MED ASSOC, V287, P1427, DOI 10.1001/jama.287.11.1427
   Mantero F, 2000, J CLIN ENDOCR METAB, V85, P637, DOI 10.1210/jc.85.2.637
   Neumann HPH, 2002, NEW ENGL J MED, V346, P1459, DOI 10.1056/NEJMoa020152
   Neumann HPH, 2004, JAMA-J AM MED ASSOC, V292, P943, DOI 10.1001/jama.292.8.943
   Pacak K, 2001, ANN INTERN MED, V134, P315, DOI 10.7326/0003-4819-134-4-200102200-00016
   Plouin PF, 2006, NAT CLIN PRACT ENDOC, V2, P60, DOI 10.1038/ncpendmet0097
   Plouin PF, 1997, HYPERTENSION, V29, P1133, DOI 10.1161/01.HYP.29.5.1133
   Plouin PF, 2001, J CLIN ENDOCR METAB, V86, P1480, DOI 10.1210/jc.86.4.1480
   Selak MA, 2005, CANCER CELL, V7, P77, DOI 10.1016/j.ccr.2004.11.022
   SENANAYAKE PD, 1995, J CLIN INVEST, V96, P2503, DOI 10.1172/JCI118310
   van der Harst E, 2002, EUR J ENDOCRINOL, V147, P85, DOI 10.1530/eje.0.1470085
   Walther MM, 2002, ANN NY ACAD SCI, V970, P41, DOI 10.1111/j.1749-6632.2002.tb04411.x
NR 29
TC 59
Z9 60
U1 0
U2 3
PU BMC
PI LONDON
PA CAMPUS, 4 CRINAN ST, LONDON N1 9XW, ENGLAND
SN 1750-1172
J9 ORPHANET J RARE DIS
JI Orphanet J. Rare Dis.
PD DEC 8
PY 2006
VL 1
AR 49
DI 10.1186/1750-1172-1-49
PG 7
WC Genetics & Heredity; Medicine, Research & Experimental
WE Science Citation Index Expanded (SCI-EXPANDED)
SC Genetics & Heredity; Research & Experimental Medicine
GA 240KR
UT WOS:000251587000001
PM 17156452
OA gold, Green Published
DA 2023-08-27
ER

PT J
AU SACKS, AD
   KOTZEN, J
   SACKS, AJ
   GOLDSCHMIDT, RP
AF SACKS, AD
   KOTZEN, J
   SACKS, AJ
   GOLDSCHMIDT, RP
TI DIAGNOSIS AND MANAGEMENT OF PEDIATRIC BRAIN-STEM GLIOMAS
SO SOUTH AFRICAN JOURNAL OF SURGERY
LA English
DT Article
AB A review of paediatric brainstem gliomas (BSGs) treated in the Department of Radiation Oncology of the University of the Witwatersrand teaching hospital group is presented.  Eleven patients between the ages of 4 years and 9 years were seen in the period 1982 - 1992.  Of these cases, 9 were diffuse, 1 focal and 1 exophytic; the radiological features classifying these primary brainstem tumours are described.  The survival from initiation of treatment was longest for the exophytic type BSG (72 weeks), while little difference in survival between the 9 diffuse type BSGs (mean 21,5 weeks) and the single focal type BSG (21 weeks) was found.  The treatment is described and the role of hyperfractionated radiotherapy is reviewed.
RP SACKS, AD (通讯作者)，HILLBROW HOSP,DEPT DIAGNOST RADIOL,HOHANNESBURG,SOUTH AFRICA.
NR 0
TC 1
Z9 1
U1 0
U2 0
PU MED ASSOC S AFRICA
PI JOHANNESBURG
PA MED HOUSE CENTRAL SQ 7430 PINELANDS JOHANNESBURG, SOUTH AFRICA
SN 0038-2361
J9 S AFR J SURG
JI South Afr. J. Surg.
PD JUN
PY 1994
VL 32
IS 2
BP 51
EP 57
PG 7
WC Surgery
WE Science Citation Index Expanded (SCI-EXPANDED)
SC Surgery
GA PM945
UT WOS:A1994PM94500005
PM 7502171
DA 2023-08-27
ER

PT J
AU Wang, LA
   Zhang, JT
   Wu, Z
   Jia, GJ
   Zhang, LW
   Hao, SY
   Geng, SM
AF Wang, Liang
   Zhang, Junting
   Wu, Zhen
   Jia, Guijun
   Zhang, Liwei
   Hao, Shuyu
   Geng, Sumin
TI Diagnosis and Management of Adult Intracranial Neurenteric Cysts
SO NEUROSURGERY
LA English
DT Article
DE Diagnosis; Enterogenous cysts; Neurenteric cysts; Therapy Microsurgery
ID ENTEROGENOUS CYST; CEREBELLOPONTINE ANGLE
AB BACKGROUND: Intracranial neurenteric cysts are rare, benign lesions of the central nervous system.
   OBJECTIVE: To analyze our experience with these lesions and conduct a review of the literature with the aim of identifying a rational approach to diagnosis and management.
   METHODS: Seven patients underwent surgical treatment for intracranial neurenteric cysts from July 2000 to December 2008. The general strategy was to perform complete resection whenever possible. The clinical, radiological, operative, and pathological findings of the patients were reviewed retrospectively.
   RESULTS: This series included 3 women and 4 men. The age of hospitalized patients ranged from 19 to 55 years, with an average age of 34 years. Headache and dizziness were the most common chief symptoms. The imaging spectrum for these lesions is broad, leading to several preoperative misdiagnoses. All 7 lesions were resected by a far lateral transcondylar or retrosigmoid approach. Total tumor resection was achieved in 3 patients. Four patients presented transient fever postoperatively. All patients were healthy and showed no signs of tumor recurrence at their latest follow-up.
   CONCLUSION: Intracranial neurenteric cysts ventral to the brainstem are rare lesions occurring in young adults. These lesions should be considered in the differential diagnosis for intracranial extra-axial cystic lesions anterior to the brainstem. Total surgical resection seems to be the most effective therapeutic method, although radical resection at the risk of impairing the brainstem is not recommended. This benign lesion has a favorable overall prognosis.
C1 [Wang, Liang; Zhang, Junting; Wu, Zhen; Jia, Guijun; Zhang, Liwei; Hao, Shuyu; Geng, Sumin] Capital Med Univ, Beijing Tiantan Hosp, Dept Neurosurg, Beijing 100050, Peoples R China.
C3 Capital Medical University
RP Zhang, JT (通讯作者)，Capital Med Univ, Beijing Tiantan Hosp, Dept Neurosurg, Tiantan Xili 6, Beijing 100050, Peoples R China.
EM
CR AOKI S, 1987, NEURORADIOLOGY, V29, P291, DOI 10.1007/BF00451770
   ARNOLD HM, 2006, NEUROSURGERY, V58, P97
   Bejjani GK, 1998, J NEUROSURG, V89, P326, DOI 10.3171/jns.1998.89.2.0326
   BROOKS BS, 1993, AM J NEURORADIOL, V14, P735
   Chaynes P, 1998, ACTA NEUROCHIR, V140, P905, DOI 10.1007/s007010050192
   Christov C, 2004, NEUROSURGERY, V54, P759, DOI 10.1227/01.NEU.0000109538.07853.7F
   Emerson RE, 2004, APPL IMMUNOHISTO M M, V12, P230, DOI 10.1097/00129039-200409000-00008
   Fuse T, 1998, SURG NEUROL, V50, P431, DOI 10.1016/S0090-3019(97)00280-2
   Gao P, 1995, INT J NEURORADIOL, V1, P17
   Hayashi Y, 1999, J COMPUT ASSIST TOMO, V23, P34, DOI 10.1097/00004728-199901000-00008
   HUTCHISON J, 1953, BRIT J SURG, V41, P15, DOI 10.1002/bjs.18004116507
   JORGE AL, 1995, CHILD NERV SYST, V11, P936
   Kapoor V, 2002, AM J NEURORADIOL, V23, P476
   KLEIHUES P, 1993, BRAIN PATHOL, V3, P255, DOI 10.1111/j.1750-3639.1993.tb00752.x
   LEVENTER DB, 1994, OPHTHALMOLOGY, V101, P1614
   LIU KJ, 2005, NEUROSURG FOCUS, V19, pE9
   Mylonas C, 1996, J CLIN NEUROSCI, V3, P79, DOI 10.1016/S0967-5868(96)90088-0
   Osborn A., 2004, DIAGNOSTIC IMAGING B, P40
   Preece MT, 2006, AM J NEURORADIOL, V27, P1211
   Rao MB, 1996, CLIN NEUROL NEUROSUR, V98, P32, DOI 10.1016/0303-8467(95)00076-3
   REWATI RS, 2001, J CLIN NEUROSCI, V8, P133
   RONY KA, 2008, OTOLARYNGOLOGY HEAD, V139, P863
   SMALL JM, 1962, J NEUROL NEUROSUR PS, V25, P184
   VAIJAYANTEE K, 2000, NEUROSURGERY, V47, P764
   VEENEKLAAS GMH, 1952, AMA AM J DIS CHILD, V83, P500, DOI 10.1001/archpedi.1952.02040080096010
   WANG YM, 2002, CHIN J NEUROSURG, V18, P122
   WOO PYC, 1982, POSTGRAD MED J, V58, P424, DOI 10.1136/pgmj.58.681.424
   ZULCH KJ, 1979, HISTOLOGICAL TYPING, P21
NR 28
TC 46
Z9 50
U1 0
U2 5
PU OXFORD UNIV PRESS INC
PI CARY
PA JOURNALS DEPT, 2001 EVANS RD, CARY, NC 27513 USA
SN 0148-396X
EI 1524-4040
J9 NEUROSURGERY
JI Neurosurgery
PD JAN
PY 2011
VL 68
IS 1
BP 44
EP 52
DI 10.1227/NEU.0b013e3181fc5ee0
PG 9
WC Clinical Neurology; Surgery
WE Science Citation Index Expanded (SCI-EXPANDED)
SC Neurosciences & Neurology; Surgery
GA 694IE
UT WOS:000285288200025
PM 21150754
DA 2023-08-27
ER

PT J
AU Cole, DE
   Lester-McCully, CM
   Widemann, BC
   Warren, KE
AF Cole, Diane E.
   Lester-McCully, Cynthia M.
   Widemann, Brigitte C.
   Warren, Katherine E.
TI Plasma and cerebrospinal fluid pharmacokinetics of the Akt inhibitor,
   perifosine, in a non-human primate model
SO CANCER CHEMOTHERAPY AND PHARMACOLOGY
LA English
DT Article
DE Pharmacology; Non-human primate; Perifosine; Pharmacokinetics; Glioma
ID PHASE-I; GLIOMAS; ALKYLPHOSPHOCHOLINE; MICRODIALYSIS; TEMOZOLOMIDE;
   EVEROLIMUS; ACTIVATION; PATHWAY; BRAIN
AB Central nervous system tumors are histologically and biologically heterogeneous. Standard treatment for malignant tumors includes surgery, radiation and chemotherapy, yet surgical resection is not always an option and chemotherapeutic agents have limited benefit. Recent investigations have focused on molecularly targeted therapies aimed at critical tumorigenic pathways. Several tumor types, including high-grade gliomas and pediatric pontine gliomas, exhibit Akt activation. Perifosine, an orally bioavailable, synthetic alkylphospholipid and potent Akt inhibitor, has demonstrated activity in some preclinical models, but absent activity in a genetically engineered mouse model of pontine glioma. We evaluated the plasma and cerebrospinal fluid pharmacokinetics of orally administered perifosine in a non-human primate model to evaluate CNS penetration.
   Perifosine was administered orally to three adult rhesus monkeys as a single dose of 7.0 mg/kg perifosine. Serial paired plasma and CSF samples were collected for up to 64 days. Perifosine was quantified with a validated HPLC/tandem mass spectrometry assay. Pharmacokinetic parameters were estimated using non-compartmental methods. CSF penetration was calculated from the areas under the concentration-time curves.
   Peak plasma concentrations (C (max)) ranged from 11.7-19.3 A mu M, and remained > 1 A mu M for > 28 days. Time to C (max) (T (max)) was 19 h. The median (range) AUC(Pl) was 3148 (2502-4705) A mu M/h, with a median (range) terminal half-life (t (1/2)) of 193 (170-221) h. Plasma clearance was 494 (329-637) mL/h/kg. Peak CSF concentrations were 4.1-10.1 nM (T (max) 64-235 h). CSF AUCs and t (1/2) were 6358 (2266-7568) nM/h and 277 (146-350) h, respectively. Perifosine concentrations in the CSF remained over nM for > 35 days. The mean CSF penetration was 0.16 %.
   CNS penetration of perifosine after systemic administration is poor. However, levels were measurable in both plasma and CSF for an extended time (> 2 months) after a single oral dose.
C1 [Cole, Diane E.; Widemann, Brigitte C.] NCI, Pharmacol & Expt Therapeut, POB, NIH, Bethesda, MD 20892 USA.
   [Lester-McCully, Cynthia M.; Warren, Katherine E.] NCI, Pediat Neurooncol Sect, POB, NIH, Bethesda, MD 20892 USA.
C3 National Institutes of Health (NIH) - USA; NIH National Cancer Institute
   (NCI); National Institutes of Health (NIH) - USA; NIH National Cancer
   Institute (NCI)
RP Warren, KE (通讯作者)，NCI, Pediat Neurooncol Sect, POB, NIH, 10 Ctr Dr,Bldg 10-Rm 1 W-5750, Bethesda, MD 20892 USA.
EM
FU Intramural Research Program of the National Institutes of Health,
   National Cancer Institute, Center for Cancer Research
FX This work was presented in part at the 2012 International Society of
   Pediatric Neuro-Oncology Meeting in Toronto. This research was supported
   in part by the Intramural Research Program of the National Institutes of
   Health, National Cancer Institute, Center for Cancer Research. The views
   herein do not necessarily represent the official views of the National
   Cancer Institute, the National Institutes of Health, the U.S. Department
   of Health and Human Services, or any other agency of the U.S.
   Government.
CR [Anonymous], 2011, GUIDE CARE USE LAB A
   Bagley RG, 2011, CANCER CHEMOTH PHARM, V68, P1537, DOI 10.1007/s00280-011-1658-0
   Becher OJ, 2010, CANCER RES, V70, P2548, DOI 10.1158/0008-5472.CAN-09-2503
   Crul M, 2002, EUR J CANCER, V38, P1615, DOI 10.1016/S0959-8049(02)00127-2
   de la Pena L, 2006, MOL CANCER THER, V5, P1504, DOI 10.1158/1535-7163.MCT-06-0091
   Fox E, 2002, J PHARMACOL EXP THER, V301, P1003, DOI 10.1124/jpet.301.3.1003
   Gills JJ, 2009, CURR ONCOL REP, V11, P102, DOI 10.1007/s11912-009-0016-4
   Hennessy BT, 2007, CLIN CANCER RES, V13, P7421, DOI 10.1158/1078-0432.CCR-07-0760
   Jacobs S, 2010, CANCER CHEMOTH PHARM, V65, P817, DOI 10.1007/s00280-009-1085-7
   Kondapaka SB, 2003, MOL CANCER THER, V2, P1093
   Konstantinov SM, 1998, INT J CANCER, V77, P778, DOI 10.1002/(SICI)1097-0215(19980831)77:5<778::AID-IJC18>3.0.CO;2-Y
   Kreisl TN, 2009, J NEURO-ONCOL, V92, P99, DOI 10.1007/s11060-008-9741-z
   Kumar A, 2009, MOL CANCER RES, V7, P1813, DOI 10.1158/1541-7786.MCR-09-0069
   Li Z, 2010, JNCI-J NATL CANCER I, V102, P759
   MCCULLY CL, 1990, LAB ANIM SCI, V40, P520
   Momota H, 2005, CANCER RES, V65, P7429, DOI 10.1158/0008-5472.CAN-05-1042
   Mueller S, 2012, NEURO-ONCOLOGY, V14, P1146, DOI 10.1093/neuonc/nos140
   Pollack IF, 2010, J NEURO-ONCOL, V99, P155, DOI 10.1007/s11060-010-0297-3
   Sarkaria JN, 2011, INT J RADIAT ONCOL, V81, P468, DOI 10.1016/j.ijrobp.2010.05.064
   Schmidt-Hieber M, 2012, INVEST NEW DRUG, V30, P480, DOI 10.1007/s10637-010-9576-2
   Stambolic V, 1998, CELL, V95, P29, DOI 10.1016/S0092-8674(00)81780-8
   Tokunaga E, 2008, CURR CANCER DRUG TAR, V8, P27, DOI 10.2174/156800908783497140
   Unger C, 2010, EUR J CANCER, V46, P920, DOI 10.1016/j.ejca.2009.12.028
   Van Ummersen L, 2004, CLIN CANCER RES, V10, P7450, DOI 10.1158/1078-0432.CCR-03-0406
   Woo EW, 2001, J CHROMATOGR B, V759, P247, DOI 10.1016/S0378-4347(01)00231-6
NR 25
TC 10
Z9 10
U1 0
U2 5
PU SPRINGER
PI NEW YORK
PA ONE NEW YORK PLAZA, SUITE 4600, NEW YORK, NY, UNITED STATES
SN 0344-5704
EI 1432-0843
J9 CANCER CHEMOTH PHARM
JI Cancer Chemother. Pharmacol.
PD MAY
PY 2015
VL 75
IS 5
BP 923
EP 928
DI 10.1007/s00280-015-2711-1
PG 6
WC Oncology; Pharmacology & Pharmacy
WE Science Citation Index Expanded (SCI-EXPANDED)
SC Oncology; Pharmacology & Pharmacy
GA CH5DU
UT WOS:000354055800006
PM 25740692
OA Green Accepted
DA 2023-08-27
ER

PT J
AU Phelps, AL
   Salguero, FJ
   Hunter, L
   Stoll, AL
   Jenner, DC
   O'Brien, LM
   Williamson, ED
   Lever, MS
   Laws, TR
AF Phelps, Amanda L.
   Salguero, Francisco J.
   Hunter, Laura
   Stoll, Alexander L.
   Jenner, Dominic C.
   O'Brien, Lyn M.
   Williamson, E. Diane
   Lever, M. Stephen
   Laws, Thomas R.
TI Tumour Necrosis Factor-& alpha;, Chemokines, and Leukocyte Infiltrate
   Are Biomarkers for Pathology in the Brains of Venezuelan Equine
   Encephalitis (VEEV)-Infected Mice
SO VIRUSES-BASEL
LA English
DT Article
DE alphavirus; VEEV; venezuelan equine encephalitis virus; inflammation;
   cytokines; chemokines; leukocytes; pathology; mouse
ID ANTIBODY PROTECTS MICE; MONOCLONAL-ANTIBODY; AIRBORNE CHALLENGE;
   GLYCOPROTEIN PROTECTS; PARTICLE VACCINE; IMMUNE-RESPONSES; VIRUS
   INFECTION; CLINICAL-TRIAL; MURINE MODELS; 24 H
AB Venezuelan equine encephalitis virus (VEEV) is a disease typically confined to South and Central America, whereby human disease is characterised by a transient systemic infection and occasionally severe encephalitis, which is associated with lethality. Using an established mouse model of VEEV infection, the encephalitic aspects of the disease were analysed to identify biomarkers associated with inflammation. Sequential sampling of lethally challenged mice (infected subcutaneously) confirmed a rapid onset systemic infection with subsequent spread to the brain within 24 h of the challenge. Changes in inflammatory biomarkers (TNF-a, CCL-2, and CCL-5) and CD45(+) cell counts were found to correlate strongly to pathology (R>0.9) and present previously unproven biomarkers for disease severity in the model, more so than viral titre. The greatest level of pathology was observed within the olfactory bulb and midbrain/thalamus. The virus was distributed throughout the brain/encephalon, often in areas not associated with pathology. The principal component analysis identified five principal factors across two independent experiments, with the first two describing almost half of the data: (1) confirmation of a systemic Th1-biased inflammatory response to VEEV infection, and (2) a clear correlation between specific inflammation of the brain and clinical signs of disease. Targeting strongly associated biomarkers of deleterious inflammation may ameliorate or even eliminate the encephalitic syndrome of this disease.
C1 [Phelps, Amanda L.; Stoll, Alexander L.; Jenner, Dominic C.; O'Brien, Lyn M.; Williamson, E. Diane; Lever, M. Stephen; Laws, Thomas R.] Def Sci & Technol Lab, Salisbury SP4 0JQ, England.
   [Salguero, Francisco J.; Hunter, Laura] UK Hlth Secur Agcy, Salisbury SP4 0JG, England.
C3 Defence Science & Technology Laboratory; UK Health Security Agency
   (UKHSA)
RP Laws, TR (通讯作者)，Def Sci & Technol Lab, Salisbury SP4 0JQ, England.
EM
FU Defense Threat Reduction Agency, USA [HDTRA117-CBMB-05-2-0008]
FX This research was funded by the Defense Threat Reduction Agency, USA,
   contract number HDTRA117-CBMB-05-2-0008.
CR AREND W P, 1990, Progress in Growth Factor Research, V2, P193, DOI 10.1016/0955-2235(90)90018-F
   Bedoui Y, 2021, PLOS NEGLECT TROP D, V15, DOI 10.1371/journal.pntd.0009115
   Blakely PK, 2016, J NEUROINFLAMM, V13, DOI 10.1186/s12974-016-0683-7
   Bocan TM, 2019, VIRUSES-BASEL, V11, DOI 10.3390/v11111052
   Burke CW, 2022, VIRUSES-BASEL, V14, DOI 10.3390/v14071502
   Burke CW, 2019, PLOS PATHOG, V15, DOI 10.1371/journal.ppat.1008157
   Cain MD, 2017, J NEUROIMMUNOL, V308, P118, DOI 10.1016/j.jneuroim.2017.04.008
   Carson MJ, 2006, IMMUNOL REV, V213, P48, DOI 10.1111/j.1600-065X.2006.00441.x
   Ceballos-Olvera I, 2011, PLOS PATHOG, V7, DOI 10.1371/journal.ppat.1002452
   Charles PC, 2001, VIROLOGY, V284, P190, DOI 10.1006/viro.2001.0878
   Coates EE, 2022, LANCET INFECT DIS, V22, P1210, DOI 10.1016/S1473-3099(22)00052-4
   Daniels BP, 2014, MBIO, V5, DOI 10.1128/mBio.01476-14
   Daza E., 1995, MMWR MORB MORTAL WKL, V44, P721
   Dhimolea E, 2010, MABS-AUSTIN, V2, P3, DOI 10.4161/mabs.2.1.10328
   Durrant DM, 2016, ACS CHEM NEUROSCI, V7, P464, DOI 10.1021/acschemneuro.6b00043
   Fei Y, 2011, J INFECT DIS, V204, P348, DOI 10.1093/infdis/jir266
   Gadani SP, 2012, J IMMUNOL, V189, P4213, DOI 10.4049/jimmunol.1202246
   Goodchild SA, 2011, ANTIVIR RES, V90, P1, DOI 10.1016/j.antiviral.2011.01.010
   Hannaman D, 2016, VACCINE, V34, P3607, DOI 10.1016/j.vaccine.2016.04.077
   HANSON RP, 1967, SCIENCE, V158, P1283, DOI 10.1126/science.158.3806.1283
   Hollidge BS, 2021, VIRULENCE, V12, P430, DOI 10.1080/21505594.2020.1870834
   Honnold S.P., 2018, MED ASPECTS BIOL WAR, P483
   Horby PW, 2022, LANCET, V400, P359, DOI 10.1016/S0140-6736(22)01109-6
   Horby PW, 2022, LANCET, V399, P143, DOI 10.1016/S0140-6736(21)01825-0
   Horby PW, 2021, LANCET, V397, P1637, DOI 10.1016/S0140-6736(21)00676-0
   Hu WG, 2010, VACCINE, V28, P5558, DOI 10.1016/j.vaccine.2010.06.038
   Hunt AR, 2006, J GEN VIROL, V87, P2467, DOI 10.1099/vir.0.81925-0
   Hunt AR, 2011, VIROLOGY, V414, P146, DOI 10.1016/j.virol.2011.03.016
   Irani DN, 2007, J NEUROPATH EXP NEUR, V66, P533, DOI 10.1097/01.jnen.0000263867.46070.e2
   Jones S.A., 2018, CONSID MED, V2, P2, DOI [10.1136/conmed-2018-000005, DOI 10.1136/CONMED-2018-000005]
   Kafai NM, 2022, J EXP MED, V219, DOI 10.1084/jem.20212532
   Laws TR, 2019, EXPERT REV ANTI-INFE, V17, P957, DOI 10.1080/14787210.2018.1496330
   Lehman CW, 2022, VIRUSES-BASEL, V14, DOI 10.3390/v14061210
   Lukaszewski RA, 2000, J VIROL, V74, P5006, DOI 10.1128/JVI.74.11.5006-5015.2000
   Luschnig P, 2021, BIOCHEM PHARMACOL, V192, DOI 10.1016/j.bcp.2021.114690
   Miossec P, 2009, NEW ENGL J MED, V361, P888, DOI 10.1056/NEJMra0707449
   Montiel Milagros, 2015, Antiviral Chemistry & Chemotherapy, V24, P99, DOI 10.1177/2040206616660851
   Newton R, 2007, MOL PHARMACOL, V72, P799, DOI 10.1124/mol.107.038794
   O'Brien LM, 2012, VIROLOGY, V426, P100, DOI 10.1016/j.virol.2012.01.038
   O'Brien LM, 2009, VIROL J, V6, DOI 10.1186/1743-422X-6-206
   Parker MD, 2010, J VIROL, V84, P12683, DOI 10.1128/JVI.01345-10
   Phillpotts RJ, 2006, VIRUS RES, V120, P107, DOI 10.1016/j.virusres.2006.02.003
   Phillpotts RJ, 2005, VACCINE, V23, P1615, DOI 10.1016/j.vaccine.2004.06.056
   Phillpotts RJ, 2003, J INTERF CYTOK RES, V23, P47, DOI 10.1089/10799900360520441
   Phillpotts RJ, 2002, VACCINE, V20, P1497, DOI 10.1016/S0264-410X(01)00505-9
   Pittman P.R., 2018, MED ASPECTS BIOL WAR, P773
   recife, ABOUT US
   Recovery Collaborative Group, 2021, NEW ENGL J MED, V384, P693, DOI [10.1056/NEJMoa2021436, DOI 10.1056/NEJMOA2021436]
   Reed DS, 2014, J VIROL, V88, P12077, DOI 10.1128/JVI.01406-14
   Risner K, 2019, VIRUSES-BASEL, V11, DOI 10.3390/v11121151
   Schafer A, 2011, J VIROL, V85, P10682, DOI 10.1128/JVI.05032-11
   Schoneboom BA, 2000, J NEUROIMMUNOL, V109, P132, DOI 10.1016/S0165-5728(00)00290-3
   Schwartz DM, 2018, NAT REV DRUG DISCOV, V17, DOI 10.1038/nrd.2017.267
   Sharma A, 2011, VIROL J, V8, DOI 10.1186/1743-422X-8-197
   Sharma A, 2008, BMC GENOMICS, V9, DOI 10.1186/1471-2164-9-289
   Steele KE, 2010, VET PATHOL, V47, P790, DOI 10.1177/0300985810372508
   Steinskog ESS, 2016, ONCOTARGET, V7, P45789, DOI 10.18632/oncotarget.9953
   Stromberg ZR, 2020, VACCINES-BASEL, V8, DOI 10.3390/vaccines8020273
   Tretyakova I, 2020, VACCINE, V38, P3378, DOI 10.1016/j.vaccine.2020.02.007
   usammda, ABOUT US
   van Vollenhoven RF, 2013, CURR OPIN RHEUMATOL, V25, P391, DOI 10.1097/BOR.0b013e32835fd828
   Weaver SC, 2004, ANNU REV ENTOMOL, V49, P141, DOI 10.1146/annurev.ento.49.061802.123422
   Zarghi A, 2011, IRAN J PHARM RES, V10, P655
NR 63
TC 0
Z9 0
U1 1
U2 1
PU MDPI
PI BASEL
PA ST ALBAN-ANLAGE 66, CH-4052 BASEL, SWITZERLAND
EI 1999-4915
J9 VIRUSES-BASEL
JI Viruses-Basel
PD JUN
PY 2023
VL 15
IS 6
AR 1307
DI 10.3390/v15061307
PG 21
WC Virology
WE Science Citation Index Expanded (SCI-EXPANDED)
SC Virology
GA L1SA3
UT WOS:001021112500001
PM 37376607
OA gold, Green Published
DA 2023-08-27
ER

PT J
AU Fukuda, T
   Takahashi, K
   Suzuki, T
   Saruta, M
   Watanabe, M
   Nakata, T
   Sasano, H
AF Fukuda, T
   Takahashi, K
   Suzuki, T
   Saruta, M
   Watanabe, M
   Nakata, T
   Sasano, H
TI Urocortin 1, urocortin 3/stresscopin, and corticotropin releasing factor
   receptors in human adrenal and its disorders
SO JOURNAL OF CLINICAL ENDOCRINOLOGY & METABOLISM
LA English
DT Article
ID IMPAIRED STRESS-RESPONSE; BEHAVIORAL-RESPONSES; ZONA GLOMERULOSA;
   CHRONIC INFUSION; MICE DEFICIENT; MESSENGER-RNA; EXPRESSION; ANXIETY;
   IDENTIFICATION; SECRETION
AB Context: Urocortin 1 (Ucn1) and urocortin 3 (Ucn3)/stresscopin are new members of the corticotropin-releasing factor (CRF) neuropeptide family. Ucn1 binds to both CRF type 1 (CRF1) and type 2 receptors (CRF2), whereas Ucn3 is a specific agonist for CRF2. Recently, direct involvement of the locally synthesized CRF family in adrenocortical function has been proposed.
   Objective, Design, and Setting: We examined in situ expression of Ucn and CRF receptors in nonpathological human adrenal gland and its disorders using immunohistochemistry and mRNA in situ hybridization.
   Results: Ucn immunoreactivity was localized in the cortex and medulla of nonpathological adrenal glands. Ucn1 immunoreactivity was marked in the medulla, whereas Ucn3 was immunostained mostly in the cortex. Both CRF type 1 and CRF2 were expressed in the cortex, particularly in the zonae fasciculata and reticularis but very weakly or undetectably in the medulla. Immunohistochemistry in serial tissue sections with mirror images revealed that both Ucn3 and CRF2 were colocalized in more than 85% of the adrenocortical cells. mRNA in situ hybridization confirmed these findings above. In fetal adrenals, Ucn and CRF receptors were expressed in both fetal and definitive zones of the cortex. Ucn and CRF receptors were all expressed in the tumor cells of pheochromocytomas, adrenocortical adenomas, and carcinomas, but its positivity was less than that in nonpathological adrenal glands, suggesting that Ucn1, Ucn3, and CRF receptors were down-regulated in these adrenal neoplasms.
   Conclusions: Ucn1, Ucn3, and CRF receptors are all expressed in human adrenal cortex and medulla and may play important roles in physiological adrenal functions.
C1 Tohoku Univ, Sch Med, Dept Pathol, Aoba Ku, Sendai, Miyagi 9808575, Japan.
   Tohoku Univ, Sch Med, Dept Mol Biol & Appl Physiol, Sendai, Miyagi 9808575, Japan.
   Tohoku Univ, Sch Med, Dept Analyt Med Technol, Sendai, Miyagi 9808575, Japan.
   Kyowa Medex Co Ltd, Res Lab, Shizuoka 4110932, Japan.
   Kyowa Hakko Kogyo Co Ltd, Pharmaceut Mkt Ctr, Tokyo 1008185, Japan.
C3 Tohoku University; Tohoku University; Tohoku University; Kyowa Kirin Ltd
RP Fukuda, T (通讯作者)，Tohoku Univ, Sch Med, Dept Pathol, Aoba Ku, 2-1 Seiryo Machi, Sendai, Miyagi 9808575, Japan.
EM
RI Saruta, Masayuki/ABL-4991-2022; Takahashi, Kazuhiro/A-4887-2010
OI Saruta, Masayuki/0000-0001-8172-3240; Takahashi,
   Kazuhiro/0000-0001-8943-1875
CR Andreis PG, 2000, ENDOCRINOLOGY, V141, P2098, DOI 10.1210/en.141.6.2098
   Bale TL, 2002, J NEUROSCI, V22, P193, DOI 10.1523/JNEUROSCI.22-01-00193.2002
   Bale TL, 2000, NAT GENET, V24, P410, DOI 10.1038/74263
   Bornstein SR, 2000, Z RHEUMATOL, V59, P12
   Chakravorty A, 1999, J CLIN ENDOCR METAB, V84, P3732, DOI 10.1210/jc.84.10.3732
   CHEN RP, 1993, P NATL ACAD SCI USA, V90, P8967, DOI 10.1073/pnas.90.19.8967
   COZZA EN, 1990, ENDOCRINOLOGY, V127, P549, DOI 10.1210/endo-127-2-549
   Donaldson CJ, 1996, ENDOCRINOLOGY, V137, P2167, DOI 10.1210/en.137.5.2167
   Dunn AJ, 1999, BRAIN RES, V845, P14, DOI 10.1016/S0006-8993(99)01912-5
   EVANS CJ, 1983, SCIENCE, V221, P957, DOI 10.1126/science.6308766
   Heinrichs SC, 1997, REGUL PEPTIDES, V71, P15, DOI 10.1016/S0167-0115(97)01005-7
   Hsu SY, 2001, NAT MED, V7, P605, DOI 10.1038/87936
   Iino K, 1999, CLIN ENDOCRINOL, V50, P107, DOI 10.1046/j.1365-2265.1999.00624.x
   Iino K, 1997, J CLIN ENDOCR METAB, V82, P3842, DOI 10.1210/jc.82.11.3842
   Ikeda K, 2002, LIFE SCI, V71, P1929, DOI 10.1016/S0024-3205(02)01945-8
   Kimura Y, 2002, J CLIN ENDOCR METAB, V87, P340, DOI 10.1210/jc.87.1.340
   Koob GF, 1999, BRAIN RES, V848, P141, DOI 10.1016/S0006-8993(99)01991-5
   Kostich WA, 1998, MOL ENDOCRINOL, V12, P1077, DOI 10.1210/me.12.8.1077
   Lewis K, 2001, P NATL ACAD SCI USA, V98, P7570, DOI 10.1073/pnas.121165198
   Li C, 2003, ENDOCRINOLOGY, V144, P3216, DOI 10.1210/en.2002-0087
   LIEBSCH G, 1995, REGUL PEPTIDES, V59, P229, DOI 10.1016/0167-0115(95)00099-W
   Liebsch G, 1999, J PSYCHIAT RES, V33, P153, DOI 10.1016/S0022-3956(98)80047-2
   LOVENBERG TW, 1995, P NATL ACAD SCI USA, V92, P836, DOI 10.1073/pnas.92.3.836
   LUNDBERG JM, 1979, P NATL ACAD SCI USA, V76, P4079, DOI 10.1073/pnas.76.8.4079
   MCLEAN M, 1995, NAT MED, V1, P460, DOI 10.1038/nm0595-460
   Muramatsu Y, 2001, J CLIN ENDOCR METAB, V86, P1362, DOI 10.1210/jc.86.3.1362
   Muramatsu Y, 2000, PEPTIDES, V21, P1799, DOI 10.1016/S0196-9781(00)00335-1
   Nawata H, 1995, ANN REPORT MINISTRY, P223
   Odagiri E, 2004, ENDOCR J, V51, P295, DOI 10.1507/endocrj.51.295
   Okuyama S, 1999, J PHARMACOL EXP THER, V289, P926
   Petraglia F, 1996, J CLIN ENDOCR METAB, V81, P3807, DOI 10.1210/jc.81.10.3807
   Reubi JC, 2003, J CLIN ENDOCR METAB, V88, P3312, DOI 10.1210/jc.2002-021853
   Reul JMHM, 2002, CURR OPIN PHARMACOL, V2, P23, DOI 10.1016/S1471-4892(01)00117-5
   Reyes TM, 2001, P NATL ACAD SCI USA, V98, P2843, DOI 10.1073/pnas.051626398
   Ruhmann A, 1998, P NATL ACAD SCI USA, V95, P15264, DOI 10.1073/pnas.95.26.15264
   Saruta M, 2005, PEPTIDES, V26, P1196, DOI 10.1016/j.peptides.2005.01.014
   Saruta M, 2004, J CLIN ENDOCR METAB, V89, P5352, DOI 10.1210/jc.2004-0195
   SASAKI A, 1988, J CLIN ENDOCR METAB, V67, P768, DOI 10.1210/jcem-67-4-768
   Seres J, 2004, J CLIN ENDOCR METAB, V89, P965, DOI 10.1210/jc.2003-031299
   Sirianni R, 2005, J CLIN ENDOCR METAB, V90, P279, DOI 10.1210/jc.2004-0865
   Smagin GN, 2001, PEPTIDES, V22, P713, DOI 10.1016/S0196-9781(01)00384-9
   Smith GW, 1998, NEURON, V20, P1093, DOI 10.1016/S0896-6273(00)80491-2
   Smith R, 1998, J CLIN ENDOCR METAB, V83, P2916, DOI 10.1210/jc.83.8.2916
   STENZELPOORE MP, 1994, J NEUROSCI, V14, P2579
   SUDA T, 1984, J CLIN ENDOCR METAB, V59, P861, DOI 10.1210/jcem-59-5-861
   Swiergiel AH, 1999, PHARMACOL BIOCHEM BE, V64, P59, DOI 10.1016/S0091-3057(99)00065-9
   Takahashi K, 1998, PEPTIDES, V19, P643, DOI 10.1016/S0196-9781(98)00012-6
   Takahashi K, 2004, J CLIN ENDOCR METAB, V89, P1897, DOI 10.1210/jc.2003-031663
   Takahashi K, 2004, PEPTIDES, V25, P1723, DOI 10.1016/j.peptides.2004.04.018
   Takahashi K, 2002, EXP CLIN ENDOCR DIAB, V110, P373, DOI 10.1055/s-2002-36423
   Takahashi K, 2001, J CLIN ENDOCR METAB, V86, P369, DOI 10.1210/jc.86.1.369
   Takahashi K, 2000, PEPTIDES, V21, P251, DOI 10.1016/S0196-9781(99)00205-3
   Uzuki M, 2001, CLIN SCI, V100, P577, DOI 10.1042/CS20000242
   VAUGHAN J, 1995, NATURE, V378, P287, DOI 10.1038/378287a0
   Venihaki M, 1998, ENDOCRINOLOGY, V139, P713, DOI 10.1210/en.139.2.713
   Vrezas I, 2003, MICROSC RES TECHNIQ, V61, P308, DOI 10.1002/jemt.10340
   WEINGER SC, 1999, P NATL ACAD SCI USA, V96, P8283
   WEISS LM, 1984, AM J SURG PATHOL, V8, P163, DOI 10.1097/00000478-198403000-00001
   Weninger SC, 1999, REGUL PEPTIDES, V84, P69, DOI 10.1016/S0167-0115(99)00070-1
   Willenberg HS, 2000, MOL PSYCHIATR, V5, P137, DOI 10.1038/sj.mp.4000720
NR 60
TC 41
Z9 41
U1 0
U2 2
PU ENDOCRINE SOC
PI WASHINGTON
PA 2055 L ST NW, SUITE 600, WASHINGTON, DC 20036 USA
SN 0021-972X
EI 1945-7197
J9 J CLIN ENDOCR METAB
JI J. Clin. Endocrinol. Metab.
PD AUG
PY 2005
VL 90
IS 8
BP 4671
EP 4678
DI 10.1210/jc.2005-0090
PG 8
WC Endocrinology & Metabolism
WE Science Citation Index Expanded (SCI-EXPANDED)
SC Endocrinology & Metabolism
GA 953HZ
UT WOS:000231068500038
PM 15914529
OA Bronze
DA 2023-08-27
ER

PT J
AU Schwab, M
AF Schwab, M
TI Human neuroblastoma: From basic science to clinical debut of cellular
   oncogenes
SO NATURWISSENSCHAFTEN
LA English
DT Review
ID N-MYC GENE; NEURO-BLASTOMA; C-MYC; ENHANCED EXPRESSION; CHROMOSOME 1P;
   HUMAN CANCER; SINGLE-COPY; TUMOR-CELLS; AMPLIFICATION; DNA
AB Neuroblastoma is a childhood embryonic tumor of migrating neuroectodermal cells derived from the neural crest and destined for the adrenal medulla and the sympathetic nervous system. It very often has a rapidly progressive clinical course, and although many advances have been made in understanding the development of this tumor, improving the survival rates particularly in patients with metastatic tumor has been a frustrating experience. The mechanisms leading to neuroblastoma are largely unclear, but nonrandom chromosomal changes discovered early suggested the involvement of genetic alterations. Most prominent among these is the amplification of the oncogene MYCN, which identifies a group of patients who have a particularly dire prognosis. Amplified MYCN is used today as a prognostic marker on which therapy design is based to a large extent. An unusual aspect of neuroblastoma is the high rate at which tumors regress spontaneously, even in infants with extensive liver involvement and numerous subcutaneous nodules. Identifying the molecular and cellular basis of spontaneous regression could result in improved therapeutic approaches. Neuroblastoma is a model tumor with many fascinating aspects but has remained a challenge to the pediatric oncologist.
C1 Deutsch Krebsforschungszentrum, Abt Zytogenet H0400, D-69120 Heidelberg, Germany.
C3 Helmholtz Association; German Cancer Research Center (DKFZ)
RP Schwab, M (通讯作者)，Deutsch Krebsforschungszentrum, Abt Zytogenet H0400, Neuenheimer Feld 280, D-69120 Heidelberg, Germany.
CR ALITALO K, 1983, P NATL ACAD SCI-BIOL, V80, P1707, DOI 10.1073/pnas.80.6.1707
   AMLER LC, 1989, MOL CELL BIOL, V9, P4903, DOI 10.1128/MCB.9.11.4903
   Amler LC, 1996, GENE CHROMOSOME CANC, V15, P134, DOI 10.1002/(SICI)1098-2264(199602)15:2<134::AID-GCC9>3.0.CO;2-4
   Berthold F, 1997, EUR J CANCER, V33, P2092, DOI 10.1016/S0959-8049(97)00237-2
   BERTHOLD F, 1994, MONATSSCHR KINDERH, V142, P296
   BERTHOLD F, 1990, NEUROBLASTOMA TUMOR, P1
   BRODEUR GM, 1993, J CLIN ONCOL, V11, P1466, DOI 10.1200/JCO.1993.11.8.1466
   Caron H, 1996, NEW ENGL J MED, V334, P225, DOI 10.1056/NEJM199601253340404
   Coquelle A, 1997, CELL, V89, P215, DOI 10.1016/S0092-8674(00)80201-9
   CORVI R, 1995, ONCOGENE, V10, P1081
   CORVI R, 1994, P NATL ACAD SCI USA, V91, P5523, DOI 10.1073/pnas.91.12.5523
   CORVI R, 1995, CANCER RES, V55, P3471
   Debatisse M, 1998, RECENT RES CANCER, V154, P216
   EKLOF O, 1983, ACTA PAEDIATR SCAND, V72, P473, DOI 10.1111/j.1651-2227.1983.tb09752.x
   FIORILLO A, 1982, J PEDIATR-US, V101, P564, DOI 10.1016/S0022-3476(82)80703-8
   FULDA S, 1999, IN PRESS ONCOGENE
   GAUDRAY P, 1992, MUTAT RES, V276, P317, DOI 10.1016/0165-1110(92)90018-5
   GEHRING M, 1995, CANCER RES, V55, P5366
   Glover TW, 1998, RECENT RES CANCER, V154, P185
   GUAN XY, 1994, NAT GENET, V8, P155, DOI 10.1038/ng1094-155
   HAYASHI Y, 1989, CANCER, V63, P126, DOI 10.1002/1097-0142(19890101)63:1<126::AID-CNCR2820630120>3.0.CO;2-Z
   IBSON JM, 1988, ONCOGENE, V2, P399
   KHATIB ZA, 1993, CANCER RES, V53, P5535
   KUSHNER BH, 1986, CANCER-AM CANCER SOC, V57, P1887, DOI 10.1002/1097-0142(19860501)57:9<1887::AID-CNCR2820570931>3.0.CO;2-7
   Lutz W, 1997, ONCOGENE, V15, P303, DOI 10.1038/sj.onc.1201195
   Lutz W, 1998, ONCOGENE, V17, P339, DOI 10.1038/sj.onc.1200201
   MARIS JM, 1995, CANCER RES, V55, P4664
   NISHI M, 1987, CANCER, V60, P433, DOI 10.1002/1097-0142(19870801)60:3<433::AID-CNCR2820600326>3.0.CO;2-H
   Praml C, 1998, CANCER RES, V58, P5014
   PRAML C, 1995, ONCOGENE, V11, P1357
   Rubie H, 1997, J CLIN ONCOL, V15, P1171, DOI 10.1200/JCO.1997.15.3.1171
   SAVELYEVA L, 1994, AM J HUM GENET, V55, P334
   SAWADA T, 1984, CANCER, V53, P2731, DOI 10.1002/1097-0142(19840615)53:12<2731::AID-CNCR2820531232>3.0.CO;2-D
   SCHIMKE RT, 1992, MUTAT RES, V276, P145, DOI 10.1016/0165-1110(92)90004-S
   SCHIMKE RT, 1984, CELL, V37, P705, DOI 10.1016/0092-8674(84)90406-9
   SCHWAB M, 1984, NATURE, V308, P288, DOI 10.1038/308288a0
   SCHWAB M, 1983, NATURE, V305, P245, DOI 10.1038/305245a0
   SCHWAB M, 1984, P NATL ACAD SCI-BIOL, V81, P4940, DOI 10.1073/pnas.81.15.4940
   Schwab M, 1998, BIOESSAYS, V20, P473, DOI 10.1002/(SICI)1521-1878(199806)20:6<473::AID-BIES5>3.0.CO;2-N
   SCHWAB M, 1990, GENE CHROMOSOME CANC, V1, P181, DOI 10.1002/gcc.2870010302
   SCHWAB M, 1983, NATURE, V303, P497, DOI 10.1038/303497a0
   SCHWAB M, 1985, NATURE, V316, P160, DOI 10.1038/316160a0
   SCHWAB M, 1985, NATURE, V315, P345, DOI 10.1038/315345a0
   SCHWAB M, 1988, P NATL ACAD SCI USA, V85, P9585, DOI 10.1073/pnas.85.24.9585
   Schwab M, 1996, GENE CHROMOSOME CANC, V16, P211, DOI 10.1002/(SICI)1098-2264(199608)16:4<211::AID-GCC1>3.0.CO;2-0
   Schwab M, 1995, NEUROSCIENTIST, V1, P277
   Schwab M, 1997, PATHOLOGY GENETICS T, P117
   SPRADLING AC, 1980, P NATL ACAD SCI-BIOL, V77, P1096, DOI 10.1073/pnas.77.2.1096
   SRIVATSAN ES, 1993, GENE CHROMOSOME CANC, V7, P32, DOI 10.1002/gcc.2870070106
   TAKITA J, 1995, ONCOGENE, V11, P1829
   VERSTEEG R, 1995, EUR J CANCER, V31A, P538, DOI 10.1016/0959-8049(95)00037-J
   WENZEL A, 1995, EUR J CANCER, V31A, P516, DOI 10.1016/0959-8049(95)00060-V
   WENZEL A, 1991, EMBO J, V10, P3703, DOI 10.1002/j.1460-2075.1991.tb04938.x
NR 53
TC 21
Z9 21
U1 0
U2 2
PU SPRINGER VERLAG
PI NEW YORK
PA 175 FIFTH AVE, NEW YORK, NY 10010 USA
SN 0028-1042
J9 NATURWISSENSCHAFTEN
JI Naturwissenschaften
PD FEB
PY 1999
VL 86
IS 2
BP 71
EP 78
DI 10.1007/s001140050574
PG 8
WC Multidisciplinary Sciences
WE Science Citation Index Expanded (SCI-EXPANDED)
SC Science & Technology - Other Topics
GA 168WL
UT WOS:000078716100003
PM 10084150
DA 2023-08-27
ER

PT J
AU Bjorklund, P
   Backman, S
AF Bjorklund, Peyman
   Backman, Samuel
TI Epigenetics of pheochromocytoma and paraganglioma
SO MOLECULAR AND CELLULAR ENDOCRINOLOGY
LA English
DT Article
DE Pheochromocytoma; Paraganglioma; Epigenetics; Adrenal; Methylation
ID CHROMATIN-REMODELING GENES; DNA METHYLATION; MALIGNANT
   PHEOCHROMOCYTOMAS; HISTONE METHYLATION; GERMLINE MUTATIONS; RET
   PROTOONCOGENE; ATRX; EXPRESSION; DAXX; SDHD
AB Pheochromocytomas and paragangliomas (PPGLs) are neuroendocrine tumors arising in the medullae of the adrenal glands or in paraganglia. The knowledge of the tumor biology of these lesions has increased dramatically during the past two decades and more than a dozen recurrently mutated genes have been identified. Different clusters have been described that share epigenetic signatures. Mutations in the succinate dehydrogenase complex subunit genes play a pivotal role in reprogramming the epigenetic state of these tumors by inhibiting epigenetic regulators such as TET enzymes and histone demethylases. Another subgroup of tumors carries hypomethylated genomes, and overexpression of several microRNAs has been described. While much remains to be investigated regarding the epigenetics of PPGLs, it is clear that it plays an important role in PPGL biology. (C) 2017 Elsevier B.V. All rights reserved.
C1 [Bjorklund, Peyman; Backman, Samuel] Uppsala Univ, Dept Surg Sci, Expt Surg, Uppsala, Sweden.
C3 Uppsala University
RP Bjorklund, P (通讯作者)，Rudbeckslab Hus R3,BV Dag Hammarskjolds V 20, S-75185 Uppsala, Sweden.
EM
RI Björklund, Peyman/A-1509-2014
OI Björklund, Peyman/0000-0002-9398-4496
FU Swedish Cancer Society [15 0130]; Selander Foundation
FX Supported by Swedish Cancer Society (15 0130) and Selander Foundation.
   PB is a Swedish Cancer Society Investigator.
CR Astuti D, 2001, ONCOGENE, V20, P7573, DOI 10.1038/sj.onc.1204968
   Astuti D, 2001, AM J HUM GENET, V69, P49, DOI 10.1086/321282
   Ayala-Ramirez M, 2012, J CLIN ENDOCR METAB, V97, P4040, DOI 10.1210/jc.2012-2356
   Backman S, 2017, SCI REP-UK, V7, DOI 10.1038/srep44943
   Bartel DP, 2004, CELL, V116, P281, DOI 10.1016/S0092-8674(04)00045-5
   Bayley JP, 2010, LANCET ONCOL, V11, P366, DOI 10.1016/S1470-2045(10)70007-3
   Baysal BE, 2000, SCIENCE, V287, P848, DOI 10.1126/science.287.5454.848
   Baysal BE, 2011, HUM MOL GENET, V20, P4452, DOI 10.1093/hmg/ddr376
   Bjorklund P, 2016, J INTERN MED, V280, P559, DOI 10.1111/joim.12507
   Brennan CW, 2013, CELL, V155, P462, DOI 10.1016/j.cell.2013.09.034
   Burnichon N, 2012, HUM MOL GENET, V21, P5397, DOI 10.1093/hmg/dds374
   Burnichon N, 2010, HUM MOL GENET, V19, P3011, DOI 10.1093/hmg/ddq206
   Carrozza MJ, 2005, CELL, V123, P581, DOI 10.1016/j.cell.2005.10.023
   Castro-Vega LJ, 2015, NAT COMMUN, V6, DOI 10.1038/ncomms7044
   Castro-Vega LJ, 2014, HUM MOL GENET, V23, P2440, DOI 10.1093/hmg/ddt639
   Cervera AM, 2009, MOL CANCER, V8, DOI 10.1186/1476-4598-8-89
   Comino-Mendez I, 2011, NAT GENET, V43, P663, DOI 10.1038/ng.861
   Crona J, 2013, J CLIN ENDOCR METAB, V98, pE1266, DOI 10.1210/jc.2012-4257
   de Cubas AA, 2015, CLIN CANCER RES, V21, P3020, DOI 10.1158/1078-0432.CCR-14-2804
   de Cubas AA, 2013, ENDOCR-RELAT CANCER, V20, P477, DOI 10.1530/ERC-12-0183
   Eden A, 2003, SCIENCE, V300, P455, DOI 10.1126/science.1083557
   Eguchi K, 1997, CANCER RES, V57, P4913
   Favier J, 2015, NAT REV ENDOCRINOL, V11, P101, DOI 10.1038/nrendo.2014.188
   Fishbein L, 2017, CANCER CELL, V31, P181, DOI 10.1016/j.ccell.2017.01.001
   Fishbein L, 2015, NAT COMMUN, V6, DOI 10.1038/ncomms7140
   Flavahan WA, 2016, NATURE, V529, P110, DOI 10.1038/nature16490
   Friedrich CA, 1999, CANCER-AM CANCER SOC, V86, P2478, DOI 10.1002/(SICI)1097-0142(19991201)86:11+<2478::AID-CNCR4>3.0.CO;2-5
   Geli J, 2008, CLIN CANCER RES, V14, P2551, DOI 10.1158/1078-0432.CCR-07-1867
   Geli J, 2007, ENDOCR-RELAT CANCER, V14, P125, DOI 10.1677/ERC-06-0031
   Gonias S, 2009, J CLIN ONCOL, V27, P4162, DOI 10.1200/JCO.2008.21.3496
   Greer EL, 2012, NAT REV GENET, V13, P343, DOI 10.1038/nrg3173
   Hacloux J., 2014, INT J CANCER, V135, P2711
   Hammons GJ, 1999, CELL BIOL TOXICOL, V15, P389, DOI 10.1023/A:1007658000971
   Heaphy CM, 2011, SCIENCE, V333, P425, DOI 10.1126/science.1207313
   Hegi ME, 2005, NEW ENGL J MED, V352, P997, DOI 10.1056/NEJMoa043331
   Hensen EF, 2004, ONCOGENE, V23, P4076, DOI 10.1038/sj.onc.1207591
   Her YF, 2015, PLOS ONE, V10, DOI 10.1371/journal.pone.0127471
   Hoekstra AS, 2015, ONCOTARGET, V6, P38777, DOI 10.18632/oncotarget.6091
   HOFSTRA RMW, 1994, NATURE, V367, P375, DOI 10.1038/367375a0
   Huang H, 2008, CANCER-AM CANCER SOC, V113, P2020, DOI 10.1002/cncr.23812
   Jenuwein T, 2001, SCIENCE, V293, P1074, DOI 10.1126/science.1063127
   Jia GF, 2011, NAT CHEM BIOL, V7, P885, DOI [10.1038/NCHEMBIO.687, 10.1038/nchembio.687]
   Jiao YC, 2011, SCIENCE, V331, P1199, DOI 10.1126/science.1200609
   Joshua AM, 2009, J CLIN ENDOCR METAB, V94, P5, DOI 10.1210/jc.2008-1836
   Juhlin CC, 2015, GENE CHROMOSOME CANC, V54, P542, DOI 10.1002/gcc.22267
   Killian JK, 2014, SCI TRANSL MED, V6, DOI 10.1126/scitranslmed.3009961
   Kohli RM, 2013, NATURE, V502, P472, DOI 10.1038/nature12750
   Letouze E, 2013, CANCER CELL, V23, P739, DOI 10.1016/j.ccr.2013.04.018
   Lewis PW, 2010, P NATL ACAD SCI USA, V107, P14075, DOI 10.1073/pnas.1008850107
   Loriot C, 2015, ONCOTARGET, V6, P32955, DOI 10.18632/oncotarget.5106
   Marinoni I, 2014, GASTROENTEROLOGY, V146, P453, DOI 10.1053/j.gastro.2013.10.020
   Maunakea AK, 2010, NATURE, V466, P253, DOI 10.1038/nature09165
   Mercer TR, 2009, NAT REV GENET, V10, P155, DOI 10.1038/nrg2521
   Meyer-Rochow GY, 2010, ENDOCR-RELAT CANCER, V17, P835, DOI 10.1677/ERC-10-0142
   Morgan CP, 2011, J NEUROSCI, V31, P11748, DOI 10.1523/JNEUROSCI.1887-11.2011
   Morison IM, 2005, TRENDS GENET, V21, P457, DOI 10.1016/j.tig.2005.06.008
   MULLIGAN LM, 1993, NATURE, V363, P458, DOI 10.1038/363458a0
   Niemann S, 2000, NAT GENET, V26, P268, DOI 10.1038/81551
   Niemeijer ND, 2014, CLIN ENDOCRINOL, V81, P642, DOI 10.1111/cen.12542
   Patterson E, 2012, ENDOCR-RELAT CANCER, V19, P157, DOI 10.1530/ERC-11-0308
   Pembrey ME, 2006, EUR J HUM GENET, V14, P159, DOI 10.1038/sj.ejhg.5201538
   Qin Y, 2014, HUM MOL GENET, V23, P2428, DOI 10.1093/hmg/ddt638
   Qin YJ, 2010, NAT GENET, V42, P229, DOI 10.1038/ng.533
   Richter S, 2016, J CLIN ENDOCR METAB, V101, P359, DOI 10.1210/jc.2015-3856
   Sandgren J, 2010, EXP MOL MED, V42, P484, DOI 10.3858/emm.2010.42.7.050
   Schwartzentruber J, 2012, NATURE, V482, P226, DOI [10.1038/nature10833, 10.1007/s13311-012-0137-6]
   Stenman A., 2016, CLIN ENDOCRINOL OXF
   Tombol Z, 2010, MODERN PATHOL, V23, P1583, DOI 10.1038/modpathol.2010.164
   Toledo RA, 2016, CLIN CANCER RES, V22, P2301, DOI 10.1158/1078-0432.CCR-15-1841
   Tsang VHM, 2014, ENDOCR-RELAT CANCER, V21, P415, DOI 10.1530/ERC-13-0519
   Turcan S, 2012, NATURE, V483, P479, DOI 10.1038/nature10866
   Waddington CH, 2012, INT J EPIDEMIOL, V41, P10, DOI 10.1093/ije/dyr184
   WALLACE MR, 1990, SCIENCE, V249, P181, DOI 10.1126/science.2134734
   Walther MM, 1999, J UROLOGY, V162, P1582, DOI 10.1016/S0022-5347(05)68171-2
   Weber M, 2007, NAT GENET, V39, P457, DOI 10.1038/ng1990
   Welander J, 2012, HUM MOL GENET, V21, P5406, DOI 10.1093/hmg/dds402
   Wu GY, 2004, J NUTR, V134, P2169, DOI 10.1093/jn/134.9.2169
   Wu TP, 2016, NATURE, V532, P329, DOI 10.1038/nature17640
   Xiao MT, 2012, GENE DEV, V26, P1326, DOI 10.1101/gad.191056.112
   Xue YT, 2003, P NATL ACAD SCI USA, V100, P10635, DOI 10.1073/pnas.1937626100
   Zemach A, 2010, SCIENCE, V328, P916, DOI 10.1126/science.1186366
   Zhang Y, 2001, GENE DEV, V15, P2343, DOI 10.1101/gad.927301
   Zhuang ZP, 2012, NEW ENGL J MED, V367, P922, DOI 10.1056/NEJMoa1205119
   Zong LG, 2015, INT J CLIN EXP PATHO, V8, P1545
NR 84
TC 8
Z9 8
U1 1
U2 8
PU ELSEVIER IRELAND LTD
PI CLARE
PA ELSEVIER HOUSE, BROOKVALE PLAZA, EAST PARK SHANNON, CO, CLARE, 00000,
   IRELAND
SN 0303-7207
EI 1872-8057
J9 MOL CELL ENDOCRINOL
JI Mol. Cell. Endocrinol.
PD JUL 5
PY 2018
VL 469
BP 92
EP 97
DI 10.1016/j.mce.2017.06.016
PG 6
WC Cell Biology; Endocrinology & Metabolism
WE Science Citation Index Expanded (SCI-EXPANDED)
SC Cell Biology; Endocrinology & Metabolism
GA GX7KQ
UT WOS:000447949400012
PM 28630023
DA 2023-08-27
ER

PT J
AU Chua, J
   Nafziger, E
   Leung, D
AF Chua, Jason
   Nafziger, Elizabeth
   Leung, Denise
TI Evidence-Based Practice: Temozolomide Beyond Glioblastoma
SO CURRENT ONCOLOGY REPORTS
LA English
DT Review
DE Temozolomide; Low grade glioma; GBM in the elderly; Ependymoma;
   Pilocytic astrocytoma; Pleomorphic xanthoastrocytoma; DIPG; Bevacizumab
ID BRAIN-STEM GLIOMAS; PHASE-II TRIAL; DOSE-INTENSE TEMOZOLOMIDE; PILOCYTIC
   ASTROCYTOMA; CONCURRENT RADIOTHERAPY; ADJUVANT TEMOZOLOMIDE;
   ELDERLY-PATIENTS; LONG-TERM; CHILDREN; THERAPY
AB Purpose of ReviewTemozolomide is a first-line treatment for newly diagnosed glioblastoma. In this review, we will examine the use of temozolomide in other contexts for treating gliomas, including recurrent glioblastoma, glioblastoma in the elderly, diffuse low- and high-grade gliomas, non-diffuse gliomas, diffuse intrinsic pontine glioma (DIPG), ependymoma, pilocytic astrocytoma, and pleomorphic xanthoastrocytoma.Recent FindingsTemozolomide improved survival in older patients with glioblastoma, anaplastic gliomas regardless of 1p/19q deletion status, and progressive ependymomas. Temozolomide afforded less toxicity and comparable efficacy to radiation in high-risk low-grade gliomas and to platinum-based chemotherapy in pediatric high-grade gliomas.SummaryThe success of temozolomide in promoting survival has expanded beyond glioblastoma to benefit patients with non-glioblastoma tumors. Identifying practical biomarkers for predicting temozolomide susceptibility, and establishing complementary agents for chemosensitizing tumors to temozolomide, will be key next steps for future success.
C1 [Chua, Jason; Nafziger, Elizabeth; Leung, Denise] Univ Michigan, Dept Neurol, Taubman Ctr 1914, 1500 E Med Ctr Dr, Ann Arbor, MI 48109 USA.
C3 University of Michigan System; University of Michigan
RP Leung, D (通讯作者)，Univ Michigan, Dept Neurol, Taubman Ctr 1914, 1500 E Med Ctr Dr, Ann Arbor, MI 48109 USA.
EM
CR Abacioglu U, 2011, J NEURO-ONCOL, V103, P585, DOI 10.1007/s11060-010-0423-2
   Agarwala S S, 2000, Oncologist, V5, P144, DOI 10.1634/theoncologist.5-2-144
   Babu R, 2013, J CLIN NEUROSCI, V20, P1382, DOI 10.1016/j.jocn.2012.12.011
   Bailey S, 2013, EUR J CANCER, V49, P3856, DOI 10.1016/j.ejca.2013.08.006
   Baumert BG, 2016, LANCET ONCOL, V17, P1521, DOI 10.1016/S1470-2045(16)30313-8
   Bell EH, 2018, JAMA ONCOL, V4, P1405, DOI 10.1001/jamaoncol.2018.1977
   Bond KM, 2018, WORLD NEUROSURG, V110, P276, DOI 10.1016/j.wneu.2017.11.102
   Brandes AA, 2006, BRIT J CANCER, V95, P1155, DOI 10.1038/sj.bjc.6603376
   BRINDLEY CJ, 1986, BRIT J CANCER, V53, P91, DOI 10.1038/bjc.1986.13
   Buckner JC, 2016, NEW ENGL J MED, V374, P1344, DOI 10.1056/NEJMoa1500925
   Cohen KJ, 2011, NEURO-ONCOLOGY, V13, P410, DOI 10.1093/neuonc/noq205
   Collins VP, 2015, ACTA NEUROPATHOL, V129, P775, DOI 10.1007/s00401-015-1410-7
   Desjardins A, 2012, CANCER-AM CANCER SOC, V118, P1302, DOI 10.1002/cncr.26381
   Dirven CMF, 1997, CHILD NERV SYST, V13, P17, DOI 10.1007/s003810050033
   Drappatz Jan, 2018, Prog Neurol Surg, V31, P152, DOI 10.1159/000467376
   Fisher B, 2018, NEURO-ONCOLOGY, V20, DOI [10.1093/neuonc/noy148.039., DOI 10.1093/NEUONC/NOY148.039]
   Fisher BJ, 2015, INT J RADIAT ONCOL, V91, P497, DOI 10.1016/j.ijrobp.2014.11.012
   Flannery PC, 2018, ONCOL REP, V39, P455, DOI 10.3892/or.2017.6122
   Freyschlag CF, 2011, ANTICANCER RES, V31, P1023
   Gramatzki D, 2016, BMC CANCER, V16, DOI 10.1186/s12885-016-2323-0
   Gururangan S, 2007, NEURO-ONCOLOGY, V9, P161, DOI 10.1215/15228517-2006-030
   Han SJ, 2014, NEURO-ONCOLOGY, V16, P1255, DOI 10.1093/neuonc/nou044
   Hart MG, 2008, COCHRANE DB SYST REV, DOI 10.1002/14651858.CD007415
   Hashizume R, 2014, NAT MED, V20, P1394, DOI 10.1038/nm.3716
   Hassan H, 2017, J NEURO-ONCOL, V135, P13, DOI 10.1007/s11060-017-2546-1
   Ironside S, 2017, CURR TREAT OPTION ON, V18, DOI 10.1007/s11864-017-0508-7
   Izquierdo C, 2018, J NEURO-ONCOL, V136, P533, DOI 10.1007/s11060-017-2677-4
   Kaley T, 2018, J CLIN ONCOL, V36, P3477, DOI 10.1200/JCO.2018.78.9990
   Katayama K, 2013, BRAIN TUMOR PATHOL, V30, P242, DOI 10.1007/s10014-013-0137-1
   Khan RB, 2002, NEURO-ONCOLOGY, V4, P39, DOI 10.1093/neuonc/4.1.39
   Khaw SL, 2007, PEDIATR BLOOD CANCER, V49, P808, DOI 10.1002/pbc.21270
   Kong DS, 2010, NEURO-ONCOLOGY, V12, P289, DOI 10.1093/neuonc/nop030
   Krieger MD, 1997, PEDIATR NEUROSURG, V27, P1, DOI 10.1159/000121218
   Krishnamurthy A, 2018, PHARMACOL THERAPEUT, V185, P122, DOI 10.1016/j.pharmthera.2017.12.002
   Lecavalier-Barsoum M, 2014, COCHRANE DB SYST REV, DOI 10.1002/14651858.CD007104.pub2
   Louis DN, 2016, ACTA NEUROPATHOL, V131, P803, DOI 10.1007/s00401-016-1545-1
   Malmstrom A, 2012, LANCET ONCOL, V13, P916, DOI 10.1016/S1470-2045(12)70265-6
   Massimino M, 2008, J NEURO-ONCOL, V87, P355, DOI 10.1007/s11060-008-9525-5
   Mount CW, 2018, NAT MED, V24, P572, DOI 10.1038/s41591-018-0006-x
   Nabors LB, CENTRAL NERVOUS SYST
   Newlands ES, 1997, CANCER TREAT REV, V23, P35, DOI 10.1016/S0305-7372(97)90019-0
   Norden AD, 2013, NEURO-ONCOLOGY, V15, P930, DOI 10.1093/neuonc/not040
   Omuro A, 2013, NEURO-ONCOLOGY, V15, P242, DOI 10.1093/neuonc/nos295
   Ostrom QT, 2014, NEURO-ONCOLOGY, V16, P1, DOI [10.1093/neuonc/nov189, 10.1093/neuonc/nou327, 10.1093/neuonc/nou223]
   Perez-Larraya JG, 2011, J CLIN ONCOL, V29, P3050, DOI 10.1200/JCO.2011.34.8086
   Perry JR, 2017, NEW ENGL J MED, V376, P1027, DOI 10.1056/NEJMoa1611977
   Perry JR, 2010, J CLIN ONCOL, V28, P2051, DOI 10.1200/JCO.2009.26.5520
   Pianko Matthew J, 2017, Stem Cell Investig, V4, P32, DOI 10.21037/sci.2017.03.04
   Reyes-Botero G, 2014, J NEURO-ONCOL, V116, P405, DOI 10.1007/s11060-013-1312-2
   Reyes-Botero G, 2012, ONCOLOGIST, V17, P388, DOI 10.1634/theoncologist.2011-0335
   Rizzo D, 2015, J CHEMOTHERAPY, V27, P106, DOI 10.1179/1973947814Y.0000000228
   Roskoski R, 2018, PHARMACOL RES, V135, P239, DOI 10.1016/j.phrs.2018.08.013
   Ruda R, 2016, NEURO-ONCOLOGY, V18, P261, DOI 10.1093/neuonc/nov167
   Salmaggi A, 2008, J NEUROL, V255, P171, DOI 10.1007/s00415-008-0589-0
   Seidel C, 2018, STRAHLENTHER ONKOL, V194, P215, DOI 10.1007/s00066-017-1218-6
   Sirachainan N, 2008, NEURO-ONCOLOGY, V10, P577, DOI 10.1215/15228517-2008-025
   Stupp R, 2005, NEW ENGL J MED, V352, P987, DOI 10.1056/NEJMoa043330
   Suzuki Y, 2016, WORLD NEUROSURG, V94, DOI 10.1016/j.wneu.2016.07.110
   Theeler BJ, 2015, J NEUROL SCI, V353, P92, DOI 10.1016/j.jns.2015.04.014
   Thompson EM, 2018, J NEURO-ONCOL, V140, P261, DOI 10.1007/s11060-018-2975-5
   van den Bent MJ, 2018, NEUROL CLIN, V36, P467, DOI 10.1016/j.ncl.2018.04.005
   van den Bent MJ, 2017, LANCET, V390, P1645, DOI 10.1016/S0140-6736(17)31442-3
   van den Bent MJ, 2017, J CLIN ONCOL, V35, P2394, DOI 10.1200/JCO.2017.72.6737
   van Zanten SEMV, 2017, J NEURO-ONCOL, V134, P231, DOI 10.1007/s11060-017-2514-9
   Verhoeff JJC, 2010, ANN ONCOL, V21, P1723, DOI 10.1093/annonc/mdp591
   Warren KE, 2018, FRONT ONCOL, V8, DOI 10.3389/fonc.2018.00239
   Weller M, 2015, CLIN CANCER RES, V21, P2057, DOI 10.1158/1078-0432.CCR-14-2737
   Wick A, 2007, J CLIN ONCOL, V25, P3357, DOI 10.1200/JCO.2007.10.7722
   Wick W, 2012, LANCET ONCOL, V13, P707, DOI 10.1016/S1470-2045(12)70164-X
   Wick W, 2009, J CLIN ONCOL, V27, P5874, DOI 10.1200/JCO.2009.23.6497
   Zhao ZJ, 2018, ACTA PHARM SIN B, V8, P539, DOI 10.1016/j.apsb.2018.03.001
   Zhu M, 2017, CHINESE NEUROSURGICA, V3, P22, DOI [10.1186/s41016-017-0087-2, DOI 10.1186/S41016-017-0087-2]
NR 72
TC 35
Z9 35
U1 3
U2 24
PU SPRINGER
PI NEW YORK
PA ONE NEW YORK PLAZA, SUITE 4600, NEW YORK, NY, UNITED STATES
SN 1523-3790
EI 1534-6269
J9 CURR ONCOL REP
JI Curr. Oncol. Rep.
PD APR
PY 2019
VL 21
IS 4
AR 30
DI 10.1007/s11912-019-0783-5
PG 9
WC Oncology
WE Science Citation Index Expanded (SCI-EXPANDED)
SC Oncology
GA HN8FP
UT WOS:000460430900003
PM 30835007
DA 2023-08-27
ER

PT J
AU HOLMES, B
   SEKHAR, L
   SOFAER, S
   HOLMES, KL
   WRIGHT, DC
AF HOLMES, B
   SEKHAR, L
   SOFAER, S
   HOLMES, KL
   WRIGHT, DC
TI OUTCOMES ANALYSIS IN CRANIAL BASE SURGERY - PRELIMINARY-RESULTS
SO ACTA NEUROCHIRURGICA
LA English
DT Article
DE SKULL BASE; OUTCOMES ANALYSIS; QUALITY OF LIFE; DATABASE
ID HEALTH SURVEY SF-36; 36-ITEM
AB A system of analysis addressing predictors of management outcomes in Cranial Base Surgery has yet to be published. We therefore report data on seventy-nine consecutive patients undergoing surgery for tumors involving thr cranial base, excluding patients with the diagnosis of pituitary microadenoma. Outcomes were defined prospectively in terms of completeness of tumor resection, complications of treatment with emphasis on neurological morbidity, and return tn work or independent living. Also, preoperative features are analyzed as influencing cost of treatment, estimated in terms of the number of surgical procedures required, duration of hospital and Intensive Care Unit stay, and time taken to return to work. Preliminary analysis of data reveals that severe brainstem compression, large tumor size (average diameter > 3 cm), high cavernous sinus grade, and tumor encasement of major cerebral arteries are associated with incomplete tumor resection (p < 0.05). Patient age greater than 65, preoperative Karnofsky Performance Score (KPS) less than 80, and severe brainstem compression are associated with increased risk of stroke (p < 0.05). Age greater than 65 and preoperative KPS less than 80 are associated with an increased length of stay (p < 0.05). Other untoward events did not occur with sufficient frequency to reach statistical significance. A model of outcomes analysis in Cranial Base Surgery is proposed utilizing a database to incorporate a group of non-operated patients and include quality of life measurements in long-term patient follow-up.
C1 GEORGE WASHINGTON UNIV,MED CTR,DEPT SURG,WASHINGTON,DC 20037.
   GEORGE WASHINGTON UNIV,MED CTR,DEPT HLTH CARE SCI,WASHINGTON,DC 20037.
C3 George Washington University; George Washington University
CR HIRSCH WL, 1988, AM J ROENTGENOL, V151, P1015, DOI 10.2214/ajr.151.5.1015
   MCHORNEY CA, 1993, MED CARE, V31, P247, DOI 10.1097/00005650-199303000-00006
   Sekhar Laligam N., 1993, P521
   SEKHAR LN, 1994, J NEUROSURG, V81, P860, DOI 10.3171/jns.1994.81.6.0860
   SPETZLER RF, 1990, J NEUROSURG, V27, P496
   WARE JE, 1992, MED CARE, V30, P473, DOI 10.1097/00005650-199206000-00002
   WINSLOW R, 1992, WALL STREET J   0225, pB1
NR 7
TC 10
Z9 10
U1 0
U2 0
PU SPRINGER-VERLAG WIEN
PI VIENNA
PA SACHSENPLATZ 4-6, PO BOX 89, A-1201 VIENNA, AUSTRIA
SN 0001-6268
J9 ACTA NEUROCHIR
JI Acta Neurochir.
PY 1995
VL 134
IS 3-4
BP 136
EP 138
DI 10.1007/BF01417680
PG 3
WC Clinical Neurology; Surgery
WE Science Citation Index Expanded (SCI-EXPANDED)
SC Neurosciences & Neurology; Surgery
GA RH828
UT WOS:A1995RH82800005
PM 8748772
DA 2023-08-27
ER

PT J
AU Picariello, S
   Cerbone, M
   D'Arco, F
   Gan, HW
   O'Hare, P
   Aquilina, K
   Opocher, E
   Hargrave, D
   Spoudeas, HA
AF Picariello, Stefania
   Cerbone, Manuela
   D'Arco, Felice
   Gan, Hoong-Wei
   O'Hare, Patricia
   Aquilina, Kristian
   Opocher, Enrico
   Hargrave, Darren
   Spoudeas, Helen A.
TI A 40-Year Cohort Study of Evolving Hypothalamic Dysfunction in Infants
   and Young Children (<3 years) with Optic Pathway Gliomas
SO CANCERS
LA English
DT Article
DE optic pathway glioma; endocrine morbidity; infancy; pituitary;
   hypothalamus; diencephalic syndrome
ID LONG-TERM SURVIVORS; DIENCEPHALIC SYNDROME; NEUROFIBROMATOSIS TYPE-1;
   GROWTH-HORMONE; CHILDHOOD CRANIOPHARYNGIOMA; FOLLOW-UP; ENDOCRINE;
   CHEMOTHERAPY; TUMORS; MULTICENTER
AB Simple Summary Serious, poorly understood health issues affect young children with optic pathway tumours. We studied the risk of developing life-limiting hormonal, metabolic, and neurobehavioural disorders by tumour position, recurrence, and treatment, in those diagnosed under 3 years. We found the highest risk for future complex health issues in those presenting with failure to thrive, under one year of age, and/or a tumour involving a deep midbrain area called the hypothalamus. Time, repeated tumour growth, and salvage therapies (radiotherapy, surgery) contributed. We highlight the need for a better understanding of tumour-induced midbrain injury and for neurobehavioural and hormonal support to inform rehabilitation in the growing years, during and beyond cure, to optimise quality of life for these survivors and their families. This might inform oncology treatment strategies and determine new areas for support and collaborative neuroscience research in this high survival group. Despite high survival, paediatric optic pathway hypothalamic gliomas are associated with significant morbidity and late mortality. Those youngest at presentation have the worst outcomes. We aimed to assess presenting disease, tumour location, and treatment factors implicated in the evolution of neuroendocrine, metabolic, and neurobehavioural morbidity in 90 infants/children diagnosed before their third birthday and followed-up for 9.5 years (range 0.5-25.0). A total of 52 (57.8%) patients experienced endo-metabolic dysfunction (EMD), the large majority (46) of whom had hypothalamic involvement (H+) and lower endocrine event-free survival (EEFS) rates. EMD was greatly increased by a diencephalic syndrome presentation (85.2% vs. 46%, p = 0.001)), H+ (OR 6.1 95% CI 1.7-21.7, p 0.005), radiotherapy (OR 16.2, 95% CI 1.7-158.6, p = 0.017) and surgery (OR 4.8 95% CI 1.3-17.2, p = 0.015), all associated with anterior pituitary disorders. Obesity occurred in 25% of cases and was clustered with the endocrinopathies. Neurobehavioural deficits occurred in over half (52) of the cohort and were associated with H+ (OR 2.5 95% C.I. 1.1-5.9, p = 0.043) and radiotherapy (OR 23.1 C.I. 2.9-182, p = 0.003). Very young children with OPHG carry a high risk of endo-metabolic and neurobehavioural comorbidities which deserve better understanding and timely/parallel support from diagnosis to improve outcomes. These evolve in complex, hierarchical patterns over time whose aetiology appears predominantly determined by injury from the hypothalamic tumour location alongside adjuvant treatment strategies.
C1 [Picariello, Stefania] Santobono Pausilipon Childrens Hosp, Dept Paediat Oncol, Neurooncol Unit, I-80123 Naples, Italy.
   [Picariello, Stefania] Univ Campania Luigi Vanvitelli, Dept Woman Child & Gen & Specialized Surg, I-80138 Naples, Italy.
   [Cerbone, Manuela; Gan, Hoong-Wei; Spoudeas, Helen A.] Great Ormond St Hosp Sick Children, Dept Endocrinol, London WC1N 3JH, England.
   [Cerbone, Manuela; Gan, Hoong-Wei; Spoudeas, Helen A.] UCL, Great Ormond St Hosp, Inst Child Hlth, Sect Mol Basis Rare Dis, London WC1N 1EH, England.
   [D'Arco, Felice] Great Ormond St Hosp Sick Children, Dept Radiol, London WC1N 3JH, England.
   [O'Hare, Patricia; Opocher, Enrico; Hargrave, Darren] Great Ormond St Hosp Sick Children, Dept Oncol, London WC1N 3JH, England.
   [Aquilina, Kristian] Great Ormond St Hosp Sick Children, Dept Neurosurg, London WC1N 3JH, England.
   [Opocher, Enrico] Padua Univ Hosp, Pediat Hematol Oncol & Stem Cell Transplant Div, I-35128 Padua, Italy.
C3 Universita della Campania Vanvitelli; University of London; University
   College London; Great Ormond Street Hospital for Children NHS Foundation
   Trust; University of London; University College London; Great Ormond
   Street Hospital for Children NHS Foundation Trust; University of London;
   University College London; Great Ormond Street Hospital for Children NHS
   Foundation Trust; University of London; University College London; Great
   Ormond Street Hospital for Children NHS Foundation Trust; University of
   London; University College London; Great Ormond Street Hospital for
   Children NHS Foundation Trust; University of Padua; Azienda Ospedaliera
   - Universita di Padova
RP Cerbone, M (通讯作者)，Great Ormond St Hosp Sick Children, Dept Endocrinol, London WC1N 3JH, England.; Cerbone, M (通讯作者)，UCL, Great Ormond St Hosp, Inst Child Hlth, Sect Mol Basis Rare Dis, London WC1N 1EH, England.
EM;
   Felice.D';
  ;
  ;
  
RI Hargrave, Darren R/H-1066-2014; Gan, Hoong-Wei/K-8031-2013
OI Hargrave, Darren R/0000-0001-8219-9807; Gan,
   Hoong-Wei/0000-0001-6198-0397; Opocher, Enrico/0000-0001-8483-1618;
   Aquilina, Kristian/0000-0001-7015-5196
FU Great Ormond Street Hospital NHS Foundation Trust; NIHR Great Ormond
   Street Hospital Biomedical Research Centre; NHS; NIHR
FX All of the studies at Great Ormond Street Hospital NHS Foundation Trust
   and UCL Great Ormond Street Institute of Child Health were made possible
   by the NIHR Great Ormond Street Hospital Biomedical Research Centre. The
   views expressed are those of the author(s) and notnecessarily those of
   the NHS, the NIHR, or the UK Department of Health
CR Acharya S, 2019, J NEURO-ONCOL, V144, P603, DOI 10.1007/s11060-019-03264-2
   Ahn Y, 2006, CHILD NERV SYST, V22, P1136, DOI 10.1007/s00381-006-0086-7
   [Anonymous], 2008, STAT REP PRIM BRAIN
   Armstrong GT, 2011, NEURO-ONCOLOGY, V13, P223, DOI 10.1093/neuonc/noq178
   Azizi AA, 2021, NEURO-ONCOLOGY, V23, P100, DOI 10.1093/neuonc/noaa153
   Benesch M, 2006, J NEURO-ONCOL, V78, P199, DOI 10.1007/s11060-005-9091-z
   Binning Mandy J, 2007, Neurosurg Focus, V23, pE2
   BRAUNER R, 1990, EUR J PEDIATR, V149, P825, DOI 10.1007/BF02072067
   Brauner R, 2006, J CLIN ENDOCR METAB, V91, P2467, DOI 10.1210/jc.2006-0322
   Cappelli C, 1998, ARCH DIS CHILD, V79, P334, DOI 10.1136/adc.79.4.334
   CollettSolberg PF, 1997, CLIN ENDOCRINOL, V47, P79, DOI 10.1046/j.1365-2265.1997.2211032.x
   Darendeliler F, 2006, ACTA PAEDIATR, V95, P1284, DOI 10.1080/08035250600577889
   de Blank PMK, 2017, J NEURO-OPHTHALMOL, V37, pS23, DOI 10.1097/WNO.0000000000000550
   DeVile CJ, 1996, ARCH DIS CHILD, V75, P108, DOI 10.1136/adc.75.2.108
   Dodgshun AJ, 2015, CANCER-AM CANCER SOC, V121, P4190, DOI 10.1002/cncr.29649
   Falzon K, 2018, BRIT J OPHTHALMOL, V102, P1367, DOI 10.1136/bjophthalmol-2017-311305
   Gan HW, 2015, J CLIN ENDOCR METAB, V100, P3787, DOI 10.1210/jc.2015-2028
   Gnekow AK, 2019, KLIN PADIATR, V231, P107, DOI 10.1055/a-0889-8256
   Hill CS, 2021, CHILD NERV SYST, V37, P1917, DOI 10.1007/s00381-021-05060-8
   Hoffmann A, 2014, J CLIN ENDOCR METAB, V99, P3972, DOI 10.1210/jc.2014-1680
   Huguenin M, 2003, J PEDIATR-US, V142, P699, DOI 10.1067/mpd.2003.234
   Kilday JP, 2014, J NEURO-ONCOL, V116, P195, DOI 10.1007/s11060-013-1284-2
   Liu APY, 2019, CANCER-AM CANCER SOC, V125, P1163, DOI 10.1002/cncr.31918
   Merchant TE, 2009, J CLIN ONCOL, V27, P3691, DOI 10.1200/JCO.2008.21.2738
   Mishra MV, 2012, J NEURO-ONCOL, V107, P591, DOI 10.1007/s11060-011-0783-2
   Muller HL, 2010, HORM RES PAEDIAT, V73, P175, DOI 10.1159/000284358
   Packer RJ, 2001, J CLIN ONCOL, V19, P480, DOI 10.1200/JCO.2001.19.2.480
   Papini C, 2020, SCI REP-UK, V10, DOI 10.1038/s41598-020-59896-2
   Poussaint TY, 1997, AM J NEURORADIOL, V18, P1499
   Pritchard-Jones K., 2007, BRIT J CANCER, V96, P1927, DOI [10.1038/sj.bjc.6603800, DOI 10.1038/SJ.BJC.6603800]
   Rakotonjanahary J, 2015, PLOS ONE, V10, DOI 10.1371/journal.pone.0127676
   Ris MD, 2019, CANCER-AM CANCER SOC, V125, P3050, DOI 10.1002/cncr.32186
   RODRIGUEZ LA, 1990, NEUROSURGERY, V26, P242, DOI 10.1227/00006123-199002000-00010
   Sani I, 2017, HORM RES PAEDIAT, V87, P179, DOI 10.1159/000458525
   Santoro C, 2020, J CLIN ENDOCR METAB, V105, pE2214, DOI 10.1210/clinem/dgaa138
   Shofty B, 2020, CHILD NERV SYST, V36, P2351, DOI 10.1007/s00381-020-04697-1
   Silva MM, 2000, PEDIATR NEUROSURG, V33, P151, DOI 10.1159/000028996
   Sklar CA, 2018, J CLIN ENDOCR METAB, V103, P2761, DOI 10.1210/jc.2018-01175
   Stokland T, 2010, NEURO-ONCOLOGY, V12, P1257, DOI 10.1093/neuonc/noq092
   Swerdlow AJ, 2017, J CLIN ENDOCR METAB, V102, P1661, DOI 10.1210/jc.2016-2046
   Taylor T, 2008, BRIT J RADIOL, V81, P761, DOI 10.1259/bjr/65246351
NR 41
TC 6
Z9 6
U1 0
U2 0
PU MDPI
PI BASEL
PA ST ALBAN-ANLAGE 66, CH-4052 BASEL, SWITZERLAND
EI 2072-6694
J9 CANCERS
JI Cancers
PD FEB
PY 2022
VL 14
IS 3
AR 747
DI 10.3390/cancers14030747
PG 16
WC Oncology
WE Science Citation Index Expanded (SCI-EXPANDED)
SC Oncology
GA YZ3WV
UT WOS:000755410300001
PM 35159015
OA gold, Green Published
DA 2023-08-27
ER

PT J
AU Kurwale, NS
   Suri, V
   Suri, A
   Sarkar, C
   Gupta, DK
   Sharma, BS
   Mahapatra, AK
AF Kurwale, Nilesh S.
   Suri, Vaishali
   Suri, Ashish
   Sarkar, Chitra
   Gupta, Deepak Kumar
   Sharma, Bhavani Shankar
   Mahapatra, Ashok Kumar
TI Predictive factors for early symptomatic recurrence in pilocytic
   astrocytoma: Does angiogenesis have a role to play?
SO JOURNAL OF CLINICAL NEUROSCIENCE
LA English
DT Article
DE Cerebellar astrocytoma; MIB-I; Pilocytic astrocytoma; Vascular
   endothelial growth factor
ID ENDOTHELIAL GROWTH-FACTOR; CEREBELLAR ASTROCYTOMAS; LONG-TERM; HUMAN
   GLIOMAS; CHILDREN; RESECTION
AB We studied predictive factors with respect to angiogenesis and proliferative indices for early symptomatic recurrences in patients with pilocytic astrocytoma (PA). One hundred and eighteen patients who underwent surgery for PA were divided into non-recurrent and early symptomatic recurrence groups to analyze clinicoradiological and immunohistopathological (n = 33) parameters. Patients with non-recurrent tumors presented with synptoms for a mean duration of 10.2 +/- 9.1 months while those with recurrent tumors presented slightly earlier (6.9 +/- 4.5 months). Common tumor locations were the cerebellum (38.1%), optic chiasm (27.9%), supratentorial region (19.4%) and brainstem (9.3%). Recurrent tumors were mostly located in the cerebellum (44%) and brainstem (33%). Strong contrast enhancement was noted in 70 (59.3%) tumors, while 48 (40.7%) showed poor contrast. Resection was complete in 53% of patients while near total excision was achieved for the remaining patients. Cellularity and plemorphism were similar in both groups. Extensive endothelial proliferation was observed in 18.1% of patients while the remainder showed a focal pattern. Diffuse vascular endothelial growth factor (VEGF) expression was observed in 36.3% of patients while 63.6% showed mild-to-moderate focal expression. Endothelial proliferation and VEGF expression were more pronounced in patients with non-recurrent tumors, but this was not statistically significant. MIB-I labeling indices were similar (1-5%) for both groups. Symptomatic recurrences were common in infratentorial PAs. Radiology, histopathology and proliferative indices did not offer any prognostic information. Angiogenesis markers such as endothelial proliferation and VEFG expression did not predict early symptomatic recurrence. Diffuse VEGF expression and endothelial proliferation were observed in tumors that showed strong contrast enhancement. (c) 2011 Published by Elsevier Ltd.
C1 [Kurwale, Nilesh S.; Suri, Vaishali; Suri, Ashish; Sarkar, Chitra; Gupta, Deepak Kumar; Sharma, Bhavani Shankar; Mahapatra, Ashok Kumar] All India Inst Med Sci, Cardio Neuro Ctr, New Delhi 110029, India.
C3 All India Institute of Medical Sciences (AIIMS) New Delhi
RP Suri, A (通讯作者)，All India Inst Med Sci, Cardio Neuro Ctr, Room 712,7th Floor, New Delhi 110029, India.
EM
OI Kurwale, Nilesh/0000-0002-7088-7167
CR AUSTIN EJ, 1988, J NEUROSURG, V68, P41, DOI 10.3171/jns.1988.68.1.0041
   Bartels U, 2006, J NEUROSURG, V104, P314, DOI 10.3171/ped.2006.104.5.314
   Benesch M, 2006, PEDIATR NEUROSURG, V42, P159, DOI 10.1159/000091859
   Bowers DC, 2003, J CLIN ONCOL, V21, P2968, DOI 10.1200/JCO.2003.01.017
   Burger PC, 2002, SURG PATHOLOGY NERVO, P203
   Burger PC, 2000, PATHOLOGY GENETICS T, P45
   DAVIS CH, 1981, J NEUROL NEUROSUR PS, V44, P820, DOI 10.1136/jnnp.44.9.820
   Dirven CMF, 1997, CHILD NERV SYST, V13, P17, DOI 10.1007/s003810050033
   GEISSINGER JD, 1971, ARCH NEUROL-CHICAGO, V24, P125, DOI 10.1001/archneur.1971.00480320053005
   Holmes K, 2007, CELL SIGNAL, V19, P2003, DOI 10.1016/j.cellsig.2007.05.013
   Koeller KK, 2004, RADIOGRAPHICS, V24, P1693, DOI 10.1148/rg.246045146
   Komotar Ricardo J, 2005, Neurosurg Focus, V18, pE7
   Krieger MD, 1997, PEDIATR NEUROSURG, V27, P1, DOI 10.1159/000121218
   Leung SY, 1997, AM J SURG PATHOL, V21, P941, DOI 10.1097/00000478-199708000-00009
   Palma L, 2004, ACTA NEUROCHIR, V146, P581, DOI 10.1007/s00701-004-0257-9
   Pencalet P, 1999, J NEUROSURG, V90, P265, DOI 10.3171/jns.1999.90.2.0265
   PLATE KH, 1992, NATURE, V359, P845, DOI 10.1038/359845a0
   Schmidt NO, 1999, INT J CANCER, V84, P10, DOI 10.1002/(SICI)1097-0215(19990219)84:1<10::AID-IJC3>3.0.CO;2-L
   Tibbetts KM, 2009, ACTA NEUROPATHOL, V117, P657, DOI 10.1007/s00401-009-0506-3
NR 19
TC 7
Z9 7
U1 0
U2 1
PU ELSEVIER SCI LTD
PI OXFORD
PA THE BOULEVARD, LANGFORD LANE, KIDLINGTON, OXFORD OX5 1GB, OXON, ENGLAND
SN 0967-5868
EI 1532-2653
J9 J CLIN NEUROSCI
JI J. Clin. Neurosci.
PD APR
PY 2011
VL 18
IS 4
BP 472
EP 477
DI 10.1016/j.jocn.2010.04.055
PG 6
WC Clinical Neurology; Neurosciences
WE Science Citation Index Expanded (SCI-EXPANDED)
SC Neurosciences & Neurology
GA 747MY
UT WOS:000289325300005
PM 21292490
DA 2023-08-27
ER

PT J
AU Indelicato, DJ
   Rotondo, RL
   Uezono, H
   Sandler, ES
   Aldana, PR
   Ranalli, NJ
   Beier, AD
   Morris, CG
   Bradley, JA
AF Indelicato, Daniel J.
   Rotondo, Ronny L.
   Uezono, Haruka
   Sandler, Eric S.
   Aldana, Philipp R.
   Ranalli, Nathan J.
   Beier, Alexandra D.
   Morris, Christopher G.
   Bradley, Julie A.
TI Outcomes Following Proton Therapy for Pediatric Low-Grade Glioma
SO INTERNATIONAL JOURNAL OF RADIATION ONCOLOGY BIOLOGY PHYSICS
LA English
DT Article; Proceedings Paper
CT Annual Meeting of the Paediatric-Radiation-Oncology-Society
CY NOV, 2018
CL Kyoto, JAPAN
SP Paediat Radiat Oncol Soc
ID OPTIC PATHWAY GLIOMAS; RADIATION-THERAPY; VISUAL OUTCOMES; RADIOTHERAPY;
   CHILDREN; TRIAL; PSEUDOPROGRESSION; VINCRISTINE; CARBOPLATIN
AB Purpose: Dosimetric studies show that proton therapy can reduce the low/intermediate radiation dose to uninvolved tissue in children with low-grade glioma (LGG). For this reason, LGG is the fourth most common pediatric tumor treated with proton therapy, yet clinical outcome data on efficacy and toxicity are limited.
   Methods and Materials: We reviewed the medical records of 174 children (<= 21 years old) with nonmetastatic LGG enrolled on a prospective protocol and treated with proton therapy between 2007 and 2017. We assessed clinical outcomes and toxicity and analyzed patient, tumor, and treatment-related variables.
   Results: The median age was 10.2 years (range, 2-21). Fifty-eight percent of tumors were World Health Organization grade 1 and 30% were grade 2; 12% were diagnosed on imaging characteristics alone. The most common histology was pilocytic astrocytoma (47%). The most common tumor subsites were diencephalon/optic pathway (52%), caudal brainstem (16%), and cerebellum (13%). Forty-two percent received chemotherapy before radiation therapy. The median follow-up was 4.4 years. The 5year actuarial rates of local control, progression-free survival, and overall survival were 85% (95% confidence interval [CI], 78%-90%), 84% (95% CI, 77%-89%), and 92% (95% CI, 85%-95%), respectively. On univariate analysis, brainstem/spinal cord tumor location (62% vs 90% elsewhere) and dose < 54 GyRBE (67% vs 91% for 54 GyRBE) were associated with inferior local control (P <.01 for both). Twenty-two patients (12.6%) experienced acute nausea or vomiting requiring ondansetron; 2 patients (1.1%) required corticosteroids. Serious toxicities (4% of patients) included brainstem necrosis requiring corticosteroids (n = 2), symptomatic vasculopathy (n = 2), radiation retinopathy (n = 1), epilepsy (n = 1), and death from radiation-induced high-grade glioma (n = 1). Thirty-nine patients (22%) developed new-onset central hormone deficiency. Pseudoprogression was observed in 32.1%. Conclusions: Compared with modern photon series, proton therapy reduces the radiation dose to developing brain tissue, diminishing acute toxicities without compromising disease control. (C) 2019 The Authors. Published by Elsevier Inc. This is an open access article under the CC BY-NC-ND license (http://creativecommons. org/ licenses/by-nc-nd/4.0/).
C1 [Indelicato, Daniel J.; Rotondo, Ronny L.; Uezono, Haruka; Morris, Christopher G.; Bradley, Julie A.] Univ Florida, Coll Med, Dept Radiat Oncol, 2015 North Jefferson St, Jacksonville, FL 32206 USA.
   [Sandler, Eric S.] Nemours Childrens Hlth Syst, Jacksonville, FL USA.
   [Aldana, Philipp R.; Ranalli, Nathan J.; Beier, Alexandra D.] Univ Florida, Coll Med, Dept Neurosurg, Jacksonville, FL USA.
C3 State University System of Florida; University of Florida; State
   University System of Florida; University of Florida
RP Indelicato, DJ (通讯作者)，Univ Florida, Coll Med, Dept Radiat Oncol, 2015 North Jefferson St, Jacksonville, FL 32206 USA.
EM
RI Indelicato, Daniel/AAL-4761-2020; Sandler, Eric/AAE-8656-2019
OI Indelicato, Daniel/0000-0001-5765-1873; sandler,
   eric/0000-0002-0083-9370; Bradley, Julie/0000-0001-7581-4605
CR Amelio D, 2013, INT J RADIAT ONCOL, V87, pS253, DOI 10.1016/j.ijrobp.2013.06.658
   Awdeh RM, 2012, INT J RADIAT ONCOL, V84, P46, DOI 10.1016/j.ijrobp.2011.10.067
   Brown AP, 2013, INT J RADIAT ONCOL, V86, P277, DOI 10.1016/j.ijrobp.2013.01.014
   Cappellano AM, 2015, J NEURO-ONCOL, V121, P405, DOI 10.1007/s11060-014-1652-6
   Cherlow JM, 2019, INT J RADIAT ONCOL, V103, P861, DOI 10.1016/j.ijrobp.2018.11.004
   Chong AL, 2013, NEURO-ONCOLOGY, V15, P391, DOI 10.1093/neuonc/nos312
   Eaton BR, 2014, CANCER J, V20, P403, DOI 10.1097/PPO.0000000000000079
   Gnekow AK, 2017, EUR J CANCER, V81, P206, DOI 10.1016/j.ejca.2017.04.019
   Gnekow AK, 2012, NEURO-ONCOLOGY, V14, P1265, DOI 10.1093/neuonc/nos202
   Greenberger BA, 2014, INT J RADIAT ONCOL, V89, P1060, DOI 10.1016/j.ijrobp.2014.04.053
   Haas-Kogan D, 2018, INT J RADIAT ONCOL, V101, P152, DOI 10.1016/j.ijrobp.2018.01.013
   Hall MD, 2018, INT J RADIAT ONCOL, V101, P854, DOI 10.1016/j.ijrobp.2018.03.027
   Hug EB, 2002, STRAHLENTHER ONKOL, V178, P10, DOI 10.1007/s00066-002-0874-2
   Indelicato DJ, 2017, PEDIATR BLOOD CANCER, V64, DOI 10.1002/pbc.26654
   Journy N, 2018, C RAD HLTH 2018 SEPT
   Karim ABMF, 1996, INT J RADIAT ONCOL, V36, P549, DOI 10.1016/S0360-3016(96)00352-5
   Lassaletta A, 2017, J CLIN ONCOL, V35, P2934, DOI 10.1200/JCO.2016.71.8726
   Lassen-Ramshad Y, 2015, ACTA ONCOL, V54, P1701, DOI 10.3109/0284186X.2015.1078498
   Mannina Edward M, 2016, Int J Part Ther, V3, P291, DOI 10.14338/IJPT-16-00020.1
   Mansur DB, 2011, INT J RADIAT ONCOL, V79, P829, DOI 10.1016/j.ijrobp.2009.11.015
   Marcus KJ, 2005, INT J RADIAT ONCOL, V61, P374, DOI 10.1016/j.ijrobp.2004.06.012
   Merchant TE, 2009, J CLIN ONCOL, V27, P3598, DOI 10.1200/JCO.2008.20.9494
   Huynh-Le MP, 2016, CHILD NERV SYST, V32, P1425, DOI 10.1007/s00381-016-3100-8
   Mishra KK, 2006, NEURO-ONCOLOGY, V8, P166, DOI 10.1215/15228517-2005-011
   PACKER RJ, 1993, J CLIN ONCOL, V11, P850, DOI 10.1200/JCO.1993.11.5.850
   Packer Roger J, 2010, Curr Neurol Neurosci Rep, V10, P10, DOI 10.1007/s11910-009-0084-8
   Perilongo G, 2005, J NEURO-ONCOL, V75, P301, DOI 10.1007/s11060-005-6754-8
   Plimpton SR, 2015, PEDIATR HEMAT ONCOL, V32, P78, DOI 10.3109/08880018.2013.791738
   Tsang DS, 2017, J NEURO-ONCOL, V135, P371, DOI 10.1007/s11060-017-2583-9
   Tsang DS, 2017, INT J RADIAT ONCOL, V99, P642, DOI 10.1016/j.ijrobp.2017.07.023
   Wan MJ, 2016, J NEURO-ONCOL, V129, P173, DOI 10.1007/s11060-016-2163-4
   Williams NL, 2018, AM J CLIN ONCOL-CANC, V41, P307, DOI 10.1097/COC.0000000000000267
NR 32
TC 57
Z9 59
U1 0
U2 6
PU ELSEVIER SCIENCE INC
PI NEW YORK
PA STE 800, 230 PARK AVE, NEW YORK, NY 10169 USA
SN 0360-3016
EI 1879-355X
J9 INT J RADIAT ONCOL
JI Int. J. Radiat. Oncol. Biol. Phys.
PD MAY 1
PY 2019
VL 104
IS 1
BP 149
EP 156
DI 10.1016/j.ijrobp.2019.01.078
PG 8
WC Oncology; Radiology, Nuclear Medicine & Medical Imaging
WE Science Citation Index Expanded (SCI-EXPANDED); Conference Proceedings Citation Index - Science (CPCI-S)
SC Oncology; Radiology, Nuclear Medicine & Medical Imaging
GA HS4KC
UT WOS:000463830100022
PM 30684665
OA hybrid
DA 2023-08-27
ER

PT J
AU Shiue, K
   Barnett, GH
   Suh, JH
   Vogelbaum, MA
   Reddy, CA
   Weil, RJ
   Angelov, L
   Neyman, G
   Chao, ST
AF Shiue, Kevin
   Barnett, Gene H.
   Suh, John H.
   Vogelbaum, Michael A.
   Reddy, Chandana A.
   Weil, Robert J.
   Angelov, Lilyana
   Neyman, Gennady
   Chao, Samuel T.
TI Using Higher Isodose Lines for Gamma Knife Treatment of 1 to 3 Brain
   Metastases Is Safe and Effective
SO NEUROSURGERY
LA English
DT Article
DE Brain metastases; Isodose line; Radiation necrosis; Radiosurgery
ID STEREOTACTIC RADIOSURGERY; MANAGEMENT
AB BACKGROUND: Higher isodose lines (IDLs) in Gamma Knife (GK) Perfexion treatment of brain metastases (BMet) could result in lower local control (LC) or higher radiation necrosis (RN) rates, but reduce treatment time.
   OBJECTIVE: To assess the impact of the heterogeneity index (HI) and conformality index (CFI) ion local failure (LF) for patients treated with GK for 1 to 3 BMet.
   METHODS: From an institutional review board-approved database, 320 patients with 496 BMet were identified, treated for 1 to 3 BMet from July 2007 to April 2011 on GK Perfexion. Cox proportional hazards regression was used to analyze significance of HI, CFI, IDL, dose, tumor diameter, recursive partitioning analysis class, tumor radioresistance, primary, smoking history, metastasis location, and whole-brain radiation therapy (WBRT) history with LF and RN.
   RESULTS: Median follow-up by lesion was 6.8 months (range, 0-49.6). The series median survival was 14.2 months. Per RECIST, 9.5% of lesions failed, 33.9% were stable, 38.3% partially responded, 17.1% responded completely, and 1.2% could not be assessed. The 12-month LC rate was 87.3%. On univariate analysis, a dose less than 20 Gy (hazard ratio [HR]: 2.940, P < .001); tumor size (HR: 1.674, P < .001); and cerebellum/brainstem location vs other (HR: 1.891, P = .043) were significant for LF. Non-small cell lung cancer (HR: 0.333, P = .0097) was associated with better LC. On multivariate analysis, tumor size (HR: 1.696, P < .001) and cerebellum/brainstem location vs other (HR: 1.959, P = .033) remained significant for LF. Variables not significant for LF included CI, IDL, and HI.
   CONCLUSION: Our study of patients with 1 to 3 BMet treated with GK demonstrated no difference in LC or RN with varying HI, indicating that physicians can treat to IDL at 70% or higher IDL to reduce treatment time without increased LF or RN.
C1 [Shiue, Kevin; Vogelbaum, Michael A.] Case Western Reserve Univ, Sch Med, Cleveland, OH USA.
   [Barnett, Gene H.; Suh, John H.; Vogelbaum, Michael A.; Weil, Robert J.; Angelov, Lilyana; Chao, Samuel T.] Cleveland Clin, Rose Ella Burkhardt Brain Tumor & Neurooncol Ctr, Cleveland, OH 44195 USA.
   [Barnett, Gene H.; Weil, Robert J.; Angelov, Lilyana] Cleveland Clin, Neurol Inst, Dept Neurosurg, Cleveland, OH 44195 USA.
   [Suh, John H.; Reddy, Chandana A.; Neyman, Gennady; Chao, Samuel T.] Cleveland Clin, Taussig Canc Inst, Dept Radiat Oncol, Cleveland, OH 44195 USA.
C3 Case Western Reserve University; Cleveland Clinic Foundation; Cleveland
   Clinic Foundation; Cleveland Clinic Foundation
RP Chao, ST (通讯作者)，Cleveland Clin, Lerner Coll Med, Dept Radiat Oncol, Desk T28,9500 Euclid Ave, Cleveland, OH 44195 USA.
EM
RI Shiue, Kevin/IZP-5505-2023; suh, */HRA-0278-2023
OI Shiue, Kevin/0000-0001-6125-9241; Reddy, Chandana/0000-0002-3832-8744
FU US Department of Defense Breast Cancer Research Program [W81XWH-062];
   American Cancer Society Joseph and Jeanette Silber Student Research
   Fellowship
FX Kevin Shiue was awarded a 2011 American Cancer Society Joseph and
   Jeanette Silber Student Research Fellowship to support his participation
   in this study. Dr Barnett has the following associations with Monteris
   Medical: consultancy, payment for lectures and manuscript preparation
   (not this manuscript) and development of educational presentations, and
   stock options. Dr Vogelbaum is a consultant for Merck, Pharmicokinesis,
   and Neuralstem and holds a patent with Infuseon Therapeutics. Dr Weil
   was supported in part by grant W81XWH-062 to 0033 from the US Department
   of Defense Breast Cancer Research Program. Dr Angelov has received
   payment for serving on the speakers' bureau of BrainLab. Dr Neyman is a
   consultant for Elekta AB and received payment for lectures for Elekta
   AB. The other authors have no personal financial or institutional
   interest in any of the drugs, materials, or devices described in this
   article.
CR Balagamwala EH, 2012, INT J RADIAT ONCOL, V83, P1406, DOI 10.1016/j.ijrobp.2011.10.024
   Chao ST, 2008, CANCER, V113, P2198, DOI 10.1002/cncr.23821
   COX DR, 1972, J R STAT SOC B, V34, P187
   Eichler AF, 2011, NAT REV CLIN ONCOL, V8, P344, DOI 10.1038/nrclinonc.2011.58
   Grandhi R, 2012, J NEUROSURG, V117, P237, DOI 10.3171/2012.4.JNS11870
   Hunter GK, 2012, INT J RADIAT ONCOL, V83, P1394, DOI 10.1016/j.ijrobp.2011.10.026
   KAPLAN EL, 1958, J AM STAT ASSOC, V53, P457, DOI 10.2307/2281868
   Koyfman SA, 2010, INT J RADIAT ONCOL, V78, P409, DOI 10.1016/j.ijrobp.2009.07.1750
   Lwu S, 2013, ONCOL REP, V29, P407, DOI 10.3892/or.2012.2139
   Rades D, 2009, EUR J CANCER, V45, P400, DOI 10.1016/j.ejca.2008.10.033
   Shaw E, 2000, INT J RADIAT ONCOL, V47, P291, DOI 10.1016/S0360-3016(99)00507-6
   Soltys SG, 2008, INT J RADIAT ONCOL, V70, P187, DOI 10.1016/j.ijrobp.2007.06.068
   Suh JH, 2010, NEW ENGL J MED, V362, P1119, DOI 10.1056/NEJMct0806951
   Vogelbaum MA, 2006, J NEUROSURG, V104, P907, DOI 10.3171/jns.2006.104.6.907
   Xu ZY, 2013, WORLD NEUROSURG, V80, P351, DOI 10.1016/j.wneu.2012.03.021
NR 15
TC 13
Z9 13
U1 0
U2 3
PU OXFORD UNIV PRESS INC
PI CARY
PA JOURNALS DEPT, 2001 EVANS RD, CARY, NC 27513 USA
SN 0148-396X
EI 1524-4040
J9 NEUROSURGERY
JI Neurosurgery
PD APR
PY 2014
VL 74
IS 4
BP 360
EP 364
DI 10.1227/NEU.0000000000000289
PG 5
WC Clinical Neurology; Surgery
WE Science Citation Index Expanded (SCI-EXPANDED)
SC Neurosciences & Neurology; Surgery
GA AJ5KW
UT WOS:000337722100021
PM 24448185
DA 2023-08-27
ER

PT J
AU Eder, U
   Fischer-Colbrie, R
   Kogner, P
   Leitner, B
   Bjellerup, P
   Winkler, H
AF Eder, U
   Fischer-Colbrie, R
   Kogner, P
   Leitner, B
   Bjellerup, P
   Winkler, H
TI Levels and molecular forms of chromogranins in human childhood
   neuroblastomas and ganglioneuromas
SO NEUROSCIENCE LETTERS
LA English
DT Article
DE chromagranin A; secretogranin II; tumor; neuropeptide; proteolytic
   processing
ID SECRETOGRANIN-II; NEUROENDOCRINE CELLS; CHROMAFFIN GRANULES;
   ADRENAL-MEDULLA; DIAGNOSIS; PHEOCHROMOCYTOMA; SECRETONEURIN; TISSUES;
   BIOSYNTHESIS; SENSITIVITY
AB The chromogranins are a class of acidic proteins found in large secretory granules of neuroendocrine tissues and tumors derived from them. We measured the relative amounts and characterized the molecular forms of two members of this family, i.e. chromogranin A and secretogranin II, in 14 neuroblastomas and five ganglioneuromas. In all the tumors investigated significant amounts of chromogranin A and secretogranin II were found. Neuroblastomas contained two times and ganglioneuromas 45 times more secretogranin II compared to chromogranin A. Both proteins were processed in these tumors to a great extent to smaller peptides, only limited amounts of intact chromogranin A or secretogranin II were present. In general, proteolytic processing of secretogranin II to the small neuropeptide secretoneurin was more complete than that of chromogranin A to the peptide GE-25. Proteolytic processing of both chromogranins as well as the total amounts of these proteins were unrelated to tumor staging. (C) 1998 Elsevier Science Ireland Ltd. All rights reserved.
C1 Univ Innsbruck, Dept Pharmacol, A-6020 Innsbruck, Austria.
   Dept Clin Chem, Stockholm, Sweden.
   Karolinska Hosp, S-10401 Stockholm, Sweden.
C3 University of Innsbruck; Karolinska Institutet; Karolinska University
   Hospital
RP Fischer-Colbrie, R (通讯作者)，Univ Innsbruck, Dept Pharmacol, Peter Mayr Str 1A, A-6020 Innsbruck, Austria.
EM
OI Kogner, Per/0000-0002-2202-9694
CR ABRAHAMSSON PA, 1993, J ANDROL, V14, P307
   BOOMSMA F, 1995, CLIN CHIM ACTA, V239, P57, DOI 10.1016/0009-8981(95)06100-R
   BRODEUR GM, 1993, J CLIN ONCOL, V11, P1466, DOI 10.1200/JCO.1993.11.8.1466
   DEFTOS LJ, 1991, ENDOCR REV, V12, P181, DOI 10.1210/edrv-12-2-181
   DISANTAGNESE PA, 1992, CANCER-AM CANCER SOC, V70, P254, DOI 10.1002/1097-0142(19920701)70:1+<254::AID-CNCR2820701312>3.0.CO;2-E
   FISCHERCOLBRIE R, 1995, PROG NEUROBIOL, V46, P49, DOI 10.1016/0301-0082(94)00060-U
   HAGN C, 1986, NEUROSCI LETT, V67, P295, DOI 10.1016/0304-3940(86)90325-3
   HSIAO RJ, 1991, MEDICINE, V70, P33, DOI 10.1097/00005792-199101000-00003
   HSIAO RJ, 1990, J CLIN INVEST, V85, P1555, DOI 10.1172/JCI114604
   KIRCHMAIR R, 1995, BIOCHEM J, V310, P331, DOI 10.1042/bj3100331
   KIRCHMAIR R, 1993, NEUROSCIENCE, V53, P359, DOI 10.1016/0306-4522(93)90200-Y
   KOGNER P, 1995, EUR J CANCER, V31A, P557, DOI 10.1016/0959-8049(95)00057-P
   LACHMUND A, 1994, NEUROSCIENCE, V62, P361, DOI 10.1016/0306-4522(94)90371-9
   LASLOP A, 1992, NEUROSCIENCE, V49, P443, DOI 10.1016/0306-4522(92)90109-F
   Leitner B, 1996, J NEUROCHEM, V66, P1312
   Pagani A, 1992, Diagn Mol Pathol, V1, P16, DOI 10.1097/00019606-199203000-00003
   ROSA P, 1994, J ENDOCRINOL INVEST, V17, P207, DOI 10.1007/BF03347721
   SCHMID KW, 1993, HISTOPATHOLOGY, V22, P527, DOI 10.1111/j.1365-2559.1993.tb00172.x
   SCHMID KW, 1994, MODERN PATHOL, V7, P347
   SCHOBER M, 1987, LAB INVEST, V57, P385
   WEILER R, 1990, FEBS LETT, V265, P27, DOI 10.1016/0014-5793(90)80875-J
   WEILER R, 1990, BRAIN RES, V532, P87, DOI 10.1016/0006-8993(90)91746-4
   WINKLER H, 1992, NEUROSCIENCE, V49, P497, DOI 10.1016/0306-4522(92)90222-N
NR 23
TC 18
Z9 18
U1 0
U2 1
PU ELSEVIER IRELAND LTD
PI CLARE
PA ELSEVIER HOUSE, BROOKVALE PLAZA, EAST PARK SHANNON, CO, CLARE, 00000,
   IRELAND
SN 0304-3940
J9 NEUROSCI LETT
JI Neurosci. Lett.
PD AUG 28
PY 1998
VL 253
IS 1
BP 17
EP 20
DI 10.1016/S0304-3940(98)00588-6
PG 4
WC Neurosciences
WE Science Citation Index Expanded (SCI-EXPANDED)
SC Neurosciences & Neurology
GA 118KB
UT WOS:000075836200005
PM 9754794
DA 2023-08-27
ER

PT J
AU Stenman, A
   Zedenius, J
   Juhlin, CC
AF Stenman, Adam
   Zedenius, Jan
   Juhlin, Carl Christofer
TI The Value of Histological Algorithms to Predict the Malignancy Potential
   of Pheochromocytomas and Abdominal Paragangliomas-A Meta-Analysis and
   Systematic Review of the Literature
SO CANCERS
LA English
DT Review
DE PASS; GAPP; histology; meta-analysis; paraganglioma; pheochromocytoma
ID GLAND SCALED SCORE; PASS; EXPRESSION; PARAMETERS; TUMOR
AB Pheochromocytomas (PCCs) and abdominal paragangliomas (PGLs), collectively abbreviated PPGLs, are neuroendocrine tumors of the adrenal medulla and paraganglia, respectively. These tumors exhibit malignant potential but seldom display evidence of metastatic spread, the latter being the only widely accepted evidence of malignancy. To counter this, pre-defined histological algorithms have been suggested to stratify the risk of malignancy: Pheochromocytoma of the Adrenal Gland Scaled Score (PASS) and the Grading system for Adrenal Pheochromocytoma and Paraganglioma (GAPP). The PASS algorithm was originally intended for PCCs whereas the GAPP model is proposed for stratification of both PCCs and PGLs. In parallel, advances in terms of coupling overtly malignant PPGLs to the underlying molecular genetics have been made, but there is yet no combined risk stratification model based on histology and the overall mutational profile of the tumor. In this review, we systematically meta-analyzed previously reported cohorts using the PASS and GAPP algorithms and acknowledge a "rule-out way of approaching these stratification models rather than a classical "rule-in" strategy. Moreover, the current genetic panorama regarding possible molecular adjunct markers for PPGL malignancy is reviewed. A combined histological and genetic approach will be needed to fully elucidate the malignant potential of these tumors.
C1 [Stenman, Adam; Juhlin, Carl Christofer] Karolinska Inst, Dept Oncol Pathol, S-17176 Stockholm, Sweden.
   [Stenman, Adam; Zedenius, Jan] Karolinska Inst, Dept Mol Med & Surg, S-17176 Stockholm, Sweden.
   [Stenman, Adam; Zedenius, Jan] Karolinska Univ Hosp, Dept Breast Endocrine Tumours & Sarcoma, S-17176 Stockholm, Sweden.
   [Juhlin, Carl Christofer] Karolinska Univ Hosp, Dept Pathol & Cytol, S-17176 Stockholm, Sweden.
C3 Karolinska Institutet; Karolinska Institutet; Karolinska Institutet;
   Karolinska University Hospital; Karolinska Institutet; Karolinska
   University Hospital
RP Juhlin, CC (通讯作者)，Karolinska Inst, Dept Oncol Pathol, S-17176 Stockholm, Sweden.; Juhlin, CC (通讯作者)，Karolinska Univ Hosp, Dept Pathol & Cytol, S-17176 Stockholm, Sweden.
EM
RI Juhlin, Carl Christofer/C-1125-2016; Stenman, Adam/C-6131-2015
OI Juhlin, Carl Christofer/0000-0002-5945-9081; Stenman,
   Adam/0000-0003-3248-7629
FU Swedish Cancer Society; Swedish Society for Medical Research
FX This research was funded by grants generously provided by the Swedish
   Cancer Society and the Swedish Society for Medical Research.
CR Agarwal A, 2010, WORLD J SURG, V34, P3022, DOI 10.1007/s00268-010-0744-5
   Aggeli C, 2017, HORM-INT J ENDOCRINO, V16, P388, DOI 10.14310/horm.2002.1759
   August C, 2004, MODERN PATHOL, V17, P1119, DOI 10.1038/modpathol.3800160
   Bialas M, 2013, POL J PATHOL, V64, P129, DOI 10.5114/PJP.2013.36004
   Bialas M, 2012, POL J PATHOL, V63, P235, DOI 10.5114/PJP.2012.32770
   Bilek R, 2008, PHYSIOL RES, V57, pS171, DOI 10.33549/physiolres.931502
   Burnichon N, 2011, HUM MOL GENET, V20, P3974, DOI 10.1093/hmg/ddr324
   Carlsen E, 2009, HORM METAB RES, V41, P715, DOI 10.1055/s-0029-1238274
   Chaux A, 2012, UROLOGY, V80, DOI 10.1016/j.urology.2012.04.032
   Crona J, 2017, ENDOCR REV, V38, P489, DOI 10.1210/er.2017-00062
   Dahia PLM, 2014, NAT REV CANCER, V14, P108, DOI 10.1038/nrc3648
   de Wailly P, 2012, LANGENBECK ARCH SURG, V397, P239, DOI 10.1007/s00423-011-0850-3
   Dwight T, 2018, ENDOCR-RELAT CANCER, V25, P1, DOI 10.1530/ERC-17-0306
   Elston CW, 2000, EUR J CANCER, V36, P1769, DOI 10.1016/S0959-8049(00)00181-7
   Fishbein L, 2017, CANCER CELL, V31, P181, DOI 10.1016/j.ccell.2017.01.001
   Gao BH, 2006, UROLOGY, V68, P282, DOI 10.1016/j.urology.2006.02.019
   Gupta G, 2017, ENDOCR PRACT, V23, P690, DOI 10.4158/EP161718.RA
   Gupta S, 2016, ENDOCR PATHOL, V27, P243, DOI 10.1007/s12022-016-9439-2
   Job S, 2019, CLIN CANCER RES, V25, P760, DOI 10.1158/1078-0432.CCR-18-0139
   Jovanovic R, 2012, Prilozi, V33, P23
   Kajor Maciej, 2005, Endokrynol Pol, V56, P911
   Kim KY, 2016, PLOS ONE, V11, DOI 10.1371/journal.pone.0168413
   Kim S, 2018, SURG ENDOSC, V32, P3401, DOI 10.1007/s00464-017-5969-z
   Kimura N, 2014, ENDOCR-RELAT CANCER, V21, P405, DOI 10.1530/ERC-13-0494
   Koh JM, 2017, PLOS ONE, V12, DOI 10.1371/journal.pone.0187398
   Konosu-Fukaya S, 2018, ENDOCR PATHOL, V29, P302, DOI 10.1007/s12022-018-9544-5
   Kulkarni MM, 2016, J CLIN DIAGN RES, V10, pEC1, DOI 10.7860/JCDR/2016/20565.8419
   Liu TT, 2014, ENDOCR-RELAT CANCER, V21, P427, DOI 10.1530/ERC-14-0016
   Lupsan N, 2016, ROM J MORPHOL EMBRYO, V57, P651
   Maignan A, 2017, LANGENBECK ARCH SURG, V402, P787, DOI 10.1007/s00423-017-1564-y
   Mlika Mona, 2013, Tunis Med, V91, P209
   Muchweti D, 2018, INT J SURG CASE REP, V50, P111, DOI 10.1016/j.ijscr.2018.07.032
   Murali R, 2009, ANN SURG, V249, P641, DOI 10.1097/SLA.0b013e31819ed973
   Netto GJ, 2011, UROLOGY, V77, P1155, DOI 10.1016/j.urology.2010.08.031
   Ocal I, 2014, ASIAN PAC J CANCER P, V15, P1751, DOI 10.7314/APJCP.2014.15.4.1751
   Oudijk L, 2017, NEUROENDOCRINOLOGY, V105, P384, DOI 10.1159/000455864
   Pedziwiatr M, 2015, BMC SURG, V15, DOI 10.1186/s12893-015-0088-z
   Rasquin L, 2018, CASE REP ENDOCRINOL, V2018, DOI 10.1155/2018/6358485
   Stenman A, 2019, AM J SURG PATHOL, V43, P409, DOI 10.1097/PAS.0000000000001190
   Stenman A, 2018, LANGENBECK ARCH SURG, V403, P785, DOI 10.1007/s00423-018-1679-9
   Strong VE, 2008, SURGERY, V143, P759, DOI 10.1016/j.surg.2008.02.007
   Suenaga S, 2016, INTERNAL MED, V55, P3611, DOI 10.2169/internalmedicine.55.7293
   Szalat A, 2011, ENDOCRINE, V39, P160, DOI 10.1007/s12020-010-9422-5
   Thompson LDR, 2002, AM J SURG PATHOL, V26, P551, DOI 10.1097/00000478-200205000-00002
   Wu D, 2009, AM J SURG PATHOL, V33, P599, DOI 10.1097/PAS.0b013e318190d12e
NR 45
TC 38
Z9 40
U1 1
U2 3
PU MDPI
PI BASEL
PA ST ALBAN-ANLAGE 66, CH-4052 BASEL, SWITZERLAND
SN 2072-6694
J9 CANCERS
JI Cancers
PD FEB
PY 2019
VL 11
IS 2
AR 225
DI 10.3390/cancers11020225
PG 12
WC Oncology
WE Science Citation Index Expanded (SCI-EXPANDED)
SC Oncology
GA HO2LS
UT WOS:000460747200099
PM 30769931
OA Green Published, Green Submitted, gold
DA 2023-08-27
ER

PT J
AU Sutton, I
   Winer, J
   Rowlands, D
   Dalmau, J
AF Sutton, I
   Winer, J
   Rowlands, D
   Dalmau, J
TI Limbic encephalitis and antibodies to Ma2: a paraneoplastic presentation
   of breast cancer
SO JOURNAL OF NEUROLOGY NEUROSURGERY AND PSYCHIATRY
LA English
DT Article
DE paraneoplastic; limbic encephalitis; anti-Ma2 antibodies; breast cancer
AB A patient with atypical medullary breast cancer is described who presented with symptoms of limbic encephalitis. The patient's serum and CSF contained antibodies that reacted with the nervous system and the tumour. These antibodies recognised Ma2, a neuronal protein related to paraneoplastic limbic and brainstem encephalitis in men with testicular tumours. This report highlights the importance of testing for paraneoplastic antineuronal antibodies in cases of unexplained limbic encephalitis and suggests screening for breast cancer in women with antibodies predominantly directed to Ma2.
C1 Queen Elizabeth Hosp, Dept Neurol, Birmingham B15 2TH, W Midlands, England.
   Univ Birmingham, Dept Pathol, Birmingham B15 2TT, W Midlands, England.
   Univ Arkansas Med Sci, Dept Neurol, Little Rock, AR 72205 USA.
C3 University of Birmingham; University of Birmingham; University of
   Arkansas System; University of Arkansas Medical Sciences
RP Sutton, I (通讯作者)，Queen Elizabeth Hosp, Dept Neurol, Birmingham B15 2TH, W Midlands, England.
EM
OI Dalmau, Josep/0000-0001-5856-2813
CR Alamowitch S, 1997, BRAIN, V120, P923, DOI 10.1093/brain/120.6.923
   Corriveau RA, 1998, NEURON, V21, P505, DOI 10.1016/S0896-6273(00)80562-0
   Dalmau J, 1999, BRAIN, V122, P27, DOI 10.1093/brain/122.1.27
   GRAUS F, 1990, NEUROLOGY, V40, P219, DOI 10.1212/WNL.40.2.219
   SLOANE JP, 1993, HISTOPATHOLOGY, V23, P225, DOI 10.1111/j.1365-2559.1993.tb01194.x
   SMITT PAES, 1995, NEUROLOGY, V45, P1873
   Voltz R, 1999, NEW ENGL J MED, V340, P1788, DOI 10.1056/NEJM199906103402303
NR 7
TC 25
Z9 26
U1 0
U2 2
PU BMJ PUBLISHING GROUP
PI LONDON
PA BRITISH MED ASSOC HOUSE, TAVISTOCK SQUARE, LONDON WC1H 9JR, ENGLAND
SN 0022-3050
EI 1468-330X
J9 J NEUROL NEUROSUR PS
JI J. Neurol. Neurosurg. Psychiatry
PD AUG
PY 2000
VL 69
IS 2
BP 266
EP 268
DI 10.1136/jnnp.69.2.266
PG 3
WC Clinical Neurology; Psychiatry; Surgery
WE Science Citation Index Expanded (SCI-EXPANDED)
SC Neurosciences & Neurology; Psychiatry; Surgery
GA 338XM
UT WOS:000088448600027
PM 10896708
OA Green Published, Bronze
DA 2023-08-27
ER

PT J
AU Xu, YW
   Lin, P
   Yao, PS
   Zheng, SF
   Kang, DZ
AF Xu, Ya-Wen
   Lin, Peng
   Yao, Pei-Sen
   Zheng, Shu-Fa
   Kang, De-Zhi
TI Structure and function of corticospinal projection originating from
   supplementary motor area
SO NEURORADIOLOGY
LA English
DT Article
DE Motor cortex; Motor evoked potentials; Neurooncology imaging;
   Corticospinal projection; Supplementary motor area
AB Purpose The importance of supplementary motor area (SMA) for motor function and compensation for primary motor area (M1) has received increased attention. Methods We used diffusion tensor imaging (DTI) and transcranial magnetic stimulation (TMS) to evaluate structure and function of corticospinal projection originating from SMA. Fibers of corticospinal projection originating from M1 (CST) and SMA (ACST) were analyzed. ACST originating from mesial SMA area formed separate white matter bundles leaving the anterior part of M1 area, which then entered the posterior limb of the internal capsule. Projection and overlap of both CST and ACST were detected on medulla. Results Fibers of contralesional ACST were more than that of ipsilesional ACST in patients with SMA tumors (p<0.05). In patients with SMA tumor, all patients experienced temporary akinesia postoperatively. Seven hundred forty-one fibers of ipsilateral ACST and no fibers of ipsilateral CST were detected in the patient with M1 glioma, while most of contralateral limb movement was preserved. MEP could be evoked by stimulating SMA area as well as M1 area. ACST originated from SMA area and projected to the medial medulla. Conclusion SMA area and ACST integrity contributed to contralateral motor function and were a compensation for M1 lesion and damaged CST.
C1 [Xu, Ya-Wen; Yao, Pei-Sen; Zheng, Shu-Fa; Kang, De-Zhi] Fujian Med Univ, Affiliated Hosp 1, Inst Neurosurg, Dept Neurosurg, 20 Chazhong Rd, Fuzhou 350004, Peoples R China.
   [Lin, Peng] Fujian Med Univ, Affiliated Hosp 1, Dept Pain Treatment, Fuzhou, Peoples R China.
   [Kang, De-Zhi] Fujian Med Univ, Affiliated Hosp 1, Fujian Key Lab Precis Med Canc, Fuzhou, Peoples R China.
C3 Fujian Medical University; Fujian Medical University; Fujian Medical
   University
RP Kang, DZ (通讯作者)，Fujian Med Univ, Affiliated Hosp 1, Inst Neurosurg, Dept Neurosurg, 20 Chazhong Rd, Fuzhou 350004, Peoples R China.; Kang, DZ (通讯作者)，Fujian Med Univ, Affiliated Hosp 1, Fujian Key Lab Precis Med Canc, Fuzhou, Peoples R China.
EM
FU National Natural Science Foundation of China [81802492]; Young and
   Middle-aged Backbone Key Research Project of National Health and Family
   Planning Commission of Fujian Province [2017-ZQN-46]; Natural Science
   Funding of Fujian Province [2018J01175, 2018J01176]; Fujian Medical
   University [2019QH1087]
FX This work was supported by National Natural Science Foundation of China
   (No. 81802492 to Pei-Sen Yao), Young and Middle-aged Backbone Key
   Research Project of National Health and Family Planning Commission of
   Fujian Province (No. 2017-ZQN-46 to Pei-Sen Yao), and Natural Science
   Funding of Fujian Province (No. 2018J01175 to Pei-Sen Yao and No.
   2018J01176 to Shu-Fa Zheng) and Startup Fund for scientific research,
   Fujian Medical University(No.2019QH1087 to Ming-xia Xu).
CR Amengual JL, 2014, J NEUROPHYSIOL, V112, P2251, DOI 10.1152/jn.00735.2013
   Baker CM, 2018, BRAIN BEHAV, V8, DOI 10.1002/brb3.926
   Bulubas L, 2016, J NEUROSURG, V125, P1431, DOI 10.3171/2015.11.JNS152103
   Chang MC, 2015, NEURAL REGEN RES, V10, P325, DOI 10.4103/1673-5374.152389
   Cona G, 2017, NEUROIMAGE, V146, P770, DOI 10.1016/j.neuroimage.2016.10.032
   Cona G, 2017, NEUROSCI BIOBEHAV R, V72, P28, DOI 10.1016/j.neubiorev.2016.10.033
   Darling WG, 2018, J NEUROSCI, V38, P6323, DOI 10.1523/JNEUROSCI.0403-18.2018
   DEMEURISSE G, 1980, EUR NEUROL, V19, P382, DOI 10.1159/000115178
   Dromerick AW, 2003, J REHABIL RES DEV, V40, P1, DOI 10.1682/JRRD.2003.01.0001
   GALEA MP, 1994, CEREB CORTEX, V4, P166, DOI 10.1093/cercor/4.2.166
   Gerloff C, 1997, BRAIN, V120, P1587, DOI 10.1093/brain/120.9.1587
   Green PE, 2018, NEUROBIOL AGING, V64, P85, DOI 10.1016/j.neurobiolaging.2017.12.016
   Kim YH, 2013, J NEUROSURG, V119, P7, DOI 10.3171/2013.3.JNS121492
   Kobayashi M, 2003, LANCET NEUROL, V2, P145, DOI 10.1016/S1474-4422(03)00321-1
   Magill Stephen T, 2018, J Neurosurg, V129, P961, DOI 10.3171/2017.5.JNS163045
   Mathew P, 2018, STROKE, V49, pE28, DOI 10.1161/STROKEAHA.117.019106
   McNeal DW, 2010, J COMP NEUROL, V518, P586, DOI 10.1002/cne.22218
   Newton JM, 2006, BRAIN, V129, P1844, DOI 10.1093/brain/awl106
   Riley JD, 2011, STROKE, V42, P421, DOI 10.1161/STROKEAHA.110.599340
   Rosenberg K, 2010, J NEUROSURG, V113, P1152, DOI 10.3171/2010.6.JNS1090
   Rosenberg K, 2008, NEUROCASE, V14, P465, DOI 10.1080/13554790802459486
   Schulz R, 2012, STROKE, V43, P2248, DOI 10.1161/STROKEAHA.112.662619
   Seo JP, 2013, AM J NEURORADIOL, V34, P1359, DOI 10.3174/ajnr.A3389
   Tate MC, 2011, J NEUROSURG, V114, P640, DOI 10.3171/2010.9.JNS10709
   Vassal M, 2017, J NEUROSURG, V126, P1181, DOI 10.3171/2016.4.JNS152484
   Yeo SS, 2012, NEURAL REGEN RES, V7, P939, DOI 10.3969/j.issn.1673-5374.2012.12.010
NR 26
TC 2
Z9 2
U1 0
U2 8
PU SPRINGER
PI NEW YORK
PA ONE NEW YORK PLAZA, SUITE 4600, NEW YORK, NY, UNITED STATES
SN 0028-3940
EI 1432-1920
J9 NEURORADIOLOGY
JI Neuroradiology
PD AUG
PY 2021
VL 63
IS 8
BP 1283
EP 1292
DI 10.1007/s00234-021-02669-z
EA FEB 2021
PG 10
WC Clinical Neurology; Neuroimaging; Radiology, Nuclear Medicine & Medical
   Imaging
WE Science Citation Index Expanded (SCI-EXPANDED)
SC Neurosciences & Neurology; Radiology, Nuclear Medicine & Medical Imaging
GA TM3KQ
UT WOS:000619905600002
PM 33611621
DA 2023-08-27
ER

PT J
AU Portychova, L
   Nyvltova, Z
   Vrankova, AB
   Bartos, M
   Pilarova, M
   Vermousek, I
   Antal, M
   Horna, A
AF Portychova, Lenka
   Nyvltova, Zora
   Vrankova, Alice Brabcova
   Bartos, Michal
   Pilarova, Michaela
   Vermousek, Ivan
   Antal, Miroslav
   Horna, Ales
TI PLASMA FREE METANEPHRINES AS DIAGNOSTIC MARKERS OF PHEOCHROMOCYTOMA
SO CHEMICKE LISTY
LA English
DT Article
AB Quantitative determination of catecholamines and their O-methyl metabolites plays an important role in the diagnosis of pheochromocytoma (PHEO) - adrenal medulla tumour. This kind of tumour synthesizes, stocks, metabolizes and mostly secretes catecholamines. For this reason it is possible to use elevated concentrations of catecholamines and their metabolic products from plasma and urine as diagnostic markers of this tumour. Determination of metanephrines is often preferred against determination of catecholamines regarding the diagnosis of PHEO. Tumour cells produce free metanephrines continuously and irrespective of the release of catecholamines.
   The project aims to develop a new kit for the determination of plasma metanephrines. Solid phase extraction (SPE) was used for the pre-treatment of plasma samples. The determination was performed by high performance liquid chromatography with electrochemical detection (HPLC-ED).
C1 [Portychova, Lenka; Pilarova, Michaela; Vermousek, Ivan; Horna, Ales] RADANAL Ltd, Pardubice, Czech Republic.
   [Nyvltova, Zora; Bartos, Michal; Antal, Miroslav] Organ Synth Inc, Res Inst, Rybitvi, Czech Republic.
   [Vrankova, Alice Brabcova] Charles Univ Prague, Fac Med 1, Internal Dept 3, Prague, Czech Republic.
   [Vrankova, Alice Brabcova] Charles Univ Prague, Gen Teaching Hosp, Prague, Czech Republic.
C3 4EU+; Charles University Prague; 4EU+; Charles University Prague;
   General University Hospital Prague
RP Portychova, L (通讯作者)，RADANAL Ltd, Pardubice, Czech Republic.
EM
RI Horna, Ales/H-6388-2012; Vrankova, Alice/C-2516-2017
OI Vrankova, Alice/0000-0002-2327-2338
FU Ministry of industry and trade of Czech Republic [FR-TI4/331]
FX This work was supported by Ministry of industry and trade of Czech
   Republic (project FR-TI4/331).
CR Eisenhofer G, 2012, EUR J CANCER, V48, P1739, DOI 10.1016/j.ejca.2011.07.016
   JOH TH, 1987, ANN NY ACAD SCI, V493, P342, DOI 10.1111/j.1749-6632.1987.tb27217.x
   Krsek M., 2011, ENDOKRINOLOGIE
   Pacak K., 2008, FEOCHROMOCYTOM
   Purves D., 2007, NEUROSCIENCE, V4
   Raggi MA, 1999, J CHROMATOGR B, V730, P201, DOI 10.1016/S0378-4347(99)00213-3
NR 6
TC 0
Z9 0
U1 0
U2 7
PU CHEMICKE LISTY
PI PRAGUE 6
PA NOVOTNEHO LAVKA 5, PRAGUE 6 116 68, CZECH REPUBLIC
SN 0009-2770
EI 1213-7103
J9 CHEM LISTY
JI Chem. Listy
PY 2013
VL 107
SU 3
SI SI
BP S420
EP S422
PG 3
WC Chemistry, Multidisciplinary
WE Science Citation Index Expanded (SCI-EXPANDED)
SC Chemistry
GA 276EL
UT WOS:000328730800056
DA 2023-08-27
ER

PT J
AU Tabarki, B
   Thabet, F
   Al Shafi, S
   Al Adwani, N
   Chehab, M
   Al Shahwan, S
AF Tabarki, Brahim
   Thabet, Farah
   Al Shafi, Shatha
   Al Adwani, Nawal
   Chehab, May
   Al Shahwan, Saad
TI Acute necrotizing encephalopathy associated with enterovirus infection
SO BRAIN & DEVELOPMENT
LA English
DT Article
DE Acute necrotizing encephalopathy; Cytokines; Brainstem lesion; Steroid
ID CHILDHOOD; LESIONS
AB Acute necrotizing encephalopathy is a rare, clinically distinct entity of acute encephalopathy triggered by acute febrile diseases, mostly viral infections. It is postulated to arise from uncontrolled cytokine release during a febrile illness, and is most often seen in East Asia. We describe a rare Saudi patient of acute necrotizing encephalopathy attributable to enterovirus in a 4 years and 6 months old girl. A work-up revealed elevations in serum and cerebrospinal fluid interleukin-6 and tumor necrosis factor-alpha. The outcome on intravenous pulse methylprednisolone was good. This case is the first, to the best of our knowledge, of acute necrotizing encephalopathy reported from Saudi Arabia with a good outcome despite severe magnetic resonance imaging findings and delay in the steroid treatment. (c) 2012 The Japanese Society of Child Neurology. Published by Elsevier B.V. All rights reserved.
C1 [Tabarki, Brahim; Al Shafi, Shatha; Al Shahwan, Saad] Riyadh Mil Hosp, Dept Pediat, Div Pediat Neurol, Riyadh 11159, Saudi Arabia.
   [Thabet, Farah; Chehab, May] Riyadh Mil Hosp, Dept Pediat, Div Pediat Crit Care, Riyadh 11159, Saudi Arabia.
   [Al Adwani, Nawal] Riyadh Mil Hosp, Dept Radiol, Riyadh 11159, Saudi Arabia.
C3 Prince Sultan Military Medical City
RP Tabarki, B (通讯作者)，Riyadh Mil Hosp, Dept Pediat, Div Pediat Neurol, POB 7897, Riyadh 11159, Saudi Arabia.
EM
CR Fowlkes AL, 2008, J INFECT DIS, V198, P1685, DOI 10.1086/592988
   Gika AD, 2010, DEV MED CHILD NEUROL, V52, P99, DOI 10.1111/j.1469-8749.2009.03405.x
   Harada M, 2000, JMRI-J MAGN RESON IM, V11, P559, DOI 10.1002/(SICI)1522-2586(200005)11:5<559::AID-JMRI12>3.0.CO;2-C
   Kawano G, 2011, ARCH DIS CHILD, V96, P936, DOI 10.1136/adc.2009.180554
   Kim JH, 2004, KOREAN J RADIOL, V5, P171, DOI 10.3348/kjr.2004.5.3.171
   Lyon JB, 2010, PEDIATR RADIOL, V40, P200, DOI 10.1007/s00247-009-1487-z
   Mizuguchi M, 2002, NEURORADIOLOGY, V44, P489, DOI 10.1007/s00234-002-0773-3
   MIZUGUCHI M, 1995, J NEUROL NEUROSUR PS, V58, P555, DOI 10.1136/jnnp.58.5.555
   Neilson DE, 2009, AM J HUM GENET, V84, P44, DOI 10.1016/j.ajhg.2008.12.009
   Okumura A, 2009, BRAIN DEV-JPN, V31, P221, DOI 10.1016/j.braindev.2008.03.005
   Sujay M, 2011, PEDIATR NEUROL, V45, P400
   Weng WC, 2010, J CHILD NEUROL, V25, P1539, DOI 10.1177/0883073810374877
   Wilken B, 1997, J PEDIATR-US, V130, P89, DOI 10.1016/S0022-3476(97)70315-9
NR 13
TC 19
Z9 27
U1 0
U2 8
PU ELSEVIER SCIENCE BV
PI AMSTERDAM
PA PO BOX 211, 1000 AE AMSTERDAM, NETHERLANDS
SN 0387-7604
J9 BRAIN DEV-JPN
JI Brain Dev.
PD MAY
PY 2013
VL 35
IS 5
BP 454
EP 457
DI 10.1016/j.braindev.2012.07.001
PG 4
WC Clinical Neurology; Pediatrics
WE Science Citation Index Expanded (SCI-EXPANDED)
SC Neurosciences & Neurology; Pediatrics
GA 136VE
UT WOS:000318390700013
PM 22832063
DA 2023-08-27
ER

PT J
AU Khong, J
   Govindaraj, R
   Ramm, D
   Edwards, S
   Roos, D
AF Khong, Jeremy
   Govindaraj, Ramkumar
   Ramm, Daniel
   Edwards, Suzanne
   Roos, Daniel
TI Cochlear sparing in LINAC-based radiosurgery for vestibular schwannoma:
   a dosimetric comparison of dynamic conformal arc, IMRT and VMAT
   treatment plans
SO RADIATION ONCOLOGY
LA English
DT Article
DE Cochlear sparing; Plan comparison; Stereotactic radiosurgery;
   Techniques; Vestibular schwannoma
ID GAMMA-KNIFE RADIOSURGERY; STEREOTACTIC RADIOSURGERY; HEARING
   PRESERVATION; RADIOTHERAPY; OUTCOMES; THERAPY
AB BackgroundStereotactic radiosurgery (SRS) is the preferred treatment for vestibular schwannoma (VS) in patients with preserved hearing and tumour diameter < 3 cm. Emerging evidence suggests restricting cochlear dose could preserve hearing. This retrospective replanning study aims to compare dynamic conformal arc therapy (DCAT), intensity modulated radiation therapy (IMRT) and volumetric modulated arc therapy (VMAT) plans for superiority of cochlear dose sparing without compromising tumour coverage. MethodsEligibility criteria included sporadic VS, serviceable hearing and availability of CT and MRI for planning. The original gross tumour volume and brainstem OAR volume were retained; the cochlea was newly contoured on the planning CT scan (bone window). Each case was replanned using the three above techniques, prescribing 12 Gy to the 80% isodose line. No dose constraint was applied to the cochlea. ResultsEighteen patients were replanned. Mean tumour volume was 2.25 cc. Tumour coverage and tumour mean dose (DCAT: 14.2, IMRT: 14.6, VMAT: 14.5 Gy) were comparable. Paddick and RTOG conformity indices were better for DCAT (0.66 and 1.6) and VMAT (0.69 and 1.5) compared to IMRT (0.56 and 1.9). DCAT had superior gradient index (3.0) compared to VMAT (3.4) and IMRT (3.4). VMAT delivered the lowest mean brainstem maximum dose (8.3 Gy) and decreased the mean cochlear dose (3.4 Gy) by 2.3 and 2.1 Gy, and the mean cochlear maximum dose (3.6 Gy) by 2.4 and 2.5 Gy relative to DCAT and IMRT, respectively. ConclusionLINAC-based SRS treatment using VMAT can achieve better cochlear dose sparing than DCAT or IMRT while maintaining tumour coverage.
C1 [Khong, Jeremy; Govindaraj, Ramkumar; Ramm, Daniel; Roos, Daniel] Royal Adelaide Hosp, Dept Radiat Oncol, Port Rd, Adelaide, SA 5000, Australia.
   [Govindaraj, Ramkumar; Roos, Daniel] Univ Adelaide, Sch Med, Adelaide, SA, Australia.
   [Edwards, Suzanne] Univ Adelaide, Sch Publ Hlth, Adelaide Hlth Technol Assessment AHTA, Adelaide, SA, Australia.
C3 Royal Adelaide Hospital; University of Adelaide; University of Adelaide
RP Khong, J (通讯作者)，Royal Adelaide Hosp, Dept Radiat Oncol, Port Rd, Adelaide, SA 5000, Australia.
EM
OI Govindaraj, Ramkumar/0000-0002-7229-7179
CR Abacioglu U, 2014, RADIAT ONCOL, V9, DOI 10.1186/1748-717X-9-119
   Babu R, 2013, J NEUROSURG, V119, P121, DOI 10.3171/2013.1.JNS121370
   Balik S, 2018, J RADIOSURGERY SBRT, V5, P237
   Baschnagel AM, 2013, J NEUROSURG, V118, P571, DOI 10.3171/2012.10.JNS12880
   Benghiat H, 2014, CLIN ONCOL-UK, V26, P309, DOI 10.1016/j.clon.2014.02.008
   Bhandare N, 2010, INT J RADIAT ONCOL, V76, pS50, DOI 10.1016/j.ijrobp.2009.04.096
   Carlson ML, 2018, NEUROSURGERY, V82, pE35, DOI 10.1093/neuros/nyx511
   Dutta D, 2012, J NEURO-ONCOL, V106, P637, DOI 10.1007/s11060-011-0703-5
   Evans DGR, 2005, OTOL NEUROTOL, V26, P93, DOI 10.1097/00129492-200501000-00016
   Frischer JM, 2019, J NEUROSURG, V130, P388, DOI 10.3171/2017.8.JNS171281
   Goldbrunner R, 2020, NEURO-ONCOLOGY, V22, P31, DOI 10.1093/neuonc/noz153
   Jacob JT, 2014, NEUROSURGERY, V74, P466, DOI 10.1227/NEU.0000000000000299
   Kim H, 2015, RADIOTHER ONCOL, V115, P229, DOI 10.1016/j.radonc.2015.03.031
   Lagerwaard FJ, 2009, INT J RADIAT ONCOL, V74, P610, DOI 10.1016/j.ijrobp.2008.12.076
   Lin RH, 2017, CLIN NEUROL NEUROSUR, V162, P16, DOI 10.1016/j.clineuro.2017.09.001
   Mahboubi H, 2017, OTOLARYNG HEAD NECK, V157, P7, DOI 10.1177/0194599817695805
   McDonald F, 2018, CLIN ONCOL-UK, V30, P1, DOI [10.1016/j.clon.2017.09.007, 10.1016/j.clon.2017.10.013]
   Ohata K, 2002, J Postgrad Med, V48, P253
   Pacholke HD, 2005, AM J CLIN ONCOL-CANC, V28, P143, DOI 10.1097/01.coc.0000143847.57027.16
   Paddick I, 2000, J NEUROSURG, V93, P219, DOI 10.3171/jns.2000.93.supplement_3.0219
   Patel KS, 2019, J NEURO-ONCOL, V145, P329, DOI 10.1007/s11060-019-03299-5
   Regis Jean, 2008, Prog Neurol Surg, V21, P142, DOI 10.1159/000156901
   Schumacher AJ, 2017, J NEUROL SURG PART B, V78, P2, DOI 10.1055/s-0036-1584231
   Sharma SD, 2014, J CANCER RES THER, V10, P29, DOI 10.4103/0973-1482.131353
   Tamura M, 2009, NEUROSURGERY, V64, P289, DOI 10.1227/01.NEU.0000338256.87936.7C
   Tolisano AM, 2019, J NEUROL SURG PART B, V80, P156, DOI 10.1055/s-0039-1677680
   Torrens M, 2014, J NEUROSURG, V121, P2, DOI 10.3171/2014.7.GKS141199
   Tsao MN, 2017, J RADIOSURGERY SBRT, V5, P5
   Wolbers John G, 2013, BMJ Open, V3, DOI 10.1136/bmjopen-2012-001345
NR 29
TC 0
Z9 0
U1 0
U2 0
PU BMC
PI LONDON
PA CAMPUS, 4 CRINAN ST, LONDON N1 9XW, ENGLAND
EI 1748-717X
J9 RADIAT ONCOL
JI Radiat. Oncol.
PD JAN 4
PY 2023
VL 18
IS 1
AR 2
DI 10.1186/s13014-022-02188-y
PG 7
WC Oncology; Radiology, Nuclear Medicine & Medical Imaging
WE Science Citation Index Expanded (SCI-EXPANDED)
SC Oncology; Radiology, Nuclear Medicine & Medical Imaging
GA 7W3VC
UT WOS:000913442100001
PM 36600254
OA gold, Green Published
DA 2023-08-27
ER

PT J
AU Zhang, L
   Shi, YY
   Wang, YX
   Zhu, FL
   Wang, Q
   Ma, CH
   Chen, YHH
   Zhang, LN
AF Zhang, Li
   Shi, Yongyu
   Wang, Yongxiang
   Zhu, Faliang
   Wang, Qun
   Ma, Chunhong
   Chen, Youhai H.
   Zhang, Lining
TI The unique expression profile of human TIPE2 suggests new functions
   beyond its role in immune regulation
SO MOLECULAR IMMUNOLOGY
LA English
DT Article
DE Human TIPE2 (tumor necrosis factor-alpha-induced protein 8-like 2);
   Expression; Immunohistochemistry (IHC); Flow cytometry; Western blot
ID NF-KAPPA-B; STRATIFIED SQUAMOUS EPITHELIA; UP-REGULATION; HOMEOSTASIS;
   EPIDERMIS; DIFFERENTIATION; KERATINOCYTES; GROWTH; DEATH; CELLS
AB TIPE2 (tumor necrosis factor-alpha-induced protein 8-like 2, or TNFAIP8L2) is a newly identified negative regulator of innate and adaptive immunity. TIPE2 deficiency in mice leads to systemic inflammation and TIPE2 down-regulation in humans is associated with autoimmunity. However, the nature of human TIPE2-expressing cells in various tissues has not been characterized due to the lack of suitable antibodies. In the present study, we generated specific rabbit antibodies against human TIPE2 and examined human TIPE2 expression in various tissues by Western blot, flow cytometry, and immunohistochemistry. We found that unlike murine TIPE2 that is preferentially expressed by hematopoietic cells, human TIPE2 was also expressed in a wide variety of non-hematopoietic cell types, including hepatocytes, neurons in brain and brainstem, squamous epithelial cells in esophagus and cervix, transitional epithelial cells in bladder and ureter, and glandular epithelial cells in stomach, colon and appendix. For squamous epithelium, the level of TIPE2 expression increased dramatically in terminally differentiated cells with the proliferating precursor cells completely devoid of TIPE2. The unique expressional profile of human TIPE2 suggests new functions beyond controlling innate and adaptive immunity. (C) 2011 Elsevier Ltd. All rights reserved.
C1 [Zhang, Li; Shi, Yongyu; Wang, Yongxiang; Zhu, Faliang; Wang, Qun; Ma, Chunhong; Zhang, Lining] Shandong Univ, Sch Med, Dept Immunol, Jinan 250012, Shandong, Peoples R China.
   [Chen, Youhai H.] Univ Penn, Dept Pathol & Lab Med, Sch Med, Philadelphia, PA 19104 USA.
   [Zhang, Li] Qianfoshan Hosp Shandong Prov, Dept Lab, Jinan, Shandong, Peoples R China.
C3 Shandong University; University of Pennsylvania; Shandong First Medical
   University & Shandong Academy of Medical Sciences
