## Supplemental Material 1 for "Bibliometric analysis of emerging trends and research foci in brainstem tumor field over thirty years (1992-2023)": savedrecs (8).txt

FN Clarivate Analytics Web of Science
VR 1.0
PT J
AU Iannalfi, A
   D'Ippolito, E
   Riva, G
   Molinelli, S
   Gandini, S
   Viselner, G
   Fiore, MR
   Vischioni, B
   Vitolo, V
   Bonora, M
   Ronchi, S
   Petrucci, R
   Barcellini, A
   Mirandola, A
   Russo, S
   Vai, A
   Mastella, E
   Magro, G
   Maestri, D
   Ciocca, M
   Preda, L
   Valvo, F
   Orecchia, R
AF Iannalfi, Alberto
   D'Ippolito, Emma
   Riva, Giulia
   Molinelli, Silvia
   Gandini, Sara
   Viselner, Gisela
   Fiore, Maria Rosaria
   Vischioni, Barbara
   Vitolo, Viviana
   Bonora, Maria
   Ronchi, Sara
   Petrucci, Rachele
   Barcellini, Amelia
   Mirandola, Alfredo
   Russo, Stefania
   Vai, Alessandro
   Mastella, Edoardo
   Magro, Giuseppe
   Maestri, Davide
   Ciocca, Mario
   Preda, Lorenzo
   Valvo, Francesca
   Orecchia, Roberto
TI Proton and carbon ion radiotherapy in skull base chordomas: a
   prospective study based on a dual particle and a patient-customized
   treatment strategy
SO NEURO-ONCOLOGY
LA English
DT Article
DE carbon ions therapy; chordoma; proton therapy; skull base
ID RADIATION-THERAPY; PROGNOSTIC-FACTORS; DOSE PRESCRIPTION;
   CERVICAL-SPINE; IRRADIATION; MANAGEMENT; SURVIVAL; OUTCOMES;
   CHONDROSARCOMAS; RECURRENCE
AB Background. The aim of this study is to evaluate results in terms of local control (LC), overall survival (OS), and toxicity profile and to better identify factors influencing clinical outcome of skull base chordoma treated with proton therapy (PT) and carbon ion radiotherapy (CIRT).
   Methods. We prospectively collected and analyzed data of 135 patients treated between November 2011 and December 2018. Total prescription dose in the PT group (70 patients) and CIRT group (65 patients) was 74 Gy relative biological effectiveness (RBE) delivered in 37 fractions and 70.4 Gy(RBE) delivered in 16 fractions, respectively (CIRT in unfavorable patients). LC and OS were evaluated using the Kaplan-Meier method. Univariate and multivariate analyses were performed, to identify prognostic factors on clinical outcomes.
   Results. After a median follow-up of 44 (range, 6-87) months, 14 (21%) and 8 (11%) local failures were observed in CIRT and PT group, respectively. Five-year LC rate was 71% in CIRT cohort and 84% in PT cohort. The estimated 5-year OS rate in the CIRT and PT group was 82% and 83%, respectively. On multivariate analysis, gross tumor volume (GTV), optic pathways, and/or brainstem compression and dose coverage are independent prognostic factors of local failure risk. High rate toxicity grade >= 3 was reported in 11% of patients.
   Conclusions. Particle radiotherapy is an effective treatment for skull base chordoma with acceptable late toxicity. GTV, optic pathways, and/or brainstem compression and target coverage were independent prognostic factors for LC.
C1 [Iannalfi, Alberto; D'Ippolito, Emma; Riva, Giulia; Molinelli, Silvia; Viselner, Gisela; Fiore, Maria Rosaria; Vischioni, Barbara; Vitolo, Viviana; Bonora, Maria; Ronchi, Sara; Petrucci, Rachele; Barcellini, Amelia; Mirandola, Alfredo; Russo, Stefania; Vai, Alessandro; Mastella, Edoardo; Magro, Giuseppe; Maestri, Davide; Ciocca, Mario; Preda, Lorenzo; Valvo, Francesca] Natl Ctr Oncol Hadrontherapy CNAO, Clin Dept, Pavia, Italy.
   [Gandini, Sara] IRCCS, Dept Expt Oncol, European Inst Oncol, Milan, Italy.
   [Orecchia, Roberto] IRCCS, European Inst Oncol, Sci Directorate, Milan, Italy.
C3 Centro Nazionale di Adroterapia Oncologica (CNAO); IRCCS European
   Institute of Oncology (IEO); IRCCS European Institute of Oncology (IEO)
RP Iannalfi, A (通讯作者)，Fdn CNAO, Str Campeggi 53, I-27100 Pavia, PV, Italy.
EM
RI Ronchi, Sara/AAC-7655-2022; PREDA, LORENZO/AAC-1969-2019; Vischioni,
   Barbara/GYJ-3050-2022; Mastella, Edoardo/ITV-2670-2023; Barcellini,
   Amelia/AAK-9012-2020; Gandini, Sara/J-4916-2018
OI Ronchi, Sara/0000-0002-5518-4121; PREDA, LORENZO/0000-0002-5479-2766;
   Vischioni, Barbara/0000-0003-4920-2239; Mastella,
   Edoardo/0000-0002-1913-3976; Barcellini, Amelia/0000-0002-1595-104X;
   Gandini, Sara/0000-0002-1348-4548; D'Ippolito, Emma/0000-0003-2899-6447;
   Galzio, Renato/0000-0003-1933-1355; Mirandola,
   Alfredo/0000-0003-2427-2329
CR Amit M, 2014, J NEUROL SURG PART B, V75, P383, DOI 10.1055/s-0034-1376197
   Ares C, 2009, INT J RADIAT ONCOL, V75, P1111, DOI 10.1016/j.ijrobp.2008.12.055
   Choy W, 2015, J CLIN NEUROSCI, V22, P1792, DOI 10.1016/j.jocn.2015.05.024
   Dale JE, 2019, RADIOTHER ONCOL, V140, P175, DOI 10.1016/j.radonc.2019.06.028
   Debus J, 1997, INT J RADIAT ONCOL, V39, P967, DOI 10.1016/S0360-3016(97)00364-7
   Durante M, 2010, NAT REV CLIN ONCOL, V7, P37, DOI 10.1038/nrclinonc.2009.183
   Flanagan A., 2013, WHO CLASSIFICATION T, P328
   Fossati P, 2012, PHYS MED BIOL, V57, P7543, DOI 10.1088/0031-9155/57/22/7543
   Fung V, 2018, RADIOTHER ONCOL, V128, P198, DOI 10.1016/j.radonc.2017.12.017
   Halperin EC, 1997, INT J RADIAT ONCOL, V38, P225, DOI 10.1016/S0360-3016(97)00041-2
   Hasegawa A, 2006, INT J RADIAT ONCOL, V64, P396, DOI 10.1016/j.ijrobp.2005.07.298
   Hug EB, 1999, J NEUROSURG, V91, P432, DOI 10.3171/jns.1999.91.3.0432
   Jahangiri A, 2015, NEUROSURGERY, V76, P179, DOI 10.1227/NEU.0000000000000611
   Kanai T, 1999, INT J RADIAT ONCOL, V44, P201, DOI 10.1016/S0360-3016(98)00544-6
   Koto M, 2014, RADIOTHER ONCOL, V111, P25, DOI 10.1016/j.radonc.2013.11.005
   Kramer M, 2000, PHYS MED BIOL, V45, P3319, DOI 10.1088/0031-9155/45/11/314
   Mayo C, 2010, INT J RADIAT ONCOL, V76, pS28, DOI 10.1016/j.ijrobp.2009.07.1753
   Mayo C, 2010, INT J RADIAT ONCOL, V76, pS36, DOI 10.1016/j.ijrobp.2009.08.078
   McDonald MW, 2016, INT J RADIAT ONCOL, V95, P304, DOI 10.1016/j.ijrobp.2015.08.011
   Mirandola A, 2015, MED PHYS, V42, P5287, DOI 10.1118/1.4928397
   Mizoe J, 2016, REP PRACT ONCOL RADI, V21, P356, DOI 10.1016/j.rpor.2015.01.008
   Mizoe JE, 2009, SKULL BASE-INTERD AP, V19, P219, DOI 10.1055/s-0028-1114295
   Molinelli S, 2019, RADIOTHER ONCOL, V141, P227, DOI 10.1016/j.radonc.2019.08.022
   Molinelli S, 2016, RADIOTHER ONCOL, V120, P307, DOI 10.1016/j.radonc.2016.05.031
   Munzenrider JE, 1999, STRAHLENTHER ONKOL, V175, P57, DOI 10.1007/BF03038890
   Noel G, 2005, ACTA ONCOL, V44, P700, DOI 10.1080/02841860500326257
   Noel G, 2004, NEUROSURGERY, V55, P1252, DOI 10.1227/01.NEU.0000143330.30405.AA
   Noel G, 2003, STRAHLENTHER ONKOL, V179, P241, DOI 10.1007/s00066-003-1065-5
   Pehlivan B, 2012, INT J RADIAT ONCOL, V83, P1432, DOI 10.1016/j.ijrobp.2011.10.042
   Potluri S, 2011, CLIN ONCOL-UK, V23, P199, DOI 10.1016/j.clon.2010.09.011
   Program CTE, COMMON TERMINOLOGY C
   Samii A, 2007, J NEUROSURG, V107, P319, DOI 10.3171/JNS-07/08/0319
   Santoni R, 1998, INT J RADIAT ONCOL, V41, P59, DOI 10.1016/S0360-3016(98)00031-5
   Scholz M, 1997, RADIAT ENVIRON BIOPH, V36, P59, DOI 10.1007/s004110050055
   Schulz-Ertner D, 2007, INT J RADIAT ONCOL, V68, P449, DOI 10.1016/j.ijrobp.2006.12.059
   Shanbhogue AKP, 2010, J COMPUT ASSIST TOMO, V34, P479, DOI 10.1097/RCT.0b013e3181db2670
   Smoll NR, 2013, CANCER-AM CANCER SOC, V119, P2029, DOI 10.1002/cncr.28032
   Stacchiotti S, 2015, LANCET ONCOL, V16, pE71, DOI 10.1016/S1470-2045(14)71190-8
   Terahara A, 1999, INT J RADIAT ONCOL, V45, P351, DOI 10.1016/S0360-3016(99)00146-7
   Tuan J, 2013, J RADIAT RES, V54, P31, DOI 10.1093/jrr/rrt036
   Uhl M, 2014, CANCER-AM CANCER SOC, V120, P3410, DOI 10.1002/cncr.28877
   Vogin G, 2019, RADIAT ONCOL, V14, DOI 10.1186/s13014-019-1224-1
   Walcott BP, 2012, LANCET ONCOL, V13, pE69, DOI 10.1016/S1470-2045(11)70337-0
   Weber DC, 2016, RADIOTHER ONCOL, V120, P169, DOI 10.1016/j.radonc.2016.05.011
   YOUDEN WJ, 1950, BIOMETRICS, V6, P172, DOI 10.1002/1097-0142(1950)3:1<32::AID-CNCR2820030106>3.0.CO;2-3
   Zou MX, 2018, WORLD NEUROSURG, V109, P307, DOI 10.1016/j.wneu.2017.10.010
NR 46
TC 27
Z9 27
U1 1
U2 5
PU OXFORD UNIV PRESS INC
PI CARY
PA JOURNALS DEPT, 2001 EVANS RD, CARY, NC 27513 USA
SN 1522-8517
EI 1523-5866
J9 NEURO-ONCOLOGY
JI Neuro-Oncology
PD SEP
PY 2020
VL 22
IS 9
BP 1348
EP 1358
DI 10.1093/neuonc/noaa067
PG 11
WC Oncology; Clinical Neurology
WE Science Citation Index Expanded (SCI-EXPANDED)
SC Oncology; Neurosciences & Neurology
GA OW8HJ
UT WOS:000593120300016
PM 32193546
OA Bronze, Green Published
DA 2023-08-27
ER

PT J
AU Filbin, MG
   Sturm, D
AF Filbin, Mariella G.
   Sturm, Dominik
TI Gliomas in Children
SO SEMINARS IN NEUROLOGY
LA English
DT Article
DE CNS; brain tumor; pediatric glioma; astrocytoma; genetics; epigenetics;
   targeted therapy
ID DIFFUSE INTRINSIC PONTINE; LOW-GRADE GLIOMAS; ACTIVATING ACVR1
   MUTATIONS; BRAIN-STEM TUMORS; PHASE-III TRIAL; ONCOLOGY-GROUP; PEDIATRIC
   GLIOBLASTOMA; HYPERFRACTIONATED RADIOTHERAPY; NEUROFIBROMATOSIS TYPE-1;
   GENETIC ALTERATIONS
AB Gliomas are the most common primary central nervous system (CNS) neoplasms in children and adolescents and are thought to arise from their glial progenitors or stem cells. Although the exact cells of origin for most pediatric gliomas remain to be identified, our current understanding is that specific cell populations during CNS development are susceptible to particular oncogenic events during certain time windows and thus give rise to pediatric gliomas with distinct histological, molecular, and clinical features. These may be roughly segregated into low-grade gliomas (WHO grades I or II; including most mixed glial-neuronal tumors) and high-grade gliomas (WHO grades III or IV) according to their clinical course when untreated, even though this is not yet entirely clear for some of the recently emerging groups. The genetic and epigenetic characterization of pediatric gliomas across ages and histologies has facilitated the delineation of biologically relevant subgroups and have revealed potentially targetable alterations in some of them. This review outlines diagnostic features and molecular alterations in pediatric low- and high-grade gliomas and how the latter might be exploited with future targeted therapeutic strategies.
C1 [Filbin, Mariella G.] Dana Farber Canc Inst, Dept Pediat Oncol, 450 Brookline Ave, Boston, MA 02215 USA.
   [Filbin, Mariella G.] Childrens Hosp, Ctr Canc, 450 Brookline Ave, Boston, MA 02215 USA.
   [Filbin, Mariella G.] Massachusetts Gen Hosp, Dept Pathol, Boston, MA 02114 USA.
   [Filbin, Mariella G.] Massachusetts Gen Hosp, Ctr Canc Res, Boston, MA 02114 USA.
   [Filbin, Mariella G.] Broad Inst MIT & Harvard, Cambridge, MA USA.
   [Sturm, Dominik] NCT Heidelberg KiTZ, Hopp Childrens Canc Ctr, Heidelberg, Germany.
   [Sturm, Dominik] German Canc Res Ctr, Div Pediat Neurooncol, Heidelberg, Germany.
   [Sturm, Dominik] German Canc Consortium DKTK, Heidelberg, Germany.
   [Sturm, Dominik] Heidelberg Univ Hosp, Dept Pediat Hematol & Oncol, Heidelberg, Germany.
C3 Harvard University; Dana-Farber Cancer Institute; Harvard University;
   Boston Children's Hospital; Harvard University; Massachusetts General
   Hospital; Harvard University; Massachusetts General Hospital; Harvard
   University; Massachusetts Institute of Technology (MIT); Broad
   Institute; Helmholtz Association; German Cancer Research Center (DKFZ);
   Helmholtz Association; German Cancer Research Center (DKFZ); Ruprecht
   Karls University Heidelberg
RP Filbin, MG (通讯作者)，Dana Farber Canc Inst, Dept Pediat Oncol, 450 Brookline Ave, Boston, MA 02215 USA.; Filbin, MG (通讯作者)，Childrens Hosp, Ctr Canc, 450 Brookline Ave, Boston, MA 02215 USA.
EM
RI Sturm, Dominik/AAL-3689-2020
OI Sturm, Dominik/0000-0003-0250-1696
CR [Anonymous], 2016, WHO CLASSIFICATION T
   Ater JL, 2016, CANCER-AM CANCER SOC, V122, P1928, DOI 10.1002/cncr.29987
   Bailey S, 2013, EUR J CANCER, V49, P3856, DOI 10.1016/j.ejca.2013.08.006
   Bandopadhayay P, 2016, NAT GENET, V48, P273, DOI 10.1038/ng.3500
   Bandopadhayay P, 2014, PEDIATR BLOOD CANCER, V61, P1173, DOI 10.1002/pbc.24958
   Banerjee A, 2017, NEURO-ONCOLOGY, V19, P1135, DOI 10.1093/neuonc/now282
   Bergthold G, 2014, BBA-REV CANCER, V1845, P294, DOI 10.1016/j.bbcan.2014.02.004
   Broniscer A, 2016, ACTA NEUROPATHOL, V131, P299, DOI 10.1007/s00401-015-1532-y
   Buczkowicz P, 2014, ACTA NEUROPATHOL, V128, P573, DOI 10.1007/s00401-014-1319-6
   Buczkowicz P, 2014, NAT GENET, V46, P451, DOI 10.1038/ng.2936
   Castel D, 2015, ACTA NEUROPATHOL, V130, P815, DOI 10.1007/s00401-015-1478-0
   Cohen KJ, 2011, NEURO-ONCOLOGY, V13, P317, DOI 10.1093/neuonc/noq191
   Dawson MA, 2012, CELL, V150, P12, DOI 10.1016/j.cell.2012.06.013
   DIRKS PB, 1994, NEUROSURGERY, V34, P68
   Dombi E, 2016, NEW ENGL J MED, V375, P2550, DOI 10.1056/NEJMoa1605943
   Dougherty MJ, 2010, NEURO-ONCOLOGY, V12, P621, DOI 10.1093/neuonc/noq007
   Filbin MG, 2016, ANNU REV PATHOL-MECH, V11, P497, DOI 10.1146/annurev-pathol-012615-044208
   FINLAY JL, 1995, J CLIN ONCOL, V13, P112, DOI 10.1200/JCO.1995.13.1.112
   Fontebasso AM, 2014, NAT GENET, V46, P462, DOI 10.1038/ng.2950
   Franz DN, 2015, ANN NEUROL, V78, P929, DOI 10.1002/ana.24523
   Gnekow AK, 2012, NEURO-ONCOLOGY, V14, P1265, DOI 10.1093/neuonc/nos202
   Grasso CS, 2015, NAT MED, V21, P555, DOI 10.1038/nm.3855
   Grill J, 1999, ANN NEUROL, V45, P393, DOI 10.1002/1531-8249(199903)45:3<393::AID-ANA17>3.0.CO;2-B
   Hashizume R, 2014, NAT MED, V20, P1394, DOI 10.1038/nm.3716
   Hoffman LM, 2016, ACTA NEUROPATHOL COM, V4, DOI 10.1186/s40478-015-0269-0
   Huang TY, 2017, ACTA NEUROPATHOL COM, V5, DOI 10.1186/s40478-017-0436-6
   Hummel TR, 2016, J NEURO-ONCOL, V127, P53, DOI 10.1007/s11060-015-2008-6
   Hummel TR, 2013, PEDIATR BLOOD CANCER, V60, P1452, DOI 10.1002/pbc.24541
   Jakacki RI, 2016, NEURO-ONCOLOGY, V18, P1442, DOI 10.1093/neuonc/now038
   Janssens GO, 2017, EUR J CANCER, V73, P38, DOI 10.1016/j.ejca.2016.12.007
   JENKIN RDT, 1987, J NEUROSURG, V66, P227, DOI 10.3171/jns.1987.66.2.0227
   Johansson G, 2016, ACTA ONCOL, V55, P401, DOI 10.3109/0284186X.2015.1107190
   Jones C, 2014, NAT REV CANCER, V14, P651, DOI 10.1038/nrc3811
   Jones C, 2017, NEURO-ONCOLOGY, V19, P153, DOI 10.1093/neuonc/now101
   Jones DTW, 2008, CANCER RES, V68, P8673, DOI 10.1158/0008-5472.CAN-08-2097
   Jones DTW, 2013, NAT GENET, V45, P927, DOI 10.1038/ng.2682
   Karajannis MA, 2014, NEURO-ONCOLOGY, V16, P1408, DOI 10.1093/neuonc/nou059
   Kieran MW, 2016, ANN ONCOL, V27, DOI 10.1093/annonc/mdw435.9
   Kieran M, 2014, NEURO-ONCOLOGY, V16, P56
   Korshunov A, 2017, ACTA NEUROPATHOL, V134, P507, DOI 10.1007/s00401-017-1710-1
   Korshunov A, 2015, ACTA NEUROPATHOL, V129, P669, DOI 10.1007/s00401-015-1405-4
   Koschmann C, 2016, ONCOTARGET, V7, P65696, DOI 10.18632/oncotarget.11602
   Krueger DA, 2010, NEW ENGL J MED, V363, P1801, DOI 10.1056/NEJMoa1001671
   Lassaletta A, 2016, J CLIN ONCOL, V34, P3537, DOI 10.1200/JCO.2016.68.1585
   Lewis PW, 2013, SCIENCE, V340, P857, DOI 10.1126/science.1232245
   Listernick R, 1999, AM J MED GENET, V89, P38, DOI 10.1002/(SICI)1096-8628(19990326)89:1<38::AID-AJMG8>3.0.CO;2-M
   Macy ME, 2017, PEDIATR BLOOD CANCER, V64, DOI 10.1002/pbc.26621
   Mandell LR, 1999, INT J RADIAT ONCOL, V43, P959, DOI 10.1016/S0360-3016(98)00501-X
   Mistry M, 2015, J CLIN ONCOL, V33, P1015, DOI 10.1200/JCO.2014.58.3922
   Mohammad F, 2017, NAT MED, V23, P483, DOI 10.1038/nm.4293
   La Madrid AM, 2017, CHILD NERV SYST, V33, P849, DOI 10.1007/s00381-017-3352-y
   Northrup H, 2013, PEDIATR NEUROL, V49, P243, DOI 10.1016/j.pediatrneurol.2013.08.001
   PACKER RJ, 1994, CANCER-AM CANCER SOC, V74, P1827, DOI 10.1002/1097-0142(19940915)74:6<1827::AID-CNCR2820740628>3.0.CO;2-Q
   Packer RJ, 2017, NEURO-ONCOLOGY, V19, P750, DOI 10.1093/neuonc/now209
   Paugh BS, 2013, CANCER RES, V73, P6219, DOI 10.1158/0008-5472.CAN-13-1491
   Piunti A, 2017, NAT MED, V23, P493, DOI 10.1038/nm.4296
   Pollack IF, 2003, NEURO-ONCOLOGY, V5, P197, DOI 10.1215/S1152851703000097
   Qaddoumi I, 2016, ACTA NEUROPATHOL, V131, P833, DOI 10.1007/s00401-016-1539-z
   Qaddoumi I, 2014, FRONT ONCOL, V4, DOI 10.3389/fonc.2014.00067
   Ramkissoon LA, 2013, P NATL ACAD SCI USA, V110, P8188, DOI 10.1073/pnas.1300252110
   Ramkissoon SH, 2017, NEURO-ONCOLOGY, V19, P986, DOI 10.1093/neuonc/now294
   Saygin C, 2017, J HEMATOL ONCOL, V10, DOI 10.1186/s13045-017-0463-6
   Schindler G, 2011, ACTA NEUROPATHOL, V121, P397, DOI 10.1007/s00401-011-0802-6
   Schwartzentruber J, 2012, NATURE, V482, P226, DOI [10.1038/nature10833, 10.1007/s13311-012-0137-6]
   Sharif S, 2006, J CLIN ONCOL, V24, P2570, DOI 10.1200/JCO.2005.03.8349
   Singleton WG, 2017, INT J NANOMED, V12, P1385, DOI 10.2147/IJN.S125300
   SPOSTO R, 1989, J NEURO-ONCOL, V7, P165, DOI 10.1007/BF00165101
   Sturm D, 2016, CELL, V164, P1060, DOI 10.1016/j.cell.2016.01.015
   Sturm D, 2014, NAT REV CANCER, V14, P92, DOI 10.1038/nrc3655
   Sturm D, 2012, CANCER CELL, V22, P425, DOI 10.1016/j.ccr.2012.08.024
   Sun Y, 2017, NEURO-ONCOLOGY, V19, P774, DOI 10.1093/neuonc/now261
   Tatevossian RG, 2010, ACTA NEUROPATHOL, V120, P731, DOI 10.1007/s00401-010-0763-1
   Taylor KR, 2014, NAT GENET, V46, P457, DOI 10.1038/ng.2925
   Williams N L, 2016, AM J CLIN ONCOL
   Worst BC, 2016, EUR J CANCER, V65, P91, DOI 10.1016/j.ejca.2016.06.009
   Wu G, 2014, NAT GENET, V46, P444, DOI 10.1038/ng.2938
   Wu G, 2012, NAT GENET, V44, P251, DOI 10.1038/ng.1102
   Zhang JH, 2013, NAT GENET, V45, P602, DOI 10.1038/ng.2611
   Zhou ZP, 2017, CURR NEUROPHARMACOL, V15, P116, DOI 10.2174/1570159X14666160614093615
NR 79
TC 13
Z9 13
U1 0
U2 1
PU THIEME MEDICAL PUBL INC
PI NEW YORK
PA 333 SEVENTH AVE, NEW YORK, NY 10001 USA
SN 0271-8235
EI 1098-9021
J9 SEMIN NEUROL
JI Semin. Neurol.
PD FEB
PY 2018
VL 38
IS 1
BP 121
EP 130
DI 10.1055/s-0038-1635106
PG 10
WC Clinical Neurology
WE Science Citation Index Expanded (SCI-EXPANDED)
SC Neurosciences & Neurology
GA FZ9IQ
UT WOS:000427924400016
PM 29548059
DA 2023-08-27
ER

PT J
AU Mobine, HR
   Baker, AB
   Wang, LB
   Wakimoto, H
   Jacobsen, KC
   Seidman, CE
   Seidman, JG
   Edelman, ER
AF Mobine, Hector R.
   Baker, Aaron B.
   Wang, Libin
   Wakimoto, Hiroko
   Jacobsen, Kurt C.
   Seidman, Christine E.
   Seidman, J. G.
   Edelman, Elazer R.
TI Pheochromocytoma-Induced Cardiomyopathy is Modulated by the Synergistic
   Effects of Cell-Secreted Factors
SO CIRCULATION-HEART FAILURE
LA English
DT Article
DE norepinephrine; cardiomyopathy; catecholamines; heartfailure
ID INDUCED MYOCARDIAL NECROSIS; PC12 CELLS; MODELS; ACTIVATION; DIAGNOSIS;
   RELEASE; PATHWAY
AB Background-Pheochromocytomas are rare tumors derived from the chromaffin cells of the adrenal medulla. Although these tumors have long been postulated to induce hypertension and cardiomyopathy through the hypersecretion of catecholamines, catecholamines alone may not fully explain the profound myocardial remodeling induced by these tumors. We sought to determine whether changes in myocardial function in pheochromocytoma-induced cardiomyopathy result solely from catecholamines secretion or from multiple pheochromocytoma-derived factors.
   Methods and Results-Isolated cardiomyocytes incubated with pheochromocytoma-conditioned growth media contracted at a higher frequency than cardiomyocytes incubated with norepinephrine (NE) only. Sprague-Dawley rats and black-6 mice were implanted with agarose-encapsulated pheochromocytoma (PC12) cells, dihydroxyphenylalanine decarboxylase knock-out PC12 cells deficient in NE (PC12-KO), or NE-secreting pumps. PC12 cell implantation increased left ventricular dilation by 3 5 +/- 6% and 9.6 +/- 1.4% and reduced left ventricular fractional shortening by 20 +/- 3 % and 2 8 +/- 4% in rats and mice compared with animals dosed only with NE, respectively. Elimination of NE secretion in PC12-KO cells induced neither cardiac dilation (3.9% +/- 1.8% increase versus control) nor changes in (1.9% +/- 0.4% reduction) fractional shortening compared to controls.
   Conclusions-Pheochromocytomas induce a greater degree of cardiomyopathy than equivalent doses of NE, suggesting pheochromocytoma-induced cardiomyopathy is not solely mediated by NE, rather pheochromocytoma secretory factors in combination with catecholamines act synergistically to induce greater cardiac damage than catecholamines alone. (Circ Heart Fail. 2009;2:121-128.)
C1 [Mobine, Hector R.; Baker, Aaron B.; Edelman, Elazer R.] MIT, Harvard Mit Div Hlth Sci & Technol, Cambridge, MA 02139 USA.
   [Wang, Libin; Wakimoto, Hiroko; Jacobsen, Kurt C.; Seidman, Christine E.; Seidman, J. G.] Harvard Univ, Dept Genet, Sch Med, Cambridge, MA 02138 USA.
   [Seidman, Christine E.] Howard Hughes Med Inst, Chevy Chase, MD USA.
   [Edelman, Elazer R.] Harvard Univ, Brigham & Womens Hosp, Sch Med, Div Cardiovasc, Boston, MA 02115 USA.
C3 Harvard University; Massachusetts Institute of Technology (MIT); Harvard
   University; Howard Hughes Medical Institute; Harvard University; Brigham
   & Women's Hospital; Harvard Medical School
RP Mobine, HR (通讯作者)，MIT, Harvard Mit Div Hlth Sci & Technol, 77 Massachusetts Ave,E25-442, Cambridge, MA 02139 USA.
EM
RI Baker, Aaron/AAT-4635-2020
OI Edelman, Elazer/0000-0002-7832-7156
FU National Institutes of Health [R01 49039]
FX This work was supported by the National Institutes of Health (R01 49039
   to E.R.E.), a Predoctoral Fellowship (to H.R.M.), and a Philip Morris
   postdoctoral research fellowship (to A.B.B.).
CR Aukrust P, 1998, CIRCULATION, V97, P1136, DOI 10.1161/01.CIR.97.12.1136
   BLAIKLOCK RG, 1981, RES COMMUN CHEM PATH, V34, P179
   Bravo EL, 2003, ENDOCR REV, V24, P539, DOI 10.1210/er.2002-0013
   Briest W, 2001, CARDIOVASC RES, V52, P265, DOI 10.1016/S0008-6363(01)00398-4
   CHAPPEL CI, 1959, CAN J BIOCHEM PHYS, V37, P35, DOI 10.1139/y59-005
   Communal C, 1998, CIRCULATION, V98, P1329, DOI 10.1161/01.CIR.98.13.1329
   Dixon DN, 2005, IN VITRO CELL DEV-AN, V41, P197
   DOWNING SE, 1985, J MOL CELL CARDIOL, V17, P377, DOI 10.1016/S0022-2828(85)80137-1
   DOWNING SE, 1983, CIRC RES, V52, P471, DOI 10.1161/01.RES.52.4.471
   Favia G, 1998, WORLD J SURG, V22, P689, DOI 10.1007/s002689900454
   Fedak PWM, 2004, CIRCULATION, V110, P2401, DOI 10.1161/01.CIR.0000134959.83967.2D
   Fedak PWM, 2003, AM J PHYSIOL-HEART C, V284, pH626, DOI 10.1152/ajpheart.00684.2002
   FERRANS VJ, 1964, AM HEART J, V68, P71, DOI 10.1016/0002-8703(64)90242-X
   GIFFORD RW, 1985, CARDIOLOGY, V72, P126
   KAHN DS, 1969, ANN NY ACAD SCI, V156, P285, DOI 10.1111/j.1749-6632.1969.tb16735.x
   Khademhosseini A, 2005, TISSUE ENG, V11, P1797, DOI 10.1089/ten.2005.11.1797
   LAYCOCK SK, 1995, J CARDIOVASC PHARM, V26, P584, DOI 10.1097/00005344-199510000-00012
   Lehmann MH, 1998, CYTOKINE, V10, P739, DOI 10.1006/cyto.1998.0354
   MANN DL, 1992, CIRCULATION, V85, P790, DOI 10.1161/01.CIR.85.2.790
   MEHES G, 1967, ACTA PHYSIOL HUNG, V32, P175
   Meijer WG, 2003, CLIN CHEM, V49, P586, DOI 10.1373/49.4.586
   Moller JC, 2006, NEUROSCI LETT, V400, P75, DOI 10.1016/j.neulet.2006.02.018
   Ohta S, 2006, ANN NY ACAD SCI, V1073, P300, DOI 10.1196/annals.1353.034
   Reisch N, 2006, J HYPERTENS, V24, P2331, DOI 10.1097/01.hjh.0000251887.01885.54
   Roberts T, 1996, BIOMATERIALS, V17, P267, DOI 10.1016/0142-9612(96)85564-5
   RONA G, 1985, J MOL CELL CARDIOL, V17, P291, DOI 10.1016/S0022-2828(85)80130-9
   Rosenbaum J S, 1988, Am J Cardiovasc Pathol, V1, P389
   ROSENBAUM JS, 1987, J PHARMACOL EXP THER, V241, P354
   Rozen S, 2000, Methods Mol Biol, V132, P365
   SIMONS M, 1985, AM HEART J, V109, P297, DOI 10.1016/0002-8703(85)90597-6
   Specht H, 2003, MOL CELL NEUROSCI, V22, P75, DOI 10.1016/S1044-7431(02)00023-4
   Swynghedauw B, 1999, PHYSIOL REV, V79, P215, DOI 10.1152/physrev.1999.79.1.215
   Taub D D, 1994, Ther Immunol, V1, P229
   Tischler AS, 2002, ANN NY ACAD SCI, V971, P366, DOI 10.1111/j.1749-6632.2002.tb04498.x
   TSUJIMOTO G, 1984, ENDOCRINOLOGY, V114, P1272, DOI 10.1210/endo-114-4-1272
   WARREN S, 1972, CANCER, V29, P327, DOI 10.1002/1097-0142(197202)29:2<327::AID-CNCR2820290210>3.0.CO;2-3
   WHEATLEY AM, 1985, J MOL CELL CARDIOL, V17, P349, DOI 10.1016/S0022-2828(85)80134-6
   WILKENFELD C, 1992, J HEART LUNG TRANSPL, V11, P363
   Yamazaki T, 2004, EUR J HEART FAIL, V6, P41, DOI 10.1016/j.ejheart.2003.05.002
   YOSHIDA K, 1990, JPN J MED, V29, P27
NR 40
TC 17
Z9 17
U1 0
U2 2
PU LIPPINCOTT WILLIAMS & WILKINS
PI PHILADELPHIA
PA TWO COMMERCE SQ, 2001 MARKET ST, PHILADELPHIA, PA 19103 USA
SN 1941-3289
EI 1941-3297
J9 CIRC-HEART FAIL
JI Circ.-Heart Fail.
PD MAR
PY 2009
VL 2
IS 2
BP 121
EP 128
DI 10.1161/CIRCHEARTFAILURE.108.813261
PG 8
WC Cardiac & Cardiovascular Systems
WE Science Citation Index Expanded (SCI-EXPANDED)
SC Cardiovascular System & Cardiology
GA 446NM
UT WOS:000266128300007
PM 19808327
OA Green Accepted, Bronze
DA 2023-08-27
ER

PT J
AU Wolff, JEA
   Gnekow, AK
   Kortmann, RD
   Pietsch, T
   Urban, C
   Graf, N
   Kuhl, J
AF Wolff, JEA
   Gnekow, AK
   Kortmann, RD
   Pietsch, T
   Urban, C
   Graf, N
   Kuhl, J
TI Preradiation chemotherapy for pediatric patients with high-grade glioma
SO CANCER
LA English
DT Article
DE histologic grading; malignant glioma; sandwich chemotherapy; total
   resection
ID MALIGNANT GLIOMAS; GLIOBLASTOMA-MULTIFORME; BRAIN-TUMORS;
   RADIATION-THERAPY; CHILDHOOD; RESECTION; CHILDREN; EXTENT; ADULTS; TRIAL
AB BACKGROUND. To evaluate the feasibility and efficacy of intensive chemotherapy given prior to irradiation in pediatric patients with malignant glioma, the Society 3 of Pediatric Oncology in Germany started a randomized trial in 1991, The high-grade glioma strata had to be closed because Of insufficient patient accrual. The follow-up data from these patients are reported.
   METHODS. Fifty-two patients with World Health Organization (WHO) Grade 4 malignant glioma (n = 27 patients) or with WHO Grade 3 anaplastic astrocytoma (n = 25 patients) between the ages of 3 years and 17 years were available for analysis. The tumor locations were supratentorial in 42 patients, the cerebellum in 8 patients,, and the spinal cord ill 2 patients (the brainstem was excluded). Tumor surgeries were biopsy in 10 patients, partial resection in 5 patients, subtotal resection in 10 patients, and macroscopic total resection in 21 patients. Patients received either 54 grays of irradiation (n = 22 patients) followed by chemotherapy with lomustine, vincristine, and cisplatin (maintenance chemotherapy) or sandwich chemotherapy (n = 30 patients), which consisted of ifosfamide, etoposide, methotrexate, cisplatin, and cytosine arabinoside followed by irradiation.
   RESULTS. The extent of resection was the most important prognostic factor. The median survival was 5.2 years for patients who underwent tumor resection of greater than or equal to 90% compared with 1.3 years for patients who underwent less than complete resection (P < 0.0005). After undergoing macroscopic total resection, sandwich chemotherapy (n = 15 patients) resulted in better overall survival (median, 5.2 years) compared with the maintenance protocol (n = 16 patients; median survival, 1.9 years; P = 0.015). A Cox multivariate regression analysis showed better survival for female patients (P = 0.025), WHO Grade 3 disease (P = 0.016), tumor resection of greater than or equal to 90%. (P = 0.003), irradiation with greater than or equal to 54 grays (P = 0.003), and sandwich chemotherapy (P = 0.006).
   CONCLUSIONS. These data suggest that early, intensive chemotherapy increases survival rates in patients, with malignant glioma who undergo complete resection. (C) 2002 American Cancer Society.
C1 Klin St Hedwig, Hamatol Onkol Abt, D-93049 Regensburg, Germany.
   Cent Hosp Augsburg, Hosp Children & Adolescents, Dept Pediat, Augsburg, Germany.
   Univ Tubingen, Dept Radiat Oncol, Tubingen, Germany.
   Univ Bonn, Dept Neuropathol, D-5300 Bonn, Germany.
   Univ Childrens Hosp, Div Pediat Hematol Oncol, Graz, Austria.
   Univ Klin Kinder & Jugendmed, Dept Pediat Hematol Oncol, Homburg, Germany.
   Univ Wurzburg, Wurzburg, Germany.
C3 Eberhard Karls University of Tubingen; University of Bonn;
   Universitatsklinikum des Saarlandes; University of Wurzburg
RP Wolff, JEA (通讯作者)，Klin St Hedwig, Hamatol Onkol Abt, Steinmetzstr 1-3, D-93049 Regensburg, Germany.
RI Graf, Norbert/H-1328-2011; urban, christian e/F-9151-2010
OI Graf, Norbert/0000-0002-2248-323X; Wolff, Johannes/0000-0003-0278-1309
CR Brandes AA, 1996, J NEURO-ONCOL, V30, P247
   FINLAY JL, 1995, J CLIN ONCOL, V13, P112, DOI 10.1200/JCO.1995.13.1.112
   Finlay JL, 1999, CHILD NERV SYST, V15, P786, DOI 10.1007/s003810050471
   Gjerris F, 1998, CHILD NERV SYST, V14, P302, DOI 10.1007/s003810050231
   Heideman RL, 1997, CANCER-AM CANCER SOC, V80, P497
   Hjalmars U, 1999, CANCER, V85, P2077, DOI 10.1002/(SICI)1097-0142(19990501)85:9<2077::AID-CNCR27>3.0.CO;2-4
   Jeremic B, 1997, Srp Arh Celok Lek, V125, P93
   Johnson JH, 1996, CANCER INVEST, V14, P609, DOI 10.3109/07357909609076905
   Keles GE, 1999, SURG NEUROL, V52, P371, DOI 10.1016/S0090-3019(99)00103-2
   Kirby S, 1996, CAN J NEUROL SCI, V23, P123, DOI 10.1017/S0317167100038841
   KLEIHUES PC, 1993, WHO INT HISTOLOGY CL
   Kortmann RD, 2000, INT J RADIAT ONCOL, V46, P269, DOI 10.1016/S0360-3016(99)00369-7
   Kreth FW, 1999, CANCER, V86, P2117, DOI 10.1002/(SICI)1097-0142(19991115)86:10<2117::AID-CNCR33>3.3.CO;2-#
   Kuhl J, 1998, KLIN PADIATR, V210, P227, DOI 10.1055/s-2008-1043883
   NITTA T, 1995, CANCER-AM CANCER SOC, V75, P2727, DOI 10.1002/1097-0142(19950601)75:11<2727::AID-CNCR2820751115>3.0.CO;2-H
   Packer, 1996, Oncologist, V1, P381
   Packer RJ, 1999, J CLIN ONCOL, V17, P2127, DOI 10.1200/JCO.1999.17.7.2127
   PARKER FG, 1996, J NEUROSURG, V84, P42
   Poppenborg H, 1997, ANTICANCER RES, V17, P1131
   Poppenborg H, 1997, ANTICANCER RES, V17, P2073
   SHELINE GE, 1977, CANCER, V39, P873, DOI 10.1002/1097-0142(197702)39:2+<873::AID-CNCR2820390725>3.0.CO;2-Y
   SPOSTO R, 1989, J NEURO-ONCOL, V7, P165, DOI 10.1007/BF00165101
   VONDEIMLING A, 1993, BRAIN PATHOL, V3, P19
   Wisoff JH, 1998, J NEUROSURG, V89, P52, DOI 10.3171/jns.1998.89.1.0052
   Wolff JEA, 1996, KLIN PADIATR, V208, P193, DOI 10.1055/s-2008-1046473
   Wolff JEA, 2000, CANCER, V89, P2131, DOI 10.1002/1097-0142(20001115)89:10<2131::AID-CNCR14>3.0.CO;2-J
NR 26
TC 77
Z9 79
U1 0
U2 2
PU JOHN WILEY & SONS INC
PI HOBOKEN
PA 111 RIVER ST, HOBOKEN, NJ 07030 USA
SN 0008-543X
J9 CANCER
JI Cancer
PD JAN 1
PY 2002
VL 94
IS 1
BP 264
EP 271
DI 10.1002/cncr.10114
PG 8
WC Oncology
WE Science Citation Index Expanded (SCI-EXPANDED)
SC Oncology
GA 509FC
UT WOS:000173134800034
PM 11815986
OA Bronze
DA 2023-08-27
ER

PT J
AU Humbert-Claude, M
   Duc, D
   Dwir, D
   Thieren, L
   von Tobel, JS
   Begka, C
   Legueux, F
   Velin, D
   Maillard, MH
   Do, KQ
   Monnet-Tschudi, F
   Tenenbaum, L
AF Humbert-Claude, Marie
   Duc, D.
   Dwir, D.
   Thieren, L.
   von Tobel, J. Sandstrom
   Begka, C.
   Legueux, F.
   Velin, D.
   Maillard, M. H.
   Do, K. Q.
   Monnet-Tschudi, F.
   Tenenbaum, L.
TI Tollip, an early regulator of the acute inflammatory response in the
   substantia nigra
SO JOURNAL OF NEUROINFLAMMATION
LA English
DT Article
DE Tollip; Substantia nigra; Lipopolysaccharide; Neuroinflammation; iNOS;
   Oxidative stress; Cytokine; Adeno-associated viral vector; Parkinson's
   disease; Toll-like interacting protein
ID NITRIC-OXIDE SYNTHASE; PARKINSONS-DISEASE; MICROGLIAL ACTIVATION;
   INTERACTING PROTEIN; INNATE IMMUNITY; GLIAL-CELLS; LIPOPOLYSACCHARIDE;
   RECEPTOR; BRAIN; LPS
AB Background: Tollip is a ubiquitously expressed protein, originally described as a modulator of the IL-1R/TLR-NF-kappa B signaling pathways. Although this property has been well characterized in peripheral cells, and despite some evidence of its expression in the central nervous system, the role of Tollip in neuroinflammation remains poorly understood. The present study sought to explore the implication of Tollip in inflammation in the substantia nigra pars compacta, the structure affected in Parkinson's disease.
   Methods: We first investigated Tollip distribution in the midbrain by immunohistochemistry. Then, we addressed TLR4-mediated response by intra-nigral injections of lipopolysaccharide (LPS), a TLR4 agonist, on inflammatory markers in Tollip knockout (KO) and wild-type (WT) mice.
   Results: We report an unexpectedly high Tollip immunostaining in dopaminergic neurons of the mice brain. Second, intra-nigral injection of LPS led to increased susceptibility to neuroinflammation in Tollip KO compared to Tollip WT mice. This was demonstrated by a significant increase of tumor necrosis factor alpha (TNF-alpha), interleukin 1 beta (IL-1 beta), interleukin 6 (IL-6), and interferon gamma (IFN-gamma) messenger RNA (mRNA) in the midbrain of Tollip KO mice upon LPS injection. Consistently, brain rAAV viral vector transduction with a nuclear factor kappa B (NF-kappa B)-inducible reporter gene confirmed increased NF-kappa B activation in Tollip KO mice. Lastly, Tollip KO mice displayed higher inducible NO synthase (iNOS) production, both at the messenger and protein level when compared to LPS-injected WT mice. Tollip deletion also aggravated LPS-induced oxidative and nitrosative damages, as indicated by an increase of 8-oxo-2'-deoxyguanosine and nitrotyrosine immunostaining, respectively.
   Conclusions: Altogether, these findings highlight a critical role of Tollip in the early phase of TLR4-mediated neuroinflammation. As brain inflammation is known to contribute to Parkinson's disease, Tollip may be a potential target for neuroprotection.
C1 [Humbert-Claude, Marie; Duc, D.; Thieren, L.; Tenenbaum, L.] Univ Lausanne Hosp, Dept Clin Neurosci, Ctr Neurosci Res, Lab Cellular & Mol Neurotherapies, Lausanne, Switzerland.
   [Dwir, D.; Do, K. Q.] Univ Lausanne, Ctr Hosp Univ Vaudois, Ctr Psychiat Neurosci, Dept Psychiat, Lausanne, Switzerland.
   [von Tobel, J. Sandstrom; Monnet-Tschudi, F.] Univ Lausanne, Dept Physiol, Lausanne, Switzerland.
   [Begka, C.; Velin, D.; Maillard, M. H.] Univ Lausanne Hosp, Dept Med, Serv Gastroenterol & Hepatol, Lausanne, Switzerland.
   [Legueux, F.] FIRALIS SA, Huningue, France.
C3 University of Lausanne; Centre Hospitalier Universitaire Vaudois (CHUV);
   University of Lausanne; Centre Hospitalier Universitaire Vaudois (CHUV);
   University of Lausanne; University of Lausanne; Centre Hospitalier
   Universitaire Vaudois (CHUV)
RP Humbert-Claude, M (通讯作者)，Univ Lausanne Hosp, Dept Clin Neurosci, Ctr Neurosci Res, Lab Cellular & Mol Neurotherapies, Lausanne, Switzerland.
EM
RI Tenenbaum, Liliane/AAW-5685-2021; Do, Kim Q./G-6694-2016; Dwir,
   Daniella/ABB-5269-2021; Tenenbaum, Liliane/O-5917-2016
OI Tenenbaum, Liliane/0000-0003-4619-9660; Do, Kim Q./0000-0003-1968-1646;
   Dwir, Daniella/0000-0003-4931-9007; Tenenbaum,
   Liliane/0000-0003-4619-9660; Sandstrom, Jenny/0000-0003-0117-6975;
   Humbert-Claude, Marie/0000-0001-6610-1662; Duc,
   Donovan/0000-0002-5079-5783; VELIN, Dominique/0000-0001-6033-5871
FU Swiss National Research Foundation [31003A-127177, 310030_141145]; EU
   FP7 Marie Curie Industry-Academy Partnerships and Pathways grant
   [286071]; Swiss Center for Applied Human Toxicology (SCAHT); Novartis
   Foundation; San Salvatore Foundation; Emma Muschamp Foundation; Swiss
   National Science Foundation (SNF) [31003A_127177, 310030_141145] Funding
   Source: Swiss National Science Foundation (SNF)
FX This work was supported by the Swiss National Research Foundation (grant
   number 31003A-127177 to LT and grant number 310030_141145 to DV), by the
   EU FP7 Marie Curie Industry-Academy Partnerships and Pathways grant
   (contract no 286071 to LT), by the Swiss Center for Applied Human
   Toxicology (SCAHT) to FTM, and by Novartis, San Salvatore, and Emma
   Muschamp Foundations to MM.
CR Aktan F, 2004, LIFE SCI, V75, P639, DOI 10.1016/j.lfs.2003.10.042
   Arai H, 2004, J BIOL CHEM, V279, P51647, DOI 10.1074/jbc.M407328200
   Begka C, 2016, INFLAMM BOWEL DIS, V22, P1723, DOI 10.1097/MIB.0000000000000838
   Benoit M, 2008, J IMMUNOL, V181, P3733, DOI 10.4049/jimmunol.181.6.3733
   Bowman CC, 2003, GLIA, V43, P281, DOI 10.1002/glia.10256
   Brissoni B, 2006, CURR BIOL, V16, P2265, DOI 10.1016/j.cub.2006.09.062
   Brown GC, 2010, MOL NEUROBIOL, V41, P242, DOI 10.1007/s12035-010-8105-9
   Bulut Y, 2001, J IMMUNOL, V167, P987, DOI 10.4049/jimmunol.167.2.987
   Burns K, 2000, NAT CELL BIOL, V2, P346, DOI 10.1038/35014038
   Capelluto DGS, 2012, MICROBES INFECT, V14, P140, DOI 10.1016/j.micinf.2011.08.018
   Carithers LJ, 2015, BIOPRESERV BIOBANK, V13, P311, DOI 10.1089/bio.2015.0032
   Chtarto A, 2013, PLOS ONE, V8, DOI 10.1371/journal.pone.0053156
   Collins LM, 2012, NEUROPHARMACOLOGY, V62, P2154, DOI 10.1016/j.neuropharm.2012.01.028
   Cribbs DH, 2012, J NEUROINFLAMM, V9, DOI 10.1186/1742-2094-9-179
   Dalkara D, 2012, GENE THER, V19, P176, DOI 10.1038/gt.2011.163
   Dauer W, 2003, NEURON, V39, P889, DOI 10.1016/S0896-6273(03)00568-3
   David S, 2011, NAT REV NEUROSCI, V12, P388, DOI 10.1038/nrn3053
   DEXTER DT, 1987, LANCET, V2, P1219
   Didierlaurent A, 2006, MOL CELL BIOL, V26, P735, DOI 10.1128/MCB.26.3.735-742.2006
   Doi H, 2004, FEBS LETT, V571, P171, DOI 10.1016/j.febslet.2004.06.077
   Fernandez E, 2013, ANTIOXID REDOX SIGN, V19, P912, DOI 10.1089/ars.2013.5250
   Forsyth CB, 2011, PLOS ONE, V6, DOI 10.1371/journal.pone.0028032
   Gao HM, 2008, TRENDS IMMUNOL, V29, P357, DOI 10.1016/j.it.2008.05.002
   Gao HM, 2011, ENVIRON HEALTH PERSP, V119, P807, DOI 10.1289/ehp.1003013
   Garry PS, 2015, EXP NEUROL, V263, P235, DOI 10.1016/j.expneurol.2014.10.017
   Gerhard A, 2006, NEUROBIOL DIS, V21, P404, DOI 10.1016/j.nbd.2005.08.002
   Hamza TH, 2010, NAT GENET, V42, P781, DOI 10.1038/ng.642
   Herrera AJ, 2000, NEUROBIOL DIS, V7, P429, DOI 10.1006/nbdi.2000.0289
   Hirsch EC, 1998, ANN NEUROL, V44, pS115, DOI 10.1002/ana.410440717
   Hirsch Etienne C, 2012, Parkinsonism Relat Disord, V18 Suppl 1, pS210, DOI 10.1016/S1353-8020(11)70065-7
   Hirsch EC, 2009, LANCET NEUROL, V8, P382, DOI 10.1016/S1474-4422(09)70062-6
   Hunot S, 1996, NEUROSCIENCE, V72, P355, DOI 10.1016/0306-4522(95)00578-1
   Ji KA, 2007, GLIA, V55, P1577, DOI 10.1002/glia.20571
   Kannarkat GT, 2015, NPJ PARKINSON DIS, V1, DOI 10.1038/npjparkd.2015.2
   Katoh Y, 2004, J BIOL CHEM, V279, P24435, DOI 10.1074/jbc.M400059200
   Knott C, 2000, MOL CELL NEUROSCI, V16, P724, DOI 10.1006/mcne.2000.0914
   Kooy NW, 1997, CRIT CARE MED, V25, P812, DOI 10.1097/00003246-199705000-00017
   Lee HJ, 2012, J NEUROINFLAMM, V9, DOI 10.1186/1742-2094-9-271
   Li MC, 2015, J PATHOL, V237, P249, DOI 10.1002/path.4565
   Li T, 2004, MOL IMMUNOL, V41, P85, DOI 10.1016/j.molimm.2004.03.009
   Liu B, 2002, ANN NY ACAD SCI, V962, P318, DOI 10.1111/j.1749-6632.2002.tb04077.x
   Liu YX, 2008, NEUROTOXICOLOGY, V29, P864, DOI 10.1016/j.neuro.2008.02.014
   Lo U, 2014, SCI REP-UK, V4, DOI 10.1038/srep07405
   Lo YLS, 2009, MAMM GENOME, V20, P305, DOI 10.1007/s00335-009-9188-3
   Lock M, 2010, HUM GENE THER, V21, P1273, DOI 10.1089/hum.2009.223
   Lu KF, 2014, CELL, V158, P549, DOI 10.1016/j.cell.2014.05.048
   MacMicking J, 1997, ANNU REV IMMUNOL, V15, P323, DOI 10.1146/annurev.immunol.15.1.323
   Maillard MH, 2014, INFLAMM BOWEL DIS, V20, P660, DOI 10.1097/MIB.0000000000000006
   Maitra U, 2012, J IMMUNOL, V189, P1014, DOI 10.4049/jimmunol.1200857
   Makioka K, 2016, J NEUROL SCI, V365, P101, DOI 10.1016/j.jns.2016.03.035
   Mallozzi C, 2013, BBA-MOL CELL RES, V1833, P110, DOI 10.1016/j.bbamcr.2012.10.022
   Mele M, 2015, SCIENCE, V348, P660, DOI 10.1126/science.aaa0355
   Mitra S, 2013, J BIOL CHEM, V288, P25780, DOI 10.1074/jbc.M113.484170
   Monnet-Tschudi Florianne, 2011, Curr Protoc Toxicol, VChapter 12, DOI 10.1002/0471140856.tx1219s50
   MonteroMenei CN, 1996, BRAIN RES, V724, P55, DOI 10.1016/0006-8993(96)00268-5
   Moore DJ, 2003, NEUROMOL MED, V4, P95, DOI 10.1385/NMM:4:1-2:95
   Murphy S, 2000, GLIA, V29, P1, DOI 10.1002/(SICI)1098-1136(20000101)29:1<1::AID-GLIA1>3.0.CO;2-N
   Netea MG, 2002, TRENDS IMMUNOL, V23, P135, DOI 10.1016/S1471-4906(01)02169-X
   Nishimura M, 2005, BIOL PHARM BULL, V28, P886, DOI 10.1248/bpb.28.886
   Norden DM, 2015, NEUROPHARMACOLOGY, V96, P29, DOI 10.1016/j.neuropharm.2014.10.028
   Oguro A, 2011, NEUROSCI LETT, V503, P234, DOI 10.1016/j.neulet.2011.08.043
   Olson JK, 2004, J IMMUNOL, V173, P3916, DOI 10.4049/jimmunol.173.6.3916
   Pacher P, 2007, PHYSIOL REV, V87, P315, DOI 10.1152/physrev.00029.2006
   Palsson-McDermott EM, 2004, IMMUNOLOGY, V113, P153, DOI 10.1111/j.1365-2567.2004.01976.x
   Paxinos G, 1998, RAT BRAIN IN STEREOTAXIC COORDINATES, FOURTH ED., pix
   Perry VH, 2014, NAT REV NEUROL, V10, P217, DOI 10.1038/nrneurol.2014.38
   Qin LY, 2007, GLIA, V55, P453, DOI 10.1002/glia.20467
   Radi R, 2004, P NATL ACAD SCI USA, V101, P4003, DOI 10.1073/pnas.0307446101
   Richards RI, 2016, FRONT NEUROSCI-SWITZ, V10, DOI 10.3389/fnins.2016.00193
   Ruano D, 2006, NEUROSCIENCE, V140, P1157, DOI 10.1016/j.neuroscience.2006.02.073
   Russo I, 2014, J NEUROINFLAMM, V11, DOI 10.1186/1742-2094-11-52
   Saha RN, 2006, ANTIOXID REDOX SIGN, V8, P929, DOI 10.1089/ars.2006.8.929
   Sasaki T, 2016, EXP NEUROL, V275, P220, DOI 10.1016/j.expneurol.2015.11.003
   Saunders JAH, 2012, J NEUROIMMUNE PHARM, V7, P927, DOI 10.1007/s11481-012-9402-z
   SCHAPIRA AHV, 1989, LANCET, V1, P1269, DOI 10.1016/S0140-6736(89)92366-0
   SIAN J, 1994, ANN NEUROL, V36, P348, DOI 10.1002/ana.410360305
   Terawaki H, 2010, THER APHER DIAL, V14, P477, DOI 10.1111/j.1744-9987.2010.00815.x
   Trotta T, 2014, J NEUROIMMUNOL, V268, P1, DOI 10.1016/j.jneuroim.2014.01.014
   Trujillo M., 2010, NITRIC OXIDE BIOL PA, V2, P1010
   Uhlen M, 2015, SCIENCE, V347, DOI 10.1126/science.1260419
   Vartanian KB, 2011, J NEUROINFLAMM, V8, DOI 10.1186/1742-2094-8-140
   Zhang GL, 2002, J BIOL CHEM, V277, P7059, DOI 10.1074/jbc.M109537200
   Zhang Y, 2014, J NEUROSCI, V34, P11929, DOI [10.1523/JNEUROSCI.1860-14.2014, 10.11772/j.issn.1001-9081.2014.07.1929]
   Zolotukhin S, 2002, METHODS, V28, P158, DOI 10.1016/S1046-2023(02)00220-7
NR 84
TC 23
Z9 24
U1 0
U2 6
PU BMC
PI LONDON
PA CAMPUS, 4 CRINAN ST, LONDON N1 9XW, ENGLAND
SN 1742-2094
J9 J NEUROINFLAMM
JI J. Neuroinflamm.
PD DEC 7
PY 2016
VL 13
AR 303
DI 10.1186/s12974-016-0766-5
PG 15
WC Immunology; Neurosciences
WE Science Citation Index Expanded (SCI-EXPANDED)
SC Immunology; Neurosciences & Neurology
GA EG0II
UT WOS:000390714800001
PM 27927222
OA Green Published, gold
DA 2023-08-27
ER

PT J
AU Sirachainan, N
   Boongird, A
   Swangsilpa, T
   Klaisuban, W
   Lusawat, A
   Hongeng, S
AF Sirachainan, Nongnuch
   Boongird, Atthaporn
   Swangsilpa, Thiti
   Klaisuban, Wipawi
   Lusawat, Apasri
   Hongeng, Suradej
TI Reported outcomes of children with newly diagnosed high-grade gliomas
   treated with nimotuzumab and irinotecan
SO CHILDS NERVOUS SYSTEM
LA English
DT Article
DE High-grade gliomas; Chemotherapy; Outcome; Nimotuzumab
ID RECURRENT MALIGNANT GLIOMA; CHEMOTHERAPY; RADIOTHERAPY; CPT-11; TUMORS;
   TRIAL
AB Purpose The outcome of children with high-grade gliomas (HGGs) treated with radiation and adjuvant chemotherapy remains poor. The expression of epidermal growth factor receptor (EGFR) has been established in children with HGGs. This report demonstrated the outcomes of adjuvant nimotuzumab, an EGFR inhibitor, with irinotecan in pediatric HGGs.
   Methods Children with newly diagnosed HGGs were enrolled. Two weeks after surgery, nimotuzumab with a dose of 150 mg/m(2) was given every week during radiation. After completion of radiation, a 4-week cycle of nimotuzumab (150 mg/m(2)) at week 1 and 3 and irinotecan (125 mg/m(2)) at week 1, 2, and 3 was given.
   Results Sixteen patients (5 females, 11 males), with a mean +/- SD age of 8.2 +/- 3.5 years were included. Tumors were located at the supratentorial region (50.0%), infratentorial region (43.8%), and both locations (6.2%). The 5-year PFS and OS were 19.9 +/- 11.6 and 31.5 +/- 13.0%, respectively. Median times of PFS and OS were 1.8 and 1.9 years, respectively. Prognostic factors related to good outcome were the location of tumor at the supratentorial region or outside brainstem and the extension of surgery. Side effects were minimal, with grade 1 anemia in three patients and diarrhea in one patient. Although, the adjuvant regimen of nimotuzumab and irinotecan slightly increases the overall outcome when compared to the historical study, the advantages of this protocol were minimal side effect, short period of hospitalization, and improved OS in patients who received extensive surgery.
C1 [Sirachainan, Nongnuch; Hongeng, Suradej] Mahidol Univ, Fac Med, Dept Pediat, Ramathibodi Hosp, Bangkok 10400, Thailand.
   [Boongird, Atthaporn] Mahidol Univ, Dept Surg, Fac Med, Ramathibodi Hosp, Bangkok 10400, Thailand.
   [Swangsilpa, Thiti] Mahidol Univ, Dept Radiol, Fac Med, Ramathibodi Hosp, Bangkok 10400, Thailand.
   [Klaisuban, Wipawi] Mahidol Univ, Dept Pathol, Fac Med, Ramathibodi Hosp, Bangkok 10400, Thailand.
   [Lusawat, Apasri] Minist Publ Hlth, Dept Pediat Neurol, Prasat Neurol Inst, Dept Med Serv, Bangkok 10400, Thailand.
C3 Mahidol University; Mahidol University; Mahidol University; Mahidol
   University; Ministry of Public Health - Thailand
RP Hongeng, S (通讯作者)，Mahidol Univ, Fac Med, Dept Pediat, Ramathibodi Hosp, Bangkok 10400, Thailand.
EM
OI Hongeng, Suradej/0000-0002-7376-9393
FU Cancer Center of Faculty of Medicine Ramathibodi Hospital; Faculty of
   Medicine Ramathibodi Hospital, Mahidol University, Bangkok
FX The authors thank other physicians including Samart Pakakasama MD, Ake
   Hansasuta MD, Usanarat Anurathapan MD, Watinee Sanpote MD, Surapong
   Lertthammakiat MD, Peerapong Lueangapapong MD, Artit Jinawath MD and
   Noppadol Larbcharoensub MD, and paramedical personnel who have been
   involved in patient treatment and funding support from the Cancer Center
   of Faculty of Medicine Ramathibodi Hospital. Nongnuch Sirachainan is a
   recipient of the Career Development Award from the Faculty of Medicine
   Ramathibodi Hospital, Mahidol University, Bangkok.
CR [Anonymous], 2006, COMMON TERMINOLOGY C
   Bode U, 2007, J CLIN ONCOL, V25
   Broniscer A, 2006, J NEURO-ONCOL, V76, P313, DOI 10.1007/s11060-005-7409-5
   Cabanas R, 2014, CANCER BIOTHER RADIO, V29, P173, DOI 10.1089/cbr.2013.1591
   Chamberlain MC, 2002, J NEURO-ONCOL, V56, P183, DOI 10.1023/A:1014532202188
   Cloughesy TF, 2003, CANCER-AM CANCER SOC, V97, P2381, DOI 10.1002/cncr.11306
   Friedman HS, 1999, J CLIN ONCOL, V17, P1516, DOI 10.1200/JCO.1999.17.5.1516
   Gottardo NG, 2008, J CHILD NEUROL, V23, P1149, DOI 10.1177/0883073808321765
   Massimino M, 2011, EXPERT OPIN BIOL TH, V11, P247, DOI 10.1517/14712598.2011.546341
   Pollack IF, 2003, NEURO-ONCOLOGY, V5, P197, DOI 10.1215/S1152851703000097
   Prados MD, 2006, NEURO-ONCOLOGY, V8, P189, DOI 10.1215/15228517-2005-010
   Qaddoumi I, 2009, CANCER-AM CANCER SOC, V115, P5761, DOI 10.1002/cncr.24663
   Santisteban M, 2009, J NEURO-ONCOL, V92, P165, DOI 10.1007/s11060-008-9749-4
   SPOSTO R, 1989, J NEURO-ONCOL, V7, P165, DOI 10.1007/BF00165101
   Solomon MT, 2014, CANCER BIOL THER, V15, P504, DOI 10.4161/cbt.28021
   Solomon MT, 2013, BMC CANCER, V13, DOI 10.1186/1471-2407-13-299
   Wolff JEA, 2010, CANCER-AM CANCER SOC, V116, P705, DOI 10.1002/cncr.24730
NR 17
TC 8
Z9 8
U1 0
U2 5
PU SPRINGER
PI NEW YORK
PA 233 SPRING ST, NEW YORK, NY 10013 USA
SN 0256-7040
EI 1433-0350
J9 CHILD NERV SYST
JI Childs Nerv. Syst.
PD JUN
PY 2017
VL 33
IS 6
BP 893
EP 897
DI 10.1007/s00381-017-3409-y
PG 5
WC Clinical Neurology; Pediatrics; Surgery
WE Science Citation Index Expanded (SCI-EXPANDED)
SC Neurosciences & Neurology; Pediatrics; Surgery
GA EZ7NI
UT WOS:000404911800006
PM 28439659
DA 2023-08-27
ER

PT J
AU Vilela, MD
   Pedrosa, HA
   Mesquita, BF
   Horiguchi, CLF
AF Vilela, Marcelo D.
   Pedrosa, Hugo AS.
   Mesquita, Braulio F.
   Horiguchi, Claudia L. F.
TI Low-Pressure Hydrocephalus and Shunt Malfunction Following a Lumbar
   Puncture in an Adult Reversed by an Epidural Blood Patch
SO WORLD NEUROSURGERY
LA English
DT Article
DE Blood patch; Hydrocephalus; Low pressure; Lumbar puncture; Shunt
   malfunction
ID CEREBRAL VENOUS OUTFLOW; SPONTANEOUS INTRACRANIAL HYPOTENSION;
   NEGATIVE-PRESSURE; ABDUCENS PALSY; CHILDREN; POSTURE; DYNAMICS; SERIES;
   BRAIN; MRI
AB BACKGROUND: Low-pressure hydrocephalus (LPH) is a relatively rare condition, and its presentation is similar to the classically seen high-pressure hydrocephalus, with headaches, cranial nerve dysfunction, ataxia, and disturbances of consciousness. Cerebral cerebrospinal fluid loss in the presence of altered brain viscoelastic properties has previously been suggested as the pathophysiologic process leading to ventriculomegaly, despite low or negative intracranial pressures and patent shunts. More recently, cerebral venous overdrainage has been proposed as a possible explanation in the pathogenesis of LPH, although its connection to lumbar punctures in patients with shunts has not been contemplated yet. The effectiveness of epidural blood patch in the management of post-lumbar puncture LPH has been shown in children but has not been reported in adults.
   CASE DESCRIPTION: Herein we detail 2 episodes of shunt malfunction in a 30-year-old female patient with a history of hydrocephalus related to a posterior fossa tumor diagnosed during childhood. In both instances, imaging studies demonstrated ventricular dilation along with perimedullary cistern enlargement and brainstem distortion, which occurred following a lumbar puncture despite a patent shunt. A lumbar blood patch was effective in both episodes, enabling resolution of the ventriculomegaly and a good outcome.
   CONCLUSIONS: A blood patch can be efficient in adults with post-lumbar puncture LPH. Some symptoms may be explained by brainstem compression caused by enlarged cerebrospinal fluid spaces at the skull base. The role of cerebral venous overdrainage in the setting of post-lumbar puncture LPH is further supported.
C1 [Vilela, Marcelo D.] Univ Washington, Dept Neurol Surg, Seattle, WA 98195 USA.
   [Vilela, Marcelo D.; Pedrosa, Hugo AS.] Mater Dei Hosp, Neurosurg, Belo Horizonte, MG, Brazil.
   [Mesquita, Braulio F.; Horiguchi, Claudia L. F.] Mater Dei Hosp, Anesthesiol, Belo Horizonte, MG, Brazil.
C3 University of Washington; University of Washington Seattle
RP Vilela, MD (通讯作者)，Univ Washington, Dept Neurol Surg, Seattle, WA 98195 USA.; Vilela, MD (通讯作者)，Mater Dei Hosp, Neurosurg, Belo Horizonte, MG, Brazil.
EM
CR Alperin N, 2005, ACT NEUR S, V95, P177
   Barami K, 2000, PEDIATR NEUROSURG, V33, P64, DOI 10.1159/000028989
   Barami K, 2016, NEUROSURG FOCUS, V41, DOI 10.3171/2016.6.FOCUS16172
   Bechard P, 2007, CAN J ANAESTH, V54, P146, DOI 10.1007/BF03022012
   Davies MJ, 2020, WORLD NEUROSURG, V137, pE242, DOI 10.1016/j.wneu.2020.01.163
   Dias MS, 1999, PEDIATR NEUROSURG, V30, P146, DOI 10.1159/000028783
   Paz RDR, 2019, WORLD NEUROSURG, V125, P6, DOI 10.1016/j.wneu.2019.01.117
   EPSTEIN HM, 1970, ANESTHESIOLOGY, V32, P332, DOI 10.1097/00000542-197004000-00007
   Faltings L, 2018, CUREUS J MED SCIENCE, V10, DOI 10.7759/cureus.3633
   Fichtner J, 2012, NEUROCRIT CARE, V16, P444, DOI 10.1007/s12028-012-9702-4
   Filippidis AS, 2011, J NEUROSURG, V115, P1031, DOI 10.3171/2011.6.JNS101504
   Franzini A, 2010, J NEUROSURG, V112, P300, DOI 10.3171/2009.6.JNS09415
   Gisolf J, 2004, J PHYSIOL-LONDON, V560, P317, DOI 10.1113/jphysiol.2004.070409
   GRABB PA, 1994, NEUROSURGERY, V35, P406, DOI 10.1227/00006123-199409000-00007
   GRADY MS, 1986, J NEUROSURG, V65, P199, DOI 10.3171/jns.1986.65.2.0199
   Hamilton MG, 2012, ACTA NEUROCHIR SUPPL, V113, P155, DOI 10.1007/978-3-7091-0923-6_31
   INSEL TR, 1980, NEW ENGL J MED, V303, P703
   Lesniak MS, 2002, BRIT J NEUROSURG, V16, P555
   LUCE JM, 1982, J APPL PHYSIOL, V53, P1496, DOI 10.1152/jappl.1982.53.6.1496
   MCCULLOUGH DC, 1986, NEUROSURGERY, V19, P617, DOI 10.1227/00006123-198610000-00019
   Nardone R, 2016, J CLIN NEUROL, V12, P21, DOI 10.3988/jcn.2016.12.1.21
   OLSEN KS, 1987, PAIN, V30, P293, DOI 10.1016/0304-3959(87)90017-0
   PANG DL, 1994, NEUROSURGERY, V35, P643, DOI 10.1227/00006123-199410000-00010
   Poca MA, 2006, NEUROSURGERY, V58, P899, DOI 10.1227/01.NEU.0000209915.16235.6D
   PORTNOY HD, 1994, CHILD NERV SYST, V10, P29, DOI 10.1007/BF00313582
   Preuss M, 2013, CHILD NERV SYST, V29, P2307, DOI 10.1007/s00381-013-2173-x
   Preuss M, 2013, CHILD NERV SYST, V29, P1263, DOI 10.1007/s00381-013-2159-8
   REKATE HL, 1992, PEDIATR NEUROSURG, V18, P257, DOI 10.1159/000120672
   REKATE HL, 1988, PEDIATR NEUROSCI, V14, P77, DOI 10.1159/000120367
   Seinfeld Joshua, 2007, Neurosurgery, V61, pE175
   Smalley ZS, 2017, NEUROSURGERY, V80, P439, DOI 10.1093/neuros/nyw046
   Valdueza JM, 2000, LANCET, V355, P200
   VASSILYADI M, 1995, J NEUROSURG, V83, P486, DOI 10.3171/jns.1995.83.3.0486
   Williams J, 2017, SEIZURE-EUR J EPILEP, V50, P189, DOI 10.1016/j.seizure.2017.06.024
NR 34
TC 0
Z9 2
U1 1
U2 2
PU ELSEVIER SCIENCE INC
PI NEW YORK
PA STE 800, 230 PARK AVE, NEW YORK, NY 10169 USA
SN 1878-8750
EI 1878-8769
J9 WORLD NEUROSURG
JI World Neurosurg.
PD NOV
PY 2020
VL 143
BP 56
EP 61
DI 10.1016/j.wneu.2020.07.134
PG 6
WC Clinical Neurology; Surgery
WE Science Citation Index Expanded (SCI-EXPANDED)
SC Neurosciences & Neurology; Surgery
GA OS2BR
UT WOS:000589970900013
PM 32712403
OA Green Published, Bronze
DA 2023-08-27
ER

PT J
AU Zhang, S
   Gong, YL
   Liang, Y
   Wang, BQ
   Gao, W
   Xu, QY
AF Zhang, Shu
   Gong, Yulin
   Liang, Yu
   Wang, Boqian
   Gao, Wei
   Xu, Qianyun
TI Cyclophosphamide inhibits the progression of Meniere's disease by
   reducing the generation of circulating immune complex
SO EXPERIMENTAL AND THERAPEUTIC MEDICINE
LA English
DT Article
DE circulating immune complex; cyclophosphamide; heat shock protein 70;
   Meniere's disease; tumor necrosis factor-alpha
ID ENDOLYMPHATIC HYDROPS; AUGMENTATION
AB Endolymphatic hydrops is a characteristic pathological manifestation of Meniere's disease (MD) that has been previously associated with autoimmunity. Interest in the circulating immune complex (CIC) has increased due to its reported role in the occurrence of MD. The present study aimed to investigate the potential value of serum CIC concentration in the diagnosis of MD and the therapeutic potential of cyclophosphamide (CTX) for the treatment of MD. In the present study, guinea pigs were immunized with isologous crude inner ear antigens to establish an autoimmune MD model. Pure tone audiometry, Vestibular-evoked myogenic potential test, electrocochleography test and auditory brainstem response was applied in this study for assessing the severity of MD in guinea pigs. ELISA was applied to measure CIC, tumor necrosis factor alpha (TNF-alpha) and heat shock protein 70 (HSP70) expression levels in the serum samples of different groups of patients. Western blotting was applied to detect the protein expression of HSP70 in inner ear tissues in guinea pigs. Hematoxylin and eosin staining was applied to visualize the spiral ganglions in spiral ganglions models. CIC expression in the inner ear was detected by immunohistochemistry. In vivo experiments were performed to confirm the therapeutic effects of CTX in MD. Serum concentrations of CIC, TNF-alpha and HSP70 were found to be significantly higher in patients with MD, which were also associated with increases in hearing classification and the severity of endolymphatic hydrops. Using a guinea pig MD model, ELISA results revealed significantly increased serum CIC, TNF-alpha and HSP70 concentrations compared with those in the control group. ABR results showed that the thresholds in the CTX group were notably decreased compared with that in the dexamethasone group, whereas CIC concentrations in the serum were reduced following dexamethasone and CTX treatments compared with those after saline treatment. In the inner ear tissues, the CIC concentration in CTX group was lower than that in the dexamethasone group. Similarly, reductions in HSP70 and TNF-alpha concentrations was also observed in a similar manner. Immunohistochemistry staining found notably lower CIC deposition in the inner ear tissues following CTX treatment than that in dexamethasone group. Taken together, higher CIC expression can be used as a biomarker for MD diagnosis. The efficacy of CTX in MD was found to be higher compared with that in dexamethasone, which may be associated with the effective inhibition of CIC, HSP70 and TNF-alpha generation.
C1 [Zhang, Shu; Wang, Boqian; Gao, Wei; Xu, Qianyun] Inner Mongolia Med Univ, Dept Otolaryngol, Affiliated Hosp, 1 Tongdao North St, Hohhot 010110, Inner Mongolia, Peoples R China.
   [Gong, Yulin] Inner Mongolia Med Univ, Dept Rheumatol & Immunol, Affiliated Hosp, Hohhot 010110, Inner Mongolia, Peoples R China.
   [Liang, Yu] Inner Mongolia Med Univ, Comp Informat & Network Management Ctr, Affiliated Hosp, Hohhot 010110, Inner Mongolia, Peoples R China.
C3 Inner Mongolia Medical University; Inner Mongolia Medical University;
   Inner Mongolia Medical University
RP Zhang, S (通讯作者)，Inner Mongolia Med Univ, Dept Otolaryngol, Affiliated Hosp, 1 Tongdao North St, Hohhot 010110, Inner Mongolia, Peoples R China.
EM
FU Natural Science Foundation of Inner Mongolia Autonomous Region
   [2019MS08123]; Inner Mongolia Medical University Youth Innovation Fund
   project [YKD2017QNCX064]
FX The present study was supported by the Natural Science Foundation of
   Inner Mongolia Autonomous Region (grant no. 2019MS08123) and Inner
   Mongolia Medical University Youth Innovation Fund project (grant no.
   YKD2017QNCX064).
CR Anft D, 2001, HNO, V49, P102, DOI 10.1007/s001060050717
   BERD D, 1982, CANCER RES, V42, P4862
   Bodmer D, 2007, OTOL NEUROTOL, V28, P1140, DOI 10.1097/MAO.0b013e31815aea05
   Caulley L, 2018, SEMIN ARTHRITIS RHEU, V48, P141, DOI 10.1016/j.semarthrit.2017.11.008
   Ciorba A, 2018, INT J IMMUNOPATH PH, V32, DOI 10.1177/2058738418808680
   Cooper MW, 2020, LARYNGOSCOPE, V130, P2738, DOI 10.1002/lary.28647
   Derebery MJ, 2010, OTOLARYNG CLIN N AM, V43, P1047, DOI 10.1016/j.otc.2010.05.004
   DEREBERY MJ, 1991, LARYNGOSCOPE, V101, P225
   Di Berardino F, 2018, J AM COLL NUTR, V37, P149, DOI 10.1080/07315724.2017.1364181
   DORNHOFFER JL, 1993, LARYNGOSCOPE, V103, P1027
   Editorial Board of Chinese Journal of Otorhinolaryngology Head and Neck Surgery and Society of Otorhinolaryngology Head and Neck Surgery Chinese Medical Association Guideline of diagnosis and treatment of Meniere disease, 2017, ZHONGHUA ER BI YAN H, V52, P167, DOI [10.3760/cma.j.issn.1673-0860.2017.03.002, DOI 10.3760/CMA.J.ISSN.1673-0860.2017.03.002]
   Emadi A, 2009, NAT REV CLIN ONCOL, V6, P638, DOI 10.1038/nrclinonc.2009.146
   Gazquez I, 2011, PLOS ONE, V6, DOI 10.1371/journal.pone.0026759
   Greco A, 2012, AUTOIMMUN REV, V11, P731, DOI 10.1016/j.autrev.2012.01.004
   Gurkov R, 2012, OTOL NEUROTOL, V33, P1040, DOI 10.1097/MAO.0b013e31825d9a95
   Guo S Y, 2018, Zhonghua Er Bi Yan Hou Tou Jing Wai Ke Za Zhi, V53, P953, DOI 10.3760/cma.j.issn.1673-0860.2018.12.017
   Gupta S, 2018, EBIOMEDICINE, V37, P366, DOI 10.1016/j.ebiom.2018.10.038
   Hallpike C S, 1938, Proc R Soc Med, V31, P1317
   HARADA T, 1984, ACTA OTO-LARYNGOL, V97, P450, DOI 10.3109/00016488409132920
   Helling K, 2007, LARYNGOSCOPE, V117, P2244, DOI 10.1097/MLG.0b013e3181453a3c
   Hornibrook J, 2017, FRONT NEUROSCI-SWITZ, V11, DOI 10.3389/fnins.2017.00301
   HSU L, 1990, ANN OTO RHINOL LARYN, V99, P535, DOI 10.1177/000348949009900707
   Ihler F, 2012, PLOS ONE, V7, DOI 10.1371/journal.pone.0039086
   Kilpatrick J K, 2000, Ear Nose Throat J, V79, P82
   Kim DY, 2018, SENSORS-BASEL, V18, DOI 10.3390/s18113651
   Kim J, 2017, AUSTRALAS J DERMATOL, V58, P5, DOI 10.1111/ajd.12406
   Kim SH, 2014, PLOS ONE, V9, DOI 10.1371/journal.pone.0111039
   Kraaij T, 2019, LUPUS SCI MED, V6, DOI 10.1136/lupus-2019-000326
   Lopez-Escamez JA, 2011, BMC MED GENET, V12, DOI 10.1186/1471-2350-12-2
   Lu Ling, 2008, Zhonghua Er Bi Yan Hou Tou Jing Wai Ke Za Zhi, V43, P596
   Mao Zhongyao, 2014, Lin Chung Er Bi Yan Hou Tou Jing Wai Ke Za Zhi, V28, P964
   Martin-Sanz E, 2015, LARYNGOSCOPE, V125, P1183, DOI 10.1002/lary.25057
   Mathews J, 2003, J LARYNGOL OTOL, V117, P212, DOI 10.1258/002221503321192548
   Matsuoka H, 2002, ANN OTO RHINOL LARYN, V111, P587, DOI 10.1177/000348940211100704
   Nakashima T, 2009, ACTA OTO-LARYNGOL, V129, P5, DOI 10.1080/00016480902729827
   Oh S, 2019, CANCERS, V11, DOI 10.3390/cancers11101534
   Pai I, 2020, J LARYNGOL OTOL, V134, P302, DOI 10.1017/S0022215120000626
   Peneda JF, 2019, ACTA OTORRINOLAR ESP, V70, P97, DOI 10.1016/j.otorri.2017.08.008
   Perry M, 1998, AUST VET J, V76, P286, DOI 10.1111/j.1751-0813.1998.tb10161.x
   Ren HM, 2015, INT J CLIN EXP MED, V8, P6016
   SALVIDIO G, 1986, CLIN EXP RHEUMATOL, V4, P281
   Savastano M, 2007, EUR ARCH OTO-RHINO-L, V264, P15, DOI 10.1007/s00405-006-0147-2
   Sun Y, 2018, CLIN EXP IMMUNOL, V192, P33, DOI 10.1111/cei.13083
   Tamai S., 2010, NIHON RINSHO, V68, pS105
   Tsuchida Y, 2021, MOD RHEUMATOL, V31, P1004, DOI 10.1080/14397595.2020.1856020
   Welgampola MS, 2005, NEUROLOGY, V64, P1682, DOI 10.1212/01.WNL.0000161876.20552.AA
   Wright Tony, 2015, BMJ Clin Evid, V2015
   Young YH, 2002, LARYNGOSCOPE, V112, P509, DOI 10.1097/00005537-200203000-00019
   Zheng NB, 2020, HUM VACC IMMUNOTHER, V16, P2333, DOI 10.1080/21645515.2019.1710413
NR 49
TC 1
Z9 1
U1 0
U2 5
PU SPANDIDOS PUBL LTD
PI ATHENS
PA POB 18179, ATHENS, 116 10, GREECE
SN 1792-0981
EI 1792-1015
J9 EXP THER MED
JI Exp. Ther. Med.
PD OCT
PY 2021
VL 22
IS 4
AR 1177
DI 10.3892/etm.2021.10611
PG 12
WC Medicine, Research & Experimental
WE Science Citation Index Expanded (SCI-EXPANDED)
SC Research & Experimental Medicine
GA UJ4VN
UT WOS:000691284800001
PM 34504622
OA gold, Green Published
DA 2023-08-27
ER

PT J
AU Takahashi, K
   Yoshinoya, A
   Murakami, O
   Totsune, K
   Shibahara, S
AF Takahashi, K
   Yoshinoya, A
   Murakami, O
   Totsune, K
   Shibahara, S
TI Secretion of endothelin-1 and adrenomedullin by SW-13 human
   adrenocortical carcinoma cells
SO JOURNAL OF CARDIOVASCULAR PHARMACOLOGY
LA English
DT Article; Proceedings Paper
CT 6th International Symposium on Endothelin
CY OCT 10-14, 1999
CL MONTREAL, CANADA
DE endothelin (ET); adrenomedullin (ADM); adrenal cortex; SW-13; adrenal
   carcinoma
ID IMMUNOREACTIVE-ENDOTHELIN; MESSENGER-RNA; GROWTH-FACTOR; LINES;
   EXPRESSION; RECEPTORS
AB The adrenal medulla and pheochromocytomas are known to secrete various neuropeptides and vasoactive peptides. On the other hand, the production and secretion of peptides by adrenocortical tumors have not been studied in detail. The study reported, here therefore set out to examine these two functions for two vasoactive peptides, endothelin-1 (ET-1) and adrenomedullin (ADM) in SW-13 human adrenocortical carcinoma cells by radioimmunoassay and Northern blot analysis. Both immunoreactive ET (irET) and irADM were detected in the culture medium of SW-13 cells. Northern blot analysis showed the expression of ET-1 and ADM mRNAs in SW-13 cells. On the other hand, no significant amounts of calcitonin-gene-related peptide, corticotropin-releasing-hormone, neuropeptide Y or urocortin were secreted by SW-13 cells. This study has shown that ET-1 and ADM are the two unique vasoactive peptides that are produced and secreted by adrenocortical carcinoma cells.
C1 Tohoku Univ, Sch Med, Dept Mol Biol & Appl Physiol, Aoba Ku, Sendai, Miyagi 9808575, Japan.
   Tohoku Univ, Sch Med, Dept Internal Med 2, Sendai, Miyagi 9808575, Japan.
C3 Tohoku University; Tohoku University
RP Takahashi, K (通讯作者)，Tohoku Univ, Sch Med, Dept Mol Biol & Appl Physiol, Aoba Ku, 2-1 Seiryo Machi, Sendai, Miyagi 9808575, Japan.
RI Shibahara, Shigeki/M-3644-2014; Takahashi, Kazuhiro/A-4887-2010
OI Takahashi, Kazuhiro/0000-0001-8943-1875
CR KOMURO I, 1988, FEBS LETT, V238, P249, DOI 10.1016/0014-5793(88)80489-7
   KUSUHARA M, 1990, CANCER RES, V50, P3257
   LEIBOVITZ A, 1973, JNCI-J NATL CANCER I, V51, P691
   Miller MJ, 1996, J BIOL CHEM, V271, P23345
   Nussdorfer GG, 1997, INT REV CYTOL, V171, P267
   Samson WK, 1998, FRONT NEUROENDOCRIN, V19, P100, DOI 10.1006/frne.1998.0164
   SHICHIRI M, 1991, J CLIN INVEST, V87, P1867, DOI 10.1172/JCI115210
   Takahashi K, 1998, PEPTIDES, V19, P1719, DOI 10.1016/S0196-9781(98)00128-4
   TAKAHASHI K, 1990, GASTROENTEROLOGY, V99, P1660, DOI 10.1016/0016-5085(90)90472-D
   TAKAHASHI K, 1993, CLIN SCI, V84, P47, DOI 10.1042/cs0840047
   TAKAHASHI K, 1991, J CLIN ENDOCR METAB, V72, P693, DOI 10.1210/jcem-72-3-693
NR 11
TC 8
Z9 9
U1 0
U2 1
PU LIPPINCOTT WILLIAMS & WILKINS
PI PHILADELPHIA
PA 530 WALNUT ST, PHILADELPHIA, PA 19106-3621 USA
SN 0160-2446
J9 J CARDIOVASC PHARM
JI J. Cardiovasc. Pharmacol.
PY 2000
VL 36
SU 1
BP S393
EP S394
PG 2
WC Cardiac & Cardiovascular Systems; Pharmacology & Pharmacy
WE Conference Proceedings Citation Index - Science (CPCI-S); Science Citation Index Expanded (SCI-EXPANDED)
SC Cardiovascular System & Cardiology; Pharmacology & Pharmacy
GA 368RF
UT WOS:000090130800113
PM 11078430
DA 2023-08-27
ER

PT J
AU Ogiwara, H
   Bowman, RM
   Tomita, T
AF Ogiwara, Hideki
   Bowman, Robin M.
   Tomita, Tadanori
TI Long-term Follow-up of Pediatric Benign Cerebellar Astrocytomas
SO NEUROSURGERY
LA English
DT Article
DE Astrocytoma; Cerebellum; Prognostic factors; Recurrence; Spontaneous
   regression
ID JUVENILE PILOCYTIC ASTROCYTOMA; LOW-GRADE ASTROCYTOMA;
   RADIATION-THERAPY; SPONTANEOUS REGRESSION; POSTERIOR-FOSSA; CHILDHOOD;
   CHILDREN; RADIOTHERAPY; IRRADIATION; MANAGEMENT
AB BACKGROUND: The long-termprognosis of cerebellar astrocytomas needs to be reviewed.
   OBJECTIVE: To elucidate the factors influencing tumor recurrence or progression and to determine how long these patient with cerebellar astrocytomas require surveillance with neuroimaging.
   METHODS: A retrospective review of 101 children surgically treated for a cerebellar astrocytoma and followed up for > 10 years was performed.
   RESULTS: Mean follow-up was 18.4 years. Total resection confirmed by postoperative imaging was performed in 51 patients (50.5%; group A). Twenty-three patients (22.8%) had surgical total resection; however, equivocal residual tumor was found on postoperative imaging (group B). Subtotal resection leaving a portion of brainstem or cerebellar peduncle was performed in 27 patients (26.7; group C). Of these 50 residual tumors, 16 (32%) showed spontaneous regression and 8 (16%) showed arrested growth. Radiographic recurrence or progression was noted in 29 patients (28.7%). Only 3 of 51 patients (5.9%) of group A with total resection had recurrence, whereas 26 of 50 residual tumors (52%; groups B and C) progressed. The only factor affecting recurrence or progression by multivariate analysis was the extent of surgical resection. All tumor recurrence or progression except for 1 (96.6%) occurred within 8 years from the original surgery (range, 2-132 months).
   CONCLUSIONS: Overall prognosis of cerebellar astrocytomas is good; the 10-year survival rate was 100% and recurrence- or progression-free rate was 71.3% in our cohort. Almost half of residual tumors showed spontaneous regression or arrested growth in the long term. Eight to 10 years is considered to be a reasonable follow-up period by neuroimaging.
C1 [Ogiwara, Hideki; Bowman, Robin M.; Tomita, Tadanori] Childrens Mem Hosp, Div Neurosurg, Chicago, IL 60614 USA.
C3 Ann & Robert H. Lurie Children's Hospital of Chicago
RP Tomita, T (通讯作者)，2300 Childrens Plaza, Chicago, IL 60614 USA.
EM
OI Bowman, Robin/0000-0003-2820-3574
CR ABDOLLAHZADEH M, 1994, CHILD NERV SYST, V10, P380, DOI 10.1007/BF00335126
   AKYOL FH, 1992, RADIOTHER ONCOL, V23, P245, DOI 10.1016/S0167-8140(92)80129-7
   AUSTIN EJ, 1988, J NEUROSURG, V68, P41, DOI 10.3171/jns.1988.68.1.0041
   BERNELL WR, 1972, J NEUROSURG, V37, P470, DOI 10.3171/jns.1972.37.4.0470
   BLOOM HJG, 1969, AMER J ROENTGENOL RA, V105, P43, DOI 10.2214/ajr.105.1.43
   BUDKA H, 1975, ACTA NEUROCHIR, V32, P139, DOI 10.1007/BF01405911
   Campbell JW, 1996, J NEURO-ONCOL, V28, P223, DOI 10.1007/bf00250201
   CONWAY PD, 1991, CANCER-AM CANCER SOC, V67, P2772, DOI 10.1002/1097-0142(19910601)67:11<2772::AID-CNCR2820671110>3.0.CO;2-#
   COX DR, 1972, J R STAT SOC B, V34, P187
   DEWIT L, 1984, ACTA RADIOL ONCOL, V23, P1
   Dirven CMF, 1997, CHILD NERV SYST, V13, P17, DOI 10.1007/s003810050033
   ELVIDGE AR, 1968, J NEUROSURG, V28, P399, DOI 10.3171/jns.1968.28.5.0399
   GARCIA DM, 1985, J NEUROSURG, V63, P382, DOI 10.3171/jns.1985.63.3.0382
   GARCIA DM, 1990, INT J RADIAT ONCOL, V18, P815, DOI 10.1016/0360-3016(90)90402-6
   GOL A, 1959, J NEUROSURG, V16, P287, DOI 10.3171/jns.1959.16.3.0287
   GRIFFIN TW, 1979, CANCER, V44, P276, DOI 10.1002/1097-0142(197907)44:1<276::AID-CNCR2820440147>3.0.CO;2-0
   GRUSZKA E, 1970, Folia Medica Cracoviensia, V12, P127
   Gunny RS, 2005, PEDIATR RADIOL, V35, P1086, DOI 10.1007/s00247-005-1546-z
   Hadjipanayis CG, 2002, J NEUROSURG, V97, P56, DOI 10.3171/jns.2002.97.1.0056
   HOJER C, 1994, ACTA NEUROCHIR, V129, P131, DOI 10.1007/BF01406492
   ILGREN EB, 1986, ACTA NEUROCHIR, V81, P11, DOI 10.1007/BF01456260
   KAPLAN EL, 1958, J AM STAT ASSOC, V53, P457, DOI 10.2307/2281868
   Kayama T, 1996, NEUROSURG REV, V19, P217, DOI 10.1007/BF00314833
   KLEIHUES P, 1993, WHO HISTOLOGICAL TYP
   KLEINMAN GM, 1978, J NEUROSURG, V49, P111, DOI 10.3171/jns.1978.49.1.0111
   LAPRAS C, 1986, CHILD NERV SYST, V2, P55
   LEIBEL SA, 1975, CANCER, V35, P1551, DOI 10.1002/1097-0142(197506)35:6<1551::AID-CNCR2820350612>3.0.CO;2-V
   LIEBNER EJ, 1964, RADIOLOGY, V82, P193, DOI 10.1148/82.2.193
   MARSA GW, 1973, CANCER-AM CANCER SOC, V32, P646, DOI 10.1002/1097-0142(197309)32:3<646::AID-CNCR2820320318>3.0.CO;2-Y
   MOROTA N, 1990, CHILD NERV SYST, V6, P155, DOI 10.1007/BF00308493
   Palma L, 2004, ACTA NEUROCHIR, V146, P581, DOI 10.1007/s00701-004-0257-9
   PALMA L, 1988, CHILD NERV SYST, V4, P296
   Pencalet P, 1999, J NEUROSURG, V90, P265, DOI 10.3171/jns.1999.90.2.0265
   Saunders DE, 2005, J NEUROSURG, V102, P172, DOI 10.3171/jns.2005.102.2.0172
   SCHNEIDER JH, 1992, NEUROSURGERY, V30, P58, DOI 10.1227/00006123-199201000-00011
   SGOUROS S, 1995, CHILD NERV SYST, V11, P89, DOI 10.1007/BF00303812
   SHAPIRO K, 1983, CHILD BRAIN, V10, P168
   Smoots DW, 1998, CHILD NERV SYST, V14, P636
   Sobel EL, 1996, NEUROSURGERY, V39, P45, DOI 10.1097/00006123-199607000-00010
   Steinbok P, 2006, CHILD NERV SYST, V22, P572, DOI 10.1007/s00381-006-0058-y
   Sutton LN, 1996, J NEUROSURG, V84, P721, DOI 10.3171/jns.1996.84.5.0721
   Tomita T, 1998, CHILD NERV SYST, V14, P537, DOI 10.1007/s003810050269
   UNDJIAN S, 1989, CHILD NERV SYST, V5, P99, DOI 10.1007/BF00571118
   Vassilyadi M, 2009, CAN J NEUROL SCI, V36, P707, DOI 10.1017/S0317167100008313
   WALLNER KE, 1988, J NEUROSURG, V69, P171, DOI 10.3171/jns.1988.69.2.0171
   WARA WM, 1977, INT J RADIAT ONCOL, V2, P549, DOI 10.1016/0360-3016(77)90166-3
NR 46
TC 45
Z9 46
U1 0
U2 6
PU LIPPINCOTT WILLIAMS & WILKINS
PI PHILADELPHIA
PA TWO COMMERCE SQ, 2001 MARKET ST, PHILADELPHIA, PA 19103 USA
SN 0148-396X
J9 NEUROSURGERY
JI Neurosurgery
PD JAN
PY 2012
VL 70
IS 1
BP 40
EP 47
DI 10.1227/NEU.0b013e31822ff0ed
PG 8
WC Clinical Neurology; Surgery
WE Science Citation Index Expanded (SCI-EXPANDED)
SC Neurosciences & Neurology; Surgery
GA 863EP
UT WOS:000298145900013
PM 21808215
DA 2023-08-27
ER

PT J
AU Pavesi, G
   Feletti, A
   Berlucchi, S
   Opocher, G
   Martella, M
   Murgia, A
   Scienza, R
AF Pavesi, G.
   Feletti, A.
   Berlucchi, S.
   Opocher, G.
   Martella, M.
   Murgia, A.
   Scienza, R.
TI Neurosurgical treatment of von Hippel-Lindau-associated
   hemangioblastomas: benefits, risks and outcome
SO JOURNAL OF NEUROSURGICAL SCIENCES
LA English
DT Article
DE Hemangioblastoma; Von Hippel-Lindau disease; Neoplasm; Microsurgery
ID CENTRAL-NERVOUS-SYSTEM; MOLECULAR-GENETIC ANALYSIS; GAMMA-KNIFE
   RADIOSURGERY; SURGICAL-MANAGEMENT; NATURAL-HISTORY; DISEASE; HEMORRHAGE;
   FEATURES; VHL; IMPACT
AB Aim. Von Hippel-Lindau (VHL) disease is a genetic syndrome predisposing to central nervous system (CNS) hemangioblastomas and several lesions in many organs. The cases of all VHL individuals operated on in the Neurosurgical Unit of Padua Hospital since year 2000 were reviewed in order to define which features lead to surgical treatment and to examine surgical outcome during postoperative follow-up.
   Methods. The authors evaluated 20 VHL subjects (7 males and 13 females, age at surgery 32 +/- 10 years) who underwent 28 operations in order to remove 48 CNS hemangioblastomas and 1 endolymphatic sac tumor. Among the 49 resected lesions, 21 (42%) were cerebellar, 9 (18%) at brainstem, 19 (38%) spinal (7 cervical, 6 dorsal, 6 at conecauda level), and 1 (2%) endolymphatic sac tumor in the petrous bone. Patients were graduated according to Karnofsky Performance Status (KPS) at admission, at discharge and during the last follow up visit. Genetic testing revealing the presence of a VHL disease-causing mutation was a prerequisite for inclusion in the study.
   Results. Nineteen individuals (95%) were symptomatic. Symptomatic hemangioblastomas were associated with a cyst or a syrinx in 22/27 circumstances (81%). Total removal, as confirmed by postoperative magnetic resonance imaging (MRI), was achieved in all but one lesion. Following surgery, at follow-up (38 +/- 20 months), patients improved their neurological status in 75% of cases, 20% remained stable and 5% worsened; 16 patients (80%) are able to carry on normal activity with or without minor symptoms, 3 patients require some grade of assistance, 1 patient died because of bronchopneumonia.
   Conclusion. VHL-associated hemangioblastomas generally affect a young adult population and can be successfully removed, either when symptomatic, or when they reach a critical volume. Microsurgery of hemangioblastomas has a favourable impact on survival and quality of life of VHL patients, although it is strongly influenced by preoperative conditions. Transient surgical complications are possible, particularly with brainstem and spinal cord hemangioblastomas.
C1 [Pavesi, G.; Berlucchi, S.; Scienza, R.] Padova Hosp, Neurosurg Operat Unit, Padua, Italy.
   [Feletti, A.] Univ Padua, Treviso Reg Hosp, Neurosurg Operat Unit, Padua, Italy.
   [Opocher, G.] Univ Padua, Endocrinol Operat Unit, Med & Surg Sci Dept, Padua, Italy.
   [Martella, M.; Murgia, A.] Univ Padua, Dept Pediat, Padua, Italy.
C3 ARQUS; University of Padua; Azienda Ospedaliera - Universita di Padova;
   ARQUS; University of Padua; ARQUS; University of Padua; ARQUS;
   University of Padua
RP Pavesi, G (通讯作者)，Padua City Hosp, Via Giustiniani 2, I-35100 Padua, Italy.
EM
RI Opocher, Giuseppe/F-3950-2014
OI Opocher, Giuseppe/0000-0002-9845-9623; pavesi,
   giacomo/0000-0002-9004-1775
CR ADEGBITE AB, 1983, NEUROSURGERY, V12, P327, DOI 10.1227/00006123-198303000-00015
   Catapano D, 2005, NEUROSURGERY, V56, P1215, DOI 10.1227/01.NEU.0000159646.15026.D6
   Conway JE, 2001, NEUROSURGERY, V48, P55, DOI 10.1097/00006123-200101000-00009
   Glasker S, 1999, J NEUROL NEUROSUR PS, V67, P758, DOI 10.1136/jnnp.67.6.758
   Glasker S, 2005, FAM CANCER, V4, P37, DOI 10.1007/s10689-004-5347-6
   Glasker S, 2005, NEUROSURGERY, V57, P71, DOI 10.1227/01.NEU.0000163250.71951.18
   GROSSNIKLAUS HE, 1992, OPHTHALMOLOGY, V99, P140
   Kaelin WG, 2002, NAT REV CANCER, V2, P673, DOI 10.1038/nrc885
   LATIF F, 1993, SCIENCE, V260, P1317, DOI 10.1126/science.8493574
   Lindau, 1926, ACTA PATH MICROBIO S, V1, P1
   LINDAU ARVID, 1927, ACTA OPHTHALMOL, V4, P193
   Lonser RR, 2003, J NEUROSURG, V98, P106, DOI 10.3171/jns.2003.98.1.0106
   Lonser RR, 2003, LANCET, V361, P2059, DOI 10.1016/S0140-6736(03)13643-4
   Maddock IR, 1996, J MED GENET, V33, P120, DOI 10.1136/jmg.33.2.120
   MAHER ER, 1991, J MED GENET, V28, P443, DOI 10.1136/jmg.28.7.443
   MAHER ER, 1990, Q J MED, V77, P1151, DOI 10.1093/qjmed/77.2.1151
   Miyagami Mitsusuke, 2004, Brain Tumor Pathol, V21, P75, DOI 10.1007/BF02484514
   NEUMANN HPH, 1992, J NEUROL NEUROSUR PS, V55, P898, DOI 10.1136/jnnp.55.10.898
   NEUMANN HPH, 1991, LANCET, V337, P1052, DOI 10.1016/0140-6736(91)91705-Y
   Niemela M, 1999, ACTA NEUROCHIR, V141, P1147, DOI 10.1007/s007010050412
   Niemela M, 1996, J NEUROSURG, V85, P591, DOI 10.3171/jns.1996.85.4.0591
   Niemela M, 2001, CLIN CANCER RES, V7, P510
   Pavesi G, 2006, NEUROLOGY, V67, P178, DOI 10.1212/01.wnl.0000223354.86636.ed
   Rajaraman C, 2004, BRIT J NEUROSURG, V18, P338, DOI 10.1080/02688690400004944
   Slater A, 2003, AM J NEURORADIOL, V24, P1570
   Tago M, 2005, J NEUROSURG, V102, P171, DOI 10.3171/jns.2005.102.s_supplement.0171
   Von Hippel E, 1911, A VONGRAEFES ARCH KL, V79, P350, DOI [10.1007/BF01966371, DOI 10.1007/BF01966371]
   WAKAI S, 1985, Neurologia Medico-Chirurgica, V25, P268, DOI 10.2176/nmc.25.268
   Wanebo JE, 2003, J NEUROSURG, V98, P82, DOI 10.3171/jns.2003.98.1.0082
   Wang CC, 2001, SURG NEUROL, V56, P218, DOI 10.1016/S0090-3019(01)00590-0
   Wang EM, 2005, J NEUROSURG, V102, P225, DOI 10.3171/jns.2005.102.s_supplement.0225
   Weil RJ, 2003, J NEUROSURG, V98, P95, DOI 10.3171/jns.2003.98.1.0095
   YU JS, 1994, J NEUROSURG, V81, P937, DOI 10.3171/jns.1994.81.6.0937
NR 33
TC 29
Z9 34
U1 0
U2 3
PU EDIZIONI MINERVA MEDICA
PI TURIN
PA CORSO BRAMANTE 83-85 INT JOURNALS DEPT., 10126 TURIN, ITALY
SN 0390-5616
J9 J NEUROSURG SCI
JI J. Neurosurg. Sci.
PD JUN
PY 2008
VL 52
IS 2
BP 29
EP 36
PG 8
WC Clinical Neurology; Surgery
WE Science Citation Index Expanded (SCI-EXPANDED)
SC Neurosciences & Neurology; Surgery
GA 371EM
UT WOS:000260814600001
PM 18500215
DA 2023-08-27
ER

PT J
AU Clements, HA
   Wilson, MSJ
   Smith, DM
AF Clements, Hollie Alice
   Wilson, Michael S. J.
   Smith, David M.
TI Incidental giant cystic pheochromocytoma: a case report and review of
   the literature
SO SCOTTISH MEDICAL JOURNAL
LA English
DT Review
DE Pheochromocytoma; cystic; metanephrines; adrenalectomy; laparoscopic
ID LAPAROSCOPIC ADRENALECTOMY; MANAGEMENT; BENIGN
AB Introduction Pheochromocytoma is a tumour arising from the adrenal medulla, which secretes catecholamines. Approximately 20% of pheochromocytomas are cystic and more likely to be asymptomatic. They should be surgically resected as all have a malignant potential and pose cardiovascular risk. Case presentation: We report the case of a 61-year-old female patient admitted electively for laparoscopic adrenalectomy for a large cystic pheochromocytoma detected incidentally. Diagnosis was confirmed preoperatively by elevated 24-h urinary metanephrines. The patient was treated preoperatively with alpha and beta blockade. Surgery was without complication; she had an uneventful postoperative recovery and no evidence of recurrence at one-year follow-up. Conclusion This case highlights the necessity of investigating for biochemical function in all adrenal lesions by measuring metanephrines, even when entirely cystic on imaging. Given the surgical and anaesthetic risk in resection of pheochromocytoma, attaining a preoperative diagnosis allows for careful preoperative planning and safe surgery.
C1 [Clements, Hollie Alice; Wilson, Michael S. J.; Smith, David M.] Ninewells Hosp, Dept Gen Surg, Dundee DD1 9SY, Scotland.
C3 University of Dundee
RP Clements, HA (通讯作者)，Ninewells Hosp, Dept Gen Surg, Dundee DD1 9SY, Scotland.
EM
OI Wilson, Michael/0000-0003-1083-4466; Clements, Hollie
   A/0000-0001-9495-7679
CR Ambati Druvtej, 2014, Can Urol Assoc J, V8, pE374, DOI 10.5489/cuaj.1719
   Andreoni C, 2008, BJU INT, V101, P345, DOI 10.1111/j.1464-410X.2007.07370.x
   Antedomenico Elena, 2005, Curr Surg, V62, P193, DOI 10.1016/j.cursur.2004.08.015
   Arcos Cristina Torres, 2009, Can Urol Assoc J, V3, pE89
   Ariton M, 2000, Endocr Pract, V6, P249
   Awada SH, 2003, SOUTH MED J, V96, P914, DOI 10.1097/01.SMJ.0000077069.95831.62
   Bellezza Guido, 2004, Arch Pathol Lab Med, V128, pe125
   Cajipe K, 2017, BMJ CASE REP, V2017
   Cavallaro G, 2015, INT J SURG, V13, P23, DOI 10.1016/j.ijsu.2014.11.023
   Almeida CEC, 2017, INT J SURG CASE REP, V30, P201, DOI 10.1016/j.ijscr.2016.12.018
   Fassnacht M, 2016, EUR J ENDOCRINOL, V175, pG1, DOI 10.1530/EJE-16-0467
   Filippou D, 2004, INTERNET J ONCOL, V2, P1
   GAGNER M, 1992, NEW ENGL J MED, V327, P1033
   GalvAo Braga C, 2014, REV PORT CARDIOL, V33, pe1
   Ginther DN, 2012, S AFR J SURG, V50, P98, DOI 10.7196/SAJS.1190
   Goldberg A, 2011, J CLIN ENDOCR METAB, V96, P2308, DOI 10.1210/jc.2011-0465
   Gupta A, 2016, UROL ANNALS, V8, P384, DOI 10.4103/0974-7796.184886
   Kakoki Katsura, 2015, BMC Res Notes, V8, P758, DOI 10.1186/s13104-015-1738-z
   Kapoor A, 2011, CUAJ-CAN UROL ASSOC, V5, P241, DOI 10.5489/cuaj.11135
   Karumanchery R, 2012, INT J SURG CASE REP, V3, P16, DOI 10.1016/j.ijscr.2011.10.006
   Klingler PJ, 2000, MAYO CLIN PROC, V75, P517
   Kondo T, 1995, Hinyokika Kiyo, V41, P669
   Korgali E, 2014, CASE REP UROL, V2014
   Lal TG, 2003, AM SURGEON, V69, P812
   Lee J, 2011, GASTROINTEST ENDOSC, V74, P712, DOI 10.1016/j.gie.2010.09.027
   Li CF, 2012, CUAJ-CAN UROL ASSOC, V6, pE267, DOI 10.5489/cuaj.11195
   Mahadevan R, 2019, CURR ISSUES TOUR, V22, P253, DOI 10.1080/13683500.2017.1375901
   Martins D, 2017, BMJ CASE REP, V2017, P1
   Melegh Z, 2002, PATHOL RES PRACT, V198, P103, DOI 10.1078/0344-0338-00194
   Mishra AK, 2001, SURG ENDOSC-ULTRAS, V15, P220, DOI 10.1007/s004640040036
   Costa SRP, 2008, SAO PAULO MED J, V126, P229, DOI 10.1590/S1516-31802008000400008
   Santarone M, 2008, J CARDIOVASC MED, V9, P971, DOI 10.2459/JCM.0b013e328303698b
   Sarveswaran V, 2015, CLIN CASE REP, V3, P64, DOI 10.1002/ccr3.149
   Sharma P. P., 2006, INDIAN J SURG, V68, P163
   Soufi M, 2012, INDIAN J SURG, V74, P504, DOI 10.1007/s12262-012-0719-x
   Sundahl N, 2016, ACTA CHIR BELG, V116, P239, DOI 10.1080/00015458.2016.1139838
   Thompson LDR, 2002, AM J SURG PATHOL, V26, P551, DOI 10.1097/00000478-200205000-00002
   Toniato A, 2007, SURGERY, V141, P723, DOI 10.1016/j.surg.2006.10.012
   Uysal E, 2017, TURK J SURG, V33, P296, DOI 10.5152/UCD.2015.3011
   Wang HL, 2015, ONCOL LETT, V10, P1444, DOI 10.3892/ol.2015.3484
   Wu JS, 2000, SURGERY, V128, P482, DOI 10.1067/msy.2000.104113
   Yalin GY, 2015, ACTA ENDOCRINOL-BUCH, V11, P195, DOI 10.4183/aeb.2015.195
   Young WF, 2000, ENDOCRIN METAB CLIN, V29, P159, DOI 10.1016/S0889-8529(05)70122-5
NR 43
TC 4
Z9 4
U1 0
U2 5
PU SAGE PUBLICATIONS LTD
PI LONDON
PA 1 OLIVERS YARD, 55 CITY ROAD, LONDON EC1Y 1SP, ENGLAND
SN 0036-9330
EI 2045-6441
J9 SCOT MED J
JI Scott. Med. J.
PD MAY
PY 2020
VL 65
IS 2
BP 64
EP 70
AR 0036933019900339
DI 10.1177/0036933019900339
EA JAN 2020
PG 7
WC Medicine, General & Internal
WE Science Citation Index Expanded (SCI-EXPANDED)
SC General & Internal Medicine
GA LN2CG
UT WOS:000506997100001
PM 31931648
DA 2023-08-27
ER

PT J
AU Zhao, Y
   Zhao, Y
   Zhang, LQ
   Feng, GD
AF Zhao, Yong
   Zhao, Yang
   Zhang, Li-Qin
   Feng, Guo-Dong
TI Postoperative infection of the skull base surgical site due to
   suppurative parotitis: A case report
SO WORLD JOURNAL OF CLINICAL CASES
LA English
DT Article
DE Postoperative infection; Lateral skull base; Suppurative parotitis;
   Vacuum sealing drainage; Two-stage surgery; Case report
ID PHEOCHROMOCYTOMA; SURGERY
AB BACKGROUND Paraganglioma occurring at the lateral skull base is a rare tumor. Surgery is the primary treatment of benign paragangliomas. Postoperative infection of the surgical site at the lateral skull base is very dangerous and hard to manage. CASE SUMMARY A 30-year-old man with a 1-year history of left-side progressive hearing loss, tinnitus, facial palsy, and choking failed conventional treatment and is the focus of this case report. Imaging revealed a mass around the left jugular foramen that was approximately 47 mm x 38 mm x 34 mm in size and had eroded the bone of the vertebral and horizontal segments of the internal carotid artery. The tumor breached the meninges and occupied the cerebella pontine region. A two-stage surgery was designed for the resection of the mass. In the first-stage, the epidural portion of the mass was removed. The abdominal fat and the temporal muscle flap were transposed within the surgical site. The surgery was successful; however, 25 d after surgery, he developed suppurative parotitis, and the infection spread to the surgical site at the skull base. Broad-spectrum antibiotics were used, and debridement was deployed. After that, the wound was cleaned daily. Five months after the first-stage surgery, the wound was still unclosed, and there was intermittent purulent exudation within the surgical site. vacuum sealing drainage (VSD) was used, and the wound healed in a month. One year after the first surgery, the second-stage of the operation was performed to remove the intracranial portion of the tumor. Recurrence of the tumor was not detected after a 6-month follow-up. CONCLUSION After a lateral skull base surgery, suppurative parotitis can spread into the operative cavity leading to infection of the surgical site. VSD can help to effectively heal the infected wound. A two-stage surgical approach offers a safer option for removing the lateral skull base paraganglioma that involves the meninges.
C1 [Zhao, Yong; Zhao, Yang; Zhang, Li-Qin; Feng, Guo-Dong] Chinese Acad Med Sci & Peking Union Med Coll, Peking Union Med Coll Hosp, Dept Otorhinolaryngol Head & Neck Surg, Beijing 100730, Peoples R China.
   [Feng, Guo-Dong] Chinese Acad Med Sci & Peking Union Med Coll, Peking Union Med Coll Hosp, Dept Otorhinolaryngol Head & Neck Surg, 1 Shuaifuyuan Wangfujing, Beijing 100730, Peoples R China.
C3 Chinese Academy of Medical Sciences - Peking Union Medical College;
   Peking Union Medical College; Peking Union Medical College Hospital;
   Chinese Academy of Medical Sciences - Peking Union Medical College;
   Peking Union Medical College; Peking Union Medical College Hospital
RP Feng, GD (通讯作者)，Chinese Acad Med Sci & Peking Union Med Coll, Peking Union Med Coll Hosp, Dept Otorhinolaryngol Head & Neck Surg, 1 Shuaifuyuan Wangfujing, Beijing 100730, Peoples R China.
EM
CR Belczak SQ, 2008, REV I MED TROP, V50, P303, DOI 10.1590/S0036-46652008000500010
   BROOK I, 1992, ARCH OTOLARYNGOL, V118, P469
   FLEISCHMANN W, 1993, UNFALLCHIRURG, V96, P488
   Grigoryan YA, 1999, SURG NEUROL, V52, P56, DOI 10.1016/S0090-3019(99)00050-6
   Huang Q, 2019, J FOOT ANKLE SURG, V58, P954, DOI 10.1053/j.jfas.2018.12.020
   LARY BG, 1964, ARCH SURG-CHICAGO, V89, P653
   Lenders JWM, 2005, LANCET, V366, P665, DOI 10.1016/S0140-6736(05)67139-5
   Nakase H, 1998, ACTA NEUROCHIR, V140, P891, DOI 10.1007/s007010050190
   Neumann HPH, 2019, NEW ENGL J MED, V381, P552, DOI 10.1056/NEJMra1806651
   Nishizawa S, 2001, J CLIN NEUROSCI, V8, P67, DOI 10.1054/jocn.2001.0881
   Pasini B, 2009, J INTERN MED, V266, P19, DOI 10.1111/j.1365-2796.2009.02111.x
   Petzina R, 2006, EUR J CARDIO-THORAC, V30, P85, DOI 10.1016/j.ejcts.2006.04.009
NR 12
TC 0
Z9 0
U1 0
U2 3
PU BAISHIDENG PUBLISHING GROUP INC
PI PLEASANTON
PA 7041 Koll Center Parkway, Suite 160, PLEASANTON, CA, UNITED STATES
SN 2307-8960
J9 WORLD J CLIN CASES
JI World J. Clin. Cases
PD MAY 26
PY 2022
VL 10
IS 15
BP 4991
EP 4997
DI 10.12998/wjcc.v10.i15.4991
PG 7
WC Medicine, General & Internal
WE Science Citation Index Expanded (SCI-EXPANDED)
SC General & Internal Medicine
GA 2E3MS
UT WOS:000812133700030
PM 35801038
OA Green Published, gold
DA 2023-08-27
ER

PT J
AU Kraker, J
AF Kraker, Jessica
TI Treatment of Anti-Ma2/Ta Paraneoplastic Syndrome
SO CURRENT TREATMENT OPTIONS IN NEUROLOGY
LA English
DT Review
ID ANTI-MA2-ASSOCIATED ENCEPHALITIS; LIMBIC ENCEPHALITIS;
   TESTICULAR-CANCER; CLINICAL ANALYSIS; ASSOCIATION; CARCINOMA; SURVIVORS;
   DISEASE; TUMOR
AB Opinion statement
   The paraneoplastic syndrome caused by Ma2/Ta antibodies alone (not in conjunction with Ma1 or Ma3 antibodies) varies in presentation from classic limbic encephalitis. The Ma2 syndrome may present with symptoms referable to the brainstem, diencephalon, and limbic system. These clinical symptoms are accompanied by MRI changes and abnormal electroencephalographic findings. It is important to recognize when the encephalitic syndrome is secondary to Ma2 paraneoplastic antibodies, as the patients improve or stabilize most often when the underlying carcinoma is treated. Treatment of the paraneoplastic syndrome begins with recognition of the symptoms, such as memory impairment, seizures, sleep disturbances, bradykinesia or hypokinesia, and eye movement abnormalities. If a primary tumor is discovered during the workup, it should be removed and treated with the most up-to-date oncologic treatment available. In addition to oncologic treatment, the syndrome may be treated with an immunosuppressant regimen to optimize the neurologic outcome. Leaving the patient untreated will result in decline and eventual death from the cancer itself or from complications of the paraneoplastic syndrome.
C1 Univ Maryland, Dept Neurol, Baltimore, MD 21201 USA.
C3 University System of Maryland; University of Maryland Baltimore
RP Kraker, J (通讯作者)，Univ Maryland, Dept Neurol, 22 S Greene St,N4W50, Baltimore, MD 21201 USA.
EM
CR Bataller L, 2004, CURR OPIN NEUROL, V17, P3, DOI 10.1097/00019052-200402000-00003
   Bennett JL, 1999, NEUROLOGY, V52, P864, DOI 10.1212/WNL.52.4.864
   BRIERLEY JB, 1960, BRAIN, V83, P357, DOI 10.1093/brain/83.3.357
   Castle J, 2006, NAT CLIN PRACT NEURO, V2, P566, DOI 10.1038/ncpneuro0287
   CORSELLIS JA, 1968, BRAIN, V91, P481, DOI 10.1093/brain/91.3.481
   Dalmau J, 2004, BRAIN, V127, P1831, DOI 10.1093/brain/awh203
   Gianelli W, 2006, FIRE MATER, V30, P333, DOI 10.1002/fam.912
   Graus F, 2004, J NEUROL NEUROSUR PS, V75, P1135, DOI 10.1136/jnnp.2003.034447
   Gultekin SH, 2000, BRAIN, V123, P1481, DOI 10.1093/brain/123.7.1481
   Mathew RM, 2007, NEUROLOGY, V68, P900, DOI 10.1212/01.wnl.0000252379.81933.80
   PETERSON K, 1992, NEUROLOGY, V42, P1931, DOI 10.1212/wnl.42.10.1931
   Schott JM, 2006, PRACT NEUROL, V6, P143
   Travis LB, 2005, JNCI-J NATL CANCER I, V97, P1354, DOI 10.1093/jnci/dji278
   Tuzun E, 2007, NEUROLOGIST, V13, P261, DOI 10.1097/NRL.0b013e31813e34a5
   van den Belt-Dusebout AW, 2007, J CLIN ONCOL, V25, P4370, DOI 10.1200/JCO.2006.10.5296
NR 15
TC 5
Z9 7
U1 0
U2 2
PU CURRENT MEDICINE GROUP
PI PHILADELPHIA
PA 400 MARKET STREET, STE 700, PHILADELPHIA, PA 19106 USA
SN 1092-8480
EI 1534-3138
J9 CURR TREAT OPTION NE
JI Curr. Treat. Options Neurol.
PD JAN
PY 2009
VL 11
IS 1
BP 46
EP 51
DI 10.1007/s11940-009-0007-7
PG 6
WC Clinical Neurology
WE Science Citation Index Expanded (SCI-EXPANDED)
SC Neurosciences & Neurology
GA 428XP
UT WOS:000264885700007
PM 19094836
DA 2023-08-27
ER

PT J
AU Gerosa, M
   Mesiano, N
   Longhi, M
   De Simone, A
   Foroni, R
   Verlicchi, A
   Zanotti, B
   Nicolato, A
AF Gerosa, Massimo
   Mesiano, Nazarena
   Longhi, Michele
   De Simone, Antonio
   Foroni, Roberto
   Verlicchi, Angela
   Zanotti, Bruno
   Nicolato, Antonio
TI Gamma Knife surgery in vestibular schwannomas: impact on the anterior
   and posterior labyrinth
SO JOURNAL OF NEUROSURGERY
LA English
DT Article; Proceedings Paper
CT 15th International Meeting of the Leksell-Gamma-Knife-Society
CY MAY, 2010
CL Athens, GREECE
SP Leksell Gamma Knife Soc
DE radiosurgery; Gamma Knife; vestibular schwannoma; labyrinth
ID ACOUSTIC NEUROMAS; RADIOSURGICAL TREATMENT; CRANIAL NERVE; HEARING-LOSS;
   PRESERVATION; PROGRESSION; MANAGEMENT; OUTCOMES; ORIGIN; TUMOR
AB Object. During the past decades, in small-to-medium size vestibular schwannomas, Gamma Knife surgery (GKS) has become a reliable therapeutic option because of either excellent local tumor control or minimal morbidity, with cranial neuropathy becoming increasingly rare. Although still insufficiently analyzed in larger cohorts of patients with long-term follow-ups, adequate chances of hearing preservation and vestibular sparing seem clinically guaranteed. However, deeper investigations are needed in this regard, expanding the number of cases and the follow-up period.
   Methods. A small group of patients with vestibular schwannomas (74 patients, including 41 men and 33 women) treated between 2003 and 2009 using GKS at the authors' institution were analyzed both before and after GKS with computerized static stabilometry and electronystagmography for balance disorders, vertigo, and ataxia on 1 side and pure tone average, vocal speech discrimination score, auditory brainstem response, and so forth for hearing impairment and tinnitus on the other side. Eligibility criteria for this prospective study included previously untreated unilateral lesions and a Gardner-Robertson hearing class of I-III. Dosimetry plans had been programmed at the lower effective dosages for these tumors (median surface dose 12.4 Gy, range 10-13 Gy), carefully avoiding even minimal toxic dosages on the most vulnerable targets: the cochlea (never > 6 Gy) and the vestibular canals (<7.5 Gy).
   Results. To date, tumor growth control rates remain satisfactory; at a mean follow-up of 50 months, the rate was 96%. The overall level of hearing preservation was 72%, with 81% having Gardner-Robertson Class 1 hearing. Tinnitus decreased, from 52% to 28% of patients (p <0.01). Significant improvements were also observed in vestibular symptoms, with computerized static stabilometry abnormalities decreasing from 62% to 32% (p <0.001) and electronystagmography abnormalities reducing from 48% to 14% (p <0.001).
   Conclusions. Using appropriate radiodosimetry planning, GKS seems to guarantee not only adequate tumor growth control rates, but also better levels of hearing preservation, with a documented, long-lasting improvement in vestibular functions. (DOI: 10.3171/2010.8.GKS101089)
C1 [Gerosa, Massimo; Longhi, Michele; De Simone, Antonio; Foroni, Roberto; Verlicchi, Angela; Nicolato, Antonio] Borgo Trento Univ Hosp, Dept Neurosurg, Verona, Italy.
   [Mesiano, Nazarena] Borgo Trento Univ Hosp, Dept ENT Surg, Verona, Italy.
   [Zanotti, Bruno] S Maria della Misericordia Univ Hosp, Neurosurg Unit, Udine, Italy.
C3 University of Verona; Azienda Ospedaliera Universitaria Integrata
   Verona; University of Verona; Azienda Ospedaliera Universitaria
   Integrata Verona; Hospital Santa Maria della Misericordia
RP Gerosa, M (通讯作者)，Univ Hosp Verona, Dept Neurosurg, Piazzale Stefani 1, I-37126 Verona, VR, Italy.
EM
OI De Simone, Antonio/0000-0003-2003-6097
CR *AC NEUR ASS, 2008, 2007 2008 PAT SURV
   ARTHURS BJ, 2010, AM J CLIN ONCOL
   Beegle RD, 2007, J NEUROSURG, V107, P913, DOI 10.3171/JNS-07/11/0913
   Bhandare N, 2010, INT J RADIAT ONCOL, V76, pS50, DOI 10.1016/j.ijrobp.2009.04.096
   Boutin P, 1998, Ann Otolaryngol Chir Cervicofac, V115, P35
   BRIDGER MWM, 1980, J LARYNGOL OTOL, V94, P1353, DOI 10.1017/S0022215100090186
   Flickinger JC, 2004, INT J RADIAT ONCOL, V60, P225, DOI 10.1016/j.ijrobp.2004.02.019
   FONCIN JF, 1979, ANN OTO-LAR CHIR C-F, V96, P11
   GEROSA M, 2002, ACOUSTIC NEUROMA CON, P129
   Iwai Y, 2003, NEUROSURGERY, V53, P282, DOI 10.1227/01.NEU.0000073416.22608.B3
   Kondziolka D, 2000, NEUROSURG CLIN N AM, V11, P651, DOI 10.1016/S1042-3680(18)30090-1
   Linskey ME, 2003, J NEUROSURG, V98, P800, DOI 10.3171/jns.2003.98.4.0800
   Litvack Zachary N, 2003, Neurosurg Focus, V14, pe3
   NEDZELSKI JM, 1986, LARYNGOSCOPE, V96, P825
   NEELY JG, 1976, LARYNGOSCOPE, V86, P984, DOI 10.1288/00005537-197607000-00012
   Niranjan Ajay, 2008, Prog Neurol Surg, V21, P192, DOI 10.1159/000156973
   Noren G, 1998, STEREOT FUNCT NEUROS, V70, P65, DOI 10.1159/000056408
   OGAWA K, 1991, ACTA OTO-LARYNGOL, P133
   Pollock BE, 2006, NEUROSURGERY, V58, P241, DOI 10.1227/01.NEU.0000194833.66593.8B
   Prasad D, 2000, J NEUROSURG, V92, P745, DOI 10.3171/jns.2000.92.5.0745
   Regis J, 2004, NEUROCHIRURGIE, V50, P301
   Regis J, 2002, NEUROCHIRURGIE, V48, P471
   Roche Pierre-Hugues, 2008, Prog Neurol Surg, V21, P24, DOI 10.1159/000156556
   THOMSEN J, 1983, AM J OTOL, V5, P20
   TOS M, 1992, ARCH OTOLARYNGOL, V118, P127
   Unger F, 2002, ACT NEUR S, V84, P77
   Vernimmen FJAI, 2010, INT J RADIAT BIOL, V86, P486, DOI 10.3109/09553001003667982
   WEBER B, 1995, ENTREES SYSTEME POST, P139
   WOOLLACOTT MH, 1986, DISORDERS POSTURE GA, P325
NR 29
TC 11
Z9 12
U1 0
U2 2
PU AMER ASSOC NEUROLOGICAL SURGEONS
PI ROLLING MEADOWS
PA 5550 MEADOWBROOK DRIVE, ROLLING MEADOWS, IL 60008 USA
SN 0022-3085
EI 1933-0693
J9 J NEUROSURG
JI J. Neurosurg.
PD DEC
PY 2010
VL 113
SU S
BP 128
EP 135
DI 10.3171/2010.8.GKS101089
PG 8
WC Clinical Neurology; Surgery
WE Science Citation Index Expanded (SCI-EXPANDED); Conference Proceedings Citation Index - Science (CPCI-S)
SC Neurosciences & Neurology; Surgery
GA 684QV
UT WOS:000284565300020
PM 21121794
DA 2023-08-27
ER

PT J
AU Lang, EK
   Hanano, A
   Rudman, E
   Thomas, R
   Myers, L
   Nguyen, Q
   Macchia, RJ
AF Lang, Erich K.
   Hanano, Amer
   Rudman, Ernest
   Thomas, Raju
   Myers, Leann
   Quan Nguyen
   Macchia, Richard J.
TI The fate of small renal masses, less then 1 cm size: outcome study
SO INTERNATIONAL BRAZ J UROL
LA English
DT Article
DE kidney; neoplasms; outcome assessment
ID NATURAL-HISTORY; SURVEILLANCE; MANAGEMENT
AB Purpose: We evaluated the outcome and etiologies of small renal masses (less than 1 cm in size) discovered incidentally on 2 consecutive CTs that investigated non-urologic abdominal complaints.
   Materials and Methods: A retrospective search for incidentally discovered small renal masses, less then 1 cm in size, was carried out in the files of 6 major US medical centers. 4822 such lesions had been reported over a 12 year period. A search of these patients' records revealed 1082 subsequent new CTs for non urologic complaints, allowing the assessment of the fate of the masses. Lesions enlarging, of ambivalent contour or enhancement were examined by a third multiphasic MDCT. The findings were interpreted by 2 blinded radiologists.
   Results: Six hundred and four masses could no longer be identified, 231 were significantly smaller, 113 unchanged in size and 134 larger. Of the disappearing lesions 448 were located in the medulla, 94 both in medulla and cortex and 62 in cortex. Multiphasic MDCTs obtained in 308 masses enlarging, unchanged in size or of ambivalent appearance, revealed 7 neoplasms, 45 inflammatory lesions, 8 abscesses and 62 renal medullary necrosis. Concurrent antibiotic therapy of GI conditions may have caused some of the 496 lesions to disappear.
   Conclusion: It is questionable whether the small number of malignant neoplasms (0.4%), inflammatory lesions (5%) and renal medullary necrosis (6%) justify routine follow-up CTs and exposure to radiation. The delay in intervention in neoplastic lesions probably didn't influence tumor-free survival potential and clinical symptoms would soon have revealed inflammatory conditions. With exception of ambivalent lesions, clinical surveillance appears adequate.
C1 [Lang, Erich K.; Hanano, Amer; Rudman, Ernest; Quan Nguyen; Macchia, Richard J.] SUNY Downstate Med Sch, Dept Radiol, Brooklyn, NY 11203 USA.
   [Lang, Erich K.; Hanano, Amer; Rudman, Ernest; Quan Nguyen; Macchia, Richard J.] SUNY Downstate Med Sch, Dept Urol, Brooklyn, NY 11203 USA.
   [Lang, Erich K.; Thomas, Raju; Myers, Leann] Tulane Hlth Sci Ctr, Dept Urol, New Orleans, LA USA.
   [Lang, Erich K.; Thomas, Raju; Myers, Leann] Tulane Hlth Sci Ctr, Dept Biostat, New Orleans, LA USA.
   [Lang, Erich K.; Thomas, Raju; Myers, Leann] Sch Publ Hlth, New Orleans, LA USA.
   [Lang, Erich K.] Johns Hopkins Med Inst, Dept Radiol, Baltimore, MD 21205 USA.
C3 State University of New York (SUNY) System; State University of New York
   (SUNY) Downstate Medical Center; State University of New York (SUNY)
   System; State University of New York (SUNY) Downstate Medical Center;
   Tulane University; Tulane University; Johns Hopkins University; Johns
   Hopkins Medicine
RP Lang, EK (通讯作者)，SUNY Downstate Med Sch, Dept Radiol, 450 Clarkson Av, Brooklyn, NY 11203 USA.
EM
OI Thomas, Raju/0000-0003-4233-9785
CR [Anonymous], 2009, RENAL MASS GUIDELINE, P15
   [Anonymous], 2007, J UROL
   Cozar JM, 2008, ADV UROL, V2008, DOI 10.1155/2008/916463
   Crispen PL, 2008, CANCER-AM CANCER SOC, V112, P1051, DOI 10.1002/cncr.23268
   DeRoche T, 2008, AM J CLIN PATHOL, V130, P560, DOI 10.1309/YR7P42XUVQHPHDWL
   Hsu RM, 2004, AM J ROENTGENOL, V182, P551, DOI 10.2214/ajr.182.3.1820551
   Jewett MAS, 2008, UROL CLIN N AM, V35, P627, DOI 10.1016/j.ucl.2008.07.004
   Kunkle DA, 2008, CANCER, V113, P2671, DOI 10.1002/cncr.23896
   Kunkle DA, 2008, J UROLOGY, V179, P1227, DOI 10.1016/j.juro.2007.11.047
   Kunkle DA, 2007, J UROLOGY, V177, P849, DOI 10.1016/j.juro.2006.10.073
   Kutikov A, 2009, CURR OPIN UROL, V19, P148, DOI 10.1097/MOU.0b013e328323f7d7
   Marszalek M, 2004, UROLOGY, V64, P38, DOI 10.1016/j.urology.2004.02.007
   Ozsoy M, 2008, ADV UROL, V2008, DOI 10.1155/2008/196701
   Russo P, 2008, NAT CLIN PRACT UROL, V5, P482, DOI 10.1038/ncpuro1177
   Schachter LR, 2007, J ENDOUROL, V21, P819, DOI 10.1089/end.2006.9937
   Srougi V, 2009, INT BRAZ J UROL, V35, P427, DOI 10.1590/S1677-55382009000400005
   Thompson RH, 2009, J UROLOGY, V181, P2033, DOI 10.1016/j.juro.2009.01.027
   Van Poppel H, 2007, EUR UROL, V52, P1323, DOI 10.1016/j.eururo.2007.07.025
   Volpe A, 2004, CANCER-AM CANCER SOC, V100, P738, DOI 10.1002/cncr.20025
   Volpe A, 2007, THESCIENTIFICWORLDJO, V7, P860, DOI 10.1100/tsw.2007.157
   Wang R, 2009, UROLOGY, V73, P586, DOI 10.1016/j.urology.2008.08.519
NR 21
TC 4
Z9 4
U1 0
U2 1
PU BRAZILIAN SOC UROL
PI RIO DE JANEIRO
PA RUA BAMBINA, 153, RIO DE JANEIRO, 2251-050, BRAZIL
SN 1677-5538
EI 1677-6119
J9 INT BRAZ J UROL
JI Int. Braz J Urol
PD JAN-FEB
PY 2012
VL 38
IS 1
BP 40
EP 47
DI 10.1590/S1677-55382012000100006
PG 8
WC Urology & Nephrology
WE Science Citation Index Expanded (SCI-EXPANDED)
SC Urology & Nephrology
GA 908JW
UT WOS:000301487700008
PM 22397785
OA gold, Green Submitted
DA 2023-08-27
ER

PT J
AU Freitas, MCS
   Uchida, Y
   Lassman, C
   Danovitch, GM
   Busuttil, RW
   Kupiec-Weglinski, JW
AF Freitas, Maria Cecilia S.
   Uchida, Yoichiro
   Lassman, Charles
   Danovitch, Gabriel M.
   Busuttil, Ronald W.
   Kupiec-Weglinski, Jerzy W.
TI Type I Interferon Pathway Mediates Renal Ischemia/Reperfusion Injury
SO TRANSPLANTATION
LA English
DT Article
DE Kidney ischemia/reperfusion injury; Innate immunity; Type I interferon;
   Inflammation
ID TOLL-LIKE RECEPTOR-4; NEUTROPHIL INFILTRATION; KIDNEY; ISCHEMIA;
   REPERFUSION; ALPHA; ACTIVATION; INNATE; CELLS; INTERLEUKIN-1
AB Background. Ischemia/reperfusion (I/R) injury is a common cause of acute renal failure after kidney transplantation. This study was designed to analyze the role of type I interferon (IFN) signaling downstream of Toll-like receptor 2/Toll-like receptor 4 activation in the mechanism of I/R-triggered kidney damage.
   Methods. Local warm ischemia was induced in groups wild-type (WT) and type I IFN receptor (IFNAR)-/- mice (C57BL/6) by clamping both kidney pedicles for 45 min. Mice were killed at 5/24/72 hr after reperfusion for serum and kidney sampling.
   Results. At 5 hr, serum creatinine and blood urea nitrogen levels were markedly reduced in IFNAR-/- mice as compared with WT. By 24 hr after reperfusion, both serum creatinine/blood urea nitrogen in WT increased further, whereas those in IFNAR-/- mice remained comparable with sham controls. Histological analyses showed significantly higher percentage of tubules in the outer medulla displaying cell necrosis, loss of the brush border, cast formation and tubular dilatation in WT mice, as compared with IFNAR-/-. Immunohistology revealed increased neutrophil and macrophage infiltration in the outer medulla in WT mice. The expression of proinflammatory tumor necrosis factor-alpha, interleukin-1, interleukin-6, and CXCL-2 was markedly reduced selectively in IFNAR-/- mice. Finally, terminal deoxynucleotide transferase-mediated dUTP nick-end labeling analysis showed significantly decreased frequency of apoptotic tubular epithelial cells in IFNAR-deficient mice, as compared with WT.
   Conclusion. This is the first report, which documents the key role of type I IFN signaling in the mechanism of kidney I/R injury. Type I IFN may thus serve as a novel target for the therapy against renal I/R injury.
C1 [Freitas, Maria Cecilia S.; Uchida, Yoichiro; Busuttil, Ronald W.; Kupiec-Weglinski, Jerzy W.] Dumont UCLA Transplantat Ctr, Dept Surg, Div Liver & Pancreas Transplantat, Los Angeles, CA 90095 USA.
   [Lassman, Charles] Univ Calif Los Angeles, David Geffen Sch Med, Dept Pathol & Lab Med, Los Angeles, CA 90095 USA.
   [Danovitch, Gabriel M.] Univ Calif Los Angeles, David Geffen Sch Med, Dept Nephrol, Los Angeles, CA 90095 USA.
C3 University of California System; University of California Los Angeles;
   University of California Los Angeles Medical Center; David Geffen School
   of Medicine at UCLA; University of California System; University of
   California Los Angeles; University of California Los Angeles Medical
   Center; David Geffen School of Medicine at UCLA
RP Kupiec-Weglinski, JW (通讯作者)，Dumont UCLA Transplantat Ctr, Dept Surg, Div Liver & Pancreas Transplantat, 77-120 CHS,10833 Le Conte Ave, Los Angeles, CA 90095 USA.
EM
RI Freitas, Maria/HZJ-0739-2023
FU NIH [RO1 DK062357]; Dumont Research Foundation; CAPES (Coordenacao de
   Aperfeicoamento de Pessoal de Nivel Superior); ISN (International
   Society of Nephrology)
FX This work was supported by NIH grant RO1 DK062357, the Dumont Research
   Foundation, CAPES (Coordenacao de Aperfeicoamento de Pessoal de Nivel
   Superior) (M.C.S.F.), and a Fellowship Grant from ISN (International
   Society of Nephrology) (M.C.S.F.).
CR Allan SM, 2005, NAT REV IMMUNOL, V5, P629, DOI 10.1038/nri1664
   Anders HJ, 2010, KIDNEY INT, V77, P848, DOI 10.1038/ki.2010.71
   Belardelli F, 2002, TRENDS IMMUNOL, V23, P201, DOI 10.1016/S1471-4906(02)02195-6
   Bonventre JV, 2004, KIDNEY INT, V66, P480, DOI 10.1111/j.1523-1755.2004.761_2.x
   Cao WP, 2008, NAT IMMUNOL, V9, P1157, DOI 10.1038/ni.1645
   Chen JL, 2011, KIDNEY INT, V79, P288, DOI 10.1038/ki.2010.381
   COLAMONICI OR, 1993, J BIOL CHEM, V268, P10895
   Decker T, 2005, NAT REV IMMUNOL, V5, P675, DOI 10.1038/nri1684
   Donnahoo KK, 2000, J UROLOGY, V163, P1328, DOI 10.1016/S0022-5347(05)67772-5
   Donnahoo KK, 1999, AM J PHYSIOL-REG I, V277, pR922, DOI 10.1152/ajpregu.1999.277.3.R922
   GUGENHEIM J, 1992, TRANSPLANT INT, V5, pS460
   Izzedine H, 2005, AM J KIDNEY DIS, V45, P804, DOI 10.1053/j.ajkd.2005.02.010
   Kelly KJ, 1996, J CLIN INVEST, V97, P1056, DOI 10.1172/JCI118498
   Kielar ML, 2005, J AM SOC NEPHROL, V16, P3315, DOI 10.1681/asn.2003090757
   Klebanoff SJ, 1999, P ASSOC AM PHYSICIAN, V111, P383, DOI 10.1111/paa.1999.111.5.383
   Kruger B, 2009, P NATL ACAD SCI USA, V106, P3390, DOI 10.1073/pnas.0810169106
   Kwon O, 2003, AM J KIDNEY DIS, V41, P1074, DOI 10.1016/S0272-6386(03)00206-3
   Lechner J, 2008, AM J PHYSIOL-CELL PH, V294, pC153, DOI 10.1152/ajpcell.00120.2007
   Leemans JC, 2005, J CLIN INVEST, V115, P2894, DOI 10.1172/JCI22832
   MULLANE KM, 1985, J PHARMACOL METHOD, V14, P157, DOI 10.1016/0160-5402(85)90029-4
   Nankivell BJ, 2006, TRANSPLANTATION, V81, P643, DOI 10.1097/01.tp.0000190423.82154.01
   Okusa MD, 2002, NEPHRON, V90, P133, DOI 10.1159/000049032
   Patel NSA, 2005, J PHARMACOL EXP THER, V312, P1170, DOI 10.1124/jpet.104.078659
   Patole PS, 2005, KIDNEY INT, V68, P2582, DOI 10.1111/j.1523-1755.2005.00729.x
   Perico N, 2004, LANCET, V364, P1814, DOI 10.1016/S0140-6736(04)17406-0
   Priceman SJ, 2010, BLOOD, V115, P1461, DOI 10.1182/blood-2009-08-237412
   Pulskens WP, 2008, PLOS ONE, V3, DOI 10.1371/journal.pone.0003596
   Rouschop KMA, 2008, KIDNEY INT, V73, P1333, DOI 10.1038/ki.2008.156
   Rusai K, 2008, TRANSPL INT, V21, P572, DOI 10.1111/j.1432-2277.2008.00651.x
   Saemann MD, 2009, AM J TRANSPLANT, V9, P2655, DOI 10.1111/j.1600-6143.2009.02832.x
   Samuel CE, 2001, CLIN MICROBIOL REV, V14, P778, DOI 10.1128/CMR.14.4.778-809.2001
   Savransky V, 2006, KIDNEY INT, V69, P233, DOI 10.1038/sj.ki.5000038
   Shigeoka AA, 2007, J IMMUNOL, V178, P6252, DOI 10.4049/jimmunol.178.10.6252
   Tovey MG, 1996, J LEUKOCYTE BIOL, V59, P512, DOI 10.1002/jlb.59.4.512
   Uchida Y, 2009, LIVER TRANSPLANT, V15, P939, DOI 10.1002/lt.21770
   WILLINGER CC, 1992, VIRCHOWS ARCH B, V62, P237, DOI 10.1007/BF02899687
   Wu HL, 2007, J CLIN INVEST, V117, P2847, DOI 10.1172/JCI31008
   Zhai Y, 2004, J IMMUNOL, V173, P7115, DOI 10.4049/jimmunol.173.12.7115
   Zhai YA, 2008, HEPATOLOGY, V47, P199, DOI 10.1002/hep.21970
NR 39
TC 40
Z9 42
U1 0
U2 6
PU LIPPINCOTT WILLIAMS & WILKINS
PI PHILADELPHIA
PA TWO COMMERCE SQ, 2001 MARKET ST, PHILADELPHIA, PA 19103 USA
SN 0041-1337
EI 1534-6080
J9 TRANSPLANTATION
JI Transplantation
PD JUL 27
PY 2011
VL 92
IS 2
BP 131
EP 138
DI 10.1097/TP.0b013e318220586e
PG 8
WC Immunology; Surgery; Transplantation
WE Science Citation Index Expanded (SCI-EXPANDED)
SC Immunology; Surgery; Transplantation
GA 791AU
UT WOS:000292633000008
PM 21577179
OA Green Accepted, Bronze
DA 2023-08-27
ER

PT J
AU O'Neill, E
   Harkin, A
AF O'Neill, Eoin
   Harkin, Andrew
TI Targeting the noradrenergic system for anti-inflammatory and
   neuroprotective effects: implications for Parkinson's disease
SO NEURAL REGENERATION RESEARCH
LA English
DT Review
DE noradrenaline; microglia; astrocytes; inflammation; Parkinson's disease;
   neuroprotection; animal model; dopamie
ID RAT-BRAIN; LOCUS-COERULEUS; MOTOR DEFICITS; IN-VIVO; NOREPINEPHRINE;
   EXPRESSION; DOPAMINE; PROTECTS; MODEL; NGF
AB Degeneration of the locus coeruleus noradrenergic system is thought to play a key role in the pathogenesis of Parkinson's disease (PD), whereas pharmacological approaches to increase noradrenaline bioavailability may provide neuroprotection. Noradrenaline inhibits microglial activation and suppresses pro-inflammatory mediator production (e.g., tumor necrosis factor-alpha, interleukin-1 beta & inducible nitric oxide synthase activity), thus limiting the cytotoxicity of midbrain dopaminergic neurons in response to an inflammatory stimulus. Neighbouring astrocyte populations promote a neurotrophic environment in response to beta(2)-adrenoceptor (beta(2)-AR) stimulation via the production of growth factors (e.g., brain derived neurotrophic factor, cerebral dopamine neurotrophic factor & glial cell derived neurotrophic factor which have shown promising neuroprotective and neuro-restorative effects in the nigrostriatal dopaminergic system. More recent findings have demonstrated a role for the beta(2)-AR in down-regulating expression levels of the human alpha-synuclein gene SNCA and relative alpha-synuclein protein abundance. Given that alpha-synuclein is a major protein constituent of Lewy body pathology, a hallmark neuropathological feature in Parkinson's disease, these findings could open up new avenues for pharmacological intervention strategies aimed at alleviating the burden of alpha-synucleinopathies in the Parkinsonian brain. In essence, the literature reviewed herein supports our hypothesis of a tripartite neuroprotective role for noradrenaline in combating PD-related neuropathology and motor dysfunction via (1) inhibiting nigral microglial activation & pro-inflammatory mediator production, (2) promoting the synthesis of neurotrophic factors from midbrain astrocytes and (3) downregulating alpha-synuclein gene expression and protein abundance in a beta(2)-AR-dependent manner. Thus, taken together, either pharmacologically enhancing extra-synaptic noradrenaline bioavailability or targeting glial beta(2)-ARs directly makes itself as a promising treatment option aimed at slowing/halting PD progression.
C1 [Harkin, Andrew] Trinity Coll Dublin, Sch Pharm & Pharmaceut Sci, Neuropsychopharmacol Res Grp, Dublin, Ireland.
   Trinity Coll Dublin, Inst Neurosci, Dublin, Ireland.
C3 Trinity College Dublin; Trinity College Dublin
RP Harkin, A (通讯作者)，Trinity Coll Dublin, Sch Pharm & Pharmaceut Sci, Neuropsychopharmacol Res Grp, Dublin, Ireland.
EM
OI O'Neill, Eoin/0000-0002-2472-7795
FU Trinity College
FX Eoin O'Neill was supported by a Trinity College postgraduate award.
CR Archer T, 2003, J NEURAL TRANSM, V110, P183, DOI 10.1007/s00702-002-0777-5
   Baloyannis SJ, 2006, J NEUROL SCI, V248, P35, DOI 10.1016/j.jns.2006.05.006
   Belujon P, 2007, J NEUROSCI, V27, P9595, DOI 10.1523/JNEUROSCI.2583-07.2007
   Cheng HC, 2010, ANN NEUROL, V67, P715, DOI 10.1002/ana.21995
   Counts SE, 2010, J NEUROCHEM, V113, P649, DOI 10.1111/j.1471-4159.2010.06622.x
   Culmsee C, 1999, EUR J PHARMACOL, V379, P33, DOI 10.1016/S0014-2999(99)00452-5
   Culmsee C, 1999, NEUROCHEM INT, V35, P47, DOI 10.1016/S0197-0186(99)00032-7
   Debeir T, 2004, EXP NEUROL, V190, P384, DOI 10.1016/j.expneurol.2004.08.023
   Dello Russo C, 2004, J NEUROINFLAMM, V1, DOI 10.1186/1742-2094-1-9
   Gayle DA, 2002, DEV BRAIN RES, V133, P27, DOI 10.1016/S0165-3806(01)00315-7
   Gesi M, 2000, NEUROSCI BIOBEHAV R, V24, P655, DOI 10.1016/S0149-7634(00)00028-2
   Gill SS, 2003, NAT MED, V9, P589, DOI 10.1038/nm850
   Grondin R, 2000, N-S ARCH PHARMACOL, V361, P181, DOI 10.1007/s002109900167
   Hertz L, 2004, CNS NEUROL DISORD-DR, V3, P239, DOI 10.2174/1568007043337535
   Jenner P, 2003, ANN NEUROL, V53, pS26, DOI 10.1002/ana.10483
   Jiang LL, 2015, GLIA, V63, P1057, DOI 10.1002/glia.22801
   JONSSON G, 1981, EUR J PHARMACOL, V72, P173, DOI 10.1016/0014-2999(81)90272-7
   Kim WG, 2000, J NEUROSCI, V20, P6309, DOI 10.1523/JNEUROSCI.20-16-06309.2000
   Liu M, 2011, PARKINSONS DIS-US, V2011, DOI 10.4061/2011/327089
   LU B, 1991, J NEUROSCI, V11, P318
   Marien MR, 2004, BRAIN RES REV, V45, P38, DOI 10.1016/j.brainresrev.2004.02.002
   McNamee EN, 2010, NEUROPHARMACOLOGY, V59, P37, DOI 10.1016/j.neuropharm.2010.03.014
   McNamee EN, 2010, BRAIN BEHAV IMMUN, V24, P660, DOI 10.1016/j.bbi.2010.02.005
   McNamee EN, 2010, EUR J PHARMACOL, V626, P219, DOI 10.1016/j.ejphar.2009.09.054
   Mittal S, 2017, SCIENCE, V357, P891, DOI 10.1126/science.aaf3934
   Mori K, 2002, NEUROPHARMACOLOGY, V43, P1026, DOI 10.1016/S0028-3908(02)00211-3
   O'Sullivan JB, 2010, J NEUROIMMUNOL, V220, P34, DOI 10.1016/j.jneuroim.2009.12.007
   O'Sullivan JB, 2009, INT J NEUROPSYCHOPH, V12, P687, DOI 10.1017/S146114570800967X
   Panaro MA, 2008, IMMUNOPHARM IMMUNOT, V30, P729, DOI [10.1080/08923970802278557, 10.1080/08923970802278557 ]
   Remy P, 2005, BRAIN, V128, P1314, DOI 10.1093/brain/awh445
   Rommelfanger KS, 2007, P NATL ACAD SCI USA, V104, P13804, DOI 10.1073/pnas.0702753104
   Rommelfanger KS, 2004, J NEUROCHEM, V91, P1116, DOI 10.1111/j.1471-4159.2004.02785.x
   Ryan KJ, 2013, NEUROCHEM INT, V63, P368, DOI 10.1016/j.neuint.2013.07.006
   Semkova I, 1996, BRAIN RES, V717, P44, DOI 10.1016/0006-8993(95)01567-1
   Simonini MV, 2010, J NEUROIMMUNE PHARM, V5, P252, DOI 10.1007/s11481-009-9182-2
   Swanson CJ, 2006, NEUROPHARMACOLOGY, V50, P755, DOI 10.1016/j.neuropharm.2005.11.022
   Vazey EM, 2012, FRONT BEHAV NEUROSCI, V6, DOI 10.3389/fnbeh.2012.00048
   Vollmar P, 2009, INT J NEUROPSYCHOPH, V12, P525, DOI 10.1017/S1461145708009425
   Wang L, 2002, GENE THER, V9, P381, DOI 10.1038/sj.gt.3301682
   Yssel JD, 2018, BRAIN BEHAV IMMUN, V69, P456, DOI 10.1016/j.bbi.2018.01.004
   Zarow C, 2003, ARCH NEUROL-CHICAGO, V60, P337, DOI 10.1001/archneur.60.3.337
NR 41
TC 29
Z9 29
U1 2
U2 6
PU WOLTERS KLUWER MEDKNOW PUBLICATIONS
PI MUMBAI
PA WOLTERS KLUWER INDIA PVT LTD , A-202, 2ND FLR, QUBE, C T S  NO 1498A-2
   VILLAGE MAROL, ANDHERI EAST, MUMBAI, 400059, INDIA
SN 1673-5374
EI 1876-7958
J9 NEURAL REGEN RES
JI Neural Regen. Res.
PD AUG
PY 2018
VL 13
IS 8
BP 1332
EP 1337
DI 10.4103/1673-5374.235219
PG 6
WC Cell Biology; Neurosciences
WE Science Citation Index Expanded (SCI-EXPANDED)
SC Cell Biology; Neurosciences & Neurology
GA GQ8SX
UT WOS:000442030300005
PM 30106035
OA Green Published, Green Submitted, gold
DA 2023-08-27
ER

PT J
AU Bryant, J
   Farmer, J
   Kessler, LJ
   Townsend, RR
   Nathanson, KL
AF Bryant, J
   Farmer, J
   Kessler, LJ
   Townsend, RR
   Nathanson, KL
TI Pheochromocytoma: The expanding genetic differential diagnosis
SO JOURNAL OF THE NATIONAL CANCER INSTITUTE
LA English
DT Review
ID VON-HIPPEL-LINDAU; ENDOCRINE NEOPLASIA TYPE-2; TUMOR-SUPPRESSOR GENE;
   GERM-LINE MUTATIONS; MEDULLARY-THYROID CARCINOMA; COMPLEX-II GENE; RET
   PROTOONCOGENE; SPORADIC PHEOCHROMOCYTOMAS; FAMILIAL PHEOCHROMOCYTOMA;
   NEUROFIBROMATOSIS TYPE-1
AB Pheochromocytomas and paragangliomas are tumors of the autonomic nervous system; pheochromocytomas are tumors of the adrenal medulla, and paragangliomas are extra-adrenal tumors arising from either the sympathetic nervous system or parasympathetic ganglia. It has previously been estimated that approximately 10%-15% of pheochromocytomas are due to hereditary causes. However, our increased understanding of the three hereditary syndromes (neurofibromatosis 1, multiple endocrine neoplasia type 2, and von Hippel-Lindau syndrome) in which pheochromocytoma is found and the recent discovery that mutations in genes in the succinate dehydrogenase family (SDHB and SDHD) predispose to pheochromocytoma have necessitated a re-evaluation of the genetic basis of pheochromocytoma. These studies indicate that the frequency of germline mutations associated with isolated pheochromocytoma is higher than previously estimated, with both hospital-based series and a large population-based series indicating that the frequency of germline mutations in RET, VHL, SDHB, and SDHD taken together approximates 20%. In all patients with pheochromocytoma, including those with known hereditary syndrome or a positive family history, the frequency of germline mutations in these four genes together approaches 30%. Given the frequency of germline mutations, consideration should be given to genetic counseling for all patients with pheochromocytoma and is particularly important for individuals with a positive family history, multifocal disease, or a diagnosis before age 50. Identification of patients with hereditary pheochromocytoma is important because it can guide medical management in mutation-positive patients and their families. This review provides an overview of the known genetic syndromes that are commonly associated with pheochromocytoma, examines recent data on the association of germline mutations in the succinate dehydrogenase gene family with pheochromocytoma, and suggests guidelines for the genetic evaluation of pheochromocytoma patients.
C1 Univ Penn, Sch Med, Div Med Genet, Philadelphia, PA 19104 USA.
   Univ Penn, Sch Med, Dept Med, Div Nephrol, Philadelphia, PA 19104 USA.
C3 University of Pennsylvania; University of Pennsylvania
RP Nathanson, KL (通讯作者)，Univ Penn, Sch Med, Div Med Genet, 513 BRB 2-3,421 Curie Blvd, Philadelphia, PA 19104 USA.
OI Nathanson, Katherine/0000-0002-6740-0901
FU NCI NIH HHS [K08 CA84030] Funding Source: Medline
CR Aguiar RCT, 2001, J CLIN ENDOCR METAB, V86, P2890, DOI 10.1210/jc.86.6.2890
   Astuti D, 2001, LANCET, V357, P1181, DOI 10.1016/S0140-6736(00)04378-6
   Astuti D, 2001, AM J HUM GENET, V69, P49, DOI 10.1086/321282
   Badenhop RF, 2001, GENE CHROMOSOME CANC, V31, P255, DOI 10.1002/gcc.1142
   Bar M, 1997, CLIN ENDOCRINOL, V47, P707, DOI 10.1046/j.1365-2265.1997.3251150.x
   Baysal BE, 2000, SCIENCE, V287, P848, DOI 10.1126/science.287.5454.848
   Baysal BE, 2002, J MED GENET, V39, P178, DOI 10.1136/jmg.39.3.178
   Baysal BE, 2002, J MED GENET, V39, P617, DOI 10.1136/jmg.39.9.617
   Baysal BE, 1997, AM J HUM GENET, V60, P121
   BELDJORD C, 1995, J CLIN ENDOCR METAB, V80, P2063, DOI 10.1210/jc.80.7.2063
   BOURGERON T, 1995, NAT GENET, V11, P144, DOI 10.1038/ng1095-144
   Brandi ML, 2001, J CLIN ENDOCR METAB, V86, P5658, DOI 10.1210/jc.86.12.5658
   Brauch H, 1997, J CLIN ENDOCR METAB, V82, P4101, DOI 10.1210/jc.82.12.4101
   BRAVO EL, 1991, KIDNEY INT, V40, P544, DOI 10.1038/ki.1991.244
   CARNEY JA, 1980, AM J MED, V68, P515, DOI 10.1016/0002-9343(80)90295-8
   CASANOVA S, 1993, CLIN ENDOCRINOL, V38, P531, DOI 10.1111/j.1365-2265.1993.tb00350.x
   Cascon A, 2002, EUR J HUM GENET, V10, P457, DOI 10.1038/sj.ejhg.5200829
   Clifford SC, 2001, HUM MOL GENET, V10, P1029, DOI 10.1093/hmg/10.10.1029
   CROSSEY PA, 1994, HUM GENET, V93, P53
   Dannenberg H, 2002, CLIN CANCER RES, V8, P2061
   DECKER RA, 1995, SURGERY, V118, P257, DOI 10.1016/S0039-6060(05)80332-0
   DEVILEE P, 1994, GENE CHROMOSOME CANC, V11, P71, DOI 10.1002/gcc.2870110202
   Dluhy RG, 2002, NEW ENGL J MED, V346, P1486, DOI 10.1056/NEJM200205093461911
   DUAN DR, 1995, SCIENCE, V269, P1402, DOI 10.1126/science.7660122
   Eisenhofer G, 2001, J CLIN ENDOCR METAB, V86, P1999, DOI 10.1210/jc.86.5.1999
   ENG C, 1995, CLIN ENDOCRINOL, V43, P123, DOI 10.1111/j.1365-2265.1995.tb01903.x
   ENG C, 1995, J MED GENET, V32, P934, DOI 10.1136/jmg.32.12.934
   ENG C, 1994, HUM MOL GENET, V3, P237, DOI 10.1093/hmg/3.2.237
   Eng C, 2001, J MED GENET, V38, P824, DOI 10.1136/jmg.38.12.824
   Eng C, 1996, JAMA-J AM MED ASSOC, V276, P1575, DOI 10.1001/jama.276.19.1575
   FrankRaue K, 1996, EUR J ENDOCRINOL, V135, P222, DOI 10.1530/eje.0.1350222
   Friedman JM, 1999, AM J MED GENET, V89, P1
   Gimm O, 2000, CANCER RES, V60, P6822
   Goldstein RE, 1999, ANN SURG, V229, P755, DOI 10.1097/00000658-199906000-00001
   GRUFFERMAN S, 1980, CANCER, V46, P2116, DOI 10.1002/1097-0142(19801101)46:9<2116::AID-CNCR2820460934>3.0.CO;2-S
   Gutmann DH, 2001, HUM MOL GENET, V10, P747, DOI 10.1093/hmg/10.7.747
   Gutmann DH, 1997, JAMA-J AM MED ASSOC, V278, P51, DOI 10.1001/jama.278.1.51
   Hansford JR, 2000, J MED GENET, V37, P817, DOI 10.1136/jmg.37.11.817
   HEIM RA, 1995, HUM MOL GENET, V4, P975, DOI 10.1093/hmg/4.6.975
   Hoffman MA, 2001, HUM MOL GENET, V10, P1019, DOI 10.1093/hmg/10.10.1019
   HUSON SM, 1989, J MED GENET, V26, P704, DOI 10.1136/jmg.26.11.704
   Iliopoulos O, 1996, P NATL ACAD SCI USA, V93, P10595, DOI 10.1073/pnas.93.20.10595
   Inabnet WB, 2000, SURGERY, V128, P1007, DOI 10.1067/msy.2000.110846
   Januszewicz A, 2000, J HYPERTENS, V18, P1019, DOI 10.1097/00004872-200018080-00005
   Jenne DE, 2001, AM J HUM GENET, V69, P516, DOI 10.1086/323043
   Kamura T, 1999, SCIENCE, V284, P657, DOI 10.1126/science.284.5414.657
   KAUFMAN JJ, 1979, J UROLOGY, V121, P801, DOI 10.1016/S0022-5347(17)56999-2
   LAIRMORE TC, 1993, ANN SURG, V217, P595, DOI 10.1097/00000658-199306000-00001
   LATIF F, 1993, SCIENCE, V260, P1317, DOI 10.1126/science.8493574
   Lenders JWM, 2002, JAMA-J AM MED ASSOC, V287, P1427, DOI 10.1001/jama.287.11.1427
   LINDOR NM, 1995, J CLIN ENDOCR METAB, V80, P627, DOI 10.1210/jc.80.2.627
   Lips CJM, 2001, ANN CLIN BIOCHEM, V38, P168, DOI 10.1258/0004563011900614
   Lubensky IA, 1996, AM J PATHOL, V149, P2089
   Maddock IR, 1996, J MED GENET, V33, P120, DOI 10.1136/jmg.33.2.120
   MAHER ER, 1991, J MED GENET, V28, P443, DOI 10.1136/jmg.28.7.443
   Milunsky JM, 2001, AM J MED GENET, V100, P311, DOI 10.1002/ajmg.1270
   MODIGLIANI E, 1995, J INTERN MED, V238, P363, DOI 10.1111/j.1365-2796.1995.tb01211.x
   Morrison PJ, 1996, J MED GENET, V33, P779, DOI 10.1136/jmg.33.9.779
   MULLIGAN LM, 1993, NATURE, V363, P458, DOI 10.1038/363458a0
   MULLIGAN LM, 1995, J INTERN MED, V238, P343, DOI 10.1111/j.1365-2796.1995.tb01208.x
   NEUMANN HPH, 1993, NEW ENGL J MED, V329, P1531, DOI 10.1056/NEJM199311183292103
   Neumann HPH, 2002, NEW ENGL J MED, V346, P1459, DOI 10.1056/NEJMoa020152
   Neumann HPH, 2001, NEW ENGL J MED, V345, P547
   Nguyen L, 2001, EUR J ENDOCRINOL, V144, P37, DOI 10.1530/eje.0.1440037
   Niemann S, 2000, NAT GENET, V26, P268, DOI 10.1038/81551
   Offit K, 1996, J CLIN ONCOL, V14, P1730, DOI 10.1200/JCO.1996.14.5.1730
   Ohh M, 1998, MOL CELL, V1, P959, DOI 10.1016/S1097-2765(00)80096-9
   Pacak K, 2001, ANN INTERN MED, V134, P315, DOI 10.7326/0003-4819-134-4-200102200-00016
   PARKIN JL, 1981, ANN OTO RHINOL LARYN, V90, P60, DOI 10.1177/000348948109000115
   Pomares FJ, 1998, CLIN ENDOCRINOL, V48, P195, DOI 10.1046/j.1365-2265.1998.3751208.x
   Ponder BAJ, 1999, CANCER RES, V59, p1736S
   Prowse AH, 1997, AM J HUM GENET, V60, P765
   Reed N, 2001, TRENDS MOL MED, V7, P157, DOI 10.1016/S1471-4914(01)01955-4
   Riccardi VM, 1992, NEUROFIBROMATOSIS PH
   RICHARDS FM, 1994, HUM MOL GENET, V3, P595, DOI 10.1093/hmg/3.4.595
   RICHARDS FM, 1993, HUM MOL GENET, V2, P879, DOI 10.1093/hmg/2.7.879
   RICHARDS FM, 2001, EXP REV MOL MED 0319
   Ruttledge MH, 1996, AM J HUM GENET, V59, P331
   SANTORO M, 1995, SCIENCE, V267, P381, DOI 10.1126/science.7824936
   Scheffler IE, 1998, PROG NUCLEIC ACID RE, V60, P267, DOI 10.1016/S0079-6603(08)60895-8
   SCHIMKE RN, 2002, GENE REV
   Shen MH, 1996, J MED GENET, V33, P2, DOI 10.1136/jmg.33.1.2
   Stebbins CE, 1999, SCIENCE, V284, P455, DOI 10.1126/science.284.5413.455
   Stolle C, 1998, HUM MUTAT, V12, P417, DOI 10.1002/(SICI)1098-1004(1998)12:6<417::AID-HUMU8>3.0.CO;2-K
   Szudek J, 2003, HUM GENET, V112, P289, DOI 10.1007/s00439-002-0871-7
   Taschner PEM, 2001, GENE CHROMOSOME CANC, V31, P274, DOI 10.1002/gcc.1144
   van der Harst E, 1998, INT J CANCER, V77, P337, DOI 10.1002/(SICI)1097-0215(19980729)77:3<337::AID-IJC5>3.0.CO;2-P
   VANDERMEY AGL, 1989, LANCET, V2, P1291
   WALLACE MR, 1990, SCIENCE, V249, P181, DOI 10.1126/science.2134734
   Walther MM, 1999, J UROLOGY, V162, P1582, DOI 10.1016/S0022-5347(05)68171-2
   Walther MM, 1999, J UROLOGY, V162, P659, DOI 10.1097/00005392-199909010-00004
   WIESNER GL, 2003, GEN REV GENE TESTS
   Woodward ER, 1997, HUM MOL GENET, V6, P1051, DOI 10.1093/hmg/6.7.1051
   Zoller MET, 1997, CANCER-AM CANCER SOC, V79, P2125
NR 94
TC 172
Z9 187
U1 0
U2 5
PU OXFORD UNIV PRESS INC
PI CARY
PA JOURNALS DEPT, 2001 EVANS RD, CARY, NC 27513 USA
SN 0027-8874
J9 J NATL CANCER I
JI J. Natl. Cancer Inst.
PD AUG 20
PY 2003
VL 95
IS 16
BP 1196
EP 1204
DI 10.1093/jnci/djg024
PG 9
WC Oncology
WE Science Citation Index Expanded (SCI-EXPANDED)
SC Oncology
GA 713FD
UT WOS:000184845700008
PM 12928344
OA Bronze
DA 2023-08-27
ER

PT J
AU Inoue, T
   Nishimura, S
   Hayashi, N
   Numagami, Y
   Kaimori, M
   Nishijima, M
AF Inoue, Tomoo
   Nishimura, Shinjitsu
   Hayashi, Nakamasa
   Numagami, Yoshihiro
   Kaimori, Mitsuomi
   Nishijima, Michiharu
TI Ectopic recurrence of dysplastic gangliocytoma of the cerebellum
   (Lhermitte-Duclos disease): a case report
SO BRAIN TUMOR PATHOLOGY
LA English
DT Article
DE Lhermitte-Duclos disease; recurrence; magnetic resonance imaging
ID HYPERTROPHY; ASSOCIATION; DIAGNOSIS; MRI
AB Lhermitte-Duclos disease is a rare clinical entity characterized by slow deformation of the cerebellar lesion. A 53-year-old woman presented with Lherimitte-Duclos disease manifesting as ataxic gait, occipital headache, and loss of consciousness. Magnetic resonance imaging demonstrated striated and laminar pattern lesions in the right cerebellar hemisphere and vermis. She underwent subtotal removal of the vermis compressing the brainstem, but the cerebellar hemisphere appeared normal and was preserved. Histological findings were consistent with Lherimitte-Duclos disease. Two years later, magnetic resonance imaging revealed enlargement of the right hemispheric lesion. Intraoperative appearance of the hemisphere demonstrated marked enlargement of the folia. Subtotal removal was performed. Histological examination showed no malignant findings. Lhermitte-Duclos disease may recur, and should be carefully followed up as a low-grade neoplasm, not as a hamartomatous disorder.
C1 Aomori Prefect Cent Hosp, Dept Neurosurg, Aomori, Aomori 0308553, Japan.
   Aomori Prefect Cent Hosp, Dept Pathol, Aomori, Aomori 0308553, Japan.
RP Inoue, T (通讯作者)，Aomori Prefect Cent Hosp, Dept Neurosurg, 2-1-1 Higashitsukurimichi, Aomori, Aomori 0308553, Japan.
EM
CR BANERJEE AK, 1979, IRISH J MED SCI, V148, P97, DOI 10.1007/BF02938059
   Buhl R, 2003, ACTA NEUROCHIR, V145, P509, DOI 10.1007/s00701-003-0040-3
   deLeon GA, 1996, J NEUROSURG, V85, P157, DOI 10.3171/jns.1996.85.1.0157
   Derrey S, 2004, SURG NEUROL, V61, P447, DOI 10.1016/S0090-3019(03)00576-7
   HAIR LS, 1992, ACTA NEUROPATHOL, V84, P570, DOI 10.1007/BF00304477
   Hashimoto Hiroyuki, 1997, Neurologia Medico-Chirurgica, V37, P692
   Kleihues P, 2002, J NEUROPATH EXP NEUR, V61, P215, DOI 10.1093/jnen/61.3.215
   Kulkantrakorn K, 1997, NEUROLOGY, V48, P725, DOI 10.1212/WNL.48.3.725
   Lhermitte J., 1920, B ASSOC FR ETUD CANC, V9, P99
   MARANO SR, 1988, J NEUROSURG, V69, P599, DOI 10.3171/jns.1988.69.4.0599
   MELTZER CC, 1995, RADIOLOGY, V194, P699, DOI 10.1148/radiology.194.3.7862965
   MILBOUW G, 1988, NEUROSURGERY, V22, P124, DOI 10.1227/00006123-198801000-00020
   Moonis G, 2004, NEURORADIOLOGY, V46, P351, DOI 10.1007/s00234-004-1190-6
   Nowak DA, 2002, ACTA NEUROL SCAND, V105, P137, DOI 10.1034/j.1600-0404.2002.1r127.x
   Perez-Nunez A, 2004, ACTA NEUROCHIR, V146, P679, DOI 10.1007/s00701-004-0264-x
   REEDER RF, 1988, NEUROSURGERY, V23, P240, DOI 10.1227/00006123-198808000-00022
   Robinson Shenandoah, 2006, Neurosurg Focus, V20, pE6
   ROSKI RA, 1981, J NEUROSURG, V55, P318, DOI 10.3171/jns.1981.55.2.0318
   STAPLETON SR, 1992, BRIT J NEUROSURG, V6, P153, DOI 10.3109/02688699209002919
   WELLS GB, 1994, J NEUROSURG, V81, P133, DOI 10.3171/jns.1994.81.1.0133
   WILLIAMS DW, 1992, AM J NEURORADIOL, V13, P287
NR 21
TC 8
Z9 10
U1 0
U2 1
PU SPRINGER JAPAN KK
PI TOKYO
PA CHIYODA FIRST BLDG EAST, 3-8-1 NISHI-KANDA, CHIYODA-KU, TOKYO, 101-0065,
   JAPAN
SN 1433-7398
EI 1861-387X
J9 BRAIN TUMOR PATHOL
JI Brain Tumor Pathol.
PY 2007
VL 24
IS 1
BP 25
EP 29
DI 10.1007/s10014-006-0211-z
PG 5
WC Oncology; Clinical Neurology; Pathology
WE Science Citation Index Expanded (SCI-EXPANDED)
SC Oncology; Neurosciences & Neurology; Pathology
GA 176FM
UT WOS:000247070000005
PM 18095141
DA 2023-08-27
ER

PT J
AU Speer, S
   Klein, A
   Kober, L
   Weiss, A
   Yohannes, I
   Bert, C
AF Speer, Stefan
   Klein, Andreas
   Kober, Lukas
   Weiss, Alexander
   Yohannes, Indra
   Bert, Christoph
TI Automation of radiation treatment planning Evaluation of head and neck
   cancer patient plans created by the Pinnacle(3) scripting and
   Auto-Planning functions
SO STRAHLENTHERAPIE UND ONKOLOGIE
LA English
DT Article
DE Radiotherapy, intensity-modulated; Organs at risk; Spinal cord;
   Brainstem; Parotid gland
ID OPTIMIZATION; THERAPY
AB Background Intensity-modulated radiotherapy (IMRT) techniques are now standard practice. IMRT or volumetric- modulated arc therapy (VMAT) allow treatment of the tumor while simultaneously sparing organs at risk. Nevertheless, treatment plan quality still depends on the physicist's individual skills, experiences, and personal preferences. It would therefore be advantageous to automate the planning process. This possibility is offered by the Pinnacle(3) treatment planning system (Philips Healthcare, Hamburg, Germany) via its scripting language or AutoPlanning (AP) module.
   Materials and methods AP module results were compared to in-house scripts and manually optimized treatment plans for standard head and neck cancer plans. Multiple treatment parameters were scored to judge plan quality (100 points = optimum plan). Patients were initially planned manually by different physicists and re-planned using scripts or AP.
   Results and discussion Script-based head and neck plans achieved a mean of 67.0 points and were, on average, superior to manually created (59.1 points) and AP plans (62.3 points). Moreover, they are characterized by reproducibility and lower standard deviation of treatment parameters. Even less experienced staff are able to create at least a good starting point for further optimization in a short time. However, for particular plans, experienced planners perform even better than scripts or AP. Experienced-user input is needed when setting up scripts or AP templates for the first time. Moreover, some minor drawbacks exist, such as the increase of monitor units (+ 35.5% for scripted plans).
   Conclusion On average, automatically created plans are superior to manually created treatment plans. For particular plans, experienced physicists were able to perform better than scripts or AP; thus, the benefit is greatest when time is short or staff inexperienced.
C1 [Speer, Stefan; Weiss, Alexander; Bert, Christoph] Friedrich Alexander Univ Erlangen Nurnberg, Univ Klinikum Erlangen, Dept Radiat Oncol, Univ Str 27, D-91054 Erlangen, Germany.
   [Klein, Andreas] EKS Engn GmbH, Dr Mack Str 88, D-90762 Furth, Germany.
   [Kober, Lukas] Strahlentherapie Tauber Franken, Uhlandstr 7, D-97980 Bad Mergentheim, Germany.
   [Yohannes, Indra] Rinecker Proton Therapy Ctr, Schaftlarnstr 133, D-81371 Munich, Germany.
C3 University of Erlangen Nuremberg
RP Speer, S (通讯作者)，Friedrich Alexander Univ Erlangen Nurnberg, Univ Klinikum Erlangen, Dept Radiat Oncol, Univ Str 27, D-91054 Erlangen, Germany.
EM
RI Bert, Christoph/C-2585-2013
OI Bert, Christoph/0000-0002-8539-6600
CR Breedveld S, 2012, MED PHYS, V39, P951, DOI 10.1118/1.3676689
   Buschmann M, 2016, PHYS MEDICA, V32, P465, DOI 10.1016/j.ejmp.2016.03.002
   De Luca P., 2010, J ICRU, V10, P1, DOI 10.1093/jicru_ndq002
   Geoghegan S, 2007, SCRIPTING PINNACLE3
   Haderlein M, 2014, STRAHLENTHER ONKOL, V190, P192, DOI 10.1007/s00066-013-0493-0
   Marks LB, 2010, INT J RADIAT ONCOL, V76, pS10, DOI 10.1016/j.ijrobp.2009.07.1754
   Quan EM, 2012, INT J RADIAT ONCOL, V84, pE69, DOI 10.1016/j.ijrobp.2012.02.017
   Riis B, 2013, PINN US M
   Theunissen EAR, 2015, HEAD NECK-J SCI SPEC, V37, P281, DOI 10.1002/hed.23551
   Thieke C, 2007, RADIOTHER ONCOL, V85, P292, DOI 10.1016/j.radonc.2007.06.020
   Wu BB, 2013, MED PHYS, V40, DOI 10.1118/1.4788671
   Wu BB, 2009, MED PHYS, V36, P5497, DOI 10.1118/1.3253464
   Xhaferllari I, 2013, J APPL CLIN MED PHYS, V14, P176, DOI 10.1120/jacmp.v14i1.4052
   Yang Y, 2004, PHYS MED BIOL, V49, P5101, DOI 10.1088/0031-9155/49/22/006
NR 14
TC 21
Z9 23
U1 0
U2 6
PU SPRINGER HEIDELBERG
PI HEIDELBERG
PA TIERGARTENSTRASSE 17, D-69121 HEIDELBERG, GERMANY
SN 0179-7158
EI 1439-099X
J9 STRAHLENTHER ONKOL
JI Strahlenther. Onkol.
PD AUG
PY 2017
VL 193
IS 8
BP 656
EP 665
DI 10.1007/s00066-017-1150-9
PG 10
WC Oncology; Radiology, Nuclear Medicine & Medical Imaging
WE Science Citation Index Expanded (SCI-EXPANDED)
SC Oncology; Radiology, Nuclear Medicine & Medical Imaging
GA FB5KE
UT WOS:000406179300008
PM 28653120
DA 2023-08-27
ER

PT J
AU Manotas, MC
   Rivera, AL
   Gomez, AM
   Abisambra, P
   Guevara, G
   Medina, V
   Tapiero, S
   Huertas, A
   Riano-Moreno, J
   Mejia, JC
   Gonzalez-Clavijo, AM
   Tapiero-Garcia, M
   Cuellar-Cuellar, AA
   Fierro-Maya, LF
   Sanabria-Salas, MC
AF Manotas, Maria Carolina
   Rivera, Ana Lucia
   Gomez, Ana Milena
   Abisambra, Patricia
   Guevara, Gonzalo
   Medina, Vilma
   Tapiero, Sandra
   Huertas, Antonio
   Riano-Moreno, Julian
   Mejia, Juan Carlos
   Gonzalez-Clavijo, Angelica Maria
   Tapiero-Garcia, Mireya
   Cuellar-Cuellar, Andres Arturo
   Fierro-Maya, Luis Felipe
   Sanabria-Salas, Maria Carolina
TI SDHB exon 1 deletion: A recurrent germline mutation in Colombian
   patients with pheochromocytomas and paragangliomas
SO FRONTIERS IN GENETICS
LA English
DT Article
DE pheochromocytomas; paragangliomas; germline mutation; succinate
   dehydrogenase subunit B; human; neoplastic syndromes; hereditary
ID GENETICS; GUIDELINES; DIAGNOSIS
AB Pheochromocytomas (PCCs) and paragangliomas (PGLs) (known as PPGL in combination) are rare neuroendocrine tumors of the adrenal medulla and extra-adrenal ganglia. About 40% of the patients with PPGL have a hereditary predisposition. Here we present a case-series of 19 unrelated Colombian patients with a clinical diagnosis of PPGL tumors that underwent germline genetic testing as part of the Hereditary Cancer Program developed at the Instituto Nacional de Cancerologia, Colombia (INC-C), the largest reference cancer center in the country. Ten of 19 patients (52.63%) were identified as carriers of a pathogenic/likely pathogenic (P/LP) germline variant in a known susceptibility gene. The majority of the P/LP variants were in the SDHB gene (9/10): one corresponded to a nonsense variant c.268C > T (p.Arg90*) and eight cases were found to be carriers of a recurrent CNV consisting of a large deletion of one copy of exon 1, explaining 42% (8/19) of all the affected cases. Only one additional case was found to be a carrier of a missense mutation in the VHL gene: c.355T > C (p.Phe119Leu). Our study highlights the major role of SDHB in Colombian patients with a clinical diagnosis of PGL/PCC tumors and supports the recommendation of including the analysis of large deletions/duplications of the SDHB gene as part of the genetic counselling to improve the detection rate of hereditary cases and their clinical care.
C1 [Manotas, Maria Carolina; Rivera, Ana Lucia; Gomez, Ana Milena; Guevara, Gonzalo; Medina, Vilma; Tapiero, Sandra; Huertas, Antonio; Riano-Moreno, Julian; Mejia, Juan Carlos; Gonzalez-Clavijo, Angelica Maria; Tapiero-Garcia, Mireya; Cuellar-Cuellar, Andres Arturo; Fierro-Maya, Luis Felipe; Sanabria-Salas, Maria Carolina] Inst Nacl Cancerol, Med Subdirect, Bogota, Colombia.
   [Abisambra, Patricia] Univ Norte, Dept Med, Barranquilla, Colombia.
   [Sanabria-Salas, Maria Carolina] Inst Nacl Cancerol, Subdirect Res, Bogota, Colombia.
C3 Universidad del Norte Colombia
RP Sanabria-Salas, MC (通讯作者)，Inst Nacl Cancerol, Med Subdirect, Bogota, Colombia.; Sanabria-Salas, MC (通讯作者)，Inst Nacl Cancerol, Subdirect Res, Bogota, Colombia.
EM
RI Fierro Maya, Luis Felipe/C-6776-2014
OI Fierro Maya, Luis Felipe/0000-0003-1661-6574; SANABRIA SALAS, MARIA
   CAROLINA/0000-0002-7946-2026; Cuellar Cuellar, Andres
   Arturo/0000-0002-1757-7952
FU Program for the Creation of a National Network of Hereditary Cancer in
   Colombia, INC-C [C19990300218]; Institutional Program for the
   Identification and Management of Families with Suspected Hereditary
   Cancer, INC-C [C19990300210]
FX MS (C19990300218-Program for the Creation of a National Network of
   Hereditary Cancer in Colombia, INC-C); MS (C19990300210-Institutional
   Program for the Identification and Management of Families with Suspected
   Hereditary Cancer, INC-C).
CR Albattal Shatha, 2019, Oncotarget, V10, P5919, DOI 10.18632/oncotarget.27194
   Andrews KA, 2018, J MED GENET, V55, P384, DOI 10.1136/jmedgenet-2017-105127
   Anuario estadistico 2019, 2021, I NAC CANC
   Astuti D, 2001, AM J HUM GENET, V69, P49, DOI 10.1086/321282
   Aygun N, 2020, MED B SISLI ETFAL HO, V54, P271, DOI 10.14744/SEMB.2020.14826
   Aygun N, 2020, MED B SISLI ETFAL HO, V54, P159, DOI 10.14744/SEMB.2020.18794
   Bausch B, 2014, ENDOCR-RELAT CANCER, V21, P17, DOI 10.1530/ERC-13-0415
   Bayley JP, 2005, BMC MED GENET, V6, DOI 10.1186/1471-2350-6-39
   Ben Aim L, 2022, J MED GENET, V59, P785, DOI 10.1136/jmedgenet-2020-107652
   Benn DE, 2015, ENDOCR-RELAT CANCER, V22, pT91, DOI 10.1530/ERC-15-0268
   Berends AMA, 2018, EUR J INTERN MED, V51, P68, DOI 10.1016/j.ejim.2018.01.015
   Boyd J, 2019, EUR J ENDOCRINOL, V181, P301, DOI 10.1530/EJE-19-0176
   Cascon A, 2008, J MED GENET, V45, P233, DOI 10.1136/jmg.2007.054965
   Cascon A, 2006, GENE CHROMOSOME CANC, V45, P213, DOI 10.1002/gcc.20283
   Cascon A, 2009, J CLIN ENDOCR METAB, V94, P1701, DOI 10.1210/jc.2008-2756
   Dahia PLM, 2014, NAT REV CANCER, V14, P108, DOI 10.1038/nrc3648
   Else T., 2018, HEREDITARY PARAGANGL
   Farrugia F A, 2017, Endocr Regul, V51, P168, DOI 10.1515/enr-2017-0018
   Favier J, 2015, NAT REV ENDOCRINOL, V11, P101, DOI 10.1038/nrendo.2014.188
   Fishbein L, 2017, ENDOCR-RELAT CANCER, V24, pL51, DOI 10.1530/ERC-17-0086
   Fishbein L, 2016, HEMATOL ONCOL CLIN N, V30, P135, DOI 10.1016/j.hoc.2015.09.006
   Fishbein L, 2012, CANCER GENET-NY, V205, P1, DOI 10.1016/j.cancergen.2012.01.009
   Furlan A, 2017, SCIENCE, V357, P46, DOI 10.1126/science.aal3753
   Garcia-Carbonero R, 2021, CLIN TRANSL ONCOL, V23, P1995, DOI 10.1007/s12094-021-02622-9
   Gimenez-Roqueplo AP, 2012, HORM METAB RES, V44, P328, DOI 10.1055/s-0031-1301302
   Gimenez-Roqueplo AP, 2006, CLIN ENDOCRINOL, V65, P699, DOI 10.1111/j.1365-2265.2006.02714.x
   Greer MLC, 2017, CLIN CANCER RES, V23, pE6, DOI 10.1158/1078-0432.CCR-17-0515
   Hes FJ, 2010, BMC MED GENET, V11, DOI 10.1186/1471-2350-11-92
   Jain A, 2020, PEDIATR NEPHROL, V35, P581, DOI 10.1007/s00467-018-4181-2
   Kastriti ME, 2019, FRONT MOL NEUROSCI, V12, DOI 10.3389/fnmol.2019.00006
   Lenders JWM, 2002, JAMA-J AM MED ASSOC, V287, P1427, DOI 10.1001/jama.287.11.1427
   McWhinney SR, 2004, J CLIN ENDOCR METAB, V89, P5694, DOI 10.1210/jc.2004-0769
   Muth A, 2019, J INTERN MED, V285, P187, DOI 10.1111/joim.12869
   Muth A, 2012, WORLD J SURG, V36, P1389, DOI 10.1007/s00268-012-1430-6
   Neumann HPH, 2002, NEW ENGL J MED, V346, P1459, DOI 10.1056/NEJMoa020152
   Oleaga Amelia, 2008, Endocrinol Nutr, V55, P202
   Plouin PF, 2016, EUR J ENDOCRINOL, V174, pG1, DOI 10.1530/EJE-16-0033
   Pryma DA, 2019, J NUCL MED, V60, P623, DOI 10.2967/jnumed.118.217463
   Richards S, 2015, GENET MED, V17, P405, DOI 10.1038/gim.2015.30
   Ricketts CJ, 2010, HUM MUTAT, V31, P41, DOI 10.1002/humu.21136
   Rijken JA, 2016, CLIN GENET, V89, P128, DOI 10.1111/cge.12591
   Roman-Gonzalez A, 2018, ANN SURG, V268, P172, DOI 10.1097/SLA.0000000000002195
   Tabebi M, 2022, INT J MOL SCI, V23, DOI 10.3390/ijms23010560
   Tanabe A, 2020, HYPERTENS RES, V43, P1141, DOI 10.1038/s41440-020-0531-0
   van Leeuwaarde R. S, 2000, GENEREVIEWS
   Welander J, 2011, ENDOCR-RELAT CANCER, V18, pR253, DOI 10.1530/ERC-11-0170
   Wiseman D, 2019, HORM METAB RES, V51, P470, DOI 10.1055/a-0926-3618
NR 47
TC 0
Z9 0
U1 1
U2 1
PU FRONTIERS MEDIA SA
PI LAUSANNE
PA AVENUE DU TRIBUNAL FEDERAL 34, LAUSANNE, CH-1015, SWITZERLAND
EI 1664-8021
J9 FRONT GENET
JI Front. Genet.
PD JAN 4
PY 2023
VL 13
AR 999329
DI 10.3389/fgene.2022.999329
PG 12
WC Genetics & Heredity
WE Science Citation Index Expanded (SCI-EXPANDED)
SC Genetics & Heredity
GA 8E1OV
UT WOS:000918752100001
PM 36685941
OA Green Published, gold
DA 2023-08-27
ER

PT J
AU Ota, Y
   Naganawa, S
   Kurokawa, R
   Bapuraj, JR
   Capizzano, A
   Kim, J
   Moritani, T
   Srinivasan, A
AF Ota, Y.
   Naganawa, S.
   Kurokawa, R.
   Bapuraj, J. R.
   Capizzano, A.
   Kim, J.
   Moritani, T.
   Srinivasan, A.
TI Assessment of MR Imaging and CT in Differentiating Hereditary and
   Nonhereditary Paragangliomas
SO AMERICAN JOURNAL OF NEURORADIOLOGY
LA English
DT Article
ID DIFFUSION-COEFFICIENT VALUES; NECK PARAGANGLIOMAS; INTEROBSERVER
   VARIABILITY; HEAD; DIAGNOSIS; BENIGN
AB BACKGROUND AND PURPOSE:
   Head and neck paragangliomas have been reported to be associated with mutations of the succinate dehydrogenase enzyme family. The aim of this study was to assess whether radiologic features could differentiate between paragangliomas in the head and neck positive and negative for the succinate dehydrogenase mutation.
   MATERIALS AND METHODS:
   This single-center retrospective review from January 2015 to January 2020 included 40 patients with 48 paragangliomas (30 tumors positive for succinate dehydrogenase mutation in 23 patients and 18 tumors negative for the succinate dehydrogenase mutation in 17 patients). ADC values and tumor characteristics on CT and MR imaging were evaluated by 2 radiologists. Differences between the 2 cohorts in the diagnostic performance of ADC and normalized ADC (ratio to ADC in the medulla oblongata) values were evaluated using the independent samples t test. P < .05 was considered significant.
   RESULTS:
   ADC(mean) (1.07 [SD, 0.25]/1.04 [SD, 0.12] versus 1.31 [SD, 0.16]/1.30 [SD, 0.20]? 10(?3) mm(2)/s by radiologists 1 and 2; P < .001), ADC(maximum) (1.49 [SD, 0.27]/1.49 [SD, 0.20] versus 2.01 [SD, 0.16]/1.87 [SD, 0.20] ? 10(?3) mm(2)/s; P < .001), normalized ADC(mean) (1.40 [SD, 0.33]/1.37 [SD, 0.16] versus 1.73 [SD, 0.22]/1.74 [SD, 0.27]; P < .001), and normalized ADC(maximum) (1.95 [SD, 0.37]/1.97 [SD, 0.27] versus 2.64 [SD, 0.22]/2.48 [SD, 0.28]; P < .001) were significantly lower in succinate dehydrogenase mutation?positive than mutation?negative tumors. ADC(minimum,) normalized ADC(minimum), and tumor characteristics were not statistically significant.
   CONCLUSIONS:
   ADC is a promising imaging biomarker that can help differentiate succinate dehydrogenase mutation?positive from mutation?negative paragangliomas in the head and neck.
C1 [Ota, Y.; Naganawa, S.; Bapuraj, J. R.; Capizzano, A.; Kim, J.; Moritani, T.; Srinivasan, A.] Univ Michigan, Dept Radiol, Div Neuroradiol, Ann Arbor, MI 48109 USA.
   [Kurokawa, R.] Univ Tokyo, Grad Sch Med, Dept Radiol, Tokyo, Japan.
C3 University of Michigan System; University of Michigan; University of
   Tokyo
RP Ota, Y (通讯作者)，1500 E Med Ctr Dr,UH B2, Ann Arbor, MI 48109 USA.
EM
RI Ota, Yoshiaki/GYD-3060-2022; Kurokawa, Ryo/ABE-9483-2021; Capizzano,
   Aristides/AAC-5963-2019; Bapuraj, Jayapalli Rajiv/P-2279-2017
OI Kurokawa, Ryo/0000-0002-1186-8900; Capizzano,
   Aristides/0000-0003-1950-8499; Bapuraj, Jayapalli
   Rajiv/0000-0003-3933-203X; Kim, John/0000-0002-9223-7640; Srinivasan,
   Ashok/0000-0002-4400-1959; naganawa, shotaro/0000-0001-8487-4853; Ota,
   Yoshiaki/0000-0001-8992-2156
CR Ahlawat S, 2016, J MAGN RESON IMAGING, V43, P446, DOI 10.1002/jmri.24994
   Aschenbach R, 2009, CLIN NEURORADIOL, V19, P215, DOI 10.1007/s00062-009-9004-1
   Chang CA, 2016, CANCER IMAGING, V16, DOI 10.1186/s40644-016-0084-2
   Farrugia F A, 2017, Endocr Regul, V51, P168, DOI 10.1515/enr-2017-0018
   Feng N, 2009, WORLD J GASTROENTERO, V15, P3003, DOI 10.3748/wjg.15.3003
   G?ne? A., 2019, ACTA ONCOLOGICA TURC, V52, P416, DOI [10.5505/aot.2019.19971, DOI 10.5505/AOT.2019.19971]
   Han X, 2017, J MAGN RESON IMAGING, V45, P722, DOI 10.1002/jmri.25405
   Hiwatashi A, 2003, AM J ROENTGENOL, V181, P1705, DOI 10.2214/ajr.181.6.1811705
   Koontz NA, 2017, AM J ROENTGENOL, V208, P1110, DOI 10.2214/AJR.16.16486
   LANDIS JR, 1977, BIOMETRICS, V33, P159, DOI 10.2307/2529310
   Neumann HPH, 2002, NEW ENGL J MED, V346, P1459, DOI 10.1056/NEJMoa020152
   Offergeld C, 2012, CLINICS, V67, P19, DOI 10.6061/clinics/2012(Sup01)05
   Patel D, 2020, ANN SURG ONCOL, V27, P1329, DOI 10.1245/s10434-020-08220-3
   Srinivasan A, 2008, AM J NEURORADIOL, V29, P40, DOI 10.3174/ajnr.A0743
   Telischi FF, 2000, OTOLARYNG HEAD NECK, V122, P358, DOI 10.1016/S0194-5998(00)70048-9
   Thoeny HC, 2012, RADIOLOGY, V263, P19, DOI 10.1148/radiol.11101821
   Tischler AS, 2015, ENDOCR-RELAT CANCER, V22, pT123, DOI 10.1530/ERC-15-0261
   VANGILS APG, 1994, AM J ROENTGENOL, V162, P147, DOI 10.2214/ajr.162.1.8273654
   Williams MD, 2017, HEAD NECK PATHOL, V11, P278, DOI 10.1007/s12105-017-0803-4
   Williams Michelle D, 2014, Surg Pathol Clin, V7, P543, DOI 10.1016/j.path.2014.08.007
   Withey SJ, 2019, RADIOGRAPHICS, V39, P1393, DOI 10.1148/rg.2019180151
   Woolen S, 2016, NEUROIMAG CLIN N AM, V26, P259, DOI 10.1016/j.nic.2015.12.005
   Yuan Y, 2016, BMC MED IMAGING, V16, DOI 10.1186/s12880-016-0114-3
NR 23
TC 10
Z9 10
U1 0
U2 2
PU AMER SOC NEURORADIOLOGY
PI DENVILLE
PA PO BOX 3000, DENVILLE, NJ 07834-9349 USA
SN 0195-6108
EI 1936-959X
J9 AM J NEURORADIOL
JI Am. J. Neuroradiol.
PD JUL
PY 2021
VL 42
IS 7
BP 1320
EP 1326
DI 10.3174/ajnr.A7166
EA MAY 2021
PG 7
WC Clinical Neurology; Neuroimaging; Radiology, Nuclear Medicine & Medical
   Imaging
WE Science Citation Index Expanded (SCI-EXPANDED)
SC Neurosciences & Neurology; Radiology, Nuclear Medicine & Medical Imaging
GA UB0SZ
UT WOS:000655517800001
PM 33985956
OA hybrid, Green Published
DA 2023-08-27
ER

PT J
AU Letizia, C
   De Toma, G
   Caliumi, C
   Cerci, S
   Massa, R
   Di Iorio, R
   Alo, P
   Marinoni, E
   Diacinti, D
   D'Erasmo, E
AF Letizia, C
   De Toma, G
   Caliumi, C
   Cerci, S
   Massa, R
   Di Iorio, R
   Alo, P
   Marinoni, E
   Diacinti, D
   D'Erasmo, E
TI Plasma adrenomedullin concentrations in patients with adrenal
   pheochromocytoma
SO HORMONE AND METABOLIC RESEARCH
LA English
DT Article
DE adrenal gland; adrenomedullin; catecholamine; hypertension;
   pheochromocytoma
ID IMMUNOREACTIVE ADRENOMEDULLIN; HYPOTENSIVE PEPTIDE; CATECHOLAMINES;
   SOMATOSTATIN; GLANDS
AB Background: The hypotensive peptide adrenomedullin was first isolated in extracts of human pheochromocytoma. There is, however, no information available on the behaviour of circulating adrenomedullin or on the correlation with catecholamines in patients with pheochromocytoma. Objectives: I)to investigate whether plasma adrenomedullin levels were changed in 10 patients with pheochromocytoma when compared to 21 healthy subjects and 16 patients with essential hypertension; 2) to determine whether or not adrenomedullin has a counterregulatory role in catecholamine excess in pheochromocytoma or is responsible for hemodynamic modifications before and after tumour resection; 3) to determine tissue distribution of iradrenomedullin in the pheochromocytoma. Methods: Plasma adrenomedullin and catecholamine levels were measured in all patients with pheochromocytoma before and four weeks after tumour removal. In the four patients undergoing resection of tumours, plasma levels of adrenomedullin were measured at different time-points during surgery. Results: The mean plasma adrenomedullin concentrations (+/- SD) in patients with pheochromocytoma (37.9 +/- 6 pg/ml) were significantly higher (p < 0.0001) than those in normal subjects (13.7 +/- 6.1 pg/ml) and patients with essential hypertension (22.5 +/- 9.1 pg/ml). Adrenomedullin levels correlated with plasma noradrenaline (r = 0.516, p = 0.0124). In all patients with pheochromocytoma, plasma adrenomedullin concentrations decreased after removal of tumours (from 37.9 +/- 6 to 10.9 +/- 4.6 pg/ml; p ( 0.0001). In the four patients studied during surgery, baseline plasma adrenomedullin and noradrenaline levels were markedly elevated, and increased significantly with tumour manipulation, decreasing 24 hours after operation. Adrenal medulla cells surrounding the pheochromocytoma site stained for ir-adrenomedullin, whereas only isolated cells of pheochromocytoma stained for the peptide. Conclusions: This study demonstrates that circulating adrenomedullin is increased in pheochromocytoma, and is also correlated with plasma noradrenaline levels. Adrenomedullin may represent an additional biochemical parameter for clinical monitoring of patients with pheochromocytoma.
C1 Univ Roma La Sapienza, Chair Internal Med, Dept Clin Sci, Rome, Italy.
   Univ Roma La Sapienza, Inst Clin Surg, Rome, Italy.
   Univ Roma La Sapienza, Inst Radiol, Rome, Italy.
   Univ Roma La Sapienza, Inst Obstet & Gynaecol 2, Rome, Italy.
   Univ Roma La Sapienza, Dept Expt Med & Pathol, Rome, Italy.
C3 CIVIS; Sapienza University Rome; CIVIS; Sapienza University Rome; CIVIS;
   Sapienza University Rome; CIVIS; Sapienza University Rome; CIVIS;
   Sapienza University Rome
RP Letizia, C (通讯作者)，Policlin Umberto 1, Dept Clin Sci, Viale Policlin 32, I-00161 Rome, Italy.
EM
OI DIACINTI, Daniele/0000-0001-6894-3433; Letizia,
   Claudio/0000-0003-4397-0624
CR Andreis PG, 1997, J CLIN ENDOCR METAB, V82, P1167, DOI 10.1210/jc.82.4.1167
   BALDI E, 1988, J ENDOCRINOL INVEST, V11, P133, DOI 10.1007/BF03350121
   Champion HC, 1996, HYPERTENSION, V28, P1041, DOI 10.1161/01.HYP.28.6.1041
   CROUT JR, 1964, J CLIN INVEST, V43, P94, DOI 10.1172/JCI104898
   Di Iorio R, 1998, HYPERTENSION, V32, P758, DOI 10.1161/01.HYP.32.4.758
   DiIorio R, 1997, LANCET, V349, P328
   HERRERA MF, 1992, ARCH SURG-CHICAGO, V127, P105
   Hinson JP, 1998, HORM METAB RES, V30, P334, DOI 10.1055/s-2007-978894
   HISHMITSU T, 1994, J CLIN INVEST, V94, P2158
   ICHIKI Y, 1994, FEBS LETT, V338, P6, DOI 10.1016/0014-5793(94)80106-1
   ISHIYAMA Y, 1993, EUR J PHARMACOL, V241, P271, DOI 10.1016/0014-2999(93)90214-3
   Januszewicz W, 1998, J HYPERTENS, V16, P543, DOI 10.1097/00004872-199816040-00017
   Kapas S, 1998, J ENDOCRINOL, V156, P477, DOI 10.1677/joe.0.1560477
   KATO F, 1994, J HYPERTENS       S3, V12, pS238
   KATOH F, 1995, J NEUROCHEM, V64, P459
   KITAMURA K, 1993, BIOCHEM BIOPH RES CO, V192, P553, DOI 10.1006/bbrc.1993.1451
   Kohno M, 1996, HYPERTENSION, V27, P102, DOI 10.1161/01.HYP.27.1.102
   Letizia C, 1998, CLIN ENDOCRINOL, V48, P145, DOI 10.1046/j.1365-2265.1998.3531170.x
   Letizia C., 1998, Blood Pressure, V7, P19
   Liu J, 1997, J ENDOCRINOL, V155, P483, DOI 10.1677/joe.0.1550483
   MARINONI E, 1995, AM J REPROD IMMUNOL, V34, P213, DOI 10.1111/j.1600-0897.1995.tb00943.x
   NAKANISHI N, 1995, J NEUROCHEM, V64, P600
   NEWELL KA, 1988, SURGERY, V104, P1064
   NISHIKIMI T, 1994, HYPERTENSION, V24, P600, DOI 10.1161/01.HYP.24.5.600
   SANO T, 1983, CANCER, V52, P282, DOI 10.1002/1097-0142(19830715)52:2<282::AID-CNCR2820520215>3.0.CO;2-J
   SATO K, 1995, LIFE SCI, V57, P189, DOI 10.1016/0024-3205(95)00259-9
   SATOH F, 1995, J CLIN ENDOCR METAB, V80, P1750, DOI 10.1210/jc.80.5.1750
NR 27
TC 24
Z9 27
U1 0
U2 1
PU GEORG THIEME VERLAG KG
PI STUTTGART
PA RUDIGERSTR 14, D-70469 STUTTGART, GERMANY
SN 0018-5043
EI 1439-4286
J9 HORM METAB RES
JI Horm. Metab. Res.
PD MAY
PY 2001
VL 33
IS 5
BP 290
EP 294
DI 10.1055/s-2001-15281
PG 5
WC Endocrinology & Metabolism
WE Science Citation Index Expanded (SCI-EXPANDED)
SC Endocrinology & Metabolism
GA 446RF
UT WOS:000169525700006
PM 11440275
DA 2023-08-27
ER

PT J
AU Yang, JY
   Kwon, JW
   Shin, SH
   Yoo, H
   Wang, KC
   Lee, SH
   Gwak, HS
AF Yang, Jeyul
   Kwon, Ji-Woong
   Shin, Sang Hoon
   Yoo, Heon
   Wang, Kyu-Chang
   Lee, Sang Heyon
   Gwak, Ho-Shin
TI Leptomeningeal Metastasis in Gliomas : Clinical Characteristics and Risk
   Factors
SO JOURNAL OF KOREAN NEUROSURGICAL SOCIETY
LA English
DT Article
DE Glioma; Leptomeningeal metastasis; Risk factors; Magnetic resonance
   imaging
ID STEREOTACTIC RADIOSURGERY; ULTRASONIC ASPIRATOR; DISSEMINATION;
   GLIOBLASTOMA; CANCER; SURVIVAL
AB Objective : Our objective is to analyze the occurrence, clinical course and risk factors for glioma patients with leptomeningeal metastasis (LM) according to different metastasis patterns and clinical variables. Methods : We retrospectively reviewed data from 376 World Health Organization (WHO) grade II-IV adult glioma patients who were treated in the National Cancer Center from 2001 to 2020. Patients who underwent surgery at other institutions, those without initial images or those with pathologically unconfirmed cases were excluded. LM was diagnosed based on magnetic resonance imaging (MRI) findings or cerebrospinal fluid (CSF) cytology. The metastasis pattern was categorized as nodular or linear according to the enhancement pattern. Tumor proximity to the CSF space was classified as involved or separated, whereas location of the tumor was dichotomized as midline, for tumors residing in the thalamus, basal ganglia and brainstem, or lateral, for tumors residing in the cerebral and cerebellar hemispheres. Results : A total of 138 patients were enrolled in the study. A total of 44 patients (38%) were diagnosed with LM during a median follow-up of 9 months (range, 0-60). Among the clinical variables, tumor proximity to CSF space, the location of the tumor and the WHO grade were significant factors for LM development in univariate analysis. In multivariate analysis, the midline location of the tumor and WHO grade IV gliomas were the most significant factor for LM development. The hazard ratio was 2.624 for midline located gliomas (95% confidence interval [CI], 1.384-4.974; p=0.003) and 3.008 for WHO grade IV gliomas (95% CI, 1.379-6.561; p=0.006). Conclusion : Midline location and histological grading are an important factor for LM in glioma patients. The proximity to the CSF circulation pathway is also an important factor for WHO grade IV glioma LM. Patients carrying high risks should be followed up more thoroughly.
C1 [Yang, Jeyul] Myongji Hosp, Dept Neurosurg, Goyang, South Korea.
   [Kwon, Ji-Woong; Shin, Sang Hoon; Yoo, Heon; Wang, Kyu-Chang; Gwak, Ho-Shin] Natl Canc Ctr, Neurooncol Clin, Goyang, South Korea.
   [Lee, Sang Heyon] Natl Canc Ctr, Dept Radiol, Goyang, South Korea.
   [Gwak, Ho-Shin] Grad Sch Canc Sci & Policy, Dept Canc Control, Goyang, South Korea.
   [Gwak, Ho-Shin] Grad Sch Canc Sci & Policy, Dept Canc Control, 323 Ilsan Ro, Goyang 10408, South Korea.
C3 Myongji Hospital; National Cancer Center - Korea (NCC); National Cancer
   Center - Korea (NCC)
RP Gwak, HS (通讯作者)，Grad Sch Canc Sci & Policy, Dept Canc Control, 323 Ilsan Ro, Goyang 10408, South Korea.
EM
FU National Cancer Center, Korea [2210650-2]
FX ? Acknowledgements This work was supported by a grant (2210650-2) from
   the National Cancer Center, Korea. The sponsor had no role in the design
   or conduct of this research.
CR Ahn JH, 2012, J NEUROSURG, V116, P984, DOI 10.3171/2012.1.JNS111560
   Andersen BM, 2019, NEUROLOGY, V92, pE2483, DOI 10.1212/WNL.0000000000007529
   AWAD I, 1986, NEUROSURGERY, V19, P247, DOI 10.1227/00006123-198608000-00010
   Bae Jin Woo, 2017, Brain Tumor Res Treat, V5, P99, DOI 10.14791/btrt.2017.5.2.99
   Bae JS, 2011, J KOREAN NEUROSURG S, V49, P334, DOI 10.3340/jkns.2011.49.6.334
   Brower JV, 2016, J CLIN NEUROSCI, V27, P130, DOI 10.1016/j.jocn.2015.11.012
   Bush NAO, 2017, NEUROSURG REV, V40, P1, DOI 10.1007/s10143-016-0709-8
   Cinalli G, 2017, J NEUROSURG-PEDIATR, V19, P325, DOI 10.3171/2016.10.PEDS16352
   Bordignon KC, 2006, SURG NEUROL, V65, P472, DOI 10.1016/j.surneu.2005.08.019
   Dardis C, 2014, FRONT NEUROL, V5, DOI 10.3389/fneur.2014.00220
   EADE OE, 1971, J PATHOL, V103, P245, DOI 10.1002/path.1711030407
   FREILICH RJ, 1995, ANN NEUROL, V38, P51, DOI 10.1002/ana.410380111
   Hansen N, 2011, EUR J MED RES, V16, P415, DOI 10.1186/2047-783X-16-9-415
   Jung JM, 2012, J KOREAN NEUROSURG S, V52, P193, DOI 10.3340/jkns.2012.52.3.193
   Louis DN, 2018, ACTA NEUROPATHOL, V135, P639, DOI 10.1007/s00401-018-1826-y
   Mandel JJ, 2014, J NEURO-ONCOL, V120, P597, DOI 10.1007/s11060-014-1592-1
   Michotte A, 2009, J NEUROL SCI, V287, P267, DOI 10.1016/j.jns.2009.08.047
   MOR V, 1984, CANCER-AM CANCER SOC, V53, P2002, DOI 10.1002/1097-0142(19840501)53:9<2002::AID-CNCR2820530933>3.0.CO;2-W
   Nandipati S, 2011, J NEURO-ONCOL, V105, P663, DOI 10.1007/s11060-011-0611-8
   ONDA K, 1989, NEUROSURGERY, V25, P533, DOI 10.1227/00006123-198910000-00005
   Parsa AT, 2005, J NEUROSURG, V102, P622, DOI 10.3171/jns.2005.102.4.0622
   Preston JK, 1999, BRIT J NEUROSURG, V13, P486, DOI 10.1080/02688699908540623
   Qiu TM, 2020, J NEUROSURG, V133, P1662, DOI 10.3171/2019.9.JNS191920
   Roelz R, 2015, SCI REP-UK, V5, DOI 10.1038/srep17758
   Roldan G, 2011, J NEURO-ONCOL, V104, P811, DOI 10.1007/s11060-011-0551-3
   Sreenivasan SA, 2016, J CANCER RES THER, V12, P161, DOI 10.4103/0973-1482.153999
   Suki D, 2008, J NEUROSURG, V108, P248, DOI 10.3171/JNS/2008/108/2/0248
   Suki D, 2009, NEUROSURGERY, V64, P664, DOI 10.1227/01.NEU.0000341535.53720.3E
NR 28
TC 0
Z9 0
U1 0
U2 0
PU KOREAN NEUROSURGICAL SOC
PI SEOUL
PA BLDG B, #402, 27, CHILPAE-RO, JUNG-GU, SEOUL, 04511, SOUTH KOREA
SN 2005-3711
EI 1598-7876
J9 J KOREAN NEUROSURG S
JI J. Korean. Neurosurg. Soc.
PD JUL
PY 2023
VL 66
IS 4
BP 465
EP 475
DI 10.3340/jkns.2022.0166
PG 11
WC Clinical Neurology; Surgery
WE Science Citation Index Expanded (SCI-EXPANDED)
SC Neurosciences & Neurology; Surgery
GA N6RS5
UT WOS:001038266400013
PM 36503217
OA Green Published, gold
DA 2023-08-27
ER

PT J
AU Alford, R
   Gargan, L
   Bowers, DC
   Klesse, LJ
   Weprin, B
   Koral, K
AF Alford, Raphael
   Gargan, Lynn
   Bowers, Daniel C.
   Klesse, Laura J.
   Weprin, Bradley
   Koral, Korgun
TI Postoperative surveillance of pediatric cerebellar pilocytic astrocytoma
SO JOURNAL OF NEURO-ONCOLOGY
LA English
DT Article
DE Pilocytic astrocytoma; Children; Cerebellar; Surveillance; Magnetic
   resonance imaging
ID LOW-GRADE ASTROCYTOMA; CHILDREN
AB The purpose of this study was to identify the optimal frequency and duration of magnetic resonance imaging follow-up in children who had gross totally resected cerebellar pilocytic astrocytomas (CPAs). Our hypothesis was that following two MR examinations, separated by at least 3 months, showing no evidence of tumor, gross totally resected CPAs did not recur and no further imaging follow-up was necessary. Retrospective review of Neuro-Oncology database from 1/2000 to 7/2013 yielded 53 patients with CPAs that had preoperative imaging and > 2 years post-operative imaging follow-up available. Pilocytic astrocytomas with brainstem involvement and patients with neurofibromatosis type I were excluded. Preoperative tumor volumes were calculated. The dates and reports of the examinations were tabulated. The median number of follow-up examinations was 9 over a median follow-up time of 6.05 years (2.07-12.28 years). Two consecutive MR examinations over at least a 3 month span demonstrated the smallest negative likelihood ratio of future recurrence (0.15). There was no association of recurrence with preoperative tumor volume. Among the 35 patients with gross total resection of their tumor and greater than two negative follow-up examinations, one recurrence (2.9 %) was identified, occurring 6.4 years after initial resection. Gross totally resected pediatric CPAs can recur, but this is exceedingly rare. Frequent surveillance (every 3-6 months) is suggested in patients with CPAs until absence of tumor is concluded on imaging and documented on two consecutive studies spaced at least 3 months apart. The likelihood of recurrence thereafter is low.
C1 [Alford, Raphael; Koral, Korgun] Univ Texas Southwestern Med Ctr Dallas, Dept Radiol, 1935 Med Dist Dr, Dallas, TX 75235 USA.
   [Koral, Korgun] Childrens Hlth, Dept Radiol, Dallas, TX 75235 USA.
   [Gargan, Lynn] Childrens Hlth, Dept Neurooncol, Dallas, TX 75235 USA.
   [Bowers, Daniel C.; Klesse, Laura J.] Univ Texas Southwestern Med Ctr Dallas, Dept Pediat, Dallas, TX 75235 USA.
   [Bowers, Daniel C.; Klesse, Laura J.] Childrens Hlth, Dept Pediat, Dallas, TX 75235 USA.
   [Weprin, Bradley] Univ Texas Southwestern Med Ctr Dallas, Dept Neurosurg, Dallas, TX 75235 USA.
   [Weprin, Bradley] Childrens Hlth, Dept Neurosurg, Dallas, TX 75235 USA.
C3 University of Texas System; University of Texas Southwestern Medical
   Center Dallas; University of Texas System; University of Texas
   Southwestern Medical Center Dallas; University of Texas System;
   University of Texas Southwestern Medical Center Dallas
RP Koral, K (通讯作者)，Univ Texas Southwestern Med Ctr Dallas, Dept Radiol, 1935 Med Dist Dr, Dallas, TX 75235 USA.; Koral, K (通讯作者)，Childrens Hlth, Dept Radiol, Dallas, TX 75235 USA.
EM
OI Klesse, Laura/0000-0003-1323-7720; Bowers, Daniel/0000-0002-3947-2481
CR Cravero JP, 2009, ANESTH ANALG, V108, P795, DOI 10.1213/ane.0b013e31818fc334
   Due-Tonnessen BJ, 2013, J NEUROSURG-PEDIATR, V11, P245, DOI 10.3171/2012.11.PEDS12265
   Gunny RS, 2005, PEDIATR RADIOL, V35, P1086, DOI 10.1007/s00247-005-1546-z
   Ing C, 2012, PEDIATRICS, V130, pE476, DOI 10.1542/peds.2011-3822
   Ogiwara H, 2012, NEUROSURGERY, V70, P40, DOI 10.1227/NEU.0b013e31822ff0ed
   Rodriguez FJ, 2013, ANNU REV PATHOL-MECH, V8, P361, DOI 10.1146/annurev-pathol-020712-164009
   Saunders DE, 2005, J NEUROSURG, V102, P172, DOI 10.3171/jns.2005.102.2.0172
   Scheithauer BW, 2007, WHO CLASSIFICATION T, P14
   Sutton LN, 1996, J NEUROSURG, V84, P721, DOI 10.3171/jns.1996.84.5.0721
   Udaka YT, 2013, J NEUROSURG-PEDIATR, V11, P119, DOI 10.3171/2012.10.PEDS12307
   Vassilyadi M, 2009, CAN J NEUROL SCI, V36, P707, DOI 10.1017/S0317167100008313
NR 11
TC 7
Z9 7
U1 0
U2 3
PU SPRINGER
PI NEW YORK
PA 233 SPRING ST, NEW YORK, NY 10013 USA
SN 0167-594X
EI 1573-7373
J9 J NEURO-ONCOL
JI J. Neuro-Oncol.
PD OCT
PY 2016
VL 130
IS 1
BP 149
EP 154
DI 10.1007/s11060-016-2222-x
PG 6
WC Oncology; Clinical Neurology
WE Science Citation Index Expanded (SCI-EXPANDED)
SC Oncology; Neurosciences & Neurology
GA EA4FH
UT WOS:000386565200016
PM 27502785
DA 2023-08-27
ER

PT J
AU Mielke, C
   Alexander, MSM
   Anand, N
AF Mielke, C
   Alexander, MSM
   Anand, N
TI Isolated bilateral trochlear nerve palsy as the first clinical sign of a
   metastasic bronchial carcinoma
SO AMERICAN JOURNAL OF OPHTHALMOLOGY
LA English
DT Article
AB PURPOSE: To report a case with isolated, nontraumatic bilateral fourth nerve palsy as the first clinical sign of a metastatic lung carcinoma.
   METHODS: Case report. A 56-year-old man presented with isolated, nontraumatic bilateral fourth nerve palsy. Magnetic resonance imaging (MRI) of the brain and orbits and, subsequently, chest x-ray and a computer tomographic (CT)-scan of the thorax, the abdomen, and the pelvis were performed.
   RESULTS: Magnetic resonance imaging confirmed the presence of a midline brain stem lesion in the region of decussation of the trochlear nerves. Computed tomographic scan of the chest revealed that the lesion was caused by a metastatic lung carcinoma.
   CONCLUSION: The findings of isolated bilateral fourth nerve palsy in the absence of trauma should alert the clinician to the possibility of a posterior fossa lesion in the region of the trochlear nerves. Besides urgent scanning of the dorsal midbrain, investigations should be directed to search for the primary tumor. (C) 2001 by Elsevier Science Inc. All rights reserved.
C1 Luton & Dunstable Hosp, Dept Ophthalmol, Luton LU4 0DZ, Beds, England.
   Luton & Dunstable Hosp, Dept Radiol, Luton LU4 0DZ, Beds, England.
RP Anand, N (通讯作者)，Luton & Dunstable Hosp, Dept Ophthalmol, Luton LU4 0DZ, Beds, England.
CR Barr D B, 1997, Eur J Ophthalmol, V7, P271
   KEANE JR, 1993, NEUROLOGY, V43, P2439, DOI 10.1212/WNL.43.12.2439
   MURRAY RS, 1985, J CLIN NEURO-OPHTHAL, V5, P57
   RICHARDS BW, 1992, AM J OPHTHALMOL, V113, P489, DOI 10.1016/S0002-9394(14)74718-X
   SAWLE GV, 1989, J ROY SOC MED, V82, P111, DOI 10.1177/014107688908200221
NR 5
TC 4
Z9 5
U1 0
U2 1
PU ELSEVIER SCIENCE INC
PI NEW YORK
PA 655 AVENUE OF THE AMERICAS, NEW YORK, NY 10010 USA
SN 0002-9394
J9 AM J OPHTHALMOL
JI Am. J. Ophthalmol.
PD OCT
PY 2001
VL 132
IS 4
BP 593
EP 594
DI 10.1016/S0002-9394(01)01027-3
PG 2
WC Ophthalmology
WE Science Citation Index Expanded (SCI-EXPANDED)
SC Ophthalmology
GA 481QJ
UT WOS:000171530700030
PM 11589894
DA 2023-08-27
ER

PT J
AU Scollo, C
   Russo, M
   De Gregorio, L
   Terranova, R
   Mangione, E
   Castoro, C
   Squatrito, S
   Pellegriti, G
AF Scollo, Claudia
   Russo, Marco
   De Gregorio, Laura
   Terranova, Rosa
   Mangione, Erika
   Castoro, Carlotta
   Squatrito, Sebastiano
   Pellegriti, Gabriella
TI A novel RET gene mutation in a patient with apparently sporadic
   pheochromocytoma
SO ENDOCRINE JOURNAL
LA English
DT Article
DE Pheochromocytoma; MEN 2A; RET mutation
ID ENDOCRINE NEOPLASIA TYPE-2; GERM-LINE MUTATIONS; CLINICAL-FEATURES;
   DIAGNOSIS; PROTOONCOGENE; GUIDELINES
AB Pheochromocytoma (Pheo) is a chromaffin tumor arising from the adrenal medulla. The recent discovery of new germline mutations in RET, SDHA, SDHB, SDHC, SDHD, VHL, NF1, TMEM127, MAX genes, increased the rate of genetic disease from 10% to 28% in patients with apparently sporadic tumor. RET germline mutations cause multiple endocrine neoplasia type 2 syndrome (MEN 2A) characterized by complete penetrance of medullary thyroid cancer (MTC), and lower prevalence of Pheo and hyperparathyroidism. We describe the genetic etiology of an apparently sporadic case of monolateral Pheo in a 42-year-old male patient. A new (not previously reported) MEN 2A-associated germline RET mutation located in exon 11 (G1u632Gly, caused by an A>G point mutation at position 1895 of the RET cDNA) was found in the patient but not in his living first-degree relatives. This observation increases the number of possible germline RET mutations. Genotype-phenotype correlation of this new genetic alteration is unknown, but this rare mutation is probably associated with a low risk for MTC (usually the first tumor diagnosed in MEN 2A syndrome) and with the development of Pheo before the onset of MTC. Since we expect MTC to occur in our patient, strict follow-up is mandatory. Our findings emphasize the relevance of genetic testing in patients with Pheo, especially when the clinical presentation (family history, young age at diagnosis, multiple locations, malignant lesions, and bilateralism) is suggestive.
C1 [Scollo, Claudia; Pellegriti, Gabriella] Garibaldi Nesima Hosp, Endocrinol, Via Palermo 636, I-95122 Catania, Italy.
   [Russo, Marco] Univ Messina, Dept Clin & Expt Med, Endocrinol, I-98100 Messina, Italy.
   [De Gregorio, Laura; Terranova, Rosa; Mangione, Erika; Castoro, Carlotta; Squatrito, Sebastiano] Univ Catania, Dept Clin & Expt Med, Endocrinol, I-95124 Catania, Italy.
C3 Presidio Ospedaliero Garibaldi-Nesima; University of Messina; University
   of Catania
RP Pellegriti, G (通讯作者)，Garibaldi Nesima Hosp, Endocrinol, Via Palermo 636, I-95122 Catania, Italy.
EM
RI Pellegriti, Gabriella/CAH-1780-2022; Pellegriti, Gabriella/J-7923-2012;
   Russo, Marco/AGY-2396-2022
OI Pellegriti, Gabriella/0000-0001-6102-379X; Pellegriti,
   Gabriella/0000-0001-6102-379X; Russo, Marco/0000-0001-5151-9417
CR [Anonymous], 2004, WHO CLASSIFICATION T
   Brandi ML, 2001, J CLIN ENDOCR METAB, V86, P5658, DOI 10.1210/jc.86.12.5658
   BRAVO EL, 1994, ENDOCR REV, V15, P356, DOI 10.1210/er.15.3.356
   Bryant J, 2003, J NATL CANCER I, V95, P1196, DOI 10.1093/jnci/djg024
   Frank-Raue K, 2007, CLIN LAB, V53, P273
   Gimenez-Roqueplo AP, 2006, CLIN ENDOCRINOL, V65, P699, DOI 10.1111/j.1365-2265.2006.02714.x
   Han Zhan-ying, 2006, Zhonghua Yixue Yichuanxue Zazhi, V23, P320
   Heshmati HM, 1997, EUR J ENDOCRINOL, V137, P572, DOI 10.1530/eje.0.1370572
   Imai T, 2013, EUR J ENDOCRINOL, V168, P683, DOI 10.1530/EJE-12-1106
   Jafri M, 2012, EUR J ENDOCRINOL, V166, P151, DOI 10.1530/EJE-11-0497
   Khorram-Manesh A, 2004, EJSO, V30, P556, DOI 10.1016/j.ejso.2004.03.006
   Kloos RT, 2009, THYROID, V19, P565, DOI 10.1089/thy.2008.0403
   Machens A, 2005, J CLIN ENDOCR METAB, V90, P3999, DOI 10.1210/jc.2005-0064
   Machens A, 2006, ANN NY ACAD SCI, V1073, P417, DOI 10.1196/annals.1353.045
   Miller DD, 2012, MODERN PATHOL, V25, P505, DOI 10.1038/modpathol.2011.196
   Moline J, 2011, GENET MED, V13, P755, DOI 10.1097/GIM.0b013e318216cc6d
   MULLIGAN LM, 1993, NATURE, V363, P458, DOI 10.1038/363458a0
   Neumann HPH, 2002, NEW ENGL J MED, V346, P1459, DOI 10.1056/NEJMoa020152
   Pacak K, 2007, NAT CLIN PRACT ENDOC, V3, P92, DOI 10.1038/ncpendmet0396
   Raue F, 2012, CLINICS, V67, P69, DOI 10.6061/clinics/2012(Sup01)13
   Romei C, 2012, J ONCOL, V2012, DOI 10.1155/2012/705036
   Thosani S, 2013, J CLIN ENDOCR METAB, V98, pE1813, DOI 10.1210/jc.2013-1653
   Wells SA, 2015, THYROID, V25, P567, DOI 10.1089/thy.2014.0335
NR 23
TC 2
Z9 10
U1 0
U2 3
PU JAPAN ENDOCRINE SOC
PI KYOTO
PA 75  YANAGINOBANBA NISHIIRU-MASUYA-CHO, SANJOU-DORI, NAKAGYOU-KU, KYOTO,
   604-8111, JAPAN
SN 0918-8959
EI 1348-4540
J9 ENDOCR J
JI Endocr. J.
PD JAN 20
PY 2016
VL 63
IS 1
BP 87
EP 91
DI 10.1507/endocrj.EJ15-0399
PG 5
WC Endocrinology & Metabolism
WE Science Citation Index Expanded (SCI-EXPANDED)
SC Endocrinology & Metabolism
GA DH3FE
UT WOS:000372672100011
PM 26497911
OA gold
DA 2023-08-27
ER

PT J
AU De Cock, HEV
   MacLachlan, NJ
AF De Cock, HEV
   MacLachlan, NJ
TI Simultaneous occurrence of multiple neoplasms and hyperplasias in the
   adrenal and thyroid gland of the horse resembling multiple endocrine
   neoplasia syndrome: Case report and retrospective identification of
   additional cases
SO VETERINARY PATHOLOGY
LA English
DT Article
DE horses; multiple endocrine neoplasia; pheochromocytoma; C-cell adenoma
ID BULL
AB Neoplastic and hyperplastic disorders that affect multiple endocrine tissues in a single individual are well described in humans but less so in domestic animals. Multiple endocrine neoplasia (MEN) in humans is a genetically determined syndrome characterized by the appearance of benign or malignant proliferations within two or more endocrine glands. The primary endocrine tumors that are characteristic of MEN arise from cells that share the capacity for amine precursor uptake and decarboxylation. Here we describe the case of a 22-year-old Thoroughbred mare that died during an unattended parturition and subsequently was presented for necropsy at the University of California, Davis, Veterinary Medical Teaching Hospital. A C-cell (medullary) thyroid adenoma, pheochromocytoma, and multicentric bilateral nodular hyperplasia of the adrenal medulla were present, findings that are remarkably similar to those of human MEN syndrome. Mortality during pregnancy in women with undiagnosed pheochromocytoma is high (approximately 50%), typically because of hypertension and/or hemorrhage associated with catecholamine release from the tumor. Similarly, the mare in this report died of hemorrhage subsequent to parturition. A retrospective evaluation of endocrine tumors in horses that underwent necropsy at the Veterinary Medical Teaching Hospital from 1987 to 1997 was undertaken to identify additional possible cases of MEN in horses. Data from this retrospective evaluation suggest that coexistence of hyperplasias and neoplasias of the thyroid and adrenal glands, similar to MEN syndrome of humans, also occurs with some frequency in the horse.
C1 Univ Calif Davis, Sch Vet Med, Dept Pathol Microbiol & Immunol, Davis, CA 95616 USA.
C3 University of California System; University of California Davis
RP De Cock, HEV (通讯作者)，Univ Calif Davis, Sch Vet Med, Dept Pathol Microbiol & Immunol, Davis, CA 95616 USA.
CR BLACK HE, 1973, CANCER, V32, P865, DOI 10.1002/1097-0142(197310)32:4&lt;865::AID-CNCR2820320418&gt;3.0.CO;2-T
   DEFTOS LJ, 1992, TXB ENDOCRINOLOGY, P753
   Gagel R.F., 1995, ENDOCRINOL METAB, P1537
   KAISER RH, 1995, PRINCIPLES PRACTICE, P1555
   Kuwamura M, 1998, J VET MED SCI, V60, P387, DOI 10.1292/jvms.60.387
   OLIVRY T, 1995, ARCH DERMATOL RES, V287, P186, DOI 10.1007/BF01262330
   PADBERG BC, 1992, HORM RES, V38, P24, DOI 10.1159/000182588
   PETERSON ME, 1982, J AM VET MED ASSOC, V180, P1476
   SIZEMORE GW, 1995, PRINCIPLES PRACTICE, P767
   SPONENBERG DP, 1983, VET PATHOL, V20, P396, DOI 10.1177/030098588302000402
   WILKIE BN, 1970, PATHOL VET, V7, P126, DOI 10.1177/030098587000700204
   YARRINGTON JT, 1981, VET PATHOL, V18, P316, DOI 10.1177/030098588101800304
NR 12
TC 34
Z9 34
U1 0
U2 8
PU AMER COLL VET PATHOLOGIST
PI LAWRENCE
PA 810 EAST 10TH STREET, LAWRENCE, KS 66044 USA
SN 0300-9858
J9 VET PATHOL
JI Vet. Pathol.
PD NOV
PY 1999
VL 36
IS 6
BP 633
EP 636
DI 10.1354/vp.36-6-633
PG 4
WC Pathology; Veterinary Sciences
WE Science Citation Index Expanded (SCI-EXPANDED)
SC Pathology; Veterinary Sciences
GA 283NH
UT WOS:000085281300020
PM 10568452
DA 2023-08-27
ER

PT J
AU Cascon, A
   Montero-Conde, C
   Ruiz-Liorente, S
   Mercadillo, F
   Leton, R
   Rodriguez-Antona, C
   Martinez-Delgado, B
   Delgado, M
   Diez, A
   Rovira, A
   Diaz, JA
   Robledo, M
AF Cascon, A
   Montero-Conde, C
   Ruiz-Liorente, S
   Mercadillo, F
   Leton, R
   Rodriguez-Antona, C
   Martinez-Delgado, B
   Delgado, M
   Diez, A
   Rovira, A
   Diaz, JA
   Robledo, M
TI Gross SDHB deletions in patients with paraganglioma detected by
   multiplex PCR: A possible hot spot?
SO GENES CHROMOSOMES & CANCER
LA English
DT Article
ID GENE-MUTATIONS; HEREDITARY PARAGANGLIOMA; PHEOCHROMOCYTOMA; EXPRESSION
AB Pheochromocytoma and paraganglioma are rare neuroendocrine tumors that arise in the adrenal medulla and the extra-adrenal paraganglia, respectively. Inheritance of these tumors is mainly a result of mutations affecting the VHL, RET NFI, and SDH genes. Germ-line mutations of the SDH genes have been found to account for nearly 10% of apparently sporadic cases. Nevertheless, alterations other than point mutations have not yet been well characterized. In this study, we investigated the frequency of gross SDH deletions in 24 patients who tested negative for point mutations and had at least one of the recommended features for genetic testing. For this purpose, we used a technique that is easy to implement in the lab to specifically detect gross deletions affecting SDHB, SDHC, and SDHD. We identified 3 heterozygous SDHB deletions (3/24) in 3 independent cases with paraganglioma: 1 whole SDHB deletion and 2 deletions exclusively affecting exon 1. These latter mutations match the unique gross deletion previously reported, indicating this region could be a hot spot for gross SDHB deletions. It seems likely that these alterations can account for a considerable number of both familial and apparently sporadic paraganglioma cases. Although this is the first report describing the presence of gross deletions in patients with apparently sporadic paragangliomas, the extra-adrenal location of the tumor seems to constitute a determining factor for whether to include these patients in genetic testing for gross deletions in the SDHB gene. (c) 2005 Wiley-Liss, Inc.
C1 Ctr Nacl Invest Oncol, Dept Human Genet, Hereditary Endocrine Canc Grp, Madrid 28029, Spain.
   Hosp Bierzo, Dept Endocrinol, Ponferrada, Spain.
   Fdn Jimenez Diaz, Dept Endocrinol, E-28040 Madrid, Spain.
   Hosp Univ Clin San Carlos, Dept Endocrinol, Madrid, Spain.
RP Robledo, M (通讯作者)，Ctr Nacl Invest Oncol, Dept Human Genet, Hereditary Endocrine Canc Grp, Melchor Fernandez Almagro 3, Madrid 28029, Spain.
EM
RI Montero-Conde, Cristina/K-7349-2014; Leton, Rocio/AAD-7896-2020;
   Rodriguez-Antona, Cristina/E-1073-2011; Cascon, Alberto/G-3160-2014;
   Martinez-Delgado, Beatriz/B-6833-2013; Robledo, Mercedes/O-2230-2013
OI Montero-Conde, Cristina/0000-0002-4452-4469; Rodriguez-Antona,
   Cristina/0000-0001-8750-7338; Cascon, Alberto/0000-0003-2119-891X;
   Martinez-Delgado, Beatriz/0000-0001-6834-350X; Robledo,
   Mercedes/0000-0001-6256-5902; Ruiz-Llorente, Sergio/0000-0002-5003-1759
CR Anderson Robert J., 1993, Current Opinion in Oncology, V5, P75
   [Anonymous], MOL CLONING
   Astuti D, 2001, AM J HUM GENET, V69, P49, DOI 10.1086/321282
   Baysal BE, 2004, J MED GENET, V41, P703, DOI 10.1136/jmg.2004.019224
   Baysal BE, 2000, SCIENCE, V287, P848, DOI 10.1126/science.287.5454.848
   Bieche I, 2004, CLIN CANCER RES, V10, P6789, DOI 10.1158/1078-0432.CCR-04-0306
   Dannenberg H, 2005, J CLIN ONCOL, V23, P1894, DOI 10.1200/JCO.2005.07.198
   Gimenez-Roqueplo AP, 2003, CANCER RES, V63, P5615
   Kolomietz E, 2002, GENE CHROMOSOME CANC, V35, P97, DOI 10.1002/gcc.10111
   LACK EE, 1997, TUMORS ADRENAL GLAND, V3
   Maher ER, 1996, J MED GENET, V33, P328, DOI 10.1136/jmg.33.4.328
   McWhinney SR, 2004, J CLIN ENDOCR METAB, V89, P5694, DOI 10.1210/jc.2004-0769
   Neumann HPH, 2002, NEW ENGL J MED, V346, P1459, DOI 10.1056/NEJMoa020152
   Neumann HPH, 2004, JAMA-J AM MED ASSOC, V292, P943, DOI 10.1001/jama.292.8.943
   Niemann S, 2000, NAT GENET, V26, P268, DOI 10.1038/81551
   SCHIMKE RN, 1990, AM J MED GENET, V37, P375, DOI 10.1002/ajmg.1320370317
   Schouten JP, 2002, NUCLEIC ACIDS RES, V30, DOI 10.1093/nar/gnf056
   Vanharanta S, 2004, AM J HUM GENET, V74, P153, DOI 10.1086/381054
NR 18
TC 63
Z9 63
U1 0
U2 9
PU WILEY
PI HOBOKEN
PA 111 RIVER ST, HOBOKEN 07030-5774, NJ USA
SN 1045-2257
EI 1098-2264
J9 GENE CHROMOSOME CANC
JI Gene Chromosomes Cancer
PD MAR
PY 2006
VL 45
IS 3
BP 213
EP 219
DI 10.1002/gcc.20283
PG 7
WC Oncology; Genetics & Heredity
WE Science Citation Index Expanded (SCI-EXPANDED)
SC Oncology; Genetics & Heredity
GA 011KT
UT WOS:000235267800001
PM 16258955
DA 2023-08-27
ER

PT J
AU Briggs, RJS
   Fabinyi, G
   Kaye, AH
AF Briggs, RJS
   Fabinyi, G
   Kaye, AH
TI Current management of acoustic neuromas: review of surgical approaches
   and outcomes
SO JOURNAL OF CLINICAL NEUROSCIENCE
LA English
DT Article
ID CEREBELLOPONTINE ANGLE; VESTIBULAR SCHWANNOMA; RADIOSURGERY; SURGERY
AB The management options for patients with acoustic neuromas is discussed with a review of 164 patients assessed and treated between 1944 and 1998. Twenty-one patients have neurofibromatosis type II. In 33 cases initial observation was undertaken with repealed imaging. Surgical removal of 122 tumours was performed in 121 patients. Eleven of these patients have NF2, of whom three underwent Auditory Brainstem Implantation. Hearing preservation tumour removal was attempted in 37 and was successful in 20 (54%). The middle cranial fossa approach was used in ten cases with 100% successful hearing preservation. The retrosigmoid approach was used in 27 cases with 36% successful hearing preservation. Non-hearing preservation tumour removal was performed in 85 cases where hearing was poor or the tumour measured more than 2 cm within the cerebellopontine angle. The translabyrinthine approach was used in 80 of these patients. Postoperative facial nerve outcome was dependent on tumour size. All 38 patients with tumours less than or equal to1.5 cm have normal (Grade 1) facial function. For all sized tumours, 90% of patients have good facial function (82% Grade 1, 8% Grade 2), 7% of patients have moderate function (6% Grade 3 and 1% Grade 4) and only 3% of patients have poor function (2% Grade 5, 1% Grade 6), Lasting complications were minimal with no operative mortality and eight patients (6.4%) suffering CSF fistulae. Seven patients in this series have had stereotactic radiation with variable outcome. The radiobiology of both single dose and fractionated stereotactic radiation is described and the current role of stereotactic radiation in the management of acoustic neuromas is discussed. Surgical tumour removal by an experienced multi-disciplinary team remains the primary treatment modality for acoustic neuromas. The middle cranial fossa approach is recommended for management of intracanalicular tumours. The translabyrinthine approach facilitates facial nerve preservation, particularly in patients with large tumours. (C) 2000 Harcourt Publishers Ltd.
C1 Univ Melbourne, Dept Otolaryngol, Parkville, Vic 3052, Australia.
   Austin & Repatriat Hosp, Dept Neurosurg, Melbourne, Vic, Australia.
   Univ Melbourne, Royal Melbourne Hosp, Dept Surg, Parkville, Vic 3052, Australia.
   Univ Melbourne, Royal Melbourne Hosp, Dept Neurosurg, Parkville, Vic 3052, Australia.
C3 University of Melbourne; Austin Research Institute; Royal Melbourne
   Hospital; University of Melbourne; Royal Melbourne Hospital; University
   of Melbourne
RP Briggs, RJS (通讯作者)，Univ Melbourne Hosp, Royal Melbourne Hosp, Dept Otolaryngol, 32 Gisborne St, E Melbourne, Vic 3002, Australia.
OI Kaye, Andrew Henry/0000-0001-5049-8861
CR BATTISTA RA, 1999, AM NEUR SOC, P11
   BRACKMANN DE, 1994, AM J OTOL, V15, P614
   BRIGGS RJ, 1997, AUST J OTOLARYNGOL, V2, P455
   BRIGGS RJS, 1994, ARCH OTOLARYNGOL, V120, P1307
   CASSELMAN JW, 1993, AM J NEURORADIOL, V14, P47
   CHARABI S, 1995, OTOLARYNG HEAD NECK, V113, P5, DOI 10.1016/S0194-5998(95)70138-9
   Comey CH, 1998, J NEUROSURG, V89, P653, DOI 10.3171/jns.1998.89.4.0653
   Deen HG, 1996, NEUROSURGERY, V39, P260, DOI 10.1097/00006123-199608000-00005
   DEEN HG, 1996, NEUROSURGERY, V36, P264
   DEW LA, 1996, ACOUSTIC TUMOURS DIA, P205
   Flickinger JC, 1996, INT J RADIAT ONCOL, V36, P275, DOI 10.1016/S0360-3016(96)00335-5
   GARDNER G, 1988, ANN OTO RHINOL LARYN, V97, P55, DOI 10.1177/000348948809700110
   GOLBY AJ, ISRS 4 WORLD C, P69
   HOUSE JW, 1985, OTOLARYNG HEAD NECK, V93, P146, DOI 10.1177/019459988509300202
   LEKSELL L, 1971, ACTA CHIR SCAND, V137, P763
   NANCE WE, 1994, ARCH NEUROL-CHICAGO, V51, P201
   NEDZELSKI JM, 1992, OTOLARYNG CLIN N AM, V25, P691
   NOREN G, 1999, 4 INT STER RAD SOC C
   RIEKE JW, ISRS 4 WORLD C, P57
   Selesnick SH, 1998, AM J OTOL, V19, P846
   SLATTERY WH, 1995, AM J OTOL, V16, P315
   TOMASEVIC P, 1998, AUST J OTOLARYNG, V31, P7
   Weber PC, 1996, AM J OTOL, V17, P669
NR 23
TC 88
Z9 91
U1 0
U2 6
PU CHURCHILL LIVINGSTONE
PI EDINBURGH
PA JOURNAL PRODUCTION DEPT, ROBERT STEVENSON HOUSE, 1-3 BAXTERS PLACE,
   LEITH WALK, EDINBURGH EH1 3AF, MIDLOTHIAN, SCOTLAND
SN 0967-5868
J9 J CLIN NEUROSCI
JI J. Clin. Neurosci.
PD NOV
PY 2000
VL 7
IS 6
BP 521
EP 526
DI 10.1054/jocn.2000.0728
PG 6
WC Clinical Neurology; Neurosciences
WE Science Citation Index Expanded (SCI-EXPANDED)
SC Neurosciences & Neurology
GA 384ME
UT WOS:000165946000009
PM 11029233
DA 2023-08-27
ER

PT J
AU Kafe, H
   Verbavatz, JM
   Cochand-Priollet, B
   Castagnet, P
   Vieillefond, A
AF Kafe, H
   Verbavatz, JM
   Cochand-Priollet, B
   Castagnet, P
   Vieillefond, A
TI Collecting duct carcinoma: an entity to be redefined?
SO VIRCHOWS ARCHIV
LA English
DT Article
DE kidney; collecting duct carcinoma; aquaporin; urothelial carcinoma
ID KIDNEY
AB Collecting duct carcinomas (CDCs) are highly aggressive tumors with poor survival at 1 year and are often metastatic at the time of diagnosis. It has been shown that patients may have better survival when treated with a chemotherapy regimen used for urothelial carcinoma. Such tumors must therefore be recognized, but their pathological diagnosis remains difficult. The two main differential diagnoses are renal pelvis urothelial carcinoma with infiltration of the kidney and/or high-grade and high-stage papillary renal cell carcinoma. The aim of our study was to compare the immunophenotype of 14 CDCs with 6 renal pelvis urothelial carcinomas (RPUC) infiltrating the medulla. The following markers were evaluated: ulex europeus aglutinin (UEA), peanuts aglutinin, vimentin and aquaporin 3 (AQP-3), a membrane component of normal collecting duct and urothelial cells. We were able to define a reproductive urothelial phenotype AQP-3+, vimentin - and UEA+. Among the 14 CDCs, 10 cases demonstrated this immunophenotype. It coincided with an urothelial-like trabecular and tubular pattern. In contrast, the 4 remaining papillary CDCs had the inverse pattern, AQP-3-, vimentin+ and UEA-. These results suggest that: ( 1) the trabecular and tubular variant of CDC with the urothelial AQP-3+, vimentin - phenotype can be included in the spectrum of urothelial diseases; ( 2) the papillary variant probably does not belong to the same entity; ( 3) AQP-3 is a marker of interest for improving the histological classification of CDC and unclassified aggressive renal tumors.
C1 Hop Lariboisiere, Serv Anat Pathol, F-75475 Paris 10, France.
   CEA Saclay, Serv Biol Cellulaire, F-91191 Gif Sur Yvette, France.
   Hop Cochin, Serv Anat Pathol, F-75679 Paris 14, France.
C3 UDICE-French Research Universities; Universite Paris Cite; Assistance
   Publique Hopitaux Paris (APHP); Hopital Universitaire
   Lariboisiere-Fernand-Widal - APHP; Hopital Universitaire Saint-Louis -
   APHP; CEA; UDICE-French Research Universities; Universite Paris Saclay;
   UDICE-French Research Universities; Universite Paris Cite; Assistance
   Publique Hopitaux Paris (APHP); Hopital Universitaire Cochin - APHP;
   Hopital Universitaire Necker-Enfants Malades - APHP
RP Kafe, H (通讯作者)，Hop Lariboisiere, Serv Anat Pathol, 2 Rue Ambroise Pare, F-75475 Paris 10, France.
EM
RI Verbavatz, Jean-Marc/HTN-2689-2023
OI Verbavatz, Jean-Marc/0000-0002-4654-1174
CR ECELBARGER CA, 1995, AM J PHYSIOL-RENAL, V269, pF663, DOI 10.1152/ajprenal.1995.269.5.F663
   KENNEDY SM, 1990, HUM PATHOL, V21, P449, DOI 10.1016/0046-8177(90)90209-N
   Kovacs G, 1997, J PATHOL, V183, P131, DOI 10.1002/(SICI)1096-9896(199710)183:2<131::AID-PATH931>3.0.CO;2-G
   Marples D, 1999, AM J PHYSIOL-RENAL, V276, pF331, DOI 10.1152/ajprenal.1999.276.3.F331
   Milowsky MI, 2002, CANCER, V94, P111, DOI 10.1002/cncr.10204
   Peyromaure M, 2003, J UROLOGY, V170, P1138, DOI 10.1097/01.ju.0000086616.40603.ad
   PRESTON GM, 1992, SCIENCE, V256, P385, DOI 10.1126/science.256.5055.385
   Roudier N, 1998, J BIOL CHEM, V273, P8407, DOI 10.1074/jbc.273.14.8407
   RUMPELT HJ, 1991, HISTOPATHOLOGY, V18, P115, DOI 10.1111/j.1365-2559.1991.tb01453.x
   Srigley JR, 1998, SEMIN DIAGN PATHOL, V15, P54
   Terris J, 1996, AM J PHYSIOL-RENAL, V271, pF414, DOI 10.1152/ajprenal.1996.271.2.F414
   ULRICH W, 1985, HISTOPATHOLOGY, V9, P1037
   Verkman AS, 2000, AM J PHYSIOL-RENAL, V278, pF13, DOI 10.1152/ajprenal.2000.278.1.F13
NR 13
TC 33
Z9 34
U1 0
U2 0
PU SPRINGER
PI NEW YORK
PA ONE NEW YORK PLAZA, SUITE 4600, NEW YORK, NY, UNITED STATES
SN 0945-6317
EI 1432-2307
J9 VIRCHOWS ARCH
JI Virchows Arch.
PD DEC
PY 2004
VL 445
IS 6
BP 637
EP 640
DI 10.1007/s00428-004-1124-z
PG 4
WC Pathology
WE Science Citation Index Expanded (SCI-EXPANDED)
SC Pathology
GA 876BM
UT WOS:000225470600013
PM 15480763
DA 2023-08-27
ER

PT J
AU Kawamori, H
   Tai, M
   Sato, M
   Yasugi, T
   Tabata, T
AF Kawamori, Haruhiko
   Tai, Miyako
   Sato, Makoto
   Yasugi, Tetsuo
   Tabata, Tetsuya
TI Fat/Hippo pathway regulates the progress of neural differentiation
   signaling in the Drosophila optic lobe
SO DEVELOPMENT GROWTH & DIFFERENTIATION
LA English
DT Article
DE Drosophila; epidermal growth factor; fat; hippo; neuroepithelium
ID TUMOR-SUPPRESSOR PATHWAY; PLANAR CELL POLARITY; GENE-EXPRESSION; BANTAM
   MICRORNA; TISSUE-GROWTH; WILD-TYPE; FAT; HIPPO; PROLIFERATION; BRAIN
AB A large number of neural and glial cell species differentiate from neuronal precursor cells during nervous system development. Two types of Drosophila optic lobe neurons, lamina and medulla neurons, are derived from the neuroepithelial (NE) cells of the outer optic anlagen. During larval development, epidermal growth factor receptor (EGFR)/Ras signaling sweeps the NE field from the medial edge and drives medulla neuroblast (NB) formation. This signal drives the transient expression of a proneural gene, lethal of scute, and we refer to its signal array as the ``proneural wave,'' as it is the marker of the EGFR /Ras signaling front. In this study, we show that the atypical cadherin Fat and the downstream Hippo pathways regulate the transduction of EGFR /Ras signaling along the NE field and, thus, ensure the progress of NB differentiation. Fat/Hippo pathway mutation also disrupts the pattern formation of the medulla structure, which is associated with the regulation of neurogenesis. A candidate for the Fat ligand, Dachsous is expressed in the posterior optic lobe, and its mutation was observed to cause a similar phenotype as fat mutation, although in a regionally restricted manner. We also show that Dachsous functions as the ligand in this pathway and genetically interacts with Fat in the optic lobe. These findings provide new insights into the function of the Fat/Hippo pathway, which regulates the ordered progression of neurogenesis in the complex nervous system.
C1 [Kawamori, Haruhiko; Tai, Miyako; Sato, Makoto; Yasugi, Tetsuo; Tabata, Tetsuya] Univ Tokyo, Inst Mol & Cellular Biosci, Bunkyo Ku, Tokyo 1130032, Japan.
   [Kawamori, Haruhiko; Tai, Miyako; Sato, Makoto; Yasugi, Tetsuo; Tabata, Tetsuya] Univ Tokyo, Greduate Program Biophys & Biochem, Grad Sch Sci, Bunkyo Ku, Tokyo 1130033, Japan.
C3 University of Tokyo; University of Tokyo
RP Tabata, T (通讯作者)，Univ Tokyo, Inst Mol & Cellular Biosci, Bunkyo Ku, 1-1-1 Yayoi, Tokyo 1130032, Japan.
EM
RI Yasugi, Tetsuo/N-4913-2015
FU Ministry of Education, Science and Culture of Japan; Global COE Program;
   MEXT, Japan; Toray Science and Technology Grant; Yamada Science
   foundation; Grants-in-Aid for Scientific Research [22700333] Funding
   Source: KAKEN
FX We thank K. Irvine for exchanging data prior to the submission and for
   fly strains. We also thank members of Tabata laboratory for discussion,
   S. Kato for the use of the confocal microscope and Y. Maeyama for her
   excellent technical help. We are grateful to A. Carmena for kindly
   providing us with the antibody against L(1) sc and fly strains. We are
   also grateful, H. J. Bellen, S. Blair, F. Garoia, S. Hayashi, J. Skeath,
   the Bloomington Stock Center, and Developmental Studies Hybridoma Bank
   for fly strains and antibodies. This work was supported by Grants-in-Aid
   from the Ministry of Education, Science and Culture of Japan, Global COE
   Program (Integrative Life Science Based on the Study of Biosignaling
   Mechanisms), MEXT, Japan, the Toray Science and Technology Grant and
   Yamada Science foundation to T. T. Support for H. K. was provided by a
   GCOE program from the Ministry of Education, Science and Culture of
   Japan.
CR Baena-Lopez LA, 2008, P NATL ACAD SCI USA, V105, P9645, DOI 10.1073/pnas.0803747105
   Bennett FC, 2006, CURR BIOL, V16, P2101, DOI 10.1016/j.cub.2006.09.045
   Brodsky MH, 1996, DEV BIOL, V173, P428, DOI 10.1006/dbio.1996.0038
   BRYANT PJ, 1988, DEV BIOL, V129, P541, DOI 10.1016/0012-1606(88)90399-5
   Cho E, 2006, NAT GENET, V38, P1142, DOI 10.1038/ng1887
   CLARK HF, 1995, GENE DEV, V9, P1530, DOI 10.1101/gad.9.12.1530
   Doe CQ, 2008, DEVELOPMENT, V135, P1575, DOI 10.1242/dev.014977
   Egger B, 2007, NEURAL DEV, V2, DOI 10.1186/1749-8104-2-1
   Egger B, 2010, DEVELOPMENT, V137, P2981, DOI 10.1242/dev.051250
   Emoto K, 2006, NATURE, V443, P210, DOI 10.1038/nature05090
   Fanto M, 2003, DEVELOPMENT, V130, P763, DOI 10.1242/dev.00304
   FISCHBACH KF, 1989, CELL TISSUE RES, V258, P441, DOI 10.1007/BF00218858
   Fung S, 2008, J COMP NEUROL, V506, P469, DOI 10.1002/cne.21539
   Gotz M, 2005, NAT REV MOL CELL BIO, V6, P777, DOI 10.1038/nrm1739
   Guichard A, 2000, DEV BIOL, V226, P255, DOI 10.1006/dbio.2000.9851
   Hamaratoglu F, 2009, J CELL SCI, V122, P2351, DOI 10.1242/jcs.046482
   Huang JB, 2005, CELL, V122, P421, DOI 10.1016/j.cell.2005.06.007
   Huang Z, 1996, CELL, V86, P411, DOI 10.1016/S0092-8674(00)80114-2
   Ishikawa HO, 2008, SCIENCE, V321, P401, DOI 10.1126/science.1158159
   Ito K, 1997, DEVELOPMENT, V124, P761
   Jiang Q, 2009, BIOCHEM BIOPH RES CO, V384, P114, DOI 10.1016/j.bbrc.2009.04.070
   Karim FD, 1998, DEVELOPMENT, V125, P1
   Lee T, 1999, NEURON, V22, P451, DOI 10.1016/S0896-6273(00)80701-1
   Ma D, 2003, NATURE, V421, P543, DOI 10.1038/nature01366
   MAHONEY PA, 1991, CELL, V67, P853, DOI 10.1016/0092-8674(91)90359-7
   Mao YP, 2006, DEVELOPMENT, V133, P2539, DOI 10.1242/dev.02427
   Matakatsu H, 2004, DEVELOPMENT, V131, P3785, DOI 10.1242/dev.01254
   Matakatsu H, 2006, DEVELOPMENT, V133, P2315, DOI 10.1242/dev.02401
   Mikeladze-Dvali T, 2005, CELL, V122, P775, DOI 10.1016/j.cell.2005.07.026
   Morante J, 2008, CURR BIOL, V18, P553, DOI 10.1016/j.cub.2008.02.075
   Nagaso H, 2001, J HISTOCHEM CYTOCHEM, V49, P1177, DOI 10.1177/002215540104900911
   Nassif C, 2003, J COMP NEUROL, V455, P417, DOI 10.1002/cne.10482
   Nolo R, 2006, CURR BIOL, V16, P1895, DOI 10.1016/j.cub.2006.08.057
   Oh H, 2008, DEVELOPMENT, V135, P1081, DOI 10.1242/dev.015255
   Otsuna H, 2006, J COMP NEUROL, V497, P928, DOI 10.1002/cne.21015
   Reddy BVVG, 2008, DEVELOPMENT, V135, P2827, DOI 10.1242/dev.020974
   Reddy BVVG, 2010, DEVELOPMENT, V137, P2397, DOI 10.1242/dev.050013
   Rogulja D, 2008, DEV CELL, V15, P309, DOI 10.1016/j.devcel.2008.06.003
   Silva E, 2006, CURR BIOL, V16, P2081, DOI 10.1016/j.cub.2006.09.004
   Simon MA, 2004, DEVELOPMENT, V131, P6175, DOI 10.1242/dev.01550
   Strutt H, 2005, BIOESSAYS, V27, P1218, DOI 10.1002/bies.20318
   Strutt H, 2004, DEVELOPMENT, V131, P881, DOI 10.1242/dev.00996
   Sturtevant MA, 1996, DEV BIOL, V174, P298, DOI 10.1006/dbio.1996.0075
   Takei Y, 2004, DEVELOPMENT, V131, P73, DOI 10.1242/dev.00913
   Takemura SY, 2008, J COMP NEUROL, V509, P493, DOI 10.1002/cne.21757
   Thompson BJ, 2006, CELL, V126, P767, DOI 10.1016/j.cell.2006.07.013
   Toriya M, 2006, DEV NEUROSCI-BASEL, V28, P142, DOI 10.1159/000090760
   VILLANO JL, 1995, DEVELOPMENT, V121, P2767
   Willecke M, 2006, CURR BIOL, V16, P2090, DOI 10.1016/j.cub.2006.09.005
   Willecke M, 2008, P NATL ACAD SCI USA, V105, P14897, DOI 10.1073/pnas.0805201105
   XU TA, 1995, DEVELOPMENT, V121, P1053
   Yang CH, 2002, CELL, V108, P675, DOI 10.1016/S0092-8674(02)00658-X
   Yasugi T, 2008, DEVELOPMENT, V135, P1471, DOI 10.1242/dev.019117
   Yasugi T, 2010, DEVELOPMENT, V137, P3193, DOI 10.1242/dev.048058
   ZAK NB, 1992, DEV BIOL, V149, P448, DOI 10.1016/0012-1606(92)90299-V
NR 55
TC 32
Z9 33
U1 0
U2 7
PU WILEY-BLACKWELL
PI MALDEN
PA COMMERCE PLACE, 350 MAIN ST, MALDEN 02148, MA USA
SN 0012-1592
J9 DEV GROWTH DIFFER
JI Dev. Growth Diff.
PD JUN
PY 2011
VL 53
IS 5
BP 653
EP 667
DI 10.1111/j.1440-169X.2011.01279.x
PG 15
WC Cell Biology; Developmental Biology
WE Science Citation Index Expanded (SCI-EXPANDED)
SC Cell Biology; Developmental Biology
GA 786JU
UT WOS:000292304300003
PM 21671914
OA Bronze
DA 2023-08-27
ER

PT J
AU Kosty, JA
   Mejia-Munne, J
   Dossani, R
   Savardekar, A
   Guthikonda, B
AF Kosty, Jennifer A.
   Mejia-Munne, Juan
   Dossani, Rimal
   Savardekar, Amey
   Guthikonda, Bharat
TI Jacques Jean Lhermitte and the syndrome of peduncular hallucinosis
SO NEUROSURGICAL FOCUS
LA English
DT Article
DE Jacques Jean Lhermitte; peduncular hallucinosis; complex visual
   hallucinations; peduncular hallucination
ID BRAIN-STEM COMPRESSION; MICROVASCULAR DECOMPRESSION; HALLUCINATIONS
   SECONDARY; TRIGEMINAL NEURALGIA; INFARCTION; COMPLEX
AB Jacques Jean Lhermitte (1877-1959) was among the most accomplished neurologists of the 20th century. In addition to working as a clinician and instructor, he authored more than 800 papers and 16 books on neurology, neuropathology, psychiatry, and mystical phenomena. In addition to the well-known "Lhermitte's sign," an electrical shock-like sensation caused by spinal cord irritation in demyelinating disease, Lhermitte was a pioneer in the study of the relationship between the physical substance of the brain and the experience of the mind. A fascinating example of this is the syndrome of peduncular hallucinosis, characterized by vivid visual hallucinations occurring in fully lucid patients. This syndrome, which was initially described as the result of a midbrain insult, also may occur with injury to the thalamus or pons. It has been reported as a presenting symptom of various tumors and as a complication of neurosurgical procedures. Here, the authors review the life of Lhermitte and provide a historical review of the syndrome of peduncular hallucinosis.
C1 [Kosty, Jennifer A.; Dossani, Rimal; Savardekar, Amey; Guthikonda, Bharat] Louisiana State Univ, Hlth Sci Ctr Shreveport, Dept Neurosurg, Shreveport, LA 71105 USA.
   [Mejia-Munne, Juan] Univ Cincinnati, Med Ctr, Dept Neurosurg, Cincinnati, OH 45267 USA.
C3 Louisiana State University System; Louisiana State University Health
   Sciences Center at Shreveport; University System of Ohio; University of
   Cincinnati
RP Kosty, JA (通讯作者)，LSU, Hlth Sci Ctr, Shreveport, LA 71105 USA.
EM
RI Savardekar, Amey/AAV-6000-2021
OI Savardekar, Amey/0000-0002-9305-7457
CR Alajouanine T, 1944, REV NEUROL-FRANCE, V76, P90
   Biswas SN, 2016, INDIAN J PEDIATR, V83, P736, DOI 10.1007/s12098-015-1935-8
   Boller F, 2005, CORTEX, V41, P740, DOI 10.1016/S0010-9452(08)70292-2
   CHEN HJ, 1995, J FORMOS MED ASSOC, V94, P503
   Chu DT, 2018, MULT SCLER, V20
   Couse M, 2018, MENT ILLN, V10, P14, DOI 10.4081/mi.2018.7586
   Critchley M, 1959, BMJ, V1, P652
   de Morsier G, 1935, REV NEUROL-FRANCE, V64, P606
   de Raykeer RP, 2016, J CLIN PSYCHOPHARM, V36, P285, DOI 10.1097/JCP.0000000000000497
   deMorsier G, 1938, REV OTO NEUROOPHTALM, V16, P244
   DUNN DW, 1983, NEUROLOGY, V33, P1360, DOI 10.1212/WNL.33.10.1360
   FEINBERG WM, 1989, NEUROLOGY, V39, P1535, DOI 10.1212/WNL.39.11.1535
   Garrabe J, 2013, ANN MED-PSYCHOL, V171, P54, DOI 10.1016/j.amp.2013.01.012
   GELLER TJ, 1987, ANN NEUROL, V21, P602, DOI 10.1002/ana.410210614
   Gokce M, 2003, ACTA NEUROPSYCHIATR, V15, P105, DOI 10.1034/j.1601-5215.2003.00019.x
   Grzybowski A, 2019, J NEUROL, V266, P2090, DOI 10.1007/s00415-018-9101-7
   Koerbel A, 2007, ACTA NEUROCHIR, V149, P831, DOI 10.1007/s00701-007-1181-6
   Kumar R, 1999, BRIT J NEUROSURG, V13, P500, DOI 10.1080/02688699943349
   Kumar R, 2000, NEUROL INDIA, V48, P183
   Lhermitte J, 1932, REV NEUROL-FRANCE, V57, P382
   Lhermitte J, 1929, ARCH NEURO PSYCHIATR, V22, P5, DOI 10.1001/archneurpsyc.1929.02220010008002
   Lhermitte J, 1926, BRAIN, V49, P157, DOI 10.1093/brain/49.2.157
   Lhermitte J., 1920, B ASSOC FR ETUD CANC, V9, P99
   Lhermitte J., 1922, REVNEUROL PARIS, V38, P1359
   Lhermitte JJ., 1924, REV NEUROL, V2, P56
   Maiuri F, 2002, CLIN NEUROL NEUROSUR, V104, P41, DOI 10.1016/S0303-8467(01)00184-6
   Manford M, 1998, BRAIN, V121, P1819, DOI 10.1093/brain/121.10.1819
   Marie P, 1914, ANN MED, V1, P18
   Miyazawa T, 2001, J NEUROSURG, V95, P500, DOI 10.3171/jns.2001.95.3.0500
   Miyazawa T, 2009, ACTA NEUROCHIR, V151, P285, DOI 10.1007/s00701-009-0211-y
   NADVI SS, 1994, SURG NEUROL, V41, P250, DOI 10.1016/0090-3019(94)90132-5
   Nadvi SS, 1998, BRIT J NEUROSURG, V12, P579, DOI 10.1080/02688699844484
   Nakamura M, 2005, NEUROSURGERY, V57, P77, DOI 10.1227/01.NEU.0000154699.29796.34
   Notas K, 2015, HIPPOKRATIA, V19, P268
   O'Neill SB, 2005, BRIT J NEUROSURG, V19, P359, DOI 10.1080/02688690500305407
   Parisis D, 2003, EUR NEUROL, V50, P107, DOI 10.1159/000072509
   Qasho R, 1998, J Neurosurg Sci, V42, P47
   ROZANSKI J, 1952, NEUROLOGY, V2, P341, DOI 10.1212/WNL.2.7-8.341
   TSUKAMOTO H, 1993, SURG NEUROL, V40, P31, DOI 10.1016/0090-3019(93)90166-X
   Van Bogaert L, 1927, REV NEUROL-FRANCE, V47, P608
   Yano Koh, 1994, Neurologia Medico-Chirurgica, V34, P593, DOI 10.2176/nmc.34.593
NR 41
TC 5
Z9 5
U1 0
U2 0
PU AMER ASSOC NEUROLOGICAL SURGEONS
PI ROLLING MEADOWS
PA 5550 MEADOWBROOK DRIVE, ROLLING MEADOWS, IL 60008 USA
SN 1092-0684
J9 NEUROSURG FOCUS
JI Neurosurg. Focus
PD SEP
PY 2019
VL 47
IS 3
AR E9
DI 10.3171/2019.6.FOCUS19342
PG 6
WC Clinical Neurology; Surgery
WE Science Citation Index Expanded (SCI-EXPANDED)
SC Neurosciences & Neurology; Surgery
GA IV1RM
UT WOS:000484055700009
PM 31473682
OA gold
DA 2023-08-27
ER

PT J
AU Wang, YK
   Zhu, WW
   Wu, MH
   Wu, YH
   Liu, ZX
   Liang, LM
   Sheng, C
   Hao, J
   Wang, L
   Li, W
   Zhou, Q
   Hu, BY
AF Wang, Yu-Kai
   Zhu, Wan-Wan
   Wu, Meng-Hua
   Wu, Yi-Hui
   Liu, Zheng-Xin
   Liang, Ling-Min
   Sheng, Chao
   Hao, Jie
   Wang, Liu
   Li, Wei
   Zhou, Qi
   Hu, Bao-Yang
TI Human Clinical-Grade Parthenogenetic ESC-Derived Dopaminergic Neurons
   Recover Locomotive Defects of Nonhuman Primate Models of Parkinson's
   Disease
SO STEM CELL REPORTS
LA English
DT Article
ID EMBRYONIC STEM-CELLS; NEURAL CELLS; TRANSPLANTATION; BRAIN;
   DIFFERENTIATION; MONKEYS; DERIVATION; EXPRESSION; PRINCIPLES; DEFICITS
AB Clinical application of stem cell derivatives requires clinical-grade cells and sufficient preclinical proof of safety and efficacy, preferably in primates. We previously successfully established a clinical-grade human parthenogenetic embryonic stem cell (hPESC) line, but the suitability of its subtype-specific progenies for therapy is not clear. Here, we compared the function of clinical-grade hPESC-derived midbrain dopaminergic (DA) neurons in two canonical protocols in a primate Parkinson's disease (PD) model. We found that the grafts did not form tumors and produced variable but apparent behavioral improvement for at least 24 months in most monkeys in both groups. In addition, a slight DA increase in the striatum correlates with significant functional improvement. These results demonstrated that clinical-grade hPESCs can serve as a reliable source of cells for PD treatment. Our proof-of-concept findings provide preclinical data for China's first ESC-based phase I/IIa clinical study of PD
C1 [Wang, Yu-Kai; Zhu, Wan-Wan; Wu, Meng-Hua; Wu, Yi-Hui; Liu, Zheng-Xin; Liang, Ling-Min; Sheng, Chao; Hao, Jie; Wang, Liu; Li, Wei; Zhou, Qi; Hu, Bao-Yang] Chinese Acad Sci, Inst Zool, State Key Lab Stem Cell & Reprod Biol, Beijing 100101, Peoples R China.
   [Wang, Yu-Kai; Wu, Yi-Hui; Hao, Jie; Wang, Liu; Li, Wei; Zhou, Qi; Hu, Bao-Yang] Chinese Acad Sci, Inst Stem Cell & Regenerat, Beijing 100101, Peoples R China.
   [Wang, Yu-Kai; Liang, Ling-Min; Hao, Jie; Wang, Liu; Li, Wei; Zhou, Qi; Hu, Bao-Yang] Chinese Acad Sci, Beijing Stem Cell Bank, Beijing 100190, Peoples R China.
   [Wu, Meng-Hua; Liang, Ling-Min; Wang, Liu; Li, Wei; Zhou, Qi; Hu, Bao-Yang] Univ Chinese Acad Sci, Beijing 100049, Peoples R China.
C3 Chinese Academy of Sciences; Institute of Zoology, CAS; Chinese Academy
   of Sciences; Chinese Academy of Sciences; Chinese Academy of Sciences;
   University of Chinese Academy of Sciences, CAS
RP Zhou, Q; Hu, BY (通讯作者)，Chinese Acad Sci, Inst Zool, State Key Lab Stem Cell & Reprod Biol, Beijing 100101, Peoples R China.; Zhou, Q; Hu, BY (通讯作者)，Chinese Acad Sci, Inst Stem Cell & Regenerat, Beijing 100101, Peoples R China.; Zhou, Q; Hu, BY (通讯作者)，Chinese Acad Sci, Beijing Stem Cell Bank, Beijing 100190, Peoples R China.; Zhou, Q; Hu, BY (通讯作者)，Univ Chinese Acad Sci, Beijing 100049, Peoples R China.
EM
RI Hu, Baoyang/R-8374-2016
OI Hu, Baoyang/0000-0003-3993-4014
FU Program of National Key Research and Development [2016YFA0101402,
   2017YFA0105002]; National Basic Research Program of China [2014CB964604,
   2012CBA01300]; Chinese Academy of Sciences [ZDRW-ZS-2017-5,
   QYZDY-SSW-SMC002]; National Natural Science Foundation of China
   [31621004, 31571533]; Beijing Science and Technology Major Project
   [Z181100001818002]
FX We thank S.-W. Li, X.-L. Zhu, and Q. Meng for their technical support in
   confocal laser-scanning microscopy and fluorescence-activated cell
   sorting. This work was supported by grants from the Program of National
   Key Research and Development (2016YFA0101402 and 2017YFA0105002); the
   National Basic Research Program of China (2014CB964604 and
   2012CBA01300); the Key Deployment Projects of the Chinese Academy of
   Sciences (ZDRW-ZS-2017-5); the National Natural Science Foundation of
   China (31621004 and 31571533); the Key Research Projects of the Frontier
   Science of the Chinese Academy of Sciences (QYZDY-SSW-SMC002); and the
   Beijing Science and Technology Major Project (Z181100001818002).
CR Brownell AL, 1998, NAT MED, V4, P1308, DOI 10.1038/3300
   Ding F, 2008, EXP NEUROL, V212, P431, DOI 10.1016/j.expneurol.2008.04.027
   Doi D, 2012, STEM CELLS, V30, P935, DOI 10.1002/stem.1060
   Emborg ME, 2008, CELL TRANSPLANT, V17, P383
   Emborg ME, 2013, CELL TRANSPLANT, V22, P831, DOI 10.3727/096368912X647144
   Emborg ME, 2013, CELL REP, V3, P646, DOI 10.1016/j.celrep.2013.02.016
   Espejel S, 2014, STEM CELLS, V32, P1983, DOI 10.1002/stem.1726
   Fink DW, 2009, SCIENCE, V324, P1662, DOI 10.1126/science.1173712
   Freed CR, 2001, NEW ENGL J MED, V345, P147
   Ghosh A, 2010, ANN NEUROSCI, V17, P191, DOI 10.5214/ans.0972.7531.1017410
   Gonzalez R, 2016, CELL TRANSPLANT, V25, P1945, DOI 10.3727/096368916X691682
   Gonzalez R, 2015, CELL TRANSPLANT, V24, P681, DOI 10.3727/096368915X687769
   Grealish S, 2014, CELL STEM CELL, V15, P653, DOI 10.1016/j.stem.2014.09.017
   Gu Q, 2017, STEM CELL REP, V9, P366, DOI 10.1016/j.stemcr.2017.04.017
   Hallett PJ, 2015, CELL STEM CELL, V16, P269, DOI 10.1016/j.stem.2015.01.018
   Hao J, 2009, SCI CHINA SER C, V52, P599, DOI 10.1007/s11427-009-0096-2
   Hu BY, 2010, P NATL ACAD SCI USA, V107, P4335, DOI 10.1073/pnas.0910012107
   Hyun I, 2008, CELL STEM CELL, V3, P607, DOI 10.1016/j.stem.2008.11.009
   Kettenmann H, 2006, NAT NEUROSCI, V9, P1463, DOI 10.1038/nn1206-1463
   Kikuchi T, 2017, NATURE, V548, P592, DOI 10.1038/nature23664
   Kim SH, 2009, KOREAN J RADIOL, V10, P112, DOI 10.3348/kjr.2009.10.2.112
   Kriks S, 2011, NATURE, V480, P547, DOI 10.1038/nature10648
   LINDVALL O, 1990, SCIENCE, V247, P574, DOI 10.1126/science.2105529
   Liu Y, 2013, NAT BIOTECHNOL, V31, P440, DOI 10.1038/nbt.2565
   Ma LX, 2012, CELL STEM CELL, V10, P455, DOI 10.1016/j.stem.2012.01.021
   Ma LX, 2011, METHODS MOL BIOL, V767, P411, DOI 10.1007/978-1-61779-201-4_30
   Mai QY, 2007, CELL RES, V17, P1008, DOI 10.1038/cr.2007.102
   Morizane A, 2013, STEM CELL REP, V1, P283, DOI 10.1016/j.stemcr.2013.08.007
   Morizane A, 2010, J NEUROSCI RES, V88, P3467, DOI 10.1002/jnr.22515
   Olanow CW, 2003, ANN NEUROL, V54, P403, DOI 10.1002/ana.10720
   Pankratz MT, 2007, STEM CELLS, V25, P1511, DOI 10.1634/stemcells.2006-0707
   Ramos-Moreno T, 2012, BEHAV BRAIN RES, V232, P225, DOI 10.1016/j.bbr.2012.04.020
   Roggendorf W, 1996, ACTA NEUROPATHOL, V92, P288, DOI 10.1007/s004010050520
   Sanchez-Pernaute R, 2005, STEM CELLS, V23, P914, DOI 10.1634/stemcells.2004-0172
   Studer L, 1996, BRAIN RES BULL, V41, P143, DOI 10.1016/S0361-9230(96)00114-1
   Turovets N, 2011, METHODS MOL BIOL, V767, P37, DOI 10.1007/978-1-61779-201-4_4
   Vogel DYS, 2013, J NEUROINFLAMM, V10, DOI 10.1186/1742-2094-10-35
   Wilkerson A, 2013, CELL STEM CELL, V12, P147, DOI 10.1016/j.stem.2013.01.002
   Xi JJ, 2012, STEM CELLS, V30, P1655, DOI 10.1002/stem.1152
   Yoo J, 2013, J CELL BIOCHEM, V114, P743, DOI 10.1002/jcb.24427
NR 40
TC 59
Z9 61
U1 5
U2 35
PU CELL PRESS
PI CAMBRIDGE
PA 50 HAMPSHIRE ST, FLOOR 5, CAMBRIDGE, MA 02139 USA
SN 2213-6711
J9 STEM CELL REP
JI Stem Cell Rep.
PD JUL 10
PY 2018
VL 11
IS 1
BP 171
EP 182
DI 10.1016/j.stemcr.2018.05.010
PG 12
WC Cell & Tissue Engineering; Cell Biology
WE Science Citation Index Expanded (SCI-EXPANDED)
SC Cell Biology
GA GM4EJ
UT WOS:000438067600015
PM 29910127
OA gold, Green Published
DA 2023-08-27
ER

PT J
AU Hammer, GD
   Parker, KL
   Schimmer, BP
AF Hammer, GD
   Parker, KL
   Schimmer, BP
TI Minireview: Transcriptional regulation of adrenocortical development
SO ENDOCRINOLOGY
LA English
DT Review
ID STEROIDOGENIC FACTOR-I; ADRENAL HYPOPLASIA CONGENITA; NUCLEAR RECEPTOR
   SF-1; XY SEX REVERSAL; EXPRESSION PROFILES; DAX1 EXPRESSION; CRITICAL
   REGION; GENE; FACTOR-1; WT1
AB The adrenal glands are comprised of two distinct endocrine organs: the outer cortex, which is derived from mesoderm and synthesizes steroid hormones, and the inner medulla, which contains neuroectodermal cells derived from the neural crest and produces the catecholamine hormones norepinephrine and epinephrine. The developmental program that gives rise to the adrenal gland begins early during embryogenesis and continues throughout gestation and well after birth. In this article, we review the molecular mechanisms of adrenal differentiation and development, focusing on the contributions of genes responsible for the development of the adrenal cortex as identified from studies of experimental animal models and human subjects with clinical diseases. These studies identify a hierarchical network of transcription factors, including Wilms' tumor-1, steroidogenic factor-1, dosage-sensitive sex reversal, adrenal hypoplasia congenita, X-linked-1, PBX1, and CITED2, that both give rise to the adrenal cortex and subsequently determine its subsequent function in steroidogenesis.
C1 Univ Michigan, Dept Internal Med, Div Metab Endocrinol & Metab, Ann Arbor, MI 48109 USA.
   Univ Michigan, Dept Mol & Integrat Physiol, Div Metab Endocrinol & Metab, Ann Arbor, MI 48109 USA.
   Univ Texas, SW Med Ctr, Dept Internal Med, Dallas, TX 75390 USA.
   Univ Texas, SW Med Ctr, Dept Pharmacol, Dallas, TX 75390 USA.
   Univ Toronto, Banting & Best Dept Med Res, Toronto, ON M5G 1L6, Canada.
C3 University of Michigan System; University of Michigan; University of
   Michigan System; University of Michigan; University of Texas System;
   University of Texas Dallas; University of Texas Southwestern Medical
   Center Dallas; University of Texas System; University of Texas Dallas;
   University of Texas Southwestern Medical Center Dallas; University of
   Toronto
RP Hammer, GD (通讯作者)，Univ Michigan, Dept Internal Med, Div Metab Endocrinol & Metab, 1150 W Med Ctr Dr, Ann Arbor, MI 48109 USA.
EM
CR Achermann JC, 2001, MOL CELL ENDOCRINOL, V185, P17, DOI 10.1016/S0303-7207(01)00619-0
   Achermann JC, 1999, NAT GENET, V22, P125, DOI 10.1038/9629
   Achermann JC, 2002, J CLIN ENDOCR METAB, V87, P1829, DOI 10.1210/jc.87.4.1829
   ANDERSON DJ, 2003, ANNU REV NEUROSCI, V16, P129
   Babu PS, 2002, ENDOCRINOLOGY, V143, P665, DOI 10.1210/en.143.2.665
   Bamforth SD, 2001, NAT GENET, V29, P469, DOI 10.1038/ng768
   BEAMER WG, 1994, J ENDOCRINOL, V141, P33, DOI 10.1677/joe.0.1410033
   Beuschlein F, 2003, MOL CELL BIOL, V23, P3951, DOI 10.1128/MCB.23.11.3951-3964.2003
   Beuschlein F, 2002, ENDOCRINOLOGY, V143, P3122, DOI 10.1210/en.143.8.3122
   Biason-Lauber A, 2000, AM J HUM GENET, V67, P1563, DOI 10.1086/316893
   Bischof LJ, 1998, J BIOL CHEM, V273, P7941, DOI 10.1074/jbc.273.14.7941
   Bland ML, 2004, MOL ENDOCRINOL, V18, P941, DOI 10.1210/me.2003-0333
   Bland ML, 2000, P NATL ACAD SCI USA, V97, P14488, DOI 10.1073/pnas.97.26.14488
   CALL KM, 1990, CELL, V60, P509, DOI 10.1016/0092-8674(90)90601-A
   Clark AJL, 1998, ENDOCR REV, V19, P828, DOI 10.1210/er.19.6.828
   Coll AP, 2004, ENDOCRINOLOGY, V145, P4721, DOI 10.1210/en.2004-0491
   Correa RV, 2004, J CLIN ENDOCR METAB, V89, P1767, DOI 10.1210/jc.2003-031240
   Crawford PA, 1998, MOL CELL BIOL, V18, P2949, DOI 10.1128/MCB.18.5.2949
   GESSLER M, 1990, NATURE, V343, P774, DOI 10.1038/343774a0
   Gummow BM, 2003, J BIOL CHEM, V278, P26572, DOI 10.1074/jbc.M212677200
   HABER DA, 1990, CELL, V61, P1257, DOI 10.1016/0092-8674(90)90690-G
   Hammer GD, 1999, MOL CELL, V3, P521, DOI 10.1016/S1097-2765(00)80480-3
   Hammes A, 2001, CELL, V106, P319, DOI 10.1016/S0092-8674(01)00453-6
   Hanley NA, 2001, MOL ENDOCRINOL, V15, P57, DOI 10.1210/me.15.1.57
   Hatano O, 1996, GENES CELLS, V1, P663, DOI 10.1046/j.1365-2443.1996.00254.x
   Heikkila M, 2002, ENDOCRINOLOGY, V143, P4358, DOI 10.1210/en.2002-220275
   Hossain A, 2004, MOL ENDOCRINOL, V18, P1428, DOI 10.1210/me.2003-0176
   Hossain A, 2003, J BIOL CHEM, V278, P26511, DOI 10.1074/jbc.M300804200
   Hoyle C, 2002, MOL ENDOCRINOL, V16, P747, DOI 10.1210/me.16.4.747
   IKEDA Y, 1995, MOL ENDOCRINOL, V9, P478, DOI 10.1210/me.9.4.478
   Ikeda Y, 1996, MOL ENDOCRINOL, V10, P1261, DOI 10.1210/me.10.10.1261
   INGRAHAM HA, 1994, GENE DEV, V8, P2302, DOI 10.1101/gad.8.19.2302
   Ito M, 1997, MOL CELL BIOL, V17, P1476, DOI 10.1128/MCB.17.3.1476
   Iyer AK, 2004, MOL GENET METAB, V83, P60, DOI 10.1016/j.ymgme.2004.07.018
   Jeays-Ward K, 2003, DEVELOPMENT, V130, P3663, DOI 10.1242/dev.00591
   Kawabe K, 1999, MOL ENDOCRINOL, V13, P1267, DOI 10.1210/me.13.8.1267
   Keegan CE, 2002, TRENDS ENDOCRIN MET, V13, P200, DOI 10.1016/S1043-2760(02)00602-1
   Kim J, 1999, MOL CELL BIOL, V19, P2289
   KREIDBERG JA, 1993, CELL, V74, P679, DOI 10.1016/0092-8674(93)90515-R
   Lalli E, 2000, MOL CELL BIOL, V20, P4910, DOI 10.1128/MCB.20.13.4910-4921.2000
   Little M, 1997, HUM MUTAT, V9, P209, DOI 10.1002/(SICI)1098-1004(1997)9:3<209::AID-HUMU2>3.0.CO;2-2
   LUO XR, 1994, CELL, V77, P481, DOI 10.1016/0092-8674(94)90211-9
   Mallet D, 2004, J CLIN ENDOCR METAB, V89, P4829, DOI 10.1210/jc.2004-0670
   Meeks JJ, 2003, NAT GENET, V34, P32, DOI 10.1038/ng1141
   Meeks JJ, 2003, DEVELOPMENT, V130, P1029, DOI 10.1242/dev.00316
   Mesiano S, 1997, ENDOCR REV, V18, P378, DOI 10.1210/er.18.3.378
   MILLER WL, 1988, ENDOCR REV, V9, P295, DOI 10.1210/edrv-9-3-295
   Mitani F, 2003, BBA-GEN SUBJECTS, V1619, P317, DOI 10.1016/S0304-4165(02)00490-7
   Mizusaki H, 2003, MOL ENDOCRINOL, V17, P507, DOI 10.1210/me.2002-0362
   Moore AW, 1999, DEVELOPMENT, V126, P1845
   Morley SD, 1996, MOL ENDOCRINOL, V10, P585, DOI 10.1210/me.10.5.585
   Morohashi K, 1999, TRENDS ENDOCRIN MET, V10, P169, DOI 10.1016/S1043-2760(98)00142-8
   Morohashi K, 1999, BLOOD, V93, P1586
   MUSCATELLI F, 1994, NATURE, V372, P672, DOI 10.1038/372672a0
   Nachtigal MW, 1998, CELL, V93, P445, DOI 10.1016/S0092-8674(00)81172-1
   Parker KL, 2002, RECENT PROG HORM RES, V57, P19, DOI 10.1210/rp.57.1.19
   Phelan JK, 2001, HUM MUTAT, V18, P472, DOI 10.1002/humu.1225
   SADOVSKY Y, 1995, P NATL ACAD SCI USA, V92, P10939, DOI 10.1073/pnas.92.24.10939
   Schnabel CA, 2003, GENESIS, V37, P123, DOI 10.1002/gene.10235
   SHINODA K, 1995, DEV DYNAM, V204, P22, DOI 10.1002/aja.1002040104
   Song KH, 2004, MOL ENDOCRINOL, V18, P1929, DOI 10.1210/me.2004-0043
   Swain A, 1996, NAT GENET, V12, P404, DOI 10.1038/ng0496-404
   Tolwinski NS, 2004, TRENDS GENET, V20, P177, DOI 10.1016/j.tig.2004.02.003
   Vainio S, 1999, NATURE, V397, P405, DOI 10.1038/17068
   Vilain E, 1999, J CLIN ENDOCR METAB, V84, P4335, DOI 10.1210/jc.84.12.4335
   Wagner KD, 2003, J CELL SCI, V116, P1653, DOI 10.1242/jcs.00405
   Wilhelm D, 2002, GENE DEV, V16, P1839, DOI 10.1101/gad.220102
   Yu RN, 1998, MOL ENDOCRINOL, V12, P1010, DOI 10.1210/me.12.7.1010
   Yu RN, 1998, NAT GENET, V20, P353, DOI 10.1038/3822
   ZANARIA E, 1994, NATURE, V372, P635, DOI 10.1038/372635a0
   Zazopoulos E, 1997, NATURE, V390, P311, DOI 10.1038/36899
NR 71
TC 84
Z9 88
U1 0
U2 6
PU ENDOCRINE SOC
PI WASHINGTON
PA 2055 L ST NW, SUITE 600, WASHINGTON, DC 20036 USA
SN 0013-7227
EI 1945-7170
J9 ENDOCRINOLOGY
JI Endocrinology
PD MAR
PY 2005
VL 146
IS 3
BP 1018
EP 1024
DI 10.1210/en.2004-1385
PG 7
WC Endocrinology & Metabolism
WE Science Citation Index Expanded (SCI-EXPANDED)
SC Endocrinology & Metabolism
GA 897WC
UT WOS:000227035400010
PM 15604207
OA Bronze
DA 2023-08-27
ER

PT J
AU Kakigi, A
   Nakatani, H
   Takeda, T
AF Kakigi, Akinobu
   Nakatani, Hiroaki
   Takeda, Taizo
TI Electrocochleographic and Pure-Tone Audiometric Findings in
   Contralateral Ear of Unilateral Acoustic Neurinoma
SO ORL-JOURNAL FOR OTO-RHINO-LARYNGOLOGY AND ITS RELATED SPECIALTIES
LA English
DT Article
DE Electrocochleography; Pure-tone audiometry; Acoustic neurinoma;
   Contralateral hearing; Delayed endolymphatic hydrops; Olivocochlear
   efferent nerve
ID DELAYED ENDOLYMPHATIC HYDROPS; HEARING PRESERVATION; NEUROMA SURGERY;
   RESECTION
AB Objective: Auditory disorders of the contralateral ear in patients with unilateral acoustic neurinoma have previously been reported using the auditory brainstem response. From our survey of the literature, there is no reported result about electrocochleography (ECoG) in the contralateral ear of acoustic neurinoma cases. In the present study, we reported ECoG and pure-tone audiometric results of the contralateral ears in cases of unilateral acoustic neurinoma and the relationship between ECoG findings and tumor size was discussed. Methods: ECoG and pure-tone audiometry were performed for the contralateral ears in 27 cases of unilateral acoustic neurinoma, and the relationships between tumor size and contralateral ECoG findings and between pure-tone threshold and ECoG findings were investigated. As for ECoG findings, the negative summating potential to the compound action potential ratio (-SP/AP ratio) was examined. The tumor size was assessed by magnetic resonance imaging. Results: Abnormal -SP/AP ratio exceeding 0.40 of the upper limit of the normal range was found in 7 (25.9%) of 27 ears. Incidences of abnormal -SP/AP ratio were 30.0% in large tumors (n = 10), 33.3% in mid-size tumors (n = 6) and 18.2% in small tumors (n = 11). There was no correlation between the incidences of abnormal -SP/AP ratio and tumor size. Moreover, there was no correlation between the -SP/AP ratio and pure-tone threshold at the ECoG recording. Conclusion:These results suggest that any size of acoustic neurinoma may affect contralateral electrocochleographic findings. One possible cause may be dysfunction of the olivocochlear efferent system and another may be endolymphatic hydrops (e.g. delayed endolymphatic hydrops). Currently, however, the causes of -SP/AP ratio increase are still unknown and further investigations are required. Copyright (C) 2010 S. Karger AG, Basel
C1 [Kakigi, Akinobu] Univ Tokyo, Dept Otolaryngol, Fac Med, Bunkyo Ku, Tokyo 1138655, Japan.
   [Kakigi, Akinobu; Nakatani, Hiroaki; Takeda, Taizo] Kochi Med Sch, Dept Otolaryngol, Kochi, Japan.
C3 University of Tokyo; Kochi University
RP Kakigi, A (通讯作者)，Univ Tokyo, Dept Otolaryngol, Fac Med, Bunkyo Ku, 7-3-1 Hongo, Tokyo 1138655, Japan.
EM
CR Fahlbusch R, 1998, ACTA NEUROCHIR, V140, P771, DOI 10.1007/s007010050178
   Hecht CS, 1997, LARYNGOSCOPE, V107, P1122, DOI 10.1097/00005537-199708000-00021
   Huygen PLM, 1996, INT J PEDIATR OTORHI, V35, P143, DOI 10.1016/0165-5876(96)83899-7
   Ishikawa K, 1998, ACTA OTO-LARYNGOL, P16
   Kanzaki J, 1998, AM J OTOL, V19, P644
   LELIEVER WC, 1980, J OTOLARYNGOL, V9, P375
   MOFFAT DA, 1989, J LARYNGOL OTOL, V103, P835, DOI 10.1017/S0022215100110242
   NADOL JB, 1975, ANN OTO RHINOL LARYN, V84, P841, DOI 10.1177/000348947508400617
   Samii M, 1997, NEUROSURGERY, V40, P696, DOI 10.1097/00006123-199704000-00007
   Samii M, 1995, Adv Tech Stand Neurosurg, V22, P343
   SCHUKNECHT HF, 1978, ANN OTO RHINOL LARYN, V87, P743, DOI 10.1177/000348947808700601
   SHEA JJ, 1995, AM J OTOL, V16, P235
   Stipkovits EM, 1998, AM J OTOL, V19, P834
   TAKEDA T, 1992, ORL J OTO-RHINO-LARY, V54, P15, DOI 10.1159/000278935
   YLIKOSKI J, 1988, AM J OTOL, V9, P282
NR 15
TC 0
Z9 0
U1 1
U2 1
PU KARGER
PI BASEL
PA ALLSCHWILERSTRASSE 10, CH-4009 BASEL, SWITZERLAND
J9 ORL-J OTO-RHIN-LARYN
JI ORL-J. Oto-Rhino-Laryngol. Relat. Spec.
PY 2009
VL 71
SU 1
BP 78
EP 84
DI 10.1159/000265116
PG 7
WC Otorhinolaryngology
WE Science Citation Index Expanded (SCI-EXPANDED)
SC Otorhinolaryngology
GA 562JA
UT WOS:000275044900013
PM 20185954
DA 2023-08-27
ER

PT J
AU Smets, G
   Lambert, J
   Tijssen, M
   Mai, C
   Decramer, T
   Vandenberghe, W
   Van Loon, J
   Demaerel, P
AF Smets, Gitte
   Lambert, Julie
   Tijssen, Maud
   Mai, Cindy
   Decramer, Thomas
   Vandenberghe, Wim
   Van Loon, Johannes
   Demaerel, Philippe
TI The dentato-rubro-olivary pathway revisited: New MR imaging observations
   regarding hypertrophic olivary degeneration
SO CLINICAL ANATOMY
LA English
DT Article
DE magnetic resonance imaging; Guillain-Mollaret triangle; hypertrophic
   olivary degeneration
ID BRAIN-STEM; CAVERNOUS MALFORMATIONS; PONTINE HEMORRHAGE; INFERIOR OLIVE;
   CEREBELLAR; RESECTION; HEMANGIOMA; TREMOR; TUMOR
AB Hypertrophic olivary degeneration (HOD) following a lesion of the dentato-rubro-olivary pathway (DROP) is a well-known imaging finding and has extensively been described in the recent literature. We reviewed our patients with HOD as a result of a lesion of the DROP in order to analyze the disruption of the DROP and the resulting HOD in comparison with the literature. We observed unusual imaging findings in four patients. In two patients it concerned new observations related to the timing and imaging appearances of HOD. HOD became only visible 6 years after a lesion in the red nucleus in one patient and a cystic degeneration of the olivary nucleus was seen 3 years after the HOD in a second patient. In two patients we found HOD that could only be explained by the existence of an afferent feedback loop between the dentate nucleus and the inferior olivary nucleus and by the knowledge that these fibers run through the ipsilateral olivary nucleus before ending in the contralateral olivary nucleus. In one of these patients the lesion was located in the inferior cerebellar peduncle. In the other patient the lesion was located on the midline in the medulla oblongata. The imaging findings in these patients reveal new observations in the stages of imaging appearances in HOD and shed light on the forgotten dentato-olivary afferent feedback loop of the DROP. Clin. Anat. 30:543-549, 2017. (c) 2017 Wiley Periodicals, Inc.
C1 [Smets, Gitte; Lambert, Julie; Mai, Cindy; Demaerel, Philippe] Univ Hosp KU Leuven, Dept Radiol, Herestr 49, B-3000 Leuven, Belgium.
   [Tijssen, Maud] Maastricht Univ, Dept Radiol, Med Ctr, POB 5800, NL-6202 AZ Maastricht, Netherlands.
   [Decramer, Thomas; Van Loon, Johannes] Univ Hosp KU Leuven, Dept Neurosurg, Herestr 49, B-3000 Leuven, Belgium.
   [Vandenberghe, Wim] Univ Hosp KU Leuven, Dept Neurol, Herestr 49, B-3000 Leuven, Belgium.
C3 KU Leuven; University Hospital Leuven; Maastricht University; KU Leuven;
   University Hospital Leuven; KU Leuven; University Hospital Leuven
RP Demaerel, P (通讯作者)，Univ Hosp KU Leuven, Dept Radiol, Herestr 49, B-3000 Leuven, Belgium.
EM
OI Mai, Cindy/0000-0003-0261-8851; Decramer, Thomas/0000-0002-2585-581X;
   Lambert, Julie/0000-0001-7385-3731
CR Akar S, 2008, J NEURO-ONCOL, V87, P341, DOI 10.1007/s11060-008-9523-7
   ANDERSON JR, 1973, J NEUROL NEUROSUR PS, V36, P826, DOI 10.1136/jnnp.36.5.826
   [Anonymous], HUMAN NERVOUS SYSTEM
   ARMSTRONG DM, 1974, PHYSIOL REV, V54, P358, DOI 10.1152/physrev.1974.54.2.358
   Bazzigaluppi P, 2012, PLOS ONE, V7, DOI 10.1371/journal.pone.0046360
   Conforto AB, 2005, ARQ NEURO-PSIQUIAT, V63, P321, DOI 10.1590/S0004-282X2005000200022
   Decramer T, 2015, JAMA NEUROL, V72, P1206, DOI 10.1001/jamaneurol.2015.1170
   Gatlin JL, 2011, J RADIOL CASE REP, V5, P24, DOI 10.3941/jrcr.v5i3.603
   Goyal M, 2000, AM J NEURORADIOL, V21, P1073
   Gu CN, 2015, J NEUROIMAGING, V25, P813, DOI 10.1111/jon.12267
   Guillain G, 1931, REV NEUROL-FRANCE, V56, P545
   Harter DH, 2004, J NEUROSURG, V100, P717, DOI 10.3171/jns.2004.100.4.0717
   Hornyak M, 2008, ACTA NEUROCHIR, V150, P149, DOI 10.1007/s00701-007-1470-0
   HOROUPIAN DS, 1971, J NEUROPATH EXP NEUR, V30, P571, DOI 10.1097/00005072-197110000-00002
   Inoue Y, 2014, J STROKE CEREBROVASC, V23, P554, DOI 10.1016/j.jstrokecerebrovasdis.2013.03.025
   Jang L, 2014, EUR NEUROL, V72, P144, DOI 10.1159/000360531
   JELLINGER K, 1973, Z NEUROL, V205, P153, DOI 10.1007/BF00316018
   KITAJIMA M, 1994, RADIOLOGY, V192, P539, DOI 10.1148/radiology.192.2.8029428
   Konno T, 2016, PARKINSONISM RELAT D, V28, P36, DOI 10.1016/j.parkreldis.2016.04.008
   Krings T, 2003, J NEUROL NEUROSUR PS, V74, P797, DOI 10.1136/jnnp.74.6.797
   LAPRESLE J, 1970, ARCH NEUROL-CHICAGO, V22, P135, DOI 10.1001/archneur.1970.00480200041004
   Nishie M, 2002, BRAIN, V125, P1348, DOI 10.1093/brain/awf126
   Phatouros CC, 1998, PEDIATR RADIOL, V28, P830, DOI 10.1007/s002470050475
   Salamon-Murayama N, 1999, RADIOLOGY, V213, P814, DOI 10.1148/radiology.213.3.r99dc43814
   Tartaglione T, 2015, RADIOL MED, V120, P474, DOI 10.1007/s11547-014-0477-x
   Tsui EYK, 1999, CLIN IMAG, V23, P215, DOI 10.1016/S0899-7071(99)00147-3
   UCHINO A, 1993, NEURORADIOLOGY, V35, P335, DOI 10.1007/BF00588362
   Van Der Giessen RS, 2008, NEURON, V58, P599, DOI 10.1016/j.neuron.2008.03.016
   Yun JH, 2013, ACTA NEUROCHIR, V155, P469, DOI 10.1007/s00701-012-1567-y
NR 29
TC 5
Z9 6
U1 0
U2 2
PU WILEY
PI HOBOKEN
PA 111 RIVER ST, HOBOKEN 07030-5774, NJ USA
SN 0897-3806
EI 1098-2353
J9 CLIN ANAT
JI Clin. Anat.
PD MAY
PY 2017
VL 30
IS 4
BP 543
EP 549
DI 10.1002/ca.22866
PG 7
WC Anatomy & Morphology
WE Science Citation Index Expanded (SCI-EXPANDED)
SC Anatomy & Morphology
GA ES9TV
UT WOS:000399905000026
PM 28247932
DA 2023-08-27
ER

PT J
AU Sanchez-Pernaute, R
   Studer, L
   Ferrari, D
   Perrier, A
   Lee, H
   Vinuela, A
   Isacson, O
AF Sanchez-Pernaute, R
   Studer, L
   Ferrari, D
   Perrier, A
   Lee, H
   Vinuela, A
   Isacson, O
TI Long-term survival of dopamine neurons derived from parthenogenetic
   primate embryonic stem cells (Cyno-1) after transplantation
SO STEM CELLS
LA English
DT Article
DE Parkinson's disease; embryonic stem (ES) cell; transplantation; primate;
   differentiation
ID NEURAL PROGENITOR CELLS; CENTRAL-NERVOUS-SYSTEM; IN-VITRO; RAT MODEL;
   PARKINSONS-DISEASE; SEROTONERGIC NEURONS; FATE SPECIFICATION; HUMAN
   BLASTOCYSTS; DEVELOPING BRAIN; ES CELLS
AB Dopamine (DA) neurons can be derived from human and primate embryonic stem (ES) cells in vitro. An ES cell-based replacement therapy for patients with Parkinson's disease requires that in vitro-generated neurons maintain their phenotype in vivo. Other critical issues relate to their proliferative capacity and risk of tumor formation, and the capability of migration and integration in the adult mammalian brain. Neural induction was achieved by coculture of primate parthenogenetic ES cells (Cyno-1) with stromal cells, followed by sequential exposure to midbrain patterning and differentiation factors to favor DA phenotypic specification. Differentiated ES cells were treated with mitomycin C and transplanted into adult immunosuppressed rodents and into a primate (allograft) without immunosuppression. A small percentage of DA neurons survived in both rodent and primate hosts for the entire term of the study (4 and 7 months, respectively). Other neuronal and glial populations derived from Cyno-1 ES cells showed, in vivo, phenotypic characteristics and growth and migration patterns similar to fetal primate transplants, and a majority of cells (> 80 %) expressed the forebrain transcription factor brain factor 1. No teratoma formation was observed. In this study, we demonstrate long-term survival of DA neurons obtained in vitro from primate ES cells. Optimization of differentiation, cell selection, and cell transfer is required for functional studies of ES-derived DA neurons for future therapeutic applications.
C1 Harvard Univ, Udall Parkinsons Dis Res Ctr Excellence, McLean Hosp, Belmont, MA 02478 USA.
   McLean Hosp, Neuroregenerat Labs, Belmont, MA 02178 USA.
   Mem Sloan Kettering Canc Ctr, Lab Stem Cell & Tumor Biol, Div Neurosurg, New York, NY 10021 USA.
C3 Harvard University; McLean Hospital; National Institutes of Health (NIH)
   - USA; NIH National Library of Medicine (NLM); Harvard University;
   McLean Hospital; Memorial Sloan Kettering Cancer Center
RP Sanchez-Pernaute, R (通讯作者)，Harvard Univ, Udall Parkinsons Dis Res Ctr Excellence, McLean Hosp, 115 Mill St, Belmont, MA 02478 USA.
EM
RI Sanchez-Pernaute, Rosario/ABD-8661-2021; Perrier, Anselme L/B-2272-2019;
   Studer, Lorenz/ABH-1491-2021; Perrier, Anselme/E-3764-2010;
   Sanchez-Pernaute, Rosario/AAZ-9566-2021; Ferrari, Daniela/AHE-7647-2022
OI Perrier, Anselme L/0000-0001-7915-1968; Studer,
   Lorenz/0000-0003-0741-7987; Sanchez-Pernaute,
   Rosario/0000-0003-1144-9025; Ferrari, Daniela/0000-0001-7251-9854;
   Isacson, Ole/0000-0001-8091-8302
FU NCRR NIH HHS [P51 RR000168, P51RR00168] Funding Source: Medline; NINDS
   NIH HHS [NS-39793, P50 NS039793-06A1, P50 NS039793] Funding Source:
   Medline
CR ABERCROMBIE M, 1946, ANAT REC, V94, P239, DOI 10.1002/ar.1090940210
   Barberi T, 2003, NAT BIOTECHNOL, V21, P1200, DOI 10.1038/nbt870
   Ben-Hur T, 2004, STEM CELLS, V22, P1246, DOI 10.1634/stemcells.2004-0094
   Bjorklund LM, 2002, P NATL ACAD SCI USA, V99, P2344, DOI 10.1073/pnas.022438099
   Cibelli JB, 2002, SCIENCE, V295, P819, DOI 10.1126/science.1065637
   Doetsch F, 1996, P NATL ACAD SCI USA, V93, P14895, DOI 10.1073/pnas.93.25.14895
   Echevarria D, 2003, BRAIN RES REV, V43, P179, DOI 10.1016/j.brainresrev.2003.08.002
   Fricker RA, 1999, J NEUROSCI, V19, P5990, DOI 10.1523/JNEUROSCI.19-14-05990.1999
   Galpern WR, 1996, EXP NEUROL, V140, P1, DOI 10.1006/exnr.1996.0109
   Goridis C, 2002, NAT REV NEUROSCI, V3, P531, DOI 10.1038/nrn871
   Goridis C, 1999, CURR OPIN NEUROBIOL, V9, P47, DOI 10.1016/S0959-4388(99)80006-3
   GrasbonFrodl EM, 1996, NEUROSCIENCE, V73, P171, DOI 10.1016/0306-4522(96)00008-5
   GrasbonFrodl EM, 1997, NEUROBIOL DIS, V3, P299, DOI 10.1006/nbdi.1996.0124
   Gunhaga L, 2003, NAT NEUROSCI, V6, P701, DOI 10.1038/nn1068
   Hanashima C, 2004, SCIENCE, V303, P56, DOI 10.1126/science.1090674
   Harper JM, 2004, P NATL ACAD SCI USA, V101, P7123, DOI 10.1073/pnas.0401103101
   Hitoshi S, 2002, DEVELOPMENT, V129, P233
   Holzschuh J, 2003, J NEUROSCI, V23, P5507
   Hurelbrink CB, 2002, EUR J NEUROSCI, V15, P1255, DOI 10.1046/j.1460-9568.2002.01959.x
   Hynes M, 1999, CURR OPIN NEUROBIOL, V9, P26, DOI 10.1016/S0959-4388(99)80004-X
   Johe KK, 1996, GENE DEV, V10, P3129, DOI 10.1101/gad.10.24.3129
   Kawasaki H, 2002, P NATL ACAD SCI USA, V99, P1580, DOI 10.1073/pnas.032662199
   Kawasaki H, 2000, NEURON, V28, P31, DOI 10.1016/S0896-6273(00)00083-0
   Kim JH, 2002, NATURE, V418, P50, DOI 10.1038/nature00900
   Kornack DR, 1998, P NATL ACAD SCI USA, V95, P1242, DOI 10.1073/pnas.95.3.1242
   Kornack DR, 2001, SCIENCE, V294, P2127, DOI 10.1126/science.1065467
   Lee SH, 2000, NAT BIOTECHNOL, V18, P675, DOI 10.1038/76536
   LEVITT P, 1982, DEV BRAIN RES, V4, P35, DOI 10.1016/0165-3806(82)90095-5
   Lois C, 1996, SCIENCE, V271, P978, DOI 10.1126/science.271.5251.978
   Mendez I, 2000, J NEUROSURG, V92, P493, DOI 10.3171/jns.2000.92.3.0493
   Ostenfeld T, 2000, EXP NEUROL, V164, P215, DOI 10.1006/exnr.2000.7427
   Reubinoff BE, 2001, NAT BIOTECHNOL, V19, P1134, DOI 10.1038/nbt1201-1134
   Reubinoff BE, 2000, NAT BIOTECHNOL, V18, P399, DOI 10.1038/74447
   ROUSSELOT P, 1995, J COMP NEUROL, V351, P51, DOI 10.1002/cne.903510106
   Sanchez-Pernaute R, 2001, J NEUROSCI RES, V65, P284, DOI 10.1002/jnr.1152
   Schuurmans C, 2002, CURR OPIN NEUROBIOL, V12, P26, DOI 10.1016/S0959-4388(02)00286-6
   Simerly C, 2003, SCIENCE, V300, P297, DOI 10.1126/science.1082091
   Simon HH, 2003, ANN NY ACAD SCI, V991, P36
   Svendsen CN, 1997, EXP NEUROL, V148, P135, DOI 10.1006/exnr.1997.6634
   Takagi Y, 2005, J CLIN INVEST, V115, P102, DOI 10.1172/JCI200521137
   Thomson JA, 1998, SCIENCE, V282, P1145, DOI 10.1126/science.282.5391.1145
   Tsai RYL, 2002, DEV CELL, V2, P707, DOI 10.1016/S1534-5807(02)00195-8
   Vrana KE, 2003, P NATL ACAD SCI USA, V100, P11911, DOI 10.1073/pnas.2034195100
   Whetton AD, 1999, TRENDS CELL BIOL, V9, P233, DOI 10.1016/S0962-8924(99)01559-7
   Winkler C, 1998, MOL CELL NEUROSCI, V11, P99, DOI 10.1006/mcne.1998.0674
   Yang M, 2004, CELL TRANSPLANT, V13, P535, DOI 10.3727/000000004783983729
   Ye WL, 1998, CELL, V93, P755, DOI 10.1016/S0092-8674(00)81437-3
   Zappone MV, 2000, DEVELOPMENT, V127, P2367
   Zhang SC, 2001, NAT BIOTECHNOL, V19, P1129, DOI 10.1038/nbt1201-1129
NR 49
TC 96
Z9 104
U1 0
U2 7
PU WILEY
PI HOBOKEN
PA 111 RIVER ST, HOBOKEN 07030-5774, NJ USA
SN 1066-5099
EI 1549-4918
J9 STEM CELLS
JI Stem Cells
PD AUG
PY 2005
VL 23
IS 7
BP 914
EP 922
DI 10.1634/stemcells.2004-0172
PG 9
WC Cell & Tissue Engineering; Biotechnology & Applied Microbiology;
   Oncology; Cell Biology; Hematology
WE Science Citation Index Expanded (SCI-EXPANDED)
SC Cell Biology; Biotechnology & Applied Microbiology; Oncology; Hematology
GA 957MN
UT WOS:000231374000007
PM 15941857
OA Green Accepted, Bronze
DA 2023-08-27
ER

PT J
AU Milicevic, NM
   Nohroudi, K
   Labudovic-Borovic, M
   Milicevic, Z
   Pfeffer, K
   Westermann, J
AF Milicevic, Novica M.
   Nohroudi, Klaus
   Labudovic-Borovic, Milica
   Milicevic, Zivana
   Pfeffer, Klaus
   Westermann, Juergen
TI Metallophilic macrophages are lacking in the thymus of lymphotoxin-beta
   receptor-deficient mice
SO HISTOCHEMISTRY AND CELL BIOLOGY
LA English
DT Article
DE lymphotoxin-beta receptor; thymus; macrophages; dendritic cells; mouse
ID PROMISCUOUS GENE-EXPRESSION; CORTICOMEDULLARY ZONE; RAT THYMUS;
   GERMINAL-CENTERS; EPITHELIAL-CELLS; DENDRITIC CELLS; NEGATIVE SELECTION;
   CLONAL DELETION; CYCLOSPORINE-A; IN-VITRO
AB Lymphotoxin-beta receptor (LT beta R) axis plays a crucial role in development and compartmentalization of peripheral lymphatic organs. But, it is also required for the appropriate function and maintenance of structural integrity of the thymus: in LT beta R-deficient animals the clonal deletion of autoreactive lymphocytes is impaired and differentiation of thymic medullary epithelial cells is disturbed. In this study, using several markers, we showed that thymic metallophilic macrophages were lacking in LT beta R-deficient mice. In tumor necrosis factor receptor-I (p55)-deficient mice (which we used as positive control) thymic metallophilic cells were located, similarly as in normal mice, in the thymic cortico-medullary zone at the junction of cortex and medulla. These findings show that LT beta R is necessary for maintenance of metallophilic macrophages in the thymus and provide further evidence that these cells may represent a factor involved in thymic negative selection.
C1 Univ Belgrade, Sch Med, Inst Histol & Embryol, Belgrade 11000, Serbia.
   Med Univ Lubeck, Inst Anat, Ctr Struct & Cell Biol Med, Lubeck, Germany.
   Univ Dusseldorf, Inst Med Microbiol, Dusseldorf, Germany.
C3 University of Belgrade; University of Lubeck; Heinrich Heine University
   Dusseldorf
RP Milicevic, NM (通讯作者)，Univ Belgrade, Sch Med, Inst Histol & Embryol, Visegradska 26, Belgrade 11000, Serbia.
EM
OI Pfeffer, Klaus/0000-0002-5652-6330
CR ANDERSON G, 1994, J EXP MED, V179, P2027, DOI 10.1084/jem.179.6.2027
   Bancroft J., 1982, THEORY PRACTICE HIST
   Boehm T, 2003, J EXP MED, V198, P757, DOI 10.1084/jem.20030794
   CHEN JW, 1985, ARCH BIOCHEM BIOPHYS, V239, P574, DOI 10.1016/0003-9861(85)90727-1
   Chin RK, 2003, NAT IMMUNOL, V4, P1121, DOI 10.1038/ni982
   Derbinski J, 2005, J EXP MED, V202, P33, DOI 10.1084/jem.20050471
   Derbinski J, 2001, NAT IMMUNOL, V2, P1032, DOI 10.1038/ni723
   Futterer A, 1998, IMMUNITY, V9, P59, DOI 10.1016/S1074-7613(00)80588-9
   Hogquist KA, 2005, NAT REV IMMUNOL, V5, P772, DOI 10.1038/nri1707
   JENKINS MK, 1988, SCIENCE, V241, P1655, DOI 10.1126/science.3262237
   JENKINSON EJ, 1992, J EXP MED, V176, P845, DOI 10.1084/jem.176.3.845
   Kabashima K, 2005, IMMUNITY, V22, P439, DOI 10.1016/j.immuni.2005.02.007
   KRAAL G, 1986, IMMUNOLOGY, V58, P665
   Leder L D, 1975, Verh Dtsch Ges Pathol, V59, P503
   Liston A, 2004, J EXP MED, V200, P1015, DOI 10.1084/jem.20040581
   Liston A, 2003, NAT IMMUNOL, V4, P350, DOI 10.1038/ni906
   MartinezPomares L, 1996, J EXP MED, V184, P1927, DOI 10.1084/jem.184.5.1927
   Matsumoto M, 1997, IMMUNOL REV, V156, P137, DOI 10.1111/j.1600-065X.1997.tb00965.x
   MILICEVIC NM, 1989, VIRCHOWS ARCH B, V57, P237, DOI 10.1007/BF02899087
   MILICEVIC NM, 1985, VIRCHOWS ARCH B, V50, P193
   MILICEVIC NM, 1983, J RETICULOENDOTH SOC, V34, P501
   MILICEVIC NM, 1987, J ANAT, V150, P89
   MILICEVIC NM, 1984, J LEUKOCYTE BIOL, V36, P761, DOI 10.1002/jlb.36.6.761
   Milicevic NM, 2000, ANN ANAT, V182, P471, DOI 10.1016/S0940-9602(00)80062-X
   Milicevic NM, 2004, INT REV CYTOL, V235, P1, DOI 10.1016/S0074-7696(04)35001-1
   MILICEVIC NM, 1989, J COMP PATHOL, V101, P143, DOI 10.1016/0021-9975(89)90062-5
   Milicevic NM, 1998, HISTOL HISTOPATHOL, V13, P1183, DOI 10.14670/HH-13.1183
   Sprent J, 2002, IMMUNOL REV, V185, P126, DOI 10.1034/j.1600-065X.2002.18512.x
   Villasenor J, 2005, IMMUNOL REV, V204, P156, DOI 10.1111/j.0105-2896.2005.00246.x
   Wang YG, 2005, J IMMUNOL, V175, P6997, DOI 10.4049/jimmunol.175.10.6997
   Yu P, 2002, J IMMUNOL, V168, P5117, DOI 10.4049/jimmunol.168.10.5117
NR 31
TC 12
Z9 12
U1 0
U2 0
PU SPRINGER
PI NEW YORK
PA 233 SPRING ST, NEW YORK, NY 10013 USA
SN 0948-6143
EI 1432-119X
J9 HISTOCHEM CELL BIOL
JI Histochem. Cell Biol.
PD DEC
PY 2006
VL 126
IS 6
BP 687
EP 693
DI 10.1007/s00418-006-0202-5
PG 7
WC Cell Biology; Microscopy
WE Science Citation Index Expanded (SCI-EXPANDED)
SC Cell Biology; Microscopy
GA 113UW
UT WOS:000242624400005
PM 16830123
DA 2023-08-27
ER

PT J
AU Pan, L
   Wang, EM
   Wang, BJ
   Zhou, LF
   Zhang, N
   Cai, PW
   Da, JZ
AF Pan, L
   Wang, EM
   Wang, BJ
   Zhou, LF
   Zhang, N
   Cai, PW
   Da, JZ
TI Gamma Knife radiosurgery for hemangioblastomas
SO STEREOTACTIC AND FUNCTIONAL NEUROSURGERY
LA English
DT Article
DE radiosurgery; hemangioblastoma; von Hippel Lindau; histopathology
ID CENTRAL-NERVOUS-SYSTEM; ARTERIOVENOUS-MALFORMATIONS
AB Thirteen patients (11 males, 2 females) with cerebral hemangioblastomas (HABs) were treated with Gamma Knife radiosurgery (GKR). Four patients had multiple lesions in the brain. The remainder had a single lesion. The total number of lesions was 20, Eight cases had recurrent or residual HABs after surgery. In one case diagnosis was confirmed following surgical resection 22 months after GKR, One case was diagnosed by computed tomography (CT), magnetic resonance imaging (MRI) and digital subtraction angiography (DSA), Three patients had von Hippel Lindau (VHL) disease. The mean tumor diameter was 20 mm (range 7.5 to .55 mm). The mean margin dose was 18 Gy (range 12 to 24 Gy),
   In 5 cases, there was an improvement of symptoms and reduction in tumor volume. In 4 cases the tumor volume and clinical status remained unchanged. In 3 patients, there was clinical deterioration, The cause of this was an increase in tumor cyst volume in 2 cases. Subsequent surgery resulted in clinical improvement. In a third patient with multiple lesions, deterioration was the result of adverse radiation effects in the medulla oblongata.
   Three patients were subjected to post GKR-surgery with subsequent histopathology, In one, this was due to cyst expansion. In one, it was at the patient's insistence in the presence of a stable clinical and radiological picture. In a third patient with a temporal lobe tumor, it was because of late-developing epileptic seizures. The histopathological findings in these patients showed varying degrees of small vessel thickening and occlusion together with loss of tumor cells. The observations varied in degree according to the time between GKR and the secondary operation. These findings indicate the effectiveness of the treatment. The reduction in vascularity suggests that GKR could make subsequent surgery less hazardous.
   The observations of this study suggest that while GKR is not adequately reliable for the control of HAB cysts, it can be an effective treatment for solid tumors, especially those in eloquent regions.
C1 Shanghai Med Univ, Shanghai Gamma Knife Hosp, Hua Shan Hosp, Dept Neurosurg, Shanghai 200233, Peoples R China.
   Shanghai Med Univ, Shanghai Gamma Knife Hosp, Hua Shan Hosp, Dept Radiol, Shanghai 200233, Peoples R China.
C3 Fudan University; Fudan University
RP Pan, L (通讯作者)，Shanghai Med Univ, Shanghai Gamma Knife Hosp, Hua Shan Hosp, Dept Neurosurg, 518 Wuzhong Dong Rd, Shanghai 200233, Peoples R China.
CR CHANDLER HC, 1994, NEUROSURGERY, V34, P353, DOI 10.1227/00006123-199402000-00021
   CHAUDHRY AP, 1978, CANCER, V42, P1834, DOI 10.1002/1097-0142(197810)42:4<1834::AID-CNCR2820420423>3.0.CO;2-Z
   DEGERBLAD M, 1986, ACTA ENDOCRINOL-COP, V112, P310, DOI 10.1530/acta.0.1120310
   FRIEDMAN WA, 1992, J NEUROSURG, V77, P832, DOI 10.3171/jns.1992.77.6.0832
   HELLE TL, 1980, NEUROSURGERY, V6, P82, DOI 10.1227/00006123-198001000-00012
   KERR DJ, 1995, NEUROSURGERY, V36, P573, DOI 10.1227/00006123-199503000-00017
   KONDZIOLKA D, 1991, J NEUROSURG, V74, P552, DOI 10.3171/jns.1991.74.4.0552
   Linskey M E, 1992, Neurosurg Clin N Am, V3, P191
   MELMON KL, 1964, AM J MED, V36, P595, DOI 10.1016/0002-9343(64)90107-X
   NEUMANN HPH, 1989, J NEUROSURG, V70, P24, DOI 10.3171/jns.1989.70.1.0024
   Niemela M, 1996, J NEUROSURG, V85, P591, DOI 10.3171/jns.1996.85.4.0591
   PAGE KA, 1993, SURG NEUROL, V40, P424, DOI 10.1016/0090-3019(93)90225-P
   SMALLEY SR, 1990, INT J RADIAT ONCOL, V18, P1165, DOI 10.1016/0360-3016(90)90454-R
   STEINER L, 1992, J NEUROSURG, V77, P1, DOI 10.3171/jns.1992.77.1.0001
NR 14
TC 52
Z9 52
U1 0
U2 2
PU KARGER
PI BASEL
PA ALLSCHWILERSTRASSE 10, CH-4009 BASEL, SWITZERLAND
SN 1011-6125
J9 STEREOT FUNCT NEUROS
JI Stereotact. Funct. Neurosurg.
PY 1998
VL 70
SU 1
BP 179
EP 186
DI 10.1159/000056420
PG 8
WC Neurosciences; Neuroimaging; Surgery
WE Science Citation Index Expanded (SCI-EXPANDED)
SC Neurosciences & Neurology; Surgery
GA 131TM
UT WOS:000076591400021
PM 9782249
DA 2023-08-27
ER

PT J
AU Unal-Cevik, I
   Onal, MZ
   Odabasi, Z
   Tan, E
AF Unal-Cevik, Isin
   Onal, Mehmet Zulkuf
   Odabasi, Zeki
   Tan, Ersin
TI IVIG- Responsive Multiple Cranial Neuropathy: a pharyngo-facial variant
   of Guillain-Barre Syndrome
SO ACTA NEUROLOGICA BELGICA
LA English
DT Article
DE Bilateral facial diplegia; Guillain-Barre syndrome; dysphagia;
   neuropathy; intravenous immunoglobulin therapy
ID INFLAMMATORY DEMYELINATING POLYNEUROPATHY; NEURONOPATHY FOSMN SYNDROME;
   NERVE PALSIES; DIPLEGIA
AB We report a case with symptoms of facial swelling, bilateral facial paralysis, dysphagia and aspiration. On electrophysiological studies, the right facial nerve was not excitable. The left facial nerve compound muscle action potential (CMAP) amplitude was severely dispersed and latency was mildly prolonged, consistent with demyelination. Cerebrospinal fluid examinations were normal. Antiganglioside antibodies and tumor markers were negative. Bickerstaff brainstem. encephalitis, stoke, diabetes mellitus, vasculitis, sarcoidosis, Sjogren's syndrome, Melkersson-Rosenthal Syndrome, trauma, infectious diseases, toxicity, neoplasm, facial onset sensory and motor neuronopathy (FOSMN) and other degenerative diseases were excluded. Intravenous immunoglobulin therapy resolved symptoms of lower cranial nerve dysfunction. Clinically incomplete improvement of bilateral facial paralysis was observed. We conclude that IVIg therapy may improve the symptoms of multiple cranial nerve palsies clue to pharyngo-facial variant of Guillain-Barre syndrome.
C1 [Unal-Cevik, Isin; Onal, Mehmet Zulkuf] Ufuk Univ, Fac Med, Dept Neurol, TR-06520 Ankara, Turkey.
   [Odabasi, Zeki] Gulhane Mil Med Acad, Dept Neurol, Ankara, Turkey.
   [Tan, Ersin] Hacettepe Univ, Fac Med, Dept Neurol, TR-06100 Ankara, Turkey.
C3 Ufuk University; Gulhane Military Medical Academy; Hacettepe University
RP Unal-Cevik, I (通讯作者)，Ufuk Univ, Fac Med, Dept Neurol, TR-06520 Ankara, Turkey.
EM
RI UNAL-CEVIK, ISIN/R-1672-2019; UNAL-CEVIK, ISIN/I-9553-2013
OI UNAL-CEVIK, ISIN/0000-0003-1851-7561; Odabasi, Zeki/0000-0001-7945-2015
CR BEAL MF, 1990, NEW ENGL J MED, V322, P461, DOI 10.1056/NEJM199002153220709
   Curtis CE, 2008, MIL MED, V173, P507, DOI 10.7205/MILMED.173.5.507
   Dididze M N, 2009, Georgian Med News, P48
   Gorman J B, 1977, Va Med, V104, P544
   Hokonohara T, 2008, J NEUROL SCI, V275, P157, DOI 10.1016/j.jns.2008.07.021
   KEANE JR, 1994, NEUROLOGY, V44, P1198, DOI 10.1212/WNL.44.7.1198
   Khandpur S, 2006, J DERMATOL, V33, P411, DOI 10.1111/j.1346-8138.2006.00097.x
   Kokubun N, 2007, MUSCLE NERVE, V35, P203, DOI 10.1002/mus.20679
   Levin KH, 2004, NEUROLOGIST, V10, P61, DOI 10.1097/01.nrl.0000117821.35196.0b
   MARIANI C, 1980, J NEUROL, V223, P269, DOI 10.1007/BF00313341
   Mas-Lazaro C, 2008, REV NEUROLOGIA, V47, P579, DOI 10.33588/rn.4711.2008074
   Mukherjee P K, 1973, Indian J Ophthalmol, V21, P36
   Nobile-Orazio E, 2005, J NEUROL, V252, P7, DOI 10.1007/s00415-005-1103-6
   Odabasi Z, 1996, MUSCLE NERVE, V19, P210, DOI 10.1002/(SICI)1097-4598(199602)19:2<210::AID-MUS13>3.0.CO;2-Y
   Odaka M, 2003, BRAIN, V126, P2279, DOI 10.1093/brain/awg233
   SHUAIB A, 1987, CAN J NEUROL SCI, V14, P611
   Sinsawaiwong Suwanna, 2000, Journal of the Medical Association of Thailand, V83, P1124
   SPILLANE L, 1991, ANN EMERG MED, V20, P208, DOI 10.1016/S0196-0644(05)81226-0
   Vucic S, 2006, BRAIN, V129, P3384, DOI 10.1093/brain/awl258
   WEINTRAUB MI, 1976, ARCH OTOLARYNGOL, V102, P311
   Willison HJ, 2002, BRAIN, V125, P2591, DOI 10.1093/brain/awf272
NR 21
TC 5
Z9 5
U1 1
U2 4
PU SPRINGER HEIDELBERG
PI HEIDELBERG
PA TIERGARTENSTRASSE 17, D-69121 HEIDELBERG, GERMANY
SN 0300-9009
EI 2240-2993
J9 ACTA NEUROL BELG
JI Acta Neurol. Belg.
PD DEC
PY 2009
VL 109
IS 4
BP 317
EP 321
PG 5
WC Clinical Neurology; Neurosciences
WE Science Citation Index Expanded (SCI-EXPANDED)
SC Neurosciences & Neurology
GA 545MM
UT WOS:000273736400012
PM 20120214
DA 2023-08-27
ER

PT J
AU Litteljohn, D
   Mangano, E
   Clarke, M
   Bobyn, J
   Moloney, K
   Hayley, S
AF Litteljohn, Darcy
   Mangano, Emily
   Clarke, Melanie
   Bobyn, Jessica
   Moloney, Kerry
   Hayley, Shawn
TI Inflammatory Mechanisms of Neurodegeneration in Toxin-Based Models of
   Parkinson's Disease
SO PARKINSONS DISEASE
LA English
DT Review
ID TUMOR-NECROSIS-FACTOR; NF-KAPPA-B; NONSTEROIDAL ANTIINFLAMMATORY DRUGS;
   ENVIRONMENTAL RISK-FACTORS; NITRIC-OXIDE SYNTHASE;
   LIPOPOLYSACCHARIDE-INDUCED NEUROTOXICITY; PROTECTS DOPAMINERGIC-NEURONS;
   TRAUMATIC BRAIN-INJURY; II-POSITIVE MICROGLIA; INTERFERON-GAMMA
AB Parkinson's disease (PD) has been associated with exposure to a variety of environmental agents, including pesticides, heavy metals, and organic pollutants; and inflammatory processes appear to constitute a common mechanistic link among these insults. Indeed, toxin exposure has been repeatedly demonstrated to induce the release of oxidative and inflammatory factors from immunocompetent microglia, leading to damage and death of midbrain dopamine (DA) neurons. In particular, proinflammatory cytokines such as tumor necrosis factor-alpha and interferon-gamma, which are produced locally within the brain by microglia, have been implicated in the loss of DA neurons in toxin-based models of PD; and mounting evidence suggests a contributory role of the inflammatory enzyme, cyclooxygenase-2. Likewise, immune-activating bacterial and viral agents were reported to have neurodegenerative effects themselves and to augment the deleterious impact of chemical toxins upon DA neurons. The present paper will focus upon the evidence linking microglia and their inflammatory processes to the death of DA neurons following toxin exposure. Particular attention will be devoted to the possibility that environmental toxins can activate microglia, resulting in these cells adopting a "sensitized" state that favors the production of proinflammatory cytokines and damaging oxidative radicals.
C1 [Litteljohn, Darcy; Mangano, Emily; Clarke, Melanie; Bobyn, Jessica; Moloney, Kerry; Hayley, Shawn] Carleton Univ, Inst Neurosci, Ottawa, ON K1S 5B6, Canada.
C3 Carleton University
RP Hayley, S (通讯作者)，Carleton Univ, Inst Neurosci, 1125 Colonel Dr, Ottawa, ON K1S 5B6, Canada.
EM
OI Rocha, Emily/0000-0003-2171-5104
FU Canadian Institutes of Health Research (CIHR); Parkinson's Society
   Canada
FX D. Litteljohn and S. Hayley were supported by funds from the Canadian
   Institutes of Health Research (CIHR) and Parkinson's Society Canada. S.
   Hayley holds a Canada Research Chair in Behavioural Neuroscience.
CR Ahmad AS, 2006, J NEUROTRAUM, V23, P1895, DOI 10.1089/neu.2006.23.1895
   Anisman H, 2008, PROG NEUROBIOL, V85, P1, DOI 10.1016/j.pneurobio.2008.01.004
   [Anonymous], PSYCHONEUROIMMUNOLOG
   Anrather J, 2006, J BIOL CHEM, V281, P5657, DOI 10.1074/jbc.M506172200
   APPLEBY SB, 1994, BIOCHEM J, V302, P723, DOI 10.1042/bj3020723
   Asanuma M, 2001, J NEUROCHEM, V76, P1895, DOI 10.1046/j.1471-4159.2001.00205.x
   Baba Y, 2005, PARKINSONISM RELAT D, V11, P493, DOI 10.1016/j.parkreldis.2005.07.005
   Baldi I, 2003, NEUROEPIDEMIOLOGY, V22, P305, DOI 10.1159/000071194
   BANKS WA, 1991, J PHARMACOL EXP THER, V259, P988
   Baquet ZC, 2005, J NEUROSCI, V25, P6251, DOI 10.1523/JNEUROSCI.4601-04.2005
   Barlow BK, 2007, REPROD TOXICOL, V23, P457, DOI 10.1016/j.reprotox.2007.01.007
   Barone FC, 1998, STROKE, V29, P1937, DOI 10.1161/01.STR.29.9.1937
   Barone FC, 1997, STROKE, V28, P1233, DOI 10.1161/01.STR.28.6.1233
   Bazan NG, 2005, ANN NY ACAD SCI, V1053, P137, DOI 10.1196/annals.1344.011
   Benveniste EN, 1998, CYTOKINE GROWTH F R, V9, P259, DOI 10.1016/S1359-6101(98)00015-X
   Bian MJ, 2009, BRAIN RES, V1302, P256, DOI 10.1016/j.brainres.2009.07.030
   Bolin LM, 2002, J NEUROCHEM, V83, P167, DOI 10.1046/j.1471-4159.2002.01131.x
   Bonifati V, 2007, NEUROCHEM RES, V32, P1700, DOI 10.1007/s11064-007-9324-y
   Bornebroek M, 2007, NEUROEPIDEMIOLOGY, V28, P193, DOI 10.1159/000108110
   Boyd JD, 2007, BRAIN RES, V1175, P107, DOI 10.1016/j.brainres.2007.07.067
   Brochard V, 2009, J CLIN INVEST, V119, P182, DOI 10.1172/JCI36470
   Brock TG, 1999, J BIOL CHEM, V274, P11660, DOI 10.1074/jbc.274.17.11660
   Brodacki B, 2008, NEUROSCI LETT, V441, P158, DOI 10.1016/j.neulet.2008.06.040
   Brooks AI, 1999, BRAIN RES, V823, P1, DOI 10.1016/S0006-8993(98)01192-5
   Bruce AJ, 1996, NAT MED, V2, P788, DOI 10.1038/nm0796-788
   Cannon JR, 2009, NEUROBIOL DIS, V34, P279, DOI 10.1016/j.nbd.2009.01.016
   Carrasco E, 2007, J NEUROSCI RES, V85, P3109, DOI 10.1002/jnr.21425
   Carrasco E, 2008, NEUROSCI LETT, V441, P44, DOI 10.1016/j.neulet.2008.05.111
   Carvey PM, 2003, FRONT BIOSCI-LANDMRK, V8, pS826, DOI 10.2741/1158
   Castano A, 2002, J NEUROCHEM, V81, P150, DOI 10.1046/j.1471-4159.2002.00799.x
   Chandrasekharan NV, 2002, P NATL ACAD SCI USA, V99, P13926, DOI 10.1073/pnas.162468699
   Chanyachukul T, 2004, TOXICOL LETT, V150, P259, DOI 10.1016/j.toxlet.2004.02.007
   Chen HL, 2005, ANN NEUROL, V58, P963, DOI 10.1002/ana.20682
   Chen P, 2008, BIOMED ENVIRON SCI, V21, P233, DOI 10.1016/S0895-3988(08)60035-5
   Chen Q, 2010, NEUROTOXICOLOGY, V31, P310, DOI 10.1016/j.neuro.2010.02.006
   Choi WS, 2010, J NEUROPATH EXP NEUR, V69, P511, DOI 10.1097/NEN.0b013e3181db8100
   Cicchetti F, 2009, TRENDS PHARMACOL SCI, V30, P475, DOI 10.1016/j.tips.2009.06.005
   Clausen BH, 2008, J NEUROINFLAMM, V5, DOI 10.1186/1742-2094-5-46
   Clausen F, 2009, EUR J NEUROSCI, V30, P385, DOI 10.1111/j.1460-9568.2009.06820.x
   Costa D, 2006, BIOL PHARM BULL, V29, P1659, DOI 10.1248/bpb.29.1659
   Costello S, 2009, AM J EPIDEMIOL, V169, P919, DOI 10.1093/aje/kwp006
   Cotrina ML, 2000, J NEUROSCI, V20, P2835
   D'Amelio M, 2004, MOVEMENT DISORD, V19, P807, DOI 10.1002/mds.20123
   Davalos D, 2005, NAT NEUROSCI, V8, P752, DOI 10.1038/nn1472
   Davies NM, 2004, J PHARM PHARM SCI, V7, P217
   Ezcurra ALD, 2010, NEUROBIOL DIS, V37, P630, DOI 10.1016/j.nbd.2009.11.018
   Decker T, 2005, NAT REV IMMUNOL, V5, P675, DOI 10.1038/nri1684
   Dhillon Amanpreet S, 2008, J Agromedicine, V13, P37, DOI 10.1080/10599240801986215
   Dick FD, 2007, OCCUP ENVIRON MED, V64, P666, DOI 10.1136/oem.2006.027003
   DINARELLO CA, 1988, FASEB J, V2, P108, DOI 10.1096/fasebj.2.2.3277884
   Drolet RE, 2009, NEUROBIOL DIS, V36, P96, DOI 10.1016/j.nbd.2009.06.017
   Durbin J, 2003, J ENDOTOXIN RES, V9, P313, DOI 10.1179/096805103225002575
   Echeverria V, 2005, EUR J NEUROSCI, V22, P2199, DOI 10.1111/j.1460-9568.2005.04427.x
   Eigler A, 1997, IMMUNOL TODAY, V18, P487, DOI 10.1016/S0167-5699(97)01118-3
   Elmquist JK, 1997, J COMP NEUROL, V381, P119, DOI 10.1002/(SICI)1096-9861(19970505)381:2<119::AID-CNE1>3.0.CO;2-6
   Esposito E, 2008, J PINEAL RES, V44, P78, DOI 10.1111/j.1600-079X.2007.00492.x
   Etminan M, 2008, J CLIN NEUROSCI, V15, P576, DOI 10.1016/j.jocn.2007.02.095
   Farooqui AA, 2007, J NEUROCHEM, V101, P577, DOI 10.1111/j.1471-4159.2006.04371.x
   Feng ZH, 2002, NEUROSCI LETT, V329, P354, DOI 10.1016/S0304-3940(02)00704-8
   Ferger B, 2004, J NEUROCHEM, V89, P822, DOI 10.1111/j.1471-4159.2004.02399.x
   Firestone JA, 2005, ARCH NEUROL-CHICAGO, V62, P91, DOI 10.1001/archneur.62.1.91
   Gagne JJ, 2010, NEUROLOGY, V74, P995, DOI 10.1212/WNL.0b013e3181d5a4a3
   Gao HM, 2003, FASEB J, V17, DOI 10.1096/fj.03-0109fje
   Gao HM, 2003, J NEUROSCI, V23, P1228, DOI 10.1523/JNEUROSCI.23-04-01228.2003
   Gao HM, 2008, TRENDS IMMUNOL, V29, P357, DOI 10.1016/j.it.2008.05.002
   Glass CK, 2010, CELL, V140, P918, DOI 10.1016/j.cell.2010.02.016
   Goehler LE, 2006, NEUROSCIENCE, V140, P1415, DOI 10.1016/j.neuroscience.2006.03.048
   Goodman JC, 2008, ACTA NEUROCHIR SUPPL, V102, P437, DOI 10.1007/978-3-211-85578-2_85
   Greenamyre JT, 2010, TRENDS PHARMACOL SCI, V31, P141, DOI 10.1016/j.tips.2009.12.006
   Gribova I. E., 2003, Biomeditsinskaya Khimiya, V49, P208
   GUTIERREZ EG, 1993, J NEUROIMMUNOL, V47, P169, DOI 10.1016/0165-5728(93)90027-V
   Hagberg H, 2005, BJOG-INT J OBSTET GY, V112, P16, DOI 10.1111/j.1471-0528.2005.00578.x
   Hancock DB, 2008, BMC NEUROL, V8, DOI 10.1186/1471-2377-8-6
   Hartlage-Rubsamen M, 1999, J NEUROSCI RES, V57, P388, DOI 10.1002/(SICI)1097-4547(19990801)57:3<388::AID-JNR11>3.0.CO;2-2
   Hashioka S., NEUROBIOLOG IN PRESS
   Hashioka S, 2009, NEUROBIOL AGING, V30, P1924, DOI 10.1016/j.neurobiolaging.2008.02.019
   Hayley S, 2008, J NEUROIMMUNOL, V197, P29, DOI 10.1016/j.jneuroim.2008.03.015
   Hernan MA, 2006, NEUROLOGY, V66, P1097, DOI 10.1212/01.wnl.0000204446.82823.28
   Hirsch EC, 2009, LANCET NEUROL, V8, P382, DOI 10.1016/S1474-4422(09)70062-6
   Hunot S, 2004, P NATL ACAD SCI USA, V101, P665, DOI 10.1073/pnas.0307453101
   Hunot S, 1999, J NEUROSCI, V19, P3440
   Hunot S, 1996, NEUROSCIENCE, V72, P355, DOI 10.1016/0306-4522(95)00578-1
   Hutchinson AJ, 2009, NEUROCHEM INT, V54, P439, DOI 10.1016/j.neuint.2009.01.018
   Iczkiewicz J, 2006, BRAIN RES, V1118, P239, DOI 10.1016/j.brainres.2006.08.036
   Imamura K, 2003, ACTA NEUROPATHOL, V106, P518, DOI 10.1007/s00401-003-0766-2
   Jana M, 2001, J BIOL CHEM, V276, P44527, DOI 10.1074/jbc.M106771200
   Jin F, 2008, EUR J PHARMACOL, V600, P78, DOI 10.1016/j.ejphar.2008.10.005
   Johnston LC, 2008, MOL THER, V16, P1392, DOI 10.1038/mt.2008.113
   Ju KR, 2006, BRAIN RES, V1122, P209, DOI 10.1016/j.brainres.2006.09.022
   Kajihara M, 2010, EUR J GASTROEN HEPAT, V22, P628, DOI 10.1097/MEG.0b013e32833383e3
   Kamm K, 2006, J TRAUMA, V60, P152, DOI 10.1097/01.ta.0000196345.81169.a1
   Kang MJ, 2010, TOXICOL LETT, V195, P127, DOI 10.1016/j.toxlet.2010.03.011
   Kawaguchi N, 1999, PARKINSONISM RELAT D, V5, P43, DOI 10.1016/S1353-8020(98)00040-6
   Kawano T, 2006, NAT MED, V12, P225, DOI 10.1038/nm1362
   Kawanokuchi J, 2006, MULT SCLER J, V12, P558, DOI 10.1177/1352458506070763
   Kes VB, 2008, CLIN BIOCHEM, V41, P1330, DOI 10.1016/j.clinbiochem.2008.08.080
   Kim WG, 2000, J NEUROSCI, V20, P6309, DOI 10.1523/JNEUROSCI.20-16-06309.2000
   Klintworth H, 2009, NEUROSCI LETT, V462, P1, DOI 10.1016/j.neulet.2009.06.065
   Klockgether T, 2004, CELL TISSUE RES, V318, P115, DOI 10.1007/s00441-004-0975-6
   Knott C, 2000, MOL CELL NEUROSCI, V16, P724, DOI 10.1006/mcne.2000.0914
   Konsman JP, 2000, NEUROSCIENCE, V101, P957, DOI 10.1016/S0306-4522(00)00403-6
   Koprich JB, 2008, J NEUROINFLAMM, V5, DOI 10.1186/1742-2094-5-8
   Kostic VS, 2010, NEUROLOGY, V75, P857, DOI 10.1212/WNL.0b013e3181f11c1d
   Kurkowska-Jastrzebska I, 1999, ACTA NEUROBIOL EXP, V59, P1
   Li X, 2005, CHINESE MED J-PEKING, V118, P1357
   Ling ZD, 2006, EXP NEUROL, V199, P499, DOI 10.1016/j.expneurol.2006.01.010
   Ling ZD, 2004, EXP NEUROL, V190, P373, DOI 10.1016/j.expneurol.2004.08.006
   Liou HH, 1997, NEUROLOGY, V48, P1583, DOI 10.1212/WNL.48.6.1583
   Litteljohn D, 2009, NEUROSCIENCE, V164, P1894, DOI 10.1016/j.neuroscience.2009.09.025
   Litteljohn D, 2008, EUR J NEUROSCI, V28, P707, DOI 10.1111/j.1460-9568.2008.06371.x
   Loeffler DA, 2006, J NEUROINFLAMM, V3, DOI 10.1186/1742-2094-3-29
   Lucas PC, 2004, J CELL SCI, V117, P31, DOI 10.1242/jcs.00904
   Madrigal Jose L. M., 2006, CNS & Neurological Disorders-Drug Targets, V5, P561
   Maetzler W, 2007, NEUROBIOL DIS, V25, P473, DOI 10.1016/j.nbd.2006.10.020
   Mandel S, 2000, ADV RES NEURODEGENER, V8, P117
   Mangano E. N., NEUROBIOLOG IN PRESS
   Mangano EN, 2009, NEUROBIOL AGING, V30, P1361, DOI 10.1016/j.neurobiolaging.2007.11.020
   Manning-Bog AB, 2002, J BIOL CHEM, V277, P1641, DOI 10.1074/jbc.C100560200
   Marie RM, 1999, NEUROSCI LETT, V260, P77, DOI 10.1016/S0304-3940(98)00928-8
   Mattson MP, 2005, NEUROCHEM RES, V30, P883, DOI 10.1007/s11064-005-6961-x
   McCormack AL, 2002, NEUROBIOL DIS, V10, P119, DOI 10.1006/nbdi.2002.0507
   McCoy MK, 2008, MOL THER, V16, P1572, DOI 10.1038/mt.2008.146
   McCullough L, 2004, J NEUROSCI, V24, P257, DOI 10.1523/JNEUROSCI.4485-03.2004
   MCGEER PL, 1988, NEUROLOGY, V38, P1285, DOI 10.1212/WNL.38.8.1285
   Meller R, 2005, J CEREBR BLOOD F MET, V25, P217, DOI 10.1038/sj.jcbfm.9600022
   Menza Matthew, 2006, Curr Psychiatry Rep, V8, P234, DOI 10.1007/s11920-006-0029-8
   Meredith GE, 2008, ACTA NEUROPATHOL, V115, P385, DOI 10.1007/s00401-008-0350-x
   Michelucci A, 2009, J NEUROIMMUNOL, V210, P3, DOI 10.1016/j.jneuroim.2009.02.003
   Minghetti L, 2004, J NEUROPATH EXP NEUR, V63, P901, DOI 10.1093/jnen/63.9.901
   Mir M, 2009, NEUROSCIENCE, V162, P959, DOI 10.1016/j.neuroscience.2009.05.049
   Mizuno T, 2008, FASEB J, V22, P1797, DOI 10.1096/fj.07-099499
   Mizuta I, 2001, J NEUROL NEUROSUR PS, V71, P818
   Mogi M, 2007, NEUROSCI LETT, V414, P94, DOI 10.1016/j.neulet.2006.12.003
   Moran LB, 2004, NEUROGENETICS, V5, P95, DOI 10.1007/s10048-004-0172-5
   Moran LB, 2007, J NEUROIMMUNOL, V183, P1, DOI 10.1016/j.jneuroim.2006.10.023
   Mount MP, 2007, J NEUROSCI, V27, P3328, DOI 10.1523/JNEUROSCI.5321-06.2007
   Nadeau S, 1999, NEUROSCIENCE, V93, P1449, DOI 10.1016/S0306-4522(99)00225-0
   Nadjar A, 2005, J CEREBR BLOOD F MET, V25, P1047, DOI 10.1038/sj.jcbfm.9600106
   Nagatsu T, 2000, ADV RES NEURODEGENER, V8, P277
   Nimmerjahn A, 2005, SCIENCE, V308, P1314, DOI 10.1126/science.1110647
   Pahan K, 2001, J BIOL CHEM, V276, P7899, DOI 10.1074/jbc.M008262200
   Peng J, 2004, J BIOL CHEM, V279, P32626, DOI 10.1074/jbc.M404596200
   Peng J, 2007, J NEUROSCI, V27, P6914, DOI 10.1523/JNEUROSCI.1569-07.2007
   Peng J, 2009, FREE RADICAL BIO MED, V46, P312, DOI 10.1016/j.freeradbiomed.2008.10.045
   Perry VH, 2007, NAT REV IMMUNOL, V7, P161, DOI 10.1038/nri2015
   Peterson AL, 2008, NEUROTHERAPEUTICS, V5, P270, DOI 10.1016/j.nurt.2008.02.003
   Phillis JW, 2006, BRAIN RES REV, V52, P201, DOI 10.1016/j.brainresrev.2006.02.002
   Poligone B, 2001, J BIOL CHEM, V276, P38658, DOI 10.1074/jbc.M106599200
   Popovich PG, 2008, NAT REV NEUROSCI, V9, P481, DOI 10.1038/nrn2398
   Godoy MCP, 2008, BRAIN, V131, P1880, DOI 10.1093/brain/awn101
   Godoy MCP, 2010, J NEUROIMMUNOL, V222, P29, DOI 10.1016/j.jneuroim.2010.02.018
   Poynter ME, 2004, J IMMUNOL, V173, P7003, DOI 10.4049/jimmunol.173.11.7003
   Prasad K, 2009, EXP NEUROL, V215, P358, DOI 10.1016/j.expneurol.2008.11.003
   Purisai MG, 2007, NEUROBIOL DIS, V25, P392, DOI 10.1016/j.nbd.2006.10.008
   Qian L, 2006, J PHARMACOL EXP THER, V319, P44, DOI 10.1124/jpet.106.106351
   Quesada A, 2008, DEV NEUROBIOL, V68, P632, DOI 10.1002/dneu.20609
   Ramirez G, 2005, NEUROBIOL DIS, V19, P243, DOI 10.1016/j.nbd.2005.01.007
   Rayamajhi M, 2010, J EXP MED, V207, P327, DOI 10.1084/jem.20091746
   Reale M, 2009, BRAIN BEHAV IMMUN, V23, P55, DOI 10.1016/j.bbi.2008.07.003
   Ribiani E, 2007, Minerva Ginecol, V59, P151
   Rivest S, 2001, PSYCHONEUROENDOCRINO, V26, P761, DOI 10.1016/S0306-4530(01)00064-6
   Rock RB, 2005, GENES IMMUN, V6, P712, DOI 10.1038/sj.gene.6364246
   Rogers RJ, 2001, J BIOL CHEM, V276, P20419, DOI 10.1074/jbc.M008915200
   Rojo AI, 2007, EXP NEUROL, V208, P120, DOI 10.1016/j.expneurol.2007.07.022
   ROTHWELL NJ, 1995, TRENDS NEUROSCI, V18, P130, DOI 10.1016/0166-2236(95)93890-A
   Rousselet E, 2002, EXP NEUROL, V177, P183, DOI 10.1006/exnr.2002.7960
   Ryu HJ, 2010, NEUROSCIENCE, V170, P711, DOI 10.1016/j.neuroscience.2010.07.048
   Sanchez-Guajardo V, 2010, PLOS ONE, V5, DOI 10.1371/journal.pone.0008784
   Sanchez-Pernaute R, 2004, J NEUROINFLAMM, V1, DOI 10.1186/1742-2094-1-6
   Sarasombath Pichaya, 2002, Hawaii Med J, V61, P57
   Saura J, 2003, J NEUROCHEM, V85, P651, DOI 10.1046/j.1471-4159.2003.01676.x
   SCHOBITZ B, 1994, CRIT REV NEUROBIOL, V8, P263
   Schroder K, 2004, J LEUKOCYTE BIOL, V75, P163, DOI 10.1189/jlb.0603252
   Shen YQ, 2005, J NEUROIMMUNOL, V158, P14, DOI 10.1016/j.jneuroim.2004.06.011
   Shi J, 2010, J IMMUNOL, V184, P7207, DOI 10.4049/jimmunol.0903487
   Shie FS, 2005, GLIA, V52, P70, DOI 10.1002/glia.20220
   Singh M, 2008, MOL CELL BIOCHEM, V312, P131, DOI 10.1007/s11010-008-9728-2
   Sriram K, 2002, FASEB J, V16, P1474, DOI 10.1096/fj.02-0216fje
   Sriram K, 2006, FASEB J, V20, P670, DOI 10.1096/fj.05-5106com
   Suzumura A, 1998, BRAIN RES, V787, P139, DOI 10.1016/S0006-8993(97)01166-9
   TANCREDI V, 1992, NEUROSCI LETT, V146, P176, DOI 10.1016/0304-3940(92)90071-E
   Tansey MG, 2007, EXP NEUROL, V208, P1, DOI 10.1016/j.expneurol.2007.07.004
   Teismann P, 2003, P NATL ACAD SCI USA, V100, P5473, DOI 10.1073/pnas.0837397100
   Thiruchelvam M, 2003, EUR J NEUROSCI, V18, P589, DOI 10.1046/j.1460-9568.2003.02781.x
   Thiruchelvam M, 2000, J NEUROSCI, V20, P9207
   THORNBERRY NA, 1992, NATURE, V356, P768, DOI 10.1038/356768a0
   Ton TG, 2006, MOVEMENT DISORD, V21, P964, DOI 10.1002/mds.20856
   Tzeng Shun-Fen, 2005, Current Drug Targets - Inflammation and Allergy, V4, P335, DOI 10.2174/1568010054022051
   Uversky VN, 2002, NEUROTOXICOLOGY, V23, P527, DOI 10.1016/S0161-813X(02)00067-0
   Uversky VN, 2001, FEBS LETT, V500, P105, DOI 10.1016/S0014-5793(01)02597-2
   Vazquez-Claverie M, 2009, J NEUROPATH EXP NEUR, V68, P26, DOI 10.1097/NEN.0b013e3181919275
   Vichai V, 2005, INFLAMM RES, V54, P163, DOI 10.1007/s00011-004-1338-1
   Vijitruth R, 2006, J NEUROINFLAMM, V3, DOI 10.1186/1742-2094-3-6
   Vilar R, 2007, EUR J NEUROL, V14, P156, DOI 10.1111/j.1468-1331.2006.01590.x
   von Boyen GBT, 2006, INFLAMM BOWEL DIS, V12, P346, DOI 10.1097/01.MIB.0000219350.72483.44
   Wahner AD, 2007, NEUROLOGY, V69, P1836, DOI 10.1212/01.wnl.0000279519.99344.ad
   WANG J, 1994, J NEUROSURG, V80, P484, DOI 10.3171/jns.1994.80.3.0484
   Wang TG, 2005, FASEB J, V19, P1134, DOI 10.1096/fj.04-2457fje
   Wang YS, 2009, J NEUROL SCI, V285, P172, DOI 10.1016/j.jns.2009.06.034
   Weisskopf MG, 2010, ENVIRON HEALTH PERSP, V118, P1609, DOI 10.1289/ehp.1002339
   Widner B, 2002, J NEURAL TRANSM, V109, P181, DOI 10.1007/s007020200014
   Wilson EH, 2010, J CLIN INVEST, V120, P1368, DOI 10.1172/JCI41911
   Wolters EC, 2006, J NEURAL TRANSM-SUPP, P309
   Wu DC, 2003, P NATL ACAD SCI USA, V100, P6145, DOI 10.1073/pnas.0937239100
   Wu XF, 2005, ANTIOXID REDOX SIGN, V7, P654, DOI 10.1089/ars.2005.7.654
   Wuchert F, 2009, J NEUROIMMUNOL, V206, P44, DOI 10.1016/j.jneuroim.2008.10.010
   Wyss-Coray T, 2002, NEURON, V35, P419, DOI 10.1016/S0896-6273(02)00794-8
   YAMADA T, 1994, NEUROSCI LETT, V181, P61, DOI 10.1016/0304-3940(94)90560-6
   YAMADA T, 1995, NEUROSCI LETT, V195, P41, DOI 10.1016/0304-3940(95)11777-T
   Yang W, 2010, TOXICOL LETT, V199, P239, DOI 10.1016/j.toxlet.2010.09.005
   Zamanian-Daryoush M, 2000, MOL CELL BIOL, V20, P1278, DOI 10.1128/MCB.20.4.1278-1290.2000
NR 211
TC 45
Z9 46
U1 0
U2 5
PU HINDAWI LTD
PI LONDON
PA ADAM HOUSE, 3RD FLR, 1 FITZROY SQ, LONDON, W1T 5HF, ENGLAND
SN 2090-8083
EI 2042-0080
J9 PARKINSONS DIS-US
JI Parkinsons Dis.
PY 2011
VL 2011
AR 713517
DI 10.4061/2011/713517
PG 18
WC Clinical Neurology
WE Science Citation Index Expanded (SCI-EXPANDED)
SC Neurosciences & Neurology
GA V29RN
UT WOS:000208765600063
PM 21234362
OA Green Published, Green Submitted, gold
DA 2023-08-27
ER

PT J
AU Ho, KMT
   Li, AZL
   Yiu, MK
   Lee, KC
   Lui, VCH
   Fung, PCW
   Yiu, TF
   Tam, PKH
AF Ho, KMT
   Li, AZL
   Yiu, MK
   Lee, KC
   Lui, VCH
   Fung, PCW
   Yiu, TF
   Tam, PKH
TI Altered expression of aquaporin-2 in human explants with chronic renal
   allograft dysfunction
SO BJU INTERNATIONAL
LA English
DT Article
DE aquaporin; renal allograft dysfunction; transplant; explant
ID MEDULLARY COLLECTING DUCT; WATER CHANNEL; PATHOPHYSIOLOGY; PHYSIOLOGY;
   KIDNEY
AB To investigate the distribution of aquaporins, a recently discovered family of transmembrane water channels, in human renal explants, with specific reference to chronic renal allograft dysfunction (CRAD).
   Immunohistochemistry for aquaporin-1 and -2 was used in 11 explants, of which five had clinically and histologically confirmed CRAD. Controls were taken from the six explants unaffected by CRAD and from histologically normal areas of six kidneys excised for renal tumours.
   In the renal tumour control group, aquaporin-1 immunoreactivity was detected in the glomerular endothelium, Bowman's capsule, the proximal convoluted tubules and the thin limb of the loop of Henle, whereas immunoreactivity for aquaporin-2 was detected in the collecting ducts only. Of the explants without CRAD, where architecture was preserved, immunoreactivity for aquaporin-1 and -2 was the same as in the renal tumour controls. In the two explants with no CRAD and loss of collecting ducts, there was no aquaporin-2 immunoreactivity. In five explants with CRAD, immunoreactivity for aquaporin-2 was decreased or absent from the medulla to the cortex. The apparent decreased immunoreactivity of aquaporin-1 in this group was secondary to a decrease in the number of viable proximal tubules.
   There was less aquaporin-2 immunoreactivity in human renal explants diagnosed with CRAD, starting from the medullary region. In explants with no CRAD and viable collecting ducts, or in normal controls, aquaporin-2 immunoreactivity remained unchanged. Aquaporins might be useful as markers for CRAD.
C1 Univ Hong Kong, Queen Mary Hosp, Dept Surg, Hong Kong, Hong Kong, Peoples R China.
   Univ Hong Kong, Queen Mary Hosp, Dept Med, Hong Kong, Hong Kong, Peoples R China.
   Princess Margaret Hosp, Dept Surg, Hong Kong, Hong Kong, Peoples R China.
   Princess Margaret Hosp, Dept Pathol, Hong Kong, Hong Kong, Peoples R China.
C3 University of Hong Kong; University of Hong Kong
RP Ho, KMT (通讯作者)，GPO Box 2513, Hong Kong, Hong Kong, Peoples R China.
EM
RI /C-4359-2009; /C-4405-2009
OI /0000-0002-1758-8854; /0000-0001-6231-3035
CR Bedford JJ, 2003, J AM SOC NEPHROL, V14, P2581, DOI 10.1097/01.ASN.0000089566.28106.F6
   BRENNER BM, 1996, KIDNEY, V1, P1133
   Brown D, 2003, AM J PHYSIOL-RENAL, V284, pF893, DOI 10.1152/ajprenal.00387.2002
   Deen PMT, 2000, EUR J CELL BIOL, V79, P523, DOI 10.1078/0171-9335-00075
   Furness PN, 2001, TRANSPLANTATION, V71, pSS31
   Kwon TH, 2001, SEMIN NEPHROL, V21, P231, DOI 10.1053/snep.2001.21647
   Lim SW, 2004, AM J PHYSIOL-RENAL, V287, pF139, DOI 10.1152/ajprenal.00240.2003
   Matsumura Y, 1997, J AM SOC NEPHROL, V8, P861
   Nicholson ML, 1999, TRANSPLANTATION, V68, P236, DOI 10.1097/00007890-199907270-00013
   Nielsen S, 2002, PHYSIOL REV, V82, P205, DOI 10.1152/physrev.00024.2001
   NIELSEN S, 1993, J CELL BIOL, V120, P371, DOI 10.1083/jcb.120.2.371
   Nielsen S, 1999, J AM SOC NEPHROL, V10, P647
   Wong NLM, 2003, METABOLISM, V52, P290, DOI 10.1053/meta.2003.50047
   Wong NLM, 2002, METABOLISM, V51, P970, DOI 10.1053/meta.2002.34015
   Yasui M, 1996, AM J PHYSIOL-RENAL, V271, pF461, DOI 10.1152/ajprenal.1996.271.2.F461
NR 15
TC 4
Z9 4
U1 0
U2 0
PU WILEY
PI HOBOKEN
PA 111 RIVER ST, HOBOKEN 07030-5774, NJ USA
SN 1464-4096
EI 1464-410X
J9 BJU INT
JI BJU Int.
PD MAY
PY 2005
VL 95
IS 7
BP 1104
EP 1108
DI 10.1111/j.1464-410X.2005.005475.x
PG 5
WC Urology & Nephrology
WE Science Citation Index Expanded (SCI-EXPANDED)
SC Urology & Nephrology
GA 916PL
UT WOS:000228397500040
PM 15839941
OA Bronze
DA 2023-08-27
ER

PT J
AU Nevo, I
   Sagi-Assif, O
   Botzer, LE
   Amar, D
   Maman, S
   Kariv, N
   Leider-Trejo, LE
   Savelyeva, L
   Schwab, M
   Yron, I
   Witz, IP
AF Nevo, Ido
   Sagi-Assif, Orit
   Botzer, Liat Edry
   Amar, Dana
   Maman, Shelly
   Kariv, Naam
   Leider-Trejo, Leonor E.
   Savelyeva, Larissa
   Schwab, Manfred
   Yron, Ilana
   Witz, Isaac P.
TI Generation and characterization of novel local and metastatic human
   neuroblastoma variants
SO NEOPLASIA
LA English
DT Article
ID IN-VIVO SELECTION; TUMOR MICROENVIRONMENT; CANCER METASTASIS;
   CELL-LINES; NUDE-MICE; MATRIX METALLOPROTEINASES; PULMONARY METASTASES;
   DRUG SENSITIVITY; NEURO-BLASTOMA; EXPRESSION
AB Neuroblastoma ( NB) is the most commonly occurring solid tumor in children. The disease usually arises in the adrenal medulla, and it is characterized by a remarkable heterogeneity in its progression. Most NB patients with an advanced disease have massive bone marrow infiltration at diagnosis. Lung metastasis represents a widely disseminated stage and is typically considered to be a terminal event. Much like other malignancies, NB progression is a complex, multistep process. The expression, function, and significance of the various factors involved in NB progression must be studied in relevant in vivo and in vitro models. Currently, models consisting of metastatic and nonmetastatic cell variants of the same genetic background exist for several types of cancer; however, none exists for NB. In the present study, we describe the generation of a NB metastasis model. SH-SY5Y and MHH-NB-11 NB cells were inoculated orthotopically into the adrenal glands of athymic nude mice. Neuroblastoma cells metastasizing to the lungs were isolated from mice bearing adrenal tumors. Lung metastatic variants were generated by repeated cycles of in vivo passage. Characterization of these variants included cellular morphology and immunophenotyping in vitro, aggressiveness in vivo, and various biologic parameters in vitro. The NB metastatic variant in each model displayed unique properties, and both metastatic variants demonstrated a metastatic phenotype in vivo. These reproducible models of human NB metastasis will serve as an unlimited source of transcriptomic and proteomic material. Such models can facilitate future studies on NB metastasis and the identification of novel NB biomarkers and targets for therapy.
C1 [Witz, Isaac P.] Tel Aviv Univ, George S Wise Fac Life Sci, Dept Cell Res & Immunol, IL-69978 Tel Aviv, Israel.
C3 Tel Aviv University
RP Witz, IP (通讯作者)，Tel Aviv Univ, George S Wise Fac Life Sci, Dept Cell Res & Immunol, IL-69978 Tel Aviv, Israel.
EM
CR Ara T, 2006, CANCER METAST REV, V25, P645, DOI 10.1007/s10555-006-9028-9
   BECTON DL, 1988, CANCER RES, V48, P7189
   Beltinger C, 2001, INT J CANCER, V92, P313, DOI 10.1002/ijc.1210
   BIEDLER JL, 1973, CANCER RES, V33, P2643
   BIEDLER JL, 1978, CANCER RES, V38, P3751
   Bjorklund M, 2005, BBA-REV CANCER, V1755, P37, DOI 10.1016/j.bbcan.2005.03.001
   BLATT J, 1987, CANCER RES, V47, P1749
   Bogenmann E, 1996, INT J CANCER, V67, P379, DOI 10.1002/(SICI)1097-0215(19960729)67:3<379::AID-IJC12>3.0.CO;2-3
   Brahimi-Horn Christiane, 2006, Bull Cancer, V93, pE73
   Brodeur Garrett M., 1997, P761
   Brodeur GM, 2003, NAT REV CANCER, V3, P203, DOI 10.1038/nrc1014
   BRODEUR GM, 1991, CLIN PEDIATRIC ONCOL, P437
   Bruns Christiane J., 1999, Neoplasia (New York), V1, P50, DOI 10.1038/sj.neo.7900005
   Cheung N-K, 2005, NEUROBLASTOMA
   Cowie F, 1997, MED PEDIATR ONCOL, V28, P429, DOI 10.1002/(SICI)1096-911X(199706)28:6<429::AID-MPO7>3.3.CO;2-X
   Deryugina EI, 2006, CANCER METAST REV, V25, P9, DOI 10.1007/s10555-006-7886-9
   Donfrancesco A, 1995, ANTICANCER RES, V15, P2347
   DONFRANCESCO A, 1995, EUR J CANCER, V31A, P612, DOI 10.1016/0959-8049(95)00068-T
   Elstner A, 2007, CLIN EXP METASTAS, V24, P57, DOI 10.1007/s10585-007-9057-y
   Fidler IJ, 2003, NAT REV CANCER, V3, P453, DOI 10.1038/nrc1098
   FULDA S, 1995, EUR J CANCER, V31A, P616, DOI 10.1016/0959-8049(95)00055-N
   Geminder H, 2001, J IMMUNOL, V167, P4747, DOI 10.4049/jimmunol.167.8.4747
   Goldberg-Bittman L, 2005, CYTOKINE, V29, P105, DOI 10.1016/j.cyto.2004.10.003
   Gupta GP, 2006, CELL, V127, P679, DOI 10.1016/j.cell.2006.11.001
   HALACHMI E, 1989, CANCER RES, V49, P2383
   Henriksson KC, 2004, CLIN EXP METASTAS, V21, P563, DOI 10.1007/s10585-004-4091-5
   Hirota K, 2006, CRIT REV ONCOL HEMAT, V59, P15, DOI 10.1016/j.critrevonc.2005.12.003
   Holland JD, 2006, CANCER RES, V66, P4117, DOI 10.1158/0008-5472.CAN-05-1631
   Kammen BF, 2001, AM J ROENTGENOL, V176, P755, DOI 10.2214/ajr.176.3.1760755
   Keshelava N, 1998, CANCER RES, V58, P5396
   Khanna C, 2002, IN VIVO, V16, P77
   Li Y, 2004, J CANCER RES CLIN, V130, P460, DOI 10.1007/s00432-004-0564-9
   Matsui T, 1998, ORAL ONCOL, V34, P253
   Minn AJ, 2005, NATURE, V436, P518, DOI 10.1038/nature03799
   Mohamed MM, 2006, NAT REV CANCER, V6, P764, DOI 10.1038/nrc1949
   MORIKAWA K, 1988, CANCER RES, V48, P1943
   National Cancer Institute, SURV EP END RES DAT
   Nevo I, 2004, IMMUNOL LETT, V92, P163, DOI 10.1016/j.imlet.2003.10.019
   Onn A, 2003, CLIN CANCER RES, V9, P5532
   Overall CM, 2006, NAT REV CANCER, V6, P227, DOI 10.1038/nrc1821
   OZATO K, 1981, J IMMUNOL, V126, P317
   Palmesino E, 2006, IMMUNOBIOLOGY, V211, P377, DOI 10.1016/j.imbio.2005.12.003
   PARHAM P, 1979, J IMMUNOL, V123, P342
   PIETSCH T, 1988, ANTICANCER RES, V8, P1329
   RETTIG WJ, 1987, CANCER RES, V47, P1383
   Ribatti D, 2001, LIFE SCI, V68, P1161, DOI 10.1016/S0024-3205(00)01030-4
   ROSS RA, 1995, CELL GROWTH DIFFER, V6, P449
   SPENGLER BA, 1986, CANCER TREAT REP, V70, P959
   Subarsky P, 2003, CLIN EXP METASTAS, V20, P237, DOI 10.1023/A:1022939318102
   Sugiura Y, 1998, CANCER RES, V58, P2209
   Sullivan R, 2007, CANCER METAST REV, V26, P319, DOI 10.1007/s10555-007-9062-2
   TOWBIN R, 1982, AM J ROENTGENOL, V138, P75, DOI 10.2214/ajr.138.1.75
   TSURUO T, 1981, CANCER RES, V41, P3058
   Vasudevan SA, 2005, WORLD J SURG, V29, P317, DOI 10.1007/s00268-004-7820-7
   WANG GL, 1993, BLOOD, V82, P3610
   Warzynski MJ, 2002, CYTOMETRY, V50, P298, DOI 10.1002/cyto.10159
   Witz IP, 2008, CANCER RES, V68, P9, DOI 10.1158/0008-5472.CAN-07-2917
   Ziegler MM, 1997, CANCER-AM CANCER SOC, V79, P1757
NR 58
TC 22
Z9 24
U1 0
U2 0
PU NEOPLASIA PRESS
PI ANN ARBOR
PA 1150 W MEDICAL CENTER DR, MSRB III, RM 9303, ANN ARBOR, MI 48109-0648
   USA
SN 1522-8002
J9 NEOPLASIA
JI Neoplasia
PD AUG
PY 2008
VL 10
IS 8
BP 817
EP U67
DI 10.1593/neo.08402
PG 13
WC Oncology
WE Science Citation Index Expanded (SCI-EXPANDED)
SC Oncology
GA 336YA
UT WOS:000258401500007
PM 18683320
OA gold, Green Published
DA 2023-08-27
ER

PT J
AU HUMBLET, C
   RUDENSKY, AY
   KYEWSKI, B
AF HUMBLET, C
   RUDENSKY, AY
   KYEWSKI, B
TI PRESENTATION AND INTERCELLULAR TRANSFER OF SELF ANTIGEN WITHIN THE
   THYMIC MICROENVIRONMENT - EXPRESSION OF THE E-ALPHA PEPTIDE I-AB COMPLEX
   BY ISOLATED THYMIC STROMAL CELLS
SO INTERNATIONAL IMMUNOLOGY
LA English
DT Article
DE ANTIGEN PRESENTATION; ANTIGEN PROCESSING; SELF TOLERANCE; T CELL
   SELECTION
ID CORTICAL EPITHELIAL-CELLS; REACTIVE T-CELLS; POSITIVE SELECTION;
   MONOCLONAL-ANTIBODIES; DENDRITIC CELLS; CLONAL DELETION; INTRATHYMIC
   PRESENTATION; TOLERANCE INDUCTION; CD4+8+ THYMOCYTES; LYMPHOCYTES-T
AB Expression of a self peptide derived from the alpha chain of MHC class II (I-E(d)) in association with I-A(b) was studied in the murine thymic microenvironment. Previous work using the mAb Y-Ae which specifically recognizes the E alpha-I-A(b) complex had reported differential expression between the thymic medulla and the cortex of this peptide-MHC complex: MHC class II-positive stromal cells in the medulla were strongly positive, whereas this complex was barely detectable on cortical epithelial cells (cEpC) in situ. This difference in presentation of an abundant self peptide is intriguing, since the self protein from which this peptide is derived and the presenting MHC molecule are strongly expressed in both compartments, In this report we show by cell surface phenotype and functional assays that isolated cEpC express the E alpha-I-A(b) complex at significant although lower levels than medullary dendritic cells (DC), when examined ex vivo. These results support the notion that cEpC and bone marrow-derived stromal cells present a similar set of self peptide-MHC complexes in situ, In addition, we detect intercellular transfer in situ of the E alpha determinant from radioresistant stromal cells to thymic DC, a mechanism which may enhance the efficacy of tolerance induction by spreading self antigens within the thymic microenvironment.
C1 GERMAN CANC RES CTR,TUMOR IMMUNOL PROGRAMME,D-69120 HEIDELBERG,GERMANY.
   UNIV WASHINGTON,HOWARD HUGHES MED INST,DEPT IMMUNOL,SEATTLE,WA 98195.
C3 Helmholtz Association; German Cancer Research Center (DKFZ); Howard
   Hughes Medical Institute; University of Washington; University of
   Washington Seattle
FU NIAID NIH HHS [AI34206] Funding Source: Medline
CR ARDAVIN C, 1993, NATURE, V362, P761, DOI 10.1038/362761a0
   ASHTONRICKARDT PG, 1993, CELL, V73, P1041, DOI 10.1016/0092-8674(93)90281-T
   BIX M, 1992, NATURE, V359, P330, DOI 10.1038/359330a0
   BONOMO A, 1993, J EXP MED, V177, P1153, DOI 10.1084/jem.177.4.1153
   DEKRUYFF RH, 1986, J IMMUNOL, V137, P1109
   FREEMAN GJ, 1993, SCIENCE, V262, P907, DOI 10.1126/science.7694362
   GOODMAN T, 1989, J EXP MED, V170, P1569, DOI 10.1084/jem.170.5.1569
   GROSS JA, 1992, J IMMUNOL, V149, P380
   HATHCOCK KS, 1993, SCIENCE, V262, P905, DOI 10.1126/science.7694361
   HOGQUIST KA, 1993, J EXP MED, V177, P1469, DOI 10.1084/jem.177.5.1469
   HOGQUIST KA, 1994, CELL, V76, P17, DOI 10.1016/0092-8674(94)90169-4
   HUGO P, 1993, P NATL ACAD SCI USA, V90, P10335, DOI 10.1073/pnas.90.21.10335
   HUISKAMP R, 1985, J IMMUNOL, V134, P2170
   JACOBS H, 1990, EUR J IMMUNOL, V20, P2333, DOI 10.1002/eji.1830201024
   JANEWAY CA, 1984, J IMMUNOL, V132, P662
   JENKINSON EJ, 1992, J EXP MED, V176, P845, DOI 10.1084/jem.176.3.845
   JONES LA, 1993, INT IMMUNOL, V5, P503, DOI 10.1093/intimm/5.5.503
   KOCH N, 1982, EUR J IMMUNOL, V12, P909, DOI 10.1002/eji.1830121103
   KYEWSKI BA, 1986, J EXP MED, V163, P231, DOI 10.1084/jem.163.2.231
   KYEWSKI BA, 1984, NATURE, V308, P196, DOI 10.1038/308196a0
   KYEWSKI BA, 1987, EUR J IMMUNOL, V17, P961, DOI 10.1002/eji.1830170711
   LINSLEY PS, 1993, ANNU REV IMMUNOL, V11, P191, DOI 10.1146/annurev.immunol.11.1.191
   LORENZ RG, 1989, NATURE, V340, P557, DOI 10.1038/340557a0
   LORENZ RG, 1989, NATURE, V337, P560, DOI 10.1038/337560a0
   MACDONALD HR, 1988, J EXP MED, V167, P2005, DOI 10.1084/jem.167.6.2005
   MARRACK P, 1993, J EXP MED, V178, P2173, DOI 10.1084/jem.178.6.2173
   MARRACK P, 1987, SCIENCE, V238, P1073, DOI 10.1126/science.3317824
   MATHIS DJ, 1983, P NATL ACAD SCI-BIOL, V80, P273, DOI 10.1073/pnas.80.1.273
   MAZDA O, 1991, J EXP MED, V173, P539, DOI 10.1084/jem.173.3.539
   METLAY JP, 1990, J EXP MED, V171, P1753, DOI 10.1084/jem.171.5.1753
   MIZUOCHI T, 1992, J EXP MED, V175, P1601, DOI 10.1084/jem.175.6.1601
   MOLLER G, 1993, IMMUNOL REV, P135
   MULLER KP, 1993, EUR J IMMUNOL, V23, P3203, DOI 10.1002/eji.1830231224
   MURPHY DB, 1989, NATURE, V338, P765, DOI 10.1038/338765a0
   NELSON AJ, 1993, J IMMUNOL, V151, P2453
   NIKOLICZUGIC J, 1990, NATURE, V344, P65, DOI 10.1038/344065a0
   Oi V T, 1978, Curr Top Microbiol Immunol, V81, P115
   OZATO K, 1980, J IMMUNOL, V124, P533
   PAGE DM, 1993, J IMMUNOL, V151, P1868
   PAWLOWSKI T, 1993, NATURE, V364, P642, DOI 10.1038/364642a0
   PIRCHER H, 1993, EUR J IMMUNOL, V23, P669, DOI 10.1002/eji.1830230315
   PRIETO J, 1989, EUR J IMMUNOL, V19, P1551, DOI 10.1002/eji.1830190906
   PULLEN AM, 1988, NATURE, V335, P796, DOI 10.1038/335796a0
   PUNT JA, 1994, J EXP MED, V179, P709, DOI 10.1084/jem.179.2.709
   RANSOM J, 1991, CELL IMMUNOL, V134, P180, DOI 10.1016/0008-8749(91)90341-8
   ROUSE RV, 1988, J HISTOCHEM CYTOCHEM, V36, P1511, DOI 10.1177/36.12.2461413
   RUDENSKY AY, 1991, NATURE, V353, P622, DOI 10.1038/353622a0
   RUDENSKY AY, 1991, NATURE, V353, P660, DOI 10.1038/353660a0
   RUDENSKY AY, 1990, EUR J IMMUNOL, V20, P2235, DOI 10.1002/eji.1830201012
   SHA WC, 1990, P NATL ACAD SCI USA, V87, P6186, DOI 10.1073/pnas.87.16.6186
   SHAHINIAN A, 1993, SCIENCE, V261, P609, DOI 10.1126/science.7688139
   SHORTMAN K, 1987, J IMMUNOL, V138, P342
   SPEISER DE, 1989, J EXP MED, V170, P595, DOI 10.1084/jem.170.2.595
   SURH CD, 1992, J EXP MED, V176, P495, DOI 10.1084/jem.176.2.495
   TAN RS, 1992, J IMMUNOL, V149, P3217
   VANDENBERGHE P, 1992, INT IMMUNOL, V5, P317
   VUKMANOVIC S, 1992, NATURE, V359, P729, DOI 10.1038/359729a0
   WINSLOW GM, 1992, CELL, V71, P719, DOI 10.1016/0092-8674(92)90549-R
NR 58
TC 32
Z9 32
U1 0
U2 0
PU OXFORD UNIV PRESS UNITED KINGDOM
PI OXFORD
PA WALTON ST JOURNALS DEPT, OXFORD, ENGLAND OX2 6DP
SN 0953-8178
J9 INT IMMUNOL
JI Int. Immunol.
PD DEC
PY 1994
VL 6
IS 12
BP 1949
EP 1958
DI 10.1093/intimm/6.12.1949
PG 10
WC Immunology
WE Science Citation Index Expanded (SCI-EXPANDED)
SC Immunology
GA PY362
UT WOS:A1994PY36200016
PM 7696212
DA 2023-08-27
ER

PT J
AU Harada, K
   Arita, K
   Kurisu, K
   Tahara, H
AF Harada, K
   Arita, K
   Kurisu, K
   Tahara, H
TI Telomerase activity and the expression of telomerase components in
   pituitary adenoma with malignant transformation
SO SURGICAL NEUROLOGY
LA English
DT Article
DE hTERT; MIB1; pituitary carcinoma; telomerase
ID MULTIPLE INTRACRANIAL METASTASES; HEPATOCELLULAR CARCINOMAS;
   PROLACTINOMA; TUMOR; DISSEMINATION
AB BACKGROUND
   Telomerase activity responsible for cellular immortality may participate the development of human cancers. Telomerase is a multisubunit ribonucleoprotein composed of at least three components: hTERT, hTERC, and TEP1. This is the first report showing telomerase activity and telomerase component expression in pituitary adenoma with histological malignant transformation.
   DESCRIPTION
   A 16-year-old male with a prolactin-producing pituitary adenoma with metastasis is presented. The patient underwent three partial resections of an intra- and suprasellar lesion over a 2-year period and received focal irradiation. Eight years after the first admission, a metastatic lesion to the subarachnoid space around the medulla oblongata was detected and the lesion was resected as the fourth operation. Furthermore, the suprasellar lesion showed regrowth and partial resection was performed as the fifth operation. The last two specimens were diagnosed as pituitary carcinoma. Radiotherapy with gamma knife was performed for the residual suprasellar lesion and a new lesion in the left temporal lobe after the fifth operation. Telomerase activity was examined by TRAP/TRAP-HPA methods, qualitatively and quantitatively. Telomere length was examined by Southern blot analysis, and the expression of telomerase components (hTERT, hTERC, and TEP1) was examined by reverse transcriptase-polymerase chain reaction (RT-PCR). The MIBI index, telomerase activity, and hTERT expression increased according to histologic malignancy chronologically in this patient. None of the specimens showed immunoreactivity for p53, EGFR, or bc12. No telomerase activity was detected in pituitary adenomas without malignant transformation, other benign brain tumors, or normal brain tissues.
   CONCLUSION
   We report a patient with pituitary adenoma transforming to carcinoma. The tumor cells acquired immortality and revealed malignant transformation during the course of the disease, that was proved by an increase of telomerase activity and hTERT expression. (C) 2000 by Elsevier Science Inc.
C1 Hiroshima Univ, Sch Med, Dept Neurosurg, Minami Ku, Hiroshima 7348551, Japan.
   Hiroshima Univ, Sch Med, Dept Cellular & Mol Biol, Minami Ku, Hiroshima 7348551, Japan.
C3 Hiroshima University; Hiroshima University
RP Harada, K (通讯作者)，Hiroshima Univ, Sch Med, Dept Neurosurg, Minami Ku, Kasumi 1-2-3, Hiroshima 7348551, Japan.
CR ALEXANDER JM, 1996, PITUITARY ADENOMAS, P15
   ASAI A, 1988, NEUROSURGERY, V22, P1091, DOI 10.1227/00006123-198806010-00023
   ASSIES J, 1991, J ENDOCRINOL INVE S4, V14, P4
   BEREZIN M, 1992, ACTA ENDOCRINOL-COP, V127, P476, DOI 10.1530/acta.0.1270476
   Bracke ME, 1996, CURR TOP MICROBIOL, V213, P123
   COHEN DL, 1983, CLIN ENDOCRINOL, V18, P259, DOI 10.1111/j.1365-2265.1983.tb03210.x
   DABRERA VSE, 1973, J PATHOL, V109, P335, DOI 10.1002/path.1711090408
   FEIRING EH, 1972, J NEUROSURG, V36, P781
   GASSER RW, 1985, CLIN ENDOCRINOL, V22, P17, DOI 10.1111/j.1365-2265.1985.tb01060.x
   HASHIMOTO N, 1986, J NEUROSURG, V64, P140, DOI 10.3171/jns.1986.64.1.0140
   KIM NW, 1994, SCIENCE, V266, P2011, DOI 10.1126/science.7605428
   KUROKI M, 1987, SURG NEUROL, V28, P71, DOI 10.1016/0090-3019(87)90210-2
   LAUREN AL, 1995, LANCET, V346, P1267
   MADONICK MJ, 1963, NEUROLOGY, V13, P836, DOI 10.1212/WNL.13.10.836
   MARTIN NA, 1981, J NEUROSURG, V55, P615, DOI 10.3171/jns.1981.55.4.0615
   Mizoue T, 1997, ACTA NEUROCHIR, V139, P426, DOI 10.1007/BF01808879
   MUHR C, 1988, NEUROSURGERY, V22, P374, DOI 10.1227/00006123-198802000-00017
   Nakayama JI, 1998, NAT GENET, V18, P65, DOI 10.1038/ng0198-65
   OGLIVY KM, 1973, J NEUROL NEUROSUR PS, V36, P199
   PLANGGER CA, 1985, NEUROCHIRURGIA, V28, P235
   SANG HU, 1984, HUM PATHOL, V15, P94
   SCHEITHAUER BW, 1985, CANCER, V55, P598, DOI 10.1002/1097-0142(19850201)55:3<598::AID-CNCR2820550320>3.0.CO;2-S
   SOLITARE GB, 1967, J NEUROSURG, V26, P624, DOI 10.3171/jns.1967.26.6.0624
   TAHARA H, 1995, CANCER RES, V55, P2734
   THAPAR K, 1995, 63 ANN M AM ASS NEUR, P170
   WILLIAM AST, 1994, J NEUROSURG, V81, P624
NR 26
TC 27
Z9 31
U1 0
U2 0
PU ELSEVIER SCIENCE INC
PI NEW YORK
PA 360 PARK AVE SOUTH, NEW YORK, NY 10010-1710 USA
SN 0090-3019
J9 SURG NEUROL
JI Surg. Neurol.
PD MAR
PY 2000
VL 53
IS 3
BP 267
EP 274
DI 10.1016/S0090-3019(00)00181-6
PG 8
WC Clinical Neurology; Surgery
WE Science Citation Index Expanded (SCI-EXPANDED)
SC Neurosciences & Neurology; Surgery
GA 308ZY
UT WOS:000086743800028
PM 10773260
DA 2023-08-27
ER

PT J
AU Unger, N
   Ueberberg, B
   Schulz, S
   Saeger, W
   Mann, K
   Petersenn, S
AF Unger, N.
   Ueberberg, B.
   Schulz, S.
   Saeger, W.
   Mann, K.
   Petersenn, S.
TI Differential Expression of Somatostatin Receptor Subtype 1-5 Proteins in
   Numerous Human Normal Tissues
SO EXPERIMENTAL AND CLINICAL ENDOCRINOLOGY & DIABETES
LA English
DT Article
DE sst1-5; physiology; regulation; immunohistochemistry
ID HUMAN BREAST-CANCER; MESSENGER-RNA EXPRESSION; HUMAN KIDNEY;
   HEPATOCELLULAR CARCINOMAS; NEUROENDOCRINE TUMORS; THYROID-CARCINOMA;
   CELL-LINES; IN-VITRO; SST(2A); OCTREOTIDE
AB 5 human somatostatin receptor subtypes (sst1-5) mediate the antisecretory and antiproliferative effects of somatostatin.
   We examined somatostatin receptor protein expression in 28 human normal tissues. Immunostaining was performed with specific polyclonal antibodies for sst1-5. Staining pattern and distribution of ssts were evaluated.
   Anterior pituitary was positively stained for all 5 ssts. Pancreatic islets exhibited a positive staining for sst1-3 and sst5. Adrenal cortex expressed all 5 receptor subtypes, while the medulla was positive for sst3 and sst5 only. The thyroid expressed sst5 only, limited to single interfollicular cells. All 5 ssts were detected in the ovary, limited to luteinized granulosa cells of the corpus luteum. In the testis, sst2A was detected in the basal parts of the tubules, while sst5 was positively stained in the luminal parts. Sst1 was found in Leydig cells only. Stomach was positively stained for all 5 ssts. Investigation of the kidney revealed differential expression, with sst2A being found in the glomerules. The tubules expressed all 5 ssts. In the bone marrow cells of the granulocytopoiesis expressed sst2A only. The cerebellum expressed sst5 in a certain cell type, representing presumably Purkinje cells, while sst2A was stained in intercellular fibers.
   The expression of somatostatin receptor sub-types in a variety of human normal tissues may indicate a physiological role in these organs. Somatostatin analogues may off er new diagnostic and therapeutic implications for tumours related to these tissues. However, treatment of defined tumours with somatostatin analogues may also alter other normal tissues.
C1 [Unger, N.; Ueberberg, B.; Mann, K.; Petersenn, S.] Univ Duisburg Essen, Dept Endocrinol, Essen, Germany.
   [Unger, N.; Ueberberg, B.; Mann, K.; Petersenn, S.] Univ Duisburg Essen, Div Lab Res, Essen, Germany.
   [Schulz, S.] Univ Jena, Inst Pharmacol & Toxicol, D-6900 Jena, Germany.
   [Saeger, W.] Marien Hosp, Inst Pathol, Hamburg, Germany.
C3 University of Duisburg Essen; University of Duisburg Essen; Friedrich
   Schiller University of Jena; St. Marien Hospital
RP Petersenn, S (通讯作者)，ENDOC Ctr Endocrine Tumors, Altonaer Str 59, D-20357 Hamburg, Germany.
EM
RI Petersenn, Stephan/AHC-4344-2022; Schulz, Stefan/A-6928-2017
OI Schulz, Stefan/0000-0002-5997-8885; Petersenn,
   Stephan/0000-0002-0618-6147
CR Ain KB, 1997, J CLIN ENDOCR METAB, V82, P1857, DOI 10.1210/jcem.82.6.4013
   Bajetta E, 2002, CANCER, V94, P299, DOI 10.1002/cncr.10239
   Balster DA, 2001, AM J PHYSIOL-RENAL, V280, pF457, DOI 10.1152/ajprenal.2001.280.3.F457
   Baou N, 2000, CARCINOGENESIS, V21, P805, DOI 10.1093/carcin/21.4.805
   Bhandari S, 2008, J HISTOCHEM CYTOCHEM, V56, P733, DOI 10.1369/jhc.2008.950998
   Blaker M, 2004, J HEPATOL, V41, P112, DOI 10.1016/j.jhep.2004.03.018
   Boscaro M, 2009, J CLIN ENDOCR METAB, V94, P115, DOI 10.1210/jc.2008-1008
   Bruns C, 2002, EUR J ENDOCRINOL, V146, P707, DOI 10.1530/eje.0.1460707
   Bruns C, 1996, METABOLISM, V45, P17, DOI 10.1016/S0026-0495(96)90072-4
   Colao A, 2000, J CLIN ENDOCR METAB, V85, P3132, DOI 10.1210/jc.85.9.3132
   Fischer T, 2008, J CLIN ENDOCRINOL ME
   Fombonne Joanna, 2003, Reprod Biol Endocrinol, V1, P19, DOI 10.1186/1477-7827-1-19
   Freda PU, 2002, J CLIN ENDOCR METAB, V87, P3013, DOI 10.1210/jc.87.7.3013
   Freudenberg LS, 2008, NUKLEARMED-NUCL MED, V47, P127, DOI 10.3413/nukmed-0119
   Goddard I, 2001, BIOL REPROD, V65, P1732, DOI 10.1095/biolreprod65.6.1732
   Hofland LJ, 2005, EUR J ENDOCRINOL, V152, P645, DOI 10.1530/eje.1.01876
   Korner M, 2005, AM J SURG PATHOL, V29, P1642
   Kreienkamp HJ, 1999, FEBS LETT, V462, P464, DOI 10.1016/S0014-5793(99)01582-3
   Kulaksiz H, 2002, GUT, V50, P52, DOI 10.1136/gut.50.1.52
   Kumar U, 2005, BREAST CANCER RES TR, V92, P175, DOI 10.1007/s10549-005-2414-0
   Kumar U, 1999, DIABETES, V48, P77, DOI 10.2337/diabetes.48.1.77
   LeRomancer M, 1996, LIFE SCI, V58, P1091, DOI 10.1016/0024-3205(96)00063-X
   Lupp A, 2011, NEUROENDOCRINOLOGY, V94, P255, DOI 10.1159/000329876
   Mato E, 1998, J CLIN ENDOCR METAB, V83, P2417, DOI 10.1210/jc.83.7.2417
   Mercado M, 2007, CLIN ENDOCRINOL, V66, P859, DOI 10.1111/j.1365-2265.2007.02825.x
   Mundschenk J, 2003, J CLIN ENDOCR METAB, V88, P5150, DOI 10.1210/jc.2003-030262
   Oberg K, 2004, ANN ONCOL, V15, P966, DOI 10.1093/annonc/mdh216
   OCARROLL AM, 1995, ENDOCRINOLOGY, V136, P5224, DOI 10.1210/en.136.11.5224
   Orlando C, 2004, ENDOCR-RELAT CANCER, V11, P323, DOI 10.1677/erc.0.0110323
   Panetta Rosemarie, 1995, Life Sciences, V56, P333
   Park S, 2003, NEUROENDOCRINOLOGY, V78, P163, DOI 10.1159/000072798
   Patel YC, 1999, FRONT NEUROENDOCRIN, V20, P157, DOI 10.1006/frne.1999.0183
   Petersenn S, 2010, J CLIN ENDOCR METAB, V95, P2781, DOI 10.1210/jc.2009-2272
   Pfeiffer M, 2001, J BIOL CHEM, V276, P14027, DOI 10.1074/jbc.M006084200
   Pfeiffer M, 2002, J BIOL CHEM, V277, P19762, DOI 10.1074/jbc.M110373200
   PREVOST G, 1991, ENDOCRINOLOGY, V129, P323, DOI 10.1210/endo-129-1-323
   Raderer M, 2000, INT J ONCOL, V16, P1197
   REUBI JC, 1993, J CLIN ENDOCR METAB, V77, P1323, DOI 10.1210/jc.77.5.1323
   Reynaert H, 2004, GUT, V53, P1180, DOI 10.1136/gut.2003.036053
   Rohrer SP, 1998, SCIENCE, V282, P737, DOI 10.1126/science.282.5389.737
   Rolleman EJ, 2007, EUR J NUCL MED MOL I, V34, P1854, DOI 10.1007/s00259-007-0457-7
   ROSSOWSKI WJ, 1994, BIOCHEM BIOPH RES CO, V205, P341, DOI 10.1006/bbrc.1994.2670
   Ruan WF, 2006, MOL ENDOCRINOL, V20, P426, DOI 10.1210/me.2005-0283
   Schopohl J, 2011, EXP CLIN ENDOCR DIAB, V119, P156, DOI 10.1055/s-0030-1267244
   Schulz S, 2000, J PHYSIOLOGY-PARIS, V94, P259, DOI 10.1016/S0928-4257(00)00212-6
   Schulz S, 1998, NEUROSCI LETT, V257, P37, DOI 10.1016/S0304-3940(98)00803-9
   Schulz S, 1998, CLIN CANCER RES, V4, P2047
   Schulz S, 2000, CLIN CANCER RES, V6, P1865
   Schulz S, 2002, BREAST CANCER RES TR, V72, P221, DOI 10.1023/A:1014972520302
   Schulz S, 2002, GYNECOL ONCOL, V84, P235, DOI 10.1006/gyno.2001.6468
   Smith WHT, 2005, J ENDOCRINOL, V187, P379, DOI 10.1677/joe.1.06082
   Taniyama Y, 2005, ENDOCR J, V52, P605, DOI 10.1507/endocrj.52.605
   Unger N, 2008, CLIN ENDOCRINOL, V68, P850, DOI 10.1111/j.1365-2265.2007.03124.x
   Varecza Z, 2009, J HISTOCHEM CYTOCHEM, V57, P1127, DOI 10.1369/jhc.2009.953919
   Verhoef C, 2008, DIGEST SURG, V25, P21, DOI 10.1159/000117819
   WECKBECKER G, 1992, CANCER RES, V52, P4973
   YAMADA Y, 1992, P NATL ACAD SCI USA, V89, P251, DOI 10.1073/pnas.89.1.251
NR 57
TC 36
Z9 38
U1 0
U2 11
PU JOHANN AMBROSIUS BARTH VERLAG MEDIZINVERLAGE HEIDELBERG GMBH
PI STUTTGART
PA RUEDIGERSTR 14, D-70469 STUTTGART, GERMANY
SN 0947-7349
J9 EXP CLIN ENDOCR DIAB
JI Exp. Clin. Endocrinol. Diabet.
PD SEP
PY 2012
VL 120
IS 8
BP 482
EP 489
DI 10.1055/s-0032-1314859
PG 8
WC Endocrinology & Metabolism
WE Science Citation Index Expanded (SCI-EXPANDED)
SC Endocrinology & Metabolism
GA 012SH
UT WOS:000309256300009
PM 22976314
DA 2023-08-27
ER

PT J
AU Hone, SW
   Commins, DJ
   Rames, P
   Chen, JM
   Rowed, D
   McLean, A
   Nedzelski, JM
AF Hone, SW
   Commins, DJ
   Rames, P
   Chen, JM
   Rowed, D
   McLean, A
   Nedzelski, JM
TI Prognostic factors in intraoperative facial nerve monitoring for
   acoustic neuroma
SO JOURNAL OF OTOLARYNGOLOGY
LA English
DT Article; Proceedings Paper
CT 50th Annual Meeting of the
   Canadian-Society-of-Otolaryngology-Head-and-Neck-Surgery
CY JUN   18, 1996
CL LONDON, CANADA
SP Canadian Soc Otolaryngol Head & Neck Surg
DE facial nerve function; intraoperative facial nerve monitoring;
   vestibular schwannoma
ID CEREBELLOPONTINE ANGLE SURGERY; TUMOR SURGERY; ELECTRONEUROGRAPHY;
   PREDICTION; REMOVAL
AB Objectives: To determine the predictive value of intraoperative threshold stimulus for facial nerve outcome and the prevalence and prognostic value of persistent trains of activity and frequent spontaneous or mechanically induced contractions during acoustic neuroma surgery.
   Study design: Prospective recording and subsequent review of facial nerve activity.
   Setting: Tertiary referral centre.
   Patients and Methods: Consecutive patients undergoing acoustic neuroma surgery. Intraoperative facial nerve activity was digitised and stored on a personal computer for future analysis. Operative events were flagged. Recordings were available in 27 patients.
   Main Outcome Measures: Frequent mechanically induced contractions (< 20), prolonged trains of facial nerve activity (total time > 199 seconds), and facial nerve brainstem stimulus threshold were correlated with facial nerve outcome.
   Results: A brainstem stimulus threshold > 0.1 mA was significantly associated with intermediate or poor facial nerve function (House-Brackmann grade > 2) on the sixth postoperative day, at 1 month and 6 months. Patients with normal or near-normal facial function on the first day and a threshold of > 0.1 mA were significantly more likely to develop a delayed facial nerve palsy. Frequent contractions were noted in 74% of patients and persistent train activity in 59%. Neither was predictive of facial nerve outcome.
   Conclusions: An elevated brainstem threshold is helpful in predicting delayed facial nerve palsy and suboptimal facial nerve outcome. Persistent train activity and frequent contractions, do not have major prognostic significance.
C1 Sunnybrook Hlth Sci Ctr, Dept Otolaryngol HNS, Toronto, ON M4N 3M5, Canada.
C3 University of Toronto; Sunnybrook Health Science Center; Sunnybrook
   Research Institute
RP Nedzelski, JM (通讯作者)，Sunnybrook Hlth Sci Ctr, Dept Otolaryngol, 2075 Bayview Ave,Suite A 27, Toronto, ON M4N 3M6, Canada.
CR BECK DL, 1991, OTOLARYNG HEAD NECK, V104, P780, DOI 10.1177/019459989110400602
   ESSES BA, 1994, AM J OTOL, V15, P810
   HAMMERSCHLAG PE, 1990, OTOLARYNG HEAD NECK, V103, P681, DOI 10.1177/019459989010300502
   HOUSE JW, 1985, OTOLARYNG HEAD NECK, V93, P146, DOI 10.1177/019459988509300202
   KARTUSH JM, 1992, OTOLARYNG CLIN N AM, V25, P623
   KARTUSH JM, 1989, OTOLARYNG HEAD NECK, V101, P496, DOI 10.1177/019459988910100416
   KIRKPATRICK PJ, 1993, BRIT J NEUROSURG, V7, P657, DOI 10.3109/02688699308995095
   KWARTLER JA, 1991, OTOLARYNG HEAD NECK, V104, P814, DOI 10.1177/019459989110400608
   Moffat DA, 1996, AM J OTOL, V17, P643
   MOFFAT DA, 1989, J LARYNGOL OTOL, V103, P169, DOI 10.1017/S0022215100108357
   NIPARKO JK, 1989, AM J OTOL, V10, P55
   Nissen AJ, 1997, AM J OTOL, V18, P249
   PRASAD S, 1993, AM J OTOL, V14, P326
   PRASAD S, 1993, AM J OTOL, V14, P330
   SILVERSTEIN H, 1994, LARYNGOSCOPE, V104, P539, DOI 10.1002/lary.5541040506
   SILVERSTEIN H, 1991, OTOLARYNG CLIN N AM, V24, P709
   WIEGAND DA, 1989, LARYNGOSCOPE, V99, P179
   YINGLING CD, 1992, OTOLARYNG CLIN N AM, V25, P413
NR 18
TC 28
Z9 28
U1 0
U2 2
PU DECKER PERIODICALS INC
PI HAMILTON
PA 4 HUGHSON STREET SOUTH PO BOX 620, LCD 1, HAMILTON, ONTARIO L8N 3K7,
   CANADA
SN 0381-6605
J9 J OTOLARYNGOL
JI J. Otolaryngol.
PD DEC
PY 1997
VL 26
IS 6
BP 374
EP 378
PG 5
WC Otorhinolaryngology
WE Conference Proceedings Citation Index - Science (CPCI-S); Science Citation Index Expanded (SCI-EXPANDED)
SC Otorhinolaryngology
GA YN386
UT WOS:000071163400005
PM 9438935
DA 2023-08-27
ER

PT J
AU Chu, CK
   Tseng, HM
   Young, YH
AF Chu, Chih-Kai
   Tseng, Ham-Min
   Young, Yi-Ho
TI Clinical presentation of posterior fossa epidermoid cysts
SO EUROPEAN ARCHIVES OF OTO-RHINO-LARYNGOLOGY
LA English
DT Article
DE epidermoid cyst; vestibular evoked myogenic potential; cerebellum;
   brainstem
ID EVOKED MYOGENIC POTENTIALS; CEREBELLOPONTINE ANGLE; TUMORS
AB The aim of this study was to investigate the audiovestibular deficits in those with posterior fossa epidermoid cyst including cerebellopontine angle in two patients and cerebellum in one patient. Prior to operation, all three patients showed bilateral gaze nystagmus. Audiometry revealed mild hearing loss in two patients, and caloric test displayed canal paresis in two patients. After operation, subsidence of gaze nystagmus and recovery of caloric responses were disclosed in all patients. Two patients had both hearing and vestibular evoked myogenic potential (VEMP) results recovered to normal. In contrast, the only one who underwent craniotomy twice displayed bilateral mild hearing loss and delayed VEMPs, possibly due to operation sequela. Hence, bilateral gaze nystagmus may present as an initial sign for posterior fossa epidermoid cyst. Audiovestibular deficits in cases of epidermoid cyst are attributable to compression neuropathy, which may resolve and return to normal responses after surgery.
C1 Natl Taiwan Univ Hosp, Dept Otolaryngol, Taipei, Taiwan.
   Natl Taiwan Univ Hosp, Dept Surg, Taipei, Taiwan.
   Natl Taiwan Univ, Coll Med, Dept Surg, Taipei 10018, Taiwan.
   Lo Tung Poh Ai Hosp, Dept Otolaryngol, Ilan, Taiwan.
C3 National Taiwan University; National Taiwan University Hospital;
   National Taiwan University; National Taiwan University Hospital;
   National Taiwan University
RP Young, YH (通讯作者)，Natl Taiwan Univ Hosp, Dept Otolaryngol, 1 Chang Te St, Taipei, Taiwan.
EM
CR AUGER RG, 1989, NEUROLOGY, V39, P577, DOI 10.1212/WNL.39.4.577
   BERGER MS, 1985, J NEUROSURG, V62, P214, DOI 10.3171/jns.1985.62.2.0214
   Chen CH, 2003, LARYNGOSCOPE, V113, P990, DOI 10.1097/00005537-200306000-00014
   Chen CW, 2002, LARYNGOSCOPE, V112, P267, DOI 10.1097/00005537-200202000-00013
   COLEBATCH JG, 1994, J NEUROL NEUROSUR PS, V57, P190, DOI 10.1136/jnnp.57.2.190
   Dechambre S, 1999, NEURORADIOLOGY, V41, P829, DOI 10.1007/s002340050850
   DESOUZA CE, 1989, J NEUROL NEUROSUR PS, V52, P986, DOI 10.1136/jnnp.52.8.986
   Dutt SN, 2002, OTOL NEUROTOL, V23, P84, DOI 10.1097/00129492-200201000-00019
   EEKHOF JL, 1985, SURG NEUROL, V23, P1189
   Hasegawa T, 1989, Neurol Med Chir (Tokyo), V29, P614, DOI 10.2176/nmc.29.614
   LALWANI AK, 1992, OTOLARYNG CLIN N AM, V25, P707
   NEDZELSKI JM, 1983, LARYNGOSCOPE, V93, P1251
   YAMAKAWA K, 1989, NEUROSURGERY, V24, P568, DOI 10.1227/00006123-198904000-00013
   Yang TL, 2004, INT J RADIAT ONCOL, V60, P295, DOI 10.1016/j.ijrobp.2004.02.033
NR 14
TC 10
Z9 11
U1 0
U2 0
PU SPRINGER
PI NEW YORK
PA 233 SPRING ST, NEW YORK, NY 10013 USA
SN 0937-4477
EI 1434-4726
J9 EUR ARCH OTO-RHINO-L
JI Eur. Arch. Oto-Rhino-Laryn.
PD JUN
PY 2006
VL 263
IS 6
BP 548
EP 551
DI 10.1007/s00405-005-0005-7
PG 4
WC Otorhinolaryngology
WE Science Citation Index Expanded (SCI-EXPANDED)
SC Otorhinolaryngology
GA 051OQ
UT WOS:000238170800010
PM 16477440
DA 2023-08-27
ER

PT J
AU Wei, Y
   Niu, Y
   Zhao, ZL
   Cao, XJ
   Peng, LL
   Li, Y
   Yu, MA
AF Wei, Ying
   Niu, Yun
   Zhao, Zhen-long
   Cao, Xiao-jing
   Peng, Li-li
   Li, Yan
   Yu, Ming-an
TI Effectiveness of Lymphatic Contrast Enhanced Ultrasound in the diagnosis
   of Cervical Lymph node metastasis from papillary thyroid carcinoma
SO SCIENTIFIC REPORTS
LA English
DT Article
ID CENTRAL NECK DISSECTION; LOCOREGIONAL RECURRENCE; PREOPERATIVE
   DIAGNOSIS; CANCER PATIENTS; ULTRASONOGRAPHY; METAANALYSIS; MANAGEMENT;
   MELANOMA; THERAPY
AB Cervical lymph node metastasis (CLNM) is common in patients with papillary thyroid carcinoma (PTC), which is responsible for tumor staging and surgical strategy. The accurate preoperative identification of CLNM is essential. In this study, twenty consecutive patients with PTC received a parenchyma injection of Sonazoid followed by contrast enhanced ultrasound (CEUS) to identify CLNM. The specific lymphatic CEUS (LCEUS) signs for diagnosing CLNM were summarized, which were further compared with the resected specimens to get the pathological basis. After the injection of contrast agent, lymphatic vessel and lymph node (LN) could be exclusively displayed as hyperperfusion on LCEUS. The dynamic perfusion process of contrast agent in CLNM over time can be clearly visualized. Perfusion defect and interruption of bright ring were the two characteristic LCEUS signs in diagnosing CLNM. After comparing with pathology, perfusion defect was correlated to the metastatic foci in medulla and interruption of bright ring was correlated to the tumor seeding in marginal sinus (all p values < 0.001). The diagnostic efficacies of these two signs were high (perfusion defect vs. interruption of bright ring: AUC, 0.899, 95% CI 0.752-1.000 vs. 0.904, 0.803-1.000). LCEUS has advantages in identifying CLNM from PTC. The typical LCEUS signs of CLNM correlated with pathology.
C1 [Wei, Ying; Zhao, Zhen-long; Cao, Xiao-jing; Peng, Li-li; Li, Yan; Yu, Ming-an] China Japan Friendship Hosp, Dept Intervent Med, 2 Ying-hua Yuan St, Beijing 100029, Peoples R China.
   [Niu, Yun] China Japan Friendship Hosp, Dept Pathol, 2 Ying Hua Yuan St, Beijing 100029, Peoples R China.
C3 China-Japan Friendship Hospital; China-Japan Friendship Hospital
RP Yu, MA (通讯作者)，China Japan Friendship Hosp, Dept Intervent Med, 2 Ying-hua Yuan St, Beijing 100029, Peoples R China.
EM
FU Beijing Municipal Science and Beijing University of Chemical
   Technology-ChinaJapan Friendship Hospital Biomedical Transformation
   Joint Fund Project [PYBZ1804]; Beijing Municipal Science & Technology
   Commission [Z181100001718135]
FX This study was funded by Beijing Municipal Science and Beijing
   University of Chemical Technology-ChinaJapan Friendship Hospital
   Biomedical Transformation Joint Fund Project (PYBZ1804) and Beijing
   Municipal Science & Technology Commission (grant Z181100001718135).
CR Adam MA, 2015, J CLIN ONCOL, V33, P2370, DOI 10.1200/JCO.2014.59.8391
   Ahuja AT, 2005, AM J ROENTGENOL, V184, P1691, DOI 10.2214/ajr.184.5.01841691
   Arturi F, 1997, J CLIN ENDOCR METAB, V82, P1638, DOI 10.1210/jc.82.5.1638
   Beasley NJP, 2002, CANCER RES, V62, P1315
   Curry JM, 2009, ANN OTO RHINOL LARYN, V118, P645, DOI 10.1177/000348940911800908
   Goldberg BB, 2011, J ULTRAS MED, V30, P441, DOI 10.7863/jum.2011.30.4.441
   Goldberg BB, 2004, RADIOLOGY, V230, P727, DOI 10.1148/radiol.2303021440
   Haugen BR, 2016, THYROID, V26, P1, DOI 10.1089/thy.2015.0020
   Hong YR, 2017, ULTRASOUND MED BIOL, V43, P2567, DOI 10.1016/j.ultrasmedbio.2017.07.010
   Kim SK, 2016, J AM COLL SURGEONS, V222, P853, DOI 10.1016/j.jamcollsurg.2016.02.001
   Kim SY, 2018, WORLD J SURG, V42, P123, DOI 10.1007/s00268-017-4158-5
   Lang BHH, 2016, J SURG ONCOL, V113, P526, DOI 10.1002/jso.24180
   Lang BHH, 2013, THYROID, V23, P1087, DOI 10.1089/thy.2012.0608
   Lang BHH, 2012, ANN SURG ONCOL, V19, P3472, DOI 10.1245/s10434-012-2385-4
   Leboulleux S, 2007, J CLIN ENDOCR METAB, V92, P3590, DOI 10.1210/jc.2007-0444
   Liu SR, 2020, ULTRASOUND MED BIOL, V46, P582, DOI 10.1016/j.ultrasmedbio.2019.12.004
   Mazzaferri EL, 2001, J CLIN ENDOCR METAB, V86, P1447, DOI 10.1210/jc.86.4.1447
   Polistena A, 2015, INT J SURG, V21, P128, DOI 10.1016/j.ijsu.2015.07.698
   Sapuppo G, 2017, J ENDOCRINOL INVEST, V40, P1355, DOI 10.1007/s40618-017-0714-y
   Sherman SI, 2003, LANCET, V361, P501, DOI 10.1016/S0140-6736(03)12488-9
   Shimazu K, 2017, CANCER MED-US, V6, P1915, DOI 10.1002/cam4.1142
   Tuttle RM, 2014, ENDOCRINE, V46, P363, DOI 10.1007/s12020-014-0213-2
   Wakisaka N, 2019, ACTA OTO-LARYNGOL, V139, P94, DOI 10.1080/00016489.2018.1535193
   Wu LM, 2012, EUR J RADIOL, V81, P1798, DOI 10.1016/j.ejrad.2011.04.028
   Xiang DP, 2014, EUR RADIOL, V24, P2513, DOI 10.1007/s00330-014-3288-5
   Zhao HQ, 2019, EUR J RADIOL, V112, P14, DOI 10.1016/j.ejrad.2019.01.006
NR 26
TC 4
Z9 4
U1 1
U2 7
PU NATURE PORTFOLIO
PI BERLIN
PA HEIDELBERGER PLATZ 3, BERLIN, 14197, GERMANY
SN 2045-2322
J9 SCI REP-UK
JI Sci Rep
PD JAN 12
PY 2022
VL 12
IS 1
AR 578
DI 10.1038/s41598-021-04503-1
PG 8
WC Multidisciplinary Sciences
WE Science Citation Index Expanded (SCI-EXPANDED)
SC Science & Technology - Other Topics
GA YF9YM
UT WOS:000742155800009
PM 35022480
OA Green Published, gold
DA 2023-08-27
ER

PT J
AU Samadani, U
   Huang, JH
   Baranov, D
   Zager, EL
   Grady, MS
AF Samadani, U
   Huang, JH
   Baranov, D
   Zager, EL
   Grady, MS
TI Intracranial hypotension after intraoperative lumbar cerebrospinal fluid
   drainage
SO NEUROSURGERY
LA English
DT Article
DE aneurysm; cerebrospinal fluid; cranial base; intracranial hypotension;
   lumbar drain; skull base
ID CHIARI-I MALFORMATION; SPINAL DRAINAGE; COMPLICATIONS; SECONDARY;
   HEMATOMA; TUMORS
AB OBJECTIVE: Intraoperative lumbar cerebrospinal fluid drainage is frequently performed in a variety of neurosurgical procedures. A retrospective review is presented of the complications of lumbar cerebrospinal fluid drainage.
   METHODS: The records from 75 consecutive operations requiring intraoperative lumbar cerebrospinal drainage during a 1-year period at the Hospital of the University of Pennsylvania were reviewed to assess the types of complications attributable to spinal drainage and their rates of occurrence. The operations were categorized into 46 aneurysm clippings, 21 craniotomies for tumors, and 8 other cranial base procedures.
   RESULTS: Two patients developed transient postoperative neurological complications as a result of intracranial hypotension that resolved after epidural blood patching, with a reexploration craniotomy to drain an epidural collection performed in one patient. A third patient in the study developed a persistent deficit when intracranial hypotension led to intraoperative transtentorial herniation, which resulted in an unusual constellation of multiple brainstem infarcts that caused cranial neuropathy.
   CONCLUSION: Complications of intraoperative lumbar cerebrospinal fluid drainage resulting in transient (2 of 75 patients, 3%) or persistent (1 of 75 patients, 1%) neurological deficits caused by intracranial hypotension occur infrequently and may be related to preexisting conditions such as hydrocephalus.
C1 Hosp Univ Penn, Dept Neurosurg, Philadelphia, PA 19104 USA.
   Hosp Univ Penn, Dept Anesthesiol, Philadelphia, PA 19104 USA.
C3 University of Pennsylvania; Pennsylvania Medicine; University of
   Pennsylvania; Pennsylvania Medicine
RP Samadani, U (通讯作者)，Hosp Univ Penn, Dept Neurosurg, 3400 Spruce St,Silverstein 5, Philadelphia, PA 19104 USA.
EM
RI Huang, Jason H/J-4739-2019
OI Huang, Jason H/0000-0002-4426-0168; Mayberg, Marc/0000-0002-6718-7143
CR Amin-Hanjani S, 1999, NEUROSURGERY, V44, P1113, DOI 10.1097/00006123-199905000-00097
   Atkinson JLD, 1998, J NEUROSURG, V88, P237, DOI 10.3171/jns.1998.88.2.0237
   CHUMAS PD, 1993, J NEUROSURG, V78, P568, DOI 10.3171/jns.1993.78.4.0568
   FISHMAN RA, 1993, NEUROLOGY, V43, P609, DOI 10.1212/WNL.43.3_Part_1.609
   Francel P C, 1992, J Craniofac Surg, V3, P145, DOI 10.1097/00001665-199211000-00006
   Grady RE, 1999, ANESTH ANALG, V88, P388
   GRAF CJ, 1981, J NEUROSURG, V54, P392, DOI 10.3171/jns.1981.54.3.0392
   GUIDO LJ, 1976, J NEUROSURG, V45, P348, DOI 10.3171/jns.1976.45.3.0348
   HUANG PP, 1994, J NEUROSURG, V80, P1099, DOI 10.3171/jns.1994.80.6.1099
   KASUYA H, 1991, NEUROSURGERY, V28, P56, DOI 10.1227/00006123-199101000-00009
   Manley GT, 2000, J NEUROSURG, V92, P469, DOI 10.3171/jns.2000.92.3.0469
   MCDONALD JV, 1986, ARCH NEUROL-CHICAGO, V43, P936, DOI 10.1001/archneur.1986.00520090064019
   PALMER JD, 1994, NEUROSURGERY, V35, P1061, DOI 10.1227/00006123-199412000-00007
   PAVLIN DJ, 1979, ANESTHESIOLOGY, V51, P338, DOI 10.1097/00000542-197910000-00012
   PAYNER TD, 1994, NEUROSURGERY, V34, P429, DOI 10.1227/00006123-199403000-00007
   ROLAND PS, 1992, OTOLARYNG HEAD NECK, V107, P564, DOI 10.1177/019459989210700409
   SATHI S, 1993, NEUROSURGERY, V32, P306, DOI 10.1227/00006123-199302000-00023
   SNOW RB, 1991, NEUROSURGERY, V28, P880, DOI 10.1227/00006123-199106000-00016
   YUKI K, 1990, Neurological Surgery, V18, P567
NR 19
TC 55
Z9 60
U1 0
U2 2
PU OXFORD UNIV PRESS INC
PI CARY
PA JOURNALS DEPT, 2001 EVANS RD, CARY, NC 27513 USA
SN 0148-396X
EI 1524-4040
J9 NEUROSURGERY
JI Neurosurgery
PD JAN
PY 2003
VL 52
IS 1
BP 148
EP 151
DI 10.1097/00006123-200301000-00019
PG 4
WC Clinical Neurology; Surgery
WE Science Citation Index Expanded (SCI-EXPANDED)
SC Neurosciences & Neurology; Surgery
GA 631YL
UT WOS:000180195800033
PM 12493112
DA 2023-08-27
ER

PT J
AU Yamamoto, I
   Azuma, Y
   Yamaguchi, M
AF Yamamoto, Itaru
   Azuma, Yumiko
   Yamaguchi, Masamitsu
TI Cancer-related genes and ALS
SO FRONTIERS IN BIOSCIENCE-LANDMARK
LA English
DT Article
DE Cancer; ALS; FTLD; FUS; Hippo pathway; p53; NPM-hMLF1; Drosophila;
   Review
ID AMYOTROPHIC-LATERAL-SCLEROSIS; YES-ASSOCIATED PROTEIN; ACUTE
   MYELOID-LEUKEMIA; TRANSGENIC MOUSE MODEL; RNA-BINDING PROTEINS;
   TUMOR-SUPPRESSOR; HIPPO PATHWAY; CELL-PROLIFERATION; CARDIOMYOCYTE
   PROLIFERATION; PROMOTER HYPERMETHYLATION
AB Amyotrophic lateral sclerosis (ALS) is a neurodegenerative disorder that is characterized by the progressive degeneration of both upper motor neurons in the motor cortex and lower motor neurons in the brainstem and spinal cord. Recent advances in human genetics have identified more than 30 ALS-causing genes or genetic loci that include the fused in sarcoma (FUS) gene. In addition, a set of studies suggested a mutual relationship between cancer and ALS. The hpo gene, Drosophila MST was newly identified as a novel genetic modifier of the cabeza (caz), Drosophila FUS. The Hippo pathway negatively regulates the control of organ growth and tumor suppression. Moreover, the p53 tumor suppressor was found to genetically interact with caz. Frontotemporal lobar degeneration (FTLD) is characterized by the degeneration of neurons in the frontal and temporal lobes, and consists of a spectrum with ALS. Fusion protein nucleophosmin-human myeloid leukemia factor 1 (NPM-hMLF1), which is associated with the pathologies of myelodysplastic syndrome and acute myeloid leukemia, was recently shown to suppress defects in the Drosophila FTLD model expressing the human FUS gene. Further studies in the field are expected to elucidate epidemiological, genetic, and histopathological links between cancer and ALS/FTLD, and will lead to the development of therapeutic strategies. We herein summarize previous and current findings that support mutual links between cancer and ALS/FTLD.
C1 [Yamamoto, Itaru; Yamaguchi, Masamitsu] Kyoto Inst Technol, Dept Appl Biol, Sakyo Ku, Kyoto 6068585, Japan.
   [Yamamoto, Itaru; Yamaguchi, Masamitsu] Kyoto Inst Technol, Ctr Adv Insect Res, Kyoto, Japan.
   [Azuma, Yumiko] Kyoto Prefectural Univ Med, Kamigyo Ku, Kyoto 6028566, Japan.
C3 Kyoto Institute of Technology; Kyoto Institute of Technology; Kyoto
   Prefectural University of Medicine
RP Yamaguchi, M (通讯作者)，Kyoto Inst Technol, Dept Appl Biol, Sakyo Ku, Kyoto 6068585, Japan.
EM
OI Yamaguchi, Masamitsu/0000-0002-6321-9750
FU Practical Research Project for Rare/Intractable Diseases from the Japan
   Agency for Medical Research and Development (AMED); JSPS; JSPS Program
   for Advancing Strategic International Networks to Accelerate the
   Circulation of Talented Researchers [S2802]; JSPS KAKENHI Grant
   [16K07346]; Grants-in-Aid for Scientific Research [16K07346] Funding
   Source: KAKEN
FX We thank the Bloomington Drosophila Stock Center and Vienna Drosophila
   Genetic Resource Center for the fly lines. This research was partially
   supported by the Practical Research Project for Rare/Intractable
   Diseases from the Japan Agency for Medical Research and Development
   (AMED), the JSPS Core-to-Core Program, Asia-Africa Science Platforms,
   the JSPS Program for Advancing Strategic International Networks to
   Accelerate the Circulation of Talented Researchers (Grant No. S2802),
   and JSPS KAKENHI Grant Number 16K07346.
CR Aypar U, 2014, J MOL DIAGN, V16, P527, DOI 10.1016/j.jmoldx.2014.05.004
   Azuma Y, 2018, EXP CELL RES, V371, P311, DOI 10.1016/j.yexcr.2018.08.001
   Barbosa LF, 2010, BBA-MOL BASIS DIS, V1802, P462, DOI 10.1016/j.bbadis.2010.01.011
   Bhinge A, 2016, STEM CELLS, V34, P124, DOI 10.1002/stem.2233
   Cai J, 2010, GENE DEV, V24, P2383, DOI 10.1101/gad.1978810
   Camargo FD, 2007, CURR BIOL, V17, P2054, DOI 10.1016/j.cub.2007.10.039
   Campos-Melo D, 2014, BIOGERONTOLOGY, V15, P587, DOI 10.1007/s10522-014-9531-2
   Chan SW, 2008, CANCER RES, V68, P2592, DOI 10.1158/0008-5472.CAN-07-2696
   Chia R, 2018, LANCET NEUROL, V17, P94, DOI 10.1016/S1474-4422(17)30401-5
   Colombo E, 2002, NAT CELL BIOL, V4, P529, DOI 10.1038/ncb814
   Colombrita C, 2011, ARCH ITAL BIOL, V149, P83, DOI 10.4449/aib.v149i1.1261
   Cordenonsi M, 2011, CELL, V147, P759, DOI 10.1016/j.cell.2011.09.048
   CROZAT A, 1993, NATURE, V363, P640, DOI 10.1038/363640a0
   de Aguilar JLG, 2000, NEUROBIOL DIS, V7, P406, DOI 10.1006/nbdi.2000.0295
   De Santis R, 2017, STEM CELL REP, V9, P1450, DOI 10.1016/j.stemcr.2017.09.004
   Dong JX, 2007, CELL, V130, P1120, DOI 10.1016/j.cell.2007.07.019
   Dumezy F, 2013, HAEMATOLOGICA, V98, pE52, DOI 10.3324/haematol.2012.082149
   Fang F, 2013, AMYOTROPH LAT SCL FR, V14, P362, DOI 10.3109/21678421.2013.775309
   Farg MA, 2017, HUM MOL GENET, V26, P2882, DOI 10.1093/hmg/ddx170
   Fernandez A, 2009, GENE DEV, V23, P2729, DOI 10.1101/gad.1824509
   Ferrari R, 2014, LANCET NEUROL, V13, P686, DOI 10.1016/S1474-4422(14)70065-1
   Freedman DM, 2013, CANCER CAUSE CONTROL, V24, P55, DOI 10.1007/s10552-012-0089-5
   Freedman DM, 2005, NEUROEPIDEMIOLOGY, V25, P176, DOI 10.1159/000087447
   Gibbs KL, 2018, CELL DEATH DIS, V9, DOI 10.1038/s41419-018-0624-8
   Gibson SB, 2016, NEUROLOGY, V87, P289, DOI 10.1212/WNL.0000000000002757
   Gomez-Deza J, 2015, ACTA NEUROPATHOL COM, V3, DOI 10.1186/s40478-015-0218-y
   Grimwade D, 2010, BLOOD, V116, P354, DOI 10.1182/blood-2009-11-254441
   Hardiman O, 2017, NAT REV DIS PRIMERS, V3, DOI 10.1038/nrdp.2017.71
   Harvey KF, 2003, CELL, V114, P457, DOI 10.1016/S0092-8674(03)00557-9
   Harvey KF, 2013, NAT REV CANCER, V13, P246, DOI 10.1038/nrc3458
   Heallen T, 2011, SCIENCE, V332, P458, DOI 10.1126/science.1199010
   Hitzler JK, 1999, AM J PATHOL, V155, P53, DOI 10.1016/S0002-9440(10)65098-5
   Hobson EV, 2016, NAT REV NEUROL, V12, P526, DOI 10.1038/nrneurol.2016.111
   Hu WW, 2010, MOL CELL, V38, P689, DOI 10.1016/j.molcel.2010.05.027
   Huang JB, 2005, CELL, V122, P421, DOI 10.1016/j.cell.2005.06.007
   Imamura K, 2017, SCI TRANSL MED, V9, DOI 10.1126/scitranslmed.aaf3962
   IMMANUEL D, 1995, MOL CELL BIOL, V15, P4562
   Irby RB, 2000, ONCOGENE, V19, P5636, DOI 10.1038/sj.onc.1203912
   Itahana K, 2003, MOL CELL, V12, P1151, DOI 10.1016/S1097-2765(03)00431-3
   Jia JH, 2003, GENE DEV, V17, P2514, DOI 10.1101/gad.1134003
   Jiang Z, 2006, NEUROSCI RES, V56, P450, DOI 10.1016/j.neures.2006.09.006
   Johnson R, 2014, NAT REV DRUG DISCOV, V13, P63, DOI 10.1038/nrd4161
   JUSTICE RW, 1995, GENE DEV, V9, P534, DOI 10.1101/gad.9.5.534
   Kim EK, 2010, BBA-MOL BASIS DIS, V1802, P396, DOI 10.1016/j.bbadis.2009.12.009
   Konsavage WM, 2012, J BIOL CHEM, V287, P11730, DOI 10.1074/jbc.M111.327767
   Korgaonkar C, 2005, MOL CELL BIOL, V25, P1258, DOI 10.1128/MCB.25.4.1258-1271.2005
   Koul Hari K, 2013, Genes Cancer, V4, P342, DOI 10.1177/1947601913507951
   Kuntz C, 2000, EXP NEUROL, V165, P184, DOI 10.1006/exnr.2000.7464
   Lavado A, 2013, DEVELOPMENT, V140, P3323, DOI 10.1242/dev.096537
   Lee JK, 2013, P NATL ACAD SCI USA, V110, P12066, DOI 10.1073/pnas.1300894110
   Lee K, 2012, NEUROSCI LETT, V506, P271, DOI 10.1016/j.neulet.2011.11.021
   Lee KP, 2010, P NATL ACAD SCI USA, V107, P8248, DOI 10.1073/pnas.0912203107
   Lee KH, 2016, CELL, V167, P774, DOI 10.1016/j.cell.2016.10.002
   Lee WH, 2012, LEUKEMIA, V26, P1110, DOI 10.1038/leu.2011.365
   Li H, 2012, LIVER INT, V32, P38, DOI 10.1111/j.1478-3231.2011.02646.x
   Lian I, 2010, GENE DEV, V24, P1106, DOI 10.1101/gad.1903310
   Lim G, 2018, CANCER GENET CYTOGEN, V199, P101, DOI [10.1038/s41582-018-0009-8, DOI 10.1038/S41582-018-0009-8]
   Liu YX, 2013, DNA REPAIR, V12, P741, DOI 10.1016/j.dnarep.2013.06.002
   Lopez-Gonzalez R, 2016, NEURON, V92, P383, DOI 10.1016/j.neuron.2016.09.015
   Lu L, 2010, P NATL ACAD SCI USA, V107, P1437, DOI 10.1073/pnas.0911427107
   Mann DMA, 2013, ACTA NEUROPATHOL COM, V1, DOI 10.1186/2051-5960-1-68
   Martin LJ, 2000, NEUROBIOL DIS, V7, P613, DOI 10.1006/nbdi.2000.0314
   Matsumoto N, 2000, LEUKEMIA, V14, P1757, DOI 10.1038/sj.leu.2401897
   McCubrey JA, 2017, ONCOTARGET, V8, P14221, DOI 10.18632/oncotarget.13991
   Overholtzer M, 2006, P NATL ACAD SCI USA, V103, P12405, DOI 10.1073/pnas.0605579103
   Pantalacci S, 2003, NAT CELL BIOL, V5, P921, DOI 10.1038/ncb1051
   Prudlo J, 2000, BRAIN RES, V879, P183, DOI 10.1016/S0006-8993(00)02745-1
   RABBITTS TH, 1993, NAT GENET, V4, P175, DOI 10.1038/ng0693-175
   Ranganathan Srikanth, 2010, Open Pathol J, V4, P11
   Raver-Shapira N, 2007, MOL CELL, V26, P731, DOI 10.1016/j.molcel.2007.05.017
   Rizzuti M, 2018, SCI REP-UK, V8, DOI 10.1038/s41598-018-28366-1
   Sakuma C, 2016, CELL REP, V16, P2289, DOI 10.1016/j.celrep.2016.07.066
   Sanhueza M, 2015, PLOS GENET, V11, DOI 10.1371/journal.pgen.1005107
   Sasayama H, 2012, PLOS ONE, V7, DOI 10.1371/journal.pone.0039483
   Sathasivam S, 2001, NEUROPATH APPL NEURO, V27, P257, DOI 10.1046/j.0305-1846.2001.00332.x
   Schlegelmilch K, 2011, CELL, V144, P782, DOI 10.1016/j.cell.2011.02.031
   SCHMIDTZACHMANN MS, 1987, EMBO J, V6, P1881, DOI 10.1002/j.1460-2075.1987.tb02447.x
   Seidel C, 2007, MOL CARCINOGEN, V46, P865, DOI 10.1002/mc.20317
   SEMENKOVICH CF, 1990, BIOCHEMISTRY-US, V29, P9708, DOI 10.1021/bi00493a028
   Shimamura M, 2014, EXP CELL RES, V326, P36, DOI 10.1016/j.yexcr.2014.06.004
   Song H, 2010, P NATL ACAD SCI USA, V107, P1431, DOI 10.1073/pnas.0911409107
   Sperling AS, 2017, NAT REV CANCER, V17, P5, DOI 10.1038/nrc.2016.112
   Steinhardt AA, 2008, HUM PATHOL, V39, P1582, DOI 10.1016/j.humpath.2008.04.012
   STOLOW DT, 1995, NUCLEIC ACIDS RES, V23, P835, DOI 10.1093/nar/23.5.835
   Sullivan KD, 2018, CELL DEATH DIFFER, V25, P133, DOI 10.1038/cdd.2017.174
   Svetoni F, 2016, RNA BIOL, V13, P1089, DOI 10.1080/15476286.2016.1211225
   Taguchi YH, 2017, GENES-BASEL, V8, DOI 10.3390/genes8100243
   Takahashi Y, 2005, CLIN CANCER RES, V11, P1380, DOI 10.1158/1078-0432.CCR-04-1773
   Takahashi Y, 1999, NUCLEIC ACIDS RES, V27, P510, DOI 10.1093/nar/27.2.510
   Tan AY, 2009, J MOL CELL BIOL, V1, P82, DOI 10.1093/jmcb/mjp025
   Tao ZT, 2015, HUM MOL GENET, V24, P2426, DOI 10.1093/hmg/ddv005
   Udan RS, 2003, NAT CELL BIOL, V5, P914, DOI 10.1038/ncb1050
   van Es MA, 2017, LANCET, V390, P2084, DOI 10.1016/S0140-6736(17)31287-4
   Van Mossevelde S, 2018, NAT REV NEUROL, V14, P363, DOI 10.1038/s41582-018-0009-8
   Vogt MA, 2018, SCI REP-UK, V8, DOI 10.1038/s41598-018-26397-2
   von Gise A, 2012, P NATL ACAD SCI USA, V109, P2394, DOI 10.1073/pnas.1116136109
   Wang JY, 2013, MOL CELL, V51, P211, DOI 10.1016/j.molcel.2013.05.013
   Wu HT, 2013, CELL REP, V3, P1663, DOI 10.1016/j.celrep.2013.04.020
   Wu S, 2003, CELL, V114, P445, DOI 10.1016/S0092-8674(03)00549-X
   Xin M, 2013, P NATL ACAD SCI USA, V110, P13839, DOI 10.1073/pnas.1313192110
   Xin M, 2011, SCI SIGNAL, V4, DOI 10.1126/scisignal.2002278
   Xu MZ, 2009, CANCER-AM CANCER SOC, V115, P4576, DOI 10.1002/cncr.24495
   XU TA, 1995, DEVELOPMENT, V121, P1053
   Yamaguchi M., 2016, J CARCINOG MUTAGEN, V7, pe22, DOI [10.4172/2157-2518.1000e122, DOI 10.4172/2157-2518.1000E122]
   Yamamoto I, 2018, SCI REP-UK, V8, DOI 10.1038/s41598-018-29716-9
   Yokoyama K, 2018, BLOOD ADV, V2, P2513, DOI 10.1182/bloodadvances.2017010744
   Yoneda-Kato N, 2005, EMBO J, V24, P1739, DOI 10.1038/sj.emboj.7600656
   Yoneda-Kato N, 1999, ONCOGENE, V18, P3716, DOI 10.1038/sj.onc.1202711
   Yoneda-Kato N, 2008, MOL CELL BIOL, V28, P422, DOI 10.1128/MCB.02335-06
   YonedaKato N, 1996, ONCOGENE, V12, P265
   Zender L, 2006, CELL, V125, P1253, DOI 10.1016/j.cell.2006.05.030
   Zhang HY, 2011, P NATL ACAD SCI USA, V108, P2270, DOI 10.1073/pnas.1019603108
   Zhang MT, 2017, J NEUROCHEM, V143, P244, DOI 10.1111/jnc.14154
   Zhang T, 2012, HEPATOLOGY, V56, P2051, DOI 10.1002/hep.25899
   Zhang Y, 2018, CANCERS, V10, DOI 10.3390/cancers10090297
   Zhao B, 2007, GENE DEV, V21, P2747, DOI 10.1101/gad.1602907
   Zhou DW, 2009, CANCER CELL, V16, P425, DOI 10.1016/j.ccr.2009.09.026
NR 117
TC 9
Z9 9
U1 1
U2 5
PU FRONTIERS IN BIOSCIENCE INC
PI IRVINE
PA 16471 SCIENTIFIC WAY, IRVINE, CA 92618 USA
SN 1093-9946
EI 1093-4715
J9 FRONT BIOSCI-LANDMRK
JI Front. Biosci.
PD JUN 1
PY 2019
VL 24
BP 1241
EP 1258
DI 10.2741/4777
PG 18
WC Biochemistry & Molecular Biology; Cell Biology
WE Science Citation Index Expanded (SCI-EXPANDED)
SC Biochemistry & Molecular Biology; Cell Biology
GA JB8JO
UT WOS:000488814500003
PM 31136977
DA 2023-08-27
ER

PT S
AU Elkahloun, AG
   Powers, JF
   Nyska, A
   Etsenhofer, G
   Tischler, AS
AF Elkahloun, Abdel G.
   Powers, James F.
   Nyska, Abraham
   Etsenhofer, Graeme
   Tischler, Arthur S.
BE Pacak, K
   Eisenhofer, G
TI Gene expression profiling of rat pheochromocytoma
SO PHEOCHROMOCYTOMA
SE Annals of the New York Academy of Sciences
LA English
DT Article; Proceedings Paper
CT 1st International Symposium on Pheochromocytoma
CY OCT 20-23, 2005
CL Bethesda, MD
SP NIH
DE pheochromocytoma; phenylethanolamine N-methyltransferase; ret
   proto-oncogene; microarray; heat-shock proteins
ID CELL-LINES; PROTEIN; ACTIVATION
AB Sporadic and syndrome-associated human pheochromocytomas exhibit a spectrum of common and distinctive phenotypic markers. Animal models may contribute to understanding of common denominators leading to development and progression of pheochromocytoma, and to mechanisms that underlie distinctive phenotypes. Rat pheochromocytomas are common, in contrast to their human counterparts, and their frequency is increased by a variety of genotoxic or nongenotoxic agents. Toxicological studies of rats are therefore a potentially rich source of information on pheochromocytoma biology. To compare the molecular profiles of rat and human pheochromocytomas and to identify pathways potentially involved in pathogenesis of rat pheochromocytomas, we conducted a gene expression profiling study comparing 31 pheochromocytomas obtained from the National Toxicology Program to normal adult rat adrenal medulla. The microarray chips were generated from 31,769-oligomer set representing over 27,200 unique Mouse Ensembl genes. The analysis showed over 1,900 genes that were up- or downregulated in the tumors. More than half of the former are involved in protein synthesis and signal transduction, including oncogenes of the RAS family and several heat shock proteins and chaperones. Downregulated genes included receptors and tumor-suppressor genes, including NF2 and Dmbt1. Specific genes related to neuroendocrine function were either upregulated or downregulated in subsets of tumors. Cross-comparison with a human pheochromocytoma database showed greater than 60% correlation. Results of this study reveal both generic and specific parallels between rat and human pheochromocytomas.
C1 NHGRI, Genome Technol Branch, NIH, Bethesda, MD 20892 USA.
   Tufts Univ, New England Med Ctr, Dept Pathol, Boston, MA 02111 USA.
   Natl Inst Environm Hlth Sci, NIH, Res Triangle Pk, NC USA.
   NINDS, Clin Neurocardiol Sect, NIH, Bethesda, MD 20892 USA.
C3 National Institutes of Health (NIH) - USA; NIH National Human Genome
   Research Institute (NHGRI); Tufts Medical Center; Tufts University;
   National Institutes of Health (NIH) - USA; NIH National Institute of
   Environmental Health Sciences (NIEHS); National Institutes of Health
   (NIH) - USA; NIH National Institute of Neurological Disorders & Stroke
   (NINDS)
RP Elkahloun, AG (通讯作者)，NHGRI, Genome Technol Branch, NIH, Bldg 50,50 South Dr, Bethesda, MD 20892 USA.
EM
OI Tischler, Arthur/0000-0002-9726-5739; Powers, James/0000-0001-8422-3938
FU Intramural NIH HHS Funding Source: Medline; NCI NIH HHS [CA48017]
   Funding Source: Medline
CR ALLANDER SV, 2003, CANCER RES, V61, P8624
   Boon K, 2001, EMBO J, V20, P1383, DOI 10.1093/emboj/20.6.1383
   Chen YD, 2002, BIOINFORMATICS, V18, P1207, DOI 10.1093/bioinformatics/18.9.1207
   Eisenhofer G, 2004, ENDOCR-RELAT CANCER, V11, P897, DOI 10.1677/erc.1.00838
   Kees UR, 2004, IMMUNOL CELL BIOL, V82, P154, DOI 10.1046/j.0818-9641.2004.01236.x
   Kim S, 2000, P NATL ACAD SCI USA, V97, P11198, DOI 10.1073/pnas.200372597
   Letizia C, 2001, HORM METAB RES, V33, P290, DOI 10.1055/s-2001-15281
   Marsee DK, 2004, J BIOL CHEM, V279, P43990, DOI 10.1074/jbc.M407503200
   Park JW, 2003, J CLIN ENDOCR METAB, V88, P3346, DOI 10.1210/jc.2002-020340
   Pooler AM, 2005, NEUROSCIENCE, V134, P207, DOI 10.1016/j.neuroscience.2005.03.050
   Powers JF, 2005, CANCER GENET CYTOGEN, V159, P27, DOI 10.1016/j.cancergencyto.2004.09.018
   Sherr CJ, 1999, GENE DEV, V13, P1501, DOI 10.1101/gad.13.12.1501
   Teng SC, 2004, J BIOL CHEM, V279, P14649, DOI 10.1074/jbc.M308842200
   TISCHLER AS, HISTOLOGY, V19, P883
   Walerych D, 2004, J BIOL CHEM, V279, P48836, DOI 10.1074/jbc.M407601200
NR 15
TC 6
Z9 6
U1 0
U2 1
PU WILEY-BLACKWELL
PI HOBOKEN
PA 111 RIVER STREET, HOBOKEN, NJ, UNITED STATES
SN 0077-8923
BN 1-57331-597-4
J9 ANN NY ACAD SCI
JI Ann.NY Acad.Sci.
PY 2006
VL 1073
BP 290
EP 299
DI 10.1196/annals.1353.033
PG 10
WC Oncology; Endocrinology & Metabolism; Multidisciplinary Sciences
WE Conference Proceedings Citation Index - Science (CPCI-S); Science Citation Index Expanded (SCI-EXPANDED)
SC Oncology; Endocrinology & Metabolism; Science & Technology - Other
   Topics
GA BFD27
UT WOS:000241140400032
PM 17102098
DA 2023-08-27
ER

PT J
AU Moog, S
   Lussey-Lepoutre, C
   Favier, J
AF Moog, Sophie
   Lussey-Lepoutre, Charlotte
   Favier, Judith
TI Epigenetic and metabolic reprogramming of SDH-deficient paragangliomas
SO ENDOCRINE-RELATED CANCER
LA English
DT Review
DE pheochromocytoma; paraganglioma; succinate dehydrogenase; epigenetics;
   metabolism; pseudohypoxia; biomarkers; targeted therapies
ID MAGNETIC-RESONANCE-SPECTROSCOPY; GENE-MUTATIONS; IN-VIVO; COMPLEX-II;
   MALIGNANT PHEOCHROMOCYTOMAS; HEREDITARY PHEOCHROMOCYTOMA; PYRUVATE
   CARBOXYLATION; GERMLINE MUTATIONS; RESPIRATORY-CHAIN; SUCCINATE
AB Pheochromocytomas and paragangliomas (PPGL) are rare neuroendocrine tumors arising from the adrenal medulla or extra-adrenal paraganglia. Around 40% of all cases are caused by a germline mutation in a susceptibility gene, half of which being found in an SDHx gene (SDHA, SDHB, SDHC, SDHD or SDHAF2). They encode the four subunits and assembly factor of succinate dehydrogenase (SDH), a mitochondrial enzyme involved both in the tricarboxylic acid cycle and electron transport chain. SDHx mutations lead to the accumulation of succinate, which acts as an oncometabolite by inhibiting iron(II) and alpha-ketoglutarate-dependent dioxygenases thereby regulating the cell's hypoxic response and epigenetic processes. Moreover, SDHx mutations induce cell metabolic reprogramming and redox imbalance. Major discoveries in PPGL pathophysiology have been made since the initial discovery of SDHD gene mutations in 2000, improving the understanding of their biology and patient management. It indeed provides new opportunities for diagnostic tools and innovative therapeutic targets in order to improve the prognosis of patients affected by these rare tumors, in particular in the context of metastatic diseases associated with SDHB mutations. This review first describes an overview of the pathophysiology and then focuses on clinical implications of the epigenetic and metabolic reprogramming of SDH-deficient PPGL.
C1 [Moog, Sophie; Lussey-Lepoutre, Charlotte; Favier, Judith] PARCC, INSERM, Equipe Labellisee Ligue Canc, UMR970, Paris, France.
   [Moog, Sophie; Favier, Judith] Univ Paris, Paris, France.
   [Lussey-Lepoutre, Charlotte] Sorbonne Univ, Pitie Salpetriere Hosp, Dept Nucl Med, Paris, France.
C3 Institut National de la Sante et de la Recherche Medicale (Inserm);
   UDICE-French Research Universities; Universite Paris Cite; UDICE-French
   Research Universities; Universite Paris Cite; Assistance Publique
   Hopitaux Paris (APHP); Hopital Universitaire Pitie-Salpetriere - APHP;
   UDICE-French Research Universities; Sorbonne Universite
RP Favier, J (通讯作者)，PARCC, INSERM, Equipe Labellisee Ligue Canc, UMR970, Paris, France.; Favier, J (通讯作者)，Univ Paris, Paris, France.
EM
RI Lussey-Lepoutre, Charlotte/AAO-7653-2020; Favier, Judith/M-7601-2017
OI Lussey-Lepoutre, Charlotte/0000-0003-2228-0106; Favier,
   Judith/0000-0001-8190-5853
FU Fondation pour la Recherche Medicale [FDM201806005916]; Plan Cancer,
   Epigenetique et Cancer [EPIG201303 METABEPIC]; Paradifference
   Foundation; la Ligue Contre le Cancer (Equipe Labellisee); the Cancer
   Research for Personalized Medicine -CARPEM project (Site de Recherche
   Integre sur le Cancer -SIRIC)
FX Sophie Moog is the recipient of a fellowship from la Fondation pour la
   Recherche Medicale (FDM201806005916). The team has received funding from
   The Plan Cancer, Epigenetique et Cancer (EPIG201303 METABEPIC), The
   Paradifference Foundation, la Ligue Contre le Cancer (Equipe Labellisee)
   and the Cancer Research for Personalized Medicine -CARPEM project (Site
   de Recherche Integre sur le Cancer -SIRIC).
CR Toledo RA, 2017, ENDOCR-RELAT CANCER, V24, pC9, DOI 10.1530/ERC-16-0479
   Amar L, 2007, J CLIN ENDOCR METAB, V92, P3822, DOI 10.1210/jc.2007-0709
   Amar L, 2016, EUR J ENDOCRINOL, V175, pR135, DOI 10.1530/EJE-16-0189
   Andronesi OC, 2012, SCI TRANSL MED, V4, DOI 10.1126/scitranslmed.3002693
   Astuti D, 2001, AM J HUM GENET, V69, P49, DOI 10.1086/321282
   Atlante S, 2018, CELL DEATH DIS, V9, DOI 10.1038/s41419-018-0802-8
   Baudin E, 2014, EUR J ENDOCRINOL, V171, pR111, DOI 10.1530/EJE-14-0113
   Bayley JP, 2010, LANCET ONCOL, V11, P366, DOI 10.1016/S1470-2045(10)70007-3
   Baysal BE, 2000, SCIENCE, V287, P848, DOI 10.1126/science.287.5454.848
   Benn DE, 2006, ANN NY ACAD SCI, V1073, P104, DOI 10.1196/annals.1353.011
   Bezawork-Geleta A, 2018, NAT COMMUN, V9, DOI 10.1038/s41467-018-04603-z
   Briere JJ, 2005, HUM MOL GENET, V14, P3263, DOI 10.1093/hmg/ddi359
   Buffet A, 2020, BEST PRACT RES CL EN, V34, DOI 10.1016/j.beem.2020.101416
   Buffet A, 2019, J CLIN ENDOCR METAB, V104, P1109, DOI 10.1210/jc.2018-02411
   Buffet A, 2018, CANCER RES, V78, P1914, DOI 10.1158/0008-5472.CAN-17-2463
   Burnichon N, 2017, J MED GENET, V54, P100, DOI 10.1136/jmedgenet-2016-104297
   Burnichon N, 2010, HUM MOL GENET, V19, P3011, DOI 10.1093/hmg/ddq206
   Burnichon N, 2009, J CLIN ENDOCR METAB, V94, P2817, DOI 10.1210/jc.2008-2504
   Cardaci S, 2015, NAT CELL BIOL, V17, P1317, DOI 10.1038/ncb3233
   Casey RE, 2018, JCO PRECIS ONCOL, V2, P1, DOI 10.1200/PO.17.00191
   Castro-Vega LJ, 2014, HUM MOL GENET, V23, P2440, DOI 10.1093/hmg/ddt639
   Cervera AM, 2009, MOL CANCER, V8, DOI 10.1186/1476-4598-8-89
   Chen WF, 2016, NATURE, V539, P112, DOI 10.1038/nature19796
   Cho H, 2016, NATURE, V539, P107, DOI 10.1038/nature19795
   Choi C, 2012, NAT MED, V18, P624, DOI 10.1038/nm.2682
   Courtney KD, 2018, J CLIN ONCOL, V36, P867, DOI 10.1200/JCO.2017.74.2627
   de Cubas AA, 2015, CLIN CANCER RES, V21, P3020, DOI 10.1158/1078-0432.CCR-14-2804
   Eisenhofer Graeme, 2017, Clin Biochem Rev, V38, P69
   Favier J, 2015, NAT REV ENDOCRINOL, V11, P101, DOI 10.1038/nrendo.2014.188
   Favier J, 2010, BEST PRACT RES CL EN, V24, P957, DOI 10.1016/j.beem.2010.10.004
   Favier J, 2009, PLOS ONE, V4, DOI 10.1371/journal.pone.0007094
   Fishbein L, 2017, CANCER CELL, V31, P181, DOI 10.1016/j.ccell.2017.01.001
   Florio R, 2018, SCI REP-UK, V8, DOI 10.1038/s41598-018-31797-5
   Gaal J, 2011, MODERN PATHOL, V24, P147, DOI 10.1038/modpathol.2010.185
   Gill AJ, 2011, NEW ENGL J MED, V364, P885, DOI 10.1056/NEJMc1012357
   Gill AJ, 2010, HUM PATHOL, V41, P805, DOI 10.1016/j.humpath.2009.12.005
   Gimenez-Roqueplo AP, 2001, AM J HUM GENET, V69, P1186, DOI 10.1086/324413
   Guzy RD, 2008, MOL CELL BIOL, V28, P718, DOI 10.1128/MCB.01338-07
   Hadoux J, 2014, INT J CANCER, V135, P2711, DOI 10.1002/ijc.28913
   Hamidi O, 2017, CLIN ENDOCRINOL, V87, P440, DOI 10.1111/cen.13434
   Hanahan D, 2011, CELL, V144, P646, DOI 10.1016/j.cell.2011.02.013
   Hescot S, 2019, J CLIN ENDOCR METAB, V104, P2367, DOI 10.1210/jc.2018-01968
   Hobert JA, 2012, GENET MED, V14, P616, DOI 10.1038/gim.2011.63
   Hoekstra AS, 2015, ONCOTARGET, V6, P38777, DOI 10.18632/oncotarget.6091
   Imperiale A, 2015, NEOPLASIA, V17, P55, DOI 10.1016/j.neo.2014.10.010
   Islam MS, 2018, ANNU REV BIOCHEM, V87, P585, DOI 10.1146/annurev-biochem-061516-044724
   Jimenez Camilo, 2020, Endocr Relat Cancer, V27, pR239, DOI 10.1530/ERC-20-0043
   Killian JK, 2013, CANCER DISCOV, V3, P648, DOI 10.1158/2159-8290.CD-13-0092
   Kim E, 2015, ENDOCR-RELAT CANCER, V22, P387, DOI 10.1530/ERC-15-0099
   Kl'uckova K, 2020, FASEB J, V34, P303, DOI 10.1096/fj.201901456R
   KNUDSON AG, 1971, P NATL ACAD SCI USA, V68, P820, DOI 10.1073/pnas.68.4.820
   Korpershoek E, 2011, J CLIN ENDOCR METAB, V96, pE1472, DOI 10.1210/jc.2011-1043
   Lam AKY, 2017, ENDOCR PATHOL, V28, P213, DOI 10.1007/s12022-017-9484-5
   Lenders JWM, 2020, J HYPERTENS, V38, P1443, DOI 10.1097/HJH.0000000000002438
   Lendvai N, 2014, ENDOCRINOLOGY, V155, P27, DOI 10.1210/en.2013-1549
   Letouze E, 2013, CANCER CELL, V23, P739, DOI 10.1016/j.ccr.2013.04.018
   Li M, 2017, MED SCI MONITOR, V23, P3253, DOI 10.12659/MSM.903348
   Liu Y, 2020, CLIN CANCER RES, V26, P3868, DOI 10.1158/1078-0432.CCR-19-2335
   Lorendeau D, 2017, METAB ENG, V43, P187, DOI 10.1016/j.ymben.2016.11.005
   Lussey-Lepoutre C, 2020, EUR J NUCL MED MOL I, V47, P1510, DOI 10.1007/s00259-019-04633-9
   Lussey-Lepoutre C, 2016, CLIN CANCER RES, V22, P1120, DOI 10.1158/1078-0432.CCR-15-1576
   Lussey-Lepoutre C, 2015, NAT COMMUN, V6, DOI 10.1038/ncomms9784
   Martiniova L, 2011, ENDOCR-RELAT CANCER, V18, P143, DOI 10.1677/ERC-10-0090
   Menara M, 2015, J CLIN ENDOCR METAB, V100, pE287, DOI 10.1210/jc.2014-1870
   Molina JR, 2018, NAT MED, V24, P1036, DOI 10.1038/s41591-018-0052-4
   Morin A, 2020, CELL REP, V30, P4551, DOI 10.1016/j.celrep.2020.03.022
   Ngo B, 2019, NAT REV CANCER, V19, P271, DOI 10.1038/s41568-019-0135-7
   Nickerson ML, 2008, CLIN CANCER RES, V14, P4726, DOI 10.1158/1078-0432.CCR-07-4921
   Niemann S, 2000, NAT GENET, V26, P268, DOI 10.1038/81551
   Niemeijer ND, 2014, CLIN ENDOCRINOL, V81, P642, DOI 10.1111/cen.12542
   O'Kane GM, 2019, BRIT J CANCER, V120, P1113, DOI 10.1038/s41416-019-0474-x
   Oudijk L, 2013, MODERN PATHOL, V26, P456, DOI 10.1038/modpathol.2012.186
   Pang Y, 2018, CLIN CANCER RES, V24, P3423, DOI 10.1158/1078-0432.CCR-17-3406
   Papadopoulos KP, 2018, J CLIN ONCOL, V36, DOI 10.1200/JCO.2018.36.15_suppl.2508
   Papathomas TG, 2015, MODERN PATHOL, V28, P807, DOI 10.1038/modpathol.2015.41
   Persson CU, 2020, EXP CELL RES, V388, DOI 10.1016/j.yexcr.2020.111845
   Pollard PJ, 2005, HUM MOL GENET, V14, P2231, DOI 10.1093/hmg/ddi227
   Rao JU, 2013, CLIN CANCER RES, V19, P3787, DOI 10.1158/1078-0432.CCR-12-3922
   Richter S, 2014, J CLIN ENDOCR METAB, V99, P3903, DOI 10.1210/jc.2014-2151
   RUSTIN P, 1994, CLIN CHIM ACTA, V228, P35, DOI 10.1016/0009-8981(94)90055-8
   Scheuerrnann TH, 2015, J MED CHEM, V58, P5930, DOI 10.1021/acs.jmedchem.5b00529
   Selak MA, 2005, CANCER CELL, V7, P77, DOI 10.1016/j.ccr.2004.11.022
   Selak MA, 2006, BBA-BIOENERGETICS, V1757, P567, DOI 10.1016/j.bbabio.2006.05.015
   Shi YF, 2019, NATURE, V567, P341, DOI 10.1038/s41586-019-0993-x
   Sulkowski PL, 2020, NATURE, V582, P586, DOI 10.1038/s41586-020-2363-0
   Sulkowski PL, 2018, NAT GENET, V50, P1086, DOI 10.1038/s41588-018-0170-4
   Thakur S, 2019, J MOL ENDOCRINOL, V63, pR17, DOI 10.1530/JME-19-0083
   Toledo RA, 2015, CURR OPIN ENDOCRINOL, V22, P169, DOI 10.1097/MED.0000000000000150
   Toledo RA, 2013, ENDOCR-RELAT CANCER, V20, P349, DOI 10.1530/ERC-13-0101
   van Nederveen FH, 2009, LANCET ONCOL, V10, P764, DOI 10.1016/S1470-2045(09)70164-0
   Varoquaux A, 2015, ENDOCR-RELAT CANCER, V22, pM1, DOI 10.1530/ERC-15-0246
   Wallace PW, 2020, J PATHOL, V251, P378, DOI 10.1002/path.5472
   WARBURG O, 1956, SCIENCE, V124, P269
   Wedekind MF, 2020, J CLIN ONCOL, V38
   Xu R, 2019, J MED CHEM, V62, P6876, DOI 10.1021/acs.jmedchem.9b00719
NR 95
TC 18
Z9 18
U1 0
U2 6
PU BIOSCIENTIFICA LTD
PI BRISTOL
PA STARLING HOUSE, 1600 BRISTOL PARKWAY N, BRISTOL, ENGLAND
SN 1351-0088
EI 1479-6821
J9 ENDOCR-RELAT CANCER
JI Endocr.-Relat. Cancer
PD DEC
PY 2020
VL 27
IS 12
BP R451
EP R463
DI 10.1530/ERC-20-0346
PG 13
WC Oncology; Endocrinology & Metabolism
WE Science Citation Index Expanded (SCI-EXPANDED)
SC Oncology; Endocrinology & Metabolism
GA SC9QI
UT WOS:000650997900002
PM 33112834
OA Bronze
DA 2023-08-27
ER

PT J
AU Tamborino, G
   Nonnekens, JA
   Struelens, LW
   De Saint-Hubert, M
   Verburg, FA
   Konijnenberg, MW
AF Tamborino, Giulia
   Nonnekens, Julie A.
   Struelens, Lara W.
   De Saint-Hubert, Marijke
   Verburg, Frederik A.
   Konijnenberg, Mark W.
TI Therapeutic efficacy of heterogeneously distributed radiolabelled
   peptides: Influence of radionuclide choice
SO PHYSICA MEDICA-EUROPEAN JOURNAL OF MEDICAL PHYSICS
LA English
DT Article
DE Dose-effect relationship; Peptide receptor radionuclide therapy; Monte
   Carlo simulation; S-value
ID BONE-MARROW; SOMATOSTATIN; MODEL; LU-177-DOTATATE; TOXICITY; RETENTION;
   DOSIMETRY; PROFILES; KIDNEY; LU-177
AB Purpose: To model dose-response relationships for in vivo experiments with radiolabelled peptides enabling maximum therapeutic efficacy while limiting toxicity to kidney and bone marrow.
   Methods: A multiregional murine kidney phantom, with a kinetic model for cortex and outer medulla distribution, were used to predict renal toxicity.
   Maximum tolerated activities to avoid nephrotoxicity (at 40 Gy Biological Effective Dose BED) and hematologic toxicity (at 2 Gy) were compared.
   The therapeutic efficacy of Y-90, Tb-161, Lu-177 and Bi-213 was assessed at their respective maximum tolerated activities based on cellular-level dosimetry accounting for activity and tumor heterogeneity. These results were compared with average tumor-dosimetry-based predictions.
   Results: The kidney was found to be the dose-limiting organ for all radionuclides, limiting the administered activity to 44 MBq Lu-177, 34 MBq Tb-161, 19 MBq Y-90 and 13 MBq Bi-213 , respectively. The average S-values for the initial heterogeneous activity distribution in the tumor volume are not significantly different from the homogeneous ones. The in vivo tumor cell survivals predicted by assuming uniform dose rate distributions are not significantly different from those for heterogeneous dose rate-based predictions. The lowest in vivo survival was found for Bi-213 (2%) followed by Tb-161 (30%), Lu-177 (37%) and Y-90 (60%). The minimal effective dose rate for cell kill is 13-14 mGy/h for beta-emitters and 2.2 mGy/h for the alpha-particle emitter Bi-213, below these values proliferation takes over.
   Conclusions: Radionuclides emitting alpha-particles have the highest potential for improving therapeutic efficacy in tumors and metastases with uniform receptor expression, after careful evaluation of their burden to the healthy organs.
C1 [Tamborino, Giulia; Struelens, Lara W.; De Saint-Hubert, Marijke] Belgian Nucl Res Ctr SCK CEN, Res Dosimetr Applicat, Boeretang 200, Mol, Belgium.
   [Tamborino, Giulia; Nonnekens, Julie A.; Verburg, Frederik A.; Konijnenberg, Mark W.] Erasmus Univ, Med Ctr, Dept Radiol & Nucl Med, Doctor Molewaterpl 40, NL-3000 CA Rotterdam, Netherlands.
   [Nonnekens, Julie A.] Erasmus Univ, Med Ctr, Canc Inst, Dept Mol Genet, Doctor Molewaterpl 40, NL-3000 CA Rotterdam, Netherlands.
C3 Belgian Nuclear Research Centre (SCK-CEN); Erasmus University Rotterdam;
   Erasmus MC; Erasmus University Rotterdam - Excl Erasmus MC; Erasmus
   University Rotterdam; Erasmus MC; Erasmus University Rotterdam - Excl
   Erasmus MC
RP Konijnenberg, MW (通讯作者)，ERASMUS MC, Dept Radiol & Nucl Med, Room Ns-561,POB 2040, NL-3000 CA Rotterdam, Netherlands.
EM
CR [Anonymous], 1980, ANN ICRP, V4, pIII, DOI [10.1016/0146-6453(80)90047-0, DOI 10.1016/0146-6453(80)90047-0]
   Baechler S, 2008, MED PHYS, V35, P1123, DOI 10.1118/1.2836421
   Barone R, 2005, J NUCL MED, V46, p99S
   Baum RP, 2021, J NUCL MED, V62, P1391, DOI 10.2967/jnumed.120.258376
   Bergsma H, 2018, J NUCL MED, V59, P452, DOI 10.2967/jnumed.117.189712
   Bodei L, 2008, EUR J NUCL MED MOL I, V35, P1847, DOI 10.1007/s00259-008-0778-1
   Borgna F, 2021, PHARMACEUTICS, V13, DOI 10.3390/pharmaceutics13040536
   Breeman WAP, 2003, EUR J NUCL MED MOL I, V30, P917, DOI 10.1007/s00259-003-1142-0
   Champion C, 2016, THERANOSTICS, V6, P1611, DOI 10.7150/thno.15132
   Chan HS, 2016, EJNMMI RES, V6, DOI 10.1186/s13550-016-0240-5
   Chan HS, 2016, EJNMMI RES, V6, DOI 10.1186/s13550-016-0162-2
   Colvin GA, 2004, LEUKEMIA, V18, P575, DOI 10.1038/sj.leu.2403268
   Dale R, 2004, CANCER BIOTHER RADIO, V19, P363, DOI 10.1089/1084978041425070
   de Jong M, 2005, J NUCL MED, V46, p13S
   de Jong M, 2001, J NUCL MED, V42, P1841
   de Jong M, 2001, INT J CANCER, V92, P628, DOI 10.1002/1097-0215(20010601)92:5<628::AID-IJC1244>3.3.CO;2-C
   EMAMI B, 1991, INT J RADIAT ONCOL, V21, P109, DOI 10.1016/0360-3016(91)90171-Y
   Feijtel D, 2021, THERANOSTICS, V11, P491, DOI 10.7150/thno.51215
   Forrer F, 2009, EUR J NUCL MED MOL I, V36, P1138, DOI 10.1007/s00259-009-1072-6
   Gracheva N, 2019, EJNMMI RADIOPHARM CH, V4, DOI 10.1186/s41181-019-0063-6
   Haller S, 2016, EJNMMI RES, V6, DOI 10.1186/s13550-016-0171-1
   Hindie E, 2016, J NUCL MED, V57, P759, DOI 10.2967/jnumed.115.170423
   Hobbs RF, 2012, PHYS MED BIOL, V57, P4403, DOI 10.1088/0031-9155/57/13/4403
   Hobbs RF, 2012, PHYS MED BIOL, V57, P3207, DOI 10.1088/0031-9155/57/10/3207
   Jan S, 2004, PHYS MED BIOL, V49, P4543, DOI 10.1088/0031-9155/49/19/007
   Kletting P, 2012, MED PHYS, V39, P5708, DOI 10.1118/1.4747266
   Kratochwil C, 2014, EUR J NUCL MED MOL I, V41, P2106, DOI 10.1007/s00259-014-2857-9
   Kwekkeboom DJ, 2003, EUR J NUCL MED MOL I, V30, P417, DOI 10.1007/s00259-002-1050-8
   Kwekkeboom DJ, 2005, J NUCL MED, V46, p62S
   Melis M, 2005, EUR J NUCL MED MOL I, V32, P1136, DOI 10.1007/s00259-005-1793-0
   Melis M, 2007, NUCL MED BIOL, V34, P633, DOI 10.1016/j.nucmedbio.2007.05.002
   Muller C, 2019, EUR J NUCL MED MOL I, V46, P1919, DOI 10.1007/s00259-019-04345-0
   Nayak TK, 2007, NUCL MED BIOL, V34, P185, DOI 10.1016/j.nucmedbio.2006.11.006
   ODONOGHUE JA, 1995, J NUCL MED, V36, P1902
   Pach D, 2012, RADIOTHER ONCOL, V102, P45, DOI 10.1016/j.radonc.2011.08.006
   Pacilio M, 2014, EUR J NUCL MED MOL I, V41, P238, DOI 10.1007/s00259-013-2552-2
   Reubi JC, 2000, EUR J NUCL MED, V27, P273, DOI 10.1007/s002590050034
   Rolleman EJ, 2007, EUR J NUCL MED MOL I, V34, P219, DOI 10.1007/s00259-006-0232-1
   Sandstrom M, 2013, J NUCL MED, V54, P33, DOI 10.2967/jnumed.112.107524
   Segars WP, 2004, MOL IMAGING BIOL, V6, P149, DOI 10.1016/j.mibio.2004.03.002
   Sgouros G, 2006, J NUCL MED, P47
   Strosberg J, 2017, NEW ENGL J MED, V376, P125, DOI 10.1056/NEJMoa1607427
   Strosberg J, 2018, J CLIN ONCOL, V36, P2578, DOI 10.1200/JCO.2018.78.5865
   Svensson J, 2012, NUCL MED BIOL, V39, P756, DOI 10.1016/j.nucmedbio.2012.02.003
   Tamborino G, 2022, J NUCL MED, V63, P100, DOI 10.2967/jnumed.121.262122
   Tamborino G, 2020, EJNMMI PHYS, V7, DOI 10.1186/s40658-020-0276-5
   Uusija?rvi H, 2006, J NUCL MED, P47
NR 47
TC 2
Z9 2
U1 0
U2 4
PU ELSEVIER SCI LTD
PI OXFORD
PA THE BOULEVARD, LANGFORD LANE, KIDLINGTON, OXFORD OX5 1GB, OXON, ENGLAND
SN 1120-1797
EI 1724-191X
J9 PHYS MEDICA
JI Phys. Medica
PD APR
PY 2022
VL 96
BP 90
EP 100
DI 10.1016/j.ejmp.2022.02.021
EA MAR 2022
PG 11
WC Radiology, Nuclear Medicine & Medical Imaging
WE Science Citation Index Expanded (SCI-EXPANDED)
SC Radiology, Nuclear Medicine & Medical Imaging
GA 0N8RV
UT WOS:000783099800009
PM 35245708
OA hybrid, Green Published
DA 2023-08-27
ER

PT J
AU Matschke, J
   Laas, R
AF Matschke, Jakob
   Laas, Rudolf
TI Sudden death due to central alveolar hypoventilation syndrome (Ondine's
   curse) in a 39-year-old woman with heterotopia of the inferior olive
SO AMERICAN JOURNAL OF FORENSIC MEDICINE AND PATHOLOGY
LA English
DT Article
DE Ondine's curse; forensic neuropathology; sudden death; nervous system
   malformation
ID BRAIN-STEM TUMOR; MULTIPLE-SCLEROSIS; CENTRAL CHEMOSENSITIVITY;
   AUTOMATIC RESPIRATION; ENCEPHALITIS; FAILURE; NEURONS; POLIOMYELITIS;
   INFARCTION; ISCHEMIA
AB Failure of automatic involuntary respiration with preservation of voluntary respiratory drive (Ondine's curse) is a rare occurrence which has been reported following a variety of morphologic lesions near respiratory centers in the lower brainstem. We report the case of a 39-year-old woman with a syndrome of fulminant respiratory failure with features of Ondine's curse in whom neuropathologic examination disclosed a preexisting malformation of the lower brainstem, as well as acute local subarachnoid bleeding. Mechanisms in the present case are discussed and a review of similar cases published so far is given. The necessity of sound investigation, including neuropathologic studies in cases of sudden unexplained death, is underlined.
C1 Univ Hamburg, Ctr Med, Inst Neuropathol, D-20246 Hamburg, Germany.
   Univ Hamburg, Inst Legal Med, D-20246 Hamburg, Germany.
C3 University of Hamburg; University of Hamburg
RP Matschke, J (通讯作者)，Univ Hamburg, Ctr Med, Inst Neuropathol, Martinistr 52, D-20246 Hamburg, Germany.
EM
RI Matschke, Jakob/F-6641-2013
OI Matschke, Jakob/0000-0001-6080-0198
CR ALTMAN J, 1987, J COMP NEUROL, V257, P490, DOI 10.1002/cne.902570403
   [Anonymous], 1962, CLIN RES
   Auer RN, 1996, CLIN NEUROPATHOL, V15, P101
   BEAL MF, 1983, NEUROLOGY, V33, P717, DOI 10.1212/WNL.33.6.717
   BELMUSTO L, 1963, J NEUROSURG, V20, P225, DOI 10.3171/jns.1963.20.3.0225
   BOGOUSSLAVSKY J, 1990, ANN NEUROL, V28, P668, DOI 10.1002/ana.410280511
   BOOR JW, 1977, ARCH NEUROL-CHICAGO, V34, P686, DOI 10.1001/archneur.1977.00500230056009
   BOURRAT F, 1988, DEV BRAIN RES, V39, P19, DOI 10.1016/0165-3806(88)90064-8
   CLARKE AS, 1994, CELL ADHES COMMUN, V2, P1, DOI 10.3109/15419069409014197
   COHN JE, 1962, ANN INTERN MED, V56, P633, DOI 10.7326/0003-4819-56-4-633
   DEVEREAUX MW, 1973, ARCH NEUROL-CHICAGO, V29, P46, DOI 10.1001/archneur.1973.00490250064007
   DOOLING EC, 1977, ARCH OPHTHALMOL-CHIC, V95, P1790
   FIELDING JW, 1975, J BONE JOINT SURG AM, V57, P1000, DOI 10.2106/00004623-197557070-00026
   FOLGERING H, 1979, B EUR PHYSIOPATH RES, V15, P659
   Gestreau C, 2005, RESP PHYSIOL NEUROBI, V147, P159, DOI 10.1016/j.resp.2005.03.015
   GIANGASPERO F, 1988, CLIN NEUROPATHOL, V7, P234
   GUTHRIE TC, 1952, ANN INTERN MED, V37, P1197, DOI 10.7326/0003-4819-37-6-1197
   HANAWAY J, 1971, J NEUROPATH EXP NEUR, V30, P380, DOI 10.1097/00005072-197107000-00006
   Harper RM, 2000, BRAIN RES BULL, V53, P125, DOI 10.1016/S0361-9230(00)00317-8
   His W., 1891, ABHANDLUNGEN KONI MP, V29, P1
   Hui SHL, 2000, CHEST, V118, P266, DOI 10.1378/chest.118.1.266
   Ioos C, 2001, J CHILD NEUROL, V16, P767, DOI 10.1177/088307380101601012
   JENSEN TH, 1988, ACTA NEUROL SCAND, V77, P505, DOI 10.1111/j.1600-0404.1988.tb05949.x
   Kalimo H, 2004, FORENSIC SCI INT, V146, P73, DOI 10.1016/j.forsciint.2004.06.022
   Kraus J, 1999, CLIN NEUROL NEUROSUR, V101, P196, DOI 10.1016/S0303-8467(99)00023-2
   Kuhn M, 1999, RESPIRATION, V66, P265, DOI 10.1159/000029388
   Kumar R, 2005, J COMP NEUROL, V487, P361, DOI 10.1002/cne.20565
   LOCKWOOD AH, 1976, ARCH NEUROL-CHICAGO, V33, P292, DOI 10.1001/archneur.1976.00500040076013
   Macchi V, 2002, J ANAT, V200, P195, DOI 10.1046/j.0021-8782.2001.00013.x
   Manconi M, 2003, ARCH NEUROL-CHICAGO, V60, P1787, DOI 10.1001/archneur.60.12.1787
   Manning HL, 2000, AM J RESP CRIT CARE, V161, P2100, DOI 10.1164/ajrccm.161.6.9904087
   Matschke Jakob, 1999, Archiv fuer Kriminologie, V203, P108
   McKay LC, 2005, NAT NEUROSCI, V8, P1142, DOI 10.1038/nn1517
   MITO T, 1992, PEDIATR NEUROSURG, V17, P80, DOI 10.1159/000120572
   Morrell MJ, 2001, RESP PHYSIOL, V129, P269, DOI 10.1016/S0034-5687(01)00296-1
   NEILDWYER G, 1994, ACTA NEUROCHIR, V131, P137, DOI 10.1007/BF01401464
   PHILIPPSON EA, 2000, TXB RESP MED
   PLUM F, 1958, ARCH NEURO PSYCHIATR, V80, P267, DOI 10.1001/archneurpsyc.1958.02340090003001
   RIZVI SS, 1974, AM J MED, V56, P433, DOI 10.1016/0002-9343(74)90472-0
   Sadler M, 2002, J NEUROL NEUROSUR PS, V73, P347, DOI 10.1136/jnnp.73.3.347
   SARNOFF SJ, 1951, JAMA-J AM MED ASSOC, V147, P30, DOI 10.1001/jama.1951.03670180036007
   SCHLAEFKE ME, 1979, PFLUG ARCH EUR J PHY, V378, P231, DOI 10.1007/BF00592741
   SOLLIDAY NH, 1974, RESPIRATION, V31, P177, DOI 10.1159/000193108
   STANKIEWICZ JA, 1989, OTOLARYNG HEAD NECK, V101, P611, DOI 10.1177/019459988910100522
   Stewart W, 2004, FORENSIC SCI INT, V146, P125, DOI 10.1016/j.forsciint.2004.06.025
   STRIEDER DJ, 1967, AM REV RESPIR DIS, V96, P501
   SUGAR O, 1978, JAMA-J AM MED ASSOC, V240, P236, DOI 10.1001/jama.240.3.236
   TENICELA R, 1968, ANESTHESIOLOGY, V29, P7, DOI 10.1097/00000542-196801000-00006
   TOOLE JF, 1989, HDB CLIN NEUROLOGY, V55, P1
   TRANMER BI, 1987, CAN J NEUROL SCI, V14, P262, DOI 10.1017/S0317167100026585
   VALENTE S, 1993, CHEST, V103, P1909, DOI 10.1378/chest.103.6.1909
NR 51
TC 5
Z9 7
U1 0
U2 1
PU LIPPINCOTT WILLIAMS & WILKINS
PI PHILADELPHIA
PA TWO COMMERCE SQ, 2001 MARKET ST, PHILADELPHIA, PA 19103 USA
SN 0195-7910
EI 1533-404X
J9 AM J FOREN MED PATH
JI Am. J. Forensic Med. Pathol.
PD JUN
PY 2007
VL 28
IS 2
BP 141
EP 144
DI 10.1097/01.paf.0000257396.79742.e9
PG 4
WC Medicine, Legal; Pathology
WE Science Citation Index Expanded (SCI-EXPANDED)
SC Legal Medicine; Pathology
GA 173JF
UT WOS:000246868600011
PM 17525565
DA 2023-08-27
ER

PT J
AU Elsaafien, K
   Korim, WS
   Setiadi, A
   May, CN
   Yao, ST
AF Elsaafien, Khalid
   Korim, Willian S.
   Setiadi, Anthony
   May, Clive N.
   Yao, Song T.
TI Chemoattraction and Recruitment of Activated Immune Cells, Central
   Autonomic Control, and Blood Pressure Regulation
SO FRONTIERS IN PHYSIOLOGY
LA English
DT Review
DE neuroinflammation; chemoattraction; immune system; autonomic nervous
   system; hypertension
ID ROSTRAL VENTROLATERAL MEDULLA; CHEMOKINE RECEPTOR 2; PARAVENTRICULAR
   NUCLEUS; CARDIAC-HYPERTROPHY; PROINFLAMMATORY CYTOKINES; VASCULAR
   INFLAMMATION; SYMPATHETIC OUTFLOW; PROTEIN-1 RECEPTOR; SUBFORNICAL
   ORGAN; CHRONIC INFUSION
AB Inflammatory mediators play a critical role in the regulation of sympathetic outflow to cardiovascular organs in hypertension. Emerging evidence highlights the involvement of immune cells in the regulation of blood pressure. However, it is still unclear how these immune cells are activated and recruited to key autonomic brain regions to regulate sympathetic outflow to cardiovascular organs. Chemokines such as C-C motif chemokine ligand 2 (CCL2), and pro-inflammatory cytokines such as tumor necrosis factor alpha (TNF-alpha) and interleukin 1 beta (IL-1 beta), are upregulated both peripherally and centrally in hypertension. More specifically, they are upregulated in key autonomic brain regions that control sympathetic activity and blood pressure such as the paraventricular nucleus of the hypothalamus and the rostral ventrolateral medulla. Furthermore, this upregulation of inflammatory mediators is associated with the infiltration of immune cells to these brain areas. Thus, expression of pro-inflammatory chemokines and cytokines is a potential mechanism promoting invasion of immune cells into key autonomic brain regions. In pathophysiological conditions, this can result in abnormal activation of brain circuits that control sympathetic nerve activity to cardiovascular organs and ultimately in increases in blood pressure. In this review, we discuss emerging evidence that helps explain how immune cells are chemoattracted to autonomic nuclei and contribute to changes in sympathetic outflow and blood pressure.
C1 [Elsaafien, Khalid; Korim, Willian S.; Setiadi, Anthony; May, Clive N.; Yao, Song T.] Florey Inst Neurosci & Mental Hlth, Discovery Sci, Parkville, Vic, Australia.
   [Yao, Song T.] Univ Melbourne, Florey Dept Neurosci & Mental Hlth, Parkville, Vic, Australia.
C3 Florey Institute of Neuroscience & Mental Health; University of
   Melbourne
RP Yao, ST (通讯作者)，Florey Inst Neurosci & Mental Hlth, Discovery Sci, Parkville, Vic, Australia.; Yao, ST (通讯作者)，Univ Melbourne, Florey Dept Neurosci & Mental Hlth, Parkville, Vic, Australia.
EM
RI Elsaafien, Khalid/AAX-3697-2021; Stefanadis, Christodoulos/ABH-2232-2020
OI Elsaafien, Khalid/0000-0001-9581-918X; Stefanadis,
   Christodoulos/0000-0001-5974-6454; Korim, Willian/0000-0002-8104-7163
FU National Health and Medical Research Council of Australia [GNT 1079680,
   GNT 1128108]; High Blood Pressure Research Council of Australia; Rebecca
   L Cooper Medical Foundation; Victorian Government through the
   Operational Infrastructure Scheme; Australian Government Research
   Training Program Scholarships; Australian Research Council Future
   Fellowship [FT170100363]; Australian Research Council [FT170100363]
   Funding Source: Australian Research Council
FX This work was supported by the National Health and Medical Research
   Council of Australia (GNT 1079680 to SY; GNT 1128108 to CM and SY), the
   High Blood Pressure Research Council of Australia, the Rebecca L Cooper
   Medical Foundation (WK), and the Victorian Government through the
   Operational Infrastructure Scheme. KE and AS are supported by the
   Australian Government Research Training Program Scholarships. SY is
   supported by an Australian Research Council Future Fellowship
   (FT170100363).
CR Abukar Y, 2018, BASIC RES CARDIOL, V113, DOI 10.1007/s00395-018-0695-9
   Aiyar N, 1999, MOL CELL BIOCHEM, V197, P179, DOI 10.1023/A:1006962221332
   Ajuebor MN, 1998, J LEUKOCYTE BIOL, V63, P108, DOI 10.1002/jlb.63.1.108
   Antonelli A, 2012, INT J IMMUNOPATH PH, V25, P387, DOI 10.1177/039463201202500208
   Banisadr G, 2005, J COMP NEUROL, V489, P275, DOI 10.1002/cne.20598
   Bartoli C, 2001, ACTA NEUROPATHOL, V102, P385
   Begley DJ, 2003, PROG DRUG RES, V61, P39
   Biancardi VC, 2014, HYPERTENSION, V63, P572, DOI 10.1161/HYPERTENSIONAHA.113.01743
   Bomfim GF, 2019, BRIT J PHARMACOL, V176, P2028, DOI 10.1111/bph.14436
   Braga VA, 2011, BRAZ J MED BIOL RES, V44, P871, DOI 10.1590/S0100-879X2011000900007
   BROWN DL, 1985, CIRC RES, V56, P359
   Bush E, 2000, HYPERTENSION, V36, P360, DOI 10.1161/01.HYP.36.3.360
   Caillon A, 2019, BRIT J PHARMACOL, V176, P1818, DOI 10.1111/bph.14427
   Carnagarin R, 2019, BRIT J PHARMACOL, V176, P1839, DOI 10.1111/bph.14481
   Chan CT, 2012, HYPERTENSION, V60, P1207, DOI 10.1161/HYPERTENSIONAHA.112.201251
   Chang AYW, 2014, NEUROBIOL DIS, V71, P292, DOI 10.1016/j.nbd.2014.08.005
   Chen XL, 1998, CIRC RES, V83, P952
   Chrysohoou C, 2004, AM J HYPERTENS, V17, P568, DOI 10.1016/j.amjhyper.2004.03.675
   Crowley SD, 2006, P NATL ACAD SCI USA, V103, P17985, DOI 10.1073/pnas.0605545103
   Crowley SD, 2010, HYPERTENSION, V55, P99, DOI 10.1161/HYPERTENSIONAHA.109.144964
   DAMPNEY RAL, 1987, J AUTONOM NERV SYST, V20, P73, DOI 10.1016/0165-1838(87)90083-X
   Deshmane SL, 2009, J INTERF CYTOK RES, V29, P313, DOI 10.1089/jir.2008.0027
   Elmarakby AA, 2007, HYPERTENSION, V50, P1069, DOI 10.1161/HYPERTENSIONAHA.107.098806
   Gao HL, 2016, SCI REP-UK, V6, DOI 10.1038/srep30301
   Goldblatt H, 1934, J EXP MED, V59, P347, DOI 10.1084/jem.59.3.347
   Hinojosa AE, 2011, BRAIN RES BULL, V86, P326, DOI 10.1016/j.brainresbull.2011.09.014
   Ishibashi M, 2004, CIRC RES, V94, P1203, DOI 10.1161/01.RES.0000126924.23467.A3
   Jia LL, 2014, PLOS ONE, V9, DOI 10.1371/journal.pone.0085481
   Kang YM, 2014, TOXICOL APPL PHARM, V274, P436, DOI 10.1016/j.taap.2013.12.001
   Kang YM, 2009, CARDIOVASC RES, V82, P503, DOI 10.1093/cvr/cvp073
   Korim WS, 2019, CARDIOVASC RES, V115, P1092, DOI 10.1093/cvr/cvy268
   Li HB, 2016, TOXICOL APPL PHARM, V305, P93, DOI 10.1016/j.taap.2016.06.014
   Li HB, 2014, TOXICOL APPL PHARM, V279, P141, DOI 10.1016/j.taap.2014.06.004
   Lu B, 1998, J EXP MED, V187, P601, DOI 10.1084/jem.187.4.601
   Lu P., 2017, PFLUGERS ARCH, V470, P1, DOI [10.1007/500424-017-2081-0, DOI 10.1007/500424-017-2081-0]
   Marketou ME, 2011, AM J CARDIOL, V107, P59, DOI 10.1016/j.amjcard.2010.08.048
   Marvar PJ, 2010, CIRC RES, V107, P263, DOI 10.1161/CIRCRESAHA.110.217299
   Matsuda S, 2015, J ATHEROSCLER THROMB, V22, P833, DOI 10.5551/jat.27292
   Morioka N, 2013, NEUROPHARMACOLOGY, V75, P116, DOI 10.1016/j.neuropharm.2013.07.017
   MULLER WA, 1993, J EXP MED, V178, P449, DOI 10.1084/jem.178.2.449
   Nunes FC, 2011, J RENIN-ANGIO-ALDO S, V12, P440, DOI 10.1177/1470320310394891
   Roberts TK, 2012, LAB INVEST, V92, P1213, DOI 10.1038/labinvest.2012.80
   Rollins BJ, 1997, BLOOD, V90, P909, DOI 10.1182/blood.V90.3.909.909_909_928
   Santisteban MM, 2015, CIRC RES, V117, P178, DOI 10.1161/CIRCRESAHA.117.305853
   SCHRAMM LP, 1993, BRAIN RES, V616, P251, DOI 10.1016/0006-8993(93)90216-A
   Setiadi A, 2018, EXP PHYSIOL, V103, P337, DOI 10.1113/EP086434
   Sharma RK, 2019, CIRC RES, V124, P727, DOI 10.1161/CIRCRESAHA.118.313882
   Shen XZ, 2015, HYPERTENSION, V66, P309, DOI 10.1161/HYPERTENSIONAHA.115.05333
   Shi P, 2010, HYPERTENSION, V56, P297, DOI 10.1161/HYPERTENSIONAHA.110.150409
   Shi Z, 2011, ACTA PHYSIOL, V203, P289, DOI 10.1111/j.1748-1716.2011.02313.x
   Song XA, 2014, TOXICOL APPL PHARM, V281, P101, DOI 10.1016/j.taap.2014.09.004
   Sriramula S, 2013, PLOS ONE, V8, DOI 10.1371/journal.pone.0063847
   Stamatovic SM, 2003, J CELL SCI, V116, P4615, DOI 10.1242/jcs.00755
   Stamatovic SM, 2009, J BIOL CHEM, V284, P19053, DOI 10.1074/jbc.M109.000521
   STRACK AM, 1989, BRAIN RES, V491, P274, DOI 10.1016/0006-8993(89)90063-2
   STRACK AM, 1989, BRAIN RES, V491, P156, DOI 10.1016/0006-8993(89)90098-X
   Su Q, 2014, TOXICOL APPL PHARM, V276, P115, DOI 10.1016/j.taap.2014.02.002
   Tsou CL, 2007, J CLIN INVEST, V117, P902, DOI 10.1172/JCI29919
   VANDERKOOY D, 1983, J COMP NEUROL, V219, P328, DOI 10.1002/cne.902190307
   Wang ML, 2018, J NEUROINFLAMM, V15, DOI 10.1186/s12974-018-1131-7
   Wang YP, 2015, EXP BIOL MED, V240, P1223, DOI 10.1177/1535370214565970
   Wei SG, 2015, HYPERTENSION, V65, P1126, DOI 10.1161/HYPERTENSIONAHA.114.05112
   Wittendorp MC, 2004, GLIA, V46, P410, DOI 10.1002/glia.20016
   Xie QY, 2006, INT J CARDIOL, V110, P60, DOI 10.1016/j.ijcard.2005.07.046
   YOSHIMURA T, 1989, FEBS LETT, V244, P487, DOI 10.1016/0014-5793(89)80590-3
   YOSHIMURA T, 1989, J IMMUNOL, V142, P1956
   Yu XJ, 2015, TOXICOL APPL PHARM, V284, P315, DOI 10.1016/j.taap.2015.02.023
   Zlotnik A, 2012, IMMUNITY, V36, P705, DOI 10.1016/j.immuni.2012.05.008
   Zubcevic J, 2017, FRONT PHYSIOL, V8, DOI 10.3389/fphys.2017.00592
NR 69
TC 11
Z9 12
U1 0
U2 5
PU FRONTIERS MEDIA SA
PI LAUSANNE
PA AVENUE DU TRIBUNAL FEDERAL 34, LAUSANNE, CH-1015, SWITZERLAND
SN 1664-042X
J9 FRONT PHYSIOL
JI Front. Physiol.
PD AUG 2
PY 2019
VL 10
AR 984
DI 10.3389/fphys.2019.00984
PG 7
WC Physiology
WE Science Citation Index Expanded (SCI-EXPANDED)
SC Physiology
GA IN3TX
UT WOS:000478600300002
PM 31427987
OA Green Published, gold
DA 2023-08-27
ER

PT J
AU Becker, J
   Wilting, J
AF Becker, Juergen
   Wilting, Joerg
TI WNT signaling, the development of the sympathoadrenal-paraganglionic
   system and neuroblastoma
SO CELLULAR AND MOLECULAR LIFE SCIENCES
LA English
DT Review
DE Neuroblastoma; Sympathoadrenal system; Paraganglion; Neural crest
ID NEURAL CREST CELLS; ADRENAL CHROMAFFIN CELLS; HIGH-RISK NEUROBLASTOMAS;
   INTERNATIONAL CRITERIA; SYMPATHETIC NEURONS; SUSTENTACULAR CELLS;
   NEURITE OUTGROWTH; TUMOR-SUPPRESSOR; RETINOIC ACID; DIFFERENTIATION
AB Neuroblastoma (NB) is a tumor of the sympathoadrenal system arising in children under 15 years of age. In Germany, NB accounts for 7% of childhood cancer cases, but 11% of cancer deaths. It originates from highly migratory progenitor cells that leave the dorsal neural tube and contribute neurons and glial cells to sympathetic ganglia, and chromaffin and supportive cells to the adrenal medulla and paraganglia. Clinically, histologically and molecularly, NBs present as extremely heterogeneous, ranging from very good to very poor prognosis. The etiology of NB still remains unclear and needs to be elucidated, however, aberrant auto- and paracrine embryonic cell communications seem to be likely candidates to initiate or facilitate the emergence, progression and regression of NB. The wingless-type MMTV integration site (WNT) family of proteins represents an evolutionary highly conserved signaling system that orchestrates embryogenesis. At least 19 ligands in the human, numerous receptors and co-receptors are known, which control not only proliferation, but also cell polarity, migration and differentiation. Here we seek to interconnect aspects of WNT signaling with sympathoadrenal and paraganglionic development to define new WNT signaling cues in the etiology and progression of NB.
C1 [Becker, Juergen; Wilting, Joerg] Univ Med Sch Gottingen, Inst Anat & Cell Biol, D-37075 Gottingen, Germany.
RP Becker, J (通讯作者)，Univ Med Sch Gottingen, Inst Anat & Cell Biol, D-37075 Gottingen, Germany.
EM
OI Wilting, Jorg/0000-0001-8337-2296
CR Anastas JN, 2013, NAT REV CANCER, V13, P11, DOI 10.1038/nrc3419
   Armstrong A, 2011, J NEUROSCI, V31, P2371, DOI 10.1523/JNEUROSCI.4243-10.2011
   Becker J, 2010, CLIN CANCER RES, V16, P1431, DOI 10.1158/1078-0432.CCR-09-1936
   BECKWITH JB, 1963, AM J PATHOL, V43, P1089
   Bilir A, 2010, J NEURO-ONCOL, V100, P385, DOI 10.1007/s11060-010-0209-6
   Blanc E, 2005, ONCOGENE, V24, P1277, DOI 10.1038/sj.onc.1208255
   Bodmer D, 2009, J NEUROSCI, V29, P7569, DOI 10.1523/JNEUROSCI.1445-09.2009
   BRODEUR GM, 1993, J CLIN ONCOL, V11, P1466, DOI 10.1200/JCO.1993.11.8.1466
   BRODEUR GM, 1988, J CLIN ONCOL, V6, P1874, DOI 10.1200/JCO.1988.6.12.1874
   Brodeur GM, 2003, NAT REV CANCER, V3, P203, DOI 10.1038/nrc1014
   Cantilena S, 2011, ONCOTARGET, V2, P976, DOI 10.18632/oncotarget.410
   Carmon KS, 2012, MOL CELL BIOL, V32, P2054, DOI 10.1128/MCB.00272-12
   CHRIST B, 1992, ANN ANAT, V174, P23, DOI 10.1016/S0940-9602(11)80337-7
   CICCARONE V, 1989, CANCER RES, V49, P219
   Cimmino F, 2016, J PROTEOME RES, V15, P3643, DOI 10.1021/acs.jproteome.6b00457
   Cimmino F, 2015, SCI REP-UK, V5, DOI 10.1038/srep11158
   Cohn SL, 2009, J CLIN ONCOL, V27, P289, DOI 10.1200/JCO.2008.16.6785
   Coulon A, 2011, NEOPLASIA, V13, P991, DOI 10.1593/neo.11800
   Davy A, 2007, DEV BIOL, V304, P182, DOI 10.1016/j.ydbio.2006.12.028
   De Brouwer S, 2012, INT J CANCER, V130, P2591, DOI 10.1002/ijc.26295
   De Calisto J, 2005, DEVELOPMENT, V132, P2587, DOI 10.1242/dev.01857
   Deng C, 2013, J CELL SCI, V126, P2060, DOI 10.1242/jcs.123471
   Diaz-Flores L, 2008, ACTA PHYSIOL, V192, P145, DOI 10.1111/j.1748-1716.2007.01811.x
   Duffy DJ, 2017, GENOME MED, V9, DOI 10.1186/s13073-017-0407-3
   Duffy DJ, 2016, ONCOTARGET, V7, P60310, DOI 10.18632/oncotarget.11203
   Duffy DJ, 2015, ONCOTARGET, V6, P43182, DOI 10.18632/oncotarget.6568
   Duffy DJ, 2014, MOL CANCER THER, V13, P454, DOI 10.1158/1535-7163.MCT-13-0560-T
   Dyberg C, 2016, BMC CANCER, V16, DOI 10.1186/s12885-016-2293-2
   Eggert A, 2000, KLIN PADIATR, V212, P200, DOI 10.1055/s-2000-9677
   Eggert A, 2000, ONCOGENE, V19, P2043, DOI 10.1038/sj.onc.1203518
   Eleveld TF, 2015, NAT GENET, V47, P864, DOI 10.1038/ng.3333
   Fagan AM, 1996, J NEUROSCI, V16, P6208
   Fan SS, 2004, MOL BRAIN RES, V132, P38, DOI 10.1016/j.molbrainres.2004.09.003
   Flahaut M, 2009, ONCOGENE, V28, P2245, DOI 10.1038/onc.2009.80
   Forgham H, 2015, FRONT CELL DEV BIOL, V3, DOI 10.3389/fcell.2015.00077
   Francis N, 1999, DEV BIOL, V210, P411, DOI 10.1006/dbio.1999.9269
   Granchi D, 2011, BONE, V48, P152, DOI 10.1016/j.bone.2010.06.028
   Gray JD, 2013, HUM MOL GENET, V22, P4267, DOI 10.1093/hmg/ddt277
   Green J, 2014, CSH PERSPECT BIOL, V6, DOI 10.1101/cshperspect.a009175
   Grumolato L, 2010, GENE DEV, V24, P2517, DOI 10.1101/gad.1957710
   Hall A, 1998, SCIENCE, V279, P509, DOI 10.1126/science.279.5350.509
   Harenza JL, 2017, SCI DATA, V4, DOI 10.1038/sdata.2017.33
   Harris ML, 2007, DEV DYNAM, V236, P1, DOI 10.1002/dvdy.20919
   Haug BH, 2011, CARCINOGENESIS, V32, P1005, DOI 10.1093/carcin/bgr073
   Hero B, 2008, J CLIN ONCOL, V26, P1504, DOI 10.1200/JCO.2007.12.3349
   Holmquist-Mengelbier L, 2006, CANCER CELL, V10, P413, DOI 10.1016/j.ccr.2006.08.026
   Howard MJ, 2005, DEV BIOL, V277, P271, DOI 10.1016/j.ydbio.2004.09.034
   Hu BL, 2016, CELL, V167, P1281, DOI 10.1016/j.cell.2016.10.039
   Huang SMA, 2009, NATURE, V461, P614, DOI 10.1038/nature08356
   Huber K, 2006, DEV BIOL, V298, P335, DOI 10.1016/j.ydbio.2006.07.010
   Huber K, 2015, CELL TISSUE RES, V359, P333, DOI 10.1007/s00441-014-1947-0
   Huber K, 2008, NEURAL DEV, V3, DOI 10.1186/1749-8104-3-28
   Huret JL, 2013, NUCLEIC ACIDS RES, V41, pD920, DOI 10.1093/nar/gks1082
   Iehara T, 2012, EUR J CANCER, V48, P1707, DOI 10.1016/j.ejca.2012.01.010
   IWANAGA T, 1984, CELL TISSUE RES, V236, P733
   Jurney WM, 2002, J NEUROSCI, V22, P6019
   Kaatsch P, 2016, ACTA ONCOL, V55, P1145, DOI 10.1080/0284186X.2016.1195509
   Kahn M, 2014, NAT REV DRUG DISCOV, V13, P513, DOI 10.1038/nrd4233
   Kasemeier-Kulesa JC, 2006, DEVELOPMENT, V133, P4839, DOI 10.1242/dev.02662
   Klaus A, 2008, NAT REV CANCER, V8, P387, DOI 10.1038/nrc2389
   Koppen A, 2008, INT J CANCER, V122, P1455, DOI 10.1002/ijc.23180
   Koppen A, 2007, CANCER LETT, V256, P218, DOI 10.1016/j.canlet.2007.06.011
   Krull CE, 1997, CURR BIOL, V7, P571, DOI 10.1016/S0960-9822(06)00256-9
   Kumawat K, 2016, CELL MOL LIFE SCI, V73, P567, DOI 10.1007/s00018-015-2076-y
   Kummer W, 2010, ANATOMIE, P766
   Kuriyama S, 2008, PHILOS T R SOC B, V363, P1349, DOI 10.1098/rstb.2007.2252
   L'Episcopo F, 2014, J MOL CELL BIOL, V6, P13, DOI 10.1093/jmcb/mjt053
   LaBonne C, 1998, DEVELOPMENT, V125, P2403
   LEBLANC G, 1986, J NEUROSCI, V6, P260
   LEDOUARIN NM, 1975, P NATL ACAD SCI USA, V72, P728
   LEDOUARINM, 1974, DEV BIOL, V41, P162, DOI DOI 10.1016/0012-1606(74)90291-7
   Leonhardt H, 1985, TASCHENLEHRBUCH GESA, V3
   Liu JP, 1998, DEVELOPMENT, V125, P5055
   Liu X, 2008, ONCOGENE, V27, P1478, DOI 10.1038/sj.onc.1210769
   Lu B, 2016, CANCERS, V8, DOI 10.3390/cancers8090082
   Lumb R, 2015, DEV GROWTH DIFFER, V57, P146, DOI 10.1111/dgd.12189
   Maj E, 2016, DEV BIOL, V417, P77, DOI 10.1016/j.ydbio.2016.06.022
   Maris JM, 2007, LANCET, V369, P2106, DOI 10.1016/S0140-6736(07)60983-0
   Maris JM, 2010, NEW ENGL J MED, V362, P2202, DOI 10.1056/NEJMra0804577
   Matthay KK, 1999, NEW ENGL J MED, V341, P1165, DOI 10.1056/NEJM199910143411601
   Mikels AJ, 2006, ONCOGENE, V25, P7461, DOI 10.1038/sj.onc.1210053
   Mikels AJ, 2006, PLOS BIOL, V4, P570, DOI 10.1371/journal.pbio.0040115
   Monclair T, 2009, J CLIN ONCOL, V27, P298, DOI 10.1200/JCO.2008.16.6876
   Nakagawara A, 2004, PROG BRAIN RES, V146, P233, DOI 10.1016/S0079-6123(03)46015-9
   Niehrs C, 2012, NAT REV MOL CELL BIO, V13, P767, DOI 10.1038/nrm3470
   NIETO MA, 1994, SCIENCE, V264, P835, DOI 10.1126/science.7513443
   Nusse R, 2012, EMBO J, V31, P2670, DOI 10.1038/emboj.2012.146
   OHEA TP, 1952, J PHYSIOL-LONDON, V118, pP1
   Ora I, 2011, SEMIN CANCER BIOL, V21, P217, DOI 10.1016/j.semcancer.2011.07.002
   Orme MH, 2003, MOL CELL NEUROSCI, V24, P673, DOI 10.1016/S1044-7431(03)00229-X
   Padovan-Merhar OM, 2016, PLOS GENET, V12, DOI 10.1371/journal.pgen.1006501
   Pietras A, 2009, P NATL ACAD SCI USA, V106, P16805, DOI 10.1073/pnas.0904606106
   Qi C, 2017, CELL MOL BIOL, V63, P12, DOI 10.14715/cmb/2017.63.7.2
   Revet I, 2010, CANCER LETT, V289, P195, DOI 10.1016/j.canlet.2009.08.019
   Ruffner H, 2012, PLOS ONE, V7, DOI 10.1371/journal.pone.0040976
   Ryu YK, 2013, DEV BIOL, V377, P79, DOI 10.1016/j.ydbio.2013.02.013
   Saito D, 2005, MECH DEVELOP, V122, P59
   Schilling FH, 2002, NEW ENGL J MED, V346, P1047, DOI 10.1056/NEJMoa012277
   Schleiermacher G, 2014, INT J CANCER, V135, P2249, DOI 10.1002/ijc.29077
   SCHRODER HD, 1986, HISTOPATHOLOGY, V10, P1023, DOI 10.1111/j.1365-2559.1986.tb02539.x
   Schulte JH, 2013, ONCOGENE, V32, P1059, DOI 10.1038/onc.2012.106
   Shellard A, 2016, SEMIN CELL DEV BIOL, V55, P111, DOI 10.1016/j.semcdb.2016.01.031
   SHIMADA H, 1984, J NATL CANCER I, V73, P405, DOI 10.1093/jnci/73.2.405
   Stuhlmiller TJ, 2012, CELL MOL LIFE SCI, V69, P3715, DOI 10.1007/s00018-012-0991-8
   Subashini C, 2017, SCI REP-UK, V7, DOI 10.1038/srep42523
   Suebsoonthron J, 2017, ANTI-CANCER DRUG, V28, P469, DOI 10.1097/CAD.0000000000000478
   Thiele CJ, 1999, HUMAN CELL CULTURE, V1, P21
   Tian XH, 2013, J EXP CLIN CANC RES, V32, DOI 10.1186/1756-9966-32-100
   TSOKOS M, 1987, AM J PATHOL, V128, P484
   Tucker RP, 1999, DEV DYNAM, V214, P312, DOI 10.1002/(SICI)1097-0177(199904)214:4<312::AID-AJA4>3.0.CO;2-A
   UNSICKER K, 1978, BRAIN RES, V152, P334, DOI 10.1016/0006-8993(78)90260-3
   Unsicker K, 2005, NEUROCHEM RES, V30, P921, DOI 10.1007/s11064-005-6966-5
   Vibert L, 2017, PIGM CELL MELANOMA R, V30, P219, DOI 10.1111/pcmr.12568
   Vieira GC, 2015, ONCOTARGET, V6, P40053, DOI 10.18632/oncotarget.5548
   VOGEL KS, 1990, DEV BIOL, V139, P1, DOI 10.1016/0012-1606(90)90273-L
   Wakamatsu Y, 2000, DEVELOPMENT, V127, P2811
   Wan WB, 2014, BIOMED RES INT, V2014, DOI 10.1155/2014/301575
   Welsch U, 2010, ANATOMIE, P378
   Westermann F, 2002, CANCER LETT, V184, P127, DOI 10.1016/S0304-3835(02)00199-4
   WESTON JC, 1963, ANAT REC, V145, P355
   Yang YZ, 2012, CELL BIOSCI, V2, DOI 10.1186/2045-3701-2-14
   Zhan T, 2017, ONCOGENE, V36, P1461, DOI 10.1038/onc.2016.304
   Zhang J, 2014, CANCER LETT, V348, P12, DOI 10.1016/j.canlet.2014.02.011
   Zins K, 2016, ONCOTARGET, V7, P46187, DOI 10.18632/oncotarget.10070
NR 124
TC 24
Z9 29
U1 1
U2 5
PU SPRINGER BASEL AG
PI BASEL
PA PICASSOPLATZ 4, BASEL, 4052, SWITZERLAND
SN 1420-682X
EI 1420-9071
J9 CELL MOL LIFE SCI
JI Cell. Mol. Life Sci.
PD MAR
PY 2018
VL 75
IS 6
BP 1057
EP 1070
DI 10.1007/s00018-017-2685-8
PG 14
WC Biochemistry & Molecular Biology; Cell Biology
WE Science Citation Index Expanded (SCI-EXPANDED)
SC Biochemistry & Molecular Biology; Cell Biology
GA FW4LM
UT WOS:000425285100008
PM 29058015
OA hybrid, Green Published
DA 2023-08-27
ER

PT J
AU Ono, S
   Saito, T
   Terui, K
   Yoshida, H
   Enomoto, H
AF Ono, Sachie
   Saito, Takeshi
   Terui, Keita
   Yoshida, Hideo
   Enomoto, Hideki
TI Generation of conditional ALK F1174L mutant mouse models for the study
   of neuroblastoma pathogenesis
SO GENESIS
LA English
DT Article
DE ALK; conditional mutation; model mouse; MYCN; neuroblastoma
ID RECEPTOR TYROSINE KINASE; ANAPLASTIC LYMPHOMA KINASE; ALK; MYCN;
   EXPRESSION; MUTATIONS; BIOLOGY; RET
AB Neuroblastoma, an embryonal tumor arising from the sympathetic ganglia and adrenal medulla, is among the most intractable pediatric cancers. Although a variety of genetic changes have been identified in neuroblastoma, how they contribute to its pathogenesis remains largely unclear. Recent studies have identified alterations of the anaplastic lymphoma kinase (ALK) gene in neuroblastoma; ALK F1174L (a phenylalanine-to-leucine substitution at codon 1174) represents one of the most frequent of these somatic mutations, and is associated with amplification of the MYCN gene, the most reliable marker for the poor survival. We engineered the mouse Alk locus so that ALK F1174L is expressed by its endogenous promoter and can be induced in a spatiotemporally controlled fashion using Cre-loxP system. Although expression of ALK F1174L resulted in enhanced proliferation of sympathetic ganglion progenitors and increased the size of the sympathetic ganglia, it was insufficient to cause neuroblastoma. However, lethal neuroblastoma frequently developed in mice co-expressing ALK F1174L and MYCN, even in a genetic background where MYCN alone does not cause overt tumors. These data reveal that physiological expression of ALK F1174L significantly potentiates the oncogenic ability of MYCN in vivo. Our conditional mutant mice provide a valuable platform for investigating the pathogenesis of neuroblastoma.
C1 [Ono, Sachie; Enomoto, Hideki] RIKEN Ctr Dev Biol, Lab Neuronal Differentiat & Regenerat, Kobe, Hyogo, Japan.
   [Ono, Sachie; Saito, Takeshi; Terui, Keita; Yoshida, Hideo] Chiba Univ, Grad Sch Med, Dept Pediat Surg, Chiba, Japan.
   [Enomoto, Hideki] Kobe Univ, Div Neural Differentiat & Regenerat, Dept Physiol & Cell Biol, Grad Sch Med, Kobe, Hyogo, Japan.
C3 RIKEN; Chiba University; Kobe University
RP Enomoto, H (通讯作者)，Kobe Univ, Div Neural Differentiat & Regenerat, Dept Physiol & Cell Biol, Grad Sch Med,Chuo Ku, 7-5-1 Kusunokicho, Kobe, Hyogo 6500017, Japan.
EM
OI Terui, Keita/0000-0003-4315-5179
CR Alam G, 2009, AM J PATHOL, V175, P856, DOI 10.2353/ajpath.2009.090019
   Berry T, 2012, CANCER CELL, V22, P117, DOI 10.1016/j.ccr.2012.06.001
   Brodeur GM, 2003, NAT REV CANCER, V3, P203, DOI 10.1038/nrc1014
   Chen YY, 2008, NATURE, V455, P971, DOI 10.1038/nature07399
   Chiarle R, 2008, NAT REV CANCER, V8, P11, DOI 10.1038/nrc2291
   De Brouwer S, 2010, CLIN CANCER RES, V16, P4353, DOI 10.1158/1078-0432.CCR-09-2660
   Enomoto H, 2004, NEURON, V44, P623, DOI 10.1016/j.neuron.2004.10.032
   Enomoto H, 2001, DEVELOPMENT, V128, P3963
   Enomoto H, 1998, NEURON, V21, P317, DOI 10.1016/S0896-6273(00)80541-3
   Gonsalvez DG, 2013, J NEUROSCI, V33, P5969, DOI 10.1523/JNEUROSCI.4350-12.2013
   Haase GM, 1999, SEMIN SURG ONCOL, V16, P91, DOI 10.1002/(SICI)1098-2388(199903)16:2<91::AID-SSU3>3.0.CO;2-1
   Iwahara T, 1997, ONCOGENE, V14, P439, DOI 10.1038/sj.onc.1200849
   Jiang MR, 2011, CURR TOP DEV BIOL, V94, P77, DOI 10.1016/B978-0-12-380916-2.00004-8
   Maris JM, 1999, J CLIN ONCOL, V17, P2264, DOI 10.1200/JCO.1999.17.7.2264
   Maris JM, 2010, NEW ENGL J MED, V362, P2202, DOI 10.1056/NEJMra0804577
   Moog-Lutz C, 2005, J BIOL CHEM, V280, P26039, DOI 10.1074/jbc.M501972200
   Mosse YP, 2008, NATURE, V455, P930, DOI 10.1038/nature07261
   Nagashimada M, 2012, J CLIN INVEST, V122, P3145, DOI 10.1172/JCI63401
   Raabe EH, 2008, ONCOGENE, V27, P469, DOI 10.1038/sj.onc.1210659
   Takahashi Y, 2013, SCIENCE, V341, P860, DOI 10.1126/science.1230717
   Vernersson E, 2006, GENE EXPR PATTERNS, V6, P448, DOI 10.1016/j.modgep.2005.11.006
   Weiss WA, 1997, EMBO J, V16, P2985, DOI 10.1093/emboj/16.11.2985
   Yamauchi Y, 1999, DEV BIOL, V212, P191, DOI 10.1006/dbio.1999.9323
   Zhu SZ, 2012, CANCER CELL, V21, P362, DOI 10.1016/j.ccr.2012.02.010
NR 24
TC 8
Z9 8
U1 1
U2 4
PU WILEY
PI HOBOKEN
PA 111 RIVER ST, HOBOKEN 07030-5774, NJ USA
SN 1526-954X
EI 1526-968X
J9 GENESIS
JI Genesis
PD OCT
PY 2019
VL 57
IS 10
AR e23323
DI 10.1002/dvg.23323
PG 8
WC Developmental Biology; Genetics & Heredity
WE Science Citation Index Expanded (SCI-EXPANDED)
SC Developmental Biology; Genetics & Heredity
GA JD0VX
UT WOS:000489695700006
PM 31218818
DA 2023-08-27
ER

PT J
AU Inoue, J
   Fujita, H
   Bando, T
   Kondo, Y
   Kumon, H
   Ohuchi, H
AF Inoue, Junji
   Fujita, Hirofumi
   Bando, Tetsuya
   Kondo, Yoichi
   Kumon, Hiromi
   Ohuchi, Hideyo
TI Expression analysis of Dickkopf-related protein 3 (Dkk3) suggests its
   pleiotropic roles for a secretory glycoprotein in adult mouse
SO JOURNAL OF MOLECULAR HISTOLOGY
LA English
DT Article
DE Dkk3; REIC; In situ hybridization; Immunohistochemistry; Mouse
ID SQUAMOUS-CELL CARCINOMA; PROSTATE-CANCER CELLS; DOWN-REGULATION;
   BETA-CATENIN; REIC/DKK-3; OVEREXPRESSION; FAMILY; CONTRIBUTES;
   APOPTOSIS; PATHWAY
AB Dickkopf-related protein 3 (Dkk3) is the third member of the Dkk gene family and identical to the gene, whose expression was reduced in immortalized cells. Therefore, its another name is reduced expression in immortalized cells. Since the intratumoral introduction of Dkk3 inhibits tumor growth in mouse models of cancers, Dkk3 is likely a tumor suppressor gene. However, the functions of Dkk3 in vivo remain unclear. As the first step to decipher the physiological roles of this gene, we examined the expression pattern of Dkk3 in various tissues from adult mice. In situ hybridization showed that Dkk3 mRNA was detected in the brain, retina, heart, gastrointestinal tract, adrenal glands, thymus, prostate glands, seminal vesicles, testes, and ovaries in a regionally specific manner. Furthermore, we raised anti-mouse Dkk3 antibody and performed immunohistochemistry. Cytoplasmic localization of Dkk3 protein was observed in the cells of the adrenal medulla, while Dkk3 immunoreactivity was observed in the lumen of the stomach and intestine, implying that the Dkk3 protein may be secreted into the lumen of the gastrointestinal tract. These results suggest that Dkk3 has pleiotropic roles for a secretory glycoprotein that acts primarily in the gastrointestinal tract, thymus, endocrine and reproductive organs of the mouse.
C1 [Inoue, Junji; Fujita, Hirofumi; Bando, Tetsuya; Kondo, Yoichi; Ohuchi, Hideyo] Okayama Univ, Grad Sch Med Dent & Pharmaceut Sci, Dept Cytol & Histol, Kita Ku, 2-5-1 Shikata Cho, Okayama 7008558, Japan.
   [Kumon, Hiromi] Okayama Univ, Innovat Ctr Okayama Nanobiotargeted Therapy ICONT, Kita Ku, 2-5-1 Shikata Cho, Okayama 7008558, Japan.
   [Kumon, Hiromi] Niimi Coll, 1263-2 Nishigata, Niimi, Okayama 7188585, Japan.
   [Kondo, Yoichi] Osaka Med Coll, Dept Anat & Cell Biol, 2-7 Daigakumachi, Takatsuki, Osaka 5698686, Japan.
C3 Okayama University; Okayama University; Osaka Medical College; Osaka
   Medical & Pharmaceutical University
RP Ohuchi, H (通讯作者)，Okayama Univ, Grad Sch Med Dent & Pharmaceut Sci, Dept Cytol & Histol, Kita Ku, 2-5-1 Shikata Cho, Okayama 7008558, Japan.
EM
FU grant for Promotion of Science and Technology in Okayama Prefecture by
   Ministry of Education, Culture, Sports, Science and Technology of Japan;
   Grants-in-Aid for Scientific Research [15K10475, 26293352] Funding
   Source: KAKEN
FX The authors thank Professor Takahisa Furukawa (Osaka University) for his
   permission to use the Dkk3 KO mouse. This study was supported by a grant
   for Promotion of Science and Technology in Okayama Prefecture by the
   Ministry of Education, Culture, Sports, Science and Technology of Japan.
CR Abarzua F, 2005, CANCER RES, V65, P9617, DOI 10.1158/0008-5472.CAN-05-0829
   Bafico A, 2001, NAT CELL BIOL, V3, P683, DOI 10.1038/35083081
   Bamberger C, 2005, WOUND REPAIR REGEN, V13, P41, DOI 10.1111/j.1067-1927.2005.130106.x
   Barrantes ID, 2006, MOL CELL BIOL, V26, P2317, DOI 10.1128/MCB.26.6.2317-2326.2006
   Byun T, 2005, J CLIN PATHOL, V58, P515, DOI 10.1136/jcp.2004.018598
   Das DS, 2013, PLOS ONE, V8, DOI 10.1371/journal.pone.0063603
   Du G, 2011, EXP DERMATOL, V20, P273, DOI 10.1111/j.1600-0625.2010.01244.x
   Fujii M, 2011, J MOL HISTOL, V42, P499, DOI 10.1007/s10735-011-9357-z
   Fujii Y, 2014, ACTA MED OKAYAMA, V68, P63
   Glinka A, 1998, NATURE, V391, P357, DOI 10.1038/34848
   Hara K, 2015, CANCER LETT, V356, P496, DOI 10.1016/j.canlet.2014.09.045
   Kataoka K, 2012, CLIN EXP DERMATOL, V37, P428, DOI 10.1111/j.1365-2230.2011.04301.x
   Krupnik VE, 1999, GENE, V238, P301, DOI 10.1016/S0378-1119(99)00365-0
   Lee EJ, 2009, INT J CANCER, V124, P287, DOI 10.1002/ijc.23913
   Li SJ, 2011, J MOL HISTOL, V42, P491, DOI 10.1007/s10735-011-9347-1
   Mao BY, 2003, GENE, V302, P179, DOI 10.1016/S0378-1119(02)01106-X
   Monaghan AP, 1999, MECH DEVELOP, V87, P45, DOI 10.1016/S0925-4773(99)00138-0
   Niehrs C, 2006, ONCOGENE, V25, P7469, DOI 10.1038/sj.onc.1210054
   Ochiai K, 2011, BIOCHEM BIOPH RES CO, V412, P391, DOI 10.1016/j.bbrc.2011.07.109
   Papatriantafyllou M, 2012, P NATL ACAD SCI USA, V109, P1631, DOI 10.1073/pnas.1115980109
   Ryu SW, 2013, INT J GYNECOL CANCER, V23, P134, DOI 10.1097/IGC.0b013e3182754feb
   SCHAERENWIEMERS N, 1993, HISTOCHEMISTRY, V100, P431, DOI 10.1007/BF00267823
   Suwa T, 2003, J ENDOCRINOL, V178, P149, DOI 10.1677/joe.0.1780149
   Tanimoto R, 2007, INT J MOL MED, V19, P363
   Tanimoto R, 2010, INT J CANCER, V126, P1562, DOI 10.1002/ijc.24764
   Tsuji T, 2000, BIOCHEM BIOPH RES CO, V268, P20, DOI 10.1006/bbrc.1999.2067
   Veeck J, 2012, BBA-REV CANCER, V1825, P18, DOI 10.1016/j.bbcan.2011.09.003
   Veeck J, 2009, BMC CANCER, V9, DOI 10.1186/1471-2407-9-217
   Wang Z, 2015, ONCOL REP, V33, P1097, DOI 10.3892/or.2014.3704
   Watanabe M, 2009, INT J ONCOL, V34, P657, DOI 10.3892/ijo_00000191
   Xiang TX, 2013, J CELL MOL MED, V17, P1236, DOI 10.1111/jcmm.12099
   Zhou GR, 2016, J MOL HISTOL, V47, P287, DOI 10.1007/s10735-016-9674-3
NR 32
TC 5
Z9 5
U1 2
U2 4
PU SPRINGER
PI DORDRECHT
PA VAN GODEWIJCKSTRAAT 30, 3311 GZ DORDRECHT, NETHERLANDS
SN 1567-2379
EI 1567-2387
J9 J MOL HISTOL
JI J. Mol. Histol.
PD FEB
PY 2017
VL 48
IS 1
BP 29
EP 39
DI 10.1007/s10735-016-9703-2
PG 11
WC Cell Biology
WE Science Citation Index Expanded (SCI-EXPANDED)
SC Cell Biology
GA EL1HN
UT WOS:000394371200003
PM 27866302
OA Green Submitted
DA 2023-08-27
ER

PT J
AU Yeh, SHH
   Lin, MH
   Kong, FL
   Chang, CW
   Hwang, LC
   Lin, CF
   Hwang, JJ
   Liu, RS
AF Yeh, Skye Hsin-Hsien
   Lin, Ming-Hsien
   Kong, Fan-Lin
   Chang, Chi-Wei
   Hwang, Li-Chung
   Lin, Chien-Feng
   Hwang, Jeng-Jong
   Liu, Ren-Shyan
TI Evaluation of Inhibitory Effect of Recreational Drugs on Dopaminergic
   Terminal Neuron by PET and Whole-Body Autoradiography
SO BIOMED RESEARCH INTERNATIONAL
LA English
DT Article
ID POSITRON-EMISSION-TOMOGRAPHY; IN-VIVO; NUCLEUS-ACCUMBENS; STRIATAL
   DOPAMINE; RAT-BRAIN; KETAMINE; RECEPTORS; COCAINE; MODEL; MICE
AB There is little investigation for the functional roles of peripheral dopamine. [F-18] FDOPA has been used in cancer imaging (i.e., neuroendocrine and tumors pancreatic tumors) and neuroimaging (i.e., Parkinson's disease and Huntington's disease). Here, we accessed side effects of recreational drugs such as ketamine, cocaine, and methamphetamine on dopamine neurons in peripheral organs by using positron emission tomography (PET) imaging and quantitative whole-body autoradiography (QWBAR) with [F-18] FDOPA. The images were applied for the measurement of specific binding ratios (SBRs) of striatum with the cerebellum as the reference region. Clear striatal [F-18] FDOPA-derived radioactivity was observed. Moderate level of radiotracer accumulation was presented in the mucosal layers of the stomach and small intestine. The medulla layers of kidney had higher radioactivity than that of the cortex. Blocking images markedly eliminated the specific binding of [F-18] FDOPA in the striatum and in peripheral organs such as stomachs, intestines, and kidney. Ketamine showed the highest inhibitory effect on striatal [F-18] FDOPA-derived radioactivity followed by cocaine and methamphetamine. The current results demonstrated a useful crossing-validating tool that enhances the capability of [F-18] FDOPA for further investigations of the alteration of dopaminergic neurons in the brain disorder or cancer diseases in peripheral tissues.
C1 [Yeh, Skye Hsin-Hsien; Lin, Ming-Hsien; Chang, Chi-Wei; Hwang, Li-Chung; Hwang, Jeng-Jong; Liu, Ren-Shyan] Natl Yang Ming Univ, Dept Biomed Imaging & Radiol Sci, Taipei 11221, Taiwan.
   [Yeh, Skye Hsin-Hsien] Taipei City Hosp, Dept Educ & Res, Taipei 10341, Taiwan.
   [Yeh, Skye Hsin-Hsien; Liu, Ren-Shyan] Natl Yang Ming Univ, Biophoton & Mol Imaging Res Ctr, Taipei 11221, Taiwan.
   [Lin, Ming-Hsien] Taipei City Hosp, Zhongxiao Branch, Div Nucl Med, Taipei 11556, Taiwan.
   [Kong, Fan-Lin] Univ Texas MD Anderson Canc Ctr, Dept Canc Syst Imaging, Houston, TX USA.
   [Chang, Chi-Wei; Liu, Ren-Shyan] Natl Comprehens Mouse Phenotyping & Drug Testing, Taiwan Mouse Clin, Mol & Genet Imaging Core, Taipei 11533, Taiwan.
   [Chang, Chi-Wei; Liu, Ren-Shyan] Taipei Vet Gen Hosp, Natl PET Cyclotron Ctr, Taipei 11221, Taiwan.
   [Lin, Chien-Feng] Taipei Vet Gen Hosp, Dept Nucl Med, Taipei 11221, Taiwan.
   [Lin, Chien-Feng] Taipei Med Univ, Shuang Ho Hosp, Minist Hlth & Welf, Nucl Med Dept, New Taipei City 23561, Taiwan.
   [Lin, Chien-Feng] Taipei Med Univ, Shuang Ho Hosp, Minist Hlth & Welf, PET CT Ctr, New Taipei City 23561, Taiwan.
   [Liu, Ren-Shyan] Natl Yang Ming Univ, Inst Clin Med, Taipei 11221, Taiwan.
C3 National Yang Ming Chiao Tung University; Taipei City Hospital; National
   Yang Ming Chiao Tung University; Taipei City Hospital; University of
   Texas System; UTMD Anderson Cancer Center; Taipei Veterans General
   Hospital; Taipei Veterans General Hospital; Taipei Medical University;
   Taipei Medical University; National Yang Ming Chiao Tung University
RP Hwang, JJ (通讯作者)，Natl Yang Ming Univ, Dept Biomed Imaging & Radiol Sci, Taipei 11221, Taiwan.
EM
FU Taipei City Hospital, Taiwan [102TPECH13]; National Science Council,
   Taiwan [NSC 91-2314-B010-078, NSC100-2923-B-010-001-MY3]; Taipei
   Veterans General Hospital [V102E3-007]; Molecular and Genetic Imaging
   Core, Taiwan Mouse Clinic (NSC) [102-2325-B-001-042]; National Research
   Program for Biopharmaceuticals (NRPB) at the National Science Council
   (NSC) of Taiwan
FX This research was supported by the Grants: 102TPECH13 (Taipei City
   Hospital, Taiwan), NSC 91-2314-B010-078, NSC100-2923-B-010-001-MY3
   (National Science Council, Taiwan), and V102E3-007 (Taipei Veterans
   General Hospital). The authors thank the technical support from
   Molecular and Genetic Imaging Core, Taiwan Mouse Clinic (NSC
   102-2325-B-001-042) which is funded by the National Research Program for
   Biopharmaceuticals (NRPB) at the National Science Council (NSC) of
   Taiwan.
CR Autry AE, 2011, NATURE, V475, P91, DOI 10.1038/nature10130
   Barr AM, 2006, J PSYCHIATR NEUROSCI, V31, P301
   Chang CW, 2000, NUCL MED COMMUN, V21, P799, DOI 10.1097/00006231-200009000-00003
   Fang CK, 2013, APPL RADIAT ISOTOPES, V77, P153, DOI 10.1016/j.apradiso.2013.03.091
   Hume SP, 1996, J NEUROSCI METH, V67, P103
   Hunt MJ, 2005, J NEUROSCI, V25, P524, DOI 10.1523/JNEUROSCI.3800-04.2005
   Imperiale A, 2013, J CLIN ENDOCR METAB, V98, P2643, DOI 10.1210/jc.2013-1942
   Ishiwata K, 1996, NUCL MED BIOL, V23, P295, DOI 10.1016/0969-8051(95)02083-7
   Ishiwata K, 2002, NUCL MED BIOL, V29, P307, DOI 10.1016/S0969-8051(01)00307-9
   Jokinen P, 2013, J NEUROL SCI, V329, P23, DOI 10.1016/j.jns.2013.03.006
   Kyono K, 2011, EJNMMI RES, V1, DOI 10.1186/2191-219X-1-25
   Lin KH, 2002, NUCL MED BIOL, V29, P643, DOI 10.1016/S0969-8051(02)00323-2
   Martiniova L, 2012, NUCL MED BIOL, V39, P215, DOI 10.1016/j.nucmedbio.2011.07.007
   Mateo Y, 2004, P NATL ACAD SCI USA, V101, P372, DOI 10.1073/pnas.0207805101
   Murray RM, 2013, AM J MED GENET B, V162, P661, DOI 10.1002/ajmg.b.32177
   National Institute on Drug Abuse (NIDA), 2006, METH AB ADD
   Nikolaus S, 2001, J NUCL MED, V42, P1691
   Paxinos CW, 1998, RAT BRAIN STEREOTAXI
   Reus GZ, 2013, BEHAV BRAIN RES, V256, P451, DOI 10.1016/j.bbr.2013.08.041
   Selby NM, 2008, CLIN KIDNEY J, V1, P310, DOI 10.1093/ndtplus/sfn054
   Shewan D, 1996, BRIT MED J, V313, P424
   Thanos PK, 2002, J NUCL MED, V43, P1570
   Tsukada H, 1996, J NEUROSCI, V16, P7670
   Tsukada H, 2000, BRAIN RES, V860, P141, DOI 10.1016/S0006-8993(00)02057-6
   Tuomela J, 2013, EJNMMI RES, V3, DOI 10.1186/2191-219X-3-18
   Usun Y, 2013, BEHAV BRAIN RES, V256, P229, DOI 10.1016/j.bbr.2013.08.017
   Vollenweider FX, 2000, J PSYCHIATR RES, V34, P35, DOI 10.1016/S0022-3956(99)00031-X
   Walker MD, 2013, J CEREBR BLOOD F MET, V33, P59, DOI 10.1038/jcbfm.2012.120
   Wang CM, 2013, FRONT NEUROANAT, V7, DOI 10.3389/fnana.2013.00023
   Wilder-Smith OHG, 1998, ANESTH ANALG, V86, P95, DOI 10.1097/00000539-199801000-00019
   Wright LKM, 2007, NEUROTOXICOL TERATOL, V29, P348, DOI 10.1016/j.ntt.2006.12.004
   Yamanaka H, 2014, TRANSL PSYCHIAT, V4, DOI 10.1038/tp.2013.112
   Yeh HH, 2011, P NATL ACAD SCI USA, V108, P1603, DOI 10.1073/pnas.1010744108
   Yentis S. M., 2003, ANAESTHESIA INTENSIV
   Yeung LY, 2009, TOXICOL LETT, V191, P275, DOI 10.1016/j.toxlet.2009.09.006
NR 35
TC 7
Z9 7
U1 0
U2 7
PU HINDAWI LTD
PI LONDON
PA ADAM HOUSE, 3RD FLR, 1 FITZROY SQ, LONDON, W1T 5HF, ENGLAND
SN 2314-6133
EI 2314-6141
J9 BIOMED RES INT
JI Biomed Res. Int.
PY 2014
VL 2014
AR 157923
DI 10.1155/2014/157923
PG 11
WC Biotechnology & Applied Microbiology; Medicine, Research & Experimental
WE Science Citation Index Expanded (SCI-EXPANDED)
SC Biotechnology & Applied Microbiology; Research & Experimental Medicine
GA AG8PX
UT WOS:000335681800001
PM 24877059
OA gold, Green Submitted, Green Published
DA 2023-08-27
ER

PT J
AU Stich, O
   Klages, E
   Bischler, P
   Jarius, S
   Rasiah, C
   Voltz, R
   Rauer, S
AF Stich, O.
   Klages, E.
   Bischler, P.
   Jarius, S.
   Rasiah, C.
   Voltz, R.
   Rauer, S.
TI SOX1 antibodies in sera from patients with paraneoplastic neurological
   syndromes
SO ACTA NEUROLOGICA SCANDINAVICA
LA English
DT Article
DE antiglial nuclear antibodies; antineuronal antibodies; cancer;
   coexisting antibodies; malignancy; paraneoplastic neurological
   syndromes; SOX1
ID EATON MYASTHENIC SYNDROME; CELL LUNG-CANCER; CEREBROSPINAL-FLUID; TUMOR;
   FREQUENCY; MARKERS
AB Objectives SOX1 antibodies have been described in patients with LambertEaton myasthenic syndrome (LEMS) in association with voltage-gated calcium channel antibodies as serological markers of small cell lung cancer (SCLC). This study was aimed to screen for additional SOX1 autoimmunity in onconeural antibody-positive sera from patients with paraneoplastic neurological syndromes (PNS) other than LEMS and to identify the clinicalimmunological profile and associated tumours of patients with coexisting SOX1 antibodies. Methods We retrospectively analysed sera from 55 patients with different PNS positive for well-characterized antineuronal antibodies for the presence of SOX1 antibodies by recombinant ELISA and immunoblot. Results Eight (14.5%) patients showed additional SOX1 antibodies in the ELISA and the recombinant immunoblot. Five patients had coexisting Hu antibodies, while the other three showed coexisting CV2/CRMP5, amphiphysin, and coexisting CV2/CRMP5 and Hu antibodies, respectively. PNS included (partially overlapping) subacute sensory neuropathy, subacute sensorimotor neuropathy, cerebellar degeneration, brainstem encephalitis, encephalomyelitis and limbic encephalitis. No tumour was detected in two patients, while the others had lung cancer (four SCLC and two non-SCLC). One patient showed SOX1-specific intrathecal antibody synthesis. Conclusions We describe SOX1 reactivity for the first time overlapping with CV2/CRMP5 and amphiphysin antibodies. SOX1 reactivity is predominantly associated with Hu antibodies and SCLC, but can occur also in other types of lung cancer. Neurological manifestations present in patients with coexisting SOX1 antibodies and well-characterized antineuronal antibodies do not differ from those previously described in patients positive for antineuronal antibodies but no SOX1-specific anti-glial antibodies.
C1 [Stich, O.; Klages, E.; Bischler, P.; Rauer, S.] Univ Freiburg, Dept Neurol, D-79106 Freiburg, Germany.
   [Jarius, S.] Heidelberg Univ, Dept Neurol, Div Mol Neuroimmunol, Heidelberg, Germany.
   [Rasiah, C.; Rauer, S.] Ravo Diagnost GmbH, Freiburg, Germany.
   [Voltz, R.] Univ Cologne, Dept Palliat Med, D-50931 Cologne, Germany.
C3 University of Freiburg; 4EU+; Ruprecht Karls University Heidelberg;
   University of Cologne
RP Stich, O (通讯作者)，Univ Freiburg, Dept Neurol, Breisacher Str 64, D-79106 Freiburg, Germany.
EM
CR Graus F, 2005, J NEUROIMMUNOL, V165, P166, DOI 10.1016/j.jneuroim.2005.03.020
   Graus F, 2004, J NEUROL NEUROSUR PS, V75, P1135, DOI 10.1136/jnnp.2003.034447
   Graus F, 2010, J NEUROL, V257, P509, DOI 10.1007/s00415-009-5431-9
   Gure AO, 2000, P NATL ACAD SCI USA, V97, P4198, DOI 10.1073/pnas.97.8.4198
   Linnoila RI, 1996, J CELL BIOCHEM, P92
   Pittock SJ, 2004, ANN NEUROL, V56, P715, DOI 10.1002/ana.20269
   Reiber H, 2001, J NEUROL SCI, V184, P101, DOI 10.1016/S0022-510X(00)00501-3
   Sabater L, 2008, NEUROLOGY, V70, P924, DOI 10.1212/01.wnl.0000281663.81079.24
   Stich O, 2007, J NEUROIMMUNOL, V183, P220, DOI 10.1016/j.jneuroim.2006.11.008
   Titulaer MJ, 2008, ANN NY ACAD SCI, V1132, P129, DOI 10.1196/annals.1405.030
   Titulaer MJ, 2009, J CLIN ONCOL, V27, P4260, DOI 10.1200/JCO.2008.20.6169
   Tschernatsch M, 2010, J NEUROIMMUNOL, V226, P177, DOI 10.1016/j.jneuroim.2010.07.005
   Vural B, 2005, CANCER-AM CANCER SOC, V103, P2575, DOI 10.1002/cncr.21088
   Zuliani L, 2007, J NEUROL NEUROSUR PS, V78, P204, DOI 10.1136/jnnp.2006.102467
NR 14
TC 25
Z9 26
U1 0
U2 7
PU WILEY
PI HOBOKEN
PA 111 RIVER ST, HOBOKEN 07030-5774, NJ USA
SN 0001-6314
EI 1600-0404
J9 ACTA NEUROL SCAND
JI Acta Neurol. Scand.
PD MAY
PY 2012
VL 125
IS 5
BP 326
EP 331
DI 10.1111/j.1600-0404.2011.01572.x
PG 6
WC Clinical Neurology
WE Science Citation Index Expanded (SCI-EXPANDED)
SC Neurosciences & Neurology
GA 919SB
UT WOS:000302347800008
PM 21751968
OA gold
DA 2023-08-27
ER

PT J
AU Sweetser, DA
   Froelick, GJ
   Matsumoto, AM
   Kafer, KE
   Marck, B
   Palmiter, RD
   Kapur, RP
AF Sweetser, DA
   Froelick, GJ
   Matsumoto, AM
   Kafer, KE
   Marck, B
   Palmiter, RD
   Kapur, RP
TI Ganglioneuromas and renal anomalies are induced by activated RETMEN2B in
   transgenic mice
SO ONCOGENE
LA English
DT Article
DE ganglioneuroma; ret proto-oncogene; multiple endocrine neoplasia; renal
   agenesis; neuroblastoma; transgenic; autonomic nervous system
ID MEDULLARY-THYROID CARCINOMA; TYROSINE KINASE DOMAIN; ENTERIC
   NERVOUS-SYSTEM; NEOPLASIA TYPE 2B; RET PROTOONCOGENE; CELL-LINE; C-RET;
   SYMPATHOADRENAL LINEAGE; SYMPATHETIC NEUROBLASTS; KIDNEY DEVELOPMENT
