## Supplemental Material 1 for "Bibliometric analysis of emerging trends and research foci in brainstem tumor field over thirty years (1992-2023)": savedrecs (9).txt

FN Clarivate Analytics Web of Science
VR 1.0
PT J
AU Schaller, B
   Probst, R
   Strebel, S
   Gratzl, O
AF Schaller, B
   Probst, R
   Strebel, S
   Gratzl, O
TI Trigeminocardiac reflex during surgery in the cerebellopontine angle
SO JOURNAL OF NEUROSURGERY
LA English
DT Article; Proceedings Paper
CT 3rd Congress of the European-Association-for-Neuro-Oncology
CY SEP 13-16, 1998
CL VERSAILLES, FRANCE
SP European Assoc Neuro Oncol
DE cerebellopontine angle; trigeminocardiac reflex; trigeminal nerve;
   bradycardia; hypotension
ID TRIGEMINAL DEPRESSOR RESPONSE; ZYGOMATIC ARCH FRACTURE; OCULOCARDIAC
   REFLEX; PERCUTANEOUS MICROCOMPRESSION; GASSERIAN GANGLION; BRADYCARDIA;
   NEURALGIA; ASYSTOLE; PROJECTIONS; ELEVATION
AB Object.In different experimental studies authors have analyzed the autonomic responses elicited by the electrical, mechanical, or chemical stimulation of the trigeminal nerve system. The trigeminocardiac reflex (TCR) is a well-recognized phenomenon that consists of bradycardia, arterial hypotension, apnea, and gastric hypermotility. It occurs during ocular surgery and during other manipulations in and around the orbit. Thus far, it has not been shown that central stimulation of the trigeminal nerve can also cause this reflex.
   Methods. The TCR was defined as clinical hypotension with a drop in mean arterial blood pressure (MABP) of more than 20% and bradycardia lower than 60 beats/minute. Pre-, intra-, and postoperative heart rate (HR) and MABP were reviewed retrospectively in 125 patients who underwent surgery for tumors of the cerebellopontine angle (CPA), and they were divided into two groups on the basis of the occurrence of the TCR during surgery. Of the 125 patients, 14 (11%) showed evidence of the TCR during dissection of the tumor near the trigeminal nerve at the brainstem. Their HRs fell 38% and their MABPs fell 48% during operative procedures as compared with pre-operative levels. After cessation of manipulation, the HRs and the MABPs returned to preoperative levels. Risk factors for the occurrence of the TCR were compared with results from the literature.
   Conclusions. The authors' results show the possibility of occurrence of a TCR during manipulation of the central part of the trigeminal nerve when performing surgery in the CPA.
C1 Univ Basel Hosp, Dept Neurol Surg, CH-4031 Basel, Switzerland.
   Univ Basel Hosp, Dept Anaesthesia, CH-4031 Basel, Switzerland.
   Univ Basel Hosp, Dept Otorhinolaryngol, CH-4031 Basel, Switzerland.
C3 University of Basel; University of Basel; University of Basel
RP Probst, R (通讯作者)，Univ Basel Hosp, Dept Otolaryngol, CH-4031 Basel, Switzerland.
RI Schaller, Bernhard/AAA-4398-2019; Probst, Rudolf Robert/HKV-5170-2023
OI Schaller, Bernhard/0000-0002-7710-8090; Probst, Rudolf
   Robert/0000-0001-5778-3905
CR ALEXANDER JP, 1975, BRIT J OPHTHALMOL, V59, P518, DOI 10.1136/bjo.59.9.518
   ANDERSEN HT, 1966, PHYSIOL REV, V46, P212, DOI 10.1152/physrev.1966.46.2.212
   ANDERSON RL, 1978, ARCH OPHTHALMOL-CHIC, V96, P1418
   ANGELL JE, 1969, P ROY SOC MED, V62, P1287, DOI 10.1177/003591576906201238
   ANGELLJAMES JE, 1975, ACTA OTO-LARYNGOL, V79, P242, DOI 10.3109/00016487509124680
   BAINTON R, 1987, ORAL SURG ORAL MED O, V64, P24, DOI 10.1016/0030-4220(87)90110-1
   BAINTON R, 1990, BRIT J ORAL MAX SURG, V28, P109, DOI 10.1016/0266-4356(90)90134-7
   BARNARD NA, 1990, J CRANIO MAXILL SURG, V18, P359, DOI 10.1016/S1010-5182(05)80056-3
   BELBER CJ, 1987, NEUROSURGERY, V20, P908, DOI 10.1227/00006123-198706000-00015
   BIETTI GB, 1973, STRABISMUS, V13, P727
   BLANC VF, 1983, CAN ANAESTH SOC J, V30, P360, DOI 10.1007/BF03007858
   BLANC VF, 1991, CAN J ANAESTH, V38, P696, DOI 10.1007/BF03008444
   BOSOMWORTH P P, 1958, Anesthesiology, V19, P7, DOI 10.1097/00000542-195801000-00002
   BROWN JA, 1988, NEUROSURGERY, V23, P745, DOI 10.1227/00006123-198812000-00010
   CHESLEY LD, 1989, J ORAL MAXIL SURG, V47, P522, DOI 10.1016/0278-2391(89)90290-5
   DOMINGUEZ J, 1994, NEUROSURGERY, V34, P422, DOI 10.1227/00006123-199403000-00006
   HOPKINS CS, 1988, ANAESTHESIA, V43, P157
   KERR WJ, 1983, ANAESTHESIA, V38, P833
   KLEIHUES P, 1993, HISTOLOGICAL TYPING, P30
   Kratschmer F, 1870, SBER AKAD WISS WIEN, V62, P147
   KUMADA M, 1977, BRAIN RES, V119, P305, DOI 10.1016/0006-8993(77)90313-4
   LANG S, 1991, CAN J ANAESTH, V38, P757, DOI 10.1007/BF03008454
   LEVY MN, 1979, CARDIOVASCULAR SYSTE, V1, P581
   LOBATO RD, 1990, J NEUROSURG, V72, P546, DOI 10.3171/jns.1990.72.4.0546
   LOEWINGER J, 1987, J ORAL MAXIL SURG, V45, P710, DOI 10.1016/0278-2391(87)90315-6
   MENETREY D, 1987, J COMP NEUROL, V255, P439, DOI 10.1002/cne.902550310
   MIRAKHUR RK, 1982, BRIT J ANAESTH, V54, P1059, DOI 10.1093/bja/54.10.1059
   MOONIE G T, 1964, Can Anaesth Soc J, V11, P621, DOI 10.1007/BF03004107
   MULLAN S, 1983, J NEUROSURG, V59, P1007, DOI 10.3171/jns.1983.59.6.1007
   NEGRUTIU S, 1988, Revista de Chirurgie Oncologie Radiologie O R L Oftalmologie Stomatologie Seria Chirurgie, V37, P309
   PFALTZ CR, 1992, ACOUSTIC NEUROMA, P423
   PRECIOUS DS, 1990, INT J ORAL MAX SURG, V19, P279, DOI 10.1016/S0901-5027(05)80420-5
   RAGNO JR, 1989, J ORAL MAXIL SURG, V47, P1082, DOI 10.1016/0278-2391(89)90186-9
   RIVARD JC, 1988, ANAESTH ANALG, V76, P907
   ROBIDEAUX V, 1978, ANESTHESIOLOGY, V49, P433, DOI 10.1097/00000542-197812000-00011
   RUGGIERO DA, 1981, BRAIN RES, V225, P225, DOI 10.1016/0006-8993(81)90832-5
   SCHMELING WT, 1989, ANAESTH ANALG, V66, P4
   SCHWARTZ H, 1971, HIGHLIGHTS CLIN ANES, P111
   SHEARER ES, 1987, ANAESTHESIA, V42, P1207, DOI 10.1111/j.1365-2044.1987.tb05231.x
   SHELLY MP, 1988, ANAESTHESIA, V43, P422, DOI 10.1111/j.1365-2044.1988.tb09042.x
   SPAZIANTE R, 1988, NEUROSURGERY, V22, P148, DOI 10.1227/00006123-198801000-00028
   STARR NJ, 1986, ANESTHESIOLOGY, V64, P521, DOI 10.1097/00000542-198604000-00023
   STOTT DG, 1989, BRIT J PLAST SURG, V42, P595, DOI 10.1016/0007-1226(89)90053-2
   TENNEY SM, 1986, HDB PHYSL 3, V2, P833
   TERUI N, 1981, J AUTONOM NERV SYST, V4, P1, DOI 10.1016/0165-1838(81)90002-3
NR 45
TC 181
Z9 188
U1 1
U2 5
PU AMER ASSOC NEUROLOGICAL SURGEONS
PI CHARLOTTESVILLE
PA UNIV VIRGINIA, 1224 WEST MAIN ST, STE 450, CHARLOTTESVILLE, VA 22903 USA
SN 0022-3085
J9 J NEUROSURG
JI J. Neurosurg.
PD FEB
PY 1999
VL 90
IS 2
BP 215
EP 220
DI 10.3171/jns.1999.90.2.0215
PG 6
WC Clinical Neurology; Surgery
WE Conference Proceedings Citation Index - Science (CPCI-S); Science Citation Index Expanded (SCI-EXPANDED)
SC Neurosciences & Neurology; Surgery
GA 168RU
UT WOS:000078707100005
PM 9950491
DA 2023-08-27
ER

PT J
AU Merchant, TE
   Li, CH
   Xiong, XP
   Kun, LE
   Boop, FA
   Sanford, RA
AF Merchant, Thomas E.
   Li, Chenghong
   Xiong, Xiaoping
   Kun, Larry E.
   Boop, Frederic A.
   Sanford, Robert A.
TI Conformal radiotherapy after surgery for paediatric ependymoma: a
   prospective study
SO LANCET ONCOLOGY
LA English
DT Article
ID REQUIRING PROLONGED OBSERVATION; PROGNOSTIC-FACTORS; RADIATION-THERAPY;
   INTRACRANIAL EPENDYMOMA; LOCALIZED EPENDYMOMA; CHILDHOOD EPENDYMOMA;
   POSTOPERATIVE CHEMOTHERAPY; COMPETING RISK; BRAIN-TUMORS; CHILDREN
AB Background Therapy for ependymoma includes aggressive surgical intervention and radiotherapy administered by use of methods that keep the risk of side-effects to a minimum. We extended this treatment approach to include children under the age of 3 years with the aim of improving tumour control.
   Methods Between July 11, 1997, and Nov 18, 2007, 153 paediatric patients (median age 2.9 years [range 0.9-22.9 months]) with localised ependymoma were treated. 85 patients had anaplastic ependymoma; the tumours of 122 were located in the infratentorial region, and 35 had received previous chemotherapy. Patients received conformal radiotherapy after definitive surgery (125 patients had undergone gross total, 17 near total, and 11 subtotal resection). Doses of 59.4 Gy (n=131) or 54.0 Gy (n=22) were prescribed to a 10 mm margin around the target volume. Disease control, patterns of failure, and complications were recorded for patients followed over 10 years. Overall survival, event-free survival (EFS), cumulative incidence of local recurrences, and cumulative incidence of distant recurrences were assessed. Variables considered included tumour grade, tumour location, ethnic origin, sex, age when undergoing conformal radiotherapy, total radiotherapy dose, number of surgical procedures, surgery extent, and preradiotherapy chemotherapy.
   Findings After a median follow-up of 5.3 years (range 0.4-10.4),23 patients had died and tumour progression noted in 36, including local (n=14), distant (n=15), and combined failure (n=7). 7-year local control, EFS, and overall survival were 87.3% (95% CI 77.5-97.1), 69.1% (56.9-81.3), and 81.0% (71.0-91.0), respectively. The cumulative incidences of local and distant failure were 16.3% (9.6-23.0) and 11.5% (5.9-17.1), respectively. In the 107 patients treated with immediate postoperative conformal radiotherapy (without delay or chemotherapy), 7-year local control, EFS, and overall survival were 88.7% (77.9-99.5), 76.9% (63.4-90.4), and 85.0% (74.2-95.8), respectively; the cumulative incidence of local and distant failure were 12.6% (5.1-20.1), and 8.6% (2.8-14.3), respectively. The incidence of secondary malignant brain tumour at 7 years was 2.3% (0-5.6) and brainstem necrosis 1.6% (0-4.0). Overall survival was affected by tumour grade (anaplastic vs differentiated: HR 3.98 [95% CI 1.51-10.48]; p=0.0052), extent of resection (gross total vs near total or subtotal: 0.16 [0.07-0.37]; p < 0.0001), and ethnic origin (non-white vs white: 3.0 [1.21-7.44]; p=0.018). EFS was affected by tumour grade (anaplastic vs differentiated: 2.52 [1.27-5.01]; p=0.008), extent of resection (gross total vs near total or subtotal: 0.20 [0.11-0.39]; p < 0.0001]), and sex (male vs female: 2.19 [1.03-4.66]; p=0.042). Local failure was affected by extent of resection (gross total vs near total or subtotal: 0.16 [0.067-0.38]; p < 0.0001), sex (male vs female: 3.85 [1.10-13.52]; p=0.035), and age (< 3 years vs >= 3 years: 3.25 [1.30-8.16]; p=0.012). Distant recurrence was only affected by tumour grade (anaplastic vs differentiated: 4.1 [1.2-14.0]; p=0.017).
   Interpretation Treatment of ependymoma should include surgery with the aim of gross-total resection and conformal, high-dose, postoperative irradiation. Future trials might consider treatment stratification based on sex and age.
C1 [Merchant, Thomas E.; Kun, Larry E.; Boop, Frederic A.; Sanford, Robert A.] St Jude Childrens Res Hosp, Dept Radiol Sci, Memphis, TN 38105 USA.
   [Li, Chenghong; Xiong, Xiaoping] St Jude Childrens Res Hosp, Dept Biostat, Memphis, TN 38105 USA.
C3 St Jude Children's Research Hospital; St Jude Children's Research
   Hospital
RP Merchant, TE (通讯作者)，St Jude Childrens Res Hosp, Dept Radiol Sci, Mail Stop 220,262 Danny Thomas Pl, Memphis, TN 38105 USA.
EM
RI Lien, Brian/AAI-7882-2020; Merchant, Thomas E/N-3344-2018
OI Merchant, Thomas E/0000-0002-0412-6255
FU American Cancer Society and American Lebanese Syrian Associated
   Charities (ALSAC)
FX Funding American Cancer Society and American Lebanese Syrian Associated
   Charities (ALSAC).
CR AKYILZ C, 2000, ACTA ONCOL, V39, P97
   Bouffet E, 1998, MED PEDIATR ONCOL, V30, P319, DOI 10.1002/(SICI)1096-911X(199806)30:6<319::AID-MPO1>3.3.CO;2-3
   Conklin HM, 2008, J CLIN ONCOL, V26, P3965, DOI 10.1200/JCO.2007.15.9970
   COX DR, 1972, J R STAT SOC B, V34, P187
   Fine JP, 1999, J AM STAT ASSOC, V94, P496, DOI 10.2307/2670170
   GRAY RJ, 1988, ANN STAT, V16, P1141, DOI 10.1214/aos/1176350951
   Grill J, 2001, J CLIN ONCOL, V19, P1288, DOI 10.1200/JCO.2001.19.5.1288
   Grundy RG, 2007, LANCET ONCOL, V8, P696, DOI 10.1016/S1470-2045(07)70208-5
   Hua CH, 2008, INT J RADIAT ONCOL, V72, P892, DOI 10.1016/j.ijrobp.2008.01.050
   *ICRU, 1978, 50 ICRU
   Jaing TH, 2004, J NEURO-ONCOL, V68, P255, DOI 10.1023/B:NEON.0000033383.84900.c1
   Kalbfleisch JD, 1980, STATISTICALANALYSIS, P163
   KAPLAN EL, 1958, J AM STAT ASSOC, V53, P457, DOI 10.2307/2281868
   Kiehna EN, 2006, J CLIN ONCOL, V24, P5283, DOI 10.1200/JCO.2005.03.8547
   MacDonald SM, 2008, INT J RADIAT ONCOL, V71, P979, DOI 10.1016/j.ijrobp.2007.11.065
   Mansur DB, 2005, INT J RADIAT ONCOL, V61, P387, DOI 10.1016/j.ijrobp.2004.06.002
   MANTEL NATHAN, 1966, CANCERCHEMOTHERAP REP, V50, P163
   Massimino M, 2004, INT J RADIAT ONCOL, V58, P1336, DOI 10.1016/j.ijrobp.2003.08.030
   Merchant TE, 2004, J CLIN ONCOL, V22, P3156, DOI 10.1200/JCO.2004.11.142
   MERCHANT TE, 2008, INT J RADIAT ONCOL, V63, P1546
   Merchant TE, 2008, INT J RADIAT ONCOL, V71, P87, DOI 10.1016/j.ijrobp.2007.09.037
   Oya N, 2002, J NEURO-ONCOL, V56, P87, DOI 10.1023/A:1014442106111
   Perilongo G, 1997, MED PEDIATR ONCOL, V29, P79
   PETO R, 1977, BRIT J CANCER, V35, P1, DOI 10.1038/bjc.1977.1
   PETO R, 1976, BRIT J CANCER, V34, P585, DOI 10.1038/bjc.1976.220
   POLLACK IF, 1995, NEUROSURGERY, V37, P655, DOI 10.1227/00006123-199510000-00008
   Robertson PL, 1998, J NEUROSURG, V88, P695, DOI 10.3171/jns.1998.88.4.0695
   Schroeder TM, 2008, INT J RADIAT ONCOL, V71, P987, DOI 10.1016/j.ijrobp.2007.11.058
   Shu HKG, 2007, CANCER-AM CANCER SOC, V110, P432, DOI 10.1002/cncr.22782
   van Veelen-Vincent MLC, 2002, J NEUROSURG, V97, P827, DOI 10.3171/jns.2002.97.4.0827
   Zacharoulis S, 2007, PEDIATR BLOOD CANCER, V49, P34, DOI 10.1002/pbc.20935
NR 31
TC 336
Z9 358
U1 0
U2 9
PU ELSEVIER SCIENCE INC
PI NEW YORK
PA 360 PARK AVE SOUTH, NEW YORK, NY 10010-1710 USA
SN 1470-2045
EI 1474-5488
J9 LANCET ONCOL
JI Lancet Oncol.
PD MAR
PY 2009
VL 10
IS 3
BP 258
EP 266
DI 10.1016/S1470-2045(08)70342-5
PG 9
WC Oncology
WE Science Citation Index Expanded (SCI-EXPANDED)
SC Oncology
GA 421BF
UT WOS:000264332200020
PM 19274783
OA Green Accepted
DA 2023-08-27
ER

PT J
AU Langlois, AM
   Iorio-Morin, C
   Faramand, A
   Niranjan, A
   Lunsford, LD
   Mohammed, N
   Sheehan, JP
   Liscak, R
   Urgosik, D
   Kondziolka, D
   Lee, CC
   Yang, HC
   Atik, AF
   Mathieu, D
AF Langlois, Anne-Marie
   Iorio-Morin, Christian
   Faramand, Andrew
   Niranjan, Ajay
   Lunsford, L. Dade
   Mohammed, Nasser
   Sheehan, Jason P.
   Liscak, Roman
   Urgosik, Dusan
   Kondziolka, Douglas
   Lee, Cheng-chia
   Yang, Huai-che
   Atik, Ahmet F.
   Mathieu, David
TI Outcomes after stereotactic radiosurgery for schwannomas of the
   oculomotor, trochlear, and abducens nerves
SO JOURNAL OF NEUROSURGERY
LA English
DT Article
DE oculomotor; trochlear; abducens; schwannomas; Gamma Knife; stereotactic
   radiosurgery
ID GAMMA-KNIFE RADIOSURGERY; MANAGEMENT; SURGERY; SAFETY
AB OBJECTIVE Cranial nerve (CN) schwannomas are intracranial tumors that are commonly managed by stereotactic radiosurgery (SRS). There is a large body of literature supporting the use of SRS for vestibular schwannomas. Schwannomas of the oculomotor nerves (CNs III, IV, and VI) are rare skull base tumors, occurring close to the brainstem and often involving the cavernous sinus. Resection can cause significant morbidity, including loss of nerve function. As for other schwannomas, SRS can be used to manage these tumors, but only a handful of cases have been published so far, often among reports of other uncommon schwannoma locations.
   METHODS The goal of this study was to collect retrospective multicenter data on tumor control, clinical evolution, and morbidity after SRS. This study was performed through the International Radiosurgery Research Foundation. Patients managed with single-session SRS for an oculomotor cranial nerve schwannoma (CN III, IV, or VI) were included. The diagnosis was based on diplopia or ptosis as the main presenting symptom and anatomical location on the trajectory of the presumed cranial nerve of origin, or prior resection confirming diagnosis. Demographic, SRS dose planning, clinical, and imaging data were collected from chart review of the treated patients. Chi-square and Kaplan-Meier analyses were performed.
   RESULTS Seven institutions submitted data for a total of 25 patients. The median follow-up time was 41 months. The median age at the time of treatment was 52 years. There were 11 CN III schwannomas, 11 CN IV schwannomas, and 3 CN VI schwannomas. The median target volume was 0.74 cm(3), and the median marginal dose delivered was 12.5 Gy. After SRS, only 2 patients (including the only patient with neurofibromatosis type 2) had continued tumor growth. Crude local control was 92% (23/25), and the 10-year actuarial control was 86%. Diplopia improved in the majority of patients (11/21), and only 3 had worsening following SRS, 2 of whom also had worsened ptosis, both in the context of tumor progression.
   CONCLUSIONS SRS for schwannomas of the oculomotor, trochlear, and abducens nerves is effective and provides tumor control rates similar to those for other cranial nerve schwannomas. SRS allows improvement of diplopia in the majority of patients. SRS should therefore be considered as a first-line treatment option for oculomotor nerve schwannomas.
C1 [Langlois, Anne-Marie; Iorio-Morin, Christian; Mathieu, David] Univ Sherbrooke, CHU Sherbrooke, Ctr Rech, Div Neurosurg,Dept Surg, Sherbrooke, PQ, Canada.
   [Faramand, Andrew; Niranjan, Ajay; Lunsford, L. Dade] Univ Pittsburgh, Dept Neurol Surg, Pittsburgh, PA 15260 USA.
   [Mohammed, Nasser; Sheehan, Jason P.] Univ Virginia Hlth Syst, Dept Neurol Surg, Charlottesville, VA USA.
   [Liscak, Roman; Urgosik, Dusan] Homolce Hosp, Dept Stereotact & Radiat Neurosurg, Prague, Czech Republic.
   [Kondziolka, Douglas] NYU, Langone Med Ctr, Dept Neurosurg, 550 1St Ave, New York, NY 10016 USA.
   [Lee, Cheng-chia; Yang, Huai-che] Taipei Vet Hosp, Dept Radiat Oncol & Neurol Surg, Taipei, Taiwan.
   [Atik, Ahmet F.] Cleveland Clin, Dept Neurol Surg, Cleveland, OH 44106 USA.
C3 University of Sherbrooke; Pennsylvania Commonwealth System of Higher
   Education (PCSHE); University of Pittsburgh; University of Virginia; Na
   Homolce Hospital; New York University; NYU Langone Medical Center;
   Cleveland Clinic Foundation
RP Mathieu, D (通讯作者)，Univ Sherbrooke, CHU Sherbrooke, Ctr Rech, Sherbrooke, PQ, Canada.
EM
RI Lunsford, L. Dade/AHI-7993-2022; Atik, Ahmet F/HZM-2299-2023; Liscak,
   Roman/ABI-5311-2020; Mohammed, Nasser/AAG-1525-2019
OI Liscak, Roman/0000-0002-0929-8320; Mohammed, Nasser/0000-0002-3811-3559
CR Acharya R, 2003, NEUROL SCI, V24, P74, DOI 10.1007/s100720300076
   Choi CYH, 2011, NEUROSURGERY, V68, P1200, DOI 10.1227/NEU.0b013e31820c0474
   El Asri AC, 2019, WORLD NEUROSURG, V129, P72, DOI 10.1016/j.wneu.2019.05.170
   Elsharkawy M, 2012, J NEUROSURG, V116, P66, DOI 10.3171/2011.8.JNS11215
   Hayashi M, 2010, J NEUROSURG, V113, P136, DOI 10.3171/2010.8.GKS10947
   Inoue T, 2015, J NEUROL SURG REP, V76, pE248, DOI 10.1055/s-0035-1564059
   Iorio-Morin C, 2019, NEUROSURGERY, V85, P535, DOI 10.1093/neuros/nyy416
   Iorio-Morin C, 2016, NEUROSURGERY, V78, P521, DOI 10.1227/NEU.0000000000001154
   Kano H, 2018, J NEUROSURG, V129, P928, DOI 10.3171/2017.5.JNS162894
   Kim IY, 2008, J NEUROSURG, V109, P149, DOI 10.3171/JNS/2008/109/12/S23
   Kimball MM, 2011, NEUROSURGERY, V68, P974, DOI 10.1227/NEU.0b013e318208f3a1
   Kondziolka D, 1998, NEW ENGL J MED, V339, P1426, DOI 10.1056/NEJM199811123392003
   Langlois AM, 2018, WORLD NEUROSURG, V110, pE1031, DOI 10.1016/j.wneu.2017.11.163
   Nishioka K, 2009, INT J RADIAT ONCOL, V75, P1415, DOI 10.1016/j.ijrobp.2008.12.063
   Nonaka Y, 2014, WORLD NEUROSURG, V81, P798, DOI 10.1016/j.wneu.2012.11.067
   Ohata K, 2006, NEUROL INDIA, V54, P437, DOI 10.4103/0028-3886.28125
   Peciu-Florianu I, 2017, ACTA NEUROCHIR, V159, P2409, DOI 10.1007/s00701-017-3348-0
   Petrela E, 2009, J NEURO-OPHTHALMOL, V29, P54, DOI 10.1097/WNO.0b013e31818f6463
   Pollock BE, 2002, INT J RADIAT ONCOL, V52, P1002, DOI 10.1016/S0360-3016(01)02711-0
   Prasad GL, 2016, J NEUROSURG, V125, P832, DOI 10.3171/2015.8.JNS151140
   Puataweepong Putipun, 2016, Asian Pac J Cancer Prev, V17, P3271
   Sheehan JP, 2015, J NEUROSURG, V123, P387, DOI 10.3171/2014.11.JNS141160
   Solberg TD, 2012, PRACT RADIAT ONCOL, V2, P2, DOI 10.1016/j.prro.2011.06.014
NR 23
TC 13
Z9 13
U1 0
U2 1
PU AMER ASSOC NEUROLOGICAL SURGEONS
PI ROLLING MEADOWS
PA 5550 MEADOWBROOK DRIVE, ROLLING MEADOWS, IL 60008 USA
SN 0022-3085
EI 1933-0693
J9 J NEUROSURG
JI J. Neurosurg.
PD OCT
PY 2021
VL 135
IS 4
BP 1044
EP 1050
DI 10.3171/2020.8.JNS20887
PG 7
WC Clinical Neurology; Surgery
WE Science Citation Index Expanded (SCI-EXPANDED)
SC Neurosciences & Neurology; Surgery
GA WF6SR
UT WOS:000706433600007
PM 33482633
OA Bronze
DA 2023-08-27
ER

PT J
AU Scharf, M
   Miske, R
   Heidenreich, F
   Giess, R
   Landwehr, P
   Blocker, IM
   Begemann, N
   Denno, Y
   Tiede, S
   Dahnrich, C
   Schlumberger, W
   Unger, M
   Teegen, B
   Stocker, W
   Probst, C
   Komorowski, L
AF Scharf, Madeleine
   Miske, Ramona
   Heidenreich, Fedor
   Giess, Ralf
   Landwehr, Peter
   Bloecker, Inga-Madeleine
   Begemann, Nora
   Denno, Yvonne
   Tiede, Stephan
   Daehnrich, Cornelia
   Schlumberger, Wolfgang
   Unger, Mandy
   Teegen, Bianca
   Stoecker, Winfried
   Probst, Christian
   Komorowski, Lars
TI Neuronal Na+/K+ ATPase is an autoantibody target in paraneoplastic
   neurologic syndrome
SO NEUROLOGY
LA English
DT Article
ID CASE SERIES; ANTIBODIES; ENCEPHALITIS; RECEPTOR; SUBUNIT;
   ALPHA-3-ISOFORM; NA+,K+-ATPASE; MUTATIONS; ANTIGEN
AB Objectives:To identify an autoreactivity in a 66-year-old woman who presented with combined brainstem and cerebellar syndrome including vertical gaze palsy, severe progressive ataxia, and spastic tetraparesis, an acute deterioration of vision, dysarthria, and dysphagia with concurrent diagnosis of a colon adenocarcinoma.Methods:Patient's serum and CSF underwent comprehensive autoantibody screening by indirect immunofluorescence assay and immunoblot. For autoantigen purification, a histo-immunoprecipitation technique was developed followed by mass spectrometrical analysis. Recombinant candidate antigens were expressed in HEK293 and used to verify the identification.Results:Indirect immunofluorescence assay screening revealed strong immunoglobulin G reactivity with neural tissues in serum and CSF, but not with a panel of 28 recombinantly expressed established neural autoantigens. The hitherto unknown target antigen was identified as the neuronal Na+/K+ ATPase. Epitope mapping and competitive inhibition experiments showed that the autoantibodies were directed against the membrane-spanning alpha 3 subunit (ATP1A3) of the enzyme but did not bind to extracellular epitopes. Immunohistochemical analysis revealed overexpression of this subunit in the patient's tumor.Conclusions:We describe a case of an anti-ATP1A3-associated neurologic disorder. Mutations in the gene encoding this neuronal surface protein have already been recognized as the cause of infantile alternating hemiplegia, rapid-onset dystonia parkinsonism, and CAPOS syndrome. Although the autoantibodies are unlikely to be pathogenic, they are likely to be rare biomarkers for the apparently paraneoplastic neurologic syndrome or for the tumor itself.
C1 [Scharf, Madeleine; Miske, Ramona; Bloecker, Inga-Madeleine; Begemann, Nora; Denno, Yvonne; Tiede, Stephan; Daehnrich, Cornelia; Schlumberger, Wolfgang; Unger, Mandy; Teegen, Bianca; Stoecker, Winfried; Probst, Christian; Komorowski, Lars] EUROIMMUN AG, Inst Expt Immunol, Lubeck, Germany.
   [Heidenreich, Fedor; Giess, Ralf] Hosp Henriettenstiftung, Dept Neurol & Clin Neurophysiol, Hannover, Germany.
   [Landwehr, Peter] Hosp Henriettenstiftung, Dept Diagnost & Intervent Radiol, Hannover, Germany.
C3 EUROIMMUN
RP Komorowski, L (通讯作者)，EUROIMMUN AG, Inst Expt Immunol, Lubeck, Germany.
EM
OI Tiede, Stephan/0000-0002-6045-3736
CR Aguiar PD, 2004, NEURON, V43, P169, DOI 10.1016/j.neuron.2004.06.028
   Baba A, 2002, J AM COLL CARDIOL, V40, P1153, DOI 10.1016/S0735-1097(02)02075-2
   Boronat A, 2013, ANN NEUROL, V73, P120, DOI 10.1002/ana.23756
   Brashear A, 2014, NEUROLOGY, V82, P468, DOI 10.1212/WNL.0000000000000113
   Dalmau J, 2008, LANCET NEUROL, V7, P1091, DOI 10.1016/S1474-4422(08)70224-2
   Demos MK, 2014, ORPHANET J RARE DIS, V9, DOI 10.1186/1750-1172-9-15
   Forrest MD, 2012, PLOS ONE, V7, DOI 10.1371/journal.pone.0051169
   Heinzen EL, 2012, NAT GENET, V44, P1030, DOI 10.1038/ng.2358
   Heo S, 2012, NEUROCHEM INT, V61, P702, DOI 10.1016/j.neuint.2012.07.006
   Hutchinson M, 2008, NEUROLOGY, V71, P1291, DOI 10.1212/01.wnl.0000327606.50322.f0
   Lennon VA, 2005, J EXP MED, V202, P473, DOI 10.1084/jem.20050304
   Petit-Pedrol M, 2014, LANCET NEUROL, V13, P276, DOI 10.1016/S1474-4422(13)70299-0
   Probst C, 2014, MULT SCLER RELAT DIS, V3, P303, DOI 10.1016/j.msard.2013.12.001
   Sabater L, 2014, LANCET NEUROL, V13, P575, DOI 10.1016/S1474-4422(14)70051-1
   Sakai H, 2004, FEBS LETT, V563, P151, DOI 10.1016/S0014-5793(04)00292-3
   Shibuya K, 2010, EUR J PHARMACOL, V638, P42, DOI 10.1016/j.ejphar.2010.04.029
   STOCKER W, 1987, SCAND J GASTROENTERO, V22, P41, DOI 10.3109/00365528709089774
   SVERDLOV ED, 1988, FEBS LETT, V239, P65, DOI 10.1016/0014-5793(88)80546-5
   Vicco MH, 2013, REV ESP CARDIOL, V66, P791, DOI [10.1016/j.rec.2013.05.028, 10.1016/j.recesp.2013.05.028]
NR 19
TC 10
Z9 13
U1 0
U2 7
PU LIPPINCOTT WILLIAMS & WILKINS
PI PHILADELPHIA
PA TWO COMMERCE SQ, 2001 MARKET ST, PHILADELPHIA, PA 19103 USA
SN 0028-3878
EI 1526-632X
J9 NEUROLOGY
JI Neurology
PD APR 21
PY 2015
VL 84
IS 16
BP 1673
EP 1679
DI 10.1212/WNL.0000000000001493
PG 7
WC Clinical Neurology
WE Science Citation Index Expanded (SCI-EXPANDED)
SC Neurosciences & Neurology
GA CG4GJ
UT WOS:000353243200014
PM 25809299
DA 2023-08-27
ER

PT J
AU Kansagra, SM
   Gallentine, WB
AF Kansagra, Sujay M.
   Gallentine, William B.
TI Cytokine Storm of Acute Necrotizing Encephalopathy
SO PEDIATRIC NEUROLOGY
LA English
DT Article
ID VIRUS-ASSOCIATED ENCEPHALOPATHY; INFLUENZA; INFECTION; CHILDHOOD
AB Acute necrotizing encephalopathy is a rare, clinically distinct entity characterized by multiple, symmetric areas of edema and necrosis in the thalamus, cerebellum, brainstem, and white matter. It is postulated to arise from uncontrolled cytokine release during a febrile illness, and is most often seen in East Asia. We describe a rare North American case of acute necrotizing encephalopathy attributable to human herpes virus-6 is a 9-month-old white male. The infant moved to the United States from Hong Kong, 3 months before disease onset. A workup revealed elevations in serum interleukin-1 beta, interleukin-2, and interleukin-10, with normal values of interleukin-6 and tumor necrosis factor-CL after the initiation of high-dose steroids. This profile is unique compared with previous cytokine profiles of this disease, possibly because of the effects of steroid therapy. A rare North American case with a history of birth in East Asia underscores the possibility of a role for environmental pathogens in this disease. (C) 2011 Elsevier Inc. All rights reserved.
C1 [Kansagra, Sujay M.; Gallentine, William B.] Duke Univ, Med Ctr, Div Pediat Neurol, Dept Pediat, Durham, NC 27710 USA.
C3 Duke University
RP Gallentine, WB (通讯作者)，Duke Univ, Med Ctr, Div Pediat Neurol, Dept Pediat, Box 3936, Durham, NC 27710 USA.
EM
CR Akiyoshi K, 2006, PEDIATR NEUROL, V34, P315, DOI 10.1016/j.pediatrneurol.2005.08.030
   Huang SM, 2004, J CHILD NEUROL, V19, P64, DOI 10.1177/08830738040190010709
   Ichiyama T, 2003, SCAND J INFECT DIS, V35, P59, DOI 10.1080/0036554021000026986
   Ichiyama T, 2003, PEDIATR INT, V45, P734, DOI 10.1111/j.1442-200X.2003.01822.x
   Ito Y, 1999, J MED VIROL, V58, P420, DOI 10.1002/(SICI)1096-9071(199908)58:4&lt;420::AID-JMV16&gt;3.0.CO;2-T
   Kubo T, 2006, SCAND J INFECT DIS, V38, P1122, DOI 10.1080/00365540600740520
   MIZUGUCHI M, 1995, J NEUROL NEUROSUR PS, V58, P555, DOI 10.1136/jnnp.58.5.555
   Neilson DE, 2009, AM J HUM GENET, V84, P44, DOI 10.1016/j.ajhg.2008.12.009
   Okumura A, 2009, MICROBIOL IMMUNOL, V53, P277, DOI 10.1111/j.1348-0421.2009.00124.x
NR 9
TC 46
Z9 55
U1 0
U2 7
PU ELSEVIER SCIENCE INC
PI NEW YORK
PA 360 PARK AVE SOUTH, NEW YORK, NY 10010-1710 USA
SN 0887-8994
J9 PEDIATR NEUROL
JI Pediatr. Neurol.
PD DEC
PY 2011
VL 45
IS 6
BP 400
EP 402
DI 10.1016/j.pediatrneurol.2011.09.007
PG 3
WC Clinical Neurology; Pediatrics
WE Science Citation Index Expanded (SCI-EXPANDED)
SC Neurosciences & Neurology; Pediatrics
GA 859OL
UT WOS:000297885600009
PM 22115004
DA 2023-08-27
ER

PT J
AU Park, HJ
   Kim, HJ
   Bae, GS
   Seo, SW
   Kim, DY
   Jung, WS
   Kim, MS
   Song, MY
   Kim, EK
   Kwon, KB
   Hwang, SY
   Song, HJ
   Park, CS
   Park, RK
   Chong, MS
   Park, SJ
AF Park, Hee-Je
   Kim, Hyung-Jin
   Bae, Gi-Sang
   Seo, Sang-Wan
   Kim, Do-Yun
   Jung, Won-Seok
   Kim, Min-Sun
   Song, Mi-Young
   Kim, Eun-Kyung
   Kwon, Kang-Beom
   Hwang, Sung-Yeon
   Song, Ho-Joon
   Park, Cheung-Seog
   Park, Rae-Kil
   Chong, Myong-Soo
   Park, Sung-Joo
TI Selective GSK-3 beta inhibitors attenuate the cisplatin-induced
   cytotoxicity of auditory cells
SO HEARING RESEARCH
LA English
DT Article
DE GSK-3; Ototoxicity; Cochlea; Hair cell; Hearing loss
ID GLYCOGEN-SYNTHASE KINASE-3; GROWTH-FACTOR WITHDRAWAL; NF-KAPPA-B;
   INDUCED APOPTOSIS; C57BL/6 MICE; ORGANOTYPIC CULTURES; HAIR-CELLS;
   PHOSPHORYLATION; ACTIVATION; DEATH
AB Glycogen synthase kinase-3 (GSK-3) plays an important role in the regulation of apoptosis. However, the role of GSK-3 in the auditory system remains unknown. Here we examined whether the GSK-3-specific inhibitors, SB 216763 and LiCl, could protect against cisplatin-induced cytotoxicity of auditory cells. GSK-3 was activated by cisplatin treatment of HEl-OC1 cells. SB 216763 or LiCl treatments inhibited cisplatin-induced apoptosis in a dose-dependent manner and activated caspase-9, -8 and -3. In rat primary explants of the organ of Corti, SB 216763 or LiCl treatments completely abrogated the cisplatin-induced destruction of outer hair cell arrays. Administration of SB 216763 or LiCl inhibited cochlear destruction and the production of tumor necrosis factor-alpha (TNF-alpha), interleukin-1 beta (IL-1 beta) and IL-6 in cisplatin-injected mice. Furthermore, administration of SB 216763 or LiCl reduced the thresholds of the auditory brainstem response (ABR) in cisplatin-injected mice. Collectively, these results suggest that cisplatin-induced otortoxicity might be associated with modulation of GSK-3 activation. (C) 2009 Elsevier B.V. All rights reserved.
C1 [Park, Hee-Je; Bae, Gi-Sang; Seo, Sang-Wan; Kim, Do-Yun; Jung, Won-Seok; Kim, Min-Sun; Hwang, Sung-Yeon; Song, Ho-Joon; Park, Sung-Joo] Wonkwang Univ, Coll Oriental Med, Dept Herbol, Iksan, Jeonbuk, South Korea.
   [Kim, Hyung-Jin; Park, Rae-Kil] Wonkwang Univ, Coll Med, Vestibulocochlear Res Ctr, Iksan, Jeonbuk, South Korea.
   [Kim, Hyung-Jin; Park, Rae-Kil] Wonkwang Univ, Coll Med, Dept Microbiol, Iksan, Jeonbuk, South Korea.
   [Seo, Sang-Wan; Park, Cheung-Seog] Kyung Hee Univ, Coll Med, Dept Microbiol, Seoul, South Korea.
   [Song, Mi-Young; Kim, Eun-Kyung; Kwon, Kang-Beom] Wonkwang Univ, Coll Oriental Med, Dept Physiol, Iksan, Jeonbuk, South Korea.
   [Chong, Myong-Soo] Wonkwang Univ, Coll Oriental Med, Dept Prevent Med, Iksan, Jeonbuk, South Korea.
C3 Wonkwang University; Wonkwang University; Wonkwang University; Kyung Hee
   University; Wonkwang University; Wonkwang University
RP Park, SJ (通讯作者)，Wonkwang Univ, Coll Oriental Med, Dept Herbol, Iksan, Jeonbuk, South Korea.
EM;
  ;
  ;
  ;
  ;
  
FU Ministry of Science & Technology (MoST)/Korea Science & Engineering
   Foundation (KOSEF) through the Vestibulocochlear Research Center (VCRC)
   [R13-2002-055-00000-0]
FX This work was supported by the Ministry of Science & Technology
   (MoST)/Korea Science & Engineering Foundation (KOSEF) through the
   Vestibulocochlear Research Center (VCRC) at Wonkwang University
   (R13-2002-055-00000-0).
CR Alvarez G, 1999, FEBS LETT, V453, P260, DOI 10.1016/S0014-5793(99)00685-7
   Bartolome MV, 2002, ADV OTO-RHINO-LARYNG, V59, P106
   Bhat RV, 2000, P NATL ACAD SCI USA, V97, P11074, DOI 10.1073/pnas.190297597
   Cai GQ, 2007, INT J ONCOL, V31, P657
   Coghlan MP, 2000, CHEM BIOL, V7, P793, DOI 10.1016/S1074-5521(00)00025-9
   Crowder RJ, 2000, J BIOL CHEM, V275, P34266, DOI 10.1074/jbc.M006160200
   Davis CA, 2001, J AM SOC NEPHROL, V12, P2683, DOI 10.1681/ASN.V12122683
   Devarajan P, 2002, HEARING RES, V174, P45, DOI 10.1016/S0378-5955(02)00634-2
   Feghali JG, 2001, LARYNGOSCOPE, V111, P1147, DOI 10.1097/00005537-200107000-00005
   Forde JE, 2007, CELL MOL LIFE SCI, V64, P1930, DOI 10.1007/s00018-007-7045-7
   Fram Robert J., 1992, Current Opinion in Oncology, V4, P1073, DOI 10.1097/00001622-199212000-00012
   Henry KR, 2002, HEARING RES, V170, P107, DOI 10.1016/S0378-5955(02)00391-X
   Hequembourg S, 2001, JARO-J ASSOC RES OTO, V2, P118, DOI 10.1007/s101620010075
   Huang T, 2000, INT J DEV NEUROSCI, V18, P259, DOI 10.1016/S0736-5748(99)00094-5
   Jeong HJ, 2007, J NEUROSCI RES, V85, P896, DOI 10.1002/jnr.21168
   Jordan P, 2000, CELL MOL LIFE SCI, V57, P1229, DOI 10.1007/PL00000762
   Kalinec GM, 2003, AUDIOL NEURO-OTOL, V8, P177, DOI 10.1159/000071059
   Kharbanda S, 2000, MOL CELL BIOL, V20, P4979, DOI 10.1128/MCB.20.14.4979-4989.2000
   Klein PS, 1996, P NATL ACAD SCI USA, V93, P8455, DOI 10.1073/pnas.93.16.8455
   Koh SH, 2003, MOL BRAIN RES, V118, P72, DOI 10.1016/j.molbrainres.2003.07.003
   Kopke RD, 1997, AM J OTOL, V18, P559
   Kwon O, 2008, CELL SIGNAL, V20, P602, DOI 10.1016/j.cellsig.2007.10.022
   LAURELL G, 1991, ACTA OTO-LARYNGOL, V111, P891, DOI 10.3109/00016489109138427
   Loberg RD, 2002, J BIOL CHEM, V277, P41667, DOI 10.1074/jbc.M206405200
   McFadden SL, 2001, AUDIOLOGY, V40, P313
   Oloumi A, 2006, ONCOGENE, V25, P7747, DOI 10.1038/sj.onc.1209752
   Price MA, 2002, CELL, V108, P823, DOI 10.1016/S0092-8674(02)00664-5
   Rosato RR, 2007, CANCER RES, V67, P9490, DOI 10.1158/0008-5472.CAN-07-0598
   Shaw M, 1997, FEBS LETT, V416, P307, DOI 10.1016/S0014-5793(97)01235-0
   Shiga A, 2005, AUDIOL NEURO-OTOL, V10, P97, DOI 10.1159/000083365
   So H, 2007, JARO-J ASSOC RES OTO, V8, P338, DOI 10.1007/s10162-007-0084-9
   So HS, 2005, HEARING RES, V204, P127, DOI 10.1016/j.heares.2005.01.011
   Somervaille TCP, 2001, BLOOD, V98, P1374, DOI 10.1182/blood.V98.5.1374
   Spongr VP, 1997, J ACOUST SOC AM, V101, P3546, DOI 10.1121/1.418315
   Stanton SE, 2006, J BIOL CHEM, V281, P28782, DOI 10.1074/jbc.M603348200
   Tan Y, 2006, J BIOL CHEM, V281, P17689, DOI 10.1074/jbc.M601978200
   Watanabe K, 2002, CHEMOTHERAPY, V48, P82, DOI 10.1159/000057667
   Yin H, 2004, HYPERTENSION, V43, P109, DOI 10.1161/01.HYP.0000103696.60047.55
   Zheng JL, 1996, EUR J NEUROSCI, V8, P1897, DOI 10.1111/j.1460-9568.1996.tb01333.x
NR 39
TC 37
Z9 41
U1 0
U2 9
PU ELSEVIER SCIENCE BV
PI AMSTERDAM
PA PO BOX 211, 1000 AE AMSTERDAM, NETHERLANDS
SN 0378-5955
EI 1878-5891
J9 HEARING RES
JI Hear. Res.
PD NOV
PY 2009
VL 257
IS 1-2
BP 53
EP 62
DI 10.1016/j.heares.2009.08.001
PG 10
WC Audiology & Speech-Language Pathology; Neurosciences;
   Otorhinolaryngology
WE Science Citation Index Expanded (SCI-EXPANDED)
SC Audiology & Speech-Language Pathology; Neurosciences & Neurology;
   Otorhinolaryngology
GA 511QB
UT WOS:000271180200006
PM 19666099
DA 2023-08-27
ER

PT J
AU Sawabe, M
   Esaki, Y
   Tsubokou, Y
   Takubo, K
   Yamada, S
   Mizutani, T
   Kanazawa, A
AF Sawabe, M
   Esaki, Y
   Tsubokou, Y
   Takubo, K
   Yamada, S
   Mizutani, T
   Kanazawa, A
TI Preclinical primary central nervous system lymphoma
SO CLINICAL NEUROPATHOLOGY
LA English
DT Article
DE central nervous system; early cancer; latent cancer; malignant lymphoma
ID ACQUIRED-IMMUNODEFICIENCY-SYNDROME; MALIGNANT-LYMPHOMA; JAPAN
AB We report an autopsy case of preclinical primary central nervous system (CNS) lymphoma. The case was an 89-year-old female who died of rupture of a thoracic aortic aneurysm. No neurological abnormalities were found throughout our clinical observation. Serum anti-human T-cell leukemia virus type 1 and anti-human immunodeficiency virus were negative. Grossly, the brain was normal except for a small solid tumor, 6 by 6 by 2.5 mm, in the lateral ventricular wall at the left hippocampal fimbria, and multiple smaller disseminated nodules. Histological examination revealed multiple microscopic disseminated foci throughout the brain, including the cerebral subcortical white matter, basal nucleus, thalamus, brainstem and cerebellum. Histological classification is of lymphoblastic type non-Hodgkin's lymphoma of high-grade malignancy. Lymphoma cells are positive for leukocyte common antigen and L-26, indicating a B cell phenotype. In situ hybridization with Epstein-Barr viral probes is negative. It is concluded that this case may represent a relatively early preclinical stage of multifocal type of primary CNS lymphoma.
C1 TOKYO METROPOLITAN GERIATR HOSP, DEPT CARDIOL, TOKYO 173, JAPAN.
   TOKYO METROPOLITAN INST GERONTOL, DEPT CLIN PATHOL, TOKYO, JAPAN.
   TOKYO METROPOLITAN INST GERONTOL, DEPT NEUROPATHOL, TOKYO, JAPAN.
   TOKYO MED & DENT UNIV, DEPT PATHOL & IMMUNOL, TOKYO 113, JAPAN.
C3 Tokyo Metropolitan Institute of Gerontology; Tokyo Metropolitan
   Institute of Gerontology; Tokyo Metropolitan Institute of Gerontology;
   Tokyo Medical & Dental University (TMDU)
RP Sawabe, M (通讯作者)，TOKYO METROPOLITAN GERIATR HOSP, DEPT PATHOL, ITABASHI KU, SAKAE CHO 35-2, TOKYO 173, JAPAN.
CR AHO R, 1993, ACTA NEUROPATHOL, V86, P71, DOI 10.1007/BF00454901
   AOZASA K, 1990, INT J CANCER, V45, P632, DOI 10.1002/ijc.2910450410
   BAUMGARTNER JE, 1990, J NEUROSURG, V73, P206, DOI 10.3171/jns.1990.73.2.0206
   Bergmann Markus, 1993, Zentralblatt fuer Pathologie, V139, P37
   BURGER PC, 1994, ATLAS TUMOR PATHOL, P321
   HAYAKAWA T, 1994, J NEURO-ONCOL, V19, P197, DOI 10.1007/BF01053274
   HENRY JM, 1974, CANCER, V34, P1293, DOI 10.1002/1097-0142(197410)34:4<1293::AID-CNCR2820340441>3.0.CO;2-P
   HOCHBERG FH, 1988, J NEUROSURG, V68, P835, DOI 10.3171/jns.1988.68.6.0835
   IOACHIM HL, 1991, HUM PATHOL, V22, P659, DOI 10.1016/0046-8177(91)90288-Z
   MACMAHON EME, 1991, LANCET, V338, P969, DOI 10.1016/0140-6736(91)91837-K
   MILLER DC, 1994, CANCER-AM CANCER SOC, V74, P1383, DOI 10.1002/1097-0142(19940815)74:4<1383::AID-CNCR2820740432>3.0.CO;2-1
   ONEILL BP, 1989, MAYO CLIN PROC, V64, P1005, DOI 10.1016/S0025-6196(12)61228-2
   PAULUS W, 1993, ACTA NEUROPATHOL, V86, P360, DOI 10.1007/BF00369448
   ROSENBERG SA, 1982, CANCER, V49, P2112
   TOMLINSON FH, 1995, J NEUROSURG, V82, P558, DOI 10.3171/jns.1995.82.4.0558
NR 15
TC 0
Z9 0
U1 0
U2 1
PU DUSTRI-VERLAG DR KARL FEISTLE
PI DEISENHOFEN-MUENCHEN
PA BAHNHOFSTRASSE 9 POSTFACH 49, D-82032 DEISENHOFEN-MUENCHEN, GERMANY
SN 0722-5091
J9 CLIN NEUROPATHOL
JI Clin. Neuropathol.
PD MAR-APR
PY 1997
VL 16
IS 2
BP 85
EP 89
PG 5
WC Clinical Neurology; Pathology
WE Science Citation Index Expanded (SCI-EXPANDED)
SC Neurosciences & Neurology; Pathology
GA WR107
UT WOS:A1997WR10700007
PM 9101110
DA 2023-08-27
ER

PT J
AU Kryzanski, JT
   Robertson, JH
   Heilman, CB
AF Kryzanski, James T.
   Robertson, Jon H.
   Heilman, Carl B.
TI A Minimal Access Far-Lateral Approach to Foramen Magnum Lesions
SO JOURNAL OF NEUROLOGICAL SURGERY PART B-SKULL BASE
LA English
DT Article
DE far-lateral; foramen magnum; meningioma; surgical approach;
   complications
ID OF-THE-LITERATURE; CRANIOCERVICAL JUNCTION; TRANSCONDYLAR APPROACH;
   SUBOCCIPITAL APPROACH; SURGICAL-MANAGEMENT; INTRADURAL LESIONS; ANTERIOR
   PORTION; BENIGN-TUMORS; MENINGIOMAS; EXPERIENCE
AB Objectives The far-lateral approach is widely used to treat pathology of the ventral foramen magnum. Numerous methods of exposure have been described, most of which utilize long skin incisions and myocutaneous flaps. Here we present our experience with gaining exposure through a small paramedian incision using a muscle-splitting technique.
   Design A cadaveric anatomical study was first performed to verify the feasibility of the approach. We then describe our experience with using the approach in 13 patients. A retrospective chart review was performed and data regarding pathology, imaging, and complications were collected.
   Results The cadaveric study confirmed that a small paramedian muscle-splitting approach allows sufficient exposure to approach many foramen magnum lesions. Our case series included 10 patients with meningioma, one brainstem glioblastoma, one posterior inferior cerebellar artery aneurysm, and one odontoid pannus. The exposure was adequate in all cases. For the meningioma patients, six had gross total resections and four had subtotal resection because of tumor adherence to neurovascular structures. Two patients experienced postoperative cardiovascular complications. There were no new neurologic deficits, cerebrospinal fluid leaks, or wound complications.
   Conclusions A small paramedian incision may be used to gain exposure and perform successful far-lateral approaches. The small exposure is likely to reduce the risk of local complications such as cerebrospinal fluid fistula and pseudomeningocele when compared with larger exposures.
C1 [Kryzanski, James T.; Heilman, Carl B.] Tufts Med Ctr, Dept Neurosurg, Boston, MA 02111 USA.
   [Robertson, Jon H.] Univ Tennessee, Dept Neurosurg, Coll Med, Semmes Murphey Neurol & Spine Inst, Memphis, TN USA.
C3 Tufts Medical Center; University of Tennessee System; University of
   Tennessee Health Science Center
RP Kryzanski, JT (通讯作者)，Tufts Med Ctr, 800 Washington St Box 178, Boston, MA 02111 USA.
EM
CR Arnautovic KI, 2000, J NEUROSURG, V92, P71, DOI 10.3171/spi.2000.92.1.0071
   Bassiouni H, 2006, NEUROSURGERY, V59, P1177, DOI 10.1227/01.NEU.0000245629.77968.37
   BERTALANFFY H, 1991, NEUROSURGERY, V29, P815, DOI 10.1227/00006123-199112000-00002
   Borba LAB, 2009, NEUROSURG REV, V32, P49, DOI 10.1007/s10143-008-0161-5
   Bruneau M, 2008, NEUROSURG REV, V31, P19, DOI 10.1007/s10143-007-0097-1
   Cappabianca P, 2010, ACTA NEUROCHIR, V152, P897, DOI 10.1007/s00701-009-0580-2
   GEORGE B, 1988, SURG NEUROL, V29, P484, DOI 10.1016/0090-3019(88)90145-0
   HEROS RC, 1986, J NEUROSURG, V64, P559, DOI 10.3171/jns.1986.64.4.0559
   J Jagannathan, 2007, NEUROSURGERY, V61, pE173
   Jagannathan Jay, 2007, Neurosurgery, V61, pE172, DOI 10.1227/01.neu.0000279741.38178.2d
   Kandenwein JA, 2009, BRIT J NEUROSURG, V23, P33, DOI 10.1080/02688690802545932
   Kano T, 2010, ACTA NEUROCHIR, V152, P79, DOI 10.1007/s00701-009-0511-2
   Karam YR, 2010, NEUROSURGERY, V66, pA135, DOI 10.1227/01.NEU.0000365828.03949.D0
   Lutjens G, 2011, EUR SPINE J, V20, pS262, DOI 10.1007/s00586-010-1665-6
   M Samii, 1996, NEUROSURGERY, V39, P1094
   Margalit Nevo S, 2005, Neurosurgery, V56, P324, DOI 10.1227/01.NEU.0000156796.28536.6D
   MEYER FB, 1984, J NEUROSURG, V61, P136, DOI 10.3171/jns.1984.61.1.0136
   Muthukumar N, 1999, SURG NEUROL, V51, P268, DOI 10.1016/S0090-3019(98)00099-8
   Nanda A, 2002, J NEUROSURG, V96, P302, DOI 10.3171/jns.2002.96.2.0302
   Nicolato A, 2001, MINIM INVAS NEUROSUR, V44, P211, DOI 10.1055/s-2001-19934
   Pamir MN, 2004, J CLIN NEUROSCI, V11, P863, DOI 10.1016/j.jocn.2004.02.007
   Pirotte BJM, 2010, NEUROSURGERY, V67, P58, DOI 10.1227/01.NEU.0000382971.63877.DD
   Pirotte BJM, 2010, NEUROSURGERY S, V67, pons58
   Raffa Scott J, 2009, Spine J, V9, pe25, DOI 10.1016/j.spinee.2008.07.002
   Samii M, 1996, NEUROSURGERY, V39, P1086, DOI 10.1097/00006123-199612000-00003
   SEN CN, 1990, NEUROSURGERY, V27, P197, DOI 10.1227/00006123-199008000-00004
   Spektor S, 2000, J NEUROSURG, V92, P824, DOI 10.3171/jns.2000.92.5.0824
   Starke RM, 2010, J CRANIOVERTEBRAL JU, V1, P23, DOI 10.4103/0974-8237.65478
   STEIN BM, 1963, J NEUROSURG, V20, P740, DOI 10.3171/jns.1963.20.9.0740
   Talacchi A, 2012, NEUROSURG REV, V35, P359, DOI 10.1007/s10143-012-0381-6
   Wanebo JE, 2001, NEUROSURGERY, V49, P934, DOI 10.1097/00006123-200110000-00027
   Wu AH, 2010, NEUROSURGERY, V66, pONS191, DOI [10.1227/10.NEU.0000369704.49958.5B, 10.1227/01.NEU.0000369704.49958.5B]
   Wu Z, 2009, SURG NEUROL, V72, P376, DOI 10.1016/j.surneu.2009.05.006
NR 33
TC 8
Z9 9
U1 0
U2 1
PU THIEME MEDICAL PUBL INC
PI NEW YORK
PA 333 SEVENTH AVE, NEW YORK, NY 10001 USA
SN 2193-6331
EI 2193-634X
J9 J NEUROL SURG PART B
JI Journal of Neurol. Surg. Part B
PD AUG
PY 2014
VL 75
IS 4
BP 236
EP 242
DI 10.1055/s-0034-1371363
PG 7
WC Clinical Neurology; Surgery
WE Science Citation Index Expanded (SCI-EXPANDED)
SC Neurosciences & Neurology; Surgery
GA AN3LG
UT WOS:000340488900004
PM 25093146
OA Green Published
DA 2023-08-27
ER

PT J
AU Wang, K
   Tian, KB
   Wang, L
   Wu, Z
   Ren, C
   Hao, SY
   Feng, J
   Li, JH
   Wan, H
   Jia, GJ
   Zhang, LW
   Zhang, JT
AF Wang, Ke
   Tian, Kaibing
   Wang, Liang
   Wu, Zhen
   Ren, Cong
   Hao, Shuyu
   Feng, Jie
   Li, Junhua
   Wan, Hong
   Jia, Guijun
   Zhang, Liwei
   Zhang, Junting
TI Brachyury: A sensitive marker, but not a prognostic factor, for skull
   base chordomas
SO MOLECULAR MEDICINE REPORTS
LA English
DT Article
DE skull base chordoma; prognosis; Brachyury; radical surgery
ID EXPRESSION; PATTERNS; CANCER; RISK
AB Patients with skull base chordomas have a poor prognosis, and the role of the protein expression of brachyury in chordomas remains to be fully elucidated. The present study used immunohistochemistry to analyze 57 cases of skull base chordoma, and analyzed the clinical data of the patients. The results demonstrated that the protein expression of brachyury was negative in 8.8% (5/57) of the cases. The weak/positive, positive and strong/positive rates were 5.3% (3/57), 21.1% (12/57) and 64.9% (37/57), respectively. The association between the expression of brachyury and recurrence was not statistically significant. Kaplan-Meier analysis revealed that the degree of surgery, rather than the expression of brachyury, was associated with tumor recurrence (P=0.001). In conclusion, the results of the present study demonstrated that the expression of Brachyury offers a sensitive marker, but not a risk factor, for skull base chordomas, and radical surgery is recommended.
C1 [Wang, Ke; Tian, Kaibing; Wang, Liang; Wu, Zhen; Ren, Cong; Hao, Shuyu; Jia, Guijun; Zhang, Liwei; Zhang, Junting] Capital Med Univ, Dept Neurosurg, Skull Base & Brainstem Tumor Div, Beijing Tian Tan Hosp,China Nat Clin Res Ctr Neur, Beijing 100050, Peoples R China.
   [Feng, Jie; Li, Junhua; Wan, Hong] Capital Med Univ, Beijing Neurosurg Inst, Beijing 100050, Peoples R China.
C3 Capital Medical University; Beijing Neurosurgical Institute; Capital
   Medical University
RP Zhang, JT (通讯作者)，Capital Med Univ, Dept Neurosurg, Skull Base & Brainstem Tumor Div, Beijing Tian Tan Hosp,China Nat Clin Res Ctr Neur, 6 Tiantan Xili, Beijing 100050, Peoples R China.
EM
RI jiang, lei/IWE-1124-2023; li, wei/IUQ-2973-2023; Wang,
   lili/IXD-9828-2023
FU Natural Science Foundation of China [81101910]; Natural Science
   Foundation of Beijing, China [7142052]
FX This study was supported, in part, by the Natural Science Foundation of
   China (grant. no. 81101910) and the Natural Science Foundation of
   Beijing, China (grant. no. 7142052).
CR Choi D, 2010, NEUROSURGERY, V66, P59, DOI 10.1227/01.NEU.0000362000.35742.3D
   Di Maio S, 2012, NEUROSURGERY, V70, P1355, DOI 10.1227/NEU.0b013e3182446783
   Di Maio S, 2011, J NEUROSURG, V115, P1094, DOI 10.3171/2011.7.JNS11355
   Hung GY, 2014, ANN SURG ONCOL, V21, P2490, DOI 10.1245/s10434-014-3697-3
   Jambhekar NA, 2010, ARCH PATHOL LAB MED, V134, P1181, DOI 10.1043/2009-0476-OA.1
   Kilic N, 2011, EUR J CANCER, V47, P1080, DOI 10.1016/j.ejca.2010.11.015
   Kitamura Y, 2013, J NEUROPATH EXP NEUR, V72, P816, DOI 10.1097/NEN.0b013e3182a065d0
   Le LP, 2011, PLOS ONE, V6, DOI 10.1371/journal.pone.0018846
   Lee J, 2012, J BONE JOINT SURG AM, V94A, P356, DOI 10.2106/JBJS.J.01784
   McMaster ML, 2001, CANCER CAUSE CONTROL, V12, P1, DOI 10.1023/A:1008947301735
   Palena C, 2014, ONCOIMMUNOLOGY, V3, DOI 10.4161/onci.27220
   Pillay N, 2012, NAT GENET, V44, P1185, DOI 10.1038/ng.2419
   Presneau N, 2011, J PATHOL, V223, P327, DOI 10.1002/path.2816
   Roselli M, 2012, CLIN CANCER RES, V18, P3868, DOI 10.1158/1078-0432.CCR-11-3211
   Shen J, 2011, J CLIN NEUROSCI, V18, P96, DOI 10.1016/j.jocn.2010.03.066
   Tirabosco R, 2008, AM J SURG PATHOL, V32, P572, DOI 10.1097/PAS.0b013e31815b693a
   Vujovic S, 2006, J PATHOL, V209, P157, DOI 10.1002/path.1969
   Wu Z, 2013, INT J MOL SCI, V14, P21258, DOI 10.3390/ijms141121258
   Wu Z, 2010, NEUROSURG REV, V33, P451, DOI 10.1007/s10143-010-0273-6
   Yang XHR, 2009, NAT GENET, V41, P1176, DOI 10.1038/ng.454
   Zhang LL, 2013, PLOS ONE, V8, DOI 10.1371/journal.pone.0075851
NR 21
TC 15
Z9 16
U1 1
U2 4
PU SPANDIDOS PUBL LTD
PI ATHENS
PA POB 18179, ATHENS, 116 10, GREECE
SN 1791-2997
EI 1791-3004
J9 MOL MED REP
JI Mol. Med. Rep.
PD SEP
PY 2015
VL 12
IS 3
BP 4298
EP 4304
DI 10.3892/mmr.2015.3976
PG 7
WC Oncology; Medicine, Research & Experimental
WE Science Citation Index Expanded (SCI-EXPANDED)
SC Oncology; Research & Experimental Medicine
GA CP5OW
UT WOS:000359933900150
PM 26099340
OA Green Published, hybrid, Green Submitted
DA 2023-08-27
ER

PT J
AU Whitmeyer, M
   Brahimaj, BC
   Beer-Furlan, A
   Alvi, S
   Epsten, MJ
   Crawford, F
   Byrne, RW
   Wiet, RM
AF Whitmeyer, Max
   Brahimaj, Bledi C.
   Beer-Furlan, Andre
   Alvi, Sameer
   Epsten, Madeline J.
   Crawford, Fred
   Byrne, Richard W.
   Wiet, R. Mark
TI Resection of vestibular schwannomas after stereotactic radiosurgery: a
   systematic review
SO JOURNAL OF NEUROSURGERY
LA English
DT Review
DE acoustic neuroma; microsurgery; salvage surgery; vestibular schwannoma;
   oncology; stereotactic radiosurgery
ID GAMMA-KNIFE RADIOSURGERY; SALVAGE SURGERY; TUMOR-CONTROL;
   RADIATION-THERAPY; ACOUSTIC NEUROMAS; NERVE FUNCTION; MICROSURGERY;
   MANAGEMENT; EPIDEMIOLOGY; PRESERVATION
AB OBJECTIVE Multiple short series have evaluated the efficacy of salvage microsurgery (MS) after stereotactic radiosurgery (SRS) for treatment of vestibular schwannomas (VSs); however, there is a lack of a large volume of patient data available for interpretation and clinical adaptation. The goal of this study was to provide a comprehensive review of tumor characteristics, management, and surgical outcomes of salvage of MS after SRS for VS. METHODS The Medline/PubMed, Scopus, CINAHL, Cochrane Library, and Google Scholar databases were queried according to PRISMA guidelines. All English-language and translated publications were included. Studies lacking adequate study characteristics and outcomes were excluded. Cases involving neurofibromatosis type 2, previous MS, or malignant transformation were excluded when possible. RESULTS Twenty studies containing 297 cases met inclusion criteria. Three additional cases from Rush University Medical Center were added for 300 total cases. Tumor growth with or without symptoms was the primary indication for salvage surgery (92.3% of cases), followed by worsening of symptoms without growth (4.6%) and cystic enlargement (3.1%). The average time to MS after SRS was 39.4 months. The average size and volume of tumor at surgery were 2.44 cm and 5.92 cm3, respectively. The surgical approach was retrosigmoid (42.8%) and translabyrinthine (57.2%); 59.5% of patients had a House-Brackmann (HB) grade of I or II. The facial nerve was preserved in 91.5% of cases. Facial nerve preservation and HB grades were lower for the translabyrinthine versus retrosigmoid approach (p = 0.31 and p = 0.18, respectively); however, fewer complications were noted in the translabyrinthine approach (p = 0.29). Gross-total resection (GTR) was completed in 55.7% of surgeries. Studies that predominantly used subtotal resection (STR) were associated with a lower rate of facial nerve injury (5.3% vs 11.3%, p = 0.07) and higher rate of HB grade I or II (72.9% vs 48.0%, p = 0.00003) versus those using predominantly GTR. However, majority STR was associated with a recurrence rate of 3.6% as compared to 1.4% for majority GTR (p = 0.29). CONCLUSIONS This study showed that the leading cause of MS after SRS was tumor growth at an average of 39.4 months after radiation. There were no significant differences in outcomes of facial nerve preservation, postoperative HB grade, or complication rate based on surgical approach. Patients who underwent STR showed statistically significant better HB outcomes compared with GTR. MS after SRS was considered by most authors to be more difficult than primary MS. These data support the notion that the surgical goals of salvage surgery are debulking of tumor mass, decreasing compression of the brainstem, and not necessarily pursuing GTR.
C1 [Whitmeyer, Max] Ohio State Univ, Coll Med, Columbus, OH USA.
   [Brahimaj, Bledi C.; Beer-Furlan, Andre; Byrne, Richard W.; Wiet, R. Mark] Rush Univ, Dept Neurol Surg, Med Ctr, Chicago, IL USA.
   [Alvi, Sameer] Northwestern Med, Geneva, NY USA.
   [Epsten, Madeline J.; Crawford, Fred] Rush Med Coll, Chicago, IL USA.
   [Wiet, R. Mark] Rush Univ, Dept Otolaryngol, Med Ctr, Chicago, IL USA.
C3 University System of Ohio; Ohio State University; Rush University; Rush
   University; Rush University
RP Wiet, RM (通讯作者)，Rush Univ, Med Ctr, Chicago, IL 60612 USA.
EM
CR Aboukais R, 2018, CLIN NEUROL NEUROSUR, V168, P77, DOI 10.1016/j.clineuro.2018.03.006
   Babu R, 2013, J NEUROSURG, V119, P121, DOI 10.3171/2013.1.JNS121370
   Battista RA, 2000, AM J OTOL, V21, P371, DOI 10.1016/S0196-0709(00)80047-2
   Breshears JD, 2017, OPER NEUROSURG, V13, P441, DOI 10.1093/ons/opx023
   Carlson ML, 2015, OTOLARYNG HEAD NECK, V153, P440, DOI 10.1177/0194599815590105
   Copeland WR, 2015, J NEUROSURG, V122, P312, DOI 10.3171/2014.10.JNS14432
   Couldwell WT, 2002, ACTA NEUROCHIR, V144, P1319, DOI 10.1007/s00701-002-1006-6
   Friedman RA, 2011, OTOL NEUROTOL, V32, P1322, DOI 10.1097/MAO.0b013e31822e5b76
   Gerganov VM, 2012, J NEUROSURG, V116, P713, DOI 10.3171/2011.12.JNS111682
   Hatch JL, 2018, OTOLARYNG HEAD NECK, V159, P102, DOI 10.1177/0194599818765717
   Hong BJ, 2014, OTOL NEUROTOL, V35, P171, DOI 10.1097/MAO.0000000000000174
   Hong WM, 2017, J KOREAN NEUROSURG S, V60, P165, DOI 10.3340/jkns.2013.0407.001
   Husseini ST, 2013, AM J OTOLARYNG, V34, P107, DOI 10.1016/j.amjoto.2012.09.012
   Iwai Y, 2016, WORLD NEUROSURG, V90, P385, DOI 10.1016/j.wneu.2016.03.014
   Kano H, 2010, INT J RADIAT ONCOL, V76, P520, DOI 10.1016/j.ijrobp.2009.01.076
   Kleijwegt M, 2016, OTOL NEUROTOL, V37, P1411, DOI 10.1097/MAO.0000000000001169
   Kondziolka D, 2012, NEUROSURG FOCUS, V33, DOI 10.3171/2012.6.FOCUS12192
   Kshettry VR, 2015, J NEURO-ONCOL, V124, P223, DOI 10.1007/s11060-015-1827-9
   Lee CC, 2014, J NEUROSURG, V121, P150, DOI 10.3171/2014.8.GKS141312
   Lee DJ, 2003, OTOL NEUROTOL, V24, P650, DOI 10.1097/00129492-200307000-00020
   Lee HJ, 2017, OTOL NEUROTOL, V38, P1339, DOI 10.1097/MAO.0000000000001536
   Limb CJ, 2005, LARYNGOSCOPE, V115, P93, DOI 10.1097/01.mlg.0000150686.93011.49
   Marinelli JP, 2018, OTOLARYNG HEAD NECK, V159, P717, DOI 10.1177/0194599818770629
   Mathieu D, 2007, NEUROSURGERY, V60, P460, DOI 10.1227/01.NEU.0000255340.26027.53
   Nagano O, 2008, J NEUROSURG, V109, P811, DOI 10.3171/JNS/2008/109/11/0811
   Nonaka Y, 2016, NEUROSURG REV, V39, P303, DOI 10.1007/s10143-015-0690-7
   Persson O, 2017, ACTA NEUROCHIR, V159, P1013, DOI 10.1007/s00701-017-3164-6
   Pollock BE, 1998, J NEUROSURG, V89, P949, DOI 10.3171/jns.1998.89.6.0949
   Roche Pierre-Hugues, 2008, Prog Neurol Surg, V21, P152, DOI 10.1159/000156902
   Schulder M, 1999, AM J OTOL, V20, P364
   Schumacher AJ, 2017, J NEUROL SURG PART B, V78, P2, DOI 10.1055/s-0036-1584231
   Shuto T, 2008, ACTA NEUROCHIR, V150, P229, DOI 10.1007/s00701-007-1486-5
   Shuto T, 2016, J CLIN NEUROSCI, V33, P239, DOI 10.1016/j.jocn.2016.05.022
   Stangerup SE, 2012, OTOLARYNG CLIN N AM, V45, P257, DOI 10.1016/j.otc.2011.12.008
   Taha I, 2020, ACTA NEUROCHIR, V162, P43, DOI 10.1007/s00701-019-04055-4
   Tsao MN, 2017, J RADIOSURGERY SBRT, V5, P5
   Wise SC, 2016, LARYNGOSCOPE, V126, P2580, DOI 10.1002/lary.25943
NR 37
TC 4
Z9 4
U1 1
U2 3
PU AMER ASSOC NEUROLOGICAL SURGEONS
PI ROLLING MEADOWS
PA 5550 MEADOWBROOK DRIVE, ROLLING MEADOWS, IL 60008 USA
SN 0022-3085
EI 1933-0693
J9 J NEUROSURG
JI J. Neurosurg.
PD SEP
PY 2021
VL 135
IS 3
BP 881
EP 889
DI 10.3171/2020.7.JNS2044
PG 9
WC Clinical Neurology; Surgery
WE Science Citation Index Expanded (SCI-EXPANDED)
SC Neurosciences & Neurology; Surgery
GA UL3EX
UT WOS:000692539600007
PM 34331121
OA Bronze
DA 2023-08-27
ER

PT J
AU El-Waseif, EG
   Sharawy, MH
   Suddek, GM
AF El-Waseif, Esraa G.
   Sharawy, Maha H.
   Suddek, Ghada M.
TI The modulatory effect of sodium molybdate against cisplatin-induced CKD:
   Role of TGF-beta/Smad signaling pathway
SO LIFE SCIENCES
LA English
DT Article
DE Sodium molybdate; Chronic kidney disease (CKD); Cisplatin; TGF-beta/Smad
   signaling; Tumor necrosis factor-alpha (TNF-alpha)
ID CHRONIC KIDNEY-DISEASE; INDUCED RENAL INJURY; GROWTH-FACTOR-BETA;
   FACTOR-KAPPA-B; MOLECULAR-MECHANISMS; LIPID-PEROXIDATION;
   ANGIOTENSIN-II; NEPHROTOXICITY; FIBROSIS; NEPHROPATHY
AB Aims: Chronic kidney disease (CKD) is a global non-communicable health problem. Fibrosis is considered the base and the fate of CKD; thus, the goal of this study was to evaluate the effects of sodium molybdate on cisplatininduced CKD model and demonstrate the possible involved mechanisms.
   Materials and methods: In cisplatin model, Wistar rats were challenged with cisplatin (1 mg/kg, i.p.) twice weekly for ten weeks. Sodium molybdate (100 and 200 mg/kg, orally) was given one-week prior cisplatin and was continued daily for the next ten weeks.
   Key findings Administration of sodium molybdate amended kidney function and significantly reduced the pathological changes in renal histopathological sections. Moreover, it modulates oxidative stress parameters by increasing the levels of SOD and GSH and decreasing the level of MDA. Likewise, in renal homogenate, sodium molybdate attenuated inflammation that was revealed by NF-kappa B and TNF-alpha levels significant reduction. Additionally, it ameliorated fibrosis that was indicated by decreased Masson's trichome staining in cortex and medulla. Immunohistochemical staining of renal sections against TGF-beta 1 showed reduction of positive glomerular and tubular protein expression. Additionally, it decreased profibmtic Smad3 level and increased antifibrotic Smad7 level.
   Significance: Administration of sodium molybdate demonstrated a hopeful suppressor effect on cisplatin-induced CKD/fibrosis in rat model. This effect may be through the inhibition pof TGF-p/Smad signaling pathway and upregulation of Smad7 expression leading to decreased extracellular collagen accumulation. Moreover, they could ameliorate the inflammation through NF-kappa B and TNF-alpha levels reduction, decrease in ROS, and retrieval of antioxidant defenses.
C1 [El-Waseif, Esraa G.; Sharawy, Maha H.; Suddek, Ghada M.] Mansoura Univ, Fac Pharm, Dept Pharmacol & Toxicol, Mansoura 35516, Egypt.
C3 Egyptian Knowledge Bank (EKB); Mansoura University
RP Sharawy, MH (通讯作者)，Mansoura Univ, Fac Pharm, Dept Pharmacol & Toxicol, Mansoura 35516, Egypt.
EM
RI Suddek, Ghada/HPE-4718-2023
OI El-Waseif, Esraa/0000-0002-3079-4441; Suddek, Ghada/0000-0002-3283-9471
CR Abdel-Daim MM, 2017, N-S ARCH PHARMACOL, V390, P301, DOI 10.1007/s00210-016-1332-5
   Ale-Ebrahim M, PROTECTIVE EFFECTS S
   Ale-Ebrahim M, 2015, J TRACE ELEM MED BIO, V29, P242, DOI 10.1016/j.jtemb.2014.07.002
   Anke M., 1985, NUTR RES, V1, pS180
   Arany I, 2003, SEMIN NEPHROL, V23, P460, DOI 10.1016/S0270-9295(03)00089-5
   Bae EH, 2014, PLOS ONE, V9, DOI 10.1371/journal.pone.0086553
   Baliga R, 1998, KIDNEY INT, V53, P394, DOI 10.1046/j.1523-1755.1998.00767.x
   Bancroft J., 1982, THEORY PRACTICE HIST
   Barnes PJ, 1997, NEW ENGL J MED, V336, P1066, DOI 10.1056/NEJM199704103361506
   Bhat ZY, 2015, PLOS ONE, V10, DOI 10.1371/journal.pone.0142225
   Border WA, 1998, HYPERTENSION, V31, P181, DOI 10.1161/01.HYP.31.1.181
   Bottinger EP, 2002, J AM SOC NEPHROL, V13, P2600, DOI 10.1097/01.ASN.0000033611.79556.AE
   BRADFORD MM, 1976, ANAL BIOCHEM, V72, P248, DOI 10.1016/0003-2697(76)90527-3
   Dasari S, 2014, EUR J PHARMACOL, V740, P364, DOI 10.1016/j.ejphar.2014.07.025
   Davis CA, 2001, J AM SOC NEPHROL, V12, P2683, DOI 10.1681/ASN.V12122683
   Dekkers IA, 2013, CLIN J AM SOC NEPHRO, V8, P922, DOI 10.2215/CJN.09980912
   Deng JP, 2001, KIDNEY INT, V60, P2118, DOI 10.1046/j.1523-1755.2001.00043.x
   Derynck R, 2003, NATURE, V425, P577, DOI 10.1038/nature02006
   Djudjaj S, 2019, MOL ASPECTS MED, V65, P16, DOI 10.1016/j.mam.2018.06.002
   Duffield JS, 2014, J CLIN INVEST, V124, P2299, DOI 10.1172/JCI72267
   Ebisawa T, 2001, J BIOL CHEM, V276, P12477, DOI 10.1074/jbc.C100008200
   Eidi A, 2011, J TRACE ELEM MED BIO, V25, P67, DOI 10.1016/j.jtemb.2010.12.003
   Eknoyan G, 2002, AM J KIDNEY DIS, V39, pS14, DOI 10.1053/ajkd.2002.30939
   ELLMAN GL, 1959, ARCH BIOCHEM BIOPHYS, V82, P70, DOI 10.1016/0003-9861(59)90090-6
   Faig J, 2018, AM J CLIN ONCOL-CANC, V41, P432, DOI 10.1097/COC.0000000000000301
   Flanders KC, 2004, INT J EXP PATHOL, V85, P47, DOI 10.1111/j.0959-9673.2004.00377.x
   Gagliardini E, 2006, CYTOKINE GROWTH F R, V17, P89, DOI 10.1016/j.cytogfr.2005.09.005
   Gajjala PR, 2015, FRONT IMMUNOL, V6, DOI 10.3389/fimmu.2015.00340
   GUTTERIDGE JMC, 1995, CLIN CHEM, V41, P1819
   Gutteridge JMC, 1999, BRIT MED BULL, V55, P49, DOI 10.1258/0007142991902295
   Hardy D., 2000, BOOK REV TIETZ TXB C, V3
   INOUE M, 1987, J PROTEIN CHEM, V6, P207
   KAGAMI S, 1994, J CLIN INVEST, V93, P2431, DOI 10.1172/JCI117251
   Karasawa T, 2015, TOXICOL LETT, V237, P219, DOI 10.1016/j.toxlet.2015.06.012
   Katagiri D, 2016, KIDNEY INT, V89, P374, DOI 10.1038/ki.2015.327
   Kavsak P, 2000, MOL CELL, V6, P1365, DOI 10.1016/S1097-2765(00)00134-9
   Kidney International Supplements, 2013, KIDNEY INT SUPPL, V3, P150, DOI [10.1038/kisup.2012.73, DOI 10.1038/KISUP.2012.74, DOI 10.1038/KISUP.2012.73]
   Klahr S, 2002, AM J PHYSIOL-RENAL, V283, pF861, DOI 10.1152/ajprenal.00362.2001
   Lan HY, 2008, FRONT BIOSCI-LANDMRK, V13, P4984, DOI 10.2741/3057
   Lan HY, 2011, INT J BIOL SCI, V7, P1056, DOI 10.7150/ijbs.7.1056
   Latcha S, 2016, CLIN J AM SOC NEPHRO, V11, P1173, DOI 10.2215/CJN.08070715
   Lee YJ, 2020, J FOOD DRUG ANAL, V28, P103, DOI 10.1016/j.jfda.2019.10.001
   Levey AS, 2015, JAMA-J AM MED ASSOC, V313, P837, DOI 10.1001/jama.2015.0602
   Levey AS, 2009, AM J KIDNEY DIS, V53, pS4, DOI 10.1053/j.ajkd.2008.07.048
   Levey AS, 2005, KIDNEY INT, V67, P2089, DOI 10.1111/j.1523-1755.2005.00365.x
   Li CZ, 2019, FREE RADICAL BIO MED, V130, P512, DOI 10.1016/j.freeradbiomed.2018.11.006
   Li JH, 2010, DIABETES, V59, P2612, DOI 10.2337/db09-1631
   Li S, 2019, EXP CELL RES, V383, DOI 10.1016/j.yexcr.2019.07.001
   Liu YM, 2012, J PHARMACOL EXP THER, V341, P725, DOI 10.1124/jpet.111.191247
   Gorriz JL, 2012, TRANSPLANT REV-ORLAN, V26, P3, DOI 10.1016/j.trre.2011.10.002
   Maghmomeh AO, 2020, N-S ARCH PHARMACOL, V393, P303, DOI 10.1007/s00210-019-01734-y
   Manohar S, 2018, J NEPHROL, V31, P15, DOI 10.1007/s40620-017-0392-z
   Mao H, 2016, COLORECTAL DIS, V18, P1172, DOI 10.1111/codi.13400
   MARCUSSEN N, 1990, APMIS, V98, P1087, DOI 10.1111/j.1699-0463.1990.tb05039.x
   MARKLUND S, 1974, EUR J BIOCHEM, V47, P469
   McCarty MF, 2006, MED HYPOTHESES, V67, P1277, DOI 10.1016/j.mehy.2004.11.048
   Meran S, 2011, INT J EXP PATHOL, V92, P158, DOI 10.1111/j.1365-2613.2011.00764.x
   Metcalfe W, 2007, NEPHROL DIAL TRANSPL, V22, P26, DOI 10.1093/ndt/gfm446
   mHenry, 1964, CLIN CHEM PRINCIPLES
   Miller RP, 2010, TOXINS, V2, P2490, DOI 10.3390/toxins2112490
   OHKAWA H, 1979, ANAL BIOCHEM, V95, P351, DOI 10.1016/0003-2697(79)90738-3
   Ozkok A, 2014, BIOMED RES INT, V2014, DOI 10.1155/2014/967826
   Pabla N, 2008, KIDNEY INT, V73, P994, DOI 10.1038/sj.ki.5002786
   Panneerselvam S, 2003, CHEM-BIOL INTERACT, V145, P159, DOI 10.1016/S0009-2797(02)00254-5
   Panneerselvam SR, 2004, CLIN CHIM ACTA, V345, P93, DOI 10.1016/j.cccn.2004.03.005
   Park JW, 2012, EUR J PHARMACOL, V683, P301, DOI 10.1016/j.ejphar.2012.03.019
   Park JS, 2013, NEPHROL DIAL TRANSPL, V28, P592, DOI 10.1093/ndt/gfs496
   PENNINGTON JAT, 1987, J AM DIET ASSOC, V87, P1644
   Peres Luis Alberto Batista, 2013, Braz. J. Nephrol., V35, P332, DOI 10.5935/0101-2800.20130052
   Razzaque MS, 1998, NEPHRON, V80, P434, DOI 10.1159/000045217
   RIES F, 1986, AM J KIDNEY DIS, V8, P368, DOI 10.1016/S0272-6386(86)80112-3
   Sasaki A, 2019, J TOXICOL PATHOL, V32, P79, DOI 10.1293/tox.2018-0049
   SCHROEDER HA, 1962, J CHRON DIS, V15, P51, DOI 10.1016/0021-9681(62)90101-7
   Shahbaz H., 2019, CREATININE CLEARANCE
   Sharawy MH, 2020, LIFE SCI, V260, DOI 10.1016/j.lfs.2020.118454
   Sharp CN, 2019, AM J PHYSIOL-RENAL, V316, pF162, DOI 10.1152/ajprenal.00463.2018
   Sharp CN, 2018, AM J PHYSIOL-RENAL, V315, pF161, DOI 10.1152/ajprenal.00636.2017
   Sharp CN, 2016, AM J PHYSIOL-RENAL, V310, pF560, DOI 10.1152/ajprenal.00512.2015
   Shiraishi F, 2000, AM J PHYSIOL-RENAL, V278, pF726, DOI 10.1152/ajprenal.2000.278.5.F726
   Tomino Y, 2014, KIDNEY BLOOD PRESS R, V39, P450, DOI 10.1159/000368458
   Torres R, 2016, J AM SOC NEPHROL, V27, P1102, DOI 10.1681/ASN.2015010079
   Tsang RY, 2009, DRUG SAFETY, V32, P1109, DOI 10.2165/11316640-000000000-00000
   UNDERWOOD E J, 1971, P543
   van de Vlekkert D, 2020, BIO-PROTOCOL, V10, DOI 10.21769/BioProtoc.3629
   Wang WS, 2005, J AM SOC NEPHROL, V16, P1371, DOI 10.1681/ASN.2004121070
   WANG XJ, 1992, J NUTR, V122, P1036, DOI 10.1093/jn/122.4.1036
   WRANA JL, 1994, NATURE, V370, P341, DOI 10.1038/370341a0
   Yamate J, 2004, EXP TOXICOL PATHOL, V56, P13, DOI 10.1016/j.etp.2004.04.008
   Yang Y, 2014, ARCH TOXICOL, V88, P1249, DOI 10.1007/s00204-014-1239-1
NR 89
TC 1
Z9 1
U1 1
U2 1
PU PERGAMON-ELSEVIER SCIENCE LTD
PI OXFORD
PA THE BOULEVARD, LANGFORD LANE, KIDLINGTON, OXFORD OX5 1GB, ENGLAND
SN 0024-3205
EI 1879-0631
J9 LIFE SCI
JI Life Sci.
PD OCT 1
PY 2022
VL 306
AR 120845
DI 10.1016/j.lfs.2022.120845
PG 12
WC Medicine, Research & Experimental; Pharmacology & Pharmacy
WE Science Citation Index Expanded (SCI-EXPANDED)
SC Research & Experimental Medicine; Pharmacology & Pharmacy
GA 6R7CB
UT WOS:000892455800002
PM 35917941
DA 2023-08-27
ER

PT J
AU Chang, JW
   Chang, JH
   Park, YG
   Chung, SS
AF Chang, Jin Woo
   Chang, Jong Hee
   Park, Yong Gou
   Chung, Sang Sup
TI Gamma knife radiosurgery for idiopathic and secondary trigeminal
   neuralgia
SO JOURNAL OF NEUROSURGERY
LA English
DT Article
DE gamma knife; radiosurgery; trigeminal neuralgia
ID STEREOTACTIC RADIOSURGERY
AB Object. The aim of this study was to identify pain relief and treatment morbidity following gamma knife radiosurgery (GKS) for idiopathic and secondary trigeminal neuralgia.
   Methods. Between May 1992 and December 1999, 15 patients with idiopathic trigeminal neuralgia and 38 patients with secondary trigeminal neuralgia were treated with GKS. Pain improvement was achieved in 13 of the patients with idiopathic pain (pain response rate 86.7%). Seven patients were pain free and another six experienced pain reduction. There were no serious complications; however, two patients suffered a mild facial sensory change 8 months and 9 months, respectively, after GKS.
   The patients with secondary trigeminal neuralgia were divided into two groups (Group 1, 32 patients in whom the trigeminal root entry zone [REZ] near the tumor could not be visualized; and Group II, six patients in whom the trigeminal REZ near the tumor or brainstem lesion could be visualized).
   In Group I, the pain subsided completely in eight patients and was reduced in seven (pain response rate 46.9%). In Group H, the pain subsided completely in one patient at 2.8 months and was reduced in three patients at a mean follow up of 0.8 months (range 0.6-1 month) after GKS. The pain response rate was 66.7%.
   Conclusions. The authors believe that GKS is an effective treatment modalities for idiopathic and secondary trigeminal neuralgia, particularly in patients with inoperable lesions.
C1 [Chang, Jin Woo; Chang, Jong Hee; Park, Yong Gou; Chung, Sang Sup] Yonsei Univ, Coll Med, Dept Neurosurg, Seoul, South Korea.
C3 Yonsei University; Yonsei University Health System
RP Park, YG (通讯作者)，Yonsei Univ, Coll Med, Dept Neurosurg, CPO Box 8044, Seoul, South Korea.
EM
CR Barker FG, 1996, J NEUROSURG, V84, P818, DOI 10.3171/jns.1996.84.5.0818
   BULLITT E, 1986, J NEUROSURG, V64, P865, DOI 10.3171/jns.1986.64.6.0865
   Chang JW, 1999, STEREOT FUNCT NEUROS, V72, P29, DOI 10.1159/000056436
   CHENG TMW, 1993, NEUROLOGY, V43, P2298, DOI 10.1212/WNL.43.11.2298
   Kondziolka D, 1996, J NEUROSURG, V84, P940, DOI 10.3171/jns.1996.84.6.0940
   Kondziolka D, 1998, PROG NEUROL, V14, P212
   LEKSELL L, 1951, ACTA CHIR SCAND, V102, P316
   Lovely TJ, 1997, NEUROSURG CLIN N AM, V8, P11
   Marco JKD, 1997, NEUROSURG CLIN N AM, V8, P103
   REGIS J, 1995, STEREOT FUNCT NEUROS, V64, P182, DOI 10.1159/000098778
   Regis J, 1999, NEUROSURG CLIN N AM, V10, P359, DOI 10.1016/S1042-3680(18)30201-8
   Regis J, 1997, STEREOTACT FUNCT S1, V64, P210
   Urgosik D, 1998, STEREOT FUNCT NEUROS, V70, P200, DOI 10.1159/000056423
   Young RF, 1997, NEUROLOGY, V48, P608, DOI 10.1212/WNL.48.3.608
NR 14
TC 0
Z9 0
U1 0
U2 0
PU AMER ASSOC NEUROLOGICAL SURGEONS
PI ROLLING MEADOWS
PA 5550 MEADOWBROOK DRIVE, ROLLING MEADOWS, IL 60008 USA
SN 0022-3085
EI 1933-0693
J9 J NEUROSURG
JI J. Neurosurg.
PD DEC
PY 2011
VL 115
SU S
BP 103
EP 107
PG 5
WC Clinical Neurology; Surgery
WE Science Citation Index Expanded (SCI-EXPANDED)
SC Neurosciences & Neurology; Surgery
GA 853TY
UT WOS:000297449800019
DA 2023-08-27
ER

PT J
AU Geist, RT
   Reddy, AJ
   Zhang, J
   Gutmann, DH
AF Geist, RT
   Reddy, AJ
   Zhang, J
   Gutmann, DH
TI Expression of the tuberous sclerosis 2 gene product, tuberin, in adult
   and developing nervous system tissues
SO NEUROBIOLOGY OF DISEASE
LA English
DT Article
ID EKER RAT MODEL; TSC2 GENE; IDENTIFICATION; HETEROGENEITY
AB Tuberous sclerosis (TS) is an autosomal dominant disorder in which affected individuals manifest mental retardation, seizures, and a variety of benign and malignant tumors. The TSC2 tumor suppressor gene was recently identified by positional cloning and its protein product, tuberin, shown to represent one member of the rap GTPase activating protein (rapGAP) family. In order to determine the contribution of tuberin to the development of mental retardation and seizures in patients with TS, we examined the expression of tuberin in adult and developing nervous system tissues. Since tuberin is the second rapGAP found in the nervous system, the expression of tuberin was compared to the expression of rapGAP, rap1, and rap2. In this study, we demonstrate that tuberin is expressed at greatest levels in the spinal cord and cerebellum as opposed to rapGAP, which is not enriched in these tissues. Tuberin expression in the adult CNS is restricted to the olfactory bulb, several CNS neuronal populations, brainstem nuclei, cerebellar Purkinje cells, and motor neurons in the ventral spinal cord. In contrast, rapGAP is expressed in many different cell types in the adult CNS, but not in cerebellar Purkinje cells or motor neurons in the ventral spinal cord. However, there is significant expression of rapGAP in astrocytes. The restricted distribution of tuberin expression relative to rap1 and rapGAP suggests that tuberin may be the primary rap1 regulator in a subpopulation of CNS neurons. (C) 1996 Academic Press, Inc.
C1 WASHINGTON UNIV,SCH MED,DEPT PEDIAT,ST LOUIS,MO 63110.
   WASHINGTON UNIV,SCH MED,DEPT GENET,ST LOUIS,MO 63110.
C3 Washington University (WUSTL); Washington University (WUSTL)
RP Geist, RT (通讯作者)，WASHINGTON UNIV,SCH MED,DEPT NEUROL,ST LOUIS,MO 63110, USA.
OI Gutmann, David/0000-0002-3127-5045
CR BASU TN, 1992, NATURE, V356, P713, DOI 10.1038/356713a0
   *EUR CHROM 16 TUB, 1993, CELL, V75, P1305
   GEIST RT, 1995, CELL GROWTH DIFFER, V6, P1477
   Gomez M. R., 1988, TUBEROUS SCLEROSIS
   GREEN AJ, 1993, AM J HUM GENET, V53, P244
   GUTMANN DH, 1993, J NEUROSCI RES, V36, P216, DOI 10.1002/jnr.490360212
   GUTMANN DH, 1991, P NATL ACAD SCI USA, V88, P9658, DOI 10.1073/pnas.88.21.9658
   HAINES JL, 1991, AM J HUM GENET, V49, P764
   HEWETT SJ, 1994, NEURON, V13, P487, DOI 10.1016/0896-6273(94)90362-X
   JANSSEN LAJ, 1991, ANN NY ACAD SCI, V615, P306
   KOBAYASHI T, 1995, NUCLEIC ACIDS RES, V23, P2608, DOI 10.1093/nar/23.14.2608
   KOBAYASHI T, 1995, NAT GENET, V9, P70, DOI 10.1038/ng0195-70
   OSBORNE JP, 1991, ANN NY ACAD SCI, V615, P125, DOI 10.1111/j.1749-6632.1991.tb37754.x
   Tsuchiya H, 1996, CANCER RES, V56, P429
   WIENECKE R, 1995, J BIOL CHEM, V270, P16409, DOI 10.1074/jbc.270.27.16409
   XIAO GH, 1995, CELL GROWTH DIFFER, V6, P1185
   XU L, 1995, GENOMICS, V27, P475, DOI 10.1006/geno.1995.1079
   YEUNG RS, 1994, P NATL ACAD SCI USA, V91, P11413, DOI 10.1073/pnas.91.24.11413
   YOSHIDA Y, 1992, MOL CELL BIOL, V12, P3407, DOI 10.1128/MCB.12.8.3407
NR 19
TC 21
Z9 21
U1 0
U2 1
PU ACADEMIC PRESS INC JNL-COMP SUBSCRIPTIONS
PI SAN DIEGO
PA 525 B ST, STE 1900, SAN DIEGO, CA 92101-4495
SN 0969-9961
J9 NEUROBIOL DIS
JI Neurobiol. Dis.
PD APR
PY 1996
VL 3
IS 2
BP 111
EP 120
DI 10.1006/nbdi.1996.0011
PG 10
WC Neurosciences
WE Science Citation Index Expanded (SCI-EXPANDED)
SC Neurosciences & Neurology
GA VU961
UT WOS:A1996VU96100002
PM 9173918
DA 2023-08-27
ER

PT J
AU Song, A
   Cho, GW
   Moon, C
   Park, I
   Jang, CH
AF Song, Anji
   Cho, Gwang-Won
   Moon, Changjong
   Park, Ilyong
   Jang, Chul Ho
TI Protective Effect of Resveratrol in an Experimental Model of
   Salicylate-Induced Tinnitus
SO INTERNATIONAL JOURNAL OF MOLECULAR SCIENCES
LA English
DT Article
DE resveratrol; NMDA receptor; salicylate; excitotoxicity; tinnitus
ID AUDITORY-CORTEX; EXPRESSION; NEUROTOXICITY; PICEATANNOL; OTOTOXICITY;
   IMPAIRMENT; METABOLITE; RECEPTORS; MEMANTINE; NEURONS
AB To date, the effect of resveratrol on tinnitus has not been reported. The attenuative effects of resveratrol (RSV) on a salicylate-induced tinnitus model were evaluated by in vitro and in vivo experiments. The gene expression of the activity-regulated cytoskeleton-associated protein (ARC), tumor necrosis factor-alpha (TNF alpha), and NMDA receptor subunit 2B (NR2B) in SH-SY5Y cells was examined using qPCR. Phosphorylated cAMP response element-binding protein (p-CREB), apoptosis markers, and reactive oxygen species (ROS) were evaluated by in vitro experiments. The in vivo experiment evaluated the gap-prepulse inhibition of the acoustic startle reflex (GPIAS) and auditory brainstem response (ABR) level. The NR2B expression in the auditory cortex (AC) was determined by immunohistochemistry. RSV significantly reduced the salicylate-induced expression of NR2B, ARC, and TNF alpha in neuronal cells; the GPIAS and ABR thresholds altered by salicylate in rats were recovered close to their normal range. RSV also reduced the salicylate-induced NR2B overexpression of the AC. These results confirmed that resveratrol exerted an attenuative effect on salicylate-induced tinnitus and may have a therapeutic potential.
C1 [Song, Anji; Cho, Gwang-Won] Chosun Univ, Coll Nat Sci, Dept Biol, Gwangju 61452, South Korea.
   [Song, Anji; Cho, Gwang-Won] Chosun Univ, Dept Integrat Biol Sci, BK21 FOUR Educ Res Grp Age Associated Disorder Co, Gwangju 61452, South Korea.
   [Moon, Changjong] Chonnam Natioanal Univ, Coll Vet Med, Dept Vet Anat, Gwanjgu 61186, South Korea.
   [Moon, Changjong] Chonnam Natioanal Univ, BK21 FOUR Program, Gwanjgu 61186, South Korea.
   [Park, Ilyong] Dankook Univ, Coll Med, Dept Biomed Engn, Cheonan 31116, South Korea.
   [Jang, Chul Ho] Chonnam Natl Univ, Med Sch, Dept Otolaryngol, Gwangju 61469, South Korea.
C3 Chosun University; Chosun University; Dankook University; Chonnam
   National University
RP Jang, CH (通讯作者)，Chonnam Natl Univ, Med Sch, Dept Otolaryngol, Gwangju 61469, South Korea.
EM
RI Jang, Chul/AAJ-8790-2020; Moon, Changjong/AAI-2005-2020
OI Jang, Chul/0000-0003-4084-2301; Moon, Changjong/0000-0003-2451-0374;
   Cho, Gwang-won/0000-0002-2149-5095; Park, Ilyong/0000-0003-1613-4209
FU National Research Foundation of Korea (NRF) - Korea government (MSIT)
   [2021RIF1A1060212]
FX This work was supported by the National Research Foundation of Korea
   (NRF) grant funded by the Korea government (MSIT) (No.
   2021RIF1A1060212).
CR Alvan G, 2017, BASIC CLIN PHARMACOL, V120, P5, DOI 10.1111/bcpt.12640
   Bing D, 2015, CELL PHYSIOL BIOCHEM, V35, P1905, DOI 10.1159/000374000
   Chang Y, 2009, NEUROCHEM INT, V54, P135, DOI 10.1016/j.neuint.2008.11.001
   Chen GD, 2013, HEARING RES, V295, P100, DOI 10.1016/j.heares.2012.11.016
   Corpas R, 2018, J PINEAL RES, V65, DOI 10.1111/jpi.12515
   Cui WM, 2019, MOL MED REP, V19, P51, DOI 10.3892/mmr.2018.9624
   Deng LL, 2013, NEUROTOX RES, V24, P307, DOI 10.1007/s12640-013-9384-5
   Galazyuk A, 2015, FRONT NEUROL, V6, DOI 10.3389/fneur.2015.00088
   Goncalves JT, 2016, CELL, V167, P897, DOI 10.1016/j.cell.2016.10.021
   Gong N, 2008, NEUROPHARMACOLOGY, V54, P454, DOI 10.1016/j.neuropharm.2007.10.017
   Gupta K, 2013, NEUROSCI LETT, V543, P95, DOI 10.1016/j.neulet.2013.03.010
   Hong J, 2018, NEURAL PLAST, V2018, DOI 10.1155/2018/3989201
   Hosoda R, 2021, J PHARMACOL EXP THER, V376, P385, DOI 10.1124/jpet.120.000096
   Jang CH, 2019, EXP NEUROBIOL, V28, P495, DOI 10.5607/en.2019.28.4.495
   JASTREBOFF PJ, 1988, BEHAV NEUROSCI, V102, P811, DOI 10.1037/0735-7044.102.6.811
   Karthick C, 2016, FRONT MOL NEUROSCI, V9, DOI 10.3389/fnmol.2016.00028
   Kim HT, 2021, KOREAN J PHYSIOL PHA, V25, P51, DOI 10.4196/kjpp.2021.25.1.51
   Lee SH, 2015, INT J PEDIATR OTORHI, V79, P58, DOI 10.1016/j.ijporl.2014.11.008
   Liu JX, 2003, HEARING RES, V175, P45, DOI 10.1016/S0378-5955(02)00708-6
   Lobarinas E, 2013, HEARING RES, V295, P150, DOI 10.1016/j.heares.2012.06.001
   Norena AJ, 2013, HEARING RES, V295, P161, DOI 10.1016/j.heares.2012.09.010
   Nunes S, 2018, NUTR RES REV, V31, P85, DOI 10.1017/S095442241700021X
   Palmer BF, 2020, NEW ENGL J MED, V382, P2544, DOI 10.1056/NEJMra2010852
   Pardo L, 2017, SCI REP-UK, V7, DOI 10.1038/s41598-017-06231-x
   Qin DX, 2019, NEUROTOX RES, V35, P838, DOI 10.1007/s12640-019-0006-8
   Ralli M, 2014, ACTA OTORHINOLARYNGO, V34, P198
   Scasso F, 2017, HELIYON, V3, DOI 10.1016/j.heliyon.2017.e00251
   Segal JA, 1999, BRAIN RES, V815, P270, DOI 10.1016/S0006-8993(98)01123-8
   Shaito A, 2020, INT J MOL SCI, V21, DOI 10.3390/ijms21062084
   Sheppard A, 2014, ACTA OTORHINOLARYNGO, V34, P79
   Son Y, 2013, AMINO ACIDS, V45, P393, DOI 10.1007/s00726-013-1518-9
   Tabuchi K, 2011, CURR MED CHEM, V18, P4866, DOI 10.2174/092986711797535254
   Tang DM, 2019, ADV EXP MED BIOL, V1130, P109, DOI 10.1007/978-981-13-6123-4_7
   Wu C, 2018, AM J TRANSL RES, V10, P3941
   Wu Y, 2020, EXP EYE RES, V200, DOI 10.1016/j.exer.2020.108249
   Xia N, 2017, BRIT J PHARMACOL, V174, P1633, DOI 10.1111/bph.13492
   Yi B, 2018, OTOL NEUROTOL, V39, pE173, DOI 10.1097/MAO.0000000000001717
   Yi B, 2016, SCI REP-UK, V6, DOI 10.1038/srep24428
   Zhao JH, 2018, MOL CELL BIOCHEM, V448, P71, DOI 10.1007/s11010-018-3314-z
NR 39
TC 1
Z9 1
U1 1
U2 1
PU MDPI
PI BASEL
PA ST ALBAN-ANLAGE 66, CH-4052 BASEL, SWITZERLAND
EI 1422-0067
J9 INT J MOL SCI
JI Int. J. Mol. Sci.
PD NOV
PY 2022
VL 23
IS 22
AR 14183
DI 10.3390/ijms232214183
PG 12
WC Biochemistry & Molecular Biology; Chemistry, Multidisciplinary
WE Science Citation Index Expanded (SCI-EXPANDED)
SC Biochemistry & Molecular Biology; Chemistry
GA 6K6ET
UT WOS:000887593500001
PM 36430660
OA gold, Green Published
DA 2023-08-27
ER

PT J
AU Paisansathan, C
   Koenig, HM
   Wheeler, PJ
   Baughman, VL
   Hoffman, WE
AF Paisansathan, C
   Koenig, HM
   Wheeler, PJ
   Baughman, VL
   Hoffman, WE
TI Loss of SSEP during sitting craniotomy
SO JOURNAL OF NEUROSURGICAL ANESTHESIOLOGY
LA English
DT Article
DE somatosensory evoked potential; sitting cramotomy; venous air embolism
ID EVOKED-POTENTIALS; SURGERY
AB A 54-year-old woman with a past medical history of asthma and depression presented with fight side hearing loss and ataxia. She was scheduled for a sitting craniotomy for cerebellopontine angle tumor resection. Somatosensory evoked potential, brainstem auditory evoked response, and facial nerve EMG were monitored intraoperatively. Approximately 30 minutes into the case, there was an episode of air embolism, which resolved after the source was identified and treated. Near the conclusion of the case, there was an abrupt loss of the right cortical somatosensory evoked potential signal, which never returned to baseline. A postoperative CT scan showed a substantial amount of subarachnoid air and intraventricular air in the frontal and temporal regions. The patient awakened in the ICU with no new neurologic deficit besides preoperative hearing loss on the right side. Despite the high specificity of somatosensory evoked potential change associated with postoperative neurodeficit when the change never returns to the baseline, there was no postoperative neurologic deficit in this patient. This case indicates the false-positive somatosensory evoked potentials caused by pneumocephalus in the sitting position.
C1 Univ Illinois, Chicago Hosp, Dept Anesthesiol, Chicago, IL 60612 USA.
C3 University of Illinois System; University of Illinois Chicago;
   University of Illinois Chicago Hospital
RP Paisansathan, C (通讯作者)，Univ Illinois, Chicago Hosp, Dept Anesthesiol, MC 515,1740 W Taylor St,Suite 3200, Chicago, IL 60612 USA.
RI Koenig, Heidi/L-6169-2019
CR Bejjani GK, 1998, NEUROSURGERY, V43, P491, DOI 10.1097/00006123-199809000-00050
   CRONIIN AJ, 2002, NEAR MISSES NEUROANE, P165
   Dawson G. D., 1947, JOUR NEUROL NEUROSURG AND PSYCHIAT, V10, P137
   GENTILI F, 1985, CAN J NEUROL SCI, V12, P336, DOI 10.1017/S0317167100035484
   HITSELBERGER WE, 1980, ARCH OTOLARYNGOL, V106, P69
   MONGAN PD, 1995, ANESTHESIOLOGY, V83, P1104, DOI 10.1097/00000542-199511000-00025
   Sonoo M, 2000, J CLIN NEUROPHYSIOL, V17, P258, DOI 10.1097/00004691-200005000-00004
NR 7
TC 5
Z9 5
U1 0
U2 0
PU LIPPINCOTT WILLIAMS & WILKINS
PI PHILADELPHIA
PA 530 WALNUT ST, PHILADELPHIA, PA 19106-3621 USA
SN 0898-4921
J9 J NEUROSURG ANESTH
JI J. Neurosurg. Anesthesiol.
PD OCT
PY 2003
VL 15
IS 4
BP 327
EP 329
DI 10.1097/00008506-200310000-00007
PG 3
WC Anesthesiology; Clinical Neurology; Surgery
WE Science Citation Index Expanded (SCI-EXPANDED)
SC Anesthesiology; Neurosciences & Neurology; Surgery
GA 728HF
UT WOS:000185711200007
PM 14508175
DA 2023-08-27
ER

PT J
AU Wang, K
   Wang, L
   Tian, KB
   Xiao, XR
   Wu, Z
   Jia, GJ
   Zhang, LW
   Zhang, JT
AF Wang, Ke
   Wang, Liang
   Tian, Kaibing
   Xiao, Xinru
   Wu, Zhen
   Jia, Guijun
   Zhang, Liwei
   Zhang, Junting
TI Surgical resection of upper-middle clivus chordomas via a modified
   anterior transpetrous approach
SO CLINICAL NEUROLOGY AND NEUROSURGERY
LA English
DT Article
DE Clivus chordomas; Modified ATPA approach; Complication; Survival
ID ENDOSCOPIC ENDONASAL APPROACH; SKULL BASE; RADIATION-THERAPY;
   MANAGEMENT; SURVIVAL; PATTERNS; OUTCOMES; SURGERY; CANCER
AB Background: Skull base chordomas are challenging and have a high rate of recurrence.
   Methods: A modified anterior transpetrous approach (ATPA) was performed in 17 upper clivus chordomas, and clinical data were retrospectively studied.
   Results: All 17 cases were radically treated via the modified ATPA, and the total removal and subtotal removal rates were 23.5% and 76.5%, respectively. The primary complaints were headaches and visual disturbances. The short-term postoperative complications were diplopia (12 cases, 70.6%) and facial numbness (7 cases, 41.2%). With a mean follow up of 44.5 months, 5 cases (29.4%) presented with tumor recurrence, and most cases had relatively good outcomes except for 2 patients who died because of rapid recurrence. The long-term complications were facial numbness (35.3%).
   Conclusions: The upper skull base chordomas could be radically removed via the modified ATPA under selected conditions, with limited complications and improved outcomes. The radical surgery treatment strategy was recommended for skull base chordomas. However, the present series included limited cases; therefore, post-operative follow-up, long-term outcomes and a larger number of cases of clivus chordomas should be observed to evaluate the effectiveness of the modified ATPA approach. (C) 2014 Elsevier B.V. All rights reserved.
C1 [Wang, Ke; Wang, Liang; Tian, Kaibing; Xiao, Xinru; Wu, Zhen; Jia, Guijun; Zhang, Liwei; Zhang, Junting] Capital Med Univ, Beijing Tian Tan Hosp, Dept Neurosurg, Skull Base & Brainstem Tumor Div, Beijing 100050, Peoples R China.
   [Wang, Ke; Wang, Liang; Tian, Kaibing; Xiao, Xinru; Wu, Zhen; Jia, Guijun; Zhang, Liwei; Zhang, Junting] China Natl Clin Res Ctr Neurol Dis, NCRC ND, Beijing 100050, Peoples R China.
C3 Capital Medical University
RP Zhang, JT (通讯作者)，Capital Med Univ, Beijing Tian Tan Hosp, Dept Neurosurg, Skull Base & Brainstem Tumor Div, Tiantan Xili 6, Beijing 100050, Peoples R China.
EM;
  ;
  
RI jiang, lei/IWE-1124-2023; li, wei/IUQ-2973-2023; Wang,
   lili/IXD-9828-2023
FU National Natural Science Foundation of China [81101910]; Natural Science
   Foundation of Beijing, China [7142052]
FX This work was supported in part by the National Natural Science
   Foundation of China (Grant No. 81101910), Natural Science Foundation of
   Beijing, China (Grant No. 7142052).
CR Alonso-Basanta M, 2011, OTOLARYNG CLIN N AM, V44, P1173, DOI 10.1016/j.otc.2011.08.012
   Amichetti M, 2009, NEUROSURG REV, V32, P403, DOI 10.1007/s10143-009-0194-4
   [Anonymous], 2014, NEUROSURGERY
   [Anonymous], 2014, NEUROSURGERY
   BOCHENEK Z, 1975, ACTA OTO-LARYNGOL, V80, P410, DOI 10.3109/00016487509121344
   Carrabba G, 2008, NEUROSURG FOCUS, V25, DOI 10.3171/FOC.2008.25.12.E7
   Chambers KJ, 2014, LARYNGOSCOPE, V124, P1097, DOI 10.1002/lary.24420
   Dassoulas K, 2009, J NEURO-ONCOL, V94, P243, DOI 10.1007/s11060-009-9846-z
   Dehdashti AR, 2008, NEUROSURGERY, V63, P299, DOI 10.1227/01.NEU.0000316414.20247.32
   Di Maio S, 2012, NEUROSURGERY, V70, P1355, DOI 10.1227/NEU.0b013e3182446783
   Di Maio S, 2011, J NEUROSURG, V115, P1094, DOI 10.3171/2011.7.JNS11355
   Diaz RJ, 2014, J NEUROSURG, V120, P528, DOI 10.3171/2013.9.JNS13671
   Gao ZY, 2012, MED ONCOL, V29, P2438, DOI 10.1007/s12032-012-0182-3
   Hung GY, 2014, ANN SURG ONCOL, V21, P2490, DOI 10.1245/s10434-014-3697-3
   Ito E, 2010, ACTA NEUROCHIR, V152, P57, DOI 10.1007/s00701-009-0535-7
   Jia GJ, 2010, NEUROSURG REV, V33, P121, DOI 10.1007/s10143-009-0221-5
   Jian BJ, 2011, BRIT J NEUROSURG, V25, P446, DOI 10.3109/02688697.2010.546896
   Jian BJ, 2010, J NEURO-ONCOL, V98, P101, DOI 10.1007/s11060-009-0068-1
   Koutourousiou M, 2012, NEUROSURGERY, V71, P614, DOI 10.1227/NEU.0b013e31825ea3e0
   Lee J, 2012, J BONE JOINT SURG AM, V94A, P356, DOI 10.2106/JBJS.J.01784
   Liu AL, 2008, NEUROL RES, V30, P557, DOI 10.1179/174313208X297878
   Nikoghosyan AV, 2010, BMC CANCER, V10, DOI 10.1186/1471-2407-10-607
   Sen C, 2010, J NEUROSURG, V113, P1059, DOI 10.3171/2009.9.JNS08596
   Tabaee A, 2010, OTOLARYNG HEAD NECK, V142, P126, DOI 10.1016/j.otohns.2009.09.021
   Wang L, 2014, WORLD NEUROSURG, V82, P791, DOI 10.1016/j.wneu.2012.12.037
   Wu Z, 2010, NEUROSURG REV, V33, P451, DOI 10.1007/s10143-010-0273-6
   Xiao XR, 2013, NEUROSURG REV, V36, P587, DOI 10.1007/s10143-013-0484-8
   Yoneoka Y, 2008, ACTA NEUROCHIR, V150, P773, DOI 10.1007/s00701-008-1600-3
NR 28
TC 9
Z9 9
U1 0
U2 4
PU ELSEVIER SCIENCE BV
PI AMSTERDAM
PA PO BOX 211, 1000 AE AMSTERDAM, NETHERLANDS
SN 0303-8467
EI 1872-6968
J9 CLIN NEUROL NEUROSUR
JI Clin. Neurol. Neurosurg.
PD MAR
PY 2015
VL 130
BP 20
EP 25
DI 10.1016/j.clineuro.2014.12.016
PG 6
WC Clinical Neurology; Surgery
WE Science Citation Index Expanded (SCI-EXPANDED)
SC Neurosciences & Neurology; Surgery
GA CC2PD
UT WOS:000350186200006
PM 25576881
DA 2023-08-27
ER

PT J
AU Moffat, DA
   Lloyd, SKW
   Macfarlane, R
   Mannion, R
   King, A
   Rutherford, S
   Axon, PR
   Donnelly, N
   Freeman, S
   Tysome, JR
   Evans, DG
   Ramsden, RT
AF Moffat, D. A.
   Lloyd, S. K. W.
   Macfarlane, R.
   Mannion, R.
   King, A.
   Rutherford, S.
   Axon, P. R.
   Donnelly, N.
   Freeman, S.
   Tysome, J. R.
   Evans, D. G.
   Ramsden, R. T.
TI Outcome of translabyrinthine surgery for vestibular schwannoma in
   neurofibromatosis type 2
SO BRITISH JOURNAL OF NEUROSURGERY
LA English
DT Article
DE Neurofibromatosis type 2; outcome; translabyrinthine surgery; vestibular
   schwannoma
ID FACIAL-NERVE FUNCTION; ACOUSTIC NEUROMAS; MANAGEMENT; REMOVAL
AB Objectives. To analyse the long-term outcome of translabyrinthine surgery for vestibular schwannoma (VS) in neurofibromatosis type 2 (NF2). Research type. Retrospective cohort study. Setting. Two tertiary referral NF2 units. Patients. One hundred and forty eight translabyrinthine operations for patients with VS were performed. Preoperative stereotactic radiotherapy had been performed on 12(9.4%) patients. Results. Mean tumour size was 3.1 cm. Total tumour excision was achieved in 66% of cases, capsular remnants were left in 24% of cases, and subtotal excision was achieved in 5% and partial removal was achieved in 5%. The radiological residual/recurrence rate was 13.9%. The perioperative mortality was 1.6%. At 2 years postoperatively, facial function was expressed in terms of House-Brackmann score (HB): HB 1 in 53.4%, HB 1/2 in 61.3%, HB 1-3 in 83.2% and HB 4-6 in 16.8%. All nine patients who underwent surgery following failed stereotactic radiotherapy had HB 3 function or better. Among 9.5% of the cases, 14 facial nerves were lost during surgery and repaired using direct anastomosis or grafting. There was no tinnitus present preoperatively in 27% of the cases, and 22% of patients developed tinnitus postoperatively. In patients with preoperative tinnitus, 61% remained the same, 17% got it resolved and only in 21% it worsened. The preoperative hydrocephalus rate was 26%, and among 15% of the cases five ventriculo-peritoneal (VP) shunts were performed. The cerebrospinal fluid leak rate was 2.5%. Fifty-six patients underwent auditory brainstem implantation (ABI) and two patients had cochlear implant (CI) sleepers inserted. Conclusions. The management of patients with NF2 presents the clinician with a formidable challenge with many patients still presenting themselves late with the neurological compromise and a large tumour load. There is still an argument for the management by observation until the neurological compromise dictates interventional treatment particularly with the option of hearing rehabilitation with ABI or CI. The translabyrinthine approach provides a very satisfactory means of reducing the overall tumour volume.
C1 [Moffat, D. A.; Macfarlane, R.; Mannion, R.; Axon, P. R.; Donnelly, N.; Tysome, J. R.] Cambridge Univ Teaching Hosp NHS Trust, Addenbrookes Hosp, Dept Skull Base Surg, Cambridge CB2 0QQ, England.
   [Lloyd, S. K. W.; King, A.; Rutherford, S.; Freeman, S.; Ramsden, R. T.] Manchester Royal Infirm, Dept Skull Base Surg, ENT, Manchester M13 9WL, Lancs, England.
   [Evans, D. G.] Manchester Royal Infirm, Dept Med Genet, Manchester M13 9WL, Lancs, England.
C3 Cambridge University Hospitals NHS Foundation Trust; Addenbrooke's
   Hospital; University of Cambridge; University of Manchester; University
   of Manchester
RP Moffat, DA (通讯作者)，Cambridge Univ Teaching Hosp NHS Trust, Addenbrookes Hosp, Dept Neurootol & Skull Base Surg, Cambridge CB2 0QQ, England.
EM
RI Rutherford, Scott/GMX-2404-2022; Evans, D Gareth/AAB-4308-2022; King,
   Andrew/AAR-1656-2020
OI Rutherford, Scott/0000-0002-6749-2295; Evans, D
   Gareth/0000-0002-8482-5784; 
CR Asthagiri AR, 2009, LANCET, V373, P1974, DOI 10.1016/S0140-6736(09)60259-2
   Baser M. E., 2002, Neurology, V59, P1759
   BRACKMANN DE, 1992, OTOLARYNG CLIN N AM, V25, P311
   Brackmann DE, 2001, NEUROSURGERY, V49, P274, DOI 10.1097/00006123-200108000-00007
   Brackmann DE, 2007, OTOLARYNG HEAD NECK, V136, P773, DOI 10.1016/j.otohns.2006.10.009
   Evans DGR, 2006, J MED GENET, V43, P289, DOI 10.1136/jmg.2005.036319
   EVANS DGR, 1993, J LARYNGOL OTOL, V107, P401, DOI 10.1017/S002221510012328X
   EVANS DGR, 1992, Q J MED, V84, P603
   Evans DGR, 2005, BRIT J NEUROSURG, V19, P5, DOI 10.1080/02688690500081206
   Fisher LM, 2009, OTOL NEUROTOL, V30, P835, DOI 10.1097/MAO.0b013e3181b2364c
   HARDY DG, 1989, J NEUROSURG, V71, P799, DOI 10.3171/jns.1989.71.6.0799
   House WF., 1979, ACOUSTIC TUMORS MANA, V2, P43
   LYE RH, 1992, BRIT J NEUROSURG, V6, P195, DOI 10.3109/02688699209002927
   Moffat DA, 2003, EUR ARCH OTO-RHINO-L, V260, P12, DOI 10.1007/s00405-002-0503-9
   MOFFAT DA, 1989, J LARYNGOL OTOL, V103, P169, DOI 10.1017/S0022215100108357
   Neary WJ, 2010, J LARYNGOL OTOL, V124, P720, DOI 10.1017/S0022215110000460
   Pollock Bruce E, 2008, Prog Neurol Surg, V21, P163, DOI 10.1159/000156904
   ROULEAU GA, 1993, NATURE, V363, P515, DOI 10.1038/363515a0
   Rowe Jeremy, 2008, Prog Neurol Surg, V21, P176, DOI 10.1159/000156907
   Samii M, 1997, NEUROSURGERY, V40, P696, DOI 10.1097/00006123-199704000-00007
   Samii M, 2008, Prog Neurol Surg, V21, P169, DOI 10.1159/000156905
   Tos M, ACOUSTIC NEUROMA C 1, P3
   Woodson EA, 2010, OTOL NEUROTOL, V31, P1144, DOI 10.1097/MAO.0b013e3181edb8b2
NR 23
TC 21
Z9 21
U1 0
U2 3
PU INFORMA HEALTHCARE
PI LONDON
PA TELEPHONE HOUSE, 69-77 PAUL STREET, LONDON EC2A 4LQ, ENGLAND
SN 0268-8697
J9 BRIT J NEUROSURG
JI Br. J. Neurosurg.
PD AUG
PY 2013
VL 27
IS 4
BP 446
EP 453
DI 10.3109/02688697.2013.771143
PG 8
WC Clinical Neurology; Surgery
WE Science Citation Index Expanded (SCI-EXPANDED)
SC Neurosciences & Neurology; Surgery
GA 190DP
UT WOS:000322319000010
PM 23472624
DA 2023-08-27
ER

PT J
AU Reisinger, SN
   Sideromenos, S
   Horvath, O
   Derdak, S
   Cicvaric, A
   Monje, FJ
   Bilban, M
   Haring, M
   Glat, M
   Pollak, DD
AF Reisinger, Sonali N.
   Sideromenos, Spyros
   Horvath, Orsolya
   Derdak, Sophia
   Cicvaric, Ana
   Monje, Francisco J.
   Bilban, Martin
   Haering, Martin
   Glat, Micaela
   Pollak, Daniela D.
TI STAT3 in the dorsal raphe gates behavioural reactivity and regulates
   gene networks associated with psychopathology
SO MOLECULAR PSYCHIATRY
LA English
DT Article
ID DEPRESSION-LIKE BEHAVIORS; TUMOR-SUPPRESSOR GENE; SIGNAL TRANSDUCERS;
   SEROTONIN; STRESS; ACTIVATION; PATHWAY; TRANSCRIPTION; AMPHETAMINE;
   SURVIVAL
AB The signal transducer and activator of transcription 3 (STAT3) signalling pathway is activated through phosphorylation by Janus kinases in response to a diverse set of immunogenic and non-immunogenic triggers. Several distinct lines of evidence propose an intricate involvement of STAT3 in neural function relevant to behaviour in health and disease. However, in part due to the pleiotropic effects resulting from its DNA binding activity and the consequent regulation of expression of a variety of genes with context-dependent cellular consequences, the precise nature of STAT3 involvement in the neural mechanisms underlying psychopathology remains incompletely understood. Here, we focused on the midbrain serotonergic system, a central hub for the regulation of emotions, to examine the relevance of STAT3 signalling for emotional behaviour in mice by selectively knocking down raphe STAT3 expression using germline genetic (STAT3 KO) and viral-mediated approaches. Mice lacking serotonergic STAT3 presented with reduced negative behavioural reactivity and a blunted response to the sensitising effects of amphetamine, alongside alterations in midbrain neuronal firing activity of serotonergic neurons and transcriptional control of gene networks relevant for neuropsychiatric disorders. Viral knockdown of dorsal raphe (DR) STAT3 phenocopied the behavioural alterations of STAT3 KO mice, excluding a developmentally determined effect and suggesting that disruption of STAT3 signalling in the DR of adult mice is sufficient for the manifestation of behavioural traits relevant to psychopathology. Collectively, these results suggest DR STAT3 as a molecular gate for the control of behavioural reactivity, constituting a mechanistic link between the upstream activators of STAT3, serotonergic neurotransmission and psychopathology.
C1 [Reisinger, Sonali N.; Sideromenos, Spyros; Horvath, Orsolya; Cicvaric, Ana; Monje, Francisco J.; Glat, Micaela; Pollak, Daniela D.] Med Univ Vienna, Ctr Physiol & Pharmacol, Dept Neurophysiol & Neuropharmacol, Schwarzspanierstr 17, A-1090 Vienna, Austria.
   [Derdak, Sophia] Med Univ Vienna, Core Facil Genom, Lazarettgasse 14, A-1090 Vienna, Austria.
   [Bilban, Martin] Med Univ Vienna, Dept Lab Med, Wahringer Gurtel 18-20, A-1090 Vienna, Austria.
   [Haering, Martin] Med Univ Vienna, Ctr Brain Res, Dept Mol Neurosci, Spitalgasse 4, A-1090 Vienna, Austria.
C3 Medical University of Vienna; Medical University of Vienna; Medical
   University of Vienna; Medical University of Vienna
RP Pollak, DD (通讯作者)，Med Univ Vienna, Ctr Physiol & Pharmacol, Dept Neurophysiol & Neuropharmacol, Schwarzspanierstr 17, A-1090 Vienna, Austria.
EM
OI Sideromenos, Spyros/0000-0002-5135-3346; Cicvaric,
   Ana/0000-0002-3560-4491; Reisinger, Sonali/0000-0002-9242-6249; POLLAK,
   Daniela/0000-0002-9584-6257
FU Austrian Science Fund (Fonds fur Wissenschaft und Forschung, FWF)
   [P28683]; Austrian Science Fund (FWF) [P28683] Funding Source: Austrian
   Science Fund (FWF)
FX This project was supported by a stand-alone grant of the Austrian
   Science Fund (Fonds fur Wissenschaft und Forschung, FWF; grant no.
   P28683) to DDP.
CR Ahmad SF, 2019, BRAIN RES BULL, V152, P27, DOI 10.1016/j.brainresbull.2019.07.006
   Ahmad SF, 2018, CELL SIGNAL, V52, P127, DOI 10.1016/j.cellsig.2018.09.006
   [Anonymous], 2012, Wkly Epidemiol Rec, V87, P1
   ANTELMAN SM, 1980, SCIENCE, V207, P329, DOI 10.1126/science.7188649
   Athos J, 2001, CURR PROTOC NEUROSCI, V14
   Bankhead P, ANAL FLUORESCENCE MI
   Bjornstrom L, 2002, MOL ENDOCRINOL, V16, P2202, DOI 10.1210/me.2002-0072
   Booij L, 2015, J PSYCHIATR NEUROSCI, V40, P5, DOI 10.1503/jpn.140099
   Bourin M, 2003, EUR J PHARMACOL, V463, P55, DOI 10.1016/S0014-2999(03)01274-3
   Brietzke E, 2009, J AFFECT DISORDERS, V116, P214, DOI 10.1016/j.jad.2008.12.001
   Brummelte S, 2017, NEUROSCIENCE, V342, P212, DOI 10.1016/j.neuroscience.2016.02.037
   Cargnello M, 2011, MICROBIOL MOL BIOL R, V75, P50, DOI 10.1128/MMBR.00031-10
   Chen EY, 2013, BMC BIOINFORMATICS, V14, DOI 10.1186/1471-2105-14-128
   Ciaglia E, 2015, ONCOTARGET, V6, P15464, DOI 10.18632/oncotarget.3895
   de Boer SF, 2017, NEUROSCI BIOBEHAV R, V74, P401, DOI 10.1016/j.neubiorev.2016.07.008
   de Kloet ER, 2016, NEURAL PLAST, V2016, DOI 10.1155/2016/6503162
   Der-Avakian A, 2012, TRENDS NEUROSCI, V35, P68, DOI 10.1016/j.tins.2011.11.005
   Enache D, 2019, BRAIN BEHAV IMMUN, V81, P24, DOI 10.1016/j.bbi.2019.06.015
   Febo M, 2011, BRAIN RES, V1382, P118, DOI 10.1016/j.brainres.2010.11.092
   Festing MFW, 2018, LAB ANIM-UK, V52, P341, DOI 10.1177/0023677217738268
   Firk C, 2007, J PSYCHOPHARMACOL, V21, P538, DOI 10.1177/0269881106075588
   Guo ZH, 2008, J BIOL CHEM, V283, P1754, DOI 10.1074/jbc.M703753200
   HAMAMURA T, 1993, EUR J PHARMACOL, V237, P65, DOI 10.1016/0014-2999(93)90094-X
   He JCJ, 2005, J BIOL CHEM, V280, P33426, DOI 10.1074/jbc.M502812200
   Hixson KM, 2019, BMC GENOMICS, V20, DOI 10.1186/s12864-019-6033-2
   Ho MF, 2019, FRONT PHARMACOL, V10, DOI 10.3389/fphar.2019.01302
   Homberg JR, 2016, NEUROSCI BIOBEHAV R, V71, P472, DOI 10.1016/j.neubiorev.2016.09.029
   Insel T, 2010, AM J PSYCHIAT, V167, P748, DOI 10.1176/appi.ajp.2010.09091379
   Jans LAW, 2007, MOL PSYCHIATR, V12, P522, DOI 10.1038/sj.mp.4001920
   KALIVAS PW, 1986, BIOL PSYCHIAT, V21, P939, DOI 10.1016/0006-3223(86)90268-4
   Khandaker GM, 2014, JAMA PSYCHIAT, V71, P1121, DOI 10.1001/jamapsychiatry.2014.1332
   Kong E, 2015, SCI REP-UK, V5, DOI 10.1038/srep09009
   Kuleshov MV, 2016, NUCLEIC ACIDS RES, V44, pW90, DOI 10.1093/nar/gkw377
   Kwon SH, 2017, NEUROPSYCHOPHARMACOL, V42, P2072, DOI 10.1038/npp.2017.93
   Lee H, 2012, P NATL ACAD SCI USA, V109, P7765, DOI 10.1073/pnas.1205132109
   Li J, 2013, ONCOL REP, V30, P165, DOI 10.3892/or.2013.2414
   Liao L, 2017, INT IMMUNOPHARMACOL, V49, P178, DOI 10.1016/j.intimp.2017.05.036
   Liu WN, 2017, BRAIN BEHAV IMMUN, V61, P297, DOI 10.1016/j.bbi.2017.01.001
   Lou DN, 2019, MOL CELL PROBE, V48, DOI 10.1016/j.mcp.2019.101470
   Love MI, 2014, GENOME BIOL, V15, DOI 10.1186/s13059-014-0550-8
   Mingmalairak S, 2010, BIOL PHARM BULL, V33, P636, DOI 10.1248/bpb.33.636
   Moh A, 2007, LAB INVEST, V87, P1018, DOI 10.1038/labinvest.3700630
   Molendijk ML, 2015, PSYCHONEUROENDOCRINO, V62, P389, DOI 10.1016/j.psyneuen.2015.08.028
   Mosienko V, 2012, TRANSL PSYCHIAT, V2, DOI 10.1038/tp.2012.44
   Ng YP, 2006, J BIOL CHEM, V281, P15636, DOI 10.1074/jbc.M601863200
   Nicolas Celine S, 2013, JAKSTAT, V2, pe22925, DOI 10.4161/jkst.22925
   Nicolas CS, 2012, NEURON, V73, P374, DOI 10.1016/j.neuron.2011.11.024
   Oki S, 2018, EMBO REP, V19, DOI 10.15252/embr.201846255
   Patton MS, 2017, NEUROPSYCHOPHARMACOL, V42, P1220, DOI 10.1038/npp.2016.236
   Paxinos G., 2001, MOUSE BRAIN STEREOTA
   Pillinger T, 2019, SCHIZOPHRENIA BULL, V45, P1120, DOI 10.1093/schbul/sby160
   Pinero J, 2020, NUCLEIC ACIDS RES, V48, pD845, DOI 10.1093/nar/gkz1021
   Potvin S, 2008, BIOL PSYCHIAT, V63, P801, DOI 10.1016/j.biopsych.2007.09.024
   Prus AJ, 2009, FRONT NEUROSCI, P59
   Puglisi-Allegra S, 2015, BEHAV BRAIN RES, V277, P58, DOI 10.1016/j.bbr.2014.07.052
   REGIER DA, 1990, JAMA-J AM MED ASSOC, V264, P2511
   Reisinger SN, 2019, GENES BRAIN BEHAV, V18, DOI 10.1111/gbb.12482
   Reisinger SN, 2020, PSYCHONEUROENDOCRINO, V111, DOI 10.1016/j.psyneuen.2019.104480
   Reisinger SN, 2019, TRANSL PSYCHIAT, V9, DOI 10.1038/s41398-019-0416-7
   Ross Christopher A, 2019, Mol Neuropsychiatry, V5, P218, DOI 10.1159/000501797
   Samuels BA., 2011, MOOD ANXIETY RELATED, V2, P107, DOI [DOI 10.1007/978-1-61779-313-4_7, 10.1007/978-1-61779-313-4_7]
   Schmittgen TD, 2008, NAT PROTOC, V3, P1101, DOI 10.1038/nprot.2008.73
   Schneider CA, 2012, NAT METHODS, V9, P671, DOI 10.1038/nmeth.2089
   Sharma N, 2019, CURR NEUROPHARMACOL, V17, P990, DOI 10.2174/1570159X17666190228114318
   Steinkellner T, 2015, J NEUROSCI, V35, P8258, DOI 10.1523/JNEUROSCI.4034-14.2015
   Subbanna M, 2020, PSYCHIAT CLIN NEUROS, V74, P64, DOI 10.1111/pcn.12938
   Subbanna M, 2018, CYTOKINE, V111, P112, DOI 10.1016/j.cyto.2018.08.016
   Tran THN, 2016, NEUROCHEM INT, V94, P9, DOI 10.1016/j.neuint.2016.02.001
   Timofeeva OA, 2013, P NATL ACAD SCI USA, V110, P1267, DOI 10.1073/pnas.1211805110
   Tyzack GE, 2014, NAT COMMUN, V5, DOI 10.1038/ncomms5294
   Vaisse C, 1996, NAT GENET, V14, P95, DOI 10.1038/ng0996-95
   Vogel TP, 2015, J CLIN IMMUNOL, V35, P615, DOI 10.1007/s10875-015-0187-8
   Walf AA, 2007, NAT PROTOC, V2, P322, DOI 10.1038/nprot.2007.44
   Wylie CJ, 2010, J NEUROSCI, V30, P670, DOI 10.1523/JNEUROSCI.4656-09.2010
   Yadav A, 2005, J BIOL CHEM, V280, P31830, DOI 10.1074/jbc.M501316200
   Zhang ZX, 1997, J BIOL CHEM, V272, P30607, DOI 10.1074/jbc.272.49.30607
   Zhou H, 2013, BRAIN RES, V1529, P154, DOI 10.1016/j.brainres.2013.07.006
   Zhuang XX, 2005, J NEUROSCI METH, V143, P27, DOI 10.1016/j.jneumeth.2004.09.020
   Zorina Y, 2010, J BIOL CHEM, V285, P1358, DOI 10.1074/jbc.M109.049841
NR 79
TC 6
Z9 6
U1 3
U2 8
PU SPRINGERNATURE
PI LONDON
PA CAMPUS, 4 CRINAN ST, LONDON, N1 9XW, ENGLAND
SN 1359-4184
EI 1476-5578
J9 MOL PSYCHIATR
JI Mol. Psychiatr.
PD JUL
PY 2021
VL 26
IS 7
BP 2886
EP 2899
DI 10.1038/s41380-020-00904-2
EA OCT 2020
PG 14
WC Biochemistry & Molecular Biology; Neurosciences; Psychiatry
WE Science Citation Index Expanded (SCI-EXPANDED)
SC Biochemistry & Molecular Biology; Neurosciences & Neurology; Psychiatry
GA WF6IT
UT WOS:000578434700001
PM 33046834
OA Green Published, hybrid
DA 2023-08-27
ER

PT J
AU Kuper, C
   Bartels, H
   Fraek, ML
   Beck, FX
   Neuhofer, W
AF Kuper, Christoph
   Bartels, Helmut
   Fraek, Maria-Luisa
   Beck, Franz X.
   Neuhofer, Wolfgang
TI Ectodomain shedding of pro-TGF-alpha is required for COX-2 induction and
   cell survival in renal medullary cells exposed to osmotic stress
SO AMERICAN JOURNAL OF PHYSIOLOGY-CELL PHYSIOLOGY
LA English
DT Article
DE pro-epidermal growth factor receptor ligand; tumor necrosis factor-alpha
   converting enzyme; epidermal growth factor receptor; prostaglandin E-2
ID GROWTH-FACTOR RECEPTOR; COLLECTING DUCT CELLS; CYTOSOLIC PHOSPHOLIPASE
   A(2); NECROSIS-FACTOR-ALPHA; THICK ASCENDING LIMB; CYCLOOXYGENASE-2
   EXPRESSION; HYPERTONIC INDUCTION; NITRIC-OXIDE; IN-VIVO; TRANSACTIVATION
AB In the renal medulla, cyclooxygenase (COX)-2 is induced by osmotic stress as present in this kidney region during antidiuresis. Increasing evidence suggests that EGF receptor ( EGFR) signaling is involved in this process. The aim of the present study was to examine the mechanisms responsible for COX-2 expression and PGE(2) production during hypertonic conditions and to identify potential autocrine/paracrine EGFR ligands. Immunohistochemisty and Western blot analysis revealed abundant expression of the pro-EGFR ligand pro-transforming growth factor (TGF)-alpha in renal medullary cells in vivo and in cultured Madin-Darby canine kidney cells. In Madin-Darby canine kidney cells, hypertonicity rapidly increased TNF-alpha converting enzyme (TACE)-dependent ectodomain shedding of pro-TGF-alpha; phosphorylation of EGFR, p38, and ERK1/2; expression of COX-2; and production of PGE2. Conversely, TACE inhibition prevented TGF-alpha release; EGFR, p38, and ERK1/2 activation; and COX-2 expression. Furthermore, cell survival was reduced substantially, a response that could be reversed by the addition of PGE2. Simultaneous addition of recombinant TGF-alpha during TACE inhibition restored EGFR and MAPK phosphorylation, COX-2 expression, PGE2 production, and cell survival during osmotic stress. These results indicate that hypertonicity induces TACE-mediated ectodomain shedding of pro-TGF-alpha, which subsequently activates COX-2 expression in an autocrine/paracrine fashion, via EGFR and MAPKs. We conclude that tonicity-induced TGF-alpha release is required for COX-2 expression, PGE2 synthesis, and survival of renal medullary cells during osmotic stress.
C1 [Kuper, Christoph; Fraek, Maria-Luisa; Beck, Franz X.; Neuhofer, Wolfgang] Univ Munich, Dept Physiol, D-80336 Munich, Germany.
   [Bartels, Helmut] Univ Munich, Dept Anat, Munich, Germany.
C3 University of Munich; University of Munich
RP Neuhofer, W (通讯作者)，Univ Munich, Dept Physiol, Pettenkoferstr 12, D-80336 Munich, Germany.
EM
CR ATHERTON JC, 1971, J PHYSIOL-LONDON, V215, pP42
   Black RA, 1997, NATURE, V385, P729, DOI 10.1038/385729a0
   Buckman SY, 1998, CARCINOGENESIS, V19, P723, DOI 10.1093/carcin/19.5.723
   Carpenter G, 1999, J CELL BIOL, V146, P697, DOI 10.1083/jcb.146.4.697
   Cheng H, 2002, J CELL PHYSIOL, V192, P234, DOI 10.1002/jcp.10134
   Cowley BD, 1995, AM J PHYSIOL-RENAL, V269, pF854, DOI 10.1152/ajprenal.1995.269.6.F854
   CROFFORD LJ, 1994, J CLIN INVEST, V93, P1095, DOI 10.1172/JCI117060
   CROWE PD, 1995, J EXP MED, V181, P1205, DOI 10.1084/jem.181.3.1205
   Ferreri NR, 1999, AM J PHYSIOL-RENAL, V277, pF360, DOI 10.1152/ajprenal.1999.277.3.F360
   Gschwind A, 2001, ONCOGENE, V20, P1594, DOI 10.1038/sj.onc.1204192
   Hazan-Halevy I, 2000, J BIOL CHEM, V275, P12416, DOI 10.1074/jbc.275.17.12416
   Honda S, 2000, BIOCHEM BIOPH RES CO, V268, P928, DOI 10.1006/bbrc.2000.2228
   JABS K, 1989, AM J PHYSIOL, V257, pF424, DOI 10.1152/ajprenal.1989.257.3.F424
   Kaji DM, 1996, AM J PHYSIOL-CELL PH, V271, pC354, DOI 10.1152/ajpcell.1996.271.1.C354
   Knebel A, 1996, EMBO J, V15, P5314, DOI 10.1002/j.1460-2075.1996.tb00916.x
   LEE DC, 1995, PHARMACOL REV, V47, P51
   Leslie CC, 1997, J BIOL CHEM, V272, P16709, DOI 10.1074/jbc.272.27.16709
   MASSAGUE J, 1993, ANNU REV BIOCHEM, V62, P515, DOI 10.1146/annurev.bi.62.070193.002503
   Mori T, 2004, HYPERTENSION, V43, P341, DOI 10.1161/01.HYP.0000113295.31481.36
   *NAT CTR BIOT INF, NAT I HLTH ENTR PROT
   Navar LG, 1996, PHYSIOL REV, V76, P425, DOI 10.1152/physrev.1996.76.2.425
   Neuhofer W, 2005, ANNU REV PHYSIOL, V67, P531, DOI 10.1146/annurev.physiol.67.031103.154456
   Neuhofer W, 2002, J PHYSIOL-LONDON, V543, P147, DOI 10.1113/jphysiol.2002.021931
   Neuhofer W, 2007, J PHYSIOL-LONDON, V583, P287, DOI 10.1113/jphysiol.2007.135178
   REYNOLDS LJ, 1992, ANAL BIOCHEM, V204, P190, DOI 10.1016/0003-2697(92)90160-9
   Riese DJ, 1998, BIOESSAYS, V20, P41, DOI 10.1002/(SICI)1521-1878(199801)20:1<41::AID-BIES7>3.0.CO;2-V
   Rodriguez I, 2002, CELL SIGNAL, V14, P839, DOI 10.1016/S0898-6568(02)00031-1
   SCHLONDORFF D, 1993, KIDNEY INT, V44, P643, DOI 10.1038/ki.1993.293
   Sunnarborg SW, 2002, J BIOL CHEM, V277, P12838, DOI 10.1074/jbc.M112050200
   ULLRICH A, 1990, CELL, V61, P203, DOI 10.1016/0092-8674(90)90801-K
   Woo SK, 2002, PFLUG ARCH EUR J PHY, V444, P579, DOI 10.1007/s00424-002-0849-2
   Yang TX, 2006, AM J PHYSIOL-RENAL, V291, pF891, DOI 10.1152/ajprenal.00512.2005
   Yang TX, 2000, J BIOL CHEM, V275, P23281, DOI 10.1074/jbc.M910237199
   Yang TX, 2005, J BIOL CHEM, V280, P34966, DOI 10.1074/jbc.M502430200
   Yang TX, 1999, AM J PHYSIOL-RENAL, V277, pF1, DOI 10.1152/ajprenal.1999.277.1.F1
   ZHANG F, 1995, BIOCHEM J, V312, P135, DOI 10.1042/bj3120135
   Zhang Z, 2004, P NATL ACAD SCI USA, V101, P9491, DOI 10.1073/pnas.0402961101
   Zhang ZL, 2001, FASEB J, V15, P303
   Zhao HY, 2003, AM J PHYSIOL-RENAL, V285, pF281, DOI 10.1152/ajprenal.00030.2003
NR 39
TC 21
Z9 21
U1 0
U2 0
PU AMER PHYSIOLOGICAL SOC
PI BETHESDA
PA 9650 ROCKVILLE PIKE, BETHESDA, MD 20814 USA
SN 0363-6143
J9 AM J PHYSIOL-CELL PH
JI Am. J. Physiol.-Cell Physiol.
PD DEC
PY 2007
VL 293
IS 6
BP C1971
EP C1982
DI 10.1152/ajpcell.00404.2007
PG 12
WC Cell Biology; Physiology
WE Science Citation Index Expanded (SCI-EXPANDED)
SC Cell Biology; Physiology
GA 240BO
UT WOS:000251562200027
PM 17942633
DA 2023-08-27
ER

PT J
AU Tsai, VWW
   Lin, S
   Brown, DA
   Salis, A
   Breit, SN
AF Tsai, V. W. W.
   Lin, S.
   Brown, D. A.
   Salis, A.
   Breit, S. N.
TI Anorexia-cachexia and obesity treatment may be two sides of the same
   coin: role of the TGF-b superfamily cytokine MIC-1/GDF15
SO INTERNATIONAL JOURNAL OF OBESITY
LA English
DT Review
ID MACROPHAGE-INHIBITORY CYTOKINE-1; GROWTH-FACTOR-BETA; DIFFERENTIATION
   FACTOR-15; SERUM CONCENTRATIONS; NUTRITIONAL-STATUS; BODY-COMPOSITION;
   WEIGHT-LOSS; ALL-CAUSE; MIC-1; EXPRESSION
AB Anorexia-cachexia associated with cancer and other diseases is a common and often fatal condition representing a large area of unmet medical need. It occurs most commonly in advanced cancer and is probably a consequence of molecules released by tumour cells, or tumour-associated interstitial or immune cells. These may then act directly on muscle to cause atrophy and/or may cause anorexia, which then leads to loss of both fat and lean mass. Although the aetiological triggers for this syndrome are not well characterized, recent data suggest that MIC-1/GDF15, a transforming growth factor-beta superfamily cytokine produced in large amounts by cancer cells and as a part of other disease processes, may be an important trigger. This cytokine acts on feeding centres in the hypothalamus and brainstem to cause anorexia leading to loss of lean and fat mass and eventually cachexia. In animal studies, the circulating concentrations of MIC-1/GDF15 required to cause this syndrome are similar to those seen in patients with advanced cancer, and at least some epidemiological studies support an association between MIC-1/GDF15 serum levels and measures of nutrition. This article will discuss its mechanisms of central appetite regulation, and the available data linking this action to anorexia-cachexia syndromes that suggest it is a potential target for therapy of cancer anorexia-cachexia and conversely may also be useful for the treatment of severe obesity.
C1 [Tsai, V. W. W.; Brown, D. A.; Breit, S. N.] St Vincents Hosp, St Vincents Ctr Appl Med Res, Sydney, NSW 2010, Australia.
   [Tsai, V. W. W.; Brown, D. A.; Breit, S. N.] Univ New S Wales, Fac Med, Sydney, NSW, Australia.
   [Lin, S.] Garvan Inst Med Res, Sydney, NSW, Australia.
   [Salis, A.] Univ Sydney, Boden Inst Obes Nutr Exercise & Eating Disorders, Sydney, NSW 2006, Australia.
C3 St Vincents Hospital Sydney; University of New South Wales Sydney;
   Garvan Institute of Medical Research; University of Sydney
RP Breit, SN (通讯作者)，St Vincents Hosp, St Vincents Ctr Appl Med Res, Sydney, NSW 2010, Australia.
EM
RI Salis, Amanda/A-3187-2011; Breit, Samuel N/B-6177-2008; Lin,
   Shu/A-4361-2010
OI Salis, Amanda/0000-0001-9176-1574; lin, shu/0000-0002-4239-2028; Brown,
   David/0000-0003-1382-9419
FU National Health and Medical Research Council of Australia (NHMRC);
   Cancer Council NSW; NHMRC
FX This work was supported by grants from the National Health and Medical
   Research Council of Australia (NHMRC) and Cancer Council NSW. AS, DAB
   and SL are recipients of NHMRC Career Development Awards or Fellowships.
CR Argiles JM, 2014, NAT REV CANCER, V14, P754, DOI 10.1038/nrc3829
   Baek SJ, 2006, GASTROENTEROLOGY, V131, P1553, DOI 10.1053/j.gastro.2006.09.015
   Baek SJ, 2004, J BIOL CHEM, V279, P6883, DOI 10.1074/jbc.M305295200
   Baek SJ, 2001, MOL PHARMACOL, V59, P901, DOI 10.1124/mol.59.4.901
   Banerjee A, 2012, CURR OPIN SUPPORT PA, V6, P451, DOI 10.1097/SPC.0b013e328359b681
   BARACNIETO M, 1978, AM J CLIN NUTR, V31, P23, DOI 10.1093/ajcn/31.1.23
   Bauskin AR, 2005, CANCER RES, V65, P2330, DOI 10.1158/0008-5472.CAN-04-3827
   Bauskin AR, 2006, CANCER RES, V66, P4983, DOI 10.1158/0008-5472.CAN-05-4067
   Bauskin AR, 2010, J INTERF CYTOK RES, V30, P389, DOI 10.1089/jir.2009.0052
   Blum D, 2011, CRIT REV ONCOL HEMAT, V80, P114, DOI 10.1016/j.critrevonc.2010.10.004
   Bootcov MR, 1997, P NATL ACAD SCI USA, V94, P11514, DOI 10.1073/pnas.94.21.11514
   Bottner M, 1999, GENE, V237, P105, DOI 10.1016/S0378-1119(99)00309-1
   Breit SN, 2012, NEPHROL DIAL TRANSPL, V27, P70, DOI 10.1093/ndt/gfr575
   Breit SN, 2011, GROWTH FACTORS, V29, P187, DOI 10.3109/08977194.2011.607137
   Brown DA, 2006, CLIN CANCER RES, V12, P89, DOI 10.1158/1078-0432.CCR-05-1331
   Brown DA, 2003, CLIN CANCER RES, V9, P2642
   Brown DA, 2002, BIOTECHNIQUES, V33, P118, DOI 10.2144/02331rr03
   Brown DA, 2002, LANCET, V359, P2159, DOI 10.1016/S0140-6736(02)09093-1
   Brown DA, 2007, ARTHRITIS RHEUM, V56, P753, DOI 10.1002/art.22410
   Brown DA, 2012, CANCER EPIDEM BIOMAR, V21, P337, DOI 10.1158/1055-9965.EPI-11-0786
   Cekanova M, 2009, CANCER PREV RES, V2, P450, DOI 10.1158/1940-6207.CAPR-09-0057
   Chrysovergis K, 2014, INT J OBESITY, V38, P1555, DOI 10.1038/ijo.2014.27
   Ding Q, 2009, ENDOCRINOLOGY, V150, P1688, DOI 10.1210/en.2008-0952
   Dostalova I, 2010, NUTR METAB, V7, DOI 10.1186/1743-7075-7-34
   Dostalova I, 2009, EUR J ENDOCRINOL, V161, P397, DOI 10.1530/EJE-09-0417
   Fairlie WD, 1999, J LEUKOCYTE BIOL, V65, P2, DOI 10.1002/jlb.65.1.2
   Fearon K, 2013, NAT REV CLIN ONCOL, V10, P90, DOI 10.1038/nrclinonc.2012.209
   Fearon K, 2011, LANCET ONCOL, V12, P489, DOI 10.1016/S1470-2045(10)70218-7
   Fearon KCH, 2012, CELL METAB, V16, P153, DOI 10.1016/j.cmet.2012.06.011
   Fuchs T, 2013, AGING CELL, V12, P882, DOI 10.1111/acel.12116
   Husaini Y, 2012, PLOS ONE, V7, DOI 10.1371/journal.pone.0043833
   Johnen H, 2007, NAT MED, V13, P1333, DOI 10.1038/nm1677
   Kempf T, 2006, CIRC RES, V98, P351, DOI 10.1161/01.RES.0000202805.73038.48
   Kempf T, 2007, J AM COLL CARDIOL, V50, P1054, DOI 10.1016/j.jacc.2007.04.091
   Kempf Tibor, 2009, Heart Fail Clin, V5, P537, DOI 10.1016/j.hfc.2009.04.006
   Kooh SW, 1996, BONE, V19, P181, DOI 10.1016/8756-3282(96)00162-7
   Koopmann J, 2004, CLIN CANCER RES, V10, P2386, DOI 10.1158/1078-0432.CCR-03-0165
   Lajer M, 2010, DIABETES CARE, V33, P1567, DOI 10.2337/dc09-2174
   Lecker SH, 2004, FASEB J, V18, P39, DOI 10.1096/fj.03-0610com
   Li PX, 2000, J BIOL CHEM, V275, P20127, DOI 10.1074/jbc.M909580199
   Lindmark F, 2004, JNCI-J NATL CANCER I, V96, P1248, DOI 10.1093/jnci/djh227
   Lu Z, 2014, PLOS ONE, V9
   Lu ZH, 2014, ASIAN PAC J CANCER P, V15, P6047, DOI 10.7314/APJCP.2014.15.15.6047
   Macia L, 2012, PLOS ONE, V7, DOI 10.1371/journal.pone.0034868
   Marjono AB, 2003, PLACENTA, V24, P100, DOI 10.1053/plac.2002.0881
   Moore AG, 2000, J CLIN ENDOCR METAB, V85, P4781, DOI 10.1210/jc.85.12.4781
   Muscaritoli M, 2009, CURR OPIN CLIN NUTR, V12, P378, DOI 10.1097/MCO.0b013e32832c7ae1
   Pfitzenmaier J, 2003, CANCER, V97, P1211, DOI 10.1002/cncr.11178
   Plauth M, 2002, INT J CARDIOL, V85, P83, DOI 10.1016/S0167-5273(02)00236-X
   Sartori R, 2013, NAT GENET, V45, P1309, DOI 10.1038/ng.2772
   Schiaffino S, 2013, FEBS J, V280, P4294, DOI 10.1111/febs.12253
   Skipworth RJE, 2010, BRIT J CANCER, V102, P665, DOI 10.1038/sj.bjc.6605532
   Staff AC, 2010, GYNECOL ONCOL, V118, P237, DOI 10.1016/j.ygyno.2010.05.032
   Strelau J, 2009, J NEUROSCI, V29, P13640, DOI 10.1523/JNEUROSCI.1133-09.2009
   Tanno T, 2010, CURR OPIN HEMATOL, V17, P184, DOI 10.1097/MOH.0b013e328337b52f
   Tong S, 2004, LANCET, V363, P129, DOI 10.1016/S0140-6736(03)15265-8
   Tsai VWW, 2012, J CACHEXIA SARCOPENI, V3, P239, DOI 10.1007/s13539-012-0082-6
   Tsai VWW, 2014, PLOS ONE, V9, DOI 10.1371/journal.pone.0100370
   Tsai VWW, 2013, PLOS ONE, V8, DOI 10.1371/journal.pone.0055174
   Vigano L.L. A, 2013, CANCER RES, V73
   von Haehling S, 2009, PHARMACOL THERAPEUT, V121, P227, DOI 10.1016/j.pharmthera.2008.09.009
   Wang XY, 2014, AGING-US, V6, P690, DOI 10.18632/aging.100687
   Welsh JB, 2003, P NATL ACAD SCI USA, V100, P3410, DOI 10.1073/pnas.0530278100
   Wiklund FE, 2010, AGING CELL, V9, P1057, DOI 10.1111/j.1474-9726.2010.00629.x
   Yang H, 2003, MOL CANCER THER, V2, P1023
   Zimmers TA, 2010, J CANCER RES CLIN, V136, P571, DOI 10.1007/s00432-009-0691-4
NR 66
TC 72
Z9 73
U1 1
U2 23
PU NATURE PUBLISHING GROUP
PI LONDON
PA MACMILLAN BUILDING, 4 CRINAN ST, LONDON N1 9XW, ENGLAND
SN 0307-0565
EI 1476-5497
J9 INT J OBESITY
JI Int. J. Obes.
PD FEB
PY 2016
VL 40
IS 2
SI SI
BP 193
EP 197
DI 10.1038/ijo.2015.242
PG 5
WC Endocrinology & Metabolism; Nutrition & Dietetics
WE Science Citation Index Expanded (SCI-EXPANDED)
SC Endocrinology & Metabolism; Nutrition & Dietetics
GA DD1OR
UT WOS:000369691500002
PM 26620888
OA Bronze
DA 2023-08-27
ER

PT J
AU Varendi, K
   Airavaara, M
   Anttila, J
   Vose, S
   Planken, A
   Saarma, M
   Mitchell, JR
   Andressoo, JO
AF Varendi, Kart
   Airavaara, Mikko
   Anttila, Jenni
   Vose, Sarah
   Planken, Anu
   Saarma, Mart
   Mitchell, James R.
   Andressoo, Jaan-Olle
TI Short-term Preoperative Dietary Restriction Is Neuroprotective in a Rat
   Focal Stroke Model
SO PLOS ONE
LA English
DT Article
ID CEREBRAL-ARTERY OCCLUSION; ISCHEMIA-REPERFUSION INJURY;
   TUMOR-NECROSIS-FACTOR; NEUROTROPHIC FACTOR; CARDIAC-SURGERY; TNF-ALPHA;
   CALORIE RESTRICTION; BRAIN ISCHEMIA; MICE; EXPRESSION
AB Stroke is a major complication of cardiovascular surgery, resulting in over 100,000 deaths and over a million postoperative encephalopathies annually in the US and Europe. While mitigating damage from stroke after it occurs has proven elusive, opportunities to reduce the incidence and/or severity of stroke prior to surgery in at-risk individuals remain largely unexplored. We tested the potential of short-term preoperative dietary restriction to provide neuroprotection in rat models of focal stroke. Rats were preconditioned with either three days of water-only fasting or six days of a protein free diet prior to induction of transient middle cerebral artery occlusion using two different methods, resulting in either a severe focal stroke to forebrain and midbrain, or a mild focal stroke localized to cortex only. Infarct volume, functional recovery and molecular markers of damage and protection were assessed up to two weeks after reperfusion. Preoperative fasting for 3 days reduced infarct volume after severe focal stroke. Neuroprotection was associated with modulation of innate immunity, including elevation of circulating neutrophil chemoattractant C-X-C motif ligand 1 prior to ischemia and suppression of striatal pro-inflammatory markers including tumor necrosis factor a, its receptor and downstream effector intercellular adhesion molecule-1 after reperfusion. Similarly, preoperative dietary protein restriction for 6 days reduced ischemic injury and improved functional recovery in a milder cortical infarction model. Our results suggest that short-term dietary restriction regimens may provide simple and translatable approaches to reduce perioperative stroke severity in high-risk elective vascular surgery.
C1 [Varendi, Kart; Airavaara, Mikko; Anttila, Jenni; Planken, Anu; Saarma, Mart; Andressoo, Jaan-Olle] Univ Helsinki, Inst Biotechnol, Helsinki, Finland.
   [Vose, Sarah; Mitchell, James R.] Harvard Univ, Sch Publ Hlth, Dept Genet & Complex Dis, Boston, MA 02115 USA.
C3 University of Helsinki; Harvard University; Harvard T.H. Chan School of
   Public Health
RP Mitchell, JR (通讯作者)，Harvard Univ, Sch Publ Hlth, Dept Genet & Complex Dis, 665 Huntington Ave, Boston, MA 02115 USA.
EM
RI Andressoo, Jaan-Olle/AFU-7846-2022; Andressoo, Jaan-Olle/AAA-1216-2021;
   Planken, Anu/N-6359-2019
OI Andressoo, Jaan-Olle/0000-0002-0271-3157; Planken,
   Anu/0000-0003-4553-1852; Anttila, Jenni E./0000-0001-6565-2323; Matlik,
   Kart/0000-0003-1777-5614; Airavaara, Mikko/0000-0002-2026-1609
FU Sigrid Juselius Foundation; Academy of Finland [250275, 256398, 136591,
   140983, 263700, 1126735]; Doctoral Program Brain Mind; National
   Institutes of Health [AG036712, DK090629]; Radiation Biology
   [T32CA009078]; Academy of Finland (AKA) [136591, 140983, 263700, 140983,
   263700, 136591] Funding Source: Academy of Finland (AKA)
FX The funders had no role in study design, data collection and analysis,
   decision to publish, or preparation of the manuscript. This work was
   supported by the Sigrid Juselius Foundation; the Academy of Finland
   [grant number 250275, 256398 to MA; 136591, 140983 and 263700 to JOA;
   1126735 to MS]; the Doctoral Program Brain & Mind to KV; JRM was
   supported by the National Institutes of Health [AG036712 and DK090629];
   and SV by Radiation Biology T32CA009078.
CR Ahmet I, 2005, CIRCULATION, V112, P3115, DOI 10.1161/CIRCULATIONAHA.105.563817
   Airavaara M, 2009, J COMP NEUROL, V515, P116, DOI 10.1002/cne.22039
   Amantea D, 2009, FEBS J, V276, P13, DOI 10.1111/j.1742-4658.2008.06766.x
   Anson RM, 2003, P NATL ACAD SCI USA, V100, P6216, DOI 10.1073/pnas.1035720100
   Arumugam TV, 2010, ANN NEUROL, V67, P41, DOI 10.1002/ana.21798
   Barone FC, 1997, STROKE, V28, P1233, DOI 10.1161/01.STR.28.6.1233
   BEDERSON JB, 1986, STROKE, V17, P472, DOI 10.1161/01.STR.17.3.472
   Bowen KK, 2006, NEUROCHEM INT, V49, P127, DOI 10.1016/j.neuint.2006.02.011
   Bruggemans EF, 2013, NETH HEART J, V21, P70, DOI 10.1007/s12471-012-0347-x
   Chandrasekar B, 2001, AM J PHYSIOL-HEART C, V280, pH2094, DOI 10.1152/ajpheart.2001.280.5.H2094
   CHEN ST, 1986, STROKE, V17, P738, DOI 10.1161/01.STR.17.4.738
   COMBS DJ, 1987, STROKE, V18, P503, DOI 10.1161/01.STR.18.2.503
   Damm J, 2012, NEUROSCIENCE, V201, P166, DOI 10.1016/j.neuroscience.2011.11.013
   Dirnagl U, 2009, LANCET NEUROL, V8, P398, DOI 10.1016/S1474-4422(09)70054-7
   Duan WZ, 2001, J MOL NEUROSCI, V16, P1, DOI 10.1385/JMN:16:1:1
   Duan WZ, 2001, J NEUROCHEM, V76, P619, DOI 10.1046/j.1471-4159.2001.00071.x
   Ferrer I, 2001, ACTA NEUROPATHOL, V101, P229
   Fond G, 2013, PSYCHIAT RES, V209, P253, DOI 10.1016/j.psychres.2012.12.018
   Gandhi GY, 2005, MAYO CLIN PROC, V80, P862, DOI 10.4065/80.7.862
   GO KG, 1988, METAB BRAIN DIS, V3, P257, DOI 10.1007/BF00999535
   Grandison RC, 2009, NATURE, V462, P1061, DOI 10.1038/nature08619
   Green AR, 2008, BRIT J PHARMACOL, V153, pS325, DOI 10.1038/sj.bjp.0707594
   Gregersen R, 2000, J CEREBR BLOOD F MET, V20, P53, DOI 10.1097/00004647-200001000-00009
   HALL MJ, 2010, NATL HLTH STAT REPOR, V26, P1
   Harvey BK, 2005, PHARMACOL THERAPEUT, V105, P113, DOI 10.1016/j.pharmthera.2004.09.003
   Heilbronn LK, 2006, JAMA-J AM MED ASSOC, V295, P1539, DOI 10.1001/jama.295.13.1539
   Kagansky N, 2001, ARCH NEUROL-CHICAGO, V58, P1209, DOI 10.1001/archneur.58.8.1209
   Kellermann K, 2010, SEMIN CARDIOTHORAC V, V14, P95, DOI 10.1177/1089253210370902
   KIRSCH JR, 1979, STROKE, V10, P288, DOI 10.1161/01.STR.10.3.288
   Koizumi J., 1986, JPN J STROKE, V8, P1, DOI DOI 10.3995/JSTROKE.8.1
   Kollmar R, 2007, STROKE, V38, P1585, DOI 10.1161/STROKEAHA.106.475897
   Lee J, 2002, J NEUROCHEM, V82, P1367, DOI 10.1046/j.1471-4159.2002.01085.x
   Lee J, 2000, J MOL NEUROSCI, V15, P99, DOI 10.1385/JMN:15:2:99
   Lee ST, 2012, ANN NEUROL, V72, P269, DOI 10.1002/ana.23588
   Lin T. N., 2009, P141, DOI 10.1007/978-1-60327-185-1_12
   MARIE C, 1990, METAB BRAIN DIS, V5, P65, DOI 10.1007/BF01001047
   Michalsen A, 2013, FORSCH KOMPLEMENTMED, V20, P444, DOI 10.1159/000357765
   Miller RA, 2005, AGING CELL, V4, P119, DOI 10.1111/j.1474-9726.2005.00152.x
   Mitchell JR, 2013, SURGERY, V153, P594, DOI 10.1016/j.surg.2012.09.007
   Mitchell JR, 2010, AGING CELL, V9, P40, DOI 10.1111/j.1474-9726.2009.00532.x
   Miura P, 2012, J NEUROCHEM, V120, P230, DOI 10.1111/j.1471-4159.2011.07583.x
   MURRY CE, 1986, CIRCULATION, V74, P1124, DOI 10.1161/01.CIR.74.5.1124
   Newman MF, 2006, LANCET, V368, P694, DOI 10.1016/S0140-6736(06)69254-4
   Pandya JD, 2011, BRAIN RES, V1384, P151, DOI 10.1016/j.brainres.2011.01.102
   Peng Wei, 2012, Sci Transl Med, V4, p118ra11, DOI 10.1126/scitranslmed.3002629
   PERDRIZET G A, 1989, Current Surgery, V46, P23
   Poldermans D, 2009, EUR HEART J, V30, P2769, DOI 10.1093/eurheartj/ehp337
   Reglodi D, 2000, EXP NEUROL, V163, P399, DOI 10.1006/exnr.2000.7367
   REHNCRONA S, 1985, EXP NEUROL, V87, P458, DOI 10.1016/0014-4886(85)90176-1
   Robertson LT, 2013, EXP GERONTOL, V48, P1043, DOI 10.1016/j.exger.2013.01.009
   Rosenzweig HL, 2007, J CEREBR BLOOD F MET, V27, P1663, DOI 10.1038/sj.jcbfm.9600464
   Schabitz WR, 1997, J CEREBR BLOOD F MET, V17, P500, DOI 10.1097/00004647-199705000-00003
   Selim M, 2007, NEW ENGL J MED, V356, P706, DOI 10.1056/NEJMra062668
   Sharp Frank R, 2004, NeuroRx, V1, P26, DOI 10.1007/BF03206565
   SIMONET WS, 1994, J CLIN INVEST, V94, P1310, DOI 10.1172/JCI117450
   Speakman JR, 2011, MOL ASPECTS MED, V32, P159, DOI 10.1016/j.mam.2011.07.001
   Thored P, 2009, GLIA, V57, P835, DOI 10.1002/glia.20810
   van Ginhoven TM, 2010, J SURG RES
   Varon J, 2008, CHEST, V133, P1267, DOI 10.1378/chest.07-2190
   Yang L, 2005, BLOOD, V106, P584, DOI 10.1182/blood-2004-12-4942
   Yenari MA, 2010, NEUROTHERAPEUTICS, V7, P378, DOI 10.1016/j.nurt.2010.07.005
   Yu ZF, 1999, J NEUROSCI RES, V57, P830, DOI 10.1002/(SICI)1097-4547(19990915)57:6&lt;830::AID-JNR8&gt;3.0.CO;2-2
   Zeynalov E, 2009, NEUROBIOL DIS, V35, P264, DOI 10.1016/j.nbd.2009.05.010
NR 63
TC 25
Z9 26
U1 0
U2 12
PU PUBLIC LIBRARY SCIENCE
PI SAN FRANCISCO
PA 1160 BATTERY STREET, STE 100, SAN FRANCISCO, CA 94111 USA
SN 1932-6203
J9 PLOS ONE
JI PLoS One
PD APR 4
PY 2014
VL 9
IS 4
AR e93911
DI 10.1371/journal.pone.0093911
PG 9
WC Multidisciplinary Sciences
WE Science Citation Index Expanded (SCI-EXPANDED)
SC Science & Technology - Other Topics
GA AE6MJ
UT WOS:000334107500093
PM 24705386
OA Green Published, gold, Green Submitted
DA 2023-08-27
ER

PT J
AU Langen, B
   Rudqvist, N
   Helou, K
   Forssell-Aronsson, E
AF Langen, Britta
   Rudqvist, Nils
   Helou, Khalil
   Forssell-Aronsson, Eva
TI Microarray Studies on At-211 Administration in BALB/c Nude Mice Indicate
   Systemic Effects on Transcriptional Regulation in Nonthyroid Tissues
SO JOURNAL OF NUCLEAR MEDICINE
LA English
DT Article
DE At-211; radionuclide therapy; normal tissue response; systemic effects;
   radiogenomics
ID MONOCLONAL-ANTIBODY; MOUSE-TISSUES; RADIATION; EXPRESSION; ASTATINE-211;
   IRRADIATION; INDUCTION; DOSIMETRY; TRANSPORT; I-131
AB Targeted alpha-therapy is a promising treatment option for various types of malignant tumors. Radiolabeled cancer-seeking agents, however, undergo degradation, resulting in a certain percentage of free radio-nuclide in the body. The radiohalogen At-211 accumulates in various tissues, with specifically high uptake in the thyroid. When normal thyroid function is disturbed because of ionizing radiation (IR) exposure, deleterious effects can occur in tissues that depend on thyroid hormone (TH) regulation for normal physiologic function. However, knowledge of systemic effects is still rudimentary. We previously reported similarities in transcriptomic regulation between the thyroid and other tissues despite large differences in absorbed dose from At-211. Here, we present supportive evidence on systemic effects after At-211 administration. Methods: Expression microarray data from the kidney cortex and medulla, liver, lungs, and spleen were used from previous studies in which mice were intravenously injected with 0.064-42 kBq of At-211 and killed after 24 h or injected with 1.7 kBq of At-211 and killed after 1, 6, or 168 h. Controls were mock-treated and killed after 24 h. Literature-based gene signatures were used to evaluate the relative impact from IR- or TH-induced regulation. Thyroid- and TH-associated upstream regulators as well as thyroid-related diseases and functions were generated using functional analysis software. Results: Responses in IR- or TH-associated gene signatures were tissue specific and varied over time, and the relative impact of each gene signature differed between the investigated tissues. The liver showed a clear dominance of TH-responding genes. In the kidney cortex, kidney medulla, and lungs, the TH-associated signature was detected to at least an extent similar to the IR-associated signature. The spleen was the single tissue showing regulation of only IR-associated signature genes. Various thyroid-associated diseases and functions were inferred from the data: L-triiodothyronine, TH, TH receptor, and triiodothyronine (reverse) were inferred as upstream regulators with differences in incidence and strength of regulation depending on tissue type. Conclusion: These findings indicate that transcriptional regulation in various nonthyroid tissues was-in part-induced by thyroid (hormone)-dependent signaling. Consideration of the systemic context between tissues could contribute to normal tissue risk assessment and planning of remedial measures.
C1 [Langen, Britta; Rudqvist, Nils; Forssell-Aronsson, Eva] Univ Gothenburg, Sahlgrenska Acad, Sahlgrenska Canc Ctr, Dept Radiat Phys,Inst Clin Sci, Gothenburg, Sweden.
   [Langen, Britta] Chalmers Univ Technol, Dept Appl Phys, Gothenburg, Sweden.
   [Helou, Khalil] Univ Gothenburg, Sahlgrenska Acad, Sahlgrenska Canc Ctr, Dept Oncol,Inst Clin Sci, Gothenburg, Sweden.
C3 University of Gothenburg; Chalmers University of Technology; University
   of Gothenburg
RP Langen, B (通讯作者)，Univ Gothenburg, Sahlgrenska Univ Hosp, Radiat Dept, SE-41345 Gothenburg, Sweden.
EM
OI Forssell-Aronsson, Eva/0000-0002-7558-4368
FU European Commission FP7 Collaborative Project TARCC
   [HEALTH-F2-2007-201962]; Swedish Research Council [21073]; Swedish
   Cancer Society [3427]; BioCARE-a National Strategic Research Program at
   the University of Gothenburg; Swedish Radiation Safety Authority; King
   Gustav V Jubilee Clinic Cancer Research Foundation; Sahlgrenska
   University Hospital Research Funds; Assar Gabrielsson Cancer Research
   Foundation; Adlerbertska Research Fund; Wilhelm och Martina Lundgren
   science trust fund
FX We thank the following organizations for financial support of this
   study: the European Commission FP7 Collaborative Project TARCC
   HEALTH-F2-2007-201962, the Swedish Research Council (grant no. 21073),
   the Swedish Cancer Society (grant no. 3427), BioCARE-a National
   Strategic Research Program at the University of Gothenburg, the Swedish
   Radiation Safety Authority, the King Gustav V Jubilee Clinic Cancer
   Research Foundation, the Sahlgrenska University Hospital Research Funds,
   the Assar Gabrielsson Cancer Research Foundation, the Adlerbertska
   Research Fund, and the Wilhelm och Martina Lundgren science trust fund.
   No other potential conflict of interest relevant to this article was
   reported.
CR ACKERMAN JL, 1962, AMER J ROENTGENOL RA, V87, P543
   Andersson H, 2000, ACTA ONCOL, V39, P741
   Andersson H, 2009, J NUCL MED, V50, P1153, DOI 10.2967/jnumed.109.062604
   BENJAMINI Y, 1995, J R STAT SOC B, V57, P289, DOI 10.1111/j.2517-6161.1995.tb02031.x
   Bolch WE, 2009, J NUCL MED, V50, P477, DOI 10.2967/jnumed.108.056036
   BROWN I, 1986, APPL RADIAT ISOTOPES, V37, P789, DOI 10.1016/0883-2889(86)90273-X
   Chaudhry MA, 2008, J BIOMED SCI, V15, P557, DOI 10.1007/s11373-008-9253-z
   GARG PK, 1990, CANCER RES, V50, P3514
   HAMILTON JG, 1954, P SOC EXP BIOL MED, V86, P366
   HAMILTON JG, 1954, J CLIN ENDOCR METAB, V14, P1161, DOI 10.1210/jcem-14-10-1161
   HARRISON A, 1989, RADIOCHIM ACTA, V47, P157
   Langen B, 2015, EJNMMI RES, V5, P1, DOI 10.1186/s13550-014-0078-7
   Langen B, 2013, J NUCL MED, V54, P990, DOI 10.2967/jnumed.112.114462
   Larsen RH, 1998, RADIAT RES, V149, P155, DOI 10.2307/3579925
   Lindegren S, 2001, APPL RADIAT ISOTOPES, V55, P157, DOI 10.1016/S0969-8043(01)00044-6
   Lindencrona U, 2001, NUCL MED BIOL, V28, P41, DOI 10.1016/S0969-8051(00)00179-7
   Lundh C, 2009, J NUCL MED, V50, P1161, DOI 10.2967/jnumed.108.061150
   Mothersill C, 1998, RADIAT RES, V149, P256, DOI 10.2307/3579958
   Mothersill C., 2005, BJR S, V27, P128
   Nagasawa H, 1999, RADIAT RES, V152, P552, DOI 10.2307/3580153
   NAGASAWA H, 1992, CANCER RES, V52, P6394
   Parris TZ, 2010, CLIN CANCER RES, V16, P3860, DOI 10.1158/1078-0432.CCR-10-0889
   Phillips R D, 1965, Res Dev Tech Rep, P1
   PHILLIPS RD, 1966, AM J PHYSIOL, V210, P1096, DOI 10.1152/ajplegacy.1966.210.5.1096
   Rudqvist N, 2015, PLOS ONE, V10, DOI 10.1371/journal.pone.0131686
   Rudqvist N, 2012, EJNMMI RES, V2, DOI 10.1186/2191-219X-2-32
   Seymour C B, 1997, Radiat Oncol Investig, V5, P106, DOI 10.1002/(SICI)1520-6823(1997)5:3<106::AID-ROI4>3.0.CO;2-1
   SKOOG W A, 1962, Stanford Med Bull, V20, P87
   Snyder AR, 2004, CANCER METAST REV, V23, P259, DOI 10.1023/B:CANC.0000031765.17886.fa
   Willers H, 2006, HEMATOL ONCOL CLIN N, V20, P1, DOI 10.1016/j.hoc.2006.01.007
   YUILE C L, 1970, Radiation Research, V44, P821, DOI 10.2307/3573159
NR 31
TC 6
Z9 6
U1 0
U2 6
PU SOC NUCLEAR MEDICINE INC
PI RESTON
PA 1850 SAMUEL MORSE DR, RESTON, VA 20190-5316 USA
SN 0161-5505
EI 1535-5667
J9 J NUCL MED
JI J. Nucl. Med.
PD FEB
PY 2017
VL 58
IS 2
BP 346
EP 353
DI 10.2967/jnumed.116.176958
PG 8
WC Radiology, Nuclear Medicine & Medical Imaging
WE Science Citation Index Expanded (SCI-EXPANDED)
SC Radiology, Nuclear Medicine & Medical Imaging
GA EJ6WH
UT WOS:000393360100032
PM 27765860
OA Bronze
DA 2023-08-27
ER

PT J
AU Hoshino, M
   Nakamura, S
   Mori, K
   Kawauchi, T
   Terao, M
   Nishimura, YV
   Fukuda, A
   Fuse, T
   Matsuo, N
   Sone, M
   Watanabe, M
   Bito, H
   Terashima, T
   Wright, CVE
   Kawaguchi, Y
   Nakao, K
   Nabeshima, YI
AF Hoshino, M
   Nakamura, S
   Mori, K
   Kawauchi, T
   Terao, M
   Nishimura, YV
   Fukuda, A
   Fuse, T
   Matsuo, N
   Sone, M
   Watanabe, M
   Bito, H
   Terashima, T
   Wright, CVE
   Kawaguchi, Y
   Nakao, K
   Nabeshima, YI
TI Ptf1a, a bHLH transcriptional gene, defines GABAergic neuronal fates in
   cerebellum
SO NEURON
LA English
DT Article
ID NUCLEOTIDE EXCHANGE FACTORS; KLOTHO GENE; CELL-DEATH; MOUSE; EXPRESSION;
   RAC1; GROWTH; STEF; IDENTIFICATION; PROLIFERATION
AB The molecular machinery governing glutamatergic-GABAergic neuronal subtype specification is unclear. Here we describe a cerebellar mutant, cerebelless, which lacks the entire cerebellar cortex in adults. The primary defect of the mutant brains was a specific inhibition of GABAergic neuron production from the cerebellar ventricular zone (VZ), resulting in secondary and complete loss of external germinal layer, pontine, and olivary nuclei during development. We identified the responsible gene, Ptf1a, whose expression was lost in the cerebellar VZ but was maintained in the pancreas in cerebelless. Lineage tracing revealed that two types of neural precursors exist in the cerebellar VZ: Ptf1a-expressing and -nonexpressing precursors, which generate GABAergic and glutamatergic neurons, respectively. Introduction of Ptf1a into glutamatergic neuron precursors in the dorsal telencephalon generated GABAergic neurons with representative morphological and migratory features. Our results suggest that Ptf1a is involved in driving neural precursors to differentiate into GABAergic neurons in the cerebellum.
C1 Kyoto Univ, Grad Sch Med, Dept Pathol & Tumor Biol, Sakyo Ku, Kyoto 6068501, Japan.
   Japan Sci & Technol Agcy, PRESTO, Kawaguchi, Saitama 3320012, Japan.
   Kyoto Univ, Grad Sch Med, Dept Med & Clin Sci, Sakyo Ku, Kyoto 6068507, Japan.
   Kyoto Univ, Grad Sch Med, Dept Surg & Surg Basic Sci, Sakyo Ku, Kyoto 6068507, Japan.
   Kyoto Univ, Grad Sch Med, Dept Gastroenterol & Hepatol, Sakyo Ku, Kyoto 6068507, Japan.
   Columbia Univ, Coll Phys & Surg, Dept Med, New York, NY 10032 USA.
   Kobe Univ, Grad Sch Med, Dept Neurosci, Div Anat & Neurobiol, Kobe, Hyogo 6500017, Japan.
   Vanderbilt Univ, Sch Med, Dept Cell & Dev Biol, Vanderbilt Dev Biol Program, Nashville, TN 37232 USA.
   Hokkaido Univ, Sch Med, Dept Anat, Sapporo, Hokkaido 0608638, Japan.
   Univ Tokyo, Grad Sch Med, Dept Neurochem, Bunkyo Ku, Tokyo 1130033, Japan.
C3 Kyoto University; Japan Science & Technology Agency (JST); Kyoto
   University; Kyoto University; Kyoto University; Columbia University;
   Kobe University; Vanderbilt University; Hokkaido University; University
   of Tokyo
RP Hoshino, M (通讯作者)，Kyoto Univ, Grad Sch Med, Dept Pathol & Tumor Biol, Sakyo Ku, Kyoto 6068501, Japan.
EM
RI WATANABE, Masahiko/A-4055-2012; Watanabe, Masahiko/S-3614-2019;
   Kawauchi, Takeshi/L-4770-2013; Bito, Haruhiko/A-8635-2008; Mori,
   Kiyoshi/N-1267-2014; Matsuo, Naoki/B-9929-2008; Sone, Masaki/Q-9058-2019
OI WATANABE, Masahiko/0000-0001-5037-7138; Watanabe,
   Masahiko/0000-0001-5037-7138; Kawauchi, Takeshi/0000-0002-8650-4474;
   Mori, Kiyoshi/0000-0002-7285-8351; Matsuo, Naoki/0000-0003-0988-4706;
   Bito, Haruhiko/0000-0001-6315-9594
CR ABERCROMBIE M, 1946, ANAT REC, V94, P239, DOI 10.1002/ar.1090940210
   AKAZAWA C, 1995, J BIOL CHEM, V270, P8730, DOI 10.1074/jbc.270.15.8730
   Ang ESBC, 2003, J NEUROSCI, V23, P5805
   BATINI C, 1992, J COMP NEUROL, V315, P74, DOI 10.1002/cne.903150106
   BenArie N, 1997, NATURE, V390, P169, DOI 10.1038/36579
   Bennett MR, 2002, AUTON NEUROSCI-BASIC, V95, P1, DOI 10.1016/S1566-0702(01)00358-7
   Casarosa S, 1999, DEVELOPMENT, V126, P525
   Chan-Palay V., 1974, CEREBELLAR CORTEX CY
   Cheng LP, 2004, NAT NEUROSCI, V7, P510, DOI 10.1038/nn1221
   Chu T, 2000, J NEUROBIOL, V43, P18, DOI 10.1002/(SICI)1097-4695(200004)43:1<18::AID-NEU2>3.0.CO;2-C
   COLLINS P, 1995, GRAYS ANATOMY
   Dahmane N, 1999, DEVELOPMENT, V126, P3089
   Fode C, 2000, GENE DEV, V14, P67
   Goldowitz D, 2000, J NEUROSCI, V20, P4129
   Gu GQ, 2002, DEVELOPMENT, V129, P2447
   HECKROTH JA, 1994, J COMP NEUROL, V343, P173, DOI 10.1002/cne.903430113
   Herrup K, 1996, DEV BRAIN RES, V96, P28, DOI 10.1016/0165-3806(96)00069-7
   Hoshinoso M, 1999, J BIOL CHEM, V274, P17837, DOI 10.1074/jbc.274.25.17837
   Hoveyda N, 1999, J MED GENET, V36, P700
   Jacobowitz, 1998, CHEMOARCHITECTONIC A
   Jankovski A, 1996, EUR J NEUROSCI, V8, P2308, DOI 10.1111/j.1460-9568.1996.tb01194.x
   KANEKO T, 1989, NEUROSCIENCE, V32, P79, DOI 10.1016/0306-4522(89)90109-7
   Kawaguchi Y, 2002, NAT GENET, V32, P128, DOI 10.1038/ng959
   Kawauchi T, 2005, BIOCHEM BIOPH RES CO, V331, P50, DOI 10.1016/j.bbrc.2005.03.132
   Kawauchi T, 2003, EMBO J, V22, P4190, DOI 10.1093/emboj/cdg413
   Krapp A, 1998, GENE DEV, V12, P3752, DOI 10.1101/gad.12.23.3752
   Kriegstein AR, 2003, GLIA, V43, P37, DOI 10.1002/glia.10250
   Kuroo M, 1997, NATURE, V390, P45, DOI 10.1038/36285
   Lewis PM, 2004, DEV BIOL, V270, P393, DOI 10.1016/j.ydbio.2004.03.007
   Mathis L, 2003, DEV BIOL, V258, P20, DOI 10.1016/S0012-1606(03)00098-8
   Matsumura Y, 1998, BIOCHEM BIOPH RES CO, V242, P626, DOI 10.1006/bbrc.1997.8019
   Matsuo N, 2003, MOL CELL NEUROSCI, V24, P69, DOI 10.1016/S1044-7431(03)00122-2
   Matsuo N, 2002, J BIOL CHEM, V277, P2860, DOI 10.1074/jbc.M106186200
   MIALE IL, 1961, EXP NEUROL, V4, P277, DOI 10.1016/0014-4886(61)90055-3
   NIWA H, 1991, GENE, V108, P193, DOI 10.1016/0378-1119(91)90434-D
   Obata J, 2001, GENES CELLS, V6, P345, DOI 10.1046/j.1365-2443.2001.00422.x
   Parras CM, 2002, GENE DEV, V16, P324, DOI 10.1101/gad.940902
   PIERCE E T, 1975, Brain Research, V95, P503, DOI 10.1016/0006-8993(75)90124-9
   PINKERT CA, 1987, GENE DEV, V1, P268, DOI 10.1101/gad.1.3.268
   Ross SE, 2003, NEURON, V39, P13, DOI 10.1016/S0896-6273(03)00365-9
   Schuurmans C, 2004, EMBO J, V23, P2892, DOI 10.1038/sj.emboj.7600278
   Sellick GS, 2004, NAT GENET, V36, P1301, DOI 10.1038/ng1475
   Sellick GS, 2003, DIABETES, V52, P2636, DOI 10.2337/diabetes.52.10.2636
   Soriano P, 1999, NAT GENET, V21, P70, DOI 10.1038/5007
   Tan SS, 1998, NEURON, V21, P295, DOI 10.1016/S0896-6273(00)80539-5
   Temple S, 2001, NATURE, V414, P112, DOI 10.1038/35102174
   Wallace VA, 1999, CURR BIOL, V9, P445, DOI 10.1016/S0960-9822(99)80195-X
   Wang VY, 2001, NAT REV NEUROSCI, V2, P484, DOI 10.1038/35081558
   Wechsler-Reya RJ, 1999, NEURON, V22, P103, DOI 10.1016/S0896-6273(00)80682-0
   Wilson SW, 2000, NEURON, V28, P641, DOI 10.1016/S0896-6273(00)00171-9
   Yoshizawa M, 2005, J NEUROSCI, V25, P4406, DOI 10.1523/JNEUROSCI.4955-04.2005
   Yoshizawa M, 2003, GENE EXPR PATTERNS, V3, P375, DOI 10.1016/S1567-133X(03)00002-4
   Yoshizawa M, 2002, MECH DEVELOP, V113, P65, DOI 10.1016/S0925-4773(01)00650-5
NR 53
TC 361
Z9 366
U1 1
U2 14
PU CELL PRESS
PI CAMBRIDGE
PA 50 HAMPSHIRE ST, FLOOR 5, CAMBRIDGE, MA 02139 USA
SN 0896-6273
EI 1097-4199
J9 NEURON
JI Neuron
PD JUL 21
PY 2005
VL 47
IS 2
BP 201
EP 213
DI 10.1016/j.neuron.2005.06.007
PG 13
WC Neurosciences
WE Science Citation Index Expanded (SCI-EXPANDED)
SC Neurosciences & Neurology
GA 948CP
UT WOS:000230693900008
PM 16039563
OA Bronze
DA 2023-08-27
ER

PT J
AU Agrawal, A
   Timothy, J
   Cincu, R
   Agarwal, T
   Waghmare, LB
AF Agrawal, Amit
   Timothy, Jake
   Cincu, Rafael
   Agarwal, Trisha
   Waghmare, Lalit B.
TI Bradycardia in neurosurgery
SO CLINICAL NEUROLOGY AND NEUROSURGERY
LA English
DT Review
DE bradycardia; cushing reflex; intracranial pressure
ID SPINAL-CORD-INJURY; ENDOSCOPIC 3RD VENTRICULOSTOMY; NEGATIVE-PRESSURE
   DRAINAGE; INCREASED INTRACRANIAL-PRESSURE; POSTERIOR CEREBELLAR CORTEX;
   TEMPORAL-LOBE EPILEPSY; HEART-RATE; AUTONOMIC DYSREFLEXIA;
   BLOOD-PRESSURE; OCULOCARDIAC REFLEX
AB Cushing reflex' is characterized by the occurrence of hypertension, bradycardia and apnoea secondary to raised increased intracranial pressure (ICP), leading to pressure on and or stretch, or both, of the brainstem. With the wide availability of monitoring facilities and advancements in investigation techniques, observation of increased intracranial pressure resulting in haemodynamic instability and bradycardia has been increasingly recognized in relation to many neurosurgical conditions and procedures. The causes of bradycardia include space occupying lesion involving or compressing the brain parenchyma (subdural haematoma, tumours, hydrocephalus), neurosurgical procedures (neuroendoscopy, placement of extradural drains), epileptic and non-epileptic seizures, trigemino-cardiac reflex, cerebellar lesions, spinal lesions (neurogenic shock, autonomic dysreflexia) and many other rare causes (Ventricular catheter obstruction in cases of hydrocephalus, colloid cysts related acute neurogenic cardiac dysfunction, Ondine's curse syndrome, etc.). This highlights that bradycardia can be a warning sign in many neurosurgical conditions and Cushing's reflex is a protective and effective action of the brain for preserving an adequate cerebral perfusion pressure despite an increased intracranial pressure. Management of these patients include identification and treatment of the underlying cause of bradycardia, anti-cholinergics and if necessary cardiac pacing, nevertheless, other causes of haemodynamic changes (i.e. anesthetic drugs, tumor manipulation) should also be considered and managed accordingly. We believe that this knowledge and understanding will help to identify the patients' at risk and will also help in the management of neurosurgical patients with bradycardia. (c) 2008 Elsevier B.V. All rights reserved.
C1 [Agrawal, Amit] Datta Meghe Inst Med Sci, Div Neurosurg, Dept Surg, Wardha 442004, Maharashtra, India.
   [Timothy, Jake] Leeds Gen Infirm, Dept Neurosurg, Leeds LS1 3EX, W Yorkshire, England.
   [Cincu, Rafael] Miguel Servet Univ Hosp, Dept Neurosurg, Zaragoza, Spain.
   [Agarwal, Trisha] No Gen Hosp, Sheffield S5 7AU, S Yorkshire, England.
   [Waghmare, Lalit B.] Datta Meghe Inst Med Sci, Dept Physiol, Wardha 442004, Maharashtra, India.
C3 Datta Meghe Institute of Higher Education & Research (Deemed to be
   University); Jawaharlal Nehru Medical College Wardha; Leeds General
   Infirmary; University of Leeds; Miguel Servet University Hospital;
   Northern General Hospital; Datta Meghe Institute of Higher Education &
   Research (Deemed to be University); Jawaharlal Nehru Medical College
   Wardha
RP Agrawal, A (通讯作者)，Datta Meghe Inst Med Sci, Div Neurosurg, Dept Surg, Wardha 442004, Maharashtra, India.
EM
RI Agrawal, Amit/C-1511-2011
OI Agrawal, Amit/0000-0002-3287-5448; Waghmare, Lalit/0000-0002-3311-7657
CR ALFERY DD, 1980, ANESTHESIOLOGY, V52, P443, DOI 10.1097/00000542-198005000-00016
   Anandh B, 2002, MINIM INVAS NEUROSUR, V45, P154, DOI 10.1055/s-2002-34339
   [Anonymous], 2000, EPILEPTIC SEIZURES P
   Ansakorpi H, 2002, J NEUROL NEUROSUR PS, V72, P26, DOI 10.1136/jnnp.72.1.26
   BAHL CP, 1967, BRIT J ANAESTH, V39, P657, DOI 10.1093/bja/39.8.657
   BLANC VF, 1983, CAN ANAESTH SOC J, V30, P360, DOI 10.1007/BF03007858
   BRADLEY DJ, 1987, J PHYSIOL-LONDON, V393, P107, DOI 10.1113/jphysiol.1987.sp016813
   BRADLEY DJ, 1987, J PHYSIOL-LONDON, V383, P537
   Britton JW, 2006, EPILEPSIA, V47, P737, DOI 10.1111/j.1528-1167.2006.00509.x
   CAEMAERT J, 1993, ACTA NEUROCHIR, V124, P11, DOI 10.1007/BF01400707
   Carvalho KS, 2004, SEIZURE-EUR J EPILEP, V13, P595, DOI 10.1016/j.seizure.2004.01.004
   Cha ST, 2002, J CRANIO MAXILL SURG, V30, P108, DOI 10.1054/jcms.2001.0264
   Chitravanshi VC, 2007, NEUROSCI LETT, V416, P188, DOI 10.1016/j.neulet.2007.02.007
   Cushing H, 1903, AM J MED SCI, V125, P1017, DOI 10.1097/00000441-190306000-00006
   Cushing H., 1901, JOHNS HOPKINS HOSP B, V126, P290
   de Oliveira GR, 2007, ARQ NEURO-PSIQUIAT, V65, P565, DOI 10.1590/S0004-282X2007000400003
   Deininger MH, 2006, J NEUROSURG, V105, P465, DOI 10.3171/jns.2006.105.3.465
   Deschamps A, 2004, CAN J ANAESTH, V51, P277, DOI 10.1007/BF03019113
   DOBA N, 1972, BRAIN RES, V47, P487, DOI 10.1016/0006-8993(72)90655-5
   Dumont RJ, 2001, CLIN NEUROPHARMACOL, V24, P254, DOI 10.1097/00002826-200109000-00002
   El-Dawlatly AA, 2000, ANESTH ANALG, V91, P1142, DOI 10.1097/00000539-200011000-00019
   El-Dawlatly AA, 1999, MINIM INVAS NEUROSUR, V42, P198, DOI 10.1055/s-2008-1053398
   EPSTEIN MA, 1992, NEUROLOGY, V42, P50, DOI 10.1212/WNL.42.1.50
   Fabregas N, 2000, J NEUROSURG ANESTH, V12, P21, DOI 10.1097/00008506-200001000-00005
   FINESTONE HM, 1993, AM J PHYS MED REHAB, V72, P395
   Franga DL, 2006, AM SURGEON, V72, P525
   FREEMAN R, 1995, SEMIN NEUROL, V15, P158, DOI 10.1055/s-2008-1041019
   GARDNER WJ, 1962, J NEUROSURG, V19, P947, DOI 10.3171/jns.1962.19.11.0947
   Gilsanz F J, 1984, Rev Esp Anestesiol Reanim, V31, P20
   GO KG, 1981, CLIN NEUROL NEUROSUR, V83, P47, DOI 10.1016/0303-8467(81)90001-9
   Goncalves V, 2005, CEREBELLUM, V4, P246, DOI 10.1080/14734220500325863
   Goncalves V, 2002, AUTON NEUROSCI-BASIC, V101, P78, DOI 10.1016/S1566-0702(02)00204-7
   GONCALVES V, 2006, CLIN AUTON RES, V16, P182
   HANDLER MH, 1994, NEUROSURGERY, V35, P525, DOI 10.1227/00006123-199409000-00025
   Harati Y., 1997, CLIN AUTONOMIC DISOR, P455
   Hernandez-Palazon J, 1997, Rev Esp Anestesiol Reanim, V44, P161
   Hernandez-Palazon J, 1998, BRIT J ANAESTH, V80, P599, DOI 10.1093/bja/80.5.599
   Heymans C, 1928, AM J PHYSIOL, V85, P498, DOI 10.1152/ajplegacy.1928.85.3.498
   HOFF JT, 1970, ARCH NEUROL-CHICAGO, V23, P228, DOI 10.1001/archneur.1970.00480270038006
   JACKA M, 1994, ANESTH ANALG, V78, P1022
   Jarquin-Valdivia AA, 2005, CLIN NEUROL NEUROSUR, V107, P361, DOI 10.1016/j.clineuro.2004.09.012
   Johnson C D, 1998, Bol Asoc Med P R, V90, P95
   Kalmar AF, 2005, BRIT J ANAESTH, V94, P791, DOI 10.1093/bja/aei121
   Karlsson AK, 2006, PROG BRAIN RES, V152, P1, DOI 10.1016/S0079-6123(05)52034-X
   Karlsson AK, 1999, SPINAL CORD, V37, P383, DOI 10.1038/sj.sc.3100867
   KAWAMOTO M, 1993, ANAESTH INTENS CARE, V21, P218, DOI 10.1177/0310057X9302100217
   KORKES H, 2006, ARQ BRAS CARDIOL, V87, P189
   Kosaka M, 2000, J CRANIO MAXILL SURG, V28, P106, DOI 10.1054/jcms.2000.0107
   KOTWICA Z, 1991, British Journal of Neurosurgery, V5, P461, DOI 10.3109/02688699108998474
   Kratschmer F, 1870, SBER AKAD WISS WIEN, V62, P147
   Kraus J, 1999, CLIN NEUROL NEUROSUR, V101, P196, DOI 10.1016/S0303-8467(99)00023-2
   LANE RD, 1992, STROKE, V23, P362, DOI 10.1161/01.STR.23.3.362
   LAURENSON VG, 1985, NEUROSURGERY, V17, P868, DOI 10.1097/00006123-198511000-00025
   LEHMANN KG, 1987, J AM COLL CARDIOL, V10, P46, DOI 10.1016/S0735-1097(87)80158-4
   Leutmezer F, 2003, EPILEPSIA, V44, P348, DOI 10.1046/j.1528-1157.2003.34702.x
   LIN RH, 1994, CLIN NEUROL NEUROSUR, V96, P152, DOI 10.1016/0303-8467(94)90051-5
   LINDAN R, 1980, PARAPLEGIA, V18, P285, DOI 10.1038/sc.1980.51
   Locatelli ER, 1999, ANN INTERN MED, V130, P581, DOI 10.7326/0003-4819-130-7-199904060-00018
   Mandal AKJ, 2005, HEART, V91, DOI 10.1136/hrt.2004.046318
   Mandal NG, 2004, CAN J ANAESTH, V51, P942, DOI 10.1007/BF03018898
   Massetani R, 1997, EPILEPSIA, V38, P363, DOI 10.1111/j.1528-1157.1997.tb01129.x
   MCCULLOCH GAJ, 1981, NEUROSURGERY, V9, P380, DOI 10.1227/00006123-198110000-00005
   Nashef L, 1996, J NEUROL NEUROSUR PS, V60, P297, DOI 10.1136/jnnp.60.3.297
   OGILVY CS, 1987, BIOL NEONATE, V52, P327
   Opherk C, 2002, NEUROLOGY, V58, P636, DOI 10.1212/WNL.58.4.636
   OPPENHEIMER SM, 1992, NEUROLOGY, V42, P1727, DOI 10.1212/WNL.42.9.1727
   Piepmeier J M, 1985, Cent Nerv Syst Trauma, V2, P153
   Rabchevsky AG, 2006, PROG BRAIN RES, V152, P265, DOI 10.1016/S0079-6123(05)52017-X
   RAGNO JR, 1989, J ORAL MAXIL SURG, V47, P1082, DOI 10.1016/0278-2391(89)90186-9
   Rehman HU, 1999, J ROY SOC MED, V92, P585, DOI 10.1177/014107689909201113
   Riley HA., 1942, T AM NEUROL ASSOC, V68, P28
   Rocha I, 2008, PHYSIOL RES, V57, P701, DOI 10.33549/physiolres.931262
   Ruiz-Arango Andres F, 2006, Cardiol Rev, V14, pe8, DOI 10.1097/01.crd.0000184453.29079.0f
   Schaller B, 2005, CLIN NEUROL NEUROSUR, V107, P468, DOI 10.1016/j.clineuro.2004.12.004
   SNOW JC, 1978, PARAPLEGIA, V15, P327, DOI 10.1038/sc.1977.49
   So EL, 2000, EPILEPSIA, V41, P1494, DOI 10.1111/j.1528-1157.2000.tb00128.x
   Teasell RW, 2000, ARCH PHYS MED REHAB, V81, P506, DOI 10.1053/mr.2000.3848
   THOMPSON ME, 1987, AM J PHYSIOL, V253, pR150, DOI 10.1152/ajpregu.1987.253.1.R150
   TOLEDO E, 1980, ACTA NEUROCHIR, V52, P55, DOI 10.1007/BF01400947
   van Aken J, 2003, MINIM INVAS NEUROSUR, V46, P198
   VOLKER KH, 1990, NEUROSURG CLIN N AM, V1, P729
   WASNICK JD, 1993, ANESTH ANALG, V76, P432
   Weant KA, 2007, NEUROCRIT CARE, V7, P250, DOI 10.1007/s12028-007-0067-z
   Webb Michael D, 2007, Anesth Prog, V54, P7, DOI 10.2344/0003-3006(2007)54[7:AMOTTR]2.0.CO;2
   ZAGON A, 1993, NEUROSCIENCE, V54, P729, DOI 10.1016/0306-4522(93)90243-9
   ZAMRINI EY, 1990, NEUROLOGY, V40, P1408, DOI 10.1212/WNL.40.9.1408
NR 86
TC 46
Z9 47
U1 0
U2 9
PU ELSEVIER SCIENCE BV
PI AMSTERDAM
PA PO BOX 211, 1000 AE AMSTERDAM, NETHERLANDS
SN 0303-8467
EI 1872-6968
J9 CLIN NEUROL NEUROSUR
JI Clin. Neurol. Neurosurg.
PD APR
PY 2008
VL 110
IS 4
BP 321
EP 327
DI 10.1016/j.clineuro.2008.01.013
PG 7
WC Clinical Neurology; Surgery
WE Science Citation Index Expanded (SCI-EXPANDED)
SC Neurosciences & Neurology; Surgery
GA 296UC
UT WOS:000255570300001
PM 18329164
DA 2023-08-27
ER

PT J
AU Tschan, C
   Gaab, MR
   Krauss, JK
   Oertel, J
AF Tschan, Christoph
   Gaab, Michael Robert
   Krauss, Joachim Kurt
   Oertel, Joachim
TI Waterjet dissection of the vestibulocochlear nerve: an experimental
   study Laboratory investigation
SO JOURNAL OF NEUROSURGERY
LA English
DT Article
DE cranial nerve; hydrojet dissection; vestibular schwannoma;
   vestibulocochlear nerve; waterjet dissection
ID SCHWANNOMAS ACOUSTIC NEUROMAS; FACIAL-NERVE; VESTIBULAR SCHWANNOMAS;
   HYDRO-JET; RETROSIGMOID APPROACH; AUDITORY-NERVE; IN-VIVO; HEARING
   PRESERVATION; LIVER RESECTION; COCHLEAR NERVE
AB Object. Waterjet dissection has been shown to protect intracerebral vessels, but no experience exists in applying this modality to the cranial nerves. To evaluate its potential, the authors examined waterjet dissection of the vestibulocochlear nerve in rats.
   Methods. Lateral suboccipital craniectomy and microsurgical preparation of the vestibulocochlear nerve were performed in 42 rats. Water pressures of 2-10 bar were applied, and the effect was microscopically evaluated. Auditory brainstem responses (ABRs) were used to define nerve function compared with preoperative values and the healthy contralateral side. The final anatomical preparation documented the morphological and histological effects of waterjet pressure on the nerve.
   Results. In using LIP to 6 bar, the cochlear nerve was preserved in all cases. Eight bar moderately damaged the nerve Surface. A 10-bar jet markedly damaged or even completely dissected the nerve. Time course analysis of the ABR demonstrated complete functional nerve preservation up to 6 bar after 6 weeks in all rats. Waterjet dissection with 8 bar was associated with a 60% recovery of ABR. In the 10-bar group, no recovery was seen.
   Conclusions. Microsurgical dissection of cranial nerves is possible using waterjet dissection while preserving both morphology and function. The aforementioned jet pressures are known to be effective in neurosurgical treatment of tumors. Thus, waterjet dissection may be useful in skull base surgery including dissection of cranial nerves from tumors. Further Studies on this subject are encouraged. (DOI: 10.3171/2008.5.17561)
C1 [Tschan, Christoph; Krauss, Joachim Kurt] Hannover Med Sch, Dept Neurosurg, D-3000 Hannover, Germany.
   [Gaab, Michael Robert; Oertel, Joachim] Hannover Nordstadt Hosp, Dept Neurosurg, Hannover, Germany.
C3 Hannover Medical School
RP Oertel, J (通讯作者)，Johannes Gutenberg Univ Mainz, Dept Neurosurg Klinikum, Langenbeckstr 1, D-55101 Mainz, Germany.
EM
RI Oertel, Joachim/AAO-5012-2021
CR Ciric I, 2005, NEUROSURGERY, V56, P560, DOI 10.1227/01.NEU.0000154059.34990.B8
   Corvin S, 2005, J ENDOUROL, V19, P823, DOI 10.1089/end.2005.19.823
   DARNER C, 2004, OTOL NEUROTOL, V25, P826
   DAVIS RL, 1992, J EXP ZOOL, V261, P254, DOI 10.1002/jez.1402610305
   Gharabaghi A, 2005, ACTA NEUROCHIR SUPPL, V93, P89
   Gordon E, 1998, J REFRACT SURG, V14, P338
   Hatayama T, 1999, NEUROL RES, V21, P599, DOI 10.1080/01616412.1999.11740983
   Honl M, 2001, Z ORTHOP GRENZGEB, V139, P45, DOI 10.1055/s-2001-11870
   Honl M, 2000, BIOMED TECH, V45, P222, DOI 10.1515/bmte.2000.45.9.222
   Hubert J, 1996, UROL RES, V24, P355, DOI 10.1007/BF00389793
   Jackson LE, 2000, AM J OTOL, V21, P249, DOI 10.1016/S0196-0709(00)80018-6
   Jakob S, 2000, ZBL NEUROCHIR, V61, P14, DOI 10.1055/s-2000-8260
   Jung S, 2000, SURG NEUROL, V53, P370, DOI 10.1016/S0090-3019(00)00196-8
   Kaduk WMH, 1999, J CRANIO MAXILL SURG, V27, P327, DOI 10.1054/jcms.1999.0076
   Kockerling F., 2004, TECH COLOPROCTOL, V8, P217, DOI DOI 10.1007/S10151-004-0163-4
   Kuhlmann C, 2005, BIOMED TECH, V50, P337, DOI 10.1515/BMT.2005.047
   Lesser T, 2000, CHIRURG, V71, P592
   Lipshitz I, 1996, J REFRACT SURG, V12, P184
   Lipshitz I, 1998, J REFRACT SURG, V14, P667
   Mayer TE, 2005, AM J NEURORADIOL, V26, P1389
   Mom T, 2002, NEUROCHIRURGIE, V48, P387
   Oertel J, 2004, MINIM INVAS NEUROSUR, V47, P154, DOI 10.1055/s-2004-818512
   Oertel J, 2004, ACTA NEUROCHIR, V146, P713, DOI 10.1007/s00701-004-0265-9
   Oertel J, 2004, J NEUROSURG, V100, P498, DOI 10.3171/jns.2004.100.3.0498
   Oertel J, 2003, NEUROSURG REV, V26, P168, DOI 10.1007/s10143-002-0244-7
   Oertel J, 2003, EUR J SURG ONCOL, V29, P407, DOI 10.1053/ejso.2002.1428
   Oertel J, 2003, NEUROSURGERY, V52, P153, DOI 10.1097/00006123-200301000-00020
   Oertel Joachim, 2005, Neurosurgery, V56, P142, DOI 10.1227/01.NEU.0000144316.87764.13
   Oertel J, 2006, J NEUROSURG, V105, P928, DOI 10.3171/jns.2006.105.6.928
   PAPACHRISTOU DN, 1982, BRIT J SURG, V69, P93, DOI 10.1002/bjs.1800690212
   Penchev RD, 1999, J UROLOGY, V162, P772, DOI 10.1097/00005392-199909010-00040
   Piek J, 1998, J NEUROSURG, V89, P861, DOI 10.3171/jns.1998.89.5.0861
   Piek J, 2002, J NEUROSURG, V96, P690, DOI 10.3171/jns.2002.96.4.0690
   Rau H G, 1995, Eur J Surg Oncol, V21, P183, DOI 10.1016/S0748-7983(95)90435-2
   RAU HG, 1995, SURG ENDOSC-ULTRAS, V9, P1009
   Rosahl SK, 2000, ACTA NEUROCHIR, V142, P1037, DOI 10.1007/s007010070060
   Samii M, 1997, NEUROSURGERY, V40, P696, DOI 10.1097/00006123-199704000-00007
   Samii M, 1997, NEUROSURGERY, V40, P684, DOI 10.1097/00006123-199704000-00006
   Samii M, 2006, J NEUROSURG, V105, P527, DOI 10.3171/jns.2006.105.4.527
   Sampath P, 2000, J NEUROSURG, V92, P70, DOI 10.3171/jns.2000.92.1.0070
   Schmolke S, 2004, BIOMED TECH, V49, P18, DOI 10.1515/BMT.2004.004
   SEKIYA T, 1990, ACTA NEUROCHIR, V102, P108, DOI 10.1007/BF01405423
   Sekiya T, 2002, HEARING RES, V173, P91, DOI 10.1016/S0378-5955(02)00633-0
   Seol HJ, 2006, NEUROL MED-CHIR, V46, P176, DOI 10.2176/nmc.46.176
   Shekarriz B, 2005, EXPERT REV MED DEVIC, V2, P287, DOI 10.1586/17434440.2.3.287
   Shekarriz B, 2004, J ENDOUROL, V18, P273, DOI 10.1089/089277904773582895
   Siegert R, 2000, LARYNGO RHINO OTOL, V79, P780, DOI 10.1055/s-2000-9140
   Spahn G, 2006, ARCH ORTHOP TRAUM SU, V126, P223, DOI 10.1007/s00402-005-0002-8
   Strauss C, 2006, NEUROSURGERY, V59, P577, DOI 10.1227/01.NEU.0000230260.95477.0A
   Tatagiba M, 2002, ACTA NEUROCHIR, V144, P181, DOI 10.1007/s007010200022
   Terzis A J, 1989, Br J Neurosurg, V3, P361, DOI 10.3109/02688698909002816
   Une Y, 1996, INT SURG, V81, P45
   Wilhelm F, 1998, OPHTHALMOLOGE, V95, P721, DOI 10.1007/s003470050342
   Wilhelm FH, 1999, OPHTHALMOLOGE, V96, P640, DOI 10.1007/s003470050466
   Wu YW, 2006, LANGENBECK ARCH SURG, V391, P355, DOI 10.1007/s00423-006-0058-0
   Yu Li-Mei, 2006, Zhonghua Er Bi Yan Hou Tou Jing Wai Ke Za Zhi, V41, P26
NR 57
TC 15
Z9 15
U1 0
U2 3
PU AMER ASSOC NEUROLOGICAL SURGEONS
PI ROLLING MEADOWS
PA 5550 MEADOWBROOK DRIVE, ROLLING MEADOWS, IL 60008 USA
SN 0022-3085
EI 1933-0693
J9 J NEUROSURG
JI J. Neurosurg.
PD APR
PY 2009
VL 110
IS 4
BP 656
EP 661
DI 10.3171/2008.5.17561
PG 6
WC Clinical Neurology; Surgery
WE Science Citation Index Expanded (SCI-EXPANDED)
SC Neurosciences & Neurology; Surgery
GA 424UR
UT WOS:000264594300009
PM 18834266
DA 2023-08-27
ER

PT J
AU Chang, JW
   Chang, JH
   Park, YG
   Chung, SS
AF Chang, JW
   Chang, JH
   Park, YG
   Chung, SS
TI Gamma knife radiosurgery for idiopathic and secondary trigeminal
   neuralgia
SO JOURNAL OF NEUROSURGERY
LA English
DT Article; Proceedings Paper
CT 10th International-Leksell-Gamma-Knife-Society Meeting
CY APR 13-17, 2000
CL LAKE TAHOE, CA
SP Int Leksell Gamma Knife Soc
DE gamma knife; radiosurgery; trigeminal neuralgia
ID STEREOTACTIC RADIOSURGERY
AB Object. The aim of this study was to identify pain relief and treatment morbidity following gamma knife radiosurgery (GKS) for idiopathic and secondary trigeminal neuralgia.
   Methods. Between May 1992 and December 1999, 15 patients with idiopathic trigeminal neuralgia and 38 patients with secondary trigeminal neuralgia were treated with GKS. Pain improvement was achieved in 13 of the patients with idiopathic pain (pain response rate 86.7%). Seven patients were pain free and another six experienced pain reduction. There were no serious complications; however, two patients suffered a mild facial sensory change 8 months and 9 months, respectively, after GKS.
   The patients with secondary trigeminal neuralgia were divided into two groups (Group I, 32 patients in whom the trigeminal root entry zone [REZ] near the tumor could not be visualized; and Group II, six patients in whom the trigeminal REZ near the tumor or brainstem lesion could be visualized).
   In Group I, the pain subsided completely in eight patients and was reduced in seven (pain response rate 46.9%). In Group II, the pain subsided completely in one patient at 2.8 months and was reduced in three patients at a mean follow up of 0.8 months (range 0.6-1 month) after GKS. The pain response rate was 66.7%.
   Conclusions. The authors believe that GKS is an effective treatment modalities for idiopathic and secondary trigeminal neuralgia, particularly in patients with inoperable lesions.
C1 Yonsei Univ, Coll Med, Dept Neurosurg, Seoul, South Korea.
C3 Yonsei University; Yonsei University Health System
RP Park, YG (通讯作者)，Yonsei Univ, Coll Med, Dept Neurosurg, CPO Box 8044, Seoul, South Korea.
EM
OI Chang, Jong Hee/0000-0003-1509-9800; Chang, Jin Woo/0000-0002-2717-0101
CR Barker FG, 1996, J NEUROSURG, V84, P818, DOI 10.3171/jns.1996.84.5.0818
   BULLITT E, 1986, J NEUROSURG, V64, P865, DOI 10.3171/jns.1986.64.6.0865
   Chang JW, 1999, STEREOT FUNCT NEUROS, V72, P29, DOI 10.1159/000056436
   CHENG TMW, 1993, NEUROLOGY, V43, P2298, DOI 10.1212/WNL.43.11.2298
   Kondziolka D, 1996, J NEUROSURG, V84, P940, DOI 10.3171/jns.1996.84.6.0940
   Kondziolka D, 1998, PROG NEUROL, V14, P212
   LEKSELL L, 1951, ACTA CHIR SCAND, V102, P316
   Lovely TJ, 1997, NEUROSURG CLIN N AM, V8, P11
   MANERA JRL, 1995, STEREOTACT FUNCT S1, V64, P182
   Marco JKD, 1997, NEUROSURG CLIN N AM, V8, P103
   Regis J, 1999, NEUROSURG CLIN N AM, V10, P359, DOI 10.1016/S1042-3680(18)30201-8
   Regis J, 1997, STEREOTACT FUNCT S1, V64, P210
   Urgosik D, 1998, STEREOT FUNCT NEUROS, V70, P200, DOI 10.1159/000056423
   Young RF, 1997, NEUROLOGY, V48, P608, DOI 10.1212/WNL.48.3.608
NR 14
TC 46
Z9 46
U1 0
U2 0
PU AMER ASSOC NEUROLOGICAL SURGEONS
PI ROLLING MEADOWS
PA 5550 MEADOWBROOK DRIVE, ROLLING MEADOWS, IL 60008 USA
SN 0022-3085
EI 1933-0693
J9 J NEUROSURG
JI J. Neurosurg.
PD DEC
PY 2000
VL 93
SU 3
BP 147
EP 151
DI 10.3171/jns.2000.93.supplement_3.0147
PG 5
WC Clinical Neurology; Surgery
WE Science Citation Index Expanded (SCI-EXPANDED); Conference Proceedings Citation Index - Science (CPCI-S)
SC Neurosciences & Neurology; Surgery
GA 380XZ
UT WOS:000165730500029
PM 11143233
DA 2023-08-27
ER

PT J
AU Vivinus-Nebot, M
   Rousselle, P
   Breittmayer, JP
   Cenciarini, C
   Berrih-Aknin, S
   Spong, S
   Nokelainen, P
   Cottrez, F
   Marinkovich, MP
   Bernard, A
AF Vivinus-Nebot, M
   Rousselle, P
   Breittmayer, JP
   Cenciarini, C
   Berrih-Aknin, S
   Spong, S
   Nokelainen, P
   Cottrez, F
   Marinkovich, MP
   Bernard, A
TI Mature human thymocytes migrate on laminin-5 with activation of
   metalloproteinase-14 and cleavage of CD44
SO JOURNAL OF IMMUNOLOGY
LA English
DT Article
ID MEMBRANE-TYPE-1 MATRIX-METALLOPROTEINASE; TUMOR-CELL MIGRATION;
   BASEMENT-MEMBRANES; EPITHELIAL-CELLS; THYMIC EMIGRANTS; SCATTER FACTOR;
   GAMMA-2 CHAIN; T-LYMPHOCYTES; IN-VIVO; SURFACE
AB We have previously shown that laminin-5 is expressed in the human thymic medulla, in which mature thymocytes are located. We now report that laminin-5 promotes migration of mature medullary thymocytes, whereas it has no effect on cortical immature thymocytes. Migration was inhibited by blocking mAbs directed against laminin-5 integrin receptors and by inhibitors of metalloproteinases. Interactions of thymocytes with laminin-5 induced a strong up-regulation of active metalloproteinase-14. However, we found that thymocytes did not cleave the laminin-5 gamma(2) chain, suggesting that they do not use the same pathway as epithelial cells to migrate on laminin-5. Interactions of thymocytes with laminin-5 also induced the release of a soluble fragment of CD44 cell surface molecule. Moreover, CD44-rich supernatants induced thymocyte migration in contrast with supernatants depleted in CD44 by immunoadsorption. CD44 cleavage was recently reported to be due to metalloproteinase-14 activation and led to increased migration in cancer cells. Thus, in this study, we show that laminin-5 promotes human mature thymocyte migration in vitro via a multimolecular mechanism involving laminin-5 integrin receptors, metalloproteinase-14 and CD44. These data suggest that, in vivo, laminin-5 may function in the migration of mature thymocytes within the medulla and be part of the thymic emigration process.
C1 Hop Archet 1, INSERM, U576, F-06202 Nice, France.
   Ctr Hosp Univ, Immunol Lab, Nice, France.
   IBCP, UMR 5086, Lyon, France.
   CNRS, Le Plessis Robinson, France.
   FibroGen, San Francisco, CA 94080 USA.
   Stanford Univ, Sch Med, Program Epithelial Biol, Stanford, CA 94305 USA.
C3 CHU Nice; Institut National de la Sante et de la Recherche Medicale
   (Inserm); UDICE-French Research Universities; Universite Cote d'Azur;
   CHU Nice; Centre National de la Recherche Scientifique (CNRS); CNRS -
   National Institute for Biology (INSB); UDICE-French Research
   Universities; Universite Claude Bernard Lyon 1; Centre National de la
   Recherche Scientifique (CNRS); FibroGen; Stanford University
RP Bernard, A (通讯作者)，Hop Archet 1, INSERM, U576, Route St Antoine de Ginestiere,BP3079, F-06202 Nice, France.
EM
RI Rousselle, Patricia/J-5290-2019; Rousselle, Patricia/CAF-3915-2022;
   Berrih‐Aknin, Sonia/F-8025-2013
OI Rousselle, Patricia/0000-0003-0275-4562; Rousselle,
   Patricia/0000-0003-0275-4562; Berrih‐Aknin, Sonia/0000-0001-9199-4175
CR Almeida ARM, 2001, J EXP MED, V194, P591, DOI 10.1084/jem.194.5.591
   Amano S, 2000, J BIOL CHEM, V275, P22728, DOI 10.1074/jbc.M002345200
   BAZIL V, 1994, J IMMUNOL, V152, P1314
   BENDELAC A, 1992, J EXP MED, V175, P731, DOI 10.1084/jem.175.3.731
   Berzins SP, 1999, P NATL ACAD SCI USA, V96, P9787, DOI 10.1073/pnas.96.17.9787
   Berzins SP, 1998, J EXP MED, V187, P1839, DOI 10.1084/jem.187.11.1839
   BURGESON RE, 1994, MATRIX BIOL, V14, P209, DOI 10.1016/0945-053X(94)90184-8
   Butler GS, 1998, J BIOL CHEM, V273, P871, DOI 10.1074/jbc.273.2.871
   CARTER WG, 1990, J CELL BIOL, V111, P3141, DOI 10.1083/jcb.111.6.3141
   DUNON D, 1993, BLOOD, V81, P1
   Gabor MJ, 1997, EUR J IMMUNOL, V27, P2010, DOI 10.1002/eji.1830270827
   Giannelli G, 1997, SCIENCE, V277, P225, DOI 10.1126/science.277.5323.225
   Henke CA, 1996, J CLIN INVEST, V97, P2541, DOI 10.1172/JCI118702
   Hotary K, 2000, J CELL BIOL, V149, P1309, DOI 10.1083/jcb.149.6.1309
   Itoh Y, 1998, J BIOL CHEM, V273, P24360, DOI 10.1074/jbc.273.38.24360
   Itoh Y, 2001, EMBO J, V20, P4782, DOI 10.1093/emboj/20.17.4782
   Kajita M, 2001, J CELL BIOL, V153, P893, DOI 10.1083/jcb.153.5.893
   KALLUNKI P, 1992, J CELL BIOL, V119, P679, DOI 10.1083/jcb.119.3.679
   KENDALL MD, 1989, THYMUS, V13, P157
   KIKKAWA Y, 1994, J BIOCHEM-TOKYO, V116, P862, DOI 10.1093/oxfordjournals.jbchem.a124608
   Kim MG, 2000, J IMMUNOL, V165, P192, DOI 10.4049/jimmunol.165.1.192
   Koshikawa N, 2000, J CELL BIOL, V148, P615, DOI 10.1083/jcb.148.3.615
   Kutlesa S, 2002, IMMUNOLOGY, V105, P407, DOI 10.1046/j.1365-2567.2002.01384.x
   Lind EF, 2001, J EXP MED, V194, P127, DOI 10.1084/jem.194.2.127
   Madri JA, 1996, BIOCHEM CELL BIOL, V74, P749, DOI 10.1139/o96-082
   MARINKOVICH MP, 1992, J BIOL CHEM, V267, P17900
   MARINKOVICH MP, 1992, J CELL BIOL, V119, P695, DOI 10.1083/jcb.119.3.695
   MATSUYAMA T, 1989, J EXP MED, V170, P1133, DOI 10.1084/jem.170.4.1133
   MIYAZAKI K, 1993, P NATL ACAD SCI USA, V90, P11767, DOI 10.1073/pnas.90.24.11767
   MOEBIUS U, 1989, LEUCOCYTE TYPING, V4, P342
   MOEBIUS U, 1989, LEUKOCYTE TYPING, V4, P314
   MOJCIK CF, 1995, BLOOD, V86, P4206, DOI 10.1182/blood.V86.11.4206.bloodjournal86114206
   Mori H, 2002, EMBO J, V21, P3949, DOI 10.1093/emboj/cdf411
   MORODOMI T, 1992, BIOCHEM J, V285, P603, DOI 10.1042/bj2850603
   Mu DZ, 2002, J CELL BIOL, V157, P493, DOI 10.1083/jcb.200109100
   Murphy G, 1999, CURR OPIN CELL BIOL, V11, P614, DOI 10.1016/S0955-0674(99)00022-8
   Nagase H, 1999, J BIOL CHEM, V274, P21491, DOI 10.1074/jbc.274.31.21491
   NAOT D, 1997, ADV CANCER RES, V71, P241
   Okamoto I, 2001, J CELL BIOL, V155, P755, DOI 10.1083/jcb.200108159
   Okamoto I, 1999, ONCOGENE, V18, P1435, DOI 10.1038/sj.onc.1202447
   PATEL DD, 1995, J CLIN IMMUNOL, V15, P80, DOI 10.1007/BF01541736
   PATEL DD, 1995, LEUKOCYTE TYPING 5, V2, P1725
   Pattaramalai S, 1996, EXP CELL RES, V222, P281, DOI 10.1006/excr.1996.0036
   Petrie HT, 2002, IMMUNOL REV, V189, P8, DOI 10.1034/j.1600-065X.2002.18902.x
   Poulin JF, 1999, J EXP MED, V190, P479, DOI 10.1084/jem.190.4.479
   Poznansky MC, 2002, J CLIN INVEST, V109, P1101, DOI 10.1172/JCI200213853
   ROUSSELLE P, 1994, J CELL BIOL, V125, P205, DOI 10.1083/jcb.125.1.205
   ROUSSELLE P, 1991, J CELL BIOL, V114, P567, DOI 10.1083/jcb.114.3.567
   Ryan MC, 1996, MATRIX BIOL, V15, P369, DOI 10.1016/S0945-053X(96)90157-2
   Sasaki T, 2001, J MOL BIOL, V314, P751, DOI 10.1006/jmbi.2001.5176
   SATO H, 1994, NATURE, V370, P61, DOI 10.1038/370061a0
   Sato H, 1996, FEBS LETT, V385, P238, DOI 10.1016/0014-5793(96)00389-4
   SAVAGNER P, 1986, J CELL BIOL, V103, P2715, DOI 10.1083/jcb.103.6.2715
   Savino W, 2002, TRENDS IMMUNOL, V23, P305, DOI 10.1016/S1471-4906(02)02224-X
   SCOLLAY R, 1995, IMMUNOL TODAY, V16, P268, DOI 10.1016/0167-5699(95)80179-0
   SONNENBERG A, 1988, NATURE, V336, P487, DOI 10.1038/336487a0
   STAUDER R, 1995, LEUKOCYTE TYPING 5, V2, P1719
   SYMINGTON BE, 1995, J CELL SCI, V108, P831
   Takahashi K, 1999, INT J CANCER, V80, P387, DOI 10.1002/(SICI)1097-0215(19990129)80:3<387::AID-IJC9>3.3.CO;2-K
   Tanchot C, 1997, J EXP MED, V186, P1099, DOI 10.1084/jem.186.7.1099
   THOMAS L, 1992, J CELL BIOL, V118, P971, DOI 10.1083/jcb.118.4.971
   Ueno T, 2002, IMMUNITY, V16, P205, DOI 10.1016/S1074-7613(02)00267-4
   Ushiki T, 1997, ARCH HISTOL CYTOL, V60, P89, DOI 10.1679/aohc.60.89
   VANEWIJK W, 1991, ANNU REV IMMUNOL, V9, P591, DOI 10.1146/annurev.iy.09.040191.003111
   Veitch DP, 2003, J BIOL CHEM, V278, P15661, DOI 10.1074/jbc.M210588200
   VIEIRA JL, 1994, IMMUNOL TODAY, V15, P192
   Virtanen I, 1996, HISTOCHEM J, V28, P643, DOI 10.1007/BF02331385
   Vivinus-Nebot M, 1999, J CELL BIOL, V144, P563, DOI 10.1083/jcb.144.3.563
   Wakkach A, 2003, IMMUNITY, V18, P605, DOI 10.1016/S1074-7613(03)00113-4
   WAYNER EA, 1993, J CELL BIOL, V121, P1141, DOI 10.1083/jcb.121.5.1141
   Werb Z, 1997, CELL, V91, P439, DOI 10.1016/S0092-8674(00)80429-8
   Wilkinson B, 1999, J IMMUNOL, V162, P3873
   Will H, 1996, J BIOL CHEM, V271, P17119, DOI 10.1074/jbc.271.29.17119
   WU L, 1998, MOL BIOL B CELL T CE, P235
   Zhang K, 1996, EXP CELL RES, V227, P309, DOI 10.1006/excr.1996.0280
NR 75
TC 34
Z9 35
U1 0
U2 2
PU AMER ASSOC IMMUNOLOGISTS
PI BETHESDA
PA 9650 ROCKVILLE PIKE, BETHESDA, MD 20814 USA
SN 0022-1767
EI 1550-6606
J9 J IMMUNOL
JI J. Immunol.
PD FEB 1
PY 2004
VL 172
IS 3
BP 1397
EP 1406
DI 10.4049/jimmunol.172.3.1397
PG 10
WC Immunology
WE Science Citation Index Expanded (SCI-EXPANDED)
SC Immunology
GA 766LW
UT WOS:000188378700009
PM 14734715
DA 2023-08-27
ER

PT J
AU Peng, S
   Zhang, J
   Tan, XT
   Huang, YQ
   Xu, J
   Silk, N
   Zhang, DZ
   Liu, QL
   Jiang, J
AF Peng, Song
   Zhang, Jun
   Tan, Xintao
   Huang, Yiqiang
   Xu, Jing
   Silk, Natalie
   Zhang, Dianzheng
   Liu, Qiuli
   Jiang, Jun
TI The VHL/HIF Axis in the Development and Treatment of
   Pheochromocytoma/Paraganglioma
SO FRONTIERS IN ENDOCRINOLOGY
LA English
DT Review
DE pheochromocytomas; paragangliomas; VHL; HIF; metabolism; inhibitor
ID INDUCIBLE FACTOR-I; HIPPEL-LINDAU DISEASE; ENDOTHELIAL GROWTH-FACTOR;
   TUMOR-SUPPRESSOR PROTEIN; CANCER-CELL METABOLISM; GERMLINE MUTATIONS;
   HIF2A MUTATIONS; MALIGNANT PHEOCHROMOCYTOMA; PRECLINICAL MODELS;
   MAMMALIAN TARGET
AB Pheochromocytomas and paragangliomas (PPGLs) are rare neuroendocrine tumors originating from chromaffin cells in the adrenal medulla (PCCs) or extra-adrenal sympathetic or parasympathetic paraganglia (PGLs). About 40% of PPGLs result from germline mutations and therefore they are highly inheritable. Although dysfunction of any one of a panel of more than 20 genes can lead to PPGLs, mutations in genes involved in the VHL/HIF axis including PHD, VHL, HIF-2A (EPAS1), and SDHx are more frequently found in PPGLs. Multiple lines of evidence indicate that pseudohypoxia plays a crucial role in the tumorigenesis of PPGLs, and therefore PPGLs are also known as metabolic diseases. However, the interplay between VHL/HIF-mediated pseudohypoxia and metabolic disorder in PPGLs cells is not well-defined. In this review, we will first discuss the VHL/HIF axis and genetic alterations in this axis. Then, we will dissect the underlying mechanisms in VHL/HIF axis-driven PPGL pathogenesis, with special attention paid to the interplay between the VHL/HIF axis and cancer cell metabolism. Finally, we will summarize the currently available compounds/drugs targeting this axis which could be potentially used as PPGLs treatment, as well as their underlying pharmacological mechanisms. The overall goal of this review is to better understand the role of VHL/HIF axis in PPGLs development, to establish more accurate tools in PPGLs diagnosis, and to pave the road toward efficacious therapeutics against metastatic PPGLs.
C1 [Peng, Song; Zhang, Jun; Tan, Xintao; Huang, Yiqiang; Xu, Jing; Liu, Qiuli; Jiang, Jun] Army Med Univ, Daping Hosp, Dept Urol, Chongqing, Peoples R China.
   [Silk, Natalie; Zhang, Dianzheng] Philadelphia Coll Osteopath Med, Dept Biomed Sci, Philadelphia, PA USA.
C3 Army Medical University
RP Liu, QL; Jiang, J (通讯作者)，Army Med Univ, Daping Hosp, Dept Urol, Chongqing, Peoples R China.
EM
RI Liu, Qiuli/ABA-8363-2021
OI Liu, Qiuli/0000-0002-9532-5654
FU National Natural Science Foundation of China [81972398]; University
   Research Project of Army Medical University [218XLC3073]
FX This work was supported by the National Natural Science Foundation of
   China (81972398, JJ) and University Research Project of Army Medical
   University (218XLC3073, JZ).
CR Aita Y, 2012, AM J PHYSIOL-ENDOC M, V303, pE1006, DOI 10.1152/ajpendo.00156.2012
   Ajith Thekkuttuparambil A., 2019, Journal of Basic and Clinical Physiology and Pharmacology, V30, P11, DOI 10.1515/jbcpp-2017-0167
   Al Tameemi W, 2019, FRONT CELL DEV BIOL, V7, DOI 10.3389/fcell.2019.00004
   Toledo RA, 2017, ENDOCR-RELAT CANCER, V24, pC9, DOI 10.1530/ERC-16-0479
   Andreasson A, 2013, EPIGENETICS-US, V8, P1347, DOI 10.4161/epi.26686
   Ang SO, 2002, NAT GENET, V32, P614, DOI 10.1038/ng1019
   Ariton M, 2000, Endocr Pract, V6, P249
   Ayala-Ramirez M, 2012, J CLIN ENDOCR METAB, V97, P4040, DOI 10.1210/jc.2012-2356
   Ayala-Ramirez M, 2011, J CLIN ENDOCR METAB, V96, P717, DOI 10.1210/jc.2010-1946
   Baba M, 2003, ONCOGENE, V22, P2728, DOI 10.1038/sj.onc.1206373
   Baek JH, 2005, MOL CELL, V17, P503, DOI 10.1016/j.molcel.2005.01.011
   Baysal BE, 2000, SCIENCE, V287, P848, DOI 10.1126/science.287.5454.848
   BEARD CM, 1983, MAYO CLIN PROC, V58, P802
   Beroud C, 1998, NUCLEIC ACIDS RES, V26, P256, DOI 10.1093/nar/26.1.256
   Bertout JA, 2008, NAT REV CANCER, V8, P967, DOI 10.1038/nrc2540
   Bertozzi D, 2014, MOL CANCER THER, V13, P239, DOI 10.1158/1535-7163.MCT-13-0729
   Bishop T, 2004, PLOS BIOL, V2, P1549, DOI 10.1371/journal.pbio.0020289
   Bohonowych JES, 2011, BMC CANCER, V11, DOI 10.1186/1471-2407-11-520
   Bommi-Reddy A, 2008, P NATL ACAD SCI USA, V105, P16484, DOI 10.1073/pnas.0806574105
   Bruick RK, 2001, SCIENCE, V294, P1337, DOI 10.1126/science.1066373
   Buffet A, 2014, J CLIN ENDOCR METAB, V99, pE369, DOI 10.1210/jc.2013-2600
   Burnichon N, 2010, HUM MOL GENET, V19, P3011, DOI 10.1093/hmg/ddq206
   Calsina B, 2019, THERANOSTICS, V9, P4946, DOI 10.7150/thno.35458
   Calsina B, 2018, GENET MED, V20, P1652, DOI 10.1038/s41436-018-0068-7
   Castro-Vega LJ, 2014, HUM MOL GENET, V23, P2440, DOI 10.1093/hmg/ddt639
   Chen WF, 2016, NATURE, V539, P112, DOI 10.1038/nature19796
   Chittiboina Prashant, 2015, Handb Clin Neurol, V132, P139, DOI 10.1016/B978-0-444-62702-5.00010-X
   Cho H, 2016, NATURE, V539, P107, DOI 10.1038/nature19795
   Choueiri TK, 2019, 14 EUR INT KIDN CANC, DOI [10.1093/annonc/mdz249.010, DOI 10.1093/ANNONC/MDZ249.010]
   Clifford SC, 2001, HUM MOL GENET, V10, P1029, DOI 10.1093/hmg/10.10.1029
   Coltella N, 2015, CLIN CANCER RES, V21, P3685, DOI 10.1158/1078-0432.CCR-14-3022
   Comino-Mendez I, 2013, HUM MOL GENET, V22, P2169, DOI 10.1093/hmg/ddt069
   Courtney KD, 2018, J CLIN ONCOL, V36, P867, DOI 10.1200/JCO.2017.74.2627
   CROSSEY PA, 1994, HUM GENET, V93, P53
   Dahia PLM, 2014, NAT REV CANCER, V14, P108, DOI 10.1038/nrc3648
   Dang L, 2009, NATURE, V462, P739, DOI 10.1038/nature08617
   Dayan F, 2006, CANCER RES, V66, P3688, DOI 10.1158/0008-5472.CAN-05-4564
   De Bels D, 2011, NEW ENGL J MED, V365, P1845, DOI 10.1056/NEJMc1110602
   Denorme M, 2014, CANCER LETT, V352, P236, DOI 10.1016/j.canlet.2014.07.005
   Eisenhofer G, 2011, ENDOCR-RELAT CANCER, V18, P97, DOI 10.1677/ERC-10-0211
   Erbel PJA, 2003, P NATL ACAD SCI USA, V100, P15504, DOI 10.1073/pnas.2533374100
   Erlic Z, 2019, EXP CLIN ENDOCR DIAB, V127, P129, DOI 10.1055/a-0649-0960
   Facchin C, 2020, THERANOSTICS, V10, P3518, DOI 10.7150/thno.40687
   Faivre S, 2007, NAT REV DRUG DISCOV, V6, P734, DOI 10.1038/nrd2380
   Favier J, 2012, NEW ENGL J MED, V367, P2161, DOI 10.1056/NEJMc1211953
   Favier J, 2009, PLOS ONE, V4, DOI 10.1371/journal.pone.0007094
   Fishbein L, 2017, CANCER CELL, V31, P181, DOI 10.1016/j.ccell.2017.01.001
   Flippot R, 2019, NAT REV UROL, V16, P484, DOI 10.1038/s41585-019-0195-1
   Fu BB, 2007, MOL CANCER THER, V6, P220, DOI 10.1158/1535-7163.MCT-06-0526
   Fukino K, 2000, J HUM GENET, V45, P47, DOI 10.1007/s100380050009
   Gallou C, 2004, HUM MUTAT, V24, P215, DOI 10.1002/humu.20082
   Garcia A, 1997, CLIN ENDOCRINOL, V46, P359, DOI 10.1046/j.1365-2265.1997.00149.x
   Gladek I, 2017, GENE CHROMOSOME CANC, V56, P439, DOI 10.1002/gcc.22449
   Gordan JD, 2007, CANCER CELL, V12, P108, DOI 10.1016/j.ccr.2007.07.006
   Gossage L, 2015, NAT REV CANCER, V15, P55, DOI 10.1038/nrc3844
   Gottlieb E, 2005, NAT REV CANCER, V5, P857, DOI 10.1038/nrc1737
   Greenberger LM, 2008, MOL CANCER THER, V7, P3598, DOI 10.1158/1535-7163.MCT-08-0510
   Gross-Goupil M, 2018, ANN ONCOL, V29, P2371, DOI 10.1093/annonc/mdy454
   Gunaldi M, 2014, CANCER RES TREAT, V46, P411, DOI 10.4143/crt.2013.093
   Heiden MGV, 2009, SCIENCE, V324, P1029, DOI 10.1126/science.1160809
   Heikkila M, 2011, CELL MOL LIFE SCI, V68, P3885, DOI 10.1007/s00018-011-0679-5
   Hong BA, 2019, FRONT GENET, V10, DOI 10.3389/fgene.2019.00867
   Horita N, 2015, SCI REP-UK, V5, DOI 10.1038/srep15437
   Hu CJ, 2003, MOL CELL BIOL, V23, P9361, DOI 10.1128/MCB.23.24.9361-9374.2003
   Hu N, 2013, J CELL SCI, V126, P532, DOI 10.1242/jcs.114231
   Huang X, 2014, ACTA BIOCH BIOPH SIN, V46, P220, DOI 10.1093/abbs/gmt141
   Hudson CC, 2002, MOL CELL BIOL, V22, P7004, DOI 10.1128/MCB.22.20.7004-7014.2002
   Hutt DM, 2014, PLOS ONE, V9, DOI 10.1371/journal.pone.0106224
   Ibrahim NO, 2005, CANCER RES, V65, P11094, DOI 10.1158/0008-5472.CAN-05-1877
   Icard P, 2012, BBA-REV CANCER, V1826, P423, DOI 10.1016/j.bbcan.2012.07.001
   Ivan M, 2002, P NATL ACAD SCI USA, V99, P13459, DOI 10.1073/pnas.192342099
   Jaakkola P, 2001, SCIENCE, V292, P468, DOI 10.1126/science.1059796
   Jang M, 2013, EXP MOL MED, V45, DOI 10.1038/emm.2013.85
   Jasim S, 2017, ENDOCRINE, V57, P220, DOI 10.1007/s12020-017-1359-5
   Jeong W, 2014, CANCER CHEMOTH PHARM, V73, P343, DOI 10.1007/s00280-013-2362-z
   Jimenez C, 2020, ENDOCR-RELAT CANCER, V27, pE239, DOI 10.1530/ERC-20-0043
   Jimenez C, 2009, J CLIN ENDOCR METAB, V94, P386, DOI 10.1210/jc.2008-1972
   Jochmanova I, 2015, HORM CANCER-US, V6, P191, DOI 10.1007/s12672-015-0231-4
   Jochmanova I, 2013, JNCI-J NATL CANCER I, V105, P1270, DOI 10.1093/jnci/djt201
   Joosten SC, 2018, NAT REV UROL, V15, P430, DOI 10.1038/s41585-018-0023-z
   Jordan BF, 2005, CLIN CANCER RES, V11, P529
   Joshi S, 2014, MOL CANCER RES, V12, P1520, DOI 10.1158/1541-7786.MCR-13-0682
   Joshua AM, 2009, J CLIN ENDOCR METAB, V94, P5, DOI 10.1210/jc.2008-1836
   Kaelin WG, 2008, NAT REV CANCER, V8, P865, DOI 10.1038/nrc2502
   Kaelin WG, 2008, MOL CELL, V30, P393, DOI 10.1016/j.molcel.2008.04.009
   Keith B, 2012, NAT REV CANCER, V12, P9, DOI 10.1038/nrc3183
   Kim HC, 2013, CANCER RES TREAT, V45, P349, DOI 10.4143/crt.2013.45.4.349
   Kim YH, 2011, CANCER CHEMOTH PHARM, V68, P405, DOI 10.1007/s00280-010-1500-0
   KNUDSON AG, 1971, P NATL ACAD SCI USA, V68, P820, DOI 10.1073/pnas.68.4.820
   Kong DH, 2005, CANCER RES, V65, P9047, DOI 10.1158/0008-5472.CAN-05-1235
   Korpershoek E, 2011, J CLIN ENDOCR METAB, V96, pE1472, DOI 10.1210/jc.2011-1043
   Kruspig B, 2014, MITOCHONDRION, V19, P8, DOI 10.1016/j.mito.2014.08.007
   Kubo T, 2004, BIOCHEM BIOPH RES CO, V318, P1006, DOI 10.1016/j.bbrc.2004.04.125
   Kung AL, 2004, CANCER CELL, V6, P33, DOI 10.1016/j.ccr.2004.06.009
   Ladroue C, 2008, NEW ENGL J MED, V359, P2685, DOI 10.1056/NEJMoa0806277
   Lee JS, 2016, BMC MED GENET, V17, DOI 10.1186/s12881-016-0306-2
   Lee K, 2009, P NATL ACAD SCI USA, V106, P17910, DOI 10.1073/pnas.0909353106
   Lee K, 2009, P NATL ACAD SCI USA, V106, P2353, DOI 10.1073/pnas.0812801106
   Lee K, 2011, ARCH PHARM RES, V34, P1583, DOI 10.1007/s12272-011-1021-3
   Lee K, 2010, BIOCHEM PHARMACOL, V80, P982, DOI 10.1016/j.bcp.2010.06.018
   Lee S, 2005, CANCER CELL, V8, P155, DOI 10.1016/j.ccr.2005.06.015
   Lee SH, 2015, J CELL PHYSIOL, V230, P853, DOI 10.1002/jcp.24813
   Lenders JWM, 2005, LANCET, V366, P665, DOI 10.1016/S0140-6736(05)67139-5
   Li L, 2014, INT J ONCOL, V44, P2111, DOI 10.3892/ijo.2014.2368
   Li SH, 2008, MOL CANCER THER, V7, P3729, DOI 10.1158/1535-7163.MCT-08-0074
   Liberti MV, 2016, TRENDS BIOCHEM SCI, V41, P211, DOI 10.1016/j.tibs.2015.12.001
   Liu QL, 2018, ENDOCR CONNECT, V7, P870, DOI 10.1530/EC-18-0167
   Liu QL, 2017, CANCER BIOL THER, V18, P944, DOI 10.1080/15384047.2017.1394553
   Liu QL, 2017, ENDOCR PATHOL, V28, P75, DOI 10.1007/s12022-017-9469-4
   Liu SJ, 2018, GENET MED, V20, P1266, DOI 10.1038/gim.2017.261
   Loboda A, 2010, MOL CELLS, V29, P435, DOI 10.1007/s10059-010-0067-2
   Lonser RR, 2003, LANCET, V361, P2059, DOI 10.1016/S0140-6736(03)13643-4
   Lorenzo FR, 2013, J MOL MED, V91, P507, DOI 10.1007/s00109-012-0967-z
   Lorusso D, 2010, CRIT REV ONCOL HEMAT, V74, P163, DOI 10.1016/j.critrevonc.2009.08.001
   Ma L, 2014, CANCER LETT, V355, P96, DOI 10.1016/j.canlet.2014.09.011
   Martin R, 1995, EMPORIO ARMANI M SET, VVII, pX, DOI DOI 10.1002/(SICI)1098-1004(1996)7:2<185::AID-HUMU22>3.0.CO;2-Y
   Masoud GN, 2015, ACTA PHARM SIN B, V5, P378, DOI 10.1016/j.apsb.2015.05.007
   Maxwell PH, 2001, CURR OPIN GENET DEV, V11, P293, DOI 10.1016/S0959-437X(00)00193-3
   Maxwell PH, 1997, P NATL ACAD SCI USA, V94, P8104, DOI 10.1073/pnas.94.15.8104
   Mayerhofer M, 2002, BLOOD, V100, P3767, DOI 10.1182/blood-2002-01-0109
   McCormack PL, 2014, DRUGS, V74, P1111, DOI 10.1007/s40265-014-0243-3
   Melillo G, 2007, METHOD ENZYMOL, V435, P385, DOI 10.1016/S0076-6879(07)35020-9
   Melillo G, 2007, CANCER METAST REV, V26, P341, DOI 10.1007/s10555-007-9059-x
   Minamishima YA, 2008, BLOOD, V111, P3236, DOI 10.1182/blood-2007-10-117812
   Minegishi H, 2013, ACS MED CHEM LETT, V4, P297, DOI 10.1021/ml3004632
   Miranda E, 2013, J AM CHEM SOC, V135, P10418, DOI 10.1021/ja402993u
   Morin A, 2020, CELL REP, V30, P4551, DOI 10.1016/j.celrep.2020.03.022
   Morris MR, 2009, ANTICANCER RES, V29, P4337
   Motzer RJ, 2013, NEW ENGL J MED, V369, P722, DOI 10.1056/NEJMoa1303989
   Musa F, 2013, EXPERT OPIN DRUG MET, V9, P215, DOI 10.1517/17425255.2013.758249
   Narita T, 2009, CLIN CANCER RES, V15, P6128, DOI 10.1158/1078-0432.CCR-08-3180
   Newman B, 2012, CANCER RES, V72, P4551, DOI 10.1158/0008-5472.CAN-11-3600
   Niemann S, 2000, NAT GENET, V26, P268, DOI 10.1038/81551
   Nomura K, 2009, J CLIN ENDOCR METAB, V94, P2850, DOI 10.1210/jc.2008-2697
   Nordstrom-O'Brien M, 2010, HUM MUTAT, V31, P521, DOI 10.1002/humu.21219
   O'Kane GM, 2019, BRIT J CANCER, V120, P1113, DOI 10.1038/s41416-019-0474-x
   Ohh M, 1998, MOL CELL, V1, P959, DOI 10.1016/S1097-2765(00)80096-9
   Ong KR, 2007, HUM MUTAT, V28, P143, DOI 10.1002/humu.20385
   Pacak K, 2013, J CLIN ONCOL, V31, P1690, DOI 10.1200/JCO.2012.47.1912
   Pan XY, 2020, OPHTHALMIC RES, V63, P182, DOI 10.1159/000504023
   Pan Y, 2007, MOL CELL BIOL, V27, P912, DOI 10.1128/MCB.01223-06
   Peng XF, 2020, J EXP CLIN CANC RES, V39, DOI 10.1186/s13046-020-1535-y
   Percy MJ, 2008, BLOOD, V111, P5400, DOI 10.1182/blood-2008-02-137703
   Percy MJ, 2008, NEW ENGL J MED, V358, P162, DOI 10.1056/NEJMoa073123
   Percy MJ, 2007, BLOOD, V110, P2193, DOI 10.1182/blood-2007-04-084434
   Philip B, 2013, CARCINOGENESIS, V34, P1699, DOI 10.1093/carcin/bgt209
   Philips George K, 2014, Am Soc Clin Oncol Educ Book, pe222, DOI 10.14694/EdBook_AM.2014.34.e222
   Pichun MEB, 2015, J CLIN ONCOL, V33
   Pouyssegur J, 2006, NATURE, V441, P437, DOI 10.1038/nature04871
   Puppo M, 2008, MOL CANCER THER, V7, P1974, DOI 10.1158/1535-7163.MCT-07-2059
   Rapisarda A, 2004, CANCER RES, V64, P1475, DOI 10.1158/0008-5472.CAN-03-3139
   Richard S, 2004, LANCET, V363, P1231, DOI 10.1016/S0140-6736(04)15957-6
   Richards F M, 2001, Expert Rev Mol Med, V2001, P1, DOI 10.1017/S1462399401002654
   Richter S, 2019, GENET MED, V21, P705, DOI 10.1038/s41436-018-0106-5
   Richter S, 2014, J CLIN ENDOCR METAB, V99, P3903, DOI 10.1210/jc.2014-2151
   Rini BI, 2011, LANCET, V378, P1931, DOI 10.1016/S0140-6736(11)61613-9
   Roskoski R, 2019, PHARMACOL RES, V144, P19, DOI 10.1016/j.phrs.2019.03.006
   Saito Y, 2012, AM J PHYSIOL-ENDOC M, V302, pE615, DOI 10.1152/ajpendo.00035.2011
   Samanta D, 2017, WIRES SYST BIOL MED, V9, DOI 10.1002/wsbm.1382
   Selak MA, 2005, CANCER CELL, V7, P77, DOI 10.1016/j.ccr.2004.11.022
   Semenza GL, 2010, ONCOGENE, V29, P625, DOI 10.1038/onc.2009.441
   Semenza GL, 2017, EXP CELL RES, V356, P128, DOI 10.1016/j.yexcr.2017.03.041
   Semenza GL, 2009, SEMIN CANCER BIOL, V19, P12, DOI 10.1016/j.semcancer.2008.11.009
   Shen C, 2013, SEMIN CANCER BIOL, V23, P18, DOI 10.1016/j.semcancer.2012.06.001
   STENSTROM G, 1986, ACTA MED SCAND, V220, P225
   Sun K, 2013, MOL CELL BIOL, V33, P904, DOI 10.1128/MCB.00951-12
   Taieb D, 2020, J NUCL MED, V61, P643, DOI 10.2967/jnumed.120.245613
   Pham TH, 2019, NUTRIENTS, V11, DOI 10.3390/nu11010079
   Toledo RA, 2013, ENDOCR-RELAT CANCER, V20, P349, DOI 10.1530/ERC-13-0101
   Triner D, 2016, J CLIN INVEST, V126, P3689, DOI 10.1172/JCI84430
   Vaidya A, 2018, NEW ENGL J MED, V378, P1259, DOI 10.1056/NEJMc1716652
   van Geel RMJM, 2012, ONCOLOGIST, V17, P1081, DOI 10.1634/theoncologist.2012-0055
   Vicha A, 2013, CURR OPIN ENDOCRINOL, V20, P186, DOI 10.1097/MED.0b013e32835fcc45
   Wallace EM, 2016, CANCER RES, V76, P5491, DOI 10.1158/0008-5472.CAN-16-0473
   Wang HR, 2018, CANCER RES, V78, P1266, DOI 10.1158/0008-5472.CAN-17-1718
   Wang L, 2010, DEV DYNAM, V239, P954, DOI 10.1002/dvdy.22213
   Wang YX, 2010, CANCER BIOL THER, V10, P376, DOI 10.4161/cbt.10.4.12441
   Weidemann A, 2008, CELL DEATH DIFFER, V15, P621, DOI 10.1038/cdd.2008.12
   Weirich G, 2002, J CLIN ENDOCR METAB, V87, P5241, DOI 10.1210/jc.2002-020651
   Welander J, 2011, ENDOCR-RELAT CANCER, V18, pR253, DOI 10.1530/ERC-11-0170
   Wigerup C, 2016, PHARMACOL THERAPEUT, V164, P152, DOI 10.1016/j.pharmthera.2016.04.009
   Wu DL, 2017, CURR OPIN STRUC BIOL, V43, P1, DOI 10.1016/j.sbi.2016.09.011
   Xia Y, 2012, EUR J MED CHEM, V49, P24, DOI 10.1016/j.ejmech.2012.01.033
   Xiao MT, 2012, GENE DEV, V26, P1326, DOI 10.1101/gad.191056.112
   Xu R, 2019, J MED CHEM, V62, P6876, DOI 10.1021/acs.jmedchem.9b00719
   Yamashita K, 2001, J BIOL CHEM, V276, P12645, DOI 10.1074/jbc.M011344200
   Yang CZ, 2015, J MOL MED, V93, P93, DOI 10.1007/s00109-014-1205-7
   Yang W, 2012, EXP CELL RES, V318, P944, DOI 10.1016/j.yexcr.2012.02.010
   Ying Q, 2011, HEPATOLOGY, V54, P2064, DOI 10.1002/hep.24614
   Yuan G, 2018, CANCER BIOL THER, V19, P766, DOI 10.1080/15384047.2018.1470732
   Zbar B, 1996, HUM MUTAT, V8, P348, DOI 10.1002/(SICI)1098-1004(1996)8:4<348::AID-HUMU8>3.3.CO;2-9
   Zhang L, 2018, INT IMMUNOPHARMACOL, V65, P429, DOI 10.1016/j.intimp.2018.10.035
   Zhao SM, 2009, SCIENCE, V324, P261, DOI 10.1126/science.1170944
   Zhong H, 1999, CANCER RES, V59, P5830
   Zhou X, 2018, RHEUMATOLOGY, V57, P1675, DOI 10.1093/rheumatology/key166
   Zhuang ZP, 2012, NEW ENGL J MED, V367, P922, DOI 10.1056/NEJMoa1205119
NR 196
TC 18
Z9 20
U1 0
U2 5
PU FRONTIERS MEDIA SA
PI LAUSANNE
PA AVENUE DU TRIBUNAL FEDERAL 34, LAUSANNE, CH-1015, SWITZERLAND
SN 1664-2392
J9 FRONT ENDOCRINOL
JI Front. Endocrinol.
PD NOV 24
PY 2020
VL 11
AR 586857
DI 10.3389/fendo.2020.586857
PG 13
WC Endocrinology & Metabolism
WE Science Citation Index Expanded (SCI-EXPANDED)
SC Endocrinology & Metabolism
GA PC3YU
UT WOS:000596941800001
PM 33329393
OA gold, Green Published
DA 2023-08-27
ER

PT J
AU Ferreri, NR
   McGiff, JC
   Vio, CP
   Carroll, MA
AF Ferreri, NR
   McGiff, JC
   Vio, CP
   Carroll, MA
TI TNF alpha-regulates renal COX-2 in the rat thick ascending limb (TAL)
SO THROMBOSIS RESEARCH
LA English
DT Article; Proceedings Paper
CT 11th Meeting of the Jagiellonian-Medical-Research-Center-Club
CY MAY 31-JUN 03, 2003
CL KRAKOW, POLAND
SP Jagiellonian Med Res Ctr Club
DE adrenalectomy (ADX); cyclooxygenase (COX)-2; prostaglandin E-2 (PGE(2));
   thick ascending limb (TAL); tumor necrosis factor alpha (TNF);
   20-hydroxyeicosatetraenoic acid (20-HETE)
ID CALCIUM-SENSING RECEPTOR; ARACHIDONIC-ACID; ANGIOTENSIN-II;
   BLOOD-PRESSURE; CYCLOOXYGENASE-2; RABBIT; IDENTIFICATION; METABOLISM;
   CELLS
AB We have examined cyclooxygenase (COX)-2-dependent mechanisms in preglomerular microvessels and the thick ascending limb (TAL). These renal structures are linchpins in the regulation of the renal circulation and extracellular fluid volume. Cytochrome P450 monooxygenases are the principal oxygenases in the TAL segment; however, COX-2 can be expressed in the TAL, as when challenged by angiotensin II. Glucocorticoids also affect the expression and activity of oxygenases in the TAL. Before adrenalectomy, <2% TAL cells expressed COX-2; after, >30% of TAL cells expressed COX-2. Recruitment of COX-2 is initiated in the renal cortex and proceeds to the medulla associated with: (1) COX-2 mRNA accumulation; (2) increased COX-2 expression; and (3) a two-fold increase in PGE(2) production by cortical microsomes. These changes were nullified by dexamethasone. COX-2 mRNA, protein expression and PGE(2) synthesis in the TAL are also increased in response to increased extracellular Ca2+. The Ca2+ sensing receptor is G-protein coupled and responds to changes in extracellular Ca2+ concentration by increasing protein kinase C activity to produce expression of COX-2. Thus, multiple signaling pathways contribute to COX-2 expression in TAL cells. (C) 2003 Elsevier Ltd. All rights reserved.
C1 New York Med Coll, Dept Pharmacol, Valhalla, NY 10595 USA.
   Pontificia Univ Catolica Chile, Fac Ciencias Biol, Dept Ciencias Fisiol, Santiago, Chile.
C3 New York Medical College; Pontificia Universidad Catolica de Chile
RP McGiff, JC (通讯作者)，New York Med Coll, Dept Pharmacol, Valhalla, NY 10595 USA.
EM
RI Vio, Carlos/JAX-7281-2023
OI Vio, Carlos/0000-0003-1850-5958
FU NHLBI NIH HHS [HL56423] Funding Source: Medline; PHS HHS [R01 25394]
   Funding Source: Medline; OAPP OPHS HHS [PPG HL34300] Funding Source:
   Medline
CR Baum MA, 1998, AM J MED SCI, V316, P321, DOI 10.1097/00000441-199811000-00005
   BONVALET JP, 1987, AM J PHYSIOL, V253, pF377, DOI 10.1152/ajprenal.1987.253.3.F377
   CAPDEVILA J, 1981, P NATL ACAD SCI-BIOL, V78, P5362, DOI 10.1073/pnas.78.9.5362
   CARROLL MA, 1991, J BIOL CHEM, V266, P12306
   DAVIDGE ST, 1995, CIRC RES, V77, P274, DOI 10.1161/01.RES.77.2.274
   El-Hajj Fuleihan G, 1998, J CLIN ENDOCR METAB, V83, P2366, DOI 10.1210/jc.83.7.2366
   ESCALANTE BA, 1994, AM J PHYSIOL, V266, pC1568, DOI 10.1152/ajpcell.1994.266.6.C1568
   Ferreri NR, 1998, AM J PHYSIOL-RENAL, V274, pF148, DOI 10.1152/ajprenal.1998.274.1.F148
   Ferreri NR, 1998, CLINICAL SIGNIFICANCE AND POTENTIAL OF SELECTIVE COX-2 INHIBITORS, P109
   FERRERI NR, 1984, J PHARMACOL EXP THER, V231, P441
   Ferreri NR, 1997, J HYPERTENS, V15, P1481, DOI 10.1097/00004872-199715120-00016
   Griffith LE, 1999, AM J HYPERTENS, V12, P84, DOI 10.1016/S0895-7061(98)00224-6
   HARDER DR, 1994, AM J PHYSIOL, V266, pH2098, DOI 10.1152/ajpheart.1994.266.5.H2098
   Hughes FJ, 1999, J BIOL CHEM, V274, P1776, DOI 10.1074/jbc.274.3.1776
   Lu M, 1996, J GEN PHYSIOL, V108, P537, DOI 10.1085/jgp.108.6.537
   MACICA CM, 1994, KIDNEY INT, V46, P113, DOI 10.1038/ki.1994.250
   McCarron DA, 1998, SCIENCE, V281, P933, DOI 10.1126/science.281.5379.933
   Pratt JH, 2000, AM J HYPERTENS, V13, P654, DOI 10.1016/S0895-7061(99)00285-X
   Qi ZH, 2002, J CLIN INVEST, V110, P61, DOI 10.1172/JCI200214752
   REEVES WB, 2000, KIDNEY PHYSL PATHOPH, P1333
   SCHWARTZMAN M, 1985, HYPERTENSION, V7, pI136, DOI 10.1161/01.HYP.7.3_Pt_2.I136
   SCHWARTZMAN M, 1985, NATURE, V314, P620, DOI 10.1038/314620a0
   SMITH WL, 1977, PROSTAGLANDINS, V13, P873, DOI 10.1016/0090-6980(77)90217-9
   VANLANSCHOT JJB, 1991, SURG GYNECOL OBSTET, V172, P293
   VASSALLI P, 1992, ANNU REV IMMUNOL, V10, P411, DOI 10.1146/annurev.immunol.10.1.411
   Vio CP, 2001, J AM SOC NEPHROL, V12, P649, DOI 10.1681/ASN.V124649
   Vio CP, 1997, HYPERTENSION, V30, P687, DOI 10.1161/01.HYP.30.3.687
   Wang D, 2000, J PHYSIOL PHARMACOL, V51, P587
   Wang DR, 2002, AM J PHYSIOL-RENAL, V283, pF963, DOI 10.1152/ajprenal.00108.2002
   Wang DR, 2001, AM J PHYSIOL-RENAL, V281, pF658, DOI 10.1152/ajprenal.2001.281.4.F658
   Zhang MZ, 1997, AM J PHYSIOL-RENAL, V273, pF994, DOI 10.1152/ajprenal.1997.273.6.F994
   ZOU AP, 1994, AM J PHYSIOL, V266, pF275, DOI 10.1152/ajprenal.1994.266.2.F275
NR 32
TC 10
Z9 10
U1 0
U2 0
PU PERGAMON-ELSEVIER SCIENCE LTD
PI OXFORD
PA THE BOULEVARD, LANGFORD LANE, KIDLINGTON, OXFORD OX5 1GB, ENGLAND
SN 0049-3848
EI 1879-2472
J9 THROMB RES
JI Thromb. Res.
PD JUN 15
PY 2003
VL 110
IS 5-6
BP 277
EP 280
DI 10.1016/S0049-3848(03)00381-5
PG 4
WC Hematology; Peripheral Vascular Disease
WE Science Citation Index Expanded (SCI-EXPANDED); Conference Proceedings Citation Index - Science (CPCI-S)
SC Hematology; Cardiovascular System & Cardiology
GA 743DV
UT WOS:000186557900007
PM 14592548
DA 2023-08-27
ER

PT J
AU Bernini, GP
   Miccoli, P
   Moretti, A
   Vivaldi, MS
   Iacconi, P
   Salvetti, A
AF Bernini, GP
   Miccoli, P
   Moretti, A
   Vivaldi, MS
   Iacconi, P
   Salvetti, A
TI Sixty adrenal masses of large dimensions: Hormonal and morphologic
   evaluation
SO UROLOGY
LA English
DT Article
ID COMPUTED-TOMOGRAPHY; CUSHINGS-SYNDROME; INCIDENTALOMAS;
   PHEOCHROMOCYTOMA; DIAGNOSIS; TUMORS; ABNORMALITIES; HYPERPLASIA;
   EXPERIENCE; SECRETION
AB Objectives. To evaluate the nature and function of adrenal masses of large dimensions (macrotumors).
   Methods. Sixty consecutive patients (31 women, 29 men, age range 15 to 84 years) with adrenal masses 4.0 cm in diameter or larger (range 4.0 to 15.0 cm) underwent morphologic study by computed tomography (CT); the majority also underwent 131-1-6beta-norcholesterol (1311-NC) or 1311-MIBG scintigraphy. Basal evaluation of glucocorticoids, mineralcorticoids, and catecholamines was performed in all patients, and in 38 cases determination of androgens was also made. In addition, on the basis of various clinical suspicions, a dynamic hormonal study was performed.
   Results. Macrotumors were benign in 78.3% of cases and included pheochromocytomas (n = 17), nonfunctioning cortical adenomas (n = 12), and cortisol-secreting tumors (n = 7, Cushing's syndrome). Malignant forms were 21.7% of the total, including pheochromocytomas (n = 3), cortical carcinomas (n = 6), and metastases (n = 4). On CT, malignant masses were larger (8.4 +/- 0.9 cm) than benign ones (5.7 +/- 0.3 cm) (P < 0.0001) and the mass size was strictly related to malignancy (P < 0.03). CT did not offer other diagnostic criteria for malignancy, except irregular margins and regional lymph node enlargement, which were more frequently (P < 0.0001) found in malignant forms. 1311-MIBG scintigraphy showed tracer uptake in all pheochromocytomas, both benign and malignant. By contrast, on 1311-NC scintigraphy, cortical malignancies never accumulated the radiotracer, whereas uptake was observed in all cases of solid cortical benign adenomas. Patients with cortical carcinomas showed plasma sex steroids above the normal range, pheochromocytomas were asymptomatic in 15% of cases, and almost half of the patients with Cushing's syndrome did not show clinical features of the disease (pre-Cushing's syndrome).
   Conclusions. Adrenal macrotumors frequently show endocrine activity and the medulla seems to be involved more than the cortex. Pheochromocytomas and cortisol-secreting adenomas are sometimes asymptomatic. Malignancy is often found in macromasses and, at least for the cortical forms, size of the tumor on CT, 1311-NC uptake on scintigraphy, and determination of levels of sex steroids seem to be useful criteria for predicting the nature of the mass. (C) 1998, Elsevier Science Inc. All rights reserved.
C1 Univ Pisa, Dipartimento Med Interna, I-56100 Pisa, Italy.
   Univ Pisa, Dipartimento Chirurg, I-56100 Pisa, Italy.
C3 University of Pisa; University of Pisa
RP Bernini, GP (通讯作者)，Univ Pisa, Dipartimento Med Interna, Via Roma 67, I-56100 Pisa, Italy.
CR AMBROSI B, 1995, EUR J ENDOCRINOL, V132, P422, DOI 10.1530/eje.0.1320422
   ASO Y, 1992, J UROLOGY, V147, P1478, DOI 10.1016/S0022-5347(17)37601-2
   BASKIN HJ, 1987, ARCH INTERN MED, V147, P847, DOI 10.1001/archinte.147.5.847
   BERNINI GP, 1994, J ENDOCRINOL INVEST, V17, P799, DOI 10.1007/BF03347780
   BEYER HS, 1986, J CLIN ENDOCR METAB, V62, P1317, DOI 10.1210/jcem-62-6-1317
   COPELAND PM, 1983, ANN INTERN MED, V98, P940, DOI 10.7326/0003-4819-98-6-940
   DERKSEN J, 1994, NEW ENGL J MED, V331, P968, DOI 10.1056/NEJM199410133311502
   DUNNICK NR, 1990, AM J ROENTGENOL, V154, P927, DOI 10.2214/ajr.154.5.2108567
   FRANCIS IR, 1992, RADIOLOGY, V184, P1
   GAJRAJ H, 1993, BRIT J SURG, V80, P422, DOI 10.1002/bjs.1800800405
   GIFFORD RW, 1994, ENDOCRIN METAB CLIN, V23, P387, DOI 10.1016/S0889-8529(18)30104-X
   GLAZER HS, 1982, AM J ROENTGENOL, V139, P81, DOI 10.2214/ajr.139.1.81
   GROSS MD, 1988, ANN INTERN MED, V109, P613, DOI 10.7326/0003-4819-109-8-613
   GROSS MD, 1994, J NUCL MED, V35, P1145
   GROSS MD, 1993, J CLIN ENDOCR METAB, V77, P885, DOI 10.1210/jc.77.4.885
   GUERRERO LA, 1985, UROLOGY, V26, P435, DOI 10.1016/0090-4295(85)90149-9
   HARTLEY L, 1985, AUST NZ J SURG, V55, P471, DOI 10.1111/j.1445-2197.1985.tb00925.x
   HEDELAND H, 1968, ACTA MED SCAND, V184, P211
   HUIRAS CM, 1989, JAMA-J AM MED ASSOC, V261, P894, DOI 10.1001/jama.261.6.894
   JARESCH S, 1992, J CLIN ENDOCR METAB, V74, P685, DOI 10.1210/jc.74.3.685
   KASSIRER JP, 1989, NEW ENGL J MED, V320, P1489, DOI 10.1056/NEJM198906013202211
   KHAFAGI FA, 1991, BRIT J SURG, V78, P828, DOI 10.1002/bjs.1800780720
   KOBAYASHI S, 1993, J UROLOGY, V150, P8, DOI 10.1016/S0022-5347(17)35384-3
   KRANE NK, 1986, ARCH INTERN MED, V146, P54, DOI 10.1001/archinte.146.1.54
   LOPEZ JM, 1990, J ENDOCRINOL INVEST, V13, P581, DOI 10.1007/BF03348629
   LUTON JP, 1990, NEW ENGL J MED, V322, P1195, DOI 10.1056/NEJM199004263221705
   MCLEOD MK, 1990, AM SURGEON, V56, P398
   MOULTON JS, 1988, SEMIN ROENTGENOL, V33, P288
   NEW MI, 1983, J CLIN ENDOCR METAB, V57, P320, DOI 10.1210/jcem-57-2-320
   OLEARY TJ, 1986, CAN J SURG, V29, P6
   OSELLA G, 1994, J CLIN ENDOCR METAB, V79, P1532, DOI 10.1210/jc.79.6.1532
   PRINZ RA, 1982, JAMA-J AM MED ASSOC, V248, P701, DOI 10.1001/jama.248.6.701
   REINCKE M, 1992, J CLIN ENDOCR METAB, V75, P826, DOI 10.1210/jc.75.3.826
   REINIG JW, 1994, RADIOLOGY, V192, P41, DOI 10.1148/radiology.192.1.8208962
   ROSEN HN, 1992, AM J MED, V92, P213, DOI 10.1016/0002-9343(92)90115-R
   ROSS NS, 1990, NEW ENGL J MED, V323, P1401
   SIEKAVIZZA JL, 1981, UROLOGY, V18, P625, DOI 10.1016/0090-4295(81)90476-3
   STENSTROM G, 1986, ACTA MED SCAND, V220, P225
   SUTTON MGS, 1981, MAYO CLIN PROC, V56, P354
   THOMPSON NW, 1987, SURG CLIN N AM, V67, P423
   TURTON DB, 1992, J ENDOCRINOL INVEST, V15, P789, DOI 10.1007/BF03348806
   VICK CW, 1984, UROL RADIOL, V6, P7, DOI 10.1007/BF02923691
   VIRKKALA A, 1989, ACTA ENDOCRINOL-COP, V121, P67, DOI 10.1530/acta.0.1210067
NR 43
TC 14
Z9 14
U1 0
U2 0
PU CAHNERS PUBL CO
PI NEW YORK
PA 249 WEST 17 STREET, NEW YORK, NY 10011 USA
SN 0090-4295
J9 UROLOGY
JI UROLOGY
PD JUN
PY 1998
VL 51
IS 6
BP 920
EP 925
DI 10.1016/S0090-4295(98)00026-0
PG 6
WC Urology & Nephrology
WE Science Citation Index Expanded (SCI-EXPANDED)
SC Urology & Nephrology
GA ZP380
UT WOS:000073746500013
PM 9609627
DA 2023-08-27
ER

PT J
AU Manley, GT
   Binder, DK
   Papadopoulos, MC
   Verkman, AS
AF Manley, GT
   Binder, DK
   Papadopoulos, MC
   Verkman, AS
TI New insights into water transport and edema in the central nervous
   system from phenotype analysis of aquaporin-4 null mice
SO NEUROSCIENCE
LA English
DT Article
DE brain injury; cytotoxic edema; epilepsy; stroke; vasogenic edema water
   channel
ID CUZN-SUPEROXIDE DISMUTASE; FOCAL CEREBRAL-ISCHEMIA; DYSTROPHIC MDX-MICE;
   INDUCED BRAIN EDEMA; ORTHOGONAL ARRAYS; CHANNEL PROTEIN; GLIAL-CELLS;
   EXTRACELLULAR-SPACE; EPILEPTIFORM BURSTS; SKELETAL-MUSCLE
AB Aquaporin-4 (AQP4) is the major water channel in the CNS. Its expression at fluid-tissue barriers (blood-brain and brain-cerebrospinal fluid barriers) throughout the brain and spinal cord suggests a role in water transport under normal and pathological conditions. Phenotype studies of transgenic mice lacking AQP4 have provided evidence for a role of AQP4 in cerebral water balance and neural signal transduction. Primary cultures of astrocytes from AQP4-null mice have greatly reduced osmotic water permeability compared with wild-type astrocytes, indicating that AQP4 is the principal water channel in these cells. AQP4-null mice have reduced brain swelling and improved neurological outcome following water intoxication and focal cerebral ischemia, establishing a role of AQP4 in the development of cytotoxic (cellular) cerebral edema. In contrast, brain swelling and clinical outcome are worse in AQP4-null mice in models of vasogenic (fluid leak) edema caused by freeze-injury and brain tumor, probably due to impaired AQP4-dependent brain water clearance. AQP4-null mice also have markedly reduced acoustic brainstem response potentials and significantly increased seizure threshold in response to chemical convulsants, implicating AQP4 in modulation of neural signal transduction. Pharmacological modulation of AQP4 function may thus provide a novel therapeutic strategy for the treatment of stroke, tumor-associated edema, epilepsy, traumatic brain injury, and other disorders of the CNS associated with altered brain water balance. (C) 2004 IBRO. Published by Elsevier Ltd. All rights reserved.
C1 Univ Calif San Francisco, Dept Neurol Surg, Cardiovasc Res Inst, San Francisco, CA 94143 USA.
   Univ Calif San Francisco, Dept Med & Physiol, Cardiovasc Res Inst, San Francisco, CA 94143 USA.
C3 University of California System; University of California San Francisco;
   University of California System; University of California San Francisco
RP Manley, GT (通讯作者)，Univ Calif San Francisco, Dept Neurol Surg, Cardiovasc Res Inst, 1001 Potrero Ave,Bldg 1,Room 101, San Francisco, CA 94143 USA.
EM
OI Papadopoulos, Marios/0000-0001-9174-4176
CR Abbott NJ, 2004, NEUROCHEM INT, V45, P545, DOI 10.1016/j.neuint.2003.11.006
   Agre P, 2002, J PHYSIOL-LONDON, V542, P3, DOI 10.1113/jphysiol.2002.020818
   Amiry-Moghaddam M, 2003, NAT REV NEUROSCI, V4, P991, DOI 10.1038/nrn1252
   Amiry-Moghaddam M, 2003, P NATL ACAD SCI USA, V100, P13615, DOI 10.1073/pnas.2336064100
   Amiry-Moghaddam M, 2003, P NATL ACAD SCI USA, V100, P2106, DOI 10.1073/pnas.0437946100
   ANDREW RD, 1989, BRAIN RES, V498, P175, DOI 10.1016/0006-8993(89)90417-4
   Badaut J, 2001, J CEREBR BLOOD F MET, V21, P477, DOI 10.1097/00004647-200105000-00001
   Badaut T, 2002, J CEREBR BLOOD F MET, V22, P367, DOI 10.1097/00004647-200204000-00001
   Binder DK, 2004, NEUROREPORT, V15, P259, DOI 10.1097/00001756-200402090-00009
   CHAN PH, 1991, ANN NEUROL, V29, P482, DOI 10.1002/ana.410290506
   CHEBABO SR, 1995, BRAIN RES, V695, P203, DOI 10.1016/0006-8993(95)00778-O
   CUEVAS P, 1985, ANAT EMBRYOL, V172, P171, DOI 10.1007/BF00319599
   Dalloz C, 2003, HUM MOL GENET, V12, P1543, DOI 10.1093/hmg/ddg170
   DUDEK FE, 1990, NEUROSCI LETT, V120, P267, DOI 10.1016/0304-3940(90)90056-F
   FISHMAN RA, 1975, NEW ENGL J MED, V293, P706, DOI 10.1056/NEJM197510022931407
   FRIGERI A, 1995, J CELL SCI, V108, P2993
   Frigeri A, 2001, FASEB J, V15, P90, DOI 10.1096/fj.00-0260com
   Furman CS, 2003, P NATL ACAD SCI USA, V100, P13609, DOI 10.1073/pnas.2235843100
   Hamann S, 2002, J FLUORESC, V12, P139, DOI 10.1023/A:1016832027325
   HASEGAWA H, 1994, J BIOL CHEM, V269, P5497
   HOCHMAN DW, 1995, SCIENCE, V270, P99, DOI 10.1126/science.270.5233.99
   Horio Y, 2001, JPN J PHARMACOL, V87, P1, DOI 10.1254/jjp.87.1
   Hossmann KA, 1998, CARDIOVASC RES, V39, P106, DOI 10.1016/S0008-6363(98)00075-3
   JOHNSTONE BM, 1989, J PHYSIOL-LONDON, V408, P77, DOI 10.1113/jphysiol.1989.sp017448
   JUNG JS, 1994, P NATL ACAD SCI USA, V91, P13052, DOI 10.1073/pnas.91.26.13052
   Kofuji P, 2000, J NEUROSCI, V20, P5733, DOI 10.1523/JNEUROSCI.20-15-05733.2000
   Kondo T, 1997, J NEUROSCI, V17, P4180
   LANDIS DMD, 1974, J CELL BIOL, V60, P316, DOI 10.1083/jcb.60.1.316
   Li J, 2001, J BIOL CHEM, V276, P31233, DOI 10.1074/jbc.M104368200
   Ma TH, 1997, J CLIN INVEST, V100, P957, DOI 10.1172/JCI231
   Manley GT, 2000, NAT MED, V6, P159, DOI 10.1038/72256
   MARMAROU A, 1994, AM J PHYSIOL, V267, pH514, DOI 10.1152/ajpheart.1994.267.2.H514
   MCBAIN CJ, 1990, SCIENCE, V249, P674, DOI 10.1126/science.2382142
   Meinild AK, 1998, J BIOL CHEM, V273, P32446, DOI 10.1074/jbc.273.49.32446
   Mhatre AN, 2002, HEARING RES, V170, P59, DOI 10.1016/S0378-5955(02)00452-5
   Nagelhus EA, 1998, J NEUROSCI, V18, P2506
   Neely JD, 1999, BIOCHEMISTRY-US, V38, P11156, DOI 10.1021/bi990941s
   Neely JD, 2001, P NATL ACAD SCI USA, V98, P14108, DOI 10.1073/pnas.241508198
   Nicchia GP, 2003, FASEB J, V17, P1508, DOI 10.1096/fj.02-1183fje
   Nicchia GP, 2000, GLIA, V31, P29, DOI 10.1002/(SICI)1098-1136(200007)31:1<29::AID-GLIA30>3.0.CO;2-3
   Nico B, 2003, GLIA, V42, P235, DOI 10.1002/glia.10216
   Nielsen S, 1997, J NEUROSCI, V17, P171
   Oshio K, 2004, NEUROSCIENCE, V127, P685, DOI 10.1016/j.neuroscience.2004.03.016
   OSHIO K, 2004, IN PRESS FASEB J
   OURY TD, 1993, J BIOL CHEM, V268, P15394
   Pan EH, 1996, NEUROSCI LETT, V207, P9, DOI 10.1016/0304-3940(96)12477-0
   Papadopoulos MC, 2004, NEUROSCIENCE, V129, P1011, DOI 10.1016/j.neuroscience.2004.05.044
   Papadopoulos MC, 2004, FASEB J, V18, P1291, DOI 10.1096/fj.04-1723fje
   Ransom B, 2003, TRENDS NEUROSCI, V26, P520, DOI 10.1016/j.tins.2003.08.006
   Rash JE, 1998, P NATL ACAD SCI USA, V95, P11981, DOI 10.1073/pnas.95.20.11981
   REULEN HJ, 1977, J NEUROSURG, V46, P24, DOI 10.3171/jns.1977.46.1.0024
   ROPER SN, 1992, ANN NEUROL, V31, P81, DOI 10.1002/ana.410310115
   SHI LB, 1995, BIOCHEMISTRY-US, V34, P8250, DOI 10.1021/bi00026a006
   Shi LB, 1996, BIOCHEMISTRY-US, V35, P538, DOI 10.1021/bi9520038
   Solenov E, 2004, AM J PHYSIOL-CELL PH, V286, pC426, DOI 10.1152/ajpcell.00298.2003
   Speake T, 2003, BBA-BIOMEMBRANES, V1609, P80, DOI 10.1016/S0005-2736(02)00658-2
   Stringer JL, 1997, BRAIN RES, V745, P328, DOI 10.1016/S0006-8993(96)01253-X
   Takumi Y, 1998, EUR J NEUROSCI, V10, P3584, DOI 10.1046/j.1460-9568.1998.00360.x
   TRAYNELIS SF, 1989, J NEUROPHYSIOL, V61, P927, DOI 10.1152/jn.1989.61.5.927
   Vajda Z, 2002, P NATL ACAD SCI USA, V99, P13131, DOI 10.1073/pnas.192457099
   Verbavatz JM, 1997, J CELL SCI, V110, P2855
   Verkman AS, 2002, ANN MED, V34, P192, DOI 10.1080/713782138
   Weed LH, 1919, AM J PHYSIOL, V48, P531, DOI 10.1152/ajplegacy.1919.48.4.531
   WOLBURG H, 1995, J BRAIN RES, V36, P239
   Yang BX, 1996, J BIOL CHEM, V271, P4577
   Yang BX, 2000, AM J PHYSIOL-CELL PH, V278, pC1108, DOI 10.1152/ajpcell.2000.278.6.C1108
NR 66
TC 250
Z9 286
U1 0
U2 20
PU PERGAMON-ELSEVIER SCIENCE LTD
PI OXFORD
PA THE BOULEVARD, LANGFORD LANE, KIDLINGTON, OXFORD OX5 1GB, ENGLAND
SN 0306-4522
EI 1873-7544
J9 NEUROSCIENCE
JI Neuroscience
PY 2004
VL 129
IS 4
BP 983
EP 991
DI 10.1016/j.neuroscience.2004.06.088
PG 9
WC Neurosciences
WE Science Citation Index Expanded (SCI-EXPANDED)
SC Neurosciences & Neurology
GA 879OB
UT WOS:000225726000012
PM 15561413
DA 2023-08-27
ER

PT J
AU Greenlee, JE
AF Greenlee, John E.
TI Treatment of Paraneoplastic Neurologic Disorders
SO CURRENT TREATMENT OPTIONS IN NEUROLOGY
LA English
DT Review
ID STIFF-PERSON SYNDROME; EATON MYASTHENIC SYNDROME; INTRAVENOUS
   IMMUNOGLOBULIN THERAPY; CEREBELLAR DEGENERATION; PLASMA-EXCHANGE; LIMBIC
   ENCEPHALITIS; ANTI-HU; NEUROIMMUNOLOGICAL DISORDERS; ANTINEURONAL
   ANTIBODIES; OPSOCLONUS-MYOCLONUS
AB Paraneoplastic neurologic disorders are rare, autoimmune disorders, which can be broken down into two groups: those in which antibody response is directed against intracellular neuronal or neuroglial proteins (Group 1) and those in which the immune response is directed against antigens within or subjacent to the neuronal cell membrane (Group 2). In both groups, detection and treatment of the underlying neoplasm is critical and carries the best chance of clinical stabilization or remission.
   Syndromes in Group 2 frequently respond to therapy. This may involve corticosteroids, plasma exchange (PE), or intravenous immunoglobulin G (IgG), depending on the specific paraneoplastic syndrome. Cyclophosphamide or rituximab may be helpful in patients who fail to stabilize or improve on less aggressive therapies.
   Treatment of syndromes in Group 1 is far more difficult, and proven treatment strategies do not exist. Younger men (< 40 years of age) with limbic or brainstem syndromes, testicular or germ cell tumors, and anti-Ma2 antibodies may respond to specific tumor treatment together with immunotherapy. Patients with paraneoplastic syndromes and anti-Ri antibodies may respond to corticosteroids and/or cyclophosphamide. Evidence-based treatment guidelines do not exist for patients with other central paraneoplastic syndromes such as cerebellar degeneration or encephalomyeloneuritis. Approaches to therapy, apart from treating the underlying tumor, are thus speculative.
   In patients with rapidly progressive symptoms classically suggestive of a paraneoplastic neurologic syndrome, time is of the essence in arresting neurologic deterioration. Clinical improvement in patients with longstanding symptoms is unlikely. At the outset, one should move rapidly to define the antibody response involved, as this may also assist tumor diagnosis. Treatment may include prednisone, intravenous IgG, and cyclophosphamide; rituximab plus prednisone may be an alternative, either initially or in the face of continued disease progression despite treatment with intravenous IgG or cyclophosphamide. Although PE is of questionable benefit, a single cycle of PE may be considered before other treatment, to achieve rapid lowering of circulating paraneoplastic autoantibodies.
C1 [Greenlee, John E.] Univ Utah, Clin Neurosci Ctr, Salt Lake City, UT 84132 USA.
C3 Utah System of Higher Education; University of Utah
RP Greenlee, JE (通讯作者)，Univ Utah, Clin Neurosci Ctr, 175 N Med Dr E, Salt Lake City, UT 84132 USA.
EM
CR Albert ML, 2000, ANN NEUROL, V47, P9
   Baker MR, 2005, J NEUROL NEUROSUR PS, V76, P999, DOI 10.1136/jnnp.2004.051144
   Bataller L, 2001, BRAIN, V124, P437, DOI 10.1093/brain/124.2.437
   Bataller L, 2010, NEUROLOGY, V74, P265, DOI 10.1212/WNL.0b013e3181cb3e52
   Batchelor TT, 1998, J NEURO-ONCOL, V40, P131, DOI 10.1023/A:1006136219490
   Butler MH, 2000, NEURON, V26, P307, DOI 10.1016/S0896-6273(00)81165-4
   Candler PM, 2004, J NEUROL NEUROSUR PS, V75, P1411, DOI 10.1136/jnnp.2003.025171
   Carson KR, 2009, BLOOD, V113, P4834, DOI 10.1182/blood-2008-10-186999
   CHER LM, 1995, CANCER-AM CANCER SOC, V75, P1678, DOI 10.1002/1097-0142(19950401)75:7<1678::AID-CNCR2820750719>3.0.CO;2-2
   Coret F, 2009, J NEURO-ONCOL, V93, P421, DOI 10.1007/s11060-008-9787-y
   DALAKAS MC, 1994, NEUROLOGY, V44, P223, DOI 10.1212/WNL.44.2.223
   Dalakas MC, 2001, NEW ENGL J MED, V345, P1870, DOI 10.1056/NEJMoa01167
   Dalmau J, 2004, BRAIN, V127, P1831, DOI 10.1093/brain/awh203
   Dalmau J, 2008, LANCET NEUROL, V7, P327, DOI 10.1016/S1474-4422(08)70060-7
   de la Sayette V, 1998, ARCH NEUROL-CHICAGO, V55, P405, DOI 10.1001/archneur.55.3.405
   Duddy ME, 2009, FRONT NEUROL NEUROSC, V26, P147, DOI 10.1159/000212375
   Dupond JL, 2010, J CLIN NEUROSCI, V17, P389, DOI 10.1016/j.jocn.2009.06.015
   Elovaara I, 2008, EUR J NEUROL, V15, P893, DOI 10.1111/j.1468-1331.2008.02246.x
   Esposito M, 2008, J NEURO-ONCOL, V86, P363, DOI 10.1007/s11060-007-9479-z
   Evangelou N, 2003, J CLIN PATHOL, V56, P308, DOI 10.1136/jcp.56.4.308
   Gilhus NE, 2009, EXPERT REV NEUROTHER, V9, P351, DOI 10.1586/14737175.9.3.351
   Graus F, 2008, NEUROLOGY, V71, P930, DOI 10.1212/01.wnl.0000325917.48466.55
   GRAUS F, 1990, NEUROLOGY, V40, P1621, DOI 10.1212/WNL.40.10.1621
   Graus F, 2001, BRAIN, V124, P1138, DOI 10.1093/brain/124.6.1138
   GRAUS F, 2009, J NEUROL        1225
   HAMMACK J, 1992, NEUROLOGY, V42, P1938
   Hoffmann LA, 2008, J NEUROL NEUROSUR PS, V79, P767, DOI 10.1136/jnnp.2007.118588
   JANIACSADI A, 2010, CURR TREAT IN PRESS
   Jayabose S, 1999, J PEDIAT HEMATOL ONC, V21, P514, DOI 10.1097/00043426-199911000-00012
   Keime-Guibert F, 2000, J NEUROL NEUROSUR PS, V68, P479, DOI 10.1136/jnnp.68.4.479
   KIRSCH GE, 1978, BIOPHYS J, V22, P507, DOI 10.1016/S0006-3495(78)85503-9
   Lai MZ, 2009, ANN NEUROL, V65, P424, DOI 10.1002/ana.21589
   Lehmann HC, 2006, ARCH NEUROL-CHICAGO, V63, P1066, DOI 10.1001/archneur.63.8.1066
   Lehmann HC, 2006, ARCH NEUROL-CHICAGO, V63, P930, DOI 10.1001/archneur.63.7.930
   LEVI ME, 2009, CLIN TRANSPLANT 0803
   LUNDH H, 1984, NEUROLOGY, V34, P1324, DOI 10.1212/WNL.34.10.1324
   LUNDH H, 1993, ACTA NEUROL SCAND, V88, P136
   LUQUE FA, 1991, ANN NEUROL, V29, P241, DOI 10.1002/ana.410290303
   MCEVOY KM, 1989, NEW ENGL J MED, V321, P1567, DOI 10.1056/NEJM198912073212303
   Meinck HM, 2001, CNS DRUGS, V15, P515, DOI 10.2165/00023210-200115070-00002
   Murinson BB, 2008, NEUROLOGY, V71, P1955, DOI 10.1212/01.wnl.0000327342.58936.e0
   Oh SJ, 2009, MUSCLE NERVE, V40, P795, DOI 10.1002/mus.21422
   Pellkofer H, 2004, BRAIN, V127, P1822, DOI 10.1093/brain/awh205
   Pellkofer HL, 2009, ARCH NEUROL-CHICAGO, V66, P655, DOI 10.1001/archneurol.2009.56
   Petereit HF, 2008, ACTA NEUROL SCAND, V117, P399, DOI 10.1111/j.1600-0404.2007.00958.x
   PETERSON K, 1992, NEUROLOGY, V42, P1931, DOI 10.1212/wnl.42.10.1931
   Phuphanich S, 2007, J NEURO-ONCOL, V81, P67, DOI 10.1007/s11060-006-9198-x
   Pittock SJ, 2003, ANN NEUROL, V53, P580, DOI 10.1002/ana.10518
   Poepel A, 2007, J NEUROL SCI, V263, P145, DOI 10.1016/j.jns.2007.07.007
   Pranzatelli MR, 2010, J CLIN IMMUNOL, V30, P106, DOI 10.1007/s10875-009-9335-3
   Punga AR, 2009, MUSCLE NERVE, V39, P724, DOI 10.1002/mus.21319
   Rojas I, 2000, NEUROLOGY, V55, P713, DOI 10.1212/WNL.55.5.713
   Sanders DB, 2000, NEUROLOGY, V54, P603, DOI 10.1212/WNL.54.3.603
   SCHLAKE HP, 1989, CLIN NEUROL NEUROSUR, V91, P329, DOI 10.1016/0303-8467(89)90010-3
   Schroeder A, 2009, EXPERT REV NEUROTHER, V9, P1331, DOI 10.1586/ERN.09.81
   SEKUL EA, 1994, ANN INTERN MED, V121, P259, DOI 10.7326/0003-4819-121-4-199408150-00004
   Shams'ili S, 2006, J NEUROL, V253, P16, DOI 10.1007/s00415-005-0882-0
   Shams'ili S, 2003, BRAIN, V126, P1409, DOI 10.1093/brain/awg133
   Sharma M, 2009, AM J HEMATOL, V84, P614, DOI 10.1002/ajh.21484
   Shemin D, 2007, J CLIN APHERESIS, V22, P270, DOI 10.1002/jca.20143
   Skeie GO, 2006, EUR J NEUROL, V13, P691, DOI 10.1111/j.1468-1331.2006.01476.x
   THOMSEN RH, 1983, J PHARMACOL EXP THER, V227, P260
   Uchuya M, 1996, J NEUROL NEUROSUR PS, V60, P388, DOI 10.1136/jnnp.60.4.388
   Vedeler CA, 2006, EUR J NEUROL, V13, P682, DOI 10.1111/j.1468-1331.2006.01266.x
   Vernino S, 2004, NEURO-ONCOLOGY, V6, P55, DOI 10.1215/S1152851703000395
   Voltz R, 2006, J NEUROL, V253, P33, DOI 10.1007/s00415-006-5005-z
   Weinstein R, 2000, J CLIN APHERESIS, V15, P74, DOI 10.1002/(SICI)1098-1101(2000)15:1/2<74::AID-JCA6>3.0.CO;2-O
   Weinstein R, 2008, J CLIN APHERESIS, V23, P196, DOI 10.1002/jca.20178
   Widdess-Walsh P, 2003, J NEURO-ONCOL, V63, P187, DOI 10.1023/A:1023931501503
   Wirtz PW, 2009, CLIN PHARMACOL THER, V86, P44, DOI 10.1038/clpt.2009.35
NR 70
TC 32
Z9 32
U1 0
U2 4
PU CURRENT MEDICINE GROUP
PI PHILADELPHIA
PA 400 MARKET STREET, STE 700, PHILADELPHIA, PA 19106 USA
SN 1092-8480
J9 CURR TREAT OPTION NE
JI Curr. Treat. Options Neurol.
PD MAY
PY 2010
VL 12
IS 3
BP 212
EP 230
DI 10.1007/s11940-010-0066-9
PG 19
WC Clinical Neurology
WE Science Citation Index Expanded (SCI-EXPANDED)
SC Neurosciences & Neurology
GA 589QB
UT WOS:000277166200005
PM 20842583
DA 2023-08-27
ER

PT J
AU Li, GR
   Cui, G
   Tzeng, NS
   Wei, SJ
   Wang, TG
   Block, ML
   Hong, JS
AF Li, GR
   Cui, G
   Tzeng, NS
   Wei, SJ
   Wang, TG
   Block, ML
   Hong, JS
TI Femtomolar concentrations of dextromethorphan protect mesencephalic
   dopaminergic neurons from inflammatory damage
SO FASEB JOURNAL
LA English
DT Article
DE DM; femtomolar; inflammation; microglia; neuroprotection; Parkinson's
   disease
ID LIPOPOLYSACCHARIDE-INDUCED NEUROTOXICITY; INJURY FOLLOWING TRANSIENT;
   NEURODEGENERATIVE DISEASES; MICROGLIAL ACTIVATION; PARKINSONS-DISEASE;
   ALZHEIMERS-DISEASE; OPIOID PEPTIDE; FOCAL ISCHEMIA; NADPH OXIDASE; RAT
AB Inflammation in the brain has increasingly been recognized to play an important role in the pathogenesis of several neuro degenerative disorders, including Parkinson's disease (PD). Progress in the search for effective therapeutic strategies that can halt this degenerative process remains limited. We previously showed that micromolar concentrations of dextromethorphan (DM), a major ingredient of widely used antitussive remedies, reduced the inflammation-mediated degeneration of dopaminergic neurons through the inhibition of microglial activation. In this study, we report that femto- and micromolar concentrations of DM (both pre- and post-treatment) showed equal efficacy in protecting lipopolysaccharide (LPS)-induced dopaminergic neuron death in midbrain neuron-glia cultures. Both concentrations of DM decreased LPS-induced release of nitric oxide, tumor necrosis factor-alpha, prostaglandin E-2 and superoxide from microglia in comparable degrees. The important role of superoxide was demonstrated by DM's failure to show a neuroprotective effect in neuron-glia cultures from NADPH oxidase-deficient mice. These results suggest that the neuroprotective effect elicited by femtomolar concentrations of DM is mediated through the inhibition of LPS-induced proinflammatory factors, especially superoxide. These findings suggest a novel therapeutic concept of using "ultra-low" drug concentrations for the intervention of inflammation-related neurodegenerative diseases.
C1 NIEHS, Neuropharmacol Sect, Lab Pharmacol & Chem, NIH, Res Triangle Pk, NC 27709 USA.
   NIEHS, Natl Ctr Toxicogenom, Res Triangle Pk, NC 27709 USA.
   Univ N Carolina, Chapel Hill, NC USA.
   Tri Serv Gen Hosp, Natl Def Med Ctr, Dept Psychiat, Taipei, Taiwan.
C3 National Institutes of Health (NIH) - USA; NIH National Institute of
   Environmental Health Sciences (NIEHS); National Institutes of Health
   (NIH) - USA; NIH National Institute of Environmental Health Sciences
   (NIEHS); University of North Carolina; University of North Carolina
   Chapel Hill; National Defense Medical Center; Tri-Service General
   Hospital
RP Li, GR (通讯作者)，NIEHS, Neuropharmacol Sect, Lab Pharmacol & Chem, NIH, POB 12233,MD-F1-01 111 Alexander Dr, Res Triangle Pk, NC 27709 USA.
EM
RI Tzeng, Nian-Sheng/AAU-4945-2021; Hong, Jau-shyong/F-1920-2019
OI Hong, Jau-shyong/0000-0002-3056-8401; Block,
   Michelle/0000-0001-8303-7914; Wei, Sung Jen/0000-0002-9810-9041
CR BASKIN DS, 1984, NATURE, V312, P551, DOI 10.1038/312551a0
   Britton P, 1997, LIFE SCI, V60, P1729, DOI 10.1016/S0024-3205(97)00132-X
   Christensen ST, 1996, CELL BIOL INT, V20, P437, DOI 10.1006/cbir.1996.0055
   Crain SM, 2001, BRAIN RES, V888, P75, DOI 10.1016/S0006-8993(00)03010-9
   DICKSON DW, 1993, GLIA, V7, P75, DOI 10.1002/glia.440070113
   EFANOV AM, 1994, FEBS LETT, V355, P114, DOI 10.1016/0014-5793(94)01109-5
   Gao HM, 2002, J NEUROCHEM, V81, P1285, DOI 10.1046/j.1471-4159.2002.00928.x
   GATLEY SJ, 1976, BIOCHEM J, V158, P307, DOI 10.1042/bj1580307
   GEORGE CP, 1988, BRAIN RES, V440, P375, DOI 10.1016/0006-8993(88)91011-6
   Giulian D, 1999, AM J HUM GENET, V65, P13, DOI 10.1086/302477
   Greenamyre JT, 1999, BIOCHEM SOC SYMP, V66, P85, DOI 10.1042/bss0660085
   Hayashi T, 2002, SYNAPSE, V43, P86, DOI 10.1002/syn.10019
   Jenner P, 1996, PATHOL BIOL, V44, P57
   Jeohn GH, 1998, J NEUROIMMUNOL, V85, P1, DOI 10.1016/S0165-5728(97)00204-X
   Kim WG, 2000, J NEUROSCI, V20, P6309, DOI 10.1523/JNEUROSCI.20-16-06309.2000
   KUTTAN SC, 1991, J CARDIOVASC PHARM, V17, P929, DOI 10.1097/00005344-199106000-00011
   Leker RR, 2002, STROKE, V33, P1085, DOI 10.1161/01.STR.0000014207.05597.D7
   Liu B, 2000, NEUROSCIENCE, V97, P749, DOI 10.1016/S0306-4522(00)00057-9
   Liu B, 2003, METH MOLEC MED, V79, P387
   Liu B, 2003, J PHARMACOL EXP THER, V304, P1, DOI 10.1124/jpet.102.035048
   Liu B, 2001, J PHARMACOL EXP THER, V298, P1133
   Liu J, 2001, LIFE SCI, V69, P1833, DOI 10.1016/S0024-3205(01)01267-X
   Liu YX, 2003, J PHARMACOL EXP THER, V305, P212, DOI 10.1124/jpet.102.043166
   Liu YX, 2002, J PHARMACOL EXP THER, V302, P1212, DOI 10.1124/jpet.102.035956
   McGeer PL, 1995, BRAIN RES REV, V21, P195, DOI 10.1016/0165-0173(95)00011-9
   MCGEER PL, 1988, NEUROLOGY, V38, P1285, DOI 10.1212/WNL.38.8.1285
   MONYER H, 1988, BRAIN RES, V446, P144, DOI 10.1016/0006-8993(88)91304-2
   PRINCE DA, 1988, NEUROSCI LETT, V85, P291, DOI 10.1016/0304-3940(88)90581-2
   Qin LY, 2004, J BIOL CHEM, V279, P1415, DOI 10.1074/jbc.M307657200
   Qin LY, 2002, J NEUROCHEM, V83, P973, DOI 10.1046/j.1471-4159.2002.01210.x
   RAMESHWAR P, 1993, J IMMUNOL, V151, P2484
   Reglodi D, 2000, STROKE, V31, P1411, DOI 10.1161/01.STR.31.6.1411
   Sharp BM, 1998, BIOCHEM PHARMACOL, V56, P289, DOI 10.1016/S0006-2952(98)00155-5
   Souza HP, 2001, AM J PHYSIOL-HEART C, V280, pH658, DOI 10.1152/ajpheart.2001.280.2.H658
   STEINBERG GK, 1988, STROKE, V19, P1112, DOI 10.1161/01.STR.19.9.1112
   Tortella FC, 1999, J PHARMACOL EXP THER, V291, P399
   Yoshida LS, 1998, J BIOL CHEM, V273, P27879, DOI 10.1074/jbc.273.43.27879
NR 37
TC 65
Z9 72
U1 0
U2 6
PU FEDERATION AMER SOC EXP BIOL
PI BETHESDA
PA 9650 ROCKVILLE PIKE, BETHESDA, MD 20814-3998 USA
SN 0892-6638
EI 1530-6860
J9 FASEB J
JI Faseb J.
PD APR
PY 2005
VL 19
IS 6
BP 489
EP 496
DI 10.1096/fj.04-2555com
PG 8
WC Biochemistry & Molecular Biology; Biology; Cell Biology
WE Science Citation Index Expanded (SCI-EXPANDED)
SC Biochemistry & Molecular Biology; Life Sciences & Biomedicine - Other
   Topics; Cell Biology
GA 922UP
UT WOS:000228865300032
PM 15790998
DA 2023-08-27
ER

PT J
AU Asarian, L
   Langhans, W
AF Asarian, Lori
   Langhans, Wolfgang
TI A new look on brain mechanisms of acute illness anorexia
SO PHYSIOLOGY & BEHAVIOR
LA English
DT Article; Proceedings Paper
CT 17th Annual Meeting of the Society-for-the-Study-of-Ingestive-Behavior
CY JUL 28-AUG 01, 2009
CL Portland, OR
SP Soc Study Ingest Behav
DE LPS; Cytokines; aPKC; EP receptor; 5-HT receptor; Macrophage;
   Perivascular cells; Blood-brain barrier; Estradiol
ID LIPOPOLYSACCHARIDE-INDUCED ANOREXIA; TUMOR-NECROSIS-FACTOR;
   PROSTAGLANDIN-E SYNTHASE-1; INFLUENZA-VIRUS INFECTION; FOOD-MOTIVATED
   BEHAVIOR; LPS-INDUCED HYPOPHAGIA; KAPPA-B ACTIVATION; RAT-BRAIN;
   SICKNESS BEHAVIOR; TNF-ALPHA
AB Bacterial lipopolysaccharide (LPS) and other microbial substances trigger the organism's acute phase response and cause acute illness anorexia. Pro-inflammatory cytokines are major endogenous mediators of acute illness anorexia, but how LPS or cytokines stimulate the brain to inhibit eating is not fully resolved. One emerging mechanism involves the activation of the enzyme cyclooxygenase-2 (COX-2) in blood-brain barrier endothelial cells and the subsequent release of prostaglandin E2 (PGE2). Serotonin neurons in the midbrain raphe are targets of PGE2, and serotonergic projections from the midbrain raphe to the hypothalamus appear to be crucial for LPS anorexia. That is, raphe projections activate (1) the corticotrophin-releasing hormone neurons in the paraventricular nucleus which then elicit the stress response and (2) the pro-opiomelanocortin neurons in the arcuate nucleus which then release alpha MSH and elicit anorexia. Here we review available data to support a role for this brain mechanism in acute illness anorexia by center staging PGE2 signaling pathways that converge on central neural circuits that control normal eating. In addition, we review interactions between gonadal hormones and immune function that lead to sex differences in acute illness anorexia.
   The paper represents an invited review by a symposium, award winner or keynote speaker at the Society for the Study of Ingestive Behavior [SSIB] Annual Meeting in Portland. July 2009. (C) 2010 Elsevier Inc. All rights reserved.
C1 [Asarian, Lori; Langhans, Wolfgang] Swiss Fed Inst Technol, Inst Food Nutr & Hlth, CH-8603 Schwerzenbach, Switzerland.
C3 Swiss Federal Institutes of Technology Domain; ETH Zurich
RP Langhans, W (通讯作者)，Swiss Fed Inst Technol, Inst Food Nutr & Hlth, Schorenstr 16, CH-8603 Schwerzenbach, Switzerland.
EM
RI Langhans, Wolfgang/C-4073-2016
CR Arsenijevic D, 2000, EUR CYTOKINE NETW, V11, P662
   Asarian L, 2006, INT REV PSYCHIATR, V17, P451, DOI 10.1080/02646830500381450
   Asarian L, 2009, APPETITE, V52, P817
   Avitsur R, 1999, PHARMACOL BIOCHEM BE, V64, P787, DOI 10.1016/S0091-3057(99)00165-3
   AVITSUR R, 1995, BRAIN BEHAV IMMUN, V9, P234, DOI 10.1006/brbi.1995.1022
   Banks WA, 1996, LIFE SCI, V59, P1923, DOI 10.1016/S0024-3205(96)00380-3
   Beutler B, 2003, ANNU REV PHARMACOL, V43, P609, DOI 10.1146/annurev.pharmtox.43.100901.135729
   Beutler B, 2002, CURR TOP MICROBIOL, V270, P109
   BLUTHE RM, 1992, BRAIN RES, V573, P318, DOI 10.1016/0006-8993(92)90779-9
   BREDER CD, 1988, SCIENCE, V240, P321, DOI 10.1126/science.3258444
   BretDibat JL, 1997, BRAIN RES BULL, V42, P443, DOI 10.1016/S0361-9230(96)00370-X
   BRETDIBAT JL, 1995, BRAIN BEHAV IMMUN, V9, P242, DOI 10.1006/brbi.1995.1023
   Butera PC, 2002, BRAIN BEHAV IMMUN, V16, P275, DOI 10.1006/brbi.2001.0621
   Cao CY, 1996, BRAIN RES, V733, P263, DOI 10.1016/0006-8993(96)00575-6
   Cuschieri J, 2004, J SURG RES, V121, P76, DOI 10.1016/j.jss.2004.04.005
   Dallaporta M, 2007, BRAIN BEHAV IMMUN, V21, P1109, DOI 10.1016/j.bbi.2007.05.003
   Dantzer R, 1996, MOL PSYCHIATR, V1, P98
   Duerschner DR, 1943, J INFECT DIS, V72, P183, DOI 10.1093/infdis/72.3.183
   Elander L, 2007, AM J PHYSIOL-REG I, V292, pR258, DOI 10.1152/ajpregu.00511.2006
   Elmquist JK, 1997, TRENDS NEUROSCI, V20, P565, DOI 10.1016/S0166-2236(97)01138-7
   Elmquist JK, 1996, J COMP NEUROL, V374, P315, DOI 10.1002/(SICI)1096-9861(19961021)374:3<315::AID-CNE1>3.0.CO;2-4
   Enomoto N, 1999, AM J PHYSIOL-GASTR L, V277, pG671, DOI 10.1152/ajpgi.1999.277.3.G671
   Ericsson A, 1997, J NEUROSCI, V17, P7166
   Fan W, 1997, NATURE, V385, P165, DOI 10.1038/385165a0
   Geary N, 2004, PHYSIOL BEHAV, V82, P251, DOI 10.1016/j.physbeh.2004.03.034
   Geary N, 2001, NUTRITION, V17, P499, DOI 10.1016/S0899-9007(01)00588-3
   Goehler LE, 1999, J NEUROSCI, V19, P2799
   HART BL, 1988, NEUROSCI BIOBEHAV R, V12, P123, DOI 10.1016/S0149-7634(88)80004-6
   Heisler LK, 2002, SCIENCE, V297, P609, DOI 10.1126/science.1072327
   Heisler LK, 2007, J NEUROSCI, V27, P6956, DOI 10.1523/JNEUROSCI.2584-06.2007
   Heisler LK, 2006, NEURON, V51, P239, DOI 10.1016/j.neuron.2006.06.004
   Hrupka BJ, 2001, PHARMACOL BIOCHEM BE, V68, P355, DOI 10.1016/S0091-3057(00)00463-9
   Huang J, 2004, BRAIN RES, V1028, P156, DOI 10.1016/j.brainres.2004.09.009
   Huang QH, 1999, AM J PHYSIOL-REG I, V276, pR864, DOI 10.1152/ajpregu.1999.276.3.R864
   JANEWAY CA, 1999, HOST DEFENSE INFECT, P316
   Jang PG, 2010, J BIOL CHEM, V285, P9706, DOI 10.1074/jbc.M109.070706
   KATSUURA G, 1990, AM J PHYSIOL, V258, pE163, DOI 10.1152/ajpendo.1990.258.1.E163
   KILPATRICK GJ, 1987, NATURE, V330, P746, DOI 10.1038/330746a0
   KLUGER MJ, 1991, PHYSIOL REV, V71, P93, DOI 10.1152/physrev.1991.71.1.93
   KOMAKI G, 1992, AM J PHYSIOL, V262, pE246, DOI 10.1152/ajpendo.1992.262.2.E246
   Kopf BS, 2010, BRAIN RES, V1306, P77, DOI 10.1016/j.brainres.2009.09.079
   Kurosawa M, 1997, J AUTONOM NERV SYST, V62, P72, DOI 10.1016/S0165-1838(96)00111-7
   Lacroix S, 1996, J NEUROIMMUNOL, V70, P163, DOI 10.1016/S0165-5728(96)00114-2
   Lacroix S, 1997, J COMP NEUROL, V387, P307, DOI 10.1002/(SICI)1096-9861(19971020)387:2<307::AID-CNE11>3.0.CO;2-R
   Laflamme N, 1999, J NEUROSCI, V19, P10923, DOI 10.1523/JNEUROSCI.19-24-10923.1999
   Lahita R G, 2000, J Gend Specif Med, V3, P19
   LANGHANS W, 1993, PHYSIOL BEHAV, V53, P643, DOI 10.1016/0031-9384(93)90168-F
   LANGHANS W, 1989, PHYSIOL BEHAV, V46, P535, DOI 10.1016/0031-9384(89)90032-2
   Langhans W, 1999, NEUROPEPTIDES, V33, P415, DOI 10.1054/npep.1999.0048
   LANGHANS W, 2004, NEUROBIOLOGY FOOD FL, P347
   Langhans W, 2007, P NUTR SOC, V66, P321, DOI 10.1017/S0029665107005587
   Laye S, 2000, AM J PHYSIOL-REG I, V279, pR93, DOI 10.1152/ajpregu.2000.279.1.R93
   Licinio J, 1997, J CLIN INVEST, V100, P2941, DOI 10.1172/JCI119846
   LIPTON JM, 1979, J PHYSIOL-LONDON, V295, P263, DOI 10.1113/jphysiol.1979.sp012967
   Lugarini F, 2002, AM J PHYSIOL-REG I, V283, pR862, DOI 10.1152/ajpregu.00200.2002
   Maier SF, 1998, PSYCHOL REV, V105, P83, DOI 10.1037/0033-295X.105.1.83
   Maier SF, 1998, ANN NY ACAD SCI, V840, P289, DOI 10.1111/j.1749-6632.1998.tb09569.x
   Marks DL, 2003, ENDOCRINOLOGY, V144, P1513, DOI 10.1210/en.2002-221099
   Marks DL, 2001, CANCER RES, V61, P1432
   Martin JT, 2000, EUR J PHARMACOL, V405, P251, DOI 10.1016/S0014-2999(00)00557-4
   Medzhitov R, 2001, NAT REV IMMUNOL, V1, P135, DOI 10.1038/35100529
   Medzhitov R, 2000, IMMUNOL REV, V173, P89, DOI 10.1034/j.1600-065X.2000.917309.x
   Miller BW, 1997, AM J PHYSIOL-CELL PH, V273, pC130, DOI 10.1152/ajpcell.1997.273.1.C130
   Mouihate A, 1998, AM J PHYSIOL-REG I, V275, pR1450, DOI 10.1152/ajpregu.1998.275.5.R1450
   MURAKAMI N, 1987, AM J PHYSIOL, V252, pR284, DOI 10.1152/ajpregu.1987.252.2.R284
   MURRAY MJ, 1979, AM J CLIN NUTR, V32, P593, DOI 10.1093/ajcn/32.3.593
   Nadeau S, 1999, NEUROSCIENCE, V93, P1449, DOI 10.1016/S0306-4522(99)00225-0
   Nadjar A, 2005, NEUROPSYCHOPHARMACOL, V30, P1492, DOI 10.1038/sj.npp.1300755
   Nakamura K, 2004, J THERM BIOL, V29, P339, DOI 10.1016/j.jtherbio.2004.08.001
   Nakamura K, 2002, J NEUROSCI, V22, P4600, DOI 10.1523/JNEUROSCI.22-11-04600.2002
   Nakamura K, 2001, NEUROSCIENCE, V103, P763, DOI 10.1016/S0306-4522(01)00027-6
   Nappi RE, 1997, ENDOCRINOLOGY, V138, P1374, DOI 10.1210/en.138.4.1374
   Niijima A, 1996, J AUTONOM NERV SYST, V61, P287, DOI 10.1016/S0165-1838(96)00098-7
   Ogimoto K, 2006, ENDOCRINOLOGY, V147, P4445, DOI 10.1210/en.2006-0465
   Oka T, 2000, J COMP NEUROL, V428, P20, DOI 10.1002/1096-9861(20001204)428:1<20::AID-CNE3>3.0.CO;2-X
   Pecchi E, 2006, PHYSIOL GENOMICS, V25, P485, DOI 10.1152/physiolgenomics.00306.2005
   Pecchi E, 2009, PHYSIOL BEHAV, V97, P279, DOI 10.1016/j.physbeh.2009.02.040
   PlataSalaman CR, 1995, NUTRITION, V11, P674
   Porter MH, 1998, AM J PHYSIOL-REG I, V275, pR384, DOI 10.1152/ajpregu.1998.275.2.R384
   Porter MH, 1998, AM J PHYSIOL-REG I, V274, pR741, DOI 10.1152/ajpregu.1998.274.3.R741
   Porter MH, 2000, AM J PHYSIOL-REG I, V279, pR2113, DOI 10.1152/ajpregu.2000.279.6.R2113
   RIETSCHEL ET, 1994, FASEB J, V8, P217, DOI 10.1096/fasebj.8.2.8119492
   Rivera HM, 2005, PHYSIOL BEHAV, V86, P331, DOI 10.1016/j.physbeh.2005.08.004
   Rivest S, 1999, MOL PSYCHIATR, V4, P501, DOI 10.1038/sj.mp.4000676
   SALAMANCA S, 1992, PHARMACOL BIOCHEM BE, V43, P953, DOI 10.1016/0091-3057(92)90431-E
   Sanz L, 2000, EMBO J, V19, P1576, DOI 10.1093/emboj/19.7.1576
   Saper CB, 2010, NEURON, V65, P4, DOI 10.1016/j.neuron.2009.12.029
   Schiltz JC, 2003, FRONT BIOSCI-LANDMRK, V8, pS1321, DOI 10.2741/1211
   SCHUURS AHWM, 1990, J STEROID BIOCHEM, V35, P157, DOI 10.1016/0022-4731(90)90270-3
   Schwartz GJ, 1997, AM J PHYSIOL-REG I, V273, pR1193, DOI 10.1152/ajpregu.1997.273.3.R1193
   Serrats J, 2010, NEURON, V65, P94, DOI 10.1016/j.neuron.2009.11.032
   Simons CT, 1998, AM J PHYSIOL-REG I, V275, pR63, DOI 10.1152/ajpregu.1998.275.1.R63
   SIRKO S, 1989, AM J PHYSIOL, V256, pR616, DOI 10.1152/ajpregu.1989.256.3.R616
   Sonti G, 1996, AM J PHYSIOL-REG I, V270, pR1394, DOI 10.1152/ajpregu.1996.270.6.R1394
   STITT JT, 1990, YALE J BIOL MED, V63, P121
   Sunday L, 2006, AM J PHYSIOL-ENDOC M, V291, pE261, DOI 10.1152/ajpendo.00550.2005
   Swiergiel AH, 1999, BRAIN BEHAV IMMUN, V13, P252, DOI 10.1006/brbi.1999.0565
   Swiergiel AH, 1997, PHARMACOL BIOCHEM BE, V57, P389, DOI 10.1016/S0091-3057(96)00335-8
   Swiergiel AH, 2002, J PHARMACOL EXP THER, V302, P1031, DOI 10.1124/jpet.102.036640
   Thaler JP, 2009, ENDOCRINOLOGY, V150, P5362, DOI 10.1210/en.2009-0509
   Turrin NP, 2004, EXP BIOL MED, V229, P996, DOI 10.1177/153537020422901003
   UEHARA A, 1989, AM J PHYSIOL, V257, pR613, DOI 10.1152/ajpregu.1989.257.3.R613
   Ulevitch RJ, 1999, NAT MED, V5, P144, DOI 10.1038/5504
   Ulevitch RJ, 1999, CURR OPIN IMMUNOL, V11, P19, DOI 10.1016/S0952-7915(99)80004-1
   UPHOUSE L, 1991, PHARMACOL BIOCHEM BE, V40, P901, DOI 10.1016/0091-3057(91)90104-A
   von Meyenburg C, 2004, AM J PHYSIOL-REG I, V287, pR298, DOI 10.1152/ajpregu.00659.2003
   von Meyenburg C, 2003, PHARMACOL BIOCHEM BE, V74, P1025, DOI 10.1016/S0091-3057(03)00030-3
   von Meyenburg C, 2003, PHARMACOL BIOCHEM BE, V74, P505, DOI 10.1016/S0091-3057(02)01029-8
   Wachulec M, 1997, AM J PHYSIOL-REG I, V272, pR1252, DOI 10.1152/ajpregu.1997.272.4.R1252
   Wisse BE, 2007, ENDOCRINOLOGY, V148, P5230, DOI 10.1210/en.2007-0394
   Wylie CJ, 2010, J NEUROSCI, V30, P670, DOI 10.1523/JNEUROSCI.4656-09.2010
   Yirmiya R, 1997, BRAIN RES, V749, P71, DOI 10.1016/S0006-8993(96)01295-4
   Zhang J, 1999, EUR J NEUROSCI, V11, P2651, DOI 10.1046/j.1460-9568.1999.00682.x
NR 113
TC 30
Z9 30
U1 0
U2 8
PU PERGAMON-ELSEVIER SCIENCE LTD
PI OXFORD
PA THE BOULEVARD, LANGFORD LANE, KIDLINGTON, OXFORD OX5 1GB, ENGLAND
SN 0031-9384
J9 PHYSIOL BEHAV
JI Physiol. Behav.
PD JUL 14
PY 2010
VL 100
IS 5
SI SI
BP 464
EP 471
DI 10.1016/j.physbeh.2010.04.009
PG 8
WC Psychology, Biological; Behavioral Sciences
WE Science Citation Index Expanded (SCI-EXPANDED); Social Science Citation Index (SSCI); Conference Proceedings Citation Index - Science (CPCI-S); Conference Proceedings Citation Index - Social Science &amp; Humanities (CPCI-SSH)
SC Psychology; Behavioral Sciences
GA 626SV
UT WOS:000279988100007
PM 20394763
DA 2023-08-27
ER

PT J
AU Hochart, A
   Gaillard, V
   Baroncini, M
   Andre, N
   Vannier, JP
   Vinchon, M
   Dubrulle, F
   Lejeune, JP
   Vincent, C
   Neve, V
   Sudour Bonnange, H
   Bonne, NX
   Leblond, P
AF Hochart, Audrey
   Gaillard, Vianney
   Baroncini, Marc
   Andre, Nicolas
   Vannier, Jean-Pierre
   Vinchon, Matthieu
   Dubrulle, Frederique
   Lejeune, Jean-Paul
   Vincent, Christophe
   Neve, Veronique
   Sudour Bonnange, Helene
   Bonne, Nicolas Xavier
   Leblond, Pierre
TI Bevacizumab decreases vestibular schwannomas growth rate in children and
   teenagers with neurofibromatosis type 2
SO JOURNAL OF NEURO-ONCOLOGY
LA English
DT Article
DE Neurofibromatosis type 2; Vestibular schwannoma; Pediatric; Bevacizumab
ID HISTONE DEACETYLASE INHIBITOR; PHASE-II; THERAPY; AR-42; DRUG; VEGF
AB Vestibular schwannoma (VS) growth in neurofibromatosis type 2 (NF2) can be responsible for brainstem compression and hearing loss. Surgical removal remains the standard therapy despite potential morbidity. Previous studies suggested that the inhibition of the VEGF-pathway with bevacizumab could result in hearing improvement, reduction of the tumor volume or both in adults. We retrospectively describe the French experience of bevacizumab treatment delivered for progressive VS in pediatric NF2 patients. Patients received Bevacizumab 5 or 10 mg/kg every 2 weeks according to the physician's choice. Follow-up included clinical assessment, audiometry and volumetric MRI every 3-6 months. Seven patients harboring 11 VS were included. The median age at inclusion was 15 years (11.4-18.8), and the median treatment duration was 11.3 months (3.2-55.6). At baseline, the median tumor volume was 1.2 cm(3) (0.52-13.5) and the median word recognition score was 90 % (0-100). We observed one major response, two minor responses and a decrease in the rate of tumor growth for the 4 other patients. The median annual growth rate before treatment was significantly higher than after 1 year of treatment (138 vs. 36 %, n = 5, p = 0.043). We noted one hearing improvement over the course of 1 year under treatment (hearing response rate was 14 %). Overall, the treatment was well tolerated. Our study supports that bevacizumab is an attractive therapeutic option for pediatric NF2 patients with growing VS. Thorough multidisciplinary evaluation is necessary to identify the best candidates prior to treatment. It is likely that a better functional outcome would be expected if targeted therapies were discussed early in the management of the disease.
C1 [Hochart, Audrey; Sudour Bonnange, Helene; Leblond, Pierre] Ctr Oscar Lambret, Pediat Oncol Unit, F-59020 Lille, France.
   [Gaillard, Vianney; Dubrulle, Frederique] Univ Hosp, C Huriez Hosp, Dept Radiol, Lille, France.
   [Baroncini, Marc; Vinchon, Matthieu; Lejeune, Jean-Paul] Lille Univ Hosp, Dept Pediat Neurosurg, Lille, France.
   [Andre, Nicolas] La Timone Hosp, Dept Pediat Hematol & Oncol, Marseille, France.
   [Andre, Nicolas] Aix Marseille Univ, Ctr Rech Oncol Biol & Oncopharmacol, INSERM UMR 911, Marseille, France.
   [Vannier, Jean-Pierre] Charles Nicolle Hosp, Dept Pediat Hematol & Oncol, Rouen, France.
   [Vincent, Christophe; Bonne, Nicolas Xavier] Lille Univ Hosp, R Salengro Hosp, Dept Otol & Neurootol, Lille, France.
   [Neve, Veronique] Lille Univ Hosp, Pediat Pulm Funct Testing Unit, Lille, France.
C3 UNICANCER; Centre Oscar Lambret; Universite de Lille - ISITE; CHU Lille;
   Universite de Lille - ISITE; CHU Lille; UDICE-French Research
   Universities; Aix-Marseille Universite; Assistance Publique-Hopitaux de
   Marseille; Institut National de la Sante et de la Recherche Medicale
   (Inserm); UDICE-French Research Universities; Aix-Marseille Universite;
   Universite de Rouen Normandie; CHU de Rouen; Universite de Lille -
   ISITE; CHU Lille; Universite de Lille - ISITE; CHU Lille
RP Hochart, A (通讯作者)，Ctr Oscar Lambret, Pediat Oncol Unit, 3 Rue F Combemale, F-59020 Lille, France.
EM
RI Vincent, Christophe/F-6261-2016
OI Vincent, Christophe/0000-0003-3201-7213; ANDRE,
   Nicolas/0000-0002-6786-4968; LEBLOND, Pierre/0000-0002-7088-2614;
   Vinchon, Matthieu/0000-0002-9122-6165; Bonne,
   Nicolas-Xavier/0000-0003-4985-0869
CR Ammoun S, 2011, NEURO-ONCOLOGY, V13, P759, DOI 10.1093/neuonc/nor056
   [Anonymous], 1995, Otolaryngol Head Neck Surg, V113, P179
   Baser ME, 2002, AM J HUM GENET, V71, P715, DOI 10.1086/342716
   Blakeley Jaishri O, 2012, Am J Med Genet A, V158A, P24, DOI 10.1002/ajmg.a.34359
   Bush ML, 2011, NEURO-ONCOLOGY, V13, P983, DOI 10.1093/neuonc/nor072
   Caye-Thomasen P, 2005, OTOL NEUROTOL, V26, P98, DOI 10.1097/00129492-200501000-00017
   Evans DGR, 2005, OTOL NEUROTOL, V26, P93, DOI 10.1097/00129492-200501000-00016
   Giovannini M, 2014, NEURO-ONCOLOGY, V16, P493, DOI 10.1093/neuonc/not242
   Harris GJ, 2008, NEUROSURGERY, V62, P1314, DOI 10.1227/01.neu.0000333303.79931.83
   Jacob A, 2012, LARYNGOSCOPE, V122, P174, DOI 10.1002/lary.22392
   Karajannis MA, 2014, NEURO-ONCOLOGY, V16, P292, DOI 10.1093/neuonc/not150
   Karajannis MA, 2012, NEURO-ONCOLOGY, V14, P1163, DOI 10.1093/neuonc/nos146
   Mathieu D, 2007, NEUROSURGERY, V60, P460, DOI 10.1227/01.NEU.0000255340.26027.53
   Mautner VF, 2010, NEURO-ONCOLOGY, V12, P14, DOI 10.1093/neuonc/nop010
   Plotkin SR, 2012, OTOL NEUROTOL, V33, P1046, DOI 10.1097/MAO.0b013e31825e73f5
   Plotkin SR, 2010, OTOL NEUROTOL, V31, P1135, DOI 10.1097/MAO.0b013e3181eb328a
   Plotkin SR, 2009, J NEURO-ONCOL, V93, P61, DOI 10.1007/s11060-009-9867-7
   Plotkin SR, 2009, NEW ENGL J MED, V361, P358, DOI 10.1056/NEJMoa0902579
   Shord SS, 2009, AM J HEALTH-SYST PH, V66, P999, DOI 10.2146/ajhp080455
   Slattery WH, 2004, OTOL NEUROTOL, V25, P811, DOI 10.1097/00129492-200409000-00027
   Slusarz KM, 2014, CANCER CHEMOTH PHARM, V73, P1197, DOI 10.1007/s00280-014-2456-2
   Subbiah V, 2012, J CLIN ONCOL, V30, pE64, DOI 10.1200/JCO.2011.38.2614
   Uesaka T, 2007, J NEURO-ONCOL, V83, P259, DOI 10.1007/s11060-007-9336-0
NR 23
TC 37
Z9 37
U1 0
U2 10
PU SPRINGER
PI NEW YORK
PA 233 SPRING ST, NEW YORK, NY 10013 USA
SN 0167-594X
EI 1573-7373
J9 J NEURO-ONCOL
JI J. Neuro-Oncol.
PD SEP
PY 2015
VL 124
IS 2
BP 229
EP 236
DI 10.1007/s11060-015-1828-8
PG 8
WC Oncology; Clinical Neurology
WE Science Citation Index Expanded (SCI-EXPANDED)
SC Oncology; Neurosciences & Neurology
GA CS2FJ
UT WOS:000361883000010
PM 26022982
DA 2023-08-27
ER

PT J
AU Vernon, V
   Naik, H
   Guha, A
AF Vernon, Velho
   Naik, Harish
   Guha, Amrita
TI Surgical management of cerebellopontine angle epidermoid cysts: an
   institutional experience of 10 years
SO BRITISH JOURNAL OF NEUROSURGERY
LA English
DT Article
DE Epidermoid cysts; cerebellopontine angle; retrosigmoidal approach;
   subtemporal approach; endoscope-assisted microsurgical technique
AB Background: Cerebellopontine angle (CPA) epidermoids, although of benign nature, are of considerable neurosurgical interest because of their close proximity and adherence to the cranial nerves and brain stem. In this paper, we describe our experience and attempt to correlate the final outcomes with the extent of surgical removal. The main objectives were to study various modes of surgical management of CPA epidermoids with regard to removal and preservation of the cranial nerves and also to evaluate the role of endoscopic assisted microsurgical excision thereby minimizing recurrences. This case series is one of the largest series reported so far worldwide.
   Materials and methods: From 2006 to 2016, 139 patients with CPA epidermoids were operated at Grant Medical College and J. J. Hospital, Mumbai. All patients underwent detailed magnetic resonance imaging (MRI) of brain. Lesions were classified according Rogelio Revuelta-Gutierrez et al. with respect to their anatomic extent: grade I- within the boundaries of the CPA, grade II- extension to the suprasellar and perimesencephalic cisterns, and grade III-parasellar and temporomesial region involvement. Retrosigmoidal and sub temporal approaches were taken to excise the lesions. Endoscopic assisted microsurgical excision was done in cases with extensions beyond the CPA. Patient follow-up was based on outpatient repeated brain MRI studies.
   Results: The mean duration of symptoms before surgery was 42 months (range, 2 months to 6 years). The mean follow-up period was 27 months (range, 2-60 months). The main presenting sympt om was headache in 69% (96/139) of the cases and trigeminal neuralgia in 30% cases was the second most common cause of consultation. Seventy-five percent of patients had some degree of cranial nerve (CN) involvement. Retrosigmoid approach was taken in 92% patients and 7 patients with supratentorial extension were operated by combined retrosigmoidal and subtemporal approach. Endoscopic assisted microsurgical excision was done in 40% cases. Use of angled views by an endoscope helped to excise residual tumor in 47 (83%) patients. Complete excision was achieved in 67% of cases. In 33% patients, small capsular remnants could not be removed completely because of their adherence to vessels, brainstem and cranial nerves. Compared with their preoperative clinical status, 74% improved and 20% had persistent cranial nerve deficits in the first year of follow up.
   Conclusions: Epidermoid cysts are challenging entities in current neurosurgery practice due to tumor adhesions to neurovascular structures. Meticulous surgical technique with the aid of neurophysiological monitoring is crucial to achieve safe and effective total or subtotal removal of these lesions. A conservative approach is indicated for patients in whom the fragments of capsule is adhered closely to blood vessels, nerves, or the brainstem, in order to avoid risk of serious neurological deficits related to an inadvertent damage of these structures. Use of angled views by endoscope at the conclusion of the surgery may assure the surgeon of total removal of the tumor.
C1 [Vernon, Velho; Naik, Harish] JJ Grp Hosp & Grant Med Coll, Dept Neurosurg, Mumbai 400008, Maharashtra, India.
   [Guha, Amrita] Tata Mem Hosp, Dept Radiol, Mumbai, Maharashtra, India.
C3 Tata Memorial Centre (TMC); Tata Memorial Hospital
RP Naik, H (通讯作者)，JJ Grp Hosp & Grant Med Coll, Dept Neurosurg, Mumbai 400008, Maharashtra, India.
EM
CR Akhavan-Sigari R, 2007, ACTA NEUROCHIR, V149, P429, DOI 10.1007/s00701-007-1117-1
   ALTSCHULER EM, 1990, NEUROSURGERY, V26, P606, DOI 10.1227/00006123-199004000-00008
   Aprile I, 1999, AM J NEURORADIOL, V20, P1259
   Apuzzo MLJ., 1993, BRAIN SURG COMPLICAT, P568
   BERGER MS, 1985, J NEUROSURG, V62, P214, DOI 10.3171/jns.1985.62.2.0214
   David EA, 2003, OTOL NEUROTOL, V24, P699, DOI 10.1097/00129492-200307000-00028
   DESOUZA CE, 1989, J NEUROL NEUROSUR PS, V52, P986, DOI 10.1136/jnnp.52.8.986
   DIAS MS, 1992, PEDIATR NEUROSURG, V18, P229, DOI 10.1159/000120670
   FEIN JM, 1981, NEUROSURGERY, V9, P179, DOI 10.1227/00006123-198108000-00014
   Fries G, 1998, NEUROSURGERY, V42, P226, DOI 10.1097/00006123-199802000-00008
   GAO PY, 1992, AM J NEURORADIOL, V13, P863
   Gopalakrishnan CV, 2014, CLIN NEUROL NEUROSUR, V117, P93, DOI 10.1016/j.clineuro.2013.11.025
   GUIDETTI B, 1977, J NEUROSURG, V47, P12, DOI 10.3171/jns.1977.47.1.0012
   Hasegawa M, 2016, NEUROSURG REV, V39, P259, DOI 10.1007/s10143-015-0684-5
   Kato Koichi, 2010, Asian J Neurosurg, V5, P32
   Kobata H, 2002, NEUROSURGERY, V50, P276, DOI 10.1097/00006123-200202000-00008
   Lewis D., 1945, PRACTICE OF SURG, P628
   Love JG, 1936, J AMER MED ASSOC, V107, P1876, DOI 10.1001/jama.1936.02770490030009
   LUNARDI P, 1990, ACTA NEUROCHIR, V103, P105, DOI 10.1007/BF01407514
   Mallucci CL, 1999, J NEUROL NEUROSUR PS, V66, P768, DOI 10.1136/jnnp.66.6.768
   Mohanty A, 1997, NEUROSURGERY, V40, P24, DOI 10.1097/00006123-199701000-00004
   NIIKAWA S, 1992, CHILD NERV SYST, V8, P453, DOI 10.1007/BF00274406
   OGLEZNEV KY, 1991, ACTA NEUROCHIR, V110, P116, DOI 10.1007/BF01400677
   Osborn, 1991, HDB NEURORADIOLOGY
   RAPPAPORT ZH, 1985, NEUROCHIRURGIA, V28, P211
   REVUELTA R, 1995, NEUROSURGERY, V37, P138, DOI 10.1227/00006123-199507000-00022
   Revuelta-Gutierrez R, 2009, CIR CIR, V77, P241
   RUBIN G, 1989, ACTA NEUROCHIR, V97, P1, DOI 10.1007/BF01577734
   SABIN HI, 1987, NEUROSURGERY, V21, P798, DOI 10.1227/00006123-198712000-00004
   Safavi-Abbasi S, 2008, SKULL BASE-INTERD AP, V18, P85, DOI 10.1055/s-2007-991108
   SALAZAR J, 1987, ACTA NEUROCHIR, V85, P34, DOI 10.1007/BF01402367
   Samii M, 1996, J NEUROSURG, V84, P14, DOI 10.3171/jns.1996.84.1.0014
   Schiefer TK, 2008, SURG NEUROL, V70, P584, DOI 10.1016/j.surneu.2007.12.021
   Schroeder HWS, 2004, J NEUROSURG, V101, P227, DOI 10.3171/jns.2004.101.2.0227
   Talacchi A, 1998, NEUROSURGERY, V42, P242, DOI 10.1097/00006123-199802000-00020
   VINCHON M, 1995, NEUROSURGERY, V36, P52, DOI 10.1227/00006123-199501000-00006
   YAMAKAWA K, 1989, NEUROSURGERY, V24, P568, DOI 10.1227/00006123-198904000-00013
   YASARGIL MG, 1989, NEUROSURGERY, V24, P561
NR 38
TC 6
Z9 7
U1 0
U2 2
PU TAYLOR & FRANCIS LTD
PI ABINGDON
PA 2-4 PARK SQUARE, MILTON PARK, ABINGDON OR14 4RN, OXON, ENGLAND
SN 0268-8697
EI 1360-046X
J9 BRIT J NEUROSURG
JI Br. J. Neurosurg.
PD MAR 4
PY 2022
VL 36
IS 2
BP 203
EP 212
DI 10.1080/02688697.2020.1867058
EA JAN 2021
PG 10
WC Clinical Neurology; Surgery
WE Science Citation Index Expanded (SCI-EXPANDED)
SC Neurosciences & Neurology; Surgery
GA 2E1LL
UT WOS:000605724500001
PM 33410366
DA 2023-08-27
ER

PT J
AU Martin-Vaquero, P
   da Costa, RC
   Aeffner, F
   Oglesbee, MJ
   Echandi, RL
AF Martin-Vaquero, Paula
   da Costa, Ronaldo C.
   Aeffner, Famke
   Oglesbee, Michael J.
   Echandi, Rita L.
TI IMAGING DIAGNOSIS-HEMORRHAGIC MENINGIOMA
SO VETERINARY RADIOLOGY & ULTRASOUND
LA English
DT Article
DE acute hemorrhage; dog; intracranial neoplasia; magnetic resonance
   imaging
ID DOGS; APPEARANCE; FEATURES; TUMORS; BRAIN
AB An 8-year-old Labrador Retriever developed acute central vestibular signs. An extra-axial mass was detected on MR images ventral to the brainstem. The mass was both T1- and T2-hypointense; there was also thin-rimmed patchy contrast enhancement. These findings were nonspecific, but the extreme T2-hypointensity was notable and suggested a hemorrhagic mass. The histologic diagnosis was anaplastic meningioma with acute hemorrhage. These findings document an unusual appearance of a meningioma in MR images due to intratumoral hemorrhage.
C1 [Martin-Vaquero, Paula; da Costa, Ronaldo C.; Echandi, Rita L.] Ohio State Univ, Coll Vet Med, Dept Vet Sci, Columbus, OH 43210 USA.
   [Aeffner, Famke; Oglesbee, Michael J.] Ohio State Univ, Coll Vet Med, Dept Biosci, Columbus, OH 43210 USA.
C3 University System of Ohio; Ohio State University; University System of
   Ohio; Ohio State University
RP Martin-Vaquero, P (通讯作者)，Ohio State Univ, Coll Vet Med, Dept Vet Sci, Columbus, OH 43210 USA.
EM
OI CASIMIRO DA COSTA, RONALDO/0000-0002-4034-9800; Aeffner,
   Famke/0000-0002-1405-9228
CR BRADLEY WG, 1993, RADIOLOGY, V189, P15, DOI 10.1148/radiology.189.1.8372185
   BUETOW MP, 1991, RADIOGRAPHICS, V11, P1087, DOI 10.1148/radiographics.11.6.1749851
   CHEN TC, 1992, NEUROSURGERY, V31, P1015, DOI 10.1227/00006123-199212000-00005
   Hasegawa D, 2008, J VET MED SCI, V70, P615, DOI 10.1292/jvms.70.615
   Hathcock JT, 1996, VET RADIOL ULTRASOUN, V37, P257, DOI 10.1111/j.1740-8261.1996.tb01227.x
   KAPLAN RD, 1992, J COMPUT ASSIST TOMO, V16, P366, DOI 10.1097/00004728-199205000-00006
   Kraft SL, 1997, J VET INTERN MED, V11, P218, DOI 10.1111/j.1939-1676.1997.tb00094.x
   Kubota Yoshinori, 1997, Neurologia Medico-Chirurgica, V37, P407, DOI 10.2176/nmc.37.407
   LeCouteur R. A., 1996, SMALL ANIMAL CLIN ON, P393
   Shaman MA, 2003, RADIOGRAPHICS, V23, P785, DOI 10.1148/rg.233025122
   Snyder JM, 2006, J VET INTERN MED, V20, P669, DOI 10.1892/0891-6640(2006)20[669:CIPNC]2.0.CO;2
   Sturges BK, 2008, J VET INTERN MED, V22, P586, DOI 10.1111/j.1939-1676.2008.00042.x
   Thomas WB, 1996, VET RADIOL ULTRASOUN, V37, P20, DOI 10.1111/j.1740-8261.1996.tb00807.x
   Thomas WB, 1997, VET RADIOL ULTRASOUN, V38, P371, DOI 10.1111/j.1740-8261.1997.tb02100.x
   Wessmann A, 2009, VET J, V180, P290, DOI 10.1016/j.tvjl.2007.12.023
   ZEE CS, 1992, SEMIN ULTRASOUND CT, V13, P154
NR 16
TC 7
Z9 7
U1 0
U2 2
PU WILEY-BLACKWELL PUBLISHING, INC
PI MALDEN
PA COMMERCE PLACE, 350 MAIN ST, MALDEN 02148, MA USA
SN 1058-8183
J9 VET RADIOL ULTRASOUN
JI Vet. Radiol. Ultrasound
PD MAR-APR
PY 2010
VL 51
IS 2
BP 165
EP 167
DI 10.1111/j.1740-8261.2009.01645.x
PG 3
WC Veterinary Sciences
WE Science Citation Index Expanded (SCI-EXPANDED)
SC Veterinary Sciences
GA 566SM
UT WOS:000275393400013
PM 20402404
DA 2023-08-27
ER

PT J
AU Ladzinski, P
   Majchrzak, H
   Kaspera, W
   Maliszewski, M
   Majchrzak, K
   Tymowski, M
   Adamczyk, P
   Blaszczyk, B
   Slaska-Kaspera, A
AF Ladzinski, Piotr
   Majchrzak, Henryk
   Kaspera, Wojciech
   Maliszewski, Mariusz
   Majchrzak, Krzysztof
   Tymowski, Michal
   Adamczyk, Piotr
   Blaszczyk, Bartlomiej
   Slaska-Kaspera, Aleksandra
TI Meningiomas of the upper and middle part of the clivus and surrounding
   structures: early and long-term outcome
SO NEUROLOGIA I NEUROCHIRURGIA POLSKA
LA English
DT Article
DE clivus; petrous pyramid; meningioma; microsurgery; skull base surgery
ID PETROCLIVAL MENINGIOMAS; TRANSTENTORIAL APPROACH; PETROSAL APPROACH;
   EXPERIENCE; RECURRENCE; RESECTION; REMOVAL
AB Background and purpose: Meningiomas of the upper and middle parts of the clivus and surrounding structures are removed using petrosal approaches: anterior, posterior, combined and complete. The purpose of this study is to show the results of treatment of these meningiomas and to present our interpretation of the treatment strategy
   Material and methods: Twenty-six patients (17 women, 9 men) were included in the study The neurological status of the patients was assessed before and after surgery as well as at the conclusion of the treatment. The following measurements and data were collected and recorded: approximate volume of the treated lesion, its relation to large blood vessels, cranial nerves and the brainstem, as well as tumour consistency and vascularisation.
   Results: Symptoms duration ranged from 1 to 60 months (median: 16 months). In 57.7% of patients, imbalance was the predominant sign. Less frequent symptoms were: headaches, dysacusis and hemiparesis. Approximate volumes of the tumours ranged from 4 to 65 mL (mean: 32 mL). Total or subtotal resection was achieved in 73.1% of patients. The patients' performance improved postoperatively in 34.5%, remained unchanged in 46.2% and deteriorated in 11.5% of patients. Two (7.8%) patients died after the surgery.
   Conclusions: The use of petrosal approaches in the surgical treatment of meningiomas of the upper and middle parts of the clivus and the surrounding structures facilitates good or at least satisfactory neurological outcome with a high proportion of complete resections and relatively low mortality
C1 [Ladzinski, Piotr; Majchrzak, Henryk; Kaspera, Wojciech; Maliszewski, Mariusz; Majchrzak, Krzysztof; Tymowski, Michal; Adamczyk, Piotr; Blaszczyk, Bartlomiej] Slaski Uniwersytet Med Katowicach, Katedra & Oddzial Klini Neurochirurg, PL-41200 Sosnowiec, Poland.
   [Slaska-Kaspera, Aleksandra] Slaski Uniwersytet Med Katowicach, Katedra & Klin Laryngol, PL-41200 Sosnowiec, Poland.
C3 Medical University Silesia; Medical University Silesia
RP Ladzinski, P (通讯作者)，Slaski Uniwersytet Med Katowicach, Katedra & Oddzial Klini Neurochirurg, Wojewodzki Szpital Specjalisty Barbary 5, Pl Medykow 1, PL-41200 Sosnowiec, Poland.
EM
RI Kaspera, Wojciech/AAF-2715-2019
OI Kaspera, Wojciech/0000-0002-7139-3409; Majchrzak,
   Krzysztof/0000-0003-4352-9337; Ladzinski, Piotr/0000-0001-8957-0162;
   Slaska-Kaspera, Aleksandra/0000-0002-4243-7847; Blaszczyk,
   Bartlomiej/0000-0001-5648-8138
CR Al-Mefty O., 1997, OPERATIVE ATLAS MENI, P209
   ALMEFTY O, 1988, NEUROSURGERY, V22, P510, DOI 10.1227/00006123-198803000-00010
   Aziz KMA, 2000, NEUROSURGERY, V47, P139, DOI 10.1097/00006123-200007000-00030
   Barnett S.L., 2008, MENINGIOMAS, P425
   Cho CW, 2002, NEUROSURGERY, V51, P708, DOI 10.1097/00006123-200209000-00017
   CIRIC I, 1993, SURG NEUROL, V39, P530, DOI 10.1016/0090-3019(93)90040-8
   Couldwell WT, 1996, J NEUROSURG, V84, P20, DOI 10.3171/jns.1996.84.1.0020
   Javed T, 1991, Acta Neurochir Suppl (Wien), V53, P171
   KAWASE T, 1994, ACTA NEUROCHIR, V129, P113, DOI 10.1007/BF01406489
   Kawase T, 1996, NEUROSURGERY, V39, P527, DOI 10.1097/00006123-199609000-00019
   KAWASE T, 1991, NEUROSURGERY, V28, P869, DOI 10.1227/00006123-199106000-00014
   Kawase T, 1999, OP TECH NEUROSURG, V2, P10
   Ladzinski P., 2011, NEUROL NEUROCHIR POL, V46, P205
   Ladzinski P, 2010, NEUROL NEUROCHIR POL, V44, P492
   Little KM, 2005, NEUROSURGERY, V56, P546, DOI 10.1227/01.NEU.0000153906.12640.62
   Mathiesen T, 1996, NEUROSURGERY, V39, P2, DOI 10.1097/00006123-199607000-00002
   MAYBERG MR, 1986, J NEUROSURG, V65, P160, DOI 10.3171/jns.1986.65.2.0160
   Nassif PS, 1997, SKULL BASE SURG, V7, P5, DOI 10.1055/s-2008-1058618
   SAMII M, 1992, ACTA NEUROCHIR, V118, P27, DOI 10.1007/BF01400723
   SAMII M, 1989, NEUROSURGERY, V24, P12, DOI 10.1227/00006123-198901000-00003
   Samii M, 2001, NEUROSURGERY, V49, P216, DOI 10.1097/00006123-200107000-00036
   Sekhar Laligam N., 1993, P605
   SEKHAR LN, 1990, NEUROSURGERY, V27, P764, DOI 10.1227/00006123-199011000-00015
   SEKHAR LN, 1994, J NEUROSURG, V81, P860, DOI 10.3171/jns.1994.81.6.0860
   SIMPSON D, 1957, J NEUROL NEUROSUR PS, V20, P22, DOI 10.1136/jnnp.20.1.22
   THEDINGER BA, 1992, AM J OTOL, V13, P408
   Van Havenbergh T, 2003, NEUROSURGERY, V52, P55, DOI 10.1097/00006123-200301000-00006
   Ware M.L., 2008, MENINGIOMAS, P403
   YASARGIL MG, 1980, ADV TECHNICAL STANDA, P1
NR 29
TC 0
Z9 0
U1 0
U2 3
PU TERMEDIA PUBLISHING HOUSE LTD
PI POZNAN
PA WENEDOW ST 9-1, POZNAN, 61-614, POLAND
SN 0028-3843
J9 NEUROL NEUROCHIR POL
JI Neurol. Neurochir. Pol.
PD JUL-AUG
PY 2013
VL 47
IS 4
BP 332
EP 344
DI 10.5114/ninp.2013.36758
PG 13
WC Clinical Neurology
WE Science Citation Index Expanded (SCI-EXPANDED)
SC Neurosciences & Neurology
GA 214RQ
UT WOS:000324153900005
PM 23986423
OA Green Submitted
DA 2023-08-27
ER

PT J
AU Ng, WH
   Mukhida, K
   Rutka, JT
AF Ng, Wai Hoe
   Mukhida, Karim
   Rutka, James T.
TI Image guidance and neuromonitoring in neurosurgery
SO CHILDS NERVOUS SYSTEM
LA English
DT Article
DE Neuromonitoring; Neuronavigation; Image guidance; DTI; fMRI; MEG;
   Neurophysiology
ID MONITORED CONSCIOUS SEDATION; PEDIATRIC EPILEPSY SURGERY; SPINAL-CORD
   TUMORS; FIBER TRACKING; WHITE-MATTER; BRAIN SHIFT;
   FUNCTIONAL-NEURONAVIGATION; ELECTRICAL-STIMULATION; LOCAL-ANESTHESIA;
   AWAKE CRANIOTOMY
AB The localization of tumors and epileptogenic foci within the somatosensory or language cortex of the brain of a child poses unique neurosurgical challenges. In the past, lesions in these regions were not treated aggressively for fear of inducing neurological deficits. As a result, while function may have been preserved, the underlying disease may not have been optimally treated, and repeat neurosurgical procedures were frequently required. Today, with the advent of preoperative brain mapping, image guidance or neuronavigation, and intraoperative monitoring, peri-Rolandic and language cortex lesions can be approached directly and definitively with a high degree of confidence that neurosurgical function will be maintained.
   The preoperative brain maps can now be achieved with magnetic resonance imaging (MRI), functional MRI, magnetoencephalography, and diffusion tensor imaging. Image guidance systems have improved significantly and include the use of the intraoperative MRI. Somatosensory, motor, and brainstem auditory-evoked potentials are used as standard neuromonitoring techniques in many centers around the world. Added to this now is the use of continuous train-of-five monitoring of the integrity of the corticospinal tract while operating in the peri-Rolandic region.
   We are in an era where continued advancements can be expected in mapping additional pathways such as visual, memory, and hearing pathways. With these new advances, neurosurgeons can expect to significantly improve their surgical outcomes further.
C1 [Mukhida, Karim; Rutka, James T.] Univ Toronto, Hosp Sick Children, Div Neurosurg, Toronto, ON M5G 1X8, Canada.
   [Ng, Wai Hoe] Natl Inst Neurosci, Dept Neurosurg, Singapore, Singapore.
C3 University of Toronto; Hospital for Sick Children (SickKids); National
   Neuroscience Institute (NNI)
RP Rutka, JT (通讯作者)，Univ Toronto, Hosp Sick Children, Div Neurosurg, Suite 1503,555 Univ Ave, Toronto, ON M5G 1X8, Canada.
EM
FU Hospital for Sick Children in Toronto; Singapore Health Services;
   National Neuroscience Institute (Singapore)
FX This work was made possible through funds from the Wiley Fund and Jack
   Beqaj Fund at the Hospital for Sick Children in Toronto. WII Ng is
   funded by the Singapore Health Services Health Manpower Development
   Programme (HMDP) Fellowship and National Neuroscience Institute
   Scholarship (Singapore).
CR [Anonymous], T AM NEUROL ASS
   Basser PJ, 2000, MAGNET RESON MED, V44, P625, DOI 10.1002/1522-2594(200010)44:4<625::AID-MRM17>3.0.CO;2-O
   Benke T, 2006, EPILEPSIA, V47, P1308, DOI 10.1111/j.1528-1167.2006.00549.x
   Berger M S, 1995, Clin Neurosurg, V42, P437
   Berman JI, 2004, J NEUROSURG, V101, P66, DOI 10.3171/jns.2004.101.1.0066
   Black PM, 1997, NEUROSURGERY, V41, P831, DOI 10.1097/00006123-199710000-00013
   Bowyer SM, 2005, EPILEPSY BEHAV, V6, P235, DOI 10.1016/j.yebeh.2004.12.002
   Breier JI, 2000, J CLIN NEUROPHYSIOL, V17, P503, DOI 10.1097/00004691-200009000-00010
   Breier JI, 1999, NEUROLOGY, V53, P938, DOI 10.1212/WNL.53.5.938
   Buchfelder M, 2002, EPILEPSIA, V43, P864, DOI 10.1046/j.1528-1157.2002.46201.x
   Buchfelder M, 2000, J MAGN RESON IMAGING, V12, P547
   Bucholz RD, 1997, LECT NOTES COMPUT SC, V1205, P459, DOI 10.1007/BFb0029268
   Carter TJ, 2005, MED ENG PHYS, V27, P893, DOI 10.1016/j.medengphy.2005.10.005
   Catani M, 2003, BRAIN, V126, P2093, DOI 10.1093/brain/awg203
   Ciccarelli O, 2003, NEUROIMAGE, V18, P348, DOI 10.1016/S1053-8119(02)00042-3
   Clark CA, 2003, NEUROIMAGE, V20, P1601, DOI 10.1016/j.neuroimage.2003.07.022
   Comeau RM, 2000, MED PHYS, V27, P787, DOI 10.1118/1.598942
   Conturo TE, 1999, P NATL ACAD SCI USA, V96, P10422, DOI 10.1073/pnas.96.18.10422
   Danks RA, 2000, J NEURO-ONCOL, V49, P131, DOI 10.1023/A:1026577518902
   Danks RA, 1998, NEUROSURGERY, V42, P28, DOI 10.1097/00006123-199801000-00006
   EMERSON RG, 1993, J CLIN NEUROPHYSIOL, V10, P404, DOI 10.1097/00004691-199310000-00002
   FOX PT, 1986, P NATL ACAD SCI USA, V83, P1140, DOI 10.1073/pnas.83.4.1140
   GALLEN CC, 1995, J NEUROSURG, V82, P988, DOI 10.3171/jns.1995.82.6.0988
   GALLEN CC, 1993, NEUROSURGERY, V33, P260, DOI 10.1097/00006123-199308000-00012
   Gasser T, 2005, NEUROIMAGE, V26, P685, DOI 10.1016/j.neuroimage.2005.02.022
   Grondin R, 2006, CHILD NERV SYST, V22, P779, DOI 10.1007/s00381-006-0124-5
   HAGLUND MM, 1996, TECH NEUROSURG, V2, P141
   Hagmann P, 2003, NEUROIMAGE, V19, P545, DOI 10.1016/S1053-8119(03)00142-3
   Hammoud MA, 1996, J NEUROSURG, V84, P737, DOI 10.3171/jns.1996.84.5.0737
   Hartov A, 2008, NEUROSURGERY, V63, P91, DOI [10.1227/01.neu.0000317377.15196.45, 10.1227/01.NEU.0000296955.22901.8F]
   Hirata M, 2004, NEUROIMAGE, V23, P46, DOI 10.1016/j.neuroimage.2004.05.009
   Hirsch J, 2000, NEUROSURGERY, V47, P711, DOI 10.1097/00006123-200009000-00037
   Isaacson B, 2003, OTOL NEUROTOL, V24, P812, DOI 10.1097/00129492-200309000-00020
   Jellison BJ, 2004, AM J NEURORADIOL, V25, P356
   Jodicke A, 1998, MINIM INVAS NEUROSUR, V41, P13, DOI 10.1055/s-2008-1052008
   Kaibara T, 2002, EPILEPSIA, V43, P425, DOI 10.1046/j.1528-1157.2002.32401.x
   Kamada Kyousuke, 2005, Neurosurgery, V56, P98
   Karatas AE, 2004, J CLIN NEUROSCI, V11, P343, DOI 10.1016/j.jocn.2003.03.005
   Kelly JJ, 2005, NEUROSURG CLIN N AM, V16, P173, DOI 10.1016/j.nec.2004.07.006
   Kim JS, 2008, NEUROIMAGE, V42, P1499, DOI 10.1016/j.neuroimage.2008.06.001
   Klimek M, 2004, ANAESTHESIA, V59, P607, DOI 10.1111/j.1365-2044.2004.03675.x
   Kolk AM, 2000, CHILD CARE HLTH DEV, V26, P251, DOI 10.1046/j.1365-2214.2000.00145.x
   Kombos T, 1999, ACTA NEUROCHIR, V141, P1295, DOI 10.1007/s007010050433
   Korvenoja A, 2006, RADIOLOGY, V241, P213, DOI 10.1148/radiol.2411050796
   Kothbauer KF, 2007, NEUROPHYSIOL CLIN, V37, P407, DOI 10.1016/j.neucli.2007.10.003
   LEWINE JD, 1995, FUNCTIONAL BRAIN IMA, P369
   Low D, 2007, ANN ACAD MED SINGAP, V36, P326
   Majos A, 2005, EUR RADIOL, V15, P1148, DOI 10.1007/s00330-004-2565-0
   Meyer RB, 2001, MAYO CLIN PROC, V76, P677, DOI 10.4065/76.7.677
   Miga MI, 1999, STEREOT FUNCT NEUROS, V72, P103, DOI 10.1159/000029707
   Minassian BA, 1999, ANN NEUROL, V46, P627, DOI 10.1002/1531-8249(199910)46:4<627::AID-ANA11>3.0.CO;2-C
   Morota N, 1997, NEUROSURGERY, V41, P1327, DOI 10.1097/00006123-199712000-00017
   MOSELEY ME, 1990, RADIOLOGY, V176, P439, DOI 10.1148/radiology.176.2.2367658
   Mueller WM, 1996, NEUROSURGERY, V39, P515, DOI 10.1097/00006123-199609000-00015
   Muragaki Y, 2006, ACTA NEUROCHIR SUPPL, V98, P67
   Nabavi A, 2001, NEUROSURGERY, V48, P787, DOI 10.1097/00006123-200104000-00019
   Nauta HJW, 1998, P SOC PHOTO-OPT INS, V3262, P229, DOI 10.1117/12.309473
   Ng WH, 2008, NEUROSURGERY, V63, P452, DOI 10.1227/01.NEU.0000325259.95571.20
   Nimsky C, 2004, NEUROSURGERY, V55, P358, DOI 10.1227/01.NEU.0000129694.64671.91
   Nimsky C, 2005, RADIOLOGY, V234, P218, DOI 10.1148/radiol.2341031984
   Nimsky C, 2005, NEUROSURGERY, V56, P130, DOI 10.1227/01.NEU.0000144842.18771.30
   Nimsky C, 2000, NEUROSURGERY, V47, P1070, DOI 10.1097/00006123-200011000-00008
   Nimsky C, 2006, NEUROIMAGE, V30, P1219, DOI 10.1016/j.neuroimage.2005.11.001
   Nimsky C, 2006, NEUROL RES, V28, P482, DOI 10.1179/016164106X115125
   OJEMANN G, 1989, J NEUROSURG, V71, P316, DOI 10.3171/jns.1989.71.3.0316
   Papanicolaou AC, 2004, J NEUROSURG, V100, P867, DOI 10.3171/jns.2004.100.5.0867
   PAPANICOLAOU AC, 1995, BRAIN COGNITION, V27, P331, DOI 10.1006/brcg.1995.1026
   Penfield W, 1937, BRAIN, V60, P389, DOI 10.1093/brain/60.4.389
   Rasmussen IA, 2007, ACTA NEUROCHIR, V149, P365, DOI 10.1007/s00701-006-1110-0
   Roessler K, 2005, J NEUROL NEUROSUR PS, V76, P1152, DOI 10.1136/jnnp.2004.050286
   Roux FE, 2003, NEUROSURGERY, V52, P1335, DOI 10.1227/01.NEU.0000064803.05077.40
   Rowley H A, 1995, Neuroimaging Clin N Am, V5, P695
   Sala F, 2002, CHILD NERV SYST, V18, P264, DOI 10.1007/s00381-002-0582-3
   Sala F, 2006, NEUROSURGERY, V58, P1129, DOI 10.1227/01.NEU.0000215948.97195.58
   Salmelin R, 2007, CLIN NEUROPHYSIOL, V118, P237, DOI 10.1016/j.clinph.2006.07.316
   Schwartz TH, 2002, EPILEPSIA, V43, P430, DOI 10.1046/j.1528-1157.2002.39101.x
   Serletis D, 2007, J NEUROSURG, V107, P1, DOI 10.3171/JNS-07/07/0001
   Shinoura N, 2005, NEUROIMAGE, V25, P936, DOI 10.1016/j.neuroimage.2004.12.060
   Soza Grzegorz, 2003, Comput Aided Surg, V8, P241, DOI 10.3109/10929080309146059
   Steinmeier R, 1998, NEUROSURGERY, V43, P739, DOI 10.1097/00006123-199810000-00005
NR 80
TC 19
Z9 20
U1 0
U2 11
PU SPRINGER
PI NEW YORK
PA 233 SPRING ST, NEW YORK, NY 10013 USA
SN 0256-7040
EI 1433-0350
J9 CHILD NERV SYST
JI Childs Nerv. Syst.
PD APR
PY 2010
VL 26
IS 4
BP 491
EP 502
DI 10.1007/s00381-010-1083-4
PG 12
WC Clinical Neurology; Pediatrics; Surgery
WE Science Citation Index Expanded (SCI-EXPANDED)
SC Neurosciences & Neurology; Pediatrics; Surgery
GA 568QI
UT WOS:000275537100010
PM 20174925
DA 2023-08-27
ER

PT J
AU KYOSHIMA, K
   KOBAYASHI, S
   GIBO, H
   KUROYANAGI, T
AF KYOSHIMA, K
   KOBAYASHI, S
   GIBO, H
   KUROYANAGI, T
TI A STUDY OF SAFE ENTRY ZONES VIA THE FLOOR OF THE 4TH VENTRICLE FOR
   BRAIN-STEM LESIONS - REPORT OF 3 CASES
SO JOURNAL OF NEUROSURGERY
LA English
DT Article
DE BRAIN STEM; SUPRAFACIAL TRIANGLE; INFRAFACIAL TRIANGLE; 4TH VENTRICLE;
   ANATOMICAL STUDY; SURGICAL APPROACH
ID SURGICAL-TREATMENT; TUMORS
AB Direct surgery for intra-axial lesions of the brain stem is considered a hazardous procedure, and morbidity of varying degrees cannot be avoided even with partial removal or biopsy. The main causes of morbidity relate to direct damage during removal of the lesion, selection of an entry route into the brain stem, and the direction of brain stem retraction. The authors examined the possibility of making a medullary incision and retracting the brain stem, taking into account the symptomatology and surgical anatomy, and found two safe entry zones into the brain stem through a suboccipital approach via the floor of the fourth ventricle. These safe entry zones are areas where important neural structures are less prominent. One is the ''suprafacial triangle,'' which is bordered medially by the medial longitudinal fascicle, caudally by the facial nerve (which runs in the brain-stem parenchyma), and laterally by the cerebellar peduncle. The second is the ''infrafacial triangle,'' which is bordered medially by the medial longitudinal fascicle, caudally by the striae medullares, and laterally by the facial nerve. In order to minimize the retraction-related damage to important brain-stem structures, the brain stem should be retracted either laterally or rostrally in the suprafacial triangle approach and only laterally in the infrafacial triangle approach.
   Three localized intra-axial brain-stem lesions were treated surgically via the safe entry zones using the suprafacial approach in two and the infrafacial approach in one. The cases are described and the approaches delineated. Both approaches are indicated for focal intra-axial lesions located unilaterally and dorsal to the medial lemniscus in the lower midbrain to the pons. Magnetic resonance imaging is useful in selecting these approaches, and intraoperative ultrasonography is helpful to confirm the exact location of a lesion before a medullary incision is made. These approaches can also be used as routes for aspiration of brain-stem hemorrhage as well as for tumor biopsy.
RP KYOSHIMA, K (通讯作者)，SHINSHU UNIV,SCH MED,DEPT NEUROSURG,ASAHI 3-1-1,MATSUMOTO,NAGANO 390,JAPAN.
CR BAGHAI P, 1982, NEUROSURGERY, V10, P574, DOI 10.1227/00006123-198205000-00005
   Duvernoy HM, 1978, HUMAN BRAINSTEM VESS, DOI 10.1007/978-3-662-02299-3
   EPSTEIN F, 1988, J NEURO-ONCOL, V6, P309
   EPSTEIN F, 1987, J NEUROSURG, V67, P483, DOI 10.3171/jns.1987.67.4.0483
   HACKER RJ, 1980, NEUROSURGERY, V6, P430, DOI 10.1227/00006123-198004000-00013
   HOFFMAN HJ, 1980, NEUROSURGERY, V7, P243, DOI 10.1227/00006123-198009000-00007
   KASHIWAGI S, 1990, J NEUROSURG, V72, P27, DOI 10.3171/jns.1990.72.1.0027
   KONOVALOV AN, 1990, J NEUROSURG, V73, P181, DOI 10.3171/jns.1990.73.2.0181
   NAKAJIMA K, 1983, STROKE, V14, P485, DOI 10.1161/01.STR.14.4.485
   Pendl G, 1988, Acta Neurochir Suppl (Wien), V42, P130
   SUGITA K, 1982, J NEUROSURG, V57, P747, DOI 10.3171/jns.1982.57.6.0747
   SUGITA K, 1985, MICROSURGICAL ATLAS, P6
   VILLANI R, 1975, CHILD BRAIN, V1, P126
   YASARGIL MG, 1988, MICROSURG, V3, P429
   YOSHIMOTO T, 1986, SURG NEUROL, V26, P72, DOI 10.1016/0090-3019(86)90067-4
NR 15
TC 133
Z9 150
U1 0
U2 2
PU AMER ASSOC NEUROLOGICAL SURGEONS
PI CHARLOTTESVILLE
PA UNIV VIRGINIA, 1224 WEST MAIN ST, STE 450, CHARLOTTESVILLE, VA 22903
SN 0022-3085
J9 J NEUROSURG
JI J. Neurosurg.
PD JUN
PY 1993
VL 78
IS 6
BP 987
EP 993
DI 10.3171/jns.1993.78.6.0987
PG 7
WC Clinical Neurology; Surgery
WE Science Citation Index Expanded (SCI-EXPANDED)
SC Neurosciences & Neurology; Surgery
GA LD500
UT WOS:A1993LD50000021
PM 8487085
DA 2023-08-27
ER

PT J
AU Kawashima, M
   Rhoton, AL
   Tanriover, N
   Ulm, AJ
   Yasuda, A
   Fujii, K
AF Kawashima, M
   Rhoton, AL
   Tanriover, N
   Ulm, AJ
   Yasuda, A
   Fujii, K
TI Microsurgical anatomy of cerebral revascularization. Part II: Posterior
   circulation
SO JOURNAL OF NEUROSURGERY
LA English
DT Article
DE microsurgical anatomy; cerebral revascularization; posterior
   circulation; bypass surgery
ID INFERIOR CEREBELLAR ARTERY; SUPERFICIAL TEMPORAL ARTERY; VEIN BYPASS
   GRAFT; SUBCLAVIAN STEAL SYNDROME; MICRO-SURGICAL TREATMENT; EXTERNAL
   CAROTID-ARTERY; TO-END ANASTOMOSIS; VERTEBRAL ARTERY; VERTEBROBASILAR
   INSUFFICIENCY; OCCIPITAL ARTERY
AB Object. Revasculatization is an important component of treatment for complex aneurysms, skull base tumors, and vertebrobasilar ischemia in the posterior circulation. In this study, the authors examined the microsurgical anatomy related to cerebral revascularization in the posterior circulation and demonstrate various procedures for bypass surgery.
   Methods. The microsurgical anatomy of cerebral and cerebellar vessels as they relate to revascularization procedure and techniques, including extracranial-to-intracranial bypass grafting, arterial interposition grafting, and side-to-side anastomosis, were examined by performing stepwise dissections in 22 adult cadaveric specimens. The arteries and veins in the specimens were perfused with colored silicone.
   Dominant cerebral and cerebellar revascularization procedures in the posterior circulations include superficial temporal artery (STA)-posterior cerebral artery (PCA), STA-superior cerebellar artery (SCA), occipital artery (OA)-anterior inferior cerebellar artery, OA-posterior inferior cerebellar artery (PICA), and PICA-PICA anastomoses. These procedures are effective in relatively small but critical areas including the brainstem and cerebellum. For revascularization of larger areas a saphenous vein graft is used to create a bypass between the PCA and the external carotid artery. Surgical procedures are generally difficult to perform in deep and narrow operative spaces near critical vital structures.
   Conclusions. Although a clear guideline for cerebral revascularization procedures has not yet been established, it is important to understand various microsurgical techniques and their related anatomical structures. This will help surgeons consider surgical indications for treatment of patients with vertebrobasilar ischemia caused by aneurysms, tumors, or atherosclerotic diseases in the posterior circulation.
C1 Kitasato Univ, Sch Med, Dept Neurosurg, Kanagawa, Japan.
   Univ Florida, Dept Neurol Surg, Gainesville, FL USA.
C3 Kitasato University; State University System of Florida; University of
   Florida
RP Kawashima, M (通讯作者)，Kitasato Univ, Sch Med, Dept Neurosurg, Kanagawa, Japan.
EM
RI TANRIOVER, NECMETTIN/HGC-5383-2022; Tanrıöver, Necmettin/AAD-3212-2020
OI Tanrıöver, Necmettin/0000-0001-7628-9443
CR ANSON JA, 1995, NEUROVASCULAR SURG, P383
   AUSMAN JI, 1990, J NEUROSURG, V73, P462, DOI 10.3171/jns.1990.73.3.0462
   AUSMAN JI, 1979, SURG NEUROL, V12, P277
   AUSMAN JI, 1976, MINN MED, V59, P223
   AUSMAN JI, 1981, SURG NEUROL, V16, P99, DOI 10.1016/0090-3019(81)90105-1
   AUSMAN JI, 1981, NEUROSURGERY, V9, P56, DOI 10.1227/00006123-198107000-00009
   AUSMAN JI, 1982, J NEUROSURG, V56, P766, DOI 10.3171/jns.1982.56.6.0766
   AUSMAN JI, 1990, J NEUROSURG, V72, P554, DOI 10.3171/jns.1990.72.4.0554
   BARNETT HJM, 1985, NEW ENGL J MED, V313, P1191, DOI 10.1056/nejm198511073131904
   BOHMFALK GL, 1979, J NEUROSURG, V51, P628, DOI 10.3171/jns.1979.51.5.0628
   BOHMFALK GL, 1979, J NEUROSURG, V51, P641, DOI 10.3171/jns.1979.51.5.0641
   Brennan JW, 1999, J NEUROSURG, V90, P571, DOI 10.3171/jns.1999.90.3.0571
   Cina CS, 2002, J VASC SURG, V35, P422, DOI 10.1067/mva.2002.120035
   Collice M, 1997, ACTA NEUROCHIR, V139, P124, DOI 10.1007/BF02747192
   CORKILL G, 1977, SURG NEUROL, V7, P109
   DIAZ FG, 1985, NEUROSURGERY, V17, P271, DOI 10.1227/00006123-198508000-00004
   DIAZ FG, 1983, NEUROSURGERY, V12, P629, DOI 10.1227/00006123-198306000-00006
   EDWARDS WH, 1994, ANN SURG, V219, P673, DOI 10.1097/00000658-199406000-00010
   Ewald C, 1998, NEUROL MED-CHIR, V38, P83, DOI 10.2176/nmc.38.suppl_83
   Ewald C, 2000, ACTA NEUROCHIR, V142, P731, DOI 10.1007/s007010070087
   FISHER CM, 1958, NEUROLOGY, V8, P311, DOI 10.1212/WNL.8.5.311
   Hamada J, 1996, J NEUROSURG, V85, P496, DOI 10.3171/jns.1996.85.3.0496
   Hamada J, 2002, J NEUROSURG, V96, P867, DOI 10.3171/jns.2002.96.5.0867
   HARDY DG, 1980, NEUROSURGERY, V6, P10, DOI 10.1227/00006123-198001000-00002
   HEROS RC, 1984, J NEUROSURG, V61, P387, DOI 10.3171/jns.1984.61.2.0387
   HOPKINS LN, 1983, NEUROSURGERY, V13, P189, DOI 10.1227/00006123-198308000-00016
   HOPKINS LN, 1989, J NEUROSURG, V70, P207, DOI 10.3171/jns.1989.70.2.0207
   HORVATH M, 1982, ACTA NEUROCHIR, V66, P43, DOI 10.1007/BF01809302
   Iihara K, 2002, J NEUROSURG, V97, P259, DOI 10.3171/jns.2002.97.2.0259
   Khodadad G, 1976, Surg Neurol, V5, P225
   Lemole GM, 2002, J NEUROSURG, V97, P219, DOI 10.3171/jns.2002.97.1.0219
   Lewis SB, 2002, J NEUROSURG, V97, P756, DOI 10.3171/jns.2002.97.4.0756
   LISTER JR, 1982, NEUROSURGERY, V10, P170
   MADSEN JR, 1988, SURG NEUROL, V30, P140, DOI 10.1016/0090-3019(88)90100-0
   Malek AM, 1999, STROKE, V30, P2073, DOI 10.1161/01.STR.30.10.2073
   MIYAMOTO S, 1992, J NEUROSURG, V77, P812, DOI 10.3171/jns.1992.77.5.0812
   Moossy J, 1966, Res Publ Assoc Res Nerv Ment Dis, V41, P1
   NAGAHIRO S, 1993, NEUROSURGERY, V33, P739, DOI 10.1227/00006123-199310000-00027
   NORTH RR, 1962, NEUROLOGY, V12, P810, DOI 10.1212/WNL.12.11.810
   OGAWA A, 1992, J NEUROSURG, V76, P955, DOI 10.3171/jns.1992.76.6.0955
   OKADA Y, 1994, J NEUROSURG, V81, P800, DOI 10.3171/jns.1994.81.5.0800
   PEARCE JE, 1985, NEUROSURGERY, V17, P626, DOI 10.1227/00006123-198510000-00016
   PRITZ MB, 1981, NEUROSURGERY, V9, P524, DOI 10.1227/00006123-198111000-00006
   REUL GJ, 1984, SURGERY, V96, P854
   ROSKI RA, 1982, NEUROSURGERY, V10, P44, DOI 10.1227/00006123-198201000-00008
   SAMSON DS, 1981, ARCH SURG-CHICAGO, V116, P1578
   Sekhar LN, 1998, NEUROSURGERY, V42, P667, DOI 10.1097/00006123-199803000-00045
   SEKHAR LN, 1993, SKULL BASE SURG, V3, P91, DOI 10.1055/s-2008-1060571
   SPETZLER RF, 1987, J NEUROSURG, V66, P648, DOI 10.3171/jns.1987.66.5.0648
   SPETZLER RF, 1980, J NEUROSURG, V53, P465, DOI 10.3171/jns.1980.53.4.0465
   Sultan Sherif, 2002, Vasc Endovascular Surg, V36, P389, DOI 10.1177/153857440203600510
   SUNDT TM, 1987, J NEUROSURG, V66, P172, DOI 10.3171/jns.1987.66.2.0172
   SUNDT TM, 1978, J NEUROSURG, V48, P916, DOI 10.3171/jns.1978.48.6.0916
   SUNDT TM, 1982, J NEUROSURG, V56, P205, DOI 10.3171/jns.1982.56.2.0205
   Takeuchi S, 1998, NEUROSURGERY, V43, P1212, DOI 10.1097/00006123-199811000-00113
   TAKIKAWA S, 1991, Neurological Surgery, V19, P571
   Terasaka S, 2002, NEUROSURGERY, V51, P1083, DOI 10.1097/00006123-200210000-00044
   TOUHO H, 1993, SURG NEUROL, V40, P164, DOI 10.1016/0090-3019(93)90130-S
   Touho H, 1999, SURG NEUROL, V52, P185, DOI 10.1016/S0090-3019(99)00067-1
   Tulleken CAF, 1998, J NEUROSURG, V88, P925, DOI 10.3171/jns.1998.88.5.0925
   WAKUI K, 1992, J NEUROSURG, V77, P624, DOI 10.3171/jns.1992.77.4.0624
   WEKSLER BB, 1983, STROKE, V14, P658, DOI 10.1161/01.STR.14.5.658
   WHISNANT JP, 1978, ANN NEUROL, V3, P107, DOI 10.1002/ana.410030204
   ZEAL AA, 1978, J NEUROSURG, V48, P534, DOI 10.3171/jns.1978.48.4.0534
NR 64
TC 84
Z9 88
U1 0
U2 7
PU AMER ASSOC NEUROLOGICAL SURGEONS
PI ROLLING MEADOWS
PA 5550 MEADOWBROOK DRIVE, ROLLING MEADOWS, IL 60008 USA
SN 0022-3085
EI 1933-0693
J9 J NEUROSURG
JI J. Neurosurg.
PD JAN
PY 2005
VL 102
IS 1
BP 132
EP 147
DI 10.3171/jns.2005.102.1.0132
PG 16
WC Clinical Neurology; Surgery
WE Science Citation Index Expanded (SCI-EXPANDED)
SC Neurosciences & Neurology; Surgery
GA 887FY
UT WOS:000226292300018
PM 15658105
DA 2023-08-27
ER

PT J
AU Stiefel, I
   Schroder, C
   Tanadini-Lang, S
   Pytko, I
   Vu, E
   Klement, RJ
   Guckenberger, M
   Andratschke, N
AF Stiefel, I
   Schroder, C.
   Tanadini-Lang, S.
   Pytko, I
   Vu, E.
   Klement, R. J.
   Guckenberger, M.
   Andratschke, N.
TI High-dose re-irradiation of intracranial lesions - Efficacy and safety
   including dosimetric analysis based on accumulated EQD2Gy dose EQD
   calculation
SO CLINICAL AND TRANSLATIONAL RADIATION ONCOLOGY
LA English
DT Article
DE Re-irradiation; Brain neoplasms; Organs at risk; Glioblastoma; Cranial
   irradiation
AB Introduction: The use of cranial re-irradiation is growing with improving overall survival and the advent of high-precision radiotherapy techniques. Still the value of re-irradiation needs careful evaluation regarding safety and efficacy. We analyzed dosimetric and clinical data of patients receiving cranial reirradiation using EQD2 sum plans.
   Methods and material: We retrospectively analyzed the data of 76 patients who received repeated cranial radiotherapy from 02/2013 to 09/2016. 34 patients suffered from recurrent primary brain tumors, 42 from brain metastases. Dosimetric analysis was performed accumulating EQD2 dose distributions based on rigid image registration. Clinical and radiological data was collected at follow-ups including toxicity, local control and overall survival.
   Results: In total 76 patients had at least 2 courses of intracranial radiotherapy. The median accumulated prescription EQD2 dose was 96.5 Gy(2) for all radiation courses combined. The median D(0.1 cc) of the brain for patients receiving more than 100 Gy(2) was 114 Gy(2) with a highest dose of 161.5 Gy(2). 74% of patients suffered from low grade (G1-G2) acute toxicity, only two high grade (>G3) toxicities were recorded. Median overall survival from the time of first re-irradiation was 57 weeks (range 4-186 weeks). The median time to local failure for patients with a primary brain tumor was not reached and 24 weeks (range 1-77 weeks) for patients with brain metastases.
   Conclusion: Repeated radiotherapy appears both safe and efficient in patients with recurrent primary or secondary brain tumors with doses to the brain up to 120 Gy(2) EQD2, doses below 100 Gy(2) for brainstem and doses below 75 Gy(2) EQD2 to chiasm and optic nerves. (C) 2021 Published by Elsevier B.V. on behalf of European Society for Radiotherapy and Oncology.
C1 [Stiefel, I; Schroder, C.; Tanadini-Lang, S.; Pytko, I; Vu, E.; Guckenberger, M.; Andratschke, N.] Univ Hosp Zurich, Dept Radiat Oncol, Zurich, Switzerland.
   [Schroder, C.] Paul Scherrer Inst, Ctr Proton Therapy, Villigen, Switzerland.
   [Klement, R. J.] Leopoldina Hosp, Dept Radiat Oncol, Schweinfurt, Germany.
   [Klement, R. J.] Univ Zurich, Zurich, Switzerland.
C3 University of Zurich; University Zurich Hospital; Swiss Federal
   Institutes of Technology Domain; Paul Scherrer Institute; University of
   Zurich
RP Andratschke, N (通讯作者)，Univ Zurich, Univ Hosp Zurich, Dept Radiat Oncol, Ramistr 100, CH-100 Zurich, Switzerland.
EM
RI Guckenberger, Matthias/AAX-4994-2020; Klement, Rainer J/H-3484-2019
OI Guckenberger, Matthias/0000-0002-7146-9071; Klement, Rainer
   J/0000-0003-1401-4270; /0000-0002-4168-2795; Stiefel,
   Isabel/0000-0002-8766-9019; Vu, Erwin/0000-0002-3298-517X
CR AbdelWahab MMR, 1997, AM J CLIN ONCOL-CANC, V20, P158, DOI 10.1097/00000421-199704000-00011
   Akiba T, 2012, JPN J CLIN ONCOL, V42, P264, DOI 10.1093/jjco/hys007
   Bernhardt D, 2016, LUNG CANCER, V101, P76, DOI 10.1016/j.lungcan.2016.09.010
   Ciammella P, 2013, RADIAT ONCOL, V8, DOI 10.1186/1748-717X-8-222
   Combs SE, 2005, J CLIN ONCOL, V23, P8863, DOI 10.1200/JCO.2005.03.4157
   Combs SE, 2005, CANCER, V104, P2168, DOI 10.1002/cncr.21429
   Combs SE, 2018, NEUROSURGERY, V83, P1241, DOI 10.1093/neuros/nyx629
   COOPER JS, 1990, RADIOLOGY, V174, P883, DOI 10.1148/radiology.174.3.2305074
   de Robles P, 2015, NEURO-ONCOLOGY, V17, P776, DOI 10.1093/neuonc/nou283
   Dincoglan F, 2015, TUMORI J, V101, P179, DOI 10.5301/tj.5000236
   Dorr W, 2018, CLIN ONCOL-UK, V30, P67, DOI 10.1016/j.clon.2017.11.014
   Greto D, 2014, RADIOL MED, V119, P721, DOI 10.1007/s11547-014-0383-2
   Henke G, 2009, STRAHLENTHER ONKOL, V185, P113, DOI 10.1007/s00066-009-1969-9
   Ho ALK, 2018, CLIN ONCOL-UK, V30, P124, DOI 10.1016/j.clon.2017.11.006
   Howard SP, 2017, J NEURO-ONCOL, V134, P523, DOI 10.1007/s11060-017-2392-1
   Huang Z, 2017, J RADIAT RES, V58, P142, DOI 10.1093/jrr/rrw087
   Hudes RS, 1999, INT J RADIAT ONCOL, V43, P293, DOI 10.1016/S0360-3016(98)00416-7
   LAING RW, 1993, RADIOTHER ONCOL, V27, P22, DOI 10.1016/0167-8140(93)90040-F
   Lee J, 2016, YONSEI MED J, V57, P824, DOI 10.3349/ymj.2016.57.4.824
   Linskey ME, 2010, J NEURO-ONCOL, V96, P45, DOI 10.1007/s11060-009-0073-4
   Mayer R, 2008, INT J RADIAT ONCOL, V70, P1350, DOI 10.1016/j.ijrobp.2007.08.015
   McKenzie JT, 2013, J NEURO-ONCOL, V113, P403, DOI 10.1007/s11060-013-1126-2
   Moller S, 2017, RADIOTHER ONCOL, V125, P223, DOI 10.1016/j.radonc.2017.09.039
   Nieder C, 2016, ANTICANCER RES, V36, P4985, DOI 10.21873/anticanres.11067
   Sadikov E, 2007, CLIN ONCOL-UK, V19, P532, DOI 10.1016/j.clon.2007.06.001
   Scharp M, 2014, RADIAT ONCOL, V9, DOI 10.1186/1748-717X-9-4
   Scoccianti S, 2018, CRIT REV ONCOL HEMAT, V126, P80, DOI 10.1016/j.critrevonc.2018.03.024
   Shaw E, 2000, INT J RADIAT ONCOL, V47, P291, DOI 10.1016/S0360-3016(99)00507-6
   Shepherd SF, 1997, INT J RADIAT ONCOL, V37, P393, DOI 10.1016/S0360-3016(96)00455-5
   Son Christina H, 2012, Int J Radiat Oncol Biol Phys, V82, pe167, DOI 10.1016/j.ijrobp.2011.03.020
   Stelzer Keith J, 2013, Surg Neurol Int, V4, pS192, DOI 10.4103/2152-7806.111296
   Straube C, 2018, BMC CANCER, V18, DOI 10.1186/s12885-017-3928-7
   Veninga T, 2001, RADIOTHER ONCOL, V59, P127, DOI 10.1016/S0167-8140(01)00299-7
   Vordermark D, 2005, BMC CANCER, V5, DOI 10.1186/1471-2407-5-55
   WALLNER KE, 1989, INT J RADIAT ONCOL, V16, P1405, DOI 10.1016/0360-3016(89)90941-3
   Wong WW, 1996, INT J RADIAT ONCOL, V34, P585, DOI 10.1016/0360-3016(95)02156-6
   Yazici G, 2014, J NEURO-ONCOL, V120, P117, DOI 10.1007/s11060-014-1524-0
NR 37
TC 3
Z9 3
U1 0
U2 0
PU ELSEVIER IRELAND LTD
PI CLARE
PA ELSEVIER HOUSE, BROOKVALE PLAZA, EAST PARK SHANNON, CO, CLARE, 00000,
   IRELAND
EI 2405-6308
J9 CLIN TRANSL RAD ONCO
JI Clin. Transl. Radiat. Oncol.
PD MAR
PY 2021
VL 27
BP 132
EP 138
DI 10.1016/j.ctro.2021.01.011
EA FEB 2021
PG 7
WC Oncology; Radiology, Nuclear Medicine & Medical Imaging
WE Science Citation Index Expanded (SCI-EXPANDED)
SC Oncology; Radiology, Nuclear Medicine & Medical Imaging
GA QU1PU
UT WOS:000627056100037
PM 33659717
OA Green Published, gold, Green Accepted
DA 2023-08-27
ER

PT J
AU Yang, WY
   Porras, JL
   Khalafallah, AM
   Sun, Y
   Bettegowda, A
   Mukherjee, D
AF Yang, Wuyang
   Porras, Jose L.
   Khalafallah, Adham M.
   Sun, Yi
   Bettegowda, Anya
   Mukherjee, Debraj
TI Comparison of adult and pediatric pilocytic astrocytomas using competing
   risk analysis: A population-based study
SO CLINICAL NEUROLOGY AND NEUROSURGERY
LA English
DT Article
DE Pilocytic astrocytoma; Adult; Pediatric; SEER; Competing risk analysis
ID MALIGNANT-TRANSFORMATION; DISSEMINATION
AB Background: Pilocytic astrocytoma(PA) is a relatively benign tumor occurring primarily in the pediatric popu-lation. Comparison of characteristics and survival of this tumor between adult and pediatric patients in a single, population-based study is yet to be performed.
   Objective: We aimed to directly compare the characteristics and survival of pilocytic astrocytoma between pe-diatric and adult patients in a single, population-based study.
   Methods: We utilized the SEER database using data from 1983 to 2016. All patients with histologically confirmed, intracranial pilocytic astrocytoma were included and divided into a pediatric(age < 18 years) or adult group. Due to lower risk of tumor-specific-mortality, we utilized a competing risk analysis to account for mortality from other causes. Univariable and multivariable competing risk analysis(CRA) was performed, and sub-distribution hazard ratio(SHR) or adjusted SHR(aSHR) was reported.
   Results: A total of 4357 patients comprised the final cohort, with 3014(69.2%) pediatric patients. As compared to the pediatric group, adult patients were predominantly White(p < 0.01), with PA less likely fully resected(p = 0.01), smaller tumor size(p < 0.01), and were less often located in the cerebellum(p < 0.01). Multivariable CRA revealed a worse prognosis for the adult group(p < 0.01), regional or distal extension(p < 0.01), and non-cerebellar locations including frontal(p < 0.01), parietal(p < 0.01), ventricular(p < 0.01) or brainstem(p < 0.01). Improved prognosis was seen with more recent year-of-diagnosis(2003-2016, p = 0.03), gross-total/total resection(p < 0.01), and biopsy only patients(p = 0.02).
   Conclusions: Pilocytic astrocytomas in adult patients have a worse prognosis than pediatric patients. Cumulative incidence of cancer-specific-mortality is higher in adults when adjusted for other factors. PAs with regional or distal extension and non-cerebellar locations carry worse outcomes. Surgery remains an effective treatment and GTR/TR should be achieved when possible.
C1 [Yang, Wuyang; Porras, Jose L.; Khalafallah, Adham M.; Bettegowda, Anya; Mukherjee, Debraj] Johns Hopkins Sch Med, Dept Neurosurg, Baltimore, MD USA.
   [Sun, Yi] Johns Hopkins Bloomberg Sch Publ Hlth, Dept Mental Hlth, Baltimore, MD USA.
   [Mukherjee, Debraj] Johns Hopkins Univ Sch Med, Dept Neurosurg, 1800 Orleans St Baltimore, Baltimore, MD 21287 USA.
C3 Johns Hopkins University; Johns Hopkins Medicine; Johns Hopkins
   University; Johns Hopkins Bloomberg School of Public Health; Johns
   Hopkins University
RP Mukherjee, D (通讯作者)，Johns Hopkins Univ Sch Med, Dept Neurosurg, 1800 Orleans St Baltimore, Baltimore, MD 21287 USA.
EM
RI Yang, Wuyang/HJP-9492-2023; Khalafallah, Adham/AAH-3136-2019
OI Yang, Wuyang/0000-0001-5934-7795; Khalafallah, Adham/0000-0002-3249-9657
CR Cyrine S, 2013, CLIN NEUROL NEUROSUR, V115, P1220, DOI 10.1016/j.clineuro.2012.11.009
   de Glas NA, 2016, JNCI-J NATL CANCER I, V108, DOI 10.1093/jnci/djv366
   Dutz A, 2019, RADIOTHER ONCOL, V130, P185, DOI 10.1016/j.radonc.2018.09.007
   Feakins Benjamin G, 2018, Diagn Progn Res, V2, P13, DOI 10.1186/s41512-018-0035-4
   Figueiredo EG, 2003, ARQ NEURO-PSIQUIAT, V61, P842, DOI 10.1590/S0004-282X2003000500025
   Georgakis MK, 2017, J NEURO-ONCOL, V131, P163, DOI 10.1007/s11060-016-2284-9
   He VYF, 2017, POPUL HEALTH METR, V15, DOI 10.1186/s12963-016-0118-9
   Johnson DR, 2012, J NEURO-ONCOL, V108, P187, DOI 10.1007/s11060-012-0829-0
   Konar SK, 2022, BRIT J NEUROSURG, V36, P90, DOI 10.1080/02688697.2018.1472214
   Lau B, 2009, AM J EPIDEMIOL, V170, P244, DOI 10.1093/aje/kwp107
   Louis DN, 2016, ACTA NEUROPATHOL, V131, P803, DOI 10.1007/s00401-016-1545-1
   Mair MJ, 2020, J NEURO-ONCOL, V148, P187, DOI 10.1007/s11060-020-03513-9
   Nelson AJ, 2019, BRIT J NEUROSURG, V33, P343, DOI 10.1080/02688697.2018.1549315
   OBANA WG, 1991, J NEUROSURG, V75, P972, DOI 10.3171/jns.1991.75.6.0972
   Park Jae Hui, 2019, Brain Tumor Res Treat, V7, P92, DOI 10.14791/btrt.2019.7.e36
   Parsons MW, 2021, J NEURO-ONCOL, V151, P93, DOI 10.1007/s11060-020-03653-y
   Sattar Sidra, 2017, Surg Neurol Int, V8, P116, DOI 10.4103/sni.sni_468_16
   Tabash MA, 2019, J NEUROL SCI, V400, P148, DOI 10.1016/j.jns.2019.03.028
   van der Wal EPJ, 2003, PEDIATR RADIOL, V33, P207, DOI 10.1007/s00247-002-0828-y
   Ye JM, 2014, J CLIN NEUROSCI, V21, P2160, DOI 10.1016/j.jocn.2014.04.015
NR 20
TC 1
Z9 1
U1 0
U2 0
PU ELSEVIER
PI AMSTERDAM
PA RADARWEG 29, 1043 NX AMSTERDAM, NETHERLANDS
SN 0303-8467
EI 1872-6968
J9 CLIN NEUROL NEUROSUR
JI Clin. Neurol. Neurosurg.
PD JAN
PY 2022
VL 212
AR 107084
DI 10.1016/j.clineuro.2021.107084
PG 5
WC Clinical Neurology; Surgery
WE Science Citation Index Expanded (SCI-EXPANDED)
SC Neurosciences & Neurology; Surgery
GA 0V6CB
UT WOS:000788428500006
PM 34875553
DA 2023-08-27
ER

PT J
AU Gora, J
   Hopfgartner, J
   Kuess, P
   Paskeviciute, B
   Georg, D
AF Gora, Joanna
   Hopfgartner, Johannes
   Kuess, Peter
   Paskeviciute, Brigita
   Georg, Dietmar
TI Is there room for combined modality treatments? Dosimetric comparison of
   boost strategies for advanced head and neck and prostate cancer
SO JOURNAL OF RADIATION RESEARCH
LA English
DT Article
DE VMAT; IMIT; IMPT; combined treatment; boost; H&N; prostate cancer
ID MODULATED RADIATION-THERAPY; ION RADIOTHERAPY; ARC THERAPY; RATIO;
   FRACTIONATION; ALPHA/BETA; MODEL; TRIAL; INDEX; IMRT
AB The purpose of the study was to determine the dosimetric difference between three emerging treatment modalities-volumetric-modulated arc therapy (VMAT), intensity-modulated proton beam therapy (IMPT) and intensity-modulated carbon ion beam therapy (IMIT)-for two tumour sites where selective boosting of the tumour is applied. For 10 patients with locally advanced head and neck (H&N) cancer and 10 with high-risk prostate cancer (PC) a VMAT plan was generated for PTVinitial that included lymph node regions(,) delivering 50 Gy (IsoE) for H&N and 50.4 Gy (IsoE) for PC patients. Furthermore, separate boost plans (VMAT, IMPT and IMIT) were created to boost PTVboost up to 70 Gy (IsoE) and 78 Gy (IsoE) for H&N and PC cases, respectively. Doses to brainstem, myelon, larynx and parotid glands were assessed for H&N cases. Additionally, various OARs (e.g. cochlea, middle ear, masticator space) were evaluated that are currently discussed with respect to quality of life after treatment. For PC cases, bladder, rectum and femoral heads were considered as OARs. For both tumour sites target goals were easily met. Looking at OAR sparing, generally VMAT + VMAT was worst. VMAT + IMIT had the potential to spare some structures in very close target vicinity (such as cochlea, middle ear, masticator space ) significantly better than VMAT + IMPT. Mean doses for rectal and bladder wall were on average 4 Gy (IsoE) and 1.5 Gy (IsoE) higher, respectively, compared to photons plus particles scenarios. Similar results were found for parotid glands and larynx. Concerning target coverage, no significant differences were observed between the three treatment concepts. Clear dosimetric benefits were observed for particle beam therapy as boost modality. However, the clinical benefit of combined modality treatments remains to be demonstrated.
C1 [Gora, Joanna; Hopfgartner, Johannes; Kuess, Peter; Paskeviciute, Brigita; Georg, Dietmar] Med Univ Vienna AKH Wien, Ctr Comprehens Canc, Dept Radiooncol, Div Med Radiat Phys, A-1090 Vienna, Austria.
   [Gora, Joanna] EBG MedAustron GmbH, A-2700 Wiener Neustadt, Austria.
   [Hopfgartner, Johannes; Georg, Dietmar] Med Univ Vienna, Christian Doppler Lab Med Radiat Res Radiat Oncol, Vienna, Austria.
C3 Medical University of Vienna; EBG MedAustron; Medical University of
   Vienna
RP Gora, J (通讯作者)，Med Univ Vienna AKH Wien, Ctr Comprehens Canc, Dept Radiooncol, Div Med Radiat Phys, Waehringer Guertel 18-20, A-1090 Vienna, Austria.
EM
RI Kuess, Peter/AAT-1178-2020; Georg, Dietmar/F-2783-2011
OI Kuess, Peter/0000-0003-2961-1692; Georg, Dietmar/0000-0002-8327-3877
FU European Community [215849-2]; Federal Ministry of Economy, Family and
   Youth; National Foundation for Research, Technology and Development
FX Financial support by The European Community's Seventh Framework
   Programme (FP7 2007-2013) under Grant Agreement No. 215849-2 (Project
   PARTNER), and the Federal Ministry of Economy, Family and Youth, and the
   National Foundation for Research, Technology and Development is
   gratefully acknowledged.
CR [Anonymous], BIOPH GROUP GSI DARM
   [Anonymous], IAEA TECHN REP SER
   [Anonymous], PHAS 3 INT RAD DOS E
   [Anonymous], 2006, KLIN STRAHLENBIOLOGI
   [Anonymous], BRIT J RADIOL
   [Anonymous], INT J RAD ONCOL BIOL
   [Anonymous], 78 ICRU
   [Anonymous], RAD THER TREAT PAT S
   [Anonymous], 2007, ENCY MEASUREMENT STA
   [Anonymous], PTCOG 51 SEOUL
   Arcangeli G, 2010, INT J RADIAT ONCOL, V78, P11, DOI 10.1016/j.ijrobp.2009.07.1691
   Bahadur YA, 2012, SAUDI MED J, V33, P353
   Bassler N, 2010, ACTA ONCOL, V49, P1170, DOI 10.3109/0284186X.2010.510640
   Bentzen SM, 2005, LANCET ONCOL, V6, P112, DOI 10.1016/S1470-2045(05)01737-7
   Bzdusek K, 2009, MED PHYS, V36, P2328, DOI 10.1118/1.3132234
   Chera BS, 2009, INT J RADIAT ONCOL, V75, P994, DOI 10.1016/j.ijrobp.2009.01.044
   Coote JH, 2009, INT J RADIAT ONCOL, V74, P1121, DOI 10.1016/j.ijrobp.2008.09.032
   Fowler J, 2001, INT J RADIAT ONCOL, V50, P1021, DOI 10.1016/S0360-3016(01)01607-8
   Georg D, 2008, INT J RADIAT ONCOL, V71, P1272, DOI 10.1016/j.ijrobp.2008.03.032
   Grun R, 2012, PHYS MED BIOL, V57, P7261, DOI 10.1088/0031-9155/57/22/7261
   Joiner M, 2009, BASIC CLIN RADIOBIOL
   Khor R, 2013, INT J RADIAT ONCOL, V85, P679, DOI 10.1016/j.ijrobp.2012.07.006
   Kramer M, 2000, PHYS MED BIOL, V45, P3319, DOI 10.1088/0031-9155/45/11/314
   Kramer M, 2000, PHYS MED BIOL, V45, P3299, DOI 10.1088/0031-9155/45/11/313
   Luhr A, 2012, RADIOTHER ONCOL, V102, pS156
   Mendenhall WM, 2010, HEAD NECK-J SCI SPEC, V32, P939, DOI 10.1002/hed.21261
   Myers MA, 2012, BRACHYTHERAPY, V11, P292, DOI 10.1016/j.brachy.2011.07.006
   Nyflot MJ, 2012, RADIOTHER ONCOL, V105, P36, DOI 10.1016/j.radonc.2012.09.012
   Paddick I, 2000, J NEUROSURG, V93, P219, DOI 10.3171/jns.2000.93.supplement_3.0219
   Palma D, 2008, INT J RADIAT ONCOL, V72, P996, DOI 10.1016/j.ijrobp.2008.02.047
   Rosenthal DI, 2008, INT J RADIAT ONCOL, V72, P747, DOI 10.1016/j.ijrobp.2008.01.012
   SCHOLZ M, 1996, ADV SPACE RES-SERIES, V18, P5, DOI 10.1016/0273-1177(95)00784-C
   Schwartz DL, 2010, INT J RADIAT ONCOL, V78, P1356, DOI 10.1016/j.ijrobp.2009.10.002
   Thorwarth D, 2010, RADIOTHER ONCOL, V96, P317, DOI 10.1016/j.radonc.2010.07.012
   Tome WA, 2000, INT J RADIAT ONCOL, V48, P593, DOI 10.1016/S0360-3016(00)00666-0
   Vicini FA, 1999, ONCOLOGY-NY, V13, P491
   Vogelius IR, 2013, INT J RADIAT ONCOL, V85, P89, DOI 10.1016/j.ijrobp.2012.03.004
   Wenzl T, 2011, PHYS MED BIOL, V56, P3251, DOI 10.1088/0031-9155/56/11/006
   Wiehle R, 2011, STRAHLENTHER ONKOL, V187, P820, DOI 10.1007/s00066-011-2267-x
   Wiezorek T, 2011, RADIAT ONCOL, V6, DOI 10.1186/1748-717X-6-20
   Yoon MG, 2007, J APPL CLIN MED PHYS, V8, P9, DOI 10.1120/jacmp.v8i2.2390
NR 41
TC 5
Z9 6
U1 0
U2 5
PU OXFORD UNIV PRESS
PI OXFORD
PA GREAT CLARENDON ST, OXFORD OX2 6DP, ENGLAND
SN 0449-3060
EI 1349-9157
J9 J RADIAT RES
JI J. Radiat. Res.
PD JUL
PY 2013
VL 54
SU 1
BP 97
EP 112
DI 10.1093/jrr/rrt067
PG 16
WC Biology; Oncology; Radiology, Nuclear Medicine & Medical Imaging
WE Science Citation Index Expanded (SCI-EXPANDED)
SC Life Sciences & Biomedicine - Other Topics; Oncology; Radiology, Nuclear
   Medicine & Medical Imaging
GA 178RA
UT WOS:000321463900014
PM 23824134
OA Green Published, Green Submitted, gold
DA 2023-08-27
ER

PT J
AU Hwang, JY
   Lee, J
   Oh, CK
   Kang, HW
   Hwang, IY
   Um, JW
   Park, HC
   Kim, S
   Shin, JH
   Park, WY
   Darnell, RB
   Um, HD
   Chung, KC
   Kim, K
   Oh, YJ
AF Hwang, J-Y
   Lee, J.
   Oh, C-K
   Kang, H. W.
   Hwang, I-Y
   Um, J. W.
   Park, H. C.
   Kim, S.
   Shin, J-H
   Park, W-Y
   Darnell, R. B.
   Um, H-D
   Chung, K. C.
   Kim, K.
   Oh, Y. J.
TI Proteolytic degradation and potential role of onconeural protein cdr2 in
   neurodegeneration
SO CELL DEATH & DISEASE
LA English
DT Article
ID PARANEOPLASTIC CEREBELLAR DEGENERATION; MESENCEPHALIC
   DOPAMINERGIC-NEURONS; SPINAL-CORD-INJURY; PARKINSONS-DISEASE; CALPAIN
   ACTIVATION; PRIMARY CULTURES; ALPHA-SYNUCLEIN; ANTIGEN CDR2; CELL-DEATH;
   B-MYB
AB Cerebellar degeneration-related protein 2 (cdr2) is expressed in the central nervous system, and its ectopic expression in tumor cells of patients with gynecological malignancies elicits immune responses by cdr2-specific autoantibodies and T lymphocytes, leading to neurological symptoms. However, little is known about the regulation and function of cdr2 in neurodegenerative diseases. Because we found that cdr2 is highly expressed in the midbrain, we investigated the role of cdr2 in experimental models of Parkinson's disease (PD). We found that cdr2 levels were significantly reduced after stereotaxic injection of 1-methyl-4-phenylpyridinium (MPP+) into the striatum. cdr2 levels were also decreased in the brains of post-mortem PD patients. Using primary cultures of mesencephalic neurons and MN9D cells, we confirmed that MPP+ reduces cdr2 in tyrosine hydroxylase-positive dopaminergic neuronal cells. The MPP+-induced decrease of cdr2 was primarily caused by calpain-and ubiquitin proteasome system-mediated degradation, and cotreatment with pharmacological inhibitors of these enzymes or overexpression of calcium-binding protein rendered cells less vulnerable to MPP+-mediated cytotoxicity. Consequently, overexpression of cdr2 rescued cells from MPP+-induced cytotoxicity, whereas knockdown of cdr2 accelerated toxicity. Collectively, our findings provide insights into the novel regulatory mechanism and potentially protective role of onconeural protein during dopaminergic neurodegeneration.
C1 [Hwang, J-Y; Lee, J.; Oh, C-K; Kang, H. W.; Hwang, I-Y; Um, J. W.; Chung, K. C.; Oh, Y. J.] Yonsei Univ, Coll Life Sci & Biotechnol, Dept Syst Biol, 134 Shinchon Dong, Seoul 120749, South Korea.
   [Hwang, J-Y] Albert Einstein Coll Med, Dominick P Purpura Dept Neurosci, New York, NY 10461 USA.
   [Park, H. C.; Kim, S.] Korea Univ, Grad Sch Med, Ansan 425707, Gyeonggi Do, South Korea.
   [Shin, J-H; Park, W-Y] Sungkyunkwan Univ, Sch Med, Dept Mol Cell Biol, Div Pharmacol, Suwon 440746, Gyeonggi Do, South Korea.
   [Darnell, R. B.] Rockefeller Univ, Howard Hughes Med Inst, Lab Mol Neurooncol, New York, NY 10065 USA.
   [Um, H-D] Korean Inst Radiol & Med Sci, Div Radiat Canc Biol, Seoul 01812, South Korea.
   [Kim, K.] DGIST, Dept Brain & Cognit Sci, Daegu 711873, South Korea.
C3 Yonsei University; Yeshiva University; Korea University; Korea
   University Medicine (KU Medicine); Sungkyunkwan University (SKKU);
   Howard Hughes Medical Institute; Rockefeller University; Daegu Gyeongbuk
   Institute of Science & Technology (DGIST)
RP Oh, YJ (通讯作者)，Yonsei Univ, Coll Life Sci & Biotechnol, Dept Syst Biol, 134 Shinchon Dong, Seoul 120749, South Korea.; Kim, K (通讯作者)，DGIST, Dept Brain & Cognit Sci, 333 Techno Jungang Daero, Daegu 42988, South Korea.
EM
RI Um, Hong-Duk/AAH-3733-2021; Shin, Joo-Ho/P-1417-2018; /ABD-1619-2021;
   Darnell, Robert B/B-9022-2008; Kim, Suhyun/ITT-7958-2023
OI Darnell, Robert B/0000-0002-5134-8088; Park,
   Woong-Yang/0000-0003-4234-0380; Kim, Suhyun/0000-0001-7425-1310; Oh,
   Changki/0000-0003-2489-4072
FU Mid-Career Research Program through the National Research Foundation
   (NRF) - Ministry of Education, Science, and Technology; Brain Research
   Program through NRF - Ministry of Science, ICT & Future Planning
   [2014M3C7A1064545, 2012M2A2A7010422]; Brain Research Program through the
   NRF - Ministry of Science, ICT, and Future Planning [2013M3C7A1056731];
   National Research Foundation of Korea [2013M3C7A1056731] Funding Source:
   Korea Institute of Science & Technology Information (KISTI), National
   Science & Technology Information Service (NTIS)
FX This research was supported by the Mid-Career Research Program through
   the National Research Foundation (NRF) funded by the Ministry of
   Education, Science, and Technology (to YJO), and by the Brain Research
   Program through NRF funded by the Ministry of Science, ICT & Future
   Planning: 2014M3C7A1064545 to KCC and 2012M2A2A7010422 to H-DU. JL is a
   recipient of the Brain Research Program through the NRF funded by the
   Ministry of Science, ICT, and Future Planning (2013M3C7A1056731).
CR Albert ML, 2004, NAT REV CANCER, V4, P36, DOI 10.1038/nrc1255
   BAIMBRIDGE KG, 1992, TRENDS NEUROSCI, V15, P303, DOI 10.1016/0166-2236(92)90081-I
   Banik NL, 1998, ANN NY ACAD SCI, V844, P131, DOI 10.1111/j.1749-6632.1998.tb08228.x
   Bilsland J, 2002, J NEUROSCI, V22, P2637, DOI 10.1523/JNEUROSCI.22-07-02637.2002
   BOTTENSTEIN JE, 1979, P NATL ACAD SCI USA, V76, P514, DOI 10.1073/pnas.76.1.514
   Camins A, 2009, CURR DRUG METAB, V10, P433, DOI 10.2174/138920009788898028
   CHOI HK, 1991, BRAIN RES, V552, P67, DOI 10.1016/0006-8993(91)90661-E
   Choi WS, 2008, BIOCHEM BIOPH RES CO, V371, P127, DOI 10.1016/j.bbrc.2008.04.020
   Choi WS, 2008, P NATL ACAD SCI USA, V105, P15136, DOI 10.1073/pnas.0807581105
   Choi WS, 2004, J BIOL CHEM, V279, P20451, DOI 10.1074/jbc.M311164200
   Choi WS, 2001, J NEUROCHEM, V77, P1531, DOI 10.1046/j.1471-4159.2001.00368.x
   Corradi JP, 1997, J NEUROSCI, V17, P1406
   CUNNINGHAM J, 1986, NEUROLOGY, V36, P1163, DOI 10.1212/WNL.36.9.1163
   Darnell JC, 2000, CANCER RES, V60, P2136
   Darnell RB, 1996, P NATL ACAD SCI USA, V93, P4529, DOI 10.1073/pnas.93.10.4529
   Darnell RB, 2003, NEW ENGL J MED, V349, P1543, DOI 10.1056/NEJMra023009
   Darnell RB, 2006, SEMIN ONCOL, V33, P270, DOI 10.1053/j.seminoncol.2006.03.008
   Dauer W, 2003, NEURON, V39, P889, DOI 10.1016/S0896-6273(03)00568-3
   Dekkers MPJ, 2013, J CELL BIOL, V203, P385, DOI 10.1083/jcb.201306136
   Diepenbroek M, 2014, HUM MOL GENET, V23, P3975, DOI 10.1093/hmg/ddu112
   FATHALLAHSHAYKH H, 1991, P NATL ACAD SCI USA, V88, P3451, DOI 10.1073/pnas.88.8.3451
   GREENLEE JE, 1983, ANN NEUROL, V14, P609, DOI 10.1002/ana.410140603
   Greenlee JE, 2010, J NEUROPATH EXP NEUR, V69, P997, DOI 10.1097/NEN.0b013e3181f0c82b
   Han BS, 2003, J NEUROSCI, V23, P5069
   Harbison RA, 2011, NEUROTOX RES, V19, P374, DOI 10.1007/s12640-010-9172-4
   Hartmann A, 2000, P NATL ACAD SCI USA, V97, P2875, DOI 10.1073/pnas.040556597
   HIDA C, 1994, ARCH NEUROL-CHICAGO, V51, P555, DOI 10.1001/archneur.1994.00540180033010
   Jankovic J, 2008, J NEUROL NEUROSUR PS, V79, P368, DOI 10.1136/jnnp.2007.131045
   Kawamata T, 1998, J NEUROSCI, V18, P7402
   Kim C, 2013, J BIOL CHEM, V288, P36717, DOI 10.1074/jbc.M113.492876
   Kristian T, 1998, STROKE, V29, P705, DOI 10.1161/01.STR.29.3.705
   Kubbutat MHG, 1997, MOL CELL BIOL, V17, P460, DOI 10.1128/MCB.17.1.460
   Lim Junghyun, 2014, Parkinsonism Relat Disord, V20 Suppl 1, pS162, DOI 10.1016/S1353-8020(13)70039-7
   Liu DX, 2004, J NEUROSCI, V24, P8720, DOI 10.1523/JNEUROSCI.1821-04.2004
   Manser C, 2008, FEBS LETT, V582, P2303, DOI 10.1016/j.febslet.2008.05.034
   Mishizen-Eberz AJ, 2003, J NEUROCHEM, V86, P836, DOI 10.1046/j.1471-4159.2003.01878.x
   Moore DJ, 2005, ANNU REV NEUROSCI, V28, P57, DOI 10.1146/annurev.neuro.28.061604.135718
   O'Donovan KJ, 2010, PLOS ONE, V5, DOI 10.1371/journal.pone.0010045
   Okano HJ, 1999, GENE DEV, V13, P2087, DOI 10.1101/gad.13.16.2087
   Park ES, 2012, BRAIN RES BULL, V89, P92, DOI 10.1016/j.brainresbull.2012.07.001
   Paxinos G, 1998, RAT BRAIN IN STEREOTAXIC COORDINATES, FOURTH ED., pix
   Peng S, 2011, MOL BIOL REP, V38, P905, DOI 10.1007/s11033-010-0183-2
   PERSECHINI A, 1989, TRENDS NEUROSCI, V12, P462, DOI 10.1016/0166-2236(89)90097-0
   PETERSON K, 1992, NEUROLOGY, V42, P1931, DOI 10.1212/wnl.42.10.1931
   Ray SK, 2003, BRAIN RES REV, V42, P169, DOI 10.1016/S0165-0173(03)00152-8
   Roberts WK, 2004, CURR OPIN IMMUNOL, V16, P616, DOI 10.1016/j.coi.2004.07.009
   Sakai K, 2004, NEUROBIOL DIS, V15, P529, DOI 10.1016/j.nbd.2003.11.003
   Sakai K, 2002, MOL CELL NEUROSCI, V19, P477, DOI 10.1006/mcne.2001.1059
   Samantaray S, 2015, MOL NEUROBIOL, V52, P1054, DOI 10.1007/s12035-015-9255-6
   Samantaray S, 2008, CNS NEUROL DISORD-DR, V7, P305, DOI 10.2174/187152708784936680
   SHEARMAN MS, 1994, P NATL ACAD SCI USA, V91, P1470, DOI 10.1073/pnas.91.4.1470
   Shin JH, 2011, CELL, V144, P689, DOI 10.1016/j.cell.2011.02.010
   Smith PD, 2006, J NEUROSCI, V26, P440, DOI 10.1523/JNEUROSCI.2875-05.2006
   Storstein A, 2007, ADV CLIN CHEM, V44, P143, DOI 10.1016/S0065-2423(07)44005-7
   Takanaga H, 1998, EXP CELL RES, V241, P363, DOI 10.1006/excr.1998.4060
   TANG L, 1994, J PHARMACOL EXP THER, V268, P495
   Tsubuki S, 1996, J BIOCHEM-TOKYO, V119, P572, DOI 10.1093/oxfordjournals.jbchem.a021280
   Viswanath V, 2001, J NEUROSCI, V21, P9519, DOI 10.1523/JNEUROSCI.21-24-09519.2001
   Yun N, 2014, J BIOL CHEM, V289, P22183, DOI 10.1074/jbc.M114.552679
NR 59
TC 9
Z9 9
U1 0
U2 5
PU NATURE PUBLISHING GROUP
PI LONDON
PA MACMILLAN BUILDING, 4 CRINAN ST, LONDON N1 9XW, ENGLAND
SN 2041-4889
J9 CELL DEATH DIS
JI Cell Death Dis.
PD JUN
PY 2016
VL 7
AR e2240
DI 10.1038/cddis.2016.151
PG 12
WC Cell Biology
WE Science Citation Index Expanded (SCI-EXPANDED)
SC Cell Biology
GA DO0XN
UT WOS:000377502400002
PM 27253404
OA gold, Green Published
DA 2023-08-27
ER

PT J
AU Cao, WH
   Khabazian, A
   Yepes, PP
   Lim, G
   Poenisch, F
   Grosshans, DR
   Mohan, R
AF Cao, Wenhua
   Khabazian, Azin
   Yepes, Pablo P.
   Lim, Gino
   Poenisch, Falk
   Grosshans, David R.
   Mohan, Radhe
TI Linear energy transfer incorporated intensity modulated proton therapy
   optimization
SO PHYSICS IN MEDICINE AND BIOLOGY
LA English
DT Article
DE IMPT; linear energy transfer; optimization
ID RELATIVE BIOLOGICAL EFFECTIVENESS; RADIATION-THERAPY; PARTICLE THERAPY;
   IMAGING CHANGES; EPENDYMOMA; TUMORS; TRACK; BEAMS; MODEL
AB The purpose of this study was to investigate the feasibility of incorporating linear energy transfer (LET) into the optimization of intensity modulated proton therapy (IMPT) plans. Because increased LET correlates with increased biological effectiveness of protons, high LETs in target volumes and low LETs in critical structures and normal tissues are preferred in an IMPT plan. However, if not explicitly incorporated into the optimization criteria, different IMPT plans may yield similar physical dose distributions but greatly different LET, specifically dose-averaged LET, distributions. Conventionally, the IMPT optimization criteria (or cost function) only includes dose-based objectives in which the relative biological effectiveness (RBE) is assumed to have a constant value of 1.1. In this study, we added LET-based objectives for maximizing LET in target volumes and minimizing LET in critical structures and normal tissues. Due to the fractional programming nature of the resulting model, we used a variable reformulation approach so that the optimization process is computationally equivalent to conventional IMPT optimization. In this study, five brain tumor patients who had been treated with proton therapy at our institution were selected. Two plans were created for each patient based on the proposed LET-incorporated optimization (LETOpt) and the conventional dose-based optimization (DoseOpt). The optimized plans were compared in terms of both dose (assuming a constant RBE of 1.1 as adopted in clinical practice) and LET. Both optimization approaches were able to generate comparable dose distributions. The LET-incorporated optimization achieved not only pronounced reduction of LET values in critical organs, such as brainstem and optic chiasm, but also increased LET in target volumes, compared to the conventional dose-based optimization. However, on occasion, there was a need to tradeoff the acceptability of dose and LET distributions. Our conclusion is that the inclusion of LET-dependent criteria in the IMPT optimization could lead to similar dose distributions as the conventional optimization but superior LET distributions in target volumes and normal tissues. This may have substantial advantages in improving tumor control and reducing normal tissue toxicities.
C1 [Cao, Wenhua; Yepes, Pablo P.; Poenisch, Falk; Mohan, Radhe] Univ Texas MD Anderson Canc Ctr, Dept Radiat Phys, Houston, TX 77030 USA.
   [Khabazian, Azin; Lim, Gino] Univ Houston, Dept Ind Engn, Houston, TX 77204 USA.
   [Yepes, Pablo P.] Rice Univ, Dept Phys & Astron, Houston, TX 77005 USA.
   [Grosshans, David R.] Univ Texas MD Anderson Canc Ctr, Dept Radiat Oncol, Houston, TX 77030 USA.
C3 University of Texas System; UTMD Anderson Cancer Center; University of
   Houston System; University of Houston; Rice University; University of
   Texas System; UTMD Anderson Cancer Center
RP Cao, WH (通讯作者)，Univ Texas MD Anderson Canc Ctr, Dept Radiat Phys, Houston, TX 77030 USA.
EM
RI Lim, Gino/J-4607-2014
OI Lim, Gino/0000-0003-2259-2310
FU National Cancer Institute of the National Institutes of Health
   [2U19CA021239-35]; Cancer Prevention and Research Institute of Texas
   [RP160232]
FX Authors would like to thank Amy Ninetto for providing editorial
   assistance to improve this manuscript. This work was supported in part
   by the National Cancer Institute of the National Institutes of Health
   (2U19CA021239-35) and the Cancer Prevention and Research Institute of
   Texas (RP160232).
CR Bassler N, 2014, ACTA ONCOL, V53, P25, DOI 10.3109/0284186X.2013.832835
   Bassler N, 2010, ACTA ONCOL, V49, P1170, DOI 10.3109/0284186X.2010.510640
   Bortfeld T, 1999, SEMIN RADIAT ONCOL, V9, P20, DOI 10.1016/S1053-4296(99)80052-6
   Cao WH, 2013, PHYS MED BIOL, V58, DOI 10.1088/0031-9155/58/15/5113
   Chan TCY, 2006, PHYS MED BIOL, V51, P2567, DOI 10.1088/0031-9155/51/10/014
   Chang AL, 2014, INT J PART THER, V1, P357, DOI DOI 10.14338/IJPT.13.00009.1
   Charnes A., 1962, NAV RES LOGIST Q, V9, P181, DOI DOI 10.1002/NAV.3800090303
   Fager M, 2015, INT J RADIAT ONCOL, V91, P1057, DOI 10.1016/j.ijrobp.2014.12.049
   Frese MC, 2011, INT J RADIAT ONCOL, V79, P80, DOI 10.1016/j.ijrobp.2009.10.022
   Giantsoudi D, 2013, INT J RADIAT ONCOL, V87, P216, DOI 10.1016/j.ijrobp.2013.05.013
   Gillin MT, 2010, MED PHYS, V37, P154, DOI 10.1118/1.3259742
   Grassberger C, 2011, PHYS MED BIOL, V56, P6677, DOI 10.1088/0031-9155/56/20/011
   Grassberger C, 2011, INT J RADIAT ONCOL, V80, P1559, DOI 10.1016/j.ijrobp.2010.10.027
   Guan F, 2015, MED PHYS, V42, P6234, DOI 10.1118/1.4932217
   Gunther JR, 2015, INT J RADIAT ONCOL, V93, P54, DOI 10.1016/j.ijrobp.2015.05.018
   International Commission on Radiation Units and Measurements, 2011, J ICRU, V11, P1, DOI [10.1093/jicru/ndr028, 10.1093/jicru/ndr011]
   Jia X, 2011, PHYS MED BIOL, V56, P6205, DOI 10.1088/0031-9155/56/19/004
   Lomax A, 1999, PHYS MED BIOL, V44, P185, DOI 10.1088/0031-9155/44/1/014
   McNamara AL, 2015, PHYS MED BIOL, V60, P8399, DOI 10.1088/0031-9155/60/21/8399
   Mitin T, 2014, J CLIN ONCOL, V32, P2855, DOI 10.1200/JCO.2014.55.1945
   Paganetti H, 2002, INT J RADIAT ONCOL, V53, P407, DOI 10.1016/S0360-3016(02)02754-2
   Peeler CR, 2016, RADIOTHER ONCOL, V121, P395, DOI 10.1016/j.radonc.2016.11.001
   Sabin ND, 2013, AM J NEURORADIOL, V34, P446, DOI 10.3174/ajnr.A3219
   Schuemann J, 2014, PHYS MED BIOL, V59, P4007, DOI 10.1088/0031-9155/59/15/4007
   Unkelbach J, 2016, INT J RADIAT ONCOL, V96, P1097, DOI 10.1016/j.ijrobp.2016.08.038
   Wilkens JJ, 2004, PHYS MED BIOL, V49, P2811, DOI 10.1088/0031-9155/49/13/004
   Wilkens JJ, 2005, MED PHYS, V32, P455, DOI 10.1118/1.1851925
   Yepes PP, 2016, PHYS MED BIOL, V61, P2633, DOI 10.1088/0031-9155/61/7/2633
NR 28
TC 55
Z9 56
U1 2
U2 12
PU IOP PUBLISHING LTD
PI BRISTOL
PA TEMPLE CIRCUS, TEMPLE WAY, BRISTOL BS1 6BE, ENGLAND
SN 0031-9155
EI 1361-6560
J9 PHYS MED BIOL
JI Phys. Med. Biol.
PD JAN
PY 2018
VL 63
IS 1
AR 015013
DI 10.1088/1361-6560/aa9a2e
PG 10
WC Engineering, Biomedical; Radiology, Nuclear Medicine & Medical Imaging
WE Science Citation Index Expanded (SCI-EXPANDED)
SC Engineering; Radiology, Nuclear Medicine & Medical Imaging
GA FQ5FR
UT WOS:000418384700001
PM 29131808
OA Green Accepted
DA 2023-08-27
ER

PT J
AU Song, B
   Cha, Y
   Ko, S
   Jeon, J
   Lee, N
   Seo, H
   Park, KJ
   Lee, IH
   Lopes, C
   Feitosa, M
   Luna, MJ
   Jung, JH
   Kim, J
   Hwang, D
   Cohen, BM
   Teicher, MH
   Leblanc, P
   Carter, BS
   Kordower, JH
   Bolshakov, VY
   Kong, SW
   Schweitzer, JS
   Kim, KS
AF Song, Bin
   Cha, Young
   Ko, Sanghyeok
   Jeon, Jeha
   Lee, Nayeon
   Seo, Hyemyung
   Park, Kyung-Joon
   Lee, In-Hee
   Lopes, Claudia
   Feitosa, Melissa
   Luna, Maria Jose
   Jung, Jin Hyuk
   Kim, Jisun
   Hwang, Dabin
   Cohen, Bruce M.
   Teicher, Martin H.
   Leblanc, Pierre
   Carter, Bob S.
   Kordower, Jeffrey H.
   Bolshakov, Vadim Y.
   Kong, Sek Won
   Schweitzer, Jeffrey S.
   Kim, Kwang-Soo
TI Human autologous iPSC-derived dopaminergic progenitors restore motor
   function in Parkinson's disease models
SO JOURNAL OF CLINICAL INVESTIGATION
LA English
DT Article
ID PLURIPOTENT STEM-CELLS; CLINICAL TRANSLATION; RAT MODEL; HUMAN ES;
   ANIMAL-MODELS; NEURONS; TRANSPLANTATION; CANCER; DIFFERENTIATION;
   THERAPY
AB Parkinson's disease (PD) is a neurodegenerative disorder associated with loss of striatal dopamine, secondary to degeneration of midbrain dopamine (mDA) neurons in the substantia nigra, rendering cell transplantation a promising therapeutic strategy. To establish human induced pluripotent stem cell-based (hiPSC-based) autologous cell therapy, we report a platform of core techniques for the production of mDA progenitors as a safe and effective therapeutic product. First, by combining metabolism-regulating microRNAs with reprogramming factors, we developed a method to more efficiently generate clinical-grade iPSCs, as evidenced by genomic integrity and unbiased pluripotent potential. Second, we established a "spotting"-based in vitro differentiation methodology to generate functional and healthy mDA cells in a scalable manner. Third, we developed a chemical method that safely eliminates undifferentiated cells from the final product. Dopaminergic cells thus express high levels of characteristic mDA markers, produce and secrete dopamine, and exhibit electrophysiological features typical of mDA cells. Transplantation of these cells into rodent models of PD robustly restores motor function and reinnervates host brain, while showing no evidence of tumor formation or redistribution of the implanted cells. We propose that this platform is suitable for the successful implementation of human personalized autologous cell therapy for PD.
C1 [Song, Bin; Cha, Young; Ko, Sanghyeok; Jeon, Jeha; Lee, Nayeon; Seo, Hyemyung; Park, Kyung-Joon; Lopes, Claudia; Feitosa, Melissa; Luna, Maria Jose; Jung, Jin Hyuk; Kim, Jisun; Hwang, Dabin; Cohen, Bruce M.; Teicher, Martin H.; Leblanc, Pierre; Bolshakov, Vadim Y.; Kim, Kwang-Soo] Harvard Med Sch, McLean Hosp, Dept Psychiat, Belmont, MA USA.
   [Song, Bin; Cha, Young; Ko, Sanghyeok; Jeon, Jeha; Lee, Nayeon; Seo, Hyemyung; Lopes, Claudia; Feitosa, Melissa; Luna, Maria Jose; Jung, Jin Hyuk; Kim, Jisun; Hwang, Dabin; Leblanc, Pierre; Kim, Kwang-Soo] Harvard Med Sch, McLean Hosp, Mol Neurobiol Lab, Belmont, MA USA.
   [Seo, Hyemyung; Kim, Jisun] Hanyang Univ, Dept Mol & Life Sci, Ansan, South Korea.
   [Lee, In-Hee; Kong, Sek Won] Boston Childrens Hosp, Dept Pediat, Boston, MA USA.
   [Lee, In-Hee; Kong, Sek Won] Boston Childrens Hosp, Computat Hlth Informat Program, Boston, MA USA.
   [Carter, Bob S.; Schweitzer, Jeffrey S.] Harvard Med Sch, Massachusetts Gen Hosp, Dept Neurosurg, Boston, MA 02115 USA.
   [Kordower, Jeffrey H.] Rush Univ, Med Ctr, Dept Neurol Sci, Chicago, IL 60612 USA.
C3 Harvard University; McLean Hospital; Harvard University; McLean
   Hospital; Hanyang University; Harvard University; Boston Children's
   Hospital; Harvard University; Boston Children's Hospital; Harvard
   University; Harvard Medical School; Massachusetts General Hospital; Rush
   University
RP Kim, KS (通讯作者)，McLean Hosp, Mol Neurobiol Lab, Room 216,115 Mill St, Belmont, MA 02478 USA.; Schweitzer, JS (通讯作者)，Massachusetts Gen Hosp, Dept Neurosurg, 15 Parkman St Wang 745, Boston, MA 02114 USA.
EM
RI Cohen, Bruce M./AAZ-4986-2020; Carter, Bob/AAB-4868-2021; Lee,
   Eun-ju/JAN-8749-2023; Teicher, Martin H/P-3906-2017; Lee,
   Nayeon/AAK-6395-2020; Jeon, Jeha/J-7648-2017; Cha, Young/AAI-3460-2020;
   lee, jung hee/H-8107-2018; Park, Kyungjoon/AAL-8086-2020
OI Carter, Bob/0000-0002-3586-1324; Teicher, Martin H/0000-0002-6036-0177;
   Jeon, Jeha/0000-0003-3779-2446; Cha, Young/0000-0001-7647-0699; Park,
   Kyungjoon/0000-0002-7518-636X; Song, Bin/0000-0002-3177-1911; Feitosa,
   Melissa/0000-0003-4991-1844; Lee, In-Hee/0000-0002-8857-1355
FU NIH [NS070577, NS084869, OD024622]; National Research Foundation (NRF)
   [2017R1A2B4008456]; Parkinson's Cell Therapy Research Fund at McLean
   Hospital; Massachusetts General Hospital; National Research Foundation
   of Korea [2017R1A2B4008456] Funding Source: Korea Institute of Science &
   Technology Information (KISTI), National Science & Technology
   Information Service (NTIS)
FX We would like to thank all members of the molecular neurobiology
   laboratory past and present who participated in the project. In
   particular, we thank Sangmi Chung and Kai Sonntag for technical input
   and helpful discussions. We also thank the Mostoslavsky's lab for
   gifting us the STEMCCA lentiviral vector. We would like to acknowledge
   the personnel of the Dana Farber Cancer Institute Cell Manipulation Core
   Facility, in particular Jerome Ritz, Sarah Nikiforow, Helene Negre, and
   Laeticia Pinte, for their technical and administrative help in setting
   up the GMP cell production. This work was supported by NIH grants
   (NS070577, NS084869, and OD024622) and a National Research Foundation
   (NRF) grant (2017R1A2B4008456 to HS) as well as the Parkinson's Cell
   Therapy Research Fund at McLean Hospital and Massachusetts General
   Hospital.
CR [Anonymous], NATURE
   [Anonymous], NIH
   [Anonymous], NAT BIOTECHNOL
   [Anonymous], J NEUROSCI
   Arenas E, 2015, DEVELOPMENT, V142, P1918, DOI 10.1242/dev.097394
   Banyard J, 2013, SCI REP-UK, V3, DOI 10.1038/srep03151
   Barker RA, 2017, CELL STEM CELL, V21, P569, DOI 10.1016/j.stem.2017.09.014
   Barker RA, 2015, NAT REV NEUROL, V11, P492, DOI 10.1038/nrneurol.2015.123
   Bjorklund A, 2013, MOVEMENT DISORD, V28, P110, DOI 10.1002/mds.25343
   Blandini F, 2012, FEBS J, V279, P1156, DOI 10.1111/j.1742-4658.2012.08491.x
   Cha Y, 2017, NAT CELL BIOL, V19, P445, DOI 10.1038/ncb3517
   Chambers SM, 2009, NAT BIOTECHNOL, V27, P275, DOI 10.1038/nbt.1529
   Chung S, 2009, CELL STEM CELL, V5, P646, DOI 10.1016/j.stem.2009.09.015
   Cummings BS, 2004, CURR PROTOC PHARM
   Darband SG, 2018, J CELL PHYSIOL, V233, P6544, DOI 10.1002/jcp.26595
   de Lau LML, 2006, LANCET NEUROL, V5, P525, DOI 10.1016/S1474-4422(06)70471-9
   Doi D, 2014, STEM CELL REP, V2, P337, DOI 10.1016/j.stemcr.2014.01.013
   Dowd E, 2005, BRAIN RES BULL, V68, P24, DOI 10.1016/j.brainresbull.2005.08.009
   Effenberg A, 2015, CELL TRANSPLANT, V24, P2099, DOI 10.3727/096368914X685591
   Grealish S, 2014, CELL STEM CELL, V15, P653, DOI 10.1016/j.stem.2014.09.017
   Guzman JN, 2009, J NEUROSCI, V29, P11011, DOI 10.1523/JNEUROSCI.2519-09.2009
   Hargus G, 2010, P NATL ACAD SCI USA, V107, P15921, DOI 10.1073/pnas.1010209107
   Joksimovic M, 2009, P NATL ACAD SCI USA, V106, P19185, DOI 10.1073/pnas.0904285106
   Joksimovic M, 2009, NAT NEUROSCI, V12, P125, DOI 10.1038/nn.2243
   Kandoth C, 2013, NATURE, V502, P333, DOI 10.1038/nature12634
   KANG U J, 1988, Rational Drug Therapy, V22, P1
   Katsukawa M, 2016, STEM CELLS DEV, V25, P815, DOI 10.1089/scd.2015.0394
   Kefalopoulou Z, 2014, JAMA NEUROL, V71, P83, DOI 10.1001/jamaneurol.2013.4749
   Kikuchi T, 2017, J NEUROSCI RES, V95, P1829, DOI 10.1002/jnr.24014
   Kilpinen H, 2017, NATURE, V546, P370, DOI 10.1038/nature22403
   Kirkeby A, 2017, CELL STEM CELL, V20, P135, DOI 10.1016/j.stem.2016.09.004
   Korpal M, 2008, J BIOL CHEM, V283, P14910, DOI 10.1074/jbc.C800074200
   Kriks S, 2011, NATURE, V480, P547, DOI 10.1038/nature10648
   Lee MO, 2013, P NATL ACAD SCI USA, V110, pE3281, DOI 10.1073/pnas.1303669110
   Lehnen D, 2017, STEM CELL REP, V9, P1207, DOI 10.1016/j.stemcr.2017.08.016
   Lindvall O, 2016, J INTERN MED, V279, P30, DOI 10.1111/joim.12415
   Lundblad M, 2002, EUR J NEUROSCI, V15, P120, DOI 10.1046/j.0953-816x.2001.01843.x
   Mandai M, 2017, NEW ENGL J MED, V376, P1038, DOI 10.1056/NEJMoa1608368
   Martincorena I, 2015, SCIENCE, V349, P1483, DOI 10.1126/science.aab4082
   Merkle FT, 2017, NATURE, V545, P229, DOI 10.1038/nature22312
   Miura K, 2009, NAT BIOTECHNOL, V27, P743, DOI 10.1038/nbt.1554
   Nakagawa M, 2010, P NATL ACAD SCI USA, V107, P14152, DOI 10.1073/pnas.1009374107
   Ohnishi K, 2014, CELL, V156, P663, DOI 10.1016/j.cell.2014.01.005
   OLSSON M, 1995, J NEUROSCI, V15, P3863
   Ostrom QT, 2014, NEURO-ONCOLOGY, V16, P896, DOI 10.1093/neuonc/nou087
   Park IH, 2008, NATURE, V451, P141, DOI 10.1038/natureO6534
   Pennartz S, 2004, MOL CELL NEUROSCI, V25, P692, DOI 10.1016/j.mcn.2003.12.011
   Pieters T, 2014, J CELL SCI, V127, P2603, DOI 10.1242/jcs.146720
   Porter AG, 1999, CELL DEATH DIFFER, V6, P99, DOI 10.1038/sj.cdd.4400476
   Rhee YH, 2011, J CLIN INVEST, V121, P2326, DOI 10.1172/JCI45794
   Samata B, 2016, NAT COMMUN, V7, DOI 10.1038/ncomms13097
   Samata B, 2015, J NEUROSCI METH, V243, P68, DOI 10.1016/j.jneumeth.2015.01.027
   Scudellari M, 2016, NATURE, V534, P310, DOI 10.1038/534310a
   Sonntag KC, 2018, PROG NEUROBIOL, V168, P1, DOI 10.1016/j.pneurobio.2018.04.005
   Swistowski A, 2010, STEM CELLS, V28, P1893, DOI 10.1002/stem.499
   Takahashi K, 2007, CELL, V131, P861, DOI 10.1016/j.cell.2007.11.019
   Takahashi K, 2006, CELL, V126, P663, DOI 10.1016/j.cell.2006.07.024
   Takahashi K, 2016, NAT REV MOL CELL BIO, V17, P183, DOI 10.1038/nrm.2016.8
   Tanabe K, 2013, P NATL ACAD SCI USA, V110, P12172, DOI 10.1073/pnas.1310291110
   Tao YL, 2016, CELL STEM CELL, V19, P573, DOI 10.1016/j.stem.2016.10.015
   Tapia N, 2016, CELL STEM CELL, V19, P298, DOI 10.1016/j.stem.2016.06.017
   Tieu K, 2011, CSH PERSPECT MED, V1, DOI 10.1101/cshperspect.a009316
   UNGERSTEDT U, 1970, Brain Research, V24, P485, DOI 10.1016/0006-8993(70)90187-3
   Valastyan S, 2011, J CELL SCI, V124, P999, DOI 10.1242/jcs.081513
   Wakeman DR, 2017, STEM CELL REP, V9, P149, DOI 10.1016/j.stemcr.2017.04.033
   Yu JY, 2007, SCIENCE, V318, P1917, DOI 10.1126/science.1151526
NR 66
TC 67
Z9 68
U1 3
U2 32
PU AMER SOC CLINICAL INVESTIGATION INC
PI ANN ARBOR
PA 2015 MANCHESTER RD, ANN ARBOR, MI 48104 USA
SN 0021-9738
EI 1558-8238
J9 J CLIN INVEST
JI J. Clin. Invest.
PD FEB 3
PY 2020
VL 130
IS 2
BP 904
EP 920
DI 10.1172/JCI130767
PG 17
WC Medicine, Research & Experimental
WE Science Citation Index Expanded (SCI-EXPANDED)
SC Research & Experimental Medicine
GA KK0BS
UT WOS:000512418100038
PM 31714896
OA Bronze, Green Published
DA 2023-08-27
ER

PT J
AU Hopfgartner, J
   Stock, M
   Knausl, B
   Georg, D
AF Hopfgartner, Johannes
   Stock, Markus
   Knaeusl, Barbara
   Georg, Dietmar
TI Robustness of IMPT treatment plans with respect to inter-fractional
   set-up uncertainties: Impact of various beam arrangements for cranial
   targets
SO ACTA ONCOLOGICA
LA English
DT Article
ID MODULATED PROTON THERAPY; RADIATION-THERAPY; NASAL CAVITY; OPTIMIZATION;
   SENSITIVITY; RANGE
AB In the current study IMPT plan robustness was evaluated with respect to inter-fractional patient positioning for various beam arrangements and two tumor indications in the cranial region. Material and methods. For 14 patients suffering from tumors in the cranial region [skull base (SB; n = 7) and paranasal sinus (PS; n = 7)] the CTV and OARs were delineated. A safety margin of 3 mm was applied to the CTV. A prescribed dose of 2 GyE was planned via three beam arrangements (alpha, beta, gamma). Beam arrangement a consisted of lateral opposed fields for both tumor groups while beam arrangement beta was optimized according to respective tumor and OAR locations, using two beams only. Beam arrangement gamma applied four beams in the SB group and three beams in the PS group. Dose distributions were recalculated subjected to virtual patient translations along the major anatomical axes. The following dosimetric indices were evaluated and compared to original plans: target coverage (TC), target dose homogeneity (HI), CTV median and average dose (D-median, D-mean). For OARs near maximum dose and average dose (D-2%, D-mean) were evaluated. Results. Dose distributions were distorted after introducing shifts. In the SB group, TC and HI were significantly different for caudal, cranial and anterior shifts for all beam arrangements. For PS patients, all but right shifts differed significantly from the original plans for all beam arrangements, although clinical relevance was not reached for arrangement gamma (Delta TC < 1.5%). In general, beam arrangement gamma exhibited the least spread of data regarding target indices and was consequently considered the most robust. Dosimetric parameters regarding the brainstem were mostly influenced by shifts along the anterio-posterior axis. Conclusion. For cranial IMPT, set-up uncertainties may lead to pronounced deterioration of dose distributions. According to our investigations, multi-beam arrangements were dosimetrically more robust and hence preferable over two beam arrangements.
C1 [Hopfgartner, Johannes; Stock, Markus; Knaeusl, Barbara; Georg, Dietmar] Med Univ Vienna, Dept Radiooncol, Div Med Radiat Phys, Ctr Comprehens Canc, A-1090 Vienna, Austria.
   [Stock, Markus; Knaeusl, Barbara; Georg, Dietmar] Med Univ Vienna, Christian Doppler Lab Med Radiat Res Radiat Oncol, A-1090 Vienna, Austria.
C3 Medical University of Vienna; Medical University of Vienna
RP Hopfgartner, J (通讯作者)，Med Univ Vienna, Dept Radiooncol, Div Med Radiat Phys, Ctr Comprehens Canc, Wahringer Gurtel 18-20, A-1090 Vienna, Austria.
EM
RI Knäusl, Barbara/AAE-9572-2020; Georg, Dietmar/F-2783-2011; Stock,
   Markus/I-5881-2012
OI Knäusl, Barbara/0000-0003-4031-1711; Georg, Dietmar/0000-0002-8327-3877;
   Stock, Markus/0000-0001-7510-7759
FU EC project ULICE [228436]; European Commission; Austrian Federal
   Ministry of Economy, Family and Youth; National Foundation for Research,
   Technology and Development
FX The current investigation was performed in the context of the EC project
   ULICE (Grant agreement number: 228436). The authors would like to thank
   the European Commission, the Austrian Federal Ministry of Economy,
   Family and Youth and the National Foundation for Research, Technology
   and Development for financial support.
CR Albertini F, 2011, PHYS MED BIOL, V56, P4399, DOI 10.1088/0031-9155/56/14/011
   Blomquist E, 2005, ACTA ONCOL, V44, P862, DOI 10.1080/02841860500355934
   Chen W, 2012, PHYS MED BIOL, V57, P591, DOI 10.1088/0031-9155/57/3/591
   Fredriksson A, 2011, MED PHYS, V38, P1672, DOI 10.1118/1.3556559
   Fukumitsu N, 2012, INT J RADIAT ONCOL, V83, P704, DOI 10.1016/j.ijrobp.2011.07.032
   Georg D, 2006, INT J RADIAT ONCOL, V66, P61
   Gora J, 2012, INT J RAD ONCOL BIOL
   Hillbrand M, 2010, J APPL CLIN MED PHYS, V11, P93, DOI 10.1120/jacmp.v11i4.3219
   Homayuni A, 2011, RADIOTHER ONCOL, V99, pS560, DOI 10.1016/S0167-8140(11)71626-7
   ICRU, 2007, 78 ICRU
   Jakel O, 2007, PHYS MED BIOL, V52, P635, DOI 10.1088/0031-9155/52/3/007
   Lassen-Ramshad Y, 2011, ACTA ONCOL, V50, P791, DOI 10.3109/0284186X.2011.590149
   Lomax AJ, 2008, PHYS MED BIOL, V53, P1043, DOI 10.1088/0031-9155/53/4/015
   Lomax AJ, 2008, PHYS MED BIOL, V53, P1027, DOI 10.1088/0031-9155/53/4/014
   Lomax Antony J, 2004, Z Med Phys, V14, P147
   Lomax NJ, 2003, INT J RADIAT ONCOL, V55, P1409, DOI 10.1016/S0360-3016(02)04599-6
   Meyer J, 2010, INT J RADIAT ONCOL, V78, P428, DOI 10.1016/j.ijrobp.2009.07.1696
   Palm A, 2007, ACTA ONCOL, V46, P462, DOI 10.1080/02841860701218626
   Park PC, 2011, INT J RADIAT ONCOL, V82, P329
   Rutz HP, 2008, INT J RADIAT ONCOL, V71, P220, DOI 10.1016/j.ijrobp.2007.09.014
   Schneider U, 1996, PHYS MED BIOL, V41, P111, DOI 10.1088/0031-9155/41/1/009
   Sejpal SV, 2009, INT J RADIAT ONCOL, V75, P40, DOI 10.1016/j.ijrobp.2008.08.042
   Weber DC, 2012, INT J RADIAT ONCOL, V83, P865, DOI 10.1016/j.ijrobp.2011.08.027
   Yoon MG, 2007, J APPL CLIN MED PHYS, V8, P9, DOI 10.1120/jacmp.v8i2.2390
   Zenda S, 2011, INT J RADIAT ONCOL, V81, P1473, DOI 10.1016/j.ijrobp.2010.08.009
NR 25
TC 14
Z9 14
U1 0
U2 2
PU TAYLOR & FRANCIS LTD
PI ABINGDON
PA 2-4 PARK SQUARE, MILTON PARK, ABINGDON OR14 4RN, OXON, ENGLAND
SN 0284-186X
EI 1651-226X
J9 ACTA ONCOL
JI Acta Oncol.
PD APR
PY 2013
VL 52
IS 3
BP 570
EP 579
DI 10.3109/0284186X.2012.744874
PG 10
WC Oncology
WE Science Citation Index Expanded (SCI-EXPANDED)
SC Oncology
GA 106DT
UT WOS:000316123200016
PM 23244675
DA 2023-08-27
ER

PT J
AU Bataller, L
   Graus, F
   Saiz, A
   Vilchez, JJ
AF Bataller, L
   Graus, F
   Saiz, A
   Vilchez, JJ
CA Spanish Opsoclonus Myoclonus Study
TI Clinical outcome in adult onset idiopathic or paraneoplastic
   opsoclonus-myoclonus
SO BRAIN
LA English
DT Article
DE paraneoplastic; opsoclonus-myoclonus; immunotherapy; autoantibodies
ID EATON-MYASTHENIC-SYNDROME; CELL LUNG-CARCINOMA; ANTI-HU ANTIBODY;
   CEREBELLAR DEGENERATION; HODGKINS-DISEASE; CANCER; ENCEPHALOPATHY;
   DISORDERS; ATAXIA; AUTOANTIBODIES
AB We analysed a series of 24 adult patients with idiopathic (10 cases) and paraneoplastic (14 cases) opsoclonus-myoclonus syndrome (OMS) to ascertain possible differences in clinical course and response to immunotherapies between both groups. Associated tumours were small-cell lung cancer (SCLC) (nine patients), non-SCLC tone patient), breast carcinoma (two patients), gastric adenocarcinoma tone patient) and kidney carcinoma tone patient). Patients with paraneoplastic OMS were older [median age: 66 years versus 40 years (P = 0.006) of those with idiopathic OMS] and had a higher frequency of encephalopathy (64% versus 10%; P = 0,02), Serum from 10/10 idiopathic and 12/14 paraneoplastic OMS patients showed no specific immunoreactivity on rat or human brainstem or cerebellum, lacked specific antineuronal antibodies (Hu, Yo, Ri, Tr, glutamic acid decarboxylase, amphiphysin or CV2) and did not contain antibodies to voltage-gated calcium channels. The two paraneoplastic exceptions were a patient with SCLC, whose serum contained both anti-Hu and anti-amphiphysin antibodies and a patient with breast cancer who had serum anti-Ri antibodies. The clinical course of idiopathic OMS was monophasic except in two elderly women who had relapses of the opsoclonus and mild residual ataxia, Most idiopathic OMS patients made a good recovery, but residual gait ataxia tended to persist in older patients. Immunotherapy mainly intravenous immunoglobulins or corticosteroids) seemed to accelerate recovery. Paraneoplastic OMS had a more severe clinical course, despite treatment with intravenous immunoglobulins or costicosteroids, and was the cause of death in five patients whose tumours were not treated. By contrast the eight patients whose tumours were treated showed a complete or partial neurological recovery. We conclude that idiopathic OMS occurs in younger patients, the clinical evolution is more benign and the effect of immunotherapy appears more effective than in paraneoplastic OMS, In patients aged 50 years and older with OMS who develop encephalopathy, early diagnosis and treatment of a probable underlying tumour, usually SCLC, is indicated to increase the chances of neurological recovery. At present, there are no immunological markers to identify the adult patients with paraneoplastic OMS.
C1 Univ Barcelona, Hosp Clin, Serv Neurol, E-08036 Barcelona, Spain.
   Hosp Univ La Fe, Serv Neurol, Valencia, Spain.
   Univ Barcelona, Hosp Clin, IDIBAPS, E-08036 Barcelona, Spain.
C3 University of Barcelona; Hospital Clinic de Barcelona; Hospital
   Universitari i Politecnic La Fe; University of Barcelona; Hospital
   Clinic de Barcelona; IDIBAPS
RP Graus, F (通讯作者)，Univ Barcelona, Hosp Clin, Serv Neurol, Villarroel 170, E-08036 Barcelona, Spain.
RI Saiz, Albert/K-8236-2014; Graus, Francesc/K-9904-2018; Vilchez,
   Juan/JAX-8314-2023
OI Saiz, Albert/0000-0002-5793-8791; Graus, Francesc/0000-0002-8924-8322;
   Vilchez, Juan J/0000-0002-0532-2872
CR Aggarwal A, 1997, J NEUROL NEUROSUR PS, V63, P687, DOI 10.1136/jnnp.63.5.687
   ANDERSON NE, 1988, MEDICINE, V67, P100, DOI 10.1097/00005792-198803000-00003
   Berger JR, 1999, J NEURO-ONCOL, V41, P43, DOI 10.1023/A:1006189210197
   CAVINESS JN, 1995, MOVEMENT DISORD, V10, P22, DOI 10.1002/mds.870100106
   CHALK CH, 1990, NEUROLOGY, V40, P1552, DOI 10.1212/WNL.40.10.1552
   Connolly AM, 1997, J PEDIATR-US, V130, P878, DOI 10.1016/S0022-3476(97)70272-5
   Corcia P, 1997, J NEUROL NEUROSUR PS, V62, P293, DOI 10.1136/jnnp.62.3.293
   DALMAU J, 1990, ANN NEUROL, V27, P544, DOI 10.1002/ana.410270515
   Dalmau JO, 1999, ARCH NEUROL-CHICAGO, V56, P405, DOI 10.1001/archneur.56.4.405
   Das A, 1999, J NEURO-ONCOL, V41, P181, DOI 10.1023/A:1006133102928
   DIGRE KB, 1986, ARCH NEUROL-CHICAGO, V43, P1165, DOI 10.1001/archneur.1986.00520110055016
   DROPCHO E, 1986, ARCH NEUROL-CHICAGO, V43, P410, DOI 10.1001/archneur.1986.00520040088029
   FURMAN JMR, 1988, NEUROLOGY, V38, P499
   Graus F, 1997, J NEUROIMMUNOL, V74, P55, DOI 10.1016/S0165-5728(96)00205-6
   HERSH B, 1994, NEUROLOGY, V44, P1754, DOI 10.1212/WNL.44.9.1754
   Honnorat J, 1997, J NEUROL, V244, P333, DOI 10.1007/s004150050097
   HORMIGO A, 1994, ANN NEUROL, V36, P896, DOI 10.1002/ana.410360615
   KAY CL, 1993, J NEUROL NEUROSUR PS, V56, P831, DOI 10.1136/jnnp.56.7.831
   KEARSLEY JH, 1985, ARCH NEUROL-CHICAGO, V42, P1208, DOI 10.1001/archneur.1985.04060110090026
   Keime-Guibert F, 1999, NEUROLOGY, V53, P1719, DOI 10.1212/WNL.53.8.1719
   Koukoulis A, 1998, J NEUROL NEUROSUR PS, V64, P137, DOI 10.1136/jnnp.64.1.137
   Lang B, 1996, CURR OPIN IMMUNOL, V8, P865, DOI 10.1016/S0952-7915(96)80017-3
   Leigh RJ, 1999, NEUROLOGY EYE MOVEME
   LUQUE FA, 1991, ANN NEUROL, V29, P241, DOI 10.1002/ana.410290303
   Mason WP, 1997, BRAIN, V120, P1279, DOI 10.1093/brain/120.8.1279
   Mitoma H, 1996, EUR NEUROL, V36, P322
   NEWSOMDAVIS J, 1984, NEUROLOGY, V34, P480, DOI 10.1212/WNL.34.4.480
   NITSCHKE M, 1995, NEW ENGL J MED, V332, P192, DOI 10.1056/NEJM199501193320317
   PETRUZZI MJ, 1995, J PEDIATR-US, V127, P328, DOI 10.1016/S0022-3476(95)70322-5
   Pless M, 1996, NEUROLOGY, V46, P583, DOI 10.1212/WNL.46.2.583-a
   PRANZATELLI MR, 1992, CLIN NEUROPHARMACOL, V15, P186, DOI 10.1097/00002826-199206000-00002
   Saiz A, 1999, J NEUROL NEUROSUR PS, V66, P214, DOI 10.1136/jnnp.66.2.214
   Saiz A, 1997, NEUROLOGY, V49, P1026, DOI 10.1212/WNL.49.4.1026
   SCHOLZ J, 1994, J NEUROL NEUROSUR PS, V57, P763, DOI 10.1136/jnnp.57.6.763-a
   SCHWARTZ M, 1990, J NEUROL NEUROSUR PS, V53, P534, DOI 10.1136/jnnp.53.6.534
   Voltz R, 1999, MUSCLE NERVE, V22, P119, DOI 10.1002/(SICI)1097-4598(199901)22:1<119::AID-MUS19>3.0.CO;2-5
NR 36
TC 245
Z9 254
U1 0
U2 6
PU OXFORD UNIV PRESS
PI OXFORD
PA GREAT CLARENDON ST, OXFORD OX2 6DP, ENGLAND
SN 0006-8950
J9 BRAIN
JI Brain
PD FEB
PY 2001
VL 124
BP 437
EP 443
DI 10.1093/brain/124.2.437
PN 2
PG 7
WC Clinical Neurology; Neurosciences
WE Science Citation Index Expanded (SCI-EXPANDED)
SC Neurosciences & Neurology
GA 403TZ
UT WOS:000167059700018
PM 11157570
OA Bronze
DA 2023-08-27
ER

PT J
AU Kim, H
   Kang, Y
   Li, YJ
   Chen, L
   Lin, L
   Johnson, ND
   Zhu, D
   Robinson, MH
   McSwain, L
   Barwick, BG
   Yuan, XR
   Liao, XB
   Zhao, J
   Zhang, ZP
   Shu, Q
   Chen, JJ
   Allen, EG
   Kenney, AM
   Castellino, RC
   Van Meir, EG
   Conneely, KN
   Vertino, PM
   Jin, P
   Li, J
AF Kim, Hyerim
   Kang, Yunhee
   Li, Yujing
   Chen, Li
   Lin, Li
   Johnson, Nicholas D.
   Zhu, Dan
   Robinson, M. Hope
   McSwain, Leon
   Barwick, Benjamin G.
   Yuan, Xianrui
   Liao, Xinbin
   Zhao, Jie
   Zhang, Zhiping
   Shu, Qiang
   Chen, Jianjun
   Allen, Emily G.
   Kenney, Anna M.
   Castellino, Robert C.
   Van Meir, Erwin G.
   Conneely, Karen N.
   Vertino, Paula M.
   Jin, Peng
   Li, Jian
TI Ten-eleven translocation protein 1 modulates medulloblastoma progression
SO GENOME BIOLOGY
LA English
DT Article
DE Medulloblastoma; 5-hydroxymethylcytosine; TET1; Stem-like property;
   NANOG; PDGF signaling pathway
ID MOLECULAR SUBGROUPS; DNA DEMETHYLATION; READ ALIGNMENT; MOUSE MODEL;
   STEM-CELLS; 5-HYDROXYMETHYLCYTOSINE; TET1; GENES; 5-METHYLCYTOSINE;
   TRANSCRIPTION
AB Background Medulloblastoma (MB) is the most common malignant pediatric brain tumor that originates in the cerebellum and brainstem. Frequent somatic mutations and deregulated expression of epigenetic regulators in MB highlight the substantial role of epigenetic alterations. 5-hydroxymethylcytosine (5hmC) is a highly abundant cytosine modification in the developing cerebellum and is regulated by ten-eleven translocation (TET) enzymes. Results We investigate the alterations of 5hmC and TET enzymes in MB and their significance to cerebellar cancer formation. We show total abundance of 5hmC is reduced in MB, but identify significant enrichment of MB-specific 5hmC marks at regulatory regions of genes implicated in stem-like properties and Nanog-binding motifs. While TET1 and TET2 levels are high in MBs, only knockout of Tet1 in the smoothened (SmoA1) mouse model attenuates uncontrolled proliferation, leading to a favorable prognosis. The pharmacological Tet1 inhibition reduces cell viability and platelet-derived growth factor signaling pathway-associated genes. Conclusions These results together suggest a potential key role of 5hmC and indicate an oncogenic nature for TET1 in MB tumorigenesis, suggesting it as a potential therapeutic target for MBs.
C1 [Kim, Hyerim; Kang, Yunhee; Li, Yujing; Lin, Li; Johnson, Nicholas D.; Allen, Emily G.; Conneely, Karen N.; Jin, Peng; Li, Jian] Emory Univ Sch Med, Dept Human Genet, Atlanta, GA 30322 USA.
   [Chen, Li] Indiana Univ Sch Med, Dept Biostat & Hlth Data Sci, Indianapolis, IN 46202 USA.
   [Zhu, Dan; Van Meir, Erwin G.] Emory Univ Sch Med, Dept Neurosurg, Lab Mol Neuro Oncol, Atlanta, GA 30322 USA.
   [Zhu, Dan; Barwick, Benjamin G.; Van Meir, Erwin G.; Vertino, Paula M.] Emory Univ Sch Med, Dept Hematol & Med Oncol, Atlanta, GA 30322 USA.
   [Robinson, M. Hope; McSwain, Leon] Emory Univ Sch Med, Dept Pediat Oncol, Atlanta, GA 30322 USA.
   [Barwick, Benjamin G.; Kenney, Anna M.; Van Meir, Erwin G.; Vertino, Paula M.] Emory Univ, Winship Canc Inst, Atlanta, GA 30322 USA.
   [Yuan, Xianrui; Liao, Xinbin; Zhao, Jie; Zhang, Zhiping; Li, Jian] Cent South Univ, Xiangya Hosp, Dept Neurosurg, Changsha 410008, Hunan, Peoples R China.
   [Liao, Xinbin; Zhao, Jie; Zhang, Zhiping; Li, Jian] Cent South Univ, Xiangya Hosp, Hydrocephalus Ctr, Changsha 410008, Hunan, Peoples R China.
   [Shu, Qiang] Zhejiang Univ, Sch Med, Childrens Hosp & Inst Translat Med, Hangzhou, Peoples R China.
   [Chen, Jianjun] City Hope Natl Med Ctr, Dept Syst Biol & Gehr Family Ctr Leukemia Res, Duarte, CA 91010 USA.
   [Castellino, Robert C.] Emory Univ Sch Med, Childrens Healthcare Atlanta, Aflac Canc & Blood Disorders Ctr, Atlanta, GA 30322 USA.
C3 Emory University; Indiana University System; Indiana University
   Bloomington; Emory University; Emory University; Emory University; Emory
   University; Central South University; Central South University; Zhejiang
   University; City of Hope; Children's Healthcare of Atlanta (CHOA); Emory
   University
RP Jin, P; Li, J (通讯作者)，Emory Univ Sch Med, Dept Human Genet, Atlanta, GA 30322 USA.
EM
RI Barwick, Benjamin/O-5185-2019; zhu, dd885/GRO-1645-2022; Castellino,
   Robert/GRF-7847-2022; Barwick, Benjamin/GON-5113-2022; Li,
   yujing/GQA-4109-2022; Zhu, Dan/HTQ-7132-2023; li, yu/HGL-9518-2022
OI Barwick, Benjamin/0000-0001-9810-3566; Zhu, Dan/0000-0002-9971-6222;
   Chen, Jianjun/0000-0003-3749-2902
FU National Institute of Health [NS079625, CA235162, NS096236,
   P30CA138292]; National Natural Science Foundation of China [81902551];
   CURE Childhood Cancer Foundation
FX This research was supported in part by National Institute of Health
   (NS079625 to PJ, CA235162 and NS096236 to EGVM), Winship Invest$ pilot
   grant (P30CA138292 to PJ), National Natural Science Foundation of China
   (No. 81902551) and the CURE Childhood Cancer Foundation (to EGVM).
CR Bonvin E, 2019, CANCER RES, V79, P482, DOI 10.1158/0008-5472.CAN-18-1214
   Broad Institute, 2016, PIC SET COMM LIN TOO
   Cavalli FMG, 2017, CANCER CELL, V31, P737, DOI 10.1016/j.ccell.2017.05.005
   Cerami E, 2012, CANCER DISCOV, V2, P401, DOI 10.1158/2159-8290.CD-12-0095
   Chen X, 2008, CELL, V133, P1106, DOI 10.1016/j.cell.2008.04.043
   Cho S, 2011, P NATL ACAD SCI USA, V108, P20778, DOI 10.1073/pnas.1019452108
   Chou WC, 2011, BLOOD, V118, P3803, DOI 10.1182/blood-2011-02-339747
   Costa Y, 2013, NATURE, V495, P370, DOI 10.1038/nature11925
   Cremers C. W. R. J., 2014, CLIN NEUROEMBRYOLOGY, V2nd
   Dawlaty MM, 2011, CELL STEM CELL, V9, P166, DOI 10.1016/j.stem.2011.07.010
   Delhommeau F, 2009, NEW ENGL J MED, V360, P2289, DOI 10.1056/NEJMoa0810069
   Donson AM., 2013, **DATA OBJECT**
   Ecke I, 2009, CANCER RES, V69, P887, DOI 10.1158/0008-5472.CAN-08-0946
   Ellison DW, 2011, ACTA NEUROPATHOL, V121, P381, DOI 10.1007/s00401-011-0800-8
   Fattet S, GSE12992 GENE EXPRES
   Ficz G, 2014, GENOMICS, V104, P352, DOI 10.1016/j.ygeno.2014.08.017
   Fischer Andrew H, 2008, CSH Protoc, V2008, DOI 10.1101/pdb.prot4986
   Gao JJ, 2013, SCI SIGNAL, V6, DOI 10.1126/scisignal.2004088
   Gilbertson RJ, 2003, NAT GENET, V35, P197, DOI 10.1038/ng1103-197
   Good CR, 2017, NUCLEIC ACIDS RES, V45, P8269, DOI 10.1093/nar/gkx435
   Hackett JA, 2013, SCIENCE, V339, P448, DOI 10.1126/science.1229277
   Hahn MA, 2013, CELL REP, V3, P291, DOI 10.1016/j.celrep.2013.01.011
   Hallahan AR, 2004, CANCER RES, V64, P7794, DOI 10.1158/0008-5472.CAN-04-1813
   Hatton BA, 2008, CANCER RES, V68, P1768, DOI 10.1158/0008-5472.CAN-07-5092
   Heinz S, 2010, MOL CELL, V38, P576, DOI 10.1016/j.molcel.2010.05.004
   Hoshino M, 2005, NEURON, V47, P201, DOI 10.1016/j.neuron.2005.06.007
   Hovestadt V, 2014, NATURE, V510, P537, DOI 10.1038/nature13268
   Huang Y, 2014, P NATL ACAD SCI USA, V111, P1361, DOI 10.1073/pnas.1322921111
   Jiang X, 2017, NAT COMMUN, V8, DOI 10.1038/s41467-017-02290-w
   Jin P., 2021, **DATA OBJECT**, DOI [10.6084/m9.figshare.14397986.v1, DOI 10.6084/M9.FIGSHARE.14397986.V1]
   Jin Peng., 2021, **DATA OBJECT**
   Jin SG, 2011, CANCER RES, V71, P7360, DOI 10.1158/0008-5472.CAN-11-2023
   Jin SG, 2011, NUCLEIC ACIDS RES, V39, P5015, DOI 10.1093/nar/gkr120
   Johnson KC, 2016, NAT COMMUN, V7, DOI 10.1038/ncomms13177
   Kim Hyerim, 2019, SRA
   Ko M, 2011, P NATL ACAD SCI USA, V108, P14566, DOI 10.1073/pnas.1112317108
   Ko M, 2010, NATURE, V468, P839, DOI 10.1038/nature09586
   Kool M, 2008, **DATA OBJECT**
   Kraus TFJ, 2012, INT J CANCER, V131, P1577, DOI 10.1002/ijc.27429
   Kriaucionis S, 2009, SCIENCE, V324, P929, DOI 10.1126/science.1169786
   Kudo Y, 2012, CANCER SCI, V103, P670, DOI 10.1111/j.1349-7006.2012.02213.x
   Lambert SR, 2013, **DATA OBJECT**
   Langmead B, 2012, NAT METHODS, V9, P357, DOI [10.1038/NMETH.1923, 10.1038/nmeth.1923]
   Li H, 2009, BIOINFORMATICS, V25, P1754, DOI 10.1093/bioinformatics/btp324
   Li WW, 2011, J NUCLEIC ACIDS, V2011, DOI 10.4061/2011/870726
   Li Y, 2017, STEM CELL REP, V9, P779, DOI 10.1016/j.stemcr.2017.07.011
   Lian CG, 2012, CELL, V150, P1135, DOI 10.1016/j.cell.2012.07.033
   Lin CY, 2016, NATURE, V530, P57, DOI 10.1038/nature16546
   Lopez-Moyado IF, 2019, P NATL ACAD SCI USA, V116, P16933, DOI 10.1073/pnas.1903059116
   Love MI, 2014, GENOME BIOL, V15, DOI 10.1186/s13059-014-0550-8
   MacDonald TJ, 2001, NAT GENET, V29, P143, DOI 10.1038/ng731
   Marson A, 2008, CELL, V134, P521, DOI 10.1016/j.cell.2008.07.020
   McLean CY, 2010, NAT BIOTECHNOL, V28, P495, DOI 10.1038/nbt.1630
   Mi HY, 2019, NUCLEIC ACIDS RES, V47, pD419, DOI 10.1093/nar/gky1038
   Mi HY, 2009, METHODS MOL BIOL, V563, P123, DOI 10.1007/978-1-60761-175-2_7
   Muller T, 2012, AM J PATHOL, V181, P675, DOI 10.1016/j.ajpath.2012.04.017
   Nestor CE, 2012, GENOME RES, V22, P467, DOI 10.1101/gr.126417.111
   Northcott PA, 2017, NATURE, V547, P311, DOI 10.1038/nature22973
   Northcott PA, 2011, J CLIN ONCOL, V29, P1408, DOI 10.1200/JCO.2009.27.4324
   Okashita N, 2014, DEVELOPMENT, V141, P269, DOI 10.1242/dev.099622
   Orr BA, 2012, PLOS ONE, V7, DOI 10.1371/journal.pone.0041036
   Palomero T, 2006, P NATL ACAD SCI USA, V103, P18261, DOI 10.1073/pnas.0606108103
   Pertea M, 2016, NAT PROTOC, V11, P1650, DOI 10.1038/nprot.2016.095
   Pfeifer GP, 2014, CELL TISSUE RES, V356, P631, DOI 10.1007/s00441-014-1896-7
   Pfister S., 2014, **DATA OBJECT**
   Po A, 2010, EMBO J, V29, P2646, DOI 10.1038/emboj.2010.131
   Pugh TJ, 2012, NATURE, V488, P106, DOI 10.1038/nature11329
   Quinlan AR, 2010, BIOINFORMATICS, V26, P841, DOI 10.1093/bioinformatics/btq033
   Ritchie ME, 2015, NUCLEIC ACIDS RES, V43, DOI 10.1093/nar/gkv007
   Robinson G, 2012, NATURE, V488, P43, DOI 10.1038/nature11213
   Roth R., 2007, **DATA OBJECT**
   Roth RB., 2006, **DATA OBJECT**
   Ruzov A, 2011, CELL RES, V21, P1332, DOI 10.1038/cr.2011.113
   Sardina JL, 2018, CELL STEM CELL, V23, P905, DOI 10.1016/j.stem.2018.11.001
   Schuller U, 2008, CANCER CELL, V14, P123, DOI 10.1016/j.ccr.2008.07.005
   Schwalbe EC, 2017, LANCET ONCOL, V18, P958, DOI 10.1016/S1470-2045(17)30243-7
   Shin HJ, 2009, BIOINFORMATICS, V25, P2605, DOI 10.1093/bioinformatics/btp479
   SIDMAN RL, 1973, BRAIN RES, V62, P1, DOI 10.1016/0006-8993(73)90617-3
   Song CX, 2012, NAT METHODS, V9, P75, DOI [10.1038/NMETH.1779, 10.1038/nmeth.1779]
   Song CX, 2011, NAT BIOTECHNOL, V29, P68, DOI 10.1038/nbt.1732
   Stroud H, 2011, GENOME BIOL, V12, DOI 10.1186/gb-2011-12-6-r54
   Supek F, 2014, PLOS GENET, V10, DOI 10.1371/journal.pgen.1004585
   Tahiliani M, 2009, SCIENCE, V324, P930, DOI 10.1126/science.1170116
   Takai H, 2014, CELL REP, V9, P48, DOI 10.1016/j.celrep.2014.08.071
   Tate JG, 2019, NUCLEIC ACIDS RES, V47, pD941, DOI 10.1093/nar/gky1015
   Taylor MD, 2012, ACTA NEUROPATHOL, V123, P465, DOI 10.1007/s00401-011-0922-z
   Uribe-Lewis S, 2015, GENOME BIOL, V16, DOI 10.1186/s13059-015-0605-5
   Vincent JJ, 2013, CELL STEM CELL, V12, P470, DOI 10.1016/j.stem.2013.01.016
   Wang J, 2018, ANNU REV NEUROSCI, V41, P207, DOI 10.1146/annurev-neuro-070815-013838
   Wang T, 2012, HUM MOL GENET, V21, P5500, DOI 10.1093/hmg/dds394
   Wickham H, 2009, USE R, P1, DOI 10.1007/978-0-387-98141-3
   Williams K, 2011, NATURE, V473, P343, DOI 10.1038/nature10066
   Wilson CL, 2005, BIOINFORMATICS, V21, P3683, DOI 10.1093/bioinformatics/bti605
   Wu H, 2011, GENE DEV, V25, P679, DOI 10.1101/gad.2036011
   Xu W, 2011, CANCER CELL, V19, P17, DOI 10.1016/j.ccr.2010.12.014
   Yang H, 2013, ONCOGENE, V32, P663, DOI 10.1038/onc.2012.67
   Yao B, 2014, HUM MOL GENET, V23, P1095, DOI 10.1093/hmg/ddt504
   Zhu D, 2018, CANCER CELL, V33, P1004, DOI 10.1016/j.ccell.2018.05.006
   Zhu GZ, 2014, STEM CELL REP, V2, P253, DOI 10.1016/j.stemcr.2014.01.012
   Zhu XD, 2016, NEURON, V89, P100, DOI 10.1016/j.neuron.2015.11.030
   Zhukova N, 2013, J CLIN ONCOL, V31, P2927, DOI 10.1200/JCO.2012.48.5052
NR 101
TC 2
Z9 3
U1 0
U2 9
PU BMC
PI LONDON
PA CAMPUS, 4 CRINAN ST, LONDON N1 9XW, ENGLAND
SN 1474-760X
J9 GENOME BIOL
JI Genome Biol.
PD APR 29
PY 2021
VL 22
IS 1
AR 125
DI 10.1186/s13059-021-02352-9
PG 25
WC Biotechnology & Applied Microbiology; Genetics & Heredity
WE Science Citation Index Expanded (SCI-EXPANDED)
SC Biotechnology & Applied Microbiology; Genetics & Heredity
GA SI5MC
UT WOS:000654867300006
PM 33926529
OA gold, Green Published
DA 2023-08-27
ER

PT J
AU Gupta, R
   Billis, A
   Shah, RB
   Moch, H
   Osunkoya, AO
   Jochum, W
   Hes, O
   Bacchi, CE
   de Castro, MG
   Hansel, DE
   Zhou, M
   Vankalakunti, M
   Salles, PG
   Cabrera, RA
   Gown, AM
   Amin, MB
AF Gupta, Ruta
   Billis, Athanase
   Shah, Rajal B.
   Moch, Holger
   Osunkoya, Adeboye O.
   Jochum, Wolfram
   Hes, Ondrej
   Bacchi, Carlos E.
   de Castro, Marilia G.
   Hansel, Donna E.
   Zhou, Ming
   Vankalakunti, Mahesha
   Salles, Paulo G.
   Cabrera, Rafael A.
   Gown, Allen M.
   Amin, Mahul B.
TI Carcinoma of the Collecting Ducts of Bellini and Renal Medullary
   Carcinoma Clinicopathologic Analysis of 52 Cases of Rare Aggressive
   Subtypes of Renal Cell Carcinoma With a Focus on Their Interrelationship
SO AMERICAN JOURNAL OF SURGICAL PATHOLOGY
LA English
DT Article
DE collecting duct carcinoma; renal medullary carcinoma; histologic
   subtypes; metastases; immunohistochemistry; prognosis; renal cell
   carcinoma; renal allograft; sickle cell trait; BK polyoma virus
ID PLACENTAL S100 S100P; UROTHELIAL CARCINOMA; CELL CARCINOMA; BK VIRUS;
   KIDNEY; EXPRESSION; ADULT; TRANSCRIPTION; CHEMOTHERAPY; NEPHROPATHY
AB Carcinoma of the collecting ducts of Bellini and renal medullary carcinoma are rare aggressive neoplasms of putative distal nephron origin. First described in 1949, case reports and review articles constitute a major source of information on collecting duct carcinoma, whereas Davis and colleagues and the pediatric tumor registry have contributed the seminal works on renal medullary carcinoma. Here we present a detailed study of collecting duct carcinoma (n = 39) and renal medullary carcinoma (n = 13), characterizing these rare neoplasms and analyzing their interrelationship. Both collecting duct carcinoma and renal medullary carcinoma exhibited significant similarities, such as predilection for the right kidney, tumor mass with an epicenter in the renal medulla, and a mean size of 7 cm. Overall, both tumors exhibited a poorly differentiated adenocarcinoma histology with desmoplastic stromal response (100%), inflammatory infiltrate (100%), frequent perinephric extension (collecting duct carcinoma: 97%; renal medullary carcinoma: 83%), lymphovascular invasion (100%), intraluminal mucin (collecting duct carcinoma: 42%; renal medullary carcinoma: 73%), high nuclear grade (97%), overlapping immunoreactivity for Ulex europaeus agglutinin 1 (collecting duct carcinoma: 75%; renal medullary carcinoma: 55%), CK7 (collecting duct carcinoma: 44%; renal medullary carcinoma: 71%), and high-molecular weight cytokeratin (collecting duct carcinoma: 26%; renal medullary carcinoma: 29%), and nonimmunoreactivity for Ksp-cadherin. Histologically, collecting duct carcinoma frequently had tubular, tubulopapillary, or irregular glandular architecture, whereas renal medullary carcinoma commonly demonstrated islands of anastomosing tubules and cords forming irregular microcystic spaces. Multiple metastases to the lymph nodes, lung, bone, and liver were observed in both categories at presentation (collecting duct carcinoma: 17%; renal medullary carcinoma: 36%). Only patients with organ-confined small tumors were disease free beyond the median survival time. Differential clinical features between collecting duct carcinoma and renal medullary carcinoma included proclivity for younger male individuals of African ancestry with hemoglobin abnormalities and a shorter median survival of 17 weeks (vs. 44 wk for collecting duct carcinoma) for renal medullary carcinoma. The markedly overlapping clinical features, histology, immuno-phenotype, metastasis patterns, and uniformly aggressive outcome in collecting duct and renal medullary carcinomas suggest that renal medullary carcinoma is a distinctive clinicopathologic subtype within the entity of collecting duct carcinoma. The extremely poor prognosis and ongoing clinical trials with specific therapeutic protocols argue for their accurate distinction from other renal cell carcinoma subtypes.
C1 [Gupta, Ruta; Vankalakunti, Mahesha; Amin, Mahul B.] Cedars Sinai Med Ctr, Dept Pathol & Lab Med, Los Angeles, CA 90048 USA.
   [Shah, Rajal B.] Miraca Life Sci, Dept Pathol, Dallas, TX USA.
   [Osunkoya, Adeboye O.] Emory Univ, Sch Med, Dept Pathol, Atlanta, GA 30322 USA.
   [Hansel, Donna E.] Cleveland Clin Fdn, Dept Pathol, Cleveland, OH 44195 USA.
   [Zhou, Ming] NYU, Dept Pathol, New York, NY 10016 USA.
   [Gown, Allen M.] PhenoPath Labs, Dept Pathol, Seattle, WA USA.
   [Billis, Athanase] Univ Estadual Campinas, Fac Ciencias Med, Dept Pathol, Campinas, Brazil.
   [Bacchi, Carlos E.] Dept Pathol, Botucatu, SP, Brazil.
   [de Castro, Marilia G.] Santa Casa Sch Med, Dept Pathol, Sao Paulo, Brazil.
   [Salles, Paulo G.] Hosp Luxemburgo, Assoc Mario Penna, Dept Pathol, Belo Horizonte, MG, Brazil.
   [Moch, Holger] Univ Zurich Hosp, Inst Clin Pathol, Dept Pathol, CH-8091 Zurich, Switzerland.
   [Jochum, Wolfram] Kantonsspital St Gallen, Inst Pathol, Dept Pathol, St Gallen, Switzerland.
   [Hes, Ondrej] Charles Univ Hosp, Dept Pathol, Plzen, Czech Republic.
   [Cabrera, Rafael A.] Portuguese Inst Oncol, Dept Pathol, Lisbon, Portugal.
C3 Cedars Sinai Medical Center; Emory University; Cleveland Clinic
   Foundation; New York University; PhenoPath Laboratories; Universidade
   Estadual de Campinas; University of Zurich; University Zurich Hospital;
   Kantonsspital St. Gallen; Charles University Prague; Universidade de
   Coimbra
RP Amin, MB (通讯作者)，Cedars Sinai Med Ctr, Dept Pathol & Lab Med, 8700 Beverly Blvd,Suite 8728, Los Angeles, CA 90048 USA.
EM
OI Gupta, Ruta/0000-0003-2940-313X; SALLES, PAULO GUILHERME DE
   OLIVERA/0000-0001-8839-3491
CR Adsay NV, 1998, AM J SURG PATHOL, V22, P260, DOI 10.1097/00000478-199802000-00017
   Albadine R, 2010, AM J SURG PATHOL, V34, P965, DOI 10.1097/PAS.0b013e3181dc5e8a
   Albadine R, 2009, J UROLOGY, V182, P735, DOI 10.1016/j.juro.2009.03.078
   Amin MB, 2009, AM J SURG PATHOL, V33, P384, DOI 10.1097/PAS.0b013e3181872d3f
   Amin MB, 1997, AM J SURG PATHOL, V21, P621, DOI 10.1097/00000478-199706000-00001
   Amin MB, 2002, AM J SURG PATHOL, V26, P281, DOI 10.1097/00000478-200203000-00001
   [Anonymous], ADV ANAT PATHOL
   Ataga KI, 2000, AM J HEMATOL, V63, P205, DOI 10.1002/(SICI)1096-8652(200004)63:4<205::AID-AJH8>3.0.CO;2-8
   Avery RA, 1996, CANCER, V78, P128, DOI 10.1002/(SICI)1097-0142(19960701)78:1<128::AID-CNCR18>3.0.CO;2-1
   BAER SC, 1993, HUM PATHOL, V24, P1017, DOI 10.1016/0046-8177(93)90118-Z
   Billis A., 2007, INT BRAZ J UROL, V33, P724
   Chao D, 2002, J UROLOGY, V167, P71, DOI 10.1016/S0022-5347(05)65385-2
   Cheng JX, 2008, MODERN PATHOL, V21, P647, DOI 10.1038/modpathol.2008.44
   DAVIS CJ, 1995, AM J SURG PATHOL, V19, P1, DOI 10.1097/00000478-199501000-00001
   Davis Jr CJ, 2004, WHO CLASSIFICATION T, P35
   DIMOPOULOS MA, 1993, BRIT J UROL, V71, P388, DOI 10.1111/j.1464-410X.1993.tb15978.x
   Downs TM, 2009, CRIT REV ONCOL HEMAT, V70, P59, DOI 10.1016/j.critrevonc.2008.08.006
   Eble JN, 1996, ADV ANAT PATHOL, V3, P233
   Elwood H, 2011, UROLOGY, V78, DOI 10.1016/j.urology.2011.04.043
   Emerson LL, 2008, PEDIATR TRANSPLANT, V12, P600, DOI 10.1111/j.1399-3046.2007.00855.x
   Esheha GE, 2009, AM J SURG PATHOL, V33, P347, DOI 10.1097/PAS.0b013e3181908e24
   FLEMING S, 1986, HISTOPATHOLOGY, V10, P1131, DOI 10.1111/j.1365-2559.1986.tb02553.x
   FOOT NC, 1949, JAMA-J AM MED ASSOC, V139, P356, DOI 10.1001/jama.1949.02900230010003
   FUZESI L, 1992, HISTOPATHOLOGY, V21, P155, DOI 10.1111/j.1365-2559.1992.tb00364.x
   Galfano A, 2008, UROL INT, V80, P113, DOI 10.1159/000112599
   Gupta R, 2008, ADV ANAT PATHOL, V15, P127, DOI 10.1097/PAP.0b013e31817145a9
   Gupta R, 2009, AM J SURG PATHOL, V33, P241, DOI 10.1097/PAS.0b013e318181b828
   Hakimi AA, 2007, UROLOGY, V70, P878, DOI 10.1016/j.urology.2007.06.1124
   Higgins JPT, 2007, AM J SURG PATHOL, V31, P673, DOI 10.1097/01.pas.0000213438.01278.5f
   Jiang BH, 1997, CANCER RES, V57, P5328
   Kalyanpur A, 1997, AM J ROENTGENOL, V169, P1037, DOI 10.2214/ajr.169.4.9308460
   Kamranvar SA, 2007, ONCOGENE, V26, P5115, DOI 10.1038/sj.onc.1210324
   Kausman JY, 2004, PEDIATR NEPHROL, V19, P459, DOI 10.1007/s00467-003-1407-7
   KENNEDY SM, 1990, HUM PATHOL, V21, P449, DOI 10.1016/0046-8177(90)90209-N
   Kobayashi N, 2008, HUM PATHOL, V39, P1350, DOI 10.1016/j.humpath.2007.11.020
   Mejean A, 2003, J UROLOGY, V169, P1287, DOI 10.1097/01.ju.0000050221.51509.f5
   Milowsky MI, 2002, CANCER, V94, P111, DOI 10.1002/cncr.10204
   MIYAMOTO H, 1992, UROL INT, V48, P460, DOI 10.1159/000282379
   Moscicki AB, 2008, J ADOLESCENT HEALTH, V43, pS26, DOI 10.1016/j.jadohealth.2008.07.010
   O'Donnell PH, 2010, NAT REV UROL, V7, P110, DOI 10.1038/nrurol.2009.255
   O'Neil JD, 2008, J GEN VIROL, V89, P2833, DOI 10.1099/vir.0.2008/003392-0
   Orsola A, 2005, UROLOGY, V65, P49, DOI 10.1016/j.urology.2004.08.012
   Oudard S, 2007, J UROLOGY, V177, P1698, DOI 10.1016/j.juro.2007.01.063
   Peyromaure M, 2003, J UROLOGY, V170, P1138, DOI 10.1097/01.ju.0000086616.40603.ad
   Polascik TJ, 1996, CANCER RES, V56, P1892
   Rathmell WK, 2008, UROLOGY, V72, P659, DOI 10.1016/j.urology.2008.05.009
   Sathyamoorthy K, 2006, NAT CLIN PRACT UROL, V3, P279, DOI 10.1038/ncpuro0483
   Schaeffer EM, 2010, BJU INT, V106, P62, DOI 10.1111/j.1464-410X.2009.09139.x
   SCHOENBERG M, 1995, GENE CHROMOSOME CANC, V12, P76, DOI 10.1002/gcc.2870120115
   Selby DM, 2000, J UROLOGY, V163, P1238, DOI 10.1016/S0022-5347(05)67733-6
   Selli C, 1997, J UROLOGY, V158, P245, DOI 10.1097/00005392-199707000-00079
   Sibony M, 2008, ANN PATHOL, V28, P381, DOI 10.1016/j.annpat.2008.07.009
   Simpson L, 2005, J UROLOGY, V173, P1883, DOI 10.1097/01.ju.0000158448.56888.09
   Srigley J., 2004, WHO CLASSIFICATION T, P33
   Srigley JR, 2009, MODERN PATHOL, V22, pS2, DOI 10.1038/modpathol.2009.70
   Srigley JR, 1998, SEMIN DIAGN PATHOL, V15, P54
   Staehler M, 2009, CLIN GENITOURIN CANC, V7, P58, DOI 10.3816/CGC.2009.n.010
   Stahlschmidt J, 1999, MED PEDIATR ONCOL, V33, P551, DOI 10.1002/(SICI)1096-911X(199912)33:6<551::AID-MPO5>3.0.CO;2-5
   Swartz MA, 2002, UROLOGY, V60, P1083, DOI 10.1016/S0090-4295(02)02154-4
   Vats A, 2008, PEDIATR TRANSPLANT, V12, P499, DOI 10.1111/j.1399-3046.2008.00960.x
   Watanabe IC, 2007, MODERN PATHOL, V20, P914, DOI 10.1038/modpathol.3800934
   WEISS LM, 1995, AM J CLIN PATHOL, V103, P624
   Wesche WA, 1998, PEDIATR PATHOL LAB M, V18, P97, DOI 10.1080/107710498174263
   Wright JL, 2009, J UROLOGY, V182, P2595, DOI 10.1016/j.juro.2009.08.049
   Wu ZS, 2009, BJU INT, V103, P1721, DOI 10.1111/j.1464-410X.2008.08290.x
   Yang XMJ, 2004, CANCER-AM CANCER SOC, V100, P976, DOI 10.1002/cncr.20049
NR 66
TC 98
Z9 102
U1 0
U2 7
PU LIPPINCOTT WILLIAMS & WILKINS
PI PHILADELPHIA
PA TWO COMMERCE SQ, 2001 MARKET ST, PHILADELPHIA, PA 19103 USA
SN 0147-5185
EI 1532-0979
J9 AM J SURG PATHOL
JI Am. J. Surg. Pathol.
PD SEP
PY 2012
VL 36
IS 9
BP 1265
EP 1278
DI 10.1097/PAS.0b013e3182635954
PG 14
WC Pathology; Surgery
WE Science Citation Index Expanded (SCI-EXPANDED)
SC Pathology; Surgery
GA 998CF
UT WOS:000308214100001
PM 22895263
DA 2023-08-27
ER

PT J
AU Shaida, AM
   McFerran, DJ
   da Cruz, M
   Hardy, DG
   Moffat, DA
AF Shaida, AM
   McFerran, DJ
   da Cruz, M
   Hardy, DG
   Moffat, DA
TI Cavernous haemangioma of the internal auditory canal
SO JOURNAL OF LARYNGOLOGY AND OTOLOGY
LA English
DT Article
DE haemangioma; facial nerve; hearing loss, sensorineural
ID TEMPORAL BONE; FACIAL-NERVE; VASCULAR MALFORMATION; HEMANGIOMA; TUMORS;
   MANAGEMENT; MRI; CT
AB Cavernous haemangiomas are rare lesions of the cerebello-pontine angle that can mimic the more commonly occurring vestibular schwannoma. A case report involving a patient with a cavernous haemangioma of the internal auditory canal (IAC) highlights this as a diagnostic possibility for lesions of the IAC by comparing and contrasting the clinical and radiological findings with the more commonly occurring vestibular nerve and facial schwannomas.
   Symptoms such as hearing loss and facial paralysis that are disproportionate to the size of the lesion or fluctuate with hormonal changes such as those seen in pregnancy are suggestive of haemangioma. Radiological imaging demonstrating a lesion enchancing with gadolinium and containing areas of calcification is also suggestive of haemangioma. It is important to consider the possible diagnosis of haemangioma as early recognition of this entity may improve the chances of preserving the functional integrity of the facial nerve.
C1 Addenbrookes Hosp, Dept Otoneurosurg & Skull Base Surg, Cambridge, England.
C3 Cambridge University Hospitals NHS Foundation Trust; Addenbrooke's
   Hospital; University of Cambridge
RP Shaida, AM (通讯作者)，Addenbrookes Hosp NHS Trust, Dept Otolaryngol, Clin 10, Hills Rd,Box 48, Cambridge CB2 2QQ, England.
OI McFerran, Don/0000-0002-4395-3405
CR ATLAS MD, 1992, ANN OTO RHINOL LARYN, V101, P620, DOI 10.1177/000348949210100715
   BATSAKIS JG, 1979, TUMOURS HEAD NECK CL
   BOTTRILL I, 1995, ARCH OTOLARYNGOL, V121, P348
   BUCHANAN DS, 1992, J LARYNGOL OTOL, V106, P1086, DOI 10.1017/S0022215100121838
   CURTIN HD, 1987, RADIOLOGY, V164, P831, DOI 10.1148/radiology.164.3.3112865
   EBY TL, 1992, AM J OTOL, V13, P223
   Escada P, 1997, J LARYNGOL OTOL, V111, P858, DOI 10.1017/S0022215100138812
   Fisch U., 1977, P 3 S FAC NERV SURG, P448
   GLASSCOCK ME, 1984, LARYNGOSCOPE, V94, P869
   HOUSE JW, 1985, OTOLARYNG HEAD NECK, V93, P146, DOI 10.1177/019459988509300202
   LINSKEY ME, 1991, J NEUROSURG, V74, P516, DOI 10.3171/jns.1991.74.3.0516
   LIPKIN AF, 1987, OTOLARYNG HEAD NECK, V96, P71, DOI 10.1177/019459988709600113
   LO WWM, 1989, RADIOLOGY, V17, P443
   MANGHAM CA, 1981, LARYNGOSCOPE, V91, P867
   MARTIN N, 1992, NEURORADIOLOGY, V34, P420, DOI 10.1007/BF00596506
   Morrison GAJ, 1996, CLIN OTOLARYNGOL, V21, P80, DOI 10.1111/j.1365-2273.1996.tb01030.x
   MUGLISTON TAH, 1985, J LARYNGOL OTOL, V99, P1309, DOI 10.1017/S0022215100098601
   Omojola MF, 1997, BRIT J RADIOL, V70, P1184, DOI 10.1259/bjr.70.839.9536913
   SALEH E, 1993, J LARYNGOL OTOL, V107, P1039, DOI 10.1017/S0022215100125216
   WAGNER BJ, 1995, SEMIN ULTRASOUND CT, V16, P253, DOI 10.1016/0887-2171(95)90021-7
NR 20
TC 17
Z9 18
U1 0
U2 1
PU HEADLEY BROTHERS LTD
PI ASHFORD
PA INVICTA PRESS, ASHFORD, KENT, ENGLAND TN24 8HH
SN 0022-2151
J9 J LARYNGOL OTOL
JI J. Laryngol. Otol.
PD JUN
PY 2000
VL 114
IS 6
BP 453
EP 455
PG 3
WC Otorhinolaryngology
WE Science Citation Index Expanded (SCI-EXPANDED)
SC Otorhinolaryngology
GA 336JF
UT WOS:000088297600011
PM 10962680
DA 2023-08-27
ER

PT J
AU Fukumitsu, N
   Okumura, T
   Mizumoto, M
   Oshiro, Y
   Hashimoto, T
   Kanemoto, A
   Hashii, H
   Ohkawa, A
   Moritake, T
   Tsuboi, K
   Tabuchi, K
   Wada, T
   Hara, A
   Sakurai, H
AF Fukumitsu, Nobuyoshi
   Okumura, Toshiyuki
   Mizumoto, Masashi
   Oshiro, Yoshiko
   Hashimoto, Takayuki
   Kanemoto, Ayae
   Hashii, Haruko
   Ohkawa, Ayako
   Moritake, Takashi
   Tsuboi, Koji
   Tabuchi, Keiji
   Wada, Tetsuro
   Hara, Akira
   Sakurai, Hideyuki
TI Outcome of T4 (International Union Against Cancer Staging System, 7th
   edition) or Recurrent Nasal Cavity and Paranasal Sinus Carcinoma Treated
   With Proton Beam
SO INTERNATIONAL JOURNAL OF RADIATION ONCOLOGY BIOLOGY PHYSICS
LA English
DT Article
DE Carcinoma; Nasal cavity; Paranasal sinus; Proton beam therapy
ID INTENSITY-MODULATED RADIOTHERAPY; RADIATION-THERAPY; MALIGNANCIES; HEAD;
   NECK; RADIOCHEMOTHERAPY; ONCOLOGY
AB Purpose: To investigate the clinical features, prognostic factors, and toxicity of treatment for unresectable carcinomas of the nasal cavity and paranasal sinus (NCPS) treated with proton beam therapy (PBT).
   Methods and Materials: Seventeen patients (13 men, 4 women) with unresectable carcinomas of the NCPS who underwent PBT at the University of Tsukuba between 2001 and 2007 were analyzed. The patients' median age was 62 years (range, 30-83 years). The tumors were located in the nasal cavity in 3 patients, the frontal sinus in 1, the ethmoid sinus in 9, and the maxillary sinus in 4. The clinical stage was Stage IVA in 5 cases, IVB in 10, and recurrent in 2. The tumors were deemed unresectable for medical reasons in 16 patients and because of refusal at a previous hospital 4 months earlier in 1 patient. All the patients received PBT irradiation dose of 22-82.5 GyE and a total of 72.4-89.6 GyE over 30-64 fractions (median 78 GyE over 36 fractions) with X-ray, with attention not exceeding the delivery of 50 GyE to the optic chiasm and brain-stem.
   Results: The overall survival rate was 47.1% at 2 years and 15.7% at 5 years, and the local control rate was 35.0% at 2 years and 17.5% at 5 years. Invasion of the frontal or sphenoid sinus was a prognostic factor for overall survival or local control. Late toxicity of more than Grade 3 was found in 2 patients (brain necrosis in 1 and ipsilateral blindness in 1); however, no mortal adverse effects were observed.
   Conclusion: Proton beam therapy enabled a reduced irradiation dose to the optic chiasm and brainstem, enabling the safe treatment of unresectable carcinomas in the NCPS. Superior or posterior extension of the tumor influenced patient outcome. (C) 2012 Elsevier Inc.
C1 [Fukumitsu, Nobuyoshi; Okumura, Toshiyuki; Mizumoto, Masashi; Oshiro, Yoshiko; Hashimoto, Takayuki; Kanemoto, Ayae; Hashii, Haruko; Ohkawa, Ayako; Moritake, Takashi; Tsuboi, Koji; Sakurai, Hideyuki] Univ Tsukuba, Proton Med Res Ctr, Grad Sch Comprehens Human Sci, Tsukuba, Ibaraki 3058575, Japan.
   [Tabuchi, Keiji; Wada, Tetsuro; Hara, Akira] Univ Tsukuba, Dept Otolaryngol, Grad Sch Comprehens Human Sci, Tsukuba, Ibaraki 3058575, Japan.
C3 University of Tsukuba; University of Tsukuba
RP Fukumitsu, N (通讯作者)，Univ Tsukuba, Proton Med Res Ctr, Grad Sch Comprehens Human Sci, 1-1-1 Tennoudai, Tsukuba, Ibaraki 3058575, Japan.
EM
RI Wada, Tetsuro/T-7917-2019; Mizumoto, Masashi/HJP-4287-2023
OI Mizumoto, Masashi/0000-0001-7837-1301
FU Grants-in-Aid for Scientific Research [24659556, 21390338, 23791390]
   Funding Source: KAKEN
CR Blanco AI, 2004, INT J RADIAT ONCOL, V59, P51, DOI 10.1016/j.ijrobp.2003.09.101
   Chen AM, 2007, INT J RADIAT ONCOL, V69, P141, DOI 10.1016/j.ijrobp.2007.02.031
   COX JD, 1995, INT J RADIAT ONCOL, V31, P1341, DOI 10.1016/0360-3016(95)00060-C
   Daly ME, 2007, INT J RADIAT ONCOL, V67, P151, DOI 10.1016/j.ijrobp.2006.07.1389
   Denis F, 2004, J CLIN ONCOL, V22, P69, DOI 10.1200/JCO.2004.08.021
   Dirix P, 2007, RADIOTHER ONCOL, V85, P385, DOI 10.1016/j.radonc.2007.10.001
   Dirix P, 2010, INT J RADIAT ONCOL, V78, P998, DOI 10.1016/j.ijrobp.2009.09.067
   Dulguerov P, 2001, CANCER-AM CANCER SOC, V92, P3012, DOI 10.1002/1097-0142(20011215)92:12<3012::AID-CNCR10131>3.0.CO;2-E
   Fortin A, 2008, AM J CLIN ONCOL-CANC, V31, P379, DOI 10.1097/COC.0b013e318165c036
   Hoppe BS, 2008, INT J RADIAT ONCOL, V72, P763, DOI 10.1016/j.ijrobp.2008.01.038
   Hoppe BS, 2007, INT J RADIAT ONCOL, V67, P691, DOI 10.1016/j.ijrobp.2006.09.023
   Madani I, 2009, INT J RADIAT ONCOL, V73, P424, DOI 10.1016/j.ijrobp.2008.04.037
   Mizoe J, 2004, INT J RADIAT ONCOL, V60, P358, DOI 10.1016/j.ijrobp.2004.02.067
   Mizumoto M, 2010, STRAHLENTHER ONKOL, V186, P36, DOI 10.1007/s00066-009-2019-3
   Pignon JP, 2000, LANCET, V355, P949, DOI 10.1016/S0140-6736(00)90011-4
   Pommier P, 2006, ARCH OTOLARYNGOL, V132, P1242, DOI 10.1001/archotol.132.11.1242
   Shirasaka T, 1996, ANTI-CANCER DRUG, V7, P548, DOI 10.1097/00001813-199607000-00010
   Silverberg E, 1970, CA Cancer J Clin, V20, P11
   SUIT H, 1992, JNCI-J NATL CANCER I, V84, P155, DOI 10.1093/jnci/84.3.155
   Tokuuye K, 2004, STRAHLENTHER ONKOL, V180, P96, DOI 10.1007/s00066-004-1132-6
   Truong MT, 2009, HEAD NECK-J SCI SPEC, V31, P1297, DOI 10.1002/hed.21092
   Weber DC, 2006, RADIOTHER ONCOL, V81, P243, DOI 10.1016/j.radonc.2006.09.009
   Zenda S, INT J RAD O IN PRESS
NR 23
TC 35
Z9 35
U1 0
U2 7
PU ELSEVIER SCIENCE INC
PI NEW YORK
PA 360 PARK AVE SOUTH, NEW YORK, NY 10010-1710 USA
SN 0360-3016
J9 INT J RADIAT ONCOL
JI Int. J. Radiat. Oncol. Biol. Phys.
PD JUN 1
PY 2012
VL 83
IS 2
BP 704
EP 711
DI 10.1016/j.ijrobp.2011.07.032
PG 8
WC Oncology; Radiology, Nuclear Medicine & Medical Imaging
WE Science Citation Index Expanded (SCI-EXPANDED)
SC Oncology; Radiology, Nuclear Medicine & Medical Imaging
GA 940VI
UT WOS:000303920800056
PM 22099036
DA 2023-08-27
ER

PT J
AU Patel, NS
   Link, MJ
   Driscoll, CLW
   Neff, BA
   Van Gompel, JJ
   Lane, JI
   Carlson, ML
AF Patel, Neil S.
   Link, Michael J.
   Driscoll, Colin L. W.
   Neff, Brian A.
   Van Gompel, Jamie J.
   Lane, John, I
   Carlson, Matthew L.
TI Involvement of the Cochlear Aqueduct by Jugular Paraganglioma Is
   Associated With Sensorineural Hearing Loss
SO OTOLOGY & NEUROTOLOGY
LA English
DT Article
DE Cochlear aqueduct; Glomus jugulare; Jugular paraganglioma; Sensorineural
   hearing loss
ID MENINGITIS
AB Objective: The etiology of sensorineural hearing loss (SNHL) in patients with jugular paraganglioma (JP) whose tumors lack inner ear fistulae or vestibulocochlear nerve involvement is unknown. Recent literature has proposed that occlusion of the inferior cochlear vein may be causative. Herein, we assess the association between radiologic involvement of the cochlear aqueduct (CA) and the development of SNHL.
   Study Design: Blinded, retrospective review of imaging and audiometry.
   Setting: Tertiary center.
   Patients: Adults with JP.
   Intervention(s): None.
   Main Outcome Measures: Asymmetric SNHL was assessed continuously as the difference in bone conduction pure-tone average (BCPTA) between ears and as a categorical variable (>= 15 dB difference at two consecutive frequencies, or a difference in speech discrimination score of >= 15%). Involvement of the CA was considered present if there was evidence of medial T2 fluid signal loss, contrast enhancement, or bony erosion/expansion.
   Results: Of 30 patients meeting inclusion criteria, 15 (50%) had asymmetric SNHL. CA involvement was observed in 87% of patients with asymmetric SNHL compared with 13% in those with symmetric hearing (p = 0.0001). Univariate analysis demonstrated that age, sex, and tumor volume were not associated with asymmetric SNHL. The median difference in BCPTA between ears in patients with CA involvement was 21.3 dB HL compared to 1.2dB HL in those without CA involvement (p <0.0001). Regression analysis demonstrates that enhancement within the CA is associated with a BCPTA difference of 19.4 dB HL (p = 0.0006).
   Conclusions: Cochlear aqueduct involvement by JP is associated with SNHL in the absence of inner ear fistula, vestibulocochlear nerve involvement, or brainstem compression. Correlation with operative findings or histopathologic evidence of tumor involvement may validate this intriguing imaging finding.
C1 [Patel, Neil S.; Link, Michael J.; Driscoll, Colin L. W.; Neff, Brian A.; Van Gompel, Jamie J.; Carlson, Matthew L.] Mayo Clin, Dept Otolaryngol Head & Neck Surg, 200 1st St SW, Rochester, MN 55905 USA.
   [Link, Michael J.; Driscoll, Colin L. W.; Van Gompel, Jamie J.; Carlson, Matthew L.] Mayo Clin, Dept Neurol Surg, Rochester, MN 55905 USA.
   [Lane, John, I] Mayo Clin, Dept Radiol, Rochester, MN 55905 USA.
C3 Mayo Clinic; Mayo Clinic; Mayo Clinic
RP Carlson, ML (通讯作者)，Mayo Clin, Dept Otolaryngol Head & Neck Surg, 200 1st St SW, Rochester, MN 55905 USA.
EM
RI Van Gompel, Jamie/AAW-4459-2020
OI Van Gompel, Jamie/0000-0001-8087-7870
CR [Anonymous], 1995, Otolaryngol Head Neck Surg, V113, P179
   Cueva RA, 2004, LARYNGOSCOPE, V114, P1686, DOI 10.1097/00005537-200410000-00003
   Gopen Q, 1997, HEARING RES, V107, P9, DOI 10.1016/S0378-5955(97)00017-8
   Gurgel RK, 2012, OTOLARYNG HEAD NECK, V147, P102, DOI 10.1177/0194599812437156
   Isaacson B, 2019, LARYNGOSCOPE, V129, P67, DOI 10.1002/lary.27343
   JACKSON CG, 1982, ARCH OTOLARYNGOL, V108, P401
   KAY R, 1991, NEUROPATH APPL NEURO, V17, P485, DOI 10.1111/j.1365-2990.1991.tb00751.x
   Merchant SN, 1996, AM J OTOL, V17, P375
   PALVA T, 1972, ARCHIV OTOLARYNGOL, V96, P130
   Patel NS, 2019, J NEUROSURG, V130, P379, DOI 10.3171/2017.10.JNS17764
NR 10
TC 0
Z9 0
U1 0
U2 1
PU LIPPINCOTT WILLIAMS & WILKINS
PI PHILADELPHIA
PA TWO COMMERCE SQ, 2001 MARKET ST, PHILADELPHIA, PA 19103 USA
SN 1531-7129
EI 1537-4505
J9 OTOL NEUROTOL
JI Otol. Neurotol.
PD OCT
PY 2019
VL 40
IS 9
BP 1230
EP 1236
DI 10.1097/MAO.0000000000002346
PG 7
WC Clinical Neurology; Otorhinolaryngology
WE Science Citation Index Expanded (SCI-EXPANDED)
SC Neurosciences & Neurology; Otorhinolaryngology
GA KG1CG
UT WOS:000509678200028
PM 31469795
DA 2023-08-27
ER

PT J
AU Kam, MKM
   Chau, RMC
   Suen, J
   Choi, PHK
   Teo, PML
AF Kam, MKM
   Chau, RMC
   Suen, J
   Choi, PHK
   Teo, PML
TI Intensity-modulated radiotherapy in nasopharyngeal carcinoma: Dosimetric
   advantage over conventional plans and feasibility of dose escalation
SO INTERNATIONAL JOURNAL OF RADIATION ONCOLOGY BIOLOGY PHYSICS
LA English
DT Article; Proceedings Paper
CT 3rd S Takahashi Memorial International Workshop on 3-dimensional
   Conformal Radiotherapy
CY DEC 08-10, 2001
CL NAGOYA, JAPAN
DE nasopharyngeal carcinoma; intensity-modulation; dosimetry; radiotherapy
ID RADIATION-THERAPY; LOCAL-CONTROL; ACCELERATED FRACTIONATION;
   RETROSPECTIVE ANALYSIS; TUMOR-CONTROL; NECK; IRRADIATION; CANCER;
   VOLUME; HEAD
AB Purpose: To compare intensity-modulated radiotherapy (IMRT) with two-dimensional RT (2D-RT) and three-dimensional conformal radiotherapy (3D-CRT) treatment plans in different stages of nasopharyngeal carcinoma and to explore the feasibility of dose escalation in locally advanced disease.
   Methods and Materials: Three patients with different stages (T1N0M0, T2bN2M0 with retrostyloid extension, and T4N2M0) were selected, and 2D-RT, 3D-CRT, and IMRT treatment plans (66 Gy) were made for each of them and compared with respect to target coverage, normal tissue sparing, and tumor control probability/normal tissue complication probability values. In the Stage T2b and T4 patients, the IMRT 66-Gy plan was combined with a 3D-CRT 14-Gy boost plan using a 3-mm micromultileaf collimator, and the dose-volume histograms of the summed plans were compared with their corresponding 66-Gy 2D-RT plans.
   Results: In the dosimetric comparison of 2D-RT, 3D-CRT, and IMRT treatment plans, the T1N0M0 patient had better sparing of the parotid glands and temporomandibular joints with IMRT (dose to 50% parotid volume, 57 Gy, 50 Gy, and 31 Gy, respectively). In the T2bN2M0 patient, the dose to 95% volume of the planning target volume improved from 57.5 Gy in 2D-RT to 64.8 Gy in 3D-CRT and 68 Gy in IMRT. In the T4N2M0 patient, improvement in both target coverage and brainstem/temporal lobe sparing was seen with IMRT planning. In the dose-escalation study for locally advanced disease, IMRT 66 Gy plus 14 Gy 3D-CRT boost achieved an improvement in the therapeutic ratio by delivering a higher dose to the target while keeping the normal organs below the maximal tolerance dose.
   Conclusion: IMRT is useful in treating all stages of nonmetastatic nasopharyngeal carcinoma because of its dosimetric advantages. In early-stage disease, it provides better parotid gland sparing. In locally advanced disease, IMRT offers better tumor coverage and normal organ sparing and allows room for dose escalation. (C) 2003 Elsevier Inc.
C1 Chinese Univ Hong Kong, Prince Wales Hosp, Dept Clin Oncol, Shatin, Hong Kong, Peoples R China.
C3 Chinese University of Hong Kong; Prince of Wales Hospital
RP Kam, MKM (通讯作者)，Chinese Univ Hong Kong, Prince Wales Hosp, Dept Clin Oncol, Shatin, Hong Kong, Peoples R China.
CR Al-Sarraf M, 1998, J CLIN ONCOL, V16, P1310, DOI 10.1200/JCO.1998.16.4.1310
   [Anonymous], RAD RES S
   Chao KSC, 2001, INT J RADIAT ONCOL, V49, P907, DOI 10.1016/S0360-3016(00)01441-3
   Chau RMC, 2001, RADIOTHER ONCOL, V58, P143, DOI 10.1016/S0167-8140(00)00336-4
   Eisbruch A, 1999, INT J RADIAT ONCOL, V45, P577, DOI 10.1016/S0360-3016(99)00247-3
   Fu KK, 2000, INT J RADIAT ONCOL, V48, P7, DOI 10.1016/S0360-3016(00)00663-5
   Ho JHC, 1982, TREATMENT CANCER, P249
   Hunt MA, 2001, INT J RADIAT ONCOL, V49, P623, DOI 10.1016/S0360-3016(00)01389-4
   King AD, 1999, HEAD NECK-J SCI SPEC, V21, P614, DOI 10.1002/(SICI)1097-0347(199910)21:7<614::AID-HED5>3.0.CO;2-I
   KUTCHER GJ, 1989, INT J RADIAT ONCOL, V16, P1623, DOI 10.1016/0360-3016(89)90972-3
   Lee AWM, 1998, INT J RADIAT ONCOL, V40, P35, DOI 10.1016/S0360-3016(97)00580-4
   LEE AWM, 1992, INT J RADIAT ONCOL, V23, P261, DOI 10.1016/0360-3016(92)90740-9
   Lee AWM, 2001, RADIOTHER ONCOL, V58, P121, DOI 10.1016/S0167-8140(00)00312-1
   Leung TW, 2000, INT J RADIAT ONCOL, V47, P405, DOI 10.1016/S0360-3016(00)00463-6
   Nowak PJCM, 1999, INT J RADIAT ONCOL, V45, P33, DOI 10.1016/S0360-3016(99)00049-8
   SHAM JST, 1990, BRIT J RADIOL, V63, P51, DOI 10.1259/0007-1285-63-745-51
   Spirou SV, 1998, MED PHYS, V25, P321, DOI 10.1118/1.598202
   Sultanem K, 2000, INT J RADIAT ONCOL, V48, P711, DOI 10.1016/S0360-3016(00)00702-1
   Tate DJ, 1999, INT J RADIAT ONCOL, V45, P915, DOI 10.1016/S0360-3016(99)00296-5
   Teo P, 1996, INT J RADIAT ONCOL, V36, P291, DOI 10.1016/S0360-3016(96)00323-9
   Teo PML, 2000, RADIOTHER ONCOL, V57, P155, DOI 10.1016/S0167-8140(00)00248-6
   Teo PML, 2000, INT J RADIAT ONCOL, V46, P445, DOI 10.1016/S0360-3016(99)00326-0
   Wolden SL, 2001, INT J RADIAT ONCOL, V49, P1229, DOI 10.1016/S0360-3016(00)01588-1
   Xia P, 2000, INT J RADIAT ONCOL, V48, P329, DOI 10.1016/S0360-3016(00)00585-X
NR 24
TC 238
Z9 290
U1 0
U2 16
PU ELSEVIER SCIENCE INC
PI NEW YORK
PA 360 PARK AVE SOUTH, NEW YORK, NY 10010-1710 USA
SN 0360-3016
J9 INT J RADIAT ONCOL
JI Int. J. Radiat. Oncol. Biol. Phys.
PD MAY 1
PY 2003
VL 56
IS 1
BP 145
EP 157
DI 10.1016/S0360-3016(03)00075-0
PG 13
WC Oncology; Radiology, Nuclear Medicine & Medical Imaging
WE Conference Proceedings Citation Index - Science (CPCI-S); Science Citation Index Expanded (SCI-EXPANDED)
SC Oncology; Radiology, Nuclear Medicine & Medical Imaging
GA 668TT
UT WOS:000182309800018
PM 12694833
DA 2023-08-27
ER

PT J
AU Peciu-Florianu, I
   Tuleasca, C
   Comps, JN
   Schiappacasse, L
   Zeverino, M
   Daniel, RT
   Levivier, M
AF Peciu-Florianu, Iulia
   Tuleasca, Constantin
   Comps, Jean-Nicolas
   Schiappacasse, Luis
   Zeverino, Michele
   Daniel, Roy Thomas
   Levivier, Marc
TI Radiosurgery in trochlear and abducens nerve schwannomas: case series
   and systematic review
SO ACTA NEUROCHIRURGICA
LA English
DT Review
DE Radiosurgery; Gamma knife surgery; Planned subtotal resection;
   Schwannoma; Trochlear; Abducens
ID GAMMA-KNIFE SURGERY; OF-THE-LITERATURE; FRACTION STEREOTACTIC
   RADIOSURGERY; INDUCED OPTIC NEUROPATHY; 6TH CRANIAL NERVES; OCULOMOTOR
   NERVE; VESTIBULAR SCHWANNOMAS; CAVERNOUS SINUS; NONVESTIBULAR
   SCHWANNOMAS; CLINICAL ARTICLE
AB Schwannomas involving the occulomotor cranial nerves (CNs; III, IV and VI), can be disabling, due to the associated diplopia and decreased quality of life and are extremely rare. We evaluated the role of Gamma Knife surgery (GKS) in these cases.
   Five patients with CN IV and VI schwannomas (three and two, respectively) were treated in Lausanne University Hospital between 2010 and 2015. Four benefitted from upfront GKS and one from a combined approach (planned subtotal resection followed by GKS), due to a large preoperative tumour volume (size, 3 x 2 x 2.5 cm; volume, 7.9 ml), with symptomatic mass effect and oedema, as well as an entrapement cyst at the brainstem interface, in a young patient. Neuro-ophtalmological evaluation was performed at baseline and during each follow-up time-point. A systematic literature review is presented and compared to the present report.
   The mean follow-up was 44.4 months (12-54). Initial clinical presentation was diplopia in four cases and cavernous sinus syndrome in one. The marginal dose was 12 Gy in all cases. The mean target volume was 1.51 cm(3) (0.086-5.8). The mean prescription isodose volume (PIV) was 1.71 cm(3) (0.131-6.7). At last follow-up, all patients presented with disappearance of the baseline symptoms. Tumour control was achieved in 100%, with decrease in volume in all cases. The systematic review analysed 11 peer-reviewed studies, with a total of 35 patients. For uniformly reported CN VI, the mean marginal radiation dose ranged between 12 and 12.5 Gy, with disappearance of symptoms in 12.5%, improvement in 31.25%, stabilisation in 6.25%, worsening in 12.5%. Tumour volume decreased in all cases.
   Our data suggest that first intention GKS is a safe and effective option for patients with small to medium size oculomotor schwannomas, providing a high rate of clinical alleviation and tumour control. When the initial tumour volume is too large for first intention GKS, a combined approach with planned subtotal resection followed by GKS can be performed, with favourable and comparable outcomes as in upfront GKS.
C1 [Peciu-Florianu, Iulia; Tuleasca, Constantin; Comps, Jean-Nicolas; Daniel, Roy Thomas; Levivier, Marc] Lausanne Univ Hosp CHUV, Neurosci Dept, Neurosurg Serv, Rue Bugnon 44-46,BH-08, CH-1011 Lausanne, Switzerland.
   [Peciu-Florianu, Iulia; Tuleasca, Constantin; Comps, Jean-Nicolas; Daniel, Roy Thomas; Levivier, Marc] Lausanne Univ Hosp CHUV, Gamma Knife Ctr, Rue Bugnon 44-46,BH-08, CH-1011 Lausanne, Switzerland.
   [Peciu-Florianu, Iulia; Tuleasca, Constantin; Daniel, Roy Thomas; Levivier, Marc] Univ Lausanne, Fac Biol & Med, Lausanne, Switzerland.
   [Peciu-Florianu, Iulia] CHRU Lille, Dept Neurosurg & Neurooncol, Lille, France.
   [Tuleasca, Constantin] Swiss Fed Inst Technol, Signal Proc Lab LTS5, Lausanne, Switzerland.
   [Schiappacasse, Luis] Lausanne Univ Hosp CHUV, Radiotherapy Dept, Lausanne, Switzerland.
   [Zeverino, Michele] Inst Radiat Phys Lausanne, Lausanne, Switzerland.
C3 University of Lausanne; Centre Hospitalier Universitaire Vaudois (CHUV);
   University of Lausanne; Centre Hospitalier Universitaire Vaudois (CHUV);
   University of Lausanne; Universite de Lille - ISITE; CHU Lille; Swiss
   Federal Institutes of Technology Domain; Ecole Polytechnique Federale de
   Lausanne; University of Lausanne; Centre Hospitalier Universitaire
   Vaudois (CHUV)
RP Tuleasca, C (通讯作者)，Lausanne Univ Hosp CHUV, Neurosci Dept, Neurosurg Serv, Rue Bugnon 44-46,BH-08, CH-1011 Lausanne, Switzerland.; Tuleasca, C (通讯作者)，Lausanne Univ Hosp CHUV, Gamma Knife Ctr, Rue Bugnon 44-46,BH-08, CH-1011 Lausanne, Switzerland.; Tuleasca, C (通讯作者)，Univ Lausanne, Fac Biol & Med, Lausanne, Switzerland.; Tuleasca, C (通讯作者)，Swiss Fed Inst Technol, Signal Proc Lab LTS5, Lausanne, Switzerland.
EM
OI Zeverino, Michele/0000-0001-9941-8252; TULEASCA,
   Constantin/0000-0001-6776-1486
FU Lausanne University Hospital (CHUV)
FX Lausanne University Hospital (CHUV).
CR Acharya R, 2003, NEUROL SCI, V24, P74, DOI 10.1007/s100720300076
   Asaoka K, 1999, NEUROSURGERY, V45, P630, DOI 10.1097/00006123-199909000-00039
   BARAT JL, 1992, NEUROCHIRURGIE, V38, P183
   Beppu Takaaki, 1997, Neurologia Medico-Chirurgica, V37, P416, DOI 10.2176/nmc.37.416
   CELLI P, 1992, SURG NEUROL, V38, P216, DOI 10.1016/0090-3019(92)90172-J
   Chakrabarti KB, 1996, BRIT J NEUROSURG, V10, P577
   CHEN BH, 1981, NEUROSURGERY, V9, P64
   Choi CYH, 2011, NEUROSURGERY, V68, P1200, DOI 10.1227/NEU.0b013e31820c0474
   Chowdhury FH, 2014, J NEUROSCI RURAL PRA, V5, P231, DOI 10.4103/0976-3147.133566
   Daniel RT, 2017, ACTA NEUROCHIR, V159, P1197, DOI 10.1007/s00701-017-3194-0
   Daniel RT, 2013, MINIMALLY INVASIVE S
   DERUTY R, 1988, NEUROCHIRURGIE, V34, P287
   Elflein HM, 2010, STRABISMUS, V18, P83, DOI 10.3109/09273972.2010.502959
   Elmalem VI, 2009, OPHTHALMOLOGY, V116, P2011, DOI 10.1016/j.ophtha.2009.03.054
   Elsharkawy M, 2012, J NEUROSURG, V116, P66, DOI 10.3171/2011.8.JNS11215
   Erlich SA, 2009, CLIN NEUROL NEUROSUR, V111, P467, DOI 10.1016/j.clineuro.2009.01.001
   GINSBERG F, 1988, J COMPUT ASSIST TOMO, V12, P482, DOI 10.1097/00004728-198805010-00024
   Haselsberger K, 2009, J NEUROL NEUROSUR PS, V80, P1172, DOI 10.1136/jnnp.2008.156745
   Hatae R, 2014, J NEUROL SURG REP, V75, DOI 10.1055/s-0034-1378156
   Hayashi M, 2006, J NEUROSURG, V105, P12, DOI 10.3171/sup.2006.105.7.12
   Hayashi M, 2010, J NEUROSURG, V113, P136, DOI 10.3171/2010.8.GKS10947
   Ichimi Kazuyoshi, 1997, Neurologia Medico-Chirurgica, V37, P197, DOI 10.2176/nmc.37.197
   Inoue T, 2015, J NEUROL SURG REP, V76, pE248, DOI 10.1055/s-0035-1564059
   Irace C, 2008, ACTA NEUROCHIR, V150, P1209, DOI 10.1007/s00701-008-0134-z
   IWABUCHI T, 1982, NEUROSURGERY, V10, P490, DOI 10.1227/00006123-198204000-00013
   Kim IY, 2008, J NEUROSURG, V109, P149, DOI 10.3171/JNS/2008/109/12/S23
   Kim R, 2015, CEPHALALGIA, V35, P825, DOI 10.1177/0333102414558886
   Kimball MM, 2011, NEUROSURGERY, V68, P974, DOI 10.1227/NEU.0b013e318208f3a1
   Korchi AM, 2014, DIAGN INTERV IMAG, V95, P849, DOI 10.1016/j.diii.2013.04.013
   Kumar LP, 2014, J CANCER RES THER, V10, P1098, DOI 10.4103/0973-1482.146111
   Lanotte M, 1992, J Neurosurg Sci, V36, P233
   Leavitt JA, 2013, INT J RADIAT ONCOL, V87, P524, DOI 10.1016/j.ijrobp.2013.06.2047
   LEUNDA G, 1982, J NEUROSURG, V57, P563, DOI 10.3171/jns.1982.57.4.0563
   Lo PA, 2001, J CLIN NEUROSCI, V8, P357, DOI 10.1054/jocn.2000.0846
   LUNARDI P, 1990, CLIN NEUROL NEUROSUR, V92, P333, DOI 10.1016/0303-8467(90)90060-I
   Mascarenhas L, 2004, ACTA NEUROCHIR, V146, P389, DOI 10.1007/s00701-003-0197-9
   MEHTA V S, 1990, British Journal of Neurosurgery, V4, P69, DOI 10.3109/02688699009000685
   Mindermann T, 2014, ACTA NEUROCHIR, V156, P1121, DOI 10.1007/s00701-014-2063-3
   Moses JE, 2011, NEUROL INDIA, V59, P471, DOI 10.4103/0028-3886.82743
   Nakagawa T, 2004, SURG NEUROL, V61, P559, DOI 10.1016/S0090-3019(03)00580-9
   Nakamura M, 2002, SURG NEUROL, V57, P183, DOI 10.1016/S0090-3019(01)00670-X
   Nishioka K, 2009, INT J RADIAT ONCOL, V75, P1415, DOI 10.1016/j.ijrobp.2008.12.063
   Okada Y, 1997, NEUROSURGERY, V40, P608, DOI 10.1097/00006123-199703000-00035
   Park JH, 2009, NEUROSURG REV, V32, P375, DOI 10.1007/s10143-009-0203-7
   Pendl G, 2000, J NEUROSURG, V93, P107, DOI 10.3171/jns.2000.93.supplement_3.0107
   Petrela E, 2009, J NEURO-OPHTHALMOL, V29, P54, DOI 10.1097/WNO.0b013e31818f6463
   Pollock BE, 2002, INT J RADIAT ONCOL, V52, P1002, DOI 10.1016/S0360-3016(01)02711-0
   Pollock BE, 2014, NEUROSURGERY, V75, P456, DOI 10.1227/NEU.0000000000000457
   Prabhu Sujit S, 2010, Surg Neurol Int, V1, P15, DOI 10.4103/2152-7806.63910
   Prasad GL, 2016, J NEUROSURG, P1
   Razek AAKA, 2009, AM J NEURORADIOL, V30, P444, DOI 10.3174/ajnr.A1398
   Samii M, 2010, J NEUROSURG, V112, P860, DOI 10.3171/2009.7.JNS0989
   SEKHAR LN, 1992, J NEUROSURG, V76, P935, DOI 10.3171/jns.1992.76.6.0935
   Showalter TN, 2008, NEUROSURGERY, V63, P734, DOI 10.1227/01.NEU.0000325496.10148.B3
   Sirin S, 2008, NEUROSURGERY, V62, P744, DOI [10.1227/01.neu.0000316278.14748.87, 10.1227/01.NEU.0000190653.42970.6B]
   Stangerup SE, 2006, OTOL NEUROTOL, V27, P547, DOI 10.1097/00129492-200606000-00018
   Suetake K, 1998, COMPUT MED IMAG GRAP, V22, P257, DOI 10.1016/S0895-6111(98)00023-8
   TAKANO S, 1990, Neurologia Medico-Chirurgica, V30, P132, DOI 10.2176/nmc.30.132
   Tanriover N, 2007, SURG NEUROL, V67, P83, DOI 10.1016/j.surneu.2006.06.012
   Tos M, 2004, ARCH OTOLARYNGOL, V130, P216, DOI 10.1001/archotol.130.2.216
   TUNG H, 1991, J NEUROSURG, V75, P638, DOI 10.3171/jns.1991.75.4.0638
   Vachata P, 2009, ACTA NEUROCHIR, V151, P1281, DOI 10.1007/s00701-009-0302-9
   van de Langenberg R, 2011, J NEUROSURG, V115, P875, DOI 10.3171/2011.6.JNS101958
NR 63
TC 15
Z9 15
U1 0
U2 1
PU SPRINGER WIEN
PI WIEN
PA SACHSENPLATZ 4-6, PO BOX 89, A-1201 WIEN, AUSTRIA
SN 0001-6268
EI 0942-0940
J9 ACTA NEUROCHIR
JI Acta Neurochir.
PD DEC
PY 2017
VL 159
IS 12
BP 2409
EP 2418
DI 10.1007/s00701-017-3348-0
PG 10
WC Clinical Neurology; Surgery
WE Science Citation Index Expanded (SCI-EXPANDED)
SC Neurosciences & Neurology; Surgery
GA FM8QP
UT WOS:000415354800027
PM 29022157
DA 2023-08-27
ER

PT J
AU Boyle, Y
   Johns, TG
   Fletcher, EV
AF Boyle, Yasmin
   Johns, Terrance G.
   Fletcher, Emily, V
TI Potassium Ion Channels in Malignant Central Nervous System Cancers
SO CANCERS
LA English
DT Review
DE potassium ion channels; high grade glioma; glioblastoma;
   medulloblastoma; cellular plasticity
ID VASCULAR SMOOTH-MUSCLE; STEM-LIKE CELLS; K+ CHANNELS; HUMAN
   OSTEOSARCOMA; CRYSTAL-STRUCTURE; NEURONAL-ACTIVITY; RANDOMIZED-TRIAL;
   GENE-EXPRESSION; HERG1 CHANNELS; BK CHANNELS
AB Simple Summary Malignant central nervous system (CNS) cancers are notoriously difficult to treat, with just one-third of patients surviving five years post-diagnosis. Their location within the brain and brainstem presents several challenges to successful treatment, including surgical inaccessibility and designing effective therapies that pass the brain's protective barrier. Furthermore, high-grade CNS cancer is also prone to recurrence, and the secondary tumours that arise are highly resistant to treatment. Potassium ion channels are a class of transmembrane proteins involved in maintaining the electrical microenvironment of cells. In cancers, these proteins are known to play a significant role in the development of cancer hallmarks like proliferation, invasion, and adaptive drug resistance. Our review explores the relationship between potassium ion channel function and the progression of malignant CNS cancers. Targeting these proteins presents a promising, novel treatment strategy, with several FDA-approved potassium ion channel-targeting drugs already in clinical use for various CNS disorders. Malignant central nervous system (CNS) cancers are among the most difficult to treat, with low rates of survival and a high likelihood of recurrence. This is primarily due to their location within the CNS, hindering adequate drug delivery and tumour access via surgery. Furthermore, CNS cancer cells are highly plastic, an adaptive property that enables them to bypass targeted treatment strategies and develop drug resistance. Potassium ion channels have long been implicated in the progression of many cancers due to their integral role in several hallmarks of the disease. Here, we will explore this relationship further, with a focus on malignant CNS cancers, including high-grade glioma (HGG). HGG is the most lethal form of primary brain tumour in adults, with the majority of patient mortality attributed to drug-resistant secondary tumours. Hence, targeting proteins that are integral to cellular plasticity could reduce tumour recurrence, improving survival. This review summarises the role of potassium ion channels in malignant CNS cancers, specifically how they contribute to proliferation, invasion, metastasis, angiogenesis, and plasticity. We will also explore how specific modulation of these proteins may provide a novel way to overcome drug resistance and improve patient outcomes.
C1 [Boyle, Yasmin; Johns, Terrance G.; Fletcher, Emily, V] Perth Childrens Hosp, Telethon Kids Inst, 15 Hosp Ave, Perth, WA 6009, Australia.
   [Boyle, Yasmin; Johns, Terrance G.; Fletcher, Emily, V] Univ Western Australia, Sch Biomed, 35 Stirling Hwy, Perth, WA 6009, Australia.
C3 Telethon Kids Institute; University of Western Australia; University of
   Western Australia
RP Boyle, Y (通讯作者)，Perth Childrens Hosp, Telethon Kids Inst, 15 Hosp Ave, Perth, WA 6009, Australia.; Boyle, Y (通讯作者)，Univ Western Australia, Sch Biomed, 35 Stirling Hwy, Perth, WA 6009, Australia.
EM
OI Fletcher, Emily/0000-0002-9786-4221
FU Telethon Kids Institute; Robert Connor Dawes Foundation; Cure Starts
   Now; Brain Tumour Charity [GN-000546]; Perth Children's Hospital
   Foundation [9896]; Australian Government Research Training Program
   Scholarship at The University of Western Australia
FX The research work related to the review in the author's lab is funded by
   multiple sources. This includes direct funding from Telethon Kids
   Institute and philanthropic donations made by the Ethan Davies
   Fellowship and The Robert Connor Dawes Foundation. Funding is also
   provided by grants from The Cure Starts Now, The Brain Tumour Charity,
   grant number GN-000546 and The Perth Children's Hospital Foundation,
   grant number 9896. At the time of writing, the first author of this
   review was receiving the support of the Australian Government Research
   Training Program Scholarship at The University ofWestern Australia.
CR Abdul M, 2002, CANCER LETT, V186, P99, DOI 10.1016/S0304-3835(02)00348-8
   Abdul M, 2006, J MEMBRANE BIOL, V214, P99, DOI 10.1007/s00232-006-0065-7
   Abdullaev IF, 2010, PLOS ONE, V5, DOI 10.1371/journal.pone.0012304
   Adam MA, 2010, ONCOLOGIST, V15, P61, DOI 10.1634/theoncologist.2009-0185
   Aissaoui D, 2018, INT J BIOL MACROMOL, V111, P1146, DOI 10.1016/j.ijbiomac.2018.01.144
   Alberts B., 2008, MOL BIOL CELL, V5th edn
   An ZY, 2018, ONCOGENE, V37, P1561, DOI 10.1038/s41388-017-0045-7
   Anumonwo JMB, 2010, J MOL CELL CARDIOL, V48, P45, DOI 10.1016/j.yjmcc.2009.08.013
   Arvind S, 2012, BRIT J NEUROSURG, V26, P16, DOI 10.3109/02688697.2011.583365
   Augustus M, 2021, CANCERS, V13, DOI 10.3390/cancers13092107
   Banderali U, 2011, CELL PHYSIOL BIOCHEM, V28, P1169, DOI 10.1159/000335850
   Barcia G, 2012, NAT GENET, V44, P1255, DOI 10.1038/ng.2441
   Batchelor TT, 2013, J CLIN ONCOL, V31, P3212, DOI 10.1200/JCO.2012.47.2464
   Bausch AE, 2015, LEARN MEMORY, V22, P323, DOI 10.1101/lm.037820.114
   Belus MT, 2018, DEV BIOL, V444, pS297, DOI 10.1016/j.ydbio.2018.02.012
   Bi D, 2013, J BIOL CHEM, V288, P15843, DOI 10.1074/jbc.M112.427187
   Bielanska J, 2012, ONCOL LETT, V4, P227, DOI 10.3892/ol.2012.718
   Bijlenga P, 1998, J PHYSIOL-LONDON, V512, P317, DOI 10.1111/j.1469-7793.1998.317be.x
   Blackiston DJ, 2009, CELL CYCLE, V8, P3527, DOI 10.4161/cc.8.21.9888
   Bordey A, 1998, J NEUROPHYSIOL, V79, P2782, DOI 10.1152/jn.1998.79.5.2782
   Bouffet E, 2022, J CLIN ONCOL, V40
   Bourkoula E, 2014, STEM CELLS, V32, P1239, DOI 10.1002/stem.1605
   Brandalise F, 2020, FRONT NEUROSCI-SWITZ, V14, DOI 10.3389/fnins.2020.595664
   Brendorp B, 2002, DRUG SAFETY, V25, P847, DOI 10.2165/00002018-200225120-00003
   Brevet M, 2009, CELL PHYSIOL BIOCHEM, V24, P25, DOI 10.1159/000227810
   Brohawn SG, 2012, SCIENCE, V335, P436, DOI 10.1126/science.1213808
   BRUGGEMANN A, 1993, NATURE, V365, P445, DOI 10.1038/365445a0
