## Supplemental Material 1 for "Bibliometric analysis of emerging trends and research foci in brainstem tumor field over thirty years (1992-2023)": savedrecs (10).txt

FN Clarivate Analytics Web of Science
VR 1.0
PT J
AU Walters, BJ
   Liu, ZY
   Crabtree, M
   Coak, E
   Cox, BC
   Zuo, J
AF Walters, Bradley J.
   Liu, Zhiyong
   Crabtree, Mark
   Coak, Emily
   Cox, Brandon C.
   Zuo, Jian
TI Auditory Hair Cell-Specific Deletion of p27(Kip1) in Postnatal Mice
   Promotes Cell-Autonomous Generation of New Hair Cells and Normal Hearing
SO JOURNAL OF NEUROSCIENCE
LA English
DT Article
DE Cdkn1b; Cip/Kip; cochlea; cyclins; regeneration; retinoblastoma
ID DEPENDENT KINASE INHIBITOR; WALTZING GUINEA-PIG; MOUSE INNER-EAR;
   IN-VIVO; RETINOBLASTOMA PROTEIN; POSTMITOTIC STATE; GENE-EXPRESSION;
   STEM-CELLS; PROLIFERATION; COCHLEA
AB Hearing in mammals relies upon the transduction of sound by hair cells (HCs) in the organ of Corti within the cochlea of the inner ear. Sensorineural hearing loss is a widespread and permanent disability due largely to a lack of HC regeneration in mammals. Recent studies suggest that targeting the retinoblastoma (Rb)/E2F pathway can elicit proliferation of auditory HCs. However, previous attempts to induce HC proliferation in this manner have resulted in abnormal cochlear morphology, HC death, and hearing loss. Here we show that cochlear HCs readily proliferate and survive following neonatal, HC-specific, conditional knock-out of p27(Kip1) (p27CKO), a tumor suppressor upstream of Rb. Indeed, HC-specific p27CKO results in proliferation of these cells without the upregulation of the supporting cell or progenitor cell proteins, Prox1 or Sox2, suggesting that they remain HCs. Furthermore, p27CKO leads to a significant addition of postnatally derived HCs that express characteristic synaptic and stereociliary markers and survive to adulthood, although a portion of the newly derived inner HCs exhibit cytocauds and lack VGlut3 expression. Despite this, p27CKO mice exhibit normal hearing as measured by evoked auditory brainstem responses, which suggests that the newly generated HCs may contribute to, or at least do not greatly detract from, function. These results show that p27(Kip1) actively maintains HC quiescence in postnatal mice, and suggest that inhibition of p27(Kip1) in residual HCs represents a potential strategy for cell-autonomous auditory HC regeneration.
C1 [Walters, Bradley J.; Liu, Zhiyong; Crabtree, Mark; Coak, Emily; Cox, Brandon C.; Zuo, Jian] St Jude Childrens Res Hosp, Dept Dev Neurobiol, Memphis, TN 38105 USA.
C3 St Jude Children's Research Hospital
RP Zuo, J (通讯作者)，St Jude Childrens Res Hosp, Dept Dev Neurobiol, 262 Danny Thomas Pl,MS 323, Memphis, TN 38105 USA.
EM
RI Zuo, Jian/HPD-5969-2023; Cox, Brandon/AAJ-1899-2020
OI Cox, Brandon/0000-0002-6989-161X
FU National Institutes of Health [DC006471, DC010310, P30CA21765]; American
   Lebanese Syrian Associated Charities of St. Jude Children's Research
   Hospital; Office of Naval Research [N000140911014, N000141210191,
   N000141210775]; National Organization for Hearing Research Foundation;
   Hearing Health Foundation [Emerging Research Grant]; Hartwell Foundation
   [Individual Biomedical Research Award]
FX This work was supported by funding from the National Institutes of
   Health [Grants DC006471 (J.Z.), DC010310 (B.C.C.), P30CA21765 (St.
   Jude)], the American Lebanese Syrian Associated Charities of St. Jude
   Children's Research Hospital, the Office of Naval Research [Grants
   N000140911014, N000141210191, N000141210775 (J.Z.)], the National
   Organization for Hearing Research Foundation (B.J.W.), the Hearing
   Health Foundation [Emerging Research Grant (B.J.W.)], and The Hartwell
   Foundation [Individual Biomedical Research Award (J.Z.)]. The authors
   thank Lingli Zhang and Mario Sauceda for assistance with genotyping,
   Jennifer Dearman for technical assistance, and Drs. Brandon Walters and
   Wanda Layman for providing helpful feedback on experimental designs and
   statistical analyses.
CR Akil O, 2012, NEURON, V75, P283, DOI 10.1016/j.neuron.2012.05.019
   Bermingham-McDonogh O, 2006, J COMP NEUROL, V496, P172, DOI 10.1002/cne.20944
   Bryda EC, 2012, MAMM GENOME, V23, P270, DOI 10.1007/s00335-011-9381-z
   Burns J, 2008, J COMP NEUROL, V511, P396, DOI 10.1002/cne.21849
   CHELLAPPAN SP, 1991, CELL, V65, P1053, DOI 10.1016/0092-8674(91)90557-F
   Chen P, 2003, NAT CELL BIOL, V5, P422, DOI 10.1038/ncb976
   Chen P, 1999, DEVELOPMENT, V126, P1581
   Chien WM, 2006, P NATL ACAD SCI USA, V103, P4122, DOI 10.1073/pnas.0509514103
   Chow LML, 2006, DEV DYNAM, V235, P2991, DOI 10.1002/dvdy.20948
   Cox BC, 2014, DEVELOPMENT, V141, P816, DOI 10.1242/dev.103036
   Cox BC, 2012, JARO-J ASSOC RES OTO, V13, P295, DOI 10.1007/s10162-012-0324-5
   Dor Y, 2004, NATURE, V429, P41, DOI 10.1038/nature02520
   Eulalio A, 2012, NATURE, V492, P376, DOI 10.1038/nature11739
   Flock A, 1979, Adv Otorhinolaryngol, V25, P12
   Goodyear RJ, 2012, J NEUROSCI, V32, P2762, DOI 10.1523/JNEUROSCI.3635-11.2012
   Gubbels SP, 2008, NATURE, V455, P537, DOI 10.1038/nature07265
   GURLEY LR, 1973, BIOCHEM BIOPH RES CO, V50, P744, DOI 10.1016/0006-291X(73)91307-7
   Henriet P, 2000, P NATL ACAD SCI USA, V97, P10026, DOI 10.1073/pnas.170290997
   Huang MQ, 2011, CELL CYCLE, V10, P337, DOI 10.4161/cc.10.2.14640
   Izumikawa M, 2005, NAT MED, V11, P271, DOI 10.1038/nm1193
   Izumikawa M, 2008, HEARING RES, V240, P52, DOI 10.1016/j.heares.2008.02.007
   Kaiser CL, 2009, LARYNGOSCOPE, V119, P1770, DOI 10.1002/lary.20557
   Kanzaki S, 2002, AUDIOL NEURO-OTOL, V7, P289, DOI 10.1159/000064447
   Kanzaki S, 2006, HEARING RES, V214, P28, DOI 10.1016/j.heares.2006.01.014
   Kawamoto K, 2003, J NEUROSCI, V23, P4395
   Kiernan AE, 2005, NATURE, V434, P1031, DOI 10.1038/nature03487
   Kraft S, 2013, LARYNGOSCOPE, V123, P992, DOI 10.1002/lary.22171
   Kusaba T, 2014, P NATL ACAD SCI USA, V111, P1527, DOI 10.1073/pnas.1310653110
   Laine H, 2007, J NEUROSCI, V27, P1434, DOI 10.1523/JNEUROSCI.4956-06.2007
   Laine H, 2010, DEV BIOL, V337, P134, DOI 10.1016/j.ydbio.2009.10.027
   Lee YS, 2006, DEVELOPMENT, V133, P2817, DOI 10.1242/dev.02453
   Liu HZ, 2014, J NEUROSCI, V34, P11085, DOI 10.1523/JNEUROSCI.1690-14.2014
   Liu ZY, 2014, PLOS ONE, V9, DOI 10.1371/journal.pone.0089377
   Liu ZY, 2012, J NEUROSCI, V32, P10530, DOI 10.1523/JNEUROSCI.0686-12.2012
   Liu ZY, 2012, PLOS ONE, V7, DOI [10.1371/journal.pone.0050367, 10.1371/journal.pone.0034123]
   Liu ZY, 2012, J NEUROSCI, V32, P6600, DOI 10.1523/JNEUROSCI.0818-12.2012
   Liu ZY, 2012, DEV DYNAM, V241, P684, DOI 10.1002/dvdy.23754
   Lowenheim H, 1999, P NATL ACAD SCI USA, V96, P4084, DOI 10.1073/pnas.96.7.4084
   Mantela J, 2005, DEVELOPMENT, V132, P2377, DOI 10.1242/dev.01834
   Marino S, 2000, GENE DEV, V14, P994
   McClatchey AI, 2012, CURR OPIN CELL BIOL, V24, P685, DOI 10.1016/j.ceb.2012.06.009
   MCGILL TJI, 1976, LARYNGOSCOPE, V86, P1293, DOI 10.1288/00005537-197609000-00001
   Mizutari K, 2013, NEURON, V77, P58, DOI 10.1016/j.neuron.2012.10.032
   Oesterle EC, 2011, CELL CYCLE, V10, P1237, DOI 10.4161/cc.10.8.15301
   Oshima K, 2010, CELL, V141, P704, DOI 10.1016/j.cell.2010.03.035
   Peng Z, 2013, BRAIN RES, V1537, P100, DOI 10.1016/j.brainres.2013.09.019
   POLYAK K, 1994, CELL, V78, P59, DOI 10.1016/0092-8674(94)90572-X
   Ronaghi M, 2014, STEM CELLS DEV, V23, P1275, DOI 10.1089/scd.2014.0033
   Ruben RJ, 1967, ACTA OTO-LARYNGOL, V220, P221
   Sage C, 2005, SCIENCE, V307, P1114, DOI 10.1126/science.1106642
   Sage C, 2006, P NATL ACAD SCI USA, V103, P7345, DOI 10.1073/pnas.0510631103
   Salic A, 2008, P NATL ACAD SCI USA, V105, P2415, DOI 10.1073/pnas.0712168105
   Sato T, 2009, ACTA OTO-LARYNGOL, V129, P12, DOI 10.1080/00016480902926464
   Seal RP, 2008, NEURON, V57, P263, DOI 10.1016/j.neuron.2007.11.032
   Senyo SE, 2013, NATURE, V493, P433, DOI 10.1038/nature11682
   Son EJ, 2012, PLOS ONE, V7, DOI 10.1371/journal.pone.0040735
   Stockinger A, 2001, J CELL BIOL, V154, P1185, DOI 10.1083/jcb.200104036
   Uttamapinant C, 2012, ANGEW CHEM INT EDIT, V51, P5852, DOI 10.1002/anie.201108181
   Van Hooser A, 1998, J CELL SCI, V111, P3497
   Weber T, 2008, P NATL ACAD SCI USA, V105, P781, DOI 10.1073/pnas.0708061105
   Wu XD, 2004, MOL BRAIN RES, V126, P30, DOI 10.1016/j.molbrainres.2004.03.020
   Yu YL, 2010, J NEUROSCI, V30, P5927, DOI 10.1523/JNEUROSCI.5989-09.2010
NR 62
TC 31
Z9 32
U1 0
U2 12
PU SOC NEUROSCIENCE
PI WASHINGTON
PA 11 DUPONT CIRCLE, NW, STE 500, WASHINGTON, DC 20036 USA
SN 0270-6474
J9 J NEUROSCI
JI J. Neurosci.
PD NOV 19
PY 2014
VL 34
IS 47
BP 15751
EP 15763
DI 10.1523/JNEUROSCI.3200-14.2014
PG 13
WC Neurosciences
WE Science Citation Index Expanded (SCI-EXPANDED)
SC Neurosciences & Neurology
GA AU9JD
UT WOS:000345907500024
PM 25411503
OA hybrid, Green Published
DA 2023-08-27
ER

PT J
AU Mengue, L
   Bertaut, A
   Mbus, LN
   Dore, M
   Ayadi, M
   Clement-Colmou, K
   Claude, L
   Carrie, C
   Laude, C
   Tanguy, R
   Blanc, J
   Sunyach, MP
AF Mengue, Laurence
   Bertaut, Aurelie
   Mbus, Louise Ngo
   Dore, Melanie
   Ayadi, Myriam
   Clement-Colmou, Karen
   Claude, Line
   Carrie, Christian
   Laude, Cecile
   Tanguy, Ronan
   Blanc, Julie
   Sunyach, Marie-Pierre
TI Brain metastases treated with hypofractionated stereotactic
   radiotherapy: 8 years experience after Cyberknife installation
SO RADIATION ONCOLOGY
LA English
DT Article
DE Hypofractionated stereotactic radiotherapy; Brain metastases;
   Radionecrosis
ID GRADED PROGNOSTIC ASSESSMENT; CELL LUNG-CANCER; RADIATION-THERAPY;
   SURGICAL RESECTION; RADIOSURGERY; MANAGEMENT; RISK; GUIDELINES;
   DIAGNOSIS; TRIAL
AB Background Hypofractionated stereotactic radiotherapy (HFSRT) is indicated for large brain metastases (BM) or proximity to critical organs (brainstem, chiasm, optic nerves, hippocampus). The primary aim of this study was to assess factors influencing BM local control after HFSRT. Then the effect of surgery plus HFSRT was compared with exclusive HFSRT on oncologic outcomes, including overall survival. Materials and methods Retrospective study conducted in Leon Berard Cancer Center, included patients over 18 years-old with BM, secondary to a tumor proven by histology and treated by HFSRT alone or after surgery. Three different dose-fractionation schedules were compared: 27 Gy (3 x 9 Gy), 30 Gy (5 x 6 Gy) and 35 Gy (5 x 7 Gy), prescribed on isodose 80%. Primary endpoint were local control (LC). Secondary endpoints were overall survival (OS) and radionecrosis (RN) rate. Results A total of 389 patients and 400 BM with regular MRI follow-up were analyzed. There was no statistical difference between the different dose-fractionations. On multivariate analysis, surgery (p = 0.049) and size (< 2.5 cm) (p = 0.01) were independent factors improving LC. The 12 months LC was 87.02% in the group Surgery plus HFSRT group vs 73.53% at 12 months in the group HFSRT. OS was 61.43% at 12 months in the group Surgery plus HFSRT group vs 50.13% at 12 months in the group HFSRT (p < 0.0085). Prior surgery (OR = 1.86; p = 0.0028) and sex (OR = 1.4; p = 0.0139) control of primary tumor (OR = 0.671, p = 0.0069) and KPS < 70 (OR = 0.769, p = 0.0094) were independently predictive of OS. The RN rate was 5% and all patients concerned were symptomatic. Conclusions This study suggests that HFSRT is an efficient and well-tolerated treatment. The optimal dose-fractionation remains difficult to determine. Smaller size and surgery are correlated to LC. These results evidence the importance of surgery for larger BM (> 2.5 cm) with a poorer prognosis. Multidisciplinary committees and prospective studies are necessary to validate these observations.
C1 [Mengue, Laurence; Ayadi, Myriam; Claude, Line; Carrie, Christian; Laude, Cecile; Tanguy, Ronan; Sunyach, Marie-Pierre] Leon Berard Canc Ctr, Dept Radiotherapy, Lyon, France.
   [Bertaut, Aurelie; Blanc, Julie] Ctr Georges Francois Leclerc, Methodol & Biostat Unit, Dijon, France.
   [Mbus, Louise Ngo] Hop Aurillac, Dept Med, Aurillac, France.
   [Dore, Melanie; Clement-Colmou, Karen] Inst Cancerol Ouest, Dept Radiat Oncol, Nantes, France.
C3 UNICANCER; Centre Leon Berard; UNICANCER; Centre Georges-Francois
   Leclerc; UNICANCER; Institut de Cancerologie de l'Ouest (ICO)
RP Mengue, L (通讯作者)，Leon Berard Canc Ctr, Dept Radiotherapy, Lyon, France.
EM
RI tanguy, ronan/AFP-1804-2022; Tanguy, Ronan/AES-9511-2022
OI tanguy, ronan/0000-0002-6442-924X; Tanguy, Ronan/0000-0002-6442-924X;
   Clement-Colmou, Karen/0000-0003-0986-2718
CR Ahmed KA, 2016, AM J CLIN ONCOL-CANC, V39, P379, DOI 10.1097/COC.0000000000000076
   Andrews DW, 2004, LANCET, V363, P1665, DOI 10.1016/S0140-6736(04)16250-8
   Arvold ND, 2016, NEURO-ONCOLOGY, V18, P1043, DOI 10.1093/neuonc/now127
   Atalar B, 2013, INT J RADIAT ONCOL, V87, P713, DOI 10.1016/j.ijrobp.2013.07.034
   Baschnagel AM, 2013, J NEUROSURG, V119, P1139, DOI 10.3171/2013.7.JNS13431
   Chamberlain MC, 2017, NEURO-ONCOLOGY, V19, pI1, DOI [10.1093/neuonc/now197, 10.1093/neuonc/now279]
   Churilla TM, 2017, ANN ONCOL, V28, P2588, DOI 10.1093/annonc/mdx332
   Ernst-Stecken A, 2006, RADIOTHER ONCOL, V81, P18, DOI 10.1016/j.radonc.2006.08.024
   Fahrig A, 2007, STRAHLENTHER ONKOL, V183, P625, DOI 10.1007/s00066-007-1714-1
   Fokas E, 2012, J NEURO-ONCOL, V109, P91, DOI 10.1007/s11060-012-0868-6
   Fox BD, 2011, NEUROSURG CLIN N AM, V22, P1, DOI 10.1016/j.nec.2010.08.007
   Gaspar L, 1997, INT J RADIAT ONCOL, V37, P745, DOI 10.1016/S0360-3016(96)00619-0
   Giubilei C, 2009, J NEURO-ONCOL, V91, P207, DOI 10.1007/s11060-008-9700-8
   Inoue HK, 2014, RADIAT ONCOL, V9, DOI 10.1186/s13014-014-0231-5
   Kirkpatrick JP, 2017, NEURO-ONCOLOGY, V19, P38, DOI 10.1093/neuonc/now301
   Kocher M, 2011, J CLIN ONCOL, V29, P134, DOI 10.1200/JCO.2010.30.1655
   Kumar AMS, 2018, J NEURO-ONCOL, V139, P449, DOI 10.1007/s11060-018-2885-6
   Lamba N, 2019, WORLD NEUROSURG, V122, pE1557, DOI 10.1016/j.wneu.2018.11.100
   Le Rhun E, 2016, EXPERT REV NEUROTHER, V16, P903, DOI 10.1080/14737175.2016.1184572
   Le Rhun E, 2015, CANCER RADIOTHER, V19, P66, DOI 10.1016/j.canrad.2014.11.006
   Lehrer EJ, 2019, INT J RADIAT ONCOL, V103, P618, DOI 10.1016/j.ijrobp.2018.10.038
   Lesueur P, 2018, RADIAT ONCOL, V13, DOI 10.1186/s13014-018-1083-1
   Mahajan A, 2017, LANCET ONCOL, V18, P1040, DOI 10.1016/S1470-2045(17)30414-X
   Marchetti M, 2011, NEUROL SCI, V32, P393, DOI 10.1007/s10072-010-0473-4
   Masucci GL, 2018, FRONT ONCOL, V8, DOI 10.3389/fonc.2018.00379
   Minniti G, 2016, INT J RADIAT ONCOL, V95, P1142, DOI 10.1016/j.ijrobp.2016.03.013
   Minniti G, 2014, RADIAT ONCOL, V9, DOI 10.1186/1748-717X-9-110
   Minniti G, 2011, RADIAT ONCOL, V6, DOI 10.1186/1748-717X-6-48
   Nieder C, 2007, INT J RADIAT ONCOL, V69, P640, DOI 10.1016/j.ijrobp.2007.06.047
   Nieder C, 2009, ACTA ONCOL, V48, P457, DOI 10.1080/02841860802342390
   Noh Thomas, 2018, Handb Clin Neurol, V149, P75, DOI 10.1016/B978-0-12-811161-1.00006-2
   Pacelli R., 2007, EUR J NUCL MED MOL I, V34, P965
   Scorsetti M, 2009, ANTICANCER RES, V29, P4259
   Soffietti R, 2017, NEURO-ONCOLOGY, V19, P162, DOI 10.1093/neuonc/now241
   Sperduto PW, 2013, INT J RADIAT ONCOL, V85, P1312, DOI 10.1016/j.ijrobp.2012.11.042
   Sperduto PW, 2012, J CLIN ONCOL, V30, P419, DOI 10.1200/JCO.2011.38.0527
   Timmerman R, 2011, FRONT RADIAT THER ON, V43, P382, DOI 10.1159/000322494
   Yamamoto M, 2014, WORLD NEUROSURG, V82, DOI 10.1016/j.wneu.2014.08.008
   Yamamoto M, 2012, INT J RADIAT ONCOL, V83, P1399, DOI 10.1016/j.ijrobp.2011.10.018
   Zindler JD, 2017, IMPROVING PATIENT SE, DOI [10.26481/dis.20171025jz, DOI 10.26481/DIS.20171025JZ]
NR 40
TC 20
Z9 20
U1 0
U2 8
PU BMC
PI LONDON
PA CAMPUS, 4 CRINAN ST, LONDON N1 9XW, ENGLAND
EI 1748-717X
J9 RADIAT ONCOL
JI Radiat. Oncol.
PD APR 17
PY 2020
VL 15
IS 1
AR 82
DI 10.1186/s13014-020-01517-3
PG 11
WC Oncology; Radiology, Nuclear Medicine & Medical Imaging
WE Science Citation Index Expanded (SCI-EXPANDED)
SC Oncology; Radiology, Nuclear Medicine & Medical Imaging
GA LH4NE
UT WOS:000528761200001
PM 32303236
OA Green Published, gold
DA 2023-08-27
ER

PT J
AU Bentley, RT
   Yanke, AB
   Miller, MA
   Heng, HG
   Cohen-Gadol, A
   Rossmeisl, JH
AF Bentley, R. Timothy
   Yanke, Amy B.
   Miller, Margaret A.
   Heng, Hock Gan
   Cohen-Gadol, Aaron
   Rossmeisl, John H.
TI Cerebrospinal Fluid Drop Metastases of Canine Glioma: Magnetic Resonance
   Imaging Classification
SO FRONTIERS IN VETERINARY SCIENCE
LA English
DT Article
DE astrocytoma; dissemination; dog; ependyma; glioblastoma; leptomeninges;
   meninges; oligodendroglioma
ID DIAGNOSIS; BRAIN; DISSEMINATION; ASTROCYTOMA; FEATURES; CRITERIA
AB Dissemination of glioma in humans can occur as leptomeningeal nodules, diffuse leptomeningeal lesions, or ependymal lesions. Cerebrospinal fluid (CSF) drop metastasis of glioma is not well-recognized in dogs. Ten dogs with at least two anatomically distinct and histologically confirmed foci of glioma were included in this study. The 10 dogs underwent 28 magnetic resonance imaging (MRI) examinations, with distant CSF drop metastasis revealed in 13 MRIs. The CSF drop metastases appeared as leptomeningeal nodules in four dogs, diffuse leptomeningeal lesions in six dogs, and ependymal lesions in seven dogs; six dogs had a combination of lesion types. Primary tumors were generally T2-heterogeneous and contrast-enhancing. Many metastases were T2-homogeneous and non-enhancing. Diffuse leptomeningeal lesions were seen as widespread extra-axial contrast-enhancement, again very dissimilar to the intra-axial primary mass. Primary masses were rostrotentorial, whereas metastases generally occurred in the direction of CSF flow, in ventricles, CSF cisterns, and the central canal or leptomeninges of the cervical or thoracolumbar spinal cord. Seven of the dogs had received therapy limited to the primary mass, such as surgery or stereotactic radiation, then developed metastasis in the following months. CSF drop metastasis of glioma may take a very different appearance on MRI to the primary mass, including periventricular lesions that are more homogeneous and less contrast-enhancing, rostral horn signal changes, or leptomeningeal enhancement ventral to the brainstem or encircling the spinal cord.
C1 [Bentley, R. Timothy; Heng, Hock Gan; Cohen-Gadol, Aaron] Purdue Univ, Dept Vet Clin Sci, Coll Vet Med, W Lafayette, IN 47907 USA.
   [Yanke, Amy B.] Auburn Univ, Coll Vet Med, Dept Clin Sci, Auburn, AL 36849 USA.
   [Miller, Margaret A.] Purdue Univ, Coll Vet Med, Dept Comparat Pathobiol, W Lafayette, IN 47907 USA.
   [Cohen-Gadol, Aaron] Indiana Univ Sch Med, Dept Neurol Surg, Indianapolis, IN 46202 USA.
   [Rossmeisl, John H.] Virginia Maryland Coll Vet Med, Dept Small Anim Clin Sci, Blacksburg, VA USA.
C3 Purdue University System; Purdue University; Purdue University West
   Lafayette Campus; Auburn University System; Auburn University; Purdue
   University System; Purdue University; Purdue University West Lafayette
   Campus; Indiana University System; Indiana University Bloomington
RP Bentley, RT (通讯作者)，Purdue Univ, Dept Vet Clin Sci, Coll Vet Med, W Lafayette, IN 47907 USA.
EM
RI Rossmeisl, John H/Q-7119-2016
OI Rossmeisl, John H/0000-0003-1655-7076
FU Agricultural Animal and Small Animal Disease Research Funds, Purdue
   University College of Veterinary Medicine; Mary Ann and Gene Zink Fund,
   Indiana University School of Medicine; NIH/NCI [R01CA139099,
   P01CA207206]
FX Management of cases in this study was supported by the Agricultural
   Animal and Small Animal Disease Research Funds, Purdue University
   College of Veterinary Medicine (to RB); Mary Ann and Gene Zink Fund,
   Indiana University School of Medicine (to RB and AC-G); and NIH/NCI
   R01CA139099 and P01CA207206 (to JR).
CR Bentley RT, 2013, VET J, V198, P463, DOI 10.1016/j.tvjl.2013.08.015
   Bentley RT, 2018, WORLD NEUROSURG, V116, pE534, DOI 10.1016/j.wneu.2018.05.027
   Bentley RT, 2015, VET J, V205, P204, DOI 10.1016/j.tvjl.2015.01.025
   BERNSTEIN JJ, 1989, J NEUROSCI RES, V22, P134, DOI 10.1002/jnr.490220205
   Bordignon KC, 2006, SURG NEUROL, V65, P472, DOI 10.1016/j.surneu.2005.08.019
   Dreha-Kulaczewski S, 2017, J NEUROSCI, V37, P2395, DOI 10.1523/JNEUROSCI.2754-16.2017
   Figueiredo EG, 2003, ARQ NEURO-PSIQUIAT, V61, P842, DOI 10.1590/S0004-282X2003000500025
   Koch MW, 2011, J AM ANIM HOSP ASSOC, V47, pE77, DOI 10.5326/JAAHA-MS-5551
   Koehler JW, 2018, J NEUROPATH EXP NEUR, V77, P2018, DOI 10.1093/jnen/nly085
   Nakamoto Y, 2018, J VET MED SCI, V80, P1590, DOI 10.1292/jvms.17-0652
   Packer RA, 2018, VET RADIOL ULTRASOUN, V59, P261, DOI 10.1111/vru.12608
   Pande A, 2021, WORLD NEUROSURG, V146, P261, DOI 10.1016/j.wneu.2020.10.023
   Rissi DR, 2015, J VET DIAGN INVEST, V27, P396, DOI 10.1177/1040638715584619
   Sharma P, 2020, NEURO-ONCOL ADV, V2, DOI 10.1093/noajnl/vdaa107
   Vigeral M, 2018, VET RADIOL ULTRASOUN, V59, pE32, DOI 10.1111/vru.12474
   Wen PY, 2010, J CLIN ONCOL, V28, P1963, DOI 10.1200/JCO.2009.26.3541
   Young BD, 2011, VET RADIOL ULTRASOUN, V52, P132, DOI 10.1111/j.1740-8261.2010.01758.x
NR 17
TC 5
Z9 5
U1 0
U2 3
PU FRONTIERS MEDIA SA
PI LAUSANNE
PA AVENUE DU TRIBUNAL FEDERAL 34, LAUSANNE, CH-1015, SWITZERLAND
EI 2297-1769
J9 FRONT VET SCI
JI Front. Vet. Sci.
PD MAY 3
PY 2021
VL 8
AR 650320
DI 10.3389/fvets.2021.650320
PG 15
WC Veterinary Sciences
WE Science Citation Index Expanded (SCI-EXPANDED)
SC Veterinary Sciences
GA SC9MP
UT WOS:000650988100001
PM 34012987
OA gold, Green Published
DA 2023-08-27
ER

PT J
AU Ko, HS
   von Coelln, R
   Sriram, SR
   Kim, SW
   Chung, KKK
   Pletnikova, O
   Troncoso, J
   Johnson, B
   Saffary, R
   Goh, EL
   Song, HJ
   Park, BJ
   Kim, MJ
   Kim, S
   Dawson, VL
   Dawson, TM
AF Ko, HS
   von Coelln, R
   Sriram, SR
   Kim, SW
   Chung, KKK
   Pletnikova, O
   Troncoso, J
   Johnson, B
   Saffary, R
   Goh, EL
   Song, HJ
   Park, BJ
   Kim, MJ
   Kim, S
   Dawson, VL
   Dawson, TM
TI Accumulation of the authentic parkin substrate aminoacyl-tRNA synthetase
   cofactor, p38/JTV-1, leads to catecholaminergic cell death
SO JOURNAL OF NEUROSCIENCE
LA English
DT Article
DE Parkinson's disease; parkin; ubiquitination; proteasome degradation;
   p38/JTV1; dopaminergic neuronal cell death
ID DISEASE GENE-PRODUCT; ALPHA-SYNUCLEIN; S-NITROSYLATION; MUTATIONS;
   UBIQUITIN; PROTEIN; LIGASE; P38; ASSOCIATION; COMPLEX
AB Autosomal-recessive juvenile parkinsonism (AR-JP) is caused by loss-of-function mutations of the parkin gene. Parkin, a RING-type E3 ubiquitin ligase, is responsible for the ubiquitination and degradation of substrate proteins that are important in the survival of dopamine neurons in Parkinson's disease (PD). Accordingly, the abnormal accumulation of neurotoxic parkin substrates attributable to loss of parkin function may be the cause of neurodegeneration in parkin-related parkinsonism. We evaluated the known parkin substrates identified to date in parkin null mice to determine whether the absence of parkin results in accumulation of these substrates. Here we show that only the aminoacyl-tRNA synthetase cofactor p38 is upregulated in the ventral midbrain/hindbrain of both young and old parkin null mice. Consistent with upregulation in parkin knock-out mice, brains of AR-JP and idiopathic PD and diffuse Lewy body disease also exhibit increased level of p38. In addition, p38 interacts with parkin and parkin ubiquitinates and targets p38 for degradation. Furthermore, overexpression of p38 induces cell death that increases with tumor necrosis factor-alpha treatment and parkin blocks the pro-cell death effect of p38, whereas the R42P, familial-linked mutant of parkin, fails to rescue cell death. We further show that adenovirus-mediated overexpression of p38 in the substantia nigra in mice leads to loss of dopaminergic neurons. Together, our study represents a major advance in our understanding of parkin function, because it clearly identifies p38 as an important authentic pathophysiologic substrate of parkin. Moreover, these results have important implications for understanding the molecular mechanisms of neurodegeneration in PD.
C1 Johns Hopkins Univ, Sch Med, Inst Cell Engn, Baltimore, MD 21205 USA.
   Johns Hopkins Univ, Sch Med, Dept Neurol, Baltimore, MD 21205 USA.
   Johns Hopkins Univ, Sch Med, Dept Neurosci, Baltimore, MD 21205 USA.
   Johns Hopkins Univ, Sch Med, Dept Physiol, Baltimore, MD 21205 USA.
   Johns Hopkins Univ, Sch Med, Dept Pathol, Baltimore, MD 21205 USA.
   Johns Hopkins Univ, Sch Med, Grad Program Cellular & Mol Med, Baltimore, MD 21205 USA.
   Seoul Natl Univ, Coll Pharm, Natl Creat Res Initiat Ctr Human Aminoacyl, tRNA Synthetases Network, Seoul 151742, South Korea.
C3 Johns Hopkins University; Johns Hopkins University; Johns Hopkins
   University; Johns Hopkins University; Johns Hopkins University; Johns
   Hopkins University; Seoul National University (SNU)
RP Dawson, TM (通讯作者)，Johns Hopkins Univ, Sch Med, Inst Cell Engn, 733 N Broadway St,Suite 731, Baltimore, MD 21205 USA.
EM
RI Goh, Eyleen EL/A-8006-2013; Dawson, Valina/Y-9757-2019; Kim,
   Sunghoon/AAE-8314-2020
OI von Coelln, Rainer/0000-0001-7651-2780; Dawson,
   Valina/0000-0002-2915-3970; Goh, Eyleen/0000-0002-8244-6959
FU NINDS NIH HHS [R01 NS048206, NS 48206, P50 NS038377, NS 38377] Funding
   Source: Medline
CR Ahn HC, 2003, FEBS LETT, V542, P119, DOI 10.1016/S0014-5793(03)00362-4
   Bonifati V, 2003, SCIENCE, V299, P256, DOI 10.1126/science.1077209
   Choi P, 2003, MOL BRAIN RES, V117, P179, DOI 10.1016/S0169-328X(03)00318-8
   Chung KKK, 2004, SCIENCE, V304, P1328, DOI 10.1126/science.1093891
   Chung KKK, 2001, NAT MED, V7, P1144, DOI 10.1038/nm1001-1144
   Cookson MR, 2005, ANNU REV BIOCHEM, V74, P29, DOI 10.1146/annurev.biochem.74.082803.133400
   Cookson MR, 2003, NEUROMOL MED, V3, P1, DOI 10.1385/NMM:3:1:1
   Corti O, 2003, HUM MOL GENET, V12, P1427, DOI 10.1093/hmg/ddg159
   Cuervo AM, 2004, TRENDS CELL BIOL, V14, P70, DOI 10.1016/j.tcb.2003.12.002
   Dawson TM, 2003, SCIENCE, V302, P819, DOI 10.1126/science.1087753
   Dong ZZ, 2003, P NATL ACAD SCI USA, V100, P12438, DOI 10.1073/pnas.2132992100
   Doss-Pepe EW, 2005, J BIOL CHEM, V280, P16619, DOI 10.1074/jbc.M413591200
   Feany MB, 2003, NEURON, V38, P13, DOI 10.1016/S0896-6273(03)00201-0
   Goldberg MS, 2003, J BIOL CHEM, V278, P43628, DOI 10.1074/jbc.M308947200
   Huynh DP, 2003, HUM MOL GENET, V12, P2587, DOI 10.1093/hmg/ddg269
   Imai Y, 2001, CELL, V105, P891, DOI 10.1016/S0092-8674(01)00407-X
   Imai Y, 2002, MOL CELL, V10, P55, DOI 10.1016/S1097-2765(02)00583-X
   Itier JM, 2003, HUM MOL GENET, V12, P2277, DOI 10.1093/hmg/ddg239
   Jiang HB, 2004, J BIOL CHEM, V279, P54380, DOI 10.1074/jbc.M409282200
   Kahle PJ, 2004, EMBO REP, V5, P681, DOI 10.1038/sj.embor.7400188
   Kim JY, 2002, P NATL ACAD SCI USA, V99, P7912, DOI 10.1073/pnas.122110199
   Kim MJ, 2003, NAT GENET, V34, P330, DOI 10.1038/ng1182
   Kitada T, 1998, NATURE, V392, P605, DOI 10.1038/33416
   Lim KL, 2005, J NEUROSCI, V25, P2002, DOI 10.1523/JNEUROSCI.4474-04.2005
   Lucking CB, 2000, NEW ENGL J MED, V342, P1560, DOI 10.1056/NEJM200005253422103
   Moore DJ, 2005, HUM MOL GENET, V14, P71, DOI 10.1093/hmg/ddi007
   Paisan-Ruiz C, 2004, NEURON, V44, P595, DOI 10.1016/j.neuron.2004.10.023
   Palacino JJ, 2004, J BIOL CHEM, V279, P18614, DOI 10.1074/jbc.M401135200
   PAXINOS G, 1985, J NEUROSCI METH, V13, P139, DOI 10.1016/0165-0270(85)90026-3
   Polymeropoulos MH, 1997, SCIENCE, V276, P2045, DOI 10.1126/science.276.5321.2045
   Quevillon S, 1999, J MOL BIOL, V285, P183, DOI 10.1006/jmbi.1998.2316
   Ren Y, 2003, J NEUROSCI, V23, P3316
   Sakata E, 2003, EMBO REP, V4, P301, DOI 10.1038/sj.embor.embor764
   Shimura H, 2000, NAT GENET, V25, P302, DOI 10.1038/77060
   Shimura H, 2001, SCIENCE, V293, P263, DOI 10.1126/science.1060627
   Staropoli JF, 2003, NEURON, V37, P735, DOI 10.1016/S0896-6273(03)00084-9
   Valente EM, 2004, SCIENCE, V304, P1158, DOI 10.1126/science.1096284
   von Coelln R, 2004, P NATL ACAD SCI USA, V101, P10744, DOI 10.1073/pnas.0401297101
   von Coelln R, 2004, CELL TISSUE RES, V318, P175, DOI 10.1007/s00441-004-0924-4
   Yao DD, 2004, P NATL ACAD SCI USA, V101, P10810, DOI 10.1073/pnas.0404161101
   Yuan H, 2005, J NEUROSCI METH, V144, P35, DOI 10.1016/j.jneumeth.2004.10.004
   Zhang Y, 2000, P NATL ACAD SCI USA, V97, P13354, DOI 10.1073/pnas.240347797
   Zhao XY, 2003, P NATL ACAD SCI USA, V100, P6777, DOI 10.1073/pnas.1131928100
   Zimprich A, 2004, NEURON, V44, P601, DOI 10.1016/j.neuron.2004.11.005
NR 44
TC 193
Z9 202
U1 1
U2 11
PU SOC NEUROSCIENCE
PI WASHINGTON
PA 11 DUPONT CIRCLE, NW, STE 500, WASHINGTON, DC 20036 USA
SN 0270-6474
J9 J NEUROSCI
JI J. Neurosci.
PD AUG 31
PY 2005
VL 25
IS 35
BP 7968
EP 7978
DI 10.1523/JNEUROSCI.2172-05.2005
PG 11
WC Neurosciences
WE Science Citation Index Expanded (SCI-EXPANDED)
SC Neurosciences & Neurology
GA 960BL
UT WOS:000231565100009
PM 16135753
OA Green Published, hybrid
DA 2023-08-27
ER

PT J
AU Dubey, D
   Wilson, MR
   Clarkson, B
   Giannini, C
   Gandhi, M
   Cheville, J
   Lennon, VA
   Eggers, S
   Devine, MF
   Mandel-Brehm, C
   Kryzer, T
   Hinson, SR
   Khazaie, K
   Hales, C
   Kattah, J
   Pavelko, KD
   Andrews, P
   Eaton, JE
   Jitprapaikulsan, J
   Mills, JR
   Flanagan, EP
   Zekeridou, A
   Leibovich, B
   Fryer, J
   Torre, M
   Kaufman, C
   Thoreson, JB
   Sagen, J
   Linnoila, JJ
   DeRisi, JL
   Howe, CL
   McKeon, A
   Pittock, SJ
AF Dubey, Divyanshu
   Wilson, Michael R.
   Clarkson, Benjamin
   Giannini, Caterina
   Gandhi, Manish
   Cheville, John
   Lennon, Vanda A.
   Eggers, Scott
   Devine, Michelle F.
   Mandel-Brehm, Caleigh
   Kryzer, Thomas
   Hinson, Shannon R.
   Khazaie, Khashayarsha
   Hales, Chadwick
   Kattah, Jorge
   Pavelko, Kevin D.
   Andrews, Patrick
   Eaton, James E.
   Jitprapaikulsan, Jiraporn
   Mills, John R.
   Flanagan, Eoin P.
   Zekeridou, Anastasia
   Leibovich, Bradley
   Fryer, James
   Torre, Matthew
   Kaufman, Charles
   Thoreson, James B.
   Sagen, Jessica
   Linnoila, Jenny J.
   DeRisi, Joseph L.
   Howe, Charles L.
   McKeon, Andrew
   Pittock, Sean J.
TI Expanded Clinical Phenotype, Oncological Associations, and
   Immunopathologic Insights of Paraneoplastic Kelch-like Protein-11
   Encephalitis
SO JAMA NEUROLOGY
LA English
DT Article
ID OUT TESTICULAR-TUMOR; ANTIGEN PRESENTATION; AUTOIMMUNE; ANTIBODIES;
   CANCER
AB Question What are the clinical, radiologic, immunopathologic, and human leukocyte antigen features of Kelch-like-protein-11 (KLHL11) encephalitis? Findings In this case series, the common presenting feature of KLHL11 encephalitis is a rhombencephalitis phenotype with ataxia, diplopia, dysarthria, vertigo, hearing loss, and tinnitus. A strong oncologic association, KLHL11-specific T-cell response, human leukocyte antigen associations, and brain histopathology supports an immunopathologic basis. Meaning Recognizing presenting features of KLHL11 encephalitis may aid in early diagnosis of this paraneoplastic syndrome and immunopathologic findings may guide immunotherapeutic management.
   This study describes expanding neurologic phenotype, cancer associations, outcomes, and immunopathologic features of Kelch-like protein-11 encephalitis.
   Importance Recognizing the presenting and immunopathological features of Kelch-like protein-11 immunoglobulin G seropositive (KLHL11 IgG+) patients may aid in early diagnosis and management. Objective To describe expanding neurologic phenotype, cancer associations, outcomes, and immunopathologic features of KLHL11 encephalitis. Design, Setting, and Participants This retrospective tertiary care center study, conducted from October 15, 1998, to November 1, 2019, prospectively identified 31 KLHL11 IgG+ cases in the neuroimmunology laboratory. Eight were identified by retrospective testing of patients with rhomboencephalitis (confirmed by tissue-based-immunofluorescence and transfected-cell-based assays). Main Outcomes and Measures Outcome variables included modified Rankin score and gait aid use. Results All 39 KLHL11 IgG+ patients were men (median age, 46 years; range, 28-73 years). Initial clinical presentations were ataxia (n = 32; 82%), diplopia (n = 22; 56%), vertigo (n = 21; 54%), hearing loss (n = 15; 39%), tinnitus (n = 14; 36%), dysarthria (n = 11; 28%), and seizures (n = 9; 23%). Atypical neurologic presentations included neuropsychiatric dysfunction, myeloneuropathy, and cervical amyotrophy. Hearing loss or tinnitus preceded other neurologic deficits by 1 to 8 months in 10 patients (26%). Among patients screened for malignancy (n = 36), testicular germ-cell tumors (n = 23; 64%) or testicular microlithiasis and fibrosis concerning for regressed germ cell tumor (n = 7; 19%) were found in 83% of the patients (n = 30). In 2 patients, lymph node biopsy diagnosed metastatic lung adenocarcinoma in one and chronic lymphocytic leukemia in the other. Initial brain magnetic resonance imaging revealed T2 hyperintensities in the temporal lobe (n = 12), cerebellum (n = 9), brainstem (n = 3), or diencephalon (n = 3). Among KLHL11 IgG+ patients who underwent HLA class I and class II genotyping (n = 10), most were found to have HLA-DQB1*02:01 (n = 7; 70%) and HLA-DRB1*03:01 (n = 6; 60%) associations. A biopsied gadolinium-enhancing temporal lobe lesion demonstrated T cell-predominant inflammation and nonnecrotizing granulomas. Cerebellar biopsy (patient with chronic ataxia) and 2 autopsied brains demonstrated Purkinje neuronal loss and Bergmann gliosis, supporting early active inflammation and later extensive neuronal loss. Compared with nonautoimmune control peripheral blood mononuclear cells, cluster of differentiation (CD) 8+ and CD4+ T cells were significantly activated when patient peripheral blood mononuclear cells were cultured with KLHL11 protein. Most patients (58%) benefitted from immunotherapy and/or cancer treatment (neurological disability stabilized [n = 10] or improved [n = 9]). Kaplan-Meier curve demonstrated significantly higher probability of wheelchair dependence among patients without detectable testicular cancer. Long-term outcomes in KLHL11-IgG+ patients were similar to Ma2 encephalitis. Conclusions and Relevance Kelch-like protein-11 IgG is a biomarker of testicular germ-cell tumor and paraneoplastic neurologic syndrome, often refractory to treatment. Described expanded neurologic phenotype and paraclinical findings may aid in its early diagnosis and treatment.
C1 [Dubey, Divyanshu; Giannini, Caterina; Gandhi, Manish; Cheville, John; Lennon, Vanda A.; Devine, Michelle F.; Mills, John R.; Flanagan, Eoin P.; Zekeridou, Anastasia; Thoreson, James B.; Sagen, Jessica; McKeon, Andrew; Pittock, Sean J.] Mayo Clin, Dept Lab Med & Pathol, Rochester, MN USA.
   [Dubey, Divyanshu; Clarkson, Benjamin; Lennon, Vanda A.; Eggers, Scott; Kryzer, Thomas; Hinson, Shannon R.; Andrews, Patrick; Flanagan, Eoin P.; Zekeridou, Anastasia; Fryer, James; Sagen, Jessica; Howe, Charles L.; McKeon, Andrew; Pittock, Sean J.] Mayo Clin, Dept Neurol, Rochester, MN USA.
   [Dubey, Divyanshu; Lennon, Vanda A.; Khazaie, Khashayarsha; Pavelko, Kevin D.; Howe, Charles L.] Mayo Clin, Dept Immunol, Rochester, MN USA.
   [Dubey, Divyanshu; Clarkson, Benjamin; Giannini, Caterina; Lennon, Vanda A.; Devine, Michelle F.; Kryzer, Thomas; Hinson, Shannon R.; Andrews, Patrick; Jitprapaikulsan, Jiraporn; Mills, John R.; Flanagan, Eoin P.; Zekeridou, Anastasia; Fryer, James; Sagen, Jessica; Howe, Charles L.; McKeon, Andrew; Pittock, Sean J.] Mayo Clin, Ctr MS & Autoimmune Neurol, Rochester, MN USA.
   [Wilson, Michael R.] Univ Calif San Francisco, Dept Neurol, Weill Inst Neurosci, San Francisco, CA USA.
   [Mandel-Brehm, Caleigh; DeRisi, Joseph L.] Univ Calif San Francisco, Dept Biochem & Biophys, San Francisco, CA USA.
   [Hales, Chadwick] Emory Univ, Sch Med, Dept Neurol, Atlanta, GA 30322 USA.
   [Kattah, Jorge] Univ Illinois, Coll Med, Dept Neurol, Peoria, IL 61656 USA.
   [Eaton, James E.] Vanderbilt Univ, Med Ctr, Dept Neurol, Nashville, TN USA.
   [Jitprapaikulsan, Jiraporn] Mahidol Univ, Dept Neurol, Bangkok, Thailand.
   [Leibovich, Bradley] Mayo Clin, Dept Urol, Rochester, MN USA.
   [Torre, Matthew] Brigham & Womens Hosp, Dept Pathol, 75 Francis St, Boston, MA 02115 USA.
   [Kaufman, Charles] Louisiana Neurol Consultants, Baton Rouge, LA USA.
   [Linnoila, Jenny J.] Massachusetts Gen Hosp, Dept Neurol, Boston, MA 02114 USA.
   [DeRisi, Joseph L.] Chan Zuckerberg Biohub, San Francisco, CA USA.
C3 Mayo Clinic; Mayo Clinic; Mayo Clinic; Mayo Clinic; University of
   California System; University of California San Francisco; University of
   California System; University of California San Francisco; Emory
   University; University of Illinois System; University of Illinois
   Peoria; Vanderbilt University; Mahidol University; Mayo Clinic; Harvard
   University; Brigham & Women's Hospital; Harvard University;
   Massachusetts General Hospital
RP Dubey, D (通讯作者)，Mayo Clin, Dept Lab Med & Pathol, Dept Neurol, 200 First St SW, Rochester, MN 55905 USA.
EM
RI Wilson, Michael R/IST-7754-2023; Howe, Charles L/B-4945-2009; Pavelko,
   Kevin D./K-7072-2019; Giannini, Caterina/AAB-7551-2022; McKeon,
   Andrew/AAY-9214-2021
OI Wilson, Michael R/0000-0002-8705-5084; Pavelko, Kevin
   D./0000-0001-7555-1315; Howe, Charles/0000-0002-3889-8739; Dubey,
   Divyanshu/0000-0001-6865-9045; Eaton, James/0000-0003-0010-921X;
   mandel-brehm, caleigh/0000-0001-7764-6529
FU NINDS NIH HHS [K08 NS096117] Funding Source: Medline
CR Adams C, 2011, ARCH NEUROL-CHICAGO, V68, P521, DOI 10.1001/archneurol.2011.56
   Balalaa N, 2011, CASE REP ONCOL, V4, P12, DOI 10.1159/000324041
   Balzer BL, 2006, AM J SURG PATHOL, V30, P858, DOI 10.1097/01.pas.0000209831.24230.56
   Basal E, 2018, NEUROLOGY, V91, pE1677, DOI 10.1212/WNL.0000000000006435
   Dalmau J, 2004, BRAIN, V127, P1831, DOI 10.1093/brain/awh203
   DALMAU J, 1992, MEDICINE, V71, P59, DOI 10.1097/00005792-199203000-00001
   Dalmau J, 2017, PHYSIOL REV, V97, P839, DOI 10.1152/physrev.00010.2016
   Darnell RB, 1996, P NATL ACAD SCI USA, V93, P4529, DOI 10.1073/pnas.93.10.4529
   Darnell RB, 2006, SEMIN ONCOL, V33, P270, DOI 10.1053/j.seminoncol.2006.03.008
   Dubey D, 2018, ANN NEUROL, V83, P166, DOI 10.1002/ana.25131
   English SW, 2018, NEUROLOGY, V90, pE1535, DOI 10.1212/WNL.0000000000005356
   Falcioni F, 1999, NAT BIOTECHNOL, V17, P562, DOI 10.1038/9865
   Flanagan EP, 2017, ANN NEUROL, V81, P298, DOI 10.1002/ana.24881
   Freifeld Y, 2018, FRONT NEUROL, V9, DOI 10.3389/fneur.2018.00020
   Hara M, 2018, NEUROLOGY, V90, pE1386, DOI 10.1212/WNL.0000000000005329
   Ishikawa H, 2016, ACTA NEUROL SCAND, V133, P398, DOI 10.1111/ane.12469
   Jones AL, 2015, JAMA NEUROL, V72, P1304, DOI 10.1001/jamaneurol.2015.2378
   Kaneko A, 2018, J NEUROL, V265, P586, DOI 10.1007/s00415-018-8749-3
   Lucchinetti CF, 1998, NEUROLOGY, V50, P652, DOI 10.1212/WNL.50.3.652
   Mandel-Brehm C, 2019, NEW ENGL J MED, V381, P47, DOI 10.1056/NEJMoa1816721
   Maudes E, 2020, NEUROL-NEUROIMMUNOL, V7, DOI 10.1212/NXI.0000000000000666
   Pittock SJ, 2004, ANN NEUROL, V56, P715, DOI 10.1002/ana.20269
   Quek AML, 2012, ARCH NEUROL-CHICAGO, V69, P582, DOI 10.1001/archneurol.2011.2985
   Rojas I, 2000, NEUROLOGY, V55, P713, DOI 10.1212/WNL.55.5.713
   Xia J, 2006, J AM CHEM SOC, V128, P1859, DOI 10.1021/ja056423o
NR 25
TC 63
Z9 65
U1 2
U2 13
PU AMER MEDICAL ASSOC
PI CHICAGO
PA 330 N WABASH AVE, STE 39300, CHICAGO, IL 60611-5885 USA
SN 2168-6149
EI 2168-6157
J9 JAMA NEUROL
JI JAMA Neurol.
PD NOV
PY 2020
VL 77
IS 11
BP 1420
EP 1429
DI 10.1001/jamaneurol.2020.2231
PG 10
WC Clinical Neurology
WE Science Citation Index Expanded (SCI-EXPANDED)
SC Neurosciences & Neurology
GA OX7SS
UT WOS:000593760500014
PM 32744608
OA Bronze, Green Published
DA 2023-08-27
ER

PT J
AU Hao, SY
   Tang, J
   Wu, Z
   Zhang, LW
   Zhang, JT
   Wang, ZC
AF Hao, Shuyu
   Tang, Jie
   Wu, Zhen
   Zhang, Liwei
   Zhang, Junting
   Wang, Zhongcheng
TI Natural Malignant Transformation of an Intracranial Epidermoid Cyst
SO JOURNAL OF THE FORMOSAN MEDICAL ASSOCIATION
LA English
DT Article
DE epidermoid cyst; intracranial neoplasms; malignant transformation
ID SQUAMOUS-CELL CARCINOMA; TUMORS
AB Malignant transformation of intracranial epidermoid cyst (EC) is very rare, and when it does occur, the clinical course is aggressive. We present an unusual case of natural malignant transformation of an intracranial EC, without a history of surgery. A 61-year-old woman was diagnosed with an EC in the right cerebellopontine angle and prepontine cistern 6 years before the operation described in this report. Her neurological symptoms deteriorated in the 2 months prior to the present admission. Magnetic resonance imaging revealed an irregular, nodular enhanced lesion in the cerebellopontine angle, prepontine cistern, and temporoparietal lobe. Subtotal resection of the lesion was performed. Pathological examination revealed malignant transformation of the EC. The patient died on postoperative day 36 from brainstem infarction, chemical meningitis, and hydrocephalus. Spontaneous rupture of the EC could have contributed to the malignant transformation. This could have been avoided by early removal of the EC.
C1 [Hao, Shuyu; Tang, Jie; Wu, Zhen; Zhang, Liwei; Zhang, Junting] Capital Med Univ, Dept Neurosurg, Beijing Tiantan Hosp, Beijing 100050, Peoples R China.
   [Hao, Shuyu; Wang, Zhongcheng] Capital Med Univ, Beijing Neurosurg Inst, Beijing Tiantan Hosp, Beijing 100050, Peoples R China.
C3 Capital Medical University; Beijing Neurosurgical Institute; Capital
   Medical University
RP Tang, J (通讯作者)，Capital Med Univ, Dept Neurosurg, Beijing Tiantan Hosp, Tiantan Xili 6, Beijing 100050, Peoples R China.
EM
CR Agarwal S, 2007, CLIN NEUROL NEUROSUR, V109, P888, DOI 10.1016/j.clineuro.2007.07.026
   Akar Z, 2003, NEUROL MED-CHIR, V43, P275, DOI 10.2176/nmc.43.275
   ALVORD EC, 1977, ANN NEUROL, V2, P367, DOI 10.1002/ana.410020504
   Asahi T, 2001, J CLIN NEUROSCI, V8, P572, DOI 10.1054/jocn.2000.0856
   DAVIDSON SI, 1960, J NEUROL NEUROSUR PS, V23, P176, DOI 10.1136/jnnp.23.2.176
   DUBOIS PJ, 1981, J COMPUT ASSIST TOMO, V5, P433, DOI 10.1097/00004728-198106000-00025
   FOX H, 1965, J NEUROL NEUROSUR PS, V28, P276, DOI 10.1136/jnnp.28.3.276
   GOLDMAN SA, 1987, J NEUROSURG, V66, P618, DOI 10.3171/jns.1987.66.4.0618
   Guan LM, 2004, CHINESE MED J-PEKING, V117, P1880
   Hamlat A, 2005, J NEURO-ONCOL, V74, P187, DOI 10.1007/s11060-004-5175-4
   Kim MS, 2008, J KOREAN NEUROSURG S, V44, P401, DOI 10.3340/jkns.2008.44.6.401
   Kodama H, 2007, J NEURO-ONCOL, V82, P171, DOI 10.1007/s11060-006-9255-5
   LANDERS JW, 1960, ARCH PATHOL, V70, P419
   LEWIS AJ, 1983, J NEUROSURG, V59, P538, DOI 10.3171/jns.1983.59.3.0538
   Link MJ, 2002, J NEUROSURG, V97, P1237, DOI 10.3171/jns.2002.97.5.1237
   Michael LM, 2005, BRIT J NEUROSURG, V19, P505, DOI 10.1080/02688690500495356
   Murase S, 1999, NEUROL MED-CHIR, V39, P49, DOI 10.2176/nmc.39.49
   NISHIO S, 1995, NEUROSURGERY, V37, P329, DOI 10.1227/00006123-199508000-00021
   Nishiura I, 1989, Neurol Med Chir (Tokyo), V29, P600, DOI 10.2176/nmc.29.600
   Pagni F, 2007, NEUROPATHOLOGY, V27, P566, DOI 10.1111/j.1440-1789.2007.00820.x
   Sawan B, 2000, ANN PATHOL, V20, P258
   Tamura K, 2006, J NEURO-ONCOL, V79, P67, DOI 10.1007/s11060-005-9117-6
   TOGNETTI F, 1991, British Journal of Neurosurgery, V5, P303, DOI 10.3109/02688699109005191
NR 23
TC 30
Z9 33
U1 0
U2 2
PU ELSEVIER TAIWAN
PI TAIPEI
PA RM N-412, 4F, CHIA HSIN BUILDING 11, NO 96, ZHONG SHAN N ROAD SEC 2,
   TAIPEI, 10449, TAIWAN
SN 0929-6646
EI 1876-0821
J9 J FORMOS MED ASSOC
JI J. Formos. Med. Assoc.
PD MAY
PY 2010
VL 109
IS 5
BP 390
EP 396
DI 10.1016/S0929-6646(10)60068-X
PG 7
WC Medicine, General & Internal
WE Science Citation Index Expanded (SCI-EXPANDED)
SC General & Internal Medicine
GA 606CD
UT WOS:000278397500010
PM 20497873
OA Bronze
DA 2023-08-27
ER

PT J
AU Zheng, LR
   Mei, QL
   Gao, YX
   Du, FL
   Xiao, L
   Wu, GF
AF Zheng, Lirong
   Mei, Qiaoli
   Gao, Yuxiang
   Du, Fenglei
   Xiao, Lin
   Wu, Gangfeng
TI Parotid Gland Stem Cell Preservation during Intensity-Modulated
   Radiotherapy for Nasopharyngeal Carcinoma: Dosimetric Analysis and
   Feasibility
SO JOURNAL OF ONCOLOGY
LA English
DT Article
ID RADIATION-THERAPY; SUBMANDIBULAR-GLANDS; NECK TUMORS; SALIVARY; HEAD;
   CANCER; MECHANISM; EDITION; INJURY; MODEL
AB Objective. Parotid gland (PG) is a radiosensitive organ, and xerostomia (XS) is a key factor affecting patients' life quality after conventional radiotherapy for head and neck tumors. In this study, dosimetry analysis was performed on PG stem cell preservation in intensity-modulated radiotherapy (IMRT) for nasopharyngeal carcinoma (NPC). Methods. All clinical data of 80 NPC patients diagnosed pathologically in the Radiotherapy Department of Taizhou Hospital of Zhejiang Province Affiliated with Wenzhou Medical University from August 2017 to September 2019 were retrospectively analyzed. Patients were assigned to a regular group and a restricted group according to different IMRT plans, in which a dose limitation for the parotid duct was added in the restricted group in addition to the conventional plan used in the regular group to minimize the parotid duct radiation dose. The differences in planning target volume (PTV) dose distribution, organ at risk (OAR) dose, and dose to the PG and its ducts were compared between the two groups. Results. Significantly higher mean irradiation doses of the brainstem, mandible, and oral cavity were determined in the restricted group compared with the regular group (P > 0.05), but there was no significant difference in the mean dose of other OARs irradiated (P > 0.05). As compared to the irradiation of bilateral PGs, no statistical differences were found in the mean irradiation dose and V-30 between regular and restricted groups (P > 0.05), but lower V-20 and higher V-45 were determined in the restricted group (P < 0.05). The mean irradiation dose, V-15, V-20, and V-26 of bilateral parotid ducts were lower in the restricted group as compared to the regular group (P < 0.05). Conclusion. IMRT for NPC can effectively reduce the mean irradiation dose and play a PG stem cell preservation role by giving specific dose limitation conditions to the parotid duct area without affecting PTV dose distribution and OAR irradiation dose, which has certain feasibility.
C1 [Zheng, Lirong; Gao, Yuxiang] Wenzhou Med Univ, Dept ENT, Taizhou Hosp Zhejiang Prov, Taizhou 317000, Zhejiang, Peoples R China.
   [Mei, Qiaoli] Wenzhou Med Univ, Dept Pharm, Taizhou Hosp Zhejiang Prov, Taizhou 317000, Zhejiang, Peoples R China.
   [Du, Fenglei] Univ Chinese Acad Sci, Dept Radiotherapy, Canc Hosp, Zhejiang Canc Hosp, Hangzhou 310000, Zhejiang, Peoples R China.
   [Xiao, Lin] Taizhou Enze Med Ctr Grp Enze Hosp, Dept Otolaryngol, Taizhou 317000, Zhejiang, Peoples R China.
   [Wu, Gangfeng] Tai Zhou Second Peoples Hosp, Dept ENT, Taizhou 317000, Zhejiang, Peoples R China.
C3 Wenzhou Medical University; Wenzhou Medical University; Chinese Academy
   of Sciences; University of Chinese Academy of Sciences, CAS; Zhejiang
   Cancer Hospital
RP Wu, GF (通讯作者)，Tai Zhou Second Peoples Hosp, Dept ENT, Taizhou 317000, Zhejiang, Peoples R China.
EM
CR [Anonymous], 2007, PATIENT TUMOR CHARAC
   Bray F, 2020, CA-CANCER J CLIN, V70, P313, DOI 10.3322/caac.21609
   Broggi S, 2010, RADIOTHER ONCOL, V94, P206, DOI 10.1016/j.radonc.2009.12.014
   Burlage FR, 2001, RADIOTHER ONCOL, V61, P271, DOI 10.1016/S0167-8140(01)00427-3
   Chang ET, 2021, CANCER EPIDEM BIOMAR, V30, P1035, DOI 10.1158/1055-9965.EPI-20-1702
   Chao KSC, 2002, SEMIN RADIAT ONCOL, V12, P20, DOI 10.1053/srao.2002.31359
   Chavaudra J, 2001, Cancer Radiother, V5, P472, DOI 10.1016/S1278-3218(01)00117-2
   Chen QY, 2011, JNCI-J NATL CANCER I, V103, P1761, DOI 10.1093/jnci/djr432
   Chen YP, 2019, LANCET, V394, P64, DOI 10.1016/S0140-6736(19)30956-0
   Cumpston E., 2021, SUBMANDIBULAR EXCISI
   de Paula F, 2017, ANAT REC, V300, P1180, DOI 10.1002/ar.23569
   Guan SZ, 2020, EUR J MED CHEM, V207, DOI 10.1016/j.ejmech.2020.112758
   Guo R, 2019, BRIT J RADIOL, V92, DOI 10.1259/bjr.20190244
   Haberer-Guillerm S, 2015, EUR ANN OTORHINOLARY, V132, P147, DOI 10.1016/j.anorl.2014.02.008
   Jensen Siri Beier, 2019, Journal of the National Cancer Institute Monographs, P95, DOI 10.1093/jncimonographs/lgz016
   Konings AWT, 2005, INT J RADIAT ONCOL, V62, P1187, DOI 10.1016/j.ijrobp.2004.12.051
   Lee AW, 2019, INT J RADIAT ONCOL, V105, P567, DOI 10.1016/j.ijrobp.2019.06.2540
   Lee N, 2009, J CLIN ONCOL, V27, P3684, DOI 10.1200/JCO.2008.19.9109
   Lou JJ, 2018, J APPL CLIN MED PHYS, V19, P251, DOI 10.1002/acm2.12327
   Lu J, 2014, J RADIAT RES, V55, P97, DOI 10.1093/jrr/rrt076
   Munter MW, 2004, INT J RADIAT ONCOL, V58, P175, DOI 10.1016/S0360-3016(03)01437-8
   Nagler RM, 2002, ORAL DIS, V8, P141, DOI 10.1034/j.1601-0825.2002.02838.x
   Osorio EMV, 2008, INT J RADIAT ONCOL, V70, P875, DOI 10.1016/j.ijrobp.2007.10.063
   Pan JJ, 2016, CANCER-AM CANCER SOC, V122, P546, DOI 10.1002/cncr.29795
   Patel VN, 2014, SEMIN CELL DEV BIOL, V25, P52, DOI 10.1016/j.semcdb.2013.12.001
   Pringle S, 2013, STEM CELLS, V31, P613, DOI 10.1002/stem.1327
   Radfar L, 2003, ORAL SURG ORAL MED O, V96, P267, DOI 10.1016/S1079-2104(03)00369-X
   Ren G, 2015, BIOMED RES INT, V2015, DOI 10.1155/2015/670327
   Ries L.A.G., 2007, CANC SURVIVAL ADULTS, P7
   Seethala RR, 2017, HEAD NECK PATHOL, V11, P55, DOI 10.1007/s12105-017-0795-0
   STEPHENS LC, 1986, AM J PATHOL, V124, P469
   Teo PML, 2006, RADIOTHER ONCOL, V79, P27, DOI 10.1016/j.radonc.2006.03.012
   Thomson DJ, 2016, ADV OTO-RHINO-LARYNG, V78, P141, DOI 10.1159/000442134
   van Luijk P, 2009, INT J RADIAT ONCOL, V74, P1002, DOI 10.1016/j.ijrobp.2009.03.039
   Wang W, 2010, INT J RADIAT ONCOL, V77, P617, DOI 10.1016/j.ijrobp.2009.08.036
   Wang ZH, 2009, LARYNGOSCOPE, V119, P1966, DOI 10.1002/lary.20601
   Zhang BL, 2015, ORAL ONCOL, V51, P1041, DOI 10.1016/j.oraloncology.2015.08.005
NR 37
TC 0
Z9 0
U1 0
U2 2
PU HINDAWI LTD
PI LONDON
PA ADAM HOUSE, 3RD FLR, 1 FITZROY SQ, LONDON, W1T 5HF, ENGLAND
SN 1687-8450
EI 1687-8469
J9 J ONCOL
JI J. Oncol.
PD JUL 12
PY 2022
VL 2022
AR 4922409
DI 10.1155/2022/4922409
PG 7
WC Oncology
WE Science Citation Index Expanded (SCI-EXPANDED)
SC Oncology
GA 5N7ZD
UT WOS:000872006300001
PM 35865088
OA Green Published, gold
DA 2023-08-27
ER

PT J
AU Hung, SY
   Liou, HC
   Kang, KH
   Wu, RM
   Wen, CC
   Fu, WM
AF Hung, Shih-Ya
   Liou, Houng-Chi
   Kang, Kai-Hsiang
   Wu, Ruey-Meei
   Wen, Chun-Chiang
   Fu, Wen-Mei
TI Overexpression of Heme Oxygenase-1 Protects Dopaminergic Neurons against
   1-Methyl-4-Phenylpyridinium-Induced Neurotoxicity
SO MOLECULAR PHARMACOLOGY
LA English
DT Article
ID PARKINSONS-DISEASE; NEUROTROPHIC FACTOR; OXIDATIVE STRESS;
   TIN-PROTOPORPHYRIN; HUMAN BRAIN; INJURY; EXPRESSION; ASTROCYTES;
   BILIRUBIN; MONOXIDE
AB Heme oxygenase-1 (HO-1) is up-regulated in response to oxidative stress and catalyzes the degradation of pro-oxidant heme to carbon monoxide ( CO), iron, and bilirubin. Intense HO-1 immunostaining in the Parkinsonian brain is demonstrated, indicating that HO-1 may be involved in the pathogenesis of Parkinsonism. We here locally injected adenovirus containing human HO-1 gene (Ad-HO-1) into rat substantia nigra concomitantly with 1-methyl-4-phenylpyridinium (MPP+). Seven days after injection of MPP+ and Ad-HO-1, the brain was isolated for immunostaining and for measurement of dopamine content and inflammatory cytokines. It was found that overexpression of HO-1 significantly increased the survival rate of dopaminergic neurons; reduced the production of tumor necrosis factor alpha(TNF-alpha) and interleukin-1 beta (IL-1 beta) in substantia nigra; antagonized the reduction of striatal dopamine content induced by MPP+; and also up-regulated brain-derived neurotrophic factor ( BDNF) and glial cell line-derived neurotrophic factor ( GDNF) expression in substantia nigra. Apomorphine-induced rotation after MPP+ treatment was also inhibited by Ad-HO-1. On the other hand, inhibition of HO enzymatic activity by zinc protoporphyrin-IX facilitated the MPP+-induced rotatory behavior and enhanced the reduction of dopamine content. HO-1 overexpression also protected dopaminergic neurons against MPP+-induced neurotoxicity in midbrain neuron-glia cocultures. Overexpression of HO-1 increased the expression of BDNF and GDNF in astrocytes and BDNF in neurons. Our results indicate that HO-1 induction exerts neuroprotection both in vitro and in vivo. Pharmacological or genetic approaches targeting HO-1 may represent a promising and novel therapeutic strategy in treating Parkinsonism.
C1 [Hung, Shih-Ya; Liou, Houng-Chi; Kang, Kai-Hsiang; Fu, Wen-Mei] Natl Taiwan Univ, Coll Med, Inst Pharmacol, Taipei 100, Taiwan.
   [Wu, Ruey-Meei] Natl Taiwan Univ Hosp, Dept Neurol, Taipei, Taiwan.
   [Wen, Chun-Chiang] Natl Taiwan Univ Hosp, Dept Psychiat, Taipei, Taiwan.
C3 National Taiwan University; National Taiwan University; National Taiwan
   University Hospital; National Taiwan University; National Taiwan
   University Hospital
RP Fu, WM (通讯作者)，Natl Taiwan Univ, Coll Med, Inst Pharmacol, 1,Sec 1,Jen Ai Rd, Taipei 100, Taiwan.
EM
OI Wu, Ruey-Meei/0000-0002-4947-5467; FU, WEN-MEI/0000-0003-4157-8987;
   Kang, Kai-Hsiang/0000-0002-6529-4543
FU National Science of Council of Taiwan
FX This work was supported by research grants from the National Science of
   Council of Taiwan.
CR Amersi F, 1999, J CLIN INVEST, V104, P1631, DOI 10.1172/JCI7903
   Baranano DE, 2002, P NATL ACAD SCI USA, V99, P16093, DOI 10.1073/pnas.252626999
   Chen K, 2000, J NEUROCHEM, V75, P304, DOI 10.1046/j.1471-4159.2000.0750304.x
   Chen PS, 2006, MOL PSYCHIATR, V11, P1116, DOI 10.1038/sj.mp.4001893
   Connor B, 1998, BRAIN RES REV, V27, P1, DOI 10.1016/S0165-0173(98)00004-6
   Dore S, 1999, P NATL ACAD SCI USA, V96, P2445, DOI 10.1073/pnas.96.5.2445
   DRUMMOND GS, 1981, P NATL ACAD SCI-BIOL, V78, P6466, DOI 10.1073/pnas.78.10.6466
   Gill SS, 2003, NAT MED, V9, P589, DOI 10.1038/nm850
   Hornykiewicz O, 1987, Adv Neurol, V45, P19
   Huang E, 2005, J NEUROSCI RES, V80, P268, DOI 10.1002/jnr.20444
   Jing CR, 2005, J NEUROSCI RES, V82, P802, DOI 10.1002/jnr.20681
   Juan SH, 2001, CIRCULATION, V104, P1519, DOI 10.1161/hc3801.095663
   Kim HP, 2004, FASEB J, V18, P1080, DOI 10.1096/fj.03-1391com
   Kirschner PB, 1996, BRAIN RES, V713, P178, DOI 10.1016/0006-8993(95)01513-2
   Lang AE, 1998, NEW ENGL J MED, V339, P1044, DOI 10.1056/NEJM199810083391506
   Le W, 1999, J NEUROSCI RES, V56, P652
   LIN LFH, 1993, SCIENCE, V260, P1130, DOI 10.1126/science.8493557
   Lo Bianco C, 2002, P NATL ACAD SCI USA, V99, P10813, DOI 10.1073/pnas.152339799
   Love S, 2005, NAT MED, V11, P703, DOI 10.1038/nm0705-703
   Motterlini R, 2002, CIRC RES, V90, pE17, DOI 10.1161/hh0202.104530
   Munoz AM, 2005, J CHEM NEUROANAT, V29, P113, DOI 10.1016/j.jchemneu.2004.10.001
   OLANOW CW, 1990, NEUROLOGY, V40, P32
   Otterbein LE, 1999, J CLIN INVEST, V103, P1047, DOI 10.1172/JCI5342
   Otterbein LE, 2003, NAT MED, V9, P183, DOI 10.1038/nm817
   Patel NK, 2005, ANN NEUROL, V57, P298, DOI 10.1002/ana.20374
   ROBERTS PJ, 1977, NEUROCHEM RES, V2, P485, DOI 10.1007/BF00966009
   Ryter SW, 2006, PHYSIOL REV, V86, P583, DOI 10.1152/physrev.00011.2005
   Schipper HM, 2004, FREE RADICAL BIO MED, V37, P1995, DOI 10.1016/j.freeradbiomed.2004.09.015
   Schipper HM, 1998, EXP NEUROL, V150, P60, DOI 10.1006/exnr.1997.6752
   Schipper HM, 2000, EXP GERONTOL, V35, P821, DOI 10.1016/S0531-5565(00)00148-0
   SCHIPPER HM, 1995, ANN NEUROL, V37, P758, DOI 10.1002/ana.410370609
   Sindhu KM, 2006, PHARMACOL BIOCHEM BE, V84, P321, DOI 10.1016/j.pbb.2006.05.017
   SMITH A, 1993, J BIOL CHEM, V268, P7365
   Storch A, 2004, J NEURAL TRANSM, V111, P1267, DOI 10.1007/s00702-004-0203-2
   Teng ZP, 2004, NEUROBIOL DIS, V17, P179, DOI 10.1016/j.nbd.2004.07.009
   TENHUNEN R, 1969, J BIOL CHEM, V244, P6388
   Treanor JJS, 1996, NATURE, V382, P80, DOI 10.1038/382080a0
   Valente EM, 2004, SCIENCE, V304, P1158, DOI 10.1126/science.1096284
   Weisiger RA, 2001, J BIOL CHEM, V276, P29953, DOI 10.1074/jbc.M104628200
NR 39
TC 114
Z9 117
U1 0
U2 2
PU AMER SOC PHARMACOLOGY EXPERIMENTAL THERAPEUTICS
PI BETHESDA
PA 9650 ROCKVILLE PIKE, BETHESDA, MD 20814-3995 USA
SN 0026-895X
EI 1521-0111
J9 MOL PHARMACOL
JI Mol. Pharmacol.
PD DEC
PY 2008
VL 74
IS 6
BP 1564
EP 1575
DI 10.1124/mol.108.048611
PG 12
WC Pharmacology & Pharmacy
WE Science Citation Index Expanded (SCI-EXPANDED)
SC Pharmacology & Pharmacy
GA 373WG
UT WOS:000261003600008
PM 18799798
DA 2023-08-27
ER

PT J
AU Miyaji, K
   Furukawa, J
   Suzuki, Y
   Yamamoto, N
   Shinohara, Y
   Yuki, N
AF Miyaji, Kazuki
   Furukawa, Jun-ichi
   Suzuki, Youichi
   Yamamoto, Naoki
   Shinohara, Yasuro
   Yuki, Nobuhiro
TI Altered gene expression of glycosyltransferases and sialyltransferases
   and total amount of glycosphingolipids following herpes simplex virus
   infection
SO CARBOHYDRATE RESEARCH
LA English
DT Article
DE Ganglioside; Guillain-Barre syndrome; Glycosphingolipid; Herpes simplex
   virus
ID GUILLAIN-BARRE-SYNDROME; MASS-SPECTROMETRY; QUANTITATIVE GLYCOMICS;
   CELLULAR GLYCOMICS; CELLS; FIBROBLASTS; GANGLIOSIDE; ANTIBODIES
AB There is a case report of a patient with overlapping Guillain Barre syndrome and Bickerstaff brainstem encephalitis after infection with herpes simplex virus type 1 (HSV-1), who carried high titers of serum anti-GQ1b IgG antibodies. Several studies have linked viral infection to the modulation of ganglioside expression such as human T-lymphotropic virus to GD2 and simian virus 40 to GM3. Also, enhancement of the expression of GM2 on the cell membrane after cytomegalovirus infection has been reported. The objective of this study was to unveil the relationship between HSV-1 infection and the alteration of cellular ganglioside expression in neuronal and glial cell lines. In addition to these cell lines, several human tumor cell lines including astrocytoma cells, neuroblastoma cells, T-cell leukemia cells and kidney cells derived from normal human and monkey were infected with HSV-1 as well as HSV-2. To measure changes in ganglioside-related gene expressions and gangliosides levels in cells, quantitative PCR and glycosphingolipid-glycomic analysis were performed. Changes in gene expression of glycosyltransferases and sialyltransferases were observed in HSV-1- and HSV-2-infected cells, although with different trends. 39 glycosphingolipid-glycans were quantitatively analyzed. HSV-1 and HSV-2 infections resulted in changes in the total amount of gangliosides depending on the cell lines used and type of virus. Qualitative changes caused by each infection of HSV-1 and HSV-2 were almost negligible. (C) 2016 Elsevier Ltd. All rights reserved.
C1 [Miyaji, Kazuki] Natl Univ Singapore, Dept Med, Yong Loo Lin Sch Med, Ctr Translat Med, 14 Med Dr, Singapore 117599, Singapore.
   [Furukawa, Jun-ichi; Shinohara, Yasuro] Hokkaido Univ, Grad Sch Adv Life Sci, Lab Med & Funct Glyc, Sapporo, Hokkaido 0010021, Japan.
   [Furukawa, Jun-ichi; Shinohara, Yasuro] Hokkaido Univ, Frontier Res Ctr Postgenome Sci & Technol, Sapporo, Hokkaido 0010021, Japan.
   [Suzuki, Youichi; Yamamoto, Naoki] Natl Univ Singapore, Yong Loo Lin Sch Med, Dept Microbiol, Ctr Translat Med, 14 Med Dr, Singapore 117599, Singapore.
   [Yuki, Nobuhiro] Natl Univ Singapore, Yong Loo Lin Sch Med, Dept Physiol, Ctr Translat Med, 14 Med Dr, Singapore 117599, Singapore.
C3 National University of Singapore; Hokkaido University; Hokkaido
   University; National University of Singapore; National University of
   Singapore
RP Shinohara, Y (通讯作者)，Hokkaido Univ, Grad Sch Adv Life Sci, Lab Med & Funct Glyc, Sapporo, Hokkaido 0010021, Japan.; Shinohara, Y (通讯作者)，Hokkaido Univ, Frontier Res Ctr Postgenome Sci & Technol, Sapporo, Hokkaido 0010021, Japan.; Yuki, N (通讯作者)，3-9-5 Kami Ikedai,Ota Ku, Tokyo 1450064, Japan.
EM
OI Suzuki, Youichi/0000-0003-3554-0755
FU Singapore National Medical Research Council [IRG 10nov086,
   CSA/047/2012]; Ministry of Education, Culture, Sports, Science and
   Technology (MEXT) of Japan
FX This work was supported by Singapore National Medical Research Council
   (IRG 10nov086 and CSA/047/2012) to N.Y. and Special Coordination Funds
   for Promoting Science and Technology from the Ministry of Education,
   Culture, Sports, Science and Technology (MEXT) of Japan to Y.S.
CR Ang CW, 2000, NEUROLOGY, V54, P1453, DOI 10.1212/WNL.54.7.1453
   BAI HW, 1992, BIOCHIM BIOPHYS ACTA, V1136, P23, DOI 10.1016/0167-4889(92)90080-U
   Bennun SV, 2013, PLOS COMPUT BIOL, V9, DOI 10.1371/journal.pcbi.1002813
   BERNSEN HJJA, 1989, J NEUROL NEUROSUR PS, V52, P788, DOI 10.1136/jnnp.52.6.788
   Cebulla CM, 2000, TRANSPLANTATION, V69, P1202, DOI 10.1097/00007890-200003270-00027
   Fujitani N, 2013, P NATL ACAD SCI USA, V110, P2105, DOI 10.1073/pnas.1214233110
   Fujitani N, 2011, J BIOL CHEM, V286, P41669, DOI 10.1074/jbc.M111.301796
   Furukawa J, 2015, J LIPID RES, V56, P2399, DOI 10.1194/jlr.D062083
   FURUKAWA K, 1993, P NATL ACAD SCI USA, V90, P1972, DOI 10.1073/pnas.90.5.1972
   Jacobs BC, 1998, NEUROLOGY, V51, P1110, DOI 10.1212/WNL.51.4.1110
   Merrill AH, 2005, GLYCOBIOLOGY, V15, p15G, DOI 10.1093/glycob/cwi070
   Miura Y, 2007, CHEM-EUR J, V13, P4797, DOI 10.1002/chem.200601872
   Mori M, 2000, BRAIN, V123, P2171, DOI 10.1093/brain/123.10.2171
   Sodeik B, 1997, J CELL BIOL, V136, P1007, DOI 10.1083/jcb.136.5.1007
   TERRYBERRY J, 1995, J CLIN LAB ANAL, V9, P308, DOI 10.1002/jcla.1860090506
   Uematsu R, 2005, MOL CELL PROTEOMICS, V4, P1977, DOI 10.1074/mcp.M500203-MCP200
   Wheeler SF, 2009, RAPID COMMUN MASS SP, V23, P303, DOI 10.1002/rcm.3867
   Yuki N, 2001, ACTA NEUROL SCAND, V104, P57, DOI 10.1034/j.1600-0404.2001.00288.x
   Yuki N, 2012, NEW ENGL J MED, V366, P2294, DOI 10.1056/NEJMra1114525
NR 19
TC 8
Z9 8
U1 0
U2 6
PU ELSEVIER SCI LTD
PI OXFORD
PA THE BOULEVARD, LANGFORD LANE, KIDLINGTON, OXFORD OX5 1GB, OXON, ENGLAND
SN 0008-6215
EI 1873-426X
J9 CARBOHYD RES
JI Carbohydr. Res.
PD NOV 3
PY 2016
VL 434
BP 37
EP 43
DI 10.1016/j.carres.2016.08.004
PG 7
WC Biochemistry & Molecular Biology; Chemistry, Applied; Chemistry, Organic
WE Science Citation Index Expanded (SCI-EXPANDED)
SC Biochemistry & Molecular Biology; Chemistry
GA EA8BX
UT WOS:000386861400007
PM 27588895
DA 2023-08-27
ER

PT J
AU Hanss, Z
   Larsen, SB
   Antony, P
   Mencke, P
   Massart, F
   Jarazo, J
   Schwamborn, JC
   Barbuti, PA
   Mellick, GD
   Kruger, R
AF Hanss, Zoe
   Larsen, Simone B.
   Antony, Paul
   Mencke, Pauline
   Massart, Francois
   Jarazo, Javier
   Schwamborn, Jens C.
   Barbuti, Peter A.
   Mellick, George D.
   Kruger, Rejko
TI Mitochondrial and Clearance Impairment in p.D620N VPS35 Patient-Derived
   Neurons
SO MOVEMENT DISORDERS
LA English
DT Article
DE VPS35; induced pluripotent stem cells; mitochondrial impairment;
   Parkinson&apos; s disease; &#945; &#8208; synuclein
ID ALPHA-SYNUCLEIN; D620N MUTATION; PARKINSONS; COMPLEX; RETROMER;
   TRAFFICKING; DEGRADATION; AUTOPHAGY; RAPAMYCIN; RECEPTOR
AB Background VPS35 is part of the retromer complex and is responsible for the trafficking and recycling of proteins implicated in autophagy and lysosomal degradation, but also takes part in the degradation of mitochondrial proteins via mitochondria-derived vesicles. The p.D620N mutation of VPS35 causes an autosomal-dominant form of Parkinson's disease (PD), clinically representing typical PD.
   Objective Most of the studies on p.D620N VPS35 were performed on human tumor cell lines, rodent models overexpressing mutant VPS35, or in patient-derived fibroblasts. Here, based on identified target proteins, we investigated the implication of mutant VPS35 in autophagy, lysosomal degradation, and mitochondrial function in induced pluripotent stem cell-derived neurons from a patient harboring the p.D620N mutation.
   Methods We reprogrammed fibroblasts from a PD patient carrying the p.D620N mutation in the VPS35 gene and from two healthy donors in induced pluripotent stem cells. These were subsequently differentiated into neuronal precursor cells to finally generate midbrain dopaminergic neurons.
   Results We observed a decreased autophagic flux and lysosomal mass associated with an accumulation of alpha-synuclein in patient-derived neurons compared to controls. Moreover, patient-derived neurons presented a mitochondrial dysfunction with decreased membrane potential, impaired mitochondrial respiration, and increased production of reactive oxygen species associated with a defect in mitochondrial quality control via mitophagy.
   Conclusion We describe for the first time the impact of the p.D620N VPS35 mutation on autophago-lysosome pathway and mitochondrial function in stem cell-derived neurons from an affected p.D620N carrier and define neuronal phenotypes for future pharmacological interventions. (c) 2020 International Parkinson and Movement Disorder Society
C1 [Hanss, Zoe; Larsen, Simone B.; Antony, Paul; Mencke, Pauline; Massart, Francois; Jarazo, Javier; Schwamborn, Jens C.; Barbuti, Peter A.; Kruger, Rejko] Univ Luxembourg, Luxembourg Ctr Syst Biomed LCSB, Esch Sur Alzette, Luxembourg.
   [Barbuti, Peter A.] Columbia Univ, Med Ctr, Dept Pathol & Cell Biol, New York, NY USA.
   [Barbuti, Peter A.; Kruger, Rejko] Luxembourg Inst Hlth LIH, Transversal Translat Med, Strassen, Luxembourg.
   [Mellick, George D.] Griffith Univ, Griffith Inst Drug Discovery, Nathan, Qld, Australia.
   [Kruger, Rejko] Ctr Hosp Luxembourg CHL, Parkinson Res Clin, Luxembourg, Luxembourg.
C3 University of Luxembourg; Columbia University; Luxembourg Institute of
   Health; Griffith University; Luxembourg Hospital Center
RP Kruger, R (通讯作者)，Univ Luxembourg, LCSB, 6 Ave Swing, L-4367 Belvaux, Luxembourg.
EM
RI Schwamborn, Jens C/F-9395-2011; Barbuti, Peter Antony/AFP-3655-2022;
   Antony, Paul MA/C-8152-2016; Mellick, George/A-7650-2010
OI Schwamborn, Jens C/0000-0003-4496-0559; Barbuti, Peter
   Antony/0000-0001-9182-1629; Antony, Paul MA/0000-0002-0450-9301; Hanss,
   Zoe/0000-0002-3659-3624; Mencke, Pauline/0000-0002-5564-0385; Jarazo,
   Javier/0000-0001-9652-3620; Krueger, Rejko/0000-0003-4258-6241; Mellick,
   George/0000-0002-7211-4651
FU local University; Hertie Institute; DZNE
FX Patient fibroblasts were obtained from Griffith University. We would
   like to thank Prof. Peter Silburn for clinical updates on this patient.
   Control fibroblasts were obtained from the NeuroBiobank of the
   University of Tuebingen, Germany
   (https://www.hihtuebingen.de/en/about-us/core-facilities/biobank/).This
   biobank is supported by the local University, the Hertie Institute, and
   the DZNE. We thank Christine Bus from the University of Tuebingen for
   the plasmids and protocol for reprogramming.
   pX330-U6-Chimeric_BB-CBh-hSpCas9 was a gift from Feng Zhang (Addgene
   plasmid #42230). AAVS1 SA-2Apuro-pA donor was a gift from Rudolf
   Jaenisch (Addgene plasmid #22075). We would like to thank Prof. T.
   Graham and A. Sargsyan from the University of Utah for kindly providing
   us with the pHluorin construct.
CR Arias-Fuenzalida J, 2019, SCI REP-UK, V9, DOI 10.1038/s41598-019-45917-2
   Arighi CN, 2004, J CELL BIOL, V165, P123, DOI 10.1083/jcb.200312055
   Bandres-Ciga S, 2019, MOVEMENT DISORD, V34, P460, DOI 10.1002/mds.27614
   Baumuratov AS, 2016, SCI REP-UK, V6, DOI 10.1038/srep33117
   Bentley SR, 2018, PARKINSONISM RELAT D, V49, P34, DOI 10.1016/j.parkreldis.2017.12.033
   Braschi E, 2009, EMBO REP, V10, P748, DOI 10.1038/embor.2009.86
   Burbulla LF, 2014, CELL DEATH DIS, V5, DOI 10.1038/cddis.2014.103
   Chen X, 2019, P NATL ACAD SCI USA, V116, P5765, DOI 10.1073/pnas.1814909116
   Cong L, 2013, SCIENCE, V339, P819, DOI 10.1126/science.1231143
   Dehay B, 2010, J NEUROSCI, V30, P12535, DOI 10.1523/JNEUROSCI.1920-10.2010
   Deng H, 2013, MOVEMENT DISORD, V28, P569, DOI 10.1002/mds.25430
   Follett J, 2014, TRAFFIC, V15, P230, DOI 10.1111/tra.12136
   Hockemeyer D, 2009, NAT BIOTECHNOL, V27, P851, DOI 10.1038/nbt.1562
   Krebiehl G, 2010, PLOS ONE, V5, DOI 10.1371/journal.pone.0009367
   Kruger R, 2017, J NEURAL TRANSM, V124, P1015, DOI 10.1007/s00702-017-1707-x
   Larsen SB, 2018, CELL TISSUE RES, V373, P21, DOI 10.1007/s00441-017-2768-8
   Larsen SB, 2020, STEM CELL RES, V45, DOI 10.1016/j.scr.2020.101776
   Little D, 2018, SCI REP-UK, V8, DOI 10.1038/s41598-018-27058-0
   MacLeod DA, 2013, NEURON, V77, P425, DOI 10.1016/j.neuron.2012.11.033
   Mali P, 2013, SCIENCE, V339, P823, DOI 10.1126/science.1232033
   Mazzulli JR, 2016, P NATL ACAD SCI USA, V113, P1931, DOI 10.1073/pnas.1520335113
   McGough IJ, 2014, CURR BIOL, V24, P1670, DOI 10.1016/j.cub.2014.06.024
   Miura E, 2014, NEUROBIOL DIS, V71, P1, DOI 10.1016/j.nbd.2014.07.014
   Mohan M, 2017, PARKINSONISM RELAT D, V36, P10, DOI 10.1016/j.parkreldis.2016.12.001
   Munsie LN, 2015, HUM MOL GENET, V24, P1691, DOI 10.1093/hmg/ddu582
   Perrett RM, 2015, MOL CELL NEUROSCI, V66, P21, DOI 10.1016/j.mcn.2015.02.009
   Popovic D, 2014, EMBO REP, V15, P392, DOI 10.1002/embr.201337995
   Reinhardt P, 2013, PLOS ONE, V8, DOI 10.1371/journal.pone.0059252
   Sarkar S, 2009, CELL DEATH DIFFER, V16, P46, DOI 10.1038/cdd.2008.110
   Seaman MNJ, 1998, J CELL BIOL, V142, P665, DOI 10.1083/jcb.142.3.665
   Tang FL, 2015, CELL REP, V12, P1631, DOI 10.1016/j.celrep.2015.08.001
   Tang FL, 2015, J NEUROSCI, V35, P10613, DOI 10.1523/JNEUROSCI.0042-15.2015
   Thoreen CC, 2009, AUTOPHAGY, V5, P725, DOI 10.4161/auto.5.5.8504
   Tsika E, 2014, HUM MOL GENET, V23, P4621, DOI 10.1093/hmg/ddu178
   Vilarino-Guell C, 2011, AM J HUM GENET, V89, P162, DOI 10.1016/j.ajhg.2011.06.001
   Wang WZ, 2016, NAT MED, V22, P54, DOI 10.1038/nm.3983
   Zambon F, 2019, HUM MOL GENET, V28, P2001, DOI 10.1093/hmg/ddz038
   Zanin M, 2020, BIORXIV, DOI 10.1038/s41540-020-00156-4
   Zavodszky E, 2014, NAT COMMUN, V5, DOI 10.1038/ncomms4828
   Zhou LP, 2017, BBA-MOL BASIS DIS, V1863, P2791, DOI 10.1016/j.bbadis.2017.07.032
   Zimprich A, 2011, AM J HUM GENET, V89, P168, DOI 10.1016/j.ajhg.2011.06.008
NR 41
TC 20
Z9 20
U1 0
U2 4
PU WILEY
PI HOBOKEN
PA 111 RIVER ST, HOBOKEN 07030-5774, NJ USA
SN 0885-3185
EI 1531-8257
J9 MOVEMENT DISORD
JI Mov. Disord.
PD MAR
PY 2021
VL 36
IS 3
BP 704
EP 715
DI 10.1002/mds.28365
EA NOV 2020
PG 12
WC Clinical Neurology
WE Science Citation Index Expanded (SCI-EXPANDED)
SC Neurosciences & Neurology
GA RW6KS
UT WOS:000584089400001
PM 33142012
OA hybrid, Green Published
DA 2023-08-27
ER

PT J
AU Alexopoulos, H
   Dalakas, MC
AF Alexopoulos, Harry
   Dalakas, Marinos C.
TI The immunobiology of autoimmune encephalitides
SO JOURNAL OF AUTOIMMUNITY
LA English
DT Review
DE Encephalitis; Autoimmunity; Autoantibodies; Immunotherapy; Synapses;
   Autoimmune neuropsychiatry
ID STIFF-PERSON-SYNDROME; CONTACTIN-ASSOCIATED PROTEIN-2; NMDA-RECEPTOR
   ENCEPHALITIS; GLIOMA-INACTIVATED 1; CEREBROSPINAL-FLUID; BIPOLAR
   DISORDER; NEUROMYELITIS-OPTICA; LIMBIC ENCEPHALITIS; ANTIBODY-TITERS;
   GAD-ANTIBODIES
AB Autoimmune encephalitides, with an estimated incidence of 1.5 per million population per year, although described only 15 years ago, have already had a remarkable impact in neurology and paved the field to autoimmune neuropsychiatry. Many patients traditionally presented with aberrant behavior, especially of acute or subacute onset, and treated with anti-psychotic therapies, turn out to have a CNS autoimmune disease with pathogenic autoantibodies against synaptic antigens responding to immunotherapies. The review describes the clinical spectrum of these disorders, and the pathogenetic role of key autoantibodies directed against: a) cell surface synaptic antigens and receptors, including NMDAR, GABAa, GABAb, AMPA and glycine receptors; b) channels such as AQP4 water-permeable channel or voltage-gated potassium channels; c) proteins that stabilize voltage-gated potassium channel complex into the membrane, like the LGI1 and CASPR2; and d) enzymes that catalyze the formation of neurotransmitters such as Glutamic Acid Decarboxylase (GAD). These antibodies, effectively target excitatory or inhibitory synapses in the limbic system, basal ganglia or brainstem altering synaptic function and resulting in uncontrolled neurological excitability disorder clinically manifested with psychosis, agitation, behavioral alterations, depression, sleep disturbances, seizure-like phenomena, movement disorders such as ataxia, chorea and dystonia, memory changes or coma. Some of the identified triggering factors include: viruses, especially herpes simplex, accounting for the majority of relapses occurring after viral encephalitis, which respond to immunotherapy rather than antiviral agents; tumors especially teratoma, SCLC and thymomas; and biological cancer therapies (immune-check-point inhibitors). As anti-synaptic antibodies persist after viral infections or tumor removal, augmentation of autoreactive B cells which release autoantigens to draining lymph nodes, molecular mimicry and infection-induced bystander immune activation products play a role in autoimmunization process or perpetuating autoimmune neuroinflammation. The review stresses the importance of early detection, clinical recognition, proper antibody testing and early therapy initiation as these disorders, regardless of a known or not trigger, are potentially treatable responding to systemic immunotherapy with intravenous steroids, IVIg, rituximab or even bortezomid
C1 [Alexopoulos, Harry; Dalakas, Marinos C.] Univ Athens, Fac Med, Neuroimmunol Unit, Dept Pathophysiol, Athens, Greece.
   [Dalakas, Marinos C.] Thomas Jefferson Univ, Dept Neurol, Philadelphia, PA 19107 USA.
C3 CIVIS; National & Kapodistrian University of Athens; Jefferson
   University
RP Dalakas, MC (通讯作者)，Univ Athens, Dept Pathophysiol, Neuroimmunol Unit, Mikras Asias 75, Athens 11527, Greece.
EM
CR Al-Diwani A, 2017, FRONT PSYCHIATRY, V8, DOI 10.3389/fpsyt.2017.00013
   Al-Diwani AAJ, 2017, IMMUNOLOGY, V152, P388, DOI 10.1111/imm.12795
   Alexopoulos H, 2018, THER ADV NEUROL DISO, V11, DOI 10.1177/1756286418768778
   Alexopoulos H, 2014, NEUROL-NEUROIMMUNOL, V1, DOI 10.1212/NXI.0000000000000039
   Alexopoulos H, 2013, EXPERT REV CLIN IMMU, V9, P1043, DOI 10.1586/1744666X.2013.845527
   Alexopoulos H, 2011, J NEUROL, V258, P1568, DOI 10.1007/s00415-011-5982-4
   Alexopoulos H, 2010, EUR J CLIN INVEST, V40, P1018, DOI 10.1111/j.1365-2362.2010.02340.x
   Ameli R, 2005, NEUROLOGY, V64, P1961, DOI 10.1212/01.WNL.0000163984.71993.FE
   Anaya JM, 2016, BMC MED, V14, DOI 10.1186/s12916-016-0601-y
   Arino H, 2015, JAMA NEUROL, V72, P874, DOI 10.1001/jamaneurol.2015.0749
   Armangue T, 2018, LANCET NEUROL, V17, P760, DOI 10.1016/S1474-4422(18)30244-8
   Armangue T, 2014, CURR OPIN NEUROL, V27, P361, DOI 10.1097/WCO.0000000000000087
   Armangue T, 2014, ANN NEUROL, V75, P317, DOI 10.1002/ana.24083
   Bale JF, 2015, PEDIATR NEUROL, V53, P3, DOI 10.1016/j.pediatrneurol.2015.03.013
   Balint B, 2018, BRAIN, V141, P13, DOI 10.1093/brain/awx189
   Bastiaansen AEM, 2017, CURR OPIN NEUROL, V30, P302, DOI 10.1097/WCO.0000000000000444
   Bauer J, 2016, HAND CLINIC, V133, P107, DOI 10.1016/B978-0-444-63432-0.00007-4
   Bechter K, 2019, FRONT PSYCHIATRY, V10, DOI 10.3389/fpsyt.2019.00169
   Benros ME, 2011, AM J PSYCHIAT, V168, P1303, DOI 10.1176/appi.ajp.2011.11030516
   Berk M, 2011, NEUROSCI BIOBEHAV R, V35, P804, DOI 10.1016/j.neubiorev.2010.10.001
   Bien CG, 2017, EUR J NEUROL, V24, P175, DOI 10.1111/ene.13180
   Bien CG, 2012, BRAIN, V135, P1622, DOI 10.1093/brain/aws082
   Binks SNM, 2018, J NEUROL NEUROSUR PS, V89, P526, DOI 10.1136/jnnp-2017-315720
   Bost C, 2018, J NEUROL, V265, P2190, DOI 10.1007/s00415-018-8970-0
   Bradshaw MJ, 2018, SEMIN NEUROL, V38, P330, DOI 10.1055/s-0038-1660821
   Bradshaw MJ, 2016, NEUROTHERAPEUTICS, V13, P493, DOI 10.1007/s13311-016-0433-7
   Bradshaw MJ, 2015, NEUROLOGY, V85, P2176, DOI 10.1212/WNL.0000000000002218
   Brietzke E, 2009, BRAIN BEHAV IMMUN, V23, P1079, DOI 10.1016/j.bbi.2009.04.008
   Brietzke E, 2009, J AFFECT DISORDERS, V116, P214, DOI 10.1016/j.jad.2008.12.001
   Brimberg L, 2015, TRENDS IMMUNOL, V36, P709, DOI 10.1016/j.it.2015.09.008
   Carvajal-Gonzalez A, 2014, BRAIN, V137, P2178, DOI 10.1093/brain/awu142
   Castillo-Gomez E, 2017, MOL PSYCHIATR, V22, P1776, DOI 10.1038/mp.2016.125
   Colijn MA, 2019, NEUROPSYCHOBIOLOGY, V78, P70, DOI 10.1159/000499714
   Coutinho E, 2014, BIOL PSYCHIAT, V75, P269, DOI 10.1016/j.biopsych.2013.07.040
   Crisp SJ, 2017, CURR OPIN NEUROL, V30, P310, DOI 10.1097/WCO.0000000000000450
   Cutforth T, 2016, FUTUR NEUROL, V11, P63, DOI 10.2217/fnl.16.4
   Dalakas MC, 2018, THER ADV NEUROL DISO, V11, DOI 10.1177/1756286418799864
   Dalakas MC, 2013, EXP NEUROL, V247, P303, DOI 10.1016/j.expneurol.2013.02.010
   Dalakas MC, 2009, CURR TREAT OPTION NE, V11, P102, DOI 10.1007/s11940-009-0013-9
   Dalakas MC, 2000, NEUROLOGY, V55, P1531, DOI 10.1212/WNL.55.10.1531
   Dalmau J, 2019, LANCET NEUROL, V18, P1045, DOI 10.1016/S1474-4422(19)30244-3
   Dalmau J, 2018, NEW ENGL J MED, V378, P840, DOI 10.1056/NEJMra1708712
   Dalmau J, 2017, PHYSIOL REV, V97, P839, DOI 10.1152/physrev.00010.2016
   Dalmau J, 2008, LANCET NEUROL, V7, P1091, DOI 10.1016/S1474-4422(08)70224-2
   Dantzer R, 2008, NAT REV NEUROSCI, V9, P46, DOI 10.1038/nrn2297
   Dinkel K, 1998, ANN NEUROL, V44, P194, DOI 10.1002/ana.410440209
   Dowlati Y, 2010, BIOL PSYCHIAT, V67, P446, DOI 10.1016/j.biopsych.2009.09.033
   Dubey D, 2018, ANN NEUROL, V83, P166, DOI 10.1002/ana.25131
   Finke C, 2019, LANCET PSYCHIAT, V6, P191, DOI 10.1016/S2215-0366(19)30038-0
   Fukata M, 2018, CURR OPIN NEUROBIOL, V48, P1, DOI 10.1016/j.conb.2017.07.012
   Gastaldi M, 2017, METHODS MOL BIOL, V1677, P117, DOI 10.1007/978-1-4939-7321-7_4
   Giannoccaro MP, 2019, BRAIN, V142, P2000, DOI 10.1093/brain/awz119
   Gonzalez-Billault C, 2017, ANN NEUROL, V81, P618, DOI 10.1002/ana.24884
   Gozzard P, 2015, NEUROLOGY, V85, P235, DOI 10.1212/WNL.0000000000001721
   Graus F, 2008, NEUROLOGY, V70, P500, DOI 10.1212/01.wnl.0000299189.43164.96
   Graus F, 2019, NAT REV CLIN ONCOL, V16, P535, DOI 10.1038/s41571-019-0194-4
   Graus F, 2016, LANCET NEUROL, V15, P391, DOI 10.1016/S1474-4422(15)00401-9
   Greenlee John E, 2012, Continuum (Minneap Minn), V18, P1271, DOI 10.1212/01.CON.0000423847.40147.06
   Greenlee JE, 2014, J NEUROINFLAMM, V11, DOI 10.1186/s12974-014-0160-0
   Gresa-Arribas N, 2014, LANCET NEUROL, V13, P167, DOI 10.1016/S1474-4422(13)70282-5
   Gurrera RJ, 2019, PSYCHIAT CLIN NEUROS, V73, P262, DOI 10.1111/pcn.12821
   Hara M, 2018, NEUROLOGY, V90, pE1386, DOI 10.1212/WNL.0000000000005329
   Hardy-Werbin M, 2018, ONCOIMMUNOLOGY, V7, DOI 10.1080/2162402X.2017.1395125
   Haselmann H, 2018, NEURON, V100, P91, DOI 10.1016/j.neuron.2018.07.048
   Hassin-Baer S, 2004, ARCH NEUROL-CHICAGO, V61, P938, DOI 10.1001/archneur.61.6.938
   Herken J, 2017, FRONT PSYCHIATRY, V8, DOI 10.3389/fpsyt.2017.00025
   Hoftberger R, 2013, NEUROLOGY, V81, P1500, DOI 10.1212/WNL.0b013e3182a9585f
   Honnorat J, 2018, REV NEUROL-FRANCE, V174, P228, DOI 10.1016/j.neurol.2017.11.004
   Irani SR, 2014, ANN NEUROL, V76, P168, DOI 10.1002/ana.24200
   Irani SR, 2012, ANN NEUROL, V72, P241, DOI 10.1002/ana.23577
   Irani SR, 2010, BRAIN, V133, P2734, DOI 10.1093/brain/awq213
   Isgren A, 2015, BRAIN BEHAV IMMUN, V43, P198, DOI 10.1016/j.bbi.2014.10.001
   Jain A, 2016, HAND CLINIC, V133, P77, DOI 10.1016/B978-0-444-63432-0.00005-0
   Jakobsson J, 2015, J PSYCHIATR NEUROSCI, V40, P250, DOI 10.1503/jpn.140183
   Jarius S, 2010, J NEUROIMMUNOL, V229, P219, DOI 10.1016/j.jneuroim.2010.07.019
   Jarius S, 2016, J NEUROINFLAMM, V13, DOI 10.1186/s12974-016-0737-x
   Jeffery OJ, 2013, NEUROLOGY, V81, P882, DOI 10.1212/WNL.0b013e3182a35271
   Jezequel J, 2017, NAT COMMUN, V8, DOI 10.1038/s41467-017-01700-3
   Joubert B, 2019, REV NEUROL-FRANCE, V175, P420, DOI 10.1016/j.neurol.2019.07.004
   Joubert B, 2016, JAMA NEUROL, V73, P1115, DOI 10.1001/jamaneurol.2016.1585
   Joubert B, 2015, BBA-BIOMEMBRANES, V1848, P2665, DOI 10.1016/j.bbamem.2015.04.003
   Karagianni P., 2019, CLIN IMMUNOLOGY ORLA
   Kelleher E, 2020, CURR TOP BEHAV NEURO, V44, P85, DOI 10.1007/7854_2019_90
   Klein CJ, 2013, JAMA NEUROL, V70, P229, DOI 10.1001/jamaneurol.2013.592
   Koch H, 2019, SCI REP-UK, V9, DOI 10.1038/s41598-019-41849-z
   Kreye J, 2016, BRAIN, V139, P2641, DOI 10.1093/brain/aww208
   Kruse JL, 2015, PSYCHOSOMATICS, V56, P227, DOI 10.1016/j.psym.2015.01.003
   Ladepeche L, 2018, CELL REP, V23, P3759, DOI 10.1016/j.celrep.2018.05.096
   Lai MZ, 2009, ANN NEUROL, V65, P424, DOI 10.1002/ana.21589
   Lancaster E, 2010, LANCET NEUROL, V9, P67, DOI 10.1016/S1474-4422(09)70324-2
   Laurence M, 2017, MULT SCLER RELAT DIS, V16, P8, DOI 10.1016/j.msard.2017.05.009
   Leis AA, 2014, MUSCLE NERVE, V49, P26, DOI 10.1002/mus.23869
   Leonard B, 2012, NEUROSCI BIOBEHAV R, V36, P764, DOI 10.1016/j.neubiorev.2011.12.005
   Levy Lucien M., 1999, Annals of Internal Medicine, V131, P522
   Leypoldt F, 2015, ANN NY ACAD SCI, V1338, P94, DOI 10.1111/nyas.12553
   Liblau R, 1998, ANN MED INTERNE, V149, P512
   Linnoila JJ, 2016, NEUROL-NEUROIMMUNOL, V3, DOI 10.1212/NXI.0000000000000245
   Linnoila JJ, 2014, SEMIN NEUROL, V34, P458, DOI 10.1055/s-0034-1390394
   Liu Y, 2012, J AFFECT DISORDERS, V139, P230, DOI 10.1016/j.jad.2011.08.003
   Ludvigsson JF, 2007, SCAND J GASTROENTERO, V42, P179, DOI 10.1080/00365520600863472
   Lungen EM, 2019, FRONT PSYCHIATRY, V10, DOI 10.3389/fpsyt.2019.00226
   Maes M, 2012, PROG NEURO-PSYCHOPH, V36, P169, DOI 10.1016/j.pnpbp.2011.09.006
   Magana SM, 2009, NEUROLOGY, V72, P712, DOI 10.1212/01.wnl.0000343001.36493.ae
   Magira EE, 2016, THER ADV NEUROL DISO, V9, P69, DOI 10.1177/1756285615614812
   Mane-Damas M, 2019, AUTOIMMUN REV, V18, DOI 10.1016/j.autrev.2019.102348
   MANLEY GT, 1995, ANN NEUROL, V38, P102, DOI 10.1002/ana.410380117
   McKeon A, 2016, HAND CLINIC, V133, P301, DOI 10.1016/B978-0-444-63432-0.00017-7
   Mikasova L, 2012, BRAIN, V135, P1606, DOI 10.1093/brain/aws092
   Miller AH, 2009, BIOL PSYCHIAT, V65, P732, DOI 10.1016/j.biopsych.2008.11.029
   Miller BJ, 2011, BMJ-BRIT MED J, V343, DOI 10.1136/bmj.d5652
   Modabbernia A, 2013, BIOL PSYCHIAT, V74, P15, DOI 10.1016/j.biopsych.2013.01.007
   Mundiyanapurath S, 2013, J NEUROIMMUNOL, V259, P88, DOI 10.1016/j.jneuroim.2013.04.004
   Musselman DL, 2001, AM J PSYCHIAT, V158, P1252, DOI 10.1176/appi.ajp.158.8.1252
   Najjar S, 2013, J NEUROINFLAMM, V10, DOI 10.1186/1742-2094-10-142
   Nemni R, 2004, NEUROLOGY, V62, P2101, DOI 10.1212/01.WNL.0000127446.61806.2F
   Niki M, 2019, MOL CLIN ONCOL, V10, P267, DOI 10.3892/mco.2018.1777
   Pape K, 2019, NAT REV NEUROL, V15, P317, DOI 10.1038/s41582-019-0174-4
   Petit-Pedrol M, 2018, BRAIN, V141, P3144, DOI 10.1093/brain/awy253
   Planaguma J, 2015, BRAIN, V138, P94, DOI 10.1093/brain/awu310
   Platt MP, 2017, FRONT IMMUNOL, V8, DOI 10.3389/fimmu.2017.00442
   POSNER JB, 1995, PERSPECT BIOL MED, V38, P167
   Postal M, 2015, AUTOIMMUN REV, V14, P30, DOI 10.1016/j.autrev.2014.09.001
   Probst C, 2015, NEUROL-NEUROIMMUNOL, V2, DOI 10.1212/NXI.0000000000000068
   Pruss H, 2018, BRAIN RES, V1701, P146, DOI 10.1016/j.brainres.2018.09.009
   Raju R, 2005, J IMMUNOL, V175, P7755, DOI 10.4049/jimmunol.175.11.7755
   Rakocevic G, 2004, ARCH NEUROL-CHICAGO, V61, P902, DOI 10.1001/archneur.61.6.902
   Sabater L, 2016, J NEUROINFLAMM, V13, DOI 10.1186/s12974-016-0689-1
   Salovin A, 2018, NEUROL-NEUROIMMUNOL, V5, DOI 10.1212/NXI.0000000000000458
   Schneider S, 2017, CASE REP ONCOL, V10, P473, DOI 10.1159/000477162
   Shah S, 2018, CASE REP NEUROL MED, V2018, DOI 10.1155/2018/2548528
   Stefansson H, 2009, NATURE, V460, P744, DOI 10.1038/nature08186
   Strik H, 2017, EUR J CANCER, V87, P205, DOI 10.1016/j.ejca.2017.09.026
   van Coevorden-Hameete MH, 2014, AUTOIMMUN REV, V13, P299, DOI 10.1016/j.autrev.2013.10.016
   VANCOEVORDENHAMEET, 2019, BRAIN J NEUROL, V142, P1631, DOI DOI 10.1093/BRAIN/AWZ094
   Varley J, 2018, NEUROPHARMACOLOGY, V132, P71, DOI 10.1016/j.neuropharm.2017.04.046
   Vincent A, 2005, CURR OPIN NEUROL, V18, P519, DOI 10.1097/01.wco.0000180660.57801.3f
   Vincent A, 2008, BRAIN, V131, P2536, DOI 10.1093/brain/awn221
   Vincent A, 2011, LANCET NEUROL, V10, P759, DOI 10.1016/S1474-4422(11)70096-5
   Waters P, 2008, ARCH NEUROL-CHICAGO, V65, P913, DOI 10.1001/archneur.65.7.913
   Wenke NK, 2019, ANN NEUROL, V85, P771, DOI 10.1002/ana.25460
   Williams JP, 2018, SEMIN NEUROL, V38, P267, DOI 10.1055/s-0038-1660501
   Willison HJ, 2016, LANCET, V388, P717, DOI 10.1016/S0140-6736(16)00339-1
   Yolken RH, 2008, MOL PSYCHIATR, V13, P470, DOI 10.1038/mp.2008.5
   Zekeridou A, 2019, THORAC CANCER, V10, P1001, DOI 10.1111/1759-7714.13009
   Zetterberg H, 2014, PSYCHIAT RES, V217, P143, DOI 10.1016/j.psychres.2014.03.045
NR 145
TC 34
Z9 38
U1 2
U2 24
PU ACADEMIC PRESS LTD- ELSEVIER SCIENCE LTD
PI LONDON
PA 24-28 OVAL RD, LONDON NW1 7DX, ENGLAND
SN 0896-8411
EI 1095-9157
J9 J AUTOIMMUN
JI J. Autoimmun.
PD NOV
PY 2019
VL 104
AR 102339
DI 10.1016/j.jaut.2019.102339
PG 11
WC Immunology
WE Science Citation Index Expanded (SCI-EXPANDED); Social Science Citation Index (SSCI)
SC Immunology
GA JL4KC
UT WOS:000495498400014
PM 31611142
DA 2023-08-27
ER

PT J
AU Ciarimboli, G
   Deuster, D
   Knief, A
   Sperling, M
   Holtkamp, M
   Edemir, B
   Pavenstadt, H
   Lanvers-Kaminsky, C
   Zehnhoff-Dinnesen, AA
   Schinkel, AH
   Koepsell, H
   Jurgens, H
   Schlatter, E
AF Ciarimboli, Giuliano
   Deuster, Dirk
   Knief, Arne
   Sperling, Michael
   Holtkamp, Michael
   Edemir, Bayram
   Pavenstaedt, Hermann
   Lanvers-Kaminsky, Claudia
   Zehnhoff-Dinnesen, Antoinette Am
   Schinkel, Alfred H.
   Koepsell, Hermann
   Juergens, Heribert
   Schlatter, Eberhard
TI Organic Cation Transporter 2 Mediates Cisplatin-Induced Oto- and
   Nephrotoxicity and Is a Target for Protective Interventions
SO AMERICAN JOURNAL OF PATHOLOGY
LA English
DT Article
ID COLORIMETRIC ASSAY; HAIR-CELLS; EXPRESSION; KIDNEY; DRUGS; TOXICITY;
   MICE; OTOTOXICITY; OXALIPLATIN; MECHANISMS
AB The use of the effective antineoplastic agent cisplatin is limited by its serious side effects, such as oto- and nephrotoxicity. Ototoxicity is a problem of special importance in children, because deafness hampers their language and psychosocial development. Recently, organic cation transporters (OCTs) were identified in vitro as cellular uptake mechanisms for cisplatin. In the present study, we investigated in an in vivo model the role of OCTs in the development of cisplatin oto- and nephrotoxicity. The functional effects of cisplatin treatment on kidney (24 hours excretion of glucose, water, and protein) and hearing (auditory brainstem response) were studied in wildtype and OCT1/2 double-knockout (KO) mice. No sign of ototoxicity and only mild nephrotoxicity were observed after cisplatin treatment of knockout mice. Comedication of wild-type mice with cisplatin and the organic cation cimetidine protected from ototoxicity and partly from nephrotoxicity. For the first time we showed that OCT2 is expressed in hair cells of the cochlea. Furthermore, cisplatin-sensitive cell lines from pediatric tumors showed no expression of mRNA for OCTs, indicating the feasibility of therapeutic approaches aimed to reduce cisplatin toxicities by competing OCT2-mediated cisplatin uptake in renal proximal tubular and cochlear hair cells. These findings are very important to establish chemotherapeutical protocols aimed to maximize the antineoplastic effect of cisplatin while reducing the risk of toxicities. (Am J Pathol 2010,176:1169-1180; DOI: 10.2353/ajpath.2010.090610)
C1 [Ciarimboli, Giuliano; Edemir, Bayram; Pavenstaedt, Hermann; Schlatter, Eberhard] Univ Klinikum Munster, Med Klin & Poliklin D, D-48149 Munster, Germany.
   [Deuster, Dirk; Knief, Arne; Zehnhoff-Dinnesen, Antoinette Am] Univ Klinikum Munster, Klin & Poliklin Phoniatrie & Padiaudiol, D-48149 Munster, Germany.
   [Lanvers-Kaminsky, Claudia; Juergens, Heribert] Univ Klinikum Munster, Klin & Poliklin Padiat Onkol, D-48149 Munster, Germany.
   [Sperling, Michael; Holtkamp, Michael] Univ Munster, Inst Anorgan Chem, D-4400 Munster, Germany.
   [Schinkel, Alfred H.] Netherlands Canc Inst, Amsterdam, Netherlands.
   [Koepsell, Hermann] Univ Wurzburg, Inst Anat & Zellbiol, Wurzburg, Germany.
C3 University of Munster; University of Munster; University of Munster;
   University of Munster; Netherlands Cancer Institute; University of
   Wurzburg
RP Ciarimboli, G (通讯作者)，Univ Klinikum Munster, Med Klin & Poliklin D, Domagkstr 3A, D-48149 Munster, Germany.
EM
RI Deuster, Dirk/A-2479-2012; Sperling, Michael/C-2584-2008; Deuster,
   Dirk/L-4635-2014; Edemir, Bayram/AAF-6845-2020; Holtkamp,
   Michael/B-2159-2017; Lanvers-Kaminsky, Claudia/AAY-2071-2021
OI Sperling, Michael/0000-0001-6676-2487; Deuster,
   Dirk/0000-0002-3476-3300; Ciarimboli, Giuliano/0000-0002-4365-3656
FU Deutsche Krebshilfe [108539]; Deutsche Forschungsgemeinschaft
   [SFB487/A4]
FX Supported by the Deutsche Krebshilfe grant 108539 (to G.C., A.a.Z.-D.,
   and H,J.) and the Deutsche Forschungsgemeinschaft grant SFB487/A4 (to
   H.K.).
CR Alnouti Y, 2006, DRUG METAB DISPOS, V34, P477, DOI 10.1121/dmd.105.006932
   Ciarimboli G, 2008, XENOBIOTICA, V38, P936, DOI [10.1080/00498250701882482, 10.1080/00498250701882482 ]
   Ciarimboli G, 2005, AM J PATHOL, V167, P1477, DOI 10.1016/S0002-9440(10)61234-5
   Filipski KK, 2009, CLIN PHARMACOL THER, V86, P396, DOI 10.1038/clpt.2009.139
   Gekle M, 2004, AM J PHYSIOL-RENAL, V287, pF469, DOI 10.1152/ajprenal.00059.2004
   Hardie NA, 2004, BRAIN RES, V1000, P200, DOI 10.1016/j.brainres.2003.10.071
   Hughes J, 2007, NEPHROLOGY, V12, P452, DOI 10.1111/j.1440-1797.2007.00854.x
   Jonker JW, 2003, MOL CELL BIOL, V23, P7902, DOI 10.1128/MCB.23.21.7902-7908.2003
   Kalow W, 1999, PHARMACOLOGY, V58, P281, DOI 10.1159/000028292
   Kerb R, 2006, CANCER LETT, V234, P4, DOI 10.1016/j.canlet.2005.06.051
   Kim SW, 2001, J AM SOC NEPHROL, V12, P875, DOI 10.1681/ASN.V125875
   Koepsell H, 2004, PFLUG ARCH EUR J PHY, V447, P666, DOI 10.1007/s00424-003-1089-9
   Koepsell H, 2004, REV PHYSIOL BIOCH P, V150, P36, DOI 10.1007/s10254-003-0017-x
   Kuo YM, 2006, J NUTR, V136, P21, DOI 10.1093/jn/136.1.21
   Larson CA, 2009, MOL PHARMACOL, V75, P324, DOI 10.1124/mol.108.052381
   Lee WK, 2009, AM J PHYSIOL-RENAL, V296, pF1504, DOI 10.1152/ajprenal.90754.2008
   Livak KJ, 2001, METHODS, V25, P402, DOI 10.1006/meth.2001.1262
   Ludwig T, 2004, KIDNEY INT, V66, P196, DOI 10.1111/j.1523-1755.2004.00720.x
   MOSMANN T, 1983, J IMMUNOL METHODS, V65, P55, DOI 10.1016/0022-1759(83)90303-4
   Motohashi H, 2002, J AM SOC NEPHROL, V13, DOI 10.1681/ASN.V134866
   Pabla N, 2008, KIDNEY INT, V73, P994, DOI 10.1038/sj.ki.5002786
   Pabla N, 2009, AM J PHYSIOL-RENAL, V296, pF505, DOI 10.1152/ajprenal.90545.2008
   PASIC TR, 1991, LARYNGOSCOPE, V101, P985
   Rabik CA, 2007, CANCER TREAT REV, V33, P9, DOI 10.1016/j.ctrv.2006.09.006
   Ramesh G, 2002, J CLIN INVEST, V110, P835, DOI 10.1172/JCI200215606
   Ramirez-Camacho R, 2008, J LARYNGOL OTOL, V122, P1151, DOI 10.1017/S002221510700134X
   Safaei R, 2005, CRIT REV ONCOL HEMAT, V53, P13, DOI 10.1016/j.critrevonc.2004.09.007
   Sha SH, 2001, HEARING RES, V155, P1, DOI 10.1016/S0378-5955(01)00224-6
   Skinner R, 2004, EUR J CANCER, V40, P2352, DOI 10.1016/j.ejca.2004.08.002
   TADA H, 1986, J IMMUNOL METHODS, V93, P157, DOI 10.1016/0022-1759(86)90183-3
   Turner JR, 2000, AM J PHYSIOL-CELL PH, V279, pC1918, DOI 10.1152/ajpcell.2000.279.6.C1918
   Wang D, 2005, NAT REV DRUG DISCOV, V4, P307, DOI 10.1038/nrd1691
   Yokoo S, 2007, BIOCHEM PHARMACOL, V74, P477, DOI 10.1016/j.bcp.2007.03.004
   Yonezawa A, 2005, BIOCHEM PHARMACOL, V70, P1823, DOI 10.1016/j.bcp.2005.09.020
   Yonezawa A, 2006, J PHARMACOL EXP THER, V319, P879, DOI 10.1124/jpet.106.110346
   Zhang SZ, 2006, CANCER RES, V66, P8847, DOI 10.1158/0008-5472.CAN-06-0769
NR 36
TC 314
Z9 329
U1 0
U2 43
PU AMER SOC INVESTIGATIVE PATHOLOGY, INC
PI BETHESDA
PA 9650 ROCKVILLE PIKE, BETHESDA, MD 20814-3993 USA
SN 0002-9440
J9 AM J PATHOL
JI Am. J. Pathol.
PD MAR
PY 2010
VL 176
IS 3
BP 1169
EP 1180
DI 10.2353/ajpath.2010.090610
PG 12
WC Pathology
WE Science Citation Index Expanded (SCI-EXPANDED)
SC Pathology
GA 565MI
UT WOS:000275297000016
PM 20110413
OA hybrid, Green Published
DA 2023-08-27
ER

PT J
AU Schick, B
   Brors, D
   Koch, O
   Schafers, M
   Kahle, G
AF Schick, B
   Brors, D
   Koch, O
   Schafers, M
   Kahle, G
TI Magnetic resonance imaging in patients with sudden hearing loss,
   tinnitus and vertigo
SO OTOLOGY & NEUROTOLOGY
LA English
DT Article; Proceedings Paper
CT 23rd Meeting of the Association-for-Research-in-Otolaryngology
CY FEB 21-23, 2000
CL ST PETERSBURG, FL
SP Assoc Res Otolaryngol
DE magnetic resonance imaging; sudden hearing loss; tinnitus; vertigo
ID INNER-EAR; DEAFNESS
AB Objective: The etiopathogenesis in audiovestibular symptoms can be elusive, despite extensive differential diagnosis. This article addresses the value of magnetic resonance imaging (MRI) in analysis of the complete audiovestibular pathway.
   Study Design: Retrospective evaluation. Setting: Tertiary referral center.
   Patients: Consecutive sample of 354 patients (mean age 49 years, range 8 to 86 years) with audiovestibular disorders.
   Intervention: Contrast-enhanced MRI of the head with thin-slice investigation of the inner ear, internal auditory meatus, and cerebellopontine angle.
   Main Outcome Measure: All MRIs were evaluated by experienced independent investigators. Statistical analysis was performed using the Statistical Package of Social Sciences data analysis 9.0.
   Results: MRI abnormalities were seen in 122 of 354 patients (34.5%). The MRIs revealed the following: 4 pathologic conditions (1.1%) of the cochlea/labyrinth, 23 abnormalities (6.5%) at the internal auditory meatus/cerebellopontine angle, 12 pathologic lesions (3.4%) that involved the central audiovestibular tract at the brainstem, 78 microangiopathic changes of the brain (22%), 3 focal hyperintensities of the brain that turned out to be the first evidence of multiple sclerosis in 2 patients and sarcoidosis in 1 patient, and 1 temporal metastasis. Other pathologic conditions, such as parotid gland or petrous bone apex tumors, were unrelated to the audiovestibular symptoms.
   Conclusions: This study indicates that contrast-enhanced MRI can be used to assess a significant number of different pathologic conditions in patients with audiovestibular disorders.
C1 Univ Hosp Homburg Saar, Dept Ear Nose & Throat Dis Head & Neck Surg, Homburg, Germany.
   Univ Marburg, Dept Ear Nose & Throat Dis Head Neck & Facial, Hosp Fulda, Acad Teaching Hosp, Marburg, Germany.
   Univ Marburg, Inst Radiol, Hosp Fulda, Acad Teaching Hosp, Marburg, Germany.
   Univ Hosp Wurzburg, Dept Neurol, Wurzburg, Germany.
C3 Universitatsklinikum des Saarlandes; Philipps University Marburg;
   Philipps University Marburg; University of Wurzburg
RP Schick, B (通讯作者)，Univ Homburg, Dept Ear Nose & Throat Dis, Kirrberger Str, D-66421 Homburg, Germany.
CR Carrier LTCDA, 1997, OTOLARYNG HEAD NECK, V116, P567
   CASSELMAN JW, 1994, ACTA OTO-LARYNGOL, P15
   Daniels RL, 1998, OTOLARYNG HEAD NECK, V119, P364, DOI 10.1016/S0194-5998(98)70080-4
   Doyle KJ, 1996, OTOLARYNG HEAD NECK, V114, P482, DOI 10.1016/S0194-5998(96)70224-3
   DRULOVIC B, 1994, AUDIOLOGY, V33, P195
   FITZGERALD DC, 1988, AJNR, V19, P1433
   Hudgins PA, 1998, AM J NEURORADIOL, V19, P1807
   Katzman GL, 1999, JAMA-J AM MED ASSOC, V282, P36, DOI 10.1001/jama.282.1.36
   Makins AE, 1998, LARYNGOSCOPE, V108, P1739, DOI 10.1097/00005537-199811000-00027
   Roob G, 1999, NEUROLOGY, V52, P991, DOI 10.1212/WNL.52.5.991
   ROSSEAU GL, 1993, AM J OTOL, V14, P392
   Stokroos RJ, 1998, EUR ARCH OTO-RHINO-L, V255, P433, DOI 10.1007/s004050050093
   Weber PC, 1997, OTOLARYNG HEAD NECK, V116, P153, DOI 10.1016/S0194-5998(97)70317-6
   WU W, 1995, ACTA RADIOL, V36, P603
NR 14
TC 45
Z9 51
U1 0
U2 7
PU LIPPINCOTT WILLIAMS & WILKINS
PI PHILADELPHIA
PA TWO COMMERCE SQ, 2001 MARKET ST, PHILADELPHIA, PA 19103 USA
SN 1531-7129
EI 1537-4505
J9 OTOL NEUROTOL
JI Otol. Neurotol.
PD NOV
PY 2001
VL 22
IS 6
BP 808
EP 812
DI 10.1097/00129492-200111000-00016
PG 5
WC Clinical Neurology; Otorhinolaryngology
WE Science Citation Index Expanded (SCI-EXPANDED); Conference Proceedings Citation Index - Science (CPCI-S)
SC Neurosciences & Neurology; Otorhinolaryngology
GA 494CG
UT WOS:000172260700016
PM 11698800
DA 2023-08-27
ER

PT J
AU Chovanec, M
   Zverina, E
   Profant, O
   Skrivan, J
   Cakrt, O
   Lisy, J
   Betka, J
AF Chovanec, Martin
   Zverina, Eduard
   Profant, Oliver
   Skrivan, Jiri
   Cakrt, Ondrej
   Lisy, Jiri
   Betka, Jan
TI Impact of video-endoscopy on the results of retrosigmoid-transmeatal
   microsurgery of vestibular schwannoma: prospective study
SO EUROPEAN ARCHIVES OF OTO-RHINO-LARYNGOLOGY
LA English
DT Article
DE Vestibular schwannoma; Acoustic neuroma; Endoscopy-assisted
   microsurgery; Retrosigmoid approach; Hearing preservation; Facial nerve
ID ACOUSTIC NEUROMA SURGERY; NERVE
AB Endoscopy-assisted microsurgery represents modern trend of treatment of the cerebellopontine angle (CPA) pathologies including vestibular schwannoma (VS). Endoscopes are used in adjunct to microscope to achieve better functional results with less morbidity. Angled optics, magnification and illumination enable superior view in the operative field. Consecutive 89 patients with untreated unilateral sporadic vestibular schwannoma undergoing tumor resection via a retrosigmoid approach during 2008-2010 were prospectively analysed. Endoscopy-assisted microsurgical (EA-MS) removal was performed in 39 cases (Grade 1: 2, Grade 2: 5, Grade 3: 9, Grade 4: 22, Grade 5: 1) and microsurgical (MS) removal was performed in 50 cases (Grade 1: 1, Grade 2: 3, Grade 3: 9, Grade 4: 34, Grade 5: 3). Minimally invasive approach with craniotomy a parts per thousand currency sign2.5 cm was employed for small tumors (Grade 1 and 2) in the EA-MS group. Endoscopic technique was used for monitoring of neuro-vascular anatomy in CPA, during dissection of the meatal portion of tumors, assessment of radicality and for identification of potential pathways for CSF leak formation. All cases in MS group were deemed as radically removed. In the EA-MS group, residual tumor tissue in the fundus of internal auditory canal not observable with microscope was identified with endoscope in four cases. Such cases were radicalized. Tumor recurrence was not observed during the follow-up in EA-MS group. There is a suspicious intrameatal tumor recurrence on the repeated MRI scan in one patient in the MS group. Neither mortality nor infection was observed. The most common complication was pseudomeningocele (EA-MS 20 cases; MS 23). It was managed with aspiration with or without tissue-gluing in all cases without the need for any surgical revision. Adjunctive use of endoscope in the EA-MS group identified potential pathways for CSF leak formation, which was not observable with the microscope in five patients. Improved cochlear nerve (EA-MS: 22, MS: 14; p = 0.012), brainstem auditory evoked potentials (EA-MS: 3 of 8, MS: 0 of 4) and hearing (EA-MS: 14 of 36, MS: 4 of 45; p = 0.001) preservation were observed in EA-MS group. Despite the trend for better useful hearing (Gardner-Robertson class 1 and 2) preservation (EA-MS: 8 of 26, MS: 1 of 16) there were no significant differences in the postoperative hearing handicap inventory in both groups. There were no differences in the postoperative tinnitus in both groups. Better facial nerve preservation (EA-MS: 39, MS: 44; p = 0.027) and excellent-very good (House-Brackmann 1 or 2) facial nerve function (EA-MS: 31, MS: 29; p = 0.035) were observed in EA-MS group. Postoperative compensation of vestibular lesion, symptoms typical for VS, patients assessed by dizziness handicap inventory, facial disability index were comparable in both studied groups. Adjunctive use of endoscope during the VS surgery due to its magnification and illumination enable superior view in the operative field. It is valuable for assessment of radicality of resection in the region of internal auditory meatus. Improved information about critical structures and tumor itself helps the surgeon to preserve facial nerve and in selected cases also hearing. These techniques can help to decrease incidence of postoperative complications.
C1 [Chovanec, Martin; Zverina, Eduard; Profant, Oliver; Skrivan, Jiri; Betka, Jan] Charles Univ Prague, Dept Otorhinolaryngol Head & Neck Surg, Fac Med 1, Fac Hosp Motol, Prague 15006 5, Czech Republic.
   [Chovanec, Martin] Charles Univ Prague, Inst Anat, Fac Med 1, Prague 15006 5, Czech Republic.
   [Zverina, Eduard] Charles Univ Prague, Dept Neurosurg, Fac Hosp Kralovske Vinohrady, Fac Med 3, Prague 15006 5, Czech Republic.
   [Profant, Oliver] Acad Sci Czech Republic, Dept Auditory Neurosci, Inst Expt Med, Prague 14220 4, Czech Republic.
   [Cakrt, Ondrej] Charles Univ Prague, Dept Rehabil & Sport Med, Fac Med 2, Univ Hosp Motol, Prague 15006 5, Czech Republic.
   [Cakrt, Ondrej] Czech Tech Univ, Joint Ctr Biomed Engn, Fac Biomed Engn, CR-16635 Prague, Czech Republic.
   [Cakrt, Ondrej] Charles Univ Prague, Prague 15006 5, Czech Republic.
   [Lisy, Jiri] Charles Univ Prague, Dept Radiol, Fac Med 2, Fac Hosp Motol, Prague 15006 5, Czech Republic.
C3 Charles University Prague; Charles University Prague; Charles University
   Prague; University Hospital Vinohrady; Czech Academy of Sciences;
   Institute of Experimental Medicine of the Czech Academy of Sciences;
   Charles University Prague; Motol University Hospital; Czech Technical
   University Prague; Charles University Prague; Charles University Prague
RP Chovanec, M (通讯作者)，Charles Univ Prague, Dept Otorhinolaryngol Head & Neck Surg, Fac Med 1, Fac Hosp Motol, V Uvalu 84, Prague 15006 5, Czech Republic.
EM
RI Zvěřina, Eduard/L-9787-2017; Chovanec, Martin/AAL-2004-2021; Skrivan,
   Jiri/G-3753-2017; Betka, Jan/H-7497-2017
OI Zvěřina, Eduard/0000-0002-1018-972X; Chovanec,
   Martin/0000-0001-9087-0269; Skrivan, Jiri/0000-0002-7044-6310; Lisy,
   Jiri/0000-0003-2255-5923; Betka, Jan/0000-0001-8772-317X
FU Internal Grant Agency of the Ministry of Health of Czech Republic
   [NS/9909-4]
FX This study was supported by the research project of Internal Grant
   Agency of the Ministry of Health of Czech Republic no. NS/9909-4. The
   authors are grateful to Ilja Merunka Bc. and Barbora Hajna Bc. for the
   help in preparation of the manuscript.
CR Cakrt O, 2010, EUR ARCH OTO-RHINO-L, V267, P1355, DOI 10.1007/s00405-010-1227-x
   Castelnuovo P, 2010, EUR ARCH OTO-RHINO-L, V267, P649, DOI 10.1007/s00405-009-1196-0
   GARDNER G, 1988, ANN OTO RHINOL LARYN, V97, P55, DOI 10.1177/000348948809700110
   Gerganov VM, 2010, EJSO-EUR J SURG ONC, V36, P422, DOI 10.1016/j.ejso.2009.11.003
   Goksu N, 2005, ORL-J OTO-RHIN-LARYN, V67, P87, DOI 10.1159/000084623
   Guevara N, 2008, EUR ARCH OTO-RHINO-L, V265, P397, DOI 10.1007/s00405-007-0471-1
   Hori T, 2006, MINIM INVAS NEUROSUR, V49, P25, DOI 10.1055/s-2006-932125
   HOUSE JW, 1985, OTOLARYNG HEAD NECK, V93, P146, DOI 10.1177/019459988509300202
   Magnan J, 2002, OTOL NEUROTOL, V23, P141, DOI 10.1097/00129492-200203000-00006
   Magnan J, 1993, Ann Otolaryngol Chir Cervicofac, V110, P259
   Roberson JB, 1996, AM J OTOL, V17, P307
   Shahinian HK, 2011, MINIM INVAS NEUROSUR, V54, P61, DOI 10.1055/s-0031-1275335
   Skrivan J, 2004, Otolaryngol Pol, V58, P69
   Tatagiba M, 1996, NEUROSURGERY, V38, P737, DOI 10.1227/00006123-199604000-00021
   Zverina Eduard, 2010, Cas Lek Cesk, V149, P269
NR 15
TC 18
Z9 18
U1 0
U2 14
PU SPRINGER
PI NEW YORK
PA 233 SPRING ST, NEW YORK, NY 10013 USA
SN 0937-4477
J9 EUR ARCH OTO-RHINO-L
JI Eur. Arch. Oto-Rhino-Laryn.
PD MAR
PY 2013
VL 270
IS 4
BP 1277
EP 1284
DI 10.1007/s00405-012-2112-6
PG 8
WC Otorhinolaryngology
WE Science Citation Index Expanded (SCI-EXPANDED)
SC Otorhinolaryngology
GA 115NC
UT WOS:000316818000015
PM 23010789
DA 2023-08-27
ER

PT J
AU Martinage, G
   Geffrelot, J
   Stefan, D
   Bogart, E
   Rault, E
   Reyns, N
   Emery, E
   Makhlouff-Martinage, S
   Mouttet-Audouard, R
   Basson, L
   Mirabel, X
   Lartigau, E
   Pasquier, D
AF Martinage, Geoffrey
   Geffrelot, Julien
   Stefan, Dinu
   Bogart, Emilie
   Rault, Erwan
   Reyns, Nicolas
   Emery, Evelyne
   Makhlouff-Martinage, Samira
   Mouttet-Audouard, Raphaelle
   Basson, Laurent
   Mirabel, Xavier
   Lartigau, Eric
   Pasquier, David
TI Efficacy and Tolerance of Post-operative Hypo-Fractionated Stereotactic
   Radiotherapy in a Large Series of Patients With Brain Metastases
SO FRONTIERS IN ONCOLOGY
LA English
DT Article
DE radiotherapy; hypofractionated stereotactic radiation therapy; brain
   metastasis; surgery; Cyberknife
ID SURGICAL RESECTION; SINGLE METASTASES; SOLID TUMORS; RADIOSURGERY;
   CAVITY; SURVIVAL; TRIAL; RISK; BED; GUIDELINE
AB Purpose: The aim of this study was to assess, in a large series, the efficacy and tolerance of post-operative adjuvant hypofractionated stereotactic radiation therapy (HFSRT) for brain metastases (BMs).
   Materials and Methods: Between July 2012 and January 2017, 160 patients from 2 centers were operated for BM and treated by HFSRT. Patients had between 1 and 3 BMs, no brainstem lesions or carcinomatous meningitis. The primary endpoint was local control. Secondary endpoints were distant brain control, overall survival (OS) and tolerance to HFSRT.
   Results: 73 patients (46%) presented with non-small cell lung cancer (NSCLC), 23 (14%) had melanoma and 21 (13%) breast cancer. Median age was 58 years (range, 22-83 years). BMs were synchronous in 50% of the cases. The most frequent prescription regimens were 24Gy in 3 fractions (n = 52, 33%) and 30Gy in 5 fractions (n = 37, 23%). Local control rates at 1 and 2 years were 88% [95% CI, 81-93%] and 81% [95% CI, 70-88%], respectively. Distant control rate at 1 year was 48% [95% CI, 81-93%]. In multivariate analysis, primary NSCLC was associated with a significant reduction in the risk of death compared to other primary sites (HR = 0.57, p = 0.007), the number of extra-cerebral metastatic sites (HR = 1.26, p = 0.003) and planning target volumes (HR = 1.15, p = 0.012) were associated with a lower OS. There was no prognostic factor of time to local progression. Median OS was 15.2 months [95% CI, 12.0-17.9 months] and the OS rate at 1 year was 58% [95% CI, 50-65%]. Salvage radiotherapy was administered to 72 patients (45%), of which 49 received new HFSRT. Ten (7%) patients presented late grade 2 and 4 (3%) patients late grade 3 toxicities. Thirteen (8.9%) patients developed radiation necrosis.
   Conclusions: This large multicenter retrospective study shows that HFSRT allows for good local control of metastasectomy tumor beds and that this technique is well-tolerated by patients.
C1 [Martinage, Geoffrey; Mouttet-Audouard, Raphaelle; Basson, Laurent; Mirabel, Xavier; Lartigau, Eric; Pasquier, David] Univ Lille II, Acad Dept Radiat Oncol, Ctr Oscar Lambret, Lille, France.
   [Geffrelot, Julien; Stefan, Dinu] Ctr Francois Baclesse, Dept Radiat Oncol, Caen, France.
   [Bogart, Emilie] Ctr Oscar Lambret, Dept Biostat, Lille, France.
   [Rault, Erwan] Ctr Oscar Lambret, Dept Med Phys, Lille, France.
   [Reyns, Nicolas] CHRU Lille, Dept Neurosurg, Lille, France.
   [Emery, Evelyne] Univ Hosp Caen, Neurosurg Dept, Caen, France.
   [Makhlouff-Martinage, Samira] Ctr Oscar Lambret, Dept Clin Res & Innovat, Lille, France.
   [Lartigau, Eric; Pasquier, David] Lille Univ, CRIStAL UMR CNRS 9189, Lille, France.
C3 UNICANCER; Centre Oscar Lambret; Universite de Lille - ISITE; Universite
   de Lille; UNICANCER; Centre Francois Baclesse; UNICANCER; Centre Oscar
   Lambret; UNICANCER; Centre Oscar Lambret; Universite de Lille - ISITE;
   CHU Lille; CHU de Caen NORMANDIE; Universite de Caen Normandie;
   UNICANCER; Centre Oscar Lambret; Universite de Lille - ISITE; Universite
   de Lille
RP Pasquier, D (通讯作者)，Univ Lille II, Acad Dept Radiat Oncol, Ctr Oscar Lambret, Lille, France.; Pasquier, D (通讯作者)，Lille Univ, CRIStAL UMR CNRS 9189, Lille, France.
EM
RI PASQUIER, David/O-8004-2014; lartigau, eric/GSI-5327-2022
OI PASQUIER, David/0000-0001-6019-7309; Rault, Erwann/0000-0003-0595-9301
CR Ahmed KA, 2014, J NEURO-ONCOL, V118, P179, DOI 10.1007/s11060-014-1417-2
   Al-Omair A, 2013, TECHNOL CANCER RES T, V12, P493, DOI 10.7785/tcrt.2012.500336
   Alexander BM, 2018, LANCET ONCOL, V19, pE33, DOI 10.1016/S1470-2045(17)30692-7
   Atalar B, 2013, INT J RADIAT ONCOL, V87, P713, DOI 10.1016/j.ijrobp.2013.07.034
   Barnholtz-Sloan JS, 2012, NEURO-ONCOLOGY, V14, P910, DOI 10.1093/neuonc/nos087
   Borghaei H, 2015, NEW ENGL J MED, V373, P1627, DOI 10.1056/NEJMoa1507643
   Brown PD, 2017, LANCET ONCOL, V18, P1049, DOI 10.1016/S1470-2045(17)30441-2
   Chang EL, 2009, LANCET ONCOL, V10, P1037, DOI 10.1016/S1470-2045(09)70263-3
   Eaton BR, 2015, J NEURO-ONCOL, V123, P103, DOI 10.1007/s11060-015-1767-4
   Eisenhauer EA, 2009, EUR J CANCER, V45, P228, DOI 10.1016/j.ejca.2008.10.026
   Gans JH, 2013, NEUROSURGERY, V72, P317, DOI 10.1227/NEU.0b013e31827fcd60
   Jarvis LA, 2012, INT J RADIAT ONCOL, V84, P943, DOI 10.1016/j.ijrobp.2012.01.067
   Keller A, 2017, INT J RADIAT ONCOL, V99, P1179, DOI 10.1016/j.ijrobp.2017.08.014
   Kocher M, 2011, J CLIN ONCOL, V29, P134, DOI 10.1200/JCO.2010.30.1655
   Ling DC, 2015, NEUROSURGERY, V76, P150, DOI 10.1227/NEU.0000000000000584
   Mahajan A, 2017, LANCET ONCOL, V18, P1040, DOI 10.1016/S1470-2045(17)30414-X
   Marchan EM, 2018, FRONT ONCOL, V8, DOI 10.3389/fonc.2018.00342
   Minniti G, 2016, INT J RADIAT ONCOL, V95, P1142, DOI 10.1016/j.ijrobp.2016.03.013
   Minniti G, 2013, INT J RADIAT ONCOL, V86, P623, DOI 10.1016/j.ijrobp.2013.03.037
   Ogiwara H, 2012, J NEURO-ONCOL, V108, P141, DOI 10.1007/s11060-012-0808-5
   Patchell RA, 2003, CANCER TREAT REV, V29, P533, DOI 10.1016/S0305-7372(03)00105-1
   PATCHELL RA, 1990, NEW ENGL J MED, V322, P494, DOI 10.1056/NEJM199002223220802
   Patchell RA, 1998, JAMA-J AM MED ASSOC, V280, P1485, DOI 10.1001/jama.280.17.1485
   Patel AJ, 2010, J NEUROSURG, V113, P181, DOI 10.3171/2009.11.JNS09659
   Pessina F, 2016, PLOS ONE, V11, DOI 10.1371/journal.pone.0157869
   Prabhu R, 2012, INT J RADIAT ONCOL, V83, pE61, DOI 10.1016/j.ijrobp.2011.12.017
   Robbins JR, 2012, NEUROSURGERY, V71, P937, DOI 10.1227/NEU.0b013e31826909f2
   Royston P, 2011, STAT MED, V30, P2409, DOI 10.1002/sim.4274
   Shaw E, 2000, INT J RADIAT ONCOL, V47, P291, DOI 10.1016/S0360-3016(99)00507-6
   Soffietti R, 2013, J CLIN ONCOL, V31, P65, DOI 10.1200/JCO.2011.41.0639
   Soliman H, 2018, INT J RADIAT ONCOL, V100, P436, DOI 10.1016/j.ijrobp.2017.09.047
   VECHT CJ, 1993, ANN NEUROL, V33, P583, DOI 10.1002/ana.410330605
   Wang CC, 2012, J NEURO-ONCOL, V106, P601, DOI 10.1007/s11060-011-0697-z
NR 33
TC 9
Z9 9
U1 0
U2 2
PU FRONTIERS MEDIA SA
PI LAUSANNE
PA AVENUE DU TRIBUNAL FEDERAL 34, LAUSANNE, CH-1015, SWITZERLAND
SN 2234-943X
J9 FRONT ONCOL
JI Front. Oncol.
PD MAR 28
PY 2019
VL 9
AR 184
DI 10.3389/fonc.2019.00184
PG 9
WC Oncology
WE Science Citation Index Expanded (SCI-EXPANDED)
SC Oncology
GA HQ8VM
UT WOS:000462704700002
PM 30984617
OA Green Published, gold
DA 2023-08-27
ER

PT J
AU Keramaris, E
   Ruzhynsky, VA
   Callaghan, SM
   Wong, E
   Davis, RJ
   Flavell, R
   Slack, RS
   Park, DS
AF Keramaris, Elizabeth
   Ruzhynsky, Vladamir A.
   Callaghan, Steve M.
   Wong, Estelle
   Davis, Roger J.
   Flavell, Richard
   Slack, Ruth S.
   Park, David S.
TI Required roles of Bax and JNKs in central and peripheral nervous system
   death of retinoblastoma-deficient mice
SO JOURNAL OF BIOLOGICAL CHEMISTRY
LA English
DT Article
ID N-TERMINAL PHOSPHORYLATION; ACTIVATED PROTEIN-KINASES; CYCLIN-DEPENDENT
   KINASES; JUN NH2-TERMINAL KINASE; C-JUN; CELL-DEATH;
   SIGNAL-TRANSDUCTION; PARKINSONS-DISEASE; CORTICAL-NEURONS;
   TUMOR-SUPPRESSOR
AB Retinoblastoma-deficient mice show massive neuronal damage and deficits in both CNS and PNS tissue. Previous work in the field has shown that death is regulated through distinct processes where CNS tissue undergoes death regulated by the tumor suppressor p53 and the apoptosome component, APAF1. Death in the PNS, however, is independent of p53 and reliant on the death protease, caspase 3. In the present study, we more carefully delineated the common and distinct mechanisms of death regulation by examining the stress-activated kinases, JNK2 and 3, the conserved Bcl-2 member Bax, and the relationship among these elements including p53. By use of genetic modeling, we show that death in various regions of the CNS and DRGs of the PNS is reliant on Bax. In the CNS, Bax acts downstream of p53. The relevance of the JNKs is more complex, however. Surprisingly, JNK3 deficiency by itself does not inhibit c-Jun phosphorylation and instead, aggravates death in both CNS and PNS tissue. However, JNK2/3 double deficiency blocks death due to Rb loss in both the PNS and CNS. Importantly, the relationships between JNKs, p53, and Bax exhibit regional differences. In the medulla region of the hindbrain in the CNS, JNK2/3 deficiency blocks p53 activation. Moreover, Bax deficiency does not affect c-Jun phosphorylation. This indicates that a JNK- p53-Bax pathway is central in the hindbrain. However, in the diencephalon regions of the forebrain (thalamus), Bax deficiency blocks c-Jun activation, indicating that a Bax-JNK pathway of death is more relevant. In the DRGs of the PNS, a third pathway is present. In this case, a JNK- Bax pathway, independent of p53, regulates damage. Accordingly, our results show that a death regulator Bax is common to death in both PNS and CNS tissue. However, it is regulated by or itself regulates different effectors including the JNKs and p53 depending upon the specific region of the nervous system.
C1 [Keramaris, Elizabeth; Ruzhynsky, Vladamir A.; Callaghan, Steve M.; Wong, Estelle; Slack, Ruth S.; Park, David S.] Univ Ottawa, Ottawa Hlth Res Inst, Dept Cellular Mol Med Neurosci E, Ottawa, ON K1H 8M5, Canada.
   [Davis, Roger J.] Univ Massachusetts, Sch Med, Howard Hughes Med Inst, Dept Biochem & Mol Biol, Worcester, MA 01605 USA.
   [Flavell, Richard] Yale Univ, Sch Med, Howard Hughes Med Inst, Immunobiol Sect, New Haven, CT 06520 USA.
C3 University of Ottawa; Ottawa Hospital Research Institute; Howard Hughes
   Medical Institute; University of Massachusetts System; University of
   Massachusetts Worcester; Howard Hughes Medical Institute; Yale
   University
RP Park, DS (通讯作者)，Univ Ottawa, Ottawa Hlth Res Inst, Dept Cellular Mol Med Neurosci E, Ottawa, ON K1H 8M5, Canada.
EM
OI Slack, Ruth/0000-0002-1552-2835; Davis, Roger/0000-0002-0130-1652; Park,
   David/0000-0002-4490-3784
CR Buschmann T, 2001, MOL CELL BIOL, V21, P2743, DOI 10.1128/MCB.21.8.2743-2754.2001
   CLARKE AR, 1992, NATURE, V359, P328, DOI 10.1038/359328a0
   Cregan SP, 2002, J CELL BIOL, V158, P507, DOI 10.1083/jcb.200202130
   Cregan SP, 1999, J NEUROSCI, V19, P7860
   Crocker SJ, 2001, P NATL ACAD SCI USA, V98, P13385, DOI 10.1073/pnas.231177098
   Davis RJ, 2000, CELL, V103, P239, DOI 10.1016/S0092-8674(00)00116-1
   Ferguson KL, 2002, EMBO J, V21, P3337, DOI 10.1093/emboj/cdf338
   Fuchs SY, 1998, P NATL ACAD SCI USA, V95, P10541, DOI 10.1073/pnas.95.18.10541
   Ghahremani MH, 2002, J BIOL CHEM, V277, P35586, DOI 10.1074/jbc.M204362200
   Giovanni A, 2000, J BIOL CHEM, V275, P11553, DOI 10.1074/jbc.275.16.11553
   Guo Z, 2001, CANCER RES, V61, P8395
   Harris CA, 2002, J NEUROSCI, V22, P103, DOI 10.1523/JNEUROSCI.22-01-00103.2002
   Hayley S, 2004, J NEUROSCI, V24, P2045, DOI 10.1523/JNEUROSCI.4564-03.2004
   Herdegen T, 2001, ONCOGENE, V20, P2424, DOI 10.1038/sj.onc.1204387
   Herdegen T, 1998, J NEUROSCI, V18, P5124, DOI 10.1523/JNEUROSCI.18-14-05124.1998
   Hunot S, 2004, P NATL ACAD SCI USA, V101, P665, DOI 10.1073/pnas.0307453101
   JACKS T, 1992, NATURE, V359, P295, DOI 10.1038/359295a0
   KARIN M, 1995, J BIOL CHEM, V270, P16483, DOI 10.1074/jbc.270.28.16483
   Karin M, 1997, CURR OPIN CELL BIOL, V9, P240, DOI 10.1016/S0955-0674(97)80068-3
   Kuan CY, 2003, P NATL ACAD SCI USA, V100, P15184, DOI 10.1073/pnas.2336254100
   LEE EYHP, 1992, NATURE, V359, P288, DOI 10.1038/359288a0
   Lei K, 2002, MOL CELL BIOL, V22, P4929, DOI 10.1128/MCB.22.13.4929-4942.2002
   Lei K, 2003, P NATL ACAD SCI USA, V100, P2432, DOI 10.1073/pnas.0438011100
   LIN AN, 1995, SCIENCE, V268, P286, DOI 10.1126/science.7716521
   Lipinski MM, 2001, EMBO J, V20, P3402, DOI 10.1093/emboj/20.13.3402
   Macleod KF, 1996, EMBO J, V15, P6178, DOI 10.1002/j.1460-2075.1996.tb01006.x
   MINDEN A, 1994, MOL CELL BIOL, V14, P6683, DOI 10.1128/MCB.14.10.6683
   Morris EJ, 2001, ADV CANCER RES, V82, P1, DOI 10.1016/S0065-230X(01)82001-7
   Morris EJ, 2001, J NEUROSCI, V21, P5017, DOI 10.1523/JNEUROSCI.21-14-05017.2001
   Mulligan G, 1998, TRENDS GENET, V14, P223, DOI 10.1016/S0168-9525(98)01470-X
   Oliva AA, 2006, J NEUROSCI, V26, P9462, DOI 10.1523/JNEUROSCI.2625-06.2006
   Pan HC, 1998, MOL CELL, V2, P283, DOI 10.1016/S1097-2765(00)80273-7
   Park DS, 1996, J BIOL CHEM, V271, P21898, DOI 10.1074/jbc.271.36.21898
   Putcha GV, 2003, NEURON, V38, P899, DOI 10.1016/S0896-6273(03)00355-6
   Raivich G, 2004, NEURON, V43, P57, DOI 10.1016/j.neuron.2004.06.005
   Simpson MTW, 2001, J NEUROSCI, V21, P7089, DOI 10.1523/JNEUROSCI.21-18-07089.2001
   Slack RS, 1996, DEV GENET, V18, P81, DOI 10.1002/(SICI)1520-6408(1996)18:1<81::AID-DVG9>3.0.CO;2-Y
   Tsai KY, 1998, MOL CELL, V2, P293, DOI 10.1016/S1097-2765(00)80274-9
   Whitfield J, 2001, NEURON, V29, P629, DOI 10.1016/S0896-6273(01)00239-2
   Ziebold U, 2001, GENE DEV, V15, P386, DOI 10.1101/gad.858801
NR 40
TC 9
Z9 10
U1 0
U2 1
PU AMER SOC BIOCHEMISTRY MOLECULAR BIOLOGY INC
PI BETHESDA
PA 9650 ROCKVILLE PIKE, BETHESDA, MD 20814-3996 USA
EI 1083-351X
J9 J BIOL CHEM
JI J. Biol. Chem.
PD JAN 4
PY 2008
VL 283
IS 1
BP 405
EP 415
DI 10.1074/jbc.M701552200
PG 11
WC Biochemistry & Molecular Biology
WE Science Citation Index Expanded (SCI-EXPANDED)
SC Biochemistry & Molecular Biology
GA 245MV
UT WOS:000251940300045
PM 17984095
OA hybrid
DA 2023-08-27
ER

PT J
AU La Rocca, G
   Sabatino, G
   Altieri, R
   Signorelli, F
   Ricciardi, L
   Gessi, M
   Della Pepa, GM
AF La Rocca, Giuseppe
   Sabatino, Giovanni
   Altieri, Roberto
   Signorelli, Francesco
   Ricciardi, Luca
   Gessi, Marco
   Della Pepa, Giuseppe Maria
TI Significance of H3K27M Mutation in "Nonmidline" High-Grade Gliomas of
   Cerebral Hemispheres
SO WORLD NEUROSURGERY
LA English
DT Article
DE Diffuse midline glioma; Glioblastoma; H3K27M-m; High-grade glioma;
   Histone mutation
ID K27M MUTATION; HISTONE H3.3
AB BACKGROUND: "Diffuse midline glioma H3K27M-mutant" (a mutation gene encoding H3 histone variants) represents a defined pathologic entity since the 2016 World Health Organization Central Nervous System classification and includes midline structures, namely the thalamus, brainstem, and spinal cord. H3K27M-mutant glioma responds more poorly and is associated with worse clinical outcome. Mutation detection is now diagnostic for "diffused midline gliomas." We report on a case arising in an atypical nonmidline localization with an exceedingly adverse course. The present case raises the question of the possible meaning of the H3K27M-mutation in high-grade gliomas arising from the cerebral hemispheres and its implications from prognostic and therapeutic views.
   CASE DESCRIPTION: A 48-year-old woman was admitted with a mass arising from the splenium of the corpus callosum with bilateral hemispheric extension, displaying magnetic resonance imaging characteristics typical of a high-grade glioma. Needle biopsy confirmed the diagnosis of "diffuse astrocytic neoplasm, World Health Organization grade IV." Immunostaining of the tumor revealed the H3K27M-mutant nuclear expression. The patient was referred to adjuvant therapy and died 3 months after biopsy.
   CONCLUSIONS: This experience highlights the possible negative prognostic impact of H3K27M-m and possibility that H3K27M-m could be a reliable prognostic indicator to be considered in not only diffuse midline gliomas but also gliomas from the cerebral hemispheres. Because of the rarity and limited knowledge of this specific mutation in this experience, reports should be encouraged.
C1 [La Rocca, Giuseppe; Sabatino, Giovanni; Signorelli, Francesco; Ricciardi, Luca; Della Pepa, Giuseppe Maria] Univ Cattolica Sacro Cuore, Inst Neurosurg, Rome, Italy.
   [Altieri, Roberto] Univ Turin, Inst Neurosurg, Turin, Italy.
   [Gessi, Marco] Univ Cattolica Sacro Cuore, Inst Neuropathol, Rome, Italy.
C3 Catholic University of the Sacred Heart; IRCCS Policlinico Gemelli;
   University of Turin; Catholic University of the Sacred Heart; IRCCS
   Policlinico Gemelli
RP Della Pepa, GM (通讯作者)，Univ Cattolica Sacro Cuore, Inst Neurosurg, Rome, Italy.
EM
RI la rocca, giuseppe/AAG-3922-2019; Gessi, Marco/K-2572-2018; Signorelli,
   Francesco/Y-9746-2019; Altieri, Roberto/AAH-9818-2021; Ricciardi,
   Luca/L-5405-2019; Della Pepa, Giuseppe Maria/AAC-1697-2019
OI la rocca, giuseppe/0000-0003-1830-4849; Ricciardi,
   Luca/0000-0001-5110-8386; Della Pepa, Giuseppe
   Maria/0000-0001-8698-3359; SABATINO, Giovanni/0000-0002-4227-0434;
   Altieri, Roberto/0000-0003-2671-4944
CR Khuong-Quang DA, 2012, ACTA NEUROPATHOL, V124, P439, DOI 10.1007/s00401-012-0998-0
   Feng J, 2015, HUM PATHOL, V46, P1626, DOI 10.1016/j.humpath.2015.07.002
   Lopez G, 2017, ACTA NEUROPATHOL COM, V5, DOI 10.1186/s40478-017-0440-x
   Pratt D, 2018, ACTA NEUROPATHOL, V135, P299, DOI 10.1007/s00401-018-1805-3
   Schwartzentruber J, 2012, NATURE, V482, P226, DOI [10.1038/nature10833, 10.1007/s13311-012-0137-6]
NR 5
TC 8
Z9 8
U1 0
U2 6
PU ELSEVIER SCIENCE INC
PI NEW YORK
PA STE 800, 230 PARK AVE, NEW YORK, NY 10169 USA
SN 1878-8750
EI 1878-8769
J9 WORLD NEUROSURG
JI World Neurosurg.
PD NOV
PY 2019
VL 131
BP 174
EP 176
DI 10.1016/j.wneu.2019.08.024
PG 3
WC Clinical Neurology; Surgery
WE Science Citation Index Expanded (SCI-EXPANDED)
SC Neurosciences & Neurology; Surgery
GA JG3GV
UT WOS:000491963300115
PM 31415896
DA 2023-08-27
ER

PT J
AU Mirza, R
   Sharma, B
AF Mirza, Roohi
   Sharma, Bhupesh
TI Benefits of Fenofibrate in prenatal valproic acid-induced autism
   spectrum disorder related phenotype in rats
SO BRAIN RESEARCH BULLETIN
LA English
DT Article
DE Autism spectrum disorder; Valproic acid; PPAR-alpha; Fenofibrate;
   Neuroinflammation; Oxidative stress
ID ACTIVATED RECEPTOR-ALPHA; INCREASED OXIDATIVE STRESS; BRAIN-BARRIER
   IMPAIRMENTS; INDUCED ANIMAL-MODEL; PPAR-ALPHA; BEHAVIORAL DEFICITS;
   AGONIST FENOFIBRATE; NEUROTROPHIC FACTOR; DOPAMINE NEURONS; TNF-ALPHA
AB Autism Spectrum Disorder (ASD) is a neurodevelopmental disorder with two major behavioral symptoms i.e. repetitive behavior and social-communication impairment. The unknown etiology of ASD is responsible for the difficulty in identifying the possible therapeutic modulators for ASD. Valproic acid (VPA) is an anticonvulsant drug in both human and rodents with teratogenic effects during pregnancy. Therefore, prenatal exposure of VPA induced autism spectrum disorder like phenotypes in both human and rodents. Peroxisome proliferator-activated receptor-alpha (PPAR-alpha) is widely localized in the brain. This research investigates the utility of fenofibrate, a selective agonist of PPAR-alpha in prenatal VPA-induced experimental ASD in Wistar rats. The prenatal VPA has induced social impairment (three chambers social behavior apparatus), repetitive behavior (Y-maze), hyperlocomotion (actophotometer), anxiety (elevated plus maze) and low exploratory activity (hole board test). Also, prenatal VPA treated rats have shown higher levels of oxidative stress (increased in thiobarbituric acid reactive species and decreased in reduced glutathione level) and inflammation (increased in interleukin-6, tumor necrosis factor-a and decreased in interleukin-10) in the cerebellum, brainstem and prefrontal cortex. Treatment with fenofibrate significantly attenuated prenatal VPA-induced social impairment, repetitive behavior, hyperactivity, anxiety, and low exploratory activity. Furthermore, fenofibrate also decreased the prenatal VPA-induced oxidative stress and inflammation in brain regions. Hence, it may be concluded that fenofibrate may provide neurobehavioral and biochemical benefits in prenatal VPA-induced autism phenotypes in rats.
C1 [Mirza, Roohi; Sharma, Bhupesh] Amity Univ Uttar Pradesh, Amity Inst Pharm, Dept Pharmacol, Sect 125, Noida 201313, Uttar Pradesh, India.
   [Sharma, Bhupesh] Consci Res, CNS Pharmacol, Delhi, India.
C3 Amity University Noida
RP Sharma, B (通讯作者)，Amity Univ Uttar Pradesh, Amity Inst Pharm, Dept Pharmacol, Sect 125, Noida 201313, Uttar Pradesh, India.
EM
RI MIRZA, ROOHI/IUQ-5297-2023; Sharma, Bhupesh/D-3037-2014
OI Sharma, Bhupesh/0000-0002-3423-007X; MIRZA, ROOHI/0000-0001-5570-6420
CR Al-Amin MM, 2015, BEHAV BRAIN RES, V286, P112, DOI 10.1016/j.bbr.2015.02.041
   American Psychiatric Association, 1987, DIAGN STAT MAN MENT, V3rd Ed
   Bhateja DK, 2012, EUR J PHARMACOL, V674, P33, DOI 10.1016/j.ejphar.2011.10.029
   Bruchhage M. M., 2017, FRONT CELL NEUROSCI, DOI [10.3389/conf.fnce1.2017.37, DOI 10.3389/CONF.FNCE1.2017.37]
   Chen J, 2013, NEUROSCI LETT, V541, P275, DOI 10.1016/j.neulet.2013.02.005
   Chen XR, 2007, J NEUROTRAUM, V24, P1119, DOI 10.1089/neu.2006.0216
   Cheng R, 2016, DIABETES, V65, P3730, DOI 10.2337/db16-0426
   Cheng YH, 2015, NEUROTOX RES, V27, P55, DOI 10.1007/s12640-014-9485-9
   Chikahisa S, 2014, NEUROPHARMACOLOGY, V79, P399, DOI 10.1016/j.neuropharm.2013.12.009
   Codagnone MG, 2015, DEV NEUROSCI-BASEL, V37, P215, DOI 10.1159/000375489
   Cullingford TE, 1998, J NEUROCHEM, V70, P1366
   D'Agostino G, 2015, MOL METAB, V4, P528, DOI 10.1016/j.molmet.2015.04.005
   Delerive P, 1999, J BIOL CHEM, V274, P32048, DOI 10.1074/jbc.274.45.32048
   Dumont M, 2012, HUM MOL GENET, V21, P5091, DOI 10.1093/hmg/dds355
   Eissa N, 2018, SCI REP-UK, V8, DOI 10.1038/s41598-018-31385-7
   Ergaz Z, 2016, REPROD TOXICOL, V64, P116, DOI 10.1016/j.reprotox.2016.04.024
   Frye Richard E, 2013, Autism Res Treat, V2013, P609705, DOI 10.1155/2013/609705
   Geva R, 2013, SOC COGN AFFECT NEUR, V8, P158, DOI 10.1093/scan/nsr082
   Gottfried C., 2013, RECENT ADV AUTISM SP, DOI [10.5772/54824, DOI 10.5772/54824]
   Grover S, 2013, PHARMACOL BIOCHEM BE, V111, P17, DOI 10.1016/j.pbb.2013.08.001
   Guldenpfennig M, 2011, PHYSIOL BEHAV, V103, P404, DOI 10.1016/j.physbeh.2011.03.008
   Gvozdjakova A, 2014, OXID MED CELL LONGEV, V2014, DOI 10.1155/2014/798957
   Ha S, 2017, BEHAV BRAIN RES, V317, P479, DOI 10.1016/j.bbr.2016.10.004
   Hanna BG, 2001, DEV MED CHILD NEUROL, V43, P847, DOI 10.1017/S0012162201221549
   Hegazy HG, 2015, CYTOKINE, V71, P173, DOI 10.1016/j.cyto.2014.10.027
   Hofvander B, 2009, BMC PSYCHIATRY, V9, DOI 10.1186/1471-244X-9-35
   Hong H, 2016, INT NEUROUROL J, V20, pS2, DOI 10.5213/inj.1632604.302
   Huang W, 2014, J CEREBR BLOOD F MET, V34, P646, DOI 10.1038/jcbfm.2013.240
   James SJ, 2004, AM J CLIN NUTR, V80, P1611, DOI 10.1093/ajcn/80.6.1611
   Jiang B, 2017, BRIT J PHARMACOL, V174, P177, DOI 10.1111/bph.13668
   Jin Yan, 2015, Mach Learn Med Imaging, V9352, P170, DOI 10.1007/978-3-319-24888-2_21
   Kapadia R, 2008, FRONT BIOSCI-LANDMRK, V13, P1813, DOI 10.2741/2802
   Kerr DM, 2016, PHARMACOL RES, V113, P228, DOI 10.1016/j.phrs.2016.08.033
   Kim JW, 2017, NEUROPHARMACOLOGY, V113, P71, DOI 10.1016/j.neuropharm.2016.09.014
   Kim KC, 2014, MOL NEUROBIOL, V49, P512, DOI 10.1007/s12035-013-8535-2
   Kraeuter AK, 2018, METHODS MOL BIOL, V1735, P403, DOI 10.1007/978-1-4939-7614-0_28
   Kumar H, 2016, BRAIN RES BULL, V124, P27, DOI 10.1016/j.brainresbull.2016.03.013
   Kumar H, 2016, BRAIN RES, V1630, P83, DOI 10.1016/j.brainres.2015.10.052
   Kumar H, 2015, NEUROCHEM INT, V91, P34, DOI 10.1016/j.neuint.2015.10.007
   Lauber E, 2016, FRONT MOL NEUROSCI, V9, DOI 10.3389/fnmol.2016.00150
   Li K, 2018, TOXICOL SCI, V162, P189, DOI 10.1093/toxsci/kfx240
   Li SY, 2013, AM J PHYSIOL-RENAL, V305, pF618, DOI 10.1152/ajprenal.00309.2013
   LOWRY OH, 1951, J BIOL CHEM, V193, P265
   Markram K, 2008, NEUROPSYCHOPHARMACOL, V33, P901, DOI 10.1038/sj.npp.1301453
   Melis M, 2013, J NEUROSCI, V33, P6203, DOI 10.1523/JNEUROSCI.4647-12.2013
   Melis M, 2008, J NEUROSCI, V28, P13985, DOI 10.1523/JNEUROSCI.3221-08.2008
   Merali Z, 2014, BEHAV BRAIN RES, V263, P60, DOI 10.1016/j.bbr.2014.01.008
   Mijangos-Moreno S, 2016, NEUROSCI RES, V105, P65, DOI 10.1016/j.neures.2015.09.005
   Mirza R, 2018, LIFE SCI, V214, P106, DOI 10.1016/j.lfs.2018.10.045
   Morakotsriwan N, 2016, OXID MED CELL LONGEV, V2016, DOI 10.1155/2016/3206561
   Neumeyer A. M., 2018, ACADPEDIATR, VS1876-2859, DOI [10.1016/j.acap.2018.06.014, DOI 10.1016/J.ACAP.2018.06.014.30456-X]
   OHKAWA H, 1979, ANAL BIOCHEM, V95, P351, DOI 10.1016/0003-2697(79)90738-3
   Olexova L, 2016, NEUROSCI LETT, V629, P9, DOI 10.1016/j.neulet.2016.06.035
   Ouk T, 2014, J CEREBR BLOOD F MET, V34, P542, DOI 10.1038/jcbfm.2013.233
   Paintlia MK, 2008, EXP NEUROL, V210, P560, DOI 10.1016/j.expneurol.2007.12.011
   Peca J, 2011, NATURE, V472, P437, DOI 10.1038/nature09965
   Pickering M, 2005, EXP PHYSIOL, V90, P663, DOI 10.1113/expphysiol.2005.030734
   Pillion JP, 2018, COGN BEHAV NEUROL, V31, P53, DOI 10.1097/WNN.0000000000000154
   Ramanan S, 2009, INT J RADIAT ONCOL, V75, P870, DOI 10.1016/j.ijrobp.2009.06.059
   Raso GM, 2011, J NEUROENDOCRINOL, V23, P591, DOI 10.1111/j.1365-2826.2011.02152.x
   Rodier PM, 1997, REPROD TOXICOL, V11, P417, DOI 10.1016/S0890-6238(97)80001-U
   Rodier PM, 1996, J COMP NEUROL, V370, P247, DOI 10.1002/(SICI)1096-9861(19960624)370:2<247::AID-CNE8>3.0.CO;2-2
   Rodriguez Juan I, 2011, Neuron Glia Biol, V7, P205, DOI 10.1017/S1740925X12000142
   Rose S, 2012, TRANSL PSYCHIAT, V2, DOI 10.1038/tp.2012.61
   Ross HE, 2013, BRAIN BEHAV IMMUN, V31, P76, DOI 10.1016/j.bbi.2012.12.021
   Roullet FI, 2013, NEUROTOXICOL TERATOL, V36, P47, DOI 10.1016/j.ntt.2013.01.004
   Salman MS, 2016, NEUROIMAG CLIN N AM, V26, P317, DOI 10.1016/j.nic.2016.03.003
   Sandhya T, 2012, NEUROCHEM RES, V37, P1121, DOI 10.1007/s11064-012-0717-1
   Sasso O, 2010, EUR NEUROPSYCHOPHARM, V20, P195, DOI 10.1016/j.euroneuro.2009.09.003
   Schneider T, 2005, NEUROPSYCHOPHARMACOL, V30, P80, DOI 10.1038/sj.npp.1300518
   Sehgal N, 2012, PLOS ONE, V7, DOI 10.1371/journal.pone.0035427
   Sengupta P, 2013, INT J PREVENTIVE MED, V4, P624
   Servadio M, 2018, EUR NEUROPSYCHOPHARM, V28, P85, DOI 10.1016/j.euroneuro.2017.11.014
   South M, 2017, FRONT HUM NEUROSCI, V11, DOI 10.3389/fnhum.2017.00020
   Stanton ME, 2007, BEHAV BRAIN RES, V176, P133, DOI 10.1016/j.bbr.2006.10.022
   Uppalapati D, 2014, PPAR RES, V2014, DOI 10.1155/2014/753587
   van Steensel FJA, 2011, CLIN CHILD FAM PSYCH, V14, P302, DOI 10.1007/s10567-011-0097-0
   Vitale C, 2004, EUR J IMMUNOL, V34, P3028, DOI 10.1002/eji.200425418
   Wang GM, 2011, NEUROSCI RES, V70, P230, DOI 10.1016/j.neures.2011.02.001
   Wei H, 2013, NEUROSCIENCE, V252, P320, DOI 10.1016/j.neuroscience.2013.08.025
   Wei HG, 2012, BBA-MOL BASIS DIS, V1822, P831, DOI 10.1016/j.bbadis.2012.01.011
   Wohr M, 2015, TRANSL PSYCHIAT, V5, DOI 10.1038/tp.2015.19
   Wu HF, 2017, MOL NEUROBIOL, V54, P5264, DOI 10.1007/s12035-016-0074-1
   Xie J, 2017, ONCOTARGET, V8, P82390, DOI 10.18632/oncotarget.19326
   Xu NG, 2015, MEDIAT INFLAMM, V2015, DOI 10.1155/2015/531518
   Yoshizawa K, 2017, SYNAPSE, V71, DOI 10.1002/syn.21978
   Zhang J, 2017, NEUROPSYCH DIS TREAT, V13, P835, DOI 10.2147/NDT.S125088
   Zhang YH, 2012, NEUROCHEM RES, V37, P1409, DOI 10.1007/s11064-012-0724-2
   Zhou H, 2017, 2016 INTERNATIONAL CONFERENCE ON INTELLIGENT TRANSPORTATION, BIG DATA & SMART CITY (ICITBS), P1, DOI 10.1109/ICITBS.2016.27
   Zoroglu SS, 2004, EUR ARCH PSY CLIN N, V254, P143, DOI 10.1007/s00406-004-0456-7
NR 90
TC 44
Z9 45
U1 2
U2 14
PU PERGAMON-ELSEVIER SCIENCE LTD
PI OXFORD
PA THE BOULEVARD, LANGFORD LANE, KIDLINGTON, OXFORD OX5 1GB, ENGLAND
SN 0361-9230
EI 1873-2747
J9 BRAIN RES BULL
JI Brain Res. Bull.
PD APR
PY 2019
VL 147
BP 36
EP 46
DI 10.1016/j.brainresbull.2019.02.003
PG 11
WC Neurosciences
WE Science Citation Index Expanded (SCI-EXPANDED)
SC Neurosciences & Neurology
GA HR0EW
UT WOS:000462802100004
PM 30769127
DA 2023-08-27
ER

PT J
AU Xiao, LM
   Xie, SH
   Tang, B
   Wu, X
   Ding, H
   Bao, YY
   Hong, T
AF Xiao, Limin
   Xie, Shenhao
   Tang, Bin
   Wu, Xiao
   Ding, Han
   Bao, Youyuan
   Hong, Tao
TI A novel technique to manage internal carotid artery injury in endoscopic
   endonasal skull base surgery in the premise of proximal and distal
   controls
SO NEUROSURGICAL REVIEW
LA English
DT Article
DE Endonasal endoscopic approach; Interdural; Internal carotid artery
   injury; Proximal and distal control; Interdural; Parasellar; Paraclival;
   Skull base
AB Intraoperative internal carotid artery injury is one of the most daunting complications in endoscopic skull base surgery. This paper proposed a novel technique to manage ICA injury after proximal and distal controls. The appropriate block sites together with the proximal and distal controls of ICA were demonstrated in six injected cadaveric specimens. The surgical outcomes of five patients with intraoperative ICA injury and managed with this concept were retrospectively reviewed. Five block sites for vascular control could be identified in all six specimens, including (1) distal to the distal dural ring, (2) proximal to the proximal dural ring, (3) anterior genu of the parasellar ICA, (4) the upper third of the paraclival ICA, and (5) just above the foramen lacerum. Both proximal and distal controls of ICA were achieved by using the block sites in combination. Gross tumor resection was achieved in all five cases after the intraoperative ICA injury was successfully managed. Three coping techniques were used, including direct coagulation to seal (three cases), endoscopic suture (one case), and coagulation to sacrifice (one case). Focal brainstem infarction occurred in one case, one patient died of intracranial infection, and the other three cases had no sequelae. No pseudoaneurysm occurred in all patients. Except the sacrificed ICA, the other ICA was intact during follow-up. It is technically feasible to manage ICA injuries after proximal and distal controls during EEA surgeries. The surgical outcomes from our case series supported the use of this novel technique.
C1 [Xiao, Limin; Xie, Shenhao; Tang, Bin; Wu, Xiao; Ding, Han; Bao, Youyuan; Hong, Tao] Nanchang Univ, Affiliated Hosp 1, Dept Neurosurg, Nanchang 330006, Jiangxi, Peoples R China.
C3 Nanchang University
RP Hong, T (通讯作者)，Nanchang Univ, Affiliated Hosp 1, Dept Neurosurg, Nanchang 330006, Jiangxi, Peoples R China.
EM
FU Natural Science Foundation of Jiangxi Science and Technology Department
   [20202BABL216057]
FX This study was funded by the Natural Science Foundation of Jiangxi
   Science and Technology Department (20202BABL216057).
CR Banerji D, 2001, NEUROL INDIA, V49, P253
   Cebula N, 2014, WORLD NEUROSURG, V82, DOI 10.1016/j.wneu.2014.09.021
   Chin OY, 2016, LARYNGOSCOPE, V126, P582, DOI 10.1002/lary.25748
   Duek I, 2017, J NEUROL SURG REP, V78, pe125, DOI 10.1055/s-0037-1608635
   Dusick JR, 2007, NEUROSURGERY, V60, P322, DOI 10.1227/01.NEU.0000255408.84269.A8
   Froelich S, 2011, NEUROSURGERY, V68, P310, DOI 10.1227/NEU.0b013e3182117063
   Gardner PA, 2016, OTOLARYNG CLIN N AM, V49, P819, DOI 10.1016/j.otc.2016.03.003
   Gardner PA, 2015, WORLD NEUROSURG, V84, P1380, DOI 10.1016/j.wneu.2015.06.032
   Gardner PA, 2013, NEUROSURGERY, V73, P261, DOI 10.1227/01.neu.0000430821.71267.f2
   Giorgianni A, 2019, WORLD NEUROSURG, V122, P376, DOI 10.1016/j.wneu.2018.10.234
   Gondim JA, 2011, PITUITARY, V14, P174, DOI 10.1007/s11102-010-0280-1
   Kassam Amin, 2005, Neurosurg Focus, V19, pE7
   Kassam AB, 2007, J NEUROSURG, V107, P1047, DOI 10.3171/JNS-07/11/1047
   Kassam AB, 2006, NEUROSURGERY, V59, P162
   Labib MA, 2014, OPER NEUROSURG, V10, P448, DOI 10.1227/NEU.0000000000000362
   LEONETTI JP, 1990, OTOLARYNG HEAD NECK, V103, P46, DOI 10.1177/019459989010300107
   LINSKEY ME, 1994, AM J NEURORADIOL, V15, P829
   Meybodi AT, 2019, J NEUROSURG, V131, P1734, DOI 10.3171/2018.6.JNS18800
   Muto J, 2017, LARYNGOSCOPE, V127, P38, DOI 10.1002/lary.26181
   SEKHAR LN, 1986, NEUROSURGERY, V19, P967, DOI 10.1227/00006123-198612000-00012
   Somanna S, 2015, ACTA NEUROCHIR, V157, P2077, DOI 10.1007/s00701-015-2610-6
   Szentirmai O, 2016, J NEUROSURG, V124, P463, DOI 10.3171/2015.1.JNS142650
   TARR RW, 1991, SKULL BASE SURG, V1, P240, DOI 10.1055/s-2008-1057104
   Valentine R, 2011, INT FORUM ALLERGY RH, V1, P118, DOI 10.1002/alr.20033
   Valentine R, 2011, LARYNGOSCOPE, V121, P562, DOI 10.1002/lary.21361
   Xiao LM, 2018, WORLD NEUROSURG, V115, pE33, DOI 10.1016/j.wneu.2018.03.093
NR 26
TC 4
Z9 4
U1 0
U2 6
PU SPRINGER
PI NEW YORK
PA ONE NEW YORK PLAZA, SUITE 4600, NEW YORK, NY, UNITED STATES
SN 0344-5607
EI 1437-2320
J9 NEUROSURG REV
JI Neurosurg. Rev.
PD DEC
PY 2021
VL 44
IS 6
BP 3437
EP 3445
DI 10.1007/s10143-021-01517-1
EA MAR 2021
PG 9
WC Clinical Neurology; Surgery
WE Science Citation Index Expanded (SCI-EXPANDED)
SC Neurosciences & Neurology; Surgery
GA WX9UP
UT WOS:000630284200001
PM 33738637
DA 2023-08-27
ER

PT J
AU Amaral, GF
   Dossa, PD
   Viebig, LB
   Konno, FTC
   Consoli, A
   Martins, MDM
   Viani, FC
   Bondan, EF
AF Amaral, Gisele Ferreira
   Dossa, Pietro Domingues
   Viebig, Ligia Bocamino
   Camargo Konno, Fabiana Toshie
   Consoli, Amanda
   Monteiro Martins, Maria de Fatima
   Viani, Flavio Cesar
   Bondan, Eduardo Fernandes
TI Astrocytic expression of GFAP and serum levels of IL-1 beta and
   TNF-alpha in rats treated with different pain relievers
SO BRAZILIAN JOURNAL OF PHARMACEUTICAL SCIENCES
LA English
DT Article
DE Neuropathic pain/treatment; Astrocytes/pain regulation;
   Analgesics/tests/treatment; Cytokines; Glial Fibrillary Acidic
   Protein/study
ID NUCLEUS-ACCUMBENS DOPAMINE; MORPHINE-TOLERANCE; MAJOR DEPRESSION;
   NEUROPATHIC PAIN; CLINICAL PAIN; GABAPENTIN; CYTOKINES; METHADONE;
   COCAINE; AMITRIPTYLINE
AB Pro-inflammatory cytokines and glial cells, especially microglial cells, have been implicated in persistent pain sensitization. Less is known about the role of astrocytes in pain regulation. This study aimed to observe the expression of the astrocytic biomarker glial fibrillary acidic protein (GFAP) and the serum levels of interleukin 1 beta (IL-1 beta) and tumor necrosis factor alpha (TNF-alpha) after short-term administration of central pain relievers in rats not submitted to noxious stimuli. Male Wistar rats were divided into five groups, receiving for nine days-(1) amitriptyline (Amt-10 mg/kg/day, by gavage); (2) gabapentin (Gb-60 mg/kg/day, by gavage; (3) methadone (Me-4.5 mg/kg/day,intraperitoneal route [IP]); (4) morphine (Mo-10 mg/kg/day, IP); or (5) 0.9% saline solution, IP. Brain samples were collected for immunohistochemical study of GFAP expression in the mesencephalon and nucleus accumbens (NAc). The area of GFAP-positive cells was calculated using MetaMorph (R). software and serum levels of IL-1 beta and TNF-alpha were measured by enzyme-linked immunosorbent assay. Serum TNF-alpha levels were decreased in the groups treated with Mo, Me and Gb, but not in the Amt-treated group. IL-1 beta decreased only in rats treated with Me. The astrocytic expression of GFAP was decreased in the brainstem with all drugs, while it was increased in the NAc with Amt, Me and Mo.
C1 [Amaral, Gisele Ferreira; Dossa, Pietro Domingues; Viebig, Ligia Bocamino; Camargo Konno, Fabiana Toshie; Monteiro Martins, Maria de Fatima; Bondan, Eduardo Fernandes] Univ Paulista, Dept Environm & Expt Pathol, Sao Paulo, SP, Brazil.
   [Consoli, Amanda] Univ Fed Sao Paulo, Dept Psychobiol, Sao Paulo, SP, Brazil.
   [Monteiro Martins, Maria de Fatima; Viani, Flavio Cesar; Bondan, Eduardo Fernandes] Univ Cruzeiro Sul, Dept Vet Med, Sao Paulo, SP, Brazil.
C3 Universidade Paulista; Universidade Federal de Sao Paulo (UNIFESP);
   Universidade Cruzeiro do Sul
RP Bondan, EF (通讯作者)，Univ Paulista, Dept Patol Ambiental & Expt, Rua Caconde 125-51, BR-01425011 Sao Paulo, SP, Brazil.
EM
RI Bondan, Eduardo F./D-5417-2016
OI Bondan, Eduardo F./0000-0002-3887-0023
CR Anisman H, 1999, BIOL PSYCHIAT, V46, P1649, DOI 10.1016/S0006-3223(99)00211-5
   Banks WA, 1995, NEUROIMMUNOMODULAT, V2, P241, DOI 10.1159/000097202
   Bao YH, 2014, NEUROREPORT, V25, P71, DOI 10.1097/WNR.0b013e328363fde8
   Bondan EF, 2013, ARQ BRAS ENDOCRINOL, V57, P431, DOI 10.1590/S0004-27302013000600004
   De Waal EJ, 1998, TOXICOLOGY, V129, P201, DOI 10.1016/S0300-483X(98)00077-8
   Di Matteo V, 2000, PSYCHOPHARMACOLOGY, V150, P45, DOI 10.1007/s002130000420
   DICHIARA G, 1988, J PHARMACOL EXP THER, V244, P1067
   Gerasimov MR, 2001, EUR J PHARMACOL, V414, P205, DOI 10.1016/S0014-2999(01)00800-7
   Gonzalez G, 2007, DRUG ALCOHOL DEPEN, V87, P1, DOI 10.1016/j.drugalcdep.2006.07.003
   Huang YN, 2012, BEHAV BRAIN RES, V229, P401, DOI 10.1016/j.bbr.2012.01.044
   Hutchinson Mark R, 2007, ScientificWorldJournal, V7, P98
   Hutchinson MR, 2004, INT IMMUNOPHARMACOL, V4, P1525, DOI 10.1016/j.intimp.2004.07.011
   Ji RR, 2013, PAIN, V154, pS10, DOI 10.1016/j.pain.2013.06.022
   John GR, 2005, GLIA, V49, P161, DOI 10.1002/glia.20109
   Kafami L, 2013, J NEUROIMMUNOL, V255, P39, DOI 10.1016/j.jneuroim.2012.10.015
   Karakaya S, 2007, J NEUROENDOCRINOL, V19, P682, DOI 10.1111/j.1365-2826.2007.01575.x
   Kenis G, 2002, INT J NEUROPSYCHOPH, V5, P401, DOI 10.1017/S1461145702003164
   Landmann R, 1997, BIOL PSYCHIAT, V41, P675, DOI 10.1016/S0006-3223(96)00176-X
   Lazriev IL, 2001, B EXP BIOL MED+, V131, P248, DOI 10.1023/A:1017699315355
   Lee BS, 2013, J KOREAN MED SCI, V28, P308, DOI 10.3346/jkms.2013.28.2.308
   Lynch Mary E, 2006, Pain Res Manag, V11, P11
   McMahon SB, 2005, EXP NEUROL, V192, P444, DOI 10.1016/j.expneurol.2004.11.001
   Middeldorp J, 2011, PROG NEUROBIOL, V93, P421, DOI 10.1016/j.pneurobio.2011.01.005
   Monibi FA, 2015, RES VET SCI, V99, P70, DOI 10.1016/j.rvsc.2015.01.010
   OLDS ME, 1982, BRAIN RES, V237, P429, DOI 10.1016/0006-8993(82)90454-1
   Ossipov MH, 2010, J CLIN INVEST, V120, P3779, DOI 10.1172/JCI43766
   Peng XQ, 2008, DRUG ALCOHOL DEPEN, V97, P207, DOI 10.1016/j.drugalcdep.2007.09.019
   Pontieri FE, 1995, P NATL ACAD SCI USA, V92, P12304, DOI 10.1073/pnas.92.26.12304
   Raghavendra V, 2004, EUR J NEUROSCI, V20, P467, DOI 10.1111/j.1460-9568.2004.03514.x
   Rossi AR, 2013, PLOS ONE, V8, DOI 10.1371/journal.pone.0078516
   Rittner HL, 2005, J LEUKOCYTE BIOL, V78, P1215, DOI 10.1189/jlb.0405223
   Rothermundt M, 2001, J AFFECT DISORDERS, V63, P93, DOI 10.1016/S0165-0327(00)00157-9
   Sacerdote P, 2006, PALLIATIVE MED, V20, pS9, DOI 10.1191/0269216306pm1124oa
   Schiepers OJG, 2005, PROG NEURO-PSYCHOPH, V29, P201, DOI 10.1016/j.pnpbp.2004.11.003
   Scholz J, 2007, NAT NEUROSCI, V10, P1361, DOI 10.1038/nn1992
   Sofroniew MV, 2010, ACTA NEUROPATHOL, V119, P7, DOI 10.1007/s00401-009-0619-8
   Sofroniew MV, 2009, TRENDS NEUROSCI, V32, P638, DOI 10.1016/j.tins.2009.08.002
   Song P, 2001, NEUROSCI RES, V39, P281, DOI 10.1016/S0168-0102(00)00226-1
   Suo JL, 1998, ADV EXP MED BIOL, V437, P177
   Tai YH, 2006, PAIN, V124, P77, DOI 10.1016/j.pain.2006.03.018
   Vallejo Ricardo, 2004, Am J Ther, V11, P354, DOI 10.1097/01.mjt.0000132250.95650.85
   VANDERLAAN JW, 1995, INT J IMMUNOPHARMACO, V17, P535, DOI 10.1016/0192-0561(95)00010-Y
   Watkins LR, 2007, BRAIN BEHAV IMMUN, V21, P131, DOI 10.1016/j.bbi.2006.10.011
   Watkins LR, 2003, ADV EXP MED BIOL, V521, P1
   Whitehead RE, 2001, J NEUROCHEM, V76, P1242, DOI 10.1046/j.1471-4159.2001.00125.x
   Zhang L, 2000, NEUROREPORT, V11, P409, DOI 10.1097/00001756-200002070-00037
NR 46
TC 4
Z9 4
U1 0
U2 7
PU UNIV SAO PAULO, CONJUNTO QUIMICAS
PI SAO PAULO
PA SERVICO PUBLICACOES E CIRCULACAO, CAIXA POSTAL 66083, SAO PAULO, 00000,
   BRAZIL
SN 1984-8250
EI 2175-9790
J9 BRAZ J PHARM SCI
JI Braz. J. Pharm. Sci.
PD OCT-DEC
PY 2016
VL 52
IS 4
BP 623
EP 633
DI 10.1590/S1984-82502016000400006
PG 11
WC Pharmacology & Pharmacy
WE Science Citation Index Expanded (SCI-EXPANDED)
SC Pharmacology & Pharmacy
GA EN1YL
UT WOS:000395806400006
OA Green Published, gold, Green Submitted
DA 2023-08-27
ER

PT J
AU Height, R
   Khoo, V
   Lawford, C
   Cox, J
   Joon, DL
   Rolfo, A
   Wada, M
AF Height, Rebecca
   Khoo, Vincent
   Lawford, Catherine
   Cox, Jennifer
   Joon, Daryl Lim
   Rolfo, Aldo
   Wada, Morikatsu
TI The dosimetric consequences of anatomic changes in head and neck
   radiotherapy patients
SO JOURNAL OF MEDICAL IMAGING AND RADIATION ONCOLOGY
LA English
DT Article
DE dosimetric verification; geometric changes; head and neck; weight loss
ID DEFORMABLE IMAGE REGISTRATION; ADAPTIVE RADIATION-THERAPY;
   COMPUTED-TOMOGRAPHY IMAGES; CANCER RADIOTHERAPY; SALIVARY-GLANDS;
   FEASIBILITY; IRRADIATION; DELINEATION; ALGORITHM; REPEAT
AB Introduction:
   To investigate anatomical response-related changes in the head and neck region during a course of radical radiotherapy and their impact on the planned dosimetry.
   Methods:
   The study consisted of 10 patients with primary mucosal carcinoma. Patients' nutritional requirements were managed as per departmental protocol to minimise weight loss during treatment. Kilovoltage computed tomography (CT) scans were acquired once 40 to 50 Gy had been delivered. Gross tumour volumes (GTV) and organs at risk were delineated and the initial optimised treatment plan was overlaid on the repeat CT. Comparisons were made between scans and absolute volume variations, centres of structures, dice similarity coefficients and the subsequent dosimetric impact were assessed.
   Results:
   Median weight loss at second scan was 3%. Primary and lymph node GTVs reduced by 49.9% (range 21.3-82%) and 73.7% (range 61.7-88.6%), respectively, yet continued to receive the prescribed dose. Maximum dose to spinal cord and brainstem changed minimally. Spared and un-spared parotid gland volumes reduced by median 23.5% and 20.5%, respectively, with no consistent translational displacement direction and minimal change in the mean dose.
   Conclusions:
   Despite some significant geometric changes, nutritional management ensured patient size and shape was maintained in these consecutively selected patients and subsequently there was no apparent under-dosing of targets or over-dosing of normal structures with this technique. Further investigations which model gradual change and allow cumulative dosimetry are required to better characterise what occurs during the treatment course.
C1 [Height, Rebecca; Khoo, Vincent; Lawford, Catherine; Joon, Daryl Lim; Wada, Morikatsu] Austin Hlth, Dept Radiat Oncol, Melbourne, Vic, Australia.
   [Height, Rebecca; Rolfo, Aldo] Peter MacCallum Canc Ctr, Dept Radiat Oncol, Melbourne, Vic, Australia.
   [Height, Rebecca; Cox, Jennifer] Univ Sydney, Fac Hlth Sci, Sydney, NSW 2006, Australia.
   [Khoo, Vincent] Royal Marsden NHS Fdn Trust, Dept Radiat Oncol, London, England.
C3 Austin Research Institute; Florey Institute of Neuroscience & Mental
   Health; Peter Maccallum Cancer Center; University of Sydney; Royal
   Marsden NHS Foundation Trust
RP Height, R (通讯作者)，Peter MacCallum Canc Ctr, Level 4,32 Erin St, Richmond, Vic 3121, Australia.
EM
CR Barker JL, 2004, INT J RADIAT ONCOL, V59, P960, DOI 10.1016/j.ijrobp.2003.12.024
   Castadot P, 2008, RADIOTHER ONCOL, V89, P1, DOI 10.1016/j.radonc.2008.04.010
   Chao M, 2008, INT J RADIAT ONCOL, V70, P599, DOI 10.1016/j.ijrobp.2007.09.057
   Ciernik IF, 2003, INT J RADIAT ONCOL, V57, P853, DOI 10.1016/S0360-3016(03)00346-8
   Court LE, 2006, PHYS MED BIOL, V51, P2493, DOI 10.1088/0031-9155/51/10/009
   Eisbruch A, 1999, INT J RADIAT ONCOL, V45, P577, DOI 10.1016/S0360-3016(99)00247-3
   Erkal HS, 2001, RADIOTHER ONCOL, V61, P247, DOI 10.1016/S0167-8140(01)00448-0
   Gregoire V, 2003, RADIOTHER ONCOL, V69, P227, DOI 10.1016/j.radonc.2003.09.011
   Hansen EK, 2006, INT J RADIAT ONCOL, V64, P355, DOI 10.1016/j.ijrobp.2005.07.957
   Khoo VS, 1997, RADIOTHER ONCOL, V42, P1, DOI 10.1016/S0167-8140(96)01866-X
   Lee C, 2008, INT J RADIAT ONCOL, V71, P1563, DOI 10.1016/j.ijrobp.2008.04.013
   Lee JH, 1998, ARCH OTOLARYNGOL, V124, P871, DOI 10.1001/archotol.124.8.871
   Lee N, 2004, INT J RADIAT ONCOL, V59, P584, DOI 10.1016/j.ijrobp.2004.02.005
   Li H, 2008, INT J RADIAT ONCOL, V71, P916, DOI 10.1016/j.ijrobp.2008.01.008
   Mercuri A, 2009, J MED IMAG RADIAT ON, V53, P500, DOI 10.1111/j.1754-9485.2009.02105.x
   Mohan R, 2005, INT J RADIAT ONCOL, V61, P1258, DOI 10.1016/j.ijrobp.2004.11.033
   Parsons JT, 2002, CANCER-AM CANCER SOC, V94, P2967, DOI 10.1002/cncr.10567
   Robar JL, 2007, INT J RADIAT ONCOL, V68, P1121, DOI 10.1016/j.ijrobp.2007.01.030
   STEPHENS LC, 1986, RADIOTHER ONCOL, V7, P165
   Wang H, 2005, INT J RADIAT ONCOL, V61, P725, DOI 10.1016/j.ijrobp.2004.07.677
   Wang H, 2008, INT J RADIAT ONCOL, V72, P210, DOI 10.1016/j.ijrobp.2008.05.008
   Wiggenraad RGJ, 2007, CLIN OTOLARYNGOL, V32, P384, DOI 10.1111/j.1749-4486.2007.01533.x
   Zhang TZ, 2007, INT J RADIAT ONCOL, V68, P522, DOI 10.1016/j.ijrobp.2007.01.038
NR 23
TC 24
Z9 25
U1 0
U2 3
PU WILEY-BLACKWELL
PI MALDEN
PA COMMERCE PLACE, 350 MAIN ST, MALDEN 02148, MA USA
SN 1754-9477
J9 J MED IMAG RADIAT ON
JI J. Med. Imag. Radiat. Oncol.
PD OCT
PY 2010
VL 54
IS 5
BP 497
EP 504
DI 10.1111/j.1754-9485.2010.02209.x
PG 8
WC Radiology, Nuclear Medicine & Medical Imaging
WE Science Citation Index Expanded (SCI-EXPANDED)
SC Radiology, Nuclear Medicine & Medical Imaging
GA 669AY
UT WOS:000283320000015
PM 20958950
DA 2023-08-27
ER

PT J
AU Crijns, CP
   Baeumlin, Y
   De Rycke, L
   Broeckx, BJG
   Vlaminck, L
   Bergman, EHJ
   van Bree, H
   Gielen, I
AF Crijns, Casper P.
   Baeumlin, Yseult
   De Rycke, Lieve
   Broeckx, Bart J. G.
   Vlaminck, Lieven
   Bergman, Erik H. J.
   van Bree, Henri
   Gielen, Ingrid
TI Intra-arterial versus intra venous contrast-enhanced computed tomography
   of the equine head
SO BMC VETERINARY RESEARCH
LA English
DT Article
DE Horse; Imaging; Computed tomography; Contrast; Head
ID HORSES; DIAGNOSIS; TUMORS; MEDIA; CT
AB Background: The anatomical complexity of the horse's head limits the abilities of radiography. Computed tomography (CT) in combination with contrast enhanced CT is used more often for diagnosing various head pathology in horses. The objective of this study was to compare intravenous and intra-arterial contrast-enhancement techniques and describe normal and abnormal contrast enhancement in the horse's head.
   Results: All 24 horses included in the study recovered without complication from the procedures. Compared to the pre-contrast studies, post-contrast studies showed significant contrast enhancement in the pituitary gland (IA: p < 0.0001; IV: p < 0.0001), IA nose septum (p = 0.002), nose mucosa (IA: p < 0.0001; IV: p = 0.02), parotid salivary gland (IA: p < 0.0001; IV p < 0.0001), cerebrum (IA: p < 0.0001; IV: p < 0.0001), rectus capitis muscle (IA: p < 0.0001; IV p = 0.001), IA temporal muscle (p < 0.0001), IA masseter muscle (p < 0.0001) and IV brainstem (p = 0.01). No significant contrast enhancement was seen in the eye (IA: p = 0.23; IV p = 0.33), tongue (IA p = 0.2; IV p = 0.57), IA brainstem (p = 0.88), IV nose septum (p = 0.26), IV temporal muscle (p = 0.09) and IV masseter muscle (p = 0.46). Three different categories of abnormal enhancement were detected: a strong vascularised mass, an enhanced rim surrounding an unenhanced structure and an inflamed anatomical structure with abnormal contrast enhancement.
   Conclusion: Using the intra-arterial technique, similar contrast enhancement is achieved using less contrast medium compared to the intravenous technique. And a potential major advantage of the IA technique is the ability to evaluate lesions that are characterized by increased blood flow. Using the intravenous technique, a symmetrical and homogenous enhancement is achieved, however timing is more crucial and the contrast dosage is more of influence in the IV protocol. And a potential major advantage of the IV technique is the ability to evaluate lesions that are characterized by increased vascular permeability. Knowing the different normal contrast enhancement patterns will facilitate the recognition of abnormal contrast enhancements.
C1 [Crijns, Casper P.; De Rycke, Lieve; van Bree, Henri; Gielen, Ingrid] Univ Ghent, Fac Vet Med, Dept Vet Med Imaging & Small Anim Orthopaed, Merelbeke, Belgium.
   [Baeumlin, Yseult] Tierarztliches Uberweisungszentrum, Tenniken, Switzerland.
   [Broeckx, Bart J. G.] Univ Ghent, Pharmaceut Sci, B-9000 Ghent, Belgium.
   [Vlaminck, Lieven] Univ Ghent, Surg & Anaesthesiol Large Anim, Merelbeke, Belgium.
   [Bergman, Erik H. J.] Lingehoeve Diergeneeskunde, Lienden, Netherlands.
C3 Ghent University; Ghent University; Ghent University
RP Crijns, CP (通讯作者)，Univ Ghent, Fac Vet Med, Dept Vet Med Imaging & Small Anim Orthopaed, Merelbeke, Belgium.
EM
RI Crijns, Casper/AAW-3965-2021; Broeckx, Bart/IYJ-3270-2023; Broeckx,
   Bart/AAH-3940-2021
OI Crijns, Casper/0000-0002-3578-6256; Broeckx, Bart/0000-0001-6742-3911
CR Ae P, 2010, AUST VET J, V88, P374
   Ap P, 2011, J VET INTERN MED, V25, P1144
   Ba B, 2007, RADIOLOGY, V242, P109
   BARONE R, 1996, ANATOMIE COMPAREE MA, V5
   Baum U, 2000, EUR J RADIOL, V33, P153, DOI 10.1016/S0720-048X(99)00139-4
   Bergman Hj, 2012, 16 C INT VET RAD ASS
   Crijns CP, 2015, EQUINE VET EDUC, V27, P11, DOI 10.1111/eve.12246
   Dd C, 2012, VET RADIOL ULTRASOUN, V53, P245
   de Vries A, 2009, J VET PHARMACOL THER, V32, P353, DOI 10.1111/j.1365-2885.2008.01051.x
   Dg M, 1999, VET J, V158, P182
   Gunkel CI, 2004, JAVMA-J AM VET MED A, V224, P1143, DOI 10.2460/javma.2004.224.1143
   Henninger W, 2003, VET RADIOL ULTRASOUN, V44, P269, DOI 10.1111/j.1740-8261.2003.tb00454.x
   Hilton H, 2009, VET RADIOL ULTRASOUN, V50, P151, DOI 10.1111/j.1740-8261.2009.01508.x
   Je S, 2002, VET RADIOL ULTRASOUN, V43, P99
   Joshi S, 2008, ANESTHESIOLOGY, V109, P543, DOI 10.1097/ALN.0b013e318182c81b
   Jr A, 1987, EQUINE VET J, V19, P552
   Kishimoto M, 2008, J VET MED SCI, V70, P981, DOI 10.1292/jvms.70.981
   Kl M, 2000, VET RADIOL ULTRASOUN, V41, P491
   Kromhout K, 2012, ACTA VET SCAND, V54, DOI 10.1186/1751-0147-54-24
   Nickel R, 2004, KREISLAUFSYSTEM HAUT, V3
   Pownder S, 2010, J VET INTERN MED, V24, P398, DOI 10.1111/j.1939-1676.2009.0456.x
   Puchalski SM, 2012, VET CLIN N AM-EQUINE, V28, P563, DOI 10.1016/j.cveq.2012.08.002
   Ra L, 1996, AM J ROENTGENOL, V167, P211
   Re P, 2008, AM J VET RES, V69, P1274
   Re P, 2011, VET RADIOL ULTRASOUN, V52, P441
   Rl E, 2007, VET RADIOL ULTRASOUN, V48, P38
   Rl T, 2001, VET CLIN N AM-EQUINE, V17, P131
   Se P, 2010, EQUINE VET J, V42, P270
   Sisson S., 1953, ANATOMY DOMESTIC ANI, V4th ed
   Sm P, 2009, VET RADIOL ULTRASOUN, V50, P292
   Sm P, 2007, VET RADIOL ULTRASOUN, V48, P21
   Sogaro-Robinson C, 2009, JAVMA-J AM VET MED A, V235, P176, DOI 10.2460/javma.235.2.176
   Sp C, 2013, VET OPHTHALMOL, V16, P352
   Tietje S, 1996, EQUINE VET J, V28, P98, DOI 10.1111/j.2042-3306.1996.tb01599.x
   Tremaine WH, 2001, EQUINE VET J, V33, P274, DOI 10.2746/042516401776249615
   Va L, 2010, EQUINE VET J, V42, P393
   Vance A, 2012, J AM ANIM HOSP ASSOC, V48, P172, DOI 10.5326/JAAHA-MS-5689
   Vanschandevijl K, 2008, JAVMA-J AM VET MED A, V233, P950, DOI 10.2460/javma.233.6.950
   Veraa S, 2009, EQUINE VET J, V41, P872, DOI 10.2746/042516409X452143
   Veraa S, 2009, EQUINE VET EDUC, V21, P284, DOI 10.2746/095777309X423031
   Vink-Nooteboom M, 1998, VET RADIOL ULTRASOUN, V39, P512, DOI 10.1111/j.1740-8261.1998.tb01641.x
   Waldenberger P, 2009, EUR RADIOL, V19, P503, DOI 10.1007/s00330-008-1161-0
NR 42
TC 9
Z9 9
U1 0
U2 8
PU BIOMED CENTRAL LTD
PI LONDON
PA 236 GRAYS INN RD, FLOOR 6, LONDON WC1X 8HL, ENGLAND
SN 1746-6148
J9 BMC VET RES
JI BMC Vet. Res.
PD JAN 7
PY 2016
VL 12
AR 6
DI 10.1186/s12917-016-0632-9
PG 13
WC Veterinary Sciences
WE Science Citation Index Expanded (SCI-EXPANDED)
SC Veterinary Sciences
GA DA3SQ
UT WOS:000367719500001
PM 26739315
OA gold, Green Published
DA 2023-08-27
ER

PT J
AU Fogt, F
   Zimmerman, RL
   Mulligan, N
   Vortmeyer, AO
   Poremba, C
   Harris, CA
   Alsaigh, N
   Tomaszewski, JE
   Nikulasson, ST
AF Fogt, F
   Zimmerman, RL
   Mulligan, N
   Vortmeyer, AO
   Poremba, C
   Harris, CA
   Alsaigh, N
   Tomaszewski, JE
   Nikulasson, ST
TI BCL-2 expression and inhibin-A in adrenal neoplasms: A comparative study
SO INTERNATIONAL JOURNAL OF MOLECULAR MEDICINE
LA English
DT Article
DE adrenal cortex; pheochromocytoma; adrenal cortical carcinoma; BCL-2;
   inhibin-A; immunohistochemistry
ID ADRENOCORTICAL CARCINOMA; DIFFERENTIAL-DIAGNOSIS; CORTICAL TUMORS; CELL
   APOPTOSIS; PHEOCHROMOCYTOMAS
AB Differential diagnosis between adrenal cortical and adrenal medullary lesions may be difficult in many cases. Different immunohistochemical, histochemical tools as well as ultrastructural diagnostic techniques have been employed to aid in differentiating between these lesions. Recently, both inhibin-A and BCL-2 have been shown to stain selectively adrenal cortical tissue and its derived neoplasms but not adrenal medulla or pheochromocytomas. In this study we compared the staining reactions of inhibin-A and BCL-2 in cases of adrenal cortical adenomas and carcinomas as well as pheochromocytomas. We found that both inhibin-A and BCL-2 stained cortical derived tissues, but not medullary derived tissues. Staining intensity for inhibin-A was significantly weaker than for BCL-2. We found that fixation techniques may influence the staining reactivity, as some cases did not immunoreact with any of the antibodies. We conclude that both inhibin-A, and, preferentially, BCL-2 are useful additions to a staining protocol to help in the differential diagnosis of cortical and medullary neoplasms.
C1 Univ Penn, Dept Pathol, Philadelphia, PA 19104 USA.
   Univ Munster, Dept Pathol, D-4400 Munster, Germany.
   Univ Iceland, Dept Pathol, Reykjavik, Iceland.
   Boston Univ, Mallory Inst Pathol, Boston, MA USA.
   NIH, Pathol Lab, Bethesda, MD 20892 USA.
   Dianon Syst, Stratford, CT USA.
C3 University of Pennsylvania; University of Munster; University of
   Iceland; Boston University; National Institutes of Health (NIH) - USA
RP Fogt, F (通讯作者)，Univ Penn, Presbyterian Med Ctr, Dept Pathol & Lab Med, 39th & Market St,551 Wright Saunders Bldg, Philadelphia, PA 19104 USA.
CR BILLIG H, 1993, ENDOCRINOLOGY, V133, P2204, DOI 10.1210/en.133.5.2204
   CHEIFETZ S, 1988, J BIOL CHEM, V263, P17225
   Chivite A, 1998, APPL IMMUNOHISTO M M, V6, P42, DOI 10.1097/00022744-199803000-00008
   COPE O, 1952, J CLIN ENDOCR METAB, V12, P875, DOI 10.1210/jcem-12-7-875
   Fogt F, 1998, MODERN PATHOL, V11, P716
   HOUGH AJ, 1979, AM J CLIN PATHOL, V72, P390
   Iczkowski KA, 1998, MODERN PATHOL, V11, P774
   KOMMINOTH P, 1995, LAB INVEST, V72, P424
   MIETTINEN M, 1992, LAB INVEST, V66, P169
   RAMSAY JA, 1987, AM J SURG PATHOL, V11, P480, DOI 10.1097/00000478-198706000-00009
   SCHRODER S, 1990, VIRCHOWS ARCH A, V417, P89, DOI 10.1007/BF02190525
   Tao XJ, 1997, J CLIN ENDOCR METAB, V82, P2738, DOI 10.1210/jc.82.8.2738
   TARTOUR E, 1993, CANCER, V72, P3296, DOI 10.1002/1097-0142(19931201)72:11<3296::AID-CNCR2820721127>3.0.CO;2-4
   VONSLOOTEN H, 1985, CANCER, V55, P766
   WEISS LM, 1984, AM J SURG PATHOL, V8, P163, DOI 10.1097/00000478-198403000-00001
NR 15
TC 7
Z9 7
U1 0
U2 0
PU SPANDIDOS PUBL LTD
PI ATHENS
PA POB 18179, ATHENS, 116 10, GREECE
SN 1107-3756
EI 1791-244X
J9 INT J MOL MED
JI Int. J. Mol. Med.
PD MAR
PY 1999
VL 3
IS 3
BP 271
EP 274
PG 4
WC Medicine, Research & Experimental
WE Science Citation Index Expanded (SCI-EXPANDED)
SC Research & Experimental Medicine
GA 169WN
UT WOS:000078772300005
PM 10028050
DA 2023-08-27
ER

PT J
AU Zindler, JD
   Bruynzeel, AME
   Eekers, DBP
   Hurkmans, CW
   Swinnen, A
   Lambin, P
AF Zindler, Jaap D.
   Bruynzeel, Anna M. E.
   Eekers, Danielle B. P.
   Hurkmans, Coen W.
   Swinnen, Ans
   Lambin, Philippe
TI Whole brain radiotherapy versus stereotactic radiosurgery for 4-10 brain
   metastases: a phase III randomised multicentre trial
SO BMC CANCER
LA English
DT Article
DE Brain metastases; Stereotactic radiosurgery; Whole brain radiotherapy;
   Quality of life
ID CANCER
AB Background: Maintenance of quality of life is the primary goal during treatment of brain metastases (BM). This is a protocol of an ongoing phase III randomised multicentre study. This study aims to determine the exact additional palliative value of stereotactic radiosurgery (SRS) over whole brain radiotherapy (WBRT) in patients with 4-10 BM.
   Methods: The study will include patients with 4-10 BM from solid primary tumours diagnosed on a high-resolution contrast-enhanced MRI scan with a maximum lesional diameter of 2.5 cm in any direction and a maximum cumulative lesional volume of 30 cm(3). Patients will be randomised between WBRT in five fractions of 4 Gy to a total dose of 20 Gy (standard arm) and single dose SRS to the BMs (study arm) in the range of 15-24 Gy. The largest BM or a localisation in the brainstem will determine the prescribed SRS dose. The primary endpoint is difference in quality of life (EQ5D EUROQOL score) at 3 months after radiotherapy with regard to baseline. Secondary endpoints are difference in quality of life (EQ5D EUROQOL questionnaire) at 6, 9 and 12 months after radiotherapy with regard to baseline. Other secondary endpoints are at 3, 6, 9 and 12 months after radiotherapy survival, Karnofsky >= 70, WHO performance status, steroid use (mg), toxicity according to CTCAE V4.0 including hair loss, fatigue, brain salvage during follow-up, type of salvage, time to salvage after randomisation and Barthel index. Facultative secondary endpoints are neurocognition with the Hopkins verbal learning test revised, quality of life EORTC QLQ-C30, quality of life EORTC BN20 brain module and fatigue scale EORTC QLQ-FA13.
   Discussion: Worldwide, most patients with more than 4 BM will be treated with WBRT. Considering the potential advantages of SRS over WBRT, i.e. limiting radiation doses to uninvolved brain and a high rate of local tumour control by just a single treatment with fewer side effects, such as hair loss and fatigue, compared to WBRT, SRS might be a suitable alternative for patients with 4-10 BM.
C1 [Zindler, Jaap D.; Eekers, Danielle B. P.; Swinnen, Ans; Lambin, Philippe] Maastricht Univ, Med Ctr, GROW Sch Oncol & Dev Biol, Dept Radiat Oncol,MAASTRO Clin, Dr Tanslaan 12, NL-6229 ET Maastricht, Netherlands.
   [Bruynzeel, Anna M. E.] Vrije Univ Amsterdam Med Ctr, Dept Radiat Oncol, Boelelaan 1117, NL-1081 HV Amsterdam, Netherlands.
   [Hurkmans, Coen W.] Catharina Hosp, Dept Radiat Oncol, Michelangelolaan 2, NL-5623 EJ Eindhoven, Netherlands.
   [Zindler, Jaap D.] MAASTRO Clin, POB 3035, NL-6202 NA Maastricht, Netherlands.
C3 Maastricht University; Maastricht University Medical Centre (MUMC);
   Vrije Universiteit Amsterdam; VU UNIVERSITY MEDICAL CENTER; Catharina
   Hospital; Maastricht University; Maastricht University Medical Centre
   (MUMC)
RP Zindler, JD (通讯作者)，Maastricht Univ, Med Ctr, GROW Sch Oncol & Dev Biol, Dept Radiat Oncol,MAASTRO Clin, Dr Tanslaan 12, NL-6229 ET Maastricht, Netherlands.; Zindler, JD (通讯作者)，MAASTRO Clin, POB 3035, NL-6202 NA Maastricht, Netherlands.
EM
RI Eekers, Danielle/IVH-6693-2023
OI Eekers, Danielle/0000-0003-1917-6279; Lambin,
   Philippe/0000-0001-7961-0191
FU MAASTRO clinic; Varian Medical Systems
FX According to the research agreement of MAASTRO clinic with Varian
   Medical Systems, this trial is mentioned and financially supported.
   Varian Medical Systems is not involved in the design of the study, and
   collection/storage/analysis of the data gathered in this study.
CR Brown PD, 2016, JAMA-J AM MED ASSOC, V316, P401, DOI 10.1001/jama.2016.9839
   Chavaudra J, 2001, Cancer Radiother, V5, P472, DOI 10.1016/S1278-3218(01)00117-2
   Mulvenna P, 2016, LANCET, V388, P2004, DOI 10.1016/S0140-6736(16)30825-X
   Pickard AS, 2007, HEALTH QUAL LIFE OUT, V5, DOI 10.1186/1477-7525-5-70
   Scoccianti S, 2015, RADIOTHER ONCOL, V114, P230, DOI 10.1016/j.radonc.2015.01.016
   Yamamoto M, 2014, LANCET ONCOL, V15, P387, DOI 10.1016/S1470-2045(14)70061-0
NR 6
TC 25
Z9 25
U1 0
U2 2
PU BIOMED CENTRAL LTD
PI LONDON
PA 236 GRAYS INN RD, FLOOR 6, LONDON WC1X 8HL, ENGLAND
SN 1471-2407
J9 BMC CANCER
JI BMC Cancer
PD JUL 25
PY 2017
VL 17
AR 500
DI 10.1186/s12885-017-3494-z
PG 5
WC Oncology
WE Science Citation Index Expanded (SCI-EXPANDED)
SC Oncology
GA FB8WX
UT WOS:000406421500003
PM 28743240
OA Green Published, gold
DA 2023-08-27
ER

PT J
AU Tai, DT
   Oanh, LT
   Phuong, PH
   Sulieman, A
   Abolaban, FA
   Omer, H
   Chow, JCL
AF Duong Thanh Tai
   Luong Thi Oanh
   Pham Hoai Phuong
   Sulieman, Abdelmoneim
   Abolaban, Fouad A.
   Omer, Hiba
   Chow, James C. L.
TI Dosimetric and radiobiological comparison in head-and-neck radiotherapy
   using JO-IMRT and 3D-CRT
SO SAUDI JOURNAL OF BIOLOGICAL SCIENCES
LA English
DT Article
DE 3D-CRT technique; JO-IMRT technique; Dosimetric and radiobiological
   evaluation; Equivalent Uniform Dose (EUD); Plan evaluation
ID MODULATED RADIATION-THERAPY; QUALITY-ASSURANCE; TREATMENT PLANS;
   COLLAPSED-CONE; ARC THERAPY; JAWS; INDEX; BEAM
AB Introduction: Dosimetric and radiobiological evaluations for the Jaws-only Intensity-modulated radiotherapy (JO-IMRT) technique for head and neck jaws-only intensity-modulated radiation therapy (JOIMRT) and 3D conformal radiation therapy (3D-CRT). To compare the head-and-neck therapeutic approaches utilizing JO-IMRT and 3D-CRT techniques, different radiation dose indices were calculated, including: conformity index (CI), homogeneity index (HI), and radiobiological variables like Niemierko's equivalent uniform dose based tumor control probability (TCP) of planning target volume (PTV), normal tissue complication probability (NTCP) of organs at risk (OAR) (brainstem, spinal cord, and parotid grand).
   Materials and methods: Twenty-five nasopharynx patients were studied using the Prowess Panther Treatment Planning System (Prowess Inc). The results were compared with the dose distribution obtained using 3D-CRT.
   Results: Regarding tumor coverage and CI, JO-IMRT showed better results than 3D-CRT. The average doses received by the PTVs were quite similar: 72.1 +/- 0.8 Gy by 3D-CRT and 72.5 +/- 0.6 Gy by JO-IMRT plans (p > 0.05). The mean doses received by the parotid gland were 56.7 +/- 0.7 Gy by 3D-CRT and 26.8 +/- 0.3 Gy by JO-IMRT (p > 0.05). The HI and CI were 0.13 +/- 0.01 and 0.14 +/- 0.05 and (p > 0.05) by 3DCRT and 0.83 +/- 0.05 and 0.73 +/- 0.10 by JO-IMRT (p < 0.05). The average TCP of PTV was 0.82 +/- 0.08 by 3D-CRT and 0.92 +/- 0.02 by JO-IMRT. Moreover, the NTCP of the parotid glands, brain stem, and spinal cord were lower using the JO-IMRT than 3D-CRT plans. In comparison to the 3D-CRT approach, the JO-IMRT technique was able to boost dose coverage to the PTV, improve the target's CI and HI, and spare the parotid glands. This suggests the power of the JO-IMRT over 3D-CRT in head-and-neck radiotherapy. (C) 2022 The Author(s). Published by Elsevier B.V. on behalf of King Saud University.
C1 [Duong Thanh Tai] Nguyen Tat Thanh Univ, Fac Med, Dept Med Phys, 298-300A Nguyen Tat Thanh St,Ward 13,Dist 4, Ho Chi Minh City, Vietnam.
   [Duong Thanh Tai] Robarts Res Inst, London, ON N6A 5K8, Canada.
   [Luong Thi Oanh] Ho Chi Minh City Oncol Hosp, Dept Radiat Technol, Ho Chi Minh City 702000, Vietnam.
   [Pham Hoai Phuong] Nguyen Tat Thanh Univ, VKTECH Res Ctr, 298-300A Nguyen Tat Thanh St,Ward 13,Dist 4, Ho Chi Minh City, Vietnam.
   [Sulieman, Abdelmoneim] Prince Sattam Bin Abdulaziz Univ, Coll Appl Med Sci, Radiol & Med Imaging Dept, Alkharj, Saudi Arabia.
   [Abolaban, Fouad A.] King Abdulaziz Univ, Dept Nucl Engn, Fac Engn, POB 80204, Jeddah 21589, Saudi Arabia.
   [Abolaban, Fouad A.] King Abdulaziz Univ, KA CARE Energy Res & Innovat Ctr, Jeddah 21589, Saudi Arabia.
   [Omer, Hiba] Imam Abdulrahman Bin Faisal Univ, Dept Basic Sci, Deanship Preparatory Year & Supporting Studies, POB 1982, Dammam 34212, Saudi Arabia.
   [Chow, James C. L.] Univ Toronto, Dept Radiat Oncol, Toronto, ON M5S 1A1, Canada.
   [Chow, James C. L.] Univ Hlth Network, Princess Margaret Canc Ctr, Radiat Med Program, Toronto, ON M5G 1X6, Canada.
C3 Nguyen Tat Thanh University (NTTU); Western University (University of
   Western Ontario); Nguyen Tat Thanh University (NTTU); Prince Sattam Bin
   Abdulaziz University; King Abdulaziz University; King Abdulaziz
   University; Imam Abdulrahman Bin Faisal University; University of
   Toronto; University of Toronto; University Health Network Toronto;
   Princess Margaret Cancer Centre
RP Tai, DT; Sulieman, A; Chow, JCL (通讯作者)，Nguyen Tat Thanh Univ, Fac Med, Dept Med Phys, 298-300A Nguyen Tat Thanh St,Ward 13,Dist 4, Ho Chi Minh City, Vietnam.; Tai, DT; Sulieman, A; Chow, JCL (通讯作者)，Univ Toronto, Dept Radiat Oncol, Toronto, ON M5T 1P5, Canada.; Tai, DT; Sulieman, A; Chow, JCL (通讯作者)，Prince Sattam Bin Abdulaziz Univ, POB 422, Alkharj 11943, Saudi Arabia.
EM;
  ;
  
RI Chow, James C. L./JBS-0697-2023; Tai, PhD, Duong Thanh/U-9445-2018
OI Chow, James C. L./0000-0003-4202-4855; Tai, PhD, Duong
   Thanh/0000-0001-7276-8105; Sulieman, Abdelmoneim/0000-0003-0830-4706
FU King Abdullah City for Atomic and Renewable Energy (K.A.CARE) under K.A.
   CARE-King Abdulaziz University Collaboration Program; Deanship of
   Scientific Research, King Abdulaziz University [RG-87-135-42]
FX The authors acknowledge the support provided by King Abdullah City for
   Atomic and Renewable Energy (K.A.CARE) under K.A. CARE-King Abdulaziz
   University Collaboration Program. The authors are also thankful to the
   Deanship of Scientific Research, King Abdulaziz University for providing
   financial support vide grant number (RG-87-135-42). The first author
   would like to extend thanks to the Director of Dong Nai General Hospital
   and the Head of Radiation Oncology for their support.
CR [Anonymous], 2010, J ICRU, V10, P27, DOI 10.1093/jicru/ndq008
   Baltas D, 1998, INT J RADIAT ONCOL, V40, P515, DOI 10.1016/S0360-3016(97)00732-3
   Banaei A, 2019, POL J MED PHYS ENG, V25, P35, DOI 10.2478/pjmpe-2019-0006
   Bray F, 2021, CANCER-AM CANCER SOC, V127, P3029, DOI 10.1002/cncr.33587
   Chow JCL, 2019, J RADIOTHER PRACT, V18, P323, DOI 10.1017/S1460396919000098
   Chow JCL, 2020, MED DOSIM, V45, P14, DOI 10.1016/j.meddos.2019.04.004
   Chow JCL, 2018, MED DOSIM, V43, P383, DOI 10.1016/j.meddos.2017.12.002
   Tai DT, 2019, REP PRACT ONCOL RADI, V24, P105, DOI 10.1016/j.rpor.2018.11.004
   Tai DT, 2018, POL J MED PHYS ENG, V24, P75, DOI 10.2478/pjmpe-2018-0010
   Tai DT, 2017, PHYS MEDICA, V38, P148, DOI 10.1016/j.ejmp.2017.05.059
   Ebert M., 2001, RADIOL ONCOL, V35, P215
   Eisbruch A., 2005, 0022 RTOG
   El-Ghoneimy E.G., 2012, CHINESE GERMAN J CLI, V11, P626
   Gay HA, 2007, PHYS MEDICA, V23, P115, DOI 10.1016/j.ejmp.2007.07.001
   Khairi Y, 2021, RADIAT PHYS CHEM, V178, DOI 10.1016/j.radphyschem.2020.109000
   Khan MI, 2016, J MED PHYS, V41, P162, DOI 10.4103/0971-6203.189479
   Kim KH, 2018, PLOS ONE, V13, DOI 10.1371/journal.pone.0207232
   Kim SJ, 2015, J KOREAN PHYS SOC, V67, P153, DOI 10.3938/jkps.67.153
   Lee S, 2015, PHYS RADIOBIOLOGICAL, P109
   Mesbahi A., 2017, IRAN J MED PHYS, V14, P190
   Mu G, 2009, PHYS MED BIOL, V54, P5613, DOI 10.1088/0031-9155/54/18/017
   Paddick I, 2000, J NEUROSURG, V93, P219, DOI 10.3171/jns.2000.93.supplement_3.0219
   Park YK, 2014, J APPL CLIN MED PHYS, V15, P316, DOI 10.1120/jacmp.v15i4.4941
   Paudel NR, 2017, J APPL CLIN MED PHYS, V18, P237, DOI 10.1002/acm2.12145
   Puzhakkal N, 2016, POL J MED PHYS ENG, V22, P61, DOI 10.1515/pjmpe-2016-0011
   Rehman JU, 2018, J MED PHYS, V43, P129, DOI 10.4103/jmp.JMP_106_17
   Sung H, 2021, CA-CANCER J CLIN, V71, P209, DOI 10.3322/caac.21660
   Tai D.T., 2017, P INT C DEV BIOMEDIC, V69, P335
   Tai D.T, 2017, SCI TECHNOL DEV J NA, V1, P79
   Zheng RF, 2015, J BUON, V20, P248
   Zugazagoitia J, 2016, CLIN THER, V38, P1551, DOI 10.1016/j.clinthera.2016.03.026
NR 31
TC 3
Z9 3
U1 0
U2 2
PU ELSEVIER
PI AMSTERDAM
PA RADARWEG 29, 1043 NX AMSTERDAM, NETHERLANDS
SN 1319-562X
EI 2213-7106
J9 SAUDI J BIOL SCI
JI Saudi J. Biol. Sci.
PD AUG
PY 2022
VL 29
IS 8
AR 103336
DI 10.1016/j.sjbs.2022.103336
PG 6
WC Biology
WE Science Citation Index Expanded (SCI-EXPANDED)
SC Life Sciences & Biomedicine - Other Topics
GA 5D0HR
UT WOS:000864632500001
PM 35754762
OA gold, Green Published
DA 2023-08-27
ER

PT J
AU MOZLEY, PD
   KIM, CK
   MOHSIN, J
   JATLOW, A
   GOSFIELD, E
   ALAVI, A
AF MOZLEY, PD
   KIM, CK
   MOHSIN, J
   JATLOW, A
   GOSFIELD, E
   ALAVI, A
TI THE EFFICACY OF IODINE-123-MIBG AS A SCREENING-TEST FOR PHEOCHROMOCYTOMA
SO JOURNAL OF NUCLEAR MEDICINE
LA English
DT Article
DE IODINE-123-METAIODOBENZYLGUANIDINE; PHEOCHROMOCYTOMA; CATECHOLAMINES
ID ADRENAL-MEDULLA; I-131 MIBG; LOCALIZATION; SCINTIGRAPHY;
   METAIODOBENZYLGUANIDINE; MANAGEMENT; DIAGNOSIS; PARAGANGLIOMAS;
   EXPERIENCE; TUMORS
AB The primary objective of this study was to characterize the effectiveness of I-123-metaiodobenzylguanidine (MIBG) as a screening test for pheochromocytoma in routine clinical practice. An attempt was made to determine why some tumors and some adrenal glands without pheochromocytoma minimally manifest increased uptake. Methods: Planar images were obtained with a standardized protocol in a diverse group of patients. The intensity of uptake in each adrenal gland was graded on a 0-3-point scale by using the intensity of activity in the liver as a reference. Follow-up data were obtained from both the patients and the referring physicians. A final diagnosis was eventually established in 120 patients who had a total of 238 adrenal glands. Results: There was an intramedullary pheochromocytoma in 24 of the 238 adrenal glands (10.1%). The uptake was very intense (Grade 3) in 21 of them (87.5%). The uptake was only mildly to moderately increased in the other three intraadrenal tumors. There was no visible uptake in 148 of the 214 (69.2%) adrenals without a pheochromocytoma, but there was mildly to moderately increased activity in 66 (30.8%). There were no other features of the clinical data base which could differentiate between mildly increased uptake in a pheochromocytoma and mildly increased activity in a gland without a tumor, including the 24-hr urinary excretion of catecholamines. Conclusions: Since every intra- and extra-adrenal tumor was visualized, the findings suggest that I-123-MIBG may be the most sensitive screening test available for diagnosing pheochromocytoma. The test results should be definitive in most patients.
C1 UNIV PENN,MED CTR,DEPT RADIOL,DIV NUCL MED,PHILADELPHIA,PA 19104.
C3 University of Pennsylvania
CR ACKERY DM, 1984, BRIT MED J, V288, P1587, DOI 10.1136/bmj.288.6430.1587
   BEARD CM, 1983, MAYO CLIN PROC, V58, P802
   BEIERWALTES WH, 1991, J NUCL MED, V32, P1627
   BOMANJI J, 1987, J NUCL MED, V28, P973
   BOMANJI J, 1988, CLIN RADIOL, V39, P502, DOI 10.1016/S0009-9260(88)80214-9
   BRAVO EL, 1979, NEW ENGL J MED, V301, P682, DOI 10.1056/NEJM197909273011302
   BRAVO EL, 1984, NEW ENGL J MED, V311, P1298, DOI 10.1056/NEJM198411153112007
   CAMPEAU RJ, 1991, SOUTHERN MED J, V84, P1221, DOI 10.1097/00007611-199110000-00015
   CHATAL JF, 1993, J NUCL MED, V34, P180
   ENGELMAN K, 1977, CLIN ENDOCRINOL META, V6, P769, DOI 10.1016/S0300-595X(77)80079-0
   FRANCIS IR, 1992, RADIOLOGY, V184, P1
   FRANCIS IR, 1983, AM J ROENTGENOL, V141, P717
   GROSS MD, 1989, SEMIN NUCL MED, V19, P122, DOI 10.1016/S0001-2998(89)80007-8
   GROSS MD, 1989, DIAGNOSTIC NUCLEAR M, P815
   KALFF V, 1982, SURGERY, V91, P374
   LANDSBERG L, 1991, HARRISONS PRINCIPLES, P1735
   LYNN MD, 1984, J NUCL MED, V25, P436
   LYNN MD, 1985, RADIOLOGY, V156, P789
   MAUREA S, 1993, J NUCL MED, V34, P173
   NAKAJO M, 1983, J NUCL MED, V24, P672
   NEUMAN HPH, 1993, NEW ENGL J MED, V329, P1521
   PARISI MT, 1992, SEMIN NUCL MED, V22, P46, DOI 10.1016/S0001-2998(05)80157-6
   QUINT LE, 1987, RADIOLOGY, V165, P89, DOI 10.1148/radiology.165.1.3628794
   REMINE WH, 1974, ANN SURG, V179, P740, DOI 10.1097/00000658-197405000-00029
   SAMAAN NA, 1988, CANCER, V62, P2451, DOI 10.1002/1097-0142(19881201)62:11<2451::AID-CNCR2820621134>3.0.CO;2-Q
   SANFIELD JA, 1989, ARCH NEUROL-CHICAGO, V46, P53, DOI 10.1001/archneur.1989.00520370055019
   SHAPIRO B, 1984, CLIN ENDOCRINOL, V20, P189, DOI 10.1111/j.1365-2265.1984.tb00074.x
   SHAPIRO B, 1985, J NUCL MED, V26, P576
   SHEPS SG, 1990, MAYO CLIN PROC, V65, P88, DOI 10.1016/S0025-6196(12)62113-2
   SISSON JC, 1981, NEW ENGL J MED, V305, P12, DOI 10.1056/NEJM198107023050103
   SWENSON SJ, 1985, MAYO CLIN P, V60, P229
   TRONCONE L, 1990, EUR J NUCL MED, V16, P325, DOI 10.1007/BF00842788
   VANGILS APG, 1990, J NUCL MED, V31, P1147
   VANHEERDEN JA, 1982, SURGERY, V91, P367
   VELCHIK MG, 1989, J NUCL MED, V30, P328
   WIELAND DM, 1979, J NUCL MED, V20, P155
   WIELAND DM, 1980, J NUCL MED, V21, P349
   ZHU RS, 1988, CHINESE MED J-PEKING, V101, P513
NR 38
TC 75
Z9 77
U1 0
U2 3
PU SOC NUCLEAR MEDICINE INC
PI RESTON
PA 1850 SAMUEL MORSE DR, RESTON, VA 20190-5316
SN 0161-5505
J9 J NUCL MED
JI J. Nucl. Med.
PD JUL
PY 1994
VL 35
IS 7
BP 1138
EP 1144
PG 7
WC Radiology, Nuclear Medicine & Medical Imaging
WE Science Citation Index Expanded (SCI-EXPANDED)
SC Radiology, Nuclear Medicine & Medical Imaging
GA NW337
UT WOS:A1994NW33700015
PM 8014671
DA 2023-08-27
ER

PT J
AU Brambilla, D
   Barajon, I
   Bianchi, S
   Opp, MR
   Imeri, L
AF Brambilla, Dario
   Barajon, Isabella
   Bianchi, Susanna
   Opp, Mark R.
   Imeri, Luca
TI Interleukin-1 Inhibits Putative Cholinergic Neurons in Vitro and REM
   Sleep when Microinjected into the Rat Laterodorsal Tegmental Nucleus
SO SLEEP
LA English
DT Article
DE Cytokines; patch clamp; acetylcholine; glutamate; GABA; adenosine
ID MEDIAL PREOPTIC AREA; CORTICOTROPIN-RELEASING HORMONE; MUSCARINIC
   RECEPTOR SUBTYPES; TUMOR-NECROSIS-FACTOR; DORSAL RAPHE NUCLEUS;
   PONTOMESENCEPHALIC TEGMENTUM; SEROTONERGIC NEURONS; SYNAPTIC INHIBITION;
   BRAIN TEMPERATURE; BODY-TEMPERATURE
AB Study Objectives: REM sleep is suppressed during infection, an effect mimicked by the administration of cytokines such as interleukin-1 (IL-1). In spite of this observation, brain sites and neurochemical systems mediating IL-1-induced suppression of REM sleep have not been identified. Cholinergic neurons in the brainstem laterodorsal tegmental nucleus (LDT) are part of the neuronal circuitry responsible for REM sleep generation. Since IL-1 inhibits acetylcholine synthesis and release, the aim of this study was to test the two different, but related hypotheses. We hypothesized that IL-1 inhibits LDT cholinergic neurons, and that, as a result of this inhibition, IL-1 suppresses REM sleep.
   Design, Measurement, and Results: To test these hypotheses, the electrophysiological activity of putative cholinergic LDT neurons was recorded in a rat brainstem slice preparation. Interleukin-1 significantly inhibited the firing rate of 76% of recorded putative cholinergic LDT neurons and reduced the amplitude of glutamatergic evoked potentials in 60% of recorded neurons. When IL-1 (1 ng) was microinjected into the LDT of freely behaving rats, REM sleep was reduced by about 50% (from 12.7% +/- 1.5% of recording time [after vehicle] to 6.1% +/- 1.4% following IL-1 administration) during post-injection hours 3-4.
   Conclusions: Results of this study support the hypothesis that IL-1 can suppress REM sleep by acting at the level of the LDT nucleus. Furthermore this effect may result from the inhibition of evoked glutamatergic responses and of spontaneous firing of putative cholinergic LDT neurons.
C1 [Brambilla, Dario; Bianchi, Susanna; Imeri, Luca] Univ Milan, Sch Med, Dept Human Physiol, I-20133 Milan, Italy.
   [Brambilla, Dario; Bianchi, Susanna; Imeri, Luca] Univ Milan, Sch Med, Giuseppe Moruzzi Ctr Expt Sleep Res, I-20133 Milan, Italy.
   [Barajon, Isabella] Univ Milan, Sch Med, Dept Human Morphol & Biomed Sci, I-20133 Milan, Italy.
   [Opp, Mark R.] Univ Michigan, Sch Med, Dept Mol & Integrat Physiol, Ann Arbor, MI USA.
   [Opp, Mark R.; Imeri, Luca] Univ Michigan, Sch Med, Dept Anesthesiol, Ann Arbor, MI USA.
C3 University of Milan; University of Milan; University of Milan;
   University of Michigan System; University of Michigan; University of
   Michigan System; University of Michigan
RP Imeri, L (通讯作者)，Univ Milan, Sch Med, Dept Human Physiol, Via Mangiagalli 32, I-20133 Milan, Italy.
EM
RI Brambilla, Dario/F-3126-2012
OI Barajon, Isabella/0000-0001-5893-0241; IMERI, LUCA/0000-0003-0753-004X;
   Opp, Mark/0000-0002-9831-9866; Brambilla, Dario/0000-0002-4838-482X
FU National Institutes of Health [MH 64843]
FX This work was supported, in part, by the National Institutes of Health
   grant MH 64843 (MRO, LI). The authors thank Dr. Francesca Amaboldi for
   technical assistance in immunofluorescence identification of recorded
   neurons.
CR Arrigoni E, 2001, J NEUROSCI, V21, P1076, DOI 10.1523/JNEUROSCI.21-03-01076.2001
   BAGHDOYAN HA, 2002, SLEEP SCI INTEGRATIN, P88
   Brambilla D, 2007, EUR J NEUROSCI, V26, P1862, DOI 10.1111/j.1460-9568.2007.05796.x
   Brambilla D, 2005, NEURON, V46, P275, DOI 10.1016/j.neuron.2005.03.016
   CARMELIET P, 1989, BRAIN RES, V491, P199, DOI 10.1016/0006-8993(89)90106-6
   CHIBA T, 1985, BRAIN RES BULL, V14, P261, DOI 10.1016/0361-9230(85)90091-7
   De A, 2002, BRAIN RES, V949, P209, DOI 10.1016/S0006-8993(02)03157-8
   Dinarello CA, 2005, J EXP MED, V201, P1355, DOI 10.1084/jem.20050640
   DINARELLO CA, 1987, J IMMUNOL, V139, P1902
   ELMANSARI M, 1989, EXP BRAIN RES, V76, P519, DOI 10.1007/BF00248908
   Feleder C, 2000, NEUROIMMUNOMODULAT, V7, P46, DOI 10.1159/000026419
   FORD B, 1995, J COMP NEUROL, V363, P177, DOI 10.1002/cne.903630203
   Gemma C, 1997, AM J PHYSIOL-REG I, V272, pR601, DOI 10.1152/ajpregu.1997.272.2.R601
   GREENE RW, 1999, HDB BEHAV STATE CONT, P277
   IMERI L, 1994, BRAIN RES, V636, P68, DOI 10.1016/0006-8993(94)90176-7
   Imeri L, 2004, BRAIN BEHAV IMMUN, V18, P399, DOI 10.1016/j.bbi.2003.12.003
   IMERI L, 1995, AM J PHYSIOL-REG I, V269, pR294, DOI 10.1152/ajpregu.1995.269.2.R294
   Imeri L, 1996, NEUROREPORT, V7, P417, DOI 10.1097/00001756-199601310-00010
   Imeri L, 1997, AM J PHYSIOL-REG I, V273, P1663
   Imeri L, 2009, NAT REV NEUROSCI, V10, P199, DOI 10.1038/nrn2576
   KAMONDI A, 1992, J NEUROPHYSIOL, V68, P1359, DOI 10.1152/jn.1992.68.4.1359
   KANG Y, 1990, BRAIN RES, V535, P79, DOI 10.1016/0006-8993(90)91826-3
   KIRKPATRICK WE, 1970, NEUROPHARMACOLOGY, V9, P195, DOI 10.1016/0028-3908(70)90067-5
   Kluger MJ, 1996, INFECT DIS CLIN N AM, V10, P1, DOI 10.1016/S0891-5520(05)70282-8
   Li YK, 2000, J NEUROSCI, V20, P149, DOI 10.1523/JNEUROSCI.20-01-00149.2000
   LIN MT, 1980, NEUROPHARMACOLOGY, V19, P561, DOI 10.1016/0028-3908(80)90027-1
   Lu J, 2006, NATURE, V441, P589, DOI 10.1038/nature04767
   LUEBKE JI, 1992, P NATL ACAD SCI USA, V89, P743, DOI 10.1073/pnas.89.2.743
   Luk WP, 1999, J NEUROSCI, V19, P4238
   Mallick BN, 1997, BRAIN RES, V750, P311, DOI 10.1016/S0006-8993(96)01400-X
   Maloney KJ, 1999, J NEUROSCI, V19, P3057
   Manfridi A, 2003, EUR J NEUROSCI, V18, P1041, DOI 10.1046/j.1460-9568.2003.02836.x
   McCarley RW, 2007, SLEEP MED, V8, P302, DOI 10.1016/j.sleep.2007.03.005
   MCGINTY D, 1994, BRAIN RES, V667, P273, DOI 10.1016/0006-8993(94)91506-7
   MILLER LG, 1991, MOL PHARMACOL, V39, P105
   MITANI A, 1988, J COMP NEUROL, V268, P527, DOI 10.1002/cne.902680405
   Obal F, 2003, FRONT BIOSCI-LANDMRK, V8, pD520, DOI 10.2741/1033
   Olivadoti MD, 2008, NEUROSCIENCE, V153, P338, DOI 10.1016/j.neuroscience.2008.02.008
   Opp MR, 2005, SLEEP MED REV, V9, P355, DOI 10.1016/j.smrv.2005.01.002
   OPP MR, 1991, AM J PHYSIOL, V260, pR52, DOI 10.1152/ajpregu.1991.260.1.R52
   Opp MR, 2001, NEUROENDOCRINOLOGY, V73, P272, DOI 10.1159/000054644
   Opp MR, 1997, PHYSIOL BEHAV, V63, P67, DOI 10.1016/S0031-9384(97)00390-9
   Parmeggiani PL, 2003, FRONT BIOSCI, V8, pS557, DOI 10.2741/1054
   Paxinos G, 2018, RAT BRAIN STEREOTAXI
   POOLE S, 1979, BRIT J PHARMACOL, V65, P43, DOI 10.1111/j.1476-5381.1979.tb17332.x
   Proescholdt MG, 2000, NEUROSCIENCE, V95, P577
   RADA P, 1991, BRAIN RES, V550, P287, DOI 10.1016/0006-8993(91)91330-4
   RAINNIE DG, 1994, SCIENCE, V263, P689, DOI 10.1126/science.8303279
   Rodrigues AG, 2004, J APPL PHYSIOL, V97, P333, DOI 10.1152/japplphysiol.00742.2003
   SCHOBITZ B, 1994, PROG NEUROBIOL, V44, P397, DOI 10.1016/0301-0082(94)90034-5
   SEMBA K, 1990, NEUROSCIENCE, V38, P643, DOI 10.1016/0306-4522(90)90058-C
   SEMBA K, 1999, HDB BEHAV STATE CONT, P161
   Sperlagh B, 2004, J NEUROIMMUNOL, V151, P33, DOI 10.1016/j.jneuroim.2004.02.004
   STERIADE M, 1990, J NEUROSCI, V10, P2560
   SZYMUSIAK R, 1981, PHYSIOL BEHAV, V26, P687, DOI 10.1016/0031-9384(81)90145-1
   Tabarean IV, 2006, NEUROSCIENCE, V141, P1685, DOI 10.1016/j.neuroscience.2006.05.007
   Taishi P, 2008, BRAIN RES, V1226, P89, DOI 10.1016/j.brainres.2008.05.067
   Takahashi A, 2001, BRAIN RES, V889, P191, DOI 10.1016/S0006-8993(00)03132-2
   Thakkar M, 1996, BRAIN RES, V723, P223, DOI 10.1016/0006-8993(96)00256-9
   Ulnas R. R., 1990, BRAIN CHOLINERGIC SY, P205
   Vitkovic L, 2000, J NEUROCHEM, V74, P457, DOI 10.1046/j.1471-4159.2000.740457.x
   WEBSTER HH, 1988, BRAIN RES, V458, P285, DOI 10.1016/0006-8993(88)90471-4
   WILLIAMS JA, 1993, J NEUROSCI, V13, P3878
   ZEISE ML, 1992, REGUL PEPTIDES, V39, P1, DOI 10.1016/0167-0115(92)90002-C
   Zhu G, 2006, EUR J PHARMACOL, V532, P246, DOI 10.1016/j.ejphar.2005.12.085
NR 65
TC 13
Z9 14
U1 0
U2 6
PU OXFORD UNIV PRESS INC
PI CARY
PA JOURNALS DEPT, 2001 EVANS RD, CARY, NC 27513 USA
SN 0161-8105
EI 1550-9109
J9 SLEEP
JI Sleep
PD JUL 1
PY 2010
VL 33
IS 7
BP 919
EP 929
DI 10.1093/sleep/33.7.919
PG 11
WC Clinical Neurology; Neurosciences
WE Science Citation Index Expanded (SCI-EXPANDED)
SC Neurosciences & Neurology
GA 618OO
UT WOS:000279365600012
PM 20614852
OA Green Published, Bronze
DA 2023-08-27
ER

PT J
AU Tai, AX
   Nayar, VV
AF Tai, Alexander X.
   Nayar, Vikram V.
TI Update on Trigeminal Neuralgia
SO CURRENT TREATMENT OPTIONS IN NEUROLOGY
LA English
DT Review
DE Trigeminal neuralgia; Facial pain; Microvascular decompression;
   Percutaneous trigeminal rhizotomy; Radiosurgery
ID GAMMA-KNIFE RADIOSURGERY; MICROVASCULAR DECOMPRESSION; NEUROVASCULAR
   COMPRESSION; CONSECUTIVE PATIENTS; PROGNOSTIC-FACTORS; EFFICACY;
   HISTORY; SERIES
AB Purpose of reviewTo review current treatments for trigeminal neuralgia, with an emphasis on determining which patients may benefit from neurosurgical procedures.Recent findingsA detailed history is the most helpful tool for diagnosing trigeminal neuralgia (TN) and predicting response to neurosurgical treatments. Patients with classic trigeminal neuralgia will describe severe, unilateral, intermittent facial pain that is triggered by innocuous sensory stimuli. In most cases, pain is caused by compression of the trigeminal nerve by a blood vessel near the brainstem. Magnetic resonance imaging is necessary to rule out TN secondary to multiple sclerosis or tumor. Modern high-resolution T2 images may demonstrate neurovascular contact, particularly when analyzed by a neurosurgeon with expertise in TN. Initial management involves a trial of medication, usually carbamazepine or oxcarbazepine. Microvascular decompression (MVD) is safe and effective surgery, for patients with classic TN related to neurovascular compression. For patients with TN secondary to multiple sclerosis, and for patients who are otherwise poor candidates for MVD, neurosurgical options include percutaneous trigeminal rhizotomy and radiosurgery. Neurosurgical procedures are less effective in relieving atypical facial pain.SummaryIn the clinical evaluation of a patient with facial pain, it is important to distinguish classic trigeminal neuralgia from atypical facial pain. A patient with classic trigeminal neuralgia would benefit from neurosurgical consultation. The advent of high-resolution MRI and MRA sequences now allows a neurosurgeon to detect when neurovascular compression is likely, and select the optimal procedure for treatment.
C1 [Tai, Alexander X.; Nayar, Vikram V.] Medstar Georgetown Univ Hosp, Dept Neurosurg, 3800 Reservoir Rd NW,PHC 7, Washington, DC 20007 USA.
C3 Georgetown University
RP Nayar, VV (通讯作者)，Medstar Georgetown Univ Hosp, Dept Neurosurg, 3800 Reservoir Rd NW,PHC 7, Washington, DC 20007 USA.
EM
OI Tai, Alexander/0000-0003-3800-6101
CR Antonini G, 2014, PAIN, V155, P1464, DOI 10.1016/j.pain.2014.04.020
   Arai YCP, 2013, J ANESTH, V27, P960, DOI 10.1007/s00540-013-1641-5
   Barker FG, 1996, NEW ENGL J MED, V334, P1077, DOI 10.1056/NEJM199604253341701
   Bendtsen L, 2019, EUR J NEUROL, V26, P831, DOI 10.1111/ene.13950
   Bes A, 2013, CEPHALALGIA, V33, P629, DOI 10.1177/0333102413485658
   Beydoun A, 2000, PHARMACOTHERAPY, V20, p152S, DOI 10.1592/phco.20.12.152S.35254
   BURCHIEL KJ, 1988, J NEUROSURG, V69, P35, DOI 10.3171/jns.1988.69.1.0035
   Cheshire WP, 1997, CNS DRUGS, V7, P98, DOI 10.2165/00023210-199707020-00002
   Devor M, 2002, CLIN J PAIN, V18, P4, DOI 10.1097/00002508-200201000-00002
   Di Stefano G, 2014, J HEADACHE PAIN, V15, DOI 10.1186/1129-2377-15-34
   Eller Jorge L, 2005, Neurosurg Focus, V18, pE3
   Fothergill J., 1773, MEDICAL OBSERVATIONS, V5, P129
   Fraioli MF, 2012, J CLIN NEUROSCI, V19, P1401, DOI 10.1016/j.jocn.2011.11.036
   Gronseth G, 2008, NEUROLOGY, V71, P1183, DOI 10.1212/01.wnl.0000326598.83183.04
   Hardaway FA, 2020, J NEUROSURG, V132, P217, DOI 10.3171/2018.9.JNS181208
   JANNETTA PJ, 1967, J NEUROSURG, V26, P159, DOI 10.3171/jns.1967.26.1part2.0159
   Jia DZ, 2010, MED HYPOTHESES, V74, P505, DOI 10.1016/j.mehy.2009.09.056
   Jo KW, 2013, J CLIN NEUROSCI, V20, P440, DOI 10.1016/j.jocn.2012.03.037
   Kano H, 2010, NEUROSURGERY, V67, P1637, DOI 10.1227/NEU.0b013e3181fa098a
   Karam SD, 2014, J RADIOL ONCOL, V3, P125, DOI 10.1007/s13566-013-0134-3
   Karam Sana D, 2014, Radiat Oncol, V9, P257, DOI 10.1186/s13014-014-0257-8
   KATUSIC S, 1990, ANN NEUROL, V27, P89, DOI 10.1002/ana.410270114
   Kaufmann AM, 2020, J NEUROSURG, V132, P639, DOI 10.3171/2018.10.JNS181983
   Leal PRL, 2010, ACTA NEUROCHIR, V152, P817, DOI 10.1007/s00701-009-0588-7
   Massager N, 2004, J NEUROSURG, V100, P597, DOI 10.3171/jns.2004.100.4.0597
   Miller JP, 2009, J NEUROSURG, V110, P627, DOI 10.3171/2008.6.17620
   Obermann M, 2007, NEUROLOGY, V69, P835, DOI 10.1212/01.wnl.0000269670.30045.6b
   Obermann M, 2009, EXPERT REV NEUROTHER, V9, P323, DOI 10.1586/14737175.9.3.323
   Park KJ, 2012, NEUROSURGERY, V70, P295, DOI 10.1227/NEU.0b013e318230218e
   Patel SK, 2016, NEUROSURG CLIN N AM, V27, P265, DOI 10.1016/j.nec.2016.02.002
   Pollock BE, 2005, INT J RADIAT ONCOL, V61, P192, DOI 10.1016/j.ijrobp.2004.03.019
   Pollock BE, 2002, J NEUROSURG, V97, P347, DOI 10.3171/jns.2002.97.2.0347
   Sindou M, 2007, J NEUROSURG, V107, P1144, DOI 10.3171/JNS-07/12/1144
   Stavropoulou Evmorfia, 2014, ISRN Pain, V2014, P853826, DOI 10.1155/2014/853826
   Toda H, 2015, NEUROL MED-CHIR, V55, P432, DOI 10.2176/nmc.ra.2014-0393
   Xu ZY, 2019, NEUROSURGERY, V84, P499, DOI 10.1093/neuros/nyy142
   Zacest AC, 2010, J NEUROSURG, V113, P511, DOI 10.3171/2009.12.JNS09977
NR 37
TC 18
Z9 22
U1 2
U2 28
PU CURRENT MEDICINE GROUP
PI PHILADELPHIA
PA 400 MARKET STREET, STE 700, PHILADELPHIA, PA 19106 USA
SN 1092-8480
EI 1534-3138
J9 CURR TREAT OPTION NE
JI Curr. Treat. Options Neurol.
PD SEP
PY 2019
VL 21
IS 9
AR 42
DI 10.1007/s11940-019-0583-0
PG 11
WC Clinical Neurology
WE Science Citation Index Expanded (SCI-EXPANDED)
SC Neurosciences & Neurology
GA IM5SX
UT WOS:000478054700001
PM 31367794
DA 2023-08-27
ER

PT J
AU Kyrnetskiy, EE
   Kun, LE
   Boop, FA
   Sanford, RA
   Khan, RB
AF Kyrnetskiy, EE
   Kun, LE
   Boop, FA
   Sanford, RA
   Khan, RB
TI Types, causes, and outcome of intracranial hemorrhage in children with
   cancer
SO JOURNAL OF NEUROSURGERY
LA English
DT Article
DE intracranial hemorrhage; cancer; radiation treatment; pediatric
   neurosurgery
ID CLINICAL FACTORS; THERAPY; SURVIVORS; BRAIN
AB Object. The aim of this study was to investigate the cause and outcome of intracranial hemorrhage (ICH) in children with cancer.
   Methods. The charts of 51 children who underwent treatment for both cancer and ICH between January 1985 and January 2003 were retrospectively reviewed. Assessment tools included the Karnofsky Performance Scale (KPS), Glasgow Coma Scale (GCS), and the Fisher exact and Student t-tests.
   Among the 51 cases, 30 involved brain tumors, 19 leukemia, and two lymphoma. The treatment group (Group 1) comprised 36 patients who suffered ICH during cancer treatment; the posttreatment group (Group 2) consisted of the 15 patients who suffered ICH after the completion of cancer treatment. The types of ICH included 22 cortical, four subcortical, 17 subdural, five brainstem, one subarachnoid, one epidural, and one ventricular. Thrombocytopenia was present in nine patients (25%) in Group 1. More patients in Group 2 (87%) than in Group 1 (44%) underwent cranial radiation treatment. Patients in Group 1 experienced a higher incidence of coagulopathy (37%) and ICH-related death (25%) than those in Group 2 (0 and 7%, respectively). Decrease in KPS and GCS scores of greater than 30 and greater than 3, respectively, at the time of ICH were indicators of increased mortality. Of the 17 children with subdural ICH, 13 suffered the hemorrhage following treatment for hydrocephalus and three patients suffered ICH associated with thrombocytopenia. In the 33 children alive at the 3-month follow-up examination after the ICH, no difference existed in the mean KPS scores pre- and post-ICH.
   Conclusions. Treatment for hydrocephalus, coagulopathy, thrombocytopenia, and hemorrhage into the tumor were the most probable causes of ICH among patients in Group 1. Radiation-induced vasculopathy was a possible cause of ICH in the patients in Group 2. Significant decline in the patient's neurological status at the time of ICH is a poor prognostic factor, but those patients who survive cancer and ICH are likely to regain neurological function.
C1 St Jude Childrens Res Hosp, Dept Radiol Sci, Div Neurol, Memphis, TN 38105 USA.
C3 St Jude Children's Research Hospital
RP Khan, RB (通讯作者)，St Jude Childrens Res Hosp, Dept Radiol Sci, Div Neurol, 332 N Lauderdale St,Mail Stop 220, Memphis, TN 38105 USA.
EM
RI Khan, Raja B/N-8150-2018
OI Khan, Raja B/0000-0003-3702-7250
FU NCI NIH HHS [CA-21765] Funding Source: Medline
CR Aguiar PH, 2000, CHILD NERV SYST, V16, P218, DOI 10.1007/s003810050501
   ALLEN JC, 1991, NEUROLOGY, V41, P148, DOI 10.1212/WNL.41.1.148
   [Anonymous], 1949, EVALUATION CHEMOTHER
   COLLINS RC, 1975, NEUROLOGY, V25, P795, DOI 10.1212/WNL.25.9.795
   Hardemark HG, 1999, CEREBROVASC DIS, V9, P10, DOI 10.1159/000015890
   HOPPEHIRSCH E, 1987, CHILD NERV SYST, V3, P97, DOI 10.1007/BF00271133
   Meyer-Heim AD, 2003, BRAIN DEV-JPN, V25, P416, DOI 10.1016/S0387-7604(03)00029-9
   Novelli PM, 1997, PEDIATR NEUROSURG, V26, P322, DOI 10.1159/000121213
   Oishi M, 2001, NEUROL MED-CHIR, V41, P382, DOI 10.2176/nmc.41.382
   PAAKKO E, 1994, NEURORADIOLOGY, V36, P652
   PACKER RJ, 1985, PEDIATRICS, V76, P194
   PRIEST JR, 1982, J PEDIATR-US, V100, P984, DOI 10.1016/S0022-3476(82)80535-0
   Russo C, 1999, INT J RADIAT ONCOL, V44, P85, DOI 10.1016/S0360-3016(98)00514-8
   Sharma RR, 1999, PEDIATR NEUROSURG, V31, P150, DOI 10.1159/000028851
   TEASDALE G, 1974, LANCET, V2, P81
NR 15
TC 32
Z9 32
U1 0
U2 0
PU AMER ASSOC NEUROLOGICAL SURGEONS
PI ROLLING MEADOWS
PA 5550 MEADOWBROOK DRIVE, ROLLING MEADOWS, IL 60008 USA
SN 0022-3085
J9 J NEUROSURG
JI J. Neurosurg.
PD JAN
PY 2005
VL 102
IS 1
SU S
BP 31
EP 35
DI 10.3171/ped.2005.102.1.0031
PG 5
WC Clinical Neurology; Surgery
WE Science Citation Index Expanded (SCI-EXPANDED)
SC Neurosciences & Neurology; Surgery
GA 899VM
UT WOS:000227173800004
PM 16206731
DA 2023-08-27
ER

PT J
AU Ferber-Viart, C
   Colleaux, B
   Laoust, L
   Dubreuil, C
   Duclaux, R
AF Ferber-Viart, C
   Colleaux, B
   Laoust, L
   Dubreuil, C
   Duclaux, R
TI Is the presence of transient evoked otoacoustic emissions in ears with
   acoustic neuroma significant?
SO LARYNGOSCOPE
LA English
DT Article
ID FACIAL-NERVE FUNCTION; HEARING PRESERVATION; TUMORS; SURGERY
AB One hundred sixty eight patients admitted for acoustic neuroma removal were involved in this study. In all cases, the size of the tumor and the presence or absence of tinnitus and vertigo or dizziness were evaluated. Investigated functional procedures included bilateral measures of pure-tone audiometry (PTA), auditory brainstem response (ABR), electronystagmography (ENG), and transient evoked otoacoustic emissions (TEOAEs). Thirty-five patients (21%) had normal preoperative TEOAEs in the affected ear, whereas 133 patients (79%) failed to show reproducible responses, The aims of this study were 1. to evaluate what distinguishes patients with preoperative TEOAEs in the pathological ear (group A) from those who had no TEOAE (group B); and 2. to determine in cases of attempted hearing preservation whether preoperative TEOAE presence in the neuroma ear (group C) was predictive of postoperative hearing preservation compared with the group of patients without TEOAEs (group D). The presence of vertigo or dizziness was significantly less frequent, the age was lower, and preoperative mean PTA loss in both ears was lower in group A compared with group B. Frequency of the other studied parameters and ABR threshold were similar in both groups. When hearing preservation was attempted, the mean preoperative PTA loss of group C patients was lower in both ears compared with group D. However, postoperative mean PTA loss did not significantly differ in the two groups. In group C, the percentage of hearing preservation (66.6%) was significantly higher than the percentage of deafness (33.4%), whereas in group D the percentage of postoperative preserved audition and deafness did not significantly differ (respectively 44.4% and 55.6%). The findings suggest that 1. TEOAEs in ears with acoustic neuromas are found in younger patients with a lower preoperative mean PTA loss and are accompanied by fewer functional complaints, perhaps because preserved TEOAEs indicate a better preservation of inner ear vasculature; and, 2. along with radiological and electrophysiological investigations, TEOAE presence in the pathological ear could provide an additional criterion or predictive factor for the successful outcome of attempted hearing-conservation surgery in ears with acoustic neuromas.
C1 Univ Lyon 1, Ctr Hosp Lyon Sud, Lab Neurosci & Syst Sensoriels, F-69495 Pierre Benite, France.
   Hop Edouard Herriot, Lyon, France.
   Ctr Hosp, Federat Explorat Audiovestibulaires, Lyon, France.
C3 CHU Lyon; UDICE-French Research Universities; Universite Claude Bernard
   Lyon 1; CHU Lyon; CHU Lyon
RP Ferber-Viart, C (通讯作者)，Univ Lyon 1, Ctr Hosp Lyon Sud, Lab Neurosci & Syst Sensoriels, F-69495 Pierre Benite, France.
CR AOYAGI M, 1994, ACTA OTO-LARYNGOL, P40
   ATLAS MD, 1992, LARYNGOSCOPE, V102, P779, DOI 10.1288/00005537-199207000-00007
   BONFILS P, 1988, AM J OTOL, V9, P412
   BRAY P, 1987, British Journal of Audiology, V21, P191, DOI 10.3109/03005368709076405
   CANE MA, 1992, SCAND AUDIOL, V21, P173, DOI 10.3109/01050399209045999
   CANE MA, 1994, AM J OTOL, V15, P207
   CERULLO LJ, 1993, SURG NEUROL, V39, P485, DOI 10.1016/0090-3019(93)90036-Z
   CLEMIS JD, 1981, LARYNGOSCOPE, V91, P1792
   CLEMIS JD, 1976, ARCH OTOLARYNGOL, V102, P654, DOI 10.1001/archotol.1976.00780160050002
   COLLEAUX B, 1994, LYON CHIR, V90, P455
   FERBERVIART C, 1994, BRAIN DEV-JPN, V16, P213, DOI 10.1016/0387-7604(94)90072-8
   FerberViart C, 1995, HEARING RES, V91, P202, DOI 10.1016/0378-5955(95)00193-X
   FerberViart C, 1996, INT J NEUROSCI, V86, P207, DOI 10.3109/00207459608986711
   HOUSE WF, 1979, ACOUSTIC TUMOR MANAG, V2, P210
   KEMINK JL, 1990, LARYNGOSCOPE, V100, P597
   KEMP DT, 1978, J ACOUST SOC AM, V64, P1386, DOI 10.1121/1.382104
   KEMP DT, 1990, EAR HEARING, V11, P93, DOI 10.1097/00003446-199004000-00004
   LENOIR M, 1987, HEARING RES, V29, P265, DOI 10.1016/0378-5955(87)90173-0
   NADOL JB, 1992, LARYNGOSCOPE, V102, P1153, DOI 10.1288/00005537-199210000-00010
   Probst R, 1990, Adv Otorhinolaryngol, V44, P1
   Ren TY, 1995, HEARING RES, V92, P30, DOI 10.1016/0378-5955(95)00192-1
   ROBINETTE MS, 1992, HEAR J, V45, P32
   STERKERS JM, 1994, OTOLARYNG HEAD NECK, V110, P146, DOI 10.1177/019459989411000202
   TELISCHI FF, 1995, LARYNGOSCOPE, V105, P675, DOI 10.1288/00005537-199507000-00002
NR 24
TC 20
Z9 20
U1 0
U2 1
PU WILEY
PI HOBOKEN
PA 111 RIVER ST, HOBOKEN 07030-5774, NJ USA
SN 0023-852X
EI 1531-4995
J9 LARYNGOSCOPE
JI Laryngoscope
PD APR
PY 1998
VL 108
IS 4
BP 605
EP 609
DI 10.1097/00005537-199804000-00026
PN 1
PG 5
WC Medicine, Research & Experimental; Otorhinolaryngology
WE Science Citation Index Expanded (SCI-EXPANDED)
SC Research & Experimental Medicine; Otorhinolaryngology
GA ZF846
UT WOS:000072939100026
PM 9546278
DA 2023-08-27
ER

PT J
AU Carbonara, R
   Di Rito, A
   Monti, A
   Rubini, G
   Sardaro, A
AF Carbonara, Roberta
   Di Rito, Alessia
   Monti, Angela
   Rubini, Giuseppe
   Sardaro, Angela
TI Proton versus Photon Radiotherapy for Pediatric Central Nervous System
   Malignancies: A Systematic Review and Meta-Analysis of Dosimetric
   Comparison Studies
SO JOURNAL OF ONCOLOGY
LA English
DT Review
ID CRANIOSPINAL IRRADIATION; RADIATION-THERAPY; 2ND CANCERS; BRAIN; TUMORS;
   RISK; CHILDREN; OUTCOMES; MEDULLOBLASTOMA; BEAMS
AB Background. Radiotherapy (RT) plays a fundamental role in the treatment of pediatric central nervous system (CNS) malignancies, but its late sequelae are still a challenging question. Despite developments in modern high-conformal photon techniques and proton beam therapy (PBT) are improving the normal tissues dose-sparing while maintaining satisfactory target coverage, clinical advantages supporting the optimal treatment strategy have to be better evaluated in long-term clinical studies and assessed in further radiobiological analyses. Our analysis aimed to systematically review current knowledge on the dosimetric advantages of PBT in the considered setting, which should be the basis for future specific studies. Materials and Methods. A PubMed and Google Scholar search was conducted in June 2019 to select dosimetric studies comparing photon versus proton RT for pediatric patients affected by CNS tumors. Then, a systematic review and meta-analysis according to the PRISMA statement was performed. Average and standard deviation values of Conformity Index, Homogeneity Index, and mean and maximum doses to intracranial and extracranial organs at risk (OARs) were specifically evaluated for secondary dosimetric comparisons. The standardized mean differences (SMDs) for target parameters and the mean differences (MDs) for OARs were summarized in forest plots (P<0.05 was considered statistically significant). Publication bias was also assessed by the funnel plot and Egger's regression test. Results. Among the 88 identified papers, a total of twelve studies were included in the meta-analysis. PBT showed dosimetric advantages in target homogeneity (significant especially in the subgroup comparing PBT and 3D conformal RT), as well as in the dose sparing of almost all analyzed OARs (significantly superior results for brainstem, normal brain, and hippocampal dose constraints and for extracranial OARs parameters, excluding the kidneys). Publication bias was observed for Conformity Index. Conclusion. Our analysis supports the evidence of dosimetric advantages of PBT over photon RT, especially in the dose sparing of normal growing tissues. Confirmations from wider well-designed studies are required.
C1 [Carbonara, Roberta; Monti, Angela; Sardaro, Angela] Univ Bari Aldo Moro, Sect Radiol & Radiat Oncol, Interdisciplinary Dept Med, Bari, Italy.
   [Di Rito, Alessia] IRCCS Ist Tumori Giovanni Paolo II, Radiat Oncol Unit, Bari, Italy.
   [Rubini, Giuseppe] Univ Bari Aldo Moro, Nucl Med Sect, Interdisciplinary Dept Med, Bari, Italy.
C3 Universita degli Studi di Bari Aldo Moro; IRCCS Istituto Tumori Bari
   Giovanni Paolo II; Universita degli Studi di Bari Aldo Moro
RP Carbonara, R (通讯作者)，Univ Bari Aldo Moro, Sect Radiol & Radiat Oncol, Interdisciplinary Dept Med, Bari, Italy.
EM;
  
RI Di Rito, Alessia/AAI-1791-2020
OI Di Rito, Alessia/0000-0001-6694-5080; Monti, Angela/0000-0002-1952-9348;
   Rubini, Giuseppe/0000-0003-1972-3308
CR Armstrong GT, 2009, JNCI-J NATL CANCER I, V101, P946, DOI 10.1093/jnci/djp148
   Beltran Chris, 2012, Int J Radiat Oncol Biol Phys, V82, pe281, DOI 10.1016/j.ijrobp.2011.01.005
   Boehling NS, 2012, INT J RADIAT ONCOL, V82, P643, DOI 10.1016/j.ijrobp.2010.11.027
   Brodin NP, 2011, ACTA ONCOL, V50, P806, DOI 10.3109/0284186X.2011.582514
   Chung CS, 2013, INT J RADIAT ONCOL, V87, P46, DOI 10.1016/j.ijrobp.2013.04.030
   Constine LS, 2019, CLIN ONCOL-UK, V31, P199, DOI 10.1016/j.clon.2019.01.002
   Correia D, 2019, CLIN TRANSL RAD ONCO, V15, P53, DOI 10.1016/j.ctro.2019.01.002
   Dennis ER, 2013, TECHNOL CANCER RES T, V12, P1, DOI 10.7785/tcrt.2012.500276
   Eaton BR, 2014, CANCER J, V20, P403, DOI 10.1097/PPO.0000000000000079
   Fagerstrom Kristensen I., 2015, INT J PART THER, V2, P367, DOI [10.14338/IJPT-14-00026.1, DOI 10.14338/IJPT-14-00026.1]
   Freund D, 2015, CANCERS, V7, P617, DOI 10.3390/cancers7020617
   Ho ESQ, 2017, ACTA ONCOL, V56, P1031, DOI 10.1080/0284186X.2017.1324207
   Hoeben BA, 2019, LANCET ONCOL, V20, pE155, DOI 10.1016/S1470-2045(19)30034-8
   Howell RM, 2012, RADIAT ONCOL, V7, DOI 10.1186/1748-717X-7-116
   Kataria T, 2012, J MED PHYS, V37, P207, DOI 10.4103/0971-6203.103606
   Laprie A, 2015, CANCER RADIOTHER, V19, P775, DOI 10.1016/j.canrad.2015.05.028
   Leroy R, 2016, INT J RADIAT ONCOL, V95, P267, DOI 10.1016/j.ijrobp.2015.10.025
   Louis DN, 2016, ACTA NEUROPATHOL, V131, P803, DOI 10.1007/s00401-016-1545-1
   Ludmir Ethan B, 2018, Bioengineering (Basel), V5, DOI 10.3390/bioengineering5040097
   MacDonald SM, 2008, INT J RADIAT ONCOL, V71, P979, DOI 10.1016/j.ijrobp.2007.11.065
   Main C, 2016, SYST REV-LONDON, V5, DOI 10.1186/s13643-016-0285-6
   Marks LB, 2010, INT J RADIAT ONCOL, V76, pS10, DOI 10.1016/j.ijrobp.2009.07.1754
   Merchant TE, 2006, INT J RADIAT ONCOL, V65, P210, DOI 10.1016/j.ijrobp.2005.10.038
   Miralbell R, 2002, INT J RADIAT ONCOL, V54, P824, DOI 10.1016/S0360-3016(02)02982-6
   Mizumoto M, 2017, NEUROL MED-CHIR, V57, P343, DOI 10.2176/nmc.ra.2017-0003
   Moher D, 2015, SYST REV-LONDON, V4, DOI [10.1186/s13643-015-0087-2, 10.1016/j.ijsu.2010.02.007, 10.1016/j.ijsu.2010.07.299, 10.1136/bmj.b2535]
   Mu XK, 2005, ACTA ONCOL, V44, P554, DOI 10.1080/02841860500218819
   Newhauser WD, 2009, PHYS MED BIOL, V54, P2277, DOI 10.1088/0031-9155/54/8/002
   Ohtakara K, 2012, BRIT J RADIOL, V85, pE640, DOI 10.1259/bjr/21015703
   Padovani L, 2019, CLIN ONCOL-UK, V31, P171, DOI 10.1016/j.clon.2019.01.001
   Schneider U, 2002, INT J RADIAT ONCOL, V53, P244, DOI 10.1016/S0360-3016(01)02826-7
   Schneider U, 2015, FRONT ONCOL, V5, DOI 10.3389/fonc.2015.00235
   Scoccianti S, 2015, RADIOTHER ONCOL, V114, P230, DOI 10.1016/j.radonc.2015.01.016
   Seravalli E, 2018, ACTA ONCOL, V57, P1240, DOI 10.1080/0284186X.2018.1465588
   Steinmeier T., 2019, CLIN ONCOLOGY, V31, DOI [10.1016/j.clon.2018.12.005, DOI 10.1016/J.CLON.2018.12.0052-S2.0-85059675838]
   Stoker J, 2018, PHYS IMAG RADIAT ONC, V8, P28, DOI 10.1016/j.phro.2018.11.001
   Takizawa D, 2017, RADIAT ONCOL, V12, DOI 10.1186/s13014-017-0775-2
   Weber DC, 2018, RADIOTHER ONCOL, V128, P44, DOI 10.1016/j.radonc.2018.05.020
   Yoon M, 2011, INT J RADIAT ONCOL, V81, P637, DOI 10.1016/j.ijrobp.2010.06.039
   Zhang R, 2014, RADIOTHER ONCOL, V113, P84, DOI 10.1016/j.radonc.2014.07.003
NR 40
TC 12
Z9 12
U1 1
U2 3
PU HINDAWI LTD
PI LONDON
PA ADAM HOUSE, 3RD FLR, 1 FITZROY SQ, LONDON, W1T 5HF, ENGLAND
SN 1687-8450
EI 1687-8469
J9 J ONCOL
JI J. Oncol.
PD NOV 27
PY 2019
VL 2019
AR 5879723
DI 10.1155/2019/5879723
PG 17
WC Oncology
WE Science Citation Index Expanded (SCI-EXPANDED)
SC Oncology
GA JU6TD
UT WOS:000501805900002
PM 31885580
OA Green Published, gold
DA 2023-08-27
ER

PT J
AU Bai, XM
   Lim, G
   Grosshans, D
   Mohan, R
   Cao, WH
AF Bai, Xuemin
   Lim, Gino
   Grosshans, David
   Mohan, Radhe
   Cao, Wenhua
TI A biological effect-guided optimization approach using beam distal-edge
   avoidance for intensity-modulated proton therapy
SO MEDICAL PHYSICS
LA English
DT Article
DE biological effect; IMPT; LET; RBE
ID EFFECTIVENESS RBE VALUES; RANGE UNCERTAINTY; MODEL; ALGORITHM; POINT
AB Purpose Linear energy transfer (LET)-guided methods have been applied to intensity-modulated proton therapy (IMPT) to improve its biological effect. However, using LET as a surrogate for biological effect ignores the topological relationship of the scanning spot to different structures of interest. In this study, we developed an optimization method that takes advantage of the continuing increase in LET beyond the physical dose Bragg peak. This method avoids placing high biological effect values in critical structures and increases biological effect in the tumor area without compromising target coverage. Methods We selected the cases of two patients with brain tumors and two patients with head and neck tumors who had been treated with proton therapy at our institution. Three plans were created for each case: a plan based on conventional dose-based optimization (DoseOpt), one based on LET-incorporating optimization (LETOpt), and one based on the proposed distal-edge avoidance-guided optimization method (DEAOpt). In DEAOpt, anL(1)-norm sparsity term, in which the penalty of each scanning spot was set according to the topological relationship between the organ positions and the location of the peak scaled LET-weighted dose (c LETxD) was added to a conventional dose-based optimization objective function. All plans were normalized to give the same target dose coverage. Dose (assuming a constant relative biological effectiveness value of 1.1, as in clinical practice), biological effect (c LETxD), and computing time consumption were evaluated and compared among the three optimization approaches for each patient case. Results For all four cases, all three optimization methods generated comparable dose coverage in both target and critical structures. The LETOpt plans and DEAOpt plans reduced biological effect hot spots in critical structures and increased biological effect in the target volumes to a similar extent. For the target, the c LETxD(98%)and c LETxD(2%)in the DEAOpt plans were on average 7.2% and 11.74% higher than in the DoseOpt plans, respectively. For the brainstem, the c LETxD(mean)in the DEAOpt plans was on average 33.38% lower than in the DoseOpt plans. In addition, the DEAOpt method saved 30.37% of the computation cost over the LETOpt method. Conclusions DEAOpt is an alternative IMPT optimization approach that correlates the location of scanning spots with biological effect distribution. IMPT could benefit from the use of DEAOpt because this method not only delivers comparable biological effects to LETOpt plans, but also is faster.
C1 [Bai, Xuemin; Lim, Gino] Univ Houston, Dept Ind Engn, Houston, TX 77004 USA.
   [Bai, Xuemin] Linking Med Technol, Beijing 100085, Peoples R China.
   [Grosshans, David; Mohan, Radhe; Cao, Wenhua] Univ Texas MD Anderson Canc Ctr, Dept Radiat Oncol, Houston, TX 77030 USA.
C3 University of Houston System; University of Houston; University of Texas
   System; UTMD Anderson Cancer Center
RP Cao, WH (通讯作者)，Univ Texas MD Anderson Canc Ctr, Dept Radiat Oncol, Houston, TX 77030 USA.
EM
RI Lim, Gino/J-4607-2014
OI Lim, Gino/0000-0003-2259-2310
FU National Cancer Institute of the National Institutes of Health
   [2U19CA021239-35]; Cancer Prevention and Research Institute of Texas
   [RP160232]
FX This work was partly supported by the National Cancer Institute of the
   National Institutes of Health (2U19CA021239-35) and the Cancer
   Prevention and Research Institute of Texas (RP160232). Editorial support
   was provided by Amy Ninetto of Scientific Publications Services,
   Research Medical Library, The University of Texas MD Anderson Cancer
   Center.
CR An Y, 2017, MED PHYS, V44, P28, DOI [10.1002/mp.12001, 10.1002/mp.12610]
   [Anonymous], 2010, J ICRU, V10, P7
   Bai XM, 2019, PHYS MED BIOL, V64, DOI 10.1088/1361-6560/aaf5e9
   Bassler N, 2010, ACTA ONCOL, V49, P1170, DOI 10.3109/0284186X.2010.510640
   Bortfeld T, 1999, SEMIN RADIAT ONCOL, V9, P20, DOI 10.1016/S1053-4296(99)80052-6
   Cao WH, 2018, PHYS MED BIOL, V63, DOI 10.1088/1361-6560/aa9a2e
   Carabe-Fernandez A, 2011, MED PHYS, V38, P3639, DOI 10.1118/1.3612611
   Carabe-Fernandez A, 2007, INT J RADIAT BIOL, V83, P27, DOI 10.1080/09553000601087176
   Chen W, 2012, PHYS MED BIOL, V57, P591, DOI 10.1088/0031-9155/57/3/591
   Fager M, 2015, INT J RADIAT ONCOL, V91, P1057, DOI 10.1016/j.ijrobp.2014.12.049
   Giantsoudi D, 2017, INT J RADIAT ONCOL, V99, pS106, DOI 10.1016/j.ijrobp.2017.06.253
   Giantsoudi D, 2013, INT J RADIAT ONCOL, V87, P216, DOI 10.1016/j.ijrobp.2013.05.013
   Giovannini G, 2016, RADIAT ONCOL, V11, DOI 10.1186/s13014-016-0642-6
   Hong L, 1996, PHYS MED BIOL, V41, P1305, DOI 10.1088/0031-9155/41/8/005
   Inaniwa T, 2017, PHYS MED BIOL, V62, P5180, DOI 10.1088/1361-6560/aa68d7
   Kohno R, 2019, INT J MED PHYS CLIN, V08, P32
   Lomax A, 1999, PHYS MED BIOL, V44, P185, DOI 10.1088/0031-9155/44/1/014
   Lomax AJ, 2001, MED PHYS, V28, P317, DOI 10.1118/1.1350587
   McNamara AL, 2015, PHYS MED BIOL, V60, P8399, DOI 10.1088/0031-9155/60/21/8399
   Oelfke U, 2001, Med Dosim, V26, P113, DOI 10.1016/S0958-3947(01)00057-7
   Paganetti H, 2002, INT J RADIAT ONCOL, V53, P407, DOI 10.1016/S0360-3016(02)02754-2
   Paganetti H, 2014, PHYS MED BIOL, V59, pR419, DOI 10.1088/0031-9155/59/22/R419
   Traneus E, 2019, INT J RADIAT ONCOL, V103, P747, DOI 10.1016/j.ijrobp.2018.10.031
   Tseung HSWC, 2016, INT J RADIAT ONCOL, V95, P1535, DOI 10.1016/j.ijrobp.2016.03.041
   Unkelbach J, 2016, INT J RADIAT ONCOL, V96, P1097, DOI 10.1016/j.ijrobp.2016.08.038
   Wachter A, 2006, MATH PROGRAM, V106, P25, DOI 10.1007/s10107-004-0559-y
   Wang QX, 2018, BIOMED PHYS ENG EXPR, V4, DOI 10.1088/2057-1976/aae039
   Wedenberg M, 2013, ACTA ONCOL, V52, P580, DOI 10.3109/0284186X.2012.705892
   Wieser HP, 2017, MED PHYS, V44, P2556, DOI 10.1002/mp.12251
   Wilkens JJ, 2003, MED PHYS, V30, P806, DOI 10.1118/1.1567852
NR 30
TC 7
Z9 7
U1 1
U2 3
PU WILEY
PI HOBOKEN
PA 111 RIVER ST, HOBOKEN 07030-5774, NJ USA
SN 0094-2405
EI 2473-4209
J9 MED PHYS
JI Med. Phys.
PD SEP
PY 2020
VL 47
IS 9
BP 3816
EP 3825
DI 10.1002/mp.14335
EA JUL 2020
PG 10
WC Radiology, Nuclear Medicine & Medical Imaging
WE Science Citation Index Expanded (SCI-EXPANDED)
SC Radiology, Nuclear Medicine & Medical Imaging
GA NT0IY
UT WOS:000547742200001
PM 32557747
DA 2023-08-27
ER

PT J
AU LENARZ, T
   ERNST, A
AF LENARZ, T
   ERNST, A
TI INTRAOPERATIVE MONITORING BY TRANSTYMPANIC ELECTROCOCHLEOGRAPHY AND
   BRAIN-STEM ELECTRICAL RESPONSE AUDIOMETRY IN ACOUSTIC NEUROMA SURGERY
SO EUROPEAN ARCHIVES OF OTO-RHINO-LARYNGOLOGY
LA English
DT Article
DE ACOUSTIC NEUROMA SURGERY; PRESERVATION OF HEARING; INTRAOPERATIVE
   MONITORING; ELECTROCOCHLEOGRAPHY; BRAIN-STEM ELECTRICAL RESPONSE
   AUDIOMETRY
ID AUDITORY EVOKED-POTENTIALS; POSTERIOR-FOSSA SURGERY; HEARING
   PRESERVATION; CEREBELLOPONTINE ANGLE; SUBOCCIPITAL REMOVAL; NERVE
   FUNCTION; TUMOR SURGERY; MANAGEMENT
AB The preservation of hearing is a major aim of contemporary temporal bone surgery. Our present findings demonstrate that intraoperative monitoring is a key method for attaining serviceable postoperative hearing after the removal of an acoustic neuroma. Both electrocochleography (ECoG) and brainstem electrical response audiometry were performed in 96 patients operated on for acoustic neuromas. The specificity of the different monitoring methods was affected by surgical manipulations in addition to such non-specific influences as CSF drainage, core body temperature and anesthesia. In the present study ECoG was found to be more reliable in assessing the intra- and postoperative course with respect to the preservation of cochlear function.
RP LENARZ, T (通讯作者)，UNIV TUBINGEN,DEPT OTOLARYNGOL,SILCHERSTR 5,W-7400 TUBINGEN 1,GERMANY.
CR COHEN NL, 1986, ANN OTO RHINOL LARYN, V95, P21, DOI 10.1177/000348948609500105
   EGGERMONT JJ, 1986, HDB SENSORY PHYSL, V5, P625
   GIBSON WPR, 1977, AUDIOLOGY, V16, P389
   GIBSON WPR, 1988, OTOLARYNGOL HEAD NEC, V5, P170
   HARNER SG, 1984, LARYNGOSCOPE, V94, P1431
   HOHMANN D, 1988, LARYN RHINOL OTOL, V67, P611
   HOHMANN D, 1991, AM J OTOL, V12, P172
   KANZAKI J, 1989, ACTA OTO-LARYNGOL, V107, P474, DOI 10.3109/00016488909127543
   KEMINK JL, 1990, LARYNGOSCOPE, V100, P597
   KVETON JF, 1990, LARYNGOSCOPE, V100, P473
   KVETON JF, 1990, LARYNGOSCOPE, V100, P1171
   LAMBERT PR, 1988, OTOLARYNG HEAD NECK, V98, P575, DOI 10.1177/019459988809800607
   LENARZ T, 1985, ACTA NEUROCHIR, V78, P21, DOI 10.1007/BF01809236
   LENARZ T, 1988, LARYN RHINOL OTOL, V76, P123
   LENARZ T, IN PRESS ACOUSTIC NE
   LENARZ T, 1991, EUR ARCH OTO-RHINO-L, V248, P258
   LENARZ T, 1989, SPR M SPEECH HEAR SC
   LLOYDTHOMAS AR, 1990, BRIT J ANAESTH, V65, P306, DOI 10.1093/bja/65.3.306
   MOLLER AR, 1989, NEUROSURGERY, V24, P257
   MOLLER AR, 1984, OTOLARYNG HEAD NECK, V92, P434
   MOLLER AR, 1988, EVOKED POTPENTIALS I, P111
   NADOL JB, 1987, LARYNGOSCOPE, V97, P1287, DOI 10.1288/00005537-198711000-00007
   OJEMANN RG, 1984, J NEUROSURG, V61, P938, DOI 10.3171/jns.1984.61.5.0938
   PRASS RL, 1987, OTOLARYNG HEAD NECK, V97, P343, DOI 10.1177/019459988709700401
   RAUDZENS PA, 1982, J NEUROSURG, V57, P341, DOI 10.3171/jns.1982.57.3.0341
   SCHRAMM J, 1989, ACTA NEUROCHIR, V98, P9, DOI 10.1007/BF01407170
   SCHRAMM J, 1985, HNO, V33, P495
   SILVERSTEIN H, 1986, OTOLARYNG HEAD NECK, V95, P285, DOI 10.1177/01945998860953P104
   SMITH MFW, 1985, AM J OTOL, P161
   STRAUSS C, 1989, HNO, V37, P281
   TOS M, 1991, ACTA OTO-LARYNGOL, V111, P616, DOI 10.3109/00016489109138392
   WATANABE E, 1989, ACTA NEUROCHIR, V98, P118, DOI 10.1007/BF01407337
   YINGLING CD, IN PRESS OTOLARYNGOL
NR 33
TC 18
Z9 18
U1 0
U2 0
PU SPRINGER VERLAG
PI NEW YORK
PA 175 FIFTH AVE, NEW YORK, NY 10010
SN 0937-4477
J9 EUR ARCH OTO-RHINO-L
JI Eur. Arch. Oto-Rhino-Laryn.
PD AUG
PY 1992
VL 249
IS 5
BP 257
EP 262
DI 10.1007/BF00714488
PG 6
WC Otorhinolaryngology
WE Science Citation Index Expanded (SCI-EXPANDED)
SC Otorhinolaryngology
GA JK171
UT WOS:A1992JK17100005
PM 1524806
DA 2023-08-27
ER

PT J
AU Adachi, K
   Hasegawa, M
   Hayakawa, M
   Tateyama, S
   Hirose, Y
AF Adachi, Kazuhide
   Hasegawa, Mituhiro
   Hayakawa, Motoharu
   Tateyama, Shinichiro
   Hirose, Yuichi
TI Susceptibility-Weighted Imaging of Deep Venous Congestion in Petroclival
   Meningioma
SO WORLD NEUROSURGERY
LA English
DT Article
DE Anterior transpetrosal approach; Deep venous congestion; Petroclival
   meningioma; Skull base meningioma; Susceptibility-weighted imaging
ID CEREBRAL MICROBLEEDS; DRAINAGE PATTERNS; PETROSECTOMY; EXPERIENCE;
   RESECTION; CONTRAST; LESIONS; GLIOMA; VEINS
AB BACKGROUND: Protecting the venous drainage route during surgery in cases of petroclival meningioma (PCM) is important. Identifying venous congestion preoperatively can be valuable in reducing the risks associated with venous congestion during surgery. In this study, we examined the utility of susceptibility-weighted imaging (SWI) in identifying the presence of venous congestion in PCM cases preoperatively and identified the factors associated with it.
   METHODS: We retrospectively examined 24 patients who had undergone surgery for primary PCM. The areas of the basal and internal cerebral veins on the affected and unaffected sides, obtained using SWI, were compared to identify venous congestion. We further examined the association between multiple candidate factors that are thought to be related to venous congestion and venous congestion using statistical analyses.
   RESULTS: SWI could successfully identify venous congestion in 11 of 24 PCM cases. Among the 12 factors examined, those associated with venous congestion were an extension of the tumor, over the midline or upward, which is known to disturb the venous flow at the brainstem surface; anastomosis of the superficial cerebral vein (i.e., bypass route for venous congestion); and a high ABC Surgical Risk Scale score, an indicator of postoperative neurologic deterioration.
   CONCLUSIONS: We showed that SWI is useful for evaluating venous congestion in PCM cases preoperatively and for identifying factors reflecting the risk of venous congestion. Taken together, our findings provide a multi-modal strategy for the preoperative prediction of venous congestion, which could facilitate the treatment of PCM.
C1 [Adachi, Kazuhide; Hasegawa, Mituhiro; Hayakawa, Motoharu; Tateyama, Shinichiro; Hirose, Yuichi] Fujita Hlth Univ, Sch Med, Dept Neurosurg, Toyoake, Aichi, Japan.
C3 Fujita Health University
RP Adachi, K (通讯作者)，Fujita Hlth Univ, Sch Med, Dept Neurosurg, Toyoake, Aichi, Japan.
EM
RI Hirose, Yuichi/P-1731-2019
OI Hirose, Yuichi/0000-0002-1305-1830; ADACHI, KAZUHIDE/0000-0002-5275-8572
CR Adachi K, 2018, WORLD NEUROSURG, V116, pE611, DOI 10.1016/j.wneu.2018.05.052
   Adachi K, 2017, NEUROL MED-CHIR, V57, P505, DOI 10.2176/nmc.ra.2016-0336
   Adachi K, 2016, WORLD NEUROSURG, V92, P339, DOI 10.1016/j.wneu.2016.05.019
   Adachi K, 2009, J NEUROSURG, V111, P1053, DOI 10.3171/2007.11.17446
   ALMEFTY O, 1988, NEUROSURGERY, V22, P510, DOI 10.1227/00006123-198803000-00010
   Almefty R, 2014, J NEUROSURG, V120, P40, DOI 10.3171/2013.8.JNS13535
   [Anonymous], 1999, NEUROSURGERY
   Ashwal S, 2010, DEV NEUROSCI-BASEL, V32, P343, DOI 10.1159/000316801
   Bambakidis NC, 2007, NEUROSURGERY, V61, P202, DOI [10.1227/01.neu.0000303218.61230.39, 10.1227/01.NIEU.0000280119.36270.BA]
   Bian W, 2014, NEURORADIOLOGY, V56, P91, DOI 10.1007/s00234-013-1297-8
   Colbert CA, 2010, RADIOLOGY, V256, P898, DOI 10.1148/radiol.10091842
   Di Ieva A, 2013, NEURORADIOLOGY, V55, P35, DOI 10.1007/s00234-012-1081-1
   Di Ieva A, 2012, WORLD NEUROSURG, V77, DOI 10.1016/j.wneu.2011.09.006
   Di Ieva A, 2011, NEUROIMAGE, V54, P74, DOI 10.1016/j.neuroimage.2010.07.045
   DiMeco F, 2004, NEUROSURGERY, V55, P1263, DOI 10.1227/01.NEU.0000143373.74160.F2
   Gasparetto E L, 2011, AJNR Am J Neuroradiol, V32, pE54, DOI 10.3174/ajnr.A2457
   Grabner G, 2012, MAGN RESON IMAGING, V30, P139, DOI 10.1016/j.mri.2011.08.004
   Haacke EM, 2009, AM J NEURORADIOL, V30, P19, DOI 10.3174/ajnr.A1400
   Haacke EM, 2004, MAGN RESON MED, V52, P612, DOI 10.1002/mrm.20198
   Hafez A, 2011, J NEUROSURG, V114, P1294, DOI 10.3171/2010.6.JNS091461
   Ichimura S, 2012, NEUROSURG REV, V35, P609, DOI 10.1007/s10143-012-0405-2
   KAWASE T, 1994, ACTA NEUROCHIR, V129, P113, DOI 10.1007/BF01406489
   Koerbel A, 2009, EJSO-EUR J SURG ONC, V35, P773, DOI 10.1016/j.ejso.2008.02.015
   Letourneau-Guillon L, 2012, AM J NEURORADIOL, V33, P301, DOI 10.3174/ajnr.A2777
   Li CT, 2010, EUR J RADIOL, V75, pE81, DOI 10.1016/j.ejrad.2009.08.003
   Little KM, 2005, NEUROSURGERY, V56, P546, DOI 10.1227/01.NEU.0000153906.12640.62
   Matsushima S, 2018, J COMPUT ASSIST TOMO, V42, P100, DOI 10.1097/RCT.0000000000000637
   Noguchi K, 2010, AM J NEURORADIOL, V31, P1903, DOI 10.3174/ajnr.A2231
   PADGET DH, 1956, AM J ANAT, V98, P307, DOI 10.1002/aja.1000980302
   Park JH, 2009, J KOREAN NEUROSURG S, V46, P365, DOI 10.3340/jkns.2009.46.4.365
   Rhoton AL, 2000, NEUROSURGERY, V47, pS69, DOI 10.1097/00006123-200009001-00012
   Saini J, 2009, AJNR Am J Neuroradiol, V30, pE6, DOI 10.3174/ajnr.A1265
   Sehgal V, 2006, J MAGN RESON IMAGING, V24, P41, DOI 10.1002/jmri.20598
   Sekhar LN, 1999, NEUROSURGERY, V44, P537, DOI 10.1097/00006123-199903000-00060
   Shibao S, 2016, J NEUROSURG, V124, P432, DOI 10.3171/2015.1.JNS141854
   Shibao S, 2015, WORLD NEUROSURG, V84, P574, DOI 10.1016/j.wneu.2015.03.055
   Siddiqui FM, 2015, STROKE, V46, P1263, DOI 10.1161/STROKEAHA.114.007465
   SPETZLER RF, 1992, J NEUROSURG, V76, P588, DOI 10.3171/jns.1992.76.4.0588
   Wang Q, 2014, MEDICINE, V93, DOI 10.1097/MD.0000000000000191
NR 39
TC 4
Z9 4
U1 0
U2 0
PU ELSEVIER SCIENCE INC
PI NEW YORK
PA 360 PARK AVE SOUTH, NEW YORK, NY 10010-1710 USA
SN 1878-8750
EI 1878-8769
J9 WORLD NEUROSURG
JI World Neurosurg.
PD FEB
PY 2019
VL 122
BP E20
EP E31
DI 10.1016/j.wneu.2018.08.218
PG 12
WC Clinical Neurology; Surgery
WE Science Citation Index Expanded (SCI-EXPANDED)
SC Neurosciences & Neurology; Surgery
GA HJ6UM
UT WOS:000457328100004
PM 30236813
DA 2023-08-27
ER

PT J
AU Grayeli, AB
   Kalamarides, M
   Fraysse, B
   Deguine, O
   Favre, G
   Martin, C
   Mom, T
   Sterkers, O
AF Grayeli, AB
   Kalamarides, M
   Fraysse, B
   Deguine, O
   Favre, G
   Martin, C
   Mom, T
   Sterkers, O
TI Comparison between intraoperative observations and electromyographic
   monitoring data for facial nerve outcome after vestibular schwannoma
   surgery
SO ACTA OTO-LARYNGOLOGICA
LA English
DT Article
DE acoustic neurinoma; facial function; facial monitoring; surgery
ID CEREBELLOPONTINE ANGLE SURGERY; ACOUSTIC NEUROMA; RESECTION; STIMULUS
AB Conclusions. A four-channel device may enhance the sensitivity of electromyography (EMG). Determination of stimulation thresholds (STs) below 0.05 mA improves facial prognostic information after vestibular schwannoma ( VS) surgery. Objective. To compare intraoperative observations with electromyographic data for predicting the immediate facial function outcome after VS surgery. Material and methods. A total of 111 VS patients operated on and intraoperatively monitored using four-channel facial EMG between October 2002 and September 2003 were included in this multicentre, prospective study. Muscular activity detection was performed in the frontal, orbicularis oculi, orbicularis oris and platysma muscles. Intraoperative observations were made concerning adhesion and nerve stretch, and facial function was graded (House-Brackmann classification) at postoperative Days 1 and 8. Results. Facial function at postoperative Days 1 and 8 was related to the intraoperative nerve STs at the brainstem and in the adhesion zone (range 0.01-3 mA for a response >100 mu V). At postoperative Day 8, good facial function (Grades 1 or 2) was observed in 93% of cases for STs in the adhesion zone of 0.01-0.04 mA, 85% for STs of 0.05-0.3 mA and 79% for STs >0.3 mA. These STs were related to the degree of tumor adhesion and not to the nerve stretch. The maximal EMG response was detected in the frontal or platysma muscles in 26% of cases, and in the orbicularis oris and orbicularis oculi muscles in 74%.
C1 Hop Beaujon, AP HP, Dept Otorhinolaryngol Head & Neck Surg, FR-92110 Clichy, France.
   Univ Paris 07, Fac Xavier Bichat, INSERM, EMI 0112, Paris, France.
   Hop Beaujon, Dept Neurosurg, Clichy, France.
   Hop Purpan, Dept Otolaryngol Head & Neck Surg, Toulouse, France.
   Hop Bellevue, Dept Otolaryngol Head & Neck Surg, St Etienne, France.
   CHU Clermont Ferrand, Clermont Ferrand, France.
C3 Assistance Publique Hopitaux Paris (APHP); Hopital Universitaire
   Paul-Brousse - APHP; UDICE-French Research Universities; Universite
   Paris Cite; Hopital Universitaire Beaujon - APHP; Hopital Universitaire
   Bichat-Claude Bernard - APHP; Hopital Universitaire Hotel-Dieu - APHP;
   Institut National de la Sante et de la Recherche Medicale (Inserm);
   UDICE-French Research Universities; Universite Paris Cite; UDICE-French
   Research Universities; Universite Paris Cite; Assistance Publique
   Hopitaux Paris (APHP); Hopital Universitaire Beaujon - APHP; CHU de
   Toulouse; Universite de Toulouse; Universite Toulouse III - Paul
   Sabatier; CHU de St Etienne; CHU Clermont Ferrand
RP Grayeli, AB (通讯作者)，Hop Beaujon, Serv ORL, 100 Bd Gen Leclerc, FR-92110 Clichy, France.
EM
RI MOM, Thierry/L-5809-2015; DEGUINE, Olivier G/A-6999-2011
OI MOM, Thierry/0000-0002-2229-2337
CR Axon PR, 1999, LARYNGOSCOPE, V109, P922, DOI 10.1097/00005537-199906000-00015
   Goldbrunner RH, 2000, NEUROSURGERY, V46, P1140, DOI 10.1097/00006123-200005000-00023
   Grayeli AB, 2005, OTOL NEUROTOL, V26, P114, DOI 10.1097/00129492-200501000-00021
   Hone SW, 1997, J OTOLARYNGOL, V26, P374
   Isaacson B, 2003, OTOL NEUROTOL, V24, P812, DOI 10.1097/00129492-200309000-00020
   Kombos T, 2000, NEUROL MED-CHIR, V40, P501, DOI 10.2176/nmc.40.501
   Magliulo G, 1997, J LARYNGOL OTOL, V111, P715, DOI 10.1017/S0022215100138447
   Nakao Y, 2002, OTOL NEUROTOL, V23, P93, DOI 10.1097/00129492-200201000-00020
   Nissen AJ, 1997, AM J OTOL, V18, P249
   Romstock J, 2000, J NEUROSURG, V93, P586, DOI 10.3171/jns.2000.93.4.0586
   Sobottka SB, 1998, ACTA NEUROCHIR, V140, P235, DOI 10.1007/s007010050090
   Zeitouni AG, 1997, AM J OTOL, V18, P494
NR 12
TC 14
Z9 14
U1 0
U2 3
PU TAYLOR & FRANCIS LTD
PI ABINGDON
PA 2-4 PARK SQUARE, MILTON PARK, ABINGDON OR14 4RN, OXON, ENGLAND
SN 0001-6489
EI 1651-2251
J9 ACTA OTO-LARYNGOL
JI Acta Oto-Laryngol.
PD OCT
PY 2005
VL 125
IS 10
BP 1069
EP 1074
DI 10.1080/00016480510038608
PG 6
WC Otorhinolaryngology
WE Science Citation Index Expanded (SCI-EXPANDED)
SC Otorhinolaryngology
GA 969AS
UT WOS:000232206400008
PM 16298788
DA 2023-08-27
ER

PT J
AU Rodier, PM
   Ingram, JL
   Tisdale, B
   Nelson, S
   Romano, J
AF Rodier, PM
   Ingram, JL
   Tisdale, B
   Nelson, S
   Romano, J
TI Embryological origin for autism: Developmental anomalies of the cranial
   nerve motor nuclei
SO JOURNAL OF COMPARATIVE NEUROLOGY
LA English
DT Article
DE teratology; valproic acid; brain stem; neural tube; Hox genes
ID EARLY INFANTILE-AUTISM; BRAIN-STEM; VALPROIC ACID; MOEBIUS SYNDROME;
   RETINOIC ACID; CELL COUNTS; RAT; TIME; NEURONS; MOUSE
AB The underlying brain injury that leads to autism has been difficult to identify. The diagnostic criteria of the disease are not readily associated with any brain region or system, nor are they mimicked by vascular accidents, tumors, or degenerative neurological diseases occurring in adults. Fortuitously, a recent report of autism induced by thalidomide exposure provides evidence that the disease originates by an injury at the time of closure of the neural tube. The human data suggest that the initiating lesion includes the motor cranial nerve nuclei. To test this hypothesis, we first examined motor nuclei in the brainstem of a human autistic case. The autopsy brain exhibited near-complete absence of the facial nucleus and superior olive along with shortening of the brainstem between the trapezoid body and the inferior olive. A similar deficit has been reported in Hoxa-1 gene knockout mice in which pattern formation of the hindbrain is disrupted during neurulation. Alternatively, exposure to antimitotic agents just after neural tube closure could produce the observed pattern of deficits. Thus, the lesions observed in the autopsy case appear to match those predicted by the thalidomide cases in both time of origin and central nervous system (CNS) location. To produce similar brain lesions experimentally, we exposed rat embryos to valproic acid, a second teratogen newly linked to autism. Dams received 350 mg/kg of valproic acid (VPA) on day 11.5 (the day of neural tube closure), day 12, or day 12.5 of gestation. Each treatment significantly reduced the number of motor neurons counted in matched sections of the earliest-forming motor nuclei (V, XII), and progressively later exposures affected tire VIth and IIIrd cranial nerve nuclei. All treatments spared the facial nucleus, which forms still later. Counts from the mesencephalic nucleus of trigeminal, the dorsal motor nucleus of the vagus, and the locus ceruleus were not affected by exposure to VPA, even though these nuclei form during the period when exposure occurred. Despite its effects on the motor nuclei, valproic acid exposure did not alter the further development of the brain in any obvious way. Treated animals were robust and had no external malformations. The autopsy data and experimental data from rats confirm that CNS injuries occurring during or just after neural tube closure can lead to a selective loss of neurons derived from the basal plate of the rhombencephalon. The results add two new lines of evidence that place the initiating injury for autism around the time of neural tube closure. (C) 1996 Wiley-Liss, Inc.
C1 UNIV ROCHESTER, SCH MED, DEPT PSYCHIAT, ROCHESTER, NY 14642 USA.
C3 University of Rochester
RP Rodier, PM (通讯作者)，UNIV ROCHESTER, SCH MED, DEPT OBSTET & GYNECOL, BOX 668, 601 ELMWOOD AVE, ROCHESTER, NY 14642 USA.
CR ALTMAN J, 1980, J COMP NEUROL, V194, P37, DOI 10.1002/cne.901940103
   ALTMAN J, 1980, J COMP NEUROL, V194, P1, DOI 10.1002/cne.901940102
   ALTMAN J, 1980, J COMP NEUROL, V194, P905, DOI 10.1002/cne.901940411
   ALTMAN J, 1978, J COMP NEUROL, V179, P23, DOI 10.1002/cne.901790104
   ANDREOLI J, 1973, AM J ANAT, V137, P87, DOI 10.1002/aja.1001370107
   [Anonymous], NEUROBIOLOGY AUTISM
   ARDINGER HH, 1988, AM J MED GENET, V29, P171, DOI 10.1002/ajmg.1320290123
   BAUMAN M, 1985, NEUROLOGY, V35, P866, DOI 10.1212/WNL.35.6.866
   BAUMAN ML, 1986, NEUROLOGY, V36, P190
   BAYER SA, 1993, NEUROTOXICOLOGY, V14, P83
   BINKERD PE, 1988, FUND APPL TOXICOL, V11, P485, DOI 10.1016/0272-0590(88)90112-1
   BOHN MC, 1980, NEUROSCIENCE, V5, P2003, DOI 10.1016/0306-4522(80)90045-7
   BOHN MC, 1978, DEV NEUROSCI-BASEL, V1, P250, DOI 10.1159/000112579
   BOUGHMAN JA, 1993, PERSPECTIVES PEDIAT, V4, P123
   BRUCE A, 1892, ILLUSTRATIONS NERVE
   BRYSON SE, 1988, J CHILD PSYCHOL PSYC, V29, P433, DOI 10.1111/j.1469-7610.1988.tb00735.x
   CARPENTER EM, 1993, DEVELOPMENT, V118, P1063
   CESARONI L, 1991, J AUTISM DEV DISORD, V21, P303, DOI 10.1007/BF02207327
   CHISAKA O, 1992, NATURE, V355, P516, DOI 10.1038/355516a0
   CHRISTIANSON AL, 1994, DEV MED CHILD NEUROL, V36, P361
   COGGESHALL RE, 1990, J NEUROSCI METH, V35, P187, DOI 10.1016/0165-0270(90)90123-W
   COLEMAN PD, 1985, J AUTISM DEV DISORD, V15, P245, DOI 10.1007/BF01531496
   COLLINS MD, 1991, TERATOLOGY, V44, P617, DOI 10.1002/tera.1420440604
   COURCHESNE E, 1994, NEUROLOGY, V44, P214, DOI 10.1212/WNL.44.2.214
   COURCHESNE E, 1988, NEW ENGL J MED, V318, P1349, DOI 10.1056/NEJM198805263182102
   DAVIGNON M, 1964, ARCHIV OTOLARYNGOL, V80, P136, DOI 10.1001/archotol.80.2.136
   DeArmond S. J., 1989, STRUCTURE HUMAN BRAI
   DELONG GR, 1981, ARCH NEUROL-CHICAGO, V38, P191, DOI 10.1001/archneur.1981.00510030085013
   DEMPSEY EE, 1983, TERATOLOGY, V28, P461, DOI 10.1002/tera.1420280318
   EHLERS K, 1992, TERATOLOGY, V46, P117, DOI 10.1002/tera.1420460205
   GAFFNEY GR, 1988, BIOL PSYCHIAT, V24, P578, DOI 10.1016/0006-3223(88)90168-0
   GAFFNEY GR, 1987, AM J DIS CHILD, V141, P1330, DOI 10.1001/archpedi.1987.04460120096044
   GILLBERG C, 1991, BRIT J PSYCHIAT, V158, P403, DOI 10.1192/bjp.158.3.403
   GILLBERG C, 1987, DEV MED CHILD NEUROL, V29, P641
   GILLBERG C, 1989, ACTA PAEDIATR SCAND, V78, P314, DOI 10.1111/j.1651-2227.1989.tb11076.x
   GILLBERG C, 1987, J AUTISM DEV DISORD, V17, P273, DOI 10.1007/BF01495061
   GILLBERG C, 1986, J AUTISM DEV DISORD, V16, P369, DOI 10.1007/BF01531665
   GILLES FH, 1969, ARCH PATHOL, V88, P32
   HARKMARK W, 1954, J COMP NEUROL, V100, P115, DOI 10.1002/cne.901000107
   HASHIMOTO T, 1995, J AUTISM DEV DISORD, V25, P1, DOI 10.1007/BF02178163
   HAYES RW, 1977, LANCET, V2, P767
   HO KC, 1980, ARCH PATHOL LAB MED, V104, P635
   HOLROYD S, 1991, BIOL PSYCHIAT, V29, P287, DOI 10.1016/0006-3223(91)91291-X
   HOTCHKISS MG, 1980, ARCH OPHTHALMOL-CHIC, V98, P870, DOI 10.1001/archopht.1980.01020030864013
   HSU M, 1991, ARCH NEUROL-CHICAGO, V48, P1160, DOI 10.1001/archneur.1991.00530230068024
   JOUBERT M, 1969, NEUROLOGY, V19, P813, DOI 10.1212/WNL.19.9.813
   KEMPER TL, 1992, INT CONGR SER, V965, P43
   KESSEL M, 1993, NEURON, V10, P379, DOI 10.1016/0896-6273(93)90328-O
   KLEIMAN MD, 1992, NEUROLOGY, V42, P753, DOI 10.1212/WNL.42.4.753
   KLIN A, 1993, J AUTISM DEV DISORD, V23, P15, DOI 10.1007/BF01066416
   Lemire R., 1975, NORMAL ABNORMAL DEV
   LENZ W, 1962, LANCET, V1, P271
   LIPSON AH, 1989, TERATOLOGY, V40, P339, DOI 10.1002/tera.1420400406
   LUFKIN T, 1991, CELL, V66, P1105, DOI 10.1016/0092-8674(91)90034-V
   MARK M, 1993, DEVELOPMENT, V119, P319
   MAY M, 1986, FACIAL NERVE, P404
   Miller M. T., 1993, Teratology, V47, P387
   Miller M T, 1991, Trans Am Ophthalmol Soc, V89, P623
   Mittal VA, 2011, PSYCHIAT RES, V189, P158, DOI 10.1016/j.psychres.2011.06.006
   NORNES HO, 1979, DEV NEUROSCI-BASEL, V2, P101, DOI 10.1159/000112444
   OLSSON I, 1988, ARCH NEUROL-CHICAGO, V45, P666, DOI 10.1001/archneur.1988.00520300086024
   OLSZEWSKI J, 1954, CYTOARCHITECTURE HUM, P80
   ORAHILLY R, 1963, J EMBRYOL EXP MORPH, V11, P741
   ORNITZ EM, 1983, INT J NEUROSCI, V19, P85, DOI 10.3109/00207458309148648
   Pierce E T, 1973, Prog Brain Res, V40, P53
   PIVEN J, 1992, BIOL PSYCHIAT, V31, P491, DOI 10.1016/0006-3223(92)90260-7
   PIVEN J, 1994, NEUROBIOLOGY AUTISM, P18
   PIVEN J, 1992, AM J PSYCHIAT, V14, P734
   RAMSAY J, 1980, BRIT J OPHTHALMOL, V64, P518, DOI 10.1136/bjo.64.7.518
   REISS AL, 1986, SCHIZOPHRENIA BULL, V12, P724, DOI 10.1093/schbul/12.4.724
   RITVO ER, 1986, AM J PSYCHIAT, V143, P862
   Rodier P.M., 1986, HDB BEHAV TERATOLOGY, P185, DOI [10.1007/978-1-4613-2189-7_9., DOI 10.1007/978-1-4613-2189-7_9]
   ROSENHALL U, 1988, J LARYNGOL OTOL, V102, P435, DOI 10.1017/S0022215100105286
   SCHARRE JE, 1992, OPTOMETRY VISION SCI, V69, P433, DOI 10.1097/00006324-199206000-00004
   SCHUMACHER HJ, 1972, TERATOLOGY, V5, P233, DOI 10.1002/tera.1420050213
   SOHAL GS, 1977, EXP NEUROL, V56, P227, DOI 10.1016/0014-4886(77)90152-2
   STREETER GL, 1948, CONTRIB EMBRYOL CARN, V30, P213
   STROMLAND K, 1994, DEV MED CHILD NEUROL, V36, P351
   TURNER S, 1990, TERATOLOGY, V41, P421, DOI 10.1002/tera.1420410408
   VORHEES CV, 1987, PSYCHOPHARMACOLOGY, V92, P173
   WALKER HA, 1977, J AUTISM CHILD SCHIZ, V7, P165, DOI 10.1007/BF01537727
   WILLIAMS RS, 1980, ARCH NEUROL-CHICAGO, V37, P749, DOI 10.1001/archneur.1980.00500610029003
   WILSON J G, 1959, J Chronic Dis, V10, P111, DOI 10.1016/0021-9681(59)90026-8
NR 83
TC 465
Z9 476
U1 0
U2 39
PU WILEY
PI HOBOKEN
PA 111 RIVER ST, HOBOKEN 07030-5774, NJ USA
SN 0021-9967
EI 1096-9861
J9 J COMP NEUROL
JI J. Comp. Neurol.
PD JUN 24
PY 1996
VL 370
IS 2
BP 247
EP 261
DI 10.1002/(SICI)1096-9861(19960624)370:2<247::AID-CNE8>3.0.CO;2-2
PG 15
WC Neurosciences; Zoology
WE Science Citation Index Expanded (SCI-EXPANDED); Social Science Citation Index (SSCI)
SC Neurosciences & Neurology; Zoology
GA UT943
UT WOS:A1996UT94300008
PM 8808733
DA 2023-08-27
ER

PT J
AU Takasuna, H
   Goto, T
   Kakizawa, Y
   Miyahara, T
   Koyama, J
   Tanaka, Y
   Kawai, T
   Hongo, K
AF Takasuna, H.
   Goto, T.
   Kakizawa, Y.
   Miyahara, T.
   Koyama, J.
   Tanaka, Y.
   Kawai, T.
   Hongo, K.
TI Use of a micromanipulator system (NeuRobot) in endoscopic neurosurgery
SO JOURNAL OF CLINICAL NEUROSCIENCE
LA English
DT Article
DE Brain surgery; Intraventricular lesion; Micromanipulator;
   Neuroendoscope; Robotics surgery
ID 3RD VENTRICULOSTOMY; TUMORS; HYDROCEPHALUS; MANAGEMENT
AB NeuRobot, a micromanipulator system with a rigid neuroendoscope and three micromanipulators, was developed for less invasive and telecontrolled neurosurgery. This system can be used to perform sophisticated surgical procedures through a small, 10-mm-diameter, window. The present study was performed to evaluate the feasibility of using NeuRobot in neuroendoscopy. Four different intraventricular neurosurgical procedures were simulated in three fixed cadaver heads using NeuRobot: (1) fenestration of the floor of the third ventricle; (2) fenestration of the septum pellucidum; (3) biopsy of the thalamus; and (4) biopsy of the choroid plexus of the lateral ventricle. Each procedure required less than 2 min, and all procedures were performed accurately. After these surgical simulations, a third ventriculostomy was carried out safely and adequately in a patient with obstructive hydrocephalus due to a midbrain venous angioma. Our results confirmed that NeuRobot is applicable to lesions in which conventional endoscopic neurosurgery is indicated. Furthermore, NeuRobot can perform more complex surgical procedures than a conventional neuroendoscope because of its maneuverability and stability. NeuRobot will become a useful neurosurgical tool for dealing with lesions that are difficult to treat by conventional neuroendoscopic surgery. (C) 2012 Elsevier Ltd. All rights reserved.
C1 [Takasuna, H.; Goto, T.; Kakizawa, Y.; Miyahara, T.; Koyama, J.; Tanaka, Y.; Hongo, K.] Shinshu Univ, Sch Med, Dept Neurosurg, Matsumoto, Nagano 3908621, Japan.
   [Kawai, T.] Hitachi Ltd, Mech Engn Res Lab, Tsuchiura, Ibaraki 300, Japan.
C3 Shinshu University; Hitachi Limited
RP Hongo, K (通讯作者)，Shinshu Univ, Sch Med, Dept Neurosurg, 3-1-1 Asahi, Matsumoto, Nagano 3908621, Japan.
EM
RI Goto, Tetsuya/HGC-7552-2022; kakizawa, yukinari/AAI-4434-2020; de Lima,
   Juliana/R-7501-2018
OI kakizawa, yukinari/0000-0002-0404-0355; 
FU Grants-in-Aid for Scientific Research [23240078] Funding Source: KAKEN
CR Ferrer E, 1997, ACTA NEUROCHIR, V139, P12, DOI 10.1007/BF01850862
   Goto T, 2003, J NEUROSURG, V98, P131, DOI 10.3171/jns.2003.98.1.0131
   Goto T, 2003, J NEUROSURG, V99, P1082, DOI 10.3171/jns.2003.99.6.1082
   Hongo K, 2002, NEUROSURGERY, V51, P985, DOI 10.1227/01.NEU.0000029082.48894.18
   Hopf NJ, 1999, NEUROSURGERY, V44, P795, DOI 10.1097/00006123-199904000-00062
   Hor F, 1992, Acta Neurochir Suppl (Wien), V54, P77
   Kan K, 2001, INT CONGR SER, V1230, P144
   Kawai T, 2001, ADV ROBOTICS, V15, P301, DOI 10.1163/156855301300235814
   Koyama J, 2002, NEUROL RES, V24, P373, DOI 10.1179/016164102101200168
   LEWIS AI, 1995, J NEUROSURG, V82, P780, DOI 10.3171/jns.1995.82.5.0780
   Oka K, 1999, J NEUROSURG, V91, P964, DOI 10.3171/jns.1999.91.6.0964
   Schroeder HWS, 2002, J NEUROSURG, V96, P1032, DOI 10.3171/jns.2002.96.6.1032
   Veto F, 1997, NEUROSURGERY, V40, P871, DOI 10.1097/00006123-199704000-00048
NR 13
TC 17
Z9 17
U1 0
U2 7
PU ELSEVIER SCI LTD
PI OXFORD
PA THE BOULEVARD, LANGFORD LANE, KIDLINGTON, OXFORD OX5 1GB, OXON, ENGLAND
SN 0967-5868
J9 J CLIN NEUROSCI
JI J. Clin. Neurosci.
PD NOV
PY 2012
VL 19
IS 11
BP 1553
EP 1557
DI 10.1016/j.jocn.2012.01.033
PG 5
WC Clinical Neurology; Neurosciences
WE Science Citation Index Expanded (SCI-EXPANDED)
SC Neurosciences & Neurology
GA 025HV
UT WOS:000310180200020
PM 22995760
OA Green Submitted
DA 2023-08-27
ER

PT J
AU Borges, A
AF Borges, A
TI Trigeminal neuralgia and facial nerve paralysis
SO EUROPEAN RADIOLOGY
LA English
DT Article
DE trigeminal neuralgia; facial nerve palsy; cranial neuropathy; CT; MRI
ID NEUROVASCULAR COMPRESSION; DIFFERENTIAL-DIAGNOSIS; CRANIAL NERVES;
   PALSY; ANATOMY; MRI; ANGIOGRAPHY; PATHOLOGY; CT
AB The trigeminal nerve is the largest of the cranial nerves. It provides sensory input from the face and motor innervation to the muscles of mastication. The facial nerve is the cranial nerve with the longest extra-cranial course, and its main functions include motor innervation to the muscles of facial expression, sensory control of lacrimation and salivation, control of the stapedial reflex and to carry taste sensation from the anterior two-thirds of the tongue. In order to be able adequately to image and follow the course of these cranial nerves and their main branches, a detailed knowledge of neuroanatomy is required. As we are dealing with very small anatomic structures, high resolution dedicated imaging studies are required to pick up normal and pathologic nerves. Whereas CT is best suited to demonstrate bony neurovascular foramina and canals, MRI is preferred to directly visualize the nerve. It is also the single technique able to detect pathologic processes afflicting the nerve without causing considerable expansion such as is usually the case in certain inflammatory/infectious conditions, perineural spread of malignancies and in very small intrinsic tumours. Because a long course from the brainstem nuclei to the peripheral branches is seen, it is useful to subdivide the nerve in several segments and then tailor the imaging modality and the imaging study to that specific segment. This is particularly true in cases where topographic diagnosis can be used to locate a lesion in the course of these nerves.
C1 IPOFG, Dept Radiol, P-1093 Lisbon, Portugal.
RP Borges, A (通讯作者)，IPOFG, Dept Radiol, R Prof Lima Basto, P-1093 Lisbon, Portugal.
EM
CR Benecke JE, 2002, OTOLARYNG CLIN N AM, V35, P357, DOI 10.1016/S0030-6665(02)00003-8
   Brisman R, 2004, J NEUROSURG, V100, P848, DOI 10.3171/jns.2004.100.5.0848
   Brisman R, 2002, NEUROSURGERY, V50, P1261, DOI 10.1097/00006123-200206000-00015
   CASSELMAN JW, 2004, UPPER LOWER CRANIAL
   Chavin JM, 2003, OTOLARYNG CLIN N AM, V36, P1079, DOI 10.1016/S0030-6665(03)00124-5
   Dandy W., 1934, AM J SURG, V24, P477
   DEMARCO JK, 1993, NEUROIMAG CLIN N AM, V3, P117
   Fukuda H, 2003, SURG NEUROL, V59, P93, DOI 10.1016/S0090-3019(02)00993-X
   Go JL, 2001, SEMIN ULTRASOUND CT, V22, P502, DOI 10.1016/S0887-2171(01)90004-6
   Grandis JR, 2004, LANCET INFECT DIS, V4, P34, DOI 10.1016/S1473-3099(03)00858-2
   Held P, 1997, BRIT J RADIOL, V70, P558, DOI 10.1259/bjr.70.834.9227246
   Jackson CG, 1999, MED CLIN N AM, V83, P179, DOI 10.1016/S0025-7125(05)70096-1
   Jager L, 2001, EUR J RADIOL, V40, P133, DOI 10.1016/S0720-048X(01)00381-3
   JAGER L, 2003, MULTISLICE CT
   Jannetta PJ, 2003, NEUROSURGERY, V52, P1511, DOI 10.1227/01.NEU.0000068352.22859.82
   JONSSON L, 1995, ACTA OTO-LARYNGOL, V115, P577, DOI 10.3109/00016489509139371
   Kaylie DM, 2003, AM J NEURORADIOL, V24, P326
   Kress B, 2004, RADIOLOGY, V230, P504, DOI 10.1148/radiol.2302021353
   KUO MJ, 1995, J LARYNGOL OTOL, V109, P777, DOI 10.1017/S0022215100131305
   Kuroiwa T, 1996, Radiat Med, V14, P235
   Laine FJ, 1998, NEUROIMAG CLIN N AM, V8, P69
   Larrier D, 2003, OTOLARYNG CLIN N AM, V36, P1041, DOI 10.1016/S0030-6665(03)00127-0
   LEBLANC A, 2001, TRIGEMINAL NERVE, P79
   LOURYAN S, 2004, ANATOMY CRANIAL NERV
   Lufkin RB, 2001, MRI HEAD NECK, V2nd
   Lufkin RB, 2000, TEACHING ATLAS HEAD
   Majoie CBLM, 1997, RADIOLOGY, V204, P455, DOI 10.1148/radiology.204.2.9240535
   MASUR H, 1995, J NEUROL, V242, P93, DOI 10.1007/BF00887823
   MAY M, 1991, OTOLARYNG CLIN N AM, V24, P613
   McLaughlin MR, 1999, J NEUROSURG, V90, P1, DOI 10.3171/jns.1999.90.1.0001
   Patel NK, 2003, BRIT J NEUROSURG, V17, P60, DOI 10.1080/0268869031000093735
   Phillips CD, 2002, SEMIN ULTRASOUND CT, V23, P202, DOI 10.1016/S0887-2171(02)90047-8
   Roob G, 1999, EUR NEUROL, V41, P3, DOI 10.1159/000007990
   Salib RJ, 2001, J LARYNGOL OTOL, V115, P510, DOI 10.1258/0022215011908090
   Stone J A, 1998, Magn Reson Imaging Clin N Am, V6, P195
   Suzuki F, 2001, OTOL NEUROTOL, V22, P549, DOI 10.1097/00129492-200107000-00023
   SWARTZ JD, 1998, IMAGING TEMPORAL BON, P383
   WILSONPAWELS L, 1998, CRANIAL NERVES ANATO
   YAMAGIDA M, 1991, FACIAL NERVE RES JPN, V11, P221
   Yamamoto M, 2004, AM J ROENTGENOL, V182, P385, DOI 10.2214/ajr.182.2.1820385
   YANAGIDA M, 1993, ACTA OTO-LARYNGOL, P58
   Yoshino N, 2003, RADIOLOGY, V228, P539, DOI 10.1148/radiol.2282020439
   Zakrzewska JM, 2002, CLIN J PAIN, V18, P14, DOI 10.1097/00002508-200201000-00003
NR 43
TC 21
Z9 23
U1 0
U2 6
PU SPRINGER
PI NEW YORK
PA 233 SPRING ST, NEW YORK, NY 10013 USA
SN 0938-7994
EI 1432-1084
J9 EUR RADIOL
JI Eur. Radiol.
PD MAR
PY 2005
VL 15
IS 3
BP 511
EP 533
DI 10.1007/s00330-004-2613-9
PG 23
WC Radiology, Nuclear Medicine & Medical Imaging
WE Science Citation Index Expanded (SCI-EXPANDED)
SC Radiology, Nuclear Medicine & Medical Imaging
GA 912OT
UT WOS:000228089500013
PM 15690205
DA 2023-08-27
ER

PT J
AU Pal, SK
   Choueiri, TK
   Wang, K
   Khaira, D
   Karam, JA
   Van Allen, E
   Palma, NA
   Stein, MN
   Johnson, A
   Squillace, R
   Elvin, JA
   Chmielecki, J
   Yelensky, R
   Yakirevich, E
   Lipson, D
   Lin, DI
   Miller, VA
   Stephens, PJ
   Ali, SM
   Ross, JS
AF Pal, Sumanta K.
   Choueiri, Toni K.
   Wang, Kai
   Khaira, Depinder
   Karam, Jose A.
   Van Allen, Eliezer
   Palma, Norma A.
   Stein, Mark N.
   Johnson, Adrienne
   Squillace, Rachel
   Elvin, Julia A.
   Chmielecki, Juliann
   Yelensky, Roman
   Yakirevich, Evgeny
   Lipson, Doron
   Lin, Douglas I.
   Miller, Vincent A.
   Stephens, Philip J.
   Ali, Siraj M.
   Ross, Jeffrey S.
TI Characterization of Clinical Cases of Collecting Duct Carcinoma of the
   Kidney Assessed by Comprehensive Genomic Profiling
SO EUROPEAN UROLOGY
LA English
DT Article
DE Collecting duct carcinoma; Genomic profiling; Medullary carcinoma; NF2
ID RENAL-CELL CARCINOMA; HIGH-FREQUENCY; CANCER; SURVIVAL; NF2/MERLIN;
   EVEROLIMUS; MUTATIONS; MTORC1; DOMAIN; MERLIN
AB Background: Collecting duct carcinoma (CDC) is a rare type of renal cell carcinoma (RCC) originating from the renal medulla. Clinical outcomes are poor, and there are no consensus guidelines to guide therapy.
   Objective: To determine genomic alterations (GAs) in a series of patients with locally advanced or metastatic CDC for whom genomic profiling was performed during the course of clinical care.
   Design, setting, and participants: Formalin-fixed, paraffin-embedded blocks or slides were obtained for 17 patients with CDC. DNA was extracted and comprehensive genomic profiling was performed in a laboratory certified under the Clinical Laboratory Improvement Amendments.
   Outcome measurements and statistical analysis: Bayesian algorithms and local alignment algorithms were used to detect substitutions and insertions/deletions, respectively. A comparison to normal control samples was used to detect copy number alterations. Clinically relevant GAs (CRGAs) were defined as those linked to approved or investigational targeted therapies.
   Results and limitations: The median age in the cohort was 53 yr (range 26-73), and 14 primary tumors and three metastatic sites assessed. A total of 36 GAs were detected in this series of patients, with an average of 2.1 GAs per case. The most common GAs were in NF2 (5/17, 29%), SETD2 (4/17, 24%), SMARCB1 (3/17, 18%), and CDKN2A (2/17, 12%). Of nine cases assessed for FH GAs, two patients had FH homozygous loss. A limitation is that targeted interrogation of genes known to be implicated in other cancers was performed, so mutations outside of these cannot be excluded.
   Conclusions: Recurrent CRGAs were detected in this series of CDC cases and suggest a possible benefit from targeted therapy. In particular, mTOR inhibitors may be of interest in patients with NF2 alterations. Alterations in FH and SMARCB1 also occurred in a mutually exclusive manner to NF2 alterations.
   Patient summary: This report provides important genomic insights into collecting duct carcinoma, a rare type of renal cell carcinoma with a very aggressive course. These insights could further rationalize the use of targeted therapies for rare tumors according to the individual genomic alterations harbored. (C) 2015 European Association of Urology. Published by Elsevier B.V. All rights reserved.
C1 [Pal, Sumanta K.] City Hope Comprehens Canc Ctr, Dept Med Oncol & Expt Therapeut, Duarte, CA USA.
   [Choueiri, Toni K.; Van Allen, Eliezer] Dana Farber Canc Inst, Div Med Oncol, Boston, MA 02115 USA.
   [Wang, Kai; Khaira, Depinder; Palma, Norma A.; Johnson, Adrienne; Squillace, Rachel; Elvin, Julia A.; Chmielecki, Juliann; Yelensky, Roman; Lipson, Doron; Miller, Vincent A.; Stephens, Philip J.; Ali, Siraj M.; Ross, Jeffrey S.] Fdn Med Inc, 150 Second St, Cambridge, MA 02141 USA.
   [Karam, Jose A.] Univ Texas MD Anderson Canc Ctr, Dept Urol, Houston, TX 77030 USA.
   [Stein, Mark N.] Rutgers Canc Inst New Jersey, Dept Med Oncol, New Brunswick, NJ USA.
   [Yakirevich, Evgeny] Brown Univ, Warren Alpert Sch Med, Dept Pathol & Lab Med, Providence, RI 02912 USA.
   [Lin, Douglas I.] Beth Israel Deaconess Med Ctr, Dept Pathol, 330 Brookline Ave, Boston, MA 02215 USA.
   [Ross, Jeffrey S.] Albany Med Coll, Dept Pathol & Lab Med, Albany, NY 12208 USA.
C3 City of Hope; Harvard University; Dana-Farber Cancer Institute;
   Foundation Medicine; University of Texas System; UTMD Anderson Cancer
   Center; Rutgers State University New Brunswick; Rutgers State University
   Medical Center; Rutgers Cancer Institute of New Jersey; Brown
   University; Harvard University; Beth Israel Deaconess Medical Center;
   Albany Medical College
RP Ali, SM (通讯作者)，Fdn Med Inc, 150 Second St, Cambridge, MA 02141 USA.
EM
RI Ali, Siraj/AAE-2958-2019; Choueiri, Toni/K-5238-2019; Wang,
   Kai/IWD-8760-2023
OI Choueiri, Toni/0000-0002-9201-3217; Wang, Kai/0000-0002-6170-4744; Lin,
   Douglas/0000-0003-0105-9760
CR Ali SM, 2015, EUR UROL, V67, P1195, DOI 10.1016/j.eururo.2015.01.015
   Amin MB, 2014, AM J SURG PATHOL, V38, P871, DOI 10.1097/PAS.0000000000000222
   [Anonymous], J CLIN ONCOL S3
   [Anonymous], MOD PATHOL
   Calderaro J, 2012, HISTOPATHOLOGY, V61, P428, DOI 10.1111/j.1365-2559.2012.04228.x
   Cheung LWT, 2011, CANCER DISCOV, V1, P170, DOI 10.1158/2159-8290.CD-11-0039
   Creighton CJ, 2013, NATURE, V499, P43, DOI 10.1038/nature12222
   Curto M, 2008, BRIT J CANCER, V98, P256, DOI 10.1038/sj.bjc.6604002
   Dason S, 2013, CURR ONCOL, V20, pE223, DOI 10.3747/co.20.1230
   Elkabets M, 2013, SCI TRANSL MED, V5, DOI 10.1126/scitranslmed.3005747
   Elwood H, 2011, UROLOGY, V78, DOI 10.1016/j.urology.2011.04.043
   Foundation Medicine, FOUNDATIONONE TECHN
   Frampton GM, 2013, NAT BIOTECHNOL, V31, P1023, DOI 10.1038/nbt.2696
   Gupta R, 2012, AM J SURG PATHOL, V36, P1265, DOI 10.1097/PAS.0b013e3182635954
   Hah JH, 2014, HEAD NECK-J SCI SPEC, V36, P1547, DOI 10.1002/hed.23499
   He L, 2013, CANCER DISCOV, V3, P324, DOI 10.1158/2159-8290.CD-12-0417
   Im AP, 2014, LEUKEMIA, V28, P1774, DOI 10.1038/leu.2014.124
   James MF, 2009, MOL CELL BIOL, V29, P4250, DOI 10.1128/MCB.01581-08
   Knutson SK, 2013, P NATL ACAD SCI USA, V110, P7922, DOI 10.1073/pnas.1303800110
   Linehan WM, 2013, CLIN CANCER RES, V19, P3345, DOI 10.1158/1078-0432.CCR-13-0304
   Lopez-Lago MA, 2009, MOL CELL BIOL, V29, P4235, DOI 10.1128/MCB.01578-08
   Motzer RJ, 2004, J CLIN ONCOL, V22, P454, DOI 10.1200/JCO.2004.06.132
   Motzer RJ, 2014, NEW ENGL J MED, V370, P1769, DOI 10.1056/NEJMc1400731
   Oudard S, 2007, J UROLOGY, V177, P1698, DOI 10.1016/j.juro.2007.01.063
   Pal SK, 2010, MOL CANCER THER, V9, P3115, DOI 10.1158/1535-7163.MCT-10-0873
   Park JH, 2015, BMC CANCER, V15, DOI 10.1186/s12885-015-1139-7
   Pepek JM, 2009, CLIN GENITOURIN CANC, V7, pE24, DOI 10.3816/CGC.2009.n.017
   Rao Q, HISTOPATHOL IN PRESS
   Ross JS, 2014, CLIN CANCER RES, V20, P68, DOI 10.1158/1078-0432.CCR-13-1992
   Srigley JR, 2013, AM J SURG PATHOL, V37, P1469, DOI 10.1097/PAS.0b013e318299f2d1
   Wetmore C, NEURO OONCO IN PRESS
   Wright JL, 2009, J UROLOGY, V182, P2595, DOI 10.1016/j.juro.2009.08.049
   Yogesha SD, 2011, PROTEIN SCI, V20, P2113, DOI 10.1002/pro.751
   Young RJ, 2014, PIGM CELL MELANOMA R, V27, DOI 10.1111/pcmr.12228
NR 34
TC 76
Z9 78
U1 1
U2 4
PU ELSEVIER
PI AMSTERDAM
PA RADARWEG 29, 1043 NX AMSTERDAM, NETHERLANDS
SN 0302-2838
EI 1873-7560
J9 EUR UROL
JI Eur. Urol.
PD SEP
PY 2016
VL 70
IS 3
BP 516
EP 521
DI 10.1016/j.eururo.2015.06.019
PG 6
WC Urology & Nephrology
WE Science Citation Index Expanded (SCI-EXPANDED)
SC Urology & Nephrology
GA DS4MA
UT WOS:000380754000030
PM 26149668
DA 2023-08-27
ER

PT J
AU FRANQUEMONT, DW
   MILLS, SE
   LACK, EE
AF FRANQUEMONT, DW
   MILLS, SE
   LACK, EE
TI IMMUNOHISTOCHEMICAL DETECTION OF NEUROBLASTOMATOUS FOCI IN COMPOSITE
   ADRENAL PHEOCHROMOCYTOMA-NEUROBLASTOMA
SO AMERICAN JOURNAL OF CLINICAL PATHOLOGY
LA English
DT Article
DE ADRENAL GLAND; IMMUNOHISTOCHEMISTRY; NEUROBLASTOMA; PHEOCHROMOCYTOMA
ID VASOACTIVE INTESTINAL PEPTIDE; S-100 PROTEIN; GANGLIONEUROMA;
   PARAGANGLIOMAS; CELLS; TUMORS; MEDULLA; GLAND; NEUROENDOCRINE; PROGNOSIS
AB To determine if immunohistochemistry might aid in the identification of neuroblastomatous foci in composite adrenal tumors, the authors analyzed two examples of composite adrenal pheochromocytoma-neuroblastoma, 18 pure pheochromocytomas, and six pure neuroblastomas using peanut agglutinin and a panel of antibodies directed against neuroendocrine and neural-associated antigens. Pure pheochromocytoma had the following immunopositivity: vimentin 14/18, chromogranin 18/18, synaptophysin 18/18, S100 protein 0/18 (tumor cells), neurofilament 14/18, J1 beta-tubulin (J1) 18/18, microtubule-associated protein-2 13/18, glial fibrillary acidic protein 13/18, and peanut agglutinin 17/18. Pure neuroblastoma reacted positively as follows: vimentin 0/6, chromogranin 5/6, synaptophysin 4/6, S100 protein 0/6 (tumor cells), neurofilament 5/6, J1 6/6, microtubule-associated protein-2 6/6, glial fibrillary acidic protein 1/6, and peanut agglutinin 6/6. Each component of both composite tumors reacted similarly to the pure neoplasms. Although the frequency of positive staining was similar for pheochromocytoma and neuroblastoma, the intensity and pattern differed for several antigens. Pheochromocytoma was diffusely positive for synaptophysin and chromogranin, whereas staining was focal and punctate in neuroblastoma. Microtubule-associated protein-2, J1, and neurofilament antibodies highlighted the fibrillar background of neuroblastoma, which pheochromocytoma lacked. Pheochromocytoma contained focal, ball-like immunoreactivity for glial fibrillary acidic protein and vimentin, which was absent in neuroblastoma. These immunohistochemical distinctions can assist the clinically important recognition of neuroblastomatous foci in composite adrenal pheochromocytoma-neuroblastoma.
C1 UNIV VIRGINIA,HLTH SCI CTR,DEPT PATHOL,DIV SURG PATHOL,CHARLOTTESVILLE,VA.
   GEORGETOWN UNIV,DEPT PATHOL,WASHINGTON,DC.
C3 University of Virginia; Georgetown University
RP FRANQUEMONT, DW (通讯作者)，WASHINGTON UNIV,SCH MED,DEPT PATHOL,DIV ANAT PATHOL,LAUREN V ACKERMAN LAB SURG PATHOL,BOX 8118,ST LOUIS,MO 63110, USA.
CR ACHILLES E, 1991, HISTOPATHOLOGY, V18, P453, DOI 10.1111/j.1365-2559.1991.tb00877.x
   AIBA M, 1988, AM J SURG PATHOL, V12, P559, DOI 10.1097/00000478-198807000-00008
   BALAZS M, 1988, HUM PATHOL, V19, P1352, DOI 10.1016/S0046-8177(88)80292-2
   BINDER LI, 1984, P NATL ACAD SCI-BIOL, V81, P5613, DOI 10.1073/pnas.81.17.5613
   CHOUTET P, 1981, SEM HOP PARIS, V57, P590
   DAWSON DW, 1969, J PATHOL, V97, P231, DOI 10.1002/path.1710970207
   DELELLIS RA, 1984, J HISTOCHEM CYTOCHEM, V32, P899, DOI 10.1177/32.8.6146648
   FERNANDO PB, 1951, AMA ARCH PATHOL, V52, P182
   FRANKFURTER A, 1986, J CELL BIOL, V103, pA273
   FURIHATA M, 1991, VIRCHOWS ARCH A, V418, P33, DOI 10.1007/BF01600242
   HACKER GW, 1988, VIRCHOWS ARCH A, V412, P399, DOI 10.1007/BF00750574
   HOEFLER H, 1986, VIRCHOWS ARCH A, V409, P609, DOI 10.1007/BF00713428
   KAHN H J, 1988, Pediatric Pathology, V8, P83
   Kimura Noriko, 1991, Endocr Pathol, V2, P139, DOI 10.1007/BF02915454
   KLIEWER KE, 1989, HUM PATHOL, V20, P29, DOI 10.1016/0046-8177(89)90199-8
   KRAGEL PJ, 1985, ARCH PATHOL LAB MED, V109, P470
   KUMURA N, 1991, ARCH PATHOL LAB MED, V114, P506
   LAURIOLA L, 1985, VIRCHOWS ARCH B, V49, P13, DOI 10.1007/BF02912081
   LAYFIELD LJ, 1987, ACTA CYTOL, V31, P33
   Lewis D, 1934, ARCH SURG-CHICAGO, V28, P16, DOI 10.1001/archsurg.1934.01170130019003
   LINNOILA RI, 1990, HUM PATHOL, V21, P1168
   MIETTINEN M, 1988, ULTRASTRUCT PATHOL, V12, P513, DOI 10.3109/01913128809032236
   MIN KW, 1988, ARCH PATHOL LAB MED, V112, P266
   MOLENAAR WM, 1990, AM J PATHOL, V136, P375
   MOORGHEN M, 1991, VIRCHOWS ARCH A, V419, P203, DOI 10.1007/BF01626349
   MURPHY GP, 1987, ONCOLOGY, V44, P383, DOI 10.1159/000226515
   NAKAGAWARA A, 1985, CANCER, V55, P2794, DOI 10.1002/1097-0142(19850615)55:12<2794::AID-CNCR2820551213>3.0.CO;2-L
   OSBORN M, 1982, VIRCHOWS ARCH B, V40, P141, DOI 10.1007/BF02932859
   SCHRODER HD, 1986, HISTOPATHOLOGY, V10, P1023, DOI 10.1111/j.1365-2559.1986.tb02539.x
   TISCHLER AS, 1976, SCIENCE, V192, P902, DOI 10.1126/science.179139
   TISCHLER AS, 1987, HUM PATHOL, V18, P909, DOI 10.1016/S0046-8177(87)80268-X
   TISCHLER AS, 1978, LAB INVEST, V39, P77
   TRIMMER PA, 1986, J CELL BIOL, V103, pA273
   TRUMP DL, 1977, CANCER, V40, P1526, DOI 10.1002/1097-0142(197710)40:4<1526::AID-CNCR2820400425>3.0.CO;2-7
   WIRNSBERGER GH, 1992, AM J SURG PATHOL, V16, P49
NR 35
TC 37
Z9 37
U1 0
U2 0
PU LIPPINCOTT-RAVEN PUBL
PI PHILADELPHIA
PA 227 EAST WASHINGTON SQ, PHILADELPHIA, PA 19106
SN 0002-9173
J9 AM J CLIN PATHOL
JI Am. J. Clin. Pathol.
PD AUG
PY 1994
VL 102
IS 2
BP 163
EP 170
DI 10.1093/ajcp/102.2.163
PG 8
WC Pathology
WE Science Citation Index Expanded (SCI-EXPANDED)
SC Pathology
GA PA500
UT WOS:A1994PA50000006
PM 8042583
DA 2023-08-27
ER

PT J
AU Sawin, PD
   Hitchon, PW
   Follett, KA
   Torner, JC
AF Sawin, PD
   Hitchon, PW
   Follett, KA
   Torner, JC
TI Computed imaging-assisted stereotactic brain biopsy - A risk analysis of
   225 consecutive cases
SO SURGICAL NEUROLOGY
LA English
DT Article
ID MASS LESIONS; EXPERIENCE; TOMOGRAPHY; TUMORS; MANAGEMENT; SERIES
AB BACKGROUND Treatment strategies for intracranial mass lesions are most effective when based upon histopathological diagnoses. Image-guided stereotaxy has provided the means to sample tissue from small or deeply seated intraparenchymal lesions with a relatively high degree of safety and accuracy. Although procedural complications are infrequent, devastating neurological sequelae may result from hemorrhage or direct trauma. This study was undertaken to identify factors that may confer an increased risk of morbidity from stereotactic brain biopsy.
   METHODS Two hundred twenty-five consecutive computer-assisted stereotactic brain biopsy procedures were reviewed. Patient age averaged 47.4 years (range, 3-84 years); gender ratio was approximately 2:1 (male:female). Pre-existing medical conditions were identified in nearly half of the cohort. 61.3% of biopsied lesions were lobar; the remainder (38.7%) were "deep-seated" (thalamus, basal ganglia, pineal, hypothalamus, cerebellum, brainstem). Glial tumors accounted for the majority (44.4%) of biopsied lesions; metastases (12.9%) and lymphoma (11.6%) were also relatively common. Demographical, anatomical, surgical, and histological data were compiled and putative risk factors for morbidity identified. These variables were then subjected to univariate and logistic regression analyses to determine their significance as independent predictors of operative risk.
   RESULTS Twelve patients suffered complications as a consequence of the biopsy procedure (eight from hemorrhage, four from direct trauma). Major morbidity (hemiparesis, aphasia, obtundation) occurred in eight patients (3.6%). Three patients (1.3%) suffered minor morbidity (transient, mild neurological deficits). One operative fatality occurred (0.4%). An increased risk of morbidity was associated with the preoperative use of antiplatelet agents, chronic corticosteroids, deep-seated lesions, malignant gliomas, and a greater number of biopsy attempts (p < 0.05). Factors not conferring increased morbidity included gender, age, pre-existing illness, extracranial malignancy, cardiac disease, hypertension, diabetes, HIV status, and instrument used to procure the specimen.
   CONCLUSIONS Complications arising from stereotactic brain biopsy are infrequent but can be disastrous. Operative risk is a function of several independent variables, including lesion properties (location, histology), preoperative pharmacological therapy (corticosteroids, antiplatelet agents), and operative technique. This analysis suggests that the morbidity of stereotactic brain biopsy may be minimized by risk factor modification. (C) 1998 by Elsevier Science Inc.
C1 Univ Iowa Hosp & Clin, Div Neurosurg, Iowa City, IA 52242 USA.
   Univ Iowa Hosp & Clin, Dept Biostat & Epidemiol, Iowa City, IA 52242 USA.
C3 University of Iowa; University of Iowa
RP Hitchon, PW (通讯作者)，Univ Iowa Hosp & Clin, Div Neurosurg, 200 Hawkins Dr, Iowa City, IA 52242 USA.
OI Hitchon, Patrick/0000-0002-8175-9136
CR ABERNATHEY CD, 1989, J NEUROSURG, V70, P195, DOI 10.3171/jns.1989.70.2.0195
   APUZZO MLJ, 1987, NEUROSURGERY, V20, P930, DOI 10.1227/00006123-198706000-00019
   BAXTER JD, 1992, HOSP PRACT, V27, P111
   BAXTER JD, 1987, ENDOCRINOL METAB, P511
   BERNSTEIN M, 1994, J NEUROSURG, V81, P165, DOI 10.3171/jns.1994.81.2.0165
   BOUVIER G, 1983, APPL NEUROPHYSIOL, V46, P227
   BURNSTEIN KL, 1992, MOL CELL ENDOCRINOL, V83, pC1, DOI 10.1016/0303-7207(92)90187-B
   Carpenter MB, 1991, CORE TEXT NEUROANATO, V4th, P250
   CHAPPELL ET, 1992, NEUROSURGERY, V30, P825, DOI 10.1227/00006123-199206000-00001
   COFFEY RJ, 1985, NEUROSURGERY, V17, P12, DOI 10.1227/00006123-198507000-00003
   COLBASSANI HJ, 1988, J NEUROL NEUROSUR PS, V51, P332, DOI 10.1136/jnnp.51.3.332
   Frank F, 1988, Acta Neurochir Suppl (Wien), V42, P177
   FRIEDMAN WA, 1989, NEUROSURGERY, V25, P180, DOI 10.1227/00006123-198908000-00005
   GREENE GM, 1989, J NEUROSURG, V71, P494, DOI 10.3171/jns.1989.71.4.0494
   HEILBRUN MP, 1983, J NEUROSURG, V59, P217, DOI 10.3171/jns.1983.59.2.0217
   HOOD TW, 1986, J NEUROSURG, V65, P172, DOI 10.3171/jns.1986.65.2.0172
   KELLY PJ, 1986, NEUROLOGY, V36, P535, DOI 10.1212/WNL.36.4.535
   KELLY PJ, 1985, MAYO CLIN PROC, V60, P223, DOI 10.1016/S0025-6196(12)60314-0
   LEVY RM, 1992, NEUROSURGERY, V30, P186, DOI 10.1227/00006123-199202000-00006
   LOBATO RD, 1982, APPL NEUROPHYSIOL, V45, P426
   LUNSFORD LD, 1984, SURG NEUROL, V22, P222
   Mundinger F, 1985, Acta Neurochir Suppl (Wien), V35, P70
   ONEILL KS, 1992, BRIT J NEUROSURG, V6, P421, DOI 10.3109/02688699208995030
   OSTERTAG CB, 1980, SURG NEUROL, V14, P275
   RANJAN A, 1993, J NEUROSURG, V79, P839, DOI 10.3171/jns.1993.79.6.0839
   REYMOND MA, 1992, NEUROSURG REV, V15, P21, DOI 10.1007/BF02352062
   Sedan R, 1984, ACTA NEUROCHIR S, V33, P207
   Thomas D G, 1989, Br J Neurosurg, V3, P321, DOI 10.3109/02688698909002810
   WEISS HJ, 1982, PLATELETS PATHOPHYSI, P45
   WILD A M, 1990, British Journal of Neurosurgery, V4, P407, DOI 10.3109/02688699008992763
   YONAS H, 1987, CEREBRAL BLOOD FLOW, P220
NR 31
TC 121
Z9 126
U1 0
U2 5
PU ELSEVIER SCIENCE INC
PI NEW YORK
PA 655 AVENUE OF THE AMERICAS, NEW YORK, NY 10010 USA
SN 0090-3019
J9 SURG NEUROL
JI Surg. Neurol.
PD JUN
PY 1998
VL 49
IS 6
BP 640
EP 649
DI 10.1016/S0090-3019(97)00435-7
PG 10
WC Clinical Neurology; Surgery
WE Science Citation Index Expanded (SCI-EXPANDED)
SC Neurosciences & Neurology; Surgery
GA ZV200
UT WOS:000074280400027
PM 9637625
DA 2023-08-27
ER

PT J
AU Serre, A
   Idri, K
   Fenoglietto, P
   Ailleres, N
   Santoro, L
   Lemanski, C
   Garrel, R
   Makeieff, M
   Allaw, A
   Dubois, JB
   Azria, D
AF Serre, Antoine
   Idri, Katia
   Fenoglietto, Pascal
   Ailleres, Norbert
   Santoro, Lore
   Lemanski, Claire
   Garrel, Renaud
   Makeieff, Marc
   Allaw, Ali
   Dubois, Jean-Bernard
   Azria, David
TI Dosimetric comparison between coplanar and non coplanar field
   radiotherapy for ethmoid sinus cancer
SO RADIATION ONCOLOGY
LA English
DT Article
ID INTENSITY-MODULATED RADIOTHERAPY; PARANASAL SINUS; RADIATION-THERAPY;
   NASAL CAVITY; MAXILLARY SINUS; CONFORMAL RADIOTHERAPY; NATURAL-HISTORY;
   CARCINOMA; IMRT; IRRADIATION
AB Background: To compare non coplanar field (NCF) with coplanar field (CF)-intensity-modulated radiotherapy (IMRT) planning for ethmoid cancer.
   Methods: Seven patients treated with NCF IMRT for ethmoid cancer were studied. A CF IMRT optimization was prepared with the same constraints as for the NCF treatment. The maximum point doses (D max) obtained for the different optic pathway structures (OPS) should differ no more than 3% from those achieved with the NCF IMRT plan. The distribution of the dose in the target volume and in the critical structures was compared between the two techniques, as well as the Conformity (CI) and the Homogeneity Indexes (HI) in the target volume.
   Results: We noted no difference between the two techniques in the OPS for the D1, D2, and D5%, in the inner ear and controlateral lens for the average Dmax, in the temporo-mandibular joints for the average mean dose, in the cord and brainstem for the average D1%. The dose-volume histograms were slightly better with the NCF treatment plan for the planning target volume (PTV) with a marginally better HI but no impact on CI. We found a great improvement in the PTV coverage with the CF treatment plan for two patients with T4 tumors.
   Conclusion: IMRT is one of the treatment options for ethmoid cancer. The PTV coverage is optimal without compromising the protection of the OPS. The impact of non coplanar versus coplanar set up is very slight.
C1 [Serre, Antoine; Lemanski, Claire; Allaw, Ali; Dubois, Jean-Bernard; Azria, David] Val Aurelle Canc Inst, Dept Radiat Oncol, Montpellier, France.
   [Idri, Katia; Fenoglietto, Pascal; Ailleres, Norbert; Santoro, Lore] Val Aurelle Canc Inst, Radiophys Unit, Montpellier, France.
   [Makeieff, Marc] Univ Hosp Gui Chauliac, Dept Head & Neck Surg, Montpellier, France.
C3 UNICANCER; Universite de Montpellier; Institut Regional du Cancer
   Montpellier / Val d'Aurelle (ICM); UNICANCER; Universite de Montpellier;
   Institut Regional du Cancer Montpellier / Val d'Aurelle (ICM);
   Universite de Montpellier; CHU de Montpellier
RP Azria, D (通讯作者)，Val Aurelle Canc Inst, Dept Radiat Oncol, Montpellier, France.
EM;
  ;
  ;
  ;
  ;
  
OI Garrel, Renaud/0000-0002-1367-363X
CR Adams EJ, 2001, INT J RADIAT ONCOL, V51, P579, DOI 10.1016/S0360-3016(01)01655-8
   AMENDOLA BE, 1981, INT J RADIAT ONCOL, V7, P743, DOI 10.1016/0360-3016(81)90467-3
   Claus F, 2001, INT J RADIAT ONCOL, V51, P318, DOI 10.1016/S0360-3016(01)01627-3
   Daly ME, 2007, INT J RADIAT ONCOL, V67, P151, DOI 10.1016/j.ijrobp.2006.07.1389
   Dulguerov P, 2001, CANCER-AM CANCER SOC, V92, P3012, DOI 10.1002/1097-0142(20011215)92:12<3012::AID-CNCR10131>3.0.CO;2-E
   Dulguerov Pavel, 2006, Curr Opin Otolaryngol Head Neck Surg, V14, P67
   Duthoy W, 2005, CANCER-AM CANCER SOC, V104, P71, DOI 10.1002/cncr.21100
   ELLINGWOOD KE, 1979, CANCER, V43, P1517, DOI 10.1002/1097-0142(197904)43:4<1517::AID-CNCR2820430445>3.0.CO;2-B
   EMAMI B, 1991, INT J RADIAT ONCOL, V21, P109, DOI 10.1016/0360-3016(91)90171-Y
   Feuvret L, 2006, INT J RADIAT ONCOL, V64, P333, DOI 10.1016/j.ijrobp.2005.09.028
   HENK JM, 1993, INT J RADIAT ONCOL, V25, P815, DOI 10.1016/0360-3016(93)90310-R
   HORDIJK GJ, 1985, CLIN OTOLARYNGOL, V10, P285, DOI 10.1111/j.1365-2273.1985.tb00256.x
   Huang D, 2003, INT J RADIAT ONCOL, V56, P158, DOI 10.1016/S0360-3016(03)00080-4
   Jansen EPM, 2000, INT J RADIAT ONCOL, V48, P27, DOI 10.1016/S0360-3016(00)00594-0
   JIANG GL, 1991, RADIOTHER ONCOL, V21, P193, DOI 10.1016/0167-8140(91)90037-H
   Jiang GL, 1998, RADIOTHER ONCOL, V49, P21, DOI 10.1016/S0167-8140(98)00075-9
   JIANG GL, 1994, RADIOTHER ONCOL, V30, P17, DOI 10.1016/0167-8140(94)90005-1
   Katz TS, 2002, HEAD NECK-J SCI SPEC, V24, P821, DOI 10.1002/hed.10143
   Mock U, 2004, INT J RADIAT ONCOL, V58, P147, DOI 10.1016/S0360-3016(03)01452-4
   NAKISSA N, 1983, CANCER, V51, P980, DOI 10.1002/1097-0142(19830315)51:6<980::AID-CNCR2820510603>3.0.CO;2-Y
   O'Daniel JC, 2004, INT J RADIAT ONCOL, V58, P876, DOI 10.1016/j.ijrobp.2003.10.031
   Pacholke HD, 2005, AM J CLIN ONCOL-CANC, V28, P474, DOI 10.1097/01.coc.0000182600.51019.de
   PARSONS JT, 1994, INT J RADIAT ONCOL, V30, P755, DOI 10.1016/0360-3016(94)90346-8
   Paulino AC, 1997, INT J RADIAT ONCOL, V39, P283, DOI 10.1016/S0360-3016(97)00293-9
   Pommier P, 2000, INT J RADIAT ONCOL, V48, P485, DOI 10.1016/S0360-3016(00)00616-7
   SHUKOVSK.LJ, 1972, RADIOLOGY, V104, P629, DOI 10.1148/104.3.629
   Takeda A, 1999, INT J RADIAT ONCOL, V44, P599, DOI 10.1016/S0360-3016(99)00057-7
   Tsien C, 2003, INT J RADIAT ONCOL, V55, P776, DOI 10.1016/S0360-3016(02)04274-8
   TSUJII H, 1986, CANCER, V57, P2261, DOI 10.1002/1097-0142(19860615)57:12<2261::AID-CNCR2820571204>3.0.CO;2-X
   Wang XC, 2005, INT J RADIAT ONCOL, V63, P594, DOI 10.1016/j.ijrobp.2005.06.006
NR 30
TC 8
Z9 8
U1 0
U2 1
PU BMC
PI LONDON
PA CAMPUS, 4 CRINAN ST, LONDON N1 9XW, ENGLAND
SN 1748-717X
J9 RADIAT ONCOL
JI Radiat. Oncol.
PD SEP 18
PY 2007
VL 2
AR 35
DI 10.1186/1748-717X-2-35
PG 9
WC Oncology; Radiology, Nuclear Medicine & Medical Imaging
WE Science Citation Index Expanded (SCI-EXPANDED)
SC Oncology; Radiology, Nuclear Medicine & Medical Imaging
GA 364OA
UT WOS:000260343800001
PM 17877793
OA gold, Green Published
DA 2023-08-27
ER

PT J
AU Chmielewska, B
   Kaminski, ML
AF Chmielewska, B
   Kaminski, ML
TI Progression of preexisting trigeminalgia to Tolose-Hunt-like syndrome.
   The importance of neuroimaging for early differential diagnosis
SO NEUROLOGICAL SCIENCES
LA English
DT Article; Proceedings Paper
CT 32nd Congress of the Italian-Neurological-Society
CY SEP   29, 2001
CL RIMINI, ITALY
SP Italian Neurol Soc
DE trigeminalgia; painful ophthalmoplegia; EPs; MR imaging
ID OPHTHALMOPLEGIC MIGRAINE; FACIAL-PAIN; NEURALGIA; HEADACHE; NERVE
AB Recurrent unbearable, paroxysmal, unilateral facial pain in the distribution of one or more branches of the trigeminal nerve often provoked by sensory stimuli is typical for idiopathic trigeminal neuralgia. The less frequent localization in the area of ophthalmic branch (5%) is particularly controversial and should be distinguished from pathological lesions in the brainstem and middle and posterior cranial fossa and from disesases of the orbit and eye. This case study presents a 79-year-old woman with typical clinical features of 1st division trigeminalgia without any neurological loss and with normal results of laryngological, ophthalmological, and stomatological examinations as well as neuroimaging CT, and MR /MRA evaluation. Only the evoked potential blink and masseter reflexes demosntrated the pathological values in the early phase of illness. After 1 year of pharmacological treatment no improvement was achieved and the pain became neuropathic and paresis of 3rd, 4th and 6th nerves developed, as observed in Tolose-Hunt syndrome. MRI of the orbit revealed a pathological mass in its apex with a connection to the superior orbital fissure. However, treatment with steroids was completely ineffective. Surgical resection of the tumor (leiomyosarcoma) only partially reversed oculomotor palsy and diminished aching. In differential diagnosis of idiopathic and symptomatic trigeminalgia, early MR and MRA imaging is the most essential and sometimes may be the best single test to evaluate lesions even in distant areas of the nervous system branches.
C1 Univ Sch Med, Dept & Clin Neurol, PL-20954 Lublin, Poland.
C3 Medical University of Lublin
RP Chmielewska, B (通讯作者)，Univ Sch Med, Dept & Clin Neurol, Jaczewski 8 Str, PL-20954 Lublin, Poland.
CR BLAKE PY, 1995, AM J NEURORADIOL, V16, P241
   de Arcaya AA, 1999, HEADACHE, V39, P321, DOI 10.1046/j.1526-4610.1999.3905321.x
   FROMM GH, 1990, NEUROLOGY, V40, P1493, DOI 10.1212/WNL.40.10.1493
   Graff-Radford SB, 2000, CURR OPIN NEUROL, V13, P291, DOI 10.1097/00019052-200006000-00010
   HAMLYN PJ, 1992, J NEUROSURG, V76, P948, DOI 10.3171/jns.1992.76.6.0948
   HANSEN SL, 1990, ACTA NEUROL SCAND, V81, P54
   HEDGES TR, 1979, HEADACHE, V19, P151, DOI 10.1111/j.1526-4610.1979.hed1903151.x
   KATUSIC S, 1990, ANN NEUROL, V27, P89, DOI 10.1002/ana.410270114
   Leandri M, 1998, NEUROLOGY, V51, P1641, DOI 10.1212/WNL.51.6.1641
   MAJOIE CB, 2000, RADIOLOGY, V209, P557
   Mark AS, 1998, AM J NEURORADIOL, V19, P1887
   SOLOMON S, 1990, NEUROL CLIN, V8, P913, DOI 10.1016/S0733-8619(18)30326-8
   SORGE F, 1988, CEPHALALGIA, V8, P1, DOI 10.1046/j.1468-2982.1988.0801001.x
   Sundaram PK, 1999, NEUROL INDIA, V47, P94
   TOMSAK RL, 1991, MED CLIN N AM, V75, P693, DOI 10.1016/S0025-7125(16)30443-6
NR 15
TC 3
Z9 4
U1 0
U2 2
PU SPRINGER-VERLAG
PI NEW YORK
PA 175 FIFTH AVE, NEW YORK, NY 10010 USA
SN 1590-1874
J9 NEUROL SCI
JI Neurol. Sci.
PD NOV
PY 2003
VL 24
IS 4
BP 281
EP 285
DI 10.1007/s10072-003-0157-4
PG 5
WC Clinical Neurology; Neurosciences
WE Conference Proceedings Citation Index - Science (CPCI-S); Science Citation Index Expanded (SCI-EXPANDED)
SC Neurosciences & Neurology
GA 749AM
UT WOS:000186896500016
PM 14658050
DA 2023-08-27
ER

PT J
AU Sanna, M
   Taibah, A
   Russo, A
   Falcioni, M
   Agarwal, M
AF Sanna, M
   Taibah, A
   Russo, A
   Falcioni, M
   Agarwal, M
TI Perioperative complications in acoustic neuroma (vestibular schwannoma)
   surgery
SO OTOLOGY & NEUROTOLOGY
LA English
DT Article
DE acoustic neuroma surgery; surgical complication; translabyrinthine
   approach; vestibular schwannoma
ID CEREBROSPINAL-FLUID LEAKS; TRANSLABYRINTHINE APPROACH; REMOVAL
AB Objective: Retrospective study and review of the complications other than those related to the facial nerve and hearing, encountered in acoustic neuroma surgery. Also, an evaluation of hospital stay and its relation with various factors.
   Study Design: Retrospective case review.
   Setting: Tertiary neurotologic and skull base referral center.
   Patients: A series of 707 patients who underwent surgical removal of acoustic neuroma from April 1987 to December 2001.
   Interventions: The surgical approaches used were the enlarged translabyrinthine approach, the enlarged middle fossa approach, and the retrosigmoid approach. In a small number of cases, the operations were performed through other approaches.
   Main Outcome Measures: The duration of hospital stay and appearance of complications in the perioperative period along with their management. Results related to the facial nerve and hearing were not considered in this study.
   Results: The most frequent complication was abdominal subcutaneous hematoma (site of fat harvest), which Occurred in 23 patients (3.2%). Cerebrospinal fluid leak was present in 20 patients (2.8%), 15 of whom needed revision surgery. Other complications included VIth cranial nerve dysfunction in 12 cases (1.68%), subdural hematoma in 3 cases (0.4%), cerebel-lopontine angle hematoma in 4 cases (0.6%), cerebellar edema in 2 cases (0.28%), brainstem hematoma in 1 case (0.14%), transitory aphasia in I case (0.14%), and lower cranial nerve dysfunction in I case (0.14%). Mortality occurred in only one case (0.14%). Medical complications seldom occurred. The postoperative hospital stay ranged from 2 to 36 days, with an average of 6.4 days. The overall hospital stay diminished over time from 10.2 days in 1987 to 1990, to 4.9 days in 2001. There was a significant relation between hospital stay and tumor size, approach used, and presence/absence of complications.
   Conclusions: Perioperative complications in acoustic neuroma surgery do exist, but this study demonstrated how low the incidence is. The authors believe that the low percentage of complications is mainly attributable to the majority of operations being carried out in specialized clinics, where they are considered routine operations. They believe that following individualized approaches, depending on tumor size and on the preoperative function of the cranial nerves, is the proper way to reach a significant reduction in complications while maintaining a high percentage of total tumor removal. The results of this study, considered as a basis of comparison with other studies, will certainly be useful in preoperative patient counseling.
C1 Grp Otol, I-29100 Rome, Italy.
RP Sanna, M (通讯作者)，Grp Otol, Via Emmanueli 42, I-29100 Rome, Italy.
EM
CR BRYCE GE, 1991, OTOLARYNG HEAD NECK, V104, P81, DOI 10.1177/019459989110400115
   CUEVA R, 1999, ACOUSTIC NEURINOMA O, P1073
   Falcioni M, 1999, AM J OTOL, V20, P660
   FALCIONI M, 2002, ACTA OTORHINOLARYNGO, V20, P151
   GLASSCOCK ME, 1982, OTOLARYNG CLIN N AM, V15, P883
   HOFFMAN RA, 1994, LARYNGOSCOPE, V104, P40
   KANZAKI J, 1991, ACTA OTO-LARYNGOL, P75
   Lanman TH, 1999, J NEUROSURG, V90, P617, DOI 10.3171/jns.1999.90.4.0617
   MAGNAN J, 1999, ENDOSCOPY NEUROTOLOG
   Mass SC, 1999, ARCH OTOLARYNGOL, V125, P801, DOI 10.1001/archotol.125.7.801
   NADOL JB, 1987, LARYNGOSCOPE, V97, P1287, DOI 10.1288/00005537-198711000-00007
   NAGUIB MB, 1994, J LARYNGOL OTOL, V108, P545, DOI 10.1017/S0022215100127392
   PICCIONI LO, 1999, ACOUSTIC NEURINOMA O, P1077
   Samii M, 1997, NEUROSURGERY, V40, P11, DOI 10.1097/00006123-199701000-00002
   SAMPATH P, 1999, ACOUSTIC NEURINOMA O, P527
   Sanna M, 2002, OTOL NEUROTOL, V23, P980, DOI 10.1097/00129492-200211000-00028
   SANNA M, 1998, ATLAS ACOUSTIC NEURI
   Slattery WH, 2001, OTOL NEUROTOL, V22, P895, DOI 10.1097/00129492-200111000-00031
   THOMSEN J, 1992, P 1 INT C AC NEUR CO, P331
   Wiegand DA, 1996, LARYNGOSCOPE, V106, P58, DOI 10.1097/00005537-199601000-00012
NR 20
TC 97
Z9 106
U1 0
U2 6
PU LIPPINCOTT WILLIAMS & WILKINS
PI PHILADELPHIA
PA TWO COMMERCE SQ, 2001 MARKET ST, PHILADELPHIA, PA 19103 USA
SN 1531-7129
EI 1537-4505
J9 OTOL NEUROTOL
JI Otol. Neurotol.
PD MAY
PY 2004
VL 25
IS 3
BP 379
EP 386
DI 10.1097/00129492-200405000-00029
PG 8
WC Clinical Neurology; Otorhinolaryngology
WE Science Citation Index Expanded (SCI-EXPANDED)
SC Neurosciences & Neurology; Otorhinolaryngology
GA 818SV
UT WOS:000221265700029
PM 15129121
DA 2023-08-27
ER

PT J
AU Aboelwafa, HR
   Ramadan, RA
   Ibraheim, SS
   Yousef, HN
AF Aboelwafa, Hanaa R.
   Ramadan, Ramadan A.
   Ibraheim, Somaya S.
   Yousef, Hany N.
TI Modulation Effects of Eugenol on Nephrotoxicity Triggered by Silver
   Nanoparticles in Adult Rats
SO BIOLOGY-BASEL
LA English
DT Article
DE silver nanoparticles; eugenol; kidney; oxidative stress; histology;
   histopathology; immunohistochemistry
ID REPEATED-DOSE TOXICITY; ANTIOXIDANT ACTIVITY; ESSENTIAL OIL; STRESS;
   IMMUNOTOXICITY; HEPATOTOXICITY; CYTOTOXICITY; MECHANISMS; PATHOLOGY;
   CURCUMIN
AB Simple Summary The main objective of this study is to assess how eugenol (Eug) affects AgNP-induced nephrotoxicity in rats. After 30 days of treatment with AgNPs, rats developed nephrotoxicity, which was characterized by disruptions in the serum levels of blood urea nitrogen, creatinine, uric acid, the total oxidant capacity, the total antioxidant capacity, and interfering levels of kidney injury molecule-1, superoxide dismutase, catalase, reduced glutathione, glutathione peroxidase, malondialdehyde, tumor necrosis factor-alpha (TNF-alpha), and interleukin-6 in kidney tissues. These biochemical alterations were accompanied by the destruction of normal renal architecture, with most renal components shedding their thickness, diameter, and quantity. Furthermore, P53, Caspase3, and TNF-alpha immunoreactivity were considerably elevated; however, Bcl-2 immunoreactivity was reduced. Most biochemical, histological, histomorphometrical, and immunohistochemical alterations in AgNP-treated rats were reversed by Eug. We may infer that Eug has a protective effect against AgNP-induced nephrotoxicity. The use of silver nanoparticles (AgNPs) is expanding. This study evaluates the modulator effect of eugenol (Eug) on AgNP-induced nephrotoxicity in rats. Sixty male rats were separated into six groups: control, Eug, AgNPs low-dose, AgNPs high-dose, Eug + AgNPs low-dose, and Eug + AgNPs high-dose. After 30 days, kidney function, antioxidative and proinflammatory status, histopathological, histomorphometrical, and immunohistochemical assessments were performed. AgNPs markedly induced oxidative stress in renal tissues, characterized by increased levels of blood urea nitrogen, creatinine, uric acid, kidney injury molecule-1, the total oxidant capacity, malondialdehyde, tumor necrosis factor-alpha (TNF-alpha), and interleukin-6, as well as decreased levels of the total antioxidant capacity, superoxide dismutase, catalase, reduced glutathione, and glutathione peroxidase. Moreover, the normal renal architecture was destroyed, and the thickness of the renal capsules, cortex, and medulla, alongside the diameter and quantity of the normal Malpighian corpuscles and the proximal and distal convoluted tubules were decreased. Immunoreactivity for P53, caspase-3, and TNF-alpha reactive proteins were significantly increased; however, Bcl-2 immunoreactivity was decreased. Eug reversed most biochemical, histological, histomorphometrical, and immunohistochemical changes in AgNP-treated animals. This study demonstrated that nephrotoxicity in AgNP-treated rats was mitigated by an Eug supplementation. Eug's antioxidant, antiapoptotic, and anti-inflammatory capabilities were the key in modulating AgNPs nephrotoxicity.
C1 [Aboelwafa, Hanaa R.; Ramadan, Ramadan A.; Ibraheim, Somaya S.; Yousef, Hany N.] Ain Shams Univ, Fac Educ, Biol & Geol Sci Dept, Cairo 11566, Egypt.
C3 Egyptian Knowledge Bank (EKB); Ain Shams University
RP Yousef, HN (通讯作者)，Ain Shams Univ, Fac Educ, Biol & Geol Sci Dept, Cairo 11566, Egypt.
EM
OI Yousef, Hany/0000-0001-9795-2363
CR Abdelhalim MAK, 2011, LIPIDS HEALTH DIS, V10, DOI 10.1186/1476-511X-10-147
   Abdelhalim MAK, 2013, SAUDI J BIOL SCI, V20, P177, DOI 10.1016/j.sjbs.2013.01.007
   AEBI H, 1984, METHOD ENZYMOL, V105, P121
   Bachiega TF, 2012, J PHARM PHARMACOL, V64, P610, DOI 10.1111/j.2042-7158.2011.01440.x
   Banu H, 2018, BIOL TRACE ELEM RES, V186, P122, DOI 10.1007/s12011-018-1287-0
   Batiha GE, 2020, BIOMOLECULES, V10, DOI 10.3390/biom10030352
   BEUTLER E, 1963, J LAB CLIN MED, V61, P882
   Bianchi S, 1999, AM J KIDNEY DIS, V34, P973, DOI 10.1016/S0272-6386(99)70002-8
   Binu P, 2018, IRAN J MED SCI, V43, P305
   Bonventre JV, 2008, SCAND J CLIN LAB INV, V68, P78, DOI 10.1080/00365510802145059
   Brieger K, 2012, SWISS MED WKLY, V142, DOI 10.4414/smw.2012.13659
   Buege J A, 1978, Methods Enzymol, V52, P302
   Chan SL, 2004, CLIN EXP PHARMACOL P, V31, P119, DOI 10.1111/j.1440-1681.2004.03975.x
   Choi CHJ, 2011, P NATL ACAD SCI USA, V108, P6656, DOI 10.1073/pnas.1103573108
   Cory S, 2002, NAT REV CANCER, V2, P647, DOI 10.1038/nrc883
   Daniel AN, 2009, REV BRAS FARMACOGN, V19, P212, DOI 10.1590/S0102-695X2009000200006
   De Jong WH, 2013, BIOMATERIALS, V34, P8333, DOI 10.1016/j.biomaterials.2013.06.048
   Dedman CJ, 2020, SCI TOTAL ENVIRON, V747, DOI 10.1016/j.scitotenv.2020.141229
   Del Monte U, 2005, MED HYPOTHESES, V64, P818, DOI 10.1016/j.mehy.2004.08.028
   El Mahdy MM, 2015, EXP TOXICOL PATHOL, V67, P21, DOI 10.1016/j.etp.2014.09.005
   Elkhateeb SA, 2020, J BIOCHEM MOL TOXIC, V34, DOI 10.1002/jbt.22593
   Epstein FH, 1997, KIDNEY INT, V51, P381, DOI 10.1038/ki.1997.50
   Evenepoel P, 2010, MANAGEMENT OF ACUTE KIDNEY PROBLEMS, P317, DOI 10.1007/978-3-540-69441-0_6.6
   Fehaid A., 2020, OPEN BIOTECHNOL J, V14, P113, DOI 10.2174/1874070702014010113
   Ferdous Z, 2020, INT J MOL SCI, V21, DOI 10.3390/ijms21072375
   Fitzgerald DC, 2007, VET IMMUNOL IMMUNOP, V116, P59, DOI 10.1016/j.vetimm.2006.12.008
   Fogo A.B., 2014, FUNDAMENTALS RENAL P, P167
   Forni C, 2019, BIOMED RES INT-UK, V2019, DOI 10.1155/2019/8748253
   Galatage ST, 2021, SILVER MICRONANOPART, P61, DOI [10.5772/intechopen:92480, DOI 10.5772/INTECHOPEN.99173]
   Gamble M., 2008, THEORY PRACTICE HIST, P83
   Garud MS, 2017, FLAVOUR FRAG J, V32, P54, DOI 10.1002/ffj.3357
   Gherkhbolagh MH, 2018, J RES MED DENT SCI, V6, P43, DOI 10.24896/jrmds.2018619
   Gibson G.G., 1986, INTRO DRUG METABOLIS, P175
   Gois PHF, 2016, FREE RADICAL BIO MED, V101, P176, DOI 10.1016/j.freeradbiomed.2016.10.012
   Gowda S, 2010, N AM J MED SCI, V2, P170
   Gulcin I, 2011, J MED FOOD, V14, P975, DOI 10.1089/jmf.2010.0197
   Guo HT, 2015, NAT MED, V21, P677, DOI 10.1038/nm.3893
   Guo H, 2016, PART FIBRE TOXICOL, V13, DOI 10.1186/s12989-016-0133-9
   Hussain SM, 2005, TOXICOL IN VITRO, V19, P975, DOI 10.1016/j.tiv.2005.06.034
   Inumaru J, 2009, GENES CELLS, V14, P703, DOI 10.1111/j.1365-2443.2009.01303.x
   Johar D, 2004, Rocz Akad Med Bialymst, V49, P31
   Khalil AA, 2017, RSC ADV, V7, P32669, DOI 10.1039/c7ra04803c
   Koracevic D, 2001, J CLIN PATHOL, V54, P356, DOI 10.1136/jcp.54.5.356
   Kunak CS, 2016, BRIT J RADIOL, V89, DOI 10.1259/bjr.20140724
   Li FJ, 2018, THORAC CANCER, V9, P25, DOI 10.1111/1759-7714.12508
   Li SQ, 2008, FOOD CHEM TOXICOL, V46, P3626, DOI 10.1016/j.fct.2008.09.012
   Liu Y, 2020, ARH HIG RADA TOKSIKO, V71, P121, DOI 10.2478/aiht-2020-71-3364
   Loh AHL, 2009, ANN ACAD MED SINGAP, V38, P240
   LOWRY OH, 1951, J BIOL CHEM, V193, P265
   Ma WR, 2015, INT J BIOL SCI, V11, P860, DOI 10.7150/ijbs.12059
   Mak KitKay, 2019, Pharmacognosy Reviews, V13, P1, DOI 10.4103/phrev.phrev_46_18
   Markakis Charalampos, 2016, HPB Surg, V2016, P3203147, DOI 10.1155/2016/3203147
   Markowitz GS, 2005, CLIN CHIM ACTA, V351, P31, DOI 10.1016/j.cccn.2004.09.005
   May P, 1999, ONCOGENE, V18, P7621, DOI 10.1038/sj.onc.1203285
   McShan D, 2014, J FOOD DRUG ANAL, V22, P116, DOI 10.1016/j.jfda.2014.01.010
   Mishra AK, 2009, ANN CLIN MICROB ANTI, V8, DOI 10.1186/1476-0711-8-9
   Moradi-Sardareh H, 2018, LIFE SCI, V211, P81, DOI 10.1016/j.lfs.2018.09.001
   NISHIKIMI M, 1972, BIOCHEM BIOPH RES CO, V46, P849, DOI 10.1016/S0006-291X(72)80218-3
   Nosrati H, 2021, BMC NEPHROL, V22, DOI 10.1186/s12882-021-02428-5
   Oeckinghaus A, 2011, NAT IMMUNOL, V12, P695, DOI 10.1038/ni.2065
   PAGLIA DE, 1967, J LAB CLIN MED, V70, P158
   Pandey Govind, 2008, Toxicology International, V15, P69
   Pannu N, 2008, CRIT CARE MED, V36, pS216, DOI 10.1097/CCM.0b013e318168e375
   Patlolla AK, 2015, MOL CELL BIOCHEM, V399, P257, DOI 10.1007/s11010-014-2252-7
   Perazella MA, 2012, KIDNEY INT, V81, P1172, DOI 10.1038/ki.2010.475
   Perazella MA, 2009, CLIN J AM SOC NEPHRO, V4, P1275, DOI 10.2215/CJN.02050309
   Petrosyan K, 2002, J HISTOTECHNOL, V25, P247
   Pondman KM, 2015, NANOMED-NANOTECHNOL, V11, P2109, DOI 10.1016/j.nano.2015.06.009
   Pourhamzeh M, 2016, AVICENNA J MED BIOCH, V4, DOI [10.17795/ajmb-35557, DOI 10.17795/AJMB-35557]
   Prasath S, 2019, ENVIRON GEOCHEM HLTH, V41, P2295, DOI 10.1007/s10653-019-00240-7
   Reddy ACP, 1996, TOXICOLOGY, V107, P39, DOI 10.1016/0300-483X(95)03199-P
   REDDY ACP, 1994, MOL CELL BIOCHEM, V137, P1, DOI 10.1007/BF00926033
   Said MM, 2011, FUND CLIN PHARMACOL, V25, P708, DOI 10.1111/j.1472-8206.2010.00900.x
   Schrand AM, 2010, WIRES NANOMED NANOBI, V2, P544, DOI 10.1002/wnan.103
   Shi YF, 2017, AM J PHYSIOL-RENAL, V312, pF502, DOI 10.1152/ajprenal.00546.2016
   Silva Fred G, 2004, Toxicol Pathol, V32 Suppl 2, P71, DOI 10.1080/01926230490457530
   Sulaiman FA, 2015, J ACUTE MED, V5, P96, DOI 10.1016/j.jacme.2015.09.005
   Syafiuddin A, 2017, J CHIN CHEM SOC-TAIP, V64, P732, DOI 10.1002/jccs.201700067
   Tatzber F, 2003, ANAL BIOCHEM, V316, P147, DOI 10.1016/S0003-2697(02)00652-8
   Temizel-Sekeryan S, 2020, RESOUR CONSERV RECY, V156, DOI [10.1016/j.resconrec.2020.104676, 10.1016/j.resconrec.2019.104676]
   Tietz NWE., 1976, FUNDAMENTALS CLIN CH
   Tiku AB, 2004, J RADIAT RES, V45, P435, DOI 10.1269/jrr.45.435
   Tucker PS, 2015, OXID MED CELL LONGEV, V2015, DOI 10.1155/2015/806358
   Van Eycke YR, 2017, SCI REP-UK, V7, P1, DOI 10.1038/srep42964
   Vandebriel RJ, 2014, PART FIBRE TOXICOL, V11, DOI 10.1186/1743-8977-11-21
   Wang F, 2009, TOXICOL IN VITRO, V23, P808, DOI 10.1016/j.tiv.2009.04.009
   Yang FC, 2016, SCI REP-UK, V6, DOI 10.1038/srep21714
   Yang Yoon Mee, 2015, Curr Pathobiol Rep, V3, P253
   Yen HJ, 2009, SMALL, V5, P1553, DOI 10.1002/smll.200900126
   Yousef HN, 2022, OXID MED CELL LONGEV, V2022, DOI 10.1155/2022/3820848
   Zahran HY, 2019, MATER RES EXPRESS, V6, DOI 10.1088/2053-1591/ab30a4
   Zhang XF, 2016, INT J MOL SCI, V17, DOI 10.3390/ijms17091534
NR 92
TC 0
Z9 0
U1 1
U2 4
PU MDPI
PI BASEL
PA ST ALBAN-ANLAGE 66, CH-4052 BASEL, SWITZERLAND
EI 2079-7737
J9 BIOLOGY-BASEL
JI Biology-Basel
PD DEC
PY 2022
VL 11
IS 12
AR 1719
DI 10.3390/biology11121719
PG 23
WC Biology
WE Science Citation Index Expanded (SCI-EXPANDED)
SC Life Sciences & Biomedicine - Other Topics
GA 7G2KO
UT WOS:000902360300001
PM 36552229
OA Green Published, gold
DA 2023-08-27
ER

PT J
AU af Rosenschold, PM
   Law, I
   Engelholm, S
   Engelholm, SA
   Muhic, A
   Lundemann, MJ
   Roed, H
   Grunnet, K
   Poulsen, HS
AF af Rosenschold, P. Munck
   Law, I
   Engelholm, S.
   Engelholm, S. A.
   Muhic, A.
   Lundemann, M. J.
   Roed, H.
   Grunnet, K.
   Poulsen, H. Skovgaard
TI Influence of volumetric modulated arc therapy and FET-PET scanning on
   treatment outcomes for glioblastoma patients
SO RADIOTHERAPY AND ONCOLOGY
LA English
DT Article
DE Glioblastoma; Radiotherapy; FET-PET; VMAT; IGRT; Technology
ID RADIOTHERAPY PLUS CONCOMITANT; RADIATION-THERAPY; CONFORMAL
   RADIOTHERAPY; SUBVENTRICULAR ZONE; PROGNOSTIC VALUE; PROTON THERAPY;
   F-18-FET PET; FAILURE; TEMOZOLOMIDE; MULTIFORME
AB Background: We sought to assess the influence of the clinical introduction of new radiotherapy technologies on glioblastoma patients' outcomes.
   Methods: Newly diagnosed glioblastoma patients treated with 60 Gy and temozolomide (2005-2014) were analyzed. The patients' GTV and CTV were defined based on MR (n = 521) or FET-PET/MR (n = 190), and were treated using conformal radiotherapy (CRT, n = 159) or image-guided volumetric modulated arc therapy with hippocampal sparing (IG-VMAT, n = 362). Progression-free survival (PFS) was assessed using the McDonald criteria. Associations between clinical data, dosimetry data, treatment technology, for PFS and overall survival (OS) were explored.
   Results: The PFS (7 months) and OS (15 months) were unaffected by CRT, IG-VMAT and FET-PET technology. Mean brain dose was correlated with tumor volume, and was lower for IG-VMAT vs. CRT (p < 0.001). Larger mean brain dose was associated with inferior PFS (univariate/multivariate Cox models, p < 0.001) and OS (univariate, p < 0.001). Multivariate Cox models revealed association of larger mean brainstem dose (p < 0.001), BTV (p = 0.045), steroid use at baseline (p = 0.003), age (p = 0.019) and MGMT status (p = 0.022) with lower OS.
   Conclusions: Introduction of hippocampal-sparing IG-VMAT technology appeared to be safe, and may have reduced toxicity and cognitive impairment. Larger mean brain dose was strongly associated with inferior PFS and OS. (C) 2018 Elsevier B.V. All rights reserved.
C1 [af Rosenschold, P. Munck; Engelholm, S.; Engelholm, S. A.; Muhic, A.; Lundemann, M. J.; Roed, H.; Grunnet, K.; Poulsen, H. Skovgaard] Rigshosp, Dept Oncol, Copenhagen, Denmark.
   [af Rosenschold, P. Munck; Lundemann, M. J.] Univ Copenhagen, Niels Bohr Inst, Copenhagen, Denmark.
   [Law, I; Lundemann, M. J.] Rigshosp, Dept Clin Physiol Nucl Med & PET, Copenhagen, Denmark.
C3 Rigshospitalet; University of Copenhagen; University of Copenhagen;
   Niels Bohr Institute; Rigshospitalet; University of Copenhagen
RP af Rosenschold, PM (通讯作者)，Skane Univ Hosp, Radiat Phys, Lund, Sweden.
EM
RI ; Munck af Rosenschold, Per/L-9368-2016
OI Lundemann, Michael/0000-0002-4216-5757; Munck af Rosenschold,
   Per/0000-0002-5988-8994
FU Lundbeck Foundation
FX The Lundbeck Foundation is acknowledged for financial support.
CR Adeberg S, 2016, STRAHLENTHER ONKOL, V192, P770, DOI 10.1007/s00066-016-1007-7
   af Rosenschold PM, 2015, NEURO-ONCOLOGY, V17, P757, DOI 10.1093/neuonc/nou316
   BRAHME A, 1982, PHYS MED BIOL, V27, P1221, DOI 10.1088/0031-9155/27/10/002
   Ellingson BM, 2013, AM J NEURORADIOL, V34, P533, DOI 10.3174/ajnr.A3253
   Gebhardt BJ, 2014, RADIAT ONCOL, V9, DOI 10.1186/1748-717X-9-130
   Gondi V, 2013, INT J RADIAT ONCOL, V85, P348, DOI 10.1016/j.ijrobp.2012.11.031
   Grosu AL, 2010, RADIOTHER ONCOL, V96, P325, DOI 10.1016/j.radonc.2010.08.001
   Grosu Anca-Ligia, 2009, Eur J Cancer, V45 Suppl 1, P461, DOI 10.1016/S0959-8049(09)70090-5
   Gzell C, 2016, CLIN ONCOL R COLL RA
   Harrabi S, 2016, STRAHLENTHER ONKOL, V192, P759, DOI 10.1007/s00066-016-1005-9
   Jafri NF, 2013, NEURO-ONCOLOGY, V15, P91, DOI 10.1093/neuonc/nos268
   Jansen NL, 2015, J NUCL MED, V56, P9, DOI 10.2967/jnumed.114.144675
   Kristensen LS, 2016, J NEUROPATH EXP NEUR, V75, P246, DOI 10.1093/jnen/nlv024
   Lawrence YR, 2010, INT J RADIAT ONCOL, V76, pS20, DOI 10.1016/j.ijrobp.2009.02.091
   Lee IH, 2009, INT J RADIAT ONCOL, V73, P479, DOI 10.1016/j.ijrobp.2008.04.050
   Lee P, 2013, INT J RADIAT ONCOL, V86, P609, DOI 10.1016/j.ijrobp.2013.01.009
   Lim DA, 2007, NEURO-ONCOLOGY, V9, P424, DOI 10.1215/15228517-2007-023
   Lundemann M, 2017, RADIOTHER ONCOL J EU
   Mayo C, 2010, INT J RADIAT ONCOL, V76, pS28, DOI 10.1016/j.ijrobp.2009.07.1753
   Mayo C, 2010, INT J RADIAT ONCOL, V76, pS36, DOI 10.1016/j.ijrobp.2009.08.078
   McDonald MW, 2011, INT J RADIAT ONCOL, V79, P130, DOI 10.1016/j.ijrobp.2009.10.048
   Michaelsen SR, 2013, BMC CANCER, V13, DOI 10.1186/1471-2407-13-402
   Minniti G, 2010, RADIOTHER ONCOL, V97, P377, DOI 10.1016/j.radonc.2010.08.020
   Moller S, 2016, RADIOTHER ONCOL, V121, P132, DOI 10.1016/j.radonc.2016.08.014
   Munck AF, 2011, ACTA ONCOL, V50, P777, DOI DOI 10.3109/0284186X.2011.584555
   Narayana A, 2006, INT J RADIAT ONCOL, V64, P892, DOI 10.1016/j.ijrobp.2005.05.067
   Navarria P, 2016, BMC CANCER, V16, DOI 10.1186/s12885-016-2399-6
   Niyazi M, 2011, RADIOTHER ONCOL, V99, P44, DOI 10.1016/j.radonc.2011.03.001
   Pauleit D, 2005, BRAIN, V128, P678, DOI 10.1093/brain/awh399
   Paulsson AK, 2014, AM J CLIN ONCOL-CANC, V37, P177, DOI 10.1097/COC.0b013e318271ae03
   Piroth MD, 2011, RADIOTHER ONCOL, V99, P218, DOI 10.1016/j.radonc.2011.03.006
   Piroth MD, 2011, INT J RADIAT ONCOL, V80, P176, DOI 10.1016/j.ijrobp.2010.01.055
   Poulsen SH, 2016, EUR J NUCL MED MOL I
   Rieken S, 2013, RADIOTHER ONCOL, V109, P487, DOI 10.1016/j.radonc.2013.06.043
   Stockhammer F, 2008, J NEURO-ONCOL, V88, P205, DOI 10.1007/s11060-008-9551-3
   Stupp R, 2005, NEW ENGL J MED, V352, P987, DOI 10.1056/NEJMoa043330
   Suchorska B, 2015, NEUROLOGY, V84, P710, DOI 10.1212/WNL.0000000000001262
   Tarnavski N, 2016, J RADIOSURGERY SBRT, V4, P15
   Urup T, 2016, MOL ONCOL, V10, P1160, DOI 10.1016/j.molonc.2016.05.005
   van Herk M, 2000, INT J RADIAT ONCOL, V47, P1121, DOI 10.1016/S0360-3016(00)00518-6
   Verellen D, 2007, NAT REV CANCER, V7, P949, DOI 10.1038/nrc2288
   Wagner D, 2009, RADIOTHER ONCOL, V93, P593, DOI 10.1016/j.radonc.2009.10.002
   Weber DC, 2009, RADIOTHER ONCOL, V93, P586, DOI 10.1016/j.radonc.2009.08.043
   Weber DC, 2008, RADIAT ONCOL, V3, DOI 10.1186/1748-717X-3-44
NR 44
TC 8
Z9 8
U1 0
U2 3
PU ELSEVIER IRELAND LTD
PI CLARE
PA ELSEVIER HOUSE, BROOKVALE PLAZA, EAST PARK SHANNON, CO, CLARE, 00000,
   IRELAND
SN 0167-8140
EI 1879-0887
J9 RADIOTHER ONCOL
JI Radiother. Oncol.
PD JAN
PY 2019
VL 130
BP 149
EP 155
DI 10.1016/j.radonc.2018.10.003
PG 7
WC Oncology; Radiology, Nuclear Medicine & Medical Imaging
WE Science Citation Index Expanded (SCI-EXPANDED)
SC Oncology; Radiology, Nuclear Medicine & Medical Imaging
GA HI8MH
UT WOS:000456709600021
PM 30446316
DA 2023-08-27
ER

PT J
AU Arts, HA
   Telian, SA
   El-Kashlan, H
   Thompson, BG
AF Arts, HA
   Telian, SA
   El-Kashlan, H
   Thompson, BG
TI Hearing preservation and facial nerve outcomes in vestibular schwannoma
   surgery: Results using the middle cranial fossa approach
SO OTOLOGY & NEUROTOLOGY
LA English
DT Article
DE facial nerve; hearing preservation; middle cranial fossa; outcomes;
   vestibular schwannoma
ID ACOUSTIC NEUROMA SURGERY
AB Objective: To evaluate surgical results using the middle cranial fossa approach for hearing preservation vestibular schwannoma surgery.
   Study Design: Retrospective case review.
   Setting: Tertiary referral academic center.
   Patients: Seventy-three consecutive patients with vestibular schwannoma operated on using the middle cranial fossa approach between February 1999 and February 2005.
   Interventions: The tumors were removed via the middle cranial fossa approach with modifications to improve exposure. Standard auditory brainstem and facial nerve monitoring were used.
   Main Outcome Measures: Pre- and postoperative hearing measures and facial function, tumor size, and postoperative complications. Hearing status was categorized into Classes A, B, C, and D as described by the American Academy of Otolaryngology-Head and Heck Surgery "Guidelines for the Evaluation of Hearing Preservation in Acoustic Neuroma, 1995."
   Results: Thirty-four patients presented with Class A hearing preoperatively. Among patients presenting with Class A hearing, a total of 27 (80%) maintained Class A or B hearing postoperatively. Of these, 21 (62%) remained in Class A, 6 (18%) deteriorated slightly to Class B, and 7 (20%) deteriorated to Class D postoperatively. Twenty-eight patients presented with Class B hearing preoperatively. Of these, 18 (64%) remained in Class B, 3 (11%) deteriorated to Class C, and 7 (25%) deteriorated to Class D. Three patients had Class C hearing preoperatively. Of these, 2 (66%) remained in Class C and 1 (33%) deteriorated to Class D. Eight patients presented in Class D and one of these improved to Class C postoperatively. Overall, 62 patients presented with useful (Class A or B) hearing and 45 (73%) remained in Class A or B. Nineteen patients had tumors larger than 10 mm in greatest dimension and had Class A or B hearing preoperatively. Of these, 11 (58%) retained Class A or B hearing postoperatively. At 4 months or greater follow-up, facial nerve outcome were excellent in 96%: House-Brackmann Grade I in 61 (85%), Grade II in 8 (11%), and Grade III in 3 (4%). There were no Grade IV, V, or VI results on final follow-up. Six (8%) patients developed cerebrospinal fluid leaks.
   Conclusion: By achieving excellent exposure and using meticulous microsurgical technique, it is possible to resect small vestibular schwannomas via the middle fossa approach, with preservation of hearing at excellent or preoperative levels in the majority of patients, with excellent or satisfactory facial nerve outcomes in 96% of patients.
C1 Univ Michigan, Dept Otolaryngol Head & Neck Surg, Div Otol Neurotol, Sch Med, Ann Arbor, MI 48109 USA.
   Univ Michigan, Dept Neurosurg, Sch Med, Ann Arbor, MI 48109 USA.
C3 University of Michigan System; University of Michigan; University of
   Michigan System; University of Michigan
RP Arts, HA (通讯作者)，Univ Michigan, Dept Otolaryngol Head & Neck Surg, Div Otol Neurotol, Sch Med, 1500 E Med Ctr Dr, Ann Arbor, MI 48109 USA.
EM
CR Arriaga MA, 1997, LARYNGOSCOPE, V107, P1043, DOI 10.1097/00005537-199708000-00007
   Betchen SA, 2005, J NEUROSURG, V102, P6, DOI 10.3171/jns.2005.102.1.0006
   BRACKMANN DE, 1994, AM J OTOL, V15, P614
   Colletti V, 2005, OTOLARYNG HEAD NECK, V132, P459, DOI 10.1016/j.otohns.2004.09.027
   Eisenman DJ, 1999, AM J OTOL, V20, P77
   Gjuric M, 2001, OTOL NEUROTOL, V22, P223, DOI 10.1097/00129492-200103000-00019
   Hecht CS, 1997, LARYNGOSCOPE, V107, P1122, DOI 10.1097/00005537-199708000-00021
   Holsinger FC, 2000, AM J OTOL, V21, P695
   HOUSE JW, 1985, OTOLARYNG HEAD NECK, V93, P146, DOI 10.1177/019459988509300202
   HOUSE W F, 1961, Laryngoscope, V71, P1363
   Kanzaki J, 2003, OTOL NEUROTOL, V24, P642, DOI 10.1097/00129492-200307000-00019
   Mohr G, 2005, J NEUROSURG, V102, P1, DOI 10.3171/jns.2005.102.1.0001
   MONSELL EM, 1995, OTOLARYNG HEAD NECK, V113, P179
   MONSELL EM, 1995, OTOLARYNG HEAD NECK, V113, P176, DOI 10.1016/S0194-5998(95)70100-1
   Rappaport JM, 1999, OTOLARYNG HEAD NECK, V121, P176, DOI 10.1016/S0194-5998(99)70167-1
   Sanna M, 2004, ANN OTO RHINOL LARYN, V113, P156, DOI 10.1177/000348940411300215
   SANNA M, 1987, AM J OTOL, V8, P500
   Slatterly W, 1997, AM J OTOL, V18, P796
   Slattery WH, 1997, AM J OTOL, V18, P596
   Staecker H, 2000, AM J OTOL, V21, P399, DOI 10.1016/S0196-0709(00)80051-4
   THORNTON AR, 1978, J SPEECH HEAR RES, V21, P507, DOI 10.1044/jshr.2103.507
   Weber PC, 1996, AM J OTOL, V17, P669
NR 22
TC 84
Z9 92
U1 0
U2 3
PU LIPPINCOTT WILLIAMS & WILKINS
PI PHILADELPHIA
PA TWO COMMERCE SQ, 2001 MARKET ST, PHILADELPHIA, PA 19103 USA
SN 1531-7129
EI 1537-4505
J9 OTOL NEUROTOL
JI Otol. Neurotol.
PD FEB
PY 2006
VL 27
IS 2
BP 234
EP 241
DI 10.1097/01.mao.0000185153.54457.16
PG 8
WC Clinical Neurology; Otorhinolaryngology
WE Science Citation Index Expanded (SCI-EXPANDED)
SC Neurosciences & Neurology; Otorhinolaryngology
GA 012NR
UT WOS:000235346400016
PM 16436995
DA 2023-08-27
ER

PT J
AU Parmer, RJ
   Mahata, M
   Mahata, S
   Sebald, MT
   OConnor, DT
   Miles, LA
AF Parmer, RJ
   Mahata, M
   Mahata, S
   Sebald, MT
   OConnor, DT
   Miles, LA
TI Tissue plasminogen activator (t-PA) is targeted to the regulated
   secretory pathway - Catecholamine storage vesicles as a reservoir for
   the rapid release of t-PA
SO JOURNAL OF BIOLOGICAL CHEMISTRY
LA English
DT Article
ID ADRENAL CHROMAFFIN GRANULES; HUMAN GROWTH-HORMONE; CHROMOGRANIN-A;
   CELL-LINE; PC12 CELLS; ENDOTHELIAL-CELLS; PROTEIN; PHEOCHROMOCYTOMA;
   INHIBITOR; PEPTIDE
AB Tissue-type plasminogen activator (t-PA) is a serine protease that plays a central role in the regulation of intravascular thrombolysis. The acute release of t-PA in vivo is induced by a variety of stimuli including exercise, trauma, and neural stimulation. These types of stimuli also result in sympathoadrenal activation and exocytotic release of amines and proteins from catecholamine storage vesicles of the adrenal medulla and sympathetic neurons. Therefore, we tested the hypothesis that t-PA is packaged in and released directly from catecholamine storage vesicles, using several chromaffin cell sources including the rat pheochromocytoma PC-12 chromaffin cell line, primary cultures of bovine adrenal chromaffin cells, and human pheochromocytoma. t-PA was expressed in chromaffin cells as detected by Northern blotting, immunoprecipitation of [S-35]Met-labeled t-PA, and specific t-PA enzyme-linked immunosorbent assay of cell homogenates. In addition, chromaffin cell t-PA was enzymatically active by fibrin zymography. To explore the subcellular localization of the expressed t-PA, PC-12 cells were labeled with [H-3]norepinephrine, homogenized, and subjected to sucrose density fractionation. [H-3]Norepinephrine and t-PA antigen were colocalized to the same subcellular fraction with a major peak at 1.4 M sucrose, consistent with the buoyant density of catecholamine storage vesicles. In addition, catecholamine storage vesicle lysates isolated from human pheochromocytoma tumors were enriched approximately 30-fold in t-PA antigen, compared with tumor homogenate. Furthermore, exposure of PC-12 cells or primary bovine adrenal chromaffin cells to chromaffin cell secretagogues (60 mu M nicotine, 55 mM KCl, or 2 mM BaCl2) resulted in co-release of t-PA in parallel with catecholamines. These data demonstrate that t-PA is expressed in chromaffin cells, is sorted into the regulated pathway of secretion, and is co released with catecholamines by chromaffin cell stimulation. Catecholamine storage vesicles may be an important reservoir and sympathoadrenal activation an important physiologic mechanism for the rapid release of t-PA. In addition, expression of t-PA by chromaffin cells suggests a role for this protease in the proteolytic processing of chromaffin cell proteins.
C1 UNIV CALIF SAN DIEGO, CTR GENET MOL, SAN DIEGO, CA 92161 USA.
   VET ADM MED CTR, SAN DIEGO, CA 92161 USA.
   SCRIPPS RES INST, LA JOLLA, CA 92037 USA.
C3 University of California System; University of California San Diego; US
   Department of Veterans Affairs; Veterans Health Administration (VHA);
   Scripps Research Institute
RP Parmer, RJ (通讯作者)，UNIV CALIF SAN DIEGO, DEPT MED 9111H, SAN DIEGO, CA 92161 USA.
RI Mahata, Sushil/AAF-8781-2021
FU NHLBI NIH HHS [HL50174, HL45934, HL50398] Funding Source: Medline
CR BANSAL DD, 1992, J CLIN ENDOCR METAB, V74, P266, DOI 10.1210/jcem.74.2.1730806
   BIGGS R, 1947, LANCET, V252, P402
   BRITTON BJ, 1976, THROMB HAEMOSTASIS, V35, P396, DOI 10.1055/s-0038-1647934
   CARMELIET P, 1994, NATURE, V368, P419, DOI 10.1038/368419a0
   CHANAT E, 1991, J CELL BIOL, V115, P1505, DOI 10.1083/jcb.115.6.1505
   CHANDLER WL, 1992, J AM COLL CARDIOL, V19, P1412, DOI 10.1016/0735-1097(92)90596-F
   CHIRGWIN JM, 1979, BIOCHEMISTRY-US, V18, P5294, DOI 10.1021/bi00591a005
   CHUNG KN, 1989, SCIENCE, V243, P192, DOI 10.1126/science.2911732
   COHEN RJ, 1968, LANCET, V2, P1264
   COLLEN D, 1991, BLOOD, V78, P3114
   DESRUISSEAUGONZALVEZ S, 1993, IN VITRO CELL DEV-AN, V29, P161
   DISDIER M, 1992, MOL BIOL CELL, V3, P309, DOI 10.1091/mbc.3.3.309
   EIDEN LE, 1987, NATURE, V325, P301, DOI 10.1038/325301a0
   EMEIS JJ, 1992, ANN NY ACAD SCI, V667, P249, DOI 10.1111/j.1749-6632.1992.tb51622.x
   EMEIS JJ, 1988, TISSUE TYPE PLASMINO, V2, P21
   FANTL P, 1948, AUST J EXP BIOL MED, V26, P521, DOI 10.1038/icb.1948.54
   Frey U, 1996, J NEUROSCI, V16, P2057
   FRIEDMAN GC, 1994, DEV BRAIN RES, V81, P41, DOI 10.1016/0165-3806(94)90066-3
   GOMBAU L, 1994, J BIOL CHEM, V269, P3875
   GREENE LA, 1976, P NATL ACAD SCI USA, V73, P2424, DOI 10.1073/pnas.73.7.2424
   Gualandris A, 1996, J NEUROSCI, V16, P2220
   HELLE KB, 1994, ACTA PHYSIOL SCAND, V152, P1, DOI 10.1111/j.1748-1716.1994.tb09779.x
   HENRY JP, 1994, J EXP BIOL, V196, P251
   HIGAZI AAR, 1994, J BIOL CHEM, V269, P25529
   HUTTNER WB, 1987, NATURE, V325, P305, DOI 10.1038/325305b0
   HUTTNER WB, 1991, TRENDS BIOCHEM SCI, V16, P27, DOI 10.1016/0968-0004(91)90012-K
   KELLY RB, 1985, SCIENCE, V230, P25, DOI 10.1126/science.2994224
   KORSANBENGTSEN K, 1974, SCAND J HAEMATOL, V13, P377
   KRISTENSEN P, 1986, HISTOCHEMISTRY, V85, P431, DOI 10.1007/BF00982674
   KRISTENSEN P, 1985, J CELL BIOL, V101, P305, DOI 10.1083/jcb.101.1.305
   KRYSTOSEK A, 1981, SCIENCE, V213, P1532, DOI 10.1126/science.7197054
   LAEMMLI UK, 1970, NATURE, V227, P680, DOI 10.1038/227680a0
   LEPRINCE P, 1991, J NEUROCHEM, V57, P665, DOI 10.1111/j.1471-4159.1991.tb03798.x
   LEVINE JD, 1982, BLOOD, V60, P531
   LIVETT BG, 1984, PHYSIOL REV, V64, P1103, DOI 10.1152/physrev.1984.64.4.1103
   LOSKUTOFF DJ, 1988, METHOD ENZYMOL, V163, P293
   MACFARLANE RG, 1946, LANCET, V251, P862
   MANNUCCI PM, 1975, BRIT J HAEMATOL, V30, P81, DOI 10.1111/j.1365-2141.1975.tb00521.x
   MEDH RD, 1992, BLOOD, V80, P981
   MILES LA, 1988, J BIOL CHEM, V263, P11928
   MOONEN G, 1982, NATURE, V298, P753, DOI 10.1038/298753a0
   MOORE HPH, 1987, ANN NY ACAD SCI, V493, P50, DOI 10.1111/j.1749-6632.1987.tb27180.x
   OBERLECHNER E, 1982, J NEUROCHEM, V38, P615, DOI 10.1111/j.1471-4159.1982.tb08675.x
   OCONNOR DT, 1994, ANN NY ACAD SCI, V733, P36, DOI 10.1111/j.1749-6632.1994.tb17254.x
   Padro T, 1990, Blood Coagul Fibrinolysis, V1, P601
   Parmer R. J., 1994, Fibrinolysis, V8, P53
   PARMER RJ, 1993, J CLIN INVEST, V92, P1042, DOI 10.1172/JCI116609
   PARMER RJ, 1988, J HYPERTENS, V6, P187, DOI 10.1097/00004872-198803000-00002
   PATZAK A, 1986, J CELL BIOL, V102, P510, DOI 10.1083/jcb.102.2.510
   PIMPLIKAR SW, 1992, J BIOL CHEM, V267, P4110
   PITTMAN RN, 1995, J NEUROCHEM, V64, P566
   PITTMAN RN, 1989, J NEUROSCI, V9, P4269
   QIAN Z, 1993, NATURE, V361, P453, DOI 10.1038/361453a0
   RODA LG, 1980, EXP CELL RES, V128, P103, DOI 10.1016/0014-4827(80)90392-4
   Sambrook J, 1989, MOL CLONING LAB MANU
   SAPPINO AP, 1993, J CLIN INVEST, V92, P679, DOI 10.1172/JCI116637
   SCHLEEF RR, 1989, J CLIN INVEST, V83, P1747, DOI 10.1172/JCI114076
   SCHNECK SA, 1961, NEUROLOGY, V11, P959, DOI 10.1212/WNL.11.11.959
   SCHRAUWEN Y, 1994, FIBRINOLYSIS, V8, P8, DOI 10.1016/0268-9499(94)90230-5
   SCHUBERT D, 1977, P NATL ACAD SCI USA, V74, P5184, DOI 10.1073/pnas.74.11.5184
   SCHWEITZER ES, 1985, J CELL BIOL, V101, P667, DOI 10.1083/jcb.101.2.667
   SCHWEITZER ES, 1990, J CELL SCI, V96, P375
   SEEDS NW, 1992, ANN NY ACAD SCI, V667, P32
   SIMON JP, 1988, P NATL ACAD SCI USA, V85, P1712, DOI 10.1073/pnas.85.5.1712
   SMITH AD, 1967, BIOCHEM J, V103, P480, DOI 10.1042/bj1030480
   TAKIYYUDDIN MA, 1990, HYPERTENSION, V15, P237, DOI 10.1161/01.HYP.15.3.237
   TATEMOTO K, 1986, NATURE, V324, P476, DOI 10.1038/324476a0
   TODD AS, 1959, J PATHOL BACTERIOL, V78, P281, DOI 10.1002/path.1700780131
   TSIRKA SE, 1995, NATURE, V377, P340, DOI 10.1038/377340a0
   VANDENEIJNDENSCHRAUWEN Y, 1995, BLOOD, V85, P3510, DOI 10.1182/blood.V85.12.3510.bloodjournal85123510
   VASSALLI JD, 1991, J CLIN INVEST, V88, P1067, DOI 10.1172/JCI115405
   WARREN BA, 1963, BRIT J EXP PATHOL, V44, P365
   Weil-Malherbe H., 1971, METHODS BIOCH ANAL S, P119
   WINKLER H, 1976, NEUROSCIENCE, V1, P65, DOI 10.1016/0306-4522(76)90001-4
   WINKLER H, 1980, NEUROSCIENCE, V5, P1803, DOI 10.1016/0306-4522(80)90031-7
NR 75
TC 137
Z9 141
U1 0
U2 3
PU AMER SOC BIOCHEMISTRY MOLECULAR BIOLOGY INC
PI BETHESDA
PA 9650 ROCKVILLE PIKE, BETHESDA, MD 20814-3996 USA
SN 0021-9258
EI 1083-351X
J9 J BIOL CHEM
JI J. Biol. Chem.
PD JAN 17
PY 1997
VL 272
IS 3
BP 1976
EP 1982
DI 10.1074/jbc.272.3.1976
PG 7
WC Biochemistry & Molecular Biology
WE Science Citation Index Expanded (SCI-EXPANDED)
SC Biochemistry & Molecular Biology
GA WD058
UT WOS:A1997WD05800087
PM 8999889
OA hybrid
DA 2023-08-27
ER

PT J
AU Xu, LL
   Zhang, M
   Zheng, XB
   Yi, PS
   Lan, C
   Xu, MQ
AF Xu, Liangliang
   Zhang, Ming
   Zheng, Xiaobo
   Yi, Pengsheng
   Lan, Chuan
   Xu, Mingqing
TI The circular RNA ciRS-7 (Cdr1as) acts as a risk factor of hepatic
   microvascular invasion in hepatocellular carcinoma
SO JOURNAL OF CANCER RESEARCH AND CLINICAL ONCOLOGY
LA English
DT Article
DE ciRS-7; microRNA-7; Microvascular invasion; PIK3CD/Akt pathway;
   Prognosis; Hepatocellular carcinoma
ID GROWTH-FACTOR RECEPTOR; ALPHA-FETOPROTEIN; VASCULAR INVASION; SURVIVAL;
   MIR-7; RECURRENCE; RESECTION; EXPRESSION; PREDICTOR; PATHWAY
AB Purpose Circular RNAs (circRNA) represent a novel class of widespread and diverse endogenous RNAs that regulate gene expression in mammals. microRNA-7 (miR-7) is a well-demonstrated suppressor of hepatocellular carcinoma (HCC). Recent studies have showed that one such circRNA, ciRS-7 (also termed as Cdr1as) was the inhibitor and sponge of miR-7 in the embryonic zebrafish midbrain and islet cells. However, the relationships among ciRS-7, miR-7 and clinical features of HCC remain to be clarified.
   Methods Expression levels of ciRS-7, miR-7 and three miR-7-targeted mRNAs in 108 pairs of HCC and their matched non-tumor tissues were examined by quantitative reverse transcription polymerase chain reaction (qRT-PCR). The protein production of these three miR-7-targeted mRNAs was further verified by Western blot. The relationship between ciRS-7 level and clinicopathological features as well as the recurrence of HCC patients was analyzed. The univariate and multivariate logistic regression analyses were used to detect the risk factors of hepatic microvascular invasion (MVI). The correlation among ciRS-7, miR-7 and miR-7-targeted mRNAs was evaluated using Spearman's correlation test.
   Results There was no significant difference of ciRS-7 expression levels between the HCC tissues and the matched non-tumor tissues (0.67 +/- 1.49 vs. 0.44 +/- 0.45, p = 0.13), and the ciRS-7 levels in more than half of HCC tissues (65 out of 108, 60.2 %) were down-regulated when compared with their matched non-tumor tissues. However, the expression of ciRS-7 was significantly correlated with the following three clinicopathological characteristics of HCC patients: age <40 years (p = 0.02), serum AFP >= 400 ng/mu l (p < 0.01) and hepatic MVI (p = 0.03). Meanwhile, up-regulated ciRS-7 expression was not only an independent risk factor of hepatic MVI but also had a capable predictive ability for MVI (AUC = 0.68, p = 0.001) at the cut-off value of 0.135. Furthermore, the expression of ciRS-7 in HCC tissues with concurrent MVI was inversely correlated with that of miR-7 (r = -0.39, p = 0.007) and positively related with that of two miR-7-targeted genes [PIK3CD (r = 0.55, p < 0.001) and p70S6K (r = 0.34, p = 0.021)]. In addition, the median recurrent time of patients from higher ciRS-7 level group was shorter than that of lower ciRS-7 group (18 vs. 25 months), but no significant difference was observed (p = 0.38).
   Conclusions The expression levels of ciRS-7 were comparable between HCC and matched non-tumor tissues. However, the highly ciRS-7 expression in HCC tissues was significantly correlated with hepatic MVI, AFP level and younger age and thus partly related with the deterioration of HCC. Especially, ciRS-7 was one of the independent factors of hepatic MVI. These data suggested that ciRS-7 may be a promising biomarker of hepatic MVI and a novel therapy target for restraining MVI.
C1 [Xu, Liangliang; Zhang, Ming; Zheng, Xiaobo; Yi, Pengsheng; Xu, Mingqing] Sichuan Univ, West China Hosp, Dept Liver Surg, Chengdu 610041, Sichuan Provinc, Peoples R China.
   [Lan, Chuan] North Sichuan Med Coll, Inst Hepatobiliary Pancreat & Intestinal Dis, Nanchong 637000, Sichuan, Peoples R China.
   [Lan, Chuan] North Sichuan Med Coll, Affiliated Hosp, Dept Hepatobiliary Surg, Nanchong 637000, Sichuan, Peoples R China.
C3 Sichuan University; North Sichuan Medical University; North Sichuan
   Medical University
RP Xu, MQ (通讯作者)，Sichuan Univ, West China Hosp, Dept Liver Surg, Chengdu 610041, Sichuan Provinc, Peoples R China.
EM
RI Xu, Ming/IWM-0504-2023; Lan, Chuan/GYQ-6675-2022
FU National Natural Science Foundation of China [71673193]; Key Technology
   Research and Development Program of the Sichuan Province [2015SZ0131]
FX This study was funded by the National Natural Science Foundation of
   China (No. 71673193), and Key Technology Research and Development
   Program of the Sichuan Province (2015SZ0131).
CR DIBISCEGLIE AM, 1989, CANCER, V64, P2117, DOI 10.1002/1097-0142(19891115)64:10<2117::AID-CNCR2820641024>3.0.CO;2-7
   Fang YX, 2012, HEPATOLOGY, V55, P1852, DOI 10.1002/hep.25576
   Fleige Simone, 2006, Molecular Aspects of Medicine, V27, P126, DOI 10.1016/j.mam.2005.12.003
   Golfieri R, 2013, J HEPATOL, V59, P753, DOI 10.1016/j.jhep.2013.05.025
   Hansen TB, 2013, NATURE, V495, P384, DOI 10.1038/nature11993
   Hansen TB, 2011, EMBO J, V30, P4414, DOI 10.1038/emboj.2011.359
   HSU MT, 1979, NATURE, V280, P339, DOI 10.1038/280339a0
   Imamura H, 2003, J HEPATOL, V38, P200, DOI 10.1016/S0168-8278(02)00360-4
   Jemal A., 2011, CA-CANCER J CLIN, V61, P69, DOI [10.3322/caac.20107, DOI 10.3322/caac.21208]
   Jiang L, 2010, BIOCHEM J, V432, P199, DOI 10.1042/BJ20100859
   Kefas B, 2008, CANCER RES, V68, P3566, DOI 10.1158/0008-5472.CAN-07-6639
   Kong D, 2012, ONCOGENE, V31, P3949, DOI 10.1038/onc.2011.558
   Lagos-Quintana M, 2001, SCIENCE, V294, P853, DOI 10.1126/science.1064921
   Lee Ji Soo, 2015, Korean J Hepatobiliary Pancreat Surg, V19, P154, DOI 10.14701/kjhbps.2015.19.4.154
   Lei ZQ, 2016, JAMA SURG, V151, P356, DOI 10.1001/jamasurg.2015.4257
   Li F, 2015, ONCOTARGET, V6, P6001, DOI 10.18632/oncotarget.3469
   Li JR, 2014, ONCOL REP, V32, P2511, DOI 10.3892/or.2014.3519
   Lim KC, 2011, ANN SURG, V254, P108, DOI 10.1097/SLA.0b013e31821ad884
   Liu Q, 2016, SCI REP-UK, V6, DOI 10.1038/srep22572
   Liu S, 2013, FEBS LETT, V587, P2247, DOI 10.1016/j.febslet.2013.05.054
   Llovet JM, 2016, NAT REV DIS PRIMERS, V2, DOI [10.1038/nrdp.2016.19, 10.1038/nrdp.2016.18]
   Ma CQ, 2013, IUBMB LIFE, V65, P1026, DOI 10.1002/iub.1231
   Matter MS, 2014, J HEPATOL, V60, P855, DOI 10.1016/j.jhep.2013.11.031
   Memczak S, 2013, NATURE, V495, P333, DOI 10.1038/nature11928
   OKA H, 1994, HEPATOLOGY, V19, P61, DOI 10.1016/0270-9139(94)90053-1
   Poon RTP, 2009, ANN SURG ONCOL, V16, P792, DOI 10.1245/s10434-009-0330-y
   Quaglia A, 2005, ARCH PATHOL LAB MED, V129, P639
   Roayaie S, 2009, GASTROENTEROLOGY, V137, P850, DOI 10.1053/j.gastro.2009.06.003
   Rodriguez-Peralvarez M, 2013, ANN SURG ONCOL, V20, P325, DOI 10.1245/s10434-012-2513-1
   Salzman J, 2012, PLOS ONE, V7, DOI 10.1371/journal.pone.0030733
   Saydam O, 2011, CANCER RES, V71, P852, DOI 10.1158/0008-5472.CAN-10-1219
   Thuluvath PJ, 2009, J CLIN GASTROENTEROL, V43, P101, DOI 10.1097/MCG.0b013e318191e64f
   Villanueva A, 2008, GASTROENTEROLOGY, V135, P1972, DOI 10.1053/j.gastro.2008.08.008
   Vora SR, 2009, ONCOLOGIST, V14, P717, DOI 10.1634/theoncologist.2009-0038
   Wang Z, 2014, BIOMED RES INT, V2014, DOI 10.1155/2014/735672
   Xu HY, 2015, SCI REP-UK, V5, DOI 10.1038/srep12453
   Yamazaki Y, 2007, DIGEST DIS SCI, V52, P1103, DOI 10.1007/s10620-006-9578-2
   Yau T, 2011, ONCOLOGIST, V16, P1270, DOI 10.1634/theoncologist.2011-0105
   Yu L, 2016, PLOS ONE, V11, DOI 10.1371/journal.pone.0158347
   Zhang HY, 2014, STEM CELLS, V32, P2858, DOI 10.1002/stem.1795
   Zhang X, 2014, BIOCHEM BIOPH RES CO, V443, P1078, DOI 10.1016/j.bbrc.2013.12.095
   Zhou LD, 2010, MED ONCOL, V27, P255, DOI 10.1007/s12032-009-9201-4
   Zimmerman MA, 2008, ARCH SURG-CHICAGO, V143, P182, DOI 10.1001/archsurg.2007.39
NR 43
TC 248
Z9 271
U1 3
U2 33
PU SPRINGER
PI NEW YORK
PA ONE NEW YORK PLAZA, SUITE 4600, NEW YORK, NY, UNITED STATES
SN 0171-5216
EI 1432-1335
J9 J CANCER RES CLIN
JI J. Cancer Res. Clin. Oncol.
PD JAN
PY 2017
VL 143
IS 1
BP 17
EP 27
DI 10.1007/s00432-016-2256-7
PG 11
WC Oncology
WE Science Citation Index Expanded (SCI-EXPANDED)
SC Oncology
GA EI6TL
UT WOS:000392629700002
PM 27614453
HC Y
HP N
DA 2023-08-27
ER

PT J
AU Woiciechowsky, C
   Schoning, B
   Lanksch, WR
   Volk, HD
   Docke, WD
AF Woiciechowsky, C
   Schoning, B
   Lanksch, WR
   Volk, HD
   Docke, WD
TI Mechanisms of brain-mediated systemic anti-inflammatory syndrome causing
   immunodepression
SO JOURNAL OF MOLECULAR MEDICINE-JMM
LA English
DT Review
DE immunodepression; brain injury; cytokines; interleukin-10;
   glucocorticoids
ID TUMOR-NECROSIS-FACTOR; PITUITARY-ADRENAL AXIS; CORTICOTROPIN-RELEASING
   FACTOR; INTERLEUKIN-1 RECEPTOR ANTAGONIST; PRIMARY INTRACRANIAL TUMORS;
   SYMPATHETIC NERVOUS-SYSTEM; STRESS-INDUCED IMMUNOSUPPRESSION;
   GROWTH-FACTOR-BETA; HLA-DR EXPRESSION; FACTOR-ALPHA
AB Overwhelming inflammatory immune response can result in systemic inflammation and septic shock. To prevent excessive and deleterious action of proinflammatory cytokines after they have produced their initial beneficial effects, the immune system can release several anti-inflammatory mediators, including interleukin-10, interleukin-1 receptor antagonist, and soluble tumor necrosis factor receptors, thus initiating a compensatory anti-inflammatory response syndrome. However, in vivo the delicate balance between pro- and antiinflammatory responses is additionally controlled by the central nervous system. Therefore, proinflammatory cytokines stimulate the hypothalamic-pituitary-adrenal axis and enhance sympathetic nerve system activity. The mediators of these neuroimmune pathways can again suppress immune cell functions to control systemic inflammation. The question is, however, what happens if the immunoinhibitory CNS pathways are activated without systemic inflammation? This can result from production of cytokines in the brain following infection, injury, or ischemia or in response to various stressors (e.g., life events, depression, anxiety) or directly from brainstem irritation. The answer is that this may generate a brain-mediated immunodepression. Many animal and clinical studies have demonstrated a stress and brain cytokine mediated decrease in the cellular immune response at the lymphocyte level. More recently, the importance of monocytes in systemic immunocapacity has been shown. Monocytic inactivation with decreased capability of antigen presentation and depressed secretion of proinflammatory cytokines increases the risk of infectious complications. Interestingly, cytokines in the brain and other stressors can also generate systemic immunodepression at the monocyte level. In this scenario the catecholamine-induced release of the potent anti-inflammatory cytokine interleukin-10 is a newly discovered mechanism of the brain-mediated monocyte deactivation in addition to the "well known" immunosuppressive action of glucocorticoids. Furthermore, other neuropeptides such as a-melanocyte-stimulating hormone and P-endorphin which can be released in stressful situations have also inhibitory effects on immune cells. Thus mediators of the CNS are implicated in the regulation of immune functions and may play a role in both conditioning the host's response to endogenous or exogenous stimuli and generating a "brain-mediated" immunodepression.
C1 Humboldt Univ, Dept Neurosurg, D-13353 Berlin, Germany.
   Humboldt Univ, Inst Med Immunol, D-10098 Berlin, Germany.
C3 Humboldt University of Berlin; Humboldt University of Berlin
RP Woiciechowsky, C (通讯作者)，Humboldt Univ, Dept Neurosurg, Augustenburger Pl 1,Campus Virchow Klinikum, D-13353 Berlin, Germany.
RI volk, hans-dieter/AAV-8053-2021
OI volk, hans-dieter/0000-0002-7743-6668
CR Aarli Johan A., 1995, Tidsskrift for den Norske Laegeforening, V115, P954
   Agarwal SK, 1998, J INTERF CYTOK RES, V18, P1059, DOI 10.1089/jir.1998.18.1059
   Arend WP, 1998, ANNU REV IMMUNOL, V16, P27, DOI 10.1146/annurev.immunol.16.1.27
   ASADULLAH K, 1995, CRIT CARE MED, V23, P1976, DOI 10.1097/00003246-199512000-00006
   ASADULLAH K, 1996, EUR J EMERG MED, V2, P184
   Banchereau J, 1998, NATURE, V392, P245, DOI 10.1038/32588
   BATUMAN OA, 1995, J IMMUNOL, V155, P4397
   BAYBUTT HN, 1990, ENDOCRINOLOGY, V127, P476, DOI 10.1210/endo-127-1-476
   BEAMER NB, 1995, ANN NEUROL, V37, P800, DOI 10.1002/ana.410370614
   Bell MJ, 1997, J NEUROTRAUM, V14, P451, DOI 10.1089/neu.1997.14.451
   Bemelmans MHA, 1996, CRIT REV IMMUNOL, V16, P1
   BERKENBOSCH F, 1987, SCIENCE, V238, P524, DOI 10.1126/science.2443979
   BERTINI R, 1988, J EXP MED, V167, P1708, DOI 10.1084/jem.167.5.1708
   BESEDOVSKY H, 1986, SCIENCE, V233, P652, DOI 10.1126/science.3014662
   Bhardwaj RS, 1996, J IMMUNOL, V156, P2517
   BLANDINI F, 1995, LIFE SCI, V56, P1113, DOI 10.1016/0024-3205(95)00048-B
   BOGDAN C, 1992, J BIOL CHEM, V267, P23301
   BOHM H, 1991, Z KARDIOL S80, V8, P85
   Bone RC, 1996, CRIT CARE MED, V24, P1125, DOI 10.1097/00003246-199607000-00010
   BROOKS WH, 1972, J EXP MED, V136, P1631, DOI 10.1084/jem.136.6.1631
   BUCKINGHAM JC, 1994, PHARMACOL RES, V30, P35, DOI 10.1016/1043-6618(94)80085-5
   CELADA A, 1994, IMMUNOL TODAY, V15, P100, DOI 10.1016/0167-5699(94)90150-3
   CHELMICKASCHORR E, 1992, ANN NY ACAD SCI, V650, P40
   Costas M, 1996, J CLIN INVEST, V98, P1409, DOI 10.1172/JCI118928
   CROISET G, 1990, LIFE SCI, V46, P419, DOI 10.1016/0024-3205(90)90085-6
   CROSS RJ, 1984, IMMUNOLOGY, V51, P399
   CUPPS TR, 1982, IMMUNOL REV, V65, P133, DOI 10.1111/j.1600-065X.1982.tb00431.x
   de Waal MR, 1991, J EXP MED, V174, P1209, DOI DOI 10.1084/JEM.174.5.1209
   DEUEL TF, 1991, ANNU REV MED, V42, P567, DOI 10.1146/annurev.me.42.020191.003031
   Docke WD, 1997, NAT MED, V3, P678, DOI 10.1038/nm0697-678
   DOCKE WD, 1996, BASIS MECH INFLAMMAT, P162
   Donnelly SC, 1997, MOL MED TODAY, V3, P502, DOI 10.1016/S1357-4310(97)01133-7
   DUNN AJ, 1992, BRAIN RES BULL, V29, P807, DOI 10.1016/0361-9230(92)90148-Q
   DUNN AJ, 1988, LIFE SCI, V43, P429, DOI 10.1016/0024-3205(88)90522-X
   ELLIOTT LH, 1984, J IMMUNOL, V132, P1208
   ELLIOTT LH, 1987, J NEUROSURG, V67, P231, DOI 10.3171/jns.1987.67.2.0231
   FASSBENDER K, 1994, STROKE, V25, P1105, DOI 10.1161/01.STR.25.6.1105
   FASSBENDER K, 1994, J NEUROL SCI, V122, P135, DOI 10.1016/0022-510X(94)90289-5
   FEUERSTEIN GZ, 1994, CEREBROVAS BRAIN MET, V6, P341
   FIORENTINO DF, 1991, J IMMUNOL, V146, P3444
   FISHER SA, 1995, AM J PHYSIOL-CELL PH, V268, pC910
   Friedman EM, 1995, ANN NY ACAD SCI, V771, P396, DOI 10.1111/j.1749-6632.1995.tb44698.x
   Ghirnikar RS, 1998, NEUROCHEM RES, V23, P329, DOI 10.1023/A:1022453332560
   Grabbe S, 1996, J IMMUNOL, V156, P473
   GUILLOU PJ, 1993, LANCET, V342, P217, DOI 10.1016/0140-6736(93)92303-B
   Hansen MK, 1998, J NEUROSCI, V18, P2247
   HASSAN NF, 1990, CLIN IMMUNOL IMMUNOP, V54, P220, DOI 10.1016/0090-1229(90)90083-3
   HOCH RC, 1993, CRIT CARE MED, V21, P839, DOI 10.1097/00003246-199306000-00010
   HOWARD M, 1992, IMMUNOL TODAY, V13, P198, DOI 10.1016/0167-5699(92)90153-X
   ICHIJO T, 1994, BRAIN RES BULL, V34, P547, DOI 10.1016/0361-9230(94)90139-2
   Joyce DA, 1997, INFLAMM RES, V46, P447, DOI 10.1007/s000110050222
   Kapcala LP, 1995, ANN NY ACAD SCI, V771, P419, DOI 10.1111/j.1749-6632.1995.tb44699.x
   KATAFUCHI T, 1993, J PHYSIOL-LONDON, V471, P209, DOI 10.1113/jphysiol.1993.sp019898
   KATSUURA G, 1988, ENDOCRINOLOGY, V122, P1773, DOI 10.1210/endo-122-5-1773
   KATSUURA G, 1990, AM J PHYSIOL, V258, pE163, DOI 10.1152/ajpendo.1990.258.1.E163
   KELLER SE, 1988, P NATL ACAD SCI USA, V85, P9297, DOI 10.1073/pnas.85.23.9297
   KELLER SE, 1983, SCIENCE, V221, P1301, DOI 10.1126/science.6612346
   KLIR JJ, 1994, AM J PHYSIOL, V266, pR1845, DOI 10.1152/ajpregu.1994.266.6.R1845
   KOSSMANN T, 1995, SHOCK, V4, P311, DOI 10.1097/00024382-199511000-00001
   KUPPNER MC, 1989, J NEUROSURG, V71, P211, DOI 10.3171/jns.1989.71.2.0211
   Kvetnansky R, 1995, ANN NY ACAD SCI, V771, P131, DOI 10.1111/j.1749-6632.1995.tb44676.x
   LAYE S, 1995, AM J PHYSIOL-REG I, V268, pR1327, DOI 10.1152/ajpregu.1995.268.5.R1327
   LENNARD TWJ, 1985, BRIT J SURG, V72, P771, DOI 10.1002/bjs.1800721002
   Linthorst ACE, 1995, EUR J NEUROSCI, V7, P2418, DOI 10.1111/j.1460-9568.1995.tb01040.x
   Linthorst ACE, 1998, ANN NY ACAD SCI, V840, P139, DOI 10.1111/j.1749-6632.1998.tb09558.x
   Lipton JM, 1997, IMMUNOL TODAY, V18, P140, DOI 10.1016/S0167-5699(97)01009-8
   LIVINGSTON DH, 1988, ARCH SURG-CHICAGO, V123, P1309
   LYSON K, 1991, NEUROENDOCRINOLOGY, V54, P262, DOI 10.1159/000125884
   MADDEN KS, 1995, ANNU REV PHARMACOL, V35, P417, DOI 10.1146/annurev.pa.35.040195.002221
   MAHALEY MS, 1977, J NEUROSURG, V46, P467, DOI 10.3171/jns.1977.46.4.0467
   MAIMONE D, 1993, J NEUROIMMUNOL, V47, P55, DOI 10.1016/0165-5728(93)90284-6
   Mastorakos G, 1995, ANN NY ACAD SCI, V771, P438, DOI 10.1111/j.1749-6632.1995.tb44700.x
   MATHIESEN T, 1993, J NEUROSURG, V78, P562, DOI 10.3171/jns.1993.78.4.0562
   Mathiesen T, 1997, J NEUROSURG, V87, P215, DOI 10.3171/jns.1997.87.2.0215
   Mazzarella G, 1998, Monaldi Arch Chest Dis, V53, P92
   Meisel C, 1996, EUR J IMMUNOL, V26, P1580, DOI 10.1002/eji.1830260726
   METCALF D, 1991, SCIENCE, V254, P529, DOI 10.1126/science.1948028
   NEEFJES JJ, 1992, IMMUNOL TODAY, V13, P179, DOI 10.1016/0167-5699(92)90123-O
   OTT L, 1994, J NEUROTRAUM, V11, P447, DOI 10.1089/neu.1994.11.447
   Ottaviani E, 1997, IMMUNOL TODAY, V18, P169, DOI 10.1016/S0167-5699(97)84663-4
   Panerai AE, 1997, IMMUNOL TODAY, V18, P317, DOI 10.1016/S0167-5699(97)01045-1
   PARROTT RF, 1994, RES VET SCI, V56, P234, DOI 10.1016/0034-5288(94)90109-0
   PAYNE LC, 1992, J IMMUNOTHER, V12, P171, DOI 10.1097/00002371-199210000-00005
   PLATZER C, 1995, INT IMMUNOL, V7, P517, DOI 10.1093/intimm/7.4.517
   Pretolani M, 1997, IMMUNOL TODAY, V18, P277, DOI 10.1016/S0167-5699(97)80023-0
   RABIN BS, 1989, CRIT REV IMMUNOL, V9, P279
   Ramirez F, 1996, J IMMUNOL, V156, P2406
   REICHLIN S, 1993, NEW ENGL J MED, V329, P1246
   RILEY V, 1981, SCIENCE, V212, P1100, DOI 10.1126/science.7233204
   RIVIER C, 1989, ENDOCRINOLOGY, V125, P3096, DOI 10.1210/endo-125-6-3096
   ROMANO TA, 1991, BRAIN BEHAV IMMUN, V5, P116, DOI 10.1016/0889-1591(91)90011-X
   ROOZEN AWN, 1995, AM J VET RES, V56, P1225
   ROSZMAN T, 1991, IMMUNOL TODAY, V12, P370, DOI 10.1016/0167-5699(91)90068-5
   ROSZMAN TL, 1987, J NEUROSURG, V67, P874, DOI 10.3171/jns.1987.67.6.0874
   Sadeghi R, 1992, Eur Cytokine Netw, V3, P373
   Saito K, 1996, NEUROSCI LETT, V206, P149, DOI 10.1016/S0304-3940(96)12460-5
   SAPERSTEIN A, 1992, ENDOCRINOLOGY, V130, P152, DOI 10.1210/en.130.1.152
   SAPOLSKY R, 1987, SCIENCE, V238, P522, DOI 10.1126/science.2821621
   SCHOBITZ B, 1994, CRIT REV NEUROBIOL, V8, P263
   SEVERN A, 1992, J IMMUNOL, V148, P3441
   Sherry RM, 1996, J TRAUMA, V40, P613, DOI 10.1097/00005373-199604000-00016
   SHIMIZU N, 1994, BRAIN BEHAV IMMUN, V8, P14, DOI 10.1006/brbi.1994.1002
   SIGURDSSON A, 1993, AM HEART J, V126, P1068, DOI 10.1016/0002-8703(93)90656-T
   Smith E., 1992, NEUROIMMUNOENDOCRINO, P154
   SUNDAR SK, 1989, P NATL ACAD SCI USA, V86, P6398, DOI 10.1073/pnas.86.16.6398
   SUNDAR SK, 1990, J NEUROSCI, V10, P3701
   SWEEP F, 1991, ACTA ENDOCRINOL-COP, V125, P84
   Tabardel Y, 1996, SURGERY, V119, P76, DOI 10.1016/S0039-6060(96)80217-0
   TADA M, 1993, J NEURO-ONCOL, V17, P261, DOI 10.1007/BF01049981
   TAUPIN V, 1993, J NEUROIMMUNOL, V42, P177, DOI 10.1016/0165-5728(93)90008-M
   TERAO A, 1994, AM J PHYSIOL, V266, pR400, DOI 10.1152/ajpregu.1994.266.2.R400
   Trinchieri G, 1998, Int Rev Immunol, V16, P365, DOI 10.3109/08830189809043002
   Trinchieri G, 1998, IMMUNOL RES, V17, P269, DOI 10.1007/BF02786451
   TURNBULL AV, 1994, PSYCHONEUROENDOCRINO, V19, P165, DOI 10.1016/0306-4530(94)90006-X
   Unanue Emil R., 1993, P111
   VANDENBRINK HR, 1994, J RHEUMATOL, V21, P430
   VANDERPOLL T, 1994, INFECT IMMUN, V62, P2046, DOI 10.1128/IAI.62.5.2046-2050.1994
   vanderPoll T, 1996, J CLIN INVEST, V97, P713, DOI 10.1172/JCI118469
   vanFurth AM, 1996, INFECT IMMUN, V64, P4883, DOI 10.1128/IAI.64.12.4883-4890.1996
   Volk HD, 1993, HOST DEFENSE DYSFUNC, P365
   WAHL SM, 1993, J IMMUNOL, V150, P3553
   WAKEFIELD CH, 1993, BRIT J SURG, V80, P205, DOI 10.1002/bjs.1800800224
   WATKINS LR, 1995, NEUROSCI LETT, V183, P27, DOI 10.1016/0304-3940(94)11105-R
   WATKINS LR, 1995, LIFE SCI, V57, P1011, DOI 10.1016/0024-3205(95)02047-M
   WEISS JM, 1991, EUR J PHARMACOL, V192, P177, DOI 10.1016/0014-2999(91)90087-7
   WEISS JM, 1994, ANN NY ACAD SCI, V741, P338, DOI 10.1111/j.1749-6632.1994.tb39676.x
   Wilckens T, 1997, IMMUNOL TODAY, V18, P418, DOI 10.1016/S0167-5699(97)01111-0
   Woiciechowsky C, 1998, NAT MED, V4, P808, DOI 10.1038/nm0798-808
   Woiciechowsky C, 1999, BRAIN RES, V816, P563, DOI 10.1016/S0006-8993(98)01238-4
   Woiciechowsky C, 1997, ACTA NEUROCHIR, V139, P619, DOI 10.1007/BF01411996
   Woiciechowsky C, 1998, EUR J NEUROSCI, V10, P168
   Woiciechowsky C, 1998, J NEUROIMMUNOL, V84, P164, DOI 10.1016/S0165-5728(97)00236-1
   WOICIECHOWSKY C, 1996, IMMUNOBIOLOGY, V196, P86
   WOICIECHOWSKY C, 1998, ZENTRALBL NEUROCHIR, V59, P52
   WOODROOFE MN, 1991, J NEUROIMMUNOL, V33, P227, DOI 10.1016/0165-5728(91)90110-S
   Zhai QH, 1997, J NEUROL SCI, V152, P119, DOI 10.1016/S0022-510X(97)00154-8
   Zhou DH, 1996, NEUROENDOCRINOLOGY, V63, P227, DOI 10.1159/000126962
NR 137
TC 135
Z9 140
U1 0
U2 7
PU SPRINGER VERLAG
PI NEW YORK
PA 175 FIFTH AVE, NEW YORK, NY 10010 USA
SN 0946-2716
J9 J MOL MED-JMM
JI J. Mol. Med.
PD NOV
PY 1999
VL 77
IS 11
BP 769
EP 780
DI 10.1007/s001099900051
PG 12
WC Genetics & Heredity; Medicine, Research & Experimental
WE Science Citation Index Expanded (SCI-EXPANDED)
SC Genetics & Heredity; Research & Experimental Medicine
GA 264MK
UT WOS:000084185900004
PM 10619437
DA 2023-08-27
ER

PT J
AU Papageorgiou, E
   Wermund, T
   Wilhelm, H
AF Papageorgiou, Eleni
   Wermund, Thomas
   Wilhelm, Helmut
TI Pupil Perimetry Demonstrates Hemifield Pupillary Hypokinesia in a
   Patient With a Pretectal Lesion Causing a Relative Afferent Pupil Defect
   but No Visual Field Loss
SO JOURNAL OF NEURO-OPHTHALMOLOGY
LA English
DT Article
ID NORMAL VISION; PATHWAY; PARESIS; TUMOR
AB Background: Lesions affecting the pretectum or the brachium of the superior colliculus (brachium) and sparing the optic tract cause a contralateral relative afferent pupil defect (RAPD) but no visual field loss, It has been assumed that the pupillomotor pathways within the brachium are a continuation of the pupillomotor pathways traveling in the optic tract. To investigate this assumption, we looked for hemihypokinesia by means of pupil perimetry.
   Methods: Pupillary hemifield stimulation was performed in a 65-year-old woman with normal visual fields and an isolated left RAPID due to a cerebral hemorrhage affecting the right dorsal midbrain. The pupil responses from light stimulation of the nasal inferior, nasal superior, and temporal inferior and temporal superior quadrants or both eyes were recorded using computerized binocular infrared pupillography. Each stimulus was presented 5 times and the mean amplitude of the pupil response was calculated for each stimulus location.
   Results: Pupil perimetry demonstrated a marked hemihypokinesia (reduced light reaction) in the hemifield contralateral to the site of the lesion.
   Conclusions: Our experiment suggests that the brachium is indeed a continuation of the afferent pupillary fibers traveling in the optic tract.
C1 [Papageorgiou, Eleni; Wilhelm, Helmut] Univ Tubingen, Ctr Ophthalmol, D-72076 Tubingen, Germany.
   [Wermund, Thomas] Helios Kliniken Schwerin, Dept Ophthalmol, Schwerin, Germany.
C3 CIVIS; Eberhard Karls University of Tubingen; Eberhard Karls University
   Hospital; Helios Kliniken
RP Papageorgiou, E (通讯作者)，Univ Tubingen, Ctr Ophthalmol, Schleichstr 12-16, D-72076 Tubingen, Germany.
EM
OI Papageorgiou, Eleni/0000-0003-1931-4168
FU European Union (PFRACT Marie Curie Early Stage Training)
   [MEST-CT-2004-504321]
FX This work was supported by the European Union (PFRACT Marie Curie Early
   Stage Training MEST-CT-2004-504321).
CR BEHR C, 1926, Z PRAKT AUGENHEILKD, V58, P398
   Chen CJ, 2004, ARCH NEUROL-CHICAGO, V61, P1451, DOI 10.1001/archneur.61.9.1451
   ELIOTT D, 1991, J CLIN NEURO-OPHTHAL, V11, P169
   ELLIS CJK, 1984, J NEUROL NEUROSUR PS, V47, P739, DOI 10.1136/jnnp.47.7.739
   FORMAN S, 1990, ARCH OPHTHALMOL-CHIC, V108, P1074, DOI 10.1001/archopht.1990.01070100030017
   Girkin CA, 1998, ARCH OPHTHALMOL-CHIC, V116, P1544, DOI 10.1001/archopht.116.11.1544
   JOHNSON RE, 1987, CAN J OPHTHALMOL, V22, P282
   Kardon R, 2006, OPHTHALMOLOGY, V113, P1345, DOI 10.1016/j.ophtha.2006.02.055
   KING JT, 1991, NEUROLOGY, V41, P945
   LOEWENFELD IE, 1999, PUPIL ANATOMY PHYSL, V1, P203
   Papageorgiou E, 2008, NEUROLOGY, V70, P956, DOI 10.1212/01.wnl.0000305962.93520.ed
   Staubach F, 2007, GRAEF ARCH CLIN EXP, V245, P321, DOI 10.1007/s00417-005-0235-9
   Taguchi H, 2000, GRAEF ARCH CLIN EXP, V238, P927, DOI 10.1007/s004170000206
   Wilhelm BJ, 2002, BRAIN, V125, P2296, DOI 10.1093/brain/awf232
   Wilhelm H, 1996, NEURO-OPHTHALMOLOGY, V16, P219, DOI 10.3109/01658109609044629
NR 15
TC 10
Z9 10
U1 0
U2 0
PU LIPPINCOTT WILLIAMS & WILKINS
PI PHILADELPHIA
PA TWO COMMERCE SQ, 2001 MARKET ST, PHILADELPHIA, PA 19103 USA
SN 1070-8022
EI 1536-5166
J9 J NEURO-OPHTHALMOL
JI J. Neuro-Ophthal.
PD MAR
PY 2009
VL 29
IS 1
BP 33
EP 36
DI 10.1097/WNO.0b013e318198cb8c
PG 4
WC Clinical Neurology; Ophthalmology
WE Science Citation Index Expanded (SCI-EXPANDED)
SC Neurosciences & Neurology; Ophthalmology
GA 418CZ
UT WOS:000264126100009
PM 19458574
OA Bronze
DA 2023-08-27
ER

PT J
AU Takamiya, R
   Missett, B
   Weinberg, V
   Akazawa, C
   Akazawa, P
   Zytkovicz, A
   Bucci, MK
   Lee, N
   Quivey, J
   Xia, P
AF Takamiya, Robert
   Missett, Brian
   Weinberg, Vivian
   Akazawa, Clayton
   Akazawa, Pam
   Zytkovicz, Andrea
   Bucci, Mary Kara
   Lee, Nancy
   Quivey, Jeanne
   Xia, Ping
TI Simplifying intensity-modulated radiotherapy plans with fewer beam
   angles for the treatment of oropharyngeal carcinoma
SO JOURNAL OF APPLIED CLINICAL MEDICAL PHYSICS
LA English
DT Article
DE oropharyngeal carcinoma; IMRT; beam angles
ID NECK-CANCER; RADIATION-THERAPY; SALIVARY FUNCTION; HEAD; IMRT; IMPACT;
   OPTIMIZATION; IRRADIATION; QUALITY; NUMBER
AB The first aim of the present study was to investigate the feasibility of using fewer beam angles to improve delivery efficiency for the treatment of oropharyngeal cancer (OPC) with inverse-planned intensity-modulated radiation therapy (IP-IMRT). A secondary aim was to evaluate whether the simplified IP-IMRT plans could reduce the indirect radiation dose. The treatment plans for 5 consecutive OPC patients previously treated with a forward-planned IMRT (FP-IMRT) technique were selected as benchmarks for this study. The initial treatment goal for these patients was to deliver 70 Gy to >= 95% of the planning gross tumor volume (PTV-70) and 59.4 Gy to >= 95% of the planning clinical tumor volume (PTV-59.4) simultaneously. Each case was re-planned using IP-IMRT with multiple beam-angle arrangements, including four complex IP-IMRT plans using 7 or more beam angles, and one simple IMRT plan using 5 beam angles. The complex IP-IMRT plans and simple IP-IMRT plans were compared to each other and to the FP-IMRT plans by analyzing the dose coverage of the target volumes, the plan homogeneity, the dose-volume histograms of critical structures, and the treatment delivery parameters including delivery time and the total number of monitor units (MUs). When comparing the plans, we found no significant difference between the complex IP-IMRT, simple IP-IMRT, and FP-IMRT plans for tumor target coverage (PTV-70: p = 0.56; PTV-59.4: p = 0.20). The plan homogeneity, measured by the mean percentage isodose, did not significantly differ between the IP-IMRT and FP-IMRT plans (p = 0.08), although we observed a trend toward greater inhomogeneity of dose in the simple IP-IMRT plans. All IP-IMRT plans either met or exceeded the quality of the FP-IMRT plans in terms of dose to adjacent critical structures, including the parotids, spinal cord, and brainstem. As compared with the complex IP-IMRT plans, the simple IP-IMRT plans significantly reduced the mean treatment time (maximum probability for four pairwise comparisons: p = 0.0003). In conclusion, our study demonstrates that, as compared with complex IP-IMRT, simple IP-IMRT can significantly improve treatment delivery efficiency while maintaining similar target coverage and sparing of critical structures. However, the improved efficiency does not significantly reduce the total number of MUs nor the indirect radiation dose.
C1 Univ Calif San Francisco, Dept Radiat Oncol, San Francisco, CA 94143 USA.
   Univ Calif San Francisco, Ctr Comprehens Canc, San Francisco, CA 94143 USA.
C3 University of California System; University of California San Francisco;
   University of California System; University of California San Francisco;
   UCSF Medical Center; UCSF Helen Diller Family Comprehensive Cancer
   Center
RP Xia, P (通讯作者)，1600 Divisadero,Suite H-1031,Box 1708, San Francisco, CA 94143 USA.
EM
CR Chao KSC, 2004, INT J RADIAT ONCOL, V59, P43, DOI 10.1016/j.ijrobp.2003.08.004
   Chao KSC, 2002, SEMIN RADIAT ONCOL, V12, P20, DOI 10.1053/srao.2002.31359
   Chao KSC, 2001, INT J RADIAT ONCOL, V49, P907, DOI 10.1016/S0360-3016(00)01441-3
   Cozzi L, 2004, INT J RADIAT ONCOL, V58, P617, DOI 10.1016/j.ijrobp.2003.09.059
   Eisbruch A, 2001, INT J RADIAT ONCOL, V50, P695, DOI 10.1016/S0360-3016(01)01512-7
   Eisbruch A, 1999, INT J RADIAT ONCOL, V45, P577, DOI 10.1016/S0360-3016(99)00247-3
   Eisbruch A, 2001, 0022 RTOG
   Hall EJ, 2003, INT J RADIAT ONCOL, V56, P83, DOI 10.1016/S0360-3016(03)00073-7
   Hunt MA, 2001, INT J RADIAT ONCOL, V49, P623, DOI 10.1016/S0360-3016(00)01389-4
   Lee N, 2004, INT J RADIAT ONCOL, V59, P584, DOI 10.1016/j.ijrobp.2004.02.005
   Lin A, 2003, INT J RADIAT ONCOL, V57, P61, DOI 10.1016/S0360-3016(03)00361-4
   Mohan R, 2000, MED PHYS, V27, P1226, DOI 10.1118/1.599000
   Pirzkall A, 2000, INT J RADIAT ONCOL, V48, P1371, DOI 10.1016/S0360-3016(00)00772-0
   Pugachev A, 2002, INT J RADIAT ONCOL, V54, P1565, DOI 10.1016/S0360-3016(02)03917-2
   Shepard DM, 2002, MED PHYS, V29, P1007, DOI 10.1118/1.1477415
   Stein J, 1997, MED PHYS, V24, P149, DOI 10.1118/1.597923
   Xia P, 2000, INT J RADIAT ONCOL, V48, P329, DOI 10.1016/S0360-3016(00)00585-X
NR 17
TC 6
Z9 6
U1 0
U2 0
PU MULTIMED INC
PI TORONTO
PA 66 MARTIN ST, TORONTO, ON L9T 2R2, CANADA
SN 1526-9914
J9 J APPL CLIN MED PHYS
JI J. Appl. Clin. Med. Phys
PY 2007
VL 8
IS 2
BP 26
EP 36
DI 10.1120/jacmp.v8i2.2412
PG 11
WC Radiology, Nuclear Medicine & Medical Imaging
WE Science Citation Index Expanded (SCI-EXPANDED)
SC Radiology, Nuclear Medicine & Medical Imaging
GA 183DP
UT WOS:000247553200005
PM 17592464
OA Green Published, Green Submitted, gold
DA 2023-08-27
ER

PT J
AU Dahl, NA
   Danis, E
   Balakrishnan, I
   Wang, D
   Pierce, A
   Walker, FM
   Gilani, A
   Serkova, NJ
   Madhavan, K
   Fosmire, S
   Green, AL
   Foreman, NK
   Venkataraman, S
   Vibhakar, R
AF Dahl, Nathan A.
   Danis, Etienne
   Balakrishnan, Ilango
   Wang, Dong
   Pierce, Angela
   Walker, Faye M.
   Gilani, Ahmed
   Serkova, Natalie J.
   Madhavan, Krishna
   Fosmire, Susan
   Green, Adam L.
   Foreman, Nicholas K.
   Venkataraman, Sujatha
   Vibhakar, Rajeev
TI Super Elongation Complex as a Targetable Dependency in Diffuse Midline
   Glioma
SO CELL REPORTS
LA English
DT Article
ID INTRINSIC PONTINE GLIOMA; RNA-POLYMERASE-II; TRANSCRIPTION ELONGATION;
   TARGETING TRANSCRIPTION; STEM-CELL; EXPRESSION; INHIBITION; H3K27M; SEC;
   DISRUPTION
AB Histone 3 gene mutations are the eponymous drivers in diffuse midline gliomas (DMGs), aggressive pediatric brain cancers for which no curative therapy currently exists. These recurrent oncohistones induce a global loss of repressive H3K27me3 residues and broad epigenetic dysregulation. In order to identify therapeutically targetable dependencies within this disease context, we performed an RNAi screen targeting epigenetic/chromatin-associated genes in patient-derived DMG cultures. This identified AFF4, the scaffold protein of the super elongation complex (SEC), as a molecular dependency in DMG. Interrogation of SEC function demonstrates a key role for maintaining clonogenic potential while promoting self-renewal of tumor stem cells. Small-molecule inhibition of SEC using clinically relevant CDK9 inhibitors restores regulatory RNA polymerase II pausing, promotes cellular differentiation, and leads to potent anti-tumor effect both in vitro and in patient-derived xenograft models. These studies present a rationale for further exploration of SEC inhibition as a promising therapeutic approach to this intractable disease.
C1 [Dahl, Nathan A.; Danis, Etienne; Balakrishnan, Ilango; Wang, Dong; Pierce, Angela; Walker, Faye M.; Madhavan, Krishna; Fosmire, Susan; Green, Adam L.; Foreman, Nicholas K.; Venkataraman, Sujatha; Vibhakar, Rajeev] Morgan Adams Fdn, Pediat Brain Tumor Res Program, Aurora, CO USA.
   [Dahl, Nathan A.; Danis, Etienne; Balakrishnan, Ilango; Wang, Dong; Pierce, Angela; Walker, Faye M.; Madhavan, Krishna; Fosmire, Susan; Green, Adam L.; Foreman, Nicholas K.; Venkataraman, Sujatha; Vibhakar, Rajeev] Univ Colorado, Dept Pediat, Sch Med, Aurora, CO 80045 USA.
   [Dahl, Nathan A.; Green, Adam L.; Foreman, Nicholas K.; Vibhakar, Rajeev] Childrens Hosp Colorado, Ctr Canc & Blood Disorders, Aurora, CO 80045 USA.
   [Gilani, Ahmed] Univ Colorado, Dept Pathol, Sch Med, Aurora, CO USA.
   [Serkova, Natalie J.] Univ Colorado, Dept Radiol, Sch Med, Aurora, CO USA.
   [Foreman, Nicholas K.; Vibhakar, Rajeev] Univ Colorado, Dept Neurosurg, Sch Med, Aurora, CO 80045 USA.
C3 University of Colorado System; University of Colorado Anschutz Medical
   Campus; Children's Hospital Colorado; University of Colorado System;
   University of Colorado Anschutz Medical Campus; University of Colorado
   System; University of Colorado Anschutz Medical Campus; University of
   Colorado System; University of Colorado Anschutz Medical Campus
RP Dahl, NA; Vibhakar, R (通讯作者)，Morgan Adams Fdn, Pediat Brain Tumor Res Program, Aurora, CO USA.; Dahl, NA; Vibhakar, R (通讯作者)，Univ Colorado, Dept Pediat, Sch Med, Aurora, CO 80045 USA.; Dahl, NA; Vibhakar, R (通讯作者)，Childrens Hosp Colorado, Ctr Canc & Blood Disorders, Aurora, CO 80045 USA.; Vibhakar, R (通讯作者)，Univ Colorado, Dept Neurosurg, Sch Med, Aurora, CO 80045 USA.
EM
RI Gilani, Ahmed/GYJ-5201-2022
FU St. Baldrick's Foundation [606095]; Golfers Against Cancer; Cancer
   League of Colorado; Morgan Adams Foundation; Curing Kid's Cancer; Luke
   Morin Family; Cancer Center Support Grant [P30 CA046934]; NCI [P30
   CA046934]; NIH [S10 OD023485]
FX We would like to thank the following individuals for generously
   providing the cell lines used for this work: Michelle Monje (Stanford
   University) for SUDIPG4 and SU-DIPG13; Angel Montero Carcaboso (Sant
   Joan de Deu) for HSJD-DIPG007; and Nalin Gupta (University of
   California, San Francisco) for SF8268. We likewise thank the University
   of Colorado Functional Genomics Facility, Genomics and Microarray Shared
   Resource, Research Histology Shared Resource, and Animal Imaging Shared
   Resource for their contributions to this work. This work was generously
   supported by grants through the St. Baldrick's Foundation (Fellowship
   Award 606095; N.A.D. and R.V.), Golfers Against Cancer (N.A.D., A.L.G.,
   and S.V.), Cancer League of Colorado (N.A.D. and R.V.), the Morgan Adams
   Foundation (N.A.D., S.V., and R.V.), Curing Kid's Cancer (N.A.D. and
   R.V.), and the Luke Morin Family (N.A.D., S.V., and R.V.). The
   University of Colorado Research Histology Shared Resource is supported
   by a Cancer Center Support Grant (P30 CA046934). The University of
   Colorado Animal Imaging Shared Resource is supported by the NCI P30
   CA046934 and NIH S10 OD023485 grants (N.J.S.). Graphical abstract
   created in part using Servier Medical Art licensed under Creative
   Commons 3.0 Attribution.
CR Amani V, 2016, BMC CANCER, V16, DOI 10.1186/s12885-016-2690-6
   Anastas JN, 2019, CANCER CELL, V36, P528, DOI 10.1016/j.ccell.2019.09.005
   Bartels U, 2011, J NEURO-ONCOL, V105, P119, DOI 10.1007/s11060-011-0704-4
   Bender S, 2013, CANCER CELL, V24, P660, DOI 10.1016/j.ccr.2013.10.006
   Boettiger AN, 2009, SCIENCE, V325, P471, DOI 10.1126/science.1173976
   Bradner JE, 2017, CELL, V168, P629, DOI 10.1016/j.cell.2016.12.013
   Chen FX, 2018, NAT REV MOL CELL BIO, V19, P464, DOI 10.1038/s41580-018-0010-5
   Chen Y, 2019, J BIOL CHEM, V294, P10663, DOI 10.1074/jbc.RA119.008577
   Choi SA, 2014, EUR J CANCER, V50, P137, DOI 10.1016/j.ejca.2013.09.004
   Chopra VS, 2009, CURR BIOL, V19, P688, DOI 10.1016/j.cub.2009.02.055
   Core LJ, 2008, SCIENCE, V322, P1845, DOI 10.1126/science.1162228
   Dickson MA, 2011, INVEST NEW DRUG, V29, P1004, DOI 10.1007/s10637-010-9447-x
   Khuong-Quang DA, 2012, ACTA NEUROPATHOL, V124, P439, DOI 10.1007/s00401-012-0998-0
   Dunham I, 2012, NATURE, V489, P57, DOI 10.1038/nature11247
   Fabre C, 2014, CANCER CHEMOTH PHARM, V74, P1057, DOI 10.1007/s00280-014-2583-9
   Filbin MG, 2018, SCIENCE, V360, P331, DOI 10.1126/science.aao4750
   Filippakopoulos P, 2014, NAT REV DRUG DISCOV, V13, P339, DOI 10.1038/nrd4286
   Fowler T, 2014, PLOS ONE, V9, DOI 10.1371/journal.pone.0087003
   Franco HL, 2015, CELL, V163, P28, DOI 10.1016/j.cell.2015.09.013
   Grasso CS, 2015, NAT MED, V21, P555, DOI 10.1038/nm.3855
   Green AL, 2015, NEURO-ONCOLOGY, V17, P697, DOI 10.1093/neuonc/nou303
   Hagenbuchner J, 2016, BIOCHEM PHARMACOL, V107, P1, DOI 10.1016/j.bcp.2015.12.006
   Hashizume R, 2014, NAT MED, V20, P1394, DOI 10.1038/nm.3716
   Heath EI, 2008, INVEST NEW DRUG, V26, P59, DOI 10.1007/s10637-007-9090-3
   Hoffman LM, 2018, J CLIN ONCOL, V36, P1963, DOI 10.1200/JCO.2017.75.9308
   Hu YF, 2009, J IMMUNOL METHODS, V347, P70, DOI 10.1016/j.jim.2009.06.008
   Huang CH, 2014, GENE DEV, V28, P1800, DOI 10.1101/gad.244368.114
   Jansen MHA, 2012, CANCER TREAT REV, V38, P27, DOI 10.1016/j.ctrv.2011.06.007
   Jones C, 2014, NAT REV CANCER, V14, P651, DOI 10.1038/nrc3811
   Jonkers I, 2015, NAT REV MOL CELL BIO, V16, P167, DOI 10.1038/nrm3953
   Karp JE, 2012, HAEMATOL-HEMATOL J, V97, P1736, DOI 10.3324/haematol.2012.062539
   Krug B, 2019, CANCER CELL, V35, P782, DOI 10.1016/j.ccell.2019.04.004
   Lanasa MC, 2015, LEUKEMIA RES, V39, P495, DOI 10.1016/j.leukres.2015.02.001
   Langmead B, 2009, GENOME BIOL, V10, DOI 10.1186/gb-2009-10-3-r25
   Larson JD, 2019, CANCER CELL, V35, P140, DOI 10.1016/j.ccell.2018.11.015
   Levine M, 2011, CELL, V145, P502, DOI 10.1016/j.cell.2011.04.021
   Lewis PW, 2013, SCIENCE, V340, P857, DOI 10.1126/science.1232245
   Li H, 2009, BIOINFORMATICS, V25, P1754, DOI 10.1093/bioinformatics/btp324
   Liang KW, 2018, CELL, V175, P766, DOI 10.1016/j.cell.2018.09.027
   Lin CQ, 2011, GENE DEV, V25, P1486, DOI 10.1101/gad.2059211
   Lin CQ, 2010, MOL CELL, V37, P429, DOI 10.1016/j.molcel.2010.01.026
   Lin GL, 2019, SCI TRANSL MED, V11, DOI 10.1126/scitranslmed.aaw0064
   Louis DN, 2016, ACTA NEUROPATHOL, V131, P803, DOI 10.1007/s00401-016-1545-1
   Loven J, 2013, CELL, V153, P320, DOI 10.1016/j.cell.2013.03.036
   Luo ZJ, 2012, NAT REV MOL CELL BIO, V13, P543, DOI 10.1038/nrm3417
   Malumbres M, 2014, GENOME BIOL, V15, DOI 10.1186/gb4184
   Morales F, 2016, CELL CYCLE, V15, P519, DOI 10.1080/15384101.2016.1138186
   Muse GW, 2007, NAT GENET, V39, P1507, DOI 10.1038/ng.2007.21
   Nagaraja S, 2019, MOL CELL, V76, P965, DOI 10.1016/j.molcel.2019.08.030
   Nagaraja S, 2017, CANCER CELL, V31, P635, DOI 10.1016/j.ccell.2017.03.011
   Nakata S, 2017, BRAIN TUMOR PATHOL, V34, P113, DOI 10.1007/s10014-017-0288-6
   Narita T, 2017, BLOOD, V130, P1114, DOI 10.1182/blood-2016-09-741983
   Olow A, 2016, CLIN CANCER RES, V22, P5312, DOI 10.1158/1078-0432.CCR-15-1101
   Peterlin BM, 2006, MOL CELL, V23, P297, DOI 10.1016/j.molcel.2006.06.014
   Piunti A, 2017, NAT MED, V23, P493, DOI 10.1038/nm.4296
   Rueden CT, 2017, BMC BIOINFORMATICS, V18, DOI 10.1186/s12859-017-1934-z
   Ryall S, 2016, ACTA NEUROPATHOL COM, V4, DOI 10.1186/s40478-016-0353-0
   Shahbazi J, 2016, CLIN CANCER RES, V22, P2534, DOI 10.1158/1078-0432.CCR-15-1666
   Sharifnia T, 2019, NAT MED, V25, P292, DOI 10.1038/s41591-018-0312-3
   Smith E, 2011, GENE DEV, V25, P661, DOI 10.1101/gad.2015411
   Subramanian A, 2005, P NATL ACAD SCI USA, V102, P15545, DOI 10.1073/pnas.0506580102
   Taylor IC, 2015, J NEUROPATH EXP NEUR, V74, P778, DOI 10.1097/NEN.0000000000000216
   Thorvaldsdottir H, 2013, BRIEF BIOINFORM, V14, P178, DOI 10.1093/bib/bbs017
   Wu G, 2014, NAT GENET, V46, P444, DOI 10.1038/ng.2938
   Wu G, 2012, NAT GENET, V44, P251, DOI 10.1038/ng.1102
   Xing R, 2019, NAT COMMUN, V10, DOI 10.1038/s41467-019-09644-6
   Yu GC, 2015, BIOINFORMATICS, V31, P2382, DOI 10.1093/bioinformatics/btv145
   Zeitlinger J, 2007, NAT GENET, V39, P1512, DOI 10.1038/ng.2007.26
   Zhang Y, 2008, GENOME BIOL, V9, DOI 10.1186/gb-2008-9-9-r137
   Zhou Q, 2012, ANNU REV BIOCHEM, V81, P119, DOI 10.1146/annurev-biochem-052610-095910
NR 70
TC 18
Z9 19
U1 0
U2 3
PU CELL PRESS
PI CAMBRIDGE
PA 50 HAMPSHIRE ST, FLOOR 5, CAMBRIDGE, MA 02139 USA
SN 2211-1247
J9 CELL REP
JI Cell Reports
PD APR 7
PY 2020
VL 31
IS 1
AR 107485
DI 10.1016/j.celrep.2020.03.049
PG 21
WC Cell Biology
WE Science Citation Index Expanded (SCI-EXPANDED)
SC Cell Biology
GA LB9UO
UT WOS:000524976500024
PM 32268092
OA Green Submitted, gold
DA 2023-08-27
ER

PT J
AU Robert, J
   Cohen, N
AF Robert, J
   Cohen, N
TI Ontogeny of CTX expression in Xenopus
SO DEVELOPMENTAL AND COMPARATIVE IMMUNOLOGY
LA English
DT Article
DE comparative immunology; CD8; CTX; cortical thymocytes; double positive
   thymocytes; larvae; ontogeny; thymus; Xenopus
ID CLASS-I ANTIGENS; MONOCLONAL-ANTIBODY; IMMUNE-SYSTEM; CROSS-LINKING;
   TUMOR-CELLS; SURFACE; MOLECULE; METAMORPHOSIS; THYMOCYTES; EVOLUTION
AB In Xenopus, the type I transmembrane protein, CTX (cortical thymocyte-specific antigen of Xenopus), is hypothesized to play a role in thymocyte differentiation and/or thymic selection. The present studies were undertaken to gather additional evidence in support of a postulated association of CTX with a differentiation step of a cell population that corresponds to the murine immature double positive (CD4(+)CD8(+)TCR(+)) thymocyte subset. During ontogeny, cell surface expression of CTX on thymocytes can first be detected by immunocytochemistry and flow cytometry at 8 days post-fertilization, about I day after CD8(+) tells first appear. By 12 days post-fertilization, T-cells in the entire cortex except for the outer most layer of cells, are intensely CTX positive, whereas those in the medulla are negative. This pattern persists throughout larval and postmetamorphic life. (C) 1998 Elsevier Science Ltd. All rights reserved.
C1 Univ Rochester, Med Ctr, Dept Microbiol & Immunol, Rochester, NY 14642 USA.
C3 University of Rochester
RP Robert, J (通讯作者)，Univ Rochester, Med Ctr, Dept Microbiol & Immunol, 601 Elmwood Ave, Rochester, NY 14642 USA.
RI Robert, Jacques/B-2179-2008
OI Robert, Jacques/0000-0003-4380-1476
FU NICHD NIH HHS [HD-07901-30] Funding Source: Medline
CR Chretien I, 1996, EUR J IMMUNOL, V26, P780, DOI 10.1002/eji.1830260409
   Du Pasquier L, 1990, Dev Immunol, V1, P85, DOI 10.1155/1990/67913
   DUPASQUIER L, 1989, ANNU REV IMMUNOL, V7, P251, DOI 10.1146/annurev.immunol.7.1.251
   DuPasquier L, 1996, RES IMMUNOL, V147, P218, DOI 10.1016/0923-2494(96)87224-5
   DuPasquier L, 1996, RES IMMUNOL, V147, P261, DOI 10.1016/0923-2494(96)87229-4
   FLAJNIK MF, 1988, DEV BIOL, V128, P198, DOI 10.1016/0012-1606(88)90282-5
   Gravenor I, 1995, DEV COMP IMMUNOL, V19, P507, DOI 10.1016/0145-305X(95)00030-W
   HSU E, 1984, DIFFERENTIATION, V28, P109, DOI 10.1111/j.1432-0436.1984.tb00273.x
   Katevuo K., 1996, Scandinavian Journal of Immunology, V43, P694
   KOBEL HR, 1975, IMMUNOGENETICS, V2, P7
   NAGATA S, 1985, EUR J IMMUNOL, V15, P837, DOI 10.1002/eji.1830150818
   NAGATA S, 1977, CELL TISSUE RES, V179, P87
   NIEUWKOOP RD, 1967, NORMAL TABLES XENOPU
   PENIT C, 1995, J IMMUNOL, V154, P5103
   Robert J, 1997, EXP CELL RES, V235, P227, DOI 10.1006/excr.1997.3672
   Robert J, 1997, MOL IMMUNOL, V34, P133, DOI 10.1016/S0161-5890(97)00006-0
   ROBERT J, UNPUB IN VITRO DIFFE
   Rollins-Smith L A, 1990, Dev Immunol, V1, P97, DOI 10.1155/1990/25197
   RollinsSmith LA, 1997, DEV IMMUNOL, V5, P133, DOI 10.1155/1997/38464
   ROLLINSSMITH LA, 1996, METAMORPHOSIS POSTEM, P626
   SALTERCID L, 1995, CRIT REV IMMUNOL, V15, P31, DOI 10.1615/CritRevImmunol.v15.i1.20
   TOCHINAI S, 1978, DEV COMP IMMUNOL, V2, P627, DOI 10.1016/S0145-305X(78)80088-3
   TOCHINAI S, 1980, DEV COMP IMMUNOL, V4, P273, DOI 10.1016/S0145-305X(80)80031-0
   TURPEN JB, 1989, J IMMUNOL, V142, P41
NR 24
TC 19
Z9 19
U1 0
U2 0
PU PERGAMON-ELSEVIER SCIENCE LTD
PI OXFORD
PA THE BOULEVARD, LANGFORD LANE, KIDLINGTON, OXFORD OX5 1GB, ENGLAND
SN 0145-305X
J9 DEV COMP IMMUNOL
JI Dev. Comp. Immunol.
PD SEP-DEC
PY 1998
VL 22
IS 5-6
BP 605
EP 612
DI 10.1016/S0145-305X(98)00028-7
PG 8
WC Fisheries; Immunology; Veterinary Sciences; Zoology
WE Science Citation Index Expanded (SCI-EXPANDED)
SC Fisheries; Immunology; Veterinary Sciences; Zoology
GA 157XX
UT WOS:000078087900011
PM 9877440
OA hybrid
DA 2023-08-27
ER

PT J
AU Bahnemann, R
   Leibold, E
   Kittel, B
   Mellert, W
   Jackh, R
AF Bahnemann, R
   Leibold, E
   Kittel, B
   Mellert, W
   Jackh, R
TI Different patterns of kidney toxicity after subacute administration of
   Na-nitrilotriacetic acid and Fe-nitrilotriacetic acid to Wistar rats
SO TOXICOLOGICAL SCIENCES
LA English
DT Article
ID RENAL-CELL CARCINOMA; FEMALE B6C3F(1) MICE; MALE F344 RATS; FERRIC
   NITRILOTRIACETATE; IRON-NITRILOTRIACETATE; LIPID-PEROXIDATION;
   DRINKING-WATER; STRAND BREAKS; LIVER DNA; PROLIFERATION
AB Na-nitrilotriacetic acid (Na(3)NTA) and Fe-nitrilotriacetic acid (FeNTA) have both been described to cause tumors in the urinary tract of rodents. However, these effects were observed using different modes of administration at extremely different dose levels and explained by different mechanisms. Whereas FeNTA causes an iron overload of cells and is genotoxic in various assays, Na(3)NTA is predominantly bound to zinc in vivo and thereby causes cytotoxic effects in the urinary tract. In contrast to FeNTA, Na(3)NTA requires high dose levels to produce tumors. The aim of this study was to compare the effects of Na(3)NTA and FeNTA on cellular proliferation, histopathology, lipid peroxidation, and 8-OH-2'-deoxyguanosine levels in the kidneys as well as on the urinary excretion of Ca, Fe, and Zn. For evaluation of DNA synthesis both compounds were administered for 1 or 4 weeks to 14-week-old male Wistar rats at a tumor causing dose, Na(3)NTA via the diet at 150 ppm and 20,000 ppm ((similar to)9 and similar to 1000 mg/kg/day) and FeNTA ip at 25 mg/kg/day. An osmotic minipump, containing 20 mg/ml BrdU, was implanted subcutaneously 7 days before necropsy. Na(3)NTA showed nearly no effect on DNA replication after 1 week but a strong reaction after 4 weeks. The increase was 10- to 18-fold in different renal compartments. The enhancement of proliferation in the proximal tubules was nearly twice that in the distal tubules. In contrast, FeNTA caused DNA replication during the first week, and this was restricted to the proximal tubules. After 4 weeks there was an 18-fold increase in the outer stripe and no effect in the inner stripe of the outer medulla. The data presented give evidence to the assumption that both substances increase cell proliferation as a compensatory mechanism, causing different pattern of tubular proliferation in terms of time course and affected cell types. Both Na(3)NTA at 20,000 ppm and FeNTA led to increased lipid peroxidation, whereas increased levels of 8-OH-2'-deoxyguanosine were observed only after treatment with FeNTA. Urinary excretion of Zn was increased 30-fold after administration of 20,000 ppm Na(3)NTA but only 2-fold after administration of FeNTA. Urinary excretion of Ca and Fe remained unchanged after treatment with either Na(3)NTA and FeNTA. These results show that the Na(3)NTA-related proliferative effects are not mediated by an internal formation of FeNTA. (C) 1998 Society Toxicology.
C1 BASF AG, Dept Toxicol, D-67056 Ludwigshafen, Germany.
C3 BASF
RP Bahnemann, R (通讯作者)，BASF AG, Dept Toxicol, D-67056 Ludwigshafen, Germany.
CR ALDEN CL, 1982, FOOD CHEM TOXICOL, V20, P441, DOI 10.1016/S0278-6915(82)80110-5
   ALDEN CL, 1982, FOOD CHEM TOXICOL, V20, P935, DOI 10.1016/S0015-6264(82)80231-9
   ALDEN CL, 1981, VET PATHOL, V18, P549, DOI 10.1177/030098588101800413
   ANDERSON RL, 1985, CRC CR REV TOXICOL, V15, P1, DOI 10.3109/10408448509023766
   ANDERSON RL, 1981, FOOD COSMET TOXICOL, V19, P639, DOI 10.1016/0015-6264(81)90516-2
   ANDERSON RL, 1982, FOOD CHEM TOXICOL, V20, P105, DOI 10.1016/S0278-6915(82)80017-3
   [Anonymous], 1982, JPN ARCH INTERN MED
   CASANOVA M, 1994, FUND APPL TOXICOL, V23, P525, DOI 10.1006/faat.1994.1137
   CHOIE DD, 1972, AM J PATHOL, V68, P359
   COSTA R, 1988, ENVIRON MOL MUTAGEN, V12, P397, DOI 10.1002/em.2860120408
   DAHLHAUS M, 1994, TOXICOL LETT, V74, P265, DOI 10.1016/0378-4274(94)90085-X
   DAHLHAUS M, 1993, MUTAT RES, V285, P295, DOI 10.1016/0027-5107(93)90118-Y
   DEGUCHI J, 1995, JPN J CANCER RES, V86, P1068, DOI 10.1111/j.1349-7006.1995.tb03022.x
   DIETRICH DR, 1991, MUTAT RES, V248, P239, DOI 10.1016/0027-5107(91)90060-2
   EBINA Y, 1986, J NATL CANCER I, V76, P107
   FIALA ES, 1989, CANCER RES, V49, P5518
   GODDARD JG, 1983, BIOCHEM PHARMACOL, V32, P3879, DOI 10.1016/0006-2952(83)90169-7
   GOLDSWORTHY TL, 1991, PROG CLIN BIOL RES, V369, P253
   GOLDSWORTHY TL, 1988, TOXICOL APPL PHARM, V96, P367, DOI 10.1016/0041-008X(88)90095-6
   GOTTSCHLING B, 1998, THESIS U LEIPZIG
   GOYER RA, 1981, J NATL CANCER I, V66, P869
   HAGIWARA A, 1989, FUND APPL TOXICOL, V13, P332, DOI 10.1016/0272-0590(89)90269-8
   HAMAZAKI S, 1985, TOXICOL APPL PHARM, V77, P267, DOI 10.1016/0041-008X(85)90326-6
   HARTWIG A, 1993, CARCINOGENESIS, V14, P107, DOI 10.1093/carcin/14.1.107
   HIASA Y, 1985, CARCINOGENESIS, V6, P907, DOI 10.1093/carcin/6.6.907
   HIASA Y, 1984, J NATL CANCER I, V73, P297
   LARSON JL, 1994, FUND APPL TOXICOL, V23, P537, DOI 10.1006/faat.1994.1138
   LARSON JL, 1994, FUND APPL TOXICOL, V22, P90, DOI 10.1006/faat.1994.1012
   LARSON JL, 1994, FUND APPL TOXICOL, V22, P431, DOI 10.1006/faat.1994.1049
   LEBOEUF RA, 1990, ENVIRON MOL MUTAGEN, V15, P117
   LI JL, 1987, CANCER RES, V47, P1867
   MIYASHIRO A, 1984, J NARA MED ASS, V35, P550
   MONTICELLO TM, 1991, TOXICOL APPL PHARM, V111, P409, DOI 10.1016/0041-008X(91)90246-B
   MOTTOLA H A, 1974, Toxicological and Environmental Chemistry Reviews, V2, P99, DOI 10.1080/02772247409356922
   NAKATSUKA S, 1989, Kawasaki Medical Journal, V15, P25
   *NIH DEP HLTH WELL, 1977, PUBL NIH DEP HLTH WE
   NIXON GA, 1972, TOXICOL APPL PHARM, V21, P244, DOI 10.1016/0041-008X(72)90067-1
   *NTP, 1990, 347 NTP
   OLSON MJ, 1987, TOXICOL APPL PHARM, V90, P43, DOI 10.1016/0041-008X(87)90304-8
   PREECE NE, 1988, TOXICOL APPL PHARM, V93, P89, DOI 10.1016/0041-008X(88)90028-2
   SHORT BG, 1993, ENVIRON HEALTH PERSP, V101, P115, DOI 10.2307/3431853
   SHORT BG, 1991, PROG CLIN BIOL RES, V369, P357
   TOYOKUNI S, 1993, CARCINOGENESIS, V14, P223, DOI 10.1093/carcin/14.2.223
   UMEMURA T, 1990, CARCINOGENESIS, V11, P345, DOI 10.1093/carcin/11.2.345
   Umemura T, 1996, JPN J CANCER RES, V87, P882, DOI 10.1111/j.1349-7006.1996.tb02115.x
   WARD JM, 1991, PROG CLIN BIOL RES, V369, P369
NR 46
TC 11
Z9 11
U1 0
U2 6
PU OXFORD UNIV PRESS
PI OXFORD
PA GREAT CLARENDON ST, OXFORD OX2 6DP, ENGLAND
SN 1096-6080
J9 TOXICOL SCI
JI Toxicol. Sci.
PD NOV
PY 1998
VL 46
IS 1
BP 166
EP 175
PG 10
WC Toxicology
WE Science Citation Index Expanded (SCI-EXPANDED)
SC Toxicology
GA 151GB
UT WOS:000077711400018
PM 9928680
DA 2023-08-27
ER

PT J
AU Zhou, JN
   Fe, DY
   Wu, QW
AF Zhou, JN
   Fe, DY
   Wu, QW
TI Potential of intensity-modulated radiotherapy to escalate doses to
   head-and-neck cancers: What is the maximal dose?
SO INTERNATIONAL JOURNAL OF RADIATION ONCOLOGY BIOLOGY PHYSICS
LA English
DT Article
DE head-and-neck cancer; intensity-modulated radiotherapy; dose escalation
ID RADIATION-THERAPY; MULTILEAF COLLIMATORS; CONFORMAL RADIOTHERAPY; VOLUME
   CONSTRAINTS; PROSTATE-CANCER; OPTIMIZATION; EXPERIENCE; COMPENSATION;
   TOMOTHERAPY; PROJECTIONS
AB Purpose: To investigate the potential of intensity-modulated radiotherapy (IMRT) to escalate doses to head-and-neck cancer and find the maximal dose that could be prescribed to the target volume with IMRT while doses to critical organs were maintained at their currently acceptable levels. The secondary goal was to search for limits in current IMRT technology.
   Methods and Materials: For a group of 12 head-and-neck cancer patients with different tumor locations and shapes, we performed IMRT planning using a simultaneous integrated boost strategy, that is, the gross tumor volume (GTV), clinical target volume (CTV), and electively treated nodes were treated simultaneously at different dose levels. The critical structures involved in the treatment field that needed to be spared included the brainstem, spinal cord, and parotid glands, depending on the disease site. Nine coplanar 6-MV photon beams were used for planning with the IMRT system developed at our institution, and dose-volume criteria were used for optimization. By varying the optimization parameters, we gradually increased the dose to the GTV while keeping the dose to the critical structures at less than the acceptable tolerance level. The criteria for accepting the plan included the following: (1) the prescription dose to the GTV had to cover 99% of the volume, and the dose homogeneity of the GTV needed to be <10%; (2) the prescription to the CTV (which was set either at 60 Gy or 10 Gy less than that of the GTV) had to cover 95% of the volume, and the same amount of normal tissue outside the CTV received the CTV prescription dose as in the current acceptable plan; (3) the prescription to the electively treated lymph nodes needed to cover 90% of the volume; and (4) the maximal dose to the brainstem and spinal cord had to be <55 Gy and 45 Gy, respectively. For parotid glands, the dose needed to be as low as possible without compromising the target doses. The deliverable plans as determined by the actual multileaf collimator leaf sequences were used for the final evaluation. To verify that the acceptable plans were deliverable, the experimental measurements of planar dose distribution were performed in phantom with film.
   Results: The maximal dose to the GTV varied from 86 to 176 Gy if the CTV dose increased with the GTV dose. It was reduced to 76-82 Gy if the CTV dose was kept at 60 Gy. The competing criteria usually are the requirements of the tolerance doses to the critical organs and target dose homogeneity, not the target prescription dose. Using more beams only increased the dose marginally. The results could change significantly if a different set of criteria for the plan evaluation were used. Dosimetric measurements confirmed that such a high dose and dose gradient could be delivered accurately with dynamic multileaf collimators. Statistical analyses showed no significant correlations between the maximal doses and the number of GTVs and volume of GTVs and CTVs.
   Conclusion: Doses to head-and-neck cancers with simultaneous integrated boost IMRT can be escalated to a greater level than currently prescribed clinically. The limit of IMRT in head-and-neck cancer has not been reached at the current prescription level of 70 Gy. Such high total and fractionated doses should be carefully evaluated before being prescribed clinically. (C) 2003 Elsevier Inc.
C1 Virginia Commonwealth Univ, Dept Radiat Oncol, Richmond, VA 23298 USA.
   Virginia Commonwealth Univ, Dept Biomed Engn, Richmond, VA 23298 USA.
C3 Virginia Commonwealth University; Virginia Commonwealth University
RP Wu, QW (通讯作者)，Virginia Commonwealth Univ, Dept Radiat Oncol, POB 980058,401 Coll St, Richmond, VA 23298 USA.
OI Wu, Qiuwen/0000-0003-0748-7280
CR [Anonymous], 50 ICRU S
   BORTFELD T, 1990, PHYS MED BIOL, V35, P1423, DOI 10.1088/0031-9155/35/10/007
   BORTFELD TR, 1994, INT J RADIAT ONCOL, V28, P723, DOI 10.1016/0360-3016(94)90200-3
   Boyer AL, 2001, INT J RADIAT ONCOL, V51, P880, DOI 10.1016/S0360-3016(01)01749-7
   Brahme A, 1999, SEMIN RADIAT ONCOL, V9, P35, DOI 10.1016/S1053-4296(99)80053-8
   Burman C, 1997, INT J RADIAT ONCOL, V39, P863, DOI 10.1016/S0360-3016(97)00458-6
   Butler EB, 1999, INT J RADIAT ONCOL, V45, P21, DOI 10.1016/S0360-3016(99)00101-7
   Chao KSC, 2003, INT J RADIAT ONCOL, V55, P312, DOI 10.1016/S0360-3016(02)03940-8
   Chao KSC, 2000, INT J CANCER, V90, P92, DOI 10.1002/(SICI)1097-0215(20000420)90:2<92::AID-IJC5>3.0.CO;2-9
   Cho PS, 1998, MED PHYS, V25, P435, DOI 10.1118/1.598218
   Dawson LA, 2000, INT J RADIAT ONCOL, V46, P1117, DOI 10.1016/S0360-3016(99)00550-7
   Eisbruch A, 1999, Acta Otorhinolaryngol Belg, V53, P271
   Eisbruch A, 1999, INT J RADIAT ONCOL, V45, P577, DOI 10.1016/S0360-3016(99)00247-3
   EISBRUCH A, 2001, RTOG H0022 PHASE I I
   HOLMES T, 1994, MED PHYS, V21, P303, DOI 10.1118/1.597291
   Hunt MA, 2001, INT J RADIAT ONCOL, V49, P623, DOI 10.1016/S0360-3016(00)01389-4
   *INT COMM RAD UN M, 1993, ICRU50 PRESCR REC RE
   Kuppersmith R B, 1999, Ear Nose Throat J, V78, P241
   Ling CC, 1996, INT J RADIAT ONCOL, V35, P721, DOI 10.1016/0360-3016(96)00174-5
   Llacer J, 2001, PHYS MED BIOL, V46, P2637, DOI 10.1088/0031-9155/46/10/309
   Mackie TR, 1999, SEMIN RADIAT ONCOL, V9, P1, DOI 10.1016/S1053-4296(99)80050-2
   Mohan R, 2000, INT J RADIAT ONCOL, V46, P619, DOI 10.1016/S0360-3016(99)00438-1
   Shepard DM, 2000, PHYS MED BIOL, V45, P69, DOI 10.1088/0031-9155/45/1/306
   SPIROU SV, 1994, MED PHYS, V21, P1031, DOI 10.1118/1.597345
   Spirou SV, 1998, MED PHYS, V25, P321, DOI 10.1118/1.598202
   STEIN J, 1994, RADIOTHER ONCOL, V32, P163, DOI 10.1016/0167-8140(94)90103-1
   Sultanem K, 2000, INT J RADIAT ONCOL, V48, P711, DOI 10.1016/S0360-3016(00)00702-1
   SVENSSON R, 1994, PHYS MED BIOL, V39, P37, DOI 10.1088/0031-9155/39/1/003
   Verellen D, 1997, INT J RADIAT ONCOL, V39, P99, DOI 10.1016/S0360-3016(97)00304-0
   WU Q, 2003, INT J RADIAT ONCOL, V56, P571
   Wu QW, 2000, MED PHYS, V27, P701, DOI 10.1118/1.598932
   Wu QW, 2000, INT J RADIAT ONCOL, V46, P195, DOI 10.1016/S0360-3016(99)00304-1
   Xia P, 2001, INT J RADIAT ONCOL, V51, P244, DOI 10.1016/S0360-3016(01)01643-1
   Zelefsky MJ, 2001, J UROLOGY, V166, P876, DOI 10.1016/S0022-5347(05)65855-7
NR 34
TC 20
Z9 22
U1 0
U2 1
PU ELSEVIER SCIENCE INC
PI NEW YORK
PA 360 PARK AVE SOUTH, NEW YORK, NY 10010-1710 USA
SN 0360-3016
J9 INT J RADIAT ONCOL
JI Int. J. Radiat. Oncol. Biol. Phys.
PD NOV 1
PY 2003
VL 57
IS 3
BP 673
EP 682
DI 10.1016/S0360-3016(03)00626-6
PG 10
WC Oncology; Radiology, Nuclear Medicine & Medical Imaging
WE Science Citation Index Expanded (SCI-EXPANDED)
SC Oncology; Radiology, Nuclear Medicine & Medical Imaging
GA 730GQ
UT WOS:000185821200009
PM 14529771
DA 2023-08-27
ER

PT J
AU Aruga, J
AF Aruga, J
TI Slitrk6 expression profile in the mouse embryo and its relationship to
   that of Nlrr3
SO GENE EXPRESSION PATTERNS
LA English
DT Article
DE neural fold; inner ear; limb bud; neural crest; external sensory organ;
   sensory neuroepithelium; skin; vibrissae; hair root; transmembrane
   protein; neural development; cerebellar development; esophagus; stomach;
   intestine; inner muscular layer; neural patterning; cerebellar
   compartmentalization; gene expression; slit; PGP9.5; brain tumor;
   medulloblastoma
ID LEUCINE-RICH REPEATS; NERVOUS-SYSTEM; PROTEIN; GENE; CLONING
AB Slitrk6 is a member of the Slitrk family of proteins, which are integral membrane proteins possessing two leucine-rich repeat (LRR) domains and a carboxy-terminal domain partially similar to that in the trk neurotrophin receptor proteins. Here, I show that Slitrk6 is uniquely expressed in various organs, different from other Slitrk genes which are predominantly expressed in neural tissues. In the developing mouse embryo, Slitrk6 expression was detected in the otic cyst, lateral trunk epidermis and its underlying mesenchymal tissue, limb bud, maxillary process, pharyngeal arches, cochlea, retina, tongue, tooth primordium, central nervous system (CNS), and the visceral organ primordia including of the lung, gastrointestinal tract (particularly in the enteric neurons) and pancreas. The expression in these organs occurred in a spatially restricted manner. In the CNS, the expression was highly compartmentalized in the dorsal thalamus, cerebellum and medulla. The expression compartment in the thalamus in which Slitrk6 was expressed was closely related to the Gbx2-expressing prosomere 2. Interestingly, the Slitrk6 expression in the CNS, cochlea, tongue, tooth primordial, and other organs was partially complementary to the expression of Nlrr3, which belongs to another family of neuronal LRR-containing transmembrane proteins. The complementary expression of the two proteins in the dorsal thalamus persisted from E13.5 to the adult stage. (C) 2003 Elsevier B.V. All rights reserved.
C1 RIKEN, Brain Sci Inst, Dev Neurobiol Lab, Wako, Saitama 3510198, Japan.
C3 RIKEN
RP Aruga, J (通讯作者)，RIKEN, Brain Sci Inst, Dev Neurobiol Lab, 2-1 Hirosawa, Wako, Saitama 3510198, Japan.
RI Aruga, Jun/H-5837-2011
OI Aruga, Jun/0000-0001-7936-1375
CR ALTMAN J, 1997, DEV CEREBELLAR SYSTE, P206
   ARUGA J, 2003, IN PRESS MOL CELL NE
   ARUGA J, 2003, IN PRESS GENE
   Bormann P, 1999, MOL CELL NEUROSCI, V13, P167, DOI 10.1006/mcne.1999.0742
   Brose K, 2000, CURR OPIN NEUROBIOL, V10, P95, DOI 10.1016/S0959-4388(99)00066-5
   BULFONE A, 1993, J NEUROSCI, V13, P3155
   Fukamachi K, 2001, BIOCHEM BIOPH RES CO, V287, P257, DOI 10.1006/bbrc.2001.5579
   KOBE B, 1995, CURR OPIN STRUC BIOL, V5, P409, DOI 10.1016/0959-440X(95)80105-7
   KRAMMER HJ, 1993, ANN ANAT, V175, P321, DOI 10.1016/S0940-9602(11)80029-4
   Patapoutian A, 2001, CURR OPIN NEUROBIOL, V11, P272, DOI 10.1016/S0959-4388(00)00208-7
   Rubenstein JLR, 1998, ANNU REV NEUROSCI, V21, P445, DOI 10.1146/annurev.neuro.21.1.445
   Taniguchi H, 1996, MOL BRAIN RES, V36, P45, DOI 10.1016/0169-328X(95)00243-L
   WILKINSON DG, 1992, IN SITU HYBRIDIZATIO, P75
   Yagi T, 2000, GENE DEV, V14, P1169
NR 14
TC 21
Z9 23
U1 0
U2 1
PU ELSEVIER SCIENCE BV
PI AMSTERDAM
PA PO BOX 211, 1000 AE AMSTERDAM, NETHERLANDS
SN 1567-133X
J9 GENE EXPR PATTERNS
JI Gene Expr. Patterns
PD DEC
PY 2003
VL 3
IS 6
BP 727
EP 733
DI 10.1016/S1567-133X(03)00141-8
PG 7
WC Developmental Biology; Genetics & Heredity
WE Science Citation Index Expanded (SCI-EXPANDED)
SC Developmental Biology; Genetics & Heredity
GA 755JA
UT WOS:000187398700005
PM 14643680
DA 2023-08-27
ER

PT J
AU Huang, YQ
   Chan, LL
   Bai, HX
   Li, XJ
   Zhang, ZS
   Wang, YY
   Cao, Y
   Karakousis, G
   Huang, R
   Xiao, B
   Zhang, PJ
   Yang, L
AF Huang, Yuqian
   Chan, Lilian
   Bai, Harrison X.
   Li, Xuejun
   Zhang, Zishu
   Wang, Yinyan
   Cao, Ya
   Karakousis, Giorgos
   Huang, Raymond
   Xiao, Bo
   Zhang, Paul J.
   Yang, Li
TI Assessment of care pattern and outcome in hemangioblastoma
SO SCIENTIFIC REPORTS
LA English
DT Article
ID HIPPEL-LINDAU-DISEASE; CENTRAL-NERVOUS-SYSTEM; BRAIN-STEM
   HEMANGIOBLASTOMAS; SPORADIC CEREBELLAR HEMANGIOBLASTOMAS; SPINAL-CORD
   HEMANGIOBLASTOMAS; GAMMA-KNIFE RADIOSURGERY; LONG-TERM OUTCOMES;
   INTRACRANIAL HEMANGIOBLASTOMAS; STEREOTACTIC RADIOSURGERY;
   SURGICAL-MANAGEMENT
AB Due to its rarity, current literature assessing prognostic factors and survival outcomes of hemangioblastoma is limited. Patients with histologically confirmed hemangioblastoma were identified from the US National Cancer Data Base. 1488 patients met inclusion criteria. 644 patients underwent gross total resection (GTR), 220 subtotal resection (STR)/biopsy, 60 stereotactic radiosurgery (SRS), 15 external beam radiotherapy (EBRT), 51 surgery followed by radiotherapy (SR + RT) and 498 no treatment. Independent predictors of shorter OS included age >= 40 (HR, 3.897; 95% CI, 2.341-6.487; p < 0.001), Charlson-Deyo score >= 1(HR, 1.756; 95% CI, 1.213-2.544; p = 0.003), tumor location in the brainstem (HR, 1.955; 95% CI, 1.129-3. 384; p = 0.017) compared to cerebellum, no treatment (HR, 2530; 95% CI, 1.533-4.177; p < 0.001) and receipt of EBRT (HR, 2.860; 95% CI, 1.073-7.618; p = 0.036) compared to STR/biopsy. GTR was associated with longer OS (HR 0.617; 95% CI, 0.391-0.974; p = 0.038), while SRS had comparable OS to STR/biopsy. The overall trend of OS by treatment modality was consistent after matching to age-and sex-matched US population data. In patients younger than 40 years, treatment was not a significant predictor of OS. In conclusion, GTR remained the optimal treatment for hemangioblastoma. SRS may perform similarly to surgery alone. Treatment was not a significant predictor of survival in younger patients.
C1 [Huang, Yuqian; Yang, Li] Cent S Univ, Xiangya Hosp 2, Dept Neurol, Changsha 410011, Hunan, Peoples R China.
   [Xiao, Bo] Cent S Univ, Xiangya Hosp, Dept Neurol, Changsha 410011, Hunan, Peoples R China.
   [Chan, Lilian] Perelman Sch Med, Philadelphia, PA 19104 USA.
   [Bai, Harrison X.] Hosp Univ Penn, Dept Radiol, 3400 Spruce St, Philadelphia, PA 19104 USA.
   [Li, Xuejun] Cent S Univ, Xiangya Hosp, Dept Neurosurg, Changsha 410011, Hunan, Peoples R China.
   [Karakousis, Giorgos] Hosp Univ Penn, Dept Surg, 3400 Spruce St, Philadelphia, PA 19104 USA.
   [Cao, Ya] Cent S Univ, Canc Res Inst, Sch Basic Med, Changsha 410078, Hunan, Peoples R China.
   [Zhang, Paul J.] Hosp Univ Penn, Dept Pathol & Lab Med, 3400 Spruce St, Philadelphia, PA 19104 USA.
   [Zhang, Zishu] Cent S Univ, Xiangya Hosp 2, Dept Radiol, Changsha 410011, Hunan, Peoples R China.
   [Huang, Raymond] Harvard Med Sch, Brigham & Womens Hosp, Dept Radiol, Boston, MA 02120 USA.
   [Wang, Yinyan] Capital Med Univ, Beijing Tiantan Hosp, Dept Neurosurg, Beijing, Peoples R China.
C3 Central South University; Central South University; University of
   Pennsylvania; Pennsylvania Medicine; Central South University;
   University of Pennsylvania; Pennsylvania Medicine; Central South
   University; University of Pennsylvania; Pennsylvania Medicine; Central
   South University; Harvard University; Brigham & Women's Hospital;
   Harvard Medical School; Capital Medical University
RP Yang, L (通讯作者)，Cent S Univ, Xiangya Hosp 2, Dept Neurol, Changsha 410011, Hunan, Peoples R China.; Xiao, B (通讯作者)，Cent S Univ, Xiangya Hosp, Dept Neurol, Changsha 410011, Hunan, Peoples R China.
EM
RI Wang, Yinyan/AAJ-1160-2020
OI Huang, Raymond/0000-0001-7661-797X
FU ShenghuaYuying Project of Central South University [81472594, 81770781];
   National Science Foundation of China [81472594, 81770781]
FX ShenghuaYuying Project of Central South University to L.Y. and National
   Science Foundation of China to XJL (81472594 and 81770781).
CR Ammerman JM, 2006, J NEUROSURG, V105, P248, DOI 10.3171/jns.2006.105.2.248
   Chen LF, 2013, J CLIN NEUROSCI, V20, P1727, DOI 10.1016/j.jocn.2013.01.027
   Conway JE, 2001, NEUROSURGERY, V48, P55, DOI 10.1097/00006123-200101000-00009
   Dickman PW, 2004, STAT MED, V23, P51, DOI 10.1002/sim.1597
   Fukuda M, 2014, WORLD NEUROSURG, V82, P815, DOI 10.1016/j.wneu.2014.06.018
   Glasker S, 2010, CENT EUR NEUROSURG, V71, P80, DOI 10.1055/s-0029-1234040
   Gossage L, 2015, NAT REV CANCER, V15, P55, DOI 10.1038/nrc3844
   Goyal N, 2016, J NEUROSCI RURAL PRA, V7, P23, DOI 10.4103/0976-3147.172165
   Hanakita S, 2014, NEURO-ONCOLOGY, V16, P429, DOI 10.1093/neuonc/not201
   Iyer JG, 2014, J AM ACAD DERMATOL, V70, P637, DOI 10.1016/j.jaad.2013.11.031
   Jagannathan J, 2008, J NEUROSURG, V108, P210, DOI 10.3171/JNS/2008/108/2/0210
   Jawahar A, 2000, ACTA NEUROCHIR, V142, P641, DOI 10.1007/s007010070107
   Kanno H, 2009, SPINAL CORD, V47, P447, DOI 10.1038/sc.2008.151
   Kano H, 2008, NEUROSURGERY, V63, P443, DOI 10.1227/01.NEU.0000313120.81565.D7
   Kano H, 2015, J NEUROSURG, V122, P1469, DOI 10.3171/2014.10.JNS131602
   Koh ES, 2007, INT J RADIAT ONCOL, V69, P1521, DOI 10.1016/j.ijrobp.2007.05.025
   Liao CC, 2014, CLIN NEUROL NEUROSUR, V125, P160, DOI 10.1016/j.clineuro.2014.08.001
   Lonser RR, 2014, J NEUROSURG, V120, P1055, DOI 10.3171/2014.1.JNS131431
   Ma DX, 2015, WORLD NEUROSURG, V84, P1030, DOI 10.1016/j.wneu.2015.05.030
   Moss JM, 2009, NEUROSURGERY, V65, P79, DOI 10.1227/01.NEU.0000348015.51685.D2
   NEUMANN HPH, 1989, J NEUROSURG, V70, P24, DOI 10.3171/jns.1989.70.1.0024
   Niu L, 2016, CLIN NEUROL NEUROSUR, V150, P59, DOI 10.1016/j.clineuro.2016.08.028
   Padhi S, 2011, J CLIN NEUROSCI, V18, P939, DOI 10.1016/j.jocn.2010.12.050
   Parker F, 2009, J NEURO-ONCOL, V93, P133, DOI 10.1007/s11060-009-9861-0
   Patrice SJ, 1996, INT J RADIAT ONCOL, V35, P493
   Pohar M, 2006, COMPUT METH PROG BIO, V81, P272, DOI 10.1016/j.cmpb.2006.01.004
   Richard S, 2000, NEUROSURG REV, V23, P1, DOI 10.1007/s101430050024
   Sayer FT, 2011, WORLD NEUROSURG, V75, P99, DOI 10.1016/j.wneu.2010.09.032
   Van Velthoven V, 2003, NEUROSURGERY, V53, P1306, DOI 10.1227/01.NEU.0000093497.81390.29
   Wanebo JE, 2003, J NEUROSURG, V98, P82, DOI 10.3171/jns.2003.98.1.0082
   Wang C, 2001, SURG NEUROL, V56, P226
   Wang CC, 2001, SURG NEUROL, V56, P218, DOI 10.1016/S0090-3019(01)00590-0
   Wang EM, 2005, J NEUROSURG, V102, P225, DOI 10.3171/jns.2005.102.s_supplement.0225
   Weil RJ, 2003, J NEUROSURG, V98, P95, DOI 10.3171/jns.2003.98.1.0095
   Westwick HJ, 2016, NEUROEPIDEMIOLOGY, V46, P14, DOI 10.1159/000441147
   Wind JJ, 2011, J NEUROSURG, V114, P1312, DOI 10.3171/2010.9.JNS10839
   Woodward ER, 2007, BRAIN, V130, P836, DOI 10.1093/brain/awl362
   Xu QW, 2010, ACTA NEUROCHIR, V152, P1331, DOI 10.1007/s00701-010-0668-8
NR 38
TC 10
Z9 10
U1 0
U2 5
PU NATURE PUBLISHING GROUP
PI LONDON
PA MACMILLAN BUILDING, 4 CRINAN ST, LONDON N1 9XW, ENGLAND
SN 2045-2322
J9 SCI REP-UK
JI Sci Rep
PD JUL 24
PY 2018
VL 8
AR 11144
DI 10.1038/s41598-018-29047-9
PG 8
WC Multidisciplinary Sciences
WE Science Citation Index Expanded (SCI-EXPANDED)
SC Science & Technology - Other Topics
GA GN9TF
UT WOS:000439550200016
PM 30042517
OA Green Published, gold
DA 2023-08-27
ER

PT J
AU Buttini, M
   Limonta, S
   Boddeke, HWGM
AF Buttini, M
   Limonta, S
   Boddeke, HWGM
TI Peripheral administration of lipopolysaccharide induces activation of
   microglial cells in rat brain
SO NEUROCHEMISTRY INTERNATIONAL
LA English
DT Article
ID TUMOR-NECROSIS-FACTOR; TRANSIENT FOREBRAIN ISCHEMIA; CEREBRAL-ARTERY
   OCCLUSION; MONONUCLEAR PHAGOCYTES; BACTERIAL-MENINGITIS;
   CEREBROSPINAL-FLUID; INTERFERON-GAMMA; NERVOUS-SYSTEM; SPINAL-CORD;
   EXPRESSION
AB Using immunocytochemistry with monoclonal antibodies against surface immunomolecules and Griffonia simplicifolia lectin histochemistry, the microglial cell reaction in rat brain was studied after intravenous injection of lipopolysaccharide (LPS). Activation of microglial cells throughout the brain became apparent within hours and peaked at 8-24 h following administration of 1, 2.5 and 5 mg/kg LPS. High doses of LPS (2.5 and 5 mg/kg) induced a morphological transition of resting ramified microglia to round, macrophage-like cells in the anterior hypothalamus, thalamus and the brainstem. After injection of 1 mg/kg LPS, this morphological transition was only detected in the brainstem. Microglial cell reactivity gradually returned to control levels within 7 days after LPS administration. Furthermore, LPS induced enhanced expression of MHC class II by microglial cells. Maximal up-regulation of MHC class II Ia-antigen was found 3 days following injection of LPS, and only a few highly Ia immunoreactive cells were detectable 7 days following injection of LPS. Despite the presence of highly activated microglial cells in the rat brain, no signs of tissue damage were observed at any time point after injection of LPS examined. In addition to the activation of microglial cells, intravenous injection of LPS induced accumulations of macrophages in blood vessels of the choroid plexus and the brain, but no disruption of vessels with subsequent invasion of parenchyma by blood macrophages was detected. Our data demonstrate that a peripheral immune challenge leads to a high and transitory activation of microglial cells in the brain which could possibly contribute to the pathology of infections and septic shock. Copyright (C) 1996 Elsevier Science Ltd.
C1 SANDOZ PHARMA LTD, PRECLIN RES, CH-4002 BASEL, SWITZERLAND.
C3 Novartis; Sandoz
OI Buttini, Manuel/0000-0003-1805-0279
CR ANDERSSON PB, 1992, NEUROSCIENCE, V48, P169, DOI 10.1016/0306-4522(92)90347-5
   BANATI RB, 1993, GLIA, V7, P111, DOI 10.1002/glia.440070117
   BUTTINI M, 1994, MOL BRAIN RES, V23, P126, DOI 10.1016/0169-328X(94)90218-6
   BUTTINI M, 1995, NEUROSCIENCE, V65, P523, DOI 10.1016/0306-4522(94)00525-A
   BUTTINI M, 1995, UNPUB EXPRESSION TUM
   CHAO CC, 1992, J IMMUNOL, V149, P2736
   COCEANI F, 1988, BRAIN RES, V446, P245, DOI 10.1016/0006-8993(88)90883-9
   DAVIS EJ, 1994, BRAIN RES BULL, V34, P73, DOI 10.1016/0361-9230(94)90189-9
   DIJKSTRA CD, 1985, IMMUNOLOGY, V54, P589
   FUKUMOTO T, 1982, EUR J IMMUNOL, V12, P237, DOI 10.1002/eji.1830120313
   GALANOS C, 1993, IMMUNOBIOLOGY, V187, P346, DOI 10.1016/S0171-2985(11)80349-9
   GANTER S, 1992, J NEUROSCI RES, V33, P218, DOI 10.1002/jnr.490330205
   GEBICKEHAERTER PJ, 1989, J NEUROSCI, V9, P183
   GEHRMANN J, 1992, J CEREBR BLOOD F MET, V12, P257, DOI 10.1038/jcbfm.1992.36
   GEHRMANN J, 1992, LAB INVEST, V67, P100
   GIULIAN D, 1987, J NEUROSCI RES, V18, P155, DOI 10.1002/jnr.490180123
   GIULIAN D, 1990, ANN NEUROL, V27, P33, DOI 10.1002/ana.410270107
   GIULIAN D, 1993, STROKE, V24, pI84
   GRAEBER MB, 1988, J NEUROSCI RES, V21, P18, DOI 10.1002/jnr.490210104
   GRAEBER MB, 1989, J NEUROSCI RES, V22, P103, DOI 10.1002/jnr.490220114
   KENT S, 1992, TRENDS PHARMACOL SCI, V13, P24, DOI 10.1016/0165-6147(92)90012-U
   LEE SC, 1993, J IMMUNOL, V150, P2659
   LOUGHLIN AJ, 1992, IMMUNOLOGY, V75, P170
   MARTY S, 1991, NEUROSCIENCE, V45, P529, DOI 10.1016/0306-4522(91)90268-S
   MCMASTER WR, 1979, EUR J IMMUNOL, V9, P426, DOI 10.1002/eji.1830090603
   MONTEROMENEI CN, 1994, BRAIN RES, V653, P101, DOI 10.1016/0006-8993(94)90377-8
   MORIOKA T, 1993, J COMP NEUROL, V327, P123, DOI 10.1002/cne.903270110
   MORIOKA T, 1991, J CEREBR BLOOD F MET, V11, P966, DOI 10.1038/jcbfm.1991.162
   MORIOKA T, 1992, ACTA NEUROPATHOL, V83, P149, DOI 10.1007/BF00308474
   MUSTAFA MM, 1990, AM J DIS CHILD, V144, P883, DOI 10.1001/archpedi.1990.02150320047024
   PIANI D, 1992, EUR J IMMUNOL, V22, P2429, DOI 10.1002/eji.1830220936
   RANSOHOFF RM, 1992, J NEUROIMMUNOL, V37, P169, DOI 10.1016/0165-5728(92)90001-2
   ROBINSON AP, 1986, IMMUNOLOGY, V57, P239
   ROSENBAUM JT, 1980, NATURE, V286, P611, DOI 10.1038/286611a0
   SAUTER A, 1986, STROKE, V17, P1228, DOI 10.1161/01.STR.17.6.1228
   SELMAJ KW, 1988, ANN NEUROL, V23, P339, DOI 10.1002/ana.410230405
   STREIT WJ, 1989, EXP NEUROL, V105, P115, DOI 10.1016/0014-4886(89)90111-8
   STREIT WJ, 1987, J NEUROCYTOL, V16, P249, DOI 10.1007/BF01795308
   THOMAS L, 1954, ANNU REV PHYSIOL, V16, P467, DOI 10.1146/annurev.ph.16.030154.002343
   TILDERS FJH, 1994, PSYCHONEUROENDOCRINO, V19, P209, DOI 10.1016/0306-4530(94)90010-8
   TSUKADA N, 1991, J NEUROL SCI, V104, P230, DOI 10.1016/0022-510X(91)90315-X
   TUNKEL AR, 1991, IN VITRO CELL DEV B, V27, P113
   ULEVITCH RJ, 1994, CURR OPIN IMMUNOL, V6, P125, DOI 10.1016/0952-7915(94)90043-4
   VANDAM AM, 1995, NEUROSCIENCE, V65, P815, DOI 10.1016/0306-4522(94)00549-K
   VASS K, 1990, AM J PATHOL, V137, P789
   WATSON RWG, 1994, J LEUKOCYTE BIOL, V56, P95, DOI 10.1002/jlb.56.1.95
   WEBER G, 1993, J IMMUNOL, V151, P415
   Wong Mal-Li, 1994, Neuroimmunomodulation, V1, P181, DOI 10.1159/000097159
   XU J, 1994, EXP BRAIN RES, V100, P287
   ZAJICEK JP, 1992, BRAIN, V115, P1611, DOI 10.1093/brain/115.6.1611-a
NR 50
TC 120
Z9 121
U1 0
U2 11
PU PERGAMON-ELSEVIER SCIENCE LTD
PI OXFORD
PA THE BOULEVARD, LANGFORD LANE, KIDLINGTON, OXFORD OX5 1GB, ENGLAND
SN 0197-0186
EI 1872-9754
J9 NEUROCHEM INT
JI Neurochem. Int.
PD JUL
PY 1996
VL 29
IS 1
BP 25
EP 35
DI 10.1016/0197-0186(95)00141-7
PG 11
WC Biochemistry & Molecular Biology; Neurosciences
WE Science Citation Index Expanded (SCI-EXPANDED)
SC Biochemistry & Molecular Biology; Neurosciences & Neurology
GA UU407
UT WOS:A1996UU40700003
PM 8808786
DA 2023-08-27
ER

PT J
AU Thakur, JD
   Khan, IS
   Shorter, CD
   Sonig, A
   Gardner, GL
   Guthikonda, B
   Nanda, A
AF Thakur, Jai Deep
   Khan, Imad Saeed
   Shorter, Cedric D.
   Sonig, Ashish
   Gardner, Gale L.
   Guthikonda, Bharat
   Nanda, Anil
TI Do cystic vestibular schwannomas have worse surgical outcomes?
   Systematic analysis of the literature
SO NEUROSURGICAL FOCUS
LA English
DT Article
DE cystic vestibular schwannoma; acoustic neuroma; solid vestibular
   schwannoma
ID FACIAL-NERVE OUTCOMES; ACOUSTIC NEURINOMAS; RETROSIGMOID APPROACH;
   GROWTH-RATE; NEUROMAS; SURGERY; SERIES; TUMORS; MANAGEMENT
AB Object. The goal of this study was to perform a systematic quantitative comparison of the surgical outcomes between cystic vestibular schwannomas (CVSs) and solid vestibular schwannomas (SVSs).
   Methods. A review of English-language literature published between 1990 and 2011 was performed using various search engines including PubMed, Google Scholar, and the Cochrane database. Only studies that reported surgical results of CVSs in comparison with SVSs were included in the analysis. The primary end point of this study was surgical outcomes, defined by the following: 1) facial nerve outcomes at latest follow-up; 2) mortality rates; or 3) non-facial nerve complication index. Secondary end points included extent of resection and brainstem adherence.
   Results. Nine studies comprising 428 CVSs and 1287 SVSs were included in the study. The mean age of patients undergoing surgery was 48.3 +/- 6.75 and 47.1 +/- 9 years for CVSs and SVSs, respectively (p = 0.8). The mean tumor diameter for CVSs was 3.9 +/- 0.84 cm and that for SVSs was 3.7 +/- 1.2 cm (p = 0.7). There was no significant difference in the extent of resection among CVSs and SVSs (81.2% vs 80.7%, p = 0.87) Facial nerve outcomes were significantly better in the cohort of patients with SVSs than in those with CVSs (52.1% vs 39%, p = 0.0001). The perioperative mortality rates for CVSs and SVSs were not significantly different (3% and 3.8%, respectively; p = 0.6). No significant difference was noted between the cumulative non-facial nerve complication rate (including mortality) among patients with CVSs and SVSs (24.5% and 25.6%, respectively; p = 0.75)
   Conclusions. Facial nerve outcomes are worse in patients undergoing resection for CVSs than in patients undergoing resection for SVSs. There were no significant differences in the extent of resection or postoperative morbidity and mortality rates between the cohorts of patients with vestibular schwannomas. (http://thejns.org/doi/abs/10.3171/2012.6.FOCUS12200)
C1 [Thakur, Jai Deep; Khan, Imad Saeed; Shorter, Cedric D.; Sonig, Ashish; Guthikonda, Bharat; Nanda, Anil] Louisiana State Univ, Hlth Sci Ctr Shreveport, Dept Neurosurg, Shreveport, LA 71103 USA.
   [Gardner, Gale L.] Louisiana State Univ, Hlth Sci Ctr Shreveport, Dept Otolaryngol, Shreveport, LA 71103 USA.
C3 Louisiana State University System; Louisiana State University Health
   Sciences Center at Shreveport; Louisiana State University System;
   Louisiana State University Health Sciences Center at Shreveport
RP Nanda, A (通讯作者)，Louisiana State Univ, Hlth Sci Ctr Shreveport, Dept Neurosurg, 1501 Kings Highway, Shreveport, LA 71103 USA.
EM
RI Khan, Imad S/E-8958-2014
CR Benech F, 2005, NEUROSURG REV, V28, P209, DOI 10.1007/s10143-005-0380-y
   Benhaiem-Sigaux N, 1999, NEURORADIOLOGY, V41, P795, DOI 10.1007/s002340050844
   CHARABI S, 1994, ARCH OTOLARYNGOL, V120, P1333
   CHARABI S, 1994, LARYNGOSCOPE, V104, P1348
   CHARABI S, 1994, J LARYNGOL OTOL, V108, P375, DOI 10.1017/S0022215100126854
   Fundova P, 2000, J LARYNGOL OTOL, V114, P935
   Gal TJ, 2010, OTOLARYNG HEAD NECK, V142, P677, DOI 10.1016/j.otohns.2010.01.037
   Ishii K, 1996, Radiat Med, V14, P65
   Jian BJ, 2011, NEUROSURGERY, V68, P874, DOI 10.1227/NEU.0b013e318208f614
   Jones SEM, 2007, SKULL BASE-INTERD AP, V17, P281, DOI 10.1055/s-2007-986436
   Kameyama S, 1996, ACTA NEUROCHIR, V138, P695, DOI 10.1007/BF01411474
   KAMEYAMA S, 1994, ACTA NEUROCHIR, V129, P127, DOI 10.1007/BF01406491
   Kanzaki J, 2003, OTOL NEUROTOL, V24, P642, DOI 10.1097/00129492-200307000-00019
   KINGSLEY DPE, 1984, NEURORADIOLOGY, V26, P319
   LEE JP, 1989, NEUROSURGERY, V24, P764, DOI 10.1227/00006123-198905000-00020
   LEE KS, 1989, SURG NEUROL, V32, P427, DOI 10.1016/0090-3019(89)90005-0
   Lohle PNM, 1999, NEURORADIOLOGY, V41, P13, DOI 10.1007/s002340050696
   LUNARDI P, 1991, ACTA NEUROCHIR, V110, P120, DOI 10.1007/BF01400678
   MAHALEY MS, 1990, CLIN NEUR, V36, P347
   Mehrotra N, 2008, BRIT J NEUROSURG, V22, P550, DOI 10.1080/02688690802159031
   Moon KS, 2007, J NEUROSURG, V106, P866, DOI 10.3171/jns.2007.106.5.866
   Muzumdar DP, 2002, J CLIN NEUROSCI, V9, P453, DOI 10.1054/jocn.2001.1003
   Park CK, 2006, J NEUROSURG, V105, P576, DOI 10.3171/jns.2006.105.4.576
   Pendl G, 1996, STEREOT FUNCT NEUROS, V66, P103, DOI 10.1159/000099775
   Piccirillo E, 2009, OTOL NEUROTOL, V30, P826, DOI 10.1097/MAO.0b013e3181b04e18
   Samii M, 2006, J NEUROSURG, V105, P527, DOI 10.3171/jns.2006.105.4.527
   SCHOBER R, 1992, VIRCHOWS ARCH A, V420, P59, DOI 10.1007/BF01605985
   Shirato H, 2000, INT J RADIAT ONCOL, V48, P1395, DOI 10.1016/S0360-3016(00)00731-8
   Sinha S, 2008, J CLIN NEUROSCI, V15, P511, DOI 10.1016/j.jocn.2007.01.007
   Stipkovits EM, 2001, OTOL NEUROTOL, V22, P215, DOI 10.1097/00129492-200103000-00017
   Su WD, 2005, J CLIN NEUROSCI, V12, P253, DOI 10.1016/j.jocn.2004.03.040
   TALI ET, 1993, AM J NEURORADIOL, V14, P1241
   Yamakami I, 2004, J NEUROL NEUROSUR PS, V75, P453, DOI 10.1136/jnnp.2003.010827
NR 33
TC 27
Z9 29
U1 0
U2 5
PU AMER ASSOC NEUROLOGICAL SURGEONS
PI ROLLING MEADOWS
PA 5550 MEADOWBROOK DRIVE, ROLLING MEADOWS, IL 60008 USA
SN 1092-0684
J9 NEUROSURG FOCUS
JI Neurosurg. Focus
PD SEP
PY 2012
VL 33
IS 3
AR E12
DI 10.3171/2012.6.FOCUS12200
PG 8
WC Clinical Neurology; Surgery
WE Science Citation Index Expanded (SCI-EXPANDED)
SC Neurosciences & Neurology; Surgery
GA 003HD
UT WOS:000308599400013
PM 22937846
DA 2023-08-27
ER

PT J
AU Kanai, M
   Funakoshi, H
   Takahashi, H
   Hayakawa, T
   Mizuno, S
   Matsumoto, K
   Nakamura, T
AF Kanai, Masaaki
   Funakoshi, Hiroshi
   Takahashi, Hisaaki
   Hayakawa, Tomoko
   Mizuno, Shinya
   Matsumoto, Kunio
   Nakamura, Toshikazu
TI Tryptophan 2,3-dioxygenase is a key modulator of physiological
   neurogenesis and anxiety-related behavior in mice
SO MOLECULAR BRAIN
LA English
DT Article
AB Although nutrients, including amino acids and their metabolites such as serotonin (5-HT), are strong modulators of anxiety-related behavior, the metabolic pathway(s) responsible for this physiological modulation is not fully understood. Regarding tryptophan (Trp), the initial rate-limiting enzymes for the kynurenine pathway of tryptophan metabolism are tryptophan 2,3-dioxygenase (TDO) and indoleamine 2,3-dioxygenase (IDO). Here, we generated mice deficient for tdo (Tdo(-/-)). Compared with wild-type littermates, Tdo(-/-)mice showed increased plasma levels of Trp and its metabolites 5-hydroxyindoleacetic acid (5-HIAA) and kynurenine, as well as increased levels of Trp, 5-HT and 5-HIAA in the hippocampus and midbrain. These mice also showed anxiolytic modulation in the elevated plus maze and open field tests, and increased adult neurogenesis, as evidenced by double staining of BrdU and neural progenitor/neuronal markers. These findings demonstrate a direct molecular link between Trp metabolism and neurogenesis and anxiety-related behavior under physiological conditions.
C1 [Kanai, Masaaki; Funakoshi, Hiroshi; Takahashi, Hisaaki; Hayakawa, Tomoko; Mizuno, Shinya; Matsumoto, Kunio; Nakamura, Toshikazu] Osaka Univ, Grad Sch Med, Dept Biochem & Mol Biol, Div Mol Regenerat Med, Suita, Osaka 5650871, Japan.
   [Takahashi, Hisaaki] Ehime Univ, Sch Med, Dept Mol & Cellular Biol, Matsuyama, Ehime 790, Japan.
