## Supplemental Material 1 for "Bibliometric analysis of emerging trends and research foci in brainstem tumor field over thirty years (1992-2023)": savedrecs (11).txt

FN Clarivate Analytics Web of Science
VR 1.0
PT J
AU Regis, J
   Merly, L
   Balossier, A
   Baumstarck, K
   Hamdi, H
   Mariani, S
   Delsanti, C
   Vincent, M
   Nigoul, JM
   Beltaifa, Y
   Muracciole, X
AF Regis, Jean
   Merly, Louise
   Balossier, Anne
   Baumstarck, Karine
   Hamdi, Hussein
   Mariani, Sarah
   Delsanti, Christine
   Vincent, Marion
   Nigoul, Jean Marc
   Beltaifa, Yassin
   Muracciole, Xavier
TI Mask-Based versus Frame-Based Gamma Knife ICON Radiosurgery in Brain
   Metastases: A Prospective Randomized Trial
SO STEREOTACTIC AND FUNCTIONAL NEUROSURGERY
LA English
DT Article
DE Comfort; Pain; Radiosurgery; Cone beam computed tomography
ID INTRACRANIAL MOTION; ACCURACY; SYSTEM; FIXATION
AB Background: Radiosurgery is performed with a diversity of instruments relying usually either on a stereotactic frame or a mask for patient head fixation. Comfort and safety efficacy of the 2 systems have never been rigorously evaluated and compared. Material and Method: Between February 2016 and January 2017, 58 patients presenting with nonsmall cell lung cancer brain metastases have been treated by Gamma Knife radiosurgery (GKS) with random use of a frame or a mask for fixation were included patients older than 18, with <5 brain metastases (at the exclusion of brainstem and optic pathway's locations) and no earlier history of radiotherapy. The primary outcome measure was the pain scale assessment (PSA) at the beginning of the GKS procedure. Results: The PSA at the beginning of the GKS procedure was not different between the 2 groups. The PSA at the day before GKS, before magnetic resonance imaging, just after frame application, and the day after radiosurgery (departure) has shown no difference between the 2 groups. At the end of the radiosurgery itself (just after frame or mask removal) and 1 h after, the mean pain scale was higher in patients treated with the frame (p < 0.05 and p < 0.001, respectively) but 2 patients were not able to tolerate the mask discomfort and had to be treated with frame. Tumor control and morbidity probability were demonstrated to be no difference between the 2 groups in this population of patients with BM not in highly functional area. The median of the extra dose to the body due to the cone-beam computed tomography was 7.5 mGy with a maximum of 35 mGy in patients treated with a mask fixation (null in the others treated with frame). Mask fixation was associated to longer treatment time although the beam on time was not different between the 2 groups. Conclusion: In selected patients, with brain oligo-metastases out of critical location, single-dose mask-based GKS can be done with a comfort and a safety efficacy comparable to frame-based GKS. There seems to be no clear patient data that confirm the value of the mask system with regards to comfort.
C1 [Regis, Jean; Merly, Louise; Balossier, Anne; Hamdi, Hussein; Mariani, Sarah; Delsanti, Christine; Vincent, Marion; Nigoul, Jean Marc; Beltaifa, Yassin] Aix Marseille Univ, INSERM, INS, Inst Neurosci Syst, Marseille, France.
   [Regis, Jean; Merly, Louise; Balossier, Anne; Hamdi, Hussein; Mariani, Sarah; Delsanti, Christine; Vincent, Marion; Nigoul, Jean Marc; Beltaifa, Yassin] Hop dAdulte Timone, Dept Funct Neurosurg, Marseille, France.
   [Baumstarck, Karine] Aix Marseille Univ, Dept Biostatist, Marseille, France.
   [Muracciole, Xavier] Aix Marseille Univ, Dept Radiotherapy, Marseille, France.
C3 Institut National de la Sante et de la Recherche Medicale (Inserm);
   UDICE-French Research Universities; Aix-Marseille Universite; Assistance
   Publique-Hopitaux de Marseille; UDICE-French Research Universities;
   Aix-Marseille Universite; Assistance Publique-Hopitaux de Marseille;
   UDICE-French Research Universities; Aix-Marseille Universite;
   UDICE-French Research Universities; Aix-Marseille Universite
RP Regis, J (通讯作者)，Aix Marseille Univ, INSERM, INS, Inst Neurosci Syst, Marseille, France.; Regis, J (通讯作者)，Hop dAdulte Timone, Dept Funct Neurosurg, Marseille, France.
EM
OI Hamdi, Hussein/0000-0001-6174-3471
FU Assistance Publique des Hopitaux de Marseille
FX The study was sponsored by our hospital (Assistance Publique des
   Hopitaux de Marseille). This sponsoring was covering the insurances of
   the trial.
CR Adler JR, 1997, STEREOT FUNCT NEUROS, V69, P124, DOI 10.1159/000099863
   AlDahlawi I, 2017, J APPL CLIN MED PHYS, V18, P67, DOI 10.1002/acm2.12073
   Carminucci A, 2018, J NEUROSURG, V129, P133, DOI 10.3171/2018.7.GKS181516
   Chung HT, 2018, J APPL CLIN MED PHYS, V19, P148, DOI 10.1002/acm2.12365
   Chung HT, 2018, PLOS ONE, V13, DOI 10.1371/journal.pone.0193809
   Gevaert T, 2013, RADIOTHER ONCOL, V106, P192, DOI 10.1016/j.radonc.2012.07.002
   Gevaert T, 2012, INT J RADIAT ONCOL, V82, P1627, DOI 10.1016/j.ijrobp.2011.01.052
   Grimm MA, 2020, RADIOTHER ONCOL, V147, P195, DOI 10.1016/j.radonc.2020.05.011
   Kim T, 2016, J APPL CLIN MED PHYS, V17, P487, DOI 10.1120/jacmp.v17i2.5851
   Knutson NC, 2019, J APPL CLIN MED PHYS, V20, P21, DOI 10.1002/acm2.12573
   LEKSELL D G, 1987, Neurological Research, V9, P60
   LEKSELL L, 1987, J NEUROSURG, V66, P626, DOI 10.3171/jns.1987.66.4.0626
   Ma LJ, 2014, TECHNOL CANCER RES T, V13, P403, DOI [10.7785/tcrt.2013.600259, 10.7785/tcrtexpress.2013.600259]
   MacDonald RL, 2020, INT J RADIAT ONCOL, V106, P413, DOI 10.1016/j.ijrobp.2019.10.030
   Nix M, 2019, J RADIOSURGERY SBRT, V6, P121
   Podgorsak MB., 2018, J APPL CLIN MED PHYS, V3, P12417
   Ramakrishna N, 2010, RADIOTHER ONCOL, V95, P109, DOI 10.1016/j.radonc.2009.12.030
   Regis Jean, 2008, Prog Neurol Surg, V21, P54, DOI 10.1159/000156594
   Regis J, 2009, NEUROSURGERY, V64, P346, DOI 10.1227/01.NEU.0000337578.00814.75
   Reiner B, 2016, J RADIOSURGERY SBRT, V4, P275
   Sarfehnia A, 2018, MED PHYS, V45, P4179, DOI 10.1002/mp.13073
   Schlesinger D, 2012, J NEUROSURG, V117, P217, DOI 10.3171/2012.6.GKS12989
   Seneviratne DS, 2020, J RADIOSURGERY SBRT, V6, P277
   Stieler F, 2018, PHYS MEDICA, V52, P93, DOI 10.1016/j.ejmp.2018.06.640
   Thorne FM, 2009, PAIN, V144, P262, DOI 10.1016/j.pain.2009.04.004
   Wright G, 2016, J RADIOSURGERY SBRT, V4, P289
   Xu AY, 2019, MED PHYS, V46, P4333, DOI 10.1002/mp.13740
   Zeverino M, 2017, MED PHYS, V44, P355, DOI 10.1002/mp.12052
NR 28
TC 4
Z9 4
U1 0
U2 0
PU KARGER
PI BASEL
PA ALLSCHWILERSTRASSE 10, CH-4009 BASEL, SWITZERLAND
SN 1011-6125
EI 1423-0372
J9 STEREOT FUNCT NEUROS
JI Stereotact. Funct. Neurosurg.
PD MAR
PY 2022
VL 100
IS 2
BP 86
EP 94
DI 10.1159/000519280
EA DEC 2021
PG 9
WC Neurosciences; Neuroimaging; Surgery
WE Science Citation Index Expanded (SCI-EXPANDED)
SC Neurosciences & Neurology; Surgery
GA 0H8ZH
UT WOS:000733189100001
PM 34933308
DA 2023-08-27
ER

PT J
AU Uldrich, AP
   Berzins, SP
   Malin, MA
   Bouillet, P
   Strasser, A
   Smyth, MJ
   Boyd, RL
   Godfrey, DI
AF Uldrich, AP
   Berzins, SP
   Malin, MA
   Bouillet, P
   Strasser, A
   Smyth, MJ
   Boyd, RL
   Godfrey, DI
TI Antigen challenge inhibits thymic emigration
SO JOURNAL OF IMMUNOLOGY
LA English
DT Article
ID REGULATORY T-CELLS; SPHINGOSINE 1-PHOSPHATE RECEPTOR;
   STAPHYLOCOCCAL-ENTEROTOXIN-B; TUMOR-NECROSIS-FACTOR; FAMILY MEMBER BIM;
   IN-VIVO; NEGATIVE SELECTION; THYMOCYTE EMIGRATION; BACTERIAL
   SUPERANTIGENS; POSITIVE SELECTION
AB T cell development in the thymus involves a series of TCR-mediated control points including TCR-beta selection and positive and negative selection. Approximately half of the thymic sojourn is spent in the medulla, where thymocytes undergo final maturation before emigrating to the periphery. Although it is acknowledged that thymic emigration is an active process, relatively little is known about how this is regulated, why it takes so long, and whether TCR-mediated signaling can influence this step. Using wild-type and TCR transgenic mice, we found that Ag injected i.v. or intrathymically led to a striking reduction in the number of recent thymic emigrants (RTE) in the periphery. This was caused by inhibition of T cell export rather than peripheral deletion, because a cohort of RTE that was already released before in vivo Ag challenge was not depleted, and similar results were observed in Bim-deficient mice, which have impaired T cell deletion. Within the thymus, the loss of RTE was associated with retention of medullary thymocytes rather than increased negative selection. In addition to Ag-specific inhibition of export, some TCR-independent suppression of emigration was also observed that appeared to be partly the result of the inflammatory cytokine TNF. Thus, in addition to its accepted role in intrathymic selection events, TCR signaling can also play an important role in the regulation of thymic emigration.
C1 Univ Melbourne, Dept Microbiol & Immunol, Parkville, Vic 3052, Australia.
   Monash Univ, Monash Immunol & Stem Cell Labs, Sci Technol Res & Innovat Project, Clayton, Vic 3168, Australia.
   Walter & Eliza Hall Inst Med Res, Parkville, Vic, Australia.
C3 University of Melbourne; Monash University; Walter & Eliza Hall
   Institute
EM
RI Uldrich, Adam/AAX-3785-2020; Smyth, Mark J/H-8709-2014; Strasser,
   Andreas/C-7581-2013; Bouillet, Philippe/E-1241-2013
OI Smyth, Mark J/0000-0001-7098-7240; Strasser,
   Andreas/0000-0002-5020-4891; Bouillet, Philippe/0000-0002-9012-6058;
   ULDRICH, ADAM/0000-0002-6350-5976; Godfrey, Dale/0000-0002-3009-5472
CR Allende ML, 2004, J BIOL CHEM, V279, P15396, DOI 10.1074/jbc.M314291200
   ALTEVOGT P, 1994, EUR J IMMUNOL, V24, P731, DOI 10.1002/eji.1830240336
   Apostolou I, 2002, NAT IMMUNOL, V3, P756, DOI 10.1038/ni816
   BARNDEN MJ, 1994, EUR J IMMUNOL, V24, P2452, DOI 10.1002/eji.1830241029
   Berzins SP, 1998, J EXP MED, V187, P1839, DOI 10.1084/jem.187.11.1839
   Bouillet P, 1999, SCIENCE, V286, P1735, DOI 10.1126/science.286.5445.1735
   Bouillet P, 2002, NATURE, V415, P922, DOI 10.1038/415922a
   Boursalian TE, 2004, NAT IMMUNOL, V5, P418, DOI 10.1038/ni1049
   Cyster JG, 2002, J CLIN INVEST, V109, P1011, DOI 10.1172/JCI200215511
   EGERTON M, 1990, P NATL ACAD SCI USA, V87, P2579, DOI 10.1073/pnas.87.7.2579
   Feng CG, 2002, INT IMMUNOL, V14, P535, DOI 10.1093/intimm/dxf020
   Fontenot JD, 2017, J IMMUNOL, V198, P986, DOI 10.1038/ni904
   Gabor MJ, 1997, EUR J IMMUNOL, V27, P2010, DOI 10.1002/eji.1830270827
   Goettelfinger P, 2000, TRANSPL IMMUNOL, V8, P39, DOI 10.1016/S0966-3274(00)00003-4
   GONZALO JA, 1995, INT IMMUNOL, V7, P1691, DOI 10.1093/intimm/7.10.1691
   Graeler M, 2002, FASEB J, V16, P1874, DOI 10.1096/fj.02-0548com
   Grech A P, 2000, Dev Immunol, V8, P61, DOI 10.1155/2000/13492
   HERRMANN T, 1992, EUR J IMMUNOL, V22, P1935, DOI 10.1002/eji.1830220739
   Herrmann T, 1993, Semin Immunol, V5, P33, DOI 10.1006/smim.1993.1005
   Hildeman DA, 2002, IMMUNITY, V16, P759, DOI 10.1016/S1074-7613(02)00322-9
   HOGQUIST KA, 1994, CELL, V76, P17, DOI 10.1016/0092-8674(94)90169-4
   Irving BA, 1998, SCIENCE, V280, P905, DOI 10.1126/science.280.5365.905
   Jordan MS, 2001, NAT IMMUNOL, V2, P301, DOI 10.1038/86302
   Kanegane H, 1996, BLOOD, V88, P230
   KELLY KA, 1990, INT IMMUNOL, V2, P419, DOI 10.1093/intimm/2.5.419
   KELLY KA, 1991, EUR J IMMUNOL, V21, P43, DOI 10.1002/eji.1830210108
   Kishimoto H, 1997, J EXP MED, V185, P263, DOI 10.1084/jem.185.2.263
   Korner H, 1997, EUR J IMMUNOL, V27, P2600, DOI 10.1002/eji.1830271020
   Le Campion A, 2002, J IMMUNOL, V168, P1664, DOI 10.4049/jimmunol.168.4.1664
   Le Campion A, 2000, EUR J IMMUNOL, V30, P738
   Lee CK, 2001, BLOOD, V97, P1360, DOI 10.1182/blood.V97.5.1360
   Li M, 2001, J IMMUNOL, V166, P6099, DOI 10.4049/jimmunol.166.10.6099
   LUCAS B, 1994, J IMMUNOL, V153, P53
   MACDONALD HR, 1993, IMMUNOL REV, V133, P105, DOI 10.1111/j.1600-065X.1993.tb01512.x
   Martin S, 1997, EUR J IMMUNOL, V27, P2726, DOI 10.1002/eji.1830271037
   Matloubian M, 2004, NATURE, V427, P355, DOI 10.1038/nature02284
   Michie AM, 2002, SEMIN IMMUNOL, V14, P311, DOI 10.1016/S1044-5323(02)00064-7
   MIETHKE T, 1992, J EXP MED, V175, P91, DOI 10.1084/jem.175.1.91
   Mintern JD, 2004, J IMMUNOL, V172, P699, DOI 10.4049/jimmunol.172.1.699
   Nakayama T, 2002, J IMMUNOL, V168, P87, DOI 10.4049/jimmunol.168.1.87
   Palmer E, 2003, NAT REV IMMUNOL, V3, P383, DOI 10.1038/nri1085
   Pellegrini M, 2003, P NATL ACAD SCI USA, V100, P14175, DOI 10.1073/pnas.2336198100
   Penit C, 1997, J IMMUNOL, V159, P4848
   Poznansky MC, 2002, J CLIN INVEST, V109, P1101, DOI 10.1172/JCI200213853
   Renno T, 1999, J IMMUNOL, V162, P6312
   Rosen H, 2003, P NATL ACAD SCI USA, V100, P10907, DOI 10.1073/pnas.1832725100
   SCOLLAY R, 1984, AM J ANAT, V170, P331, DOI 10.1002/aja.1001700308
   SCOLLAY R, 1995, IMMUNOL TODAY, V16, P268, DOI 10.1016/0167-5699(95)80179-0
   SCOLLAY R, 1984, J IMMUNOL, V132, P25
   SCOLLAY RG, 1980, EUR J IMMUNOL, V10, P210, DOI 10.1002/eji.1830100310
   Starr TK, 2003, ANNU REV IMMUNOL, V21, P139, DOI 10.1146/annurev.immunol.21.120601.141107
   Tough DF, 1997, J EXP MED, V185, P2089, DOI 10.1084/jem.185.12.2089
   Ueno T, 2002, IMMUNITY, V16, P205, DOI 10.1016/S1074-7613(02)00267-4
   Vianello F, 2005, J IMMUNOL, V175, P5115, DOI 10.4049/jimmunol.175.8.5115
   Villunger A, 2004, P NATL ACAD SCI USA, V101, P7052, DOI 10.1073/pnas.0305757101
NR 55
TC 12
Z9 13
U1 0
U2 1
PU AMER ASSOC IMMUNOLOGISTS
PI BETHESDA
PA 9650 ROCKVILLE PIKE, BETHESDA, MD 20814 USA
SN 0022-1767
J9 J IMMUNOL
JI J. Immunol.
PD APR 15
PY 2006
VL 176
IS 8
BP 4553
EP 4561
DI 10.4049/jimmunol.176.8.4553
PG 9
WC Immunology
WE Science Citation Index Expanded (SCI-EXPANDED)
SC Immunology
GA 059YV
UT WOS:000238769000011
PM 16585545
OA Bronze
DA 2023-08-27
ER

PT J
AU Pronto-Laborinho, AC
   Pinto, S
   de Carvalho, M
AF Pronto-Laborinho, Ana Catarina
   Pinto, Susana
   de Carvalho, Mamede
TI Roles of Vascular Endothelial Growth Factor in Amyotrophic Lateral
   Sclerosis
SO BIOMED RESEARCH INTERNATIONAL
LA English
DT Review
ID HYPOXIA-RESPONSE ELEMENT; NEURAL STEM-CELLS; STIMULATES AXONAL
   OUTGROWTH; UBIQUITIN-PROTEASOME SYSTEM; LINDAU TUMOR-SUPPRESSOR;
   TRANSGENIC MOUSE MODEL; FACTOR MESSENGER-RNA; FACTOR VEGF; FACTOR GENE;
   IN-VIVO
AB Amyotrophic lateral sclerosis (ALS) is a fatal devastating neurodegenerative disorder, involving progressive degeneration of motor neurons in spinal cord, brainstem, and motor cortex. Riluzole is the only drug approved in ALS but it only confers a modest improvement in survival. In spite of a high number of clinical trials no other drug has proved effectiveness. Recent studies support that vascular endothelial growth factor (VEGF), originally described as a key angiogenic factor, also plays a key role in the nervous system, including neurogenesis, neuronal survival, neuronal migration, and axon guidance. VEGF has been used in exploratory clinical studies with promising results in ALS and other neurological disorders. Although VEGF is a very promising compound, translating the basic science breakthroughs into clinical practice is the major challenge ahead. VEGF-B, presenting a single safety profile, protects motor neurons from degeneration in ALS animal models and, therefore, it will be particularly interesting to test its effects in ALS patients. In the present paper the authors make a brief description of the molecular properties of VEGF and its receptors and review its different features and therapeutic potential in the nervous system/neurodegenerative disease, particularly in ALS.
C1 [Pronto-Laborinho, Ana Catarina; Pinto, Susana; de Carvalho, Mamede] Univ Lisbon, Fac Med, Inst Physiol, P-1649028 Lisbon, Portugal.
   [Pronto-Laborinho, Ana Catarina; Pinto, Susana; de Carvalho, Mamede] IMM, Translat Clin Physiol Unit, P-1649028 Lisbon, Portugal.
   [de Carvalho, Mamede] Ctr Hos Lisboa Norte, Hosp Santa Maria, Dept Neurosci, P-1649028 Lisbon, Portugal.
C3 Universidade de Lisboa; Universidade de Lisboa; Hospital Santa Maria
RP de Carvalho, M (通讯作者)，Univ Lisbon, Fac Med, Inst Physiol, Ave Prof Egas Moniz, P-1649028 Lisbon, Portugal.
EM
RI de Carvalho, Mamede/I-6295-2019; Pinto, Susana C/K-9435-2014; Pinto,
   Susana/L-8394-2019; Pronto-Laborinho, Ana Catarina/ABE-8944-2021
OI de Carvalho, Mamede/0000-0001-7556-0158; Pinto, Susana
   C/0000-0002-0727-5897; Pinto, Susana/0000-0002-0727-5897; Pronto
   Laborinho, Ana Catarina/0000-0002-2816-9698
CR Aase K, 1999, DEV DYNAM, V215, P12, DOI 10.1002/(SICI)1097-0177(199905)215:1<12::AID-DVDY3>3.3.CO;2-E
   Abrous DN, 2005, PHYSIOL REV, V85, P523, DOI 10.1152/physrev.00055.2003
   Achen MG, 2012, GROWTH FACTORS, V30, P283, DOI 10.3109/08977194.2012.704917
   Achen MG, 1998, P NATL ACAD SCI USA, V95, P548, DOI 10.1073/pnas.95.2.548
   Achen MG, 2005, CANCER CELL, V7, P121, DOI 10.1016/j.ccr.2005.01.017
   Alitalo K, 2002, CANCER CELL, V1, P219, DOI 10.1016/S1535-6108(02)00051-X
   Andersen PM, 2011, NAT REV NEUROL, V7, P603, DOI 10.1038/nrneurol.2011.150
   Andrews J, 2009, CURR NEUROL NEUROSCI, V9, P59, DOI 10.1007/s11910-009-0010-0
   Autiero M, 2003, NAT MED, V9, P936, DOI 10.1038/nm884
   Avakyan GN, 2002, ZH NEVROPATOL PSIKH, V102, P22
   Azimi-Nezhad M, 2012, BIOPRESERV BIOBANK, V10, P454, DOI 10.1089/bio.2012.0016
   Azzouz M, 2004, NATURE, V429, P413, DOI 10.1038/nature02544
   Bagri A, 2002, ADV EXP MED BIOL, V515, P13
   Bearden SE, 2004, MICROCIRCULATION, V11, P633, DOI 10.1080/10739680490517659
   Beazley-Long N, 2013, AM J PATHOL, V183, P918, DOI 10.1016/j.ajpath.2013.05.031
   Beck H, 2002, J NEUROPATH EXP NEUR, V61, P339, DOI 10.1093/jnen/61.4.339
   Bellon A, 2010, NEURON, V66, P205, DOI 10.1016/j.neuron.2010.04.006
   BENAV P, 1995, FEBS LETT, V372, P83, DOI 10.1016/0014-5793(95)00956-A
   Bendotti C, 2012, PROG NEUROBIOL, V97, P101, DOI 10.1016/j.pneurobio.2011.10.001
   Biselli-Chicote PM, 2012, J CANCER RES CLIN, V138, P363, DOI 10.1007/s00432-011-1073-2
   Bogaert E, 2006, MUSCLE NERVE, V34, P391, DOI 10.1002/mus.20609
   Bogaert E, 2010, NEUROBIOL AGING, V31, P2185, DOI 10.1016/j.neurobiolaging.2008.12.007
   Breen EC, 1996, J APPL PHYSIOL, V81, P355, DOI 10.1152/jappl.1996.81.1.355
   Brockington A, 2005, NEUROGENETICS, V6, P101, DOI 10.1007/s10048-004-0201-4
   Brockington A, 2006, J NEUROPATH EXP NEUR, V65, P26, DOI 10.1097/01.jnen.0000196134.51217.74
   Brockington A, 2007, BMC MED GENET, V8, DOI 10.1186/1471-2350-8-23
   Bruijn LI, 2004, ANNU REV NEUROSCI, V27, P723, DOI 10.1146/annurev.neuro.27.070203.144244
   Calvo CF, 2011, GENE DEV, V25, P831, DOI 10.1101/gad.615311
   Cao GD, 2002, J CEREBR BLOOD F MET, V22, P534, DOI 10.1097/00004647-200205000-00005
   Cao L, 2004, NAT GENET, V36, P827, DOI 10.1038/ng1395
   Carilho R, 2014, MUSCLE NERVE, V49, P545, DOI 10.1002/mus.23955
   Carmeliet P, 2013, CELL MOL LIFE SCI, V70, P1763, DOI 10.1007/s00018-013-1283-7
   Cebe-Suarez S, 2006, CELL MOL LIFE SCI, V63, P601, DOI 10.1007/s00018-005-5426-3
   Chen W, 2006, NEUROLOGY, V67, P508, DOI 10.1212/01.wnl.0000227926.42370.04
   Chen YZ, 2004, AM J HUM GENET, V74, P1128, DOI 10.1086/421054
   Cleveland DW, 2001, NAT REV NEUROSCI, V2, P806, DOI 10.1038/35097565
   Cohen T, 1996, J BIOL CHEM, V271, P736, DOI 10.1074/jbc.271.2.736
   Constantin G, 2009, STEM CELLS, V27, P2624, DOI 10.1002/stem.194
   Corti S, 2010, HUM MOL GENET, V19, P3782, DOI 10.1093/hmg/ddq293
   Costa J, 2010, CNS NEUROL DISORD-DR, V9, P764, DOI 10.2174/187152710793237502
   Da Costa MMJ, 2014, DIS MODEL MECH, V7, P73, DOI 10.1242/dmm.012013
   Dachs GU, 2000, EUR J CANCER, V36, P1649, DOI 10.1016/S0959-8049(00)00159-3
   Dantz D, 2002, J CLIN ENDOCR METAB, V87, P835, DOI 10.1210/jc.87.2.835
   Daroff R., 2012, BRADLEYS NEUROLOGY C, P1855
   de Almodovar CR, 2011, NEURON, V70, P966, DOI 10.1016/j.neuron.2011.04.014
   De Almodovar CR, 2009, PHYSIOL REV, V89, P607, DOI 10.1152/physrev.00031.2008
   De Carvalho M, 2005, AMYOTROPH LATERAL SC, V6, P202, DOI 10.1080/14660820510011997
   de Carvalho M, 2005, NEUROLOGY, V64, P1783, DOI 10.1212/01.WNL.0000162036.76024.AB
   De Falco S, 2002, TRENDS CARDIOVAS MED, V12, P241, DOI 10.1016/S1050-1738(02)00168-8
   DeJesus-Hernandez M, 2011, NEURON, V72, P245, DOI 10.1016/j.neuron.2011.09.011
   Del Bo R, 2008, NEUROBIOL AGING, V29, P314, DOI 10.1016/j.neurobiolaging.2006.10.008
   Deroanne CF, 1997, CANCER RES, V57, P5590
   Devos D, 2004, NEUROLOGY, V62, P2127, DOI 10.1212/01.WNL.0000129913.44351.A3
   Dewil M, 2007, NEUROPATH APPL NEURO, V33, P499, DOI 10.1111/j.1365-2990.2007.00850.x
   Dobrogowska DH, 1998, J NEUROCYTOL, V27, P163, DOI 10.1023/A:1006907608230
   Dodge JC, 2010, MOL THER, V18, P2075, DOI 10.1038/mt.2010.206
   Du HA, 2010, BRAIN RES, V1357, P1, DOI 10.1016/j.brainres.2010.07.015
   Dumont DJ, 1998, SCIENCE, V282, P946, DOI 10.1126/science.282.5390.946
   Dvorak HF, 2002, J CLIN ONCOL, V20, P4368, DOI 10.1200/JCO.2002.10.088
   DVORAK HF, 1995, AM J PATHOL, V146, P1029
   Enholm B, 1997, ONCOGENE, V14, P2475, DOI 10.1038/sj.onc.1201090
   Erskine L, 2011, NEURON, V70, P951, DOI 10.1016/j.neuron.2011.02.052
   Falk T, 2010, INT J MOL SCI, V11, P2875, DOI 10.3390/ijms11082875
   Fernandez-Santiago R, 2006, NEUROLOGY, V66, P1929, DOI 10.1212/01.wnl.0000219756.71928.25
   Ferraiuolo L, 2013, MOL THER, V21, P503, DOI 10.1038/mt.2013.24
   Ferraiuolo L, 2011, NAT REV NEUROL, V7, P616, DOI 10.1038/nrneurol.2011.152
   Ferrara N, 2004, ENDOCR REV, V25, P581, DOI 10.1210/er.2003-0027
   Ferrara N, 2004, ONCOLOGIST, V9, P2, DOI 10.1634/theoncologist.9-suppl_1-2
   Ferrara N, 2003, NAT MED, V9, P669, DOI 10.1038/nm0603-669
   Ferrara N, 1997, ENDOCR REV, V18, P4, DOI 10.1210/er.18.1.4
   Ferrara N, 2004, MED INTELL UNIT, P1
   Ferrara N, 1999, NAT MED, V5, P1359, DOI 10.1038/70928
   Finkenzeller G, 1997, ONCOGENE, V15, P669, DOI 10.1038/sj.onc.1201219
   Foehring D, 2012, CELLS TISSUES ORGANS, V196, P195, DOI 10.1159/000334600
   Forstreuter F, 2002, J NEUROIMMUNOL, V132, P93, DOI 10.1016/S0165-5728(02)00315-6
   Forsythe JA, 1996, MOL CELL BIOL, V16, P4604
   FRANK S, 1995, J BIOL CHEM, V270, P12607, DOI 10.1074/jbc.270.21.12607
   Gerber HP, 1997, J BIOL CHEM, V272, P23659, DOI 10.1074/jbc.272.38.23659
   Geretti E, 2008, ANGIOGENESIS, V11, P31, DOI 10.1007/s10456-008-9097-1
   Gille H, 2001, J BIOL CHEM, V276, P3222, DOI 10.1074/jbc.M002016200
   Gora-Kupilas K, 2005, FOLIA NEUROPATHOL, V43, P31
   Graham SH, 2001, J CEREBR BLOOD F MET, V21, P99, DOI 10.1097/00004647-200102000-00001
   Greenberg DA, 2005, NATURE, V438, P954, DOI 10.1038/nature04481
   Greenberg DA, 2004, TRENDS MOL MED, V10, P1, DOI 10.1016/j.molmed.2003.11.006
   Greenway MJ, 2006, NAT GENET, V38, P411, DOI 10.1038/ng1742
   Greenway MJ, 2004, NEUROLOGY, V63, P1936, DOI 10.1212/01.WNL.0000144344.39103.F6
   Grimmond S, 1996, GENOME RES, V6, P124, DOI 10.1101/gr.6.2.124
   Gros-Louis F, 2003, MUSCLE NERVE, V28, P774, DOI 10.1002/mus.10498
   GRUGEL S, 1995, J BIOL CHEM, V270, P25915, DOI 10.1074/jbc.270.43.25915
   Gu R, 2010, BIOCHEM BIOPH RES CO, V393, P481, DOI 10.1016/j.bbrc.2010.02.029
   Gupta PK, 2011, J NEUROINFLAMM, V8, DOI 10.1186/1742-2094-8-97
   Hadano S, 2001, NAT GENET, V29, P166, DOI 10.1038/ng1001-166
   Hamada K, 2000, BLOOD, V96, P3793, DOI 10.1182/blood.V96.12.3793.h8003793_3793_3800
   HARADA S, 1994, J CLIN INVEST, V93, P2490, DOI 10.1172/JCI117258
   Hardiman O, 2011, NAT REV NEUROL, V7, P639, DOI 10.1038/nrneurol.2011.153
   Harrigan MR, 2003, ACTA NEUROCHIR, V145, P49, DOI 10.1007/s00701-002-1035-1
   Hashimoto T, 2006, DEVELOPMENT, V133, P2201, DOI 10.1242/dev.02385
   Herran E, 2013, J CONTROL RELEASE, V170, P111, DOI 10.1016/j.jconrel.2013.04.028
   Hetland ML, 2008, DIS MARKERS, V24, P1
   Hiratsuka S, 1998, P NATL ACAD SCI USA, V95, P9349, DOI 10.1073/pnas.95.16.9349
   Ho QT, 2007, INT J BIOCHEM CELL B, V39, P1349, DOI 10.1016/j.biocel.2007.04.010
   Hoeben A, 2004, PHARMACOL REV, V56, P549, DOI 10.1124/pr.56.4.3
   Hormbrey E, 2002, CLIN EXP METASTAS, V19, P651, DOI 10.1023/A:1021379811308
   HOUCK KA, 1991, MOL ENDOCRINOL, V5, P1806, DOI 10.1210/mend-5-12-1806
   Hwang DH, 2009, GENE THER, V16, P1234, DOI 10.1038/gt.2009.80
   Iliopoulos O, 1996, P NATL ACAD SCI USA, V93, P10595, DOI 10.1073/pnas.93.20.10595
   Ilzecka J, 2004, CLIN NEUROL NEUROSUR, V106, P289, DOI 10.1016/j.clineuro.2003.12.007
   Ingre C, 2013, AMYOTROPH LAT SCL FR, V14, P620, DOI 10.3109/21678421.2013.822515
   Ivan M, 2001, SCIENCE, V292, P464, DOI 10.1126/science.1059817
   Jeltsch M, 1997, SCIENCE, V276, P1423, DOI 10.1126/science.276.5317.1423
   Jin KL, 2000, NEUROSCIENCE, V100, P713, DOI 10.1016/S0306-4522(00)00331-6
   Jin KL, 2000, J MOL NEUROSCI, V14, P197, DOI 10.1385/JMN:14:3:197
   Jin KL, 2002, P NATL ACAD SCI USA, V99, P11946, DOI 10.1073/pnas.182296499
   Jin KL, 2000, P NATL ACAD SCI USA, V97, P10242, DOI 10.1073/pnas.97.18.10242
   Jin KL, 2001, P NATL ACAD SCI USA, V98, P4710, DOI 10.1073/pnas.081011098
   Joukov V, 1996, EMBO J, V15, P290
   Jung KH, 2006, BRAIN RES, V1073, P190, DOI 10.1016/j.brainres.2005.12.037
   Junqueira de Azevedo ILM, 2001, J BIOL CHEM, V276, P39836
   Just N, 2007, NEUROMUSCULAR DISORD, V17, P169, DOI 10.1016/j.nmd.2006.10.004
   KAIPAINEN A, 1995, P NATL ACAD SCI USA, V92, P3566, DOI 10.1073/pnas.92.8.3566
   Kawasaki T, 1999, DEVELOPMENT, V126, P4895
   Keifer OP, 2014, PHARMACOL THERAPEUT, V141, P261, DOI 10.1016/j.pharmthera.2013.10.009
   KENDALL RL, 1993, P NATL ACAD SCI USA, V90, P10705, DOI 10.1073/pnas.90.22.10705
   Kim BW, 2008, CELL SIGNAL, V20, P714, DOI 10.1016/j.cellsig.2007.12.009
   Kitsukawa T, 1995, DEVELOPMENT, V121, P4309
   Klagsbrun M, 2002, ADV EXP MED BIOL, V515, P33
   Kliem MA, 2011, AMYOTROPH LATERAL SC, V12, P331, DOI 10.3109/17482968.2011.574142
   Koch S, 2012, CSH PERSPECT MED, V2, DOI 10.1101/cshperspect.a006502
   Kolodkin AL, 1997, CELL, V90, P753, DOI 10.1016/S0092-8674(00)80535-8
   Koppers M, 2011, AMYOTROPHIC LATERAL SCLEROSIS, P479
   Krakora D, 2013, MOL THER, V21, P1602, DOI 10.1038/mt.2013.108
   Kranich S, 2009, NEUROCHEM INT, V55, P747, DOI 10.1016/j.neuint.2009.07.007
   KRANTZ SB, 1991, BLOOD, V77, P419
   Kwiatkowski TJ, 2009, SCIENCE, V323, P1205, DOI 10.1126/science.1166066
   Lambrechts D, 2003, NAT GENET, V34, P383, DOI 10.1038/ng1211
   Lambrechts D, 2009, J MED GENET, V46, P840, DOI 10.1136/jmg.2008.058222
   Le Bras B, 2006, NAT NEUROSCI, V9, P340, DOI 10.1038/nn1646
   LEE JJ, 1988, KOREAN J CHEM ENG, V5, P5, DOI 10.1007/BF02697470
   Lee P, 2002, P NATL ACAD SCI USA, V99, P10470, DOI 10.1073/pnas.162366299
   Lemmens R, 2007, HUM MOL GENET, V16, P2359, DOI 10.1093/hmg/ddm193
   LEUNG DW, 1989, SCIENCE, V246, P1306, DOI 10.1126/science.2479986
   Levine RJ, 2004, NEW ENGL J MED, V350, P672, DOI 10.1056/NEJMoa031884
   LEVY AP, 1995, J BIOL CHEM, V270, P13333, DOI 10.1074/jbc.270.22.13333
   Li BL, 2003, MOL BRAIN RES, V111, P155, DOI 10.1016/S0169-328X(03)00025-1
   LI J, 1995, J BIOL CHEM, V270, P308, DOI 10.1074/jbc.270.1.308
   Li X, 2008, ARTERIOSCL THROM VAS, V28, P1614, DOI 10.1161/ATVBAHA.107.158725
   Li XR, 2011, NEURODEGENERATIVE DISEASES - PROCESSES, PREVENTION, PROTECTION AND MONITORING, P387
   Li XR, 2012, TRENDS MOL MED, V18, P119, DOI 10.1016/j.molmed.2011.11.006
   Li XR, 2010, CELL RES, V20, P741, DOI 10.1038/cr.2010.77
   Li XR, 2009, CELL ADHES MIGR, V3, P322, DOI 10.4161/cam.3.4.9459
   Li Y, 2008, J CLIN INVEST, V118, P913, DOI 10.1172/JCI33673
   Licht T, 2013, CELL MOL LIFE SCI, V70, P1727, DOI 10.1007/s00018-013-1280-x
   Licht T, 2011, P NATL ACAD SCI USA, V108, P5081, DOI 10.1073/pnas.1007640108
   Licht T, 2010, DEVELOPMENT, V137, P261, DOI 10.1242/dev.039636
   Llado J, 2013, FRONT CELL NEUROSCI, V7, DOI 10.3389/fncel.2013.00181
   Lohela M, 2003, THROMB HAEMOSTASIS, V90, P167, DOI 10.1160/TH03-04-0200
   Louissaint A, 2002, NEURON, V34, P945, DOI 10.1016/S0896-6273(02)00722-5
   Loureiro RMB, 2005, CYTOKINE GROWTH F R, V16, P77, DOI 10.1016/j.cytogfr.2005.01.005
   Lu L, 2007, J NEUROSCI, V27, P7929, DOI 10.1523/JNEUROSCI.1877-07.2007
   Lu L, 2009, J BIOL CHEM, V284, P33989, DOI 10.1074/jbc.M109.067918
   Ma YH, 2011, BRAIN RES BULL, V86, P441, DOI 10.1016/j.brainresbull.2011.07.007
   Mackenzie F, 2012, DEVELOPMENT, V139, P1371, DOI 10.1242/dev.072348
   MAGLIONE D, 1991, P NATL ACAD SCI USA, V88, P9267, DOI 10.1073/pnas.88.20.9267
   Mani N, 2005, EXP NEUROL, V192, P394, DOI 10.1016/j.expneurol.2004.12.022
   Marconi S, 2013, NEUROSCIENCE, V248, P333, DOI 10.1016/j.neuroscience.2013.05.034
   Marconi S, 2012, TISSUE ENG PT A, V18, P1264, DOI [10.1089/ten.tea.2011.0491, 10.1089/ten.TEA.2011.0491]
   Maruyama H, 2010, NATURE, V465, P223, DOI 10.1038/nature08971
   Matsuzaki H, 2001, FASEB J, V15, P1218, DOI 10.1096/fj.00-0495fje
   Mauceri D, 2011, NEURON, V71, P117, DOI 10.1016/j.neuron.2011.04.022
   Maxwell PH, 2002, SEMIN CELL DEV BIOL, V13, P29, DOI 10.1006/scdb.2001.0287
   Meyer M, 1999, EMBO J, V18, P363, DOI 10.1093/emboj/18.2.363
   Miller RG, 2007, COCHRANE DB SYST REV, DOI 10.1002/14651858.CD001447.pub2
   Mole DR, 2001, IUBMB LIFE, V52, P43
   Moreau C, 2006, J NEUROL NEUROSUR PS, V77, P255, DOI 10.1136/jnnp.2005.070904
   Moreau C, 2011, NEUROSCIENCE, V172, P110, DOI 10.1016/j.neuroscience.2010.10.040
   Moreau C, 2009, AMYOTROPH LATERAL SC, V10, P175, DOI 10.1080/17482960802651725
   MORRISON KE, 1994, BAILLIERE CLIN NEUR, V3, P431
   Motterlini R, 2000, J BIOL CHEM, V275, P13613, DOI 10.1074/jbc.275.18.13613
   Murakami T, 2003, BRAIN RES, V989, P231, DOI 10.1016/S0006-8993(03)03374-2
   Nalini A, 2008, J NEUROL SCI, V272, P60, DOI 10.1016/j.jns.2008.04.034
   Neufeld G, 2002, TRENDS CARDIOVAS MED, V12, P13, DOI 10.1016/S1050-1738(01)00140-2
   Neufeld G, 2002, ADV EXP MED BIOL, V515, P81
   Neufeld G, 1999, FASEB J, V13, P9, DOI 10.1096/fasebj.13.1.9
   Neumann M, 2006, SCIENCE, V314, P130, DOI 10.1126/science.1134108
   Nishie A, 1999, CLIN CANCER RES, V5, P1107
   Nishimura AL, 2004, AM J HUM GENET, V75, P822, DOI 10.1086/425287
   Nobile-Orazio E, 2009, NEUROLOGY, V72, P1024, DOI 10.1212/01.wnl.0000344569.13496.ff
   Nygren I, 2002, NEUROREPORT, V13, P2199, DOI 10.1097/00001756-200212030-00007
   Ogunshola OO, 2002, J BIOL CHEM, V277, P11410, DOI 10.1074/jbc.M111085200
   Oh SJ, 1997, DEV BIOL, V188, P96, DOI 10.1006/dbio.1997.8639
   Olofsson B, 1999, CURR OPIN BIOTECH, V10, P528, DOI 10.1016/S0958-1669(99)00024-5
   Olsson AK, 2006, NAT REV MOL CELL BIO, V7, P359, DOI 10.1038/nrm1911
   Oosthuyse B, 2001, NAT GENET, V28, P131, DOI 10.1038/88842
   Orr HT, 2011, NEURON, V72, P189, DOI 10.1016/j.neuron.2011.10.001
   Paavonen K, 1996, CIRCULATION, V93, P1079, DOI 10.1161/01.CIR.93.6.1079
   Pages G, 2005, CARDIOVASC RES, V65, P564, DOI 10.1016/j.cardiores.2004.09.032
   PAJUSOLA K, 1994, ONCOGENE, V9, P3545
   Palmer TD, 2000, J COMP NEUROL, V425, P479, DOI 10.1002/1096-9861(20001002)425:4<479::AID-CNE2>3.0.CO;2-3
   Pandya Rachna S., 2012, Central Nervous System Agents in Medicinal Chemistry, V12, P15
   PARK JE, 1994, J BIOL CHEM, V269, P25646
   Pasinelli P, 2006, NAT REV NEUROSCI, V7, P710, DOI 10.1038/nrn1971
   Persico MG, 1999, CURR TOP MICROBIOL, V237, P31
   PERTOVAARA L, 1994, J BIOL CHEM, V269, P6271
   Pitzer MR, 2003, EXP NEUROL, V182, P435, DOI 10.1016/S0014-4886(03)00100-6
   Poesen K, 2008, J NEUROSCI, V28, P10451, DOI 10.1523/JNEUROSCI.1092-08.2008
   Proescholdt MA, 1999, J NEUROPATH EXP NEUR, V58, P613, DOI 10.1097/00005072-199906000-00006
   Proia P, 2008, INT J MOL MED, V21, P63
   Pugh CW, 2003, NAT MED, V9, P677, DOI 10.1038/nm0603-677
   Renton AE, 2011, NEURON, V72, P257, DOI 10.1016/j.neuron.2011.09.010
   Rissanen TT, 2003, CIRC RES, V92, P1098, DOI 10.1161/01.RES.0000073584.46059.E3
   Rizvanov AA, 2008, NEUROCHEM INT, V53, P389, DOI 10.1016/j.neuint.2008.09.011
   Robberecht W, 2013, NAT REV NEUROSCI, V14, P248, DOI 10.1038/nrn3430
   Ropper AH, 2009, ANN NEUROL, V65, P386, DOI 10.1002/ana.21675
   ROSEN DR, 1993, NATURE, V362, P59, DOI 10.1038/362059a0
   Rosenstein JM, 2010, ORGANOGENESIS, V6, P107, DOI 10.4161/org.6.2.11687
   Rosenstein JM, 2003, J NEUROSCI, V23, P11036
   Roskoski R, 2007, CRIT REV ONCOL HEMAT, V62, P179, DOI 10.1016/j.critrevonc.2007.01.006
   Roy H, 2006, FEBS LETT, V580, P2879, DOI 10.1016/j.febslet.2006.03.087
   Ruhrberg C, 2003, BIOESSAYS, V25, P1052, DOI 10.1002/bies.10351
   Ryan HE, 2000, CANCER RES, V60, P4010
   Ryter SW, 2000, AM J PHYSIOL-HEART C, V279, pH2889, DOI 10.1152/ajpheart.2000.279.6.H2889
   Ryuto M, 1996, J BIOL CHEM, V271, P28220, DOI 10.1074/jbc.271.45.28220
   Saharinen P, 2004, TRENDS IMMUNOL, V25, P387, DOI 10.1016/j.it.2004.05.003
   SAIT SNJ, 1995, CYTOGENET CELL GENET, V70, P145, DOI 10.1159/000134081
   Sakowski SA, 2009, GENE THER, V16, P1292, DOI 10.1038/gt.2009.111
   Salceda S, 1997, J BIOL CHEM, V272, P22642, DOI 10.1074/jbc.272.36.22642
   Salven P, 1998, AM J PATHOL, V153, P103, DOI 10.1016/S0002-9440(10)65550-2
   Sathasivam S, 2008, NEUROSCI RES, V62, P71, DOI 10.1016/j.neures.2008.06.008
   SATOH H, 1987, JPN J CANCER RES, V78, P772
   Schanzer A, 2004, BRAIN PATHOL, V14, P237
   Schiera G, 2007, J CELL MOL MED, V11, P1384, DOI 10.1111/j.1582-4934.2007.00100.x
   Schwartz CM, 2012, J NEUROSCI RES, V90, P1367, DOI 10.1002/jnr.23064
   Semenza GL, 2010, ONCOGENE, V29, P625, DOI 10.1038/onc.2009.441
   Semenza GL, 2001, CURR OPIN CELL BIOL, V13, P167, DOI 10.1016/S0955-0674(00)00194-0
   SHALABY F, 1995, NATURE, V376, P62, DOI 10.1038/376062a0
   Shibuya M, 2006, EXP CELL RES, V312, P549, DOI 10.1016/j.yexcr.2005.11.012
   Shibuya M., 2008, ANGIOGENESIS, V12, P15
   Shibuya M, 2006, J BIOCHEM MOL BIOL, V39, P469
   Shin YJ, 2008, ACTA NEUROPATHOL, V116, P517, DOI 10.1007/s00401-008-0423-x
   Shiote M, 2005, NEUROSCIENCE, V132, P175, DOI 10.1016/j.neuroscience.2004.12.031
   Siemeister G, 1996, CANCER RES, V56, P2299
   Silverman WF, 1999, NEUROSCIENCE, V90, P1529, DOI 10.1016/S0306-4522(98)00540-5
   Skene JHP, 2001, NAT GENET, V28, P107, DOI 10.1038/88805
   Soker S, 1998, CELL, V92, P735, DOI 10.1016/S0092-8674(00)81402-6
   Soker S, 2002, J CELL BIOCHEM, V85, P357, DOI 10.1002/jcb.10140
   Sondell M, 1999, J NEUROSCI, V19, P5731, DOI 10.1523/JNEUROSCI.19-14-05731.1999
   Sondell M, 2000, EUR J NEUROSCI, V12, P4243, DOI 10.1046/j.0953-816X.2000.01326.x
   Storkebaum E, 2004, J CLIN INVEST, V113, P14, DOI 10.1172/JCI200420682
   Storkebaum E, 2005, NAT NEUROSCI, V8, P85, DOI 10.1038/nn1360
   Storkebaum E, 2004, BIOESSAYS, V26, P943, DOI 10.1002/bies.20092
   Sun YJ, 2004, J CEREBR BLOOD F MET, V24, P1146, DOI 10.1097/01.WCB.0000134477.38980.38
   Sun YJ, 2003, J CLIN INVEST, V111, P1843, DOI 10.1172/JCI200317977
   Sun YJ, 2006, DEV BIOL, V289, P329, DOI 10.1016/j.ydbio.2005.10.016
   Sutter CH, 2000, P NATL ACAD SCI USA, V97, P4748, DOI 10.1073/pnas.080072497
   Svensson B, 2002, J CEREBR BLOOD F MET, V22, P1170, DOI 10.1097/01.wcb.0000037988.07114.98
   Takahashi H, 2005, CLIN SCI, V109, P227, DOI 10.1042/CS20040370
   Takashima S, 2002, P NATL ACAD SCI USA, V99, P3657, DOI 10.1073/pnas.022017899
   Tammela T, 2005, CARDIOVASC RES, V65, P550, DOI 10.1016/j.cardiores.2004.12.002
   Tammela T, 2008, NATURE, V454, P656, DOI 10.1038/nature07083
   Teng YD, 2012, SCI TRANSL MED, V4, DOI 10.1126/scitranslmed.3004579
   TERMAN BI, 1991, ONCOGENE, V6, P1677
   Terry PD, 2004, J NEUROGENET, V18, P429, DOI 10.1080/01677060490894450
   Thadhani R, 2004, J CLIN ENDOCR METAB, V89, P770, DOI 10.1210/jc.2003-031244
   Tolosa L, 2008, J NEUROCHEM, V105, P1080, DOI 10.1111/j.1471-4159.2007.05206.x
   Tovar-y-Romo LB, 2010, J NEUROCHEM, V115, P1090, DOI 10.1111/j.1471-4159.2010.06766.x
   Tsuzuki Y, 2000, CANCER RES, V60, P6248
   Turner BJ, 2008, PROG NEUROBIOL, V85, P94, DOI 10.1016/j.pneurobio.2008.01.001
   Turner MR, 2013, LANCET NEUROL, V12, P310, DOI 10.1016/S1474-4422(13)70036-X
   van Blitterswijk M., 2012, COMPLEXITY FAMILIAL, P135
   Van Damme P, 2002, J NEUROPHYSIOL, V88, P1279, DOI 10.1152/jn.2002.88.3.1279
   Van Damme P, 2007, P NATL ACAD SCI USA, V104, P14825, DOI 10.1073/pnas.0705046104
   Van Den Bosch L, 2004, NEUROBIOL DIS, V17, P21, DOI 10.1016/j.nbd.2004.06.004
   Van Vught PWJ, 2005, NEUROLOGY, V65, P1643, DOI 10.1212/01.wnl.0000184514.39853.56
   Vance C, 2009, SCIENCE, V323, P1208, DOI 10.1126/science.1165942
   Vincenti V, 1996, CIRCULATION, V93, P1493, DOI 10.1161/01.CIR.93.8.1493
   Volm M, 1999, ANTICANCER RES, V19, P1669
   WANG GL, 1995, J BIOL CHEM, V270, P1230, DOI 10.1074/jbc.270.3.1230
   WANG GL, 1995, P NATL ACAD SCI USA, V92, P5510, DOI 10.1073/pnas.92.12.5510
   Wang YM, 2007, J NEUROSCI, V27, P304, DOI 10.1523/JNEUROSCI.4433-06.2007
   Wang ZG, 2010, BIOCHEM J, V428, P347, DOI 10.1042/BJ20100379
   Warner-Schmidt JL, 2008, CURR OPIN PHARMACOL, V8, P14, DOI 10.1016/j.coph.2007.10.013
   Warner-Schmidt JL, 2007, P NATL ACAD SCI USA, V104, P4647, DOI 10.1073/pnas.0610282104
   Warner-Schmidt JL, 2006, HIPPOCAMPUS, V16, P239, DOI 10.1002/hipo.20156
   Wijesekera LC, 2009, ORPHANET J RARE DIS, V4, DOI 10.1186/1750-1172-4-3
   Xie KP, 2004, CYTOKINE GROWTH F R, V15, P297, DOI 10.1016/j.cytogfr.2004.04.003
   Xie L, 2013, VASC CELL, V5, DOI 10.1186/2045-824X-5-8
   Yamada Y, 1997, GENOMICS, V42, P483, DOI 10.1006/geno.1997.4774
   Yang JP, 2009, NEUROSCI LETT, V449, P108, DOI 10.1016/j.neulet.2008.10.090
   Yang Y, 2001, NAT GENET, V29, P160, DOI 10.1038/ng1001-160
   Yuan L, 2002, DEVELOPMENT, V129, P4797
   Zachary I, 1998, EXP NEPHROL, V6, P480
   Zachary I, 2001, CARDIOVASC RES, V49, P568, DOI 10.1016/S0008-6363(00)00268-6
   Zhang C, 2009, CYTOTHERAPY, V11, P299, DOI 10.1080/14653240902806986
   Zhang F, 2009, P NATL ACAD SCI USA, V106, P6152, DOI 10.1073/pnas.0813061106
   Zhang YS, 2006, AMYOTROPH LATERAL SC, V7, P119, DOI 10.1080/14660820600600657
   Zheng CY, 2004, ANN NEUROL, V56, P564, DOI 10.1002/ana.20223
   Zhong ZH, 2008, NAT NEUROSCI, V11, P420, DOI 10.1038/nn2073
NR 297
TC 37
Z9 39
U1 0
U2 12
PU HINDAWI LTD
PI LONDON
PA ADAM HOUSE, 3RD FLR, 1 FITZROY SQ, LONDON, W1T 5HF, ENGLAND
SN 2314-6133
EI 2314-6141
J9 BIOMED RES INT
JI Biomed Res. Int.
PY 2014
VL 2014
AR 947513
DI 10.1155/2014/947513
PG 24
WC Biotechnology & Applied Microbiology; Medicine, Research & Experimental
WE Science Citation Index Expanded (SCI-EXPANDED)
SC Biotechnology & Applied Microbiology; Research & Experimental Medicine
GA AH1SI
UT WOS:000335900800001
PM 24987705
OA Green Published, gold, Green Submitted
DA 2023-08-27
ER

PT J
AU Elmquist, JK
   Scammell, TE
   Jacobson, CD
   Safer, CB
AF Elmquist, JK
   Scammell, TE
   Jacobson, CD
   Safer, CB
TI Distribution of Fos-like immunoreactivity in the rat brain following
   intravenous lipopolysaccharide administration
SO JOURNAL OF COMPARATIVE NEUROLOGY
LA English
DT Article
DE endotoxin; autonomic regulation; neuroimmune interactions; fever; acute
   phase response
ID CORTICOTROPIN-RELEASING FACTOR; TUMOR-NECROSIS-FACTOR;
   PHASEOLUS-VULGARIS-LEUKOAGGLUTININ; PROSTAGLANDIN-E2 BINDING-SITES;
   CENTRAL-NERVOUS-SYSTEM; C-FOS; PARABRACHIAL NUCLEUS; HYPOTHALAMIC
   NUCLEUS; 3RD VENTRICLE; ORGANIZATION
AB The central nervous system, particularly the hypothalamus, is intimately involved in the coordination of various aspects of the inflammatory response, including the generation of fever. We used intravenous injections of bacterial cell wall lipopolysaccharide (LPS; 5 or 125 mu g/kg) to stimulate the acute phase response and mapped the resultant distribution of Fos-like immunoreactivity in the rat brain. In addition, we compared the patterns of Fos distribution with the thermoregulatory responses elicited by the LPS. Administration of LPS resulted in a dose- and time-dependent pattern of Fos-like immunoreactivity throughout the rat brain consistent with a coordinated autonomic, endocrine, and behavioral response to the LPS challenge that was most pronounced 2 hours following injection. Specifically, Fos-like immunoreactivity was observed in key autonomic regulatory nuclear groups, including the insular and prelimbic cortices, paraventricular hypothalamic nucleus, parabrachial nucleus, nucleus of the solitary tract, and the rostral and caudal levels of the ventrolateral medulla. In addition, a significant sustained elevation of Fos-like immunoreactivity was observed in a cell group adjacent to the organum vasculosum of the lamina terminalis, which we termed the ventromedial preoptic area. This sustained elevation of Fos-like immunoreactivity coupled with the alterations in body temperature elicited by LPS leads us to hypothesize that the ventromedial preoptic area may be a key site for the initiation of fever during endotoxemia. (C) 1996 Wiley-Liss, Inc.
C1 HARVARD UNIV, BETH ISRAEL HOSP, SCH MED, PROGRAM NEUROSCI, BOSTON, MA 02115 USA.
   IOWA STATE UNIV SCI & TECHNOL, DEPT VET ANAT, AMES, IA 50011 USA.
   IOWA STATE UNIV SCI & TECHNOL, NEUROSCI PROGRAM, AMES, IA 50011 USA.
C3 Harvard University; Beth Israel Deaconess Medical Center; Harvard
   Medical School; Iowa State University; Iowa State University
RP Elmquist, JK (通讯作者)，HARVARD UNIV, BETH ISRAEL HOSP, SCH MED, DEPT NEUROL, 77 AVE LOUIS PASTEUR, BOSTON, MA 02115 USA.
FU NIMH NIH HHS [MH10832] Funding Source: Medline; NINDS NIH HHS [NS22835,
   NS09474] Funding Source: Medline
CR ABERCROMBIE M, 1946, ANAT REC, V94, P239, DOI 10.1002/ar.1090940210
   ARMSTRONG DM, 1982, J COMP NEUROL, V212, P173, DOI 10.1002/cne.902120207
   BANKS WA, 1989, BRAIN RES BULL, V23, P433, DOI 10.1016/0361-9230(89)90185-8
   BLATTEIS CM, 1983, BRAIN RES BULL, V11, P519, DOI 10.1016/0361-9230(83)90124-7
   Boulant JA., 1991, FEVER BASIC MECHANIS, P1
   BRADY LS, 1994, J NEUROSCI, V14, P4951
   BREDER CD, 1995, J COMP NEUROL, V355, P296, DOI 10.1002/cne.903550208
   BREDER CD, 1994, P NATL ACAD SCI USA, V91, P11393, DOI 10.1073/pnas.91.24.11393
   BREDER CD, 1993, J COMP NEUROL, V337, P543, DOI 10.1002/cne.903370403
   Breder CD, 1996, BRAIN RES, V713, P64, DOI 10.1016/0006-8993(95)01474-8
   BREDER CD, 1992, J COMP NEUROL, V322, P409, DOI 10.1002/cne.903220309
   BURD RS, 1992, IMMUNOTHERAPY GRAM N, P3
   CANTERAS NS, 1994, J COMP NEUROL, V348, P41, DOI 10.1002/cne.903480103
   CHAN RKW, 1993, J NEUROSCI, V13, P5126, DOI 10.1523/JNEUROSCI.13-12-05126.1993
   CHAN RKW, 1994, J COMP NEUROL, V348, P433, DOI 10.1002/cne.903480309
   Chen T Y, 1992, Curr Top Microbiol Immunol, V181, P169
   CHROUSOS GP, 1995, NEW ENGL J MED, V332, P1351, DOI 10.1056/NEJM199505183322008
   COGGESHALL RE, 1992, TRENDS NEUROSCI, V15, P9, DOI 10.1016/0166-2236(92)90339-A
   CULLINAN WE, 1995, NEUROSCIENCE, V64, P477, DOI 10.1016/0306-4522(94)00355-9
   DANTZER R, 1994, EUR CYTOKINE NETW, V5, P271
   DERIJK RH, 1994, AM J PHYSIOL, V266, pR9, DOI 10.1152/ajpregu.1994.266.1.R9
   Elmquist J. K., 1995, Society for Neuroscience Abstracts, V21, P879
   ELMQUIST JK, 1993, ENDOCRINOLOGY, V133, P3054, DOI 10.1210/en.133.6.3054
   ERICSSON A, 1994, J NEUROSCI, V14, P897
   FEIL K, 1995, J COMP NEUROL, V353, P506, DOI 10.1002/cne.903530404
   FULWILER CE, 1984, BRAIN RES REV, V7, P229, DOI 10.1016/0165-0173(84)90012-2
   GORDON CJ, 1990, PHYSIOL BEHAV, V47, P963, DOI 10.1016/0031-9384(90)90025-Y
   HERBERT H, 1995, J COMP NEUROL, V354, P377, DOI 10.1002/cne.903540307
   HERBERT H, 1992, J COMP NEUROL, V315, P34, DOI 10.1002/cne.903150104
   HERBERT H, 1990, J COMP NEUROL, V293, P581, DOI 10.1002/cne.902930405
   HERDEGEN T, 1995, J COMP NEUROL, V354, P39, DOI 10.1002/cne.903540105
   HOFFMAN GE, 1993, FRONT NEUROENDOCRIN, V14, P173, DOI 10.1006/frne.1993.1006
   INIGUEZ C, 1985, J NEUROSCI METH, V13, P77, DOI 10.1016/0165-0270(85)90045-7
   KATSUURA G, 1990, AM J PHYSIOL, V258, pE163, DOI 10.1152/ajpendo.1990.258.1.E163
   KEY BJ, 1994, J AUTONOM NERV SYST, V48, P79, DOI 10.1016/0165-1838(94)90162-7
   KLUGER MJ, 1991, PHYSIOL REV, V71, P93, DOI 10.1152/physrev.1991.71.1.93
   KONONEN J, 1990, NEUROSCI LETT, V120, P105, DOI 10.1016/0304-3940(90)90179-D
   KOVACS KJ, 1993, P NATL ACAD SCI USA, V90, P7681, DOI 10.1073/pnas.90.16.7681
   LONG NC, 1991, PHYSIOL BEHAV, V50, P625, DOI 10.1016/0031-9384(91)90556-4
   LYNN WA, 1992, IMMUNOL TODAY, V13, P271, DOI 10.1016/0167-5699(92)90009-V
   MATSUMURA K, 1990, BRAIN RES, V533, P147, DOI 10.1016/0006-8993(90)91808-T
   MOGA MM, 1985, J COMP NEUROL, V241, P275, DOI 10.1002/cne.902410304
   MOGA MM, 1989, J COMP NEUROL, V283, P315, DOI 10.1002/cne.902830302
   Oldfield Brian J., 1995, P391
   PEZZONE MA, 1992, BRAIN RES, V597, P41, DOI 10.1016/0006-8993(92)91503-7
   RISOLD PY, 1994, J COMP NEUROL, V348, P1, DOI 10.1002/cne.903480102
   SAGAR SM, 1988, SCIENCE, V240, P1328, DOI 10.1126/science.3131879
   SAGAR SM, 1994, ANN NY ACAD SCI, V739, P282, DOI 10.1111/j.1749-6632.1994.tb19830.x
   SAGAR SM, 1995, BRAIN RES BULL, V36, P381, DOI 10.1016/0361-9230(94)00217-O
   SAGAR SM, 1993, ADV NEUROL, V59, P273
   SAPER CB, 1994, NEW ENGL J MED, V330, P1880
   SAPER CB, 1992, PROG BRAIN RES, V93, P419
   SAPER CB, 1976, BRAIN RES, V117, P305, DOI 10.1016/0006-8993(76)90738-1
   Saper Clifford B., 1995, P107
   SCAMMELL TE, 1995, ANN NEUROL, V38, P339
   SCAMMELL TE, 1996, SOC NEUR ABSTR
   Sherin JE, 1996, SCIENCE, V271, P216, DOI 10.1126/science.271.5246.216
   SIMERLY RB, 1987, BRAIN RES, V400, P11, DOI 10.1016/0006-8993(87)90649-4
   Simerly Richard B., 1995, P353
   STANDAERT DG, 1988, J NEUROSCI, V8, P1940
   STANDAERT DG, 1986, J COMP NEUROL, V253, P315, DOI 10.1002/cne.902530304
   SVED AF, 1994, AM J PHYSIOL, V266, pR361, DOI 10.1152/ajpregu.1994.266.2.R361
   SWANSON LW, 1983, ANNU REV NEUROSCI, V6, P269, DOI 10.1146/annurev.ne.06.030183.001413
   TKACS NC, 1995, J AUTONOM NERV SYST, V51, P1, DOI 10.1016/0165-1838(95)80001-Q
   VEENING JG, 1993, J CHEM NEUROANAT, V6, P391, DOI 10.1016/0891-0618(93)90013-T
   WAN WH, 1994, BRAIN RES BULL, V34, P7, DOI 10.1016/0361-9230(94)90179-1
   WAN WH, 1993, BRAIN RES BULL, V32, P581, DOI 10.1016/0361-9230(93)90158-8
   WATANABE Y, 1988, J NEUROSCI, V8, P2003
   WATKINS LR, 1995, NEUROSCI LETT, V183, P27, DOI 10.1016/0304-3940(94)11105-R
NR 69
TC 283
Z9 291
U1 0
U2 8
PU WILEY
PI HOBOKEN
PA 111 RIVER ST, HOBOKEN 07030-5774, NJ USA
SN 0021-9967
EI 1096-9861
J9 J COMP NEUROL
JI J. Comp. Neurol.
PD JUL 15
PY 1996
VL 371
IS 1
BP 85
EP 103
DI 10.1002/(SICI)1096-9861(19960715)371:1<85::AID-CNE5>3.0.CO;2-H
PG 19
WC Neurosciences; Zoology
WE Science Citation Index Expanded (SCI-EXPANDED)
SC Neurosciences & Neurology; Zoology
GA UX962
UT WOS:A1996UX96200006
PM 8835720
DA 2023-08-27
ER

PT J
AU Perez-Bovet, J
   Rimbau-Munoz, J
   Martin-Ferrer, S
AF Perez-Bovet, Jordi
   Rimbau-Munoz, Jordi
   Martin-Ferrer, Secundino
TI Anaplastic Ependymoma With Holocordal and Intracranial Meningeal
   Carcinomatosis and Holospinal Bone Metastases FSD
SO NEUROSURGERY
LA English
DT Article
DE Anaplastic ependymoma; Bone metastases; Extramedullary neoplasm;
   Leptomeningeal carcinomatosis
ID SPINAL-CORD TUMORS; INTRADURAL EXTRAMEDULLARY EPENDYMOMA; CAUDA-EQUINA;
   EXTRANEURAL METASTASES; PROGNOSTIC-FACTORS; SURGICAL-TREATMENT;
   DISSEMINATION; CHEMOTHERAPY; CHILDHOOD; SURGERY
AB BACKGROUND AND IMPORTANCE: Ependymomas are the most frequent intramedullary neoplasms in adult patients. Anaplastic histology, extramedullary location, meningeal dissemination at initial diagnosis, and extraneural metastases are rare findings. We describe a case of extramedullary anaplastic ependymoma that presented with holocordal and intracranial leptomeningeal carcinomatosis and bone metastases in all the vertebral bodies and the sternum. Such an aggressive dissemination at initial diagnosis has not been previously reported.
   CLINICAL PRESENTATION: A 36-year-old woman presented with headache, multiple cranial nerve palsies, visual hallucinations, confusion, hemiparesis, hemihipoestesia, episodes of disconnection, and toxic syndrome. Magnetic resonance imaging and positron emission tomography scan revealed leptomeningeal carcinomatosis in the brainstem, the cerebellum, and along the whole spinal cord. Various nodular, intradural extramedullary lesions were present at multiple dorsal and lumbar levels. Metastatic bone disease affected all the vertebral bodies and various extraspinal bones. An intradural and bone biopsy was performed at L4, providing the diagnosis of anaplastic ependymoma (World Health Organization grade III) with focal neuronal differentiation. Despite chemotherapy, the patient's symptoms quickly progressed, and she died 7 weeks after diagnosis.
   CONCLUSION: To our knowledge, there are no previous descriptions of ependymomas with this extensive leptomeningeal, spinal, intracranial, and extraneural dissemination at clinical onset. Bone metastases in spinal ependymoma have not been previously reported.
C1 [Perez-Bovet, Jordi; Rimbau-Munoz, Jordi; Martin-Ferrer, Secundino] Univ Hosp Dr Josep Trueta, Dept Neurosurg, Girona, Girona, Spain.
C3 Universitat de Girona; Girona University Hospital Dr. Josep Trueta
RP Perez-Bovet, J (通讯作者)，Avinguda Franca S-N, Girona 17007, Girona, Spain.
EM
CR Amirian ES, 2012, J NEURO-ONCOL, V107, P183, DOI 10.1007/s11060-011-0730-2
   Benes V, 2009, EUR SPINE J, V18, P1397, DOI 10.1007/s00586-009-1076-8
   Benzagmout M, 2008, SURG NEUROL, V70, P536, DOI 10.1016/j.surneu.2007.05.055
   Bonfield CM, 2011, NEUROSURGERY, V68, pE831, DOI 10.1227/NEU.0b013e318209257e
   Bostrom A, 2011, NEUROSURGERY, V68, P302, DOI 10.1227/NEU.0b013e3182004c1e
   Duffau H, 2000, SPINE, V25, P1993, DOI 10.1097/00007632-200008010-00021
   DUFFNER PK, 1981, ANN NEUROL, V10, P261, DOI 10.1002/ana.410100311
   Eroes CA, 2010, ACTA NEUROCHIR, V152, P611, DOI 10.1007/s00701-009-0577-x
   FARRIER SE, 1994, SOUTHERN MED J, V87, P1262, DOI 10.1097/00007611-199412000-00010
   Graf M, 1999, Pathol Oncol Res, V5, P56, DOI 10.1053/paor.1999.0056
   Hussain M, 2010, PEDIATR BLOOD CANCER, V55, P1204, DOI 10.1002/pbc.22604
   Innocenzi G, 1997, CLIN NEUROL NEUROSUR, V99, P1, DOI 10.1016/S0303-8467(96)00555-0
   Iunes EA, 2011, J NEUROSURG-SPINE, V14, P65, DOI 10.3171/2010.9.SPINE09963
   Kara-Terki R, 2000, JOINT BONE SPINE, V67, P471
   Lin YH, 2005, J NEURO-ONCOL, V71, P205, DOI 10.1007/s11060-004-1386-y
   Martinez-Perez Rafael, 2012, Surg Neurol Int, V3, P33, DOI 10.4103/2152-7806.93865
   Matsuyama Y, 2009, J NEUROSURG-SPINE, V10, P404, DOI 10.3171/2009.2.SPINE08698
   MAVROUDIS C, 1977, J NEUROSURG, V47, P771, DOI 10.3171/jns.1977.47.5.0771
   MCCORMICK PC, 1990, J NEUROSURG, V72, P523, DOI 10.3171/jns.1990.72.4.0523
   MORRIS DM, 1983, J SURG ONCOL, V22, P33, DOI 10.1002/jso.2930220109
   Nakamura M, 2008, SPINAL CORD, V46, P282, DOI 10.1038/sj.sc.3102130
   NEWTON HB, 1992, J NEURO-ONCOL, V14, P135
   PATTERSON R, 1961, J NEUROSURG, V18, P145, DOI 10.3171/jns.1961.18.2.0145
   Plans G, 2006, ACTA NEUROCHIR, V148, P343, DOI 10.1007/s00701-005-0693-1
   Raco A, 2005, NEUROSURGERY, V56, P972, DOI 10.1227/01.NEU.0000158318.66568.CC
   Rickert CH, 1999, ACTA NEUROCHIR, V141, P781, DOI 10.1007/s007010050376
   RUBINSTEIN LJ, 1970, J NEUROL NEUROSUR PS, V33, P763, DOI 10.1136/jnnp.33.6.763
   Ruda R, 2008, CURR OPIN NEUROL, V21, P754, DOI 10.1097/WCO.0b013e328317efe8
   Schuurmans M, 2006, J NEURO-ONCOL, V79, P57, DOI 10.1007/s11060-005-9114-9
   Sharma KD, 1956, INDIAN J MED SCI, V10, P639
   Timmermann B, 2000, INT J RADIAT ONCOL, V46, P287, DOI 10.1016/S0360-3016(99)00414-9
   Vural M, 2010, J NEUROSURG-SPINE, V12, P467, DOI 10.3171/2009.11.SPINE08780
   Wang ZY, 2012, NEUROSURG REV, V35, P85, DOI 10.1007/s10143-011-0345-2
   WATT V, 1968, J NEUROSURG, V29, P424, DOI 10.3171/jns.1968.29.4.0424
   WEISS L, 1955, CANCER, V8, P161, DOI 10.1002/1097-0142(1955)8:1<161::AID-CNCR2820080123>3.0.CO;2-K
   WIGHT DGD, 1973, J CLIN PATHOL, V26, P929, DOI 10.1136/jcp.26.12.929
   Yang SY, 2009, SPINE, V34, P2705, DOI 10.1097/BRS.0b013e3181b43484
NR 37
TC 6
Z9 7
U1 1
U2 6
PU LIPPINCOTT WILLIAMS & WILKINS
PI PHILADELPHIA
PA 530 WALNUT ST, PHILADELPHIA, PA 19106-3621 USA
SN 0148-396X
J9 NEUROSURGERY
JI Neurosurgery
PD MAR
PY 2013
VL 72
IS 3
BP E497
EP E503
DI 10.1227/NEU.0b013e31827d102e
PG 7
WC Clinical Neurology; Surgery
WE Science Citation Index Expanded (SCI-EXPANDED)
SC Neurosciences & Neurology; Surgery
GA 093LQ
UT WOS:000315193800001
PM 23422903
DA 2023-08-27
ER

PT J
AU Senescende, L
   Bitolog, PL
   Auberger, E
   Le Bian, AZ
   Cesaretti, M
AF Senescende, L.
   Bitolog, P. L.
   Auberger, E.
   Le Bian, A. Zarzavadjian
   Cesaretti, M.
TI Adrenal ectopy of adult groin region: a systematic review of an
   unexpected anatomopathologic diagnosis
SO HERNIA
LA English
DT Review
DE Adrenal gland; Groin region; Adrenal ectopy; Adult
ID REST TUMOR; ADRENOCORTICAL CARCINOMA; BROAD LIGAMENT; HERNIA SACS;
   TISSUE; ASSOCIATION; CHILDREN; PATIENT; ADENOMA; CORTEX
AB Adrenal ectopy is a rare condition, caused by abnormal embryological development of the adrenal glands and testis. This anomaly is asymptomatic and is generally diagnosed in childhood. Most cases of adrenal ectopy of the groin region have been found incidentally during surgery. This review aims to evaluate the incidence of adrenal ectopy of the adult groin region, as well as its short- and long-term significance.
   A systematic literature search was performed using the EMBASE, MEDLINE, PubMed and Cochrane Central databases to identify all studies published until December 2015. Articles written in English containing the keywords: ''ectopic adrenal tissue'' or ''aberrant adrenal tissue'' or ''adrenal heterotopy'' combined with ''groin region'' or ''inguinal'' or ''testicular" or "scrotum" and "adult" were selected.
   Fifteen articles matched the review criteria. A total of 21 cases (100 % male) were included. No cases of bilateral nodules have been reported. The mean age at diagnosis was 43.5 years old (range 19-78 years). A predominance of right-sided lesions was observed (71.4 %). The mean size reached 7.6 mm (range 2-40 mm) and hernia sacs were the most common location (47.6 %). All histological analyses of harvested tissues consisted of adrenal cortex without medulla.
   Our results suggest that routine pathological examination of any groin tissue excised during inguinal hernia repairs is required for the diagnosis of adrenal ectopy in the groin. Although there is no current evidence that endocrine or oncologic complications can occur from excision of ectopic adrenal tissue, further confirmatory studies may be required.
C1 [Senescende, L.; Le Bian, A. Zarzavadjian; Cesaretti, M.] Ctr Hosp Simone Veil, Serv Chirurg Digest, 14 Rue St Prix, F-95600 Eaubonne, France.
   [Bitolog, P. L.; Auberger, E.] Ctr Hosp Simone Veil, Serv Anatomopathol, 14 Rue St Prix, F-95600 Eaubonne, France.
   [Le Bian, A. Zarzavadjian] Univ Paris 05, Lab Eth Med & Med Legale, Paris, France.
   [Cesaretti, M.] Beaujon Hosp, Dept HPB Surg & Liver Transplantat, 100 Blvd Gen Leclerc, F-92110 Clichy, France.
C3 UDICE-French Research Universities; Universite Paris Cite; UDICE-French
   Research Universities; Universite Paris Cite; Assistance Publique
   Hopitaux Paris (APHP); Hopital Universitaire Beaujon - APHP
RP Cesaretti, M (通讯作者)，Ctr Hosp Simone Veil, Serv Chirurg Digest, 14 Rue St Prix, F-95600 Eaubonne, France.; Cesaretti, M (通讯作者)，Beaujon Hosp, Dept HPB Surg & Liver Transplantat, 100 Blvd Gen Leclerc, F-92110 Clichy, France.
EM
RI Cesaretti, Manuela/AAK-6044-2020
OI Cesaretti, Manuela/0000-0001-6419-1481; Zarzavadjian Le Bian,
   Alban/0000-0002-7384-1148
CR Akishima-Fukasawa Y, 2011, ENDOCR PATHOL, V22, P112, DOI 10.1007/s12022-011-9152-0
   Al Nemer AM, 2015, HERNIA, V19, P915, DOI 10.1007/s10029-014-1338-1
   ANDERSON JR, 1980, POSTGRAD MED J, V56, P806, DOI 10.1136/pgmj.56.661.806
   Arai K, 2000, PATHOL INT, V50, P244, DOI 10.1046/j.1440-1827.2000.01029.x
   Bani-Hani KE, 2003, SAUDI MED J, V24, P301
   Cariou J, 2008, PROG UROL, V18, P68, DOI 10.1016/j.purol.2007.12.004
   CHROUSOS GP, 1981, J UROLOGY, V126, P127, DOI 10.1016/S0022-5347(17)54411-0
   CZAPLICKI M, 1985, International Urology and Nephrology, V17, P177, DOI 10.1007/BF02082491
   El Demellawy D, 2009, HERNIA, V13, P659, DOI 10.1007/s10029-009-0501-6
   Floyd MS, 2014, CUAJ-CAN UROL ASSOC, V8, pE176, DOI 10.5489/cuaj.1584
   GOREN E, 1991, UROLOGY, V38, P187, DOI 10.1016/S0090-4295(05)80085-8
   GRAHAM LS, 1953, CANCER, V6, P149, DOI 10.1002/1097-0142(195301)6:1<149::AID-CNCR2820060115>3.0.CO;2-8
   GUALTIERI T, 1949, J UROLOGY, V61, P949, DOI 10.1016/S0022-5347(17)69167-5
   GUTOWSKI WT, 1979, J UROLOGY, V121, P353, DOI 10.1016/S0022-5347(17)56783-X
   Iyengar V, 2007, ANN DIAGN PATHOL, V11, P291, DOI 10.1016/j.anndiagpath.2006.08.001
   Jaffe HL, 1927, ARCH PATHOL LAB MED, V3, P415
   Jain SH, 2008, NAT CLIN PRACT ENDOC, V4, P695, DOI 10.1038/ncpendmet0985
   Kasajima A, 2014, PATHOL INT, V64, P183, DOI 10.1111/pin.12154
   Konishi, 2014, OPEN J UROLOGY, V4, P151, DOI [10.4236/oju.2014.412026, DOI 10.4236/OJU.2014.412026]
   Lack EE, 1997, ATLAS TUMOR PATHOL, V19, P33
   Lodeville D, 1026, PATHOLOGICA, V1981, P629
   Mari M, 2004, UROL INT, V73, P191, DOI 10.1159/000079705
   Mendez Roberto, 2006, Int. braz j urol., V32, P202
   MORIMOTO Y, 1971, J CLIN ENDOCR METAB, V32, P201, DOI 10.1210/jcem-32-2-201
   Niveditha SR, 2014, J EVID BASED MED HEA, V3, P157
   O'Crowley CR, 1943, J UROLOGY, V50, P756, DOI 10.1016/S0022-5347(17)70508-3
   Ozel SK, 2007, PEDIATR SURG INT, V23, P171, DOI 10.1007/s00383-006-1826-1
   Oguzkurt Pelin, 2002, Hernia, V6, P62, DOI 10.1007/s10029-002-0056-2
   OKUR H, 1995, PEDIATR PATHOL LAB M, V15, P763, DOI 10.3109/15513819509027011
   Rabie ME, 2013, UROL ANNALS, V5, P302, DOI 10.4103/0974-7796.120307
   Raman RT, 2014, J CLIN DIAGN RES, V8, pFD11, DOI 10.7860/JCDR/2014/8331.4845
   Ren PT, 2013, WORLD J GASTROENTERO, V19, P778, DOI 10.3748/wjg.v19.i5.778
   Rich MA, 1998, J UROLOGY, V160, P1838, DOI 10.1016/S0022-5347(01)62429-7
   ROGGIA A, 1991, Archivos Espanoles de Urologia, V44, P1165
   Sasano H, 1997, PATHOL INT, V47, P493, DOI 10.1111/j.1440-1827.1997.tb04529.x
   Savas Cagri, 2001, International Urology and Nephrology, V32, P681, DOI 10.1023/A:1014429119226
   SCHECHTE.DC, 1968, ANN SURG, V167, P421, DOI 10.1097/00000658-196803000-00017
   Shin Yong Moon, 2010, Korean J Hepatol, V16, P338, DOI 10.3350/kjhep.2010.16.3.338
   Souverijns G, 2000, EUR RADIOL, V10, P1165, DOI 10.1007/s003309900263
   Sullivan JG, 2005, BJU INT, V95, P407, DOI 10.1111/j.1464-410X.2005.05310.x
   Tajima T, 2001, J COMPUT ASSIST TOMO, V25, P98, DOI 10.1097/00004728-200101000-00018
   Ventura L, 1998, Arch Ital Urol Androl, V70, P15
   Wang T, 2013, J CLIN PATHOL, V66, P1084, DOI 10.1136/jclinpath-2013-201734
   WOLLOCH Y, 1986, PANMINERVA MED, V28, P47
   Yokoyama H, 2013, TOHOKU J EXP MED, V229, P267, DOI 10.1620/tjem.229.267
NR 45
TC 8
Z9 9
U1 0
U2 2
PU SPRINGER
PI NEW YORK
PA 233 SPRING ST, NEW YORK, NY 10013 USA
SN 1265-4906
EI 1248-9204
J9 HERNIA
JI Hernia
PD DEC
PY 2016
VL 20
IS 6
BP 879
EP 885
DI 10.1007/s10029-016-1535-1
PG 7
WC Surgery
WE Science Citation Index Expanded (SCI-EXPANDED)
SC Surgery
GA EC5OS
UT WOS:000388186100014
PM 27601037
DA 2023-08-27
ER

PT J
AU Renard, D
   Labauge, P
   Vandenberghe, R
AF Renard, Dimitri
   Labauge, Pierre
   Vandenberghe, Rik
TI REVERSIBLE POSTERIOR LEUKOENCEPHALOPATHY SYNDROME
SO TRANSLATIONAL NEUROSCIENCE
LA English
DT Article
DE Drug-induced; Hypertensive; White matter; Cyclosporine; MRI; Brain;
   Immunosuppression
ID NERVOUS-SYSTEM TOXICITY; ENCEPHALOPATHY SYNDROME; CEREBRAL VASOSPASM;
   HYPERTENSIVE ENCEPHALOPATHY; CYCLOSPORINE NEUROTOXICITY; MR
   SPECTROSCOPY; TUMOR LYSIS; PATIENT; BRAIN; ABNORMALITIES
AB Reversible posterior leukoencephalopathy syndrome (RPLS) is characterized by subacute onset of headache, decreased alertness, vomiting, seizures, visuoperceptual disturbances, together with bilateral white matter lesions in posterior brain regions on brain imaging. The most frequently associated conditions related to RPLS are arterial hypertension and the use of immunosuppressive or cytotoxic treatment. T2-, Fluid Attenuation Inversion Recovery (FLAIR), and Apparent Diffusion Coefficient (ADC)-weighted magnetic resonance imaging (MRI) reveal hyperintensities in parietooccipital white matter but grey matter and other regions including frontal and temporal lobes, brainstem, cerebellum, basal ganglia, or even spinal cord may also be involved. According to ADC findings, the underlying pathophysiologic mechanism is probably one of vasogenic (rather than cytotoxic) oedema. These MRI findings help in differentiating RPLS from ischaemic events and other diseases resembling RPLS. Failure of cerebral autoregulation, endothelial dysfunction, disrupted blood-brain barrier, vasospasm, and direct toxic drug effects may all play a role in the pathophysiology of RPLS. Treatment consists of discontinuation of the causal drug, treatment of high blood pressure, and antiepileptic therapy. Clinical recovery and regression of radiological abnormalities are typically seen after early treatment. However, delay in diagnosis and treatment can result in irreversible brain damage, often in association with complicating cerebral infarction or haemorrhage.
C1 [Renard, Dimitri; Labauge, Pierre] Hop Caremeau, CHU Nimes, Dept Neurol, F-30029 Nimes, France.
   [Vandenberghe, Rik] Univ Ziekenhuis Gasthuisberg, Dept Neurol, B-3000 Louvain, Belgium.
C3 CHARMEU; Universite de Montpellier; CHU de Nimes; 1EUROPE; KU Leuven;
   University Hospital Leuven
RP Renard, D (通讯作者)，Hop Caremeau, CHU Nimes, Dept Neurol, Pl Pr Debre, F-30029 Nimes, France.
EM
RI Ottaviani, Giulia/B-4961-2014; Vandenberghe, Rik/AAR-7485-2020;
   Vandenberghe, Rik/K-2145-2014
OI Ottaviani, Giulia/0000-0002-6982-170X; Vandenberghe,
   Rik/0000-0001-6237-2502; Vandenberghe, Rik/0000-0001-6237-2502
CR Antunes NL, 2003, PEDIATR NEUROL, V28, P190, DOI 10.1016/S0887-8994(02)00508-8
   Bakshi R, 1999, NEUROLOGY, V52, P1285, DOI 10.1212/WNL.52.6.1285
   BHATT BD, 1988, NEW ENGL J MED, V318, P788
   Bianco F, 2005, NEURORADIOLOGY, V47, P703, DOI 10.1007/s00234-005-1403-7
   BLACK KS, 1995, AM J NEURORADIOL, V16, P1650
   Casey SO, 2000, AM J NEURORADIOL, V21, P243
   Casey SO, 2000, AM J NEURORADIOL, V21, P1199
   Chang GY, 1999, NEUROLOGY, V53, P652, DOI 10.1212/WNL.53.3.652
   Chen SP, 2006, CEPHALALGIA, V26, P530, DOI 10.1111/j.1468-2982.2005.01063.x
   Covarrubias DJ, 2002, AM J NEURORADIOL, V23, P1038
   Cruz-Flores S, 2004, NEUROLOGY, V62, P1417, DOI 10.1212/01.WNL.0000120668.73677.5F
   DEGROEN PC, 1988, NEW ENGL J MED, V318, P789
   DEGROEN PC, 1988, MAYO CLIN PROC, V63, P1012, DOI 10.1016/S0025-6196(12)64916-7
   DEGROEN PC, 1987, NEW ENGL J MED, V317, P861, DOI 10.1056/NEJM198710013171404
   DEKLIPPEL N, 1992, LANCET, V339, P1114, DOI 10.1016/0140-6736(92)90707-A
   Dodick DW, 2003, CEPHALALGIA, V23, P994, DOI 10.1046/j.1468-2982.2003.00577.x
   Doss-Esper CE, 2005, J NEUROIMAGING, V15, P188, DOI 10.1177/1051228404273820
   Eichler FS, 2002, AM J NEURORADIOL, V23, P833
   Filley CM, 2001, NEW ENGL J MED, V345, P425, DOI 10.1056/NEJM200108093450606
   GHALIE R, 1990, BONE MARROW TRANSPL, V6, P147
   Gijtenbeek JMM, 1999, J NEUROL, V246, P339, DOI 10.1007/s004150050360
   Greenwood MJ, 2003, LEUKEMIA LYMPHOMA, V44, P719, DOI 10.1080/1042819031000060582
   Hawley J S, 2004, J Postgrad Med, V50, P197
   Hefzy HM, 2009, AM J NEURORADIOL, V30, P1371, DOI 10.3174/ajnr.A1588
   Henderson RD, 2003, NEUROLOGY, V60, P326, DOI 10.1212/01.WNL.0000042095.49520.1E
   Hinchey J, 1996, NEW ENGL J MED, V334, P494, DOI 10.1056/NEJM199602223340803
   HUMPHREYS TR, 1993, J AM ACAD DERMATOL, V29, P490, DOI 10.1016/S0190-9622(08)82005-1
   Immink RV, 2004, CIRCULATION, V110, P2241, DOI 10.1161/01.CIR.0000144472.08647.40
   JOHANSSON BB, 1983, ACTA MED SCAND, P107
   Kaito E, 2005, PEDIATR RADIOL, V35, P722, DOI 10.1007/s00247-005-1434-6
   Kassem-Moussa H, 2000, AM J ROENTGENOL, V174, P1304, DOI 10.2214/ajr.174.5.1741304
   Kastrup O, 2002, J NEUROL, V249, P1563, DOI 10.1007/s00415-002-0895-x
   Kiemeneij IM, 2003, J NEUROL NEUROSUR PS, V74, P1126, DOI 10.1136/jnnp.74.8.1126
   Kinoshita T, 2003, CLIN IMAG, V27, P307, DOI 10.1016/S0899-7071(02)00592-2
   Lee SY, 2008, J CLIN NEUROSCI, V15, P457, DOI 10.1016/j.jocn.2006.12.019
   Lin JT, 2003, AM J NEURORADIOL, V24, P102
   LINDENBAUM J, 1996, CECIL TXB MED, P823
   Magnano MD, 2006, SEMIN ARTHRITIS RHEU, V35, P396, DOI 10.1016/j.semarthrit.2006.01.002
   MARK AL, 1990, NEW ENGL J MED, V323, P748, DOI 10.1056/NEJM199009133231109
   Masuhr E, 2004, J NEUROL, V251, P11, DOI 10.1007/s00415-004-0321-7
   MATSUMOTO K, 1995, INT J RADIAT ONCOL, V32, P913, DOI 10.1016/0360-3016(95)00565-G
   McKinney AM, 2009, AM J ROENTGENOL, V193, P192, DOI 10.2214/AJR.08.1176
   Min LL, 2006, SEMIN ARTHRITIS RHEU, V35, P388, DOI 10.1016/j.semarthrit.2006.01.003
   Miyazaki Y, 2004, INTERNAL MED, V43, P1005, DOI 10.2169/internalmedicine.43.1005
   NOZUE T, 1993, TRANSPLANTATION, V55, P346, DOI 10.1097/00007890-199302000-00022
   Obeid T, 1999, NEUROLOGY, V53, P1372, DOI 10.1212/WNL.53.6.1372
   Obeid T, 2004, SEIZURE-EUR J EPILEP, V13, P277, DOI 10.1016/S1059-1311(03)00152-3
   Ohta T, 2004, PEDIATR NEPHROL, V19, P442, DOI 10.1007/s00467-003-1286-y
   Omuro AMP, 2005, ARCH NEUROL-CHICAGO, V62, P1595, DOI 10.1001/archneur.62.10.1595
   Poon WL, 2005, ANN RHEUM DIS, V64, P1803, DOI 10.1136/ard.2005.038273
   Provenzale JM, 2001, AM J NEURORADIOL, V22, P1455
   Renard D, 2004, J NEUROL, V251, P226, DOI 10.1007/s00415-004-0287-5
   RENARD D, 2008, DRUG RELATED ENCEPHA, P293
   Russell MT, 2001, MAGN RESON IMAGING, V19, P129, DOI 10.1016/S0730-725X(01)00217-X
   Sengar AR, 1997, AM J NEURORADIOL, V18, P1485
   Sibai BM, 1996, NEW ENGL J MED, V335, P257, DOI 10.1056/NEJM199607253350407
   Singhal AB, 2004, ARCH NEUROL-CHICAGO, V61, P411, DOI 10.1001/archneur.61.3.411
   Tang-Wai DF, 2001, HEADACHE, V41, P198, DOI 10.1046/j.1526-4610.2001.111006198.x
   Thaipisuttikul I, 2005, J NEUROL, V252, P230, DOI 10.1007/s00415-005-0613-6
   THOMPSON CB, 1984, LANCET, V2, P1116
   TROMMER BL, 1988, STROKE, V19, P326, DOI 10.1161/01.STR.19.3.326
   Turkeltaub PE, 2009, NEUROLOGY, V73, pE30, DOI 10.1212/WNL.0b013e3181b1641e
   Ugurel MS, 2005, EUR J RADIOL, V53, P441, DOI 10.1016/j.ejrad.2004.05.015
   Vaughan CJ, 2000, LANCET, V356, P411, DOI 10.1016/S0140-6736(00)02539-3
   Voltz R, 1996, NEUROLOGY, V46, P250, DOI 10.1212/WNL.46.1.250
   Wennberg RA, 1998, EPILEPSIA, V39, P1381, DOI 10.1111/j.1528-1157.1998.tb01343.x
   Woolfenden AR, 1998, PEDIATR NEUROL, V19, P74, DOI 10.1016/S0887-8994(98)00027-7
   WORTHLEY SG, 1995, J RHEUMATOL, V22, P335
NR 68
TC 0
Z9 0
U1 0
U2 4
PU DE GRUYTER POLAND SP Z O O
PI WARSAW
PA BOGUMILA ZUGA 32A STR, 01-811 WARSAW, MAZOVIA, POLAND
SN 2081-3856
EI 2081-6936
J9 TRANSL NEUROSCI
JI Transl. Neurosci.
PD JUN
PY 2010
VL 1
IS 2
BP 115
EP 123
DI 10.2478/v10134-010-0016-9
PG 9
WC Neurosciences
WE Science Citation Index Expanded (SCI-EXPANDED)
SC Neurosciences & Neurology
GA V20PY
UT WOS:000208153100004
OA hybrid
DA 2023-08-27
ER

PT J
AU Zhu, BQ
   Sun, MY
   Yang, T
   Yu, HZ
   Wang, LM
AF Zhu, Bingqing
   Sun, Mengyang
   Yang, Ting
   Yu, Haizhen
   Wang, Limei
TI Clinical, imaging features and outcomes of patients with anti-GFAP
   antibodies: a retrospective study
SO FRONTIERS IN IMMUNOLOGY
LA English
DT Article
DE glial fibrillary astrocytic protein antibodies; clinical
   characteristics; imaging features; overlapping antibodies; prognosis
ID ACIDIC PROTEIN ASTROCYTOPATHY; CENTRAL-NERVOUS-SYSTEM; AUTOIMMUNITY
AB ObjectiveTo evaluate and compare the clinical features, imaging, overlapping antibodies, and prognosis of pediatric and adult patients with anti-GFAP antibodies. MethodsThis study included 59 patients with anti-GFAP antibodies (28 females and 31 males) who were admitted between December 2019 and September 2022. ResultsOut of 59 patients, 18 were children (under 18 years old), and 31 were adults. The overall cohort's median age at onset was 32 years old, 7 for children, and 42 for adults. There were 23 (41.1%) patients with prodromic infection, 1 (1.7%) patient with a tumor, 29 (53.7%) patients with other non-neurological autoimmune diseases, and 17 (22.8%) patients with hyponatremia. Fourteen (23.7%) patients had multiple neural autoantibodies, with the AQP4 antibody being the most common. Encephalitis (30.5%) was the most common phenotypic syndrome. Common clinical symptoms included fever (59.3%), headache (47.5%), nausea and vomiting (35.6%), limb weakness (35.6%), and disturbance of consciousness (33.9%). Brain MRI lesions were primarily located in the cortex/subcortex (37.3%), brainstem (27.1%), thalamus (23.7%), and basal ganglia (22.0%). Spinal cord MRI lesions often involved the cervical and thoracic spinal cord. There was no statistically significant difference in the MRI lesion site between children and adults. Out of 58 patients, 47 (81.0%) had a monophasic course, and 4 died. The last follow-up showed that 41/58 (80.7%) patients had an improved functional outcome (mRS <3), and children were more likely than adults to have no residual disability symptoms (p = 0.001). ConclusionThere was no statistically significant difference in clinical symptoms and imaging findings between children and adult patients with anti-GFAP antibodies; Patients with anti-GFAP antibodies may present with normal MRI findings or delayed MRI abnormalities, and patients with overlapping antibodies were common. Most patients had monophasic courses, and those with overlapping antibodies were more likely to relapse. Children were more likely than adults to have no disability. Finally, we hypothesize that the presence of anti-GFAP antibodies is a non-specific witness of inflammation.
C1 [Zhu, Bingqing; Sun, Mengyang; Yang, Ting; Yu, Haizhen; Wang, Limei] Zhengzhou Univ, Affiliated Hosp 1, Dept Neurol, Zhengzhou, Henan, Peoples R China.
C3 Zhengzhou University
RP Wang, LM (通讯作者)，Zhengzhou Univ, Affiliated Hosp 1, Dept Neurol, Zhengzhou, Henan, Peoples R China.
EM
RI Sun, Mengyang/AAP-5033-2020
OI Sun, Mengyang/0000-0002-3998-8744
CR Deng B, 2022, NEUROL-NEUROIMMUNOL, V9, DOI 10.1212/NXI.0000000000200029
   Dubey D, 2018, J NEUROIMMUNOL, V321, P157, DOI 10.1016/j.jneuroim.2018.04.016
   Fang BY, 2016, JAMA NEUROL, V73, P1297, DOI 10.1001/jamaneurol.2016.2549
   Fang HJ, 2021, FRONT IMMUNOL, V12, DOI 10.3389/fimmu.2021.761354
   Flanagan EP, 2017, ANN NEUROL, V81, P298, DOI 10.1002/ana.24881
   Gravier-Dumonceau A, 2022, NEUROLOGY, V98, pE653, DOI 10.1212/WNL.0000000000013087
   Heraud C, 2022, NEUROL THER, V11, P499, DOI 10.1007/s40120-021-00302-y
   Imanaka Shohei, 2022, eNeurologicalSci, V27, P100403, DOI 10.1016/j.ensci.2022.100403
   Iorio R, 2018, J NEUROL NEUROSUR PS, V89, P138, DOI 10.1136/jnnp-2017-316583
   Izumi M, 2023, INTERNAL MED, V62, P465, DOI 10.2169/internalmedicine.9724-22
   Kimura A, 2019, J NEUROIMMUNOL, V332, P91, DOI 10.1016/j.jneuroim.2019.04.004
   Kunchok A, 2019, CURR OPIN NEUROL, V32, P452, DOI 10.1097/WCO.0000000000000676
   Li DY, 2020, GLIA, V68, P878, DOI 10.1002/glia.23734
   Li JA, 2021, MEDICINE, V100, DOI 10.1097/MD.0000000000026448
   Li J, 2018, MULT SCLER RELAT DIS, V21, P84, DOI 10.1016/j.msard.2018.02.020
   Long Y, 2018, EUR J NEUROL, V25, P477, DOI 10.1111/ene.13531
   Natori T, 2022, INTERNAL MED, V61, P2947, DOI 10.2169/internalmedicine.8964-21
   Oger V, 2020, EUR J PAEDIATR NEURO, V26, P89, DOI 10.1016/j.ejpn.2020.03.002
   Olabarria M, 2017, ANNU REV PATHOL-MECH, V12, P131, DOI 10.1146/annurev-pathol-052016-100218
   Quek AML, 2022, INT J INFECT DIS, V124, P164, DOI 10.1016/j.ijid.2022.09.029
   Sakashita Y, 2022, CLIN NEUROL NEUROSUR, V218, DOI 10.1016/j.clineuro.2022.107272
   Sasaki K, 2014, J IMMUNOL, V192, P3029, DOI 10.4049/jimmunol.1302911
   Shan FL, 2018, FRONT IMMUNOL, V9, DOI 10.3389/fimmu.2018.02802
   Wang KKW, 2016, J NEUROTRAUM, V33, P1270, DOI 10.1089/neu.2015.3881
   Xiao J, 2021, J NEUROIMMUNOL, V360, DOI 10.1016/j.jneuroim.2021.577718
   Yamakawa M, 2021, NEUROL-NEUROIMMUNOL, V8, DOI 10.1212/NXI.0000000000001081
   Yang X, 2020, MULT SCLER RELAT DIS, V45, DOI 10.1016/j.msard.2020.102350
   Yang XG, 2018, FRONT NEUROL, V9, DOI 10.3389/fneur.2018.00251
   Yang XG, 2017, NEUROIMMUNOMODULAT, V24, P113, DOI 10.1159/000479948
   Yuan ZM, 2021, EUR J NEUROL, V28, P2121, DOI 10.1111/ene.14778
NR 30
TC 0
Z9 0
U1 0
U2 0
PU FRONTIERS MEDIA SA
PI LAUSANNE
PA AVENUE DU TRIBUNAL FEDERAL 34, LAUSANNE, CH-1015, SWITZERLAND
SN 1664-3224
J9 FRONT IMMUNOL
JI Front. Immunol.
PD MAY 2
PY 2023
VL 14
AR 1106490
DI 10.3389/fimmu.2023.1106490
PG 14
WC Immunology
WE Science Citation Index Expanded (SCI-EXPANDED)
SC Immunology
GA G4ND4
UT WOS:000988932600001
PM 37205100
OA Green Published, gold
DA 2023-08-27
ER

PT J
AU Babbage, MJ
   Feldman, MB
   O'Beirne, GA
   MacFarlane, MR
   Bird, PA
AF Babbage, Melissa J.
   Feldman, Melanie B.
   O'Beirne, Greg A.
   MacFarlane, Martin R.
   Bird, Philip A.
TI Patterns of Hearing Loss Following Retrosigmoid Excision of Unilateral
   Vestibular Schwannoma
SO JOURNAL OF NEUROLOGICAL SURGERY PART B-SKULL BASE
LA English
DT Article
DE acoustic neuroma; vestibular schwannoma; hearing preservation
ID AUDITORY-EVOKED-POTENTIALS; CEREBELLOPONTINE ANGLE MANIPULATIONS;
   ACOUSTIC TUMOR SURGERY; COCHLEAR NERVE; POSTOPERATIVE HEARING;
   PRESERVATION; COMPRESSION; MECHANISMS
AB Objectives To determine the pattern of auditory responses, time-course of hearing deterioration, and possible site of lesion following retrosigmoid excision of unilateral vestibular schwannomas.
   Design Prospective, nonrandomized, observational pilot study.
   Setting Tertiary referral medical center.
   Main outcome measures Preoperative and postoperative pure-tone and speech audiometry, auditory brainstem response testing, and distortion product otoacoustic emissions were performed in 20 patients. Testing was conducted every 24 hours for the duration of hospitalization. Transtympanic electrocochleography was performed if delayed deterioration of auditory responses was documented.
   Results Of the 20 patients, 7 had no discernible cochlear nerve at the end of the procedure. Of the 13 patients with an intact nerve, 6 retained hearing, 3 with evidence of reduced neural function. Of the 7 who lost hearing despite an intact nerve, 5 lost at least cochlear and possibly also neural function, and 1 had reduced neural function but retained cochlear function. There were two examples of delayed deterioration of cochlear nerve responses.
   Conclusions Hearing loss following retrosigmoid removal of vestibular schwannomas most often involves loss of at least cochlear function, possibly in addition to neural damage. In a smaller number of cases anacusis or hearing deterioration can be attributed to purely neural trauma.
C1 [Babbage, Melissa J.; Feldman, Melanie B.; O'Beirne, Greg A.; Bird, Philip A.] Univ Canterbury, Dept Commun Disorders, Christchurch 8140, New Zealand.
   [MacFarlane, Martin R.] Univ Otago, Dept Neurosurg, Christchurch, New Zealand.
   [Bird, Philip A.] Christchurch Publ Hosp, Dept Otolaryngol Head & Neck Surg, Christchurch, New Zealand.
C3 University of Canterbury; University of Otago; Christchurch Hospital New
   Zealand
RP O'Beirne, GA (通讯作者)，Univ Canterbury, Dept Commun Disorders, Private Bag 4800, Christchurch 8140, New Zealand.
EM
RI O'Beirne, Greg A/I-3838-2012
OI O'Beirne, Greg A/0000-0002-3545-4630
FU Oticon Foundation in New Zealand
FX This work was supported by grants from the Oticon Foundation in New
   Zealand. The authors would like to thank Mr Jeremy Hornibrook and John
   Gourley for carrying out the transtympanic ECochG measurements, and Dr
   Angela Butler for her assistance in the early stages of this work.
CR Brackmann DE, 2000, AM J OTOL, V21, P417, DOI 10.1016/S0196-0709(00)80054-X
   CACACE AT, 1994, EAR HEARING, V15, P287, DOI 10.1097/00003446-199408000-00003
   Colletti V, 1997, OTOLARYNG HEAD NECK, V117, P596, DOI 10.1016/S0194-5998(97)70039-1
   Colletti V, 1998, AUDIOLOGY, V37, P27
   DORNHOFFER JL, 1995, LARYNGOSCOPE, V105, P184, DOI 10.1288/00005537-199502000-00014
   Friedman RA, 1998, OTOLARYNG HEAD NECK, V119, P588, DOI 10.1016/S0194-5998(98)70016-6
   Galambos R, 1978, EVOKED ELECTRICAL AC, P405
   GARDNER G, 1988, ANN OTO RHINOL LARYN, V97, P55, DOI 10.1177/000348948809700110
   HASHIMOTO I, 1981, BRAIN, V104, P841, DOI 10.1093/brain/104.4.841
   Hatayama T, 1999, NEUROL RES, V21, P599, DOI 10.1080/01616412.1999.11740983
   JOSEY AF, 1988, ANN OTO RHINOL LARYN, V97, P626, DOI 10.1177/000348948809700609
   Kim AH, 2006, OTOL NEUROTOL, V27, P372, DOI 10.1097/00129492-200604000-00014
   LEVINE RA, 1984, ANN OTO RHINOL LARYN, V93, P116, DOI 10.1177/000348948409300203
   MONSELL EM, 1995, OTOLARYNG HEAD NECK, V113, P179
   Morlet T, 2003, AM J OTOLARYNG, V24, P297, DOI 10.1016/S0196-0709(03)00057-7
   PERLMAN H B, 1959, Laryngoscope, V69, P591
   Robinette MS, 1997, AM J OTOL, V18, P738
   SABIN HI, 1987, SCAND AUDIOL, V16, P109, DOI 10.3109/01050398709042164
   SEKIYA T, 1987, J NEUROSURG, V67, P244, DOI 10.3171/jns.1987.67.2.0244
   SEKIYA T, 1986, ACTA NEUROCHIR, V81, P142, DOI 10.1007/BF01401237
   SEKIYA T, 1985, J NEUROSURG, V63, P598, DOI 10.3171/jns.1985.63.4.0598
   Sekiya T, 2002, NEUROL RES, V24, P775, DOI 10.1179/016164102101200889
   Shimamura N, 2002, J NEUROSURG, V97, P929, DOI 10.3171/jns.2002.97.4.0929
   Strauss C, 2001, J NEUROSURG, V95, P771, DOI 10.3171/jns.2001.95.5.0771
NR 24
TC 3
Z9 3
U1 1
U2 6
PU THIEME MEDICAL PUBL INC
PI NEW YORK
PA 333 SEVENTH AVE, NEW YORK, NY 10001 USA
SN 2193-6331
EI 2193-634X
J9 J NEUROL SURG PART B
JI Journal of Neurol. Surg. Part B
PD JUN
PY 2013
VL 74
IS 3
BP 166
EP 175
DI 10.1055/s-0033-1342921
PG 10
WC Clinical Neurology; Surgery
WE Science Citation Index Expanded (SCI-EXPANDED)
SC Neurosciences & Neurology; Surgery
GA 175ZQ
UT WOS:000321272900011
PM 24436908
OA Green Published
DA 2023-08-27
ER

PT J
AU Edmondson, MC
   Day, R
   Wood, D
AF Edmondson, M. C.
   Day, R.
   Wood, D.
TI Vancomycin iontophoresis of allograft bone
SO BONE & JOINT RESEARCH
LA English
DT Article
DE Vancomycin; Iontophoresis; Allograft; Bone; Infection
ID TOTAL HIP-REPLACEMENT; SEGMENTAL ALLOGRAFTS; CANCELLOUS BONE; INFECTION;
   SURGERY; RECONSTRUCTION; REVISION; TUMOR; ARTHROPLASTY; MANAGEMENT
AB Objectives
   The most concerning infection of allografts and operative procedures is methicillin resistant Staphylococcus aureus (MRSA) and no current iontophoresed antibiotics effectively combat this microbe. It was initially hypothesised that iontophoresis of vancomycin through bone would not be effective due to its large molecular size and lack of charge. The aim of this study was to determine whether this was a viable procedure and to find the optimum conditions for its use.
   Methods
   An iontophoresis cell was set up with varying concentrations of Vancomycin within the medulla of a section of sheep tibia, sealed from an external saline solution. The cell was run for varying times, Vancomycin concentrations and voltages, to gain information on optimisation of conditions for impregnating the graft. Each graft was then sectioned and dust ground from the exposed surface. The dust was serially washed to extract the Vancomycin and concentrations measured and plotted for all variables tested.
   Results
   Vancomycin was successfully delivered and impregnated to the graft using the iontophoresis technique. The first order fit to the whole data set gave a significant result (p = 0.0233), with a significant concentration (p = 0.02774) component. The time component was the next most significant (p = 0.0597), but did not exceed the 95% confidence level.
   Conclusions
   Iontophoresis is an effective method for delivering Vancomycin to allograft bone. The concentrations of the vancomycin solution affected the bone concentration, but results were highly variable. Further study should be done on the effectiveness of delivering different antibiotics using this method.
C1 [Edmondson, M. C.; Day, R.; Wood, D.] Royal Perth Hosp, Dept Bioengn, Perth, WA 6001, Australia.
C3 Royal Perth Hospital; University of Western Australia
RP Edmondson, MC (通讯作者)，Royal Perth Hosp, Perth Bone & Tissue Bank, Wellington St, Perth, WA 6001, Australia.
EM
RI Day, Robert/O-6379-2019; Day, Robert E/B-5325-2016
OI Day, Robert/0000-0003-3442-2298; Day, Robert E/0000-0003-3442-2298
FU NHMRC [513865]
FX This work was partially supported by NH&MRC grant 513865
CR Alexeeff M, 1996, J BONE JOINT SURG BR, V78B, P213, DOI 10.1302/0301-620X.78B2.0780213
   Ammon P, 2004, J BONE JOINT SURG BR, V86B, P962, DOI 10.1302/0301-620X.86B7.14292
   BERREY BH, 1990, J BONE JOINT SURG AM, V72A, P825, DOI 10.2106/00004623-199072060-00005
   BERRY DJ, 1991, J BONE JOINT SURG AM, V73A, P1460, DOI 10.2106/00004623-199173100-00003
   CARA JA, 1994, ACTA ORTHOP SCAND, V65, P42, DOI 10.3109/17453679408993716
   Chan YS, 1998, J TRAUMA, V45, P758, DOI 10.1097/00005373-199810000-00023
   CLOHISY DR, 1994, J BONE JOINT SURG AM, V76A, P549, DOI 10.2106/00004623-199404000-00009
   Day RE, 2005, J BONE JOINT SURG BR, V87B, P1568, DOI 10.1302/0301-620X.87B11.16384
   DELLOYE C, 1988, ARCH ORTHOP TRAUM SU, V107, P31
   DICK HM, 1994, CLIN ORTHOP RELAT R, P46
   Dion N, 2002, J BONE JOINT SURG AM, V84A, P644, DOI 10.2106/00004623-200204000-00024
   Edwards SA, 2003, J ARTHROPLASTY, V18, P852, DOI 10.1016/S0883-5403(03)00281-X
   Haddad FS, 2000, J BONE JOINT SURG BR, V82B, P830, DOI 10.1302/0301-620X.82B6.10485
   HENRY SL, 1995, CLIN PHARMACOKINET, V29, P36, DOI 10.2165/00003088-199529010-00005
   JOFE MH, 1988, J BONE JOINT SURG AM, V70A, P507, DOI 10.2106/00004623-198870040-00005
   Khoo PPC, 2006, J BONE JOINT SURG BR, V88B, P1149, DOI 10.1302/0301-620X.88B9.17500
   LEIGH DA, 1982, J ANTIMICROB CHEMOTH, V9, P303, DOI 10.1093/jac/9.4.303
   Lenth RV, 2009, J STAT SOFTW, V32, P1
   LORD CF, 1988, J BONE JOINT SURG AM, V70A, P369, DOI 10.2106/00004623-198870030-00008
   Mankin HJ, 1996, CLIN ORTHOP RELAT R, P86
   MCGANN W, 1986, J BONE JOINT SURG AM, V68A, P4, DOI 10.2106/00004623-198668010-00002
   Michalak KA, 2006, J BONE JOINT SURG BR, V88B, P1430, DOI 10.1302/0301-620X.88B11.18335
   MUSCOLO DL, 1993, J BONE JOINT SURG AM, V75A, P1656, DOI 10.2106/00004623-199311000-00010
   Toms AD, 2004, J BONE JOINT SURG AM, V86A, P2050, DOI 10.2106/00004623-200409000-00028
   Witso E, 2000, ACTA ORTHOP SCAND, V71, P80, DOI 10.1080/00016470052943955
NR 25
TC 3
Z9 3
U1 1
U2 3
PU BRITISH EDITORIAL SOC BONE JOINT SURGERY
PI LONDON
PA 22 BUCKINGHAM STREET, LONDON WC2N 6ET, ENGLAND
SN 2046-3758
J9 BONE JOINT RES
JI Bone Jt. Res.
PD APR
PY 2014
VL 3
IS 4
BP 101
EP 107
DI 10.1302/2046-3758.34.2000223
PG 7
WC Cell & Tissue Engineering; Orthopedics
WE Science Citation Index Expanded (SCI-EXPANDED)
SC Cell Biology; Orthopedics
GA AT2KG
UT WOS:000344762900003
PM 24729101
OA gold, Green Published, Green Submitted
DA 2023-08-27
ER

PT J
AU Kuntz, L
   Matthis, R
   Wegner, N
   Lutz, S
AF Kuntz, L.
   Matthis, R.
   Wegner, N.
   Lutz, S.
TI Dosimetric comparison of mono-isocentric and multi-isocentric plans for
   oligobrain metastases: A single institutional experience
SO CANCER RADIOTHERAPIE
LA English
DT Article
DE Brain metastases; Stereotactic radiotherapy; VMAT; Arctherapy;
   Monoisocenter
ID BRAIN METASTASES; STEREOTACTIC RADIOSURGERY; CYBERKNIFE RADIOTHERAPY;
   RADIATION NECROSIS; RADIONECROSIS; VOLUME; RISK; EPIDEMIOLOGY;
   CONFORMITY; TOLERANCE
AB Pupose. - To compare dosimetric plans for the treatments of oligobrains metastases (26) using mono-isocentric arc therapy and multi-isocentric volumetric modulated arc therapy (VMAT).
   Material and methods. - A total of sixteen patients with multiple brain metastases were selected. Prescription dose was between 24 and 15 Gy depending on the tumor size. For every patient,arctherapy and VMAT plans were generated respectively, with Elements Multiple Brain Mets SRS version 2.0 (BrainLab) and Eclipse SRS Treatment Planning Systeme version 15.5. The conformity index (CI), homogeinity index (HI), gradient index (GI), dose volume histogram for each organs at risk, total Monitor Units were evaluated.
   Results. - For coverage of the PTV, mono-isocentric plans showed a better CI and a better GI than multi-isocentric plans, respectively CI of 1.18 +/- 0.11 vs 1.41 +/- 0.20 (P < 0.01), and GI of 3.55 +/- 0.59 vs 4.03 +/- 1.20 (P < 0.01). Homogeneity index was not better with mono-isocentric plans, with respectively HI 24.32 +/- 3.87 vs 14.05 +/- 4.46 (P = 1). For organs at risk, there were no statistical differences between mono and multi-isocentric plans for both eyes, both lenses, both optic nerves, chiasma, brainstem, and hippocampi. V-12Gy and V-10Gy of normal brain were statistically lower with mono-isocentric plans than with multi-isocentric plans, respectivelly V-12Gy of 3.06 Gy 95% CI [2.25;3.86]vs 5.18 Gy 95% CI [3.43;6.93] (P < 0,01) and V-10Gy 4.66 Gy 95% CI [3.33;5.98] vs7.30 Gy 95% CI [4.73;9.87] (P < 0.03). Total number of MU was significantly lower with mono-isocentric plans than with multi-isocentric plans, respectively 6668 +/- 1463 vs 12403 +/- 4941 (P < 0.01), then treatment time was lower with mono-isocentric plans.
   Conclusion. - Mono-isocentric plans had a better conformity index and gradient index than multi-isocentric plans for the treatment of multiple brain metastases. Moreover, mono-isocentric techniques gave fewer doses to normal brain and used less monitor units than multi-isocentric techniques. (C) 2020 Societe francaise de radiotherapie oncologique (SFRO). Published by Elsevier Masson SAS. Allrights reserved.
C1 [Kuntz, L.] Univ Strasbourg, Ctr Paul Strauss, Dept Radiat Oncol, Strasbourg, France.
   [Matthis, R.; Wegner, N.; Lutz, S.] Klinikum Stuttgart, Dept Radiat Oncol, Stuttgart, Germany.
C3 UDICE-French Research Universities; Universites de Strasbourg
   Etablissements Associes; Universite de Strasbourg; UNICANCER; Centre
   Paul Strauss; Klinikum Stuttgart
RP Kuntz, L (通讯作者)，Univ Strasbourg, Ctr Paul Strauss, Dept Radiat Oncol, Strasbourg, France.
EM
CR [Anonymous], 2010, J ICRU, V10, P83, DOI 10.1093/jicru/ndq012
   [Anonymous], 2014, J INT COMM RAD UNITS, V14, P101, DOI [10.1093/jicru_ndx010, DOI 10.1093/JICRU_NDX010]
   [Anonymous], [No title captured]
   Benedict SH, 2010, MED PHYS, V37, P4078, DOI 10.1118/1.3438081
   Blonigen BJ, 2010, INT J RADIAT ONCOL, V77, P996, DOI 10.1016/j.ijrobp.2009.06.006
   Chargari C, 2016, CANCER RADIOTHER, V20, pS1
   Chin LS, 2001, J NEUROSURG, V94, P899, DOI 10.3171/jns.2001.94.6.0899
   Grimm J, 2011, J APPL CLIN MED PHYS, V12, P267, DOI 10.1120/jacmp.v12i2.3368
   Inoue HK, 2014, J RADIAT RES, V55, P334, DOI 10.1093/jrr/rrt127
   Inoue HK, 2013, J RADIAT RES, V54, P727, DOI 10.1093/jrr/rrt006
   Kirkpatrick JP, 2011, J RADIOSURGERY SBRT, V1, P95
   Kirkpatrick JP, 2015, INT J RADIAT ONCOL, V91, P100, DOI 10.1016/j.ijrobp.2014.09.004
   Kohutek ZA, 2015, J NEURO-ONCOL, V125, P149, DOI 10.1007/s11060-015-1881-3
   Le Rhun E, 2016, EXPERT REV NEUROTHER, V16, P903, DOI 10.1080/14737175.2016.1184572
   Mayo C, 2010, INT J RADIAT ONCOL, V76, pS36, DOI 10.1016/j.ijrobp.2009.08.078
   Minniti G, 2011, RADIAT ONCOL, V6, DOI 10.1186/1748-717X-6-48
   Nabors LB, 2014, J NATL COMPR CANC NE, V12, P1517, DOI 10.6004/jnccn.2014.0151
   Narayanasamy G, 2013, MED PHYS, V40, DOI 10.1118/1.4818825
   Nayak L, 2012, CURR ONCOL REP, V14, P48, DOI 10.1007/s11912-011-0203-y
   Paddick I, 2000, J NEUROSURG, V93, P219, DOI 10.3171/jns.2000.93.supplement_3.0219
   Paddick I, 2006, J NEUROSURG, V105, P194, DOI 10.3171/sup.2006.105.7.194
   Scoccianti S, 2015, RADIOTHER ONCOL, V114, P230, DOI 10.1016/j.radonc.2015.01.016
   Shaw E, 2000, INT J RADIAT ONCOL, V47, P291, DOI 10.1016/S0360-3016(99)00507-6
   Taillibert S, 2015, CANCER RADIOTHER, V19, P3, DOI 10.1016/j.canrad.2014.11.001
   TISHLER RB, 1993, INT J RADIAT ONCOL, V27, P215, DOI 10.1016/0360-3016(93)90230-S
   Tsao MN, 2018, COCHRANE DB SYST REV, DOI 10.1002/14651858.CD003869.pub4
   Yamamoto M, 2014, LANCET ONCOL, V15, P387, DOI 10.1016/S1470-2045(14)70061-0
NR 27
TC 7
Z9 7
U1 1
U2 2
PU ELSEVIER
PI BRIDGEWATER
PA 685 ROUTE 202-206, BRIDGEWATER, NJ 08807 USA
SN 1278-3218
EI 1769-6658
J9 CANCER RADIOTHER
JI Cancer Radiother.
PD FEB
PY 2020
VL 24
IS 1
BP 53
EP 59
DI 10.1016/j.canrad.2019.10.003
PG 7
WC Oncology; Radiology, Nuclear Medicine & Medical Imaging
WE Science Citation Index Expanded (SCI-EXPANDED)
SC Oncology; Radiology, Nuclear Medicine & Medical Imaging
GA LG3FI
UT WOS:000527990500008
PM 32046915
OA Bronze, Green Published
DA 2023-08-27
ER

PT J
AU Howarth, A
   Madureira, PA
   Lockwood, G
   Storer, LCD
   Grundy, R
   Rahman, R
   Pilkington, GJ
   Hill, R
AF Howarth, Alison
   Madureira, Patricia A.
   Lockwood, George
   Storer, Lisa C. D.
   Grundy, Richard
   Rahman, Ruman
   Pilkington, Geoffrey J.
   Hill, Richard
TI Modulating autophagy as a therapeutic strategy for the treatment of
   paediatric high-grade glioma
SO BRAIN PATHOLOGY
LA English
DT Review
DE autophagy; paediatric high grade glioma
ID DEPENDENT PROTEIN-KINASE; DNA-DAMAGE RESPONSE; COMPREHENSIVE GENOMIC
   CHARACTERIZATION; CHAPERONE-MEDIATED AUTOPHAGY; INTRINSIC PONTINE
   GLIOMAS; CELL-DEATH; MALIGNANT GLIOMA; PHASE-I; PROMOTER METHYLATION;
   TARGETING AUTOPHAGY
AB Paediatric high-grade gliomas (pHGG) represent a therapeutically challenging group of tumors. Despite decades of research, there has been minimal improvement in treatment and the clinical prognosis remains poor. Autophagy, a highly conserved process for recycling metabolic substrates is upregulated in pHGG, promoting tumor progression and evading cell death. There is significant crosstalk between autophagy and a plethora of critical cellular pathways, many of which are dysregulated in pHGG. The following article will discuss our current understanding of autophagy signaling in pHGG and the potential modulation of this network as a therapeutic target.
C1 [Howarth, Alison; Madureira, Patricia A.; Pilkington, Geoffrey J.; Hill, Richard] Univ Portsmouth, IBBS, Brain Tumour Res Ctr, Portsmouth PO1 2DT, Hants, England.
   [Madureira, Patricia A.] Univ Algarve, Ctr Biomed Res CBMR, Faro, Portugal.
   [Lockwood, George; Storer, Lisa C. D.; Grundy, Richard; Rahman, Ruman] Univ Nottingham, Sch Med, Childrens Brain Tumour Res Ctr, Queens Med Ctr, Nottingham, England.
C3 University of Portsmouth; Universidade do Algarve; University of
   Nottingham
RP Hill, R (通讯作者)，Univ Portsmouth, IBBS, Brain Tumour Res Ctr, Portsmouth PO1 2DT, Hants, England.
EM
RI Rahman, Ruman/FFR-5799-2022; Hill, Richard J/F-4875-2012; Madureira,
   Patricia A/C-9666-2013
OI Rahman, Ruman/0000-0002-6541-9983; Madureira, Patricia
   A/0000-0002-4856-3908; Hill, Richard/0000-0003-0394-6048; Grundy,
   Richard/0000-0002-2585-2539
FU Ollie Young Foundation; Brain Tumour Research; FCT Investigator contract
   from the Foundation for Science and Technology (FCT), Portugal
   [IF/00614/2014]; FCT exploratory grant [IF/00614/2014/CP12340006]; FCT
   Research Center Grant [UID/BIM/04773/2013CBMR1334]
FX GJP was awarded grant funding from the Ollie Young Foundation and Brain
   Tumour Research, AH was funded by the Ollie Young Foundation and RH,
   Brain Tumour Research. P.A.M. is funded by FCT Investigator contract
   from the Foundation for Science and Technology (FCT), Portugal
   (ref:IF/00614/2014) and FCT exploratory grant,
   ref:IF/00614/2014/CP12340006. CBMR is financed by FCT Research Center
   Grant ref:UID/BIM/04773/2013CBMR1334.
CR Alers S, 2012, MOL CELL BIOL, V32, P2, DOI 10.1128/MCB.06159-11
   Ballou Lisa M, 2008, J Chem Biol, V1, P27, DOI 10.1007/s12154-008-0003-5
   Bao SD, 2006, NATURE, V444, P756, DOI 10.1038/nature05236
   Basto D, 2005, ACTA NEUROPATHOL, V109, P207, DOI 10.1007/s00401-004-0936-x
   Bax DA, 2010, CLIN CANCER RES, V16, P3368, DOI 10.1158/1078-0432.CCR-10-0438
   Bax DA, 2009, CLIN CANCER RES, V15, P5753, DOI 10.1158/1078-0432.CCR-08-3210
   Bax DA, 2009, PLOS ONE, V4, DOI 10.1371/journal.pone.0005209
   Beagle B, 2011, CANCER CELL, V19, P693, DOI 10.1016/j.ccr.2011.05.020
   Becher OJ, 2008, J CHILD NEUROL, V23, P1205, DOI 10.1177/0883073808321766
   Bednarczyk Martyna, 2018, Oncotarget, V9, P34413, DOI 10.18632/oncotarget.26126
   Booth LA, 2014, CELL SIGNAL, V26, P549, DOI 10.1016/j.cellsig.2013.11.028
   Cancer Genome Atlas Research Network, 2011, Nature, V474, P609, DOI 10.1038/nature10166
   Cao YJ, 2019, CLIN CHIM ACTA, V489, P10, DOI 10.1016/j.cca.2018.11.028
   Castel D, 2015, ACTA NEUROPATHOL, V130, P815, DOI 10.1007/s00401-015-1478-0
   Chalhoub N, 2009, ANNU REV PATHOL-MECH, V4, P127, DOI 10.1146/annurev.pathol.4.110807.092311
   Cheong H, 2011, P NATL ACAD SCI USA, V108, P11121, DOI 10.1073/pnas.1107969108
   Chin L, 2008, NATURE, V455, P1061, DOI 10.1038/nature07385
   Cohen KJ, 2011, NEURO-ONCOLOGY, V13, P317, DOI 10.1093/neuonc/noq191
   Cuervo AM, 2014, CELL RES, V24, P92, DOI 10.1038/cr.2013.153
   Cully M, 2006, NAT REV CANCER, V6, P184, DOI 10.1038/nrc1819
   Czarny P, 2015, INT J MOL SCI, V16, P2641, DOI 10.3390/ijms16022641
   Dahiya S, 2014, NEURO-ONCOLOGY, V16, P318, DOI 10.1093/neuonc/not146
   Daido S, 2005, CANCER RES, V65, P4368, DOI 10.1158/0008-5472.CAN-04-4202
   de Narvajas AAM, 2013, MOL CELL BIOL, V33, P3983, DOI 10.1128/MCB.00813-13
   Diaz AK, 2014, SEMIN RADIAT ONCOL, V24, P240, DOI 10.1016/j.semradonc.2014.06.003
   Donson AM, 2007, PEDIATR BLOOD CANCER, V48, P403, DOI 10.1002/pbc.20803
   Eliopoulos AG, 2016, FRONT GENET, V7, DOI 10.3389/fgene.2016.00204
   Fangusaro J, 2012, FRONT ONCOL, V2, DOI 10.3389/fonc.2012.00105
   Feng DR, 2018, J CLIN INVEST, V128, P3941, DOI 10.1172/JCI99217
   Feng YC, 2014, CELL RES, V24, P24, DOI 10.1038/cr.2013.168
   Fitzwalter BE, 2015, FEBS J, V282, P4279, DOI 10.1111/febs.13515
   Fouladi M, 2015, CLIN CANCER RES, V21, P1558, DOI 10.1158/1078-0432.CCR-14-0595
   Fruman DA, 2014, NAT REV DRUG DISCOV, V13, P140, DOI 10.1038/nrd4204
   Fullgrabe J, 2014, NAT REV MOL CELL BIO, V15, P65, DOI 10.1038/nrm3716
   Gallia GL, 2006, MOL CANCER RES, V4, P709, DOI 10.1158/1541-7786.MCR-06-0172
   Geoerger B, 2012, EUR J CANCER, V48, P253, DOI 10.1016/j.ejca.2011.09.021
   Giatromanolaki A, 2014, CANCER BIOL THER, V15, P1468, DOI 10.4161/15384047.2014.955719
   Gottardo NG, 2008, J CHILD NEUROL, V23, P1149, DOI 10.1177/0883073808321765
   Green DR, 2014, CELL, V157, P65, DOI 10.1016/j.cell.2014.02.049
   Grundy RG, 2010, EUR J CANCER, V46, P120, DOI 10.1016/j.ejca.2009.09.013
   Gurung RL, 2014, MOL CANCER, V13, DOI 10.1186/1476-4598-13-232
   Gururangan S, 2010, J CLIN ONCOL, V28, P3069, DOI 10.1200/JCO.2009.26.8789
   Hakem R, 2008, EMBO J, V27, P589, DOI 10.1038/emboj.2008.15
   Halazonetis TD, 2008, SCIENCE, V319, P1352, DOI 10.1126/science.1140735
   Hammerman PS, 2012, NATURE, V489, P519, DOI 10.1038/nature11404
   Hargrave D, 2008, BRIT J NEUROSURG, V22, P624, DOI 10.1080/02688690802484405
   Harhaji-Trajkovic L, 2009, J CELL MOL MED, V13, P3644, DOI 10.1111/j.1582-4934.2009.00663.x
   Harper JW, 2007, MOL CELL, V28, P739, DOI 10.1016/j.molcel.2007.11.015
   He RN, 2015, AUTOPHAGY, V11, P740, DOI 10.1080/15548627.2015.1034404
   Hegi ME, 2005, NEW ENGL J MED, V352, P997, DOI 10.1056/NEJMoa043331
   Hennessy BT, 2005, NAT REV DRUG DISCOV, V4, P988, DOI 10.1038/nrd1902
   Hill R, 2011, ONCOTARGET, V2, P1094, DOI 10.18632/oncotarget.378
   Hoffman LM, 2015, NEURO-ONCOLOGY, V17, P39
   Hou W, 2010, AUTOPHAGY, V6, P891, DOI 10.4161/auto.6.7.13038
   Hu YL, 2012, CANCER RES, V72, P1773, DOI 10.1158/0008-5472.CAN-11-3831
   Huang QC, 2016, AUTOPHAGY, V12, P999, DOI 10.1080/15548627.2016.1166318
   Jackson SP, 2009, NATURE, V461, P1071, DOI 10.1038/nature08467
   Jennewein L, 2016, ONCOTARGET, V7, P20016, DOI 10.18632/oncotarget.7910
   Jones C, 2014, NAT REV CANCER, V14, P651, DOI 10.1038/nrc3811
   Jones C, 2017, NEURO-ONCOLOGY, V19, P153, DOI 10.1093/neuonc/now101
   Jones C, 2012, NAT REV CLIN ONCOL, V9, P400, DOI 10.1038/nrclinonc.2012.87
   Jung CH, 2010, FEBS LETT, V584, P1287, DOI 10.1016/j.febslet.2010.01.017
   Jutten B, 2014, CELL CYCLE, V13, P42, DOI 10.4161/cc.27518
   Karajannis M, 2008, J CELL PHYSIOL, V217, P584, DOI 10.1002/jcp.21544
   Kaur J, 2015, NAT REV MOL CELL BIO, V16, P461, DOI 10.1038/nrm4024
   Kaushik S, 2012, TRENDS CELL BIOL, V22, P407, DOI 10.1016/j.tcb.2012.05.006
   Kepp O, 2011, NAT REV DRUG DISCOV, V10, P221, DOI 10.1038/nrd3373
   Kessel D, 2015, PHOTOCHEM PHOTOBIOL, V91, P931, DOI 10.1111/php.12456
   Kessel DH, 2012, AUTOPHAGY, V8, P1333, DOI 10.4161/auto.20792
   Klionsky DJ, 2008, AUTOPHAGY, V4, P151, DOI 10.4161/auto.5338
   Koukourakis MI, 2016, BRIT J CANCER, V114, P485, DOI 10.1038/bjc.2016.19
   Lambelet M, 2018, CELL DEATH DIS, V9, DOI 10.1038/s41419-017-0121-5
   Laplante M, 2012, CELL, V149, P274, DOI 10.1016/j.cell.2012.03.017
   Lempiainen H, 2009, EMBO J, V28, P3067, DOI 10.1038/emboj.2009.281
   Levy JMM, 2017, ELIFE, V6, DOI 10.7554/eLife.19671
   Levy JMM, 2014, AUTOPHAGY, V10, P2077, DOI 10.4161/auto.36138
   Levy JMM, 2012, J NEURO-ONCOL, V106, P281, DOI 10.1007/s11060-011-0684-4
   Li WW, 2012, CELL MOL LIFE SCI, V69, P1125, DOI 10.1007/s00018-011-0865-5
   Liang ML, 2008, J NEURO-ONCOL, V87, P247, DOI 10.1007/s11060-007-9513-1
   Liu C, 2015, CURR MOL MED, V15, P663, DOI 10.2174/1566524015666150831141112
   Liu PX, 2009, NAT REV DRUG DISCOV, V8, P627, DOI 10.1038/nrd2926
   Liu Y, 2015, CELL DEATH DIFFER, V22, P367, DOI 10.1038/cdd.2014.143
   Long WY, 2017, FRONT PHARMACOL, V8, DOI 10.3389/fphar.2017.00495
   MacDonald TJ, 2011, NEURO-ONCOLOGY, V13, P1049, DOI 10.1093/neuonc/nor092
   Mackay A, 2017, CANCER CELL, V32, P520, DOI 10.1016/j.ccell.2017.08.017
   Marino G, 2014, NAT REV MOL CELL BIO, V15, P81, DOI 10.1038/nrm3735
   Martini M, 2014, ANN MED, V46, P372, DOI 10.3109/07853890.2014.912836
   Mathiassen SG, 2017, FRONT ONCOL, V7, DOI 10.3389/fonc.2017.00051
   Mellinghoff IK, 2011, CURR TOP MICROBIOL I, V1, P135
   Mora R, 2009, AUTOPHAGY, V5, P419, DOI 10.4161/auto.5.3.7881
   Morgenstern DA, 2014, PEDIATR BLOOD CANCER, V61, P128, DOI 10.1002/pbc.24656
   Mossmann D, 2018, NAT REV CANCER, V18, P744, DOI 10.1038/s41568-018-0074-8
   Mueller S, 2012, NEURO-ONCOLOGY, V14, P1146, DOI 10.1093/neuonc/nos140
   Munoz-Gomez JA, 2009, AUTOPHAGY, V5, P61, DOI 10.4161/auto.5.1.7272
   Narayana A, 2010, NEURO-ONCOLOGY, V12, P985, DOI 10.1093/neuonc/noq033
   Nezis IP, 2010, AUTOPHAGY, V6, P1214, DOI 10.4161/auto.6.8.13694
   Nezis IP, 2010, J CELL BIOL, V190, P523, DOI 10.1083/jcb.201002035
   Norden AD, 2009, NAT REV NEUROL, V5, P610, DOI 10.1038/nrneurol.2009.159
   Oh WJ, 2011, CELL CYCLE, V10, P2305, DOI 10.4161/cc.10.14.16586
   Ostrom QT, 2014, NEURO-ONCOLOGY, V16, P1, DOI [10.1093/neuonc/nov189, 10.1093/neuonc/nou327, 10.1093/neuonc/nou223]
   Papandreou I, 2008, CELL DEATH DIFFER, V15, P1572, DOI 10.1038/cdd.2008.84
   Parekh C, 2011, J NEURO-ONCOL, V103, P673, DOI 10.1007/s11060-010-0444-x
   Paugh BS, 2010, J CLIN ONCOL, V28, P3061, DOI 10.1200/JCO.2009.26.7252
   Pawlowska E, 2018, INT J MOL SCI, V19, DOI 10.3390/ijms19030889
   Peng X, 2014, CELL DEATH DIS, V5, DOI 10.1038/cddis.2014.297
   Pollack IF, 2006, J NEUROSURG, V105, P418, DOI 10.3171/ped.2006.105.5.418
   Populo H, 2012, INT J MOL SCI, V13, P1886, DOI 10.3390/ijms13021886
   Puget S, 2012, PLOS ONE, V7, DOI 10.1371/journal.pone.0030313
   Rizzo D, 2015, BIOMED RES INT, V2015, DOI 10.1155/2015/215135
   Robert T, 2011, NATURE, V471, P74, DOI 10.1038/nature09803
   Rodriguez-Rocha H, 2011, MUTAT RES-FUND MOL M, V711, P158, DOI 10.1016/j.mrfmmm.2011.03.007
   Rogers HA, 2017, ONCOTARGET, V8, P2083, DOI 10.18632/oncotarget.13781
   Roos WP, 2016, NAT REV CANCER, V16, P20, DOI 10.1038/nrc.2015.2
   Samuels Y, 2006, CURR OPIN ONCOL, V18, P77, DOI 10.1097/01.cco.0000198021.99347.b9
   Sanders RP, 2007, PEDIATR BLOOD CANCER, V49, P888, DOI 10.1002/pbc.21272
   Sands SA, 2012, J CLIN ONCOL, V30, P943, DOI 10.1200/JCO.2011.35.7533
   Schwartzentruber J, 2012, NATURE, V482, P226, DOI [10.1038/nature10833, 10.1007/s13311-012-0137-6]
   Shchors K, 2015, CANCER CELL, V28, DOI 10.1016/j.ccell.2015.08.012
   Shen J, 2013, BRIT J CANCER, V109, P164, DOI 10.1038/bjc.2013.306
   Shin HJR, 2016, AUTOPHAGY, V12, P2248, DOI 10.1080/15548627.2016.1214780
   Shingu T, 2016, J TRANSL MED, V14, DOI 10.1186/s12967-016-0803-2
   Song MS, 2012, NAT REV MOL CELL BIO, V13, P283, DOI 10.1038/nrm3330
   SPOSTO R, 1989, J NEURO-ONCOL, V7, P165, DOI 10.1007/BF00165101
   Spunt SL, 2011, J CLIN ONCOL, V29, P2933, DOI 10.1200/JCO.2010.33.4649
   Strickland M, 2017, FRONT CELL DEV BIOL, V5, DOI 10.3389/fcell.2017.00043
   Sturm D, 2014, NAT REV CANCER, V14, P92, DOI 10.1038/nrc3655
   Sun F, 2016, TUMOR BIOL, V37, P15467, DOI 10.1007/s13277-016-5353-y
   Takeuchi H, 2005, CANCER RES, V65, P3336, DOI 10.1158/0008-5472.CAN-04-3640
   Taylor TE, 2012, CURR CANCER DRUG TAR, V12, P197
   Thobe K, 2017, CELL COMMUN SIGNAL, V15, DOI 10.1186/s12964-016-0159-5
   Thorburn A, 2018, J BIOL CHEM, V293, P5425, DOI 10.1074/jbc.R117.810739
   Thorburn A, 2015, CANCER DISCOV, V5, P353, DOI 10.1158/2159-8290.CD-15-0222
   van Tellingen O, 2015, DRUG RESIST UPDATE, V19, P1, DOI 10.1016/j.drup.2015.02.002
   Vanhaesebroeck B, 2010, NAT REV MOL CELL BIO, V11, P329, DOI 10.1038/nrm2882
   Varghese F, 2014, PLOS ONE, V9, DOI 10.1371/journal.pone.0096801
   Vinci M, 2018, NAT MED, V24, P1204, DOI 10.1038/s41591-018-0086-7
   Wang DW, 2015, ONCOTARGET, V6, P37098, DOI 10.18632/oncotarget.5776
   Warren KE, 2012, TRANSL PEDIATR, V1, P116, DOI 10.3978/j.issn.2224-4336.2012.09.01
   Weihua Z, 2008, CANCER CELL, V13, P385, DOI 10.1016/j.ccr.2008.03.015
   Woo RA, 2002, EMBO J, V21, P3000, DOI 10.1093/emboj/cdf307
   Woo RA, 1998, NATURE, V394, P700, DOI 10.1038/29343
   Wu G, 2014, NAT GENET, V46, P444, DOI 10.1038/ng.2938
   Wu G, 2012, NAT GENET, V44, P251, DOI 10.1038/ng.1102
   Yan YL, 2016, J EXP CLIN CANC RES, V35, DOI 10.1186/s13046-016-0303-5
   Yang ZF, 2010, CURR OPIN CELL BIOL, V22, P124, DOI 10.1016/j.ceb.2009.11.014
   Yang ZNJ, 2011, MOL CANCER THER, V10, P1533, DOI 10.1158/1535-7163.MCT-11-0047
   Yao TW, 2017, ONCOTARGET, V8, P583, DOI 10.18632/oncotarget.11882
   Yap TA, 2015, CURR OPIN PHARMACOL, V23, P98, DOI 10.1016/j.coph.2015.05.016
   Ye HX, 2016, BMC NEUROL, V16, DOI 10.1186/s12883-016-0700-6
   Youle RJ, 2011, NAT REV MOL CELL BIO, V12, P9, DOI 10.1038/nrm3028
   Yuan HX, 2013, AUTOPHAGY, V9, P1983, DOI 10.4161/auto.26058
   Zarghooni M, 2010, J CLIN ONCOL, V28, P1337, DOI 10.1200/JCO.2009.25.5463
   Zhang RJ, 2015, MOL MED REP, V12, P1727, DOI 10.3892/mmr.2015.3588
   Zhao L, 2008, ONCOGENE, V27, P5486, DOI 10.1038/onc.2008.244
   Zhuang WZ, 2009, ACTA BIOCH BIOPH SIN, V41, P341, DOI 10.1093/abbs/gmp028
   Zou ZP, 2016, CURR CANCER DRUG TAR, V16, P288, DOI 10.2174/1568009616666151113120830
NR 156
TC 9
Z9 10
U1 0
U2 11
PU WILEY
PI HOBOKEN
PA 111 RIVER ST, HOBOKEN 07030-5774, NJ USA
SN 1015-6305
EI 1750-3639
J9 BRAIN PATHOL
JI Brain Pathol.
PD NOV
PY 2019
VL 29
IS 6
BP 707
EP 725
DI 10.1111/bpa.12729
PG 19
WC Clinical Neurology; Neurosciences; Pathology
WE Science Citation Index Expanded (SCI-EXPANDED)
SC Neurosciences & Neurology; Pathology
GA JF5RW
UT WOS:000491447100001
PM 31012506
OA Green Published, Green Accepted
DA 2023-08-27
ER

PT J
AU Sikio, M
   Holli, KK
   Harrison, LCV
   Ruottinen, H
   Rossi, M
   Helminen, MT
   Ryymin, P
   Paalavuo, R
   Soimakallio, S
   Eskola, HJ
   Elovaara, I
   Dastidar, P
AF Sikio, Minna
   Holli, Kirsi K.
   Harrison, Lara C. V.
   Ruottinen, Hanna
   Rossi, Maija
   Helminen, Mika T.
   Ryymin, Pertti
   Paalavuo, Raija
   Soimakallio, Seppo
   Eskola, Hannu J.
   Elovaara, Irina
   Dastidar, Prasun
TI Parkinson's Disease: Interhemispheric Textural Differences in MR Images
SO ACADEMIC RADIOLOGY
LA English
DT Article
DE Texture analysis; Parkinson's disease; co-occurrence matrix
ID BASAL GANGLIA IRON; MAGNETIC-RESONANCE; INTRACRANIAL TUMORS;
   MULTIPLE-SCLEROSIS; SUBSTANTIA-NIGRA; VENTRICULAR ENLARGEMENT; TISSUE
   CHARACTERIZATION; ESSENTIAL TREMOR; NORMAL BRAIN; DEPOSITION
AB Rationale and Objectives: Early-stage diagnosis of Parkinson's disease (PD) is essential in making decisions related to treatment and prognosis. However, there is no specific diagnostic test for the diagnosis of PD. The aim of this study was to evaluate the role of texture analysis (TA) of magnetic resonance images in detecting subtle changes between the hemispheres in various brain structures in patients with early symptoms of parkinsonism. In addition, functional TA parameters for detecting textural changes are presented.
   Materials and Methods: Fifty-one patients with symptoms of PD and 20 healthy controls were imaged using a 3-T magnetic resonance device. Co-occurrence matrix-based TA was applied to detect changes in textures between the hemispheres in the following clinically interesting areas: dentate nucleus, basilar pons, substantia nigra, globus pallidus, thalamus, putamen, caudate nucleus, corona radiata, and centrum semiovale. The TA results were statistically evaluated using the Mann-Whitney U test.
   Results: The results showed interhemispheric textural differences among the patients, especially in the area of basilar pons and midbrain. Concentrating on this clinically interesting area, the four most discriminant parameters were defined: co-occurrence matrix correlation, contrast, difference variance, and sum variance. With these parameters, differences were also detected in the dentate nucleus, globus pallidus, and corona radiata.
   Conclusions: On the basis of this study, interhemispheric differences in the magnetic resonance images of patients with PD can be identified by the means of co-occurrence matrix-based TA. The detected areas correlate with the current pathophysiologic and neuroanatomic knowledge of PD.
C1 [Sikio, Minna; Holli, Kirsi K.; Rossi, Maija; Ryymin, Pertti; Eskola, Hannu J.; Dastidar, Prasun] Tampere Univ Hosp, Med Imaging Ctr, FIN-33521 Tampere, Finland.
   [Ruottinen, Hanna; Elovaara, Irina] Tampere Univ Hosp, Dept Neurol & Rehabil, FIN-33521 Tampere, Finland.
   [Sikio, Minna; Holli, Kirsi K.; Harrison, Lara C. V.; Rossi, Maija; Eskola, Hannu J.] Tampere Univ Technol, Dept Biomed Engn, FIN-33101 Tampere, Finland.
   [Harrison, Lara C. V.; Paalavuo, Raija; Soimakallio, Seppo; Elovaara, Irina; Dastidar, Prasun] Univ Tampere, Tampere Med Sch, FIN-33101 Tampere, Finland.
   [Helminen, Mika T.] Univ Tampere, Tampere Sch Publ Hlth, FIN-33101 Tampere, Finland.
   [Helminen, Mika T.] Pirkanmaa Hosp Dist, Ctr Sci, Tampere, Finland.
C3 Tampere University; Tampere University Hospital; Tampere University;
   Tampere University Hospital; Tampere University; Tampere University;
   Tampere University; Pirkanmaa Hospital District
RP Sikio, M (通讯作者)，Tampere Univ Hosp, Med Imaging Ctr, Teiskontie 35,POB 2000, FIN-33521 Tampere, Finland.
EM
RI Helminen, Mika T/I-3900-2016; Eskola, Hannu J/G-4279-2014
OI Helminen, Mika T/0000-0001-5695-2084; Eskola, Hannu
   J/0000-0003-1673-2305; Rossi, Maija/0000-0002-7920-2531
FU City of Tampere; Finnish Cultural Science Foundation; Finnish Foundation
   for Technology Promotion
FX This research was supported by grants from City of Tampere, the
   Pirkanmaa Regional Fund of the Finnish Cultural Science Foundation, and
   the Finnish Foundation for Technology Promotion.
CR ANTONINI A, 1993, NEUROLOGY, V43, P697, DOI 10.1212/WNL.43.4.697
   Bonilha L, 2003, EPILEPSIA, V44, P1546, DOI 10.1111/j.0013-9580.2003.27103.x
   Brar S, 2009, ARCH NEUROL-CHICAGO, V66, P371, DOI 10.1001/archneurol.2008.586
   Chen H, 2010, CHINESE MED J-PEKING, V123, P695, DOI 10.3760/cma.j.issn.0366-6999.2010.06.011
   Collewet G, 2004, MAGN RESON IMAGING, V22, P81, DOI 10.1016/j.mri.2003.09.001
   Damier P, 1999, BRAIN, V122, P1437, DOI 10.1093/brain/122.8.1437
   DEXTER DT, 1991, BRAIN, V114, P1953, DOI 10.1093/brain/114.4.1953
   DRAYER B, 1986, AM J ROENTGENOL, V147, P103, DOI 10.2214/ajr.147.1.103
   Eapen M, 2011, AM J NEURORADIOL, V32, P688, DOI 10.3174/ajnr.A2355
   FEARNLEY JM, 1991, BRAIN, V114, P2283, DOI 10.1093/brain/114.5.2283
   Freeborough PA, 1998, IEEE T MED IMAGING, V17, P475, DOI 10.1109/42.712137
   Gibbs P, 2003, MAGN RESON MED, V50, P92, DOI 10.1002/mrm.10496
   Graham JM, 2000, BRAIN, V123, P2423, DOI 10.1093/brain/123.12.2423
   Griffiths PD, 1999, BRAIN, V122, P667, DOI 10.1093/brain/122.4.667
   Hajek M, 2006, TEXTURE ANAL MAGNETI
   HARALICK RM, 1979, P IEEE, V67, P786, DOI 10.1109/PROC.1979.11328
   Harrison LCV, 2010, ACAD RADIOL, V17, P696, DOI 10.1016/j.acra.2010.01.005
   Harrison LCV, 2009, J EXP CLIN CANC RES, V28, DOI 10.1186/1756-9966-28-87
   Herlidou-Meme S, 2003, MAGN RESON IMAGING, V21, P989, DOI 10.1016/S0730-725X(03)00212-1
   Holli K, 2010, ACAD RADIOL, V17, P135, DOI 10.1016/j.acra.2009.08.012
   Holli KK, 2010, ACAD RADIOL, V17, P1096, DOI 10.1016/j.acra.2010.04.009
   Holli Kirsi K, 2010, BMC Med Imaging, V10, P8, DOI 10.1186/1471-2342-10-8
   HORNYKIE.O, 1973, FED PROC, V32, P183
   Hornykiewicz O, 1998, NEUROLOGY, V51, pS2, DOI 10.1212/WNL.51.2_Suppl_2.S2
   Huang X, 2007, MOVEMENT DISORD, V22, P1657, DOI 10.1002/mds.21626
   Jirak D, 2002, J MAGN RESON IMAGING, V15, P68, DOI 10.1002/jmri.10042
   Kassner A, 2010, AM J NEURORADIOL, V31, P809, DOI 10.3174/ajnr.A2061
   Kassner A, 2009, J MAGN RESON IMAGING, V30, P933, DOI 10.1002/jmri.21940
   KJAER L, 1995, ACTA RADIOL, V36, P127
   Kosta P, 2006, J NEUROL, V253, P26, DOI 10.1007/s00415-005-0914-9
   LEE CS, 1995, NEUROLOGY, V45, P435, DOI 10.1212/WNL.45.3.435
   Lees AJ, 2009, LANCET, V373, P2055, DOI 10.1016/S0140-6736(09)60492-X
   Lewis MM, 2009, EUR J NEUROL, V16, P475, DOI 10.1111/j.1468-1331.2008.02430.x
   Loizou CP, 2011, IEEE T INF TECHNOL B, V15, P119, DOI 10.1109/TITB.2010.2091279
   Mahmoud-Ghoneim D, 2003, MAGN RESON IMAGING, V21, P983, DOI 10.1016/S0730-725X(03)00201-7
   Mandir AS, 2000, INT REV PSYCHIATR, V12, P270, DOI 10.1080/09540260020002497
   Martin WRW, 2008, NEUROLOGY, V70, P1411, DOI 10.1212/01.wnl.0000286384.31050.b5
   Martin WRW, 1998, CAN J NEUROL SCI, V25, P44, DOI 10.1017/S0317167100033473
   Mathias JM, 1999, MAGN RESON MED, V42, P929, DOI 10.1002/(SICI)1522-2594(199911)42:5<929::AID-MRM13>3.0.CO;2-2
   Menke RA, 2009, NEUROIMAGE, V47, P435, DOI 10.1016/j.neuroimage.2009.05.017
   Molnar GF, 2005, J NEUROPHYSIOL, V93, P3094, DOI 10.1152/jn.00881.2004
   Peran P, 2010, BRAIN, V133, P3423, DOI 10.1093/brain/awq212
   Piccini P, 2006, MOVEMENT DISORD, V21, P2035, DOI 10.1002/mds.20845
   RYVLIN P, 1995, ARCH NEUROL-CHICAGO, V52, P583, DOI 10.1001/archneur.1995.00540300057013
   SCHAD LR, 1993, MAGN RESON IMAGING, V11, P889, DOI 10.1016/0730-725X(93)90206-S
   Vymazal J, 1999, RADIOLOGY, V211, P489, DOI 10.1148/radiology.211.2.r99ma53489
   Wallis LI, 2008, J MAGN RESON IMAGING, V28, P1061, DOI 10.1002/jmri.21563
   YOUDIM MBH, 1993, J NEURAL TRANSM-GEN, P57
   Zhang J, 2008, MAGN RESON IMAGING, V26, P1160, DOI 10.1016/j.mri.2008.01.016
   Zhang JQ, 2010, BRAIN RES, V1330, P124, DOI 10.1016/j.brainres.2010.03.036
NR 50
TC 7
Z9 8
U1 0
U2 5
PU ELSEVIER SCIENCE INC
PI NEW YORK
PA 360 PARK AVE SOUTH, NEW YORK, NY 10010-1710 USA
SN 1076-6332
EI 1878-4046
J9 ACAD RADIOL
JI Acad. Radiol.
PD OCT
PY 2011
VL 18
IS 10
BP 1217
EP 1224
DI 10.1016/j.acra.2011.06.007
PG 8
WC Radiology, Nuclear Medicine & Medical Imaging
WE Science Citation Index Expanded (SCI-EXPANDED)
SC Radiology, Nuclear Medicine & Medical Imaging
GA 826HF
UT WOS:000295344500003
PM 21784670
DA 2023-08-27
ER

PT J
AU MOLLER, P
   EICHELMANN, A
   KORETZ, K
   MECHTERSHEIMER, G
AF MOLLER, P
   EICHELMANN, A
   KORETZ, K
   MECHTERSHEIMER, G
TI ADHESION MOLECULES VLA-1 TO VLA-6 DEFINE DISCRETE STAGES OF PERIPHERAL
   LYMPHOCYTE-B DEVELOPMENT AND CHARACTERIZE DIFFERENT TYPES OF B-CELL
   NEOPLASIA
SO LEUKEMIA
LA English
DT Article
ID INTEGRIN; LYMPHOMAS; MEMORY; TUMOR
AB VLA-1 to VLA-6 are cell-surface molecules binding to matrix molecules such as collagen, fibronectin, epiligrin, and laminin. In addition, VLA-4 binds to VCAM-1 and ICAM-2, thus mediating intercellular adhesion prerogative for lymphocyte extravasation or 'homing'. Using frozen tissue of normal lymphoid organs and of 100 morphologically and immunologically typed B cell neoplasias, monoclonal antibodies to all six VAL-alpha and to the common beta-chain were applied to serial sections. VLAs were found differentially expressed in cytologically and microtopographically defined B-cell subsets [follicular mantle zone cells (MZ), follicular center cells (FC), extrafollicular cells (EF), and plasma cells (PC)] of normal spleen, lymph node, and thymic medulla (which contains an EF compartment). Thus, these cell types, which correspond to discrete stages of 8 cell development, can also be defined by their VLA status. Acute B lymphoblastic leukemia (ALL) was VLA-1-, 2-, 3+/-, 4+/-, 5+/-, 6-. The VLA-1-, -2+/-, 3+, -4+, -5+, -6-phenotype of chronic B lymphocytic leukemia (CLL) resembled that of MZ. Hairy cell leukemia (HCL) differed from CLL in its tendency to lack VLA-2, in its consistent lack of VLA-3, and altogether resembled splenic EF in its VLA profile. Mantle zone lymphoma (MZL) consistently expressed VLA-3 and -4 and frequently VLA-5. Nodal follicular center cell lymphomas(FCCL) were VLA-1 - and -2 - and very rarely expressed VLA-5 and -6. Thus, FCCL although roughly corresponding to FC, tended to aberrantly express VLA-3 and/or VLA-4. Burkitt's lymphoma resembled FCCL but expressed VLA-4 more frequently and at higher levels. Mediastinal clear cell lymphoma of B-cell type differed from FCCL in its regular lack of VLA-3, -5, and -6 and in frequently lacking VLA-4. Medullary plasmacytoma was VLA-1-, -2-, -3+/-, -4+/-, -5-, -6+, thus being the only B cell neoplasia which was consistently VLA-6+. With respect to the well-known clinical characteristics of the B cell malignancies examined, the leukemic phenotype might crucially depend on the presence of VLA-5.
RP MOLLER, P (通讯作者)，UNIV HEIDELBERG,INST PATHOL,NEUENHEIMER FELD 220,W-6900 HEIDELBERG,GERMANY.
CR ADDIS BJ, 1986, HISTOPATHOLOGY, V10, P379, DOI 10.1111/j.1365-2559.1986.tb02491.x
   BALLARD LL, 1991, CLIN EXP IMMUNOL, V84, P336
   BRANDTER LB, 1989, LEUKEMIA, V3, P122
   CARTER WG, 1991, CELL, V65, P599, DOI 10.1016/0092-8674(91)90092-D
   DAVIS RE, 1990, HUM PATHOL, V21, P1262, DOI 10.1016/S0046-8177(06)80040-7
   GARCIAPARDO A, 1990, J IMMUNOL, V144, P3361
   HEMLER ME, 1990, IMMUNOL REV, V114, P45, DOI 10.1111/j.1600-065X.1990.tb00561.x
   HEMLER ME, 1984, J IMMUNOL, V132, P3011
   HEMLER ME, 1990, ANNU REV IMMUNOL, V8, P365, DOI 10.1146/annurev.iy.08.040190.002053
   HOLZMANN B, 1989, EMBO J, V8, P1735, DOI 10.1002/j.1460-2075.1989.tb03566.x
   HOLZMANN B, 1989, CELL, V56, P37, DOI 10.1016/0092-8674(89)90981-1
   HUMPHRIES MJ, 1990, J CELL SCI, V97, P2585
   KLINGEMANN HG, 1989, BLOOD, V74, P1348
   KOHN FR, 1991, IMMUNOL LETT, V28, P27, DOI 10.1016/0165-2478(91)90123-R
   LIU YJ, 1988, EUR J IMMUNOL, V18, P355, DOI 10.1002/eji.1830180306
   MIELKE B, 1991, INT J CANCER, V47, P334, DOI 10.1002/ijc.2910470304
   MOLLER P, 1986, VIRCHOWS ARCH A, V409, P79, DOI 10.1007/BF00705408
   MOLLER P, 1987, BLOOD, V69, P1087
   MOLLER P, 1989, PATHOLOGE, V10, P234
   MOLLER P, 1989, INT J CANCER, V43, P10, DOI 10.1002/ijc.2910430104
   MOLLER P, 1989, LEUCOCYTE TYPING, V4, P213
   OSBORN L, 1989, CELL, V59, P1203, DOI 10.1016/0092-8674(89)90775-7
   PULIDO R, 1991, J BIOL CHEM, V266, P10241
   REYNAUD CA, 1991, CELL, V64, P995, DOI 10.1016/0092-8674(91)90323-Q
   REYNOLDS JD, 1991, EUR J IMMUNOL, V21, P283, DOI 10.1002/eji.1830210207
   RYAN DH, 1991, J CLIN INVEST, V88, P995, DOI 10.1172/JCI115403
   SCARPA A, 1987, VIRCHOWS ARCH A, V412, P17, DOI 10.1007/BF00750725
   SETH R, 1991, FEBS LETT, V282, P193, DOI 10.1016/0014-5793(91)80475-I
   STANSFELD AG, 1988, LANCET, V1, P292
   STANSFELD AG, 1988, LANCET, V2, P372
   STANSFELD AG, 1988, LANCET, V1, P603
   YOUSEM SA, 1985, AM J CLIN PATHOL, V83, P676, DOI 10.1093/ajcp/83.6.676
   ZHANG J, 1988, EUR J IMMUNOL, V18, P1417, DOI 10.1002/eji.1830180918
   ZUTTER MM, 1991, BLOOD, V77, P2231
NR 34
TC 45
Z9 46
U1 0
U2 2
PU STOCKTON PRESS
PI BASINGSTOKE
PA HOUNDMILLS, BASINGSTOKE, HAMPSHIRE, ENGLAND RG21 6XS
SN 0887-6924
J9 LEUKEMIA
JI Leukemia
PD APR
PY 1992
VL 6
IS 4
BP 256
EP 264
PG 9
WC Oncology; Hematology
WE Science Citation Index Expanded (SCI-EXPANDED)
SC Oncology; Hematology
GA HX318
UT WOS:A1992HX31800005
PM 1588789
DA 2023-08-27
ER

PT J
AU Jang, BG
   Huh, KH
   Kang, JH
   Kim, JE
   Yi, WJ
   Heo, MS
   Lee, SS
AF Jang, B. G.
   Huh, K. H.
   Kang, J. H.
   Kim, J. E.
   Yi, W. J.
   Heo, M. S.
   Lee, S. S.
TI Imaging features of chondrosarcoma of the temporomandibular joint:
   report of nine cases and literature review
SO CLINICAL RADIOLOGY
LA English
DT Review
ID SYNOVIAL CHONDROSARCOMA; CARTILAGINOUS TUMORS; MANDIBULAR CONDYLE;
   CHONDROMATOSIS; HEAD; CT; MR
AB AIM: To investigate the imaging features of chondrosarcoma of the temporomandibular joint (TMJ) and review the literature.
   MATERIALS AND METHODS: Computed tomography (CT), magnetic resonance imaging (MRI), and integrated positron-emission tomography (PET)/CT images of nine patients with histopathologically confirmed chondrosarcoma of the TMJ were reviewed retrospectively. Imaging features regarding the direction of lesion growth, bone destruction, infiltration into the tendon of the lateral pterygoid muscle (LPM) in the pterygoid fovea, enhancement pattern, calcification, periosteal reaction, markedly hyperintense T2 signal area, and qualitative PET signal intensity were evaluated.
   RESULTS: Seven of nine patients (77.8%) presented with lesion growth that was outward from the medulla of the mandibular condyle. Infiltration into the tendon of LPM in the pter-ygoid fovea was observed in all cases, and 77.8% (7/9) of them demonstrated >50% infiltration. All the lesions showed a mixed peripheral and internal enhancement, and revealed a markedly hyperintense T2 signal intensity area, which showed no enhancement. Although five of nine cases demonstrated higher FDG uptake compared with that of the liver, the other four cases showed less FDG uptake than that of the liver.
   CONCLUSION: Chondrosarcoma of the TMJ demonstrated several imaging features, including outward growth from the mandibular condyle, resultant infiltration into the tendon of LPM in the pterygoid fovea, various patterns of internal enhancement, and a markedly hyperintense T2 signal intensity area. These imaging features may be helpful to differentiate chondrosarcoma from other lesions of the TMJ. (c) 2020 The Royal College of Radiologists. Published by Elsevier Ltd. This is an open access article under the CC BY-NC-ND license (http://creativecommons.org/licenses/by-nc-nd/4.0/).
C1 [Jang, B. G.; Huh, K. H.; Yi, W. J.; Heo, M. S.; Lee, S. S.] Seoul Natl Univ, Sch Dent, Dept Oral & Maxillofacial Radiol, 101 Daehak Ro, Seoul 03080, South Korea.
   [Jang, B. G.; Huh, K. H.; Yi, W. J.; Heo, M. S.; Lee, S. S.] Seoul Natl Univ, Sch Dent, Dent Res Inst, Seoul, South Korea.
   [Kang, J. H.; Kim, J. E.] Seoul Natl Univ, Dept Oral & Maxillofacial Radiol, Dent Hosp, Seoul, South Korea.
C3 Seoul National University (SNU); Seoul National University (SNU); Seoul
   National University (SNU)
RP Huh, KH (通讯作者)，Seoul Natl Univ, Sch Dent, Dept Oral & Maxillofacial Radiol, 101 Daehak Ro, Seoul 03080, South Korea.
EM
FU Seoul National University [860-20200059]
FX This work was supported by the New Faculty Startup Fund (860-20200059)
   from Seoul National University.
CR Abu-Serriah M, 2013, J ORAL MAXIL SURG, V71, P208, DOI 10.1016/j.joms.2012.04.005
   Angiero F, 2007, QUINTESSENCE INT, V38, P349
   AOKI J, 1991, J COMPUT ASSIST TOMO, V15, P1011, DOI 10.1097/00004728-199111000-00021
   Balliu E, 2007, DENTOMAXILLOFAC RAD, V36, P55, DOI 10.1259/dmfr/89319561
   Batra PS, 1999, OTOLARYNG HEAD NECK, V120, P951, DOI 10.1016/S0194-5998(99)70348-7
   Bernasconi G, 2004, CLIN IMAG, V28, P64, DOI 10.1016/S0899-7071(03)00100-1
   BURKEY BB, 1990, LARYNGOSCOPE, V100, P1301
   CADENAT H, 1979, REV STOMATOL CHIR, V80, P20
   Coleman H, 2013, HEAD NECK PATHOL, V7, P304, DOI 10.1007/s12105-013-0439-y
   DAHLIN DC, 1971, CANCER, V28, P461, DOI 10.1002/1097-0142(197108)28:2<461::AID-CNCR2820280227>3.0.CO;2-U
   de Oliveira RC, 2009, J OROFAC PAIN, V23, P275
   DeBeuckeleer LHL, 1995, EUR J RADIOL, V21, P34, DOI 10.1016/0720-048X(96)81067-9
   Deyhimi P, 2012, J DENT SHIRAZ U MED, V13, P176
   Douis H, 2013, SKELETAL RADIOL, V42, P611, DOI 10.1007/s00256-012-1521-3
   EVANS HL, 1977, CANCER-AM CANCER SOC, V40, P818, DOI 10.1002/1097-0142(197708)40:2<818::AID-CNCR2820400234>3.0.CO;2-B
   Fukada K, 2018, J ORAL MAX SURG MED, V30, P123, DOI 10.1016/j.ajoms.2017.07.001
   Gallego L, 2009, MED ORAL PATOL ORAL, V14, pE39
   Garzino-Demo P, 2010, J ORAL MAXIL SURG, V68, P2005, DOI 10.1016/j.joms.2009.09.077
   GEIRNAERDT MJA, 1993, RADIOLOGY, V186, P813, DOI 10.1148/radiology.186.3.8430192
   GINGRASS R P, 1954, J Oral Surg (Chic), V12, P61
   Giorgione C, 2015, ACTA OTORHINOLARYNGO, V35, P208
   Giraud O., 1997, Revue de Stomatologie et de Chirurgie Maxillo-Faciale, V98, P2
   Gonzalez-Perez LM, 2011, J CRANIO MAXILL SURG, V39, P79, DOI 10.1016/j.jcms.2010.03.018
   Goutzanis L, 2013, J CRANIO MAXILL SURG, V41, pE170, DOI 10.1016/j.jcms.2012.11.045
   Ichikawa T, 1998, J ORAL MAXIL SURG, V56, P890, DOI 10.1016/S0278-2391(98)90022-2
   Kato Y, 1975, ORTHOPEDIC SURG TOKY, V26, P313
   Koch BB, 2000, HEAD NECK-J SCI SPEC, V22, P408, DOI 10.1002/1097-0347(200007)22:4<408::AID-HED15>3.0.CO;2-H
   Kumar Reddy D Sathya, 2014, Ann Maxillofac Surg, V4, P85, DOI 10.4103/2231-0746.133076
   LANIER VC, 1971, SOUTH MED J, V64, P711, DOI 10.1097/00007611-197106000-00016
   Lee K, 2016, J KOR ASSOC ORAL MAX, V42, P288, DOI 10.5125/jkaoms.2016.42.5.288
   Lee SY, 2005, YONSEI MED J, V46, P228, DOI 10.3349/ymj.2005.46.2.228
   MacIntosh RB, 2015, J ORAL MAXIL SURG, V73, P465, DOI 10.1016/j.joms.2014.09.020
   Merrill RG, 1997, J ORAL MAXIL SURG, V55, P1312, DOI 10.1016/S0278-2391(97)90191-9
   MORRIS MR, 1987, HEAD NECK-J SCI SPEC, V10, P113, DOI 10.1002/hed.2890100210
   Mostafapour SP, 2000, OTOLARYNG HEAD NECK, V123, P459, DOI 10.1067/mhn.2000.109662
   MURAYAMA S, 1988, J CRANIO MAXILL SURG, V16, P287, DOI 10.1016/S1010-5182(88)80063-5
   Murphey MD, 2003, RADIOGRAPHICS, V23, P1245, DOI 10.1148/rg.235035134
   NITZAN DW, 1993, J ORAL MAXIL SURG, V51, P312, DOI 10.1016/S0278-2391(10)80182-X
   Nomura T, 2015, CASE REP OTOLARYNGOL, V2015, P1
   NORTJE CJ, 1976, BRIT J ORAL SURG, V14, P101, DOI 10.1016/0007-117X(76)90025-1
   Oda Y, 1998, ORAL MED PATHOL, V3, P118
   Oh KY, 2016, CRANIO, V34, P270, DOI 10.1179/2151090315Y.0000000016
   Ramos-Murguialday M, 2012, J CRANIOFAC SURG, V23, P1216, DOI 10.1097/SCS.0b013e3182565b80
   Rath R, 2014, J CLIN DIAGN RES, V8, pZD09, DOI 10.7860/JCDR/2014/9261.4790
   RICHTER KJ, 1974, J ORAL SURG, V32, P777
   SATO K, 1977, J ORAL SURG, V35, P892
   Sesenna E, 1997, J ORAL MAXIL SURG, V55, P1348, DOI 10.1016/S0278-2391(97)90200-7
   Slimani F, 2019, EC DENT SCI, V18, P1495
   Sudo Y, 1972, JJ SS, V211, P417
   Tullio G, 1974, Ann Stomatol (Roma), V23, P191
   WASENKO JJ, 1990, J COMPUT ASSIST TOMO, V14, P1002, DOI 10.1097/00004728-199011000-00028
   Weber Kristy L, 2002, Iowa Orthop J, V22, P75
   Xu B, 2011, DENTOMAXILLOFAC RAD, V40, P320, DOI 10.1259/dmfr/17575463
   Ye ZX, 2015, INT J CLIN EXP MED, V8, P11589
   Yun KI, 2008, J KOR ASSOC ORAL MAX, V34, P95
   Ziai P, 2016, RADIOGRAPHICS, V36, P481, DOI 10.1148/rg.2016150102
NR 56
TC 3
Z9 3
U1 1
U2 2
PU W B SAUNDERS CO LTD
PI LONDON
PA 32 JAMESTOWN RD, LONDON NW1 7BY, ENGLAND
SN 0009-9260
EI 1365-229X
J9 CLIN RADIOL
JI Clin. Radiol.
PD NOV
PY 2020
VL 75
IS 11
DI 10.1016/j.crad.2020.07.016
PG 12
WC Radiology, Nuclear Medicine & Medical Imaging
WE Science Citation Index Expanded (SCI-EXPANDED)
SC Radiology, Nuclear Medicine & Medical Imaging
GA OC1ED
UT WOS:000578904900020
PM 32843140
OA hybrid
DA 2023-08-27
ER

PT J
AU Martin-Lacave, I
   Utrilla, JC
AF Martin-Lacave, I
   Utrilla, JC
TI Expression of a neu/c-erbB-2-like product in neuroendocrine cells of
   mammals
SO HISTOLOGY AND HISTOPATHOLOGY
LA English
DT Article
DE neu/c-erbB-2 oncoprotein; neuroendocrine cells; immunohistochemistry; C
   cells; chromaffin cells
ID THYROID PARAFOLLICULAR CELLS; NEU-ONCOGENE; C-ERBB-2 ONCOPROTEIN;
   SECRETORY GRANULES; TUMOR-ANTIGEN; EGF RECEPTOR; PROTEIN; GROWTH;
   TISSUE; IMMUNOREACTIVITY
AB The neu/c-erbB-2 oncogen encodes a 185 kDa protein closely homologous to the epidermal growth factor receptor. The protein product (p185) is a glycoprotein with an external domain and an internal domain with tyrosine kinase activity. Amplification and/or overexpression of p185 is related to several human adenocarcinomas. Subsequent studies demonstrated its presence in certain neuroendocrine (NE) neoplasms, including phaeochromocytomas, insulinomas and medullary thyroid carcinomas. However, relatively little is known about its role in normal cell growth regulation and development. Therefore, our objetive was to determine whether neu/c-erbB-2 was expressed in normal NE tissues of different mammals, specially in humans, as it was in their neoplasms. We have examined by immunohistochemistry different endocrine glands (thyroid, pancreas, suprarrenal and hypophysis) and the small intestine of human beings, rats and guinea pigs, using two polyclonal antibodies raised against the intracytoplasmic part of the protein, and specific antigen absorption controls. We have found that a neu/c-erbB-2-like product occurs in all normal NE tissues examined: C cells of the thyroid gland, chromaffin cells of the adrenal medulla, pancreatic islets, enteroendocrine cells of the small intestine and, finally, scattered cells of the adenohypophysis, according to a typical granular immunohistochemical pattern. Our results indicate that normal NE cells share a new common antigen in their cytoplasms, a neu/c-erbB-2-like product, with a similar immunostaining pattern to that presented by the neoplasms derived from them.
C1 Univ Sevilla, Sch Med, Dept Cytol & Histopathol, E-41009 Seville, Spain.
C3 University of Sevilla
RP Martin-Lacave, I (通讯作者)，Univ Sevilla, Fac Med, Dept Citol & Histol Normal & Patol, Avda Sanchez Pizjuan Sn, E-41009 Seville, Spain.
OI Utrilla, Jose Carmelo/0000-0001-5438-604X; Martin-Lacave,
   Ines/0000-0003-4645-0755
CR AKIJAMA T, 1986, SCIENCE, V232, P1644
   BARASCH JM, 1987, J NEUROSCI, V7, P4017
   BARGMANN CI, 1986, NATURE, V319, P226, DOI 10.1038/319226a0
   BUFFA R, 1987, MOL CELL PROBE, V1, P367, DOI 10.1016/0890-8508(87)90018-1
   CastillaGuerra L, 1997, HISTOPATHOLOGY, V31, P144, DOI 10.1046/j.1365-2559.1997.2240838.x
   COOMBS LM, 1991, BRIT J CANCER, V63, P601, DOI 10.1038/bjc.1991.139
   CORBETT IP, 1990, J PATHOL, V161, P15, DOI 10.1002/path.1711610105
   COUSSENS L, 1985, SCIENCE, V230, P1132, DOI 10.1126/science.2999974
   DEPOTTER CR, 1989, HISTOPATHOLOGY, V15, P351
   DOUGALL WC, 1994, ONCOGENE, V9, P2109
   DREBIN JA, 1986, P NATL ACAD SCI USA, V83, P9129, DOI 10.1073/pnas.83.23.9129
   GOULD VE, 1986, HUM PATHOL, V17, P979, DOI 10.1016/S0046-8177(86)80080-6
   HALL PA, 1990, J PATHOL, V161, P195, DOI 10.1002/path.1711610305
   JAHN R, 1985, P NATL ACAD SCI USA, V82, P4137, DOI 10.1073/pnas.82.12.4137
   KOKAI Y, 1987, P NATL ACAD SCI USA, V84, P8498, DOI 10.1073/pnas.84.23.8498
   MILLER K, 1986, J CELL BIOL, V102, P500, DOI 10.1083/jcb.102.2.500
   MORI S, 1987, VIRCHOWS ARCH B, V54, P8, DOI 10.1007/BF02899192
   PRESS MF, 1990, ONCOGENE, V5, P953
   QUIRKE P, 1989, BRIT J CANCER, V60, P64, DOI 10.1038/bjc.1989.221
   RONCALLI M, 1991, J PATHOL, V163, P117, DOI 10.1002/path.1711630207
   SCHECHTER AL, 1984, NATURE, V312, P513, DOI 10.1038/312513a0
   SHI SR, 1991, J HISTOCHEM CYTOCHEM, V39, P741, DOI 10.1177/39.6.1709656
   SLAMON DJ, 1989, SCIENCE, V244, P707, DOI 10.1126/science.2470152
   SOARES P, 1994, INT J CANCER, V56, P459, DOI 10.1002/ijc.2910560330
   STIEBER A, 1995, BRAIN RES, V700, P261, DOI 10.1016/0006-8993(95)00981-U
   Utrilla JC, 1999, HISTOPATHOLOGY, V34, P60
   WEILER R, 1989, FEBS LETT, V257, P457, DOI 10.1016/0014-5793(89)81595-9
NR 27
TC 8
Z9 8
U1 0
U2 1
PU F HERNANDEZ
PI MURCIA
PA PLAZA FUENSANTA 2-7 C, 30008 MURCIA, SPAIN
SN 0213-3911
J9 HISTOL HISTOPATHOL
JI Histol. Histopath.
PD OCT
PY 2000
VL 15
IS 4
BP 1027
EP 1033
PG 7
WC Cell Biology; Pathology
WE Science Citation Index Expanded (SCI-EXPANDED)
SC Cell Biology; Pathology
GA 355NC
UT WOS:000089390900004
PM 11005226
DA 2023-08-27
ER

PT J
AU Bernardeschi, D
   Meskine, N
   Al Otaibi, N
   Ablonczy, R
   Kalamarides, M
   Grayeli, AB
   Sterkers, O
AF Bernardeschi, Daniele
   Meskine, Nawel
   Al Otaibi, Naif
   Ablonczy, Reka
   Kalamarides, Michel
   Grayeli, Alexis Bozorg
   Sterkers, Olivier
TI Continuous Facial Nerve Stimulating Burr for Otologic Surgeries
SO OTOLOGY & NEUROTOLOGY
LA English
DT Article
DE Facial nerve; Intraoperative monitoring; Mastoidectomy; Vestibular
   schwannoma
ID ACOUSTIC NEUROMA SURGERY; CHRONIC EAR SURGERY; MIDDLE-EAR; VESTIBULAR
   SCHWANNOMA; DEHISCENCE
AB Objective: To evaluate a continuous facial nerve (FN) stimulating burr (the StimBurGard) during otologic/neurotologic procedures in terms of safety and reliability when drilling in contact with the Fallopian canal (FC) of the mastoid segment of the FN.
   Study Design: Prospective clinical trial.
   Setting: Tertiary referral center.
   Patients: Thirty-five patients operated through translabyrinthine (TL) approach for vestibular schwannoma removal were divided into 3 groups. Group 1 (5 patients): the stimulation current was set at 3 and then at 2 mA visualizing the localization of the burr when the first response at 100-mu V threshold was obtained in the mastoid cavity. Group 2 (15 patients): exposure of the FC in the mastoid segment during TL approach was stopped when the first response was obtained at 1-mA stimulation; FC thickness in the second genu and mastoid segment of the FC was evaluated on a postoperative computed tomographic (CT) scan, and FC dehiscence observed on CT scan was compared with surgical observation. Group 3 (15 patients), exposure of the FC was performed as routinely done during a TL approach and surgical observation of FC dehiscence; stimulation values in mA at the 100 mu V threshold and FC thickness on postoperative CT scan were evaluated. In all cases, the stimulation value at the cerebellopontine angle root of the FN with a 100-mu V response threshold was measured before tumor resection.
   Results: Group 1: stimulation at 3 mA occurred in aditus ad antrum and at 2 mA near the FC. Group 2: mean thickness of 1.09 +/- 0.69 mm with 2 cases of radiologic dehiscence of the FN. Group 3: the stimulation threshold was 0.6 +/- 0.37 mA, and the thickness was 0.41 +/- 0.56 mm with 9 cases of uncovered FN (p = 0.0082). In all patients, FN at brainstem was stimulated at 0.03 mA before VS dissection.
   Conclusion: Continuous FN stimulating burr by means of the StimBurGard system is a safe and effective tool for FN stimulation and identification. The integrity of FC is preserved in most cases when the stimulation intensity is 1 mA.
C1 [Bernardeschi, Daniele; Al Otaibi, Naif; Ablonczy, Reka; Grayeli, Alexis Bozorg; Sterkers, Olivier] Beaujon Hosp, AP HP, ENT Dept, F-92100 Clichy, France.
   [Bernardeschi, Daniele; Grayeli, Alexis Bozorg; Sterkers, Olivier] INSERM, Paris, France.
   [Bernardeschi, Daniele; Grayeli, Alexis Bozorg; Sterkers, Olivier] Univ Paris Diderot, Paris, France.
   [Meskine, Nawel] Beaujon Hosp, AP HP, Dept Radiol, F-92100 Clichy, France.
   [Kalamarides, Michel] Beaujon Hosp, AP HP, Dept Neurosurg, F-92100 Clichy, France.
C3 Assistance Publique Hopitaux Paris (APHP); Hopital Universitaire
   Paul-Brousse - APHP; UDICE-French Research Universities; Universite
   Paris Cite; Hopital Universitaire Beaujon - APHP; Hopital Universitaire
   Bichat-Claude Bernard - APHP; Hopital Universitaire Hotel-Dieu - APHP;
   Institut National de la Sante et de la Recherche Medicale (Inserm);
   UDICE-French Research Universities; Universite Paris Cite; Assistance
   Publique Hopitaux Paris (APHP); Hopital Universitaire Paul-Brousse -
   APHP; UDICE-French Research Universities; Universite Paris Cite; Hopital
   Universitaire Beaujon - APHP; Hopital Universitaire Bichat-Claude
   Bernard - APHP; Hopital Universitaire Hotel-Dieu - APHP; Assistance
   Publique Hopitaux Paris (APHP); Hopital Universitaire Paul-Brousse -
   APHP; UDICE-French Research Universities; Universite Paris Cite; Hopital
   Universitaire Beaujon - APHP; Hopital Universitaire Bichat-Claude
   Bernard - APHP; Hopital Universitaire Hotel-Dieu - APHP
RP Bernardeschi, D (通讯作者)，Beaujon Hosp, AP HP, ENT Dept, 100 Bd Gen Leclerc, F-92100 Clichy, France.
EM
CR Barnes G, 2001, OTOL NEUROTOL, V22, P931, DOI 10.1097/00129492-200111000-00038
   Bayazit YA, 2002, J LARYNGOL OTOL, V116, P800, DOI 10.1258/00222150260293600
   Choung YH, 2006, OTOLARYNG HEAD NECK, V135, P872, DOI 10.1016/j.otohns.2006.04.008
   Colletti V, 1998, AM J OTOL, V19, P503
   DELGADO TE, 1979, NEUROSURGERY, V4, P418, DOI 10.1227/00006123-197905000-00007
   Di Martino E, 2005, EUR ARCH OTO-RHINO-L, V262, P120, DOI 10.1007/s00405-004-0867-0
   Grayeli AB, 2005, ACTA OTO-LARYNGOL, V125, P1069, DOI 10.1080/00016480510038608
   Greenberg JS, 2002, OTOLARYNG HEAD NECK, V126, P108, DOI 10.1067/mhn.2002.121861
   JACKLER RK, 1994, AM J OTOL, V15, P611
   JACKLER RK, 1996, ATLAS SKULL BASE SUR, P17
   Kartush JM, 1998, NEUROL RES, V20, P593
   KWARTLER JA, 1991, OTOLARYNG HEAD NECK, V104, P814, DOI 10.1177/019459989110400608
   LEONETTI JP, 1990, ANN OTO RHINOL LARYN, V99, P911, DOI 10.1177/000348949009901111
   MOREANO EH, 1994, LARYNGOSCOPE, V104, P309
   Nakao Y, 2001, OTOL NEUROTOL, V22, P554, DOI 10.1097/00129492-200107000-00024
   *NIH, 1991, NIH CONS DEV C BETH
   Noss RS, 2001, LARYNGOSCOPE, V111, P831, DOI 10.1097/00005537-200105000-00014
   Olds MJ, 1997, AM J OTOL, V18, P507
   PENSAK ML, 1994, AM J OTOL, V15, P108
   SILVERSTEIN H, 1993, AM J OTOL, V14, P524
   SILVERSTEIN H, 1988, AM J OTOL, V9, P269
   SILVERSTEIN H, 1990, OTOLARYNG HEAD NECK, V103, P493, DOI 10.1177/019459989010300327
   SILVERSTEIN H, 1988, LARYNGOSCOPE, V98, P726
   Sughrue ME, 2010, BRIT J NEUROSURG, V24, P666, DOI 10.3109/02688697.2010.520761
   WIET RJ, 1982, OTOLARYNG CLIN N AM, V15, P773
NR 25
TC 21
Z9 22
U1 0
U2 1
PU LIPPINCOTT WILLIAMS & WILKINS
PI PHILADELPHIA
PA 530 WALNUT ST, PHILADELPHIA, PA 19106-3621 USA
SN 1531-7129
J9 OTOL NEUROTOL
JI Otol. Neurotol.
PD OCT
PY 2011
VL 32
IS 8
BP 1347
EP 1351
DI 10.1097/MAO.0b013e31822ec097
PG 5
WC Clinical Neurology; Otorhinolaryngology
WE Science Citation Index Expanded (SCI-EXPANDED)
SC Neurosciences & Neurology; Otorhinolaryngology
GA 821BD
UT WOS:000294948900032
PM 21892117
DA 2023-08-27
ER

PT J
AU Sato, H
   Mizumoto, M
   Okumura, T
   Sakurai, H
   Sakamoto, N
   Akutsu, H
   Ishikawa, E
   Tsuboi, K
AF Sato, Hiroshige
   Mizumoto, Masashi
   Okumura, Toshiyuki
   Sakurai, Hideyuki
   Sakamoto, Noriaki
   Akutsu, Hiroyoshi
   Ishikawa, Eiichi
   Tsuboi, Koji
TI Long-term outcomes of patients with unresectable benign meningioma
   treated with proton beam therapy
SO JOURNAL OF RADIATION RESEARCH
LA English
DT Article
DE benign meningioma; long-term control; proton beam; radiotherapy
ID INTENSITY-MODULATED RADIOTHERAPY; SKULL-BASE; EXPERIENCE; RECURRENT;
   EPIDEMIOLOGY; MORBIDITY; TOXICITY; FEATURES; CLIVUS; IMPACT
AB This study aimed to evaluate the long-term efficacy of proton beam therapy (PBT) for unresectable benign meningiomas at the University of Tsukuba, Japan. From 1986-1998, 10 patients were treated at the Particle Radiation Medical Science Center (PRMSC) with a relative biological effectiveness (RBE) value of 1.0 using an accelerator built for physics experiments. The total dose was compensated with an X-ray in three patients. Following that, from 2002-2017, 17 patients were treated with a RBE value of 1.1 at the Proton Medical Research Center (PMRC) which was built for medical use. At the PRMSC, the total dose ranged from 50.4-66 Gy (median: 54 Gy). During the followup, which lasted between 3.8 and 31.6 years (median: 25.1 years), the 5-, 10-, 15-, 20- and 30-year local control rates were 100%, and the 5-, 10-, 15-, 20- and 30-year survival rates were 90, 80, 70, 70 and 36%, respectively. One patient died of brainstem radiation necrosis 5.1 years after PBT. At PMRC, the total dose ranged from 45.0-61.2 GyE, with a median of 50.4 GyE. During the follow-up, which lasted between 3 and 17 years with a median of 10.5 years, the 5-, 10- and 15-year local control rates were 94.1%, and the 5-, 10- and 15-year survival rates were 100, 100 and 88.9%, respectively. Neither malignant transformation nor secondary malignancy was observed, indicating that fractionated PBT may be effective and safely control benign unresectable meningioma even for the lifelong period of time.
C1 [Sato, Hiroshige] Univ Tsukuba, Grad Sch Comprehens Human Sci, Biomed Sci, 1-1-1 Tennohdai, Tsukuba, Ibaraki 3058575, Japan.
   [Mizumoto, Masashi; Okumura, Toshiyuki; Sakurai, Hideyuki] Univ Tsukuba Hosp, Proton Beam Therapy Ctr, Dept Radiat Oncol, 2-1-1 Amakubo, Tsukuba, Ibaraki 3058576, Japan.
   [Sakamoto, Noriaki] Univ Tsukuba, Fac Med, Dept Diagnost Pathol, 2-1-1 Amakubo, Tsukuba, Ibaraki 3058576, Japan.
   [Akutsu, Hiroyoshi; Ishikawa, Eiichi] Univ Tsukuba, Fac Med, Dept Neurosurg, 2-1-1 Amakubo, Tsukuba, Ibaraki 3058576, Japan.
   [Tsuboi, Koji] Tsukuba Cent Hosp, Tumor Therapy Ctr, 1589-3 Kashiwada, Ushiku, Ibaraki 3001211, Japan.
C3 University of Tsukuba; University of Tsukuba; University of Tsukuba;
   University of Tsukuba
RP Tsuboi, K (通讯作者)，Tsukuba Cent Hosp, Tumor Therapy Ctr, 1589-3 Kashiwada, Ushiku, Ibaraki 3001211, Japan.
EM
RI Mizumoto, Masashi/HJP-4287-2023
FU Ministry of Education, Science, Sports and Culture of Japan [18 K19465]
FX Grant-in-Aid from the Ministry of Education, Science, Sports and Culture
   of Japan (18 K19465).
CR ANDO K, 1985, Nippon Acta Radiologica, V45, P531
   Apra C, 2018, EXPERT REV NEUROTHER, V18, P241, DOI 10.1080/14737175.2018.1429920
   Claus EB, 2005, NEUROSURGERY, V57, P1088, DOI 10.1227/01.NEU.0000188281.91351.B9
   Commins Deborah L, 2007, Neurosurg Focus, V23, pE3
   Condra KS, 1997, INT J RADIAT ONCOL, V39, P427, DOI 10.1016/S0360-3016(97)00317-9
   Couldwell WT, 1996, J NEUROSURG, V84, P20, DOI 10.3171/jns.1996.84.1.0020
   Davidson SE, 2002, CLIN ONCOL-UK, V14, P68, DOI 10.1053/clon.2001.0029
   Dennis ER, 2013, TECHNOL CANCER RES T, V12, P1, DOI 10.7785/tcrt.2012.500276
   El Shafie RA, 2018, RADIAT ONCOL, V13, DOI 10.1186/s13014-018-1002-5
   Euskirchen P, 2018, PRESSE MED, V47, pE247, DOI 10.1016/j.lpm.2018.05.016
   Gerelchuluun A, 2011, INT J RADIAT BIOL, V87, P57, DOI 10.3109/09553002.2010.518201
   Ichimura S, 2019, J NEUROL SURG PART B, V80, P474, DOI 10.1055/s-0038-1676373
   Kagei Kenji, 2004, Nippon Acta Radiologica, V64, P225
   Kosaki K, 2012, RADIAT ONCOL, V7, DOI 10.1186/1748-717X-7-44
   Kuratsu J, 2000, J NEUROSURG, V92, P766, DOI 10.3171/jns.2000.92.5.0766
   Lesueur P, 2019, CANCER TREAT REV, V72, P56, DOI 10.1016/j.ctrv.2018.11.004
   LONGSTRETH WT, 1993, CANCER, V72, P639, DOI 10.1002/1097-0142(19930801)72:3<639::AID-CNCR2820720304>3.0.CO;2-P
   Maclean J, 2014, CLIN ONCOL-UK, V26, P51, DOI 10.1016/j.clon.2013.10.001
   Maclean J, 2013, INT J RADIAT ONCOL, V85, pE179, DOI 10.1016/j.ijrobp.2012.10.032
   Matsumoto Y, 2014, J RADIAT RES, V55, P816, DOI 10.1093/jrr/rrt230
   MAYBERG MR, 1986, J NEUROSURG, V65, P160, DOI 10.3171/jns.1986.65.2.0160
   Milker-Zabel S, 2007, INT J RADIAT ONCOL, V68, P858, DOI 10.1016/j.ijrobp.2006.12.073
   Mirian C, 2020, J NEUROL NEUROSUR PS, V91, P378, DOI 10.1136/jnnp-2019-322257
   MIRIMANOFF RO, 1985, J NEUROSURG, V62, P18, DOI 10.3171/jns.1985.62.1.0018
   Murray FR, 2017, INT J RADIAT ONCOL, V99, P1190, DOI 10.1016/j.ijrobp.2017.08.005
   Noel G, 2005, INT J RADIAT ONCOL, V62, P1412, DOI 10.1016/j.ijrobp.2004.12.048
   Ostrom Quinn T, 2019, Neuro Oncol, V21, pv1, DOI 10.1093/neuonc/noz150
   Palm A, 2007, ACTA ONCOL, V46, P462, DOI 10.1080/02841860701218626
   Pirzkall A, 2003, INT J RADIAT ONCOL, V55, P362, DOI 10.1016/S0360-3016(02)03809-9
   Pollock BE, 2017, INT J RADIAT ONCOL, V97, P919, DOI 10.1016/j.ijrobp.2017.01.004
   Rockhill J., 2007, NEUROSURG, V23, pE1, DOI DOI 10.3171/FOC-07/10/E1
   Rogers L, 2007, NEUROSURG FOCUS, P23, DOI DOI 10.3171/FOC-07/10/E4
   SAMII M, 1992, ACTA NEUROCHIR, V118, P27, DOI 10.1007/BF01400723
   Schneider U, 2007, INT J RADIAT ONCOL, V68, P892, DOI 10.1016/j.ijrobp.2007.02.029
   SEKHAR LN, 1994, J NEUROSURG, V81, P860, DOI 10.3171/jns.1994.81.6.0860
   TSUJII H, 1993, INT J RADIAT ONCOL, V25, P49, DOI 10.1016/0360-3016(93)90144-K
   U.S. Department of Health and Human Services National Institutes of Health NCI, COMMON TERMINOLOGY C
   Vernimmen FJ, 2001, INT J RADIAT ONCOL, V49, P99, DOI 10.1016/S0360-3016(00)01457-7
   Weber DC, 2012, INT J RADIAT ONCOL, V83, P865, DOI 10.1016/j.ijrobp.2011.08.027
   Weber DC, 2004, RADIOTHER ONCOL, V71, P251, DOI 10.1016/j.radonc.2004.02.011
   Wenkel E, 2000, INT J RADIAT ONCOL, V48, P1363, DOI 10.1016/S0360-3016(00)01411-5
   World Med Assoc, 2013, JAMA-J AM MED ASSOC, V310, P2191, DOI 10.1001/jama.2013.281053
NR 42
TC 0
Z9 0
U1 0
U2 0
PU OXFORD UNIV PRESS
PI OXFORD
PA GREAT CLARENDON ST, OXFORD OX2 6DP, ENGLAND
SN 0449-3060
EI 1349-9157
J9 J RADIAT RES
JI J. Radiat. Res.
PD MAY
PY 2021
VL 62
IS 3
BP 427
EP 437
DI 10.1093/jrr/rrab017
EA APR 2021
PG 11
WC Biology; Oncology; Radiology, Nuclear Medicine & Medical Imaging
WE Science Citation Index Expanded (SCI-EXPANDED)
SC Life Sciences & Biomedicine - Other Topics; Oncology; Radiology, Nuclear
   Medicine & Medical Imaging
GA US9VV
UT WOS:000697774500007
OA Green Published, Green Submitted, gold
DA 2023-08-27
ER

PT J
AU Chance, PF
   Cavalier, L
   Satran, D
   Pellegrino, JE
   Koenig, M
   Dobyns, WB
AF Chance, PF
   Cavalier, L
   Satran, D
   Pellegrino, JE
   Koenig, M
   Dobyns, WB
TI Clinical nosologic and genetic aspects of Joubert and related syndromes
SO JOURNAL OF CHILD NEUROLOGY
LA English
DT Article
ID JUVENILE NEPHRONOPHTHISIS; CHROMOSOMAL LOCALIZATION; CEREBELLAR VERMIS;
   MOLECULAR-CLONING; WNT GENES; SEQUENCE; ATAXIA; RETARDATION; EXPRESSION;
   DELETIONS
AB Joubert syndrome is an autosomal-recessive disorder characterized by cerebellar hypoplasia, hypotonia, developmental delay, abnormal respiratory patterns, and abnormal eye movements. The biochemical and genetic basis of Joubert syndrome is unknown and a specific chromosomal locus for this disorder has not been identified. Review of this disorder and related syndromes suggests that (1) hypoplasia of the cerebellar vermis in Joubert syndrome is frequently associated with a complex brain stem malformation represented as the "molar tooth sign" on magnetic resonance imaging, (2) the "molar tooth sign" could be present in association with the Dandy-Walker malformation and occipital encephalocele, (3) cerebellar hypoplasia is present in conditions related to Joubert syndrome such as Arima syndrome; Senior-Loken syndrome; cerebellar vermian hypoplasia, oligophrenia, congenital ataxia, coloboma, and hepatic fibrosis syndrome; and juvenile nephronophthisis due to NPH1 mutations, and (4) the brainstem-vermis malformation spectrum is probably caused by at least two and probably several genetic loci. We have ascertained previously a cohort of 50 patients with a putative diagnosis of Joubert syndrome in order to evaluate the presence of associated malformations, and to initiate studies leading to the identification of genes causing Joubert and related syndromes. Among the associated malformations found in patients ascertained as having Joubert syndrome, 8% of patients had polydactyly, 4% had ocular colobomas, 2% had renal cysts, and 2% had soft-tissue tumors of the tongue. The WNT1 gene has been tested as a candidate gene for Joubert syndrome based on its expression in the developing cerebellum and an associated mutation in the swaying mouse. A search for mutations in WNT1 in a series of patients with Joubert syndrome did not detect mutations at this locus. This analysis suggested that mutations in WNT1 might; not have a significant role in Joubert; syndrome, and other functional candidate genes related to development of the cerebellum need to be examined. A genome-wide linkage analysis carried out in 10 Joubert syndrome pedigrees did not identify a specific chromosomal locus for this disorder. This observation, along with those from clinical studies, provides further evidence that Joubert and related syndromes are genetically heterogeneous.
C1 Univ Washington, Sch Med, Dept Pediat, Div Genet & Dev, Seattle, WA 98195 USA.
   Univ Washington, Sch Med, Dept Neurol, Seattle, WA USA.
   Childrens Hosp & Reg Med Ctr, Seattle, WA USA.
   Inst Genet & Biol Mol & Cellulaire, Illkirch Graffenstaden, France.
   Case Western Reserve Univ, Sch Med, Dept Human Genet, Cleveland, OH USA.
   Univ Med & Dent New Jersey, Robert Wood Johnson Med Sch, Dept Obstet Gynecol & Reprod Sci, Div Clin Genet, New Brunswick, NJ USA.
C3 University of Washington; University of Washington Seattle; University
   of Washington; University of Washington Seattle; Seattle Children's
   Hospital; Institut National de la Sante et de la Recherche Medicale
   (Inserm); UDICE-French Research Universities; Universites de Strasbourg
   Etablissements Associes; Universite de Strasbourg; Case Western Reserve
   University; Rutgers State University New Brunswick; Rutgers State
   University Medical Center
RP Chance, PF (通讯作者)，Univ Washington, Sch Med, Dept Pediat, Div Genet & Dev, Box 356320, Seattle, WA 98195 USA.
RI Dobyns, William/AAI-2768-2020; Chance, Phillip/AAX-5826-2020; Dobyns,
   William Brian/ABC-6525-2021
OI Dobyns, William/0000-0002-7681-2844; 
FU PHS HHS [R0130804] Funding Source: Medline
CR Arima M, 1971, NO TO HATTATSU, V3, P330
   BOLTSHAUSER E, 1977, NEUROPADIATRIE, V8, P57, DOI 10.1055/s-0028-1091505
   BOUKNIGHT DP, 1994, P GREENWOOD GENET CT, V13, P15
   CLARK CC, 1993, GENOMICS, V18, P249, DOI 10.1006/geno.1993.1463
   DEKABAN AS, 1969, AM J OPHTHALMOL, V68, P1029, DOI 10.1016/0002-9394(69)93443-6
   Dib C, 1996, NATURE, V380, P152, DOI 10.1038/380152a0
   FLANNERY DB, 1994, D SMITH WORKSH, P97
   GYAPAY G, 1994, NAT GENET, V7, P246, DOI 10.1038/ng0694supp-246
   Gyapay Gabor, 1996, Methods (Orlando), V9, P91, DOI 10.1006/meth.1996.0012
   JOUBERT M, 1969, NEUROLOGY, V19, P813, DOI 10.1212/WNL.19.9.813
   JOYNER AL, 1987, GENE DEV, V1, P29, DOI 10.1101/gad.1.1.29
   JOYNER AL, 1985, CELL, V43, P29, DOI 10.1016/0092-8674(85)90009-1
   KOENIG M, 1987, CELL, V50, P509, DOI 10.1016/0092-8674(87)90504-6
   KOHLER A, 1993, GENOMICS, V15, P233, DOI 10.1006/geno.1993.1045
   Konrad M, 1996, HUM MOL GENET, V5, P367, DOI 10.1093/hmg/5.3.367
   LATHROP GM, 1985, AM J HUM GENET, V37, P482
   Leber T., 1871, GRAEFES ARCH CLIN EX, V17, P314
   Leber T.G., 1869, A VONGRAEFES ARCH OP, V15, P1, DOI DOI 10.1007/BF02721213
   LOKEN AC, 1961, ACTA PAEDIATR, V50, P177, DOI 10.1111/j.1651-2227.1961.tb08037.x
   Maria BL, 1997, J CHILD NEUROL, V12, P423, DOI 10.1177/088307389701200703
   MATSUZAKA T, 1986, J CHILD NEUROL, V1, P338, DOI 10.1177/088307388600100404
   MEDHIOUB M, 1994, GENOMICS, V22, P296, DOI 10.1006/geno.1994.1387
   MENENZES M, 1990, PEDIATR NEUROL, V6, P428, DOI 10.1016/0887-8994(90)90015-S
   MORTON NE, 1992, ANN HUM GENET, V56, P223, DOI 10.1111/j.1469-1809.1992.tb01147.x
   MUENKE M, 1990, AM J MED GENET, V35, P360
   NADAL N, 1995, HUM MOL GENET, V4, P1963, DOI 10.1093/hmg/4.10.1963
   NOBLE KG, 1978, ARCH OPHTHALMOL-CHIC, V96, P818, DOI 10.1001/archopht.1978.03910050424004
   NUSSE R, 1992, CELL, V69, P1073, DOI 10.1016/0092-8674(92)90630-U
   NUSSE R, 1982, CELL, V31, P99, DOI 10.1016/0092-8674(82)90409-3
   Nystuen A, 1996, HUM MOL GENET, V5, P525, DOI 10.1093/hmg/5.4.525
   OTT J, 1909, ANAL HUMAN GENETIC L, P111
   PARR BA, 1993, DEVELOPMENT, V119, P247
   Pellegrino JE, 1997, AM J MED GENET, V72, P59, DOI 10.1002/(SICI)1096-8628(19971003)72:1<59::AID-AJMG12>3.0.CO;2-T
   POOLE SJ, 1989, GENOMICS, V4, P225, DOI 10.1016/0888-7543(89)90324-8
   REARDON W, 1989, J MED GENET, V26, P659, DOI 10.1136/jmg.26.10.659
   ROELINK H, 1993, GENOMICS, V17, P790, DOI 10.1006/geno.1993.1412
   Saar K, 1998, EUR J HUM GENET, V6, P501, DOI 10.1038/sj.ejhg.5200241
   SARAIVA JM, 1992, AM J MED GENET, V43, P726, DOI 10.1002/ajmg.1320430415
   Saunier S, 1997, AM J HUM GENET, V61, pA346
   Semina EV, 1996, NAT GENET, V14, P392, DOI 10.1038/ng1296-392
   SENIOR B, 1961, AM J OPHTHALMOL, V52, P625, DOI 10.1016/0002-9394(61)90147-7
   Steinlin M, 1997, NEUROPEDIATRICS, V28, P204, DOI 10.1055/s-2007-973701
   THOMAS KR, 1991, CELL, V67, P969, DOI 10.1016/0092-8674(91)90369-A
   Toriello H V, 1993, Clin Dysmorphol, V2, P95
   VANOOYEN A, 1985, EMBO J, V4, P2905, DOI 10.1002/j.1460-2075.1985.tb04021.x
   VARADI V, 1980, J MED GENET, V17, P119, DOI 10.1136/jmg.17.2.119
   VERLOES A, 1989, AM J MED GENET, V32, P227, DOI 10.1002/ajmg.1320320217
   VISKOCHIL D, 1990, CELL, V62, P187, DOI 10.1016/0092-8674(90)90252-A
   WAINWRIGHT BJ, 1988, EMBO J, V7, P1743, DOI 10.1002/j.1460-2075.1988.tb03003.x
   Wayne S, 1996, HUM MOL GENET, V5, P1689, DOI 10.1093/hmg/5.10.1689
   WURST W, 1994, DEVELOPMENT, V120, P2065
NR 51
TC 70
Z9 70
U1 0
U2 2
PU DECKER PERIODICALS INC
PI HAMILTON
PA 4 HUGHSON STREET SOUTH PO BOX 620, LCD 1, HAMILTON, ONTARIO L8N 3K7,
   CANADA
SN 0883-0738
J9 J CHILD NEUROL
JI J. Child Neurol.
PD OCT
PY 1999
VL 14
IS 10
BP 660
EP 666
DI 10.1177/088307389901401007
PG 7
WC Clinical Neurology; Pediatrics
WE Science Citation Index Expanded (SCI-EXPANDED)
SC Neurosciences & Neurology; Pediatrics
GA 240DE
UT WOS:000082809900007
PM 10511339
DA 2023-08-27
ER

PT J
AU Berg, AL
   Spitzer, JB
   Garvin, JH
AF Berg, AL
   Spitzer, JB
   Garvin, JH
TI Ototoxic impact of cisplatin in pediatric oncology patients
SO LARYNGOSCOPE
LA English
DT Article
DE hearing loss; sensorineural hearing loss; middle ear disease; cisplatin
   ototoxicity; pediatric
ID BRAIN-STEM RESPONSE; HIGH-DOSE CISPLATIN; CANCER-STUDY-GROUP;
   HEARING-LOSS; RECEIVING CISPLATIN; RISK FACTOR; CHILDREN; CHEMOTHERAPY;
   TUMORS; SENSITIVITY
AB Objective: To describe hearing changes in a group of 28 children (age range, 8-180 mo) undergoing protocol-based cisplatin therapy, Methods: Conventional, play audiometry, visual reinforcement audiometry (VRA), immittance audiometry, transient click evoked otoacoustic emissions (OAEs), and auditory brainstem response (ABR) evoked potentials were used to assess peripheral sensitivity and for threshold determination. Results: Bilateral symmetrical high-frequency sensorineural hearing loss was noted in 9 of the 28 children (26%). Hearing loss was evident as early as 1 month after chemotherapy and as late as 50 months and was not dependent on individual or cumulative dosage of cisplatin, Conclusions: 1) Presence of sensorineural hearing loss was independent of individual and/or cumulative dosage of cisplatin; 2) audiologic assessment should be incorporated into a child's periodic medical evaluations after chemotherapy treatment, as onset of sensorineural hearing loss cannot be predicted; 3) personal hearing aids may be indicated for those children with hearing loss affecting the low- to mid-frequencies; a personal assistive listening device (frequency modulated system) may be more appropriate for losses above 3000 Hz; and 4) evaluation and intervention by a speech-language pathologist may be indicated to address possible articulation or language development problems consequent to hearing loss.
C1 Pace Univ, Dyson Coll Arts & Sci, Dept Speech Commun Studies, New York, NY 10038 USA.
   Presbyterian Hosp, Dept Otolaryngol Head & Neck Surg, Div Audiol & Speech Language Pathol, New York, NY USA.
   Presbyterian Hosp, Dept Pediat, Div Hematol & Oncol, New York, NY USA.
   Columbia Univ Coll Phys & Surg, New York, NY 10032 USA.
C3 Pace University; NewYork-Presbyterian Hospital; NewYork-Presbyterian
   Hospital; Columbia University
RP Berg, AL (通讯作者)，Pace Univ, Dyson Coll Arts & Sci, Dept Speech Commun Studies, 1 Pace Plaza, New York, NY 10038 USA.
CR ABLIN AR, 1991, J CLIN ONCOL, V9, P1782, DOI 10.1200/JCO.1991.9.10.1782
   American Speech-Language-Hearing Association, 1997, GUID AUD SCREEN
   *ASHA PROF SERV BO, 1991, ASHA GUID ANSI STAND
   BARRHAMILTON RM, 1991, J LARYNGOL OTOL, V105, P7, DOI 10.1017/S0022215100114689
   BAUCH CD, 1986, EAR HEARING, V7, P314, DOI 10.1097/00003446-198610000-00004
   BAUM ES, 1979, CANCER TREAT REP, V63, P1621
   BLAKLEY BW, 1994, ARCH OTOLARYNGOL, V120, P541
   BLAKLEY BW, 1993, OTOLARYNG HEAD NECK, V109, P385, DOI 10.1177/019459989310900302
   BROCK PR, 1991, MED PEDIATR ONCOL, V19, P295, DOI 10.1002/mpo.2950190415
   CASSELBRANT ML, 1985, LARYNGOSCOPE, V95, P428
   COATS AC, 1977, ARCH OTOLARYNGOL, V103, P605
   COHEN BH, 1991, PEDIATR NEUROSURG, V16, P292, DOI 10.1159/000120545
   DINI G, 1987, MED PEDIATR ONCOL, V15, P18, DOI 10.1002/mpo.2950150105
   FAUSTI SA, 1993, ARCH OTOLARYNGOL, V119, P661
   FAUSTI SA, 1984, CANCER, V53, P224, DOI 10.1002/1097-0142(19840115)53:2<224::AID-CNCR2820530207>3.0.CO;2-D
   KHAN AB, 1982, CANCER TREAT REP, V66, P2013
   KONISHI T, 1988, AM J OTOLARYNG, V4, P18
   KOPELMAN J, 1988, LARYNGOSCOPE, V98, P858
   KRETSCHMAR CS, 1990, J CLIN ONCOL, V8, P1191, DOI 10.1200/JCO.1990.8.7.1191
   LAURELL G, 1990, LARYNGOSCOPE, V100, P724
   LAUTERMANN J, 1995, HEARING RES, V88, P47, DOI 10.1016/0378-5955(95)00097-N
   Lautermann J, 1995, LARYNGO RHINO OTOL, V74, P724, DOI 10.1055/s-2007-997833
   MARGOLIS RH, 1991, HEARING ASSESSMENT, P170
   MAXON AB, 1982, EAR HEARING, V3, P301, DOI 10.1097/00003446-198211000-00003
   MCALPINE D, 1990, HEARING RES, V47, P191, DOI 10.1016/0378-5955(90)90151-E
   MCHANEY VA, 1983, J PEDIATR-US, V102, P314, DOI 10.1016/S0022-3476(83)80551-4
   MOKOTOFF B, 1977, ARCH OTOLARYNGOL, V103, P38
   MURAKAMI T, 1990, SELECT CANCER THER, V6, P145, DOI 10.1089/sct.1990.6.145
   OATES P, 1992, EAR HEARING, V13, P28, DOI 10.1097/00003446-199202000-00007
   Olsen W. O., 1991, HEARING ASSESSMENT, P39
   PACKER RJ, 1994, J NEUROSURG, V81, P690, DOI 10.3171/jns.1994.81.5.0690
   PASIC TR, 1991, LARYNGOSCOPE, V101, P985
   POLLERA CF, 1988, CANCER CHEMOTH PHARM, V21, P61
   REDDEL RR, 1982, CANCER TREAT REP, V66, P19
   ROSEN G, 1982, CANCER-AM CANCER SOC, V49, P1221, DOI 10.1002/1097-0142(19820315)49:6<1221::AID-CNCR2820490625>3.0.CO;2-E
   SCHAEFER SD, 1985, CANCER, V56, P1934, DOI 10.1002/1097-0142(19851015)56:8<1934::AID-CNCR2820560807>3.0.CO;2-F
   SCHEUREN F, 1989, STATISTICS INCOME B, V8, P1
   Sie KCY, 1997, OTOLARYNG HEAD NECK, V116, P585, DOI 10.1016/S0194-5998(97)70232-8
   SKINNER R, 1990, BRIT J CANCER, V61, P927, DOI 10.1038/bjc.1990.208
   STAPELLS DR, 1982, J ACOUST SOC AM, V72, P74, DOI 10.1121/1.388026
   Stram DO, 1996, J CLIN ONCOL, V14, P2417, DOI 10.1200/JCO.1996.14.9.2417
   TANGE RA, 1985, ARCH OTO-RHINO-LARYN, V242, P77, DOI 10.1007/BF00464411
   Tharpe A M, 1993, J Am Acad Audiol, V4, P80
   THOMPSON G, 1984, SEMIN HEAR, V15, P85
   TODD NW, 1995, J LARYNGOL OTOL, V109, P926, DOI 10.1017/S002221510013169X
   WATERS GS, 1991, EAR HEARING, V12, P91, DOI 10.1097/00003446-199104000-00003
   WEATHERLY RA, 1991, LARYNGOSCOPE, V101, P917
   Weber BA., 1994, HDB CLIN AUDIOLOGY, P375
   WIDEN J E, 1990, Seminars in Hearing, V11, P342, DOI 10.1055/s-0028-1085514
   WILBUR LA, 1991, HEARING ASSESSMENT, P1
NR 50
TC 45
Z9 52
U1 0
U2 2
PU WILEY
PI HOBOKEN
PA 111 RIVER ST, HOBOKEN 07030-5774, NJ USA
SN 0023-852X
EI 1531-4995
J9 LARYNGOSCOPE
JI Laryngoscope
PD NOV
PY 1999
VL 109
IS 11
BP 1806
EP 1814
DI 10.1097/00005537-199911000-00016
PG 9
WC Medicine, Research & Experimental; Otorhinolaryngology
WE Science Citation Index Expanded (SCI-EXPANDED)
SC Research & Experimental Medicine; Otorhinolaryngology
GA 253LA
UT WOS:000083557000016
PM 10569412
DA 2023-08-27
ER

PT J
AU McGirt, MJ
   Woodworth, GF
   Coon, AL
   Frazier, JM
   Amundson, E
   Garonzik, I
   Olivi, A
   Weingart, JD
AF McGirt, MJ
   Woodworth, GF
   Coon, AL
   Frazier, JM
   Amundson, E
   Garonzik, I
   Olivi, A
   Weingart, JD
TI Independent predictors of morbidity after image-guided stereotactic
   brain biopsy: a risk assessment of 270 cases
SO JOURNAL OF NEUROSURGERY
LA English
DT Article
DE stereotactic biopsy; glioma grade; craniotomy; pathology
ID SPACE-OCCUPYING LESIONS; HEAD-INJURY; HYPERGLYCEMIA; TUMORS; TOMOGRAPHY;
   EXPERIENCE; GLIOMAS; YIELD
AB Object. Image-guided stereotactic brain biopsy is associated with transient and permanent incidences of morbidity in 9 and 4.5% of patients, respectively. The goal of this study was to perform a critical analysis of risk factors predictive of an enhanced operative risk in frame-based and frarneless stereotactic brain biopsy.
   Methods. The authors reviewed the clinical and neuroirriaging records of 270 patients who underwent consecutive frame-based and frameless image-guided stereotactic brain biopsies. The association between preoperative variables and biopsy-related morbidity was assessed by performing a multivariate logistic regression analysis.
   Transient and permanent stereotactic biopsy-related morbidity was observed in 23 (9%) and 13 (5%) patients, respectively. A hematoma occurred at the biopsy site in 25 patients (9%); 10 patients (4%) were symptomatic. Diabetes mellitus (odds ratio [OR] 3.73, 95% confidence interval [CI] 1.37-10.17, p = 0.01), thalamic lesions (OR 4.06, 95% CI 163-10.11, p = 0.002), and basal ganglia lesions (OR 3.29, 95% CI 1.05-10.25, p = 0.04) were independent risk factors for morbidity. In diabetic patients, a serum level of glucose that was greater than 200 rng/dl on the day of biopsy had a 100% positive predictive value and a glucose level lower than 200 mg/dl on the same day had a 95% negative predictive value for biopsy-related morbidity. Pontine biopsy was not a risk factor for morbidity. Only two (4%) of 45 patients who had epilepsy before the biopsy experienced seizures postoperatively. The creation of more than one needle trajectory increased the incidence of neurological deficits from 17 to 44% when associated with the treatment of deep lesions (those in the basal ganglia or thalamus-, p = 0.05), but was not associated with morbidity when associated with the treatment of cortex lesions.
   Conclusions.. Basal ganglia lesions, thalamic lesions, and patients with diabetes were independent risk factors for biopsy-associated morbidity. Hyperglycemia oil the day of biopsy predicted morbidity in the diabetic population. Epilepsy did not predispose to biopsy-associated seizure. For deep-seated lesions, increasing the number of biopsy samples along an established track rather than performing a second trajectory may minimize the incidence of morbidity. Close perioperative observation of glucose levels may be warranted.
C1 Johns Hopkins Sch Med, Dept Neurosurg, Baltimore, MD 21287 USA.
   Johns Hopkins Sch Med, Dept Oncol, Baltimore, MD 21287 USA.
C3 Johns Hopkins University; Johns Hopkins Medicine; Johns Hopkins
   University; Johns Hopkins Medicine
RP McGirt, MJ (通讯作者)，Johns Hopkins Sch Med, Dept Neurosurg, 600 N Wolfe St,Meyer 8-161, Baltimore, MD 21287 USA.
EM
RI Olivi, Alessandro/K-6531-2016
OI Olivi, Alessandro/0000-0002-4489-7564
CR APUZZO MLJ, 1983, NEUROSURGERY, V12, P277, DOI 10.1227/00006123-198303000-00005
   APUZZO MLJ, 1987, NEUROSURGERY, V20, P930, DOI 10.1227/00006123-198706000-00019
   Bernays RL, 2002, J NEUROSURG, V97, P354, DOI 10.3171/jns.2002.97.2.0354
   Blaauw G, 1988, Acta Neurochir Suppl (Wien), V42, P161
   BOETHIUS J, 1978, ACTA NEUROCHIR, V40, P223, DOI 10.1007/BF01774748
   BOUVIER G, 1983, APPL NEUROPHYSIOL, V46, P227
   Boviatsis EJ, 2003, NEUROL SCI, V24, P97, DOI 10.1007/s10072-003-0093-3
   Bruno A, 2002, NEUROLOGY, V59, P669, DOI 10.1212/WNL.59.5.669
   Cherian L, 1998, J NEUROTRAUM, V15, P307, DOI 10.1089/neu.1998.15.307
   Dorward NL, 2002, BRIT J NEUROSURG, V16, P110, DOI 10.1080/02688690220131705
   Field P, 2001, J NEUROSURG, V94, P545, DOI 10.3171/jns.2001.94.4.0545
   FRIEDMAN WA, 1989, NEUROSURGERY, V25, P180, DOI 10.1227/00006123-198908000-00005
   Gralla J, 2003, ZBL NEUROCHIR, V64, P166
   GREENE GM, 1989, J NEUROSURG, V71, P494, DOI 10.3171/jns.1989.71.4.0494
   Hall WA, 1998, CANCER, V82, P1749, DOI 10.1002/(SICI)1097-0142(19980501)82:9<1756::AID-CNCR23>3.0.CO;2-2
   Heilbrun M P, 1983, Clin Neurosurg, V31, P564
   Kawai N, 1998, ACT NEUR S, V71, P219
   KELLY PJ, 1985, MAYO CLIN PROC, V60, P223, DOI 10.1016/S0025-6196(12)60314-0
   Kulkarni AV, 1998, J NEUROSURG, V89, P31, DOI 10.3171/jns.1998.89.1.0031
   LAM AM, 1991, J NEUROSURG, V75, P545, DOI 10.3171/jns.1991.75.4.0545
   LEE T, 1991, British Journal of Neurosurgery, V5, P331, DOI 10.3109/02688699109002859
   LOBATO RD, 1982, APPL NEUROPHYSIOL, V45, P426
   Mundinger F, 1985, Acta Neurochir Suppl (Wien), V35, P70
   Niizuma H, 1988, Acta Neurochir Suppl (Wien), V42, P157
   OSTERTAG CB, 1980, SURG NEUROL, V14, P275
   Ritter AM, 1996, J NEUROTRAUM, V13, P473, DOI 10.1089/neu.1996.13.473
   Rovlias A, 2000, NEUROSURGERY, V46, P335, DOI 10.1097/00006123-200002000-00015
   Sawin PD, 1998, SURG NEUROL, V49, P640, DOI 10.1016/S0090-3019(97)00435-7
   WILD A M, 1990, British Journal of Neurosurgery, V4, P407, DOI 10.3109/02688699008992763
   Yamada K, 2004, J CLIN NEUROSCI, V11, P263, DOI 10.1016/j.jocn.2003.03.004
   Yu X, 2000, STEREOT FUNCT NEUROS, V75, P103, DOI 10.1159/000048390
NR 31
TC 165
Z9 170
U1 0
U2 1
PU AMER ASSOC NEUROLOGICAL SURGEONS
PI ROLLING MEADOWS
PA 5550 MEADOWBROOK DRIVE, ROLLING MEADOWS, IL 60008 USA
SN 0022-3085
EI 1933-0693
J9 J NEUROSURG
JI J. Neurosurg.
PD MAY
PY 2005
VL 102
IS 5
BP 897
EP 901
DI 10.3171/jns.2005.102.5.0897
PG 5
WC Clinical Neurology; Surgery
WE Science Citation Index Expanded (SCI-EXPANDED)
SC Neurosciences & Neurology; Surgery
GA 954KO
UT WOS:000231154200015
PM 15926716
DA 2023-08-27
ER

PT J
AU Eshraghi, AA
   Wolfovitz, A
   Yilmazer, R
   Garnham, C
   Yilmazer, AB
   Bas, E
   Ashman, P
   Roell, J
   Bohorquez, J
   Mittal, R
   Hessler, R
   Sieber, D
   Mittal, J
AF Eshraghi, Adrien A.
   Wolfovitz, Amit
   Yilmazer, Rasim
   Garnham, Carolyn
   Yilmazer, Ayca Baskadem
   Bas, Esperanza
   Ashman, Peter
   Roell, Jonathan
   Bohorquez, Jorge
   Mittal, Rahul
   Hessler, Roland
   Sieber, Daniel
   Mittal, Jeenu
TI Otoprotection to Implanted Cochlea Exposed to Noise Trauma With
   Dexamethasone Eluting Electrode
SO FRONTIERS IN CELLULAR NEUROSCIENCE
LA English
DT Article
DE drug-eluting electrodes; otoprotection; noise trauma; hearing loss;
   local drug delivery; dexamethasone; cochlear implantation
ID GROWTH-FACTOR 1; INDUCED HEARING-LOSS; RESIDUAL HEARING; DRUG-DELIVERY;
   RAT MODEL; PRESERVATION; MECHANISMS; PROTECTION; CELLS; REGENERATION
AB Cochlear implantation (CI) is now widely used to provide auditory rehabilitation to individuals having severe to profound sensorineural hearing loss (SNHL). However, CI can lead to electrode insertion trauma (EIT) that can cause damage to sensory cells in the inner ear resulting in loss of residual hearing. Even with soft surgical techniques where there is minimal macroscopic damage, we can still observe the generation of molecular events that may initiate programmed cell death via various mechanisms such as oxidative stress, the release of pro-inflammatory cytokines, and activation of the caspase pathway. In addition, individuals with CI may be exposed to noise trauma (NT) due to occupation and leisure activities that may affect their hearing ability. Recently, there has been an increased interest in the auditory community to determine the efficacy of drug-eluting electrodes for the protection of residual hearing. The objective of this study is to determine the effect of NT on implanted cochlea as well as the otoprotective efficacy of dexamethasone eluting electrode to implanted cochlea exposed to NT in a guinea pig model of CI. Animals were divided into five groups: EIT with dexamethasone eluting electrode exposed to NT; EIT exposed to NT; NT only; EIT only and naive animals (control group). The hearing thresholds were determined by auditory brainstem recordings (ABRs). The cochlea was harvested and analyzed for transcript levels of inflammation, apoptosis and fibrosis genes. We observed that threshold shifts were significantly higher in EIT, NT or EIT + NT groups compared to naive animals at all the tested frequencies. The dexamethasone eluting electrode led to a significant decrease in hearing threshold shifts in implanted animals exposed to NT. Proapoptotic tumor necrosis factor-alpha [TNF-alpha, TNF-alpha receptor 1a (TNF alpha R1a)] and pro-fibrotic transforming growth factor beta 1 (TGF beta) genes were more than two-fold up-regulated following EIT and EIT + NT compared to the control group. The use of dexamethasone releasing electrode significantly decreased the transcript levels of pro-apoptotic and pro-fibrotic genes. The dexamethasone releasing electrode has shown promising results for hearing protection in implanted animals exposed to NT. The results of this study suggest that dexamethasone releasing electrode holds great potential in developing effective treatment modalities for NT in the implanted cochlea.
C1 [Eshraghi, Adrien A.; Wolfovitz, Amit; Yilmazer, Rasim; Yilmazer, Ayca Baskadem; Bas, Esperanza; Ashman, Peter; Roell, Jonathan; Mittal, Rahul; Mittal, Jeenu] Univ Miami, Miller Sch Med, Dept Otolaryngol, Hearing Res Lab, Miami, FL 33136 USA.
   [Eshraghi, Adrien A.] Miller Sch Med, Dept Neurol Surg, Miami, FL 33136 USA.
   [Eshraghi, Adrien A.; Bohorquez, Jorge] Univ Miami, Dept Biomed Engn, Coral Gables, FL 33124 USA.
   [Garnham, Carolyn; Hessler, Roland; Sieber, Daniel] MED EL Hearing Implants, Innsbruck, Austria.
C3 University of Miami; University of Miami
RP Eshraghi, AA (通讯作者)，Univ Miami, Miller Sch Med, Dept Otolaryngol, Hearing Res Lab, Miami, FL 33136 USA.; Eshraghi, AA (通讯作者)，Miller Sch Med, Dept Neurol Surg, Miami, FL 33136 USA.; Eshraghi, AA (通讯作者)，Univ Miami, Dept Biomed Engn, Coral Gables, FL 33124 USA.
EM
RI Yılmazer, Rasim/AAB-2648-2021; Baskadem Yilmazer, Ayca/JAX-8872-2023
OI Baskadem Yilmazer, Ayca/0000-0001-9967-8046; Wolfovitz,
   Amit/0000-0001-5714-5452; Sieber, Daniel/0000-0001-7126-9160
FU MED-EL Corporation
FX This work was supported by a research grant from MED-EL Corporation to
   AE.
CR Aggarwal BB, 2003, NAT REV IMMUNOL, V3, P745, DOI 10.1038/nri1184
   Akcakaya H, 2019, COCHLEAR IMPLANTS IN, V20, P62, DOI 10.1080/14670100.2019.1565077
   Bach LA, 2015, PEDIATR ENDOCR REV P, V13, P521
   Bas E, 2020, ANAT REC, V303, P608, DOI 10.1002/ar.24064
   Bas E, 2016, HEARING RES, V337, P12, DOI 10.1016/j.heares.2016.02.003
   Bas E, 2012, ANAT REC, V295, P1944, DOI 10.1002/ar.22585
   Camarero G, 2001, J NEUROSCI, V21, P7630, DOI 10.1523/JNEUROSCI.21-19-07630.2001
   Chen HC, 2019, NEUROSCIENCE, V400, P62, DOI 10.1016/j.neuroscience.2018.12.048
   Dedhia K, 2016, OTOL NEUROTOL, V37, P478, DOI 10.1097/MAO.0000000000001011
   Digicaylioglu M, 2004, P NATL ACAD SCI USA, V101, P9855, DOI 10.1073/pnas.0403172101
   Dinh CT, 2015, FRONT CELL NEUROSCI, V9, DOI 10.3389/fncel.2015.00096
   Douchement D, 2015, Cochlear Implants Int, V16, P195, DOI 10.1179/1754762813Y.0000000053
   Eshraghi A A, 2011, Cochlear Implants Int, V12 Suppl 1, pS51, DOI 10.1179/146701011X13001035753254
   Eshraghi AA, 2006, ANAT REC PART A, V288A, P473, DOI 10.1002/ar.a.20305
   Eshraghi AA, 2005, OTOL NEUROTOL, V26, P442, DOI 10.1097/01.mao.0000169791.53201.e1
   Eshraghi AA, 2017, ACTA OTO-LARYNGOL, V137, P384, DOI 10.1080/00016489.2016.1256499
   Eshraghi AA, 2015, ACTA OTO-LARYNGOL, V135, P328, DOI 10.3109/00016489.2015.1012276
   Eshraghi AA, 2013, LARYNGOSCOPE, V123, pS1, DOI 10.1002/lary.23902
   Eshraghi AA, 2012, ANAT REC, V295, P1967, DOI 10.1002/ar.22580
   Eshraghi Adrien A, 2010, Cochlear Implants Int, V11 Suppl 1, P104, DOI 10.1179/146701010X12671177544104
   Farhadi Mohammad, 2013, Cochlear Implants Int, V14, P45
   Fujioka M, 2006, J NEUROSCI RES, V83, P575, DOI 10.1002/jnr.20764
   Ghavi FF, 2010, J BIOMED MATER RES B, V94B, P388, DOI 10.1002/jbm.b.31666
   Hayashi Y, 2014, NEUROSCI LETT, V560, P7, DOI 10.1016/j.neulet.2013.11.062
   Hayashi Y, 2013, MOL CELL NEUROSCI, V56, P29, DOI 10.1016/j.mcn.2013.03.003
   Hayden MS, 2014, SEMIN IMMUNOL, V26, P253, DOI 10.1016/j.smim.2014.05.004
   Housley GD, 2013, P NATL ACAD SCI USA, V110, P7494, DOI 10.1073/pnas.1222295110
   Iwai K, 2006, LARYNGOSCOPE, V116, P529, DOI 10.1097/01.mlg.0000200791.77819.eb
   Jia H, 2013, ANN OTO RHINOL LARYN, V122, P33, DOI 10.1177/000348941312200107
   Khater A, 2017, INT ARCH OTORHINOLAR, V21, P297, DOI 10.1055/s-0036-1585094
   Lazard DS, 2012, PLOS ONE, V7, DOI 10.1371/journal.pone.0048739
   Lee KY, 2007, OTOL NEUROTOL, V28, P976, DOI 10.1097/MAO.0b013e31811f40db
   Lenarz T, 2013, CURR PHARM BIOTECHNO, V14, P112
   Liu Y, 2015, HEARING RES, V327, P89, DOI 10.1016/j.heares.2015.04.019
   Livak KJ, 2001, METHODS, V25, P402, DOI 10.1006/meth.2001.1262
   Macherey O, 2014, CURR BIOL, V24, pR878, DOI 10.1016/j.cub.2014.06.053
   Miranda Priscila Carvalho, 2014, Int J Otolaryngol, V2014, P468515, DOI 10.1155/2014/468515
   Morrill Seth, 2017, J Otol, V12, P151, DOI 10.1016/j.joto.2017.08.001
   Paasche G, 2003, OTOL NEUROTOL, V24, P222, DOI 10.1097/00129492-200303000-00016
   Quesnel AM, 2016, HEARING RES, V333, P225, DOI 10.1016/j.heares.2015.08.018
   Raveh E, 2015, EUR ARCH OTO-RHINO-L, V272, P2261, DOI 10.1007/s00405-014-3184-2
   Schmitt H, 2018, OTOL NEUROTOL, V39, P37, DOI 10.1097/MAO.0000000000001625
   SIMPSON TH, 1993, EAR HEARING, V14, P322, DOI 10.1097/00003446-199310000-00003
   Tuerdi A, 2017, NEUROSCI LETT, V642, P123, DOI 10.1016/j.neulet.2017.02.003
   Vlajkovic SM, 2017, HEARING RES, V345, P43, DOI 10.1016/j.heares.2016.12.015
   Warnecke A, 2019, FRONT NEUROL, V10, DOI 10.3389/fneur.2019.00665
   Wilk M, 2016, PLOS ONE, V11, DOI 10.1371/journal.pone.0147552
   Yamahara K, 2019, HEARING RES, V374, P5, DOI 10.1016/j.heares.2019.01.008
   Yamahara K, 2015, HEARING RES, V330, P2, DOI 10.1016/j.heares.2015.04.009
   Yamamoto N, 2014, FRONT PHARMACOL, V5, DOI 10.3389/fphar.2014.00208
   Yang D, 2015, ACTA OTO-LARYNGOL, V135, P622, DOI 10.3109/00016489.2014.1003092
   Yu Y, 2009, J BIOMED SCI, V16, DOI 10.1186/1423-0127-16-79
NR 52
TC 9
Z9 9
U1 1
U2 6
PU FRONTIERS MEDIA SA
PI LAUSANNE
PA AVENUE DU TRIBUNAL FEDERAL 34, LAUSANNE, CH-1015, SWITZERLAND
EI 1662-5102
J9 FRONT CELL NEUROSCI
JI Front. Cell. Neurosci.
PD NOV 22
PY 2019
VL 13
AR 492
DI 10.3389/fncel.2019.00492
PG 11
WC Neurosciences
WE Science Citation Index Expanded (SCI-EXPANDED)
SC Neurosciences & Neurology
GA JT8MB
UT WOS:000501236600001
PM 31824265
OA gold, Green Published
DA 2023-08-27
ER

PT J
AU Packer, RJ
   Gajjar, A
   Vezina, G
   Rorke-Adams, L
   Burger, PC
   Robertson, PL
   Bayer, L
   LaFond, D
   Donahue, BR
   Marymont, MH
   Muraszko, K
   Langston, J
   Sposto, R
AF Packer, Roger J.
   Gajjar, Amar
   Vezina, Gilbert
   Rorke-Adams, Lucy
   Burger, Peter C.
   Robertson, Patricia L.
   Bayer, Lisa
   LaFond, Deborah
   Donahue, Bernadine R.
   Marymont, MaryAnne H.
   Muraszko, Karin
   Langston, James
   Sposto, Richard
TI Phase III study of craniospinal radiation therapy followed by adjuvant
   chemotherapy for newly diagnosed average-risk medulloblastoma
SO JOURNAL OF CLINICAL ONCOLOGY
LA English
DT Article; Proceedings Paper
CT 11th International Symposium on Pediatric Neuro Oncology
CY JUN 13-16, 2004
CL Boston, MA
ID CHILDRENS CANCER GROUP; PROSPECTIVE RANDOMIZED-TRIAL;
   OF-PEDIATRIC-ONCOLOGY; POSTERIOR-FOSSA; CHILDHOOD MEDULLOBLASTOMA;
   PRERADIATION CHEMOTHERAPY; INTERNATIONAL-SOCIETY; BRAIN-TUMORS; CNS
   TUMORS; C-MYC
AB Purpose
   To determine the event-free survival (EFS) and overall survival of children with average-risk medulloblastoma and treated with reduced-dose craniospinal radiotherapy (CSRT) and one of two postradiotherapy chemotherapies.
   Methods
   Four hundred twenty-one patients between 3 years and 21 years of age with nondisseminated medulloblastoma (MB) were prospectively randomly assigned to treatment with 23.4 Gy of CSRT, 55.8 Gy of posterior fossa RT, plus one of two adjuvant chemotherapy regimens: lomustine (CCNU), cisplatin, and vincristine; or cyclophosphamide, cisplatin, and vincristine.
   Results
   Forty-two of 421 patients enrolled were excluded from analysis. Sixty-six of the remaining 379 patients had incompletely assessable postoperative studies. Five-year EFS and survival for the cohort of 379 patients was 81% +/- 2.1% and 86% +/- 9%, respectively (median follow-up over 5 years). EFS was unaffected by sex, race, age, treatment regimen, brainstem involvement, or excessive anaplasia. EFS was detrimentally affected by neuroradiographic unassessability. Patients with areas of frank dissemination had a 5-year EFS of 36% +/- 15%. Sixty-seven percent of progressions had some component of dissemination. There were seven second malignancies. Infections occurred more frequently on the cyclophosphamide arm and electrolyte abnormalities were more common on the CCNU regimen.
   Conclusion This study discloses an encouraging EFS rate for children with nondisseminated MB treated with reduced-dose craniospinal radiation and chemotherapy. Additional, careful, step-wise reductions in CSRT in adequately staged patients may be possible.
C1 Childrens Natl Med Ctr, Dept Neurol, Div Neurol, Washington, DC 20010 USA.
   Childrens Natl Med Ctr, Div Pediat, Washington, DC 20010 USA.
   Childrens Natl Med Ctr, Div Radiol, Washington, DC 20010 USA.
   Childrens Natl Med Ctr, Div Oncol, Washington, DC 20010 USA.
   George Washington Univ, Dept Neurol, Washington, DC USA.
   George Washington Univ, Dept Pediat, Washington, DC 20052 USA.
   George Washington Univ, Dept Radiol, Washington, DC 20052 USA.
   St Jude Childrens Res Hosp, Dept Oncol & Radiol, Memphis, TN USA.
   Childrens Hosp Philadelphia, Div Neuropathol, Philadelphia, PA 19104 USA.
   Univ Penn, Dept Pathol, Philadelphia, PA 19104 USA.
   Johns Hopkins Univ, Dept Pathol, Baltimore, MD USA.
   Univ Michigan, Dept Neurol Pediat & Neurosurg, Ann Arbor, MI 48109 USA.
   Cornell Univ, Weill Med Coll, Div Pediat Hematol Oncol, Ithaca, NY 14853 USA.
   NYU, Sch Med, Dept Radiat Oncol, New York, NY USA.
   Maimonides Canc Ctr, Dept Radiat Oncol, Brooklyn, NY USA.
   Childrens Mem Hosp, Dept Radiat Oncol, Chicago, IL USA.
   Northwestern Univ, Dept Radiat Oncol, Chicago, IL 60611 USA.
   Childrens Hosp Los Angeles, Los Angeles, CA 90027 USA.
   Univ So Calif, Keck Sch Med, Los Angeles, CA USA.
   Childrens Oncol Grp, Arcadia, CA USA.
C3 Children's National Health System; Children's National Health System;
   Children's National Health System; Children's National Health System;
   George Washington University; George Washington University; George
   Washington University; St Jude Children's Research Hospital; University
   of Pennsylvania; Pennsylvania Medicine; Childrens Hospital of
   Philadelphia; University of Pennsylvania; Johns Hopkins University;
   University of Michigan System; University of Michigan; Cornell
   University; Weill Cornell Medicine; New York University; Maimonides
   Medical Center; Ann & Robert H. Lurie Children's Hospital of Chicago;
   Northwestern University; Children's Hospital Los Angeles; University of
   Southern California; Children's Oncology Group (COG)
RP Packer, RJ (通讯作者)，Childrens Natl Med Ctr, Dept Neurol, Div Neurol, 111 Michigan Ave NW, Washington, DC 20010 USA.
EM
RI Gajjar, Amar/N-8059-2018
FU NCI NIH HHS [5UI0CA098543-02] Funding Source: Medline
CR ALLEN JC, 1981, J NEUROSURG, V55, P749, DOI 10.3171/jns.1981.55.5.0749
   [Anonymous], 2011, STAT ANAL FAILURE TI
   BAILEY CC, 1995, MED PEDIATR ONCOL, V25, P166, DOI 10.1002/mpo.2950250303
   CBTRUS, 2002, 1995 1999 STAT REP P
   Eberhart CG, 2004, J NEUROPATH EXP NEUR, V63, P441, DOI 10.1093/jnen/63.5.441
   FRIEDMAN HS, 1985, NEUROL CLIN, V3, P843, DOI 10.1016/S0733-8619(18)31013-2
   Gajjar A, 2004, J CLIN ONCOL, V22, P984, DOI 10.1200/JCO.2004.06.032
   GILBERTSON RJ, 1995, BRIT J CANCER, V71, P473, DOI 10.1038/bjc.1995.96
   Grotzer MA, 2001, CLIN CANCER RES, V7, P2425
   Grotzer MA, 2000, J CLIN ONCOL, V18, P1027, DOI 10.1200/JCO.2000.18.5.1027
   Herms J, 2000, INT J CANCER, V89, P395
   Kortmann RD, 2000, INT J RADIAT ONCOL, V46, P269, DOI 10.1016/S0360-3016(99)00369-7
   Kuhl J, 1998, KLIN PADIATR, V210, P227, DOI 10.1055/s-2008-1043883
   Lamont JM, 2004, CLIN CANCER RES, V10, P5482, DOI 10.1158/1078-0432.CCR-03-0721
   MacDonald TJ, 2003, ONCOLOGIST, V8, P174, DOI 10.1634/theoncologist.8-2-174
   MacDonald TJ, 2001, NAT GENET, V29, P143, DOI 10.1038/ng731
   Oyharcabal-Bourden V, 2005, J CLIN ONCOL, V23, P4726, DOI 10.1200/JCO.2005.00.760
   Packer R J, 1999, Neuro Oncol, V1, P232, DOI 10.1093/neuonc/1.3.232
   PACKER RJ, 1989, J NEUROSURG, V70, P707, DOI 10.3171/jns.1989.70.5.0707
   PACKER RJ, 1994, J NEUROSURG, V81, P690, DOI 10.3171/jns.1994.81.5.0690
   Packer RJ, 1999, J CLIN ONCOL, V17, P2127, DOI 10.1200/JCO.1999.17.7.2127
   PACKER RJ, 1990, ANN NEUROL, V28, P823, DOI 10.1002/ana.410280615
   Packer RJ, 2001, J CLIN ONCOL, V19, P480, DOI 10.1200/JCO.2001.19.2.480
   POLLACK IF, 1995, NEUROSURGERY, V37, P885, DOI 10.1227/00006123-199511000-00006
   Pomeroy SL, 2002, NATURE, V415, P436, DOI 10.1038/415436a
   Ray A, 2004, CLIN CANCER RES, V10, P7613, DOI 10.1158/1078-0432.CCR-04-0499
   Ris MD, 2001, J CLIN ONCOL, V19, P3470, DOI 10.1200/JCO.2001.19.15.3470
   Robertson PL, 2002, ANN NEUROL, V52, pS115
   Rood BR, 2002, CANCER RES, V62, P3794
   SEGAL RA, 1994, P NATL ACAD SCI USA, V91, P12867, DOI 10.1073/pnas.91.26.12867
   SPOSTO R, 1985, J CHRON DIS, V38, P683, DOI 10.1016/0021-9681(85)90022-0
   Sposto R, 2002, STAT MED, V21, P293, DOI 10.1002/sim.987
   Taylor RE, 2003, J CLIN ONCOL, V21, P1581, DOI 10.1200/JCO.2003.05.116
   Thomas PRM, 2000, J CLIN ONCOL, V18, P3004, DOI 10.1200/JCO.2000.18.16.3004
   Zeltzer PM, 1999, J CLIN ONCOL, V17, P832, DOI 10.1200/JCO.1999.17.3.832
NR 35
TC 643
Z9 666
U1 0
U2 17
PU AMER SOC CLINICAL ONCOLOGY
PI ALEXANDRIA
PA 2318 MILL ROAD, STE 800, ALEXANDRIA, VA 22314 USA
SN 0732-183X
J9 J CLIN ONCOL
JI J. Clin. Oncol.
PD SEP 1
PY 2006
VL 24
IS 25
BP 4202
EP 4208
DI 10.1200/JCO.2006.06.4980
PG 7
WC Oncology
WE Science Citation Index Expanded (SCI-EXPANDED); Conference Proceedings Citation Index - Science (CPCI-S)
SC Oncology
GA 081LV
UT WOS:000240320000026
PM 16943538
DA 2023-08-27
ER

PT J
AU Browne, CA
   O'Brien, FE
   Connor, TJ
   Dinan, TG
   Cryan, JF
AF Browne, C. A.
   O'Brien, F. E.
   Connor, T. J.
   Dinan, T. G.
   Cryan, J. F.
TI DIFFERENTIAL LIPOPOLYSACCHARIDE-INDUCED IMMUNE ALTERATIONS IN THE
   HIPPOCAMPUS OF TWO MOUSE STRAINS: EFFECTS OF STRESS
SO NEUROSCIENCE
LA English
DT Article
DE BALB/c; C57BL/6J; IDO; TPH; LPS; Swim-stress
ID DORSAL RAPHE NUCLEUS; TRYPTOPHAN-HYDROXYLASE ISOFORM;
   CORTICOTROPIN-RELEASING-FACTOR; BRAIN-SEROTONIN SYNTHESIS; INDOLEAMINE
   2,3-DIOXYGENASE; DEPRESSIVE SYMPTOMS; INTERFERON-ALPHA; MESSENGER-RNA;
   IFN-GAMMA; CANCER-PATIENTS
AB Immunological activation may result in the development of depressive-like symptoms in a large percentage of patients treated with cytokine-based therapies. The mechanisms underlying susceptibility to cytokine-induced depression are currently unknown; however activation of the tryptophan catabolising enzyme indoleamine 2,3-dioxygenase (IDO) is associated with the induction of cytokine-induced depression. Peripheral administration of lipopolysaccharide (LPS) is one of the most commonly used immunological challenges in animal models of cytokine-induced depression. Inbred mouse strains are useful tools in the investigation of the neurobiology of psychiatric illnesses. In this study we hypothesised that two strains which differ in stress susceptibility, namely the BALB/c and C57BL/6J mice, would respond differentially to LPS and swim-stress in cytokine profile, corticosterone concentrations and mRNA expression of genes coding for the tryptophan metabolising enzymes, 1001,1002, Tph1 and Tph2. The stress-sensitive BALB/c strain exhibited increased depressive-like behaviour and enhanced corticosterone concentrations in response to LPS. Furthermore, swim-stress attenuated the LPS-induced corticosterone response in BALB/c mice only. LPS significantly increased plasma interleukin (IL)-1 beta and tumour necrosis factor alpha (TNF alpha) concentrations to a greater extent in BALB/c mice. The LPS-induced increase in IL-1 beta mRNA expression was significantly attenuated by swim-stress in the hippocampus of C57BL/6J but not in BALB/c mice. TNF alpha mRNA expression was significantly increased in BALB/c mice only; this increase was attenuated by swim-stress. Tph1 mRNA expression was upregulated in the brainstem of C57BL/6J mice post-LPS and following the combination of swim-stress and LPS in BALB/c mice. In the hippocampus Tph1 and Tph2 mRNA expression was increased in C57BL/6J but not BALB/c mice in response to LPS challenge and swim-stress. Conversely, 1002 but not 1001 mRNA expression was significantly altered following swim-stress and LPS, particularly in the hippocampus of BALB/c mice. These data indicate altered central mRNA expression of tryptophan metabolising enzymes and immune activation in BALB/c mice compared to the normo-sensitive C57BL/6J strain. (C) 2012 IBRO. Published by Elsevier Ltd. All rights reserved.
C1 [Cryan, J. F.] Univ Coll Cork, Dept Anat & Neurosci, Cork, Ireland.
   [Browne, C. A.] Univ Coll Cork, Dept Pharmacol & Therapeut, Neuropharmacol Res Grp, Cork, Ireland.
   [O'Brien, F. E.; Dinan, T. G.; Cryan, J. F.] Univ Coll Cork, Biosci Inst, Alimentary Pharmabiot Ctr, Cork, Ireland.
   [O'Brien, F. E.] Univ Coll Cork, Sch Pharm, Cork, Ireland.
   [Connor, T. J.] Trinity Coll Dublin, Sch Med, Dublin 2, Ireland.
   [Connor, T. J.] Trinity Coll Dublin, Inst Neurosci, Dublin 2, Ireland.
   [Dinan, T. G.] Univ Coll Cork, Dept Psychiat, Cork, Ireland.
C3 University College Cork; University College Cork; University College
   Cork; University College Cork; Trinity College Dublin; Trinity College
   Dublin; University College Cork
RP Cryan, JF (通讯作者)，Univ Coll Cork, Dept Anat & Neurosci, Cork, Ireland.
EM
RI Browne, Caroline/ADZ-5051-2022; Cryan, John F./A-6950-2013; Dinan,
   Timothy/ABA-8284-2020
OI Cryan, John F./0000-0001-5887-2723; Browne,
   Caroline/0000-0001-7463-7870; Dinan, Timothy/0000-0002-2316-7220
FU European Community [FP7/2007-2013, 201714]; Science Foundation Ireland
   through the Irish Government [02/CE/B124, 07/CE/B1368]
FX J.F.C. is funded by European Community's Seventh Framework Programme;
   Grant Number: FP7/2007-2013, Grant Agreement 201714. J.F.C. and T.G.D.
   are also supported in part by Science Foundation Ireland in the form of
   a centre grant for the Alimentary Pharmabiotic Centre, through the Irish
   Government's National Development plan (Grant number: 02/CE/B124 and
   07/CE/B1368). We would like to acknowledge Dr. Olivia O'Leary and Dr.
   Andrew Harkin for the helpful advice in the preparation of this
   manuscript.
CR Abumaria N, 2008, CELL MOL NEUROBIOL, V28, P331, DOI 10.1007/s10571-007-9259-5
   Amat J, 2005, NAT NEUROSCI, V8, P365, DOI 10.1038/nn1399
   Andre C, 2008, J NEUROIMMUNOL, V200, P90, DOI 10.1016/j.jneuroim.2008.06.011
   [Anonymous], 2007, WHATS WRONG MY MOUSE
   [Anonymous], 2011, PSYCHIAT CLIN NEUROS
   Austin CJD, 2010, AMINO ACIDS, V39, P565, DOI 10.1007/s00726-010-0475-9
   Bach H, 2011, J NEUROCHEM, V118, P1067, DOI 10.1111/j.1471-4159.2011.07379.x
   Ball HJ, 2009, INT J BIOCHEM CELL B, V41, P467, DOI 10.1016/j.biocel.2008.01.005
   Ball HJ, 2007, GENE, V396, P203, DOI 10.1016/j.gene.2007.04.010
   Browne CA, 2012, NEUROPHARMACOLOGY, V62, P1903, DOI 10.1016/j.neuropharm.2011.12.009
   Browne CA, 2011, NEUROPHARMACOLOGY, V60, P683, DOI 10.1016/j.neuropharm.2010.11.020
   Capuron L, 2004, BIOL PSYCHIAT, V56, P819, DOI 10.1016/j.biopsych.2004.02.009
   Capuron L, 2004, BRAIN BEHAV IMMUN, V18, P205, DOI 10.1016/j.bbi.2003.11.004
   Capuron L, 2002, MOL PSYCHIATR, V7, P468, DOI 10.1038/sj.mp.4000995
   Capuron L, 2002, NEUROPSYCHOPHARMACOL, V26, P643, DOI 10.1016/S0893-133X(01)00407-9
   Capuron L, 2001, PSYCHONEUROENDOCRINO, V26, P797, DOI 10.1016/S0306-4530(01)00030-0
   CARROLL BJ, 1980, LANCET, V2, P1249
   Chamas F, 2002, J NEUROCHEM, V82, P645, DOI 10.1046/j.1471-4159.2002.00989.x
   Charney Dennis S, 2004, Sci STKE, V2004, pre5, DOI 10.1126/stke.2252004re5
   Clark JA, 2008, MOL PSYCHIATR, V13, P498, DOI 10.1038/sj.mp.4002041
   Clark JA, 2005, BIOL PSYCHIAT, V57, P943, DOI 10.1016/j.biopsych.2005.01.013
   Connor TJ, 2008, NEUROSCI LETT, V441, P29, DOI 10.1016/j.neulet.2008.06.007
   Connor TJ, 1997, PHARMACOL BIOCHEM BE, V58, P961, DOI 10.1016/S0091-3057(97)00028-2
   Connor TJ, 2005, J NEUROIMMUNOL, V159, P119, DOI 10.1016/j.jneuroim.2004.10.016
   Cryan JF, 2005, NAT REV DRUG DISCOV, V4, P775, DOI 10.1038/nrd1825
   Curtin NM, 2009, BRAIN BEHAV IMMUN, V23, P535, DOI 10.1016/j.bbi.2009.02.003
   DELRUE C, 1994, BRAIN BEHAV IMMUN, V8, P137, DOI 10.1006/brbi.1994.1013
   Dhabhar FS, 1997, BRAIN BEHAV IMMUN, V11, P286
   DINAN TG, 1994, BRIT J PSYCHIAT, V164, P365, DOI 10.1192/bjp.164.3.365
   DUNN AJ, 1992, J PHARMACOL EXP THER, V261, P964
   Fu X, 2010, J NEUROINFLAMM, V7, DOI 10.1186/1742-2094-7-43
   Fujigaki S, 2001, EUR J IMMUNOL, V31, P2313, DOI 10.1002/1521-4141(200108)31:8<2313::AID-IMMU2313>3.0.CO;2-S
   Gibb J, 2008, BRAIN BEHAV IMMUN, V22, P573, DOI 10.1016/j.bbi.2007.12.001
   Gibb J, 2011, BRAIN BEHAV IMMUN, V25, P468, DOI 10.1016/j.bbi.2010.11.008
   Gizatullin R, 2008, J AFFECT DISORDERS, V107, P175, DOI 10.1016/j.jad.2007.07.005
   Gutknecht L, 2009, EUR NEUROPSYCHOPHARM, V19, P266, DOI 10.1016/j.euroneuro.2008.12.005
   Hansen AM, 2000, REDOX REP, V5, P112, DOI 10.1179/135100000101535384
   Jacobson LH, 2007, BEHAV GENET, V37, P171, DOI 10.1007/s10519-006-9106-3
   Kanner AM, 2004, CLIN EEG NEUROSCI, V35, P46, DOI 10.1177/155005940403500111
   Kiank C, 2010, PLOS ONE, V5, DOI 10.1371/journal.pone.0011825
   Kirby LG, 2000, NEUROPSYCHOPHARMACOL, V22, P148, DOI 10.1016/S0893-133X(99)00093-7
   Kirby LG, 2008, J NEUROSCI, V28, P12927, DOI 10.1523/JNEUROSCI.2887-08.2008
   Lanfumey L, 2008, NEUROSCI BIOBEHAV R, V32, P1174, DOI 10.1016/j.neubiorev.2008.04.006
   Leonard BE, 2005, EUR PSYCHIAT, V20, pS302, DOI 10.1016/S0924-9338(05)80180-4
   Leonard BE, 2009, HUM PSYCHOPHARM CLIN, V24, P165, DOI 10.1002/hup.1011
   Lestage J, 2002, BRAIN BEHAV IMMUN, V16, P596, DOI 10.1016/S0889-1591(02)00014-4
   Lob S, 2009, CANCER IMMUNOL IMMUN, V58, P153, DOI 10.1007/s00262-008-0513-6
   Maes M, 1999, NEUROPSYCHOPHARMACOL, V20, P370, DOI 10.1016/S0893-133X(98)00088-8
   MAES M, 1995, PROG NEURO-PSYCHOPH, V19, P11, DOI 10.1016/0278-5846(94)00101-M
   Maes M, 2011, PROG NEURO-PSYCHOPH, V35, P702, DOI 10.1016/j.pnpbp.2010.12.017
   Malek ZS, 2005, EUR J NEUROSCI, V22, P895, DOI 10.1111/j.1460-9568.2005.04264.x
   McEwen BS, 1998, ANN NY ACAD SCI, V840, P33, DOI 10.1111/j.1749-6632.1998.tb09546.x
   McEwen BS, 2005, METABOLISM, V54, P20, DOI 10.1016/j.metabol.2005.01.008
   Metz R, 2007, CANCER RES, V67, P7082, DOI 10.1158/0008-5472.CAN-07-1872
   Miller AH, 2009, BIOL PSYCHIAT, V65, P732, DOI 10.1016/j.biopsych.2008.11.029
   Nolan Y, 2000, J NEURAL TRANSM, V107, P1393, DOI 10.1007/s007020070003
   O'Mahony CM, 2010, BEHAV BRAIN RES, V213, P148, DOI 10.1016/j.bbr.2010.04.038
   Pitossi F, 1997, J NEUROSCI RES, V48, P287, DOI 10.1002/(SICI)1097-4547(19970515)48:4<287::AID-JNR1>3.0.CO;2-7
   Raison CL, 2006, TRENDS IMMUNOL, V27, P24, DOI 10.1016/j.it.2005.11.006
   Raison CL, 2005, BRAIN BEHAV IMMUN, V19, P23, DOI 10.1016/j.bbi.2004.05.001
   Robinson CM, 2005, J INTERF CYTOK RES, V25, P20, DOI 10.1089/jir.2005.25.20
   Roche M, 2003, J NEUROSCI, V23, P970
   SAITO K, 1991, BRAIN RES, V546, P151, DOI 10.1016/0006-8993(91)91171-V
   Sapolsky RM, 2000, ARCH GEN PSYCHIAT, V57, P925, DOI 10.1001/archpsyc.57.10.925
   Shishkina GT, 2007, NEUROSCIENCE, V150, P404, DOI 10.1016/j.neuroscience.2007.09.017
   Siesser WB, 2010, NEUROSCI LETT, V481, P6, DOI 10.1016/j.neulet.2010.06.035
   Smith A.K., 2011, MOL PSYCHIAT
   Sugden K, 2009, BMC NEUROSCI, V10, DOI 10.1186/1471-2202-10-50
   Takikawa O, 1999, ADV EXP MED BIOL, V467, P553
   URADE Y, 1983, J BIOL CHEM, V258, P6621
   Walther DJ, 2003, BIOCHEM PHARMACOL, V66, P1673, DOI 10.1016/S0006-2952(03)00556-2
   Walther DJ, 2003, SCIENCE, V299, P76, DOI 10.1126/science.1078197
   Wang Y, 2009, BRAIN BEHAV IMMUN
   Zhang XD, 2004, SCIENCE, V305, P217, DOI 10.1126/science.1097540
   Zill P, 2007, J PSYCHIATR RES, V41, P168, DOI 10.1016/j.jpsychires.2005.05.004
NR 75
TC 25
Z9 27
U1 2
U2 22
PU PERGAMON-ELSEVIER SCIENCE LTD
PI OXFORD
PA THE BOULEVARD, LANGFORD LANE, KIDLINGTON, OXFORD OX5 1GB, ENGLAND
SN 0306-4522
J9 NEUROSCIENCE
JI Neuroscience
PD DEC 6
PY 2012
VL 225
BP 237
EP 248
DI 10.1016/j.neuroscience.2012.08.031
PG 12
WC Neurosciences
WE Science Citation Index Expanded (SCI-EXPANDED)
SC Neurosciences & Neurology
GA 039SP
UT WOS:000311267100020
PM 22917616
DA 2023-08-27
ER

PT J
AU Garcia-Zaragoza, E
   Hernandez, C
   Barrachina, MD
   Esplugues, JV
AF Garcia-Zaragoza, E
   Hernandez, C
   Barrachina, MD
   Esplugues, JV
TI Interleukin 1 beta-induced inhibition of gastric acid secretion involves
   glutamate, NO and cGMP synthesis in the brain
SO NAUNYN-SCHMIEDEBERGS ARCHIVES OF PHARMACOLOGY
LA English
DT Article
DE brain-gut axis; central nervous system; interleukin 1 beta; gastric acid
   secretion; glutamate; nitric oxide; soluble guanylyl cyclase; endotoxin
ID CENTRAL-NERVOUS-SYSTEM; NITRIC-OXIDE SYNTHASE; TUMOR-NECROSIS-FACTOR;
   RECEPTOR ANTAGONIST; MESSENGER-RNA; ENDOTOXIN; CYTOKINE; RATS;
   LIPOPOLYSACCHARIDE; EXPRESSION
AB This study examines the role of a central pathway involving glutamate receptors, nitric oxide (NO) and cGMP in the acute inhibitory effects of central interleukin 1beta on pentagastrin-stimulated acid production.
   The acid-inhibitory effect of central interleukin 1beta was prevented by intracisternal (i.c.) microinjections of interleukin 1beta together with the NMDA receptor antagonist, dizocilpine maleate (MK-801). Intracisternal co-administration of the nitric oxide synthase inhibitor, N-G-nitro-L-arginine methyl esther (L-NAME) or 1H-[1,2,4]oxazodiolo[4,3-a]quinoxalin-1-one (ODQ), a soluble guanylyl cyclase (sGC) blocker, both reversed the hyposecretory effect of central interleukin 1beta. Peripheral administration of endotoxin significantly reduced pentagastrin-stimulated gastric acid secretion. I.c. pre-treatment with the interleukin 1 receptor antagonist, IL-1ra, failed to restore acid secretory responses in these rats. In addition, endotoxin did not modify the levels of endogenous mRNA for IL-1beta in the brainstem.
   We conclude that central glutamate receptors are involved in the acid inhibitory effect of centrally administered interleukin 1beta. This central pathway involves synthesis of NO, which acts on the enzyme sGC. However, endogenous interleukin 1beta does not seem to be involved in the inhibition of gastric acid secretion elicited by peripheral endotoxin.
C1 Univ Valencia, Dept Pharmacol, Fac Med, Valencia 46010, Spain.
C3 University of Valencia
RP Barrachina, MD (通讯作者)，Univ Valencia, Dept Pharmacol, Fac Med, Avda Blasco Ibanez 15, Valencia 46010, Spain.
RI Esplugues, Juan/ABC-1488-2021; Barrachina, Maria/ABB-9683-2021; SAEZ,
   CARLOS HERNANDEZ/M-3029-2014
OI SAEZ, CARLOS HERNANDEZ/0000-0001-8536-429X; Barrachina, Maria
   Dolores/0000-0002-4332-2929; Esplugues Mota, Juan
   Vicente/0000-0001-8205-021X
CR AREND WP, 1990, J CLIN INVEST, V85, P1694, DOI 10.1172/JCI114622
   Beales ILP, 1998, GASTROENTEROLOGY, V114, P618, DOI 10.1016/S0016-5085(98)70558-X
   BREDT DS, 1989, P NATL ACAD SCI USA, V86, P9030, DOI 10.1073/pnas.86.22.9030
   DASCOMBE MJ, 1979, BRIT J PHARMACOL, V66, P565, DOI 10.1111/j.1476-5381.1979.tb13695.x
   DINARELLO CA, 1991, BLOOD, V77, P1627
   El-Omar EM, 2001, GUT, V48, P743, DOI 10.1136/gut.48.6.743
   Eriksson C, 2000, CYTOKINE, V12, P423, DOI 10.1006/cyto.1999.0582
   ESPLUGUES JV, 1993, BRIT J PHARMACOL, V108, P9, DOI 10.1111/j.1476-5381.1993.tb13431.x
   ESPLUGUES JV, 1994, J GASTROEN HEPATOL, V9, pS45, DOI 10.1111/j.1440-1746.1994.tb01301.x
   Esplugues JV, 1996, P NATL ACAD SCI USA, V93, P14839, DOI 10.1073/pnas.93.25.14839
   Garcia-Zaragoza E, 2000, BRIT J PHARMACOL, V130, P1283, DOI 10.1038/sj.bjp.0703436
   Konsman JP, 1999, NEUROSCIENCE, V89, P535, DOI 10.1016/S0306-4522(98)00368-6
   Konturek PC, 1999, ALIMENT PHARM THER, V13, P333
   LIEBERMAN AP, 1989, P NATL ACAD SCI USA, V86, P6348, DOI 10.1073/pnas.86.16.6348
   Mascarucci P, 1998, NEUROSCIENCE, V86, P1285, DOI 10.1016/S0306-4522(98)00105-5
   Oberholzer A, 2000, CRIT CARE MED, V28, pN3, DOI 10.1097/00003246-200004001-00002
   OPP MR, 1992, P NATL ACAD SCI USA, V89, P3726, DOI 10.1073/pnas.89.9.3726
   Quan N, 1999, J NEUROIMMUNOL, V93, P72, DOI 10.1016/S0165-5728(98)00193-3
   ROBERT A, 1991, LIFE SCI, V48, P123, DOI 10.1016/0024-3205(91)90405-Z
   ROWLEY D, 1956, LANCET, V1, P366
   SAPERAS E, 1992, PEPTIDES, V13, P221, DOI 10.1016/0196-9781(92)90100-H
   SAPERAS E, 1993, LIFE SCI, V52, P785, DOI 10.1016/0024-3205(93)90076-F
   SAPERAS E, 1994, DIGEST DIS SCI, V39, P152, DOI 10.1007/BF02090075
   SAPERAS ES, 1990, GASTROENTEROLOGY, V99, P1599, DOI 10.1016/0016-5085(90)90463-B
   Satta MA, 1998, NEUROENDOCRINOLOGY, V67, P109, DOI 10.1159/000054305
   Streit WJ, 1998, EXP NEUROL, V152, P74, DOI 10.1006/exnr.1998.6835
   SUTO G, 1994, GASTROENTEROLOGY, V106, P1568
   TACHE Y, 1993, METH NEUROSCI, V17, P169
   Turrin NP, 2001, BRAIN RES BULL, V54, P443, DOI 10.1016/S0361-9230(01)00445-2
   UEHARA A, 1990, BIOCHEM BIOPH RES CO, V173, P585, DOI 10.1016/S0006-291X(05)80075-0
   UEHARA A, 1989, BIOCHEM BIOPH RES CO, V162, P1578, DOI 10.1016/0006-291X(89)90855-3
   ULICH TR, 1990, AM J PATHOL, V137, P1173
   WALLACE JL, 1991, AM J PHYSIOL, V261, pG559, DOI 10.1152/ajpgi.1991.261.4.G559
   WALLACE JL, 1992, GASTROENTEROLOGY, V102, P1176
NR 34
TC 5
Z9 5
U1 0
U2 2
PU SPRINGER-VERLAG
PI NEW YORK
PA 175 FIFTH AVE, NEW YORK, NY 10010 USA
SN 0028-1298
J9 N-S ARCH PHARMACOL
JI Naunyn-Schmiedebergs Arch. Pharmacol.
PD JAN
PY 2003
VL 367
IS 1
BP 22
EP 27
DI 10.1007/s00210-002-0654-7
PG 6
WC Pharmacology & Pharmacy
WE Science Citation Index Expanded (SCI-EXPANDED)
SC Pharmacology & Pharmacy
GA 646BD
UT WOS:000181014200003
PM 12616337
DA 2023-08-27
ER

PT J
AU I'Allemand, D
   Biason-Lauber, A
AF I'Allemand, D
   Biason-Lauber, A
TI Intra-adrenal regulation of androgen synthesis
SO EUROPEAN JOURNAL OF CLINICAL INVESTIGATION
LA English
DT Article; Proceedings Paper
CT Conference on Adrenal 2000 - From Molecular Basics to New Clinical
   Dimensions
CY 2000
CL HAMBURG, GERMANY
DE steroid synthesis; 17/20-lyase regulation; growth factors; adrenal
   gland; adrenarche
ID HUMAN ADRENOCORTICAL-CELLS; ISOLATED 17,20-LYASE DEFICIENCY;
   MESSENGER-RIBONUCLEIC-ACID; POLYCYSTIC-OVARY-SYNDROME;
   TUMOR-NECROSIS-FACTOR; GROWTH-FACTORS; DEHYDROEPIANDROSTERONE-SULFATE;
   HUMAN P450C17; 3-BETA-HYDROXYSTEROID DEHYDROGENASE; STEROIDOGENIC ENZYME
AB The adrenal cortex encloses the neuroendocrine medulla and is itself subdivided into three distinct zones, each having a specific function and regulation. While the glomerulosa and the fasciculata control vital systems of mineral and energy supply, which are stringently regulated by higher control factors, the function of the reticularis is less clear, beyond supplying a pool of weak androgens, and consequently we do not understand its redundant regulation. The following questions need to be answered:
   1 How is the formation of dehydroepiandrosterone (DHEA) and androstenedione differently regulated from glucocorticoid synthesis in normally functioning adrenals?
   2 How might growth factors, which increase prepubertally [1], prime the adrenarche?
   3 The regulation of the 17/20-lyase enzyme activity is one of the key factors of adrenal androgen secretion (review [2]). How can the two activities of the P450c17 enzyme be differently regulated in the same cell in a developmentally dependent fashion? This review focuses on the intra-adrenal growth factor system and on the role of 17/20-lyase regulation, as well as on their possible interactions. The increase of activity of the 17/20-lyase enzymatic activity is necessary for the rise of C19 steroids [3], while the relative increase of formation of DHEA is only possible in the presence of a low 3 beta -hydroxysteroid-dehydrogenase (3 beta HSD) activity [4].
C1 Humboldt Univ, Dept Paediat, Berlin, Germany.
   Univ Zurich, CH-8006 Zurich, Switzerland.
C3 Humboldt University of Berlin; University of Zurich
RP I'Allemand, D (通讯作者)，Humboldt Univ, Dept Paediat, Berlin, Germany.
CR Auchus RJ, 1998, J BIOL CHEM, V273, P3158, DOI 10.1074/jbc.273.6.3158
   AZZIZ R, 1991, J CLIN ENDOCR METAB, V72, P1025, DOI 10.1210/jcem-72-5-1025
   Balducci R, 1998, HORM RES, V49, P210, DOI 10.1159/000023173
   BiasonLauber A, 1997, J CLIN ENDOCR METAB, V82, P3807, DOI 10.1210/jc.82.11.3807
   Chambaz EM, 1996, HORM RES, V45, P222
   Clarke D, 1996, J ENDOCRINOL, V151, P301, DOI 10.1677/joe.0.1510301
   Conley AJ, 1997, BIOL REPROD, V56, P789, DOI 10.1095/biolreprod56.4.789
   Dunaif A, 1997, ENDOCR REV, V18, P774, DOI 10.1210/er.18.6.774
   Endoh A, 1996, J CLIN ENDOCR METAB, V81, P3558, DOI 10.1210/jc.81.10.3558
   Fearon U, 1998, EUR J ENDOCRINOL, V138, P567, DOI 10.1530/eje.0.1380567
   Feige JJ, 1998, HORM METAB RES, V30, P421, DOI 10.1055/s-2007-978908
   Geller DH, 1997, NAT GENET, V17, P201, DOI 10.1038/ng1097-201
   GonzalezHernandez JA, 1996, J CLIN ENDOCR METAB, V81, P807, DOI 10.1210/jc.81.2.807
   Haidan A, 1998, ENDOCRINOLOGY, V139, P772, DOI 10.1210/en.139.2.772
   Hall PF, 1995, J STEROID BIOCHEM, V55, P601, DOI 10.1016/0960-0760(95)00211-1
   HIGUCHI K, 1984, J CLIN ENDOCR METAB, V59, P714, DOI 10.1210/jcem-59-4-714
   HORNSBY PJ, 1987, J STEROID BIOCHEM, V27, P1161, DOI 10.1016/0022-4731(87)90203-2
   JAATTELA M, 1991, ENDOCRINOLOGY, V128, P623
   JUUL A, 1994, J CLIN ENDOCR METAB, V78, P744, DOI 10.1210/jc.78.3.744
   KATAGIRI M, 1995, ARCH BIOCHEM BIOPHYS, V317, P343, DOI 10.1006/abbi.1995.1173
   Kristiansen SB, 1997, STEROIDS, V62, P258, DOI 10.1016/S0039-128X(96)00223-1
   l'Allemand D, 1998, MOL CELL ENDOCRINOL, V140, P169, DOI 10.1016/S0303-7207(98)00046-X
   LAllemand D, 1996, J CLIN ENDOCR METAB, V81, P3892, DOI 10.1210/jc.81.11.3892
   LEBRETHON MC, 1994, J CLIN ENDOCR METAB, V79, P1033, DOI 10.1210/jc.79.4.1033
   MESIANO S, 1993, J CLIN ENDOCR METAB, V77, P754, DOI 10.1210/jc.77.3.754
   Mesiano S, 1997, J CLIN ENDOCR METAB, V82, P1390, DOI 10.1210/jc.82.5.1390
   Mesiano S, 1997, STEROIDS, V62, P62, DOI 10.1016/S0039-128X(96)00161-4
   Miller WL, 1997, STEROIDS, V62, P133, DOI 10.1016/S0039-128X(96)00172-9
   MUNARISILEM Y, 1999, CLIN INVESTIGATOR, V95, P1429
   Penhoat A, 1996, STEROIDS, V61, P176, DOI 10.1016/0039-128X(96)00009-8
   PENHOAT A, 1991, J MOL ENDOCRINOL, V7, P234
   SCHIEBINGER RJ, 1981, J CLIN INVEST, V67, P1177, DOI 10.1172/JCI110132
   Smith R, 1998, J CLIN ENDOCR METAB, V83, P2916, DOI 10.1210/jc.83.8.2916
   Voutilainen R, 1998, HORM METAB RES, V30, P432, DOI 10.1055/s-2007-978910
   Weber A, 1997, CLIN ENDOCRINOL, V46, P431, DOI 10.1046/j.1365-2265.1997.1580969.x
   YOUNG J, 1994, J CLIN ENDOCR METAB, V78, P299, DOI 10.1210/jc.78.2.299
   ZHANG LH, 1995, P NATL ACAD SCI USA, V92, P10619, DOI 10.1073/pnas.92.23.10619
NR 37
TC 8
Z9 8
U1 0
U2 2
PU BLACKWELL SCIENCE LTD
PI OXFORD
PA P O BOX 88, OSNEY MEAD, OXFORD OX2 0NE, OXON, ENGLAND
SN 0014-2972
J9 EUR J CLIN INVEST
JI Eur. J. Clin. Invest.
PD DEC
PY 2000
VL 30
SU 3
BP 28
EP 33
DI 10.1046/j.1365-2362.2000.0300s3028.x
PG 6
WC Medicine, General & Internal; Medicine, Research & Experimental
WE Conference Proceedings Citation Index - Science (CPCI-S); Science Citation Index Expanded (SCI-EXPANDED)
SC General & Internal Medicine; Research & Experimental Medicine
GA 405RD
UT WOS:000167172200007
PM 11281364
DA 2023-08-27
ER

PT J
AU Redmond, KJ
   Hildreth, M
   Sair, HI
   Terezakis, S
   McNutt, T
   Kleinberg, L
   Cohen, KJ
   Wharam, M
   Horska, A
   Mahone, EM
AF Redmond, Kristin J.
   Hildreth, Meghan
   Sair, Haris I.
   Terezakis, Stephanie
   McNutt, Todd
   Kleinberg, Lawrence
   Cohen, Kenneth J.
   Wharam, Moody
   Horska, Alena
   Mahone, E. Mark
TI Association of Neuronal Injury in the Genu and Body of Corpus Callosum
   After Cranial Irradiation in Children With Impaired Cognitive Control: A
   Prospective Study
SO INTERNATIONAL JOURNAL OF RADIATION ONCOLOGY BIOLOGY PHYSICS
LA English
DT Article; Proceedings Paper
CT 19th Annual Scientific Meeting of the Society-for-Neuro-Oncology (SNO)
CY NOV 13-16, 2014
CL Miami, FL
SP Soc Neuro Oncol
ID WHOLE-BRAIN IRRADIATION; WHITE-MATTER INTEGRITY; MULTIPLE-SCLEROSIS;
   RADIATION-THERAPY; NASOPHARYNGEAL CARCINOMA; PROCESSING SPEED; TUMOR
   SURVIVORS; POSTERIOR-FOSSA; WORKING-MEMORY; RADIOTHERAPY
AB Purpose: Brain radiation is associated with functional deficits in children. The purpose of this study was to examine white matter integrity as measured by diffusion tensor imaging and associations with region-specific radiation dose and neuropsychological functioning in children treated with cranial irradiation.
   Methods and Materials: A total of 20 patients and 55 age-and sex-matched controls were included in the present study. Diffusion tensor imaging and neuropsychological assessments were conducted at baseline and 6, 15, and 27 months after treatment. The neuropsychological assessment included motor dexterity, working memory, and processing speed. White matter regions were contoured, and the apparent diffusion coefficient (ADC) and fractional anisotropy (FA) were recorded for each participant. Linear mixed effects regression models were used to prospectively compare the associations among ADC, FA, radiation dose to contoured structures, and performance on the neuropsychological assessments over time.
   Results: The mean prescription dose was 44 Gy (range 12-54). Across visits, compared with the controls, the patients showed a significantly increased ADC across all selected regions and alterations in FA in the dorsal midbrain and corpus callosum (genu, splenium, body). An increased radiation dose to the genu and body of the corpus callosum was associated with alterations in ADC and FA and reduced neuropsychological performance, most notably motor speed and processing.
   Conclusions: These prospective data suggest that subcortical white matter, especially the genu and body of the corpus callosum, could be regions with increased susceptibility to radiation-induced injury, with implications for cognitive function. (C) 2018 Elsevier Inc. All rights reserved.
C1 [Redmond, Kristin J.; Hildreth, Meghan; Terezakis, Stephanie; McNutt, Todd; Kleinberg, Lawrence; Wharam, Moody] Johns Hopkins Univ, Dept Radiat Oncol & Mol Radiat Sci, 401 North Broadway,Suite 1440, Baltimore, MD 21231 USA.
   [Sair, Haris I.; Horska, Alena] Johns Hopkins Univ, Russell H Morgan Dept Radiol & Radiol Sci, Baltimore, MD USA.
   [Cohen, Kenneth J.] Sidney Kimmel Comprehens Canc Ctr, Div Pediat Oncol, Baltimore, MD USA.
   [Mahone, E. Mark] Kennedy Krieger Inst, Dept Neuropsychol, Baltimore, MD USA.
C3 Johns Hopkins University; Johns Hopkins University; Johns Hopkins
   University; Johns Hopkins Medicine; Kennedy Krieger Institute
RP Redmond, KJ (通讯作者)，Johns Hopkins Univ, Dept Radiat Oncol & Mol Radiat Sci, 401 North Broadway,Suite 1440, Baltimore, MD 21231 USA.
EM
OI Mahone, E. Mark/0000-0002-5022-1499
FU National Institutes of Health [R01 NS042851]; Kennedy Krieger
   Institute/Johns Hopkins University Intellectual and Developmental
   Disabilities Research Center [U54 HD-079123, U54 HD-09123]; Johns
   Hopkins University School of Medicine; National Institutes of Health,
   National Center for Research Resources, Clinical and Translational
   Sciences Awards Program [UL1RR025005]; Institute for Clinical and
   Translational Research
FX The present study was supported by the National Institutes of Health
   (grant R01 NS042851); the Kennedy Krieger Institute/Johns Hopkins
   University Intellectual and Developmental Disabilities Research Center
   (grants U54 HD-079123 and U54 HD-09123); the Institute for Clinical and
   Translational Research, the Johns Hopkins University School of Medicine;
   and the National Institutes of Health, National Center for Research
   Resources, Clinical and Translational Sciences Awards Program (grant
   UL1RR025005).
CR Agbahiwe H, 2017, CANCER-AM CANCER SOC, V123, P161, DOI 10.1002/cncr.30313
   Agosta F, 2014, HUM BRAIN MAPP, V35, P1921, DOI 10.1002/hbm.22302
   Alexander AL, 2007, NEUROTHERAPEUTICS, V4, P316, DOI 10.1016/j.nurt.2007.05.011
   [Anonymous], 2001, WOODCOCK JOHNSON TES
   [Anonymous], 1997, DIAGNOSTIC INTERVIEW
   ASAI A, 1989, CANCER-AM CANCER SOC, V63, P1962, DOI 10.1002/1097-0142(19890515)63:10<1962::AID-CNCR2820631016>3.0.CO;2-V
   Basser PJ, 2000, MAGNET RESON MED, V44, P625, DOI 10.1002/1522-2594(200010)44:4<625::AID-MRM17>3.0.CO;2-O
   Beery KE, 1968, BEERY BUKTENICA DEV
   Bester M, 2013, J NEUROL SCI, V330, P61, DOI 10.1016/j.jns.2013.04.005
   Bonekamp D, 2007, NEUROIMAGE, V34, P733, DOI 10.1016/j.neuroimage.2006.09.020
   Brinkman TM, 2016, J CLIN ONCOL, V34, P1358, DOI 10.1200/JCO.2015.62.2589
   Caligiuri ME, 2015, NEUROIMAGE-CLIN, V7, P28, DOI 10.1016/j.nicl.2014.11.008
   Chen WS, 2015, TUMORI, V101, P487, DOI 10.5301/tj.5000348
   Connor M, 2017, RADIOTHER ONCOL, V123, P209, DOI 10.1016/j.radonc.2017.04.006
   Conturo TE, 1999, P NATL ACAD SCI USA, V96, P10422, DOI 10.1073/pnas.96.18.10422
   Correa DG, 2015, NEURORADIOLOGY, V57, P475, DOI 10.1007/s00234-015-1489-5
   Ellenberg L, 2009, NEUROPSYCHOLOGY, V23, P705, DOI 10.1037/a0016674
   Fling BW, 2011, NEUROPSYCHOLOGIA, V49, P2564, DOI 10.1016/j.neuropsychologia.2011.05.004
   Gazdzinski LM, 2012, INT J RADIAT ONCOL, V84, pE631, DOI 10.1016/j.ijrobp.2012.06.053
   Gondi V, 2014, J CLIN ONCOL, V32, P3810, DOI 10.1200/JCO.2014.57.2909
   Gondi V, 2013, INT J RADIAT ONCOL, V85, P348, DOI 10.1016/j.ijrobp.2012.11.031
   Hofer S, 2006, NEUROIMAGE, V32, P989, DOI 10.1016/j.neuroimage.2006.05.044
   HOPPEHIRSCH E, 1995, CHILD NERV SYST, V11, P340, DOI 10.1007/BF00301666
   Johannesen TB, 2003, RADIOTHER ONCOL, V69, P169, DOI 10.1016/S0167-8140(03)00192-0
   Leng X, 2017, ONCOL LETT, V14, P6091, DOI 10.3892/ol.2017.6968
   Ma TM, 2017, RADIOTHER ONCOL, V125, P234, DOI 10.1016/j.radonc.2017.09.035
   McKenna BS, 2015, J INT NEUROPSYCH SOC, V21, P330, DOI 10.1017/S1355617715000314
   Mori S, 1999, ANN NEUROL, V45, P265, DOI 10.1002/1531-8249(199902)45:2<265::AID-ANA21>3.0.CO;2-3
   Nieman BJ, 2015, INT J RADIAT ONCOL, V93, P882, DOI 10.1016/j.ijrobp.2015.07.2293
   Penitzka S, 2002, STRAHLENTHER ONKOL, V178, P252, DOI 10.1007/s00066-002-0878-y
   Pierpaoli C, 1996, RADIOLOGY, V201, P637, DOI 10.1148/radiology.201.3.8939209
   POZZILLI C, 1991, CORTEX, V27, P441, DOI 10.1016/S0010-9452(13)80039-1
   Prust MJ, 2015, NEUROLOGY, V85, P683, DOI 10.1212/WNL.0000000000001861
   Rashid A, 2017, CHILD NERV SYST, V33, P965, DOI 10.1007/s00381-017-3412-3
   Reddick WE, 2000, MAGN RESON IMAGING, V18, P787, DOI 10.1016/S0730-725X(00)00182-X
   REDDON JR, 1988, PERCEPT MOTOR SKILL, V66, P503, DOI 10.2466/pms.1988.66.2.503
   Redmond KJ, 2017, INT J RADIAT ONCOL, V98, P603, DOI 10.1016/j.ijrobp.2017.03.009
   Redmond KJ, 2013, NEURO-ONCOLOGY, V15, P360, DOI 10.1093/neuonc/nos303
   Ris MD, 2001, J CLIN ONCOL, V19, P3470, DOI 10.1200/JCO.2001.19.15.3470
   Rueckriegel SM, 2015, PEDIATR BLOOD CANCER, V62, P1252, DOI 10.1002/pbc.25485
   Schatz J, 2000, NEUROPSYCHOLOGY, V14, P189, DOI 10.1037//0894-4105.14.2.189
   SILBER JH, 1992, J CLIN ONCOL, V10, P1390, DOI 10.1200/JCO.1992.10.9.1390
   Sorg SF, 2016, J HEAD TRAUMA REHAB, V31, P297, DOI 10.1097/HTR.0000000000000189
   Tannazi F., 2009, ISMRM 17 ANN SCI M E
   Thorndike R. L, 1986, TECHNICAL MANUAL STA
   Thust SC, 2014, J MED IMAG RADIAT ON, V58, P683, DOI 10.1111/1754-9485.12232
   Tuladhar AM, 2015, NEUROIMAGE-CLIN, V7, P518, DOI 10.1016/j.nicl.2015.02.003
   Van Hecke W, 2010, J MAGN RESON IMAGING, V31, P1492, DOI 10.1002/jmri.22198
   Welton T, 2015, RADIOLOGY, V275, P89, DOI 10.1148/radiol.14140925
   Wu Y, 2006, AM J NEURORADIOL, V27, P656
   Yaldizli O, 2014, MULT SCLER J, V20, P356, DOI 10.1177/1352458513496880
NR 51
TC 23
Z9 23
U1 0
U2 8
PU ELSEVIER SCIENCE INC
PI NEW YORK
PA 360 PARK AVE SOUTH, NEW YORK, NY 10010-1710 USA
SN 0360-3016
EI 1879-355X
J9 INT J RADIAT ONCOL
JI Int. J. Radiat. Oncol. Biol. Phys.
PD AUG 1
PY 2018
VL 101
IS 5
BP 1234
EP 1242
DI 10.1016/j.ijrobp.2018.04.037
PG 9
WC Oncology; Radiology, Nuclear Medicine & Medical Imaging
WE Science Citation Index Expanded (SCI-EXPANDED); Social Science Citation Index (SSCI); Conference Proceedings Citation Index - Science (CPCI-S)
SC Oncology; Radiology, Nuclear Medicine & Medical Imaging
GA GM7RN
UT WOS:000438391600033
PM 29908790
OA Green Accepted
DA 2023-08-27
ER

PT J
AU Gregor, KM
   Knebel, A
   Haverkamp, AK
   Baumgartner, W
   Volk, H
AF Gregor, Katharina M.
   Knebel, Anna
   Haverkamp, Ann-Kathrin
   Baumgaertner, Wolfgang
   Volk, Holger
TI Metastatic Canine Phaeochromocytoma with Unusual Manifestation
SO JOURNAL OF COMPARATIVE PATHOLOGY
LA English
DT Article
DE adrenal gland; dog; phaeochromocytoma; spinal column
ID RESPONSIVE MENINGITIS-ARTERITIS; ADRENAL-GLAND TUMORS;
   IMMUNOHISTOCHEMICAL CHARACTERIZATION; DOGS; DIAGNOSIS; DISEASE;
   DISCOSPONDYLITIS; METANEPHRINES; PAIN; NECK
AB We report the clinical, pathological and immunohistochemical characteristics of a phaeochromocytoma (PCC) in a 9-year-old male neutered Golden Retriever dog. The dog presented with acute onset of deteriorating cervical pain but was otherwise normal on general physical and neurological examinations. Magnetic resonance imaging of the cervical spine revealed a focal, extramedullary, infiltrative, poorly demarcated, heterogeneous mass with moderate contrast enhancement at the left cranial articular process of C2, associated with osteolysis and pathological fractures of C2, and marked soft tissue trauma. Due to the severe lesions and grave prognosis, the dog was euthanized. Post-mortem examination revealed severe enlargement of the right adrenal gland due to a neoplasm of the adrenal medulla. C2 was lytic and there was a white, well-demarcated, firm neoplastic mass in the surrounding musculature. Neoplasms were also present in, and adjacent to, the prostate gland and in pulmonary lymph nodes. Histologically, the neoplasms were composed of dense sheets and nests of small, round to polyhedral cells with frequent palisading along fine connective tissue septa, karyomegaly, multinucleated cells and frequent mitotic figures. Immunohistochemically, neoplastic cells expressed chromogranin A and synaptophysin. Gross, microscopic and immunohistochemical findings support the diagnosis of PCC, originating from the right adrenal gland, with multiple metastases in the cervical spine, prostate gland and pulmonary lymph nodes. This case highlights the difficulty of intra-vitam diagnosis of PCC as its manifestation can be highly variable. PCC should be considered as a rare but possible differential diagnosis for painful vertebral masses in elderly dogs. (C) 2022 Elsevier Ltd. All rights reserved.
C1 [Gregor, Katharina M.; Haverkamp, Ann-Kathrin; Baumgaertner, Wolfgang] Univ Vet Med Fdn Hannover, Dept Pathol, Hannover, Germany.
   [Knebel, Anna; Volk, Holger] Univ Vet Med Fdn Hannover, Small Anim Clin, Hannover, Germany.
   [Gregor, Katharina M.; Baumgaertner, Wolfgang] Ctr Syst Neurosci Hannover ZSN, Hannover, Germany.
C3 University of Veterinary Medicine Hannover; University of Veterinary
   Medicine Hannover
RP Baumgartner, W (通讯作者)，Univ Vet Med Fdn Hannover, Dept Pathol, Hannover, Germany.; Baumgartner, W (通讯作者)，Ctr Syst Neurosci Hannover ZSN, Hannover, Germany.
EM
CR Arenas-Gamboa AM, 2014, J COMP PATHOL, V150, P361, DOI 10.1016/j.jcpa.2013.11.211
   Barrera JS, 2013, JAVMA-J AM VET MED A, V242, P1715, DOI 10.2460/javma.242.12.1715
   Barthez PY, 1997, J VET INTERN MED, V11, P272, DOI 10.1111/j.1939-1676.1997.tb00464.x
   BERZON JL, 1981, VET MED SM ANIM CLIN, V76, P675
   Boes K, 2009, VET CLIN PATH, V38, P511, DOI 10.1111/j.1939-165X.2009.00117.x
   Carrera I, 2011, VET RADIOL ULTRASOUN, V52, P125, DOI 10.1111/j.1740-8261.2010.01756.x
   Cook AK, 2014, JAVMA-J AM VET MED A, V244, P1181, DOI 10.2460/javma.244.10.1181
   Cornelis I, 2017, J SMALL ANIM PRACT, V58, P174, DOI 10.1111/jsap.12622
   da Costa RMG, 2011, VET PATHOL, V48, P427, DOI 10.1177/0300985810381909
   De Strobel F, 2019, J SMALL ANIM PRACT, V60, P404, DOI 10.1111/jsap.12987
   DELASMULAS JM, 1995, RES VET SCI, V59, P124, DOI 10.1016/0034-5288(95)90045-4
   DeLellis RA, 2003, ACTA HISTOCHEM CYTOC, V36, P293, DOI 10.1267/ahc.36.293
   Eisenhofer G, 2004, PHARMACOL REV, V56, P331, DOI 10.1124/pr.56.3.1
   Farrugia Frederick-Anthony, 2019, Endocr Regul, V53, P191, DOI 10.2478/enr-2019-0020
   Fenn J, 2020, FRONT VET SCI, V7, DOI 10.3389/fvets.2020.579025
   Galac S, 2017, VET COMP ONCOL, V15, P1158, DOI 10.1111/vco.12291
   GILSON SD, 1994, J VET INTERN MED, V8, P228, DOI 10.1111/j.1939-1676.1994.tb03222.x
   Gostelow R, 2013, J VET INTERN MED, V27, P83, DOI 10.1111/jvim.12009
   Green BA, 2013, VET CLIN PATH, V42, P499, DOI 10.1111/vcp.12093
   Griffin JF, 2008, J SMALL ANIM PRACT, V49, P509, DOI 10.1111/j.1748-5827.2008.00588.x
   Head LL, 2004, VET RADIOL ULTRASOUN, V45, P574, DOI 10.1111/j.1740-8261.2004.04099.x
   Hermeyer K, 2007, VET DERMATOL, V18, P360, DOI 10.1111/j.1365-3164.2007.00612.x
   Holland CT, 2000, AUST VET J, V78, P676, DOI 10.1111/j.1751-0813.2000.tb10402.x
   Lai CL, 2008, PROSTATE, V68, P477, DOI 10.1002/pros.20720
   Lamagna B, 2006, J AM ANIM HOSP ASSOC, V42, P71, DOI 10.5326/0420071
   Lavely JA, 2006, VET SURG, V35, P176, DOI 10.1111/j.1532-950X.2006.00129.x
   Lowrie M, 2009, J VET INTERN MED, V23, P862, DOI 10.1111/j.1939-1676.2009.0337.x
   Maher ER, 1997, VET CLIN N AM-SMALL, V27, P359, DOI 10.1016/S0195-5616(97)50037-4
   Platt SR, 1998, J AM ANIM HOSP ASSOC, V34, P365, DOI 10.5326/15473317-34-5-365
   Rosol TJ, 2020, TUMORS DOMESTIC ANIM, P787
   Rusbridge C, 2020, IN PRACTICE, V42, DOI 10.1136/inp.m1869
   Sako T, 2003, VET PATHOL, V40, P212, DOI 10.1354/vp.40-2-212
   Salesov E, 2015, J VET INTERN MED, V29, P597, DOI 10.1111/jvim.12569
   Santamarina G, 2003, J VET INTERN MED, V17, P917, DOI 10.1892/0891-6640(2003)017&lt;0917:ATARHA&gt;2.3.CO;2
   Schwartz P, 2008, JAVMA-J AM VET MED A, V232, P77, DOI 10.2460/javma.232.1.77
   Spall B, 2011, VET RADIOL ULTRASOUN, V52, P534, DOI 10.1111/j.1740-8261.2011.01828.x
   Thangapandiyan M., 2014, International Journal of Veterinary Science, V3, P189
   Trevail R, 2014, IN PRACTICE, V36, DOI 10.1136/inp.g4953
   VANBREE H, 1981, J SMALL ANIM PRACT, V22, P59, DOI 10.1111/j.1748-5827.1981.tb01393.x
   Wagner A, 2018, VET IMMUNOL IMMUNOP, V197, P49, DOI 10.1016/j.vetimm.2018.01.011
   Webb AA, 2003, VET J, V165, P193, DOI 10.1016/S1090-0233(02)00249-6
   Webb AA, 2002, J VET INTERN MED, V16, P269, DOI 10.1892/0891-6640(2002)016&lt;0269:SMIDWN&gt;2.3.CO;2
   WHITE RAS, 1977, J SMALL ANIM PRACT, V18, P579, DOI 10.1111/j.1748-5827.1977.tb05933.x
   Wyatt S, 2019, VET RADIOL ULTRASOUN, V60, P409, DOI 10.1111/vru.12759
NR 44
TC 0
Z9 0
U1 0
U2 4
PU ELSEVIER SCI LTD
PI OXFORD
PA THE BOULEVARD, LANGFORD LANE, KIDLINGTON, OXFORD OX5 1GB, OXON, ENGLAND
SN 0021-9975
EI 1532-3129
J9 J COMP PATHOL
JI J. Comp. Pathol.
PD APR
PY 2022
VL 192
BP 33
EP 40
DI 10.1016/j.jcpa.2022.01.006
EA FEB 2022
PG 8
WC Pathology; Veterinary Sciences
WE Science Citation Index Expanded (SCI-EXPANDED)
SC Pathology; Veterinary Sciences
GA 0Z1XO
UT WOS:000790872500005
PM 35305712
DA 2023-08-27
ER

PT J
AU Domanskyi, A
   Geissler, C
   Vinnikov, IA
   Alter, H
   Schober, A
   Vogt, MA
   Gass, P
   Parlato, R
   Schutz, G
AF Domanskyi, Andrii
   Geissler, Christin
   Vinnikov, Ilya A.
   Alter, Heike
   Schober, Andreas
   Vogt, Miriam A.
   Gass, Peter
   Parlato, Rosanna
   Schuetz, Guenther
TI Pten ablation in adult dopaminergic neurons is neuroprotective in
   Parkinson's disease models
SO FASEB JOURNAL
LA English
DT Article
DE neurodegeneration; mTOR; mouse
ID TUMOR-SUPPRESSOR GENE; NIGROSTRIATAL SYSTEM; NUCLEOLAR DISRUPTION;
   MAMMALIAN TARGET; CRE RECOMBINASE; MOTOR DEFICITS; ANIMAL-MODELS;
   CELL-DEATH; MICE; BRAIN
AB Parkinson's disease (PD) is a progressive age-related movement disorder that results primarily from the selective loss of midbrain dopaminergic (DA) neurons. Symptoms of PD can be induced by genetic mutations or by DA neuron-specific toxins. A specific ablation of an essential factor controlling ribosomal RNA transcription, TifIa, in adult mouse DA neurons represses mTOR signaling and leads to progressive neurodegeneration and PD-like phenotype. Using an inducible Cre system in adult mice, we show here that the specific ablation of Pten in adult mouse DA neurons leads to activation of mTOR pathway and is neuroprotective in genetic (TifIa deletion) and neurotoxin-induced (MPTP or 6OHDA) mouse models of PD. Adult mice with DA neuron-specific Pten deletion exhibit elevated expression of tyrosine hydroxylase, a rate-limiting enzyme in the dopamine biosynthesis pathway, associated with increased striatal dopamine content, and increased mRNA levels of Foxa2, Pitx3, En1, Nurr1, and Lmx1b-the essential factors for maintaining physiological functions of adult DA neurons. Pten deletion attenuates the loss of tyrosine hydroxylase-positive cells after 6OHDA treatment, restores striatal dopamine in TifIa-knockout and MPTP-treated mice, and rescues locomotor impairments caused by TifIa loss. Inhibition of Pten-dependent functions in adult DA neurons may represent a promising PD therapy.-Domanskyi, A., Geissler, C., Vinnikov, I. A., Alter, H., Schober, A., Vogt, M. A., Gass, P., Parlato, R., Schutz, G. Pten ablation in adult dopaminergic neurons is neuroprotective in Parkinson's disease models. FASEB J. 25, 2898-2910 (2011). www.fasebj.org
C1 [Domanskyi, Andrii; Geissler, Christin; Vinnikov, Ilya A.; Alter, Heike; Parlato, Rosanna; Schuetz, Guenther] German Canc Res Ctr, Div Mol Biol Cell 1, D-69120 Heidelberg, Germany.
   [Schober, Andreas] Univ Freiburg, Dept Mol Embryol, Inst Anat & Cell Biol 2, Freiburg, Germany.
   [Vogt, Miriam A.; Gass, Peter] Univ Heidelberg, Cent Inst Mental Hlth, RG Anim Models Psychiat, D-6800 Mannheim, Germany.
C3 Helmholtz Association; German Cancer Research Center (DKFZ); University
   of Freiburg; Central Institute of Mental Health; Ruprecht Karls
   University Heidelberg
RP Schutz, G (通讯作者)，German Canc Res Ctr, Div Mol Biol Cell 1, Neuenheimer Feld 280, D-69120 Heidelberg, Germany.
EM
RI Parlato, Rosanna/G-8768-2015; Parlato, Rosanna/AAH-4429-2020; Domanskyi,
   Andrii/H-3437-2012
OI Parlato, Rosanna/0000-0001-6682-9645; Domanskyi,
   Andrii/0000-0002-4755-5981; Vogt, Miriam Annika/0000-0002-3400-5067;
   Vinnikov, Ilya/0000-0003-0488-8484
FU Deutsche Forschungsgemeinschaft [SFB 488, SFB 636]; Fonds der Chemischen
   Industrie; European Union [LSHM-CT-2005-018652 (CRESCENDO)];
   Bundesministerium fur Bildung und Forschung [FZK 01GS08153, 01GS08142,
   0313074C (HepatoSys)]; Helmholtz Gemeinschaft Deutscher
   Forschungszentren, through Initiative CoReNe and Alliance HelMA;
   Deutsche Krebshilfe [108567]
FX The authors thank R. Hertel for HPLC-ED. This work was supported by the
   Deutsche Forschungsgemeinschaft, through Collaborative Research Centers
   SFB 488 and SFB 636; the Fonds der Chemischen Industrie; the European
   Union, through grant LSHM-CT-2005-018652 (CRESCENDO); the
   Bundesministerium fur Bildung und Forschung, through NGFNplus grants FZK
   01GS08153 and 01GS08142 and project 0313074C (HepatoSys); the Helmholtz
   Gemeinschaft Deutscher Forschungszentren, through Initiative CoReNe and
   Alliance HelMA; and the Deutsche Krebshilfe, through project 108567.
CR Abou-Sleiman PM, 2006, NAT REV NEUROSCI, V7, P207, DOI 10.1038/nrn1868
   Backman SA, 2001, NAT GENET, V29, P396, DOI 10.1038/ng782
   Backman CM, 2006, GENESIS, V44, P383, DOI 10.1002/dvg.20228
   Brooks SP, 2009, NAT REV NEUROSCI, V10, P519, DOI 10.1038/nrn2652
   Burke RE, 2004, CELL TISSUE RES, V318, P63, DOI 10.1007/s00441-004-0908-4
   Carter R J, 2001, Curr Protoc Neurosci, VChapter 8, DOI 10.1002/0471142301.ns0812s15
   Carter RJ, 1999, J NEUROSCI, V19, P3248
   Chang N, 2007, TRENDS NEUROSCI, V30, P581, DOI 10.1016/j.tins.2007.08.006
   Cheng HC, 2011, J NEUROSCI, V31, P2125, DOI 10.1523/JNEUROSCI.5519-10.2011
   Cherra SJ, 2008, FUTUR NEUROL, V3, P309, DOI 10.2217/14796708.3.3.309
   Cully M, 2006, NAT REV CANCER, V6, P184, DOI 10.1038/nrc1819
   Dawson TM, 2010, NEURON, V66, P646, DOI 10.1016/j.neuron.2010.04.034
   Diaz-Ruiz O, 2009, PLOS ONE, V4, DOI 10.1371/journal.pone.0007027
   Ekstrand MI, 2009, PARKINSONISM RELAT D, V15, pS185, DOI 10.1016/S1353-8020(09)70811-9
   Engblom D, 2008, NEURON, V59, P497, DOI 10.1016/j.neuron.2008.07.010
   Fahn S, 2003, ANN NY ACAD SCI, V991, P1, DOI 10.1111/j.1749-6632.2003.tb07458.x
   Ferri ALM, 2007, DEVELOPMENT, V134, P2761, DOI 10.1242/dev.000141
   Fraser MM, 2008, NEUROSCIENCE, V151, P476, DOI 10.1016/j.neuroscience.2007.10.048
   Geisler S, 2010, NAT CELL BIOL, V12, P119, DOI 10.1038/ncb2012
   Gelb DJ, 1999, ARCH NEUROL-CHICAGO, V56, P33, DOI 10.1001/archneur.56.1.33
   Groszer M, 2001, SCIENCE, V294, P2186, DOI 10.1126/science.1065518
   Indra AK, 1999, NUCLEIC ACIDS RES, V27, P4324, DOI 10.1093/nar/27.22.4324
   Inzelberg R, 2007, NEUROLOGY, V69, P1542, DOI 10.1212/01.wnl.0000277638.63767.b8
   Jacobs FMJ, 2009, DEVELOPMENT, V136, P531, DOI 10.1242/dev.029769
   Kadkhodaei B, 2009, J NEUROSCI, V29, P15923, DOI 10.1523/JNEUROSCI.3910-09.2009
   Kim RH, 2006, BRIT J CANCER, V94, P620, DOI 10.1038/sj.bjc.6602994
   Kim RH, 2005, CANCER CELL, V7, P263, DOI 10.1016/j.ccr.2005.02.010
   Kittappa R, 2007, PLOS BIOL, V5, P2875, DOI 10.1371/journal.pbio.0050325
   Klein C, 2007, CURR OPIN NEUROL, V20, P453, DOI 10.1097/WCO.0b013e3281e6692b
   Le WD, 2008, J NEUROL SCI, V273, P29, DOI 10.1016/j.jns.2008.06.007
   Lesche R, 2002, GENESIS, V32, P148, DOI 10.1002/gene.10036
   Levine B, 2005, J CLIN INVEST, V115, P2679, DOI 10.1172/JCI26390
   Lewek MD, 2010, GAIT POSTURE, V31, P256, DOI 10.1016/j.gaitpost.2009.10.013
   Lin W, 2009, DEV BIOL, V333, P386, DOI 10.1016/j.ydbio.2009.07.006
   Liu BS, 2010, STROKE, V41, P1018, DOI 10.1161/STROKEAHA.110.579011
   Malagelada C, 2006, J NEUROSCI, V26, P9996, DOI 10.1523/JNEUROSCI.3292-06.2006
   Matsuda N, 2010, J CELL BIOL, V189, P211, DOI 10.1083/jcb.200910140
   Monville C, 2006, J NEUROSCI METH, V158, P219, DOI 10.1016/j.jneumeth.2006.06.001
   Narendra DP, 2010, PLOS BIOL, V8, DOI 10.1371/journal.pbio.1000298
   OTTO D, 1990, J NEUROSCI, V10, P1912
   Parkitna Jan Rodriguez, 2009, V530, P325, DOI 10.1007/978-1-59745-471-1_17
   Parlato R, 2008, J NEUROSCI, V28, P12759, DOI 10.1523/JNEUROSCI.2439-08.2008
   Ravikumar B, 2004, NAT GENET, V36, P585, DOI 10.1038/ng1362
   Rieker C, 2011, J NEUROSCI, V31, P453, DOI 10.1523/JNEUROSCI.0590-10.2011
   Ries V, 2006, P NATL ACAD SCI USA, V103, P18757, DOI 10.1073/pnas.0606401103
   Schober A, 2004, CELL TISSUE RES, V318, P215, DOI 10.1007/s00441-004-0938-y
   Schober A, 2007, NEUROBIOL DIS, V25, P378, DOI 10.1016/j.nbd.2006.10.005
   Sgado P, 2006, P NATL ACAD SCI USA, V103, P15242, DOI 10.1073/pnas.0602116103
   Sha D, 2010, HUM MOL GENET, V19, P352, DOI 10.1093/hmg/ddp501
   Smidt MP, 2007, NAT REV NEUROSCI, V8, P21, DOI 10.1038/nrn2039
   Sonnier L, 2007, J NEUROSCI, V27, P1063, DOI 10.1523/JNEUROSCI.4583-06.2007
   Spilman P, 2010, PLOS ONE, V5, DOI 10.1371/journal.pone.0009979
   Tain LS, 2009, NAT NEUROSCI, V12, P1129, DOI 10.1038/nn.2372
   Terzioglu M, 2008, FEBS J, V275, P1384, DOI 10.1111/j.1742-4658.2008.06302.x
   Unoki M, 2001, ONCOGENE, V20, P4457, DOI 10.1038/sj.onc.1204608
   Van Diepen MT, 2008, DEV NEUROSCI-BASEL, V30, P59, DOI 10.1159/000109852
   Xu PY, 2002, NEUROLOGY, V58, P881, DOI 10.1212/WNL.58.6.881
   Yogev G, 2007, EXP BRAIN RES, V177, P336, DOI 10.1007/s00221-006-0676-3
   Yuan XJ, 2005, MOL CELL, V19, P77, DOI 10.1016/j.molcel.2005.05.023
   Zaborszky L, 2001, BEHAV GENET, V31, P47, DOI 10.1023/A:1010257808945
   Zhang C, 2005, MOL CELL BIOL, V25, P6899, DOI 10.1128/MCB.25.16.6899-6911.2005
   Zheng KN, 2003, ARCH NEUROL-CHICAGO, V60, P722, DOI 10.1001/archneur.60.5.722
   Zhu Y, 2006, APOPTOSIS, V11, P197, DOI 10.1007/s10495-006-3714-5
   Zhu Y, 2007, NEUROCHEM INT, V50, P507, DOI 10.1016/j.neuint.2006.10.010
NR 64
TC 93
Z9 97
U1 0
U2 16
PU FEDERATION AMER SOC EXP BIOL
PI BETHESDA
PA 9650 ROCKVILLE PIKE, BETHESDA, MD 20814-3998 USA
SN 0892-6638
J9 FASEB J
JI Faseb J.
PD SEP
PY 2011
VL 25
IS 9
BP 2898
EP 2910
DI 10.1096/fj.11-181958
PG 13
WC Biochemistry & Molecular Biology; Biology; Cell Biology
WE Science Citation Index Expanded (SCI-EXPANDED)
SC Biochemistry & Molecular Biology; Life Sciences & Biomedicine - Other
   Topics; Cell Biology
GA 814GD
UT WOS:000294435200006
PM 21593433
OA Bronze
DA 2023-08-27
ER

PT J
AU CHOILUNDBERG, DL
   BOHN, MC
AF CHOILUNDBERG, DL
   BOHN, MC
TI ONTOGENY AND DISTRIBUTION OF GLIAL-CELL LINE-DERIVED NEUROTROPHIC FACTOR
   (GDNF) MESSENGER-RNA IN RAT
SO DEVELOPMENTAL BRAIN RESEARCH
LA English
DT Article
DE DOPAMINERGIC NEURON; ASTROCYTE; BRAIN DEVELOPMENT; CEREBELLUM; GROWTH
   FACTOR
ID DOPAMINERGIC-NEURONS; GROWTH-FACTOR; CULTURES
AB Glial cell line-derived neurotrophic factor (GDNF) is a member of the transforming growth factor-beta family isolated from the rat glial tumor cell line, B49. In embryonic dopaminergic (DA) neurons in vitro, GDNF promotes survival, high-affinity dopamine uptake, and neurite outgrowth. We have used a semi-quantitative reverse transcriptase-polymerase chain reaction (RT-PCR) with primers specific to GDNF to study the developmental expression of GDNF mRNA in central nervous system (CNS) and peripheral organs of embryonic rat on gestational days E11.5, E13.5 and E18, neonatal rat on postnatal days P0 and P10, and adult rat. GDNF mRNA is expressed throughout the CNS, with highest levels in P0 spinal cord and in P0 and P10 striatum. Lower levels are present in the brainstem (including the ventral mesencephalon, which contains the DA neurons of the substantia nigra), cerebellum, diencephalon, and telencephalon, as well as in primary cultures of cerebellar granule cells prepared from P7 cerebellum and astrocytes prepared from P1 cortex. The cerebellum has an unusual temporal pattern of expression, high at birth and in the adult, but undetectable at P10. GDNF: mRNA is also expressed in many peripheral tissues at higher levels than in brain. These include embryonic limb bud, kidney and gut; neonatal kidney, gut, lung and testis; and adult lung, liver and ovary. In addition to the predicted RT-PCR product, we also observed a minor band which was shown to be identical to GDNF in the mature peptide sequence, but which has a 78 base pair deletion in the preproprotein sequence. This shortened form of GDNF mRNA has an essentially identical pattern of distribution as GDNF and may represent an alternatively spliced form. A homologous short form of GDNF mRNA was also amplified from human fetal astrocytes. These observations suggest that GDNF may be a target derived and/or locally acting neurotrophic factor for other classes of CNS neurons, as well as for DA neurons of the substantia nigra. Additionally, GDNF likely plays a role as a neurotrophic factor in the perpheral nervous system and/or serves as a growth factor for non-neuronal cells.
C1 UNIV ROCHESTER,SCH MED & DENT,DEPT NEUROBIOL & ANAT,ROCHESTER,NY 14642.
C3 University of Rochester
RI Choi-Lundberg, Derek L/J-7670-2014
OI Choi-Lundberg, Derek L/0000-0002-3594-8585
CR ALEXI T, 1993, NEUROSCIENCE, V55, P903, DOI 10.1016/0306-4522(93)90307-2
   ALTMAN J, 1969, J COMP NEUROL, V136, P269, DOI 10.1002/cne.901360303
   DAVIS LG, 1986, BASIC METHODS MOL BI, P130
   HUMPEL C, 1994, NEUROSCIENCE, V59, P791, DOI 10.1016/0306-4522(94)90284-4
   HYMAN C, 1994, J NEUROSCI, V14, P335
   HYNES MA, 1994, J NEUROSCI RES, V37, P144, DOI 10.1002/jnr.490370118
   KIKUCHI S, 1993, NEUROSCI LETT, V160, P9, DOI 10.1016/0304-3940(93)90904-Y
   KNUSEL B, 1990, J NEUROSCI, V10, P558
   LIN LFH, 1993, SCIENCE, V260, P1130, DOI 10.1126/science.8493557
   MCCARTHY KD, 1980, J CELL BIOL, V85, P890, DOI 10.1083/jcb.85.3.890
   PENG LA, 1991, DEV BRAIN RES, V63, P1, DOI 10.1016/0165-3806(91)90061-M
   SCHAAR DG, 1993, EXP NEUROL, V124, P368, DOI 10.1006/exnr.1993.1207
   SCHUBERT D, 1974, NATURE, V249, P224, DOI 10.1038/249224a0
   SPRINGER JE, 1994, EXP NEUROL, V127, P167, DOI 10.1006/exnr.1994.1091
   STROMBERG I, 1993, EXP NEUROL, V124, P401, DOI 10.1006/exnr.1993.1214
   VIELKIND U, 1990, J NEUROSCI RES, V27, P360, DOI 10.1002/jnr.490270315
NR 16
TC 220
Z9 236
U1 0
U2 2
PU ELSEVIER SCIENCE BV
PI AMSTERDAM
PA PO BOX 211, 1000 AE AMSTERDAM, NETHERLANDS
SN 0165-3806
J9 DEV BRAIN RES
JI Dev. Brain Res.
PD MAR 16
PY 1995
VL 85
IS 1
BP 80
EP 88
DI 10.1016/0165-3806(94)00197-8
PG 9
WC Developmental Biology; Neurosciences
WE Science Citation Index Expanded (SCI-EXPANDED)
SC Developmental Biology; Neurosciences & Neurology
GA QM545
UT WOS:A1995QM54500010
DA 2023-08-27
ER

PT J
AU Bhardwaj, RD
   Bernstein, M
AF Bhardwaj, RD
   Bernstein, M
TI Prospective feasibility study of outpatient stereotactic brain lesion
   biopsy
SO NEUROSURGERY
LA English
DT Article
DE complications; day surgery; outcome; stereotactic biopsy
ID EARLY DISCHARGE; SAFETY; CRANIOTOMY; EFFICACY; TUMOR; STAY
AB OBJECTIVE:To assess the safety and feasibility of performing computed tomography-guided stereotactic brain lesion biopsy as an outpatient day-surgery procedure.
   METHODS: In late 1996, a prospective trial of outpatient stereotactic biopsies was initiated. The protocol consists of preadmission education of the patient, computed tomography-guided biopsy with local anesthesia (using a Brown-Roberts-Wells or Cosman-Roberts-Wells frame), postoperative observation in the postanesthetic care unit for 2 hours ands in the day surgery unit for 2 hours, and then discharge home 4 hours after the procedure.
   RESULTS; Seventy-six patients constituted the intent-to-treat group, of whom two were not discharged on the same day (97.4% success rate). The two patients underwent inpatient admission because one required intravenous antibiotic treatment of a brain abscess and the other had a hard lesion in the brainstem that precluded biopsy needle penetration; admission for further investigation of the lesion was elected. Two patients experienced complications (2.6%), i.e., one small area of intraventricular hemorrhage that produced only a mild headache and one case of mild worsening of preexisting leg weakness, with negative computed tomographic results.
   CONCLUSION: Discharging patients home after 4 hours of observation after stereotactic biopsies seems to be a safe, well-tolerated practice. In this series, there was no major morbidity and no patient was disadvantaged by participating in this protocol. This approach would be expected to result in health care resource and cost savings, with a potential increase in patient satisfaction because of shorter hospital stays.
C1 Univ Toronto, Toronto Western Hosp, Univ Hlth Network, Div Neurosurg, Toronto, ON M5T 2S8, Canada.
   Univ Toronto, Div Neurosurg, Toronto, ON, Canada.
C3 University of Toronto; University Health Network Toronto; University of
   Toronto
RP Bernstein, M (通讯作者)，Univ Toronto, Toronto Western Hosp, Univ Hlth Network, Div Neurosurg, 399 Bathurst St,4W451, Toronto, ON M5T 2S8, Canada.
EM
CR APUZZO MLJ, 1987, NEUROSURGERY, V20, P930, DOI 10.1227/00006123-198706000-00019
   Bednar DA, 1999, J SPINAL DISORD, V12, P359
   BERNSTEIN M, 1994, J NEUROSURG, V81, P165, DOI 10.3171/jns.1994.81.2.0165
   Bernstein M, 2001, CAN J NEUROL SCI, V28, P120, DOI 10.1017/S0317167100052781
   Blanshard HJ, 2001, ANESTH ANALG, V92, P89
   Field P, 2001, J NEUROSURG, V94, P545, DOI 10.3171/jns.2001.94.4.0545
   Goldstein RE, 2000, ANN SURG, V231, P732, DOI 10.1097/00000658-200005000-00014
   Kaakaji W, 2001, J NEUROSURG, V94, P892, DOI 10.3171/jns.2001.94.6.0892
   Khajehdehi P, 1999, AM J KIDNEY DIS, V34, P92, DOI 10.1016/S0272-6386(99)70113-7
   Kondziolka D, 1998, NEUROL CLIN, V16, P35, DOI 10.1016/S0733-8619(05)70366-2
   Kulkarni AV, 1998, J NEUROSURG, V89, P31, DOI 10.3171/jns.1998.89.1.0031
   Lalakea ML, 1999, ARCH OTOLARYNGOL, V125, P749, DOI 10.1001/archotol.125.7.749
   Mezei G, 1999, ANN SURG, V230, P721, DOI 10.1097/00000658-199911000-00016
   Moreno P, 1998, AM J MED, V105, P176, DOI 10.1016/S0002-9343(98)00243-5
   Sawin PD, 1998, SURG NEUROL, V49, P640, DOI 10.1016/S0090-3019(97)00435-7
   TAYLOR WAS, 1995, J NEUROSURG, V82, P48, DOI 10.3171/jns.1995.82.1.0048
   White PF, 1997, AMBULATORY ANESTHESI, P3
NR 17
TC 32
Z9 35
U1 0
U2 0
PU OXFORD UNIV PRESS INC
PI CARY
PA JOURNALS DEPT, 2001 EVANS RD, CARY, NC 27513 USA
SN 0148-396X
EI 1524-4040
J9 NEUROSURGERY
JI Neurosurgery
PD AUG
PY 2002
VL 51
IS 2
BP 358
EP 361
DI 10.1097/00006123-200208000-00011
PG 4
WC Clinical Neurology; Surgery
WE Science Citation Index Expanded (SCI-EXPANDED)
SC Neurosciences & Neurology; Surgery
GA 581CK
UT WOS:000177273400015
PM 12182773
DA 2023-08-27
ER

PT J
AU Dlouhy, BJ
   Chae, MP
   Teo, C
AF Dlouhy, Brian J.
   Chae, Michael P.
   Teo, Charles
TI The supraorbital eyebrow approach in children: clinical outcomes,
   cosmetic results, and complications
SO JOURNAL OF NEUROSURGERY-PEDIATRICS
LA English
DT Article
DE supraorbital craniotomy; keyhole; minimally invasive surgery; eyebrow;
   neuroendoscopy; craniopharyngioma; complications; cosmesis; technique
ID FRONTOLATERAL KEYHOLE CRANIOTOMY; COMMUNICATING ARTERY ANEURYSMS;
   SUPERCILIARY SKIN INCISION; ASSISTED BRAIN SURGERY; ANTERIOR CRANIAL
   FOSSA; TECHNICAL NOTE; SUPRATENTORIAL ANEURYSMS; SUPRABROW APPROACH;
   PTERIONAL APPROACH; SURGICAL-TREATMENT
AB OBJECT The supraorbital eyebrow approach utilizes an eyebrow skin incision to fashion a supraorbital craniotomy for exposure of the subfrontal corridor. This provides anterolateral access to surgical lesions in the anterior cranial fossa, parasellar regions, brainstem, and medial temporal lobe. With use of the endoscope, further areas can be accessed. This approach has been applied effectively in adults, but questions remain about its use in children-specifically with regard to adequate working space, effectiveness for achieving the desired results, cosmesis, and complications.
   METHODS The authors conducted a retrospective review of more than 450 cases involving patients of all ages who had undergone a supraorbital eyebrow approach performed by the senior author (C.T.) from 1995 to 2013. Only cases involving patients younger than 18 years with a minimum follow-up of 6 weeks were included in this study. All inpatient and outpatient records were retrospectively reviewed and clinical/operative outcomes, cosmetic results, and complications were recorded. In the present article, the authors briefly describe the surgical approach and highlight any differences in applying it in children.
   RESULTS Fifty-four pediatric patients who had undergone a supraorbital eyebrow approach met inclusion criteria. The pathological conditions consisted mostly of tumors or other resectable lesions. In a total of 51 resectable lesions, 44 surgeries resulted in a gross-total (100%) resection and 7 cases resulted in subtotal (50%-99%) resection. The endoscope assisted and expanded visualization or provided access to areas not reached by standard microscopic visualization in all cases. Cosmetic outcomes were excellent. In all cases, the incisional scar was barely visible at 6 weeks. In 3 cases a minor bone defect was observed on the forehead. Given the small size of the frontal sinus in children, no frontal sinus breaches occurred. Additionally, no CSF leak or wound infection was identified.
   CONCLUSIONS The supraorbital eyebrow approach is extremely effective in achieving desired results in properly selected cases in patients of all pediatric age ranges, from infants to teenagers. There is sufficient working space for the endoscope and all instruments, allowing for endoscopic assistance and bimanual surgical technique. Cosmetic results are excellent, and complications related to the approach are minimal.
C1 [Dlouhy, Brian J.] Univ Iowa, Hosp & Clin, Dept Neurosurg, Iowa City, IA 52242 USA.
   [Dlouhy, Brian J.; Chae, Michael P.; Teo, Charles] Prince Wales Hosp, Ctr Minimally Invas Neurosurg, Sydney, NSW, Australia.
C3 University of Iowa
RP Dlouhy, BJ (通讯作者)，Univ Iowa, Hosp & Clin, Dept Neurosurg, 200 Hawkins Dr, Iowa City, IA 52242 USA.
EM
OI Dlouhy, Brian/0000-0001-6724-688X
CR ALMEFTY O, 1985, SURG NEUROL, V23, P609, DOI 10.1016/0090-3019(85)90012-6
   Bognar L, 2003, CHILD NERV SYST, V19, P765, DOI 10.1007/s00381-003-0755-8
   Cavalcanti DD, 2010, NEUROSURGERY, V66, pONS205, DOI 10.1227/01.NEU.0000369948.37233.70
   Chalouhi N, 2013, NEUROSURGERY, V72, P437, DOI 10.1227/NEU.0b013e3182804e9c
   Cheng CM, 2013, J NEUROSURG, V118, P264, DOI 10.3171/2012.9.JNS09186
   Czirjak S, 2001, NEUROSURGERY, V48, P145, DOI 10.1097/00006123-200101000-00025
   Dare AO, 2001, J NEUROSURG, V95, P714, DOI 10.3171/jns.2001.95.4.0714
   DELASHAW JB, 1993, J NEUROSURG, V79, P615, DOI 10.3171/jns.1993.79.4.0615
   El Shafei HI, 2011, PEDIATR NEUROSURG, V47, P248, DOI 10.1159/000335399
   Fatemi N, 2009, NEUROSURGERY, V64, P269, DOI 10.1227/01.NEU.0000327857.22221.53
   Figueiredo EG, 2007, NEUROSURGERY, V61, P256, DOI 10.1227/01.neu.0000303978.11752.45
   Figueiredo EG, 2006, NEUROSURGERY, V59, P212, DOI 10.1227/01.NEU.0000223365.55701.F2
   Fries G, 1998, NEUROSURGERY, V42, P226, DOI 10.1097/00006123-199802000-00008
   Hayhurst C, 2011, OTOLARYNG CLIN N AM, V44, P953, DOI 10.1016/j.otc.2011.06.012
   Hernesniemi J, 2005, ACT NEUR S, V94, P17
   Jallo GI, 2005, J NEUROSURG, V103, P88, DOI 10.3171/ped.2005.103.1.0088
   JALLO GI, 2006, NEUROSURGERY S, V59
   JANE JA, 1982, NEUROSURGERY, V11, P537, DOI 10.1227/00006123-198210000-00016
   Jho HD, 1997, MINIM INVAS NEUROSUR, V40, P91, DOI 10.1055/s-2008-1053424
   Kanaan IN, 2005, MINIM INVAS NEUROSUR, V48, P34, DOI 10.1055/s-2004-830182
   Little AS, 2010, CRANIAL, CRANIOFACIAL AND SKULL BASE SURGERY, P27, DOI 10.1007/978-88-470-1167-0_4
   Maus M, 1999, OPHTHALMIC PLAST REC, V15, P166, DOI 10.1097/00002341-199905000-00005
   Park J, 2011, J NEUROSURG, V115, P700, DOI 10.3171/2011.5.JNS102087
   Park J, 2011, NEUROSURGERY, V68, P300, DOI 10.1227/NEU.0b013e3182124810
   Perneczky A, 1998, NEUROSURGERY, V42, P219, DOI 10.1097/00006123-199802000-00001
   Ramos-Zuniga R, 2002, NEUROSURGERY, V51, P125, DOI 10.1097/00006123-200207000-00019
   Reisch R, 2003, SURG NEUROL, V59, P223, DOI 10.1016/S0090-3019(02)01037-6
   Reisch R, 2009, MINIM INVAS NEUROSUR, V52, P163, DOI 10.1055/s-0029-1238285
   Reisch R, 2005, NEUROSURGERY, V57, P242, DOI 10.1227/01.NEU.0000178353.42777.2C
   Shanno G, 2001, NEUROSURGERY, V48, P559, DOI 10.1097/00006123-200103000-00020
   Steiger HJ, 2001, NEUROSURGERY, V48, P347, DOI 10.1097/00006123-200102000-00021
   Teo C, 2005, CHILD NERV SYST, V21, P696, DOI 10.1007/s00381-005-1204-7
   van Lindert E, 1998, SURG NEUROL, V49, P481, DOI 10.1016/S0090-3019(96)00539-3
   Warren WL, 2009, NEUROSURGERY, V64, P324, DOI 10.1227/01.NEU.0000338951.29171.07
   Wilson DA, 2014, WORLD NEUROSURG, V82, pE243, DOI 10.1016/j.wneu.2013.02.002
   Yasargil M G, 1976, Surg Neurol, V6, P83
NR 36
TC 22
Z9 25
U1 0
U2 3
PU AMER ASSOC NEUROLOGICAL SURGEONS
PI ROLLING MEADOWS
PA 5550 MEADOWBROOK DRIVE, ROLLING MEADOWS, IL 60008 USA
SN 1933-0707
EI 1933-0715
J9 J NEUROSURG-PEDIATR
JI J. Neurosurg.-Pediatr.
PD JAN
PY 2015
VL 15
IS 1
BP 12
EP 19
DI 10.3171/2014.10.PEDS1430
PG 8
WC Clinical Neurology; Pediatrics; Surgery
WE Science Citation Index Expanded (SCI-EXPANDED)
SC Neurosciences & Neurology; Pediatrics; Surgery
GA AX5DP
UT WOS:000346947500004
PM 25396702
OA Bronze
DA 2023-08-27
ER

PT J
AU Shintaku, M
   Matsumoto, R
   Sawa, H
   Nagashima, K
AF Shintaku, M
   Matsumoto, R
   Sawa, H
   Nagashima, K
TI Infection with JC virus and possible dysplastic ganglion-like
   transformation of the cerebral cortical neurons in a case of progressive
   multifocal leukoencephalopathy
SO JOURNAL OF NEUROPATHOLOGY AND EXPERIMENTAL NEUROLOGY
LA English
DT Article
DE dysplastic change; JC virus; neuronal infection; progressive multifocal
   leukoencephalopathy
ID NUCLEAR ANTIGEN EXPRESSION; HUMAN POLYOMAVIRUS JC; INSITU HYBRIDIZATION;
   BRAIN-TUMORS; CEREBROSPINAL-FLUID; TRANSGENIC MICE; CHOROID-PLEXUS;
   DNA-SEQUENCES; PAPOVAVIRUS; PROTEIN
AB Infection of the cerebral cortical neurons with JC virus (JCV) with possible dysplastic ganglion-like alteration of the infected neurons found in a case of progressive multifocal leukoencephalopathy (PML) is described. The patient was a 21-year-old man with common variable immunodeficiency who died of PML after a 9-month clinical course. Ar autopsy, the white matter of the cerebrum, brainstem, cerebellum, and spinal cord exhibited extensive demyelination and necrosis. Numerous inclusion-bearing oligodendrocytes and bizarre astrocytes were found. In the occipital and temporal cortex, thick bandlike aggregates of dysplastic ganglion-like cells (DGLCs) were found. These DGLCs showed immunohistochemical properties of neurons, and nuclei of some DGLCs were immunoreactive for large T antigen of SV40/JCV and p53, but not for capsid protein JCV VP1. In situ hybridization for mRNA of JCV large T antigen revealed positive signals in the nuclei of some DGLCs. These results indicate that JCV infected neurons and it is suggested that binding of the large T antigen with cellular proteins could have resulted in the dysplastic, ganglion cell-like change of the infected neurons, although the possibility that the aggregates of DGLCs represent a pre-existent malformative lesion of the cortex cannot be excluded completely.
C1 Osaka Red Cross Hosp, Dept Pathol, Osaka 5438555, Japan.
   Osaka Red Cross Hosp, Dept Neurol, Osaka 5438555, Japan.
   Hokkaido Univ, Sch Med, CREST,Lab Mol & Cellular Pathol, Japan Sci & Technol Corp, Sapporo, Hokkaido 060, Japan.
C3 Osaka Red Cross Hospital; Osaka Red Cross Hospital; Hokkaido University;
   Japan Science & Technology Agency (JST)
RP Shintaku, M (通讯作者)，Osaka Red Cross Hosp, Dept Pathol, 5-53 Fudegasaki, Osaka 5438555, Japan.
RI Matsumoto, Riki/I-8397-2016; Sawa, Hirofumi/AAW-8816-2021; Sawa,
   Hirofumi/F-6954-2012
OI Matsumoto, Riki/0000-0003-3985-9210; Sawa, Hirofumi/0000-0003-2569-2755
CR AKATANI K, 1994, J MED VIROL, V43, P13, DOI 10.1002/jmv.1890430104
   AKSAMIT AJ, 1985, ANN NEUROL, V18, P490, DOI 10.1002/ana.410180412
   AKSAMIT AJ, 1986, NEUROLOGY, V36, P499, DOI 10.1212/WNL.36.4.499
   AKSAMIT AJ, 1990, NEUROLOGY, V40, P1073, DOI 10.1212/WNL.40.7.1073
   ARIZA A, 1994, HUM PATHOL, V25, P1341, DOI 10.1016/0046-8177(94)90095-7
   Ariza A, 1996, J NEUROPATH EXP NEUR, V55, P144, DOI 10.1097/00005072-199602000-00002
   BERGSAGEL DJ, 1992, NEW ENGL J MED, V326, P988, DOI 10.1056/NEJM199204093261504
   Bezrodnik L, 1998, CLIN INFECT DIS, V27, P181, DOI 10.1086/514617
   Boldorini R, 1998, J NEUROVIROL, V4, P242, DOI 10.3109/13550289809114524
   CASTAIGNE P, 1974, REV NEUROL-FRANCE, V130, P379
   CHOU SM, 1975, J NEUROPATH EXP NEUR, V34, P81
   EGAN JD, 1980, TRANSPLANTATION, V29, P84, DOI 10.1097/00007890-198001000-00021
   Franks RR, 1996, ONCOGENE, V12, P2573
   Gallia GL, 1998, J NEUROVIROL, V4, P175, DOI 10.3109/13550289809114517
   GIARUSSO MH, 1978, J NEUROL SCI, V35, P391, DOI 10.1016/0022-510X(78)90019-9
   GREENLEE JE, 1978, ANN NEUROL, V3, P479, DOI 10.1002/ana.410030604
   GULLOTTA F, 1992, PATHOL RES PRACT, V188, P964, DOI 10.1016/S0344-0338(11)81239-2
   Herbarth B, 1998, GLIA, V22, P415, DOI 10.1002/(SICI)1098-1136(199804)22:4<415::AID-GLIA11>3.0.CO;2-Z
   HIROSE T, 1995, ACTA NEUROPATHOL, V90, P387
   HO K, 1980, ACTA NEUROPATHOL, V52, P81, DOI 10.1007/BF00687233
   HOUFF SA, 1988, NEW ENGL J MED, V318, P301, DOI 10.1056/NEJM198802043180507
   IRONSIDE JW, 1989, J PATHOL, V157, P291, DOI 10.1002/path.1711570405
   ITOYAMA Y, 1982, ANN NEUROL, V11, P396, DOI 10.1002/ana.410110414
   KOSAKA H, 1980, J NEUROSURG, V52, P367, DOI 10.3171/jns.1980.52.3.0367
   Krynska B, 1999, P NATL ACAD SCI USA, V96, P11519, DOI 10.1073/pnas.96.20.11519
   KUCHELMEISTER K, 1993, NEUROPATH APPL NEURO, V19, P398, DOI 10.1111/j.1365-2990.1993.tb00460.x
   LAMMIE GA, 1994, ACTA NEUROPATHOL, V88, P465
   LEDNICKY JA, 1995, VIROLOGY, V212, P710, DOI 10.1006/viro.1995.1529
   LONDON WT, 1978, SCIENCE, V201, P1246, DOI 10.1126/science.211583
   MAJOR EO, 1992, CLIN MICROBIOL REV, V5, P49, DOI 10.1128/CMR.5.1.49-73.1992
   MATSUDA M, 1987, JNCI-J NATL CANCER I, V79, P585
   MAZLO M, 1989, J NEUROPATH EXP NEUR, V48, P315
   MAZLO M, 1982, ACTA NEUROPATHOL, V56, P45, DOI 10.1007/BF00691181
   NAGASHIMA K, 1984, AM J PATHOL, V116, P455
   NAGASHIMA K, 1991, BRAIN TUMOR PATHOL, V8, P127
   NAKAMURA S, 1985, CLIN NEUROL TOKYO, V25, P1148
   PADGETT BL, 1971, LANCET, V1, P1257
   REDFEARN A, 1993, PEDIATR INFECT DIS J, V12, P399, DOI 10.1097/00006454-199305000-00010
   Rencic A, 1996, P NATL ACAD SCI USA, V93, P7352, DOI 10.1073/pnas.93.14.7352
   RHEIN GMZ, 1965, SCIENCE, V148, P1477, DOI 10.1126/science.148.3676.1477
   RICHARDSON E, 1961, NEW ENGL J MED, V265, P815, DOI 10.1056/NEJM196110262651701
   Rudge P, 1996, BRAIN, V119, P1, DOI 10.1093/brain/119.1.1
   Seemayer TA, 1996, ANDERSONS PATHOLOGY, P615
   Shishido-Hara Y, 2000, J VIROL, V74, P1840, DOI 10.1128/JVI.74.4.1840-1853.2000
   SIMA AAF, 1983, ANN NEUROL, V14, P183, DOI 10.1002/ana.410140205
   SMALL JA, 1986, P NATL ACAD SCI USA, V83, P8288, DOI 10.1073/pnas.83.21.8288
   STONER GL, 1986, P NATL ACAD SCI USA, V83, P2271, DOI 10.1073/pnas.83.7.2271
   Sugimoto C, 1998, ARCH VIROL, V143, P249, DOI 10.1007/s007050050284
   Suzuki G, 1999, J IMMUNOL, V162, P5981
   Takahashi H, 1999, CLIN DIAGN LAB IMMUN, V6, P983, DOI 10.1128/CDLI.6.6.983-985.1999
   TAYLOR DC, 1971, J NEUROL NEUROSUR PS, V34, P369, DOI 10.1136/jnnp.34.4.369
   WALDMANN TA, 1988, IMMUNOLOGICAL DISEAS, P411
   WALKER DL, 1973, SCIENCE, V181, P674, DOI 10.1126/science.181.4100.674
   WATANABE I, 1976, ACTA NEUROPATHOL, V36, P101, DOI 10.1007/BF00685273
   Weggen S, 2000, BRAIN PATHOL, V10, P85
   Zu Rhein G M, 1983, Prog Clin Biol Res, V105, P205
NR 56
TC 33
Z9 33
U1 0
U2 0
PU OXFORD UNIV PRESS INC
PI CARY
PA JOURNALS DEPT, 2001 EVANS RD, CARY, NC 27513 USA
SN 0022-3069
EI 1554-6578
J9 J NEUROPATH EXP NEUR
JI J. Neuropathol. Exp. Neurol.
PD OCT
PY 2000
VL 59
IS 10
BP 921
EP 929
DI 10.1093/jnen/59.10.921
PG 9
WC Clinical Neurology; Neurosciences; Pathology
WE Science Citation Index Expanded (SCI-EXPANDED)
SC Neurosciences & Neurology; Pathology
GA 364MK
UT WOS:000089896000009
PM 11079782
OA Bronze
DA 2023-08-27
ER

PT J
AU Kawasaki, T
   Ago, Y
   Kitao, T
   Nashida, T
   Takagi, A
   Takuma, K
   Matsuda, T
AF Kawasaki, Toshiyuki
   Ago, Yukio
   Kitao, Tatsuya
   Nashida, Tetsuaki
   Takagi, Akiko
   Takuma, Kazuhiro
   Matsuda, Toshio
TI A neuroprotective agent, T-817MA (1-{3-[2-(1-benzothiophen-5-yl)
   ethoxy]propyl} azetidin-3-ol maleate), prevents
   1-methyl-4-phenyl-1,2,3,6-tetrahydropyridine-induced neurotoxicity in
   mice
SO NEUROPHARMACOLOGY
LA English
DT Article
DE 1-Methyl-4-phenyl-1,2,3,6-tetrahydropyricline (MPTP); T-817MA; Dopamine;
   Neurotoxicity; Parkinson's disease; Microglia
ID TUMOR-NECROSIS-FACTOR; PARKINSONS-DISEASE; DOPAMINERGIC NEUROTOXICITY;
   MICROGLIAL ACTIVATION; NITRIC-OXIDE; FACTOR-ALPHA; CEREBROSPINAL-FLUID;
   NEUROTROPHIC AGENT; RADICAL SCAVENGER; OXIDATIVE STRESS
AB T-817MA (1-{3-[2-(1-benzothiophen-5-yl)ethoxy]propyl] azetidin-3-ol maleate) is a candidate therapeutic agent for Alzheimer's disease that inhibits oxidative stress and nitric oxide-induced neurotoxicity and acts as a neurotrophic factor. The present study examines the effect of T-817MA on 1-methyl-4phenyl-1,2,3,6-tetrahydropyridine (MPTP)-induced dopaminergic neurotoxicity in C57BL/6J mice. MPTP treatment (10 mg/kg, s.c. x 4 at 2-h intervals) impaired rotarod performance, and T-817MA improved this deficit. MPTP treatment also decreased dopamine levels and tyrosine hydroxylase immunostaining in the substantia nigra (SNc) and striatum. Pretreatment with T-817MA (10 and 30 mg/kg as T-817, p.o.) attenuated these decreases in dopamine levels and tyrosine hydroxylase immunoreactivity, but did not affect brain levels of 1-methyl-4-phenylpyridinium ion, an active metabolite of MPTP. The protective effect was almost complete in the SNc, but only partial in the striatum. MPTP increased levels of the lipid peroxidation product, thiobarbituric acid reactive substance, only in the midbrain, which could be blocked by T-817MA. MPTP caused microglial activation both in the SNc and striatum, but T-817MA did not affect the activation of microglia. These results suggest that T-817MA protects against MPTP-induced neurotoxicity by blocking lipid peroxidation in the SNc, and imply that this compound may be useful for treating neurodegenerative disorders related to oxidative stress, such as Parkinson's disease. (c) 2008 Elsevier Ltd. All rights reserved.
C1 [Kawasaki, Toshiyuki; Ago, Yukio; Kitao, Tatsuya; Nashida, Tetsuaki; Takuma, Kazuhiro; Matsuda, Toshio] Osaka Univ, Lab Med Pharmacol, Grad Sch Pharmaceut Sci, Suita, Osaka 5650871, Japan.
   [Takagi, Akiko] Toyama Chem Co Ltd, Res Labs, Res Dept 2, Toyama, Japan.
   [Matsuda, Toshio] Osaka Univ, Grad Sch Med, Dept Expt Dis Model, Osaka Hamamstsu Joint Res Ctr Child Mental Dev, Suita, Osaka 5650871, Japan.
C3 Osaka University; Toyama Chemical Co., Ltd.; Osaka University
RP Matsuda, T (通讯作者)，Osaka Univ, Lab Med Pharmacol, Grad Sch Pharmaceut Sci, 1-6 Yamada Oka, Suita, Osaka 5650871, Japan.
EM
RI Ago, Yukio/H-9142-2019
OI Ago, Yukio/0000-0001-7562-4000
FU Ministry of Education, Science, Sports and Culture of Japan; Toyama
   Chemical Co., Ltd
FX This work was supported in part by grants from the Ministry of
   Education, Science, Sports and Culture of Japan, and from Toyama
   Chemical Co., Ltd.
CR BOKA G, 1994, NEUROSCI LETT, V172, P151, DOI 10.1016/0304-3940(94)90684-X
   Du YS, 2001, P NATL ACAD SCI USA, V98, P14669, DOI 10.1073/pnas.251341998
   Fukushima T, 2006, NEUROCHEM INT, V48, P124, DOI 10.1016/j.neuint.2005.08.012
   Gao HM, 2003, J NEUROSCI, V23, P1228, DOI 10.1523/JNEUROSCI.23-04-01228.2003
   Gao HM, 2002, J NEUROCHEM, V81, P1285, DOI 10.1046/j.1471-4159.2002.00928.x
   Halliwell B, 2006, J NEUROCHEM, V97, P1634, DOI 10.1111/j.1471-4159.2006.03907.x
   Hirata K, 2005, J PHARMACOL EXP THER, V314, P252, DOI 10.1124/jpet.105.083543
   Kato H, 2004, BRAIN RES, V1030, P66, DOI 10.1016/j.brainres.2004.09.046
   Kawasaki T, 2007, J PHARMACOL EXP THER, V322, P274, DOI 10.1124/jpet.106.119206
   Kawasaki T, 2006, EUR J PHARMACOL, V542, P92, DOI 10.1016/j.ejphar.2006.05.012
   Liberatore GT, 1999, NAT MED, V5, P1403
   Lin X, 2003, BBA-MOL BASIS DIS, V1639, P195, DOI 10.1016/j.bbadis.2003.09.007
   McGeer PL, 2003, ANN NEUROL, V54, P599, DOI 10.1002/ana.10728
   Mogi M, 1996, NEUROSCI LETT, V211, P13, DOI 10.1016/0304-3940(96)12706-3
   MOGI M, 1994, NEUROSCI LETT, V165, P208, DOI 10.1016/0304-3940(94)90746-3
   Moore DJ, 2005, ANNU REV NEUROSCI, V28, P57, DOI 10.1146/annurev.neuro.28.061604.135718
   Nguyen PTH, 2007, HIPPOCAMPUS, V17, P443, DOI 10.1002/hipo.20281
   Satoh K, 2002, REDOX REP, V7, P219, DOI 10.1179/135100002125000587
   SCHAPIRA AHV, 1990, J NEUROCHEM, V54, P823, DOI 10.1111/j.1471-4159.1990.tb02325.x
   Sedelis M, 2001, BEHAV BRAIN RES, V125, P109, DOI 10.1016/S0166-4328(01)00309-6
   Sriram K, 2006, J NEUROCHEM, V96, P706, DOI 10.1111/j.1471-4159.2005.03566.x
   Teismann P, 2003, P NATL ACAD SCI USA, V100, P5473, DOI 10.1073/pnas.0837397100
   Tieu K, 2003, IUBMB LIFE, V55, P329, DOI 10.1080/1521654032000114320
   Whitton PS, 2007, BRIT J PHARMACOL, V150, P963, DOI 10.1038/sj.bjp.0707167
   Wu DC, 2002, J NEUROSCI, V22, P1763, DOI 10.1523/JNEUROSCI.22-05-01763.2002
   Wu DC, 2003, P NATL ACAD SCI USA, V100, P6145, DOI 10.1073/pnas.0937239100
   Yang LC, 2003, J NEUROSCI RES, V74, P278, DOI 10.1002/jnr.10709
NR 27
TC 14
Z9 19
U1 0
U2 0
PU PERGAMON-ELSEVIER SCIENCE LTD
PI OXFORD
PA THE BOULEVARD, LANGFORD LANE, KIDLINGTON, OXFORD OX5 1GB, ENGLAND
SN 0028-3908
J9 NEUROPHARMACOLOGY
JI Neuropharmacology
PD OCT
PY 2008
VL 55
IS 5
BP 654
EP 660
DI 10.1016/j.neuropharm.2008.05.032
PG 7
WC Neurosciences; Pharmacology & Pharmacy
WE Science Citation Index Expanded (SCI-EXPANDED)
SC Neurosciences & Neurology; Pharmacology & Pharmacy
GA 361VF
UT WOS:000260154600003
PM 18573265
DA 2023-08-27
ER

PT J
AU Binukumar, BK
   Bal, A
   Gill, KD
AF Binukumar, B. K.
   Bal, Amanjit
   Gill, Kiran Dip
TI Chronic Dichlorvos Exposure: Microglial Activation, Proinflammatory
   Cytokines and Damage to Nigrostriatal Dopaminergic System
SO NEUROMOLECULAR MEDICINE
LA English
DT Article
DE Parkinson's disease; Neurodegeneration; Microglia; Neuroinflammation;
   Dichlorvos
ID TUMOR-NECROSIS-FACTOR; ENVIRONMENTAL RISK-FACTORS; ALPHA TNF-ALPHA;
   PARKINSONS-DISEASE; NADPH OXIDASE; SUBSTANTIA-NIGRA; MITOCHONDRIAL
   DYSFUNCTION; OXIDATIVE STRESS; IN-VITRO; MODEL
AB Dopaminergic cells in the substantia nigra are highly vulnerable to the neurodegenerative process of Parkinson's disease. Therefore, mechanisms that enhance their susceptibility to injury bear important implications for disease pathogenesis. We have previously shown that chronic dichlorvos exposure caused nigrostriatal dopaminergic degeneration and significant behavioral impairments. In this study, we analyzed the relationship between microglial activation and dopaminergic neurodegeneration to examine the possibility that neuroinflammation may induce dopaminergic neuronal loss in the nigrostriatal system. Chronic dichlorvos exposure causes microglial activation including induction of NADPH oxidase and a selective loss of dopaminergic neurons in rat. Microglial marker expression was increased at transcription as well as translational levels in the substantia nigra (SN) and corpus striatum (CS) of rats exposed to dichlorvos. Activated microglia were seen in SN and CS of dichlorvos-treated animals but were rarely observed in controls. Immunostaining revealed lesser number of TH-positive neurons and higher number of microglia in SN and CS regions after dichlorvos treatment. The mRNA and protein levels of the NADPH oxidase main subunit gp91(phox) were significantly increased after dichlorvos administration. Dichlorvos exposure also leads to increased level of microglial noxious mediators such as IL-1 beta, TNF-alpha and IL-6 in ventral midbrain and CS at transcription as well as translational levels. Data indicate that microglial activation and consequent induction of NADPH oxidase and proinflammatory cytokines such as TNF-alpha, IL-1 beta and IL-6 may act as risk factors for Parkinson's disease by increasing the vulnerability of dopaminergic cells to dichlorvos toxic injury.
C1 [Binukumar, B. K.; Gill, Kiran Dip] Postgrad Inst Med Educ & Res, Dept Biochem, Chandigarh 160012, India.
   [Bal, Amanjit] Postgrad Inst Med Educ & Res, Dept Histopathol, Chandigarh 160012, India.
C3 Post Graduate Institute of Medical Education & Research (PGIMER),
   Chandigarh; Post Graduate Institute of Medical Education & Research
   (PGIMER), Chandigarh
RP Gill, KD (通讯作者)，Postgrad Inst Med Educ & Res, Dept Biochem, Chandigarh 160012, India.
EM
OI BK, Binukumar/0000-0003-0933-4691
FU Indian Council of Medical Research, New Delhi, India
FX The financial assistance provided to Binukumar B.K. by Indian Council of
   Medical Research, New Delhi, India, is greatly acknowledged.
CR Banati RB, 1998, MOVEMENT DISORD, V13, P221, DOI 10.1002/mds.870130205
   Barcia C, 2005, PARKINSONISM RELAT D, V11, P435, DOI 10.1016/j.parkreldis.2005.05.006
   Binukumar BK, 2010, MOL BRAIN, V3, DOI 10.1186/1756-6606-3-35
   Block ML, 2007, NAT REV NEUROSCI, V8, P57, DOI 10.1038/nrn2038
   BOKA G, 1994, NEUROSCI LETT, V172, P151, DOI 10.1016/0304-3940(94)90684-X
   Brodacki B, 2008, NEUROSCI LETT, V441, P158, DOI 10.1016/j.neulet.2008.06.040
   Cardona AE, 2006, NAT PROTOC, V1, P1947, DOI 10.1038/nprot.2006.327
   Carvey PM, 2005, J NEURAL TRANSM, V112, P601, DOI 10.1007/s00702-004-0222-z
   Chen HL, 2005, ANN NEUROL, V58, P963, DOI 10.1002/ana.20682
   Fahn S, 2003, ANN NY ACAD SCI, V991, P1, DOI 10.1111/j.1749-6632.2003.tb07458.x
   Ferger B, 2004, J NEUROCHEM, V89, P822, DOI 10.1111/j.1471-4159.2004.02399.x
   Ferrer I, 2001, J NEURAL TRANSM, V108, P1383, DOI 10.1007/s007020100015
   Gao HM, 2003, FASEB J, V17, DOI 10.1096/fj.03-0109fje
   Gao HM, 2003, J NEUROSCI, V23, P6181
   Gerhard A, 2006, NEUROBIOL DIS, V21, P404, DOI 10.1016/j.nbd.2005.08.002
   Green SP, 2001, J CEREBR BLOOD F MET, V21, P374, DOI 10.1097/00004647-200104000-00006
   Greenamyre JT, 1999, BIOCHEM SOC SYMP, V66, P85, DOI 10.1042/bss0660085
   Hald A, 2005, EXP NEUROL, V193, P279, DOI 10.1016/j.expneurol.2005.01.013
   Hartmann A, 2001, J NEUROSCI, V21, P2247, DOI 10.1523/JNEUROSCI.21-07-02247.2001
   HASEGAWA E, 1990, BIOCHEM BIOPH RES CO, V170, P1049, DOI 10.1016/0006-291X(90)90498-C
   Hunot S, 1999, J NEUROSCI, V19, P3440
   Kaur P, 2007, NEUROTOXICOLOGY, V28, P1208, DOI 10.1016/j.neuro.2007.08.001
   Kim WG, 2000, J NEUROSCI, V20, P6309, DOI 10.1523/JNEUROSCI.20-16-06309.2000
   Langston JW, 1999, ANN NEUROL, V46, P598, DOI 10.1002/1531-8249(199910)46:4<598::AID-ANA7>3.0.CO;2-F
   LAWSON LJ, 1990, NEUROSCIENCE, V39, P151, DOI 10.1016/0306-4522(90)90229-W
   Ling ZD, 2002, MOVEMENT DISORD, V17, P116, DOI 10.1002/mds.10078
   Liou HH, 1997, NEUROLOGY, V48, P1583, DOI 10.1212/WNL.48.6.1583
   LOWRY OH, 1951, J BIOL CHEM, V193, P265
   Manning-Bog AB, 2002, J BIOL CHEM, V277, P1641, DOI 10.1074/jbc.C100560200
   Mayeux R, 2003, ANNU REV NEUROSCI, V26, P81, DOI 10.1146/annurev.neuro.26.043002.094919
   McCormack AL, 2002, NEUROBIOL DIS, V10, P119, DOI 10.1006/nbdi.2002.0507
   McGeer PL, 2004, PARKINSONISM RELAT D, V10, pS3, DOI 10.1016/j.parkreldis.2004.01.005
   McGeer PL, 2002, NEUROSCI LETT, V326, P67, DOI 10.1016/S0304-3940(02)00300-2
   MCGEER PL, 1988, NEUROLOGY, V38, P1285, DOI 10.1212/WNL.38.8.1285
   MOGI M, 1994, NEUROSCI LETT, V165, P208, DOI 10.1016/0304-3940(94)90746-3
   Mogi M, 2000, J NEURAL TRANSM, V107, P335, DOI 10.1007/s007020050028
   Ouchi Y, 2005, ANN NEUROL, V57, P168, DOI 10.1002/ana.20338
   Parvathenani LK, 2003, J BIOL CHEM, V278, P13309, DOI 10.1074/jbc.M209478200
   Priyadarshi A, 2001, ENVIRON RES, V86, P122, DOI 10.1006/enrs.2001.4264
   RAJPUT AH, 1987, CAN J NEUROL SCI, V14, P414, DOI 10.1017/S0317167100037823
   Rousselet E, 2002, EXP NEUROL, V177, P183, DOI 10.1006/exnr.2002.7960
   Sawada M, 2006, J NEURAL TRANSM-SUPP, P373
   Schapira AHV, 2007, CELL DEATH DIFFER, V14, P1261, DOI 10.1038/sj.cdd.4402160
   Schulte T, 2002, NEUROSCI LETT, V326, P70, DOI 10.1016/S0304-3940(02)00301-4
   SEMCHUK KM, 1992, NEUROLOGY, V42, P1328, DOI 10.1212/WNL.42.7.1328
   Sherer TB, 2003, NEUROSCI LETT, V341, P87, DOI 10.1016/S0304-3940(03)00172-1
   Spillantini MG, 1997, NATURE, V388, P839, DOI 10.1038/42166
   Sriram K, 2002, FASEB J, V16, P1474, DOI 10.1096/fj.02-0216fje
   Stone TW, 2007, J NEUROSCI RES, V85, P1077, DOI 10.1002/jnr.21212
   SWANSON LW, 1980, NEUROENDOCRINOLOGY, V31, P410, DOI 10.1159/000123111
   Teismann P, 2004, CELL TISSUE RES, V318, P149, DOI 10.1007/s00441-004-0944-0
   Thiruchelvam M, 2003, EUR J NEUROSCI, V18, P589, DOI 10.1046/j.1460-9568.2003.02781.x
   Thiruchelvam M, 2002, NEUROTOXICOLOGY, V23, P621, DOI 10.1016/S0161-813X(02)00092-X
   Thiruchelvam M, 2000, J NEUROSCI, V20, P9207
   TURRENS JF, 1980, BIOCHEM J, V191, P421, DOI 10.1042/bj1910421
   Van Den Eeden SK, 2003, AM J EPIDEMIOL, V157, P1015, DOI 10.1093/aje/kwg068
   Wooten GF, 1997, MOVEMENT DISORDERS N, P153
   Wu DC, 2003, P NATL ACAD SCI USA, V100, P6145, DOI 10.1073/pnas.0937239100
NR 58
TC 24
Z9 25
U1 0
U2 4
PU HUMANA PRESS INC
PI TOTOWA
PA 999 RIVERVIEW DRIVE SUITE 208, TOTOWA, NJ 07512 USA
SN 1535-1084
EI 1559-1174
J9 NEUROMOL MED
JI Neuromol. Med.
PD DEC
PY 2011
VL 13
IS 4
BP 251
EP 265
DI 10.1007/s12017-011-8156-8
PG 15
WC Neurosciences
WE Science Citation Index Expanded (SCI-EXPANDED)
SC Neurosciences & Neurology
GA 893KS
UT WOS:000300355400003
PM 21964614
DA 2023-08-27
ER

PT J
AU Huang, XT
   Kou, XX
   Zhan, T
   Wei, GK
   He, FA
   Mao, XL
   Yang, HD
AF Huang, Xiaotong
   Kou, Xiaoxing
   Zhan, Ting
   Wei, Guokun
   He, Feinan
   Mao, Xueli
   Yang, Haidi
TI Apoptotic vesicles resist oxidative damage in noise-induced hearing loss
   through activation of FOXO3a-SOD2 pathway
SO STEM CELL RESEARCH & THERAPY
LA English
DT Article
DE Mesenchymal stem cell; Apoptotic vesicles; Oxidative stress;
   Noise-induced hearing loss; Mitochondrial superoxide dismutase 2 (SOD2)
ID MESENCHYMAL STROMAL CELLS; STEM-CELLS; SUPEROXIDE-DISMUTASE; COCHLEAR;
   INFLAMMATION; TRANSPLANTATION; SURVIVAL; TARGET; STRESS; MODEL
AB BackgroundMesenchymal stem cell (MSC) transplantation is a promising therapeutic approach for noise-induced hearing loss (NIHL). As the indispensable role of apoptosis in MSC transplantation was raised, the benefits of MSC-derived apoptotic vesicles (apoVs) in several disease models have been proved. However, whether apoVs benefit in NIHL have not been studied yet.MethodsFemale CBA/J mice and HEI-OC1 cells were used in this study. Flow cytometry, nanoparticle tracking analysis (NTA) and transmission electron microscopy (TEM) were used to characterize apoVs. Proteomic analysis was used to identify function proteins in apoVs. Immunofluorescence was used to reveal distribution pattern. Auditory brainstem response (ABR) test was used to measure the effect of apoVs treatment. DCFH-DA staining and MitoSOX staining were used to indicate oxidative damage. Western-blot and qRT-PCR were used to study the signaling pathways.ResultsWe found that apoVs can be endocytosed by hair cells through systemic administration. Importantly, apoVs administration effectively attenuated NIHL and reduced hair cell loss by resisting oxidative damage in vivo. Further, apoVs application activated forkhead box o3 (FOXO3a)-mitochondrial superoxide dismutase 2(SOD2) pathway, which may relate to signal transduction and activators of transcription 3 (STAT3) in apoVs.ConclusionsThese findings uncovered the role of apoVs in preventing NIHL and resisting oxidative damage, indicating that apoVs is a promising way for inner ear delivery and a prospective cell-free therapy for NIHL.
C1 [Huang, Xiaotong; Wei, Guokun; Yang, Haidi] Sun Yat Sen Univ, Sun Yat Sen Mem Hosp, Dept Otolaryngol, Guangzhou 510120, Peoples R China.
   [Huang, Xiaotong; Wei, Guokun; Yang, Haidi] Sun Yat Sen Univ, Sun Yat Sen Mem Hosp, Med Res Ctr, Guangdong Prov Key Lab Malignant Tumor Epigenet &, Guangzhou 510120, Peoples R China.
   [Kou, Xiaoxing; Mao, Xueli] Sun Yat Sen Univ, Hosp Stomatol, Guanghua Sch Stomatol, South China Ctr Craniofacial Stem Cell Res,Guangdo, Guangzhou 510055, Peoples R China.
   [Kou, Xiaoxing] Sun Yat Sen Univ, Minist Educ, Key Lab Stem Cells & Tissue Engn, Guangzhou 510080, Peoples R China.
   [Zhan, Ting] Southern Med Univ, Zhujiang Hosp, Dept Otolaryngol, Guangzhou 510285, Peoples R China.
   [He, Feinan] Second Hosp Jilin Univ, Dept Otorhinolaryngol Head & Neck Surg, Changchun 130041, Peoples R China.
   [Yang, Haidi] Guangzhou Xinhua Univ, Dept Hearing & Speech Sci, Guangzhou 510310, Peoples R China.
C3 Sun Yat Sen University; Sun Yat Sen University; Sun Yat Sen University;
   Sun Yat Sen University; Southern Medical University - China; Jilin
   University
RP Yang, HD (通讯作者)，Sun Yat Sen Univ, Sun Yat Sen Mem Hosp, Dept Otolaryngol, Guangzhou 510120, Peoples R China.; Yang, HD (通讯作者)，Sun Yat Sen Univ, Sun Yat Sen Mem Hosp, Med Res Ctr, Guangdong Prov Key Lab Malignant Tumor Epigenet &, Guangzhou 510120, Peoples R China.; Mao, XL (通讯作者)，Sun Yat Sen Univ, Hosp Stomatol, Guanghua Sch Stomatol, South China Ctr Craniofacial Stem Cell Res,Guangdo, Guangzhou 510055, Peoples R China.; Yang, HD (通讯作者)，Guangzhou Xinhua Univ, Dept Hearing & Speech Sci, Guangzhou 510310, Peoples R China.
EM
RI huang, xiaotong/GSJ-1295-2022
CR Abd El Raouf HHH, 2019, J CHEM NEUROANAT, V98, P139, DOI 10.1016/j.jchemneu.2019.04.008
   Bae WY, 2008, LARYNGOSCOPE, V118, P1659, DOI 10.1097/MLG.0b013e31817c1303
   Baglio SR, 2015, STEM CELL RES THER, V6, DOI 10.1186/s13287-015-0116-z
   Bodmer D, 2017, EXPERT OPIN DRUG DIS, V12, P1161, DOI 10.1080/17460441.2017.1372744
   Bolduc JA, 2019, FREE RADICAL BIO MED, V132, P73, DOI 10.1016/j.freeradbiomed.2018.08.038
   Chang GD, 2019, INT IMMUNOPHARMACOL, V71, P361, DOI 10.1016/j.intimp.2019.03.056
   Chang NC, 2009, AM J OTOLARYNG, V30, P396, DOI 10.1016/j.amjoto.2008.08.001
   Chen HC, 2019, INT J PEDIATR OTORHI, V117, P67, DOI 10.1016/j.ijporl.2018.11.018
   Chen J, 2017, ACTA OTO-LARYNGOL, V137, P1136, DOI 10.1080/00016489.2017.1342045
   Chen T, 2022, NUCLEIC ACIDS RES, V50, pD1522, DOI 10.1093/nar/gkab1081
   Chen Y, 2009, CELL DEATH DIFFER, V16, P1040, DOI 10.1038/cdd.2009.49
   Cousins Rick, 2022, Prog Med Chem, V61, P1, DOI 10.1016/bs.pmch.2022.05.001
   Dominici M, 2006, CYTOTHERAPY, V8, P315, DOI 10.1080/14653240600855905
   Elliott RO, 2021, PHARMACEUTICS, V13, DOI 10.3390/pharmaceutics13010122
   Fetoni AR, 2019, FREE RADICAL BIO MED, V135, P46, DOI 10.1016/j.freeradbiomed.2019.02.022
   Fetoni AR, 2013, J NEUROSCI, V33, P4011, DOI 10.1523/JNEUROSCI.2282-12.2013
   Fortunato G, 2004, CLIN CHEM, V50, P2012, DOI 10.1373/clinchem.2004.037788
   Fridovich I, 1997, J BIOL CHEM, V272, P18515, DOI 10.1074/jbc.272.30.18515
   Fu BB, 2017, BIOCHEM BIOPH RES CO, V486, P198, DOI 10.1016/j.bbrc.2017.03.027
   Fu Y, 2021, CELL DEATH DIS, V12, DOI 10.1038/s41419-021-03883-6
   Graydon K, 2019, J LARYNGOL OTOL, V133, P18, DOI 10.1017/S0022215118001275
   Gyorgy B, 2017, MOL THER, V25, P379, DOI 10.1016/j.ymthe.2016.12.010
   Haile LM, 2021, LANCET, V397, P996, DOI 10.1016/S0140-6736(21)00516-X
   Han C, 2016, NEUROBIOL AGING, V43, P58, DOI 10.1016/j.neurobiolaging.2016.03.023
   Karp JM, 2009, CELL STEM CELL, V4, P206, DOI 10.1016/j.stem.2009.02.001
   Katsuda T, 2015, STEM CELL RES THER, V6, DOI 10.1186/s13287-015-0214-y
   Lee CH, 2020, NEUROSCI LETT, V722, DOI 10.1016/j.neulet.2020.134838
   Lee YY, 2020, BBA-MOL CELL RES, V1867, DOI 10.1016/j.bbamcr.2020.118850
   Lei Q, 2022, ACS CHEM NEUROSCI, V13, P112, DOI 10.1021/acschemneuro.1c00599
   Leibacher J, 2017, CYTOTHERAPY, V19, P61, DOI 10.1016/j.jcyt.2016.09.010
   Li Q, 2021, MOL MED REP, V24
   Liang WQ, 2021, FRONT CELL DEV BIOL, V9, DOI 10.3389/fcell.2021.766512
   Liang Z, 2021, J NEUROPHYSIOL
   Lin QY, 2022, FRONT BIOENG BIOTECH, V9, DOI 10.3389/fbioe.2021.809443
   Liu DW, 2018, CELL RES, V28, P918, DOI 10.1038/s41422-018-0070-2
   Liu HW, 2021, STEM CELL RES THER, V12, DOI 10.1186/s13287-021-02382-x
   Liu H, 2020, AUTOPHAGY, V16, P2140, DOI 10.1080/15548627.2020.1717128
   Liu K, 2021, INT J NANOMED, V16, P1423, DOI 10.2147/IJN.S289914
   Liu SS, 2022, FRONT NEUROSCI-SWITZ, V16, DOI 10.3389/fnins.2022.867453
   Ma J, 2019, NUCLEIC ACIDS RES, V47, pD1211, DOI 10.1093/nar/gky869
   Mi HY, 2019, NAT PROTOC, V14, P703, DOI 10.1038/s41596-019-0128-8
   Miao L, 2021, INT J ENV RES PUB HE, V19
   Minami SB, 2004, J NEUROSCI RES, V78, P383, DOI 10.1002/jnr.20267
   Nikfarjam S, 2020, J TRANSL MED, V18, DOI 10.1186/s12967-020-02622-3
   Oh HM, 2012, J BIOL CHEM, V287, P30436, DOI 10.1074/jbc.M112.359661
   Oh HM, 2011, J BIOL CHEM, V286, P30888, DOI 10.1074/jbc.M111.253500
   Ohinata Y, 2000, BRAIN RES, V878, P163, DOI 10.1016/S0006-8993(00)02733-5
   Ohlemiller KK, 1999, AUDIOL NEURO-OTOL, V4, P229, DOI 10.1159/000013846
   Palma FR, 2020, ANTIOXID REDOX SIGN, V32, P701, DOI 10.1089/ars.2019.7962
   Pang SHM, 2021, NAT COMMUN, V12, DOI 10.1038/s41467-021-26834-3
   Poon IKH, 2019, J EXTRACELL VESICLES, V8, DOI 10.1080/20013078.2019.1608786
   Pouraghaei S, 2020, ACS BIOMATER SCI ENG, V6, P2263, DOI 10.1021/acsbiomaterials.9b01795
   Qu Y, 2022, ACTA BIOMATER
   Rhee SG, 2006, SCIENCE, V312, P1882, DOI 10.1126/science.1130481
   Rong X, 2021, FOOD FUNCT, V12, P1305, DOI [10.1039/d0fo02141e, 10.1039/D0FO02141E]
   Sarvar DP, 2016, ADV PHARM BULL, V6, P293, DOI 10.15171/apb.2016.041
   Schendzielorz P, 2017, CYTOTHERAPY, V19, P1197, DOI 10.1016/j.jcyt.2017.07.007
   Schieber M, 2014, CURR BIOL, V24, pR453, DOI 10.1016/j.cub.2014.03.034
   Shi X, 2022, SIGNAL TRANSDUCT TAR, V7, DOI 10.1038/s41392-022-00995-z
   Tan WJT, 2016, HISTOCHEM CELL BIOL, V146, P219, DOI 10.1007/s00418-016-1436-5
   Wang J, 2021, ACS NANO, V15, P14360, DOI 10.1021/acsnano.1c03517
   Winterbourn CC, 2008, FREE RADICAL BIO MED, V45, P549, DOI 10.1016/j.freeradbiomed.2008.05.004
   Wu F, 2020, REDOX BIOL, V29, DOI 10.1016/j.redox.2019.101406
   Xu YP, 2016, NEUROSCI LETT, V616, P19, DOI 10.1016/j.neulet.2016.01.016
   Yamashita D, 2005, NEUROSCIENCE, V134, P633, DOI 10.1016/j.neuroscience.2005.04.015
   Yang HD, 2022, FREE RADICAL BIO MED, V179, P229, DOI 10.1016/j.freeradbiomed.2021.11.020
   Yao J, 2019, FASEB J, V33, P1695, DOI 10.1096/fj.201800131RR
   Zhang LJ, 2022, NEURAL PLAST, V2022, DOI 10.1155/2022/5567174
   Zhang X, 2022, J EXTRACELL VESICLES, V11, DOI 10.1002/jev2.12240
   Zheng CX, 2021, J EXTRACELL VESICLES, V10, DOI 10.1002/jev2.12109
   Zhou YX, 2011, IMMUNOLOGY, V133, P133, DOI 10.1111/j.1365-2567.2011.03421.x
   Zhu GX, 2020, NEURAL PLAST, V2020, DOI 10.1155/2020/8823785
NR 72
TC 0
Z9 0
U1 4
U2 4
PU BMC
PI LONDON
PA CAMPUS, 4 CRINAN ST, LONDON N1 9XW, ENGLAND
EI 1757-6512
J9 STEM CELL RES THER
JI Stem Cell Res. Ther.
PD APR 15
PY 2023
VL 14
IS 1
AR 88
DI 10.1186/s13287-023-03314-7
PG 20
WC Cell & Tissue Engineering; Cell Biology; Medicine, Research &
   Experimental
WE Science Citation Index Expanded (SCI-EXPANDED)
SC Cell Biology; Research & Experimental Medicine
GA D6EY0
UT WOS:000969654500005
PM 37061707
OA gold, Green Published
DA 2023-08-27
ER

PT J
AU STEINHARDT, GF
   SALINASMADRIGAL, L
   DEMELLO, D
   FARBER, R
   PHILLIPS, B
   VOGLER, G
AF STEINHARDT, GF
   SALINASMADRIGAL, L
   DEMELLO, D
   FARBER, R
   PHILLIPS, B
   VOGLER, G
TI EXPERIMENTAL URETERAL OBSTRUCTION IN THE FETAL OPOSSUM - HISTOLOGIC
   ASSESSMENT
SO JOURNAL OF UROLOGY
LA English
DT Article
DE OPOSSUMS; URETERAL OBSTRUCTION
ID CONGENITAL HYDRONEPHROSIS INUTERO; NEWBORN RATS; WILMS-TUMOR; GROWTH;
   KIDNEY
AB In the North American opossum, Didelphis virginiana unilateral complete ureteral obstruction (ECO) and partial unilateral (EPO) ureteral obstruction were created during the early metanephric stage of kidney development in pups attached to the teat (approximately 4.5 cm. long, 20 days old). At 70 days of pouch life (full-term equivalent in the human) some completely obstructed ureters were unobstructed with reanastomosis to the bladder (RECO). Other pups underwent unilateral complete ureteral ligation at this full-term equivalent (70 days of pouch life) and constituted the late obstruction (LCO) group. Unoperated animals constituted the control (C) group. All animals were harvested when full grown (.7 to 2.2 kg.), and paraffin sections of the kidneys were stained with hematoxylin and eosin and Masson's Trichrome. In each a semiquantitative assessment of 24 histologic features was made and a digital score assigned. All experimental groups except EPO demonstrated significant epithelial and mesenchymal alterations. The changes can be broadly categorized as those secondary to obstruction of urine flow (dilation and cystic changes), those affecting epithelial differentiation in both cortex and medulla and those affecting mesenchymal differentiation.
   The EPO group demonstrated significantly fewer glomerular generations than did control kidneys. For all other characteristics studied, the EPO group did not differ from control. The ECO group demonstrated significant changes when compared with control for most parameters studied. Medullary dysplasia was more prominent in kidneys obstructed early. Reanastomosis (RECO) at a full-term equivalent did ameliorate the changes of medullary dysplasia and cortical atrophy. More collecting duct hyperplasia, cortical and medullary aplasia were present in the LCO group compared with the ECO. The LCO group also had less primitive duct formation and less medullary dysplasia than the ECO group. Renal blastema, previously unreported in experimental obstruction, was present in 41% of experimental kidneys.
C1 ST LOUIS UNIV,SCH MED,DEPT SURG,DIV UROL,ST LOUIS,MO 63103.
   ST LOUIS UNIV,SCH MED,DEPT PATHOL,ST LOUIS,MO 63104.
   ST LOUIS UNIV,SCH MED,DEPT COMPARAT MED,ST LOUIS,MO.
C3 Saint Louis University; Saint Louis University; Saint Louis University
RP STEINHARDT, GF (通讯作者)，CARDINAL GLENNON MEM HOSP CHILDREN,1465 S GRAND BLVD,ST LOUIS,MO 63104, USA.
CR BERNSTEIN J, 1971, Pediatric Clinics of North America, V18, P395
   CLAESSON G, 1989, J UROLOGY, V142, P1602, DOI 10.1016/S0022-5347(17)39177-2
   CRAVER R, 1986, J UROLOGY, V136, P305, DOI 10.1016/S0022-5347(17)44850-6
   CUTTS JH, 1978, BIOL NEONATE, V33, P264, DOI 10.1159/000241082
   GLICK PL, 1983, J PEDIATR SURG, V18, P681, DOI 10.1016/S0022-3468(83)80003-7
   HARRISON MR, 1982, J PEDIATR SURG, V17, P965, DOI 10.1016/S0022-3468(82)80476-4
   HUSER J, 1990, MODERN PATHOL, V3, P321
   JOSEPHSON S, 1980, INVEST UROL, V17, P478
   KLAHR S, 1991, SEMIN NEPHROL, V11, P156
   KRAUSE WJ, 1979, J ANAT, V129, P459
   KUSHNER L, 1992, J UROLOGY, V148, P555, DOI 10.1016/S0022-5347(17)36652-1
   MCVARY KT, 1989, J UROLOGY, V142, P646, DOI 10.1016/S0022-5347(17)38843-2
   NAGLE RB, 1976, LAB INVEST, V35, P18
   RANSLEY PG, 1990, J UROLOGY, V144, P584, DOI 10.1016/S0022-5347(17)39528-9
   STEINHARDT G, 1990, J UROLOGY, V144, P564, DOI 10.1016/S0022-5347(17)39524-1
   STEINHARDT GF, 1988, J PEDIATR SURG, V23, P1127, DOI 10.1016/S0022-3468(88)80327-0
   STEINHARDT GF, 1988, J UROLOGY, V140, P1316, DOI 10.1016/S0022-5347(17)42034-9
   STEINHARDT GF, 1990, DIALOG PEDIATR UROL, V13, P1
   STEINHARDT GF, 1989, PEDIATR UROL, V12, P7
   STEINHARDT GF, 1992, J MAT FETAL MED, V1, P277
NR 20
TC 18
Z9 19
U1 0
U2 1
PU WILLIAMS & WILKINS
PI BALTIMORE
PA 351 WEST CAMDEN ST, BALTIMORE, MD 21201-2436
SN 0022-5347
J9 J UROLOGY
JI J. Urol.
PD DEC
PY 1994
VL 152
IS 6
BP 2133
EP 2138
DI 10.1016/S0022-5347(17)32338-8
PN 1
PG 6
WC Urology & Nephrology
WE Science Citation Index Expanded (SCI-EXPANDED)
SC Urology & Nephrology
GA PR245
UT WOS:A1994PR24500076
PM 7966702
DA 2023-08-27
ER

PT J
AU Sun, D
   Feng, JM
   Dai, C
   Sun, L
   Jin, T
   Ma, JF
   Wang, LN
AF Sun, Dong
   Feng, Jiangmin
   Dai, Chun
   Sun, Li
   Jin, Tao
   Ma, Jianfei
   Wang, Lining
TI Role of peritubular capillary loss and hypoxia in progressive
   tubulointerstitial fibrosis in a rat model of aristolochic acid
   nephropathy
SO AMERICAN JOURNAL OF NEPHROLOGY
LA English
DT Article
DE peritubular capillary loss; aristolochic acid nephropathy;
   tubulointerstitial fibrosis
ID ENDOTHELIAL GROWTH-FACTOR; POTENTIAL ROLE; MOUSE KIDNEYS; UP-REGULATION;
   EXPRESSION; MYOFIBROBLASTS; INJURY; ANGIOGENESIS; TUMORS; ALPHA
AB Background/Aims: To investigate the effects of peritubular capillary (PTC) loss and hypoxia on the progression of tubulointerstitial fibrosis in a rat model of aristolochic acid nephropathy (AAN). Methods: Female Wistar rats received Caulis aristolochiae manshuriensis (CAM) decoction by gavage for 8 weeks, and were sacrificed at 8,12 and 16 weeks, respectively, after ad ministration. Blood urea nitrogen (BUN), serum creatinine (Scr) and urinary protein were monitored prior to sacrifice. PTC loss and tubulointerstitial hypoxia were assessed by CD34 immunostaining and hypoxia-inducible factor-alpha subunit 1 (HIF-1 alpha) expression, respectively. Myofibroblasts were assessed by alpha-smooth muscle actin (alpha-SMA) expression. The expression of angiogenic factor was assessed by vascular endothelial growth factor (VEGF). Results: AAN rats differed from controls by increased BUN, Scr and 24-hour urinary protein excretion rates. There was a progressive loss of PTCs in the AAN model, which was associated with the decreased expression of VEGF. A significant increase in nuclear localization of HIF-1 alpha was seen 16 weeks after treatment with CAM decoction in the context of severe tubulointerstitial damage. Multifocal tubulointerstitial fibrosis was seen in AAN rats at weeks 12 and 16, predominantly in the area of the outer stripe and outer medulla. No significant pathologic changes were found in control rats. Conclusion: Following the reduction of PTCs density and up-regulation of HIF-1 alpha, the tubulointerstitial fibrosis area increased. Ischemia and hypoxia are the important causes of severe tubulointerstitial fibrosis in AAN rats. Copyright (c) 2006 S. Karger AG, Basel.
C1 China Med Univ, Affiliated Hosp 1, Dept Nephrol, Shenyang 110001, Liaoning Prov, Peoples R China.
   Xuzhou Med Univ, Affiliated Hosp, Dept Nephrol, Xuzhou, Peoples R China.
C3 China Medical University; Xuzhou Medical University
RP Feng, JM (通讯作者)，China Med Univ, Affiliated Hosp 1, Dept Nephrol, 155 N Nanjing St, Shenyang 110001, Liaoning Prov, Peoples R China.
EM
RI Jiang, Tao/IWM-7503-2023
CR Bohle A, 1996, KIDNEY BLOOD PRESS R, V19, P191, DOI 10.1159/000174072
   Choi YJ, 2000, HUM PATHOL, V31, P1491, DOI 10.1053/hupa.2000.20373
   Cosyns JP, 2003, DRUG SAFETY, V26, P33, DOI 10.2165/00002018-200326010-00004
   DEPIERREUX M, 1994, AM J KIDNEY DIS, V24, P172, DOI 10.1016/S0272-6386(12)80178-8
   DIAMOND JR, 1995, AM J PATHOL, V146, P121
   EPSTEIN FH, 1994, ANN NY ACAD SCI, V718, P72
   Essawy M, 1997, NEPHROL DIAL TRANSPL, V12, P43, DOI 10.1093/ndt/12.1.43
   Ferrara N, 1999, KIDNEY INT, V56, P794, DOI 10.1046/j.1523-1755.1999.00610.x
   Fine LG, 1998, KIDNEY INT, pS74
   Gerber HP, 1998, J BIOL CHEM, V273, P30336, DOI 10.1074/jbc.273.46.30336
   GOUMENOS DS, 1994, NEPHROL DIAL TRANSPL, V9, P1418
   HARRIS DCH, 1988, AM J PHYSIOL, V254, pF267, DOI 10.1152/ajprenal.1988.254.2.F267
   Hopfl G, 2004, AM J PHYSIOL-REG I, V286, pR608, DOI 10.1152/ajpregu.00538.2003
   Jiang BH, 1996, AM J PHYSIOL-CELL PH, V271, pC1172, DOI 10.1152/ajpcell.1996.271.4.C1172
   Kairaitis LK, 2005, AM J PHYSIOL-RENAL, V288, pF198, DOI 10.1152/ajprenal.00244.2003
   Kang DH, 2001, J AM SOC NEPHROL, V12, P1434, DOI 10.1681/ASN.V1271434
   Kim YG, 2000, KIDNEY INT, V58, P2390, DOI 10.1046/j.1523-1755.2000.00422.x
   Moriyama T, 1998, KIDNEY INT, V54, P110, DOI 10.1046/j.1523-1755.1998.00964.x
   Moriyama T, 2001, KIDNEY INT, V59, P2095, DOI 10.1046/j.1523-1755.2001.00724.x
   Norman JT, 2000, KIDNEY INT, V58, P2351, DOI 10.1046/j.1523-1755.2000.00419.x
   Ohashi R, 2000, J AM SOC NEPHROL, V11, P47, DOI 10.1681/ASN.V11147
   Rodriguez-Pena A, 2002, HYPERTENSION, V40, P713, DOI 10.1161/01.HYP.0000037429.73954.27
   Safran M, 2003, J CLIN INVEST, V111, P779, DOI 10.1172/JCI200318181
   SERON D, 1990, NEPHROL DIAL TRANSPL, V5, P889, DOI 10.1093/ndt/5.10.889
   Suga SI, 2001, AM J PHYSIOL-RENAL, V281, pF620, DOI 10.1152/ajprenal.2001.281.4.F620
   Thomas SE, 1998, J AM SOC NEPHROL, V9, P231
   Utsunomiya Y, 1999, AM J KIDNEY DIS, V34, P902, DOI 10.1016/S0272-6386(99)70049-1
   Wojcikowski K, 2004, NEPHROLOGY, V9, P313, DOI 10.1111/j.1440-1797.2004.00310.x
   Yang JH, 2005, COMMUN THEOR PHYS, V44, P525, DOI 10.1088/6102/44/3/525
   Zagzag D, 2000, CANCER-AM CANCER SOC, V88, P2606, DOI 10.1002/1097-0142(20000601)88:11<2606::AID-CNCR25>3.3.CO;2-N
   Zhu Shaoming, 2005, Am J Nephrol, V25, P23, DOI 10.1159/000083655
   Zoja C, 1995, AM J KIDNEY DIS, V26, P934, DOI 10.1016/0272-6386(95)90058-6
NR 32
TC 48
Z9 59
U1 1
U2 13
PU KARGER
PI BASEL
PA ALLSCHWILERSTRASSE 10, CH-4009 BASEL, SWITZERLAND
SN 0250-8095
EI 1421-9670
J9 AM J NEPHROL
JI Am. J. Nephrol.
PY 2006
VL 26
IS 4
BP 363
EP 371
DI 10.1159/000094778
PG 9
WC Urology & Nephrology
WE Science Citation Index Expanded (SCI-EXPANDED)
SC Urology & Nephrology
GA 084SI
UT WOS:000240554300007
PM 16873992
DA 2023-08-27
ER

PT J
AU Jelaso, AM
   Acevedo, S
   Dang, T
   Lepere, A
   Ide, CF
AF Jelaso, AM
   Acevedo, S
   Dang, T
   Lepere, A
   Ide, CF
TI Interleukin-1 beta and its type 1 receptor are expressed in developing
   neural circuits in the frog, Xenopus laevis
SO JOURNAL OF COMPARATIVE NEUROLOGY
LA English
DT Article
DE cytokine; amphibian; Mauthner neuron; identified neurons; neuronal
   survival
ID CENTRAL-NERVOUS-SYSTEM; TUMOR-NECROSIS-FACTOR; PITUITARY-ADRENAL AXIS;
   PROGRAMMED CELL-DEATH; ROHON-BEARD NEURONS; GROWTH-FACTOR; SPINAL-CORD;
   NEUROTROPHIC FACTOR; CONVERTING-ENZYME; MESSENGER-RNA
AB The cytokine interleukin-1 beta (IL-1 beta) is an evolutionarily conserved molecule that was originally identified in the immune system. In addition to regulating peripheral immune responses, IL-1 beta plays an important role in mediating neural-immune interactions and regulating glial activities during healing and repair in the damaged nervous system. Active IL-1 beta is produced by interleukin-converting enzyme (ICE), a caspase thought to be involved in the induction of apoptosis. We report that, in the developing frog, Xenopus laevis, IL-1 beta and the IL-1 type 1 receptor proteins are coexpressed in specific neurons that comprise early sensory-motor circuits. IL-1 beta and IL-1 type 1 receptor proteins are colocalized in specific midbrain and hindbrain reticular cells, including Mauthner's neuron; specific cells in the trigeminal (fifth), lateral line (seventh), and vestibular (eighth) cranial ganglia; oculomotor neurons; and the primordial Purkinje cells of the lateral cerebellar auricle, In the spinal cord, Rohon-Beard sensory neurons, dorsal root ganglion cells, and primary motoneurons are immunopositive. Anteriorly, the olfactory pits, olfactory nerves, and olfactory bulbs are labeled, as are retinal cells, especially photoreceptor inner segments. With regard to the function of IL-1 beta during neural development. IL-1 beta and its type 1 receptor are present throughout the course of neural development in identifiable, long-lived neurons, such as Mauthner's neuron. These and other data suggest that IL-1 beta and its type 1 receptor may be involved in the maintenance of cell survival rather than induction of neuronal death. (C) 1998 Wiley-Liss, Inc.
C1 Tulane Univ, Dept Cell & Mol Biol, New Orleans, LA 70118 USA.
   Tulane Univ, Ctr Bioenvironm Res, New Orleans, LA 70118 USA.
   Xavier Univ, New Orleans, LA 70118 USA.
C3 Tulane University; Tulane University; Xavier University of Louisiana
RP Jelaso, AM (通讯作者)，Tulane Univ, Dept Cell & Mol Biol, New Orleans, LA 70118 USA.
EM
CR Alnemri ES, 1996, CELL, V87, P171, DOI 10.1016/S0092-8674(00)81334-3
   ALNEMRI ES, 1995, J BIOL CHEM, V270, P4312, DOI 10.1074/jbc.270.9.4312
   ALOISI F, 1994, J IMMUNOL, V152, P5022
   ARAUJO DM, 1995, INT J DEV NEUROSCI, V13, P201, DOI 10.1016/0736-5748(94)00072-B
   BANDTLOW CE, 1990, J CELL BIOL, V111, P1701, DOI 10.1083/jcb.111.4.1701
   BARTELMEZ GW, 1915, J COMP NEUROL, V194, P291
   BECK G, 1989, BIOESSAYS, V11, P62, DOI 10.1002/bies.950110206
   Benveniste EN, 1992, NEUROIMMUNOENDOCRINO, P106
   BEREZOVSKAYA O, 1995, INT J DEV NEUROSCI, V13, P285, DOI 10.1016/0736-5748(95)00013-7
   BESEDOVSKY HO, 1991, J STEROID BIOCHEM, V40, P613, DOI 10.1016/0960-0760(91)90284-C
   BLACK RA, 1988, J BIOL CHEM, V263, P9437
   BREDER CD, 1988, SCIENCE, V240, P321, DOI 10.1126/science.3258444
   BRENNEMAN DE, 1992, J NEUROCHEM, V58, P454, DOI 10.1111/j.1471-4159.1992.tb09743.x
   BRENNEMAN DE, 1995, INT J DEV NEUROSCI, V13, P187, DOI 10.1016/0736-5748(95)00014-8
   CLARKE JDW, 1984, J PHYSIOL-LONDON, V348, P511, DOI 10.1113/jphysiol.1984.sp015122
   Cohen N, 1991, PHYLOGENESIS IMMUNE, P241
   CohenCory S, 1996, DEV BIOL, V179, P102, DOI 10.1006/dbio.1996.0244
   CURTIS BM, 1990, J IMMUNOL, V144, P1295
   delRey A, 1996, BRAIN BEHAV IMMUN, V10, P235
   DINARELLO CA, 1991, BLOOD, V77, P1627
   DINARELLO CA, 1987, J IMMUNOL, V139, P1902
   EATON RC, 1988, J NEUROSCI, V8, P2758
   EATON RC, 1973, COPEIA, P673
   EATON RC, 1978, NEUROBIOLOGY MAUTHNE, P221
   ELLIS HM, 1986, CELL, V44, P817, DOI 10.1016/0092-8674(86)90004-8
   ERICSSON A, 1994, J NEUROSCI, V14, P897
   GADIENT RA, 1994, BRAIN RES, V637, P10, DOI 10.1016/0006-8993(94)91211-4
   GADIENT RA, 1990, NEUROSCI LETT, V117, P335, DOI 10.1016/0304-3940(90)90687-5
   GAGLIARDINI V, 1994, SCIENCE, V263, P826, DOI 10.1126/science.8303301
   GATTI S, 1993, BRAIN RES, V624, P291, DOI 10.1016/0006-8993(93)90090-A
   GIULIAN D, 1988, J NEUROSCI, V8, P709
   GIULIAN D, 1986, J EXP MED, V164, P594, DOI 10.1084/jem.164.2.594
   GIULIAN D, 1985, SCIENCE, V228, P497, DOI 10.1126/science.3872478
   GIULIAN D, 1988, J NEUROSCI, V8, P2485
   HAMA T, 1989, NEUROSCI LETT, V104, P340, DOI 10.1016/0304-3940(89)90600-9
   HUGHES A, 1957, J ANAT, V91, P323
   HUGHES TK, 1990, P NATL ACAD SCI USA, V87, P4426, DOI 10.1073/pnas.87.12.4426
   Jarskog LF, 1997, INT J DEV NEUROSCI, V15, P711, DOI 10.1016/S0736-5748(97)00029-4
   Jelaso AM, 1997, NEUROTOXICOLOGY, V18, P841
   Katz LC, 1996, SCIENCE, V274, P1133, DOI 10.1126/science.274.5290.1133
   KIMMEL CB, 1985, J COMP NEUROL, V233, P365, DOI 10.1002/cne.902330306
   KIMMEL CB, 1981, J COMP NEUROL, V198, P101, DOI 10.1002/cne.901980110
   KIMMEL CB, 1982, J COMP NEUROL, V205, P112, DOI 10.1002/cne.902050203
   KIMMEL CB, 1982, PROG BRAIN RES, V57, P1, DOI 10.1016/S0079-6123(08)64122-9
   KIMMEL CB, 1988, SIGNAL SENSE LOCAL G, P561
   KWON YW, 1995, J NEUROBIOL, V28, P35, DOI 10.1002/neu.480280105
   LAMBORGHINI JE, 1987, J COMP NEUROL, V264, P47, DOI 10.1002/cne.902640105
   LAMBORGHINI JE, 1980, J COMP NEUROL, V189, P323, DOI 10.1002/cne.901890208
   Leghissa Silvano, 1941, ARCH ZOOL ITAL, V29, P213
   LINDHOLM D, 1987, NATURE, V330, P658, DOI 10.1038/330658a0
   MARTINOU JC, 1990, DEV BRAIN RES, V52, P175, DOI 10.1016/0165-3806(90)90233-O
   MATTA SG, 1993, NEUROENDOCRINOLOGY, V57, P14, DOI 10.1159/000126336
   MERRILL JE, 1992, DEV NEUROSCI, V145, P1
   METCALFE WK, 1986, J COMP NEUROL, V251, P147, DOI 10.1002/cne.902510202
   METCALFE WK, 1985, J COMP NEUROL, V233, P377, DOI 10.1002/cne.902330307
   Michaelson MD, 1996, DEVELOPMENT, V122, P2661
   MILLIGAN CE, 1995, NEURON, V15, P385, DOI 10.1016/0896-6273(95)90042-X
   MINAMI M, 1992, J NEUROCHEM, V58, P390, DOI 10.1111/j.1471-4159.1992.tb09324.x
   MIURA M, 1993, CELL, V75, P653, DOI 10.1016/0092-8674(93)90486-A
   NEFF NT, 1993, J NEUROBIOL, V24, P1578, DOI 10.1002/neu.480241203
   NICHOLSON DW, 1995, NATURE, V376, P37, DOI 10.1038/376037a0
   Nieuwkoop P.D, 1994, NORMAL TABLE XENOPUS, V252, DOI 10.1201/9781003064565-29
   NORDLANDER RH, 1985, J COMP NEUROL, V231, P519, DOI 10.1002/cne.902310409
   OPPENHEIM RW, 1993, J NEUROBIOL, V24, P1065, DOI 10.1002/neu.480240806
   PATTERSON PH, 1993, CR ACAD SCI III-VIE, V316, P1150
   PATTERSON PH, 1993, CELL, V72, P123, DOI 10.1016/S0092-8674(05)80032-7
   POUSSET F, 1994, DEV BRAIN RES, V81, P143, DOI 10.1016/0165-3806(94)90078-7
   QUI LQ, 1994, DEV BIOL, V163, P516, DOI 10.1006/dbio.1994.1167
   RAY CA, 1992, CELL, V69, P597, DOI 10.1016/0092-8674(92)90223-Y
   ROTHWELL NJ, 1995, INT J DEV NEUROSCI, V13, P179, DOI 10.1016/0736-5748(95)00018-C
   SAWADA M, 1995, INT J DEV NEUROSCI, V13, P253, DOI 10.1016/0736-5748(94)00076-F
   Scripter JL, 1997, EXP NEUROL, V145, P329, DOI 10.1006/exnr.1997.6470
   SEI Y, 1995, NEUROIMMUNOMODULAT, V2, P121, DOI 10.1159/000096881
   SIGEL MM, 1986, VET IMMUNOL IMMUNOP, V12, P47, DOI 10.1016/0165-2427(86)90109-1
   SOFFE SR, 1984, J NEUROPHYSIOL, V51, P1257, DOI 10.1152/jn.1984.51.6.1257
   STEFANELLI A, 1951, Q REV BIOL, V26, P17, DOI 10.1086/397880
   Stoop R, 1996, J NEUROSCI, V16, P3256
   TABIRA T, 1995, INT J DEV NEUROSCI, V13, P241, DOI 10.1016/0736-5748(94)00020-4
   THORNBERRY NA, 1992, NATURE, V356, P768, DOI 10.1038/356768a0
   WANG T, 1995, J NEUROSCI, V15, P4796
   WARNER SJC, 1987, J EXP MED, V165, P1316, DOI 10.1084/jem.165.5.1316
   WATKINS D, 1987, IMMUNOLOGY, V62, P669
   WILSON KP, 1994, NATURE, V370, P270, DOI 10.1038/370270a0
   YUAN JY, 1993, CELL, V75, P641, DOI 10.1016/0092-8674(93)90485-9
   ZOTTOLI SJ, 1977, J EXP BIOL, V66, P243
NR 85
TC 19
Z9 20
U1 0
U2 2
PU WILEY-LISS
PI NEW YORK
PA DIV JOHN WILEY & SONS INC, 605 THIRD AVE, NEW YORK, NY 10158-0012 USA
SN 0021-9967
J9 J COMP NEUROL
JI J. Comp. Neurol.
PD MAY 4
PY 1998
VL 394
IS 2
BP 242
EP 251
DI 10.1002/(SICI)1096-9861(19980504)394:2<242::AID-CNE8>3.0.CO;2-2
PG 10
WC Neurosciences; Zoology
WE Science Citation Index Expanded (SCI-EXPANDED)
SC Neurosciences & Neurology; Zoology
GA ZE987
UT WOS:000072851500008
PM 9552129
DA 2023-08-27
ER

PT J
AU Petti, PL
   Larson, DA
   Kunwar, S
AF Petti, Paula L.
   Larson, David A.
   Kunwar, Sandeep
TI Use of hybrid shots in planning Perfexion Gamma Knife treatments for
   lesions close to critical structures
SO JOURNAL OF NEUROSURGERY
LA English
DT Article; Proceedings Paper
CT 14th International Meeting of the Leksell-Gamma-Knife-Society
CY MAY, 2008
CL Quebec City, CANADA
SP Leksell Gamma Knife Soc
DE Gamma Knife; hybrid shot; Perfexion; stereotactic radiosurgery
ID CONFORMITY; INDEX
AB Object. The authors investigated the use of different collimator values in different sectors (hybrid shots) when treating patients with lesions close to critical structures with the Perfexion model Gamma Knife.
   Methods. Twelve patients with various tumors (6 with a pituitary tumor, 3 with vestibular schwannoma, 2 with meningioma, and I with metastatic lesion) that were within 4 mm of the brainstem, optic nerve, pituitary stalk, or cochlea were considered. All patients were treated at the authors' institution between June 2007 and March 2008. The patients treatments were replanned in 2 different ways. In the first plan, hybrid shots were used such that the steepest dose gradient was aligned with the junction between the target and the critical structure(s). This was accomplished by placing low-value collimators in appropriate sectors. In the second plan, no hybrid shots were used. Sector blocking (either manual or dynamic) was required for all plans to reduce the critical structure doses to acceptable levels. Prescribed doses ranged from 12 to 30 Gy at the periphery of the target. The plans in each pair were designed to be equally conformal in terms of both target coverage (as measured by the Paddick conformity index) and critical structure sparing.
   Results. The average number of shots required was roughly the same using either planning technique (16.7 vs 16.6 shots with and without hybrids). However, for all patients, the number of blocked sectors required to protect critical areas was larger when hybrid shots were not used. On average. nearly twice as many blocked sectors (14.8 vs 7.0) were required for the plans that did not use hybrid shots. The number of high-value collimators used in each plan was also evaluated. For small targets (<= 1 cm(3)), for which 8 mm was considered a high value for the collimator. plans employing hybrids used an average of 2.3 times as many 8-mm sectors as did their nonhybrid counterparts (7.4 vs 3.2 sectors). For large targets (> 1 cm(3)), for which 16 mm was considered a high value for the collimator, hybrid plans used all average of 1.4 times as many 16-mm sectors as did the plans without hybrids (10.7 vs 7.7 sectors). Decreasing the number of blocked sectors and increasing the number of high-value collimator sectors led to use of shorter beam-on times. Beam-on times were 1-39% higher (average 17%) when hybrid shots were not allowed. The average beam-on time for plans with and without hybrid shots was 67.4 versus 78.4 minutes.
   Conclusions. The judicious use of hybrid shots in patients for whom the target is close to a critical structure is all efficient way to achieve conformal treatments while minimizing the beam-on time. The reduction in beam-on time with hybrid shots is attributed to a reduced use of blocked sectors and an increased number of high-value collimator sectors. (DOI: 10.3171/JNS/2008/109/12/S7)
C1 [Petti, Paula L.; Larson, David A.; Kunwar, Sandeep] Washington Hosp Healthcare Syst, Gamma Knife Ctr, Taylor McAdam Bell Neurosci Inst, Fremont, CA USA.
RP Petti, PL (通讯作者)，Washington Hosp, Gamma Knife Ctr, Fremont, CA 94538 USA.
EM
CR Lindquist C, 2007, NEUROSURGERY, V61, P130, DOI 10.1227/01.NEU.0000279989.86544.B6
   Paddick I, 2000, J NEUROSURG, V93, P219, DOI 10.3171/jns.2000.93.supplement_3.0219
   Paddick I, 2006, J NEUROSURG, V105, P194, DOI 10.3171/sup.2006.105.7.194
NR 3
TC 19
Z9 19
U1 0
U2 1
PU AMER ASSOC NEUROLOGICAL SURGEONS
PI ROLLING MEADOWS
PA 5550 MEADOWBROOK DRIVE, ROLLING MEADOWS, IL 60008 USA
SN 0022-3085
J9 J NEUROSURG
JI J. Neurosurg.
PD DEC
PY 2008
VL 109
SU S
BP 34
EP 40
DI 10.3171/JNS/2008/109/12/S7
PG 7
WC Clinical Neurology; Surgery
WE Science Citation Index Expanded (SCI-EXPANDED); Conference Proceedings Citation Index - Science (CPCI-S)
SC Neurosciences & Neurology; Surgery
GA 376TE
UT WOS:000261205100007
PM 19123886
DA 2023-08-27
ER

PT J
AU Tecimer, T
   Dlott, J
   Chuntharapai, A
   Martin, AW
   Peiper, SC
AF Tecimer, T
   Dlott, J
   Chuntharapai, A
   Martin, AW
   Peiper, SC
TI Expression of the chemokine receptor CXCR2 in normal and neoplastic
   neuroendocrine cells
SO ARCHIVES OF PATHOLOGY & LABORATORY MEDICINE
LA English
DT Article
ID NEURON-SPECIFIC ENOLASE; I CHROMOGRANIN-B; ENDOCRINE CELLS;
   INTERLEUKIN-8 FAMILY; TUMORS; LUNG; SYNAPTOPHYSIN; TISSUES;
   ANGIOGENESIS; NEUTROPHIL
AB Background-Chemokines effect their proinflammatory and growth regulatory roles through interaction with serpentine receptors. One such receptor, CXCR2, binds multiple CXC chemokines, including interleukin 8, GRO-alpha,, CRO-beta, GRO-gamma, and NAP-2, We have previously identified CXCR2 expression on myeloid cells, notably mature granulocytes, and projection neurons.
   Objective.-To determine the expression of CXCR2 by cells of the neuroendocrine system.
   Design,Archival specimens from normal neuroendocrine tissues and their malignant counterparts were analyzed by immunohistochemistry with monoclonal antibodies specific for CXCR1 and CXCR2.
   Results-Immunohistochemical analysis revealed high-level expression of CXCR2 by cells in the pituitary, adrenal medulla, pancreatic islets, thyroid C cells, scattered Kulchitsky cells in the bronchi, and counterpart neuroendocrine cells in the stomach, small bowel, colon, and appendix. Neuroendocrine neoplasms that demonstrated high-level CXCR2 expression included (1) primary carcinoids localized to the stomach, small bowel, colon, appendix, fallopian tube, ovary, and lung; (2) atypical carcinoids of the lung; (3) metastatic carcinoids; (4) pituitary adenomas; (5) pheochromocytomas; and (6) medullary carcinomas of the thyroid. Small cell lung carcinomas, large cell neuroendocrine carcinomas of the lung, small cell carcinoma of the cervix, Merkel cell carcinomas, neuroblastomas, and malignant melanomas lacked evidence of CXCR2 expression.
   Conclusions-The expression of CXCR2 by normal neuroendocrine cells and neoplastic counterparts that have retained phenotypic features of this differentiation program suggests that chemokines may play an important role in functions that are characteristic of this cell type. In addition, this raises the possibility that chemokines may modulate secretion of biologically active products of these cells and their neoplastic counterparts.
C1 Univ Louisville, James Graham Brown Canc Ctr, Henry Vogt Canc Res Inst, Louisville, KY 40202 USA.
   Univ Louisville, Dept Pathol, Louisville, KY 40202 USA.
   Genentech Inc, S San Francisco, CA 94080 USA.
C3 University of Louisville; University of Louisville; Roche Holding;
   Genentech
RP Tecimer, T (通讯作者)，Marmara Univ, Fac Med, Dept Pathol, TR-81190 Istanbul, Turkey.
CR Baggiolini M, 1998, NATURE, V392, P565, DOI 10.1038/33340
   Baggiolini M, 1997, ANNU REV IMMUNOL, V15, P675, DOI 10.1146/annurev.immunol.15.1.675
   BARNETT ML, 1993, BIOCHIM BIOPHYS ACTA, V1177, P275, DOI 10.1016/0167-4889(93)90123-7
   BERGH J, 1985, AM J CLIN PATHOL, V84, P1
   BONATO M, 1992, VIRCHOWS ARCH A, V420, P201, DOI 10.1007/BF01600272
   CACALANO G, 1994, SCIENCE, V265, P682, DOI 10.1126/science.8036519
   CHUNTHARAPAI A, 1994, J IMMUNOL, V153, P5682
   Collini P, 1998, VIRCHOWS ARCH, V432, P505, DOI 10.1007/s004280050198
   FUJITA T, 1979, ARCH HISTOL JPN S, V40, P1
   GOULD VE, 1983, PATHOL ANNU, V18, P287
   GOULD VE, 1987, AM J PATHOL, V126, P243
   GOULD VE, 1986, HUM PATHOL, V17, P979, DOI 10.1016/S0046-8177(86)80080-6
   HAIMOTO H, 1985, LAB INVEST, V52, P257
   HOGUEANGELETTI R, 1985, SCIENCE, V230, P89
   Horuk R, 1997, J IMMUNOL, V158, P2882
   JIN L, 1991, AM J PATHOL, V138, P961
   Kitadai Y, 1998, AM J PATHOL, V152, P93
   KOCH AE, 1992, SCIENCE, V258, P1798, DOI 10.1126/science.1281554
   KOIKE K, 1994, BIOCHEM BIOPH RES CO, V202, P161, DOI 10.1006/bbrc.1994.1907
   Kulke R, 1998, J INVEST DERMATOL, V110, P90, DOI 10.1046/j.1523-1747.1998.00074.x
   Kulke R, 1996, J INVEST DERMATOL, V106, P526, DOI 10.1111/1523-1747.ep12343916
   Lantuejoul S, 1998, AM J SURG PATHOL, V22, P1267, DOI 10.1097/00000478-199810000-00012
   LARSEN WJ, 1993, HUMAN EMBRYOLOGY, P65
   Lloyd Ricardo V., 1994, P561
   LLOYD RV, 1988, AM J PATHOL, V130, P296
   Luan J, 1997, J LEUKOCYTE BIOL, V62, P588, DOI 10.1002/jlb.62.5.588
   Luca M, 1997, AM J PATHOL, V151, P1105
   Luster AD, 1998, NEW ENGL J MED, V338, P436, DOI 10.1056/NEJM199802123380706
   MIETTINEN M, 1987, ARCH PATHOL LAB MED, V111, P813
   Moore BB, 1998, J INVEST MED, V46, P113
   NAGLE RB, 1986, AM J CLIN PATHOL, V85, P289, DOI 10.1093/ajcp/85.3.289
   NANNEY LB, 1995, AM J PATHOL, V147, P1248
   PAHLMAN S, 1986, LAB INVEST, V54, P554
   PEARSE AGE, 1979, FED PROC, V38, P2288
   PERASE AGE, 1974, PATHOL ANNU, V9, P27
   PERNICONE PJ, 1997, HISTOLOGY PATHOLOGIS, P1053
   PINTO A, 1989, CANCER-AM CANCER SOC, V64, P1266, DOI 10.1002/1097-0142(19890915)64:6<1266::AID-CNCR2820640617>3.0.CO;2-0
   SAWADA T, 1994, BIOCHEM BIOPH RES CO, V202, P155, DOI 10.1006/bbrc.1994.1906
   THOMAS P, 1987, AM J CLIN PATHOL, V88, P146, DOI 10.1093/ajcp/88.2.146
   TRAVIS WD, 1991, AM J SURG PATHOL, V15, P529, DOI 10.1097/00000478-199106000-00003
   VINORES SA, 1984, ARCH PATHOL LAB MED, V108, P536
   WIEDENMANN B, 1988, GASTROENTEROLOGY, V95, P1364, DOI 10.1016/0016-5085(88)90374-5
   WOOD JD, 1978, NATURE, V276, P834
NR 43
TC 31
Z9 34
U1 0
U2 2
PU COLLEGE AMER PATHOLOGISTS
PI NORTHFIELD
PA C/O KIMBERLY GACKI, 325 WAUKEGAN RD, NORTHFIELD, IL 60093-2750 USA
SN 0003-9985
J9 ARCH PATHOL LAB MED
JI Arch. Pathol. Lab. Med.
PD APR
PY 2000
VL 124
IS 4
BP 520
EP 525
PG 6
WC Medical Laboratory Technology; Medicine, Research & Experimental;
   Pathology
WE Science Citation Index Expanded (SCI-EXPANDED)
SC Medical Laboratory Technology; Research & Experimental Medicine;
   Pathology
GA 304XG
UT WOS:000086508900009
PM 10747307
DA 2023-08-27
ER

PT J
AU SCHMID, KW
   HELPAP, B
   TOTSCH, M
   KIRCHMAIR, R
   DOCKHORNDWORNICZAK, B
   BOCKER, W
   FISCHERCOLBRIE, R
AF SCHMID, KW
   HELPAP, B
   TOTSCH, M
   KIRCHMAIR, R
   DOCKHORNDWORNICZAK, B
   BOCKER, W
   FISCHERCOLBRIE, R
TI IMMUNOHISTOCHEMICAL LOCALIZATION OF CHROMOGRANIN-A AND CHROMOGRANIN-B
   AND SECRETOGRANIN-II IN NORMAL, HYPERPLASTIC AND NEOPLASTIC PROSTATE
SO HISTOPATHOLOGY
LA English
DT Article
DE CHROMOGRANIN A; CHROMOGRANIN B; SECRETOGRANIN II; SECRETONEURIN;
   PROSTATE CARCINOMA; NEUROENDOCRINE; DIFFERENTIATION
ID NEURO-ENDOCRINE CELLS; HUMAN ADRENAL-MEDULLA; NEUROENDOCRINE
   DIFFERENTIATION; CHROMAFFIN GRANULES; ACID-PHOSPHATASE; CARCINOMA;
   PEPTIDE; TUMORS; SEROTONIN; TISSUE
AB Routinely processed normal, hyperplastic and neoplastic prostatic tissue was immunohistochemically investigated with antibodies against chromogranin A and B and secretogranin II. In normal and hyperplastic prostates all three peptides were immunolocalized in scattered neuroendocrine cells situated within the glandular epithelium. In 17 prostatic carcinomas with pronounced neuroendocrine differentiation and in a case of prostatic carcinoid, chromogranin B was the major component whereas chromogranin A and secretogranin II were virtually absent in poorly differentiated (grade III) tumours. Neuroendocrine differentiation in prostatic cancer is most likely to be associated with a poor clinical outcome; thus, chromogranin B appears to be a useful marker in the histopathological diagnosis of these neoplasms.
C1 STADT KRANKENHAUS SINGEN, SINGEN, GERMANY.
   UNIV INNSBRUCK, DEPT PHARMACOL, INNSBRUCK, AUSTRIA.
C3 University of Innsbruck
RP SCHMID, KW (通讯作者)，UNIV MUNSTER, DEPT PATHOL, DOMAGKSTR 17, D-48149 MUNSTER, GERMANY.
CR ABRAHAMSSON PA, 1989, PROSTATE, V14, P71, DOI 10.1002/pros.2990140109
   ABRAHAMSSON PA, 1987, PATHOL RES PRACT, V182, P298, DOI 10.1016/S0344-0338(87)80065-1
   ABRAHAMSSON PA, 1986, PATHOL RES PRACT, V181, P675, DOI 10.1016/S0344-0338(86)80043-7
   ABRAHAMSSON PA, 1989, PATHOL RES PRACT, V185, P373, DOI 10.1016/S0344-0338(89)80016-0
   ALMAGRO UA, 1985, CANCER, V55, P608, DOI 10.1002/1097-0142(19850201)55:3<608::AID-CNCR2820550322>3.0.CO;2-S
   APRIKIAN AG, 1993, CANCER, V71, P3952, DOI 10.1002/1097-0142(19930615)71:12<3952::AID-CNCR2820711226>3.0.CO;2-X
   AUMULLER G, 1972, ACTA HISTOCHEM, V42, P29
   AZUMI N, 1984, AM J SURG PATHOL, V8, P545, DOI 10.1097/00000478-198407000-00007
   AZUMI N, 1991, AM J SURG PATHOL, V15, P785, DOI 10.1097/00000478-199108000-00009
   AZZOPARDI JG, 1971, J PATHOL, V104, P247, DOI 10.1002/path.1711040406
   BISHOP AE, 1989, HISTOCHEMISTRY, V90, P475, DOI 10.1007/BF00494359
   COHEN RJ, 1992, ARCH PATHOL LAB MED, V116, P65
   DAVIS NS, 1989, J UROLOGY, V142, P884, DOI 10.1016/S0022-5347(17)38936-X
   DISANTAGNESE PA, 1987, HUM PATHOL, V18, P849
   DISANTAGNESE PA, 1992, HUM PATHOL, V23, P287
   DISANTAGNESE PA, 1985, ARCH PATHOL LAB MED, V109, P607
   DISANTAGNESE PA, 1988, ARCH PATHOL LAB MED, V112, P1097
   DISANTAGNESE PA, 1992, CANCER-AM CANCER SOC, V70, P254, DOI 10.1002/1097-0142(19920701)70:1+<254::AID-CNCR2820701312>3.0.CO;2-E
   FALKMER S, 1989, ARCH HISTOL CYTOL, V52, P225, DOI 10.1679/aohc.52.Suppl_225
   FETISSOF F, 1983, VIRCHOWS ARCH B, V42, P53, DOI 10.1007/BF02890370
   FISCHERCOLBRIE R, 1990, J BIOL CHEM, V265, P9208
   FISCHERCOLBRIE R, 1987, ANN NY ACAD SCI, V493, P120, DOI 10.1111/j.1749-6632.1987.tb27189.x
   FISCHERCOLBRIE R, 1986, J NEUROCHEM, V47, P318
   FISCHERCOLBRIE R, 1985, J NEUROCHEM, V44, P1854, DOI 10.1111/j.1471-4159.1985.tb07179.x
   FISCHERCOLBRIE R, 1987, J NEUROCHEM, V48, P262, DOI 10.1111/j.1471-4159.1987.tb13157.x
   GERDES HH, 1988, NUCLEIC ACIDS RES, V16, P11811, DOI 10.1093/nar/16.24.11811
   HAGN C, 1986, LAB INVEST, V55, P405
   HELLE KB, 1966, MOL PHARMACOL, V2, P298
   HELPAP B, 1990, PATHOLOGE, V11, P18
   HELPAP B, 1989, PATHOLOGIE ABLEITEND
   JAZZAZ BA, 1974, J PATHOL, V112, P189
   KADMON D, 1991, J UROLOGY, V146, P358, DOI 10.1016/S0022-5347(17)37793-5
   KIRCHMAIR R, 1993, NEUROSCIENCE, V53, P359, DOI 10.1016/0306-4522(93)90200-Y
   OESTERLING JE, 1992, J UROLOGY, V147, P804, DOI 10.1016/S0022-5347(17)37390-1
   Pagani A, 1992, Diagn Mol Pathol, V1, P165, DOI 10.1097/00019606-199209000-00002
   RINDI G, 1986, HISTOCHEMISTRY, V85, P19, DOI 10.1007/BF00508649
   RODE J, 1990, J CLIN PATHOL, V43, P13, DOI 10.1136/jcp.43.1.13
   ROSA P, 1981, MOL CELL ENDOCRINOL, V24, P181, DOI 10.1016/0303-7207(81)90058-7
   Schmid K W, 1987, Verh Dtsch Ges Pathol, V71, P311
   SCHMID KW, 1989, HISTOCHEM J, V21, P365, DOI 10.1007/BF01798500
   SCHMID KW, 1991, AM J SURG PATHOL, V15, P1072, DOI 10.1097/00000478-199111000-00006
   SCHMID KW, 1993, HISTOPATHOLOGY, V22, P527, DOI 10.1111/j.1365-2559.1993.tb00172.x
   SCHMID KW, 1992, HISTOPATHOLOGY, V21, P225, DOI 10.1111/j.1365-2559.1992.tb00380.x
   SCHMID KW, 1987, AM J SURG PATHOL, V11, P551, DOI 10.1097/00000478-198707000-00007
   SCHMID KW, 1991, VIRCHOWS ARCH A, V418, P295, DOI 10.1007/BF01600157
   SCHMID KW, 1994, IN PRESS HISTOCHEMIS
   SCHMID KW, 1991, VERH DTSCH GES PATHO, V75, P519
   SCHOBER M, 1987, LAB INVEST, V57, P385
   TOTSCH M, 1992, HUM PATHOL, V23, P312, DOI 10.1016/0046-8177(92)90113-H
   TURBATHERRERA EA, 1988, ARCH PATHOL LAB MED, V112, P1100
   WEAVER MG, 1992, AM J SURG PATHOL, V16, P1013, DOI 10.1097/00000478-199210000-00013
   WEILER R, 1988, AM J SURG PATHOL, V12, P877, DOI 10.1097/00000478-198811000-00008
   WEILER R, 1987, VIRCHOWS ARCH A, V412, P103, DOI 10.1007/BF00716181
   WIEDENMANN B, 1988, GASTROENTEROLOGY, V95, P1364, DOI 10.1016/0016-5085(88)90374-5
   WINKLER H, 1986, NEUROSCIENCE, V18, P261, DOI 10.1016/0306-4522(86)90154-5
   WINKLER H, 1992, NEUROSCIENCE, V49, P497, DOI 10.1016/0306-4522(92)90222-N
NR 56
TC 42
Z9 43
U1 0
U2 0
PU WILEY
PI HOBOKEN
PA 111 RIVER ST, HOBOKEN 07030-5774, NJ USA
SN 0309-0167
EI 1365-2559
J9 HISTOPATHOLOGY
JI Histopathology
PD MAR
PY 1994
VL 24
IS 3
BP 233
EP 239
DI 10.1111/j.1365-2559.1994.tb00515.x
PG 7
WC Cell Biology; Pathology
WE Science Citation Index Expanded (SCI-EXPANDED)
SC Cell Biology; Pathology
GA NB187
UT WOS:A1994NB18700005
PM 7515371
DA 2023-08-27
ER

PT J
AU Fletcher, AL
   Seach, N
   Reiseger, JJ
   Lowen, TE
   Hammett, MV
   Scott, HS
   Boyd, RL
AF Fletcher, Anne L.
   Seach, Natalie
   Reiseger, Jessica J.
   Lowen, Tamara E.
   Hammett, Maree V.
   Scott, Hamish S.
   Boyd, Richard L.
TI Reduced Thymic Aire Expression and Abnormal NF-kappa B2 Signaling in a
   Model of Systemic Autoimmunity
SO JOURNAL OF IMMUNOLOGY
LA English
DT Article
ID NEW-ZEALAND MICE; STEM-CELL TRANSPLANTATION; MESSENGER-RNA EXPRESSION;
   TUMOR-NECROSIS-FACTOR; EPITHELIAL-CELLS; NZB MICE; BLACK MICE; LYMPHOID
   NEOGENESIS; ENDOTHELIAL-CELLS; DENDRITIC CELLS
AB The thymic stromal niche normally directs the production and export of a self-tolerant T cell repertoire. Many models of spontaneous autoimmunity, however, develop thymic architectural abnormalities before disease onset. Although this is suspected to affect central tolerance induction, creating an autoimmune predisposition, in-depth analysis of the microenvironment within these thymi is lacking, such that the mechanisms and likely direct effects on the T cell repertoire are unknown or speculative. Here we show that NZB mice, the first described model for systemic autoimmunity, demonstrate a complex thymic phenotype, including a lack of the autoimmune regulator (Aire), early defects in thymic epithelial cell (TEC) expansion, and evidence for altered NF-kappa B2 signaling. Analysis of medullary TEC revealed a numerical loss of the Aire-expressing MHC class II(high) (mTEC-high) subset as well reduced Aire protein and mRNA per cell. ROB expression was also reduced, while chemokines CCL19 and CCL21 were increased. Unexpectedly, the proportion of cortex and medulla in the NZB mice was normal from 36 wk, despite worsening architectural abnormalities. These data show that the NZB defect is more complex than previously appreciated, segregating into early numerical TEC deficiencies that correct with age, late degeneration of the niche architecture that does not affect TEC number, and a persistent reduction in Aire and RelB expression per cell acquired upon mTEC-high differentiation. The Journal of Immunology, 2009, 182: 2690-2699.
C1 [Fletcher, Anne L.; Seach, Natalie; Reiseger, Jessica J.; Lowen, Tamara E.; Hammett, Maree V.; Boyd, Richard L.] Monash Univ, Monash Immunol & Stem Cell Labs, Immune Regenerat Lab, Clayton, Vic 3800, Australia.
   [Scott, Hamish S.] Univ Adelaide, Sch Med, Adelaide, SA, Australia.
   [Scott, Hamish S.] Inst Med & Vet Sci, Div Mol Pathol, Adelaide, SA 5000, Australia.
   [Scott, Hamish S.] Hanson Inst Adelaide, Adelaide, SA, Australia.
C3 Monash University; University of Adelaide; Institute Medical &
   Veterinary Science Australia; Hanson Institute
RP Boyd, RL (通讯作者)，Monash Univ, Monash Immunol & Stem Cell Labs, Immune Regenerat Lab, Wellington Rd, Clayton, Vic 3800, Australia.
EM
RI Scott, Hamish/B-2122-2009; Fletcher, Anne L/A-1590-2011
OI Scott, Hamish/0000-0002-5813-631X; Fletcher, Anne/0000-0003-1534-1142;
   Lister, Natalie/0000-0001-9223-5875
CR AALTONEN J, 1994, NAT GENET, V8, P83, DOI 10.1038/ng0994-83
   Anderson G, 2001, NAT REV IMMUNOL, V1, P31, DOI 10.1038/35095500
   Anderson MS, 2005, IMMUNITY, V23, P227, DOI 10.1016/j.immuni.2005.07.005
   Anderson MS, 2002, SCIENCE, V298, P1395, DOI 10.1126/science.1075958
   Boehm T, 2003, J EXP MED, V198, P757, DOI 10.1084/jem.20030794
   BORCHERS A, 2000, SEMIN ARTHRITIS RHEU, V10, P111
   BURKLY L, 1995, NATURE, V373, P531, DOI 10.1038/373531a0
   CLAYTON PE, 2005, BROOKS CLIN PEDIAT E
   COHENKAMINSKY S, 1993, ANN NY ACAD SCI, V681, P97, DOI 10.1111/j.1749-6632.1993.tb22873.x
   Colomb E, 1996, DIABETES, V45, P1535, DOI 10.2337/diabetes.45.11.1535
   Drayton DL, 2003, J EXP MED, V197, P1153, DOI 10.1084/jem.20021761
   Fan L, 2000, J IMMUNOL, V164, P3955, DOI 10.4049/jimmunol.164.8.3955
   FITZPATRICK FTA, 1985, J ENDOCRINOL, V106, pR17, DOI 10.1677/joe.0.106R017
   Font J, 2004, SEMIN ARTHRITIS RHEU, V33, P217, DOI 10.1053/S0049-0172(03)00133-1
   Gabler J, 2007, EUR J IMMUNOL, V37, P3363, DOI 10.1002/eji.200737131
   GERSHWIN ME, 1978, J IMMUNOL, V120, P971
   Gillard GO, 2007, J IMMUNOL, V178, P3007, DOI 10.4049/jimmunol.178.5.3007
   Goldberg GL, 2007, J IMMUNOL, V178, P7473, DOI 10.4049/jimmunol.178.11.7473
   Goldberg GL, 2005, TRANSPLANTATION, V80, P1604, DOI 10.1097/01.tp.0000183962.64777.da
   Gray D, 2007, J EXP MED, V204, P2521, DOI 10.1084/jem.20070795
   Gray DHD, 2008, J IMMUNOL METHODS, V329, P56, DOI 10.1016/j.jim.2007.09.010
   Gray DHD, 2007, J IMMUNOL, V178, P4956, DOI 10.4049/jimmunol.178.8.4956
   Gray DHD, 2006, BLOOD, V108, P3777, DOI 10.1182/blood-2006-02-004531
   GREENSTEIN BD, 1986, J ENDOCRINOL, V110, P417, DOI 10.1677/joe.0.1100417
   Hashimoto Y, 2002, J IMMUNOL, V168, P81, DOI 10.4049/jimmunol.168.1.81
   Hashimoto Y, 2000, J IMMUNOL, V164, P1569, DOI 10.4049/jimmunol.164.3.1569
   Heino M, 2000, EUR J IMMUNOL, V30, P1884, DOI 10.1002/1521-4141(200007)30:7<1884::AID-IMMU1884>3.0.CO;2-P
   Heng TSP, 2005, J IMMUNOL, V175, P2982, DOI 10.4049/jimmunol.175.5.2982
   Hubert FX, 2008, J IMMUNOL, V180, P3824, DOI 10.4049/jimmunol.180.6.3824
   ISHII N, 1989, IMMUNOLOGY, V68, P215
   Ishikawa S, 2007, FRONT BIOSCI-LANDMRK, V12, P1790, DOI 10.2741/2188
   Ito T, 2004, J IMMUNOL, V172, P3628, DOI 10.4049/jimmunol.172.6.3628
   Jenkinson WE, 2003, J EXP MED, V198, P325, DOI 10.1084/jem.20022135
   JYONOUCHI H, 1982, J EXP MED, V155, P1665, DOI 10.1084/jem.155.6.1665
   Kajiura F, 2004, J IMMUNOL, V172, P2067, DOI 10.4049/jimmunol.172.4.2067
   Kinoshita D, 2006, J IMMUNOL, V176, P3995, DOI 10.4049/jimmunol.176.7.3995
   Kratz A, 1996, J EXP MED, V183, P1461, DOI 10.1084/jem.183.4.1461
   Kwan J, 2004, J IMMUNOL, V172, P3999, DOI 10.4049/jimmunol.172.7.3999
   Liston A, 2004, J EXP MED, V200, P1015, DOI 10.1084/jem.20040581
   Livak KJ, 2001, METHODS, V25, P402, DOI 10.1006/meth.2001.1262
   MILICH DR, 1980, SEMIN ARTHRITIS RHEU, V10, P111, DOI 10.1016/0049-0172(80)90004-9
   MonsoHinard C, 1997, IMMUNOLOGY, V92, P53, DOI 10.1046/j.1365-2567.1997.00297.x
   Ngo VN, 1999, J EXP MED, V189, P403, DOI 10.1084/jem.189.2.403
   Olsen NJ, 2001, ENDOCRINOLOGY, V142, P1278, DOI 10.1210/en.142.3.1278
   Page G, 2002, J IMMUNOL, V168, P5333, DOI 10.4049/jimmunol.168.10.5333
   QUERE P, 1985, EUR J IMMUNOL, V15, P112, DOI 10.1002/eji.1830150203
   Rossi SW, 2007, J EXP MED, V204, P1267, DOI 10.1084/jem.20062497
   Roy V, 2005, J IMMUNOL, V175, P7363, DOI 10.4049/jimmunol.175.11.7363
   Savinov AY, 2003, J EXP MED, V197, P643, DOI 10.1084/jem.20021378
   Scaglione BJ, 2007, BRIT J HAEMATOL, V139, P645, DOI 10.1111/j.1365-2141.2007.06851.x
   Schneider K, 2004, IMMUNOL REV, V202, P49, DOI 10.1111/j.0105-2896.2004.00206.x
   Seach N, 2008, J IMMUNOL, V180, P5384, DOI 10.4049/jimmunol.180.8.5384
   Sediva A, 2002, J PEDIATR ENDOCR MET, V15, P1491
   Sempowski GD, 2000, J IMMUNOL, V164, P2180, DOI 10.4049/jimmunol.164.4.2180
   STENIUS U, 1990, CARCINOGENESIS, V11, P69, DOI 10.1093/carcin/11.1.69
   Strobel P, 2007, J PATHOL, V211, P563, DOI 10.1002/path.2141
   Subrata LS, 2005, LIVER INT, V25, P633, DOI 10.1111/j.1478-3231.2005.01080.x
   Sutherland JS, 2008, CLIN CANCER RES, V14, P1138, DOI 10.1158/1078-0432.CCR-07-1784
   Sutherland JS, 2005, J IMMUNOL, V175, P2741, DOI 10.4049/jimmunol.175.4.2741
   Taguchi N, 1999, J AUTOIMMUN, V13, P393, DOI 10.1006/jaut.1999.0332
   Taguchi N, 2001, J AUTOIMMUN, V16, P401, DOI 10.1006/jaut.2001.0511
   Taguchi N, 2001, J AUTOIMMUN, V16, P393, DOI 10.1006/jaut.2001.0515
   TAKEOKA Y, 1995, CLIN IMMUNOL IMMUNOP, V76, P297, DOI 10.1006/clin.1995.1129
   TAKEOKA Y, 1995, J AUTOIMMUN, V8, P145, DOI 10.1006/jaut.1995.0012
   Takeoka Y, 1997, DEV IMMUNOL, V5, P79, DOI 10.1155/1997/69270
   Takeoka Y, 1999, CLIN IMMUNOL, V90, P388, DOI 10.1006/clim.1998.4655
   Timmer TCG, 2007, ARTHRITIS RHEUM-US, V56, P2492, DOI 10.1002/art.22748
   Ueno T, 2002, IMMUNITY, V16, P205, DOI 10.1016/S1074-7613(02)00267-4
   Valero R, 2002, J IMMUNOL, V169, P185, DOI 10.4049/jimmunol.169.1.185
   WANG Y, 1995, J NEUROIMMUNOL, V61, P231, DOI 10.1016/0165-5728(95)00096-K
   Ware C F, 1995, Curr Top Microbiol Immunol, V198, P175
   WATANABE Y, 1993, J IMMUNOL, V150, P4702
   YOSHIDA S, 1990, SEMIN ARTHRITIS RHEU, V19, P224, DOI 10.1016/0049-0172(90)90002-W
   Zhang BC, 2006, J BIOL CHEM, V281, P38617, DOI 10.1074/jbc.M606705200
   Zhang J, 1997, INVEST OPHTH VIS SCI, V38, P1848
   Zhu MZ, 2006, J CLIN INVEST, V116, P2964, DOI 10.1172/JCI28326
   Zuklys S, 2000, J IMMUNOL, V165, P1976, DOI 10.4049/jimmunol.165.4.1976
   2001, ARTHRITIS RHEUM, V44, P408
NR 78
TC 21
Z9 23
U1 0
U2 2
PU AMER ASSOC IMMUNOLOGISTS
PI BETHESDA
PA 9650 ROCKVILLE PIKE, BETHESDA, MD 20814 USA
SN 0022-1767
J9 J IMMUNOL
JI J. Immunol.
PD MAR 1
PY 2009
VL 182
IS 5
BP 2690
EP 2699
DI 10.4049/jimmunol.0801752
PG 10
WC Immunology
WE Science Citation Index Expanded (SCI-EXPANDED)
SC Immunology
GA 411MG
UT WOS:000263653100018
PM 19234163
OA Bronze
DA 2023-08-27
ER

PT J
AU McGregor, D
AF McGregor, Douglas
TI Hydroquinone: An evaluation of the human risks from its carcinogenic and
   mutagenic properties
SO CRITICAL REVIEWS IN TOXICOLOGY
LA English
DT Review
DE chronic progressive nephropathy; human kidney; hydroquinone; IPCS
   framework; metabolism; Mutagens; rodent carcinogens
ID CHRONIC PROGRESSIVE NEPHROPATHY; SISTER-CHROMATID EXCHANGES;
   SPRAGUE-DAWLEY RATS; PERIPHERAL-BLOOD LYMPHOCYTES;
   NATIONAL-TOXICOLOGY-PROGRAM; CULTURED HUMAN-LYMPHOCYTES; SUSPECTED
   SPINDLE POISONS; OXIDATIVE DNA-DAMAGE; BONE-MARROW CELLS; BENZENE
   METABOLITES
AB The toxicology of hydroquinone has been reviewed on a number of previous occasions. This review targets its potential for carcinogenicity and possible modes of carcinogenic action. The evaluation made by IARC (1999) of its carcinogenic risk to humans was that hydroquinone is not classifiable as to its carcinogenicity to humans (Group 3). This evaluation was based on inadequate evidence in humans and limited evidence in experimental animals. The epidemiological information comes from four cohort studies involving occupational exposures. A cohort of lithographers, some of whom had worked with hydroquinone, had an excess of malignant melanoma based on five cases, but only two of the cases had reported exposure to hydroquinone. In a study of photographic processors the number of exposed individuals was uncertain and the numbers of cases of individual cancer sites were small. In view of the statistical power limitations of these studies for individual diagnostic categories of cancers, they are not considered to be informative with regard to the carcinogenicity of hydroquinone. A cohort of workers with definite and lengthy exposure to hydroquinone, during either its manufacture or its use, had low cancer rates compared with two comparison populations; the reason for the lower than expected rates is unclear. In a motion picture film processing cohort there were significant excess malignancies of the respiratory system among workers engaged in developing, where there was exposure to hydroquinone as well as other chemicals. There was no information on tobacco smoking habits and no dose-response relationship. Hydroquinone has been shown reproducibly to induce benign neoplasms in the kidneys of male F344 rats dosed orally either by gavage (25 and 50 mg/kg body weight) or diet (0.8%). The gavage study has been evaluated in considerable detail. This evaluation showed that all renal tubule adenomas and all cases of renal tubule atypical hyperplasia occurred in areas of severe or end-stage chronic progressive nephropathy and that the neoplasms were not otherwise confined to any particular part of the kidney. It is likely that the mode of carcinogenic action of hydroquinone is exacerbation of this natural disease process. Hydroquinone is mutagenic in vitro and in vivo, having caused genotoxicity or chromosomal aberrations in rodent bone-marrowcells. At least a portion, if not all, of the chromosomal effects are caused by interference by hydroquinone or its metabolites with chromosomal segregation, probably due to interaction with mitotic spindle proteins. However, the dose routes used to demonstrate these effects in almost all of the studies in vivo were intraperitoneal or subcutaneous injection, which were considered inappropriate. There were five studies by the oral route. These included a mouse bone-marrow cell micronucleus test in which a weak, marginally positive response was obtained following a single oral dose of 80 mg/kg body weight. The remaining oral route studies all showed no significant effect. They included a mouse bone-marrow cell micronucleus test in which there was no genotoxic activity after exposure to a diet containing 0.8% hydroquinone for 6 days; two 32P-post-labeling assays, one with targets of Zymbal gland, liver, and spleen in Sprague-Dawley rats, the other with the kidney as target in F344 rats; and the last oral assay was for 8-hydroxydeoxyguanosine adducts in F344 rat kidney DNA. Thus, the evidence (and the database) for any genotoxic effect in vivo is sparse and none has been observed in kidney.
   While glutathione conjugaes could be responsible for the tumor induction, careful histology seems to show that the most actively toxic of several glutathione compounds tested, 2,3,5-triglutathion-S-yl hydroquinone, targets a very specific region of the kidney, the outer stripe of the outer medulla (OSOM), whereas hydroquinone-associated adenomas are more randomly distributed and occur in the cortex as well as the medulla. A nongenotoxic mode of action that involves exacerbation of a spontaneously occurring rodent renal disease, chronic progressive nephropathy (CPN), is proposed and evaluated. This disease is particularly prominent in male rats and the evidence is consistent with an absence of any human counterpart; therefore, the increased incidence of renal tubule adenomas in hydroquinone-dosed male rats is without human consequence.
C1 Tox Evaluat Consultants, Aberdour KY3 0TU, Scotland.
RP McGregor, D (通讯作者)，Tox Evaluat Consultants, 38 Shore Rd, Aberdour KY3 0TU, Scotland.
EM
CR ADLER ID, 1990, MUTAT RES, V234, P115, DOI 10.1016/0165-1161(90)90002-6
   ADLER ID, 1991, MUTAGENESIS, V6, P47, DOI 10.1093/mutage/6.1.47
   Anderson D, 1995, ENVIRON MOL MUTAGEN, V26, P305, DOI 10.1002/em.2850260406
   [Anonymous], IARC MON EV CARC R 2
   ANTOCCIA A, 1991, MUTAGENESIS, V6, P319, DOI 10.1093/mutage/6.4.319
   ANVER MR, 1982, EXP AGING RES, V8, P1, DOI 10.1080/03610738208258390
   Baltherwick NR, 1937, ARCH INTERN MED, V59, P572, DOI 10.1001/archinte.1937.00170200014002
   BARALE R, 1990, MUTAT RES, V244, P15, DOI 10.1016/0165-7992(90)90101-O
   BARBER ED, 1995, TOXICOL LETT, V80, P167, DOI 10.1016/0378-4274(95)03393-Y
   BARTH JL, 1979, TOXICOL PATHOL, V7, P1
   BAYLIS C, 1994, J CLIN INVEST, V94, P1823, DOI 10.1172/JCI117531
   Boatman RJ, 2000, CHEM RES TOXICOL, V13, P861, DOI 10.1021/tx000038p
   Boatman RJ, 1996, J TOXICOL ENV HEALTH, V47, P159, DOI 10.1080/009841096161861
   Boatman RJ, 2004, ARCH TOXICOL, V78, P443, DOI 10.1007/s00204-004-0563-2
   Boobis AR, 2006, CRIT REV TOXICOL, V36, P781, DOI 10.1080/10408440600977677
   BRUNNER M, 1991, MUTAGENESIS, V6, P65, DOI 10.1093/mutage/6.1.65
   BUCHER JR, 1990, J APPL TOXICOL, V10, P369, DOI 10.1002/jat.2550100510
   BUCHER JR, 1990, J APPL TOXICOL, V10, P359, DOI 10.1002/jat.2550100509
   BUCKS DAW, 1988, J TOXICOL ENV HEALTH, V24, P279, DOI 10.1080/15287398809531162
   CARLSON AJ, 1953, P SOC EXP BIOL MED, V84, P684
   CHEN HW, 1995, CARCINOGENESIS, V16, P2301, DOI 10.1093/carcin/16.10.2301
   CHEN HW, 1995, CARCINOGENESIS, V16, P1963, DOI 10.1093/carcin/16.8.1963
   CIRANNI R, 1988, MUTAT RES, V209, P23, DOI 10.1016/0165-7992(88)90105-4
   CIRANNI R, 1991, MUTAT RES, V263, P223, DOI 10.1016/0165-7992(91)90005-O
   DeCaprio AP, 1999, CRIT REV TOXICOL, V29, P283, DOI 10.1080/10408449991349221
   Deisinger PJ, 1999, DRUG METAB DISPOS, V27, P442
   Deisinger PJ, 1996, J TOXICOL ENV HEALTH, V47, P31, DOI 10.1080/009841096161915
   DEVOGEL N, 2000, V99208
   Dillon D, 1998, MUTAGENESIS, V13, P19, DOI 10.1093/mutage/13.1.19
   DIVINCENZO GD, 1984, TOXICOLOGY, V33, P9, DOI 10.1016/0300-483X(84)90012-X
   DOBO KL, 1994, ENVIRON MOL MUTAGEN, V24, P293, DOI 10.1002/em.2850240406
   Doepker CL, 2000, MUTAGENESIS, V15, P479, DOI 10.1093/mutage/15.6.479
   EASTMOND DA, 1994, MUTAT RES, V322, P9, DOI 10.1016/0165-1218(94)90028-0
   EASTMOND DA, 1987, TOXICOL APPL PHARM, V91, P85, DOI 10.1016/0041-008X(87)90196-7
   ELEMA JD, 1975, LAB INVEST, V33, P554
   ELLARD S, 1993, MUTAT RES, V287, P87, DOI 10.1016/0027-5107(93)90147-8
   ENGLISH JC, 1994, FUND APPL TOXICOL, V23, P397, DOI 10.1006/faat.1994.1121
   English JC, 2005, FOOD CHEM TOXICOL, V43, P483, DOI 10.1016/j.fct.2004.11.015
   ENGLISH JC, 1994, FUND APPL TOXICOL, V23, P391, DOI 10.1006/faat.1994.1120
   ENGLISH JC, 1988, TX8884
   EREXSON GL, 1985, CANCER RES, V45, P2471
   FAHRIG R, 1984, MOL GEN GENET, V194, P7, DOI 10.1007/BF00383490
   Felix CA, 2006, DNA REPAIR, V5, P1093, DOI 10.1016/j.dnarep.2006.05.031
   FERGUSON LR, 1993, MUTAT RES, V287, P101, DOI 10.1016/0027-5107(93)90149-A
   FOLEY WA, 1964, LAB INVEST, V13, P439
   FOUREMAN P, 1994, ENVIRON MOL MUTAGEN, V23, P51, DOI 10.1002/em.2850230109
   FRANTZ CE, 1998, ENVIRON HEALTH PER S, V104, P1319
   FRIEDLANDER BR, 1982, J OCCUP ENVIRON MED, V24, P605, DOI 10.1097/00043764-198208000-00016
   FROCHT A, 1984, J AM GERIATR SOC, V32, P28, DOI 10.1111/j.1532-5415.1984.tb05148.x
   Fryzek JP, 2005, J OCCUP ENVIRON MED, V47, P278, DOI 10.1097/01.jom.0000155712.22617.42
   GALLOWAY SM, 1987, ENVIRON MOL MUTAGEN, V10, P1, DOI 10.1002/em.2850100502
   Glockl I, 2001, J CHROMATOGR B, V761, P261, DOI 10.1016/S0378-4347(01)00322-X
   Hakura A, 1996, MUTAT RES-GENET TOX, V371, P293, DOI 10.1016/S0165-1218(96)90118-4
   Hard G. C., 1995, GUIDES TOXICOLOGIC P
   Hard GC, 2005, TOXICOL PATHOL, V33, P641, DOI 10.1080/01926230500299716
   Hard GC, 2004, TOXICOL PATHOL, V32, P171, DOI 10.1080/01926230490422574
   Hard GC, 1998, TOXICOL PATHOL, V26, P104, DOI 10.1177/019262339802600112
   Hard GC, 1997, TOXICOL PATHOL, V25, P132, DOI 10.1177/019262339702500202
   Hard GC, 2002, TOXICOL SCI, V69, P30, DOI 10.1093/toxsci/69.1.30
   Hard GC, 2006, TOXICOL PATHOL, V34, P941, DOI 10.1080/01926230601083381
   HASEGAWA R, 1990, JPN J CANCER RES, V81, P871, DOI 10.1111/j.1349-7006.1990.tb02660.x
   HAWORTH S, 1983, ENVIRON MUTAGEN, V5, P3, DOI 10.1002/em.2860050703
   HILL BA, 1993, CHEM RES TOXICOL, V6, P459, DOI 10.1021/tx00034a012
   Hillebrand E, 2005, J ECONOMETRICS, V129, P121, DOI 10.1016/j.jeconom.2004.09.005
   Hiraku Y, 1996, CANCER RES, V56, P5172
   HIROSE M, 1989, CANCER RES, V49, P5143
   HOOD RD, 2005, DEV REPRODUTIVE TOXI
   Hutt AM, 1996, ENVIRON HEALTH PERSP, V104, P1265, DOI 10.2307/3433173
   *IARC, 2004, IARC MON EV CARC RIS, V82, P339
   *IARC, 1980, IARC MON EV CARC RIS, V24, P135
   *IARC, 1992, INT CLASS ROD TUM
   INOUE O, 1989, BRIT J IND MED, V46, P559
   International Agency for Research on Cancer (IARC), 2002, IARC MON EV CARC RIS, V82, P69
   KALF G, 1990, BIOL REACT INTERMED, P443
   KARI FW, 1992, FOOD CHEM TOXICOL, V30, P737, DOI 10.1016/0278-6915(92)90075-V
   KASAI H, 1984, JPN J CANCER RES, V75, P565
   KISHIZAWA F, 1956, JPN J CANC RES GANN, V47, P601
   KISHIZAWA F, 1954, JPN J CANC RES GANN, V45, P389
   KNADLE S, 1985, CANCER RES, V45, P4853
   KOLACHANA P, 1993, CANCER RES, V53, P1023
   KONISHI N, 1989, VET PATHOL, V26, P6, DOI 10.1177/030098588902600102
   Kooyers TJ, 2006, J EUR ACAD DERMATOL, V20, P777, DOI 10.1111/j.1468-3083.2005.01218.x
   KRASAVAGE WJ, 1984, HYDROQUIONONE DOMINA
   KURATA Y, 1990, JPN J CANCER RES, V81, P754, DOI 10.1111/j.1349-7006.1990.tb02641.x
   LAU SS, 1988, MOL PHARMACOL, V34, P829
   Lau SS, 1996, ADV EXP MED BIOL, V387, P267
   Lau SS, 2001, CHEM RES TOXICOL, V14, P25, DOI 10.1021/tx000161g
   LEANDERSON P, 1990, CHEM-BIOL INTERACT, V75, P71, DOI 10.1016/0009-2797(90)90023-G
   LEOPARDI P, 1993, MUTAT RES, V287, P119, DOI 10.1016/0027-5107(93)90151-5
   LEVAY G, 1993, CARCINOGENESIS, V14, P2329
   LEVAY G, 1991, CARCINOGENESIS, V12, P1181, DOI 10.1093/carcin/12.7.1181
   Lock EA, 2004, CRIT REV TOXICOL, V34, P211, DOI 10.1080/10408440490265210
   Makropoulos V, 2006, J OCCUP MED TOXICOL, V1, DOI 10.1186/1745-6673-1-19
   MARRAZZINI A, 1994, MUTAT RES-GENET TOX, V341, P29, DOI 10.1016/0165-1218(94)90022-1
   MARRAZZINI A, 1994, MUTAGENESIS, V9, P505, DOI 10.1093/mutage/9.6.505
   MARUYAMA H, 1991, CARCINOGENESIS, V12, P1331, DOI 10.1093/carcin/12.7.1331
   McDonald TA, 2001, LEUKEMIA, V15, P10, DOI 10.1038/sj.leu.2401981
   MCGREGOR DB, 1988, ENVIRON MOL MUTAGEN, V11, P523, DOI 10.1002/em.2850110413
   MCGREGOR DB, 1988, ENVIRON MOL MUTAGEN, V11, P91, DOI 10.1002/em.2850110110
   Meek ME, 2003, CRIT REV TOXICOL, V33, P591, DOI 10.1080/713608373
   MILLER BM, 1991, MUTAGENESIS, V6, P297, DOI 10.1093/mutage/6.4.297
   MIYATA Y, 1985, JPN J CANCER RES, V76, P828
   Monks TJ, 1997, CHEM RES TOXICOL, V10, P1296, DOI 10.1021/tx9700937
   MONKS TJ, 1994, CHEM RES TOXICOL, V7, P495, DOI 10.1021/tx00040a004
   MORIMOTO K, 1983, MUTAT RES, V119, P355, DOI 10.1016/0165-7992(83)90185-9
   MORIMOTO K, 1980, CANCER RES, V40, P1189
   Morton LD, 2002, TOXICOL PATHOL, V30, P559, DOI 10.1080/01926230290105794
   NAKAMURA S, 1987, MUTAT RES, V192, P239, DOI 10.1016/0165-7992(87)90063-7
   Nielsen H, 1996, SCAND J WORK ENV HEA, V22, P108, DOI 10.5271/sjweh.118
   *NIOSH, 1978, PUBLICATION DHEW
   Nyska A, 1999, TOXICOL PATHOL, V27, P456, DOI 10.1177/019262339902700410
   O'Donoghue J, 1999, FOOD CHEM TOXICOL, V37, P931, DOI 10.1016/S0278-6915(99)00084-8
   ODONAGHUE JL, 1995, LANCET, V346, P1427, DOI 10.1016/S0140-6736(95)92444-2
   OGLESBY FL, 1947, J IND HYG TOXICOL, V29, P74
   OKAZAKI S, 1993, TERATOGEN CARCIN MUT, V13, P127, DOI 10.1002/tcm.1770130303
   PACCHIEROTTI F, 1991, MUTAGENESIS, V6, P307, DOI 10.1093/mutage/6.4.307
   Peters MMCG, 1997, CARCINOGENESIS, V18, P2393, DOI 10.1093/carcin/18.12.2393
   PIFER JW, 1995, INT ARCH OCC ENV HEA, V67, P267, DOI 10.1007/BF00409409
   PIFER JW, 1986, J OCCUP ENVIRON MED, V28, P438, DOI 10.1097/00043764-198606000-00011
   Rao GN, 2002, TOXICOL PATHOL, V30, P651, DOI 10.1080/01926230290166733
   REDDY MV, 1990, CARCINOGENESIS, V11, P1349, DOI 10.1093/carcin/11.8.1349
   REDDY MV, 1997, P AM ASS CANC RES AP, V38
   ROBERTSON ML, 1991, MUTAT RES, V249, P201, DOI 10.1016/0027-5107(91)90147-G
   ROE FJC, 1955, BRIT J CANCER, V9, P177, DOI 10.1038/bjc.1955.14
   ROSSI L, 1986, ARCH BIOCHEM BIOPHYS, V251, P25, DOI 10.1016/0003-9861(86)90047-0
   Roza L, 2003, FOOD CHEM TOXICOL, V41, P1299, DOI 10.1016/S0278-6915(03)00118-2
   Ruggenenti P, 2001, LANCET, V357, P1601, DOI 10.1016/S0140-6736(00)04728-0
   SAKAI M, 1985, MUTAT RES, V156, P61, DOI 10.1016/0165-1218(85)90007-2
   SAWADA Y, 1975, BIOCHEMISTRY-US, V14, P3761, DOI 10.1021/bi00688a007
   Schindler G, 2002, J CLIN PHARMACOL, V42, P920, DOI 10.1177/009127002401102740
   SCHLOSSER MJ, 1990, CHEM RES TOXICOL, V3, P333, DOI 10.1021/tx00016a010
   SCHLOSSER MJ, 1989, CHEM-BIOL INTERACT, V72, P191, DOI 10.1016/0009-2797(89)90027-6
   SEELBACH A, 1993, MUTAT RES, V303, P163, DOI 10.1016/0165-7992(93)90018-Q
   Seely JC, 2002, TOXICOL PATHOL, V30, P681, DOI 10.1080/01926230290166779
   SHIBATA MA, 1991, JPN J CANCER RES, V82, P1211, DOI 10.1111/j.1349-7006.1991.tb01783.x
   SHORT BG, 1989, TOXICOL APPL PHARM, V101, P414, DOI 10.1016/0041-008X(89)90191-9
   Siegers C. P., 1997, Pharmaceutical and Pharmacological Letters, V7, P90
   STENIUS U, 1989, CARCINOGENESIS, V10, P593, DOI 10.1093/carcin/10.3.593
   SUBRAHMANYAM VV, 1991, ARCH BIOCHEM BIOPHYS, V286, P76, DOI 10.1016/0003-9861(91)90010-G
   SUZUKI J, 1995, FREE RADICAL BIO MED, V18, P431, DOI 10.1016/0891-5849(94)00152-A
   TAKIZAWA N, 1940, JPN J CANC RES GANN, V34, P158
   Tanaka Akifumi, 1995, In Vivo (Attiki), V9, P495
   Tash JS, 2002, J APPL PHYSIOL, V92, P1191, DOI 10.1152/japplphysiol.00931.2001
   Topping DC, 2007, FOOD CHEM TOXICOL, V45, P70, DOI 10.1016/j.fct.2006.07.019
   Tsutsui T, 1997, MUTAT RES-FUND MOL M, V373, P113, DOI 10.1016/S0027-5107(96)00196-0
   TUNEK A, 1982, CHEM-BIOL INTERACT, V39, P129, DOI 10.1016/0009-2797(82)90116-8
   UMEDA M, 1957, JPN J CANC RES GANN, V48, P139
   UMEMURA T, 1993, J TOXICOL PATHOL, V6, P81
   *US ENV PROT AG, 1987, HLTH ENV EFF DOC P H
   *US NAT TOX PROGR, 1999, TOX CARC STUD ETH BE
   *US NAT TOX PROGR, 1989, TOX CARC STUD HYDR
   VANDELFT JHM, 1997, V97476
   VANG O, 1993, CARCINOGENESIS, V14, P2365, DOI 10.1093/carcin/14.11.2365
   VANHUMMELEN P, 1992, MUTAGENESIS, V7, P447
   VIAN L, 1995, MUTAT RES-ENVIR MUTA, V334, P1, DOI 10.1016/0165-1161(95)90024-1
   WALLES SAS, 1992, CANCER LETT, V63, P47
   WALLIN M, 1993, MUTAT RES, V287, P17, DOI 10.1016/0027-5107(93)90141-2
   WANG X, 1990, MUTAGENESIS, V5, P371, DOI 10.1093/mutage/5.4.371
   Wester RC, 1998, J TOXICOL ENV HEAL A, V54, P301
   *WHO, 1994, HYDROQUINONE
   WHYSNER J, 1995, REGUL TOXICOL PHARM, V21, P158, DOI 10.1006/rtph.1995.1020
   WILLIAMS GM, 2007, FOOD CHEM TOXICOL
   Winn LM, 2003, TOXICOL SCI, V72, P143, DOI 10.1093/toxsci/kfg008
   WINTERBOURN CC, 1981, ARCH BIOCHEM BIOPHYS, V209, P159, DOI 10.1016/0003-9861(81)90268-X
   WYNDER EL, 1967, TOBACCO TOBACCO SMOK, P388
   YAGER JW, 1990, CANCER RES, V50, P393
   YAMAGUCHI S, 1989, CANCER RES, V49, P6015
   YAMAZAKI I, 1960, J BIOL CHEM, V235, P2444
NR 168
TC 97
Z9 105
U1 1
U2 34
PU TAYLOR & FRANCIS LTD
PI ABINGDON
PA 2-4 PARK SQUARE, MILTON PARK, ABINGDON OR14 4RN, OXON, ENGLAND
SN 1040-8444
EI 1547-6898
J9 CRIT REV TOXICOL
JI Crit. Rev. Toxicol.
PY 2007
VL 37
IS 10
BP 887
EP 914
DI 10.1080/10408440701638970
PG 28
WC Toxicology
WE Science Citation Index Expanded (SCI-EXPANDED)
SC Toxicology
GA 237UF
UT WOS:000251401200002
PM 18027166
DA 2023-08-27
ER

PT J
AU Lee, N
   Akazawa, C
   Akazawa, P
   Quivey, JM
   Tang, C
   Verhey, LJ
   Xia, P
AF Lee, N
   Akazawa, C
   Akazawa, P
   Quivey, JM
   Tang, C
   Verhey, LJ
   Xia, P
TI A forward-planned treatment technique using multisegments in the
   treatment of head-and-neck cancer
SO INTERNATIONAL JOURNAL OF RADIATION ONCOLOGY BIOLOGY PHYSICS
LA English
DT Article; Proceedings Paper
CT 44th Annual Meeting of the
   American-Society-for-Therapeutic-Radiology-and-Oncology
CY OCT 06-10, 2002
CL NEW ORLEANS, LA
SP Amer Soc Therapeut Radiol & Oncol
DE IMRT; head and neck; forward planning; multisegment
ID INTENSITY-MODULATED RADIOTHERAPY; 3-DIMENSIONAL CONFORMAL RADIOTHERAPY;
   RADIATION-THERAPY; NASOPHARYNGEAL CARCINOMA; IMRT; BOOST
AB Purpose: To describe in detail a forward-planned multisegment technique (FPMS) as an alternative treatment method for patients who are not suitable for inverse-planned intensity-modulated radiation therapy (IP-IMRT), or for situations where IP-IMRT is not available in a medical clinic.
   Methods and Materials: Between April 1995 and February 2002, 38 primary head-and-neck patients were treated using tie FPMS technique, which has evolved over the past 7 years at our medical center. In the most recent version of the FPMS technique, which includes 5 patients examined in this analysis, the primary tumor and the upper neck nodes were treated with 7 gantry angles, including an anterior, 2 lateral, 2 anterior oblique, and 2 posterior oblique beams with a total of 13 beam shapes formed by multileaf collimators (MLC), called MLC segments. The shape of each MLC segment was carefully designed, and the associated weights were optimized through manual iterations. The lower neck nodes and the supraclavicular nodes were treated with a split-beam anterior field, matched to the inferior border of the FPMS plan at the isocenter. With an autosequencing delivery system, all fields, including dynamic wedges, can be automatically treated. The dosimetric accuracy of this technique was verified with a phantom plan and measured with an ionization chamber, as well as film dosimetry. A sample FPMS plan is described in detail, and the average results for the 5 patients treated with FPMS are retrospectively compared to results for similar patients treated with IP-IMRT.
   Results: The gross tumor volume was prescribed to 70 Gy (2.12 Gy/fraction) at the 88% isodose line, whereas the clinical target volume received a dose of 59.4 Gy (1.8 Gy/fraction) at the 75% isodose line. The maximum dose to the brainstem and spinal cord was below 54 and 45 Gy, respectively, comparable to IP-IMRT. The mean dose to the parotid glands was 32 Gy with FPMS vs. 26 Gy with IP-IMRT. Average delivery time was shorter for FPMS (15 min) than IP-IMRT (30 min), whereas the planning time depended on the expertise of the planner. Dosimetric accuracy for FPMS and IP-IMRT plans using phantom measurements was similar, within 1% of the phantom plan. With a median follow-up of 31 months, there was no local-regional recurrence, and the incidence of xerostomia is reduced compared to conventional techniques.
   Conclusion: FPMS achieved plans comparable to those for IP-IMRT and is an ideal alternative treatment technique for a center without the capabilities of IP-IMRT or for a patient who is not a suitable candidate, because of prolonged treatment time. The treatment outcomes from our clinical experience indicate that FPMS can achieve excellent local freedom from progression rates without causing excessive toxicity. Lastly, IP-IMRT plans should be comparable to, if not better than, FPMS plans in the treatment of head-and-neck cancer. (C) 2004 Elsevier Inc.
C1 Univ Calif San Francisco, Med Ctr, Dept Radiat Oncol, San Francisco, CA 94143 USA.
C3 University of California System; University of California San Francisco
RP Xia, P (通讯作者)，Univ Calif San Francisco, Ctr Comprehens Canc, Dept Radiat Oncol, 1600 Divisadero St,H-1031, San Francisco, CA 94143 USA.
EM
CR Butler EB, 1999, INT J RADIAT ONCOL, V45, P21, DOI 10.1016/S0360-3016(99)00101-7
   Chao KSC, 2003, INT J RADIAT ONCOL, V55, P312, DOI 10.1016/S0360-3016(02)03940-8
   Cheng JCH, 2001, INT J CANCER, V96, P126, DOI 10.1002/ijc.1004
   Chuang CF, 2002, MED PHYS, V29, P1109, DOI 10.1118/1.1481520
   Claus F, 2002, RADIOTHER ONCOL, V64, P205, DOI 10.1016/S0167-8140(02)00172-X
   Dawson LA, 2000, INT J RADIAT ONCOL, V46, P1117, DOI 10.1016/S0360-3016(99)00550-7
   Hsiung CY, 2002, INT J RADIAT ONCOL, V53, P638, DOI 10.1016/S0360-3016(02)02760-8
   Hunt MA, 2001, INT J RADIAT ONCOL, V49, P623, DOI 10.1016/S0360-3016(00)01389-4
   Kam MKM, 2003, INT J RADIAT ONCOL, V56, P145, DOI 10.1016/S0360-3016(03)00075-0
   Lee N, 2003, INT J RADIAT ONCOL, V57, P49, DOI 10.1016/S0360-3016(03)00405-X
   Lee N, 2002, INT J RADIAT ONCOL, V53, P12, DOI 10.1016/S0360-3016(02)02724-4
   Nutting C, 2000, BRIT J RADIOL, V73, P459, DOI 10.1259/bjr.73.869.10884741
   Sultanem K, 2000, INT J RADIAT ONCOL, V48, P711, DOI 10.1016/S0360-3016(00)00702-1
   van Dieren EB, 2000, INT J RADIAT ONCOL, V47, P1299, DOI 10.1016/S0360-3016(00)00564-2
   Xia P, 2000, INT J RADIAT ONCOL, V48, P329, DOI 10.1016/S0360-3016(00)00585-X
   Xia P, 2001, INT J RADIAT ONCOL, V51, P244, DOI 10.1016/S0360-3016(01)01643-1
   Zabel A, 2002, BRIT J RADIOL, V75, P356, DOI 10.1259/bjr.75.892.750356
NR 17
TC 19
Z9 19
U1 1
U2 4
PU ELSEVIER SCIENCE INC
PI NEW YORK
PA STE 800, 230 PARK AVE, NEW YORK, NY 10169 USA
SN 0360-3016
EI 1879-355X
J9 INT J RADIAT ONCOL
JI Int. J. Radiat. Oncol. Biol. Phys.
PD JUN 1
PY 2004
VL 59
IS 2
BP 584
EP 594
DI 10.1016/j.ijrobp.2004.02.005
PG 11
WC Oncology; Radiology, Nuclear Medicine & Medical Imaging
WE Science Citation Index Expanded (SCI-EXPANDED); Conference Proceedings Citation Index - Science (CPCI-S)
SC Oncology; Radiology, Nuclear Medicine & Medical Imaging
GA 821DS
UT WOS:000221440800036
PM 15145180
DA 2023-08-27
ER

PT J
AU Sturm, D
   Bender, S
   Jones, DTW
   Lichter, P
   Grill, J
   Becher, O
   Hawkins, C
   Majewski, J
   Jones, C
   Costello, JF
   Iavarone, A
   Aldape, K
   Brennan, CW
   Jabado, N
   Pfister, SM
AF Sturm, Dominik
   Bender, Sebastian
   Jones, David T. W.
   Lichter, Peter
   Grill, Jacques
   Becher, Oren
   Hawkins, Cynthia
   Majewski, Jacek
   Jones, Chris
   Costello, Joseph F.
   Iavarone, Antonio
   Aldape, Kenneth
   Brennan, Cameron W.
   Jabado, Nada
   Pfister, Stefan M.
TI Paediatric and adult glioblastoma: multiform (epi)genomic culprits
   emerge
SO NATURE REVIEWS CANCER
LA English
DT Review
ID GROWTH-FACTOR RECEPTOR; TERT PROMOTER MUTATIONS; HIGH-GRADE GLIOMAS;
   PHASE-II TRIAL; INTEGRATED GENOMIC ANALYSIS; INTRINSIC PONTINE GLIOMAS;
   BRAIN-TUMOR CONSORTIUM; TYROSINE KINASE GENES; DNA MISMATCH REPAIR;
   NEURAL STEM-CELLS
AB We have extended our understanding of the molecular biology that underlies adult glioblastoma over many years. By contrast, high-grade gliomas in children and adolescents have remained a relatively under-investigated disease. The latest large-scale genomic and epigenomic profiling studies have yielded an unprecedented abundance of novel data and provided deeper insights into gliomagenesis across all age groups, which has highlighted key distinctions but also some commonalities. As we are on the verge of dissecting glioblastomas into meaningful biological subgroups, this Review summarizes the hallmark genetic alterations that are associated with distinct epigenetic features and patient characteristics in both paediatric and adult disease, and examines the complex interplay between the glioblastoma genome and epigenome.
C1 [Sturm, Dominik; Bender, Sebastian; Jones, David T. W.; Pfister, Stefan M.] German Canc Res Ctr DKFZ Heidelberg, Div Pediat Neurooncol, D-69120 Heidelberg, Germany.
   [Sturm, Dominik; Bender, Sebastian; Pfister, Stefan M.] Univ Heidelberg Hosp, Dept Pediat Oncol Hematol & Immunol, D-69120 Heidelberg, Germany.
   [Lichter, Peter] German Canc Res Ctr DKFZ Heidelberg, Div Mol Genet, D-69120 Heidelberg, Germany.
   [Grill, Jacques] Univ Paris 11, Gustave Roussy Canc Inst, Dept Pediat & Adolescent Oncol, Brain Tumor Program, F-94805 Villejuif, France.
   [Becher, Oren] Duke Univ, Med Ctr, Div Pediat Hematol Oncol, Durham, NC 27710 USA.
   [Hawkins, Cynthia] Hosp Sick Children, Arthur & Sonia Labatt Brain Tumour Res Ctr, Toronto, ON M5G 1X8, Canada.
   [Majewski, Jacek; Jabado, Nada] McGill Univ, Div Expt Med, Montreal, PQ H3H 2R9, Canada.
   [Majewski, Jacek; Jabado, Nada] McGill Univ, Dept Human Genet, Montreal, PQ H3H 2R9, Canada.
   [Majewski, Jacek; Jabado, Nada] McGill Univ, Ctr Hlth, Montreal, PQ H3H 2R9, Canada.
   [Jones, Chris] Inst Canc Res, Div Mol Pathol, Sutton SM2 5NG, Surrey, England.
   [Jones, Chris] Inst Canc Res, Div Canc Therapeut, Sutton SM2 5NG, Surrey, England.
   [Costello, Joseph F.] Univ Calif San Francisco, Dept Neurosurg, Brain Tumor Res Ctr, San Francisco, CA 94143 USA.
   [Iavarone, Antonio] Columbia Univ, Med Ctr, Inst Canc Genet, New York, NY 10032 USA.
   [Iavarone, Antonio] Columbia Univ, Med Ctr, Dept Pathol, New York, NY 10032 USA.
   [Iavarone, Antonio] Columbia Univ, Med Ctr, Dept Neurol, New York, NY 10032 USA.
   [Aldape, Kenneth] Univ Texas MD Anderson Canc Ctr, Dept Neurooncol, Houston, TX 77030 USA.
   [Brennan, Cameron W.] Mem Sloan Kettering Canc Ctr, Brain Tumor Ctr, Human Oncol & Pathogenesis Program, New York, NY 10065 USA.
   [Brennan, Cameron W.] Mem Sloan Kettering Canc Ctr, Brain Tumor Ctr, Dept Neurosurg, New York, NY 10065 USA.
C3 Helmholtz Association; German Cancer Research Center (DKFZ); Ruprecht
   Karls University Heidelberg; Helmholtz Association; German Cancer
   Research Center (DKFZ); UDICE-French Research Universities; Universite
   Paris Saclay; UNICANCER; Gustave Roussy; Duke University; University of
   Toronto; Hospital for Sick Children (SickKids); McGill University;
   McGill University; McGill University; University of London; Institute of
   Cancer Research - UK; University of London; Institute of Cancer Research
   - UK; University of California System; University of California San
   Francisco; Columbia University; Columbia University; Columbia
   University; University of Texas System; UTMD Anderson Cancer Center;
   Memorial Sloan Kettering Cancer Center; Memorial Sloan Kettering Cancer
   Center
RP Jabado, N (通讯作者)，McGill Univ, Div Expt Med, 2155 Guy St, Montreal, PQ H3H 2R9, Canada.
EM
RI Hawkins, Cynthia/M-2571-2019; Jones, David TW/G-9588-2013; Jabado,
   Nada/AAN-4026-2020; Pfister, Stefan M/F-6860-2013; Brennan,
   Cameron/R-7082-2019; Jones, David T. W./Z-1963-2019; Becher,
   Oren/GXN-3269-2022; Grill, Jacques/R-8093-2019; Brennan,
   Cameron/IWU-9193-2023; Sturm, Dominik/AAL-3689-2020; Jones,
   Chris/ABE-6059-2021; Becher, Oren/AAI-7495-2020
OI Jones, David TW/0000-0002-2036-5141; Pfister, Stefan
   M/0000-0002-5447-5322; Brennan, Cameron/0000-0003-4064-8891; Jones,
   David T. W./0000-0002-2036-5141; Becher, Oren/0000-0003-4016-8492;
   Sturm, Dominik/0000-0003-0250-1696; Jones, Chris/0000-0001-8118-2296;
   Iavarone, Antonio/0000-0002-0683-4634
FU PedBrain Tumor Project; German Cancer Aid [109252]; German Federal
   Ministry of Education and Research (BMBF) [01KU1201A, 0315416C,
   01GS0883]; Genome Canada; Genome Quebec; Canadian Institutes for Health
   Research (CIHR); McGill University, Montreal, Quebec, Canada; Montreal
   Children's Hospital Foundation; Cancer Research UK [11736, 13982]
   Funding Source: researchfish; Rosetrees Trust [M200-F1] Funding Source:
   researchfish; The Brain Tumour Charity [6/78] Funding Source:
   researchfish
FX This work was principally supported by the PedBrain Tumor Project
   contributing to the International Cancer Genome Consortium, funded by
   German Cancer Aid (109252) and by the German Federal Ministry of
   Education and Research (BMBF; grants #01KU1201A, MedSys #0315416C and
   NGFNplus #01GS0883). This work was further performed within the context
   of the I-CHANGE consortium supported by Genome Canada, Genome Quebec,
   the Canadian Institutes for Health Research (CIHR), McGill University,
   Montreal, Quebec, Canada, and the Montreal Children's Hospital
   Foundation.
CR Alaminos M, 2005, CANCER RES, V65, P2565, DOI 10.1158/0008-5472.CAN-04-4283
   Amatya VJ, 2005, ACTA NEUROPATHOL, V110, P178, DOI 10.1007/s00401-005-1041-5
   [Anonymous], 2007, WHO HISTOLOGICAL CLA
   Appolloni I, 2012, INT J CANCER, V131, pE1078, DOI 10.1002/ijc.27606
   Arita H, 2013, ACTA NEUROPATHOL, V126, P267, DOI 10.1007/s00401-013-1141-6
   Baeza N, 2003, ACTA NEUROPATHOL, V106, P479, DOI 10.1007/s00401-003-0748-4
   Bajenaru ML, 2002, MOL CELL BIOL, V22, P5100, DOI 10.1128/MCB.22.14.5100-5113.2002
   Barker CA, 2013, INT J RADIAT ONCOL, V86, P504, DOI 10.1016/j.ijrobp.2013.02.012
   Barrett LE, 2012, CANCER CELL, V21, P11, DOI 10.1016/j.ccr.2011.11.025
   Barrow J, 2011, NEURO-ONCOLOGY, V13, P212, DOI 10.1093/neuonc/noq158
   Batchelor TT, 2013, J CLIN ONCOL, V31, P3212, DOI 10.1200/JCO.2012.47.2464
   Bax DA, 2010, CLIN CANCER RES, V16, P3368, DOI 10.1158/1078-0432.CCR-10-0438
   Bax DA, 2009, CLIN CANCER RES, V15, P5753, DOI 10.1158/1078-0432.CCR-08-3210
   Becher OJ, 2006, CANCER RES, V66, P3355, DOI 10.1158/0008-5472.CAN-05-3827
   Becher OJ, 2010, CANCER RES, V70, P2548, DOI 10.1158/0008-5472.CAN-09-2503
   Bender S, 2013, CANCER CELL, V24, P660, DOI 10.1016/j.ccr.2013.10.006
   Bhat KPL, 2013, CANCER CELL, V24, P331, DOI 10.1016/j.ccr.2013.08.001
   Bhat KPL, 2011, GENE DEV, V25, P2594, DOI 10.1101/gad.176800.111
   Bielen A, 2011, MOL CANCER THER, V10, P1407, DOI 10.1158/1535-7163.MCT-11-0205
   Biernat W, 2004, BRAIN PATHOL, V14, P131, DOI 10.1111/j.1750-3639.2004.tb00045.x
   Bjerke L, 2013, CANCER DISCOV, V3, P512, DOI 10.1158/2159-8290.CD-12-0426
   Boldrini L, 2006, INT J ONCOL, V28, P1555
   Bozdag S, 2013, PLOS ONE, V8, DOI 10.1371/journal.pone.0062982
   Brennan CW, 2013, CELL, V155, P462, DOI 10.1016/j.cell.2013.09.034
   Brown PD, 2008, J CLIN ONCOL, V26, P5603, DOI 10.1200/JCO.2008.18.0612
   Bruna A, 2007, CANCER CELL, V11, P147, DOI 10.1016/j.ccr.2006.11.023
   Buttarelli FR, 2010, CHILD NERV SYST, V26, P1051, DOI 10.1007/s00381-010-1191-1
   Cadieux B, 2006, CANCER RES, V66, P8469, DOI 10.1158/0008-5472.CAN-06-1547
   Cage TA, 2013, CHILD NERV SYST, V29, P1313, DOI 10.1007/s00381-013-2101-0
   Cahill DP, 2007, CLIN CANCER RES, V13, P2038, DOI 10.1158/1078-0432.CCR-06-2149
   Carro MS, 2010, NATURE, V463, P318, DOI 10.1038/nature08712
   Chan KM, 2013, GENE DEV, V27, P985, DOI 10.1101/gad.217778.113
   Charest A., 2013, STEM CELLS
   Chen L, 2013, BBA-REV CANCER, V1836, P158, DOI 10.1016/j.bbcan.2013.04.003
   Chen YJ, 2006, CANCER RES, V66, P6473, DOI 10.1158/0008-5472.CAN-06-0910
   Cheng ZX, 2013, CLIN CANCER RES, V19, P1748, DOI 10.1158/1078-0432.CCR-12-3066
   Chin L, 2008, NATURE, V455, P1061, DOI 10.1038/nature07385
   Cho J, 2011, CANCER RES, V71, P7587, DOI 10.1158/0008-5472.CAN-11-0821
   Chow LML, 2011, CANCER CELL, V19, P305, DOI 10.1016/j.ccr.2011.01.039
   Chowdhury R, 2011, EMBO REP, V12, P463, DOI 10.1038/embor.2011.43
   Cohen KJ, 2011, NEURO-ONCOLOGY, V13, P317, DOI 10.1093/neuonc/noq191
   COSTELLO JF, 1994, J BIOL CHEM, V269, P17228
   COSTELLO JF, 1994, MOL CELL BIOL, V14, P6515, DOI 10.1128/MCB.14.10.6515
   Costello JF, 1996, CANCER RES, V56, P2405
   Dang L, 2009, NATURE, V462, P739, DOI 10.1038/nature08617
   Danussi C, 2013, CANCER RES, V73, P5140, DOI 10.1158/0008-5472.CAN-13-1168-T
   Dawson MA, 2012, CELL, V150, P12, DOI 10.1016/j.cell.2012.06.013
   Ding H, 2003, CANCER RES, V63, P1106
   Dolecek TA, 2012, NEURO-ONCOLOGY, V14, pv1, DOI 10.1093/neuonc/nos218
   Khuong-Quang DA, 2012, ACTA NEUROPATHOL, V124, P439, DOI 10.1007/s00401-012-0998-0
   Donson AM, 2007, PEDIATR BLOOD CANCER, V48, P403, DOI 10.1002/pbc.20803
   Downing JR, 2012, NAT GENET, V44, P619, DOI 10.1038/ng.2287
   Drappatz J, 2012, J NEURO-ONCOL, V107, P133, DOI 10.1007/s11060-011-0717-z
   Dresemann G, 2010, J NEURO-ONCOL, V96, P393, DOI 10.1007/s11060-009-9976-3
   Esteller M, 1999, CANCER RES, V59, P793
   Esteller M, 2000, NEW ENGL J MED, V343, P1350, DOI 10.1056/NEJM200011093431901
   Faury D, 2007, J CLIN ONCOL, V25, P1196, DOI 10.1200/JCO.2006.07.8626
   Felsberg J, 2011, INT J CANCER, V129, P659, DOI 10.1002/ijc.26083
   Fenton TR, 2012, P NATL ACAD SCI USA, V109, P14164, DOI 10.1073/pnas.1211962109
   Figueroa ME, 2010, CANCER CELL, V18, P553, DOI 10.1016/j.ccr.2010.11.015
   Fontebasso AM, 2013, ACTA NEUROPATHOL, V125, P659, DOI 10.1007/s00401-013-1095-8
   Fontebasso AM, 2013, BRAIN PATHOL, V23, P210, DOI 10.1111/bpa.12023
   Franceschi E, 2007, BRIT J CANCER, V96, P1047, DOI 10.1038/sj.bjc.6603669
   Frattini V, 2013, NAT GENET, V45, P1141, DOI 10.1038/ng.2734
   Freije WA, 2004, CANCER RES, V64, P6503, DOI 10.1158/0008-5472.CAN-04-0452
   Friday BB, 2012, NEURO-ONCOLOGY, V14, P215, DOI 10.1093/neuonc/nor198
   Friedman HS, 2009, J CLIN ONCOL, V27, P4733, DOI 10.1200/JCO.2008.19.8721
   Friedmann-Morvinski D, 2012, SCIENCE, V338, P1080, DOI 10.1126/science.1226929
   Gallia GL, 2006, MOL CANCER RES, V4, P709, DOI 10.1158/1541-7786.MCR-06-0172
   Gan HK, 2013, FEBS J, V280, P5350, DOI 10.1111/febs.12393
   Godard S, 2003, CANCER RES, V63, P6613
   Gravendeel LAM, 2009, CANCER RES, V69, P9065, DOI 10.1158/0008-5472.CAN-09-2307
   Greaves M, 2012, NATURE, V481, P306, DOI 10.1038/nature10762
   Gross S., 2012, PROBE REPORTS NIH MO
   Guilhamon P, 2013, NAT COMMUN, V4, DOI 10.1038/ncomms3166
   Hambardzumyan D, 2011, GLIA, V59, P1155, DOI 10.1002/glia.21142
   Hambardzumyan D, 2009, TRANSL ONCOL, V2, P89, DOI 10.1593/tlo.09100
   Haque T, 2007, CLIN CANCER RES, V13, P6284, DOI 10.1158/1078-0432.CCR-07-0525
   Harada K, 1998, CANCER RES, V58, P4694
   Hargrove D, 2006, LANCET ONCOL, V7, P241, DOI 10.1016/S1470-2045(06)70615-5
   Hartmann C, 2013, CLIN CANCER RES, V19, P5146, DOI 10.1158/1078-0432.CCR-13-0017
   Hartmann C, 2010, ACTA NEUROPATHOL, V120, P707, DOI 10.1007/s00401-010-0781-z
   Heaphy CM, 2011, SCIENCE, V333, P425, DOI 10.1126/science.1207313
   Hede SM, 2009, GLIA, V57, P1143, DOI 10.1002/glia.20837
   Hegi ME, 2005, NEW ENGL J MED, V352, P997, DOI 10.1056/NEJMoa043331
   Hegi ME, 2008, J CLIN ONCOL, V26, P4189, DOI 10.1200/JCO.2007.11.5964
   Hegi ME, 2011, MOL CANCER THER, V10, P1102, DOI 10.1158/1535-7163.MCT-11-0048
   Holland EC, 2000, TOXICOL PATHOL, V28, P171, DOI 10.1177/019262330002800122
   Holland EC, 1998, GENE DEV, V12, P3675, DOI 10.1101/gad.12.23.3675
   Hovestadt V, 2013, ACTA NEUROPATHOL, V125, P913, DOI 10.1007/s00401-013-1126-5
   Hunter C, 2006, CANCER RES, V66, P3987, DOI 10.1158/0008-5472.CAN-06-0127
   Huse JT, 2011, GLIA, V59, P1190, DOI 10.1002/glia.21165
   Ichimura K, 2012, BRAIN TUMOR PATHOL, V29, P131, DOI 10.1007/s10014-012-0090-4
   Ichimura K, 2009, NEURO-ONCOLOGY, V11, P341, DOI 10.1215/15228517-2009-025
   Inda MD, 2010, GENE DEV, V24, P1731, DOI 10.1101/gad.1890510
   Jacques TS, 2010, EMBO J, V29, P222, DOI 10.1038/emboj.2009.327
   Jansen MHA, 2012, CANCER TREAT REV, V38, P27, DOI 10.1016/j.ctrv.2011.06.007
   Janssens GO, 2013, INT J RADIAT ONCOL, V85, P315, DOI 10.1016/j.ijrobp.2012.04.006
   Jiao YC, 2012, ONCOTARGET, V3, P709, DOI 10.18632/oncotarget.588
   Jones C, 2012, NAT REV CLIN ONCOL, V9, P400, DOI 10.1038/nrclinonc.2012.87
   Jones DTW, 2013, NAT GENET, V45, P927, DOI 10.1038/ng.2682
   Jordan CT, 2006, NEW ENGL J MED, V355, P1253, DOI 10.1056/NEJMra061808
   Jung V, 1999, J NEUROPATH EXP NEUR, V58, P993, DOI 10.1097/00005072-199909000-00009
   Kang HJ, 2011, NATURE, V478, P483, DOI 10.1038/nature10523
   Killela PJ, 2013, P NATL ACAD SCI USA, V110, P6021, DOI 10.1073/pnas.1303607110
   Kim W, 2012, NEUROSURG CLIN N AM, V23, P471, DOI 10.1016/j.nec.2012.04.009
   Koelsche C, 2013, ACTA NEUROPATHOL, V126, P907, DOI 10.1007/s00401-013-1195-5
   Koso H, 2012, P NATL ACAD SCI USA, V109, pE2998, DOI 10.1073/pnas.1215899109
   Kwon CH, 2008, CANCER RES, V68, P3286, DOI 10.1158/0008-5472.CAN-07-6867
   Lai A, 2011, J CLIN ONCOL, V29, P4482, DOI 10.1200/JCO.2010.33.8715
   Lai A, 2011, J CLIN ONCOL, V29, P142, DOI 10.1200/JCO.2010.30.2729
   Larjavaara S, 2007, NEURO-ONCOLOGY, V9, P319, DOI 10.1215/15228517-2007-016
   Lass U, 2012, PLOS ONE, V7, DOI 10.1371/journal.pone.0041298
   Lassman AB, 2005, CLIN CANCER RES, V11, P7841, DOI 10.1158/1078-0432.CCR-05-0421
   Lee J, 2006, CANCER CELL, V9, P391, DOI 10.1016/j.ccr.2006.03.030
   Lee JY, 2011, CHILD NERV SYST, V27, P1877, DOI 10.1007/s00381-011-1525-7
   Lei L, 2011, PLOS ONE, V6, DOI 10.1371/journal.pone.0020041
   Lewis PW, 2013, SCIENCE, V340, P857, DOI 10.1126/science.1232245
   Li F, 2013, CELL, V153, P590, DOI 10.1016/j.cell.2013.03.025
   Liang Y, 2005, P NATL ACAD SCI USA, V102, P5814, DOI 10.1073/pnas.0402870102
   Little SE, 2012, CANCER RES, V72, P1614, DOI 10.1158/0008-5472.CAN-11-4069
   Liu C, 2011, CELL, V146, P209, DOI 10.1016/j.cell.2011.06.014
   Liu XY, 2012, ACTA NEUROPATHOL, V124, P615, DOI 10.1007/s00401-012-1031-3
   Llaguno SA, 2009, CANCER CELL, V15, P45, DOI 10.1016/j.ccr.2008.12.006
   Lu QR, 2001, P NATL ACAD SCI USA, V98, P10851, DOI 10.1073/pnas.181340798
   Luchman HA, 2012, NEURO-ONCOLOGY, V14, P184, DOI 10.1093/neuonc/nor207
   Maher EA, 2006, CANCER RES, V66, P11502, DOI 10.1158/0008-5472.CAN-06-2072
   Malhotra A, 2013, GENOME RES, V23, P762, DOI 10.1101/gr.143677.112
   Malmstrom A, 2012, LANCET ONCOL, V13, P916, DOI 10.1016/S1470-2045(12)70265-6
   Marian CO, 2010, CLIN CANCER RES, V16, P154, DOI 10.1158/1078-0432.CCR-09-2850
   Marie Y, 2001, LANCET, V358, P298, DOI 10.1016/S0140-6736(01)05499-X
   Marusyk Andriy, 2010, Biochim Biophys Acta, V1805, P105, DOI 10.1016/j.bbcan.2009.11.002
   McCabe MT, 2012, P NATL ACAD SCI USA, V109, P2989, DOI 10.1073/pnas.1116418109
   Mellinghoff IK, 2005, NEW ENGL J MED, V353, P2012, DOI 10.1056/NEJMoa051918
   Monje M, 2011, P NATL ACAD SCI USA, V108, P4453, DOI 10.1073/pnas.1101657108
   Nakamura M, 2001, LAB INVEST, V81, P77, DOI 10.1038/labinvest.3780213
   Neyns B, 2009, ANN ONCOL, V20, P1596, DOI 10.1093/annonc/mdp032
   Nicolaides TP, 2011, CLIN CANCER RES, V17, P7595, DOI 10.1158/1078-0432.CCR-11-1456
   Nobusawa S, 2010, BRAIN PATHOL, V20, P936, DOI 10.1111/j.1750-3639.2010.00395.x
   Nonoguchi N, 2013, ACTA NEUROPATHOL, V126, P931, DOI 10.1007/s00401-013-1163-0
   Noushmehr Houtan, 2010, Cancer Cell, V17, P510, DOI 10.1016/j.ccr.2010.03.017
   Nutt CL, 2003, CANCER RES, V63, P1602
   Ohgaki H, 2007, AM J PATHOL, V170, P1445, DOI 10.2353/ajpath.2007.070011
   Ohgaki H, 2013, CLIN CANCER RES, V19, P764, DOI 10.1158/1078-0432.CCR-12-3002
   Ohnishi A, 2003, J NEUROPATH EXP NEUR, V62, P1052, DOI 10.1093/jnen/62.10.1052
   Olson RA, 2011, J NEURO-ONCOL, V105, P325, DOI 10.1007/s11060-011-0594-5
   Ozawa T, 2010, GENE DEV, V24, P2205, DOI 10.1101/gad.1972310
   Parker BC, 2013, J CLIN INVEST, V123, P855, DOI 10.1172/JCI67144
   Parsons DW, 2008, SCIENCE, V321, P1807, DOI 10.1126/science.1164382
   Paugh BS, 2013, CANCER RES, V73, P6219, DOI 10.1158/0008-5472.CAN-13-1491
   Paugh BS, 2011, J CLIN ONCOL, V29, P3999, DOI 10.1200/JCO.2011.35.5677
   Paugh BS, 2010, J CLIN ONCOL, V28, P3061, DOI 10.1200/JCO.2009.26.7252
   Phillips HS, 2006, CANCER CELL, V9, P157, DOI 10.1016/j.ccr.2006.02.019
   Phillips JJ, 2013, BRAIN PATHOL, V23, P565, DOI 10.1111/bpa.12043
   Pollack IF, 2006, J NEUROSURG, V105, P418, DOI 10.3171/ped.2006.105.5.418
   Pollack IF, 2011, CHILD NERV SYST, V27, P87, DOI 10.1007/s00381-010-1264-1
   Pollack IF, 2001, CANCER RES, V61, P7404
   Prados MD, 2009, J CLIN ONCOL, V27, P579, DOI 10.1200/JCO.2008.18.9639
   Puget S, 2012, PLOS ONE, V7, DOI 10.1371/journal.pone.0030313
   Puissant A, 2013, CANCER DISCOV, V3, P308, DOI 10.1158/2159-8290.CD-12-0418
   Qu HQ, 2010, NEURO-ONCOLOGY, V12, P153, DOI 10.1093/neuonc/nop001
   Radaelli E, 2009, HISTOL HISTOPATHOL, V24, P879, DOI 10.14670/HH-24.879
   Raffel C, 1999, CLIN CANCER RES, V5, P4085
   Raymond E, 2008, J CLIN ONCOL, V26, P4659, DOI 10.1200/JCO.2008.16.9235
   Reardon DA, 2009, BRIT J CANCER, V101, P1995, DOI 10.1038/sj.bjc.6605411
   Reilly KM, 2000, NAT GENET, V26, P109, DOI 10.1038/79075
   Reilly KM, 2004, P NATL ACAD SCI USA, V101, P13008, DOI 10.1073/pnas.0401236101
   Ren ZP, 2007, J NEUROPATH EXP NEUR, V66, P944, DOI 10.1097/nen.0b013e318156bc05
   Rohle D, 2013, SCIENCE, V340, P626, DOI 10.1126/science.1236062
   Ryan RJH, 2011, PLOS ONE, V6, DOI 10.1371/journal.pone.0028585
   Sanborn JZ, 2013, CANCER RES, V73, P6036, DOI 10.1158/0008-5472.CAN-13-0186
   Sasaki M, 2012, GENE DEV, V26, P2038, DOI 10.1101/gad.198200.112
   Schiffman JD, 2010, CANCER RES, V70, P512, DOI 10.1158/0008-5472.CAN-09-1851
   Schwartzentruber J, 2012, NATURE, V482, P226, DOI [10.1038/nature10833, 10.1007/s13311-012-0137-6]
   Shai R, 2003, ONCOGENE, V22, P4918, DOI 10.1038/sj.onc.1206753
   Shen H, 2013, CELL, V153, P38, DOI 10.1016/j.cell.2013.03.008
   Shirahata M, 2007, CLIN CANCER RES, V13, P7341, DOI 10.1158/1078-0432.CCR-06-2789
   Singh D, 2012, SCIENCE, V337, P1231, DOI 10.1126/science.1220834
   Sneeringer CJ, 2010, P NATL ACAD SCI USA, V107, P20980, DOI 10.1073/pnas.1012525107
   Snuderl M, 2011, CANCER CELL, V20, P810, DOI 10.1016/j.ccr.2011.11.005
   Sottoriva A, 2013, P NATL ACAD SCI USA, V110, P4009, DOI 10.1073/pnas.1219747110
   Srivastava A, 2010, CHILD NERV SYST, V26, P1613, DOI 10.1007/s00381-010-1214-y
   Stommel JM, 2007, SCIENCE, V318, P287, DOI 10.1126/science.1142946
   Stupp R, 2005, NEW ENGL J MED, V352, P987, DOI 10.1056/NEJMoa043330
   Stupp R, 2009, LANCET ONCOL, V10, P459, DOI 10.1016/S1470-2045(09)70025-7
   Sturm D, 2012, CANCER CELL, V22, P425, DOI 10.1016/j.ccr.2012.08.024
   Sugiarto S, 2011, CANCER CELL, V20, P328, DOI 10.1016/j.ccr.2011.08.011
   Swartling FJ, 2012, CANCER CELL, V21, P601, DOI 10.1016/j.ccr.2012.04.012
   Szerlip NJ, 2012, P NATL ACAD SCI USA, V109, P3041, DOI 10.1073/pnas.1114033109
   Tanaka S, 2013, NAT REV CLIN ONCOL, V10, P14, DOI 10.1038/nrclinonc.2012.204
   Tonjes M, 2013, NAT MED, V19, P901, DOI 10.1038/nm.3217
   Tso CL, 2006, CANCER RES, V66, P159, DOI 10.1158/0008-5472.CAN-05-0077
   Turcan S, 2012, NATURE, V483, P479, DOI 10.1038/nature10866
   van den Bent MJ, 2009, J CLIN ONCOL, V27, P1268, DOI 10.1200/JCO.2008.17.5984
   Venneti S, 2013, BRAIN PATHOL, V23, P558, DOI 10.1111/bpa.12042
   Verhaak RGW, 2010, CANCER CELL, V17, P98, DOI 10.1016/j.ccr.2009.12.020
   Vinagre J, 2013, NAT COMMUN, V4, DOI 10.1038/ncomms3185
   Vogt N, 2004, P NATL ACAD SCI USA, V101, P11368, DOI 10.1073/pnas.0402979101
   Waha A, 2005, NEOPLASIA, V7, P193, DOI 10.1593/neo.04490
   Wang F, 2013, SCIENCE, V340, P622, DOI 10.1126/science.1234769
   Wang Y, 2009, CANCER CELL, V15, P514, DOI 10.1016/j.ccr.2009.04.001
   Ward PS, 2010, CANCER CELL, V17, P225, DOI 10.1016/j.ccr.2010.01.020
   Warren KE, 2012, FRONT ONCOL, V2, DOI 10.3389/fonc.2012.00205
   Wei QX, 2006, CANCER RES, V66, P7429, DOI 10.1158/0008-5472.CAN-06-0712
   Weiss WA, 2003, CANCER RES, V63, P1589
   Weller M, 2011, NEUROLOGY, V77, P1156, DOI 10.1212/WNL.0b013e31822f02e1
   Wen PY, 2006, CLIN CANCER RES, V12, P4899, DOI 10.1158/1078-0432.CCR-06-0773
   Wick W, 2012, LANCET ONCOL, V13, P707, DOI 10.1016/S1470-2045(12)70164-X
   Wu G, 2012, NAT GENET, V44, P251, DOI 10.1038/ng.1102
   Xu W, 2011, CANCER CELL, V19, P17, DOI 10.1016/j.ccr.2010.12.014
   Yan H, 2009, NEW ENGL J MED, V360, P765, DOI 10.1056/NEJMoa0808710
   Yang WW, 2012, NAT CELL BIOL, V14, P1295, DOI 10.1038/ncb2629
   Yang WW, 2012, CELL, V150, P685, DOI 10.1016/j.cell.2012.07.018
   Yip S, 2009, CLIN CANCER RES, V15, P4622, DOI 10.1158/1078-0432.CCR-08-3012
   Zardo G, 2002, NAT GENET, V32, P453, DOI 10.1038/ng1007
   Zarghooni M, 2010, J CLIN ONCOL, V28, P1337, DOI 10.1200/JCO.2009.25.5463
   Zhang CZ, 2013, NEURO-ONCOLOGY, V15, P1114, DOI 10.1093/neuonc/not087
   Zhang JH, 2013, NAT GENET, V45, P602, DOI 10.1038/ng.2611
   Zhao YG, 2010, AM J PATHOL, V176, P3032, DOI 10.2353/ajpath.2010.090571
   Zheng SY, 2013, GENE DEV, V27, P1462, DOI 10.1101/gad.213686.113
   Zheng SY, 2012, CANCER J, V18, P107, DOI 10.1097/PPO.0b013e3182431c57
   Zhou H, 2007, CLIN CANCER RES, V13, P2344, DOI 10.1158/1078-0432.CCR-06-2303
   Zhu H, 2009, P NATL ACAD SCI USA, V106, P2712, DOI 10.1073/pnas.0813314106
   Zhu Y, 2005, CANCER CELL, V8, P119, DOI 10.1016/j.ccr.2005.07.004
NR 224
TC 384
Z9 392
U1 1
U2 63
PU NATURE PORTFOLIO
PI BERLIN
PA HEIDELBERGER PLATZ 3, BERLIN, 14197, GERMANY
SN 1474-175X
EI 1474-1768
J9 NAT REV CANCER
JI Nat. Rev. Cancer
PD FEB
PY 2014
VL 14
IS 2
BP 92
EP 107
DI 10.1038/nrc3655
PG 16
WC Oncology
WE Science Citation Index Expanded (SCI-EXPANDED)
SC Oncology
GA AA4AW
UT WOS:000331037400007
PM 24457416
OA Green Submitted, Green Accepted
HC Y
HP N
DA 2023-08-27
ER

PT J
AU Glasow, A
   Haidan, A
   Hilbers, U
   Breidert, M
   Gillespie, J
   Scherbaum, WA
   Chrousos, GP
   Bornstein, SR
AF Glasow, A
   Haidan, A
   Hilbers, U
   Breidert, M
   Gillespie, J
   Scherbaum, WA
   Chrousos, GP
   Bornstein, SR
TI Expression of Ob receptor in normal human adrenals: Differential
   regulation of adrenocortical and adrenomedullary function by leptin
SO JOURNAL OF CLINICAL ENDOCRINOLOGY & METABOLISM
LA English
DT Article
ID PANCREATIC BETA-CELLS; LASER CAPTURE MICRODISSECTION;
   TUMOR-NECROSIS-FACTOR; GENE-EXPRESSION; PLASMA LEPTIN; ADIPOSE-TISSUE;
   INSULIN-RESISTANCE; MESSENGER-RNA; FOOD-INTAKE; OBESE GENE
AB The major effects of leptin, an adipostatic hormone produced in fat tissue, are exerted through the hypothalamic-pituitary-adrenal axis and the systemic sympathetic/adrenomedullary system at the level of the central nervous system. Here, we examined the direct effects of leptin on the adrenal gland, a peripheral end organ of both the hypothalamic-pituitary-adrenal axis and the sympathetic/adrenomedullary system. As cortical and chromaffin tissues are intermingled in the human adrenal, we employed the novel technique of laser capture microdissection to analyze these systems separately. Functional full-length leptin receptor messenger ribonucleic acid and all human isoforms Ob219.1-3 were demonstrated by RT-PCR in both cortical and medullary tissue. Immunohistochemical staining of leptin receptor protein, however, demonstrated a strong signal only in the adrenal cortex, whereas there was weak positive staining in the medulla. Corticotropin (ACTH)-induced adrenal aldosterone, cortisol, and dehydroepiandrosterone secretion was inhibited by leptin in a concentration-dependent manner, whereas this hormone had no significant effect on catecholamine release by primary cultures of human adrenal chromaffin cells. Leptin itself was not expressed in human adrenal tissue, excluding a local paracrine or autocrine function of this peptide.
   In conclusion, this is the first report identifying functional leptin receptor in human adrenal tissue and showing a differential action of leptin on human adrenocortical and chromaffin hormone production. This peripheral action of leptin on the adrenal gland provides an additional important link between the human stress response and body weight regulation.
C1 Univ Leipzig, Dept Internal Med 3, D-04103 Leipzig, Germany.
   Carl Gustav Carus TU Dresden, Fac Med, Dresden, Germany.
   Univ Dusseldorf, Diabet Res Inst, Dusseldorf, Germany.
   NCI, Dept Pathol, NIH, Bethesda, MD 20892 USA.
   NICHHD, Dev Endocrinol Branch, NIH, Bethesda, MD 20892 USA.
C3 ARQUS; Leipzig University; Technische Universitat Dresden; Carl Gustav
   Carus University Hospital; Heinrich Heine University Dusseldorf;
   National Institutes of Health (NIH) - USA; NIH National Cancer Institute
   (NCI); National Institutes of Health (NIH) - USA; NIH Eunice Kennedy
   Shriver National Institute of Child Health & Human Development (NICHD)
RP Glasow, A (通讯作者)，Univ Leipzig, Med Klin & Poliklin 3, Philipp Rosenthal Str 27, D-04103 Leipzig, Germany.
EM
CR Ahima RS, 1996, NATURE, V382, P250, DOI 10.1038/382250a0
   ANANDSRIVASTAVA MB, 1985, FEBS LETT, V181, P199, DOI 10.1016/0014-5793(85)80259-3
   Bennett BD, 1996, CURR BIOL, V6, P1170, DOI 10.1016/S0960-9822(02)70684-2
   Blum WF, 1997, J CLIN ENDOCR METAB, V82, P2904, DOI 10.1210/jc.82.9.2904
   Bonner RF, 1997, SCIENCE, V278, P1481, DOI 10.1126/science.278.5342.1481
   BORNSTEIN SR, 1992, ENDOCRINOLOGY, V131, P3126, DOI 10.1210/en.131.6.3126
   BORNSTEIN SR, 1994, J CLIN ENDOCR METAB, V78, P225, DOI 10.1210/jc.78.1.225
   Bornstein SR, 1997, DIABETES, V46, P1235, DOI 10.2337/diabetes.46.7.1235
   Bornstein SR, 1998, J CLIN ENDOCR METAB, V83, P280, DOI 10.1210/jc.83.1.280
   Bornstein SR, 1997, NAT MED, V3, P937, DOI 10.1038/nm0997-937
   BRANDS MW, 1989, AM J PHYSIOL, V257, pR1423, DOI 10.1152/ajpregu.1989.257.6.R1423
   Campfield LA, 1996, HORM METAB RES, V28, P619, DOI 10.1055/s-2007-979867
   Cao GY, 1997, AM J PHYSIOL-ENDOC M, V273, pE448, DOI 10.1152/ajpendo.1997.273.2.E448
   Cinti S, 1997, ENDOCRINOLOGY, V138, P797, DOI 10.1210/en.138.2.797
   Cioffi JA, 1996, NAT MED, V2, P585, DOI 10.1038/nm0596-585
   Cizza G, 1997, Cancer Treat Res, V89, P25
   Cohen B, 1996, SCIENCE, V274, P1185, DOI 10.1126/science.274.5290.1185
   Cohen SL, 1996, NATURE, V382, P589, DOI 10.1038/382589a0
   Collins S, 1996, NATURE, V380, P677, DOI 10.1038/380677a0
   Costa A, 1997, NEUROREPORT, V8, P1131, DOI 10.1097/00001756-199703240-00014
   Dallman MF, 1995, ANN NY ACAD SCI, V771, P730, DOI 10.1111/j.1749-6632.1995.tb44724.x
   DEVOS P, 1995, J BIOL CHEM, V270, P15958, DOI 10.1074/jbc.270.27.15958
   Ehrhart-Bornstein M, 1998, ENDOCR REV, V19, P101, DOI 10.1210/er.19.2.101
   EHRHARTBORNSTEIN M, 1991, J ENDOCRINOL, V131, pR5, DOI 10.1677/joe.0.131R005
   Emilsson V, 1997, DIABETES, V46, P313, DOI 10.2337/diabetes.46.2.313
   EmmertBuck MR, 1996, SCIENCE, V274, P998, DOI 10.1126/science.274.5289.998
   Friedman TC, 1996, ENDOCR J, V43, P645, DOI 10.1507/endocrj.43.645
   Glasow A, 1996, J CLIN ENDOCR METAB, V81, P3103, DOI 10.1210/jc.81.8.3103
   GonzalezHernandez JA, 1996, J CLIN ENDOCR METAB, V81, P807, DOI 10.1210/jc.81.2.807
   GONZALEZHERNANDEZ JA, 1995, CLIN EXP IMMUNOL, V99, P137
   GONZALEZHERNANDEZ JA, 1994, CELL TISSUE RES, V278, P201, DOI 10.1007/BF00414161
   GUILLAUMEGENTIL C, 1993, DIABETOLOGIA, V36, P899, DOI 10.1007/BF02374470
   GUSEBEHLING H, 1992, J ENDOCRINOL, V135, P229, DOI 10.1677/joe.0.1350229
   Hardie LJ, 1996, HORM METAB RES, V28, P685, DOI 10.1055/s-2007-979878
   HAUNER H, 1989, J CLIN INVEST, V84, P1663, DOI 10.1172/JCI114345
   Havel PJ, 1996, NAT MED, V2, P949, DOI 10.1038/nm0996-949b
   Haynes WG, 1997, HYPERTENSION, V30, P619, DOI 10.1161/01.HYP.30.3.619
   Haynes WG, 1997, J CLIN INVEST, V100, P270, DOI 10.1172/JCI119532
   Heiman ML, 1997, ENDOCRINOLOGY, V138, P3859, DOI 10.1210/en.138.9.3859
   Hoggard N, 1997, BIOCHEM BIOPH RES CO, V232, P383, DOI 10.1006/bbrc.1997.6245
   Jackson Edwin K., 1996, Hypertension (Dallas), V28, P517
   JUDD AM, 1995, AM J PHYSIOL-ENDOC M, V268, pE114, DOI 10.1152/ajpendo.1995.268.1.E114
   Karlsson C, 1997, J CLIN ENDOCR METAB, V82, P4144, DOI 10.1210/jc.82.12.4144
   Kieffer TJ, 1996, BIOCHEM BIOPH RES CO, V224, P522, DOI 10.1006/bbrc.1996.1059
   Kieffer TJ, 1997, DIABETES, V46, P1087, DOI 10.2337/diabetes.46.6.1087
   Kielar D, 1998, METABOLISM, V47, P844, DOI 10.1016/S0026-0495(98)90124-X
   Kutoh E, 1997, LIFE SCI, V62, P445, DOI 10.1016/S0024-3205(97)01138-7
   LAEMMLI UK, 1970, NATURE, V227, P680, DOI 10.1038/227680a0
   LealCerro A, 1996, HORM METAB RES, V28, P711, DOI 10.1055/s-2007-979884
   Lee GH, 1996, NATURE, V379, P632, DOI 10.1038/379632a0
   Licinio J, 1997, NAT MED, V3, P575, DOI 10.1038/nm0597-575
   Mantzoros CS, 1996, DIABETES, V45, P909, DOI 10.2337/diabetes.45.7.909
   Martin LJ, 1998, CAN J PHYSIOL PHARM, V76, P237, DOI 10.1139/cjpp-76-2-237
   Masuzaki H, 1997, NAT MED, V3, P1029, DOI 10.1038/nm0997-1029
   Miell JP, 1996, HORM METAB RES, V28, P704, DOI 10.1055/s-2007-979882
   Miller SG, 1996, P NATL ACAD SCI USA, V93, P5507, DOI 10.1073/pnas.93.11.5507
   Murakami T, 1997, BIOCHEM BIOPH RES CO, V231, P26, DOI 10.1006/bbrc.1996.6030
   NOSADINI R, 1983, J CLIN ENDOCR METAB, V57, P529, DOI 10.1210/jcem-57-3-529
   PapaspyrouRao S, 1997, J CLIN ENDOCR METAB, V82, P1635, DOI 10.1210/jc.82.5.1635
   Path G, 1997, J CLIN ENDOCR METAB, V82, P2343, DOI 10.1210/jc.82.7.2343
   Reaven GM, 1996, NEW ENGL J MED, V334, P374, DOI 10.1056/NEJM199602083340607
   Salbe AD, 1997, J CLIN INVEST, V99, P592, DOI 10.1172/JCI119200
   Schwartz MW, 1997, J AM DIET ASSOC, V97, P54, DOI 10.1016/S0002-8223(97)00018-7
   Senaris R, 1997, ENDOCRINOLOGY, V138, P4501, DOI 10.1210/en.138.10.4501
   Shek EW, 1998, HYPERTENSION, V31, P409, DOI 10.1161/01.HYP.31.1.409
   Slieker LJ, 1996, J BIOL CHEM, V271, P5301, DOI 10.1074/jbc.271.10.5301
   SOWERS JR, 1984, LIFE SCI, V34, P2339, DOI 10.1016/0024-3205(84)90420-X
   STEPHENS TW, 1995, NATURE, V377, P530, DOI 10.1038/377530a0
   Tanizawa Y, 1997, ENDOCRINOLOGY, V138, P4513, DOI 10.1210/en.138.10.4513
   Tartaglia LA, 1995, CELL, V83, P1263, DOI 10.1016/0092-8674(95)90151-5
   Tataranni PA, 1996, AM J PHYSIOL-ENDOC M, V271, pE317, DOI 10.1152/ajpendo.1996.271.2.E317
   TRAYHURN P, 1995, BIOCHEM J, V311, P729, DOI 10.1042/bj3110729
   Tuominen JA, 1997, ACTA PHYSIOL SCAND, V160, P83, DOI 10.1046/j.1365-201X.1997.00102.x
   Vaisse C, 1996, NAT GENET, V14, P95, DOI 10.1038/ng0996-95
   Wabitsch M, 1996, DIABETES, V45, P1435, DOI 10.2337/diabetes.45.10.1435
   Wang JL, 1998, NATURE, V393, P684, DOI 10.1038/31474
   Wang YP, 1997, J BIOL CHEM, V272, P16216, DOI 10.1074/jbc.272.26.16216
   White DW, 1997, J BIOL CHEM, V272, P4065, DOI 10.1074/jbc.272.7.4065
   Widjaja A, 1997, J CLIN ENDOCR METAB, V82, P654, DOI 10.1210/jc.82.2.654
   WURTMAN RJ, 1966, J BIOL CHEM, V241, P2301
   YOUNG JB, 1984, AM J PHYSIOL, V247, pE35, DOI 10.1152/ajpendo.1984.247.1.E35
   Zachow RJ, 1997, ENDOCRINOLOGY, V138, P847, DOI 10.1210/en.138.2.847
   ZHANG YY, 1994, NATURE, V372, P425, DOI 10.1038/372425a0
   ZUBER MX, 1986, J BIOL CHEM, V261, P2475
   ZYSSER RD, 1981, J CLIN ENDOCR METAB, V53, P867
NR 85
TC 156
Z9 162
U1 0
U2 3
PU ENDOCRINE SOC
PI WASHINGTON
PA 2055 L ST NW, SUITE 600, WASHINGTON, DC 20036 USA
SN 0021-972X
EI 1945-7197
J9 J CLIN ENDOCR METAB
JI J. Clin. Endocrinol. Metab.
PD DEC
PY 1998
VL 83
IS 12
BP 4459
EP 4466
DI 10.1210/jc.83.12.4459
PG 8
WC Endocrinology & Metabolism
WE Science Citation Index Expanded (SCI-EXPANDED)
SC Endocrinology & Metabolism
GA 145FB
UT WOS:000077359400049
PM 9851794
DA 2023-08-27
ER

PT J
AU Taniai, E
   Hayashi, H
   Yafune, A
   Watanabe, M
   Akane, H
   Suzuki, K
   Mitsumori, K
   Shibutani, M
AF Taniai, Eriko
   Hayashi, Hitomi
   Yafune, Atsunori
   Watanabe, Maiko
   Akane, Hirotoshi
   Suzuki, Kazuhiko
   Mitsumori, Kunitoshi
   Shibutani, Makoto
TI Cellular distribution of cell cycle-related molecules in the renal
   tubules of rats treated with renal carcinogens for 28 days: relationship
   between cell cycle aberration and carcinogenesis
SO ARCHIVES OF TOXICOLOGY
LA English
DT Article
DE Renal carcinogenesis; Karyomegaly; Cell cycle; Cell proliferation;
   Immunohistochemistry
ID TOPOISOMERASE-II-ALPHA; IN-VIVO MUTAGENICITY; OCHRATOXIN-A; MADDER
   COLOR; TUMORS; TOXICITY; PROTEIN; PROLIFERATION; DISSOCIATION;
   NEPHROPATHY
AB Some renal carcinogens can induce karyomegaly, which reflects aberrant cell division in the renal tubules, from the early stages of exposure. To clarify the cell cycle-related changes during the early stages of renal carcinogenesis, we performed immunohistochemical analysis of tubular cells in male F344 rats treated with carcinogenic doses of representative renal carcinogens for 28 days. For this purpose, the karyomegaly-inducing carcinogens ochratoxin A (OTA), ferric nitrilotriacetic acid, and monuron, and the non-karyomegaly-inducing carcinogens tris(2-chloroethyl) phosphate and potassium bromate were examined. For comparison, a karyomegaly-inducing non-carcinogen, p-nitrobenzoic acid, and a non-carcinogenic non-karyomegaly-inducing renal toxicant, acetaminophen, were also examined. The outer stripe of the outer medulla (OSOM) and the cortex + OSOM were subjected to morphometric analysis of immunoreactive proximal tubular cells. Renal carcinogens, irrespective of their karyomegaly-inducing potential, increased proximal tubular cell proliferation accompanied by an increase in topoisomerase II alpha-immunoreactive cells, suggesting a reflection of cell proliferation. Karyomegaly-inducing carcinogens increased nuclear Cdc2-, gamma H2AX-, and phosphorylated Chk2-immunoreactive cells in both areas, the former two acting in response to DNA damage and the latter one suggestive of sustained G(2). OTA, an OSOM-targeting carcinogen, could easily be distinguished from untreated controls and non-carcinogens by evaluation of molecules responding to DNA damage and G(2)/M transition in the OSOM. Thus, all renal carcinogens examined facilitated proximal tubular proliferation by repeated short-term treatment. Among these, karyomegaly-inducing carcinogens may cause DNA damage and G(2) arrest in the target tubular cells.
C1 [Taniai, Eriko; Hayashi, Hitomi; Yafune, Atsunori; Akane, Hirotoshi; Suzuki, Kazuhiko; Mitsumori, Kunitoshi; Shibutani, Makoto] Tokyo Univ Agr & Technol, Lab Vet Pathol, Fuchu, Tokyo 1838509, Japan.
   [Taniai, Eriko; Hayashi, Hitomi; Yafune, Atsunori] Gifu Univ, United Grad Sch Vet Sci, Gifu 5011193, Japan.
   [Watanabe, Maiko] Natl Inst Hlth Sci, Div Microbiol, Setagaya Ku, Tokyo 1588501, Japan.
C3 Tokyo University of Agriculture & Technology; Gifu University; National
   Institute of Health Sciences - Japan
RP Shibutani, M (通讯作者)，Tokyo Univ Agr & Technol, Lab Vet Pathol, 3-5-8 Saiwai Cho, Fuchu, Tokyo 1838509, Japan.
EM
RI Suzuki, Kazuhiko/C-8348-2013; Shibutani, Makoto/C-2510-2013; Shibutani,
   Makoto/AGK-2801-2022
OI Suzuki, Kazuhiko/0000-0001-6047-8127; Shibutani,
   Makoto/0000-0003-3417-9697; MITSUMORI, KUNITOSHI/0000-0002-0754-572X
FU Ministry of Education, Culture, Sports, Science and Technology of Japan;
   Grants-in-Aid for Scientific Research [11J03954, 22380163] Funding
   Source: KAKEN
FX The authors thank Mrs. Shigeko Suzuki for her technical assistance in
   preparing the histological specimens. This work was supported by a
   Grant-in-Aid for Scientific Research (B) from the Ministry of Education,
   Culture, Sports, Science and Technology of Japan. Eriko Taniai is a
   Research Fellow of the Japan Society for the Promotion of Science.
CR Adler M, 2009, CARCINOGENESIS, V30, P711, DOI 10.1093/carcin/bgp049
   Allen DG, 2004, TOXICOL PATHOL, V32, P393, DOI 10.1080/01926230490440934
   Ashby J, 1992, IARC Sci Publ, P135
   BOORMAN GA, 1992, TOXICOL PATHOL, V20, P236, DOI 10.1177/019262339202000210
   Bower JJ, 2010, ONCOGENE, V29, P4787, DOI 10.1038/onc.2010.232
   Brizova H, 2010, VIRCHOWS ARCH, V456, P671, DOI 10.1007/s00428-010-0922-8
   Brown AL, 2007, EXP TOXICOL PATHOL, V59, P85, DOI 10.1016/j.etp.2007.05.001
   Chan TA, 1999, NATURE, V401, P616, DOI 10.1038/44188
   CLAWSON GA, 1992, CANCER RES, V52, P1304
   Doi AM, 2007, TOXICOL PATHOL, V35, P533, DOI 10.1080/01926230701338941
   Eastin WC, 1998, ENVIRON HEALTH PERSP, V106, P81, DOI 10.2307/3433914
   Hard GC, 2000, TOXICOL SCI, V53, P237, DOI 10.1093/toxsci/53.2.237
   Hibi D, 2011, TOXICOL SCI, V122, P406, DOI 10.1093/toxsci/kfr139
   Hirota T, 2005, NATURE, V438, P1176, DOI 10.1038/nature04254
   Hu ML, 2002, FREE RADICAL RES, V36, P203, DOI 10.1080/10715760290006420
   Inoue K, 2008, FOOD CHEM TOXICOL, V46, P241, DOI 10.1016/j.fct.2007.08.002
   Inoue K, 2009, FOOD CHEM TOXICOL, V47, P752, DOI 10.1016/j.fct.2009.01.003
   Inoue K, 2009, FOOD CHEM TOXICOL, V47, P184, DOI 10.1016/j.fct.2008.10.031
   Kobayashi J, 2008, J RADIAT RES, V49, P93, DOI 10.1269/jrr.07130
   Kumata K., 1980, SHOKUHIN EISEIGAKU Z, V21, P171
   KUROKAWA Y, 1990, ENVIRON HEALTH PERSP, V87, P309, DOI 10.2307/3431039
   LI JL, 1987, CANCER RES, V47, P1867
   Lock EA, 2004, CRIT REV TOXICOL, V34, P211, DOI 10.1080/10408440490265210
   Luo KT, 2009, NAT CELL BIOL, V11, P204, DOI 10.1038/ncb1828
   Mattila R, 2007, ARCH DERMATOL RES, V298, P381, DOI 10.1007/s00403-006-0711-z
   National Toxicology Program, 1996, Natl Toxicol Program Tech Rep Ser, V383, P1
   National Toxicology Program, 1988, Natl Toxicol Program Tech Rep Ser, V266, P1
   National Toxicology Program, 1994, Natl Toxicol Program Tech Rep Ser, V442, P1
   National Toxicology Program, 1993, Natl Toxicol Program Tech Rep Ser, V394, P1
   National Toxicology Program, 1989, Natl Toxicol Program Tech Rep Ser, V358, P1
   Niida H, 2006, MUTAGENESIS, V21, P3, DOI 10.1093/mutage/gei063
   Nishiyama Y, 1995, JPN J CANCER RES, V86, P1150, DOI 10.1111/j.1349-7006.1995.tb03308.x
   Ntp, 1991, NATL TOXICOL PROGRAM, V391, P1
   PARODI S, 1992, TOXICOL LETT, V64-5, P621, DOI 10.1016/0378-4274(92)90240-K
   Pentheroudakis G, 2010, LEUKEMIA LYMPHOMA, V51, P1260, DOI 10.3109/10428194.2010.483749
   Perez de Castro I, 2007, CARCINOGENESIS, V28, P899, DOI 10.1093/carcin/bgm019
   Rached E, 2007, TOXICOL SCI, V97, P288, DOI 10.1093/toxsci/kfm042
   Ruchaud S, 2007, NAT REV MOL CELL BIO, V8, P798, DOI 10.1038/nrm2257
   Scholzen T, 2000, J CELL PHYSIOL, V182, P311, DOI 10.1002/(SICI)1097-4652(200003)182:3<311::AID-JCP1>3.0.CO;2-9
   Seely JC, 2008, J TOXICOL PATHOL, V21, P199, DOI 10.1293/tox.21.199
   SHERR CJ, 1995, TRENDS BIOCHEM SCI, V20, P187, DOI 10.1016/S0968-0004(00)89005-2
   Snyderwine EG, 1999, CANCER LETT, V143, P211, DOI 10.1016/S0304-3835(99)00127-5
   Sugita-Konishi Y, 2006, TALANTA, V69, P650, DOI 10.1016/j.talanta.2005.10.036
   Tamano Seiko, 2010, Asian Pac J Cancer Prev, V11, P4
   Tanaka T, 2000, ONCOL REP, V7, P501
   TOYOKUNI S, 1994, P NATL ACAD SCI USA, V91, P2616, DOI 10.1073/pnas.91.7.2616
   Uhlmann F, 2004, EXP CELL RES, V296, P80, DOI 10.1016/j.yexcr.2004.03.005
   Umemura T, 2006, CANCER SCI, V97, P829, DOI 10.1111/j.1349-7006.2006.00248.x
   Wang Y, 2008, MOL BIOL CELL, V19, P4076, DOI 10.1091/mbc.E07-12-1296
   Williams GM, 2002, EUR J CANCER PREV, V11, P39, DOI 10.1097/00008469-200202000-00006
   Wilson DW, 2000, TOXICOL APPL PHARM, V166, P69, DOI 10.1006/taap.2000.8966
   Woo RA, 2003, CELL CYCLE, V2, P316, DOI 10.4161/cc.2.4.468
   Yoshida K, 2006, MOL CELL BIOL, V26, P3414, DOI 10.1128/MCB.26.9.3414-3431.2006
NR 53
TC 15
Z9 15
U1 0
U2 6
PU SPRINGER HEIDELBERG
PI HEIDELBERG
PA TIERGARTENSTRASSE 17, D-69121 HEIDELBERG, GERMANY
SN 0340-5761
EI 1432-0738
J9 ARCH TOXICOL
JI Arch. Toxicol.
PD SEP
PY 2012
VL 86
IS 9
BP 1453
EP 1464
DI 10.1007/s00204-012-0829-z
PG 12
WC Toxicology
WE Science Citation Index Expanded (SCI-EXPANDED)
SC Toxicology
GA 992HP
UT WOS:000307766500014
PM 22411272
DA 2023-08-27
ER

PT J
AU Braun, TP
   Marks, DL
AF Braun, Theodore P.
   Marks, Daniel L.
TI Hypothalamic regulation of muscle metabolism
SO CURRENT OPINION IN CLINICAL NUTRITION AND METABOLIC CARE
LA English
DT Article
DE cachexia; hypothalamus; leptin; muscle; sympathetic
ID ACTIVATED PROTEIN-KINASE; TUMOR-NECROSIS-FACTOR; CELL LUNG-CANCER;
   SKELETAL-MUSCLE; PROOPIOMELANOCORTIN NEURONS; LEPTIN ACTIVATION;
   GENE-PRODUCT; FACTOR-ALPHA; OBESE GENE; CACHEXIA
AB Purpose of review
   The interest in obesity research has produced a large body of data describing the impact of neuronal signaling in the hypothalamus and brainstem on metabolic regulation in the periphery. Studies have historically focused on central regulation of metabolism in adipose and hepatic tissue. Recent studies highlight an important role for these same central regulatory centers in the control of muscle metabolism. This review will focus on these new studies, and will highlight the implications of these new data for the study of muscle catabolism in disease states.
   Recent findings
   The balance of anabolism and catabolism in muscle requires activation of the hypothalamic-pituitary-adrenal axis as well as changes in energy-dependent signaling pathways in the muscle. It is now apparent that the sympathetic nervous system conveys much of this information between key metabolism-regulating nuclei in the hypothalamus and skeletal muscle.
   Summary
   Peripheral signals conveying information regarding the metabolic status of the animal appear to alter the function of metabolic centers in the brain that in turn regulate energy partitioning in muscle via a sympathetic relay. Our understanding of how this system is regulated in normal physiological states and in obesity is providing important clues for understanding muscle catabolism in disease.
C1 [Braun, Theodore P.; Marks, Daniel L.] Oregon Hlth & Sci Univ, Dept Pediat, Pape Family Pediat Res Inst, Portland, OR 97201 USA.
C3 Oregon Health & Science University
RP Marks, DL (通讯作者)，Mailcode L 481,3181 SW Sam Jackson Pk Rd, Portland, OR 97239 USA.
EM
RI Marks, Daniel/C-3712-2009
OI Braun, Theodore/0000-0002-6248-2143
CR Ahima RS, 1996, NATURE, V382, P250, DOI 10.1038/382250a0
   Babic T, 2010, BRAIN RES, V1345, P146, DOI 10.1016/j.brainres.2010.05.042
   BARACOS V, 1983, NEW ENGL J MED, V308, P553, DOI 10.1056/NEJM198303103081002
   Baskin DG, 1999, DIABETES, V48, P828, DOI 10.2337/diabetes.48.4.828
   Belgardt BF, 2010, ANN NY ACAD SCI, V1212, P97, DOI 10.1111/j.1749-6632.2010.05799.x
   Bjorbaek C, 2009, J INVEST MED, V57, P789, DOI 10.2310/JIM.0b013e3181bb0d49
   Cheung CC, 1997, ENDOCRINOLOGY, V138, P4489, DOI 10.1210/en.138.10.4489
   Elmquist JK, 1997, ENDOCRINOLOGY, V138, P839, DOI 10.1210/en.138.2.839
   Evans WJ, 2008, CLIN NUTR, V27, P793, DOI 10.1016/j.clnu.2008.06.013
   EVANS WK, 1985, CANCER RES, V45, P3347
   Fan W, 1997, NATURE, V385, P165, DOI 10.1038/385165a0
   Finck BN, 1998, ENDOCRINOLOGY, V139, P2278, DOI 10.1210/en.139.5.2278
   Finck BN, 1999, NEUROREPORT, V10, P153, DOI 10.1097/00001756-199901180-00029
   GELIN J, 1991, CANCER RES, V51, P415
   Glaum SR, 1996, MOL PHARMACOL, V50, P230
   Grill HJ, 2010, FRONT NEUROENDOCRIN, V31, P61, DOI 10.1016/j.yfrne.2009.10.005
   Grossberg AJ, 2010, PHYSIOL BEHAV, V100, P478, DOI 10.1016/j.physbeh.2010.03.011
   Grossberg AJ, 2010, ENDOCRINOLOGY, V151, P606, DOI 10.1210/en.2009-1135
   Grunfeld C, 1996, J CLIN INVEST, V97, P2152, DOI 10.1172/JCI118653
   Halaas JL, 1997, P NATL ACAD SCI USA, V94, P8878, DOI 10.1073/pnas.94.16.8878
   Hu ZY, 2009, J CLIN INVEST, V119, P3059, DOI 10.1172/JCI38770
   Jacob RJ, 1997, DIABETES, V46, P150, DOI 10.2337/diabetes.46.1.150
   Kahn BB, 2005, CELL METAB, V1, P15, DOI 10.1016/j.cmet.2004.12.003
   Kishi K, 2000, BIOCHEM BIOPH RES CO, V276, P16, DOI 10.1006/bbrc.2000.3417
   Krawiec BJ, 2005, AM J PHYSIOL-ENDOC M, V289, pE969, DOI 10.1152/ajpendo.00126.2005
   Krawiec BJ, 2007, AM J PHYSIOL-ENDOC M, V292, pE1555, DOI 10.1152/ajpendo.00622.2006
   Lee SJ, 2005, P NATL ACAD SCI USA, V102, P18117, DOI 10.1073/pnas.0505996102
   Lee SJ, 1999, CURR OPIN GENET DEV, V9, P604, DOI 10.1016/S0959-437X(99)00004-0
   Lee SJ, 2001, P NATL ACAD SCI USA, V98, P9306, DOI 10.1073/pnas.151270098
   Li W, 2009, AM J PHYSIOL-CELL PH, V297, pC706, DOI 10.1152/ajpcell.00626.2008
   Minokoshi Y, 2002, NATURE, V415, P339, DOI 10.1038/415339a
   MOLDAWER LL, 1987, AM J PHYSIOL, V253, pC766, DOI 10.1152/ajpcell.1987.253.6.C766
   MOLDAWER LL, 1988, AM J PHYSIOL, V254, pG450, DOI 10.1152/ajpgi.1988.254.3.G450
   Morley JE, 2006, AM J CLIN NUTR, V83, P735, DOI 10.1093/ajcn/83.4.735
   Moylan JS, 2008, AM J PHYSIOL-CELL PH, V295, pC986, DOI 10.1152/ajpcell.00041.2008
   Munzberg H, 2010, FORUM NUTR, V63, P123, DOI 10.1159/000264400
   Muscaritoli M, 2010, CLIN NUTR, V29, P154, DOI 10.1016/j.clnu.2009.12.004
   Niswender KD, 2001, NATURE, V413, P794, DOI 10.1038/35101657
   Nogueiras R, 2007, J CLIN INVEST, V117, P3475, DOI 10.1172/JCI31743
   PLATA-SALAMAN C R, 1989, Brain Behavior and Immunity, V3, P193, DOI 10.1016/0889-1591(89)90036-6
   Sanchez-Lasheras C, 2010, FRONT NEUROENDOCRIN, V31, P4, DOI 10.1016/j.yfrne.2009.08.002
   Sarraf P, 1997, J EXP MED, V185, P171, DOI 10.1084/jem.185.1.171
   Sayk F, 2010, J CLIN ENDOCR METAB, V95, P1998, DOI 10.1210/jc.2009-2297
   Scarlett JM, 2008, ENDOCRINOLOGY, V149, P4837, DOI 10.1210/en.2007-1680
   Scarlett JM, 2007, ENDOCRINOLOGY, V148, P4217, DOI 10.1210/en.2007-0017
   Scott HR, 1996, BRIT J CANCER, V73, P1560, DOI 10.1038/bjc.1996.294
   Shiuchi T, 2009, CELL METAB, V10, P466, DOI 10.1016/j.cmet.2009.09.013
   Souza TA, 2008, MOL ENDOCRINOL, V22, P2689, DOI 10.1210/me.2008-0290
   Tanaka T, 2007, CELL METAB, V5, P395, DOI 10.1016/j.cmet.2007.04.004
   Toda C, 2009, DIABETES, V58, P2757, DOI 10.2337/db09-0638
   Vaisse C, 1996, NAT GENET, V14, P95, DOI 10.1038/ng0996-95
   Wisse BE, 2001, ENDOCRINOLOGY, V142, P3292, DOI 10.1210/en.142.8.3292
   Woods AJ, 1996, NATURE, V381, P745, DOI 10.1038/381745a0
   Zhang FM, 1997, NATURE, V387, P206, DOI 10.1038/387206a0
   ZHANG YY, 1994, NATURE, V372, P425, DOI 10.1038/372425a0
   Zhou XL, 2010, CELL, V142, P531, DOI 10.1016/j.cell.2010.07.011
NR 56
TC 13
Z9 13
U1 0
U2 3
PU LIPPINCOTT WILLIAMS & WILKINS
PI PHILADELPHIA
PA 530 WALNUT ST, PHILADELPHIA, PA 19106-3621 USA
SN 1363-1950
J9 CURR OPIN CLIN NUTR
JI Curr. Opin. Clin. Nutr. Metab. Care
PD MAY
PY 2011
VL 14
IS 3
BP 237
EP 242
DI 10.1097/MCO.0b013e328345bbcd
PG 6
WC Endocrinology & Metabolism; Nutrition & Dietetics
WE Science Citation Index Expanded (SCI-EXPANDED)
SC Endocrinology & Metabolism; Nutrition & Dietetics
GA 749XR
UT WOS:000289505700004
PM 21502918
DA 2023-08-27
ER

PT J
AU Pearson, ADJ
   Herold, R
   Rousseau, R
   Copland, C
   Bradley-Garelik, B
   Binner, D
   Capdeville, R
   Caron, H
   Carleer, J
   Chesler, L
   Geoerger, B
   Kearns, P
   Marshall, LV
   Pfister, SM
   Schleiermacher, G
   Skolnik, J
   Spadoni, C
   Sterba, J
   van den Berg, H
   Uttenreuther-Fischer, M
   Witt, O
   Norga, K
   Vassal, G
AF Pearson, Andrew D. J.
   Herold, Ralf
   Rousseau, Raphael
   Copland, Chris
   Bradley-Garelik, Brigid
   Binner, Debbie
   Capdeville, Renaud
   Caron, Hubert
   Carleer, Jacqueline
   Chesler, Louis
   Geoerger, Birgit
   Kearns, Pamela
   Marshall, Lynley V.
   Pfister, Stefan M.
   Schleiermacher, Gudrun
   Skolnik, Jeffrey
   Spadoni, Cesare
   Sterba, Jaroslav
   van den Berg, Hendrick
   Uttenreuther-Fischer, Martina
   Witt, Olaf
   Norga, Koen
   Vassal, Gilles
CA Working Grp Paediat Platform ACCEL
TI Implementation of mechanism of action biology-driven early drug
   development for children with cancer
SO EUROPEAN JOURNAL OF CANCER
LA English
DT Article
DE Paediatric oncology; Mechanisrn of action; Targeted cancer drug
   development
ID ACTIVATING ACVR1 MUTATIONS; INTRINSIC PONTINE GLIOMA; LOW-GRADE GLIOMAS;
   CHILDHOOD-CANCER; PEDIATRIC-PATIENTS; ADOLESCENTS; LANDSCAPE; RELAPSE;
   TRIALS; TUMORS
AB An urgent need remains for new paediatric oncology drugs to cure children who die from cancer and to reduce drug-related sequelae in survivors. In 2007, the European Paediatric Regulation came into law requiring industry to create paediatric drug (all types of medicinal products) development programmes alongside those for adults. Unfortunately, paediatric drug development is still largely centred on adult conditions and not a mechanism of action (MoA)-based model, even though this would be more logical for childhood tumours as these have much fewer non-synonymous coding mutations than adult malignancies. Recent large-scale sequencing by International Genome Consortium and Paediatric Cancer Genome Project has further shown that the genetic and epigenetic repertoire of driver mutations in specific childhood malignancies differs from more common adult-type malignancies. To bring about much needed change, a Paediatric Platform, ACCELERATE, was proposed in 2013 by the Cancer Drug Development Forum, Innovative Therapies for Children with Cancer, the European Network for Cancer Research in Children and Adolescents and the European Society for Paediatric Oncology. The Platform, comprising multiple stakeholders in paediatric oncology, has three working groups, one with responsibility for promoting and developing high-quality MoA-informed paediatric drug development programmes, including specific measures for adolescents. Key is the establishment of a freely accessible aggregated database of paediatric biological tumour drug targets to be aligned with an aggregated pipeline of drugs. This will enable prioritisation and conduct of early phase clinical paediatric trials to evaluate these drugs against promising therapeutic targets and to generate clinical paediatric efficacy and safety data in an accelerated time frame. Through this work, the Platform seeks to ensure that potentially effective drugs, where the MoA is known and thought to be relevant to paediatric malignancies, are evaluated in early phase clinical trials, and that this approach to generate pre-clinical and clinical data is systematically pursued by academia, sponsors, industry, and regulatory bodies to bring new paediatric oncology drugs to front-line therapy more rapidly. (C) 2016 Elsevier Ltd. All rights reserved.
C1 [Pearson, Andrew D. J.; Marshall, Lynley V.] Royal Marsden NHS Fdn Trust, Inst Canc Res, London, England.
   [Herold, Ralf; van den Berg, Hendrick] European Med Agcy, Prod Dev Sci Support Dept, London, England.
   [Rousseau, Raphael] Genentech Inc, 1 DNA Way, San Francisco, CA 94080 USA.
   [Copland, Chris] Univ York, Ctr English Language Teaching, York YO10 5DD, N Yorkshire, England.
   [Bradley-Garelik, Brigid] Bristol Myers Squibb Co, 5 Res Pkwy, Wallingford, CT 06492 USA.
   [Binner, Debbie] Create Chloe & UK Representat aPODD, Birmingham, W Midlands, England.
   [Capdeville, Renaud] Novartis Pharma AG, CH-4002 Basel, Switzerland.
   [Caron, Hubert] Hoffmann La Roche AG, Basel, Switzerland.
   [Caron, Hubert] Univ Amsterdam, Acad Med Ctr, Emma Childrens Hosp, Dept Pediat Oncol, Meibergdreef 9, NL-1105 AZ Amsterdam, Netherlands.
   [Carleer, Jacqueline] Belgian Fed Agcy Med & Hlth Prod, Brussels, Belgium.
   [Chesler, Louis] Inst Canc Res, Div Clin Studies, London SW3 6JB, England.
   [Geoerger, Birgit] Gustave Roussy, Dept Pediat & Adolescent Oncol, Villejuif, France.
   [Kearns, Pamela] Univ Birmingham, Inst Canc & Genom Sci, Canc Res UK Clin Trials Unit CRCTU, Birmingham, W Midlands, England.
   [Marshall, Lynley V.] Royal Marsden NHS Fdn Trust, Children & Young Peoples Unit, Sutton, Surrey, England.
   [Pfister, Stefan M.] German Canc Consortium DKTK, German Canc Res Ctr DKFZ, D-69120 Heidelberg, Germany.
   [Pfister, Stefan M.] Univ Heidelberg Hosp, D-69120 Heidelberg, Germany.
   [Schleiermacher, Gudrun] Inst Curie, RTOP, INSERM, U830, Paris, France.
   [Schleiermacher, Gudrun] Inst Curie, Dept Pediat Oncol, Paris, France.
   [Skolnik, Jeffrey] TetraLog Pharmaceut, Malvern, PA 19355 USA.
   [Spadoni, Cesare] aPODD Fdn, London, England.
   [Sterba, Jaroslav] Univ Hosp Brno, Fac Med, Dept Paediat Oncol, Brno, Czech Republic.
   [Sterba, Jaroslav] Masaryk Univ, Brno, Czech Republic.
   [Sterba, Jaroslav] ICRC Brno, Masaryk Mem Canc Inst, Reg Ctr Appl Mol Oncol, Brno, Czech Republic.
   [Uttenreuther-Fischer, Martina] Boehringer, Ingelheim, Germany.
   [Witt, Olaf] German Canc Res Ctr, Clin Cooperat Unit Pediat Oncol G340, Heidelberg, Germany.
   [Norga, Koen] Univ Antwerp, Paediat Haematol Oncol Unit, Univ Antwerp Hosp, B-2020 Antwerp, Belgium.
   [Vassal, Gilles] Univ Paris 11, Inst Gustave Roussy, Dept Clin Res, Paris, France.
C3 Royal Marsden NHS Foundation Trust; University of London; Institute of
   Cancer Research - UK; Roche Holding; Genentech; University of York - UK;
   Bristol-Myers Squibb; Novartis; Roche Holding; Emma Children's Hospital;
   University of Amsterdam; Royal Marsden NHS Foundation Trust; University
   of London; Institute of Cancer Research - UK; UNICANCER; Gustave Roussy;
   Cancer Research UK; University of Birmingham; Royal Marsden NHS
   Foundation Trust; Helmholtz Association; German Cancer Research Center
   (DKFZ); Ruprecht Karls University Heidelberg; UDICE-French Research
   Universities; Universite Paris Cite; Universite PSL; UNICANCER; Institut
   Curie; Institut National de la Sante et de la Recherche Medicale
   (Inserm); UDICE-French Research Universities; Universite PSL; UNICANCER;
   Institut Curie; Masaryk University Brno; University Hospital Brno;
   Masaryk University Brno; Masaryk Memorial Cancer Institute; St Anne's
   University Hospital Brno (FNUSA-ICRC); Helmholtz Association; German
   Cancer Research Center (DKFZ); University of Antwerp; UDICE-French
   Research Universities; Universite Paris Saclay; UNICANCER; Gustave
   Roussy
RP Pearson, ADJ (通讯作者)，Royal Marsden NHS Fdn Trust, Inst Canc Res, London, England.
EM
RI Pfister, Stefan M/F-6860-2013; Schleiermacher, Gudrun/AAN-4711-2021;
   Kearns, Pamela/AAD-6011-2021; Sterba, Jaroslav/N-6156-2016; Norga,
   Koenraad/D-1137-2017
OI Pfister, Stefan M/0000-0002-5447-5322; Schleiermacher,
   Gudrun/0000-0002-0133-5879; Herold, Ralf/0000-0002-8148-6748; Chesler,
   Louis/0000-0001-7842-2068; Norga, Koenraad/0000-0002-9231-0350;
   geoerger, birgit/0000-0003-4361-3643; Rousseau,
   Raphael/0000-0001-7288-5213; Kearns, Pamela/0000-0003-2756-5813; Sterba,
   Jaroslav/0000-0002-8482-8352
FU European Union [HEALTH-F2-2011-261474]; Cancer Research UK [18339,
   15958] Funding Source: researchfish; Rosetrees Trust [M571] Funding
   Source: researchfish
FX This publication has received funding from the European Union's Seventh
   Framework Programme for research, technological development, and
   demonstration under the project ENCCA-grant agreement no.
   HEALTH-F2-2011-261474.
CR Bautista F, 2014, PEDIATR BLOOD CANCER, V61, P1101, DOI 10.1002/pbc.24891
   Berk DR, 2010, PEDIATR DERMATOL, V27, P244, DOI 10.1111/j.1525-1470.2009.01078.x
   Buczkowicz P, 2014, NAT GENET, V46, P451, DOI 10.1038/ng.2936
   CRAFT AW, 1989, CANCER SURV, V8, P605
   EMA, 2013, 5 YEAR REP EUR COMM
   FDA, 2013, PED ONC SUBC ODAC
   Hill RM, 2015, CANCER CELL, V27, P72, DOI 10.1016/j.ccell.2014.11.002
   Hovestadt V, 2014, NATURE, V510, P537, DOI 10.1038/nature13268
   Kieran MW, 2015, J CLIN ONCOL, V33
   Kool M, 2014, CANCER CELL, V25, P393, DOI 10.1016/j.ccr.2014.02.004
   Middleton G, 2015, ANN ONCOL, V26, P2464, DOI 10.1093/annonc/mdv394
   Paoletti X, 2013, EUR J CANCER, V49, P2392, DOI 10.1016/j.ejca.2013.02.028
   Pritchard-Jones K, 2013, LANCET ONCOL, V14, pE95, DOI 10.1016/S1470-2045(13)70010-X
   Rossig C, 2013, PEDIATR BLOOD CANCER, V60, P1574, DOI 10.1002/pbc.24598
   Schleiermacher G, 2014, J CLIN ONCOL, V32, P2727, DOI 10.1200/JCO.2013.54.0674
   Taylor KR, 2014, NAT GENET, V46, P457, DOI 10.1038/ng.2925
   Vassal G, CLASS WAIVER O UNPUB
   Vassal G, 2015, EUR J CANCER, V51, P218, DOI 10.1016/j.ejca.2014.10.029
   Vassal G, 2013, CLIN CANCER RES, V19, P1315, DOI 10.1158/1078-0432.CCR-12-2551
   Vassal G, 2013, LANCET ONCOL, V14, pE117, DOI 10.1016/S1470-2045(13)70013-5
   Wu G, 2014, NAT GENET, V46, P444, DOI 10.1038/ng.2938
   Young D, 2015, SCRIP INTELLIGENCE
   Zhang JH, 2013, NAT GENET, V45, P602, DOI 10.1038/ng.2611
NR 23
TC 50
Z9 50
U1 0
U2 21
PU ELSEVIER SCI LTD
PI OXFORD
PA THE BOULEVARD, LANGFORD LANE, KIDLINGTON, OXFORD OX5 1GB, OXON, ENGLAND
SN 0959-8049
EI 1879-0852
J9 EUR J CANCER
JI Eur. J. Cancer
PD JUL
PY 2016
VL 62
BP 124
EP 131
DI 10.1016/j.ejca.2016.04.001
PG 8
WC Oncology
WE Science Citation Index Expanded (SCI-EXPANDED)
SC Oncology
GA DN9FO
UT WOS:000377385100014
PM 27258969
OA Green Submitted
DA 2023-08-27
ER

PT J
AU Wakerley, BR
   Yuki, N
AF Wakerley, Benjamin R.
   Yuki, Nobuhiro
TI Infectious and noninfectious triggers in Guillain-Barre syndrome
SO EXPERT REVIEW OF CLINICAL IMMUNOLOGY
LA English
DT Review
DE acute inflammatory demyelinating polyneuropathy; acute motor axonal
   neuropathy; Bickerstaff brainstem encephalitis; Campylobacter jejuni;
   Fisher syndrome; Guillain-Barre syndrome; molecular mimicry
ID MOTOR AXONAL NEUROPATHY; SYSTEMIC-LUPUS-ERYTHEMATOSUS; BRAIN-STEM
   ENCEPHALITIS; ANTIGANGLIOSIDE ANTIBODY INDUCTION; CAMPYLOBACTER-JEJUNI
   INFECTION; LOCAL HEALTH DISTRICT; TUMOR-NECROSIS-FACTOR;
   HAEMOPHILUS-INFLUENZAE; FISHER-SYNDROME; CYTOMEGALOVIRUS-INFECTION
AB Guillain-Barre syndrome (GBS) is the commonest cause of acquired flaccid paralysis in the world and regarded by many as the prototype for postinfectious autoimmunity. Here the authors consider both infectious and noninfectious triggers of GBS and determine where possible what immunological mechanisms may account for this association. In approximately two-thirds of cases, an infectious trigger is reported in the weeks that lead up to disease onset, indicating that the host's response to infection must play an important role in disease pathogenesis. The most frequently identified bacteria, Campylobacter jejuni, through a process known as molecular mimicry, has been shown to induce cross-reactive anti-ganglioside antibodies, which can lead to the development of axonal-type GBS in some patients. Whether this paradigm can be extended to other infectious organisms or vaccines remains an important area of research and has public health implications. GBS has also been reported rarely in patients with underlying systemic diseases and immunocompromised states and although the exact mechanism is yet to be established, increased susceptibility to known infectious triggers should be considered most likely.
C1 [Wakerley, Benjamin R.; Yuki, Nobuhiro] Natl Univ Singapore Hosp, Dept Med, Singapore 117548, Singapore.
C3 National University of Singapore
RP Wakerley, BR (通讯作者)，Natl Univ Singapore Hosp, Dept Med, 1E Kent Ridge Rd, Singapore 117548, Singapore.
EM
OI Wakerley, Benjamin/0000-0003-1016-5207
FU National Medical Research Council in Singapore [IRG 10nov086]; Medical
   Research Council [G0501747] Funding Source: researchfish; MRC [G0501747]
   Funding Source: UKRI
FX This study was supported by the National Medical Research Council (IRG
   10nov086 to N Yuki) in Singapore. The authors have no other relevant
   affiliations or financial involvement with any organization or entity
   with a financial interest in or financial conflict with the subject
   matter or materials discussed in the manuscript apart from those
   disclosed.
CR Allos BM, 1997, J INFECT DIS, V176, pS125, DOI 10.1086/513783
   Ang CW, 2004, TRENDS IMMUNOL, V25, P61, DOI 10.1016/j.it.2003.12.004
   Angele MK, 2002, CRIT CARE, V6, P298, DOI 10.1186/cc1514
   [Anonymous], 2000, CAMPYLOBACTER
   ASBURY AK, 1969, MEDICINE, V48, P173, DOI 10.1097/00005792-196905000-00001
   Awad A, 2010, MUSCLE NERVE, V42, P828, DOI 10.1002/mus.21830
   Baxter R, 2012, CLIN INFECT DIS, V54, P800, DOI 10.1093/cid/cir960
