## Supplemental Material 1 for "Bibliometric analysis of emerging trends and research foci in brainstem tumor field over thirty years (1992-2023)": savedrecs.txt

FN Clarivate Analytics Web of Science
VR 1.0
PT J
AU Recinos, PF
   Sciubba, DM
   Jallo, GI
AF Recinos, Pablo F.
   Sciubba, Daniel M.
   Jallo, George I.
TI Brainstem tumors: Where are we today?
SO PEDIATRIC NEUROSURGERY
LA English
DT Article
DE brainstem tumors; cervicomedullary tumors; dorsal exophytic tumors;
   glioma; medulla; midbrain tumors; pontine glioma; tectal tumors
ID CONVECTION-ENHANCED DELIVERY; LONG-TERM; SURGICAL INDICATIONS;
   HYPERFRACTIONATED RADIOTHERAPY; CERVICOMEDULLARY JUNCTION; INTRAAXIAL
   TUMORS; GENE-THERAPY; MOTOR NUCLEI; GLIOMAS; CHILDREN
AB Brainstem tumors comprise 10-20% of all pediatric central nervous system tumors. The management of these tumors has evolved dramatically in the past century. Once considered uniformly fatal, it is now known that brainstem tumors have distinguishing characteristics and do not behave identically. The focality and location of the lesion is determined from the clinical history, presentation, and associated imaging. Based on these findings, it is possible to predict the behavior of the tumor and choose an appropriate intervention. Focal lesions have a good prognosis and are treated operatively while diffuse lesions have a poor prognosis and are managed medically. This article reviews the current classification of brainstem tumors, current management options and future directions for the treatment of these rare tumors. Copyright (c) 2007 S. Karger AG, Basel.
C1 Johns Hopkins Univ, Sch Med, Dept Neurosurg, Baltimore, MD 21205 USA.
C3 Johns Hopkins University
RP Jallo, GI (通讯作者)，Johns Hopkins Univ Hosp, Div Pediat Neurosurg, 600 N Wolfe St,Harvey 811, Baltimore, MD 21287 USA.
EM
RI Recinos, Pablo/AAU-7661-2020; Recinos, Pablo/ABB-5351-2021
OI Recinos, Pablo/0000-0002-9748-2784; 
CR Abbott R, 1993, Neurosurg Clin N Am, V4, P519
   Abbott R, 1996, PEDIATR NEUROSURG, V25, P41, DOI 10.1159/000121095
   ABBOTT R, 1992, PEDIATR NEUROSURG, V17, P239, DOI 10.1159/000120604
   Albright A L, 1993, Neurosurg Clin N Am, V4, P529
   ALBRIGHT AL, 1983, CANCER, V52, P2313, DOI 10.1002/1097-0142(19831215)52:12<2313::AID-CNCR2820521226>3.0.CO;2-I
   Albright AL, 1996, PEDIATR NEUROSURG, V24, P252
   ALBRIGHT AL, 1986, J NEUROSURG, V65, P751, DOI 10.3171/jns.1986.65.6.0751
   Albright AL, 2004, YOUMANS NEUROLOGICAL, V3, P3663
   ALBRIGHT AL, 1996, NEUROLOGICAL SURG, P2603
   ALLEN J, 1983, BRAIN STEM GLIOMA, P2
   Allen JC, 1996, PEDIATR NEUROSURG, V24, P98, DOI 10.1159/000121024
   ALVISI C, 1985, ACTA NEUROCHIR, V76, P12, DOI 10.1007/BF01403823
   Amundson EW, 2005, J NEUROSURG, V102, P565, DOI 10.3171/jns.2005.102.3.0565
   [Anonymous], 2000, OPERATIVE TECHNIQUES
   BARKOVICH AJ, 1991, PEDIATR NEUROSURG, V16, P73, DOI 10.1159/000120511
   BERGER MS, 1983, NEUROSURGERY, V12, P298, DOI 10.1227/00006123-198303000-00008
   BOBO RH, 1994, P NATL ACAD SCI USA, V91, P2076, DOI 10.1073/pnas.91.6.2076
   Bowers DC, 2000, PEDIATR NEUROSURG, V32, P24, DOI 10.1159/000028893
   BREM H, 1994, J NEUROSURG, V80, P283, DOI 10.3171/jns.1994.80.2.0283
   Bricolo A, 2000, OPER TECH NEUROSURG, V3, P137, DOI DOI 10.1053/oy.2000.6559
   Burrows FJ, 2002, CANCER GENE THER, V9, P87, DOI 10.1038/sj.cgt.7700401
   Choux M, 2000, PEDIAT NEUROSURGERY, P471
   De Bouard S, 2003, HUM GENE THER, V14, P883, DOI 10.1089/104303403765701178
   EPSTEIN F, 1985, CANCER, V56, P1804, DOI 10.1002/1097-0142(19851001)56:7+<1804::AID-CNCR2820561316>3.0.CO;2-5
   EPSTEIN F, 1988, J NEURO-ONCOL, V6, P309
   EPSTEIN F, 1986, J NEUROSURG, V64, P11, DOI 10.3171/jns.1986.64.1.0011
   EPSTEIN F, 1987, PROG EXP TUMOR RES, V30, P160
   EPSTEIN F, 1987, J NEUROSURG, V67, P483, DOI 10.3171/jns.1987.67.4.0483
   EPSTEIN FJ, 1993, J NEUROSURG, V78, P408, DOI 10.3171/jns.1993.78.3.0408
   Fischbein NJ, 1996, PEDIATR NEUROSURG, V24, P9, DOI 10.1159/000121010
   Fisher PG, 2000, CANCER, V89, P1569, DOI 10.1002/1097-0142(20001001)89:7<1569::AID-CNCR22>3.0.CO;2-0
   GILDAY DL, 1975, RADIOLOGY, V117, P93, DOI 10.1148/117.1.93
   Gomez-Gosalvez FA, 2001, REV NEUROLOGIA, V33, P605, DOI 10.33588/rn.3307.2001115
   Groothuis DR, 1999, J NEUROSURG, V90, P321, DOI 10.3171/jns.1999.90.2.0321
   HOFFMAN HJ, 1980, NEUROSURGERY, V7, P243, DOI 10.1227/00006123-198009000-00007
   Jallo GI, 2004, CHILD NERV SYST, V20, P143, DOI 10.1007/s00381-003-0870-6
   Jallo GI, 2005, CHILD NERV SYST, V21, P399, DOI 10.1007/s00381-004-1100-6
   Jallo GI, 2005, NEUROSURGERY, V56, P74, DOI 10.1227/01.NEU.0000144782.39430.12
   Kaplan AM, 1996, PEDIATR NEUROSURG, V24, P185, DOI 10.1159/000121036
   KHATIB ZA, 1994, PEDIATR NEUROSURG, V20, P2, DOI 10.1159/000120759
   Lee James, 2005, Neurosurg Focus, V18, pE11
   Li Khan W, 2005, Neurosurg Focus, V18, pE2
   LITTMAN P, 1980, CANCER, V45, P2787, DOI 10.1002/1097-0142(19800601)45:11<2787::AID-CNCR2820451113>3.0.CO;2-V
   Mandell LR, 1999, INT J RADIAT ONCOL, V43, P959, DOI 10.1016/S0360-3016(98)00501-X
   Matson DD, 1969, NEUROSURGERY INFANCY, P469
   Miller CR, 2002, CANCER RES, V62, P773
   Miura F, 2002, GENE THER, V9, P1653, DOI 10.1038/sj.gt.3301860
   Morota N, 1996, NEUROSURGERY, V39, P787, DOI 10.1097/00006123-199610000-00028
   MOROTA N, 1995, NEUROSURGERY, V37, P922, DOI 10.1227/00006123-199511000-00011
   Morota N, 2006, ACTA NEUROCHIR, V148, P499, DOI 10.1007/s00701-005-0672-6
   Occhiogrosso G, 2003, NEUROSURGERY, V52, P388, DOI 10.1227/01.NEU.0000043696.83722.8D
   PACKER RJ, 1994, CANCER-AM CANCER SOC, V74, P1827, DOI 10.1002/1097-0142(19940915)74:6<1827::AID-CNCR2820740628>3.0.CO;2-Q
   POLLACK IF, 1995, NEUROSURGERY, V37, P885, DOI 10.1227/00006123-199511000-00006
   POLLACK IF, 1993, J NEUROSURG, V78, P859, DOI 10.3171/jns.1993.78.6.0859
   RAJSHEKHAR V, 1995, J NEUROSURG, V82, P976, DOI 10.3171/jns.1995.82.6.0976
   Rand V, 2005, P NATL ACAD SCI USA, V102, P14344, DOI 10.1073/pnas.0507200102
   ROBERTSON PL, 1994, NEUROLOGY, V44, P1798, DOI 10.1212/WNL.44.10.1798
   Ruan SB, 1999, CLIN CANCER RES, V5, P197
   Sala F, 2002, CHILD NERV SYST, V18, P264, DOI 10.1007/s00381-002-0582-3
   Shiminski-Maher T, 1991, J Neurosci Nurs, V23, P356
   ShiminskiMaher T, 1996, PEDIATR NEUROSURG, V24, P267, DOI 10.1159/000121050
   Stark AM, 2005, PEDIATR NEUROL, V33, P33, DOI 10.1016/j.pediatrneurol.2004.12.014
   STECK J, 1995, SURG NEUROL, V43, P563, DOI 10.1016/0090-3019(95)00156-5
   Steinbok P, 2003, PEDIATR NEUROSURG, V39, P179, DOI 10.1159/000072468
   STROINK AR, 1987, NEUROSURGERY, V20, P439, DOI 10.1227/00006123-198703000-00014
   Todo T, 2000, CANCER GENE THER, V7, P939, DOI 10.1038/sj.cgt.7700182
   TOMITA T, 1984, J NEURO-ONCOL, V2, P117
   Walker DA, 1999, ARCH DIS CHILD, V80, P558, DOI 10.1136/adc.80.6.558
   Weiner HL, 1997, PEDIATR NEUROSURG, V27, P12, DOI 10.1159/000121219
   Wisoff J H, 1984, Neurosurgery, V15, P707
   Yamanaka R, 2002, J NEUROSURG, V97, P1184, DOI 10.3171/jns.2002.97.5.1184
NR 71
TC 55
Z9 60
U1 0
U2 4
PU KARGER
PI BASEL
PA ALLSCHWILERSTRASSE 10, CH-4009 BASEL, SWITZERLAND
SN 1016-2291
EI 1423-0305
J9 PEDIATR NEUROSURG
JI Pediatr. Neurosurg.
PY 2007
VL 43
IS 3
BP 192
EP 201
DI 10.1159/000098831
PG 10
WC Clinical Neurology; Pediatrics; Surgery
WE Science Citation Index Expanded (SCI-EXPANDED)
SC Neurosciences & Neurology; Pediatrics; Surgery
GA 161TG
UT WOS:000246038300005
PM 17409788
DA 2023-08-27
ER

PT J
AU Jia, HY
   Zhang, P
   Gu, GC
   Li, T
   Jiang, Z
   Wu, Z
   Wang, L
   Zhang, JT
   Duan, YY
   Liu, YO
   Yang, F
   Qin, SZ
   Zhang, LW
AF Jia, Heyuan
   Zhang, Peng
   Gu, Guocan
   Li, Tian
   Jiang, Zhuang
   Wu, Zhen
   Wang, Liang
   Zhang, Junting
   Duan, Yunyun
   Liu, Yaou
   Yang, Feng
   Qin, Shaozheng
   Zhang, Liwei
TI Brainstem tumors may increase the impairment of behavioral emotional
   cognition in children
SO JOURNAL OF NEURO-ONCOLOGY
LA English
DT Article
DE Brainstem tumors; Diffuse intrinsic pontine glioma; Behavioral problems;
   Emotional problems; Child behavior checklist
ID CLASSIFICATION; PROJECTIONS; DISORDER; ANATOMY; NETWORK
AB Purpose It remains unclear as to whether patients with brainstem tumor experience complex neuropsychiatric problems. In this cohort study, we specifically investigated behavioral, emotional and cognitive symptoms in pediatric patients with brainstem glioma and healthy individuals. Methods A total of 146 patients with pediatric brainstem tumors (aged 4-18 years old) and 46 age-matched healthy children were recruited to assess their behaviors and emotions examined by the Child Behavior Checklist. A variety of clinical factors were also analyzed. Results There were significant differences in most behavioral and emotional symptoms between pediatric patients and healthy subjects. Moreover, patients with pons tumors exhibited significantly higher scores than patients with medulla oblongata tumors (p = 0.012), particularly in concerning the syndrome categories of Withdrawn (p = 0.043), Anxious/depressed symptoms (p = 0.046), Thought Problems (p = 0.004), Attention deficits (p = 0.008), Externalizing problems (p = 0.013), and Aggressive behavior (p = 0.004). A tumor body located in the pontine (p = 0.01, OR = 4.5, 95% CI = 1.4-14.059) or DIPG in the midbrain (p = 0.002, OR = 3.818, 95% CI = 1.629-8.948) appears to act as a risk factor that is associated with more problems in patients with neuropsychiatric symptoms. Conclusions Pediatric patients with brainstem tumors exhibit severe behavioral and emotional problems. Tumor invades the pontine and midbrain act a risk factor with more problems. It suggests that structural and functional abnormalities in the brainstem will cause prolonged behavioral problems and emotional-cognitive dysfunctions in young children.
C1 [Jia, Heyuan] Beihang Univ, Sch Instrumentat & Optoelect Engn, 37 BeiHang Univ XueYuan Rd, Beijing 100083, Peoples R China.
   [Jia, Heyuan; Zhang, Liwei] Beihang Univ, Beijing Adv Innovat Ctr Big Data Based Precis Med, BeiHang Univ XueYuan Rd, Beijing 100083, Peoples R China.
   [Zhang, Peng; Gu, Guocan; Li, Tian; Jiang, Zhuang; Wu, Zhen; Wang, Liang; Zhang, Junting; Zhang, Liwei] Capital Med Univ, Beijing Tiantan Hosp, Dept Neurosurg, 119 West Southern 4Th Ring Rd, Beijing 100073, Peoples R China.
   [Duan, Yunyun; Liu, Yaou] Capital Med Univ, Beijing Tiantan Hosp, Dept Radiol, 119 West Southern 4Th Ring Rd, Beijing 100073, Peoples R China.
   [Yang, Feng; Zhang, Liwei] Capital Med Univ, Beijing Tiantan Hosp, China Natl Clin Res Ctr Neurol Dis, 119 West Southern 4Th Ring Rd, Beijing 100073, Peoples R China.
   [Yang, Feng] Capital Med Univ, Adv Innovat Ctr Human Brain Protect, Beijing 100070, Peoples R China.
   [Qin, Shaozheng] Beijing Normal Univ, State Key Lab Cognit Neurosci & Learning, 19 XinJieKouWai St, Beijing 100875, Peoples R China.
   [Qin, Shaozheng] Beijing Normal Univ, IDG McGovern Inst Brain Res, 19 XinJieKouWai St, Beijing 100875, Peoples R China.
C3 Beihang University; Beihang University; Capital Medical University;
   Capital Medical University; Capital Medical University; Capital Medical
   University; Beijing Normal University; Beijing Normal University
RP Zhang, LW (通讯作者)，Beihang Univ, Beijing Adv Innovat Ctr Big Data Based Precis Med, BeiHang Univ XueYuan Rd, Beijing 100083, Peoples R China.; Zhang, LW (通讯作者)，Capital Med Univ, Beijing Tiantan Hosp, Dept Neurosurg, 119 West Southern 4Th Ring Rd, Beijing 100073, Peoples R China.; Yang, F; Zhang, LW (通讯作者)，Capital Med Univ, Beijing Tiantan Hosp, China Natl Clin Res Ctr Neurol Dis, 119 West Southern 4Th Ring Rd, Beijing 100073, Peoples R China.; Yang, F (通讯作者)，Capital Med Univ, Adv Innovat Ctr Human Brain Protect, Beijing 100070, Peoples R China.; Qin, SZ (通讯作者)，Beijing Normal Univ, State Key Lab Cognit Neurosci & Learning, 19 XinJieKouWai St, Beijing 100875, Peoples R China.; Qin, SZ (通讯作者)，Beijing Normal Univ, IDG McGovern Inst Brain Res, 19 XinJieKouWai St, Beijing 100875, Peoples R China.
EM
RI Yang, Feng/IWU-7487-2023
FU National Natural Science Foundation of China [81872048, 32130045];
   Beijing Municipal Natural Science Foundation [7214214]; Beijing Health
   Commission [JingCaiKeWenZhi [2019] 2348]
FX This study was supported by the National Natural Science Foundation of
   China (Grant Numbers [81872048] and [32130045]), the Beijing Municipal
   Natural Science Foundation (Grant Numbers [7214214]), and the Beijing
   Health Commission (Grant Numbers [JingCaiKeWenZhi [2019] 2348]).
CR Achenbach T M, 2000, Pediatr Rev, V21, P265, DOI 10.1542/pir.21-8-265
   Althoff RR, 2010, PSYCHOL ASSESSMENT, V22, P609, DOI 10.1037/a0019699
   Althoff RR, 2010, PSYCHIAT RES, V178, P550, DOI 10.1016/j.psychres.2010.05.002
   Ammari R, 2020, NEURON, V106, P556, DOI 10.1016/j.neuron.2020.04.029
   CAIRNS H, 1950, Folia Psychiatr Neurol Neurochir Neerl, V53, P193
   Cheng X, 2019, J COMPUT ASSIST TOMO, V43, P547, DOI 10.1097/RCT.0000000000000881
   Cui YH, 2021, EUR CHILD ADOLES PSY, V30, P233, DOI 10.1007/s00787-020-01507-6
   D'aes T, 2015, CEREBELLUM, V14, P317, DOI 10.1007/s12311-014-0626-8
   DEVISSER BWO, 1981, CLIN NEUROL NEUROSUR, V83, P137, DOI 10.1016/0303-8467(81)90015-9
   Etard O, 2019, NEUROIMAGE, V200, P1, DOI 10.1016/j.neuroimage.2019.06.029
   Falkner AL, 2020, NEURON, V106, P637, DOI 10.1016/j.neuron.2020.02.014
   Fangusaro J, 2009, J CHILD NEUROL, V24, P1409, DOI 10.1177/0883073809338960
   Faraone SV, 2005, BIPOLAR DISORD, V7, P518, DOI 10.1111/j.1399-5618.2005.00271.x
   Fischbein NJ, 1996, PEDIATR NEUROSURG, V24, P9, DOI 10.1159/000121010
   Fisher PG, 2000, CANCER, V89, P1569, DOI 10.1002/1097-0142(20001001)89:7<1569::AID-CNCR22>3.0.CO;2-0
   Geva R, 2017, J CHILD PSYCHOL PSYC, V58, P1351, DOI 10.1111/jcpp.12746
   Green AL, 2015, CURR ONCOL REP, V17, DOI 10.1007/s11912-014-0436-7
   GREENBERG DB, 1985, J NERV MENT DIS, V173, P434, DOI 10.1097/00005053-198507000-00008
   Hurley RA, 2010, J NEUROPSYCH CLIN N, V22, pIV, DOI 10.1176/appi.neuropsych.22.1.iv
   Jansen MH, 2015, NEURO-ONCOLOGY, V17, P160, DOI 10.1093/neuonc/nou104
   Keschner M, 1937, ARCH NEURO PSYCHIATR, V37, P1
   Leach JL, 2020, NEURO-ONCOLOGY, V22, P1647, DOI 10.1093/neuonc/noaa140
   Liu XC, 1999, J AM ACAD CHILD PSY, V38, P708, DOI 10.1097/00004583-199906000-00018
   Lo LC, 2019, P NATL ACAD SCI USA, V116, P7503, DOI 10.1073/pnas.1817503116
   Lv WY, 2021, NEUROPSYCH DIS TREAT, V17, P91, DOI 10.2147/NDT.S287438
   NAUTA WJH, 1958, BRAIN, V81, P319, DOI 10.1093/brain/81.3.319
   NETSKY MG, 1952, AMA ARCH NEUROL PSY, V68, P116, DOI 10.1001/archneurpsyc.1952.02320190122012
   New PW, 2005, ARCH PHYS MED REHAB, V86, P338, DOI 10.1016/j.apmr.2004.09.005
   Palesi F, 2017, SCI REP-UK, V7, DOI 10.1038/s41598-017-13079-8
   Recinos PF, 2007, PEDIATR NEUROSURG, V43, P192, DOI 10.1159/000098831
   SCHMAHMANN JD, 1995, NEUROSCI LETT, V199, P175, DOI 10.1016/0304-3940(95)12056-A
   Schmahmann JD, 1998, BRAIN, V121, P561, DOI 10.1093/brain/121.4.561
   Shine JM, 2016, NEURON, V92, P544, DOI 10.1016/j.neuron.2016.09.018
   Sreenivasan V, 2019, P NATL ACAD SCI USA, V116, P19711, DOI 10.1073/pnas.1902704116
   Sturm W, 1999, NEUROPSYCHOLOGIA, V37, P797, DOI 10.1016/S0028-3932(98)00141-9
   Sun T, 2013, NEUROSURG REV, V36, P311, DOI 10.1007/s10143-012-0428-8
   Tang WK, 2017, J NEUROPSYCH CLIN N, V29, P172, DOI 10.1176/appi.neuropsych.15110400
   Van Zandvoort M, 2003, J INT NEUROPSYCH SOC, V9, P490, DOI 10.1017/S1355617703000146
   Venkatraman A, 2017, FRONT NEUROANAT, V11, DOI 10.3389/fnana.2017.00015
   XIN R, 1992, CAN J PSYCHIAT, V37, P250, DOI 10.1177/070674379203700408
NR 40
TC 0
Z9 0
U1 0
U2 0
PU SPRINGER
PI NEW YORK
PA ONE NEW YORK PLAZA, SUITE 4600, NEW YORK, NY, UNITED STATES
SN 0167-594X
EI 1573-7373
J9 J NEURO-ONCOL
JI J. Neuro-Oncol.
PD NOV
PY 2022
VL 160
IS 2
BP 423
EP 432
DI 10.1007/s11060-022-04161-x
EA NOV 2022
PG 10
WC Oncology; Clinical Neurology
WE Science Citation Index Expanded (SCI-EXPANDED)
SC Oncology; Neurosciences & Neurology
GA 7C8SY
UT WOS:000878938800001
PM 36333568
OA Green Published, hybrid
DA 2023-08-27
ER

PT J
AU Koh, EJ
   Phi, JH
   Park, SH
   Kim, IO
   Cheon, JE
   Wang, KC
   Cho, BK
   Kim, SK
AF Koh, Eun Jung
   Phi, Ji Hoon
   Park, Sung-Hye
   Kim, In-One
   Cheon, Jung-Eun
   Wang, Kyu-Chang
   Cho, Byung-Kyu
   Kim, Seung-Ki
TI Mixed germ cell tumor of the midbrain
SO JOURNAL OF NEUROSURGERY-PEDIATRICS
LA English
DT Article
DE mixed germ cell tumor; mature teratoma; germinoma; brainstem; midbrain
ID PRIMARY INTRACRANIAL GERMINOMA; MEDULLA-OBLONGATA GERMINOMA; BRAIN-STEM
   TUMORS; RESECTION; TERATOMA
AB This 14-year-old boy presented with left hemiparesis, gait disturbance, and multiple cranial nerve palsies. Magnetic resonance imaging revealed a multicystic mass with hemorrhagic fluid-fluid levels in the right midbrain, suggesting the presence of a cavernous malformation. Diffusion tensor imaging showed the mass to be close to the right corticospinal tract and ipsilateral medial lemniscus. Subtotal removal of the mass was performed via a right subtemporal approach. The histopathological diagnosis was of a mixed germ cell tumor (GCT) comprising mature teratoma and germinoma cells with syncytiotrophoblastic giant cells. The patient underwent postoperative chemotherapy and radiotherapy, and no tumor progression was found during 1 year of follow-up.
   Intracranial GCTs arise mainly in the pineal and the suprasellar area. Germ cell tumors in the brainstem are rare, with only 12 reported cases. Among these, 4 were in the midbrain and histologically were pure germinomas. To the authors' knowledge, this is the first reported case of a mixed GCT in the midbrain combining mature teratoma and germinoma cells. (DOI: 10.3171/2009.3.PEDS08290)
C1 [Koh, Eun Jung; Phi, Ji Hoon; Wang, Kyu-Chang; Cho, Byung-Kyu; Kim, Seung-Ki] Seoul Natl Univ, Coll Med, Childrens Hosp, Div Pediat Neurosurg, Seoul 110744, South Korea.
   [Park, Sung-Hye] Seoul Natl Univ, Coll Med, Childrens Hosp, Dept Pathol, Seoul 110744, South Korea.
   [Kim, In-One; Cheon, Jung-Eun] Seoul Natl Univ, Coll Med, Childrens Hosp, Dept Radiol, Seoul 110744, South Korea.
C3 Seoul National University (SNU); Seoul National University Hospital;
   Seoul National University (SNU); Seoul National University Hospital;
   Seoul National University (SNU); Seoul National University Hospital
RP Kim, SK (通讯作者)，Seoul Natl Univ, Coll Med, Childrens Hosp, Div Pediat Neurosurg, 101 Daehangno, Seoul 110744, South Korea.
EM
RI Kim, In-One/J-5427-2012; Park, Sung-Hye/B-5325-2011; Wang,
   Kyu-Chang/J-5545-2012; Kim, Seung-Ki/J-2742-2012; Cheon,
   Jung-Eun/J-5684-2012
OI Park, Sung-Hye/0000-0002-8681-1597; Cheon, Jung-Eun/0000-0003-1479-2064
FU National R&D Program for Cancer Control, Ministry of Health & Welfare,
   Republic of Korea [0520300]; Korea government (MEST)
   [R01-2008-000-20268-0]; National Research Foundation of Korea
   [R01-2008-000-20268-0] Funding Source: Korea Institute of Science &
   Technology Information (KISTI), National Science & Technology
   Information Service (NTIS)
FX This study was supported by a grant from the National R&D Program for
   Cancer Control, Ministry of Health & Welfare, Republic of Korea, Grant
   No. 0520300 (to B. K. Cho), and by the Korea Science and Engineering
   Foundation grant funded by the Korea government (MEST), Grant no.
   R01-2008-000-20268-0 (to S. K. Kim).
CR Ben Amor S, 2004, BRIT J NEUROSURG, V18, P310, DOI 10.1080/02688690410001732832
   Chen XL, 2007, SURG NEUROL, V68, P285, DOI 10.1016/j.surneu.2007.05.005
   Cho KT, 2002, CHILD NERV SYST, V18, P30, DOI 10.1007/s00381-001-0547-y
   HASHIMOTO M, 1992, SURG NEUROL, V37, P384, DOI 10.1016/0090-3019(92)90009-C
   Helton KJ, 2008, J NEUROSURG-PEDIATR, V1, P270, DOI 10.3171/PED/2008/1/4/270
   Koizumi H, 2006, ACTA NEUROCHIR, V148, P1197, DOI 10.1007/s00701-006-0872-8
   Liang L, 2002, NEURORADIOLOGY, V44, P382, DOI 10.1007/s00234-001-0752-0
   Matsumoto K, 1998, CLIN NEUROL NEUROSUR, V100, P292, DOI 10.1016/S0303-8467(98)00050-X
   Matsutani M, 1997, J NEUROSURG, V86, P446, DOI 10.3171/jns.1997.86.3.0446
   Miyamoto S, 2005, ACTA NEUROCHIR, V147, P93, DOI 10.1007/s00701-004-0339-8
   Nakajima H, 2000, SURG NEUROL, V53, P448, DOI 10.1016/S0090-3019(00)00224-X
   Ogawa K, 2003, CANCER-AM CANCER SOC, V98, P369, DOI 10.1002/cncr.11495
   Oosterhuis JW, 2007, INT J ANDROL, V30, P256, DOI 10.1111/j.1365-2605.2007.00793.x
   Ozelame RV, 2006, PEDIATR RADIOL, V36, P325, DOI 10.1007/s00247-005-0063-4
   POUNGVARIN N, 1991, Journal of the Medical Association of Thailand, V74, P55
   Recinos PF, 2007, PEDIATR NEUROSURG, V43, P192, DOI 10.1159/000098831
   Saito A, 1983, SHONI NO NOSHINKEI, V8, P43
   Sano K, 1999, J NEUROSURG, V90, P258, DOI 10.3171/jns.1999.90.2.0258
   Seol HJ, 2001, CHILD NERV SYST, V17, P758, DOI 10.1007/s003810100464
   Sugiyama K, 1994, Neurol Med Chir (Tokyo), V34, P291, DOI 10.2176/nmc.34.291
   Tashiro Takashi, 1993, Neurologia Medico-Chirurgica, V33, P251, DOI 10.2176/nmc.33.251
   Tharin S, 2007, NEUROSURGERY, V60, P185, DOI 10.1227/01.0000255386.95464.52
   Tsuzuki N, 2001, ACTA NEUROCHIR, V143, P1303, DOI 10.1007/PL00010098
   Uchino M, 2006, J NEURO-ONCOL, V79, P255, DOI 10.1007/s11060-006-9138-9
   Yen PS, 2003, J NEURO-ONCOL, V62, P339, DOI 10.1023/A:1023331700995
NR 25
TC 6
Z9 6
U1 0
U2 3
PU AMER ASSOC NEUROLOGICAL SURGEONS
PI ROLLING MEADOWS
PA 5550 MEADOWBROOK DRIVE, ROLLING MEADOWS, IL 60008 USA
SN 1933-0707
EI 1933-0715
J9 J NEUROSURG-PEDIATR
JI J. Neurosurg.-Pediatr.
PD AUG
PY 2009
VL 4
IS 2
BP 137
EP 142
DI 10.3171/2009.3.PEDS08290
PG 6
WC Clinical Neurology; Pediatrics; Surgery
WE Science Citation Index Expanded (SCI-EXPANDED)
SC Neurosciences & Neurology; Pediatrics; Surgery
GA 475EO
UT WOS:000268341400009
PM 19645547
DA 2023-08-27
ER

PT J
AU Chen, YW
   Lee, YY
   Lin, CF
   Huang, TY
   Ke, SH
   Mu, PF
   Pan, PS
   Chen, JK
   Lan, TL
   Hsu, PC
   Liang, ML
   Chen, HH
   Chang, FC
   Wu, CC
   Lin, SC
   Lee, JC
   Chen, SK
   Liu, HM
   Peir, JJ
   Tsai, HY
   Lin, KH
   Peng, NJ
   Chen, KH
   Wu, YH
   Kang, YM
   Yang, WC
   Liou, SC
   Huang, WH
   Tanaka, H
   Wong, TT
   Chao, Y
   Chou, FI
AF Chen, Yi-Wei
   Lee, Yi-Yen
   Lin, Chun-Fu
   Huang, Ting-Yu
   Ke, Shih-Hung
   Mu, Pei-Fan
   Pan, Po-Shen
   Chen, Jen-Kun
   Lan, Tien-Li
   Hsu, Ping-Chuan
   Liang, Muh-Lii
   Chen, Hsin-Hung
   Chang, Feng-Chi
   Wu, Chih-Chun
   Lin, Shih-Chieh
   Lee, Jia-Cheng
   Chen, Shih-Kuan
   Liu, Hong-Ming
   Peir, Jinn-Jer
   Tsai, Hui-Yu
   Lin, Ko-Han
   Peng, Nan-Jing
   Chen, Kuan-Hsuan
   Wu, Yuan-Hung
   Kang, Yu-Mei
   Yang, Wan-Chin
   Liou, Shueh-Chun
   Huang, Wei-Hsuan
   Tanaka, Hiroki
   Wong, Tai-Tong
   Chao, Yee
   Chou, Fong-In
TI Compassionate Treatment of Brainstem Tumors with Boron Neutron Capture
   Therapy: A Case Series
SO LIFE-BASEL
LA English
DT Article
DE boron neutron capture therapy; brainstem tumor; diffuse midline glioma;
   compassionate use; bevacizumab
ID INTRINSIC PONTINE GLIOMA; RECURRENT HEAD; NECK-CANCER; BNCT;
   RADIOTHERAPY; TEMOZOLOMIDE; CYCLOTRON
AB Brainstem tumors are heterogenous and cancerous glioma tumors arising from the midbrain, pons, and the medulla that are relatively common in children, accounting for 10% to 20% of all pediatric brain tumors. However, the prognosis of aggressive brainstem gliomas remains extremely poor despite aggressive treatment with chemotherapy and radiotherapy. That means there are many life-threatening patients who have exhausted all available treatment options and are beginning to face end-of-life stage. Therefore, the unique properties of highly selective heavy particle irradiation with boron neutron capture therapy (BNCT) may be well suited to prolong the lives of patients with end-stage brainstem gliomas. Herein, we report a case series of life-threatening patients with end-stage brainstem glioma who eligible for Emergency and Compassionate Use, in whom we performed a scheduled two fractions of salvage BNCT strategy with low treatment dosage each time. No patients experienced acute or late adverse events related to BNCT. There were 3 patients who relapsed after two fractionated BNCT treatment, characterized by younger age, lower T/N ratio, and receiving lower treatment dose. Therefore, two fractionated low-dose BNCT may be a promising treatment for end-stage brainstem tumors. For younger patients with low T/N ratios, more fractionated low-dose BNCT should be considered.
C1 [Chen, Yi-Wei; Lee, Yi-Yen; Wu, Yuan-Hung; Kang, Yu-Mei; Yang, Wan-Chin; Liou, Shueh-Chun] Natl Yang Ming Chiao Tung Univ, Fac Med, Taipei 112304, Taiwan.
   [Chen, Yi-Wei; Lan, Tien-Li; Lee, Jia-Cheng; Chen, Shih-Kuan; Wu, Yuan-Hung; Kang, Yu-Mei; Yang, Wan-Chin; Chao, Yee] Taipei Vet Gen Hosp, Dept Oncol, Taipei 11217, Taiwan.
   [Chen, Yi-Wei; Liou, Shueh-Chun] Yuanpei Univ Med Technol, Dept Med Imaging & Radiol Technol, Hsinchu 30015, Taiwan.
   [Chen, Yi-Wei] Natl Tsing Hua Univ, Coll Nucl Sci, Hsinchu 300044, Taiwan.
   [Lee, Yi-Yen; Lin, Chun-Fu; Hsu, Ping-Chuan] Taipei Vet Gen Hosp, Dept Neurosurg, Taipei 11217, Taiwan.
   [Huang, Ting-Yu; Ke, Shih-Hung] Taipei Vet Gen Hosp, Nursing Dept, Taipei 11217, Taiwan.
   [Mu, Pei-Fan] Natl Yang Ming Chiao Tung Univ, Sch Nursing, Taipei 112304, Taiwan.
   [Pan, Po-Shen] Tamkang Univ, Dept Chem, New Taipei 251301, Taiwan.
   [Chen, Jen-Kun] Natl Hlth Res Inst, Inst Biomed Engn & Nanomed, Zhunan Town 350, Miaoli County, Taiwan.
   [Liang, Muh-Lii] Mackay Mem Hosp, Dept Neurosurg, Taipei 104217, Taiwan.
   [Chen, Hsin-Hung] Taipei Vet Gen Hosp, Dept Pediat Neurosurg, Taipei 11217, Taiwan.
   [Chang, Feng-Chi; Wu, Chih-Chun] Taipei Vet Gen Hosp, Dept Radiol, Taipei 11217, Taiwan.
   [Lin, Shih-Chieh] Taipei Vet Gen Hosp, Dept Pathol & Lab Med, Taipei 11217, Taiwan.
   [Liu, Hong-Ming; Peir, Jinn-Jer; Tsai, Hui-Yu; Chou, Fong-In] Natl Tsing Hua Univ, Nucl Sci & Technol Dev Dept, Hsinchu 300044, Taiwan.
   [Lin, Ko-Han; Peng, Nan-Jing] Taipei Vet Gen Hosp, Dept Nucl Med, Taipei 11217, Taiwan.
   [Chen, Kuan-Hsuan] Natl Yang Ming Chiao Tung Univ, Sch Pharmaceut Sci, Dept Pharm, Hsinchu 112304, Taiwan.
   [Chen, Kuan-Hsuan] Taipei Vet Gen Hosp, Dept Pharm, Taipei 11217, Taiwan.
   [Huang, Wei-Hsuan] Mackay Mem Hosp, Dept Radiat Oncol, Taipei 104217, Taiwan.
   [Tanaka, Hiroki] Kyoto Univ, Inst Integrated Radiat & Nucl Sci, Kyoto 6068501, Japan.
   [Wong, Tai-Tong] Taipei Med Univ, Dept Neurosurg, Taipei 110301, Taiwan.
C3 National Yang Ming Chiao Tung University; Taipei Veterans General
   Hospital; National Tsing Hua University; Taipei Veterans General
   Hospital; Taipei Veterans General Hospital; National Yang Ming Chiao
   Tung University; Tamkang University; National Health Research Institutes
   - Taiwan; Mackay Memorial Hospital; Taipei Veterans General Hospital;
   Taipei Veterans General Hospital; Taipei Veterans General Hospital;
   National Tsing Hua University; Taipei Veterans General Hospital; Taipei
   Veterans General Hospital; Mackay Memorial Hospital; Kyoto University
RP Chen, YW (通讯作者)，Natl Yang Ming Chiao Tung Univ, Fac Med, Taipei 112304, Taiwan.; Chen, YW (通讯作者)，Taipei Vet Gen Hosp, Dept Oncol, Taipei 11217, Taiwan.; Chen, YW (通讯作者)，Yuanpei Univ Med Technol, Dept Med Imaging & Radiol Technol, Hsinchu 30015, Taiwan.; Chen, YW (通讯作者)，Natl Tsing Hua Univ, Coll Nucl Sci, Hsinchu 300044, Taiwan.
EM;
  ;
  ;
  ;
  ;
  ;
  ;
  ;
  ;
  ;
  
RI Chen, Yi/HIR-2608-2022; chen, ye Xiao/HSF-9650-2023; Chen,
   Yuxuan/IWL-8267-2023; Chen, Yi/HPD-0595-2023; 張, 豐基/IXN-6983-2023; ,
   jjpeir/ISS-2122-2023; Chen, Jen-Kun/C-6225-2011; Pan,
   Po-Shen/H-6781-2013
OI Wu, Yuan-Hung/0000-0003-2908-7704; Wong, Tai-Tong/0000-0003-4075-6227;
   Chao, Yee/0000-0001-5578-340X; Jinn-Jer, Peir/0000-0002-9196-4897; Chen,
   Shih-Kuan/0000-0001-8174-8077; Tsai, Hui-Yu/0000-0002-9661-5848; Chen,
   Kuan-Hsuan/0000-0003-2489-0055
CR [Anonymous], 2019, R COLL RADIOL RADIOT
   [Anonymous], 2017, COMMON TERMINOLOGY C
   Aziz-Bose R, 2019, CURR OPIN ONCOL, V31, P522, DOI 10.1097/CCO.0000000000000577
   Bailey S, 2013, EUR J CANCER, V49, P3856, DOI 10.1016/j.ejca.2013.08.006
   Barth RF, 2018, CANCER COMMUN, V38, DOI 10.1186/s40880-018-0299-7
   Cavalheiro S, 2015, CHILD NERV SYST, V31, P1815, DOI 10.1007/s00381-015-2799-y
   Chen Y.-W., 2020, THER RADIOL ONCOL, V4, P30, DOI [10.21037/tro-20-20, DOI 10.21037/TRO-20-20]
   Chen YW, 2021, BIOLOGY-BASEL, V10, DOI 10.3390/biology10040334
   Cohen KJ, 2011, NEURO-ONCOLOGY, V13, P410, DOI 10.1093/neuonc/noq205
   Cooney TM, 2020, J NEURO-ONCOL, V150, P27, DOI 10.1007/s11060-020-03553-1
   Fukuda H, 2021, CELLS-BASEL, V10, DOI 10.3390/cells10112881
   Furuse M, 2022, JPN J CLIN ONCOL, V52, P433, DOI 10.1093/jjco/hyac004
   Gallitto M, 2019, ADV RADIAT ONCOL, V4, P520, DOI 10.1016/j.adro.2019.03.009
   Gonzalez J, 2007, INT J RADIAT ONCOL, V67, P323, DOI 10.1016/j.ijrobp.2006.10.010
   Grimm SA, 2013, CURR NEUROL NEUROSCI, V13, DOI 10.1007/s11910-013-0346-3
   Guillamo JS, 2001, BRAIN, V124, P2528, DOI 10.1093/brain/124.12.2528
   Haginomori SI, 2009, HEAD NECK-J SCI SPEC, V31, P412, DOI 10.1002/hed.20895
   Hiratsuka J, 2020, J RADIAT RES, V61, P945, DOI 10.1093/jrr/rraa068
   Hirose K, 2021, RADIOTHER ONCOL, V155, P182, DOI 10.1016/j.radonc.2020.11.001
   Hu J, 2016, FRONT ONCOL, V6, DOI 10.3389/fonc.2016.00180
   Huang TY, 2022, CANCER NURS, V45, P201, DOI 10.1097/NCC.0000000000000979
   Jackson AC, 2007, J PEDIATR ONCOL NURS, V24, P95, DOI 10.1177/1043454206296030
   Kageji T, 2014, J MED INVESTIG, V61, P254, DOI 10.2152/jmi.61.254
   Kiyanagi Y, 2019, AIP CONF PROC, V2160, DOI 10.1063/1.5127704
   Lan TL, 2020, APPL RADIAT ISOTOPES, V160, DOI 10.1016/j.apradiso.2020.109105
   Leibetseder A, 2022, J NEUROL, V269, P1574, DOI 10.1007/s00415-021-10725-0
   Louis DN, 2021, NEURO-ONCOLOGY, V23, P1231, DOI 10.1093/neuonc/noab106
   Malouff TD, 2021, FRONT ONCOL, V11, DOI 10.3389/fonc.2021.601820
   Massimino M, 2016, NEURO-ONCOLOGY, V18, P1451, DOI 10.1093/neuonc/now108
   Miyatake SI, 2009, J NEURO-ONCOL, V91, P199, DOI 10.1007/s11060-008-9699-x
   Nobakht E, 2019, REP PRACT ONCOL RADI, V24, P644, DOI 10.1016/j.rpor.2019.10.005
   Ono K., 2006, P 12 INT C NEUTRON C, P27
   Rashed WM, 2019, CANCER METAST REV, V38, P759, DOI 10.1007/s10555-019-09824-2
   Reithmeier T, 2014, BMC CANCER, V14, DOI 10.1186/1471-2407-14-115
   Stenstam BH, 2007, ACTA NEUROL SCAND, V115, P243, DOI 10.1111/j.1600-0404.2006.00776.x
   Suzuki M, 2020, INT J CLIN ONCOL, V25, P43, DOI 10.1007/s10147-019-01480-4
   Suzuki M, 2014, J RADIAT RES, V55, P146, DOI 10.1093/jrr/rrt098
   Tsai YL, 2013, CHILD NERV SYST, V29, P1123, DOI 10.1007/s00381-013-2099-3
   Wang LW, 2016, INT J RADIAT ONCOL, V95, P396, DOI 10.1016/j.ijrobp.2016.02.028
   Wen PY, 2010, J CLIN ONCOL, V28, P1963, DOI 10.1200/JCO.2009.26.3541
   Yamaji N, 2020, J PEDIATR ONCOL NURS, V37, P431, DOI 10.1177/1043454220919711
   Zaky W, 2013, PEDIATR HEMAT ONCOL, V30, P623, DOI 10.3109/08880018.2013.829895
   Zhuang HQ, 2019, MOL CANCER, V18, DOI 10.1186/s12943-019-0950-1
NR 43
TC 3
Z9 3
U1 1
U2 4
PU MDPI
PI BASEL
PA ST ALBAN-ANLAGE 66, CH-4052 BASEL, SWITZERLAND
EI 2075-1729
J9 LIFE-BASEL
JI Life-Basel
PD APR
PY 2022
VL 12
IS 4
AR 566
DI 10.3390/life12040566
PG 13
WC Biology; Microbiology
WE Science Citation Index Expanded (SCI-EXPANDED)
SC Life Sciences & Biomedicine - Other Topics; Microbiology
GA 0R5UD
UT WOS:000785658700001
PM 35455057
OA gold, Green Published
DA 2023-08-27
ER

PT J
AU Raybaud, C
   Ramaswamy, V
   Taylor, MD
   Laughlin, S
AF Raybaud, Charles
   Ramaswamy, Vijay
   Taylor, Michael D.
   Laughlin, Suzanne
TI Posterior fossa tumors in children: developmental anatomy and diagnostic
   imaging
SO CHILDS NERVOUS SYSTEM
LA English
DT Article
DE Posterior fossa tumors; MR imaging; Brainstem glioma; DIPG;
   Medulloblastoma; Ependymoma
ID BRAIN-STEM GLIOMAS; MOLECULAR SUBGROUPS; MIDBRAIN TUMORS;
   MEDULLOBLASTOMA; CLASSIFICATION; EPENDYMOMAS; CELLS; SPECTROSCOPY;
   CEREBELLUM; DIFFUSION
AB Modern understanding of the relation between the mutated cancer stem cell and its site of origin and of its interaction with the tissue environment is enhancing the importance of developmental anatomy in the diagnostic assessment of posterior fossa tumors in children. The aim of this review is to show how MR imaging can improve on the exact identification of the tumors in the brainstem and in the vicinity of the fourth ventricle in children, using both structural imaging data and a precise topographical assessment guided by the developmental anatomy.
   The development of the hindbrain results from complex processes of brainstem segmentation, ventro-dorsal patterning, multiple germinative zones, and diverse migration pathways of the neural progenitors. Depending on their origin in the brainstem, gliomas may be infiltrative or not, as well as overwhelmingly malignant (pons), or mostly benign (cervicomedullary, medullo-pontine tegmental, gliomas of the cerebellar peduncles). In the vicinity of the fourth ventricles, the prognosis of the medulloblastomas (MB) correlates the molecular subtyping as well as the site of origin: WNT MB develop from the Wnt-expressing lower rhombic lip and have a good prognosis; SHH MB develop from the Shh-modulated cerebellar cortex with an intermediate prognosis (dependent on age); recurrences are local mostly. The poor prognosis group 3 MB is radiologically heterogeneous: some tumors present classic features but are juxtaventricular (rather than intraventricular); others have highly malignant features with a small principal tumor and an early dissemination. Group 4 MB has classic features, but characteristically usually does not enhance; dissemination is common. Although there is as yet no clear molecular subgrouping of the ependymomas, their sites of origin and their development can be clearly categorized, as most develop in an exophytic way from the ventricular surface of the medulla in clearly specific locations: the obex region with expansion in the cistern magna, or the lateral recess region with expansion in the CPA and prepontine cisterns (cerebellar ependymomas, and still more intra-brainstem ependymomas are rare). Finally, almost all cerebellar gliomas are pilocytic astrocytomas.
   A developmental and anatomic approach to the posterior fossa tumors in children (together with diffusion imaging data) provides a reliable pre-surgical identification of the tumor and of its aggressiveness.
C1 [Raybaud, Charles; Laughlin, Suzanne] Hosp Sick Children, Pediat Neuroradiol, Toronto, ON M5G 1X8, Canada.
   [Ramaswamy, Vijay] Hosp Sick Children, Neurooncol, Toronto, ON M5G 1X8, Canada.
   [Taylor, Michael D.] Hosp Sick Children, Neurosurg, Toronto, ON M5G 1X8, Canada.
C3 University of Toronto; Hospital for Sick Children (SickKids); University
   of Toronto; Hospital for Sick Children (SickKids); University of
   Toronto; Hospital for Sick Children (SickKids)
RP Raybaud, C (通讯作者)，Hosp Sick Children, Pediat Neuroradiol, 555 Univ Ave, Toronto, ON M5G 1X8, Canada.
EM
OI Ramaswamy, Vijay/0000-0002-6557-895X; Taylor,
   Michael/0000-0001-7009-3466
CR BOYDSTON WR, 1992, PEDIATR NEUROSURG, V17, P234, DOI 10.1159/000120603
   Broniscer A, 2006, CANCER, V106, P1364, DOI 10.1002/cncr.21749
   Cambronero F, 2000, J COMP NEUROL, V427, P522, DOI 10.1002/1096-9861(20001127)427:4<522::AID-CNE3>3.0.CO;2-Y
   Conway AE, 2014, NEURORADIOLOGY, V56, P405, DOI 10.1007/s00234-014-1348-9
   Curless RG, 2002, PEDIATR NEUROL, V26, P219, DOI 10.1016/S0887-8994(01)00363-0
   Daglioglu E, 2003, PEDIATR NEUROSURG, V38, P223, DOI 10.1159/000069823
   EPSTEIN F, 1987, J NEUROSURG, V67, P483, DOI 10.3171/jns.1987.67.4.0483
   EPSTEIN FJ, 1993, J NEUROSURG, V78, P408, DOI 10.3171/jns.1993.78.3.0408
   Gibson P, 2010, NATURE, V468, P1095, DOI 10.1038/nature09587
   Gilbertson RJ, 2007, CANCER RES, V67, P5579, DOI 10.1158/0008-5472.CAN-07-0760
   Godfraind C, 2012, ACTA NEUROPATHOL, V124, P247, DOI 10.1007/s00401-012-0981-9
   Gutierrez DR, 2014, AM J NEURORADIOL, V35, P1009, DOI 10.3174/ajnr.A3784
   IKEZAKI K, 1993, NEUROSURGERY, V32, P38, DOI 10.1227/00006123-199301000-00006
   Jandial R, 2008, NEUROSURG FOCUS, V24, DOI 10.3171/FOC/2008/24/3-4/E26
   KONNO H, 1985, ACTA NEUROPATHOL, V68, P155, DOI 10.1007/BF00688638
   Kool M, 2012, ACTA NEUROPATHOL, V123, P473, DOI 10.1007/s00401-012-0958-8
   Koral K, 2013, RADIOLOGY, V268, P532, DOI 10.1148/radiol.13121362
   Krieger Mark D, 2003, Neurosurg Focus, V15, pE5
   Laprie A, 2005, INT J RADIAT ONCOL, V62, P20, DOI 10.1016/j.ijrobp.2004.09.027
   Lobel U, 2011, AM J NEURORADIOL, V32, P315, DOI 10.3174/ajnr.A2277
   Louis DN, 2007, ACTA NEUROPATHOL, V114, P547, DOI 10.1007/s00401-007-0278-6
   Mack SC, 2014, NATURE, V506, P445, DOI 10.1038/nature13108
   Marcorelles P, 2005, CLIN NEUROPATHOL, V24, P155
   Martinez S, 2013, FRONT NEUROANAT, V7, DOI 10.3389/fnana.2013.00018
   Nakamura H, 2005, INT J DEV BIOL, V49, P231, DOI 10.1387/ijdb.041964hn
   Northcott PA, 2012, NAT REV NEUROL, V8, P340, DOI 10.1038/nrneurol.2012.78
   Northcott PA, 2011, J CLIN ONCOL, V29, P1408, DOI 10.1200/JCO.2009.27.4324
   Pajtler K, 2015, CANC CELL IN PRESS
   Panigrahy A, 2008, NEURO-ONCOLOGY, V10, P32, DOI 10.1215/15228517-2007-042
   Panigrahy A, 2010, PEDIATR RADIOL, V40, P3, DOI 10.1007/s00247-009-1450-z
   Perreault S, 2014, AM J NEURORADIOL, V35, P1263, DOI 10.3174/ajnr.A3990
   Raghunathan A, 2013, BRAIN PATHOL, V23, P584, DOI 10.1111/bpa.12050
   Ramaswamy V, 2015, NEUROONCOLOGY
   Ramaswamy V, 2014, PEDIATR BLOOD CANCER, V61, P1190, DOI [10.1002/pbc.24979, 10.1002/pbc.25002]
   Ramaswamy V, 2013, LANCET ONCOL, V14, P1200, DOI 10.1016/S1470-2045(13)70449-2
   Rauscher A, 2005, MAGNET RESON MED, V54, P87, DOI 10.1002/mrm.20520
   ROBERTSON PL, 1995, PEDIATR NEUROSURG, V22, P65, DOI 10.1159/000120879
   Roussel MF, 2011, CURR TOP DEV BIOL, V94, P235, DOI 10.1016/B978-0-12-380916-2.00008-5
   SANFORD RA, 1982, J NEUROSURG, V57, P690, DOI 10.3171/jns.1982.57.5.0690
   Sehgal V, 2005, J MAGN RESON IMAGING, V22, P439, DOI 10.1002/jmri.20404
   Sehgal V, 2006, J MAGN RESON IMAGING, V24, P41, DOI 10.1002/jmri.20598
   Sgaier SK, 2005, NEURON, V45, P27, DOI 10.1016/j.neuron.2004.12.021
   Sun B, 1999, NEURORADIOLOGY, V41, P158, DOI 10.1007/s002340050723
   Taylor MD, 2005, CANCER CELL, V8, P323, DOI 10.1016/j.ccr.2005.09.001
   Taylor MD, 2012, ACTA NEUROPATHOL, V123, P465, DOI 10.1007/s00401-011-0922-z
   ten Donkelaar HJ, 2006, CLIN NEUROEMBRYOLOGY, P274
   Ternier J, 2006, J NEUROSURG, V104, P369, DOI 10.3171/ped.2006.104.6.369
   Tihan T, 2008, MODERN PATHOL, V21, P165, DOI 10.1038/modpathol.3800999
   Tumpel S, 2009, CURR TOP DEV BIOL, V88, P103, DOI 10.1016/S0070-2153(09)88004-6
   U-King-Im JM, 2010, CHILD NERV SYST, V26, P1765, DOI 10.1007/s00381-010-1251-6
   VANDERTOP WP, 1992, NEUROSURGERY, V31, P186, DOI 10.1227/00006123-199208000-00003
   Witt H, 2012, CURR OPIN NEUROL, V25, P745, DOI 10.1097/WCO.0b013e328359cdf5
   Witt H, 2011, CANCER CELL, V20, P143, DOI 10.1016/j.ccr.2011.07.007
   Wullimann MF, 2011, FRONT NEUROANAT, V5, DOI 10.3389/fnana.2011.00027
   Xu QL, 2013, WIRES DEV BIOL, V2, P735, DOI 10.1002/wdev.106
   Yeom KW, 2013, AM J ROENTGENOL, V200, P895, DOI 10.2214/AJR.12.9249
   Zhukova N, 2013, J CLIN ONCOL, V31, P2927, DOI 10.1200/JCO.2012.48.5052
NR 57
TC 44
Z9 46
U1 0
U2 11
PU SPRINGER
PI NEW YORK
PA ONE NEW YORK PLAZA, SUITE 4600, NEW YORK, NY, UNITED STATES
SN 0256-7040
EI 1433-0350
J9 CHILD NERV SYST
JI Childs Nerv. Syst.
PD OCT
PY 2015
VL 31
IS 10
BP 1661
EP 1676
DI 10.1007/s00381-015-2834-z
PG 16
WC Clinical Neurology; Pediatrics; Surgery
WE Science Citation Index Expanded (SCI-EXPANDED)
SC Neurosciences & Neurology; Pediatrics; Surgery
GA CR0FF
UT WOS:000360993100005
PM 26351220
DA 2023-08-27
ER

PT J
AU Colosimo, C
   Tartaglione, T
   DiLella, GM
   Cerase, A
AF Colosimo, C
   Tartaglione, T
   DiLella, GM
   Cerase, A
TI Neuroradiology of brain stem and cervicomedullary tumours
SO RIVISTA DI NEURORADIOLOGIA
LA English
DT Article
DE brain tumors; brainstem; cervico-medullary junction; gliomas; magnetic
   resonance imaging
ID TUMORS; CHILDREN; GLIOMAS; PET
AB Brain stem and cervicomedullary tumours are typical of paediatric age, 80% of them occurring in patients under 18 years of age, and comprising 10-15% of all childhood and adolescent brain tumours, as well as 20-25% of infratentorial locations.
   They are characteristically pontine tumours (60% of the cases), but they commonly extend to involve the medulla, midbrain and cerebellum. Although most brain stem tumours are low grade gliomas, their prognosis is extremely severe (no more than 20% of patients are alive 3 years after diagnosis and the 5-year survival rate is 5%) because both the typical infiltrating nature and the neuro-anatomical location usually make them surgically unresectable. Surgery is generally limited to biopsy, partial decompression, or excission of the exophytic components, because of the extremely severe functional sequelae of even minor resections. Thus, the mainstay of therapy has been based on irradiation alone or combined with chemotherapy, doses of 5000-5500 cGy being usually adequate for tumour shrinkage or remission, even if recurrence is common after 10-15 months.
   At present, it is very important to establish reliable, homogeneous, objective, and reproducible diagnostic criteria for the identification of patient subsets with predictable histology, prognosis and possible therapeutic management, in some cases histology, site and relationship of tumour enabling total or subtotal resection with a lower operative risk. Since its introduction, magnetic resonance imaging (MRI) has appeared the procedure of choice for the neuroradiologic study of the brain stem and brain stem tumours, enabling a more precise definition of their margins, a correct assessment of intrinsic and exophytic components, as well as a satisfactory characterization of pathologic tissue. MRI studies should include good quality T1-, PD- and T2-weighted images and T1-weighted images after gadolinium i.v., T2-weighted sagittal images being required for complete evaluation of tumour extent. Computed tomography is still superior in the identification of calcifications and acute intratumoral haemorrhage; it is rapidly performed, thus representing the first choice procedure in emergency, i.e. the diagnosis of hydrocephalus. Epstein has proposed the most widely accepted classification system of brain stem tumours, essentially based on neuroradiological findings, surgical and stereotactic biopsy and histology generally resulting in understaging. This classification system separates intrinsic (diffuse, focal, cervico-medullary), exophytic (anterolateral into cerebellopontine angle, posterolateral into brachium pontis, posterior into fourth ventricle) and cerebrospinal fluid seeding (positive cytology or myelography) tumors.
   More recently, Barkovich - based on a multicentric study of some of the most important paediatric neurosurgery and neuro-oncology centres of the United States - has clearly defined the neuroradiological parameters which must be considered for an objective and reproducible assessment of brain stem gliomas, in order to identify patient subsets characterized by predictable histology, prognosis and possible therapeutic management. We agree with him, emphasizing that the evaluation of brain stem tumours must include a careful interpretation of all MRI findings (tumour site and origin; dimensions/degree of brain stem enlargement; tumour caudo-cranial and transverse extension; exophytic components; tumour characteristics as defined by MRI signal intensity; cysts, haemorrhage, necrosis, calcifications; ventricular dimensions and hydrocephalus; leptomeninengeal seeding) that can help in the definition of the following tumour subsets: diffuse pontine tumours, medullary tumours, cervicomedullary tumours, focal brain stem tumours.
   With regard to the neuroradiological follow-up, in patients undergoing surgery (for biopsy decompression or less frequently for radicality), the role of the neuroradiologist is similar to that in other fields of neurosurgery, and concerns the quantitation of the extent of the resection and the identification of possible parenchymal injuries or postoperative haemorrhage, always keeping in mind the negative effects of postoperative reactive phenomena and the blood-brain disruption 24-48 hours to 30-40 days after surgery. However, the major contribution of the neuroradiologist is the objective evaluation of irradiation effects and recently of combined irradiation and chemotherapy. In our experience, the efficacy of irradiation is well evaluated by MRI only 3 or 6 months after the end of treatment, even if a clinical improvement is possible after 30-60 days. However, tumour shrinkage is rarely drastic, and its disappearance, as well as that of signal alterations, is exceptional. Qualitative modifications occurring within the tumour are more difficult to interpret, because of the appearance of cystic or pseudocystic areas, markedly increased enhancement, and small areas of haemorrhage may be related both to irradiation-induced regressive modifications and disease progression,
   In conclusion, MRI represents the gold standard in the evaluation of brain stem and cervicomedullary tumors, always enabling a precise definition of tumour site and extent, and in most cases the diagnosis of nature, thus allowing the identification of patients who can undergo radical microsurgery. MRI follow-up controls the extent of resection and the effect of combined irradiation and chemotherapy; disease progression is evidenced and spinal seeding can be diagnosed.
   However, the differentiation between tumour recurrence and irradiation-induced injury may be difficult if only based on morphological data. These limitations of MRI will probably be reduced by the advances in ultra-fast MRI technology and 18F-fluoro-deoxy-glucose positron emission tomography which supplies in vivo metabolic and functional information.
C1 UNIV GABRIELE DANNUNZIO,IST SCI RADIOL,CHIETI,ITALY.
   UNIV CATTOLICA SACRO CUORE,IST RADIOL,POLICLIN AGOSTINO GEMELLI,I-00168 ROME,ITALY.
C3 G d'Annunzio University of Chieti-Pescara; Catholic University of the
   Sacred Heart; IRCCS Policlinico Gemelli
RI TARTAGLIONE, TOMMASO/K-5390-2017; Cerase, Alfonso/AFJ-8833-2022
OI TARTAGLIONE, TOMMASO/0000-0003-3896-4078; Cerase,
   Alfonso/0000-0003-4753-6649
CR BARKOVICH AJ, 1991, PEDIATR NEUROSURG, V16, P73, DOI 10.1159/000120511
   BRUGGERS CS, 1993, MED PEDIATR ONCOL, V21, P301, DOI 10.1002/mpo.2950210414
   BRUNO L, 1982, PEDIATRIC NEUROSURGE, P361
   Colosimo C, 1993, Rays, V18, P541
   EPSTEIN F, 1988, J NEURO-ONCOL, V6, P309
   EPSTEIN FJ, 1993, J NEUROSURG, V78, P408, DOI 10.3171/jns.1993.78.3.0408
   EPSTEIN FJ, 1989, CANCER, V56, P1804
   FARWELL JR, 1977, CANCER, V40, P3123, DOI 10.1002/1097-0142(197712)40:6<3123::AID-CNCR2820400656>3.0.CO;2-6
   FREEMAN CR, 1986, INT J RADIAT ONCOL, V12, P1823, DOI 10.1016/0360-3016(86)90325-1
   FULTON DS, 1981, J NEUROSURG, V54, P721, DOI 10.3171/jns.1981.54.6.0721
   GUINEY MJ, 1993, INT J RADIAT ONCOL, V21, P235
   HOFFMAN JM, 1992, J COMPUT ASSIST TOMO, V16, P62, DOI 10.1097/00004728-199201000-00011
   KONE AG, 1993, AM J NEURORADIOL, V14, P941
   NELSON MD, 1994, RADIOLOGY, V191, P279, DOI 10.1148/radiology.191.1.8134588
   Packer R J, 1992, Neurosurg Clin N Am, V3, P863
   PACKER RJ, 1990, NEUROLOGY, V40, P1237
   ROBERTSON PL, 1994, NEUROLOGY, V44, P1798, DOI 10.1212/WNL.44.10.1798
   SMITH RR, 1980, NEURORADIOLOGY, V32, P265
   ZELTZER PM, 1985, CANCER, V56, P1824, DOI 10.1002/1097-0142(19851001)56:7+<1824::AID-CNCR2820561321>3.0.CO;2-H
NR 19
TC 3
Z9 3
U1 1
U2 6
PU EDIZIONI CENTAURO
PI BOLOGNA
PA VIA DEL PRATELLO, 8, 40122 BOLOGNA, ITALY
SN 1120-9976
J9 RIV NEURORADIOL
JI Riv. Neuroradiol.
PD FEB
PY 1997
VL 10
IS 1
BP 41
EP 54
DI 10.1177/197140099701000104
PG 14
WC Neurosciences; Neuroimaging; Radiology, Nuclear Medicine & Medical
   Imaging
WE Science Citation Index Expanded (SCI-EXPANDED)
SC Neurosciences & Neurology; Radiology, Nuclear Medicine & Medical Imaging
GA XB755
UT WOS:A1997XB75500004
DA 2023-08-27
ER

PT J
AU Klimo, P
   Goumnerova, LC
AF Klimo, Paul, Jr.
   Goumnerova, Liliana C.
TI Endoscopic third ventriculocisternostomy for brainstem tumors
SO JOURNAL OF NEUROSURGERY
LA English
DT Article
DE pontine glioma; brainstem tumor; endoscopic third ventriculostomy;
   pediatric neurosurgery
ID OBSTRUCTIVE HYDROCEPHALUS; VENTRICULOSTOMY; 3RD-VENTRICULOSTOMY;
   COMPLICATIONS; MANAGEMENT; SHUNT
AB Object. The authors retrospectively reviewed the charts of all patients harboring brainstem tumors treated at their institution, excluding those with tectal gliomas, who underwent an endoscopic third ventriculocisternostomy.
   Methods. Endoscopic third ventriculocisternostomy was performed in 13 patients with tumors involving the brainstem: nine patients with diffuse pontine gliomas, two with posterior fossa ependymomas, one with a cervicomedullary tumor, and one with a pontine primitive neuroectodermal tumor. No technical difficulties attributable to the location of the tumors or surgery-related complications were encountered. Immediate symptomatic relief of hydrocephalus was achieved in all patients, and there was an associated decrease in steroid and analgesic agent requirements. Only one patient eventually required a shunt.
   Conclusions. Endoscopic third ventriculocisternostomy can be used in the terminal treatment of patients with brainstem tumors, yielding good results without significant surgical morbidity.
C1 Harvard Univ, Sch Med, Childrens Hosp, Dept Neurosurg, Boston, MA 02115 USA.
C3 Harvard University; Boston Children's Hospital; Harvard Medical School
RP Goumnerova, LC (通讯作者)，Harvard Univ, Sch Med, Childrens Hosp, Dept Neurosurg, Bader 3,300 Longwood Ave, Boston, MA 02115 USA.
EM
CR Amano T, 2002, CHILD NERV SYST, V18, P599, DOI 10.1007/s00381-002-0637-5
   [Anonymous], BOSTON MED SURG J
   BLUM KS, 2001, TUMORS PEDIAT CENTRA, P206
   Boschert J, 2003, J NEUROSURG, V98, P1032, DOI 10.3171/jns.2003.98.5.1032
   Buxton N, 2000, NEUROSURGERY, V46, P999
   Dandy WE, 1922, B JOHNS HOPKINS HOSP, V33, P189
   Feng HL, 2004, J NEUROSURG, V100, P626, DOI 10.3171/jns.2004.100.4.0626
   Garton HJL, 2002, NEUROSURGERY, V51, P69, DOI 10.1097/00006123-200207000-00012
   Goumnerova LC, 1997, PEDIATR NEUROSURG, V27, P149, DOI 10.1159/000121242
   Grunert P, 2003, MINIM INVAS NEUROSUR, V46, P16, DOI 10.1055/s-2003-37957
   Iantosca MR, 2004, NEUROSURG CLIN N AM, V15, P67, DOI 10.1016/S1042-3680(03)00067-6
   Kadrian D, 2005, NEUROSURGERY, V56, P1271, DOI 10.1227/01.NEU.0000159712.48093.AD
   MASUZAWA H, 1993, CHILD NERV SYST, V9, P256, DOI 10.1007/BF00306266
   Rekate HL, 2004, NEUROSURG CLIN N AM, V15, P39, DOI 10.1016/S1042-3680(03)00074-3
   Santamarta D, 2005, ACTA NEUROCHIR, V147, P377, DOI 10.1007/s00701-005-0484-8
   Schroeder HWS, 2002, J NEUROSURG, V96, P1032, DOI 10.3171/jns.2002.96.6.1032
   Tuli S, 1999, PEDIATR NEUROSURG, V30, P11, DOI 10.1159/000028753
   Walker ML, 2004, NEUROSURG CLIN N AM, V15, P61, DOI 10.1016/S1042-3680(03)00070-6
NR 18
TC 22
Z9 22
U1 0
U2 0
PU AMER ASSOC NEUROLOGICAL SURGEONS
PI CHARLOTTESVILLE
PA UNIV VIRGINIA, 1224 WEST MAIN ST, STE 450, CHARLOTTESVILLE, VA 22903 USA
SN 0022-3085
J9 J NEUROSURG
JI J. Neurosurg.
PD OCT
PY 2006
VL 105
IS 4
SU S
BP 271
EP 274
DI 10.3171/ped.2006.105.4.271
PG 4
WC Clinical Neurology; Surgery
WE Science Citation Index Expanded (SCI-EXPANDED)
SC Neurosciences & Neurology; Surgery
GA 093WO
UT WOS:000241201200007
PM 17328276
DA 2023-08-27
ER

PT J
AU Dellaretti, M
   Reyns, N
   Touzet, G
   Dubois, F
   Gusmao, S
   Pereira, JLB
   Blond, S
AF Dellaretti, Marcos
   Reyns, Nicolas
   Touzet, Gustavo
   Dubois, Francois
   Gusmao, Sebastiao
   Barbosa Pereira, Julio Leonardo
   Blond, Serge
TI Stereotactic Biopsy for Brainstem Tumors: Comparison of Transcerebellar
   with Transfrontal Approach
SO STEREOTACTIC AND FUNCTIONAL NEUROSURGERY
LA English
DT Article
DE Brainstem tumors; Biopsy; Transcerebellar approach; Transfrontal
   approach
ID GLIOMAS; DIAGNOSIS; CHILDREN; LESIONS
AB Background: An important aspect of evaluating patients submitted to stereotactic biopsy of the brainstem is the trajectory used. The literature describes two principal approaches:the suboccipital transcerebellar and the transfrontal; however, no studies exist comparing these two techniques. Objective:The purpose of this study was to compare diagnosis success rates and complications between the suboccipital transcerebellar and transfrontal trajectories. Methods: The study evaluated 142 patients submitted to stereotactic biopsy. The patients presented brainstem tumors in the following areas: pons (n = 31), midbrain (n = 36), medulla (n = 2), pons-medulla (n = 30), pons-nnidbrain (n = 33), and midbrain-pons-medulla (n = 10). On 123 patients, the transfrontal approach was used, and on 19 the suboccipital transcerebellar approach. Results: Comparing success rates between the two approaches, it was observed that in the group of patients submitted to the transfrontal approach, 95.1% (117 cases) were successful, while in those submitted to the suboccipital transcerebellar approach, 84.2% (16 cases) were successful. Despite a higher success rate among patients in the first group, the difference was not statistically significant. Regarding complications, in patients who were biopsied via the transfrontal trajectory, the morbidity rate was 9.8% (12 cases), while in patients submitted to the suboccipital transcerebellar approach, the morbidity rate was 5.3% (1 case) and the mortality rate 5.3% (1 case). Conclusions:This study verified a higher diagnosis rate in patients submitted to the transfrontal approach than in those submitted to the suboccipital transcerebellar approach (95.1 vs. 84.2%); however, the difference was not statistically significant. Regarding complications, the rate was similar in both groups of patients. Copyright (c) 2012 S. Karger AG, Basel
C1 [Dellaretti, Marcos; Barbosa Pereira, Julio Leonardo] Santa Casa Hosp, Dept Neurosurg, Belo Horizonte, MG, Brazil.
   [Dellaretti, Marcos; Reyns, Nicolas; Touzet, Gustavo; Dubois, Francois; Blond, Serge] Hosp Roger Salengro, Dept Neurosurg, Lille, France.
   [Dellaretti, Marcos; Gusmao, Sebastiao] Univ Fed Minas Gerais, Dept Neurosurg, Belo Horizonte, MG, Brazil.
C3 Universite de Lille - ISITE; CHU Lille; Universidade Federal de Minas
   Gerais
RP Dellaretti, M (通讯作者)，Santa Casa Hosp, Dept Neurosurg, Belo Horizonte, MG, Brazil.
EM
RI Pereira, Julio/H-3324-2011; Dellaretti, Marcos/G-4664-2012
OI Pereira, Julio/0000-0003-0303-7506; 
CR Albright AL, 1996, PEDIATR NEUROSURG, V24, P252
   ALBRIGHT AL, 1993, NEUROSURGERY, V33, P1026
   Frank F, 1988, Acta Neurochir Suppl (Wien), V42, P177
   Frazier JL, 2009, J NEUROSURG-PEDIATR, V3, P259, DOI 10.3171/2008.11.PEDS08281
   GLEASON CA, 1978, NEUROSURGERY, V2, P217, DOI 10.1227/00006123-197805000-00006
   HOOD TW, 1986, J NEUROSURG, V65, P172, DOI 10.3171/jns.1986.65.2.0172
   KONDZIOLKA D, 1995, SURG NEUROL, V43, P558, DOI 10.1016/0090-3019(95)00009-7
   Laigle-Donadey F, 2008, CURR OPIN ONCOL, V20, P662, DOI 10.1097/CCO.0b013e32831186e0
   Pincus DW, 2006, J NEUROSURG, V104, P108, DOI 10.3171/ped.2006.104.2.108
   Pirotte BJM, 2007, J NEUROSURG, V107, P392, DOI 10.3171/PED-07/11/392
   Rachinger W, 2009, J NEUROL NEUROSUR PS, V80, P1134, DOI 10.1136/jnnp.2009.174250
   Reni M, 2001, ANN HEMATOL, V80, pB113
   Roujeau T, 2007, J NEUROSURG, V107, P1, DOI 10.3171/PED-07/07/001
   Sanai N, 2008, NEUROSURGERY, V63, P460, DOI 10.1227/01.NEU.0000324731.68843.74
   Schumacher M, 2007, J NEUROSURG, V106, P111, DOI 10.3171/ped.2007.106.2.111
NR 15
TC 35
Z9 35
U1 0
U2 3
PU KARGER
PI BASEL
PA ALLSCHWILERSTRASSE 10, CH-4009 BASEL, SWITZERLAND
SN 1011-6125
EI 1423-0372
J9 STEREOT FUNCT NEUROS
JI Stereotact. Funct. Neurosurg.
PY 2012
VL 90
IS 2
BP 79
EP 83
DI 10.1159/000335502
PG 5
WC Neurosciences; Neuroimaging; Surgery
WE Science Citation Index Expanded (SCI-EXPANDED)
SC Neurosciences & Neurology; Surgery
GA 935BA
UT WOS:000303490800002
PM 22286495
DA 2023-08-27
ER

PT J
AU Jallo, GI
   Biser-Rohrbaugh, A
   Freed, D
AF Jallo, GI
   Biser-Rohrbaugh, A
   Freed, D
TI Brainstem gliomas
SO CHILDS NERVOUS SYSTEM
LA English
DT Review
DE brainstem glioma; cervicomedullary; midbrain tumors; pontine glioma;
   surgery
ID POSTERIOR-FOSSA TUMORS; CHILDRENS CANCER GROUP; PEDIATRIC-PATIENTS;
   HYPERFRACTIONATED RADIOTHERAPY; CERVICOMEDULLARY JUNCTION; SURGICAL
   INDICATIONS; INTRAAXIAL TUMORS; CHILDHOOD; NEUROFIBROMATOSIS;
   CHEMOTHERAPY
AB Introduction. Brainstem gliomas have historically been one of the most difficult pediatric cancers to treat. Tumors arising in the brainstem were once uniformly discounted as surgically unresectable lesions. Early neurosurgeons thought this location to be inoperable and fraught with disaster. The advent of computed tomography (CT), magnetic resonance imaging (MRI), and sophisticated neurophysiological monitoring techniques have significantly advanced the surgical treatment of these precarious lesions. Review. Brainstem gliomas are now recognized as a heterogenous group of tumors. They have been broadly classified into several categories depending upon the classification scheme. All these classification systems provide a framework to predict growth patterns, surgical resectability, and overall prognosis of these tumors. These systems allow the surgeon to obtain a better understanding of the distinction between low-grade tumors and diffuse inoperable tumor types. The authors review the current literature and management of brainstem tumors.
C1 Johns Hopkins Univ Hosp, Div Pediat Neurosurg, Baltimore, MD 21287 USA.
C3 Johns Hopkins University; Johns Hopkins Medicine
RP Jallo, GI (通讯作者)，Johns Hopkins Univ Hosp, Div Pediat Neurosurg, 600 N Wolfe St,Harvey 811, Baltimore, MD 21287 USA.
EM
CR Abbott R, 1993, Neurosurg Clin N Am, V4, P519
   Abbott R, 1996, PEDIATR NEUROSURG, V25, P41, DOI 10.1159/000121095
   ABBOTT R, 1992, PEDIATR NEUROSURG, V18, P153, DOI 10.1159/000120656
   ABBOTT R, 1992, PEDIATR NEUROSURG, V17, P239, DOI 10.1159/000120604
   ALBRIGHT AL, 1986, J NEUROSURG, V65, P751, DOI 10.3171/jns.1986.65.6.0751
   ALBRIGHT AL, 1993, NEUROSURGERY, V33, P1026
   ALBRIGHT AL, 1996, NEUROLOGICAL SURG, P2603
   Allen J., 1983, NEUROL NEUROSURG UPD, V4, P2
   Allen JC, 1996, PEDIATR NEUROSURG, V24, P98, DOI 10.1159/000121024
   ALVISI C, 1985, ACTA NEUROCHIR, V76, P12, DOI 10.1007/BF01403823
   Ammirati M, 2002, J NEUROSURG, V97, P922, DOI 10.3171/jns.2002.97.4.0922
   [Anonymous], 2000, OPERATIVE TECHNIQUES
   BARKOVICH AJ, 1991, PEDIATR NEUROSURG, V16, P73, DOI 10.1159/000120511
   Behnke J, 1997, CHILD NERV SYST, V13, P122, DOI 10.1007/s003810050058
   Beltramello A, 2000, OPERATIVE TECHNIQUES, V3, P87
   BERGER MS, 1983, NEUROSURGERY, V12, P298, DOI 10.1227/00006123-198303000-00008
   Blaney S, 2001, CLIN CANCER RES, V7, P32
   Bowers DC, 2000, PEDIATR NEUROSURG, V32, P24, DOI 10.1159/000028893
   Bowers DC, 2001, PEDIATR NEUROSURG, V34, P229, DOI 10.1159/000056027
   Bricolo A, 2000, OPER TECH NEUROSURG, V3, P137, DOI DOI 10.1053/oy.2000.6559
   Broniscer A, 1997, NEUROSURGERY, V40, P331, DOI 10.1097/00006123-199702000-00018
   Broniscer A, 2000, J CLIN ONCOL, V18, P1246, DOI 10.1200/JCO.2000.18.6.1246
   Choux M, 2000, PEDIAT NEUROSURGERY, P471
   CLARSON BSS, 2002, ADV CLIN NEUROSCI, V12, P89
   CONSTANTINI S, 1996, NEUROSURGERY, P2714
   Dechambre S, 1999, NEURORADIOLOGY, V41, P829, DOI 10.1007/s002340050850
   Deletis V, 2000, OPERATIVE TECHNIQUES, V3, P109, DOI DOI 10.1053/oy.2000.6562
   EPSTEIN F, 1985, CANCER, V56, P1804, DOI 10.1002/1097-0142(19851001)56:7+<1804::AID-CNCR2820561316>3.0.CO;2-5
   EPSTEIN F, 1988, J NEURO-ONCOL, V6, P309
   EPSTEIN F, 1986, J NEUROSURG, V64, P11, DOI 10.3171/jns.1986.64.1.0011
   EPSTEIN F, 1987, J NEUROSURG, V67, P483, DOI 10.3171/jns.1987.67.4.0483
   EPSTEIN FJ, 1993, J NEUROSURG, V78, P408, DOI 10.3171/jns.1993.78.3.0408
   EPSTEIN FJ, 1993, J NEUROSURG, V78, P505, DOI 10.3171/jns.1993.78.3.0505
   Fischbein NJ, 1996, PEDIATR NEUROSURG, V24, P9, DOI 10.1159/000121010
   Freeman CR, 1996, CANCER-AM CANCER SOC, V77, P555
   Freeman CR, 2000, INT J RADIAT ONCOL, V47, P561, DOI 10.1016/S0360-3016(00)00471-5
   GILDAY DL, 1975, RADIOLOGY, V117, P93, DOI 10.1148/117.1.93
   Gnanalingham KK, 2002, J NEUROSURG, V97, P821, DOI 10.3171/jns.2002.97.4.0821
   HOFFMAN HJ, 1980, NEUROSURGERY, V7, P243, DOI 10.1227/00006123-198009000-00007
   Jallo GI, 2002, CHILD NERV SYST, V18, P333, DOI 10.1007/s00381-002-0600-5
   Jallo GI, 2001, NEUROSURGERY, V48, P695, DOI 10.1097/00006123-200103000-00054
   Kaplan AM, 1996, PEDIATR NEUROSURG, V24, P185, DOI 10.1159/000121036
   KEDAR A, 1994, MED PEDIATR ONCOL, V23, P428, DOI 10.1002/mpo.2950230507
   KHATIB ZA, 1994, PEDIATR NEUROSURG, V20, P2, DOI 10.1159/000120759
   Kurpad SN, 1999, PEDIATR NEUROSURG, V31, P54, DOI 10.1159/000028833
   LITTMAN P, 1980, CANCER, V45, P2787, DOI 10.1002/1097-0142(19800601)45:11<2787::AID-CNCR2820451113>3.0.CO;2-V
   Lonser RR, 2002, J NEUROSURG, V97, P905, DOI 10.3171/jns.2002.97.4.0905
   Matson DD, 1969, NEUROSURGERY INFANCY, P469
   MILSTEIN JM, 1989, J NEURO-ONCOL, V7, P367, DOI 10.1007/BF02147094
   MOROTA N, 1995, NEUROSURGERY, V37, P922, DOI 10.1227/00006123-199511000-00011
   Murakami N, 1999, NEUROSURG REV, V22, P159
   POLLACK IF, 1995, NEUROSURGERY, V37, P885, DOI 10.1227/00006123-199511000-00006
   POLLACK IF, 1993, J NEUROSURG, V78, P859, DOI 10.3171/jns.1993.78.6.0859
   RAFFEL C, 1989, NEUROSURGERY, V25, P959, DOI 10.1227/00006123-198912000-00018
   Rhoton AL, 2000, NEUROSURGERY, V47, pS7, DOI 10.1097/00006123-200009001-00007
   ROBERTSON PL, 1994, NEUROLOGY, V44, P1798, DOI 10.1212/WNL.44.10.1798
   ROSE E, 1998, AESTHETIC FACIAL RES
   Sandberg DI, 2002, J NEUROSURG, V96, P885, DOI 10.3171/jns.2002.96.5.0885
   STEIN BM, 1971, J NEUROSURG, V35, P197, DOI 10.3171/jns.1971.35.2.0197
   Steinbok P, 2003, PEDIATR NEUROSURG, V39, P179, DOI 10.1159/000072468
   Storm PB, 2003, NEUROSURGERY, V52, P1169, DOI 10.1227/01.NEU.0000057835.70364.34
   STROINK AR, 1987, NEUROSURGERY, V20, P439, DOI 10.1227/00006123-198703000-00014
   Walker DA, 1999, ARCH DIS CHILD, V80, P558, DOI 10.1136/adc.80.6.558
   Walter AW, 1998, MED PEDIATR ONCOL, V30, P28, DOI 10.1002/(SICI)1096-911X(199801)30:1<28::AID-MPO9>3.0.CO;2-2
   Wang GH, 2001, NEUROSURG QUART, V11, P302, DOI 10.1097/00013414-200112000-00006
   Weiner HL, 1997, PEDIATR NEUROSURG, V27, P12, DOI 10.1159/000121219
   Wisoff J H, 1984, Neurosurgery, V15, P707
   Zagzag D, 2000, PEDIATRICS, V106, P1045, DOI 10.1542/peds.106.5.1045
NR 68
TC 100
Z9 110
U1 0
U2 8
PU SPRINGER
PI NEW YORK
PA 233 SPRING ST, NEW YORK, NY 10013 USA
SN 0256-7040
EI 1433-0350
J9 CHILD NERV SYST
JI Childs Nerv. Syst.
PD MAR
PY 2004
VL 20
IS 3
BP 143
EP 153
DI 10.1007/s00381-003-0870-6
PG 11
WC Clinical Neurology; Pediatrics; Surgery
WE Science Citation Index Expanded (SCI-EXPANDED)
SC Neurosciences & Neurology; Pediatrics; Surgery
GA 802VN
UT WOS:000220191100002
PM 14669023
DA 2023-08-27
ER

PT J
AU Klimo, P
   Nesvick, CL
   Broniscer, A
   Orr, BA
   Choudhri, AF
AF Klimo, Paul, Jr.
   Nesvick, Cody L.
   Broniscer, Alberto
   Orr, Brent A.
   Choudhri, Asim F.
TI Malignant brainstem tumors in children, excluding diffuse intrinsic
   pontine gliomas
SO JOURNAL OF NEUROSURGERY-PEDIATRICS
LA English
DT Article
DE brainstem tumor; malignant; glioma; embryonal; outcomes; resection;
   oncology
ID PRIMITIVE NEUROECTODERMAL TUMORS; GROSS TOTAL RESECTION; SAFE ENTRY
   ZONES; PEDIATRIC-PATIENTS; ABUNDANT NEUROPIL; EMBRYONAL TUMOR;
   NERVOUS-SYSTEM; TRUE ROSETTES; MEDULLOBLASTOMA; MANAGEMENT
AB OBJECTIVE Malignant tumors of the brainstem, excluding classic diffuse intrinsic pontine gliomas (DIPGs), are a very rare, heterogeneous group of neoplasms that have been infrequently described in the literature. In this paper, the authors present their experiences with treating these unique cancers.
   METHODS A retrospective chart review was conducted to identify eligible cases over a 15-year period. All tumors involving the pons were, by consensus, felt not to be DIPGs based on their neuroimaging features. Demographic information, pathological specimens, neuroimaging characteristics, surgical and nonsurgical management plans, and survival data were gathered for analysis.
   RESULTS Between January 2000 and December 2014, 29 patients were identified. The mean age at diagnosis was 8.4 years (range 2 months to 25 years), and 17 (59%) patients were male. The most common presenting signs and symptoms were cranial neuropathies (n = 24; 83%), hemiparesis (n = 12; 41%), and ataxia or gait disturbance (n = 10; 34%). There were 18 glial and 11 embryonal tumors. Of the glial tumors, 5 were radiation-induced and 1 was a malignant transformation of a previously known low-grade tumor. Surgical intervention consisted of biopsy alone in 12 patients and some degree of resection in another 15 patients. Two tumors were diagnosed postmortem. The median overall survival for all patients was 196 days (range 15 to 3999 days). There are currently 5 (17%) patients who are still alive: 1 with an anaplastic astrocytoma and the remaining with embryonal tumors.
   CONCLUSIONS In general, malignant non-DIPG tumors of the brainstem carry a poor prognosis. However, maximal cytoreductive surgery may be an option for select patients with focal tumors. Long-term survival is possible in patients with nonmetastatic embryonal tumors after multimodal treatment, most importantly maximal resection.
C1 [Klimo, Paul, Jr.; Choudhri, Asim F.] Le Bonheur Childrens Hosp, Le Bonheur Neurosci Inst, Memphis, TN USA.
   [Klimo, Paul, Jr.] St Jude Childrens Res Hosp, Dept Surg, Memphis, TN 38105 USA.
   [Broniscer, Alberto] St Jude Childrens Res Hosp, Dept Oncol, Memphis, TN 38105 USA.
   [Orr, Brent A.] St Jude Childrens Res Hosp, Dept Pathol, Memphis, TN 38105 USA.
   [Klimo, Paul, Jr.] Semmes Murphey Neurol & Spine Inst, Memphis, TN USA.
   [Klimo, Paul, Jr.; Choudhri, Asim F.] Univ Tennessee, Hlth Sci Ctr, Dept Neurosurg, Memphis, TN USA.
   [Broniscer, Alberto] Univ Tennessee, Hlth Sci Ctr, Dept Pediat, Memphis, TN USA.
   [Choudhri, Asim F.] Univ Tennessee, Hlth Sci Ctr, Dept Radiol, Memphis, TN USA.
   [Nesvick, Cody L.] Univ Tennessee, Hlth Sci Ctr, Memphis, TN USA.
C3 St Jude Children's Research Hospital; St Jude Children's Research
   Hospital; St Jude Children's Research Hospital; University of Tennessee
   System; University of Tennessee Health Science Center; University of
   Tennessee System; University of Tennessee Health Science Center;
   University of Tennessee System; University of Tennessee Health Science
   Center; University of Tennessee System; University of Tennessee Health
   Science Center
RP Klimo, P (通讯作者)，Semmes Murphey Neurol & Spine Clin, 6325 Humphreys Blvd, Memphis, TN 38120 USA.
EM
RI Orr, Brent/N-8068-2018; Nesvick, Cody/AAS-5327-2020
OI Orr, Brent/0000-0003-0997-4728
CR Adamek D, 2013, CHILD NERV SYST, V29, P849, DOI 10.1007/s00381-013-2037-4
   Albright AL, 1996, NEUROSURGERY, V38, P265, DOI 10.1097/00006123-199602000-00007
   Behdad A, 2010, BRAIN PATHOL, V20, P441, DOI 10.1111/j.1750-3639.2009.00314.x
   Behnke J, 1996, CHILD NERV SYST, V12, P125, DOI 10.1007/BF00266812
   Cage TA, 2013, CHILD NERV SYST, V29, P1313, DOI 10.1007/s00381-013-2101-0
   Camacho CV, 2015, ONCOGENE, V34, P1064, DOI 10.1038/onc.2014.29
   Chan KM, 2013, GENE DEV, V27, P985, DOI 10.1101/gad.217778.113
   Choudhri AF, 2014, AM J NEURORADIOL, V35, P2382, DOI 10.3174/ajnr.A4040
   Choudhri AF, 2014, NEURORADIOLOGY, V56, P169, DOI 10.1007/s00234-013-1316-9
   Khuong-Quang DA, 2012, ACTA NEUROPATHOL, V124, P439, DOI 10.1007/s00401-012-0998-0
   Ellison DW, 2011, ACTA NEUROPATHOL, V121, P381, DOI 10.1007/s00401-011-0800-8
   Fangusaro J, 2009, J CHILD NEUROL, V24, P1409, DOI 10.1177/0883073809338960
   Fangusaro JR, 2008, PEDIATR BLOOD CANCER, V50, P715, DOI 10.1002/pbc.21032
   Fleming AJ, 2012, BRAIN PATHOL, V22, P625, DOI 10.1111/j.1750-3639.2011.00561.x
   Friedrich C, 2015, J NEUROSURG-PEDIATR, V15, P227, DOI 10.3171/2014.9.PEDS14213
   Gajjar A, 1996, PEDIATR NEUROSURG, V25, P182, DOI 10.1159/000121121
   Gibson P, 2010, NATURE, V468, P1095, DOI 10.1038/nature09587
   Hargrave D, 2008, J NEURO-ONCOL, V86, P313, DOI 10.1007/s11060-007-9473-5
   Helton KJ, 2006, AM J NEURORADIOL, V27, P786
   Jansen MHA, 2012, CANCER TREAT REV, V38, P27, DOI 10.1016/j.ctrv.2011.06.007
   Judkins AR, 2004, AM J SURG PATHOL, V28, P644, DOI 10.1097/00000478-200405000-00013
   Kebudi R, 2013, PEDIATR DRUGS, V15, P351, DOI 10.1007/s40272-013-0033-5
   Khatua S, 2011, CHILD NERV SYST, V27, P1391, DOI 10.1007/s00381-011-1468-z
   Klimo P, 2006, J NEUROSURG, V105, P271, DOI 10.3171/ped.2006.105.4.271
   Klimo P, 2013, J NEUROSURG-PEDIATR, V11, P274, DOI 10.3171/2012.11.PEDS12317
   Korshunov A, 2014, ACTA NEUROPATHOL, V128, P279, DOI 10.1007/s00401-013-1228-0
   KYOSHIMA K, 1993, J NEUROSURG, V78, P987, DOI 10.3171/jns.1993.78.6.0987
   Lewis PW, 2013, SCIENCE, V340, P857, DOI 10.1126/science.1232245
   Louis DN, 2007, ACTA NEUROPATHOL, V114, P547, DOI 10.1007/s00401-007-0278-6
   Lundar T, 2014, J NEUROSURG-PEDIATR, V14, P598, DOI 10.3171/2014.9.PEDS1462
   McCrea J, 2015, PRINCIPLES PRACTICE, P553
   Mehta VS, 2009, NEUROL INDIA, V57, P274, DOI 10.4103/0028-3886.53272
   Northcott PA, 2012, NAT REV NEUROL, V8, P340, DOI 10.1038/nrneurol.2012.78
   Ogiwara H, 2013, CHILD NERV SYST, V29, P833, DOI 10.1007/s00381-013-2042-7
   Picard D, 2012, LANCET ONCOL, V13, P838, DOI 10.1016/S1470-2045(12)70257-7
   Rutka JT, 2012, J NEUROSURG-PEDIATR, V10, P79, DOI 10.3171/2012.2.PEDS1237
   SMITH TW, 1993, CLIN NEUROPATHOL, V12, P335
   Sufit A, 2012, J NEUROSURG-PEDIATR, V10, P81, DOI 10.3171/2012.3.PEDS11316
   Teo C, 2008, CHILD NERV SYST, V24, P1307, DOI 10.1007/s00381-008-0647-z
   Udaka YT, 2013, CASE REP ONCOL MED, V2013, DOI 10.1155/2013/815923
   Versteege I, 1998, NATURE, V394, P203, DOI 10.1038/28212
   Yagmurlu K, 2014, OPER NEUROSURG, V10, P602, DOI 10.1227/NEU.0000000000000466
   Yang T, 2013, WORLD NEUROSURG, V79, P537, DOI 10.1016/j.wneu.2012.09.015
   Zagzag D, 2000, PEDIATRICS, V106, P1045, DOI 10.1542/peds.106.5.1045
NR 44
TC 8
Z9 9
U1 0
U2 10
PU AMER ASSOC NEUROLOGICAL SURGEONS
PI ROLLING MEADOWS
PA 5550 MEADOWBROOK DRIVE, ROLLING MEADOWS, IL 60008 USA
SN 1933-0707
EI 1933-0715
J9 J NEUROSURG-PEDIATR
JI J. Neurosurg.-Pediatr.
PD JAN
PY 2016
VL 17
IS 1
BP 57
EP 65
DI 10.3171/2015.6.PEDS15166
PG 9
WC Clinical Neurology; Pediatrics; Surgery
WE Science Citation Index Expanded (SCI-EXPANDED)
SC Neurosciences & Neurology; Pediatrics; Surgery
GA CZ3TW
UT WOS:000367027900010
PM 26474099
OA Bronze
DA 2023-08-27
ER

PT J
AU Hirano, Y
   Shinya, Y
   Aono, T
   Hasegawa, H
   Kawashima, M
   Shin, M
   Takami, H
   Takayanagi, S
   Umekawa, M
   Ikemura, M
   Ushiku, T
   Taoka, K
   Tanaka, S
   Saito, N
AF Hirano, Yudai
   Shinya, Yuki
   Aono, Toshiya
   Hasegawa, Hirotaka
   Kawashima, Mariko
   Shin, Masahiro
   Takami, Hirokazu
   Takayanagi, Shunsaku
   Umekawa, Motoyuki
   Ikemura, Masako
   Ushiku, Tetsuo
   Taoka, Kazuki
   Tanaka, Shota
   Saito, Nobuhito
TI The Role of Stereotactic Frame-Based Biopsy for Brainstem Tumors in the
   Era of Molecular-Based Diagnosis and Treatment Decisions
SO CURRENT ONCOLOGY
LA English
DT Article
DE stereotactic biopsy; brainstem tumor; frame-based
ID CENTRAL-NERVOUS-SYSTEM; UNITED-STATES; LESIONS; TRANSCEREBELLAR; SAFETY;
   METAANALYSIS
AB Stereotactic frame-based brain tumor biopsy (SFB) is a potent diagnostic tool considering its minimal invasiveness, though its diagnostic power and safety for brainstem lesions remain to be discussed. Here, we aimed to examine the usefulness of SFB for brainstem tumors. Twenty-two patients with brainstem tumors underwent 23 SFBs at our institution during 2002-2021. We retrospectively analyzed patient characteristics, tumor pathology, surgical procedures, and outcomes, including surgery-related complications and the diagnostic value. Seven (32%) tumors were located from the midbrain to the pons, eleven (50%) in the pons only, and four (18%) from the pons to the medulla oblongata. The target lesions were in the middle cerebellar peduncles in sixteen procedures (70%), the cerebellum in four (17%), the inferior cerebellar peduncles in two (9%), and the superior cerebellar peduncles in one (4%). A definitive diagnosis was made in 21 patients (95%) at the first SFB. The diagnoses were glioma in seventeen (77%) cases, primary central nervous system lymphoma in four (18%), and a metastatic brain tumor in one (5%). The postoperative complications (cranial nerve palsy in three [13%] cases, ataxia in one [4%]) were all transient. SFB for brainstem tumors yields a high diagnostic rate with a low risk of morbidity.
C1 [Hirano, Yudai; Shinya, Yuki; Aono, Toshiya; Hasegawa, Hirotaka; Kawashima, Mariko; Takami, Hirokazu; Takayanagi, Shunsaku; Umekawa, Motoyuki; Tanaka, Shota; Saito, Nobuhito] Univ Tokyo Hosp, Dept Neurosurg, Bunkyo Ku, 7-3-1 Hongo, Tokyo 1138655, Japan.
   [Hirano, Yudai; Shin, Masahiro] Teikyo Univ, Sch Med, Dept Neurosurg, Itabashi Ku, 2-11-1 Kaga, Tokyo 1730003, Japan.
   [Ikemura, Masako; Ushiku, Tetsuo] Univ Tokyo Hosp, Dept Pathol, Bunkyo Ku, 7-3-1 Hongo, Tokyo 1138655, Japan.
   [Taoka, Kazuki] Univ Tokyo Hosp, Dept Hematol & Oncol, Bunkyo Ku, 7-3-1 Hongo, Tokyo 1138655, Japan.
C3 Teikyo University; University of Tokyo; University of Tokyo
RP Shinya, Y; Tanaka, S (通讯作者)，Univ Tokyo Hosp, Dept Neurosurg, Bunkyo Ku, 7-3-1 Hongo, Tokyo 1138655, Japan.
EM;
  ;
  ;
  ;
  ;
  ;
  
RI Tanaka, Shota/HLP-6965-2023; Hirano, Yudai/GZL-7599-2022
OI Hasegawa, Hirotaka/0000-0002-3585-2188; Hirano,
   Yudai/0000-0003-4043-0593; Umekawa, Motoyuki/0000-0002-7722-9861;
   Shinya, Yuki/0000-0003-0097-5804
FU JSPS KAKENHI [20K17919]
FX This work was supported by JSPS KAKENHI (grant number 20K17919 to
   Shinya).
CR Amundson EW, 2005, J NEUROSURG, V102, P565, DOI 10.3171/jns.2005.102.3.0565
   Carai A, 2017, WORLD NEUROSURG, V101, P584, DOI 10.1016/j.wneu.2017.02.088
   Dellaretti M, 2012, STEREOT FUNCT NEUROS, V90, P79, DOI 10.1159/000335502
   Furtak J, 2021, BRAIN SCI, V11, DOI 10.3390/brainsci11111432
   Goncalves-Ferreira AJ, 2003, SURG NEUROL, V60, P311, DOI 10.1016/S0090-3019(03)00379-3
   Hamisch C, 2017, J NEUROSURG-PEDIATR, V20, P261, DOI 10.3171/2017.2.PEDS1665
   He L, 2021, CANCER CONTROL, V28, DOI 10.1177/10732748211059858
   Jaradat A, 2021, ACTA NEUROCHIR, V163, P1957, DOI 10.1007/s00701-021-04733-2
   Jung IH, 2021, CANCER MED-US, V10, P7514, DOI 10.1002/cam4.4272
   Kickingereder P, 2013, NEUROSURGERY, V72, P873, DOI 10.1227/NEU.0b013e31828bf445
   Labuschagne J, 2021, CHILD NERV SYST, V37, P1515, DOI 10.1007/s00381-021-05101-2
   Larner AJ, 1999, EUR J NEUROL, V6, P721, DOI 10.1046/j.1468-1331.1999.660721.x
   Louis DN, 2021, NEURO-ONCOLOGY, V23, P1231, DOI 10.1093/neuonc/noab106
   Manoj N, 2014, J NEUROSCI RURAL PRA, V5, P32, DOI 10.4103/0976-3147.127869
   MCCUE MP, 1993, NEUROLOGY, V43, P377, DOI 10.1212/WNL.43.2.377
   Ostrom QT, 2019, NEURO-ONCOLOGY, V21, pV1, DOI 10.1093/neuonc/noz150
   Patel P, 2009, J PEDIATR NEUROSCI, V4, P17, DOI 10.4103/1817-1745.49101
   Peciu-Florianu I, 2022, J NEURO-ONCOL, V157, P109, DOI 10.1007/s11060-022-03952-6
   Quick-Weller J, 2016, J NEURO-ONCOL, V129, P243, DOI 10.1007/s11060-016-2166-1
   Rachinger W, 2009, J NEUROL NEUROSUR PS, V80, P1134, DOI 10.1136/jnnp.2009.174250
   Sanai N, 2008, NEUROSURGERY, V63, P460, DOI 10.1227/01.NEU.0000324731.68843.74
   Tanaka S, 2012, NEUROSURGERY, V71, P321, DOI 10.1227/NEU.0b013e31826a88a9
   Zada G, 2012, WORLD NEUROSURG, V77, P518, DOI 10.1016/j.wneu.2011.05.051
NR 23
TC 1
Z9 1
U1 0
U2 0
PU MDPI
PI BASEL
PA ST ALBAN-ANLAGE 66, CH-4052 BASEL, SWITZERLAND
SN 1198-0052
EI 1718-7729
J9 CURR ONCOL
JI Curr. Oncol.
PD JUL
PY 2022
VL 29
IS 7
BP 4558
EP 4565
DI 10.3390/curroncol29070360
PG 8
WC Oncology
WE Science Citation Index Expanded (SCI-EXPANDED)
SC Oncology
GA 3G6NL
UT WOS:000831468700001
PM 35877220
OA Green Published, gold
DA 2023-08-27
ER

PT J
AU Sufit, A
   Donson, AM
   Birks, DK
   Knipstein, JA
   Fenton, LZ
   Jedlicka, P
   Hankinson, TC
   Handler, MH
   Foreman, NK
AF Sufit, Alexandra
   Donson, Andrew M.
   Birks, Diane K.
   Knipstein, Jeffrey A.
   Fenton, Laura Z.
   Jedlicka, Paul
   Hankinson, Todd C.
   Handler, Michael H.
   Foreman, Nicholas K.
TI Diffuse intrinsic pontine tumors: a study of primitive neuroectodermal
   tumors versus the more common diffuse intrinsic pontine gliomas
   Laboratory investigation
SO JOURNAL OF NEUROSURGERY-PEDIATRICS
LA English
DT Article
DE pediatric brainstem tumors; diffuse intrinsic pontine glioma; primitive
   neuroectodermal tumor; brainstem biopsy; GATA3; oncology
ID BRAIN-STEM GLIOMA; DIAGNOSIS; CHILDREN; PROTEIN; ALEX1; GENE
AB Object. The diagnosis of diffuse pontine tumors has largely been made on the basis of MRI since the early 1990s. In cases of tumors considered "typical," as a rule, no biopsy specimen has been obtained, and the tumors have been considered diffuse intrinsic pontine gliomas (DIPGs). There have been sporadic reports that primitive neuroectodermal tumors (PNETs) of the pons may not be distinguishable from the DIPGs by radiological imaging. This study presents 2 cases of diffuse pontine PNETs with molecular evidence that these are indeed PNETs, distinct from DIPGs, thus supporting biopsy of diffuse pontine tumors as a standard of care.
   Methods. Biopsy specimens were obtained from 7 diffuse pontine tumors and snap frozen. Two of these 7 tumors were identified on the basis of pathological examination as PNETs. All 7 of the diffuse pontine tumors were analyzed for gene expression using the Affymetrix HG-U133 Plus 2.0 GeneChip microarray. Gene expression was compared with that of supratentorial PNETs, medulloblastomas, and low- and high-grade gliomas outside the brainstem.
   Results. Unsupervised hierarchical clustering analysis of gene expression demonstrated that pontine PNETs are most closely related to PNETs of the supratentorial region and not with gliomas. They do not cluster with the 5 DIPGs in the study. Thirty-eight genes, including GATA3, are uniquely differentially expressed in pontine PNETs compared with other types of pediatric brain tumors, including DIPGs and other PNETs at a false discovery rate statistical significance of less than 0.05.
   Conclusions. The cluster and individual gene expression analyses indicate that pontine PNETs are intrinsically different from DIPGs. The 2 pontine PNET cases cluster with supratentorial PNETs, rather than with DIPGs, suggesting that these tumors should be treated with a PNET regimen, not with DIPG therapy. Since diagnosis by imaging is not reliable and the biology of the tumors is disparate, a biopsy should be performed to enable accurate diagnosis and direct potentially more effective treatments. (http://thejns.org/doi/abs/10.3171/2012.3.PEDS11316)
C1 [Sufit, Alexandra; Donson, Andrew M.; Knipstein, Jeffrey A.; Foreman, Nicholas K.] Univ Colorado, Childrens Hosp Colorado, Div Neurooncol, Dept Pediat, Aurora, CO 80045 USA.
   [Birks, Diane K.; Hankinson, Todd C.; Handler, Michael H.] Univ Colorado, Childrens Hosp Colorado, Div Pediat Neurosurg, Dept Neurosurg, Aurora, CO 80045 USA.
   [Fenton, Laura Z.] Univ Colorado, Childrens Hosp Colorado, Div Radiol, Aurora, CO 80045 USA.
   [Jedlicka, Paul] Univ Colorado, Childrens Hosp Colorado, Dept Pathol, Aurora, CO 80045 USA.
C3 Children's Hospital Colorado; University of Colorado System; University
   of Colorado Anschutz Medical Campus; Children's Hospital Colorado;
   University of Colorado System; University of Colorado Anschutz Medical
   Campus; Children's Hospital Colorado; University of Colorado System;
   University of Colorado Anschutz Medical Campus; Children's Hospital
   Colorado; University of Colorado System; University of Colorado Anschutz
   Medical Campus
RP Sufit, A (通讯作者)，Univ Colorado, Anschutz Med Campus,Pediat Mail Stop 8302,POB 651, Aurora, CO 80045 USA.
EM
OI Foreman, Nicholas/0000-0003-4848-4801; Knipstein,
   Jeffrey/0000-0002-0632-2751
FU Morgan Adams Foundation
FX This project was funded by the Morgan Adams Foundation. The authors
   report no conflict of interest concerning the materials or methods used
   in this study or the findings specified in this paper.
CR ALBRIGHT AL, 1993, NEUROSURGERY, V33, P1026
   Angelini P, 2011, J NEURO-ONCOL, V101, P75, DOI 10.1007/s11060-010-0224-7
   [Anonymous], 2004, STAT APPL GENET MOL, DOI DOI 10.2202/1544-6115.1027
   Donaldson SS, 2006, J CLIN ONCOL, V24, P1266, DOI 10.1200/JCO.2005.04.6599
   Ellison DW, 2011, ACTA NEUROPATHOL, V121, P381, DOI 10.1007/s00401-011-0800-8
   Epstein F, 1996, PEDIATR NEUROSURG, V24, P24, DOI 10.1159/000121011
   Handler MH, 2010, J NEUROS-PEDIATR, V5, P140, DOI 10.3171/2009.5.PEDS09265
   Hankinson TC, 2011, J NEUROSURG-PEDIATR, V8, P97, DOI 10.3171/2011.4.PEDS1180
   Hargrove D, 2006, LANCET ONCOL, V7, P241, DOI 10.1016/S1470-2045(06)70615-5
   Iseki H, 2010, CANCER SCI, V101, P1361, DOI 10.1111/j.1349-7006.2010.01541.x
   Kleinschmidt-DeMasters BK, 2011, ACTA NEUROPATHOL, V122, P783, DOI 10.1007/s00401-011-0894-z
   Korones DN, 2008, PEDIATR BLOOD CANCER, V50, P227, DOI 10.1002/pbc.21154
   Kurochkin IV, 2001, BIOCHEM BIOPH RES CO, V280, P340, DOI 10.1006/bbrc.2000.4125
   Lindsey JC, 2011, J CLIN ONCOL, V29, pE344, DOI 10.1200/JCO.2010.33.8590
   MacDonald TJ, 2003, ONCOLOGIST, V8, P174, DOI 10.1634/theoncologist.8-2-174
   MCLENDON RE, 2007, WHO CLASSIFICATION T, P141
   Negretti L, 2011, J NEURO-ONCOL, V104, P773, DOI 10.1007/s11060-011-0542-4
   Paugh BS, 2010, J CLIN ONCOL, V28, P3061, DOI 10.1200/JCO.2009.26.7252
   Pomeroy SL, 2002, NATURE, V415, P436, DOI 10.1038/415436a
   Rorke LB, 2003, PRINCIPLES PRACTICE, P383
   Rutkowski S, 2005, NEW ENGL J MED, V352, P978, DOI 10.1056/NEJMoa042176
   Smyth GK, 2005, STAT BIOL HEALTH, P397, DOI 10.1007/0-387-29362-0_23
   Wu ZJ, 2004, J AM STAT ASSOC, V99, P909, DOI 10.1198/016214504000000683
   Zagzag D, 2000, PEDIATRICS, V106, P1045, DOI 10.1542/peds.106.5.1045
   Zarghooni M, 2010, J CLIN ONCOL, V28, P1337, DOI 10.1200/JCO.2009.25.5463
NR 25
TC 22
Z9 23
U1 0
U2 8
PU AMER ASSOC NEUROLOGICAL SURGEONS
PI ROLLING MEADOWS
PA 5550 MEADOWBROOK DRIVE, ROLLING MEADOWS, IL 60008 USA
SN 1933-0707
EI 1933-0715
J9 J NEUROSURG-PEDIATR
JI J. Neurosurg.-Pediatr.
PD AUG
PY 2012
VL 10
IS 2
BP 81
EP 88
DI 10.3171/2012.3.PEDS11316
PG 8
WC Clinical Neurology; Pediatrics; Surgery
WE Science Citation Index Expanded (SCI-EXPANDED)
SC Neurosciences & Neurology; Pediatrics; Surgery
GA 978SV
UT WOS:000306767000003
PM 22747092
OA Green Accepted
DA 2023-08-27
ER

PT J
AU Jallo, GI
   Volkov, A
   Wong, C
   Carson, BS
   Penno, MB
AF Jallo, George I.
   Volkov, Andrey
   Wong, Cyrus
   Carson, Benjamin S., Sr.
   Penno, Margaret B.
TI A novel brainstem tumor model: functional and histopathological
   characterization
SO CHILDS NERVOUS SYSTEM
LA English
DT Article
DE brainstem; animal model; pontine glioma; tumor
ID GLIOMAS; DELIVERY; THERAPY; 9L; CHEMOTHERAPY; SYSTEM; CANCER
AB Introduction Diffuse pontine gliomas remain a challenging and frustrating disease to treat. The survival rates for these high-grade brainstem tumors (BSTs) is dismal and optimal therapy has yet to be determined. The development of a satisfactory brainstem tumor model is necessary for testing new therapeutic paradigms that may prolong survival.
   Materials and methods We report the surgical technique, functional testing, and histopathological features of a novel brainstem tumor model in rats. Female Fischer 344 rats (n=45) were randomized to receive an injection of either 3 mu l of 9L gliosarcoma cells (100,000 cells, n=), 3 mu l of F98 glioma cells (100,000 cells, n=10), or 3 mu l of medium (Dulbecco's modified eagle medium) into the pontine tegmentum. Using a cannulated guide screw system, implanted in the skull of the animal, we injected each group at coordinates 1.4 mm right of the sagittal and 1.0 mm anterior of the lambdoid sutures, at a depth of 7.0 mm from the dura. The head was positioned 5 degrees from horizontal before injection. The rats were post-operatively evaluated for neurological deficits using an automated test. Kaplan-Meier curves were generated for survival and disease progression, and brains were processed postmortem for histopathology. Results and discussion 9L and F98 tumor cells grew in 100% of animals injected and resulted in a statistically significant mean onset of hemiparesis of 16.5 +/- 0.56 days (P=0.001, log-rank test), compared to animals in the control group which lacked neurological deficits by day 60. The animals with tumor cells implanted demonstrated significant deterioration of function on the automated rod testing. Animals in the control group showed no functional or pathological signs of tumor. Progression to hemiparesis was consistent in all tumor-injected animals, with predictable onset of symptoms occurring approximately 17 days post-surgery. The histopathological characteristics of the 9L and F98 BSTs were comparable to those of aggressive human BSTs.
   Conclusion The establishment of this animal tumor model will facilitate the testing of new therapeutic paradigms for the treatment of BSTs.
C1 Johns Hopkins Univ, Sch Med, Dept Neurosurg, Baltimore, MD 21205 USA.
   Johns Hopkins Univ, Sch Med, Dept Pediat, Baltimore, MD 21205 USA.
   Johns Hopkins Univ, Sch Med, Dept Oncol, Baltimore, MD 21205 USA.
   Johns Hopkins Univ, Sch Med, Dept Med, Baltimore, MD 21205 USA.
   Johns Hopkins Univ Hosp, Div Pediat Neurosurg, Baltimore, MD 21287 USA.
C3 Johns Hopkins University; Johns Hopkins University; Johns Hopkins
   University; Johns Hopkins University; Johns Hopkins University; Johns
   Hopkins Medicine
RP Jallo, GI (通讯作者)，Johns Hopkins Univ, Sch Med, Dept Neurosurg, Baltimore, MD 21205 USA.
EM
CR Albright AL, 1996, PEDIATR NEUROSURG, V24, P252
   ALBRIGHT AL, 1986, J NEUROSURG, V65, P751, DOI 10.3171/jns.1986.65.6.0751
   Allen JC, 1996, PEDIATR NEUROSURG, V24, P98, DOI 10.1159/000121024
   BARKER M, 1973, CANCER RES, V33, P976
   Barth RF, 1998, J NEURO-ONCOL, V36, P91, DOI 10.1023/A:1005805203044
   BENDA P, 1971, J NEUROSURG, V34, P310, DOI 10.3171/jns.1971.34.3.0310
   CARSON BS, 2002, ADV CLIN NEUROSCI, V12, P89
   EPSTEIN F, 1988, J NEURO-ONCOL, V6, P309
   Griffitt W, 1999, J NEURO-ONCOL, V41, P117, DOI 10.1023/A:1006115221761
   Guarnieri M, 2005, J NEUROSCI METH, V144, P147, DOI 10.1016/j.jneumeth.2004.10.019
   Guerin C, 2004, INVEST NEW DRUG, V22, P27, DOI 10.1023/B:DRUG.0000006172.65135.3e
   Jallo GI, 2004, CHILD NERV SYST, V20, P143, DOI 10.1007/s00381-003-0870-6
   Jallo GI, 2005, CHILD NERV SYST, V21, P399, DOI 10.1007/s00381-004-1100-6
   Jounaidi Y, 2004, CANCER RES, V64, P292, DOI 10.1158/0008-5472.CAN-03-1798
   Lee FT, 1997, RADIOLOGY, V203, P465, DOI 10.1148/radiology.203.2.9114106
   LEE J, 2005, NEUROSURG FOCUS, V18, DOI ARTN 11
   LITTMAN P, 1980, CANCER, V45, P2787, DOI 10.1002/1097-0142(19800601)45:11<2787::AID-CNCR2820451113>3.0.CO;2-V
   Madajewicz S, 2000, CANCER-AM CANCER SOC, V88, P2350, DOI 10.1002/(SICI)1097-0142(20000515)88:10<2350::AID-CNCR20>3.0.CO;2-R
   Occhiogrosso G, 2003, NEUROSURGERY, V52, P388, DOI 10.1227/01.NEU.0000043696.83722.8D
   RAND RW, 1982, J UROLOGY, V128, P618, DOI 10.1016/S0022-5347(17)53078-5
   Rozas G, 1997, BRAIN RES PROTOC, V2, P75, DOI 10.1016/S1385-299X(97)00034-2
   SALCMAN M, 1982, NEUROSURGERY, V11, P372, DOI 10.1227/00006123-198209000-00007
   SCHMIDEK HH, 1971, J NEUROSURG, V34, P335, DOI 10.3171/jns.1971.34.3.0335
   Storm PB, 2002, J NEURO-ONCOL, V56, P209, DOI 10.1023/A:1015003232713
   WECHSLER H, 1979, J NATL CANCER I, V62, P811
   WEIZSAECKER M, 1981, J NEUROL, V224, P183, DOI 10.1007/BF00313280
   Wu Q, 2002, VET PATHOL, V39, P293, DOI 10.1354/vp.39-3-293
NR 27
TC 30
Z9 32
U1 0
U2 5
PU SPRINGER
PI NEW YORK
PA 233 SPRING STREET, NEW YORK, NY 10013 USA
SN 0256-7040
J9 CHILD NERV SYST
JI Childs Nerv. Syst.
PD DEC
PY 2006
VL 22
IS 12
BP 1519
EP 1525
DI 10.1007/s00381-006-0174-8
PG 7
WC Clinical Neurology; Pediatrics; Surgery
WE Science Citation Index Expanded (SCI-EXPANDED)
SC Neurosciences & Neurology; Pediatrics; Surgery
GA 118UT
UT WOS:000242968900003
PM 17021732
DA 2023-08-27
ER

PT J
AU Klimo, P
   Panandiker, ASP
   Thompson, CJ
   Boop, FA
   Qaddoumi, I
   Gajjar, A
   Armstrong, GT
   Ellison, DW
   Kun, LE
   Ogg, RJ
   Sanford, RA
AF Klimo, Paul, Jr.
   Panandiker, Atmaram S. Pai
   Thompson, Clinton J.
   Boop, Frederick A.
   Qaddoumi, Ibrahim
   Gajjar, Amar
   Armstrong, Gregory T.
   Ellison, David W.
   Kun, Larry E.
   Ogg, Robert J.
   Sanford, Robert A.
TI Management and outcome of focal low-grade brainstem tumors in pediatric
   patients: the St. Jude experience
SO JOURNAL OF NEUROSURGERY-PEDIATRICS
LA English
DT Article
DE focal brainstem glioma; low grade; radiation treatment; outcomes;
   oncology; event-free survival
ID PROGNOSTIC-FACTORS; PILOCYTIC ASTROCYTOMA; SURGICAL INDICATIONS;
   CLINICAL ARTICLE; GLIOMAS; CHILDHOOD; CLASSIFICATION; CHILDREN;
   DIAGNOSIS; RESECTION
AB Object. Whereas diffuse intrinsic pontine gliomas generally have a short symptom duration and more cranial nerve involvement, focal brainstem gliomas are commonly low grade, with fewer cranial neuropathies. Although these phenotypic distinctions are not absolute predictors of outcome, they do demonstrate correlation in most cases. Because there is a limited literature on focal brainstem gliomas in pediatric patients, the objective of this paper was to report the management and outcome of these tumors.
   Methods. The authors reviewed the records Of all children diagnosed with radiographically confirmed low-grade focal brainstem gliomas from 1986 to 2010. Each patient underwent biopsy or resection for tissue diagnosis. Event-free survival (EFS) and overall survival were evaluated. Univariate analysis was conducted to identify demographic and treatment variables that may affect EFS.
   Results. Fifty-two patients (20 girls, 32 boys) with follow-up data were identified. Median follow-up was 10.0 years, and the median age at diagnosis was 6.5 years (range 1-17 years). The tumor locations were midbrain (n = 22, 42%), pons (n = 15, 29%), and medulla (n = 15, 29%). Surgical extirpation was the primary treatment in 25 patients (48%). The 5- and 10-year EFS and overall survival were 59%/98% and 52%/90%, respectively. An event or treatment failure occurred in 24 patients (46%), including 5 deaths. Median time to treatment failure was 3.4 years. Disease progression in the other 19 patients transpired within 25.1 months of diagnosis. Thirteen of these patients received radiation, including 11 within 2 months of primary treatment failure. Although children with intrinsic tumors had slightly better EFS at 5 years compared with those with exophytic tumors (p = 0.054), this difference was not significant at 10 years (p = 0.147). No other variables were predictive of EFS.
   Conclusions. Surgery suffices in many children with low-grade focal brainstem gliomas. Radiation treatment is often reserved for disease progression but offers comparable disease control following biopsy. In the authors' experience, combining an assessment of clinical course, imaging, and tumor biopsy yields a reasonable model for managing children with focal brainstem tumors. (http://thejns.org/doi/abs/10.3171/2012.11.PEDS12317)
C1 [Klimo, Paul, Jr.; Boop, Frederick A.] Univ Tennessee, Ctr Hlth Sci, Semmes Murphey Neurol & Spine Inst, Memphis, TN 38163 USA.
   [Klimo, Paul, Jr.; Boop, Frederick A.] Univ Tennessee, Ctr Hlth Sci, Dept Neurosurg, Memphis, TN 38163 USA.
   [Klimo, Paul, Jr.; Boop, Frederick A.; Sanford, Robert A.] St Jude Childrens Res Hosp, Dept Surg, Memphis, TN 38105 USA.
   [Panandiker, Atmaram S. Pai; Kun, Larry E.; Ogg, Robert J.] St Jude Childrens Res Hosp, Dept Radiol Sci, Memphis, TN 38105 USA.
   [Qaddoumi, Ibrahim; Gajjar, Amar] St Jude Childrens Res Hosp, Dept Oncol, Memphis, TN 38105 USA.
   [Armstrong, Gregory T.] St Jude Childrens Res Hosp, Dept Epidemiol & Canc Control, Memphis, TN 38105 USA.
   [Ellison, David W.] St Jude Childrens Res Hosp, Dept Pathol, Memphis, TN 38105 USA.
   [Thompson, Clinton J.] George Washington Univ, Sch Publ Hlth & Hlth Serv, Washington, DC USA.
C3 University of Tennessee System; University of Tennessee Health Science
   Center; University of Tennessee System; University of Tennessee Health
   Science Center; St Jude Children's Research Hospital; St Jude Children's
   Research Hospital; St Jude Children's Research Hospital; St Jude
   Children's Research Hospital; St Jude Children's Research Hospital;
   George Washington University
RP Panandiker, ASP (通讯作者)，St Jude Childrens Res Hosp, Dept Radiol Sci, MS 220,262 Danny Thomas Pl, Memphis, TN 38105 USA.
EM
RI Qaddoumi, Ibrahim/N-8101-2018; Armstrong, Gregory/N-8096-2018; Gajjar,
   Amar/N-8059-2018; Pai Panandiker, Atmaram Shambhu/GVU-0525-2022
FU American Lebanese Syrian Associated Charities (ALSAC); Cancer Center
   Support Grants from the NIH [CA23099, CA21765]
FX This work was supported in part by the American Lebanese Syrian
   Associated Charities (ALSAC) and by Cancer Center Support Grants CA23099
   and CA21765 from the NIH. Dr. Boop is Secretary of the AANS and
   Treasurer of the International Society for Pediatric Neurosurgery.
CR BARKOVICH AJ, 1991, PEDIATR NEUROSURG, V16, P73, DOI 10.1159/000120511
   BOYDSTON WR, 1992, PEDIATR NEUROSURG, V17, P234, DOI 10.1159/000120603
   Bricolo A, 1991, Acta Neurochir Suppl (Wien), V53, P148
   Broadway SJ, 2011, J NEUROSURG-PEDIATR, V7, P589, DOI 10.3171/2011.4.PEDS119
   Choux M, 2000, PEDIAT NEUROSURGERY, P471
   Dellaretti M, 2011, J NEUROSURG-PEDIATR, V8, P539, DOI 10.3171/2011.9.PEDS1167
   Donaldson SS, 2006, J CLIN ONCOL, V24, P1266, DOI 10.1200/JCO.2005.04.6599
   EPSTEIN F, 1988, J NEURO-ONCOL, V6, P309
   EPSTEIN F, 1986, J NEUROSURG, V64, P11, DOI 10.3171/jns.1986.64.1.0011
   Fangusaro J, 2009, J CHILD NEUROL, V24, P1409, DOI 10.1177/0883073809338960
   Farmer JP, 2001, PEDIATR NEUROSURG, V34, P206, DOI 10.1159/000056021
   Fisher PG, 2000, CANCER, V89, P1569, DOI 10.1002/1097-0142(20001001)89:7<1569::AID-CNCR22>3.0.CO;2-0
   Fried I, 2012, PEDIATR BLOOD CANCER, V58, P556, DOI 10.1002/pbc.23200
   Goncalves-Ferreira AJ, 2003, SURG NEUROL, V60, P311, DOI 10.1016/S0090-3019(03)00379-3
   Jallo GI, 2004, CHILD NERV SYST, V20, P143, DOI 10.1007/s00381-003-0870-6
   Kestle J, 2004, J NEUROSURG, V101, P1, DOI 10.3171/ped.2004.101.2.0001
   Kwon JW, 2006, PEDIATR RADIOL, V36, P959, DOI 10.1007/s00247-006-0256-5
   Lesniak MS, 2003, PEDIATR NEUROSURG, V39, P314, DOI 10.1159/000075260
   Liao CH, 2012, CHILD NERV SYST, V28, P175, DOI 10.1007/s00381-011-1620-9
   Mauffrey C, 2006, J CLIN NEUROSCI, V13, P431, DOI 10.1016/j.jocn.2005.05.015
   Mehta VS, 2009, NEUROL INDIA, V57, P274, DOI 10.4103/0028-3886.53272
   Perez-Gomez JL, 2010, CHILD NERV SYST, V26, P29, DOI 10.1007/s00381-009-1000-x
   Recinos PF, 2007, PEDIATR NEUROSURG, V43, P192, DOI 10.1159/000098831
   Sandri A, 2006, CHILD NERV SYST, V22, P1127, DOI 10.1007/s00381-006-0083-x
   Schild SE, 1998, J NEURO-ONCOL, V40, P171, DOI 10.1023/A:1006193306286
   STROINK AR, 1986, J NEUROSURG, V65, P745, DOI 10.3171/jns.1986.65.6.0745
   Teo C, 2008, CHILD NERV SYST, V24, P1307, DOI 10.1007/s00381-008-0647-z
   Ueoka DI, 2009, J NEUROL SCI, V281, P20, DOI 10.1016/j.jns.2009.03.009
   Weiner HL, 1997, PEDIATR NEUROSURG, V27, P12, DOI 10.1159/000121219
   Yen CP, 2007, J NEUROSURG, V106, P8, DOI 10.3171/jns.2007.106.1.8
NR 30
TC 26
Z9 26
U1 0
U2 6
PU AMER ASSOC NEUROLOGICAL SURGEONS
PI ROLLING MEADOWS
PA 5550 MEADOWBROOK DRIVE, ROLLING MEADOWS, IL 60008 USA
SN 1933-0707
EI 1933-0715
J9 J NEUROSURG-PEDIATR
JI J. Neurosurg.-Pediatr.
PD MAR
PY 2013
VL 11
IS 3
BP 274
EP 281
DI 10.3171/2012.11.PEDS12317
PG 8
WC Clinical Neurology; Pediatrics; Surgery
WE Science Citation Index Expanded (SCI-EXPANDED)
SC Neurosciences & Neurology; Pediatrics; Surgery
GA 094DV
UT WOS:000315244100007
PM 23289916
OA Green Accepted
DA 2023-08-27
ER

PT J
AU Shah, NC
   Ray, A
   Bartels, U
   Rutka, J
   Bouffet, E
   Drake, J
   Hawkins, CE
   Huang, A
AF Shah, Niketa C.
   Ray, Amit
   Bartels, Ute
   Rutka, James
   Bouffet, Eric
   Drake, James
   Hawkins, Cynthia E.
   Huang, Annie
TI Diffuse intrinsic brainstem tumors in neonates
SO JOURNAL OF NEUROSURGERY-PEDIATRICS
LA English
DT Article
DE congenital disease; diffuse intrinsic brainstem glioma; primitive
   neuroectodermal tumor
ID PRIMITIVE NEUROECTODERMAL TUMORS; CENTRAL-NERVOUS-SYSTEM; CHILDRENS
   CANCER GROUP; GLIOMAS; LESION; CHEMOTHERAPY; BIOPSIES; INFANTS; PONS
AB The authors report on 2 newborn infants with the unusual presentation of intrinsic brainstem tumors. Both nondysmorphic, full-term neonates had cranial nerve palsies and hypotonia. Diagnoses of diffuse intrinsic brainstem gliomas were made on the basis of magnetic resonance imaging, which showed large expansive, nonenhancing intrinsic pontine masses. Intrinsic pontine tumors, characteristically seen in school-age children, are most often high-grade gliomas that are almost invariably fatal. However, the microanatomy and natural history of pontine tumors in neonates are unknown. With parental consent, both newborns were treated expectantly with supportive care but died of progressive disease by 2 weeks of age. In one child, postmortem examination revealed a primary brainstem primitive neuroectodermal tumor. The authors conclude that. as in older children, neonatal intrinsic brainstem tumors may be of a highly malignant nature. The rapid tumor progression in both cases indicates that where a diagnostic procedure may pose significant risks, Supportive observation can aid in distinguishing malignant from benign tumor growth.
C1 [Shah, Niketa C.; Bartels, Ute; Bouffet, Eric; Huang, Annie] Hosp Sick Children, Div Hematol & Oncol, Dept Paediat, Toronto, ON M5G 1X8, Canada.
   [Ray, Amit; Rutka, James; Drake, James] Hosp Sick Children, Div Neurosurg, Dept Paediat, Lab Med, Toronto, ON M5G 1X8, Canada.
   [Hawkins, Cynthia E.] Hosp Sick Children, Div Pathol, Dept Paediat, Lab Med, Toronto, ON M5G 1X8, Canada.
C3 University of Toronto; Hospital for Sick Children (SickKids); University
   of Toronto; Hospital for Sick Children (SickKids); University of
   Toronto; Hospital for Sick Children (SickKids)
RP Huang, A (通讯作者)，Hosp Sick Children, Div Pediat Hematol Oncol, 555 Univ Ave, Toronto, ON M5G 1X8, Canada.
EM
RI Bouffet, Eric/AAB-8999-2020
OI Ray, Amitava/0000-0001-5576-2086
CR Albright AL, 1996, PEDIATR NEUROSURG, V24, P252
   ALBRIGHT AL, 1993, NEUROSURGERY, V33, P1026
   Behnke J, 1996, CHILD NERV SYST, V12, P125, DOI 10.1007/BF00266812
   de Leon FCP, 2003, CHILD NERV SYST, V19, P305, DOI 10.1007/s00381-003-0737-x
   Fangusaro JR, 2008, PEDIATR BLOOD CANCER, V50, P715, DOI 10.1002/pbc.21032
   FRAUMENI JF, 1969, AM J DIS CHILD, V117, P186, DOI 10.1001/archpedi.1969.02100030188011
   Freeman CR, 1998, INT J RADIAT ONCOL, V40, P265, DOI 10.1016/S0360-3016(97)00572-5
   Geyer JR, 2005, J CLIN ONCOL, V23, P7621, DOI 10.1200/JCO.2005.09.095
   Isaacs H, 2002, PEDIATR NEUROL, V27, P249, DOI 10.1016/S0887-8994(02)00472-1
   JANISCH W, 1984, J NEURO-ONCOL, V2, P113
   Kaplan AM, 1996, PEDIATR NEUROSURG, V24, P185, DOI 10.1159/000121036
   Larouche V, 2007, PEDIATR BLOOD CANCER, V49, P1074, DOI 10.1002/pbc.21351
   Molloy PT, 1996, J NEUROSURG, V84, P430, DOI 10.3171/jns.1996.84.3.0430
   Rao SJ, 1995, PEDIATR NEUROSURG, V23, P279, DOI 10.1159/000120972
   Schomerus L, 2007, PEDIATR RADIOL, V37, P399, DOI 10.1007/s00247-007-0424-2
   Thompson WD, 2005, J NEUROSURG, V102, P65, DOI 10.3171/ped.2005.102.1.0065
   WAKAI S, 1984, SURG NEUROL, V21, P597, DOI 10.1016/0090-3019(84)90277-5
   Zagzag D, 2000, PEDIATRICS, V106, P1045, DOI 10.1542/peds.106.5.1045
   Zimmerman RA, 1996, PEDIATR NEUROSURG, V25, P83, DOI 10.1159/000121102
NR 19
TC 8
Z9 9
U1 0
U2 1
PU AMER ASSOC NEUROLOGICAL SURGEONS
PI ROLLING MEADOWS
PA 5550 MEADOWBROOK DRIVE, ROLLING MEADOWS, IL 60008 USA
SN 1933-0707
J9 J NEUROS-PEDIATR
JI J. Neurosurg.-Pediatr.
PD MAY
PY 2008
VL 1
IS 5
BP 382
EP 385
DI 10.3171/PED/2008/1/5/382
PG 4
WC Clinical Neurology; Pediatrics; Surgery
WE Science Citation Index Expanded (SCI-EXPANDED)
SC Neurosciences & Neurology; Pediatrics; Surgery
GA 307TV
UT WOS:000256342100010
PM 18447673
DA 2023-08-27
ER

PT J
AU Friedrich, C
   Warmuth-Metz, M
   von Bueren, AO
   Nowak, J
   Bison, B
   von Hoff, K
   Pietsch, T
   Kortmann, RD
   Rutkowski, S
AF Friedrich, Carsten
   Warmuth-Metz, Monika
   von Bueren, Andre O.
   Nowak, Johannes
   Bison, Brigitte
   von Hoff, Katja
   Pietsch, Torsten
   Kortmann, Rolf D.
   Rutkowski, Stefan
TI Primitive neuroectodermal tumors of the brainstem in children treated
   according to the HIT trials: clinical findings of a rare disease
SO JOURNAL OF NEUROSURGERY-PEDIATRICS
LA English
DT Article
DE primitive neuroectodermal tumor; brain tumors; brainstem; children;
   chemotherapy; radiotherapy; oncology
ID INTRAAXIAL ENDOPHYTIC TUMORS; AND/OR MEDULLA-OBLONGATA; YOUNG-CHILDREN;
   RISK MEDULLOBLASTOMA; CHEMOTHERAPY; GLIOMAS; RADIOTHERAPY; ABERRATIONS;
   CHILDHOOD; DISTINCT
AB OBJECT Primitive neuroectodermal tumors of the central nervous system (CNS-PNET) arising in the brainstem are extremely rare, and knowledge about-them is limited. The few existing case series report fatal outcomes. The purpose of this study was to analyze clinical characteristics of and outcome for brainstem CNS-PNET patients treated according to the consecutive, population-based HIT studies covering a 19-year time period.
   METHODS Between September 1992 and November 2011,6 eligible children with histologically proven brainstem CNS-PNET not otherwise specified and 2 children with brainstem ependymoblastomas (3, partial resection; 3, subtotal resection; 2, biopsy), median age 3.3 years (range 1.2-10.6 years), were treated according to consecutive multimodal HIT protocols for CNS-PNET/medulloblastoma. Postoperative treatment was according to maintenance chemotherapy protocols (3, craniospinal irradiation [CSI] followed by maintenance chemotherapy), sandwich chemotherapy protocols (2, neoadjuvant chemotherapy, CSI, maintenance chemotherapy), or a therapy protocol for children younger than 4 years (3, postoperative chemotherapy followed by CSI).
   RESULTS The median duration of prediagnostic symptoms, predominantly cranial nerve deficits (n = 7), pyramidal tract signs (n = 5), or ataxia (n = 5), was 5 weeks (range 1-13 weeks). The tumors were all located in the pons. Most involved more than half of the pontine axial diameter and were sharply marginated. All patients had postoperative residual disease, including metastasis in 1 case. With 1 exception all tumors progressed early during treatment within 3.9 months (range 2.5-10.4 months), leading to a 1-year event-free survival rate (+/- standard error) of13% +/- 12%. After progression, patients succumbed early to their disease resulting in a 1-year overall survival rate of 25% +/- 15%. The only surviving patient had a partially resected CNS-PNET, received a sandwich chemotherapy protocol, and is without disease progression 14 months after diagnosis.
   CONCLUSIONS CNS-PNET is a rare but important,differential diagnosis-in childhood brainstem tumors. So far, efficient therapies are lacking. The sampling of tumor material for improved biological understanding and identification of new therapeutic targets is important.
C1 [Friedrich, Carsten; von Bueren, Andre O.; von Hoff, Katja; Rutkowski, Stefan] Univ Med Ctr Hamburg Eppendorf, Dept Pediat Hematol & Oncol, D-20246 Hamburg, Germany.
   [Friedrich, Carsten] Univ Hosp Leipzig, Dept Womans & Childrens Hlth, Div Pediat Oncol Hematol & Hemostaseol, Leipzig, Germany.
   [Warmuth-Metz, Monika; Nowak, Johannes; Bison, Brigitte] Univ Wurzburg, Dept Neuroradiol, Wurzburg, Germany.
   [von Bueren, Andre O.] Univ Med Ctr Goettingen, Dept Pediat & Adolescent Med, Div Pediat Hematol & Oncol, Gottingen, Germany.
   [Pietsch, Torsten] Univ Bonn, Dept Neuropathol, Bonn, Germany.
   [Kortmann, Rolf D.] Univ Leipzig, Dept Radiat Oncol, D-04109 Leipzig, Germany.
C3 University of Hamburg; University Medical Center Hamburg-Eppendorf;
   Leipzig University; University of Wurzburg; University of Gottingen;
   University of Bonn; Leipzig University
RP Rutkowski, S (通讯作者)，Univ Med Ctr Hamburg Eppendorf, Dept Pediat Hematol & Oncol, Martinistr 52, D-20246 Hamburg, Germany.
EM
RI von Bueren, Andre/AAF-2065-2020
OI von Bueren, Andre/0000-0003-4197-6264; von Hoff,
   Katja/0000-0002-5669-8546
FU German Children's Cancer Foundation (Deutsche Kinderkrebsstiftung);
   German Cancer Aid (Deutsche Krebshilfe)
FX The authors declare that there is no conflict of interest. The HIT 2000
   trial office and the reference institutions are supported by the German
   Children's Cancer Foundation (Deutsche Kinderkrebsstiftung). The HIT '91
   trial was supported by German Cancer Aid (Deutsche Krebshilfe).
CR Angelini P, 2011, J NEURO-ONCOL, V101, P75, DOI 10.1007/s11060-010-0224-7
   [Anonymous], 2007, WHO HISTOLOGICAL CLA
   BARKOVICH AJ, 1991, PEDIATR NEUROSURG, V16, P73, DOI 10.1159/000120511
   Behnke J, 1996, CHILD NERV SYST, V12, P125, DOI 10.1007/BF00266812
   Behnke J, 1997, CHILD NERV SYST, V13, P122, DOI 10.1007/s003810050058
   Behnke J, 1997, CHILD NERV SYST, V13, P135, DOI 10.1007/s003810050059
   Benesch M, 2005, J NEURO-ONCOL, V72, P179, DOI 10.1007/s11060-004-3546-5
   Benesch M, 2011, J NEURO-ONCOL, V104, P279, DOI 10.1007/s11060-010-0485-1
   BERGER MS, 1983, NEUROSURGERY, V12, P298, DOI 10.1227/00006123-198303000-00008
   COHEN BH, 1995, J CLIN ONCOL, V13, P1687, DOI 10.1200/JCO.1995.13.7.1687
   Fangusaro JR, 2008, PEDIATR BLOOD CANCER, V50, P715, DOI 10.1002/pbc.21032
   Farmer JP, 2001, PEDIATR NEUROSURG, V34, P206, DOI 10.1159/000056021
   Freeman CR, 1998, INT J RADIAT ONCOL, V40, P265, DOI 10.1016/S0360-3016(97)00572-5
   Friedrich C, 2013, NEURO-ONCOLOGY, V15, P224, DOI 10.1093/neuonc/nos292
   Gerber NU, 2012, NEURO-ONCOLOGY, V14, P95
   Gibson P, 2010, NATURE, V468, P1095, DOI 10.1038/nature09587
   GNEKOW AK, 1995, MED PEDIATR ONCOL, V24, P104, DOI 10.1002/mpo.2950240209
   Grammel D, 2012, ACTA NEUROPATHOL, V123, P601, DOI 10.1007/s00401-012-0961-0
   Jakacki RI, 2005, J NEUROSURG, V102, P44, DOI 10.3171/ped.2005.102.1.0044
   Kaatsch P, 2001, CANCER-AM CANCER SOC, V92, P3155, DOI 10.1002/1097-0142(20011215)92:12<3155::AID-CNCR10158>3.0.CO;2-C
   Lannering B, 2012, J CLIN ONCOL, V30, P3187, DOI 10.1200/JCO.2011.39.8719
   Li Mei Hua, 2005, Neurosurg Focus, V19, pE3
   LITTMAN P, 1980, CANCER, V45, P2787, DOI 10.1002/1097-0142(19800601)45:11<2787::AID-CNCR2820451113>3.0.CO;2-V
   Marec-Berard P, 2002, MED PEDIATR ONCOL, V38, P83, DOI 10.1002/mpo.1277
   Massimino M, 2006, INT J RADIAT ONCOL, V64, P1031, DOI 10.1016/j.ijrobp.2005.09.026
   Pfister S, 2007, GENE CHROMOSOME CANC, V46, P839, DOI 10.1002/gcc.20471
   PIERREKAHN A, 1993, J NEUROSURG, V79, P845, DOI 10.3171/jns.1993.79.6.0845
   Pizer BL, 2006, EUR J CANCER, V42, P1120, DOI 10.1016/j.ejca.2006.01.039
   Rutkowski S, 2005, NEW ENGL J MED, V352, P978, DOI 10.1056/NEJMoa042176
   Sturm D, 2013, NEURO-ONCOLOGY, V15, P176
   Sufit A, 2012, J NEUROSURG-PEDIATR, V10, P81, DOI 10.3171/2012.3.PEDS11316
   Timmermann B, 2006, J CLIN ONCOL, V24, P1554, DOI 10.1200/JCO.2005.04.8074
   Timmermann B, 2002, J CLIN ONCOL, V20, P842, DOI 10.1200/JCO.20.3.842
   von Bueren AO, 2011, NEURO-ONCOLOGY, V13, P669, DOI 10.1093/neuonc/nor025
   Warmuth-Metz M, 2009, CLIN NEURORADIOL, V19, P263, DOI 10.1007/s00062-009-9029-5
   Zagzag D, 2000, PEDIATRICS, V106, P1045, DOI 10.1542/peds.106.5.1045
NR 36
TC 7
Z9 12
U1 0
U2 2
PU AMER ASSOC NEUROLOGICAL SURGEONS
PI ROLLING MEADOWS
PA 5550 MEADOWBROOK DRIVE, ROLLING MEADOWS, IL 60008 USA
SN 1933-0707
EI 1933-0715
J9 J NEUROSURG-PEDIATR
JI J. Neurosurg.-Pediatr.
PD MAR
PY 2015
VL 15
IS 3
BP 227
EP 235
DI 10.3171/2014.9.PEDS14213
PG 9
WC Clinical Neurology; Pediatrics; Surgery
WE Science Citation Index Expanded (SCI-EXPANDED)
SC Neurosciences & Neurology; Pediatrics; Surgery
GA CC3RL
UT WOS:000350266900001
PM 25555122
OA Bronze
DA 2023-08-27
ER

PT J
AU Shah, A
   Jhawar, SS
   Goel, A
AF Shah, Abhidha
   Jhawar, Sukhdeep Singh
   Goel, Atul
TI The lateral cerebral peduncle approach to ventrally placed intra-axial
   midbrain tumors: A technical note
SO JOURNAL OF CLINICAL NEUROSCIENCE
LA English
DT Article
DE Brainstem; Cruscerebri; Midbrain; Interpeduncular cistern; Safe entry
   zone
ID SAFE ENTRY ZONES; BRAIN-STEM; MICROSURGICAL ANATOMY
AB We describe the anatomical landmarks and surgical feasibility of a novel 'safe' brainstem entry zone to approach ventrally placed intra-axial midbrain tumors. The anatomy of the brainstem was specifically studied to evaluate safe surgical entry zone in the mid brain on two formalin fixed silicon injected cadaver head specimens. A novel entry point through the lateral one - fifth of the cerebral peduncle was identified to be 'safe' to approach lesions of the ventral midbrain. Three patients, having oculomotor schwannoma, peduncular glioma and a peduncular cavernoma were operated using this safe entry zone. To approach the midbrain, retrosigmoid lateral supracerebellar route was used in two patients and a basal subtemporal avenue was deployed in one patient. On the basis of fine microanatomical dissection on cadavers, a novel entry point through the lateral one-fifth of the cerebral peduncle, 5 mm anterior to the lateral mesencephalic sulcus and approximately 5 mm superior to the fourth nerve was identified. The proposed brainstem entry point traverses the parieto-temporo-occipital pontine fibers and the trajectory is between the corticospinal tracts ventrally and the substantia nigra dorsally. Three patients were operated successfully using the approach. There were no post-operative motor, sensory or extra-pyramidal deficits. The corridor through the lateral one-fifth of the cerebral peduncle presents a safe and relative 'easy' surgical route to approach ventrally placed intra-axial midbrain tumors. (c) 2021 Elsevier Ltd. All rights reserved.
C1 [Shah, Abhidha; Jhawar, Sukhdeep Singh; Goel, Atul] KEM Hosp & Seth GS Med Coll, Dept Neurosurg, Mumbai 400012, Maharashtra, India.
   [Goel, Atul] Lilavati Hosp & Res Ctr, Mumbai, Maharashtra, India.
C3 Seth Gordhandas Sunderdas Medical College & King Edward Memorial
   Hospital
RP Goel, A (通讯作者)，KEM Hosp & Seth GS Med Coll, Dept Neurosurg, Mumbai 400012, Maharashtra, India.
EM
CR Ramos AA, 2013, PEDIATR NEUROL, V48, P472, DOI 10.1016/j.pediatrneurol.2013.02.009
   Bertalanffy H, 2002, NEUROSURG REV, V25, P1, DOI 10.1007/s101430100179
   Cavalcanti DD, 2018, WORLD NEUROSURG, V119, pE818, DOI 10.1016/j.wneu.2018.07.272
   Cavalcanti DD, 2016, J NEUROSURG, V124, P1359, DOI 10.3171/2015.4.JNS141945
   Cavalcanti DD, 2020, J NEUROSURG, V132, P1653, DOI 10.3171/2019.1.JNS182036
   Giliberto G, 2010, NEUROSURG FOCUS, V29, DOI 10.3171/2010.6.FOCUS10133
   Goel A, 2010, ACTA NEUROCHIR, V152, P1721, DOI 10.1007/s00701-010-0690-x
   Kalani MYS, 2018, J NEUROSURG, V128, P834, DOI 10.3171/2016.9.JNS161680
   Kalani MYS, 2016, J NEUROSURG, V125, P1596, DOI 10.3171/2016.6.JNS161043
   Meybodi AT, 2020, WORLD NEUROSURG, VS1878, P31178
   Mukherjee D, 2020, NEUROSURG REV, V43, P153, DOI 10.1007/s10143-018-1023-4
   Parraga RG, 2016, J NEUROSURG, V124, P1377, DOI 10.3171/2015.4.JNS132778
   Recalde RJ, 2008, NEUROSURGERY, V63, P9, DOI [10.1227/01.NEU.0000297062.52433.3F, 10.1227/01.neu.0000317368.69523.40]
   Shah A, 2020, WORLD NEUROSURG, V134, pE826, DOI 10.1016/j.wneu.2019.11.016
   Tanabe Michiharu, 1996, Neurologia Medico-Chirurgica, V36, P880, DOI 10.2176/nmc.36.880
   Yagmurlu K, 2014, OPER NEUROSURG, V10, P602, DOI 10.1227/NEU.0000000000000466
   Yang Y, 2019, NEUROSURG REV, V42, P721, DOI 10.1007/s10143-019-01081-9
NR 17
TC 1
Z9 1
U1 1
U2 4
PU ELSEVIER SCI LTD
PI OXFORD
PA THE BOULEVARD, LANGFORD LANE, KIDLINGTON, OXFORD OX5 1GB, OXON, ENGLAND
SN 0967-5868
EI 1532-2653
J9 J CLIN NEUROSCI
JI J. Clin. Neurosci.
PD JUL
PY 2021
VL 89
BP 226
EP 231
DI 10.1016/j.jocn.2021.04.036
EA MAY 2021
PG 6
WC Clinical Neurology; Neurosciences
WE Science Citation Index Expanded (SCI-EXPANDED)
SC Neurosciences & Neurology
GA SP8KR
UT WOS:000659912500036
PM 34119272
DA 2023-08-27
ER

PT J
AU Madden, J
   Foreman, NK
   Liu, AK
AF Madden, Jenny
   Foreman, Nicholas K.
   Liu, Arthur K.
TI Germ cell tumors of the brainstem: report on two cases with pulmonary
   complications and a review of the literature
SO JOURNAL OF NEURO-ONCOLOGY
LA English
DT Article
DE Germ cell tumor; Brainstem
ID PRIMARY INTRACRANIAL GERMINOMA; MEDULLA-OBLONGATA GERMINOMA; MIDBRAIN;
   CHEMOTHERAPY; GLIOMA
AB Intracranial germ cell tumors most commonly present in the pineal and suprasellar regions. Rare cases of brainstem germ cell tumors have been described. Here, we present two cases with brainstem germ cell tumors and review the current literature. The two patients did well, in terms of disease control, but have significant morbidity related to respiratory issues. One patient remains ventilator dependent and the second died from respiratory arrest.
C1 [Liu, Arthur K.] Univ Colorado Denver, Dept Radiat Oncol, Aurora, CO 80045 USA.
   [Madden, Jenny; Foreman, Nicholas K.] Childrens Hosp, Dept Neurooncol, Aurora, CO USA.
C3 Children's Hospital Colorado; University of Colorado System; University
   of Colorado Anschutz Medical Campus; Children's Hospital Colorado
RP Liu, AK (通讯作者)，Univ Colorado Denver, Dept Radiat Oncol, Anschutz Canc Pavil,MS F706,1665 N Ursula St,Suit, Aurora, CO 80045 USA.
EM
OI Foreman, Nicholas/0000-0003-4848-4801
CR Ben Amor S, 2004, BRIT J NEUROSURG, V18, P310, DOI 10.1080/02688690410001732832
   BJORNSSON J, 1985, J NEUROPATH EXP NEUR, V44, P32, DOI 10.1097/00005072-198501000-00003
   *CBTRUS, 2000, STAT REP PRIM BRAIN
   Douglas JG, 2006, J PEDIAT HEMATOL ONC, V28, P36
   FELIX I, 1991, PEDIATR NEUROSURG, V16, P156, DOI 10.1159/000120517
   HASHIMOTO M, 1992, SURG NEUROL, V37, P384, DOI 10.1016/0090-3019(92)90009-C
   HO DM, 1992, CANCER, V70, P1577, DOI 10.1002/1097-0142(19920915)70:6<1577::AID-CNCR2820700622>3.0.CO;2-X
   Hui SHL, 2000, CHEST, V118, P266, DOI 10.1378/chest.118.1.266
   JENNINGS MT, 1985, J NEUROSURG, V63, P155, DOI 10.3171/jns.1985.63.2.0155
   Koizumi H, 2006, ACTA NEUROCHIR, V148, P1197, DOI 10.1007/s00701-006-0872-8
   Kretschmar C, 2007, PEDIATR BLOOD CANCER, V48, P285, DOI 10.1002/pbc.20815
   Manning HL, 2000, AM J RESP CRIT CARE, V161, P2100, DOI 10.1164/ajrccm.161.6.9904087
   Matsumoto K, 1998, CLIN NEUROL NEUROSUR, V100, P292, DOI 10.1016/S0303-8467(98)00050-X
   Nakajima H, 2000, SURG NEUROL, V53, P448, DOI 10.1016/S0090-3019(00)00224-X
   POUNGVARIN N, 1991, Journal of the Medical Association of Thailand, V74, P55
   Sugiyama K, 1994, Neurol Med Chir (Tokyo), V34, P291, DOI 10.2176/nmc.34.291
   Tashiro Takashi, 1993, Neurologia Medico-Chirurgica, V33, P251, DOI 10.2176/nmc.33.251
   Uchino M, 2006, J NEURO-ONCOL, V79, P255, DOI 10.1007/s11060-006-9138-9
   VALENTE S, 1993, CHEST, V103, P1909, DOI 10.1378/chest.103.6.1909
   Yen PS, 2003, J NEURO-ONCOL, V62, P339, DOI 10.1023/A:1023331700995
NR 20
TC 1
Z9 1
U1 0
U2 2
PU SPRINGER
PI NEW YORK
PA 233 SPRING ST, NEW YORK, NY 10013 USA
SN 0167-594X
EI 1573-7373
J9 J NEURO-ONCOL
JI J. Neuro-Oncol.
PD JUL
PY 2009
VL 93
IS 3
BP 405
EP 408
DI 10.1007/s11060-008-9780-5
PG 4
WC Oncology; Clinical Neurology
WE Science Citation Index Expanded (SCI-EXPANDED)
SC Oncology; Neurosciences & Neurology
GA 451PS
UT WOS:000266482700016
PM 19139823
DA 2023-08-27
ER

PT J
AU Wu, Q
   Tyler, B
   Sukay, L
   Rhines, L
   DiMeco, F
   Clatterbuck, RE
   Guarnieri, M
   Carson, BS
AF Wu, Q
   Tyler, B
   Sukay, L
   Rhines, L
   DiMeco, F
   Clatterbuck, RE
   Guarnieri, M
   Carson, BS
TI Experimental rodent models of brainstem tumors
SO VETERINARY PATHOLOGY
LA English
DT Article
DE animal model; brainstem tumors; central nervous system cancer; medulla;
   pons
ID GENE-THERAPY; CHEMOTHERAPY; GLIOMAS; RADIOTHERAPY; DELIVERY; CELLS
AB Despite recent advances in surgical technology, resection is not an option for many brainstem tumors. Experimental models have played essential roles in examining new approaches to therapy. The objective of the present study was to generate models by determining coordinates for safe inoculation into the brainstem of mice and rats, and to establish whether the implantation of heterotopic cells would create reproducible survival curves. Morbidity and survival studies were used to map stereotactic coordinates allowing successful inoculation of tumor cells. Survival studies were used to investigate the time course of tumor growth. Tumor location was examined by light microscopy and magnetic resonance imaging. Mice survived injections of 2 muL of saline at interaural, lateral, and depth coordinates of -2.5, 1.0, and 3.5 mm and -1.5, 1.0, and 3.5 mm. Rats survived injections at interaural, lateral, and depth coordinates of -2.0, 2.0, and 7.0 mm and -3.0, 0, and 7.0 mm. Median survival of mice challenged with 5 X 101 EMT6 and 104 B 16 tumor cells was I I and 10 days, respectively. Median survival for rats challenged with 10(4) 9L and F98 cells was 14 and 13 days, respectively. The present study demonstrates a feasible approach to preparing models of brainstem tumors. Limitations of these models are discussed.
C1 Johns Hopkins Neurol Surg, Div Pediat Neurosurg, Baltimore, MD USA.
   Johns Hopkins Neurol Surg, Hunterian Brain Tumor Labs, Baltimore, MD USA.
C3 Johns Hopkins University; Johns Hopkins Medicine; Johns Hopkins
   University; Johns Hopkins Medicine
RP Guarnieri, M (通讯作者)，Johns Hopkins Neurol Surg, Div Pediat Neurosurg, Baltimore, MD USA.
EM
RI DiMeco, Francesco/Z-5431-2019; DiMeco, Francesco/Q-3785-2016
OI DiMeco, Francesco/0000-0001-6213-5793
CR Adams DM, 2000, ANTICANCER RES, V20, P3395
   Ambar BB, 1999, HUM GENE THER, V10, P1641, DOI 10.1089/10430349950017644
   Bhatnagar R., 1995, Cytobios, V82, P239
   CAHAN MA, 1994, CANCER CHEMOTH PHARM, V33, P441, DOI 10.1007/BF00686276
   Cartmill M, 1999, CHILD NERV SYST, V15, P235
   CHEN L, 1995, CANCER RES, V55, P581
   Clatterbuck RE, 1997, NEUROSURGERY, V40, P339, DOI 10.1097/00006123-199702000-00019
   Ewend MG, 1996, CANCER RES, V56, P5217
   Ewend MG, 1996, SEMIN SURG ONCOL, V12, P429, DOI 10.1002/(SICI)1098-2388(199611/12)12:6<429::AID-SSU8>3.0.CO;2-C
   Fisher PG, 2000, CANCER, V89, P1569, DOI 10.1002/1097-0142(20001001)89:7<1569::AID-CNCR22>3.0.CO;2-0
   Fisher PG, 1996, CONT NEUROSURG, V18, P1
   Freeman CR, 1999, CHILD NERV SYST, V15, P545, DOI 10.1007/s003810050542
   Freeman CR, 1998, INT J RADIAT ONCOL, V40, P265, DOI 10.1016/S0360-3016(97)00572-5
   Freeman CR, 1996, PEDIATR NEUROSURG, V24, P103, DOI 10.1159/000121025
   JACKOWSKI A, 1995, BRIT J NEUROSURG, V9, P581
   Kalifa C, 1999, CHILD NERV SYST, V15, P498, DOI 10.1007/s003810050538
   Kaplan AM, 1996, PEDIATR NEUROSURG, V24, P185, DOI 10.1159/000121036
   Klopp LS, 2000, VET SURG, V29, P533, DOI 10.1053/jvet.2000.17851
   Packer RJ, 1999, PRINCIPLES AND PRACTICE OF PEDIATRIC NEUROSURGERY, P779
   PANISCH W, 1990, PATHOLOGY TUMORS LAB, V1, P677
   Paxinos, 2008, MOUSE BRAIN STEREOTA
   PENN RD, 1983, APPL NEUROPHYSIOL, V46, P240
   POLLACK IF, 1993, J NEUROSURG, V78, P859, DOI 10.3171/jns.1993.78.6.0859
   ROSSRIVEROS P, 1979, RADIAT RES, V78, P296, DOI 10.2307/3575046
   Rubin G, 1998, CHILD NERV SYST, V14, P167, DOI 10.1007/s003810050205
   Schabet M, 1997, MED PEDIATR ONCOL, V29, P92, DOI 10.1002/(SICI)1096-911X(199708)29:2<92::AID-MPO5>3.0.CO;2-M
   Schild SE, 1998, J NEURO-ONCOL, V40, P171, DOI 10.1023/A:1006193306286
   STARK M, 1992, NEUROTOXICOL TERATOL, V14, P247, DOI 10.1016/0892-0362(92)90003-S
   STECK J, 1995, SURG NEUROL, V43, P563, DOI 10.1016/0090-3019(95)00156-5
   SWANSON LW, 1980, NEUROENDOCRINOLOGY, V31, P410, DOI 10.1159/000123111
   WECHSLER W, 1989, ACTA NEUROCHIR, V98, P77, DOI 10.1007/BF01407181
   WHEELER KT, 1983, RADIAT ENVIRON BIOPH, V22, P3, DOI 10.1007/BF01323757
   YACHNIS AT, 2000, SEMIN PEDIAT NEUROL, V4, P282
   Zlokovic BV, 1997, NEUROSURGERY, V40, P805, DOI 10.1097/00006123-199704000-00028
NR 34
TC 19
Z9 21
U1 0
U2 1
PU SAGE PUBLICATIONS INC
PI THOUSAND OAKS
PA 2455 TELLER RD, THOUSAND OAKS, CA 91320 USA
SN 0300-9858
EI 1544-2217
J9 VET PATHOL
JI Vet. Pathol.
PD MAY
PY 2002
VL 39
IS 3
BP 293
EP 299
DI 10.1354/vp.39-3-293
PG 7
WC Pathology; Veterinary Sciences
WE Science Citation Index Expanded (SCI-EXPANDED)
SC Pathology; Veterinary Sciences
GA 548RP
UT WOS:000175403400001
PM 12014493
OA Green Submitted
DA 2023-08-27
ER

PT J
AU Baroni, LV
   Solano-Paez, P
   Nobre, L
   Michaeli, O
   Hawkins, C
   Laughlin, S
   Bartels, U
   Ramaswamy, V
   Bouffet, E
AF Baroni, Lorena V.
   Solano-Paez, Palma
   Nobre, Liana
   Michaeli, Orli
   Hawkins, Cynthia
   Laughlin, Susanne
   Bartels, Ute
   Ramaswamy, Vijay
   Bouffet, Eric
TI Indolent course of brainstem tumors with K27M-H3.3 mutation
SO PEDIATRIC BLOOD & CANCER
LA English
DT Article
DE brainstem; diffuse intrinsic pontine glioma; DIPG; indolent course;
   K27M; tumor
ID HIGH-GRADE; PONTINE; GLIOMA; H3F3A
AB Diffuse intrinsic pontine glioma (DIPG) is characterized by a short history of brainstem symptoms and well-known magnetic resonance imaging features with a fatal outcome. However, we report three unusual cases of brainstem tumors with an initial indolent and protracted course, which subsequently developed the classical imaging and clinical features of DIPG. Our findings support this notion that K27M is an early event in development and suggest that the emergence of additional events resulted in rapid progression after a long period of latency. Identification of such markers of aggressive behavior in the context of an indolent course is needed for better characterization and treatment management.
C1 [Baroni, Lorena V.; Solano-Paez, Palma; Nobre, Liana; Michaeli, Orli; Bartels, Ute; Ramaswamy, Vijay; Bouffet, Eric] Hosp Sick Children, Div Haematol Oncol, 555 Univ Ave, Toronto, ON M5G 1X8, Canada.
   [Baroni, Lorena V.] Hosp JP Garrahan, Serv Hematol Oncol, Buenos Aires, DF, Argentina.
   [Hawkins, Cynthia] Hosp Sick Children, Dept Pathol, Toronto, ON, Canada.
   [Laughlin, Susanne] Hosp Sick Children, Dept Diagnost Imaging, Toronto, ON, Canada.
   [Ramaswamy, Vijay] Univ Toronto, Dept Med Biophys, Toronto, ON, Canada.
C3 University of Toronto; Hospital for Sick Children (SickKids); Hospital
   de Pediatria Doctor Juan Garrahan; University of Buenos Aires;
   University of Toronto; Hospital for Sick Children (SickKids); University
   of Toronto; Hospital for Sick Children (SickKids); University of Toronto
RP Bouffet, E (通讯作者)，Hosp Sick Children, Div Haematol Oncol, 555 Univ Ave, Toronto, ON M5G 1X8, Canada.
EM
RI michaeli, orli/AAY-7086-2021; Bouffet, Eric/AAB-8999-2020; SOLANO,
   PALMA/ABH-2475-2020
OI SOLANO, PALMA/0000-0002-0763-7552; Bouffet, Eric/0000-0002-6832-6539;
   Michaeli, Orli/0000-0002-6097-6457; Ramaswamy, Vijay/0000-0002-6557-895X
FU Meagan's Walk Foundation Fellowship in Pediatric Neuro-Oncology; Terry
   Fox Foundation International Fellowship
FX Meagan's Walk Foundation Fellowship in Pediatric Neuro-Oncology and The
   Terry Fox Foundation International Fellowship (to L.B.).
CR Castel D, 2016, ACTA NEUROPATHOL, V131, P795, DOI 10.1007/s00401-016-1568-7
   Castel D, 2015, ACTA NEUROPATHOL, V130, P815, DOI 10.1007/s00401-015-1478-0
   El Ahmadieh TY, 2019, WORLD NEUROSURG, V124, P87, DOI 10.1016/j.wneu.2018.12.147
   Hochart A, 2015, ANN CLIN TRANSL NEUR, V2, P439, DOI 10.1002/acn3.184
   Hoffman LM, 2018, J CLIN ONCOL, V36, P1963, DOI 10.1200/JCO.2017.75.9308
   Ishibashi K, 2016, CHILD NERV SYST, V32, P2433, DOI 10.1007/s00381-016-3161-8
   Joyon N, 2017, NEUROPATH APPL NEURO, V43, P271, DOI 10.1111/nan.12329
   Larson JD, 2019, CANCER CELL, V35, P140, DOI 10.1016/j.ccell.2018.11.015
   Mackay A, 2017, CANCER CELL, V32, P520, DOI 10.1016/j.ccell.2017.08.017
   Nagaraja S, 2017, CANCER CELL, V31, P635, DOI 10.1016/j.ccell.2017.03.011
   Nakano Y, 2019, BRAIN TUMOR PATHOL, V36, P162, DOI 10.1007/s10014-019-00347-w
   Orillac C, 2016, ACTA NEUROPATHOL COM, V4, DOI 10.1186/s40478-016-0361-0
   Pathania M, 2017, CANCER CELL, V32, P684, DOI 10.1016/j.ccell.2017.09.014
   Ramaswamy V, 2017, CANCER CELL, V32, P548, DOI 10.1016/j.ccell.2017.10.013
NR 14
TC 4
Z9 4
U1 0
U2 5
PU WILEY
PI HOBOKEN
PA 111 RIVER ST, HOBOKEN 07030-5774, NJ USA
SN 1545-5009
EI 1545-5017
J9 PEDIATR BLOOD CANCER
JI Pediatr. Blood Cancer
PD MAR
PY 2020
VL 67
IS 3
AR e28102
DI 10.1002/pbc.28102
EA DEC 2019
PG 4
WC Oncology; Hematology; Pediatrics
WE Science Citation Index Expanded (SCI-EXPANDED)
SC Oncology; Hematology; Pediatrics
GA KF0IN
UT WOS:000499823800001
PM 31793190
DA 2023-08-27
ER

PT J
AU Yen, CP
   Sheehan, J
   Patterson, G
   Steiner, L
AF Yen, Chun Po
   Sheehan, Jason
   Patterson, Greg
   Steiner, Ladislau
TI Gamma Knife surger, for metastatic brainstem tumors
SO JOURNAL OF NEUROSURGERY
LA English
DT Article
DE brainstem; cance; metastasis; radiosurgery; Gamma Knife
ID RADIATION-THERAPY; STEREOTACTIC RADIOSURGERY; CEREBRAL METASTASES;
   PROGNOSTIC-FACTORS; RANDOMIZED-TRIAL; IRRADIATION; RADIOTHERAPY;
   MANAGEMENT; CARCINOMA; MELANOMA
AB Object. The authors review imaging and clinical outcomes in patients with metastatic brainstem tumors treated using Gamma Knife surgery (GKS).
   Methods. Between March 1989 and March 2005, 53 patients (24 men and 29 women) with metastatic brainstem lesions underwent GKS. The metastatic deposits were located in the midbrain in eight patients, the pons in 42, and the medulla oblongata in three. Lung cancer was the most common primary malignancy, followed by breast cancer, melanoma, and renal cell carcinoma. The mean volume of the metastatic deposits at the time of treatment was 2.8 cm(3) (range 0.05-21 cm(3)). The prescription doses varied from 9 to 25 Gy (mean 17.6 Gy).
   Imaging follow-up studies were not completed in 16 patients, because of the short-term survival in 11 and patient refusal in five. Of the remaining 37 patients, who underwent an imaging follow-up evaluation at a mean of 9.8 months (range 1-25 months), the tumors disappeared in seven, shrank in 22, remained unchanged in three, and grew in five. All but one of 18 patients with asymptomatic brainstem deposits remained free of symptoms. In 35 patients with symptomatic brainstem deposits, neurological symptoms improved in 21, remained stable in 11, and worsened in three. At the time of this study, 10 patients were alive, and their survival ranged from 3 to 52 months after treatment. Thirty-four patients died of extracranial disease, three of the progressing metastatic brainstem lesion, and six of additional progressing intracranial deposits in other parts of the brain. The overall median survival period was 11 months after GKS. In terms of survival, the absence of active extracranial disease was the only favorable prognostic factor. Neither previous whole-brain radiation therapy nor a single brainstem metastasis was statistically related to the duration of survival.
   Conclusions. Compared with allowing a metastatic brainstem lesion to take its natural course, GKS prolongs survival. The risks associated with such treatment are low. The severity of systemic diseases largely determines the prognosis of metastases to the brainstem.
C1 Univ Virginia, Hlth Syst, Dept Neurol Surg, Lars Leksell Ctr Gamma Surg, Charlottesville, VA 22908 USA.
C3 University of Virginia
RP Steiner, L (通讯作者)，Univ Virginia, Hlth Syst, Dept Neurol Surg, Lars Leksell Ctr Gamma Surg, 1215 Lee St,Room G-512, Charlottesville, VA 22908 USA.
EM
CR Andrews DW, 2004, LANCET, V363, P1665, DOI 10.1016/S0140-6736(04)16250-8
   Barker FG, 2004, CANCER-AM CANCER SOC, V100, P999, DOI 10.1002/cncr.20058
   BODEN G, 1950, J FAC RADIOL-LONDON, V2, P79, DOI 10.1016/S0368-2242(50)80056-8
   Borden JA, 2000, J NEUROSURG, V93, P228, DOI 10.3171/jns.2000.93.supplement_3.0228
   CAIRNCROSS JG, 1980, ANN NEUROL, V7, P529, DOI 10.1002/ana.410070606
   DELATTRE JY, 1988, ARCH NEUROL-CHICAGO, V45, P741, DOI 10.1001/archneur.1988.00520310047016
   Ellerhorst J, 2001, INT J RADIAT ONCOL, V49, P93, DOI 10.1016/S0360-3016(00)01355-9
   EMAMI B, 1991, INT J RADIAT ONCOL, V21, P109, DOI 10.1016/0360-3016(91)90171-Y
   HAAR F, 1972, CANCER, V30, P1241, DOI 10.1002/1097-0142(197211)30:5<1241::AID-CNCR2820300515>3.0.CO;2-5
   Huang CF, 1999, J NEUROSURG, V91, P563, DOI 10.3171/jns.1999.91.4.0563
   HUNTER KMF, 1968, CAN MED ASSOC J, V98, P1
   Jawahar A, 2004, J NEUROSURG, V100, P842, DOI 10.3171/jns.2004.100.5.0842
   Julow Jeno, 2004, Strahlenther Onkol, V180, P449
   Lagerwaard FJ, 1999, INT J RADIAT ONCOL, V43, P795, DOI 10.1016/S0360-3016(98)00442-8
   LANG EF, 1964, SURG CLIN N AM, V44, P865
   MARKESBERY WR, 1978, ARCH NEUROL-CHICAGO, V35, P754, DOI 10.1001/archneur.1978.00500350058012
   Matsumoto K, 1998, J NEUROSURG, V88, P141, DOI 10.3171/jns.1998.88.1.0141
   Mingione V, 2002, J NEUROSURG, V96, P544, DOI 10.3171/jns.2002.96.3.0544
   Muacevic A, 1999, J NEUROSURG, V91, P35, DOI 10.3171/jns.1999.91.1.0035
   Pan HC, 2005, J NEUROSURG, V102, P128, DOI 10.3171/jns.2005.102.s_supplement.0128
   PATCHELL RA, 1990, NEW ENGL J MED, V322, P494, DOI 10.1056/NEJM199002223220802
   Payne BR, 2000, J NEUROSURG, V92, P760, DOI 10.3171/jns.2000.92.5.0760
   Serizawa T, 2000, J NEUROSURG, V93, P32, DOI 10.3171/jns.2000.93.supplement_3.0032
   SHELINE GE, 1980, INT J RADIAT ONCOL, V6, P1215, DOI 10.1016/0360-3016(80)90175-3
   Shiau CY, 1997, INT J RADIAT ONCOL, V37, P375, DOI 10.1016/S0360-3016(96)00497-X
   Shuto T, 2003, ACTA NEUROCHIR, V145, P755, DOI 10.1007/s00701-003-0034-1
   Simonova G, 2000, RADIOTHER ONCOL, V57, P207, DOI 10.1016/S0167-8140(00)00267-X
   Soffietti R, 2002, J NEUROL, V249, P1357, DOI 10.1007/s00415-002-0870-6
   Suzuki S, 2000, J NEUROSURG, V93, P30, DOI 10.3171/jns.2000.93.supplement_3.0030
   Tosoni A, 2004, CRIT REV ONCOL HEMAT, V52, P199, DOI 10.1016/j.critrevonc.2004.08.006
   Varlotto JM, 2005, INT J RADIAT ONCOL, V62, P1125, DOI 10.1016/j.ijrobp.2004.12.092
   VECHT CJ, 1993, ANN NEUROL, V33, P583, DOI 10.1002/ana.410330605
NR 32
TC 53
Z9 55
U1 0
U2 5
PU AMER ASSOC NEUROLOGICAL SURGEONS
PI ROLLING MEADOWS
PA 5550 MEADOWBROOK DRIVE, ROLLING MEADOWS, IL 60008 USA
SN 0022-3085
EI 1933-0693
J9 J NEUROSURG
JI J. Neurosurg.
PD AUG
PY 2006
VL 105
IS 2
BP 213
EP 219
DI 10.3171/jns.2006.105.2.213
PG 7
WC Clinical Neurology; Surgery
WE Science Citation Index Expanded (SCI-EXPANDED)
SC Neurosciences & Neurology; Surgery
GA 070PF
UT WOS:000239534400012
PM 17219825
DA 2023-08-27
ER

PT J
AU Perez-Gomez, JL
   Rodriguez-Alvarez, CA
   Marhx-Bracho, A
   Rueda-Franco, F
AF Perez-Gomez, Jose L.
   Rodriguez-Alvarez, Carlos A.
   Marhx-Bracho, Alfonso
   Rueda-Franco, Fernando
TI Stereotactic biopsy for brainstem tumors in pediatric patients
SO CHILDS NERVOUS SYSTEM
LA English
DT Article
DE Brainstem tumors; Brainstem gliomas; Stereotactic biopsy; Brainstem
ID SUBOCCIPITAL TRANSCEREBELLAR BIOPSY; MASS LESIONS; SURGICAL INDICATIONS;
   POSTERIOR-FOSSA; GLIOMAS; CHILDHOOD; CHILDREN
AB Our aim is to describe clinical and paraclinical features in patients who underwent stereotactic-guided biopsy for brainstem tumors.
   A study of case series was made by reviewing the records of patients who underwent stereotactic biopsy for brainstem tumors.
   Stereotactic biopsy for brainstem tumors was performed (between 2000 and 2008) in 20 children (11 girls, and 9 boys), mean age 7.95 +/- 3.12 years at the time of diagnosis. The mean time from onset of symptoms to diagnosis was 6.59 +/- 13.58 months (0.50-60 months). The most frequent symptoms and signs at onset were related to disturbance of cerebellar function and cranial nerve nuclei. Location was pontomesencephalic (35%), pontine (30%), pontomedullar (25%), and in the whole brainstem (10%). The most common type of image was intrinsic-diffused (55%). The histopathology was anaplastic astrocytoma (30%), followed by fibrillary and pilocytic types (25% each), low-grade astrocytoma (5%), high-grade astrocytoma (5%), and normal tissue (10%). Mild complications were observed in only two cases.
   Stereotactic biopsy done for clarifiying a diagnostic imaging in brainstem tumors is important in obtaining a definitive diagnosis with a low rate of complications.
C1 [Perez-Gomez, Jose L.; Rodriguez-Alvarez, Carlos A.; Marhx-Bracho, Alfonso; Rueda-Franco, Fernando] Inst Nacl Pediat, Dept Neurocirugia Pediat, Mexico City 04530, DF, Mexico.
RP Rodriguez-Alvarez, CA (通讯作者)，Inst Nacl Pediat, Dept Neurocirugia Pediat, Insurgentes 3700-C, Mexico City 04530, DF, Mexico.
EM
CR ABERNATHEY CD, 1989, J NEUROSURG, V70, P195, DOI 10.3171/jns.1989.70.2.0195
   ALVISI C, 1985, ACTA NEUROCHIR, V76, P12, DOI 10.1007/BF01403823
   BERGER MS, 1983, NEUROSURGERY, V12, P298, DOI 10.1227/00006123-198303000-00008
   Choux M, 1999, PEDIAT NEUROSURGERY, P471
   de Leon FCP, 2003, CHILD NERV SYST, V19, P305, DOI 10.1007/s00381-003-0737-x
   EPSTEIN F, 1988, J NEURO-ONCOL, V6, P309
   EPSTEIN F, 1986, J NEUROSURG, V64, P11, DOI 10.3171/jns.1986.64.1.0011
   Giunta F, 1989, Acta Neurochir Suppl (Wien), V46, P86
   Goncalves-Ferreira AJ, 2003, SURG NEUROL, V60, P311, DOI 10.1016/S0090-3019(03)00379-3
   HOFFMAN HJ, 1980, NEUROSURGERY, V7, P243, DOI 10.1227/00006123-198009000-00007
   Jallo G, 2006, CHILD NERV SYST, V22, P1, DOI 10.1007/s00381-005-1267-5
   Jallo GI, 2004, CHILD NERV SYST, V20, P143, DOI 10.1007/s00381-003-0870-6
   Jallo GI, 2003, ANN NEUROSURG, V3, P1
   KONDZIOLKA D, 1995, SURG NEUROL, V43, P558, DOI 10.1016/0090-3019(95)00009-7
   KRATIMENOS GP, 1992, ACTA NEUROCHIR, V116, P164, DOI 10.1007/BF01540871
   LITTMAN P, 1980, CANCER, V45, P2787, DOI 10.1002/1097-0142(19800601)45:11<2787::AID-CNCR2820451113>3.0.CO;2-V
   LOBATO RD, 1989, J NEUROSURG, V71, P466
   Massager N, 2000, J NEUROSURG, V93, P951, DOI 10.3171/jns.2000.93.6.0951
   MATHISEN JR, 1987, SURG NEUROL, V28, P100, DOI 10.1016/0090-3019(87)90080-2
   Pincus DW, 2006, J NEUROSURG, V104, P108, DOI 10.3171/ped.2006.104.2.108
   STECK J, 1995, SURG NEUROL, V43, P563, DOI 10.1016/0090-3019(95)00156-5
   STROINK AR, 1987, NEUROSURGERY, V20, P439, DOI 10.1227/00006123-198703000-00014
   Valdes-Gorcia J, 1998, ACTA NEUROCHIR, V140, P899, DOI 10.1007/s007010050191
   Varan A, 2007, CHILD NERV SYST, V23, P315, DOI 10.1007/s00381-006-0243-z
   VILLAREJO OF, 2001, NEUROCIRUGIA PEDIAT, P366
   Walker DA, 1999, ARCH DIS CHILD, V80, P558, DOI 10.1136/adc.80.6.558
NR 26
TC 27
Z9 27
U1 0
U2 2
PU SPRINGER
PI NEW YORK
PA 233 SPRING ST, NEW YORK, NY 10013 USA
SN 0256-7040
EI 1433-0350
J9 CHILD NERV SYST
JI Childs Nerv. Syst.
PD JAN
PY 2010
VL 26
IS 1
BP 29
EP 34
DI 10.1007/s00381-009-1000-x
PG 6
WC Clinical Neurology; Pediatrics; Surgery
WE Science Citation Index Expanded (SCI-EXPANDED)
SC Neurosciences & Neurology; Pediatrics; Surgery
GA 518UZ
UT WOS:000271721700007
PM 19784659
DA 2023-08-27
ER

PT J
AU Fried, I
   Hawkins, C
   Scheinemann, K
   Tsangaris, E
   Hesselson, L
   Bartels, U
   Huang, A
   Laperriere, N
   Dirks, P
   Bouffet, E
   Tabori, U
AF Fried, Iris
   Hawkins, Cynthia
   Scheinemann, Katrin
   Tsangaris, Elena
   Hesselson, Leila
   Bartels, Ute
   Huang, Annie
   Laperriere, Normand
   Dirks, Peter
   Bouffet, Eric
   Tabori, Uri
TI Favorable outcome with conservative treatment for children with low
   grade brainstem tumors
SO PEDIATRIC BLOOD & CANCER
LA English
DT Article
DE CNS tumors; brainstem tumors; low grade glioma; pediatric
ID GLIOMAS; CLASSIFICATION; SYSTEM; MANAGEMENT; CHILDHOOD; SURVIVAL;
   SURGERY
AB Background Pediatric brainstem tumors (BST) comprise a heterogeneous group of entities. Data regarding treatment options and outcome of BST, specifically brainstem low grade tumors (BSLGT) are limited. In order to better define risk groups and evaluate treatment options for pediatric BST, we performed a comprehensive analysis of all BST patients treated in our hospital during the MRI era. Procedures. We retrospectively analyzed clinical, imaging, and pathology data at presentation, treatment, and outcome of all BST patients followed at the Hospital for Sick Children in Toronto over the last 25 years. Results. Of 1,801 children with brain tumors, 223 (12%) had a brainstem primary location. Tumors without pontine involvement were BSLGT in 98.3%, whereas 75% of tumors involving the pons were high grade (P = 0.0001). Patients with BSLGT had 5-year progression-free survival (PFS) and overall survival (OS) of 57 +/- 3% and 89 +/- 5%, respectively. Upfront observation of tumor residual conferred no survival disadvantage with 5-year PFS and OS of 57 +/- 5% and 93 +/- 3%, respectively. In the group of patients requiring further treatment, 5-year PFS and OS were comparable between chemotherapy and radiotherapy with 53 +/- 12% and 93 +/- 4% and 66 +/- 11% and 83 +/- 6%, respectively (P = 0.26 and 0.3, respectively). Conclusion. BST without pontine involvement are almost invariably BSLGT. Children with BSLGT have an excellent outcome even with careful initial observation. No clear benefit was observed for radiotherapy over chemotherapy when adjuvant treatment was needed. A conservative approach may be warranted for children with non-pontine brainstem lesions. Pediatr Blood Cancer 2012; 58: 556-560. (C) 2011 Wiley Periodicals, Inc.
C1 [Fried, Iris; Scheinemann, Katrin; Tsangaris, Elena; Hesselson, Leila; Bartels, Ute; Huang, Annie; Bouffet, Eric; Tabori, Uri] Hosp Sick Children, Div Haematol Oncol, Pediat Brain Tumor Program, Toronto, ON M5G 1X8, Canada.
   [Fried, Iris] Hadassah Med Ctr, Dept Pediat Hematol & Oncol, IL-91120 Jerusalem, Israel.
   [Hawkins, Cynthia] Hosp Sick Children, Dept Pathol, Toronto, ON M5G 1X8, Canada.
   [Scheinemann, Katrin] McMaster Childrens Hosp, Dept Pediat, Div Pediat Hematol & Oncol, Hamilton, ON, Canada.
   [Laperriere, Normand] Princess Margaret Hosp, Dept Radiat Oncol, Toronto, ON M4X 1K9, Canada.
   [Dirks, Peter] Hosp Sick Children, Dept Neurosurg, Toronto, ON M5G 1X8, Canada.
C3 University of Toronto; Hospital for Sick Children (SickKids); Hebrew
   University of Jerusalem; Hadassah University Medical Center; University
   of Toronto; Hospital for Sick Children (SickKids); McMaster University;
   University of Toronto; University Health Network Toronto; Princess
   Margaret Cancer Centre; University of Toronto; Hospital for Sick
   Children (SickKids)
RP Tabori, U (通讯作者)，Hosp Sick Children, Div Haematol Oncol, Pediat Brain Tumor Program, 555 Univ Ave, Toronto, ON M5G 1X8, Canada.
EM
RI Bouffet, Eric/AAB-8999-2020
OI Tabori, Uri/0000-0002-5019-2683; Laperriere, Normand/0000-0001-8930-9650
CR [Anonymous], 2007, WHO HISTOLOGICAL CLA
   BARKOVICH AJ, 1991, PEDIATR NEUROSURG, V16, P73, DOI 10.1159/000120511
   BERGER MS, 1983, NEUROSURGERY, V12, P298, DOI 10.1227/00006123-198303000-00008
   Bowers DC, 2000, PEDIATR NEUROSURG, V32, P24, DOI 10.1159/000028893
   Combs SE, 2009, RADIOTHER ONCOL, V91, P60, DOI 10.1016/j.radonc.2009.02.012
   EDWARDS MSB, 1994, J NEUROSURG, V80, P20, DOI 10.3171/jns.1994.80.1.0020
   EPSTEIN F, 1985, CANCER, V56, P1804, DOI 10.1002/1097-0142(19851001)56:7+<1804::AID-CNCR2820561316>3.0.CO;2-5
   EPSTEIN F, 1988, J NEURO-ONCOL, V6, P309
   Fangusaro J, 2009, J CHILD NEUROL, V24, P1409, DOI 10.1177/0883073809338960
   FARWELL JR, 1977, CANCER, V40, P3123, DOI 10.1002/1097-0142(197712)40:6<3123::AID-CNCR2820400656>3.0.CO;2-6
   Fischbein NJ, 1996, PEDIATR NEUROSURG, V24, P9, DOI 10.1159/000121010
   Fisher PG, 2000, CANCER, V89, P1569, DOI 10.1002/1097-0142(20001001)89:7<1569::AID-CNCR22>3.0.CO;2-0
   Fouladi M, 2003, CANCER-AM CANCER SOC, V97, P1084, DOI 10.1002/cncr.11119
   Frazier JL, 2009, J NEUROSURG-PEDIATR, V3, P259, DOI 10.3171/2008.11.PEDS08281
   Hargrove D, 2006, LANCET ONCOL, V7, P241, DOI 10.1016/S1470-2045(06)70615-5
   Jallo GI, 2005, NEUROSURGERY, V56, P74, DOI 10.1227/01.NEU.0000144782.39430.12
   JANSS AJ, 1995, CANCER-AM CANCER SOC, V75, P1051, DOI 10.1002/1097-0142(19950215)75:4<1051::AID-CNCR2820750423>3.0.CO;2-S
   LITTMAN P, 1980, CANCER, V45, P2787, DOI 10.1002/1097-0142(19800601)45:11<2787::AID-CNCR2820451113>3.0.CO;2-V
   Marcus KJ, 2005, INT J RADIAT ONCOL, V61, P374, DOI 10.1016/j.ijrobp.2004.06.012
   Merchant TE, 2009, J CLIN ONCOL, V27, P3691, DOI 10.1200/JCO.2008.21.2738
   Merchant TE, 2009, LANCET ONCOL, V10, P258, DOI 10.1016/S1470-2045(08)70342-5
   Mohanty A, 2009, NEUROL INDIA, V57, P231, DOI 10.4103/0028-3886.53258
   POLLACK IF, 1994, J NEUROSURG, V80, P681, DOI 10.3171/jns.1994.80.4.0681
   POLLACK IF, 1993, J NEUROSURG, V78, P859, DOI 10.3171/jns.1993.78.6.0859
   ROBERTSON PL, 1994, NEUROLOGY, V44, P1798, DOI 10.1212/WNL.44.10.1798
   Stark AM, 2005, PEDIATR NEUROL, V33, P33, DOI 10.1016/j.pediatrneurol.2004.12.014
   STROINK AR, 1986, J NEUROSURG, V65, P745, DOI 10.3171/jns.1986.65.6.0745
   TOMITA T, 1984, J NEURO-ONCOL, V2, P117
   Ueoka DI, 2009, J NEUROL SCI, V281, P20, DOI 10.1016/j.jns.2009.03.009
   Weiner HL, 1997, PEDIATR NEUROSURG, V27, P12, DOI 10.1159/000121219
NR 30
TC 20
Z9 22
U1 0
U2 2
PU WILEY
PI HOBOKEN
PA 111 RIVER ST, HOBOKEN 07030-5774, NJ USA
SN 1545-5009
EI 1545-5017
J9 PEDIATR BLOOD CANCER
JI Pediatr. Blood Cancer
PD APR
PY 2012
VL 58
IS 4
BP 556
EP 560
DI 10.1002/pbc.23200
PG 5
WC Oncology; Hematology; Pediatrics
WE Science Citation Index Expanded (SCI-EXPANDED)
SC Oncology; Hematology; Pediatrics
GA 895NK
UT WOS:000300502800015
PM 21618421
OA Bronze
DA 2023-08-27
ER

PT J
AU Jallo, GI
   Penno, M
   Sukay, L
   Liu, JY
   Tyler, B
   Lee, J
   Carson, BS
   Guarnieri, M
AF Jallo, GI
   Penno, M
   Sukay, L
   Liu, JY
   Tyler, B
   Lee, J
   Carson, BS
   Guarnieri, M
TI Experimental models of brainstem tumors: development of a neonatal rat
   model
SO CHILDS NERVOUS SYSTEM
LA English
DT Article
DE neonatal; brainstem tumor; model; animal
ID CONVECTION-ENHANCED DELIVERY; MICE
AB Objective: Brainstem tumor models are required to advance the treatment for diffuse pontine gliomas in children. The feasibility of creating an experimental rodent model by inoculating newborn pups with tumor cells was examined. The study was performed to create an animal model for diffuse brainstem tumors. Methods: Eighty-two Fischer rat pups aged 12-24 h were anesthetized by hypothermia. The brainstem was injected with saline to identify anatomical coordinates for subsequent tumor cell challenges. The newborn pups were then inoculated with F98 (n=30) or 9L (n=30) glioma cells. Animals were returned to their mother for nursing. Tumor growth was assessed by survival and histopathology. Results: Twenty-one percent of the saline-treated animals (17 out of 82) and 5% of the tumor cell-challenged pups (3 out of 60) were eliminated by their mothers. Inoculations with 9L and F98 cells produced brainstem tumors in 83% (24 out of 29) and 93% of animals (26 out of 28) respectively that were evaluated. Conclusions: Our results demonstrate that neonatal rat models for brainstem tumors can be prepared using known injection coordinates and orthotopic cell lines. Decreasing rates of maternal removal during the course of the work suggests that the method involves a learning curve.
C1 Johns Hopkins Univ Hosp, Div Pediat Neurosurg, Baltimore, MD 21287 USA.
   Johns Hopkins Univ, Hunterian Brain Tumor Labs, Baltimore, MD 21287 USA.
C3 Johns Hopkins University; Johns Hopkins Medicine; Johns Hopkins
   University
RP Jallo, GI (通讯作者)，Johns Hopkins Univ Hosp, Div Pediat Neurosurg, 600 N Wolfe St,HArvey 811, Baltimore, MD 21287 USA.
EM
CR Abbott R, 1999, PRINCIPLES AND PRACTICE OF PEDIATRIC NEUROSURGERY, P629
   CARSON BS, 2002, ADV CLIN NEUROSCI, V12, P89
   Fisher PG, 1996, CONT NEUROSURG, V18, P1
   Groothuis DR, 2000, NEURO-ONCOLOGY, V2, P45, DOI 10.1093/neuonc/2.1.45
   Klopp LS, 2000, VET SURG, V29, P533, DOI 10.1053/jvet.2000.17851
   Occhiogrosso G, 2003, NEUROSURGERY, V52, P388, DOI 10.1227/01.NEU.0000043696.83722.8D
   Paxinos G., 1986, RAT BRAIN STEREOTAXI, Vsecond
   PETERSON DL, 1994, J NEUROSURG, V80, P865, DOI 10.3171/jns.1994.80.5.0865
   Sandberg DI, 2002, J NEUROSURG, V96, P885, DOI 10.3171/jns.2002.96.5.0885
   TAMARGO RJ, 1988, J NEUROSURG, V69, P928, DOI 10.3171/jns.1988.69.6.0928
   Wetmore C, 2001, CANCER RES, V61, P513
   Wu Q, 2002, VET PATHOL, V39, P293, DOI 10.1354/vp.39-3-293
   Xiao A, 2002, CANCER CELL, V1, P157, DOI 10.1016/S1535-6108(02)00029-6
NR 13
TC 20
Z9 23
U1 0
U2 2
PU SPRINGER
PI NEW YORK
PA 233 SPRING STREET, NEW YORK, NY 10013 USA
SN 0256-7040
J9 CHILD NERV SYST
JI Childs Nerv. Syst.
PD MAY
PY 2005
VL 21
IS 5
BP 399
EP 403
DI 10.1007/s00381-004-1100-6
PG 5
WC Clinical Neurology; Pediatrics; Surgery
WE Science Citation Index Expanded (SCI-EXPANDED)
SC Neurosciences & Neurology; Pediatrics; Surgery
GA 920XL
UT WOS:000228728700011
PM 15702357
DA 2023-08-27
ER

PT J
AU Oka, H
   Utsuki, S
   Tanizaki, Y
   Hagiwara, H
   Miyajima, Y
   Sato, K
   Kusumi, M
   Kijima, C
   Fujii, K
AF Oka, Hidehiro
   Utsuki, Satoshi
   Tanizaki, Yoshinori
   Hagiwara, Hiroyuki
   Miyajima, Yoshiteru
   Sato, Kimitoshi
   Kusumi, Mari
   Kijima, Chihiro
   Fujii, Kiyotaka
TI Clinicopathological features of human brainstem gliomas
SO BRAIN TUMOR PATHOLOGY
LA English
DT Article
DE Astrocytoma; Brainstem glioma; IDH1; p53; MGMT
ID CODON 132 MUTATION; IDH1 MUTATIONS; EXPRESSION; EGFR; AMPLIFICATION;
   ASTROCYTOMAS; CHILDREN
AB We describe the clinicopathological features of 25 brainstem gliomas (BSGs). Twenty BSGs located in the pons and were all in children. Four BSGs located in the medulla oblongata were in 2 children and 2 adults. One (in a child) was located in the midbrain. Radiological findings on MR images were low-intensity on T1 weighted images and high-intensity on T2 weighted images. Mean survival when pontine glioma was treated by radiotherapy and/or use of temozolomide was 14 months, although 4 patients (3 cervicomedullary types and one focal type arising from midbrain) are alive. Follow up was from 5 months to 6 years. Histopathological features of 10 cases of the diffuse type were: 4 grade II astrocytomas, 4 grade III astrocytomas, and 2 glioblastomas. MIB-1 index was from 0.8 to 38 %. P53 was positive for 80 % of 15 tumors and there were no negative results. MGMT was positive in 60 % of 15 tumors and negative in 12.4 %. IDH1 was negative in 61.6 %. There was no positive result for IDH1 in this study. Thus, our histopathological results were indicative of high p53 immunoreactivity and no IDH1 immunoreactivity related to secondary malignant change.
C1 [Oka, Hidehiro; Utsuki, Satoshi; Tanizaki, Yoshinori; Hagiwara, Hiroyuki; Miyajima, Yoshiteru; Sato, Kimitoshi; Kusumi, Mari; Kijima, Chihiro; Fujii, Kiyotaka] Kitasato Univ, Sch Med, Dept Neurosurg, Minimi Ku, Sagamihara, Kanagawa 225318, Japan.
C3 Kitasato University
RP Oka, H (通讯作者)，Kitasato Univ, Sch Med, Dept Neurosurg, Minimi Ku, 1-15-1 Kitasato, Sagamihara, Kanagawa 225318, Japan.
EM
FU Japan Society for the Promotion of Science [19591700, 19590365];
   Parent's Association Grant from Kitasato University School of Medicine,
   Japan; Grants-in-Aid for Scientific Research [19591700, 19590365]
   Funding Source: KAKEN
FX This work was supported in part by Grants-in-Aid for Scientific Research
   from the Japan Society for the Promotion of Science (nos 19591700 and
   19590365) and a Parent's Association Grant from Kitasato University
   School of Medicine, Japan. We wish to thank Ms Yoshie Yasui for
   providing technical assistance.
CR Badhe Prerna B, 2004, Indian J Cancer, V41, P170
   Balss J, 2008, ACTA NEUROPATHOL, V116, P597, DOI 10.1007/s00401-008-0455-2
   Bleeker FE, 2009, HUM MUTAT, V30, P7, DOI 10.1002/humu.20937
   Cheng Y, 1999, HUM PATHOL, V30, P1284, DOI 10.1016/S0046-8177(99)90057-6
   Di Sapio A, 2002, J NEURO-ONCOL, V59, P117, DOI 10.1023/A:1019697117253
   Geoerger B, 2009, EUR J CANCER, V45, P2342, DOI 10.1016/j.ejca.2009.03.007
   Guillamo JS, 2001, BRAIN, V124, P2528, DOI 10.1093/brain/124.12.2528
   Hargrove D, 2006, LANCET ONCOL, V7, P241, DOI 10.1016/S1470-2045(06)70615-5
   Hartmann C, 2009, ACTA NEUROPATHOL, V118, P469, DOI 10.1007/s00401-009-0561-9
   Hegi ME, 2005, NEW ENGL J MED, V352, P997, DOI 10.1056/NEJMoa043331
   Ichimura K, 2009, NEURO-ONCOLOGY, V11, P341, DOI 10.1215/15228517-2009-025
   Leach PA, 2008, BRIT J NEUROSURG, V22, P619, DOI 10.1080/02688690802366198
   Liang ML, 2008, J NEURO-ONCOL, V87, P247, DOI 10.1007/s11060-007-9513-1
   LOUIS DN, 1993, J NEUROPATH EXP NEUR, V52, P507, DOI 10.1097/00005072-199309000-00009
   Nakamura M, 2007, NEURO-ONCOLOGY, V9, P113, DOI 10.1215/15228517-2006-036
   Nobusawa S, 2009, CLIN CANCER RES, V15, P6002, DOI 10.1158/1078-0432.CCR-09-0715
   Parsons DW, 2008, SCIENCE, V321, P1807, DOI 10.1126/science.1164382
   Puputti M, 2006, MOL CANCER RES, V4, P927, DOI 10.1158/1541-7786.MCR-06-0085
   Sanson M, 2009, J CLIN ONCOL, V27, P4150, DOI 10.1200/JCO.2009.21.9832
   Sung T, 2000, BRAIN PATHOL, V10, P249, DOI 10.1111/j.1750-3639.2000.tb00258.x
   Sure U, 1997, J NEUROPATH EXP NEUR, V56, P782
   Thorarinsdottir HK, 2008, CLIN CANCER RES, V14, P3386, DOI 10.1158/1078-0432.CCR-07-1616
   van den Bent MJ, 2009, J CLIN ONCOL, V27, P5881, DOI 10.1200/JCO.2009.24.1034
   Wakabayashi T, 2009, NEUROSURGERY, V64, P455, DOI 10.1227/01.NEU.0000340683.19920.E3
   Watanabe T, 2009, AM J PATHOL, V174, P1149, DOI 10.2353/ajpath.2009.080958
   Wick W, 2009, J CLIN ONCOL, V27, P5874, DOI 10.1200/JCO.2009.23.6497
   Wong KK, 2006, CANCER RES, V66, P11172, DOI 10.1158/0008-5472.CAN-06-2438
   Yan H, 2009, NEW ENGL J MED, V360, P765, DOI 10.1056/NEJMoa0808710
   ZHANG SJ, 1993, BIOCHEM BIOPH RES CO, V196, P851, DOI 10.1006/bbrc.1993.2327
NR 29
TC 12
Z9 12
U1 0
U2 2
PU SPRINGER JAPAN KK
PI TOKYO
PA SHIROYAMA TRUST TOWER 5F, 4-3-1 TORANOMON, MINATO-KU, TOKYO, 105-6005,
   JAPAN
SN 1433-7398
EI 1861-387X
J9 BRAIN TUMOR PATHOL
JI Brain Tumor Pathol.
PD JAN
PY 2013
VL 30
IS 1
BP 1
EP 7
DI 10.1007/s10014-012-0099-8
PG 7
WC Oncology; Clinical Neurology; Pathology
WE Science Citation Index Expanded (SCI-EXPANDED)
SC Oncology; Neurosciences & Neurology; Pathology
GA 077IR
UT WOS:000314021600001
PM 22484454
DA 2023-08-27
ER

PT J
AU Lee, S
   Kambhampati, M
   Yadavilli, S
   Gordish-Dressman, H
   Santi, M
   Cruz, CR
   Packer, RJ
   Almira-Suarez, MI
   Hwang, EI
   Nazarian, J
AF Lee, Sulgi
   Kambhampati, Madhuri
   Yadavilli, Sridevi
   Gordish-Dressman, Heather
   Santi, Mariarita
   Cruz, Conrad R.
   Packer, Roger J.
   Almira-Suarez, M. Isabel
   Hwang, Eugene I.
   Nazarian, Javad
TI Differential Expression of Wilms' Tumor Protein in Diffuse Intrinsic
   Pontine Glioma
SO JOURNAL OF NEUROPATHOLOGY AND EXPERIMENTAL NEUROLOGY
LA English
DT Article
DE Brainstem glioma; Childhood midline glioma; Diffuse intrinsic pontine
   glioma (DIPG); Wilms tumor protein (WT1)
ID PEDIATRIC HIGH-GRADE; SUPPRESSOR GENE; WT1 PROTEIN; GLIOBLASTOMA;
   MUTATIONS; SUBGROUPS; CELLS
AB Diffuse intrinsic pontine gliomas (DIPGs) are deadly tumors comprising 10%-15% of all childhood CNS cancers. Standard treatment is considered palliative and prognosis is near universal mortality. DIPGs have been classified into genomic subtypes based on histone variants with the lysine to methionine mutation on position 27 of histone tails (K27M). Given the increasing promise of immunotherapy, there have been ongoing efforts to identify tumor-specific antigens to serve as immunologic targets. We evaluated a large cohort of CNS specimens for Wilms' tumor protein (WT1) expression. These specimens include primary pediatric CNS tumors (n = 38 midline gliomas and n = 3 non-midline gliomas; n = 23 DIPG, n = 10 low-grade gliomas, n = 8 high-grade gliomas), and DIPG primary cells. Here, we report the validation of WT1 as a tumor-associated antigen in DIPGs. We further report that WT1 expression is significantly correlated with specific oncohistone variants, with the highest expression detected in the H3.3K27M subgroup. WT1 expression was absent in all control specimens (n = 21). Western blot assays using DIPG primary cells (n = 6) showed a trend of higher WT1 expression in H3.3K27M cells when compared with H3.1 K27M cells and H3 wildtype cells. Our data are the first indication of the association between WT1 and DIPG, with specific upregulation in those harboring oncohistone H3.3K27M.
C1 [Lee, Sulgi; Kambhampati, Madhuri; Yadavilli, Sridevi; Gordish-Dressman, Heather; Nazarian, Javad] Childrens Natl Hlth Syst, Med Genet Res Ctr, 111 Michigan Ave NW, Washington, DC 20010 USA.
   [Lee, Sulgi; Nazarian, Javad] George Washington Univ, Sch Med & Hlth Sci, Inst Biomed Sci, Washington, DC 20052 USA.
   [Santi, Mariarita] Childrens Hosp Philadelphia, Dept Pathol, Philadelphia, PA 19104 USA.
   [Cruz, Conrad R.] Childrens Natl Hlth Syst, Ctr Canc & Immunol Res, Washington, DC USA.
   [Packer, Roger J.; Hwang, Eugene I.; Nazarian, Javad] Childrens Natl Hlth Syst, Brain Tumor Inst, Washington, DC USA.
   [Almira-Suarez, M. Isabel] Childrens Natl Hlth Syst, Dept Pathol & Lab Med, Washington, DC USA.
   [Nazarian, Javad] George Washington Univ, Sch Med & Hlth Sci, Dept Genom & Precis Med, Washington, DC 20052 USA.
C3 Children's National Health System; George Washington University;
   University of Pennsylvania; Pennsylvania Medicine; Childrens Hospital of
   Philadelphia; Children's National Health System; Children's National
   Health System; Children's National Health System; George Washington
   University
RP Nazarian, J (通讯作者)，Childrens Natl Hlth Syst, Med Genet Res Ctr, 111 Michigan Ave NW, Washington, DC 20010 USA.
EM
OI Nazarian, Javad/0000-0002-1951-9828
FU Smashing Walnuts; DIPG Collaborative; Goldwin Foundation; Cure Starts
   Now Foundation; CNHS CTSI; Zickler Family Foundation; Childhood Brain
   Tumor Foundation; Isabella Kerr Molina Foundation; Al Musella
   Foundation; Brain Tumor Foundation for Children; Kisses 4 Kayla; Matthew
   Larson Foundation; Brad Kaminsky Foundation; iDance4aCURE; Rally
   Foundation
FX Smashing Walnuts, DIPG Collaborative, The Goldwin Foundation, The Cure
   Starts Now Foundation, CNHS CTSI, Zickler Family Foundation, The
   Childhood Brain Tumor Foundation, Isabella Kerr Molina Foundation, Al
   Musella Foundation, Brain Tumor Foundation for Children, Kisses 4 Kayla,
   Matthew Larson Foundation, The Brad Kaminsky Foundation, iDance4aCURE,
   and Rally Foundation.
CR Ahmed N, 2017, JAMA ONCOL, V3, P1094, DOI 10.1001/jamaoncol.2017.0184
   Barbaux S, 1997, NAT GENET, V17, P467, DOI 10.1038/ng1297-467
   Bender S, 2013, CANCER CELL, V24, P660, DOI 10.1016/j.ccr.2013.10.006
   Bourne TD, 2010, BRAIN PATHOL, V20, P1090, DOI 10.1111/j.1750-3639.2010.00415.x
   Brennan CW, 2013, CELL, V155, P462, DOI 10.1016/j.cell.2013.09.034
   Buczkowicz P, 2014, NAT GENET, V46, P451, DOI 10.1038/ng.2936
   Chassot A, 2012, J NEURO-ONCOL, V106, P399, DOI 10.1007/s11060-011-0681-7
   Cheever MA, 2009, CLIN CANCER RES, V15, P5323, DOI 10.1158/1078-0432.CCR-09-0737
   Chiba Y, 2010, BRAIN TUMOR PATHOL, V27, P29, DOI 10.1007/s10014-010-0265-9
   Cohen KJ, 2011, NEURO-ONCOLOGY, V13, P410, DOI 10.1093/neuonc/noq205
   Griesinger AM, 2013, J IMMUNOL, V191, P4880, DOI 10.4049/jimmunol.1301966
   HABER DA, 1992, NEW BIOL, V4, P97
   Hastie ND, 2017, DEVELOPMENT, V144, P2862, DOI 10.1242/dev.153163
   Hohenstein P, 2006, HUM MOL GENET, V15, pR196, DOI 10.1093/hmg/ddl196
   Kambhampati M, 2015, ONCOTARGET, V6, P12740, DOI 10.18632/oncotarget.3374
   Kijima N, 2014, ANTICANCER RES, V34, P61
   Mackay A, 2018, CANCER CELL, V33, P829, DOI 10.1016/j.ccell.2018.04.004
   Mackay A, 2017, CANCER CELL, V32, P520, DOI 10.1016/j.ccell.2017.08.017
   Nakatsuka S, 2006, MODERN PATHOL, V19, P804, DOI 10.1038/modpathol.3800588
   Nikbakht H, 2016, NAT COMMUN, V7, DOI 10.1038/ncomms11185
   Niksic M, 2004, HUM MOL GENET, V13, P463, DOI 10.1093/hmg/ddh040
   O'Rourke DM, 2017, SCI TRANSL MED, V9, DOI 10.1126/scitranslmed.aaa0984
   Oji Y, 2004, CANCER SCI, V95, P822, DOI 10.1111/j.1349-7006.2004.tb02188.x
   Oji Y, 2016, INT J CANCER, V139, P1391, DOI 10.1002/ijc.30182
   Paugh BS, 2010, J CLIN ONCOL, V28, P3061, DOI 10.1200/JCO.2009.26.7252
   PELLETIER J, 1991, CELL, V67, P437, DOI 10.1016/0092-8674(91)90194-4
   Pollack IF, 2016, J NEURO-ONCOL, V130, P517, DOI 10.1007/s11060-016-2245-3
   Qi XW, 2015, SCI REP-UK, V5, DOI 10.1038/srep08924
   Ramos A, 2013, SEMIN ULTRASOUND CT, V34, P104, DOI 10.1053/j.sult.2013.01.001
   Saratsis AM, 2014, ACTA NEUROPATHOL, V127, P881, DOI 10.1007/s00401-013-1218-2
   Saratsis AM, 2012, NEURO-ONCOLOGY, V14, P547, DOI 10.1093/neuonc/nos067
   Schittenhelm J, 2008, BRAIN PATHOL, V18, P344, DOI 10.1111/j.1750-3639.2008.00127.x
   Schwartzentruber J, 2012, NATURE, V482, P226, DOI [10.1038/nature10833, 10.1007/s13311-012-0137-6]
   Sturm D, 2012, CANCER CELL, V22, P425, DOI 10.1016/j.ccr.2012.08.024
   Sugiyama H, 2010, JPN J CLIN ONCOL, V40, P377, DOI 10.1093/jjco/hyp194
   Wildes TJ, 2018, CLIN CANCER RES, V24, P3955, DOI 10.1158/1078-0432.CCR-17-3061
   Wu G, 2012, NAT GENET, V44, P251, DOI 10.1038/ng.1102
   Yadavilli S, 2015, ONCOTARGET, V6, P12141, DOI 10.18632/oncotarget.3716
NR 38
TC 4
Z9 4
U1 0
U2 3
PU OXFORD UNIV PRESS INC
PI CARY
PA JOURNALS DEPT, 2001 EVANS RD, CARY, NC 27513 USA
SN 0022-3069
EI 1554-6578
J9 J NEUROPATH EXP NEUR
JI J. Neuropathol. Exp. Neurol.
PD MAY
PY 2019
VL 78
IS 5
BP 380
EP 388
DI 10.1093/jnen/nlz021
PG 9
WC Clinical Neurology; Neurosciences; Pathology
WE Science Citation Index Expanded (SCI-EXPANDED)
SC Neurosciences & Neurology; Pathology
GA IF7KX
UT WOS:000473264100001
PM 30990879
OA Green Submitted, hybrid, Green Published
DA 2023-08-27
ER

PT J
AU Kobayashi, H
   Sawamura, Y
   Ikeda, J
AF Kobayashi, H
   Sawamura, Y
   Ikeda, J
TI A tumor in the medulla oblongata producing beta-HCG and AFP
SO JOURNAL OF CLINICAL NEUROSCIENCE
LA English
DT Article
DE tumor marker; germ cell tumor; medulla oblongata; chemotherapy;
   radiotherapy
ID GERM-CELL TUMORS; CENTRAL-NERVOUS-SYSTEM; SPINAL-CORD GERMINOMA; SEPTUM
   PELLUCIDUM; PINEAL REGION
AB A 23 year-old woman presented with dysarthria, hoarseness, dysphasia, ataxia and dyspnoea. MRI showed a mass of heterogeneous intensity at the dorsal medulla oblongata. Laboratory studies revealed high serum alpha-fetoprotein (AFP) and beta-subunit human chorionic gonadotropin (beta-HCG) levels. No other tumours were found on systemic investigation. An intracranial nongerminomatous germ cell tumour (NGGCT) was strongly suspected. The patient received combination chemotherapy using ifosfamide, cisplatin, and etoposide and local irradiation to a total of 52 Gy. Serum AFP and beta-HCG levels normalized after four cycles of chemotherapy and she became asymptomatic apart from mild postural hypotension. A follow-up MRI showed only a tiny residual lesion in the medulla oblongata, which has been stable for more than three years. Surgical resection should be carefully considered in patients with brainstem tumours with elevation of serum tumour markers as chemo- and radiotherapy may be effective for brainstem NGGCT. (C) 2005 Elsevier Ltd. All rights reserved.
C1 Hokkaido Univ, Sch Med, Dept Neurosurg, Kita Ku, Sapporo, Hokkaido 0608638, Japan.
C3 Hokkaido University
RP Kobayashi, H (通讯作者)，Hokkaido Univ, Sch Med, Dept Neurosurg, Kita Ku, North 15,West 7, Sapporo, Hokkaido 0608638, Japan.
EM
RI KOBAYASHI, Hiroyuki/G-8990-2012
CR Balmaceda C, 1996, J CLIN ONCOL, V14, P2908, DOI 10.1200/JCO.1996.14.11.2908
   BRUCE JN, 1995, ACTA NEUROCHIR, V134, P130, DOI 10.1007/BF01417679
   CALAMINUS G, 1994, NEUROPEDIATRICS, V25, P26, DOI 10.1055/s-2008-1071577
   HASHIMOTO M, 1992, SURG NEUROL, V37, P384, DOI 10.1016/0090-3019(92)90009-C
   KAMIYA K, 1988, SURG NEUROL, V30, P462, DOI 10.1016/0090-3019(88)90032-8
   Kiyuna M, 1999, PATHOL INT, V49, P633, DOI 10.1046/j.1440-1827.1999.00878.x
   KOYAMA S, 1991, SURG NEUROL, V35, P478, DOI 10.1016/0090-3019(91)90184-B
   MATSUOKA S, 1991, SURG NEUROL, V35, P122, DOI 10.1016/0090-3019(91)90262-8
   Matsutani M, 1997, J NEUROSURG, V86, P446, DOI 10.3171/jns.1997.86.3.0446
   MATSUTANI M, 1998, NEUROSURG FOCUS, V5
   Miyauchi A, 1996, J NEUROSURG, V84, P1060, DOI 10.3171/jns.1996.84.6.1060
   Nakajima H, 2000, SURG NEUROL, V53, P448, DOI 10.1016/S0090-3019(00)00224-X
   Sawamura Y, 1998, J NEUROSURG, V89, P728, DOI 10.3171/jns.1998.89.5.0728
   Sawamura Y, 1998, INTRACRANIAL GERM CELL TUMORS, P169
   Sawamura Y, 1998, INTRACRANIAL GERM CELL TUMORS, P155
   Sawamura Y, 1998, EUR J CANCER, V34, P104, DOI 10.1016/S0959-8049(97)10045-4
   Sawamura Y, 1996, CURR OPIN NEUROL, V9, P419, DOI 10.1097/00019052-199612000-00004
   Sugiyama K, 1994, Neurol Med Chir (Tokyo), V34, P291, DOI 10.2176/nmc.34.291
   Tada M, 1998, INTRACRANIAL GERM CELL TUMORS, P146
   TANAKA R, 1979, SURG NEUROL, V12, P239
   Tashiro Takashi, 1993, Neurologia Medico-Chirurgica, V33, P251, DOI 10.2176/nmc.33.251
   Ushio Y, 1999, J NEUROSURG, V90, P133, DOI 10.3171/jns.1999.90.1.0133
   Yen PS, 2003, J NEURO-ONCOL, V62, P339, DOI 10.1023/A:1023331700995
NR 23
TC 1
Z9 1
U1 0
U2 0
PU ELSEVIER SCI LTD
PI OXFORD
PA THE BOULEVARD, LANGFORD LANE, KIDLINGTON, OXFORD OX5 1GB, OXON, ENGLAND
SN 0967-5868
EI 1532-2653
J9 J CLIN NEUROSCI
JI J. Clin. Neurosci.
PD AUG
PY 2005
VL 12
IS 6
BP 709
EP 711
DI 10.1016/j.jocn.2004.05.023
PG 3
WC Clinical Neurology; Neurosciences
WE Science Citation Index Expanded (SCI-EXPANDED)
SC Neurosciences & Neurology
GA 016TB
UT WOS:000235642200026
PM 16098752
DA 2023-08-27
ER

PT J
AU Muzumdar, DP
   Goel, A
   Desai, KI
AF Muzumdar, DP
   Goel, A
   Desai, KI
TI Pontine glioma and cerebellopontine angle epidermoid tumour occurring as
   collision tumours
SO BRITISH JOURNAL OF NEUROSURGERY
LA English
DT Article
DE brainstem glioma; cerebellopontine angle; epidermoid tumour
AB A 36-year-old male was admitted with a 10-year history of gradually progressive left ear hearing loss, diplopia and right hemiparesis. Magnetic resonance imaging showed a left lateral pontine enhancing tumour and an additional cerebellopontine angle epidermoid tumour in close proximity forming a collision tumour. On exploration, the epidermoid tumour and the intraaxial pontine glioma were completely resected. The literature on collision tumours is reviewed.
C1 King Edward Mem Hosp, Dept Neurosurg, Parel 400012, Mumbai, India.
   Seth GS Med Coll, Parel 400012, Mumbai, India.
C3 Seth Gordhandas Sunderdas Medical College & King Edward Memorial
   Hospital; Seth Gordhandas Sunderdas Medical College & King Edward
   Memorial Hospital
RP Goel, A (通讯作者)，King Edward Mem Hosp, Dept Neurosurg, Parel 400012, Mumbai, India.
CR BILANIUK LT, 1980, RADIOLOGY, V134, P89, DOI 10.1148/radiology.134.1.7350640
   Dubois P., 1985, NEUROSURGERY, P704
   GOODMAN RR, 1991, NEUROSURGERY, V28, P433, DOI 10.1227/00006123-199103000-00017
   KITAOKA K, 1986, Neurological Surgery, V14, P1243
   NETSKY MG, 1988, SURG NEUROL, V29, P477, DOI 10.1016/0090-3019(88)90144-9
   TOLGLIA JU, 1965, J NEUROSURG, V23, P384
   ULRICH J, 1964, J NEUROSURG, V21, P1051, DOI 10.3171/jns.1964.21.12.1051
   WHITE HH, 1963, NEUROLOGY, V13, P292, DOI 10.1212/WNL.13.4.292
NR 8
TC 17
Z9 17
U1 0
U2 0
PU CARFAX PUBLISHING
PI BASINGSTOKE
PA RANKINE RD, BASINGSTOKE RG24 8PR, HANTS, ENGLAND
SN 0268-8697
J9 BRIT J NEUROSURG
JI Br. J. Neurosurg.
PD FEB
PY 2001
VL 15
IS 1
BP 68
EP 71
PG 5
WC Clinical Neurology; Surgery
WE Science Citation Index Expanded (SCI-EXPANDED)
SC Neurosciences & Neurology; Surgery
GA 406GP
UT WOS:000167206800016
PM 11303668
DA 2023-08-27
ER

PT J
AU Schild, SE
   Stafford, SL
   Brown, PD
   Wood, CP
   Scheithauer, BW
   Schomberg, PJ
   Wong, WW
   Lyons, MK
   Shaw, EG
AF Schild, SE
   Stafford, SL
   Brown, PD
   Wood, CP
   Scheithauer, BW
   Schomberg, PJ
   Wong, WW
   Lyons, MK
   Shaw, EG
TI The results of radiotherapy for brainstem tumors
SO JOURNAL OF NEURO-ONCOLOGY
LA English
DT Article
DE brainstem tumors; surgery; radiation therapy; chemotherapy
ID HYPERFRACTIONATED RADIATION-THERAPY; PEDIATRIC-ONCOLOGY-GROUP; LONG-TERM
   SURVIVAL; STEM GLIOMAS; RANDOMIZED TRIAL; MIDBRAIN TUMORS; CHILDREN;
   CHILDHOOD; CHEMOTHERAPY; IRRADIATION
AB Objective: This analysis was performed to examine the outcome of adult and pediatric patients with brainstem tumors. Methods and materials: Forty patients with brainstem glioma were evaluated retrospectively. Included were 24 females and 16 males ranging in age from 3 to 81. years (median, 29.5 years). These patients were treated with various combinations of surgery, chemotherapy, and radiotherapy (RT). The length of follow-up in survivors ranged from 0.6 to 20 years (median: 3.2 years, mean: 6 years). Survival rates were calculated with the Kaplan Meier method and differences between survival curves were calculated using the log-rank test. Results: The overall 2 and 5-year survival rates were 44% and 34%,respectively. The median survival time was 19 months. The 5-year survival rate was 54% for patients with tumors outside the pens compared to 21% for those with tumors involving the pens (p = 0.04). The 5-year survival rate was 59% for patients with exophytic tumors as compared to 23% for those with intrinsic tumors (p = 0.05). Patients undergoing subtotal resection had a 5-year survival rate of 53% compared to 28% for those having only a biopsy or no surgical intervention (p = 0.04). None of the other potential prognostic or treatment related factors evaluated [patient age, tumor grade, tumor histology, radiotherapy parameters (including BID fractionation, 3-D treatment planning, or the use of doses > 55 Gy), or the administration of adjuvant chemotherapy] evaluated were associated with patient survival. Conclusions: Brainstem gliomas generally occur in younger individuals. The survival rates were better for patients with exophytic tumors, those involving sites other than the pens, and tumors amenable to subtotal resection. Improvements in the outcome of patients with brainstem gliomas will require new therapeutic approaches.
C1 Mayo Clin Scottsdale, Dept Radiat Oncol, Scottsdale, AZ 85259 USA.
   Mayo Clin Scottsdale, Dept Neurosurg, Scottsdale, AZ 85259 USA.
   Mayo Clin Rochester, Dept Radiat Oncol, Rochester, MN USA.
   Mayo Clin Rochester, Dept Radiol, Rochester, MN USA.
   Mayo Clin Rochester, Dept Pathol, Rochester, MN USA.
   Wake Forest Univ, Bowman Gray Sch Med, Winston Salem, NC USA.
C3 Mayo Clinic; Mayo Clinic Phoenix; Mayo Clinic; Mayo Clinic Phoenix; Mayo
   Clinic; Mayo Clinic; Mayo Clinic; Wake Forest University; Wake Forest
   Baptist Medical Center
RP Schild, SE (通讯作者)，Mayo Clin Scottsdale, Dept Radiat Oncol, 13400 E Shea Blvd, Scottsdale, AZ 85259 USA.
EM
OI schild, steven/0000-0002-2850-4126
CR ALBRIGHT AL, 1983, CANCER, V52, P2313, DOI 10.1002/1097-0142(19831215)52:12<2313::AID-CNCR2820521226>3.0.CO;2-I
   BOYDSTON WR, 1992, PEDIATR NEUROSURG, V17, P234, DOI 10.1159/000120603
   EDWARDS MSB, 1994, J NEUROSURG, V80, P20, DOI 10.3171/jns.1994.80.1.0020
   EIFEL PJ, 1987, INT J RADIAT ONCOL, V13, P847, DOI 10.1016/0360-3016(87)90098-8
   EPSTEIN F, 1987, J NEUROSURG, V67, P483, DOI 10.3171/jns.1987.67.4.0483
   FREEMAN CR, 1986, INT J RADIAT ONCOL, V12, P1823, DOI 10.1016/0360-3016(86)90325-1
   Freeman CR, 1996, PEDIATR NEUROSURG, V24, P103, DOI 10.1159/000121025
   FREEMAN CR, 1995, INT J RADIAT ONCOL, V32, P549, DOI 10.1016/0360-3016(95)00141-K
   HALPERIN EC, 1985, INT J RADIAT ONCOL, V11, P1293, DOI 10.1016/0360-3016(85)90244-5
   HOFFMAN HJ, 1980, NEUROSURGERY, V7, P243, DOI 10.1227/00006123-198009000-00007
   JENKIN D, 1995, INT J RADIAT ONCOL, V31, P445, DOI 10.1016/0360-3016(94)00393-Y
   Jenkin D, 1996, Oncology (Williston Park), V10, P715
   JENKIN RDT, 1987, J NEUROSURG, V66, P227, DOI 10.3171/jns.1987.66.2.0227
   KHATIB ZA, 1994, PEDIATR NEUROSURG, V20, P2, DOI 10.1159/000120759
   KIM TH, 1980, INT J RADIAT ONCOL, V6, P51, DOI 10.1016/0360-3016(80)90203-5
   KRETSCHMAR CS, 1993, CANCER-AM CANCER SOC, V72, P1404, DOI 10.1002/1097-0142(19930815)72:4<1404::AID-CNCR2820720441>3.0.CO;2-G
   KUN LE, 1995, MOSS RAD ONCOLOGY RA, pCH29
   LINSTADT DE, 1991, INT J RADIAT ONCOL, V20, P757, DOI 10.1016/0360-3016(91)90019-Z
   Mandell L, 1997, INT J RADIAT ONCOL, V39, P143, DOI 10.1016/S0360-3016(97)80575-5
   MANTRAVADI RVP, 1982, CANCER, V49, P1294, DOI 10.1002/1097-0142(19820315)49:6<1294::AID-CNCR2820490636>3.0.CO;2-V
   POLLACK IF, 1993, J NEUROSURG, V78, P859, DOI 10.3171/jns.1993.78.6.0859
   PRADOS MD, 1995, INT J RADIAT ONCOL, V32, P85, DOI 10.1016/0360-3016(95)00563-E
   ROBERTSON PL, 1995, PEDIATR NEUROSURG, V22, P65, DOI 10.1159/000120879
   Shaw E G, 1993, Oncology (Williston Park), V7, P97
   SHELINE GE, 1977, CANCER, V39, P873, DOI 10.1002/1097-0142(197702)39:2+<873::AID-CNCR2820390725>3.0.CO;2-Y
   SHELINE GE, 1975, CANCER, V35, P957, DOI 10.1002/1097-0142(197503)35:3+<957::AID-CNCR2820350717>3.0.CO;2-Y
   SPOSTO R, 1989, J NEURO-ONCOL, V7, P165, DOI 10.1007/BF00165101
   STROINK AR, 1987, NEUROSURGERY, V20, P439, DOI 10.1227/00006123-198703000-00014
   STROINK AR, 1986, J NEUROSURG, V65, P745, DOI 10.3171/jns.1986.65.6.0745
   VANDERTOP WP, 1992, NEUROSURGERY, V31, P186, DOI 10.1227/00006123-199208000-00003
NR 30
TC 15
Z9 15
U1 0
U2 0
PU SPRINGER
PI NEW YORK
PA 233 SPRING ST, NEW YORK, NY 10013 USA
SN 0167-594X
EI 1573-7373
J9 J NEURO-ONCOL
JI J. Neuro-Oncol.
PD NOV
PY 1998
VL 40
IS 2
BP 171
EP 177
DI 10.1023/A:1006193306286
PG 7
WC Oncology; Clinical Neurology
WE Science Citation Index Expanded (SCI-EXPANDED)
SC Oncology; Neurosciences & Neurology
GA 156FV
UT WOS:000077991700009
PM 9892099
DA 2023-08-27
ER

PT J
AU Hashizume, R
   Ozawa, T
   Dinca, EB
   Banerjee, A
   Prados, MD
   James, CD
   Gupta, N
AF Hashizume, Rintaro
   Ozawa, Tomoko
   Dinca, Eduard B.
   Banerjee, Anuradha
   Prados, Michael D.
   James, Charles D.
   Gupta, Nalin
TI A human brainstem glioma xenograft model enabled for bioluminescence
   imaging
SO JOURNAL OF NEURO-ONCOLOGY
LA English
DT Article
DE Brainstem tumor; Animal model; Bioluminescence; Temozolomide
ID HUMAN GLIOBLASTOMA XENOGRAFTS; TELOMERASE INHIBITOR GRN163; CELLS
   IN-VITRO; TUMOR-MODEL; DELIVERY; VIVO; THERAPY; GROWTH; CANCER;
   TEMOZOLOMIDE
AB Despite the use of radiation and chemotherapy, the prognosis for children with diffuse brainstem gliomas is extremely poor. There is a need for relevant brainstem tumor models that can be used to test new therapeutic agents and delivery systems in pre-clinical studies. We report the development of a brainstem-tumor model in rats and the application of bioluminescence imaging (BLI) for monitoring tumor growth and response to therapy as part of this model. Luciferase-modified human glioblastoma cells from five different tumor cell sources (either cell lines or serially-passaged xenografts) were implanted into the pontine tegmentum of athymic rats using an implantable guide-screw system. Tumor growth was monitored by BLI and tumor volume was calculated by three-dimensional measurements from serial histopathologic sections. To evaluate if this model would allow detection of therapeutic response, rats bearing brainstem U-87 MG or GS2 glioblastoma xenografts were treated with the DNA methylating agent temozolomide (TMZ). For each of the tumor cell sources tested, BLI monitoring revealed progressive tumor growth in all animals, and symptoms caused by tumor burden were evident 26-29 days after implantation of U-87 MG, U-251 MG, GBM6, and GBM14 cells, and 37-47 days after implantation of GS2 cells. Histopathologic analysis revealed tumor growth within the pons in all rats and BLI correlated quantitatively with tumor volume. Variable infiltration was evident among the different tumors, with GS2 tumor cells exhibiting the greatest degree of infiltration. TMZ treatment groups were included for experiments involving U-87 MG and GS2 cells, and in each case TMZ delayed tumor growth, as indicated by BLI monitoring, and significantly extended survival of animal subjects. Our results demonstrate the development of a brainstem tumor model in athymic rats, in which tumor growth and response to therapy can be accurately monitored by BLI. This model is well suited for pre-clinical testing of therapeutics that are being considered for treatment of patients with brainstem tumors.
C1 [Hashizume, Rintaro; Ozawa, Tomoko; Dinca, Eduard B.; Prados, Michael D.; James, Charles D.; Gupta, Nalin] Univ Calif San Francisco, Dept Neurol Surg, Brain Tumor Res Ctr, San Francisco, CA 94143 USA.
   [Banerjee, Anuradha] Univ Calif San Francisco, Div Pediat Oncol, Dept Pediat, San Francisco, CA 94143 USA.
C3 University of California System; University of California San Francisco;
   University of California System; University of California San Francisco
RP Gupta, N (通讯作者)，Univ Calif San Francisco, Dept Neurol Surg, Brain Tumor Res Ctr, 505 Parnassus Ave,Room M779, San Francisco, CA 94143 USA.
EM
RI Prados, Michael/N-5220-2019; James, Charles D/E-2721-2012
OI James, Charles D/0000-0002-1027-203X
FU Pediatric Brain Tumor Foundation; Brain Tumor SPORE at UCSF; Timmy's
   Rainbow Foundation; National Institution of Health [RO1 CA107268];
   NIH/NINDS [K08 NS055061]
FX This work was supported by the Pediatric Brain Tumor Foundation
   Pre-Institute Award, a Career Development Award to AB) provided by the
   Brain Tumor SPORE at UCSF, Timmy's Rainbow Foundation for Brainstem
   Tumor Research, and National Institution of Health RO1 CA107268. NG is
   supported by K08 NS055061 NIH/NINDS). The authors thank Lily J. Hu for
   technical assistance and Ilona Garner for editorial assistance.
CR Avgeropoulos N G, 1999, Oncologist, V4, P209
   Broniscer A, 2005, CANCER-AM CANCER SOC, V103, P133, DOI 10.1002/cncr.20741
   Carson BS, 2002, J NEURO-ONCOL, V60, P151, DOI 10.1023/A:1020626419269
   Dinca EB, 2007, J NEUROSURG, V107, P610, DOI 10.3171/JNS-07/09/0610
   Edinger M, 2002, EUR J CANCER, V38, P2128, DOI 10.1016/S0959-8049(02)00410-0
   Finlay JL, 2005, J NEURO-ONCOL, V75, P253, DOI 10.1007/s11060-005-6747-7
   Giannini C, 2005, NEURO-ONCOLOGY, V7, P164, DOI 10.1215/S1152851704000821
   Gilbertson RJ, 2003, CLIN CANCER RES, V9, P3620
   Gross S, 2005, CANCER CELL, V7, P5, DOI 10.1016/j.ccr.2004.12.011
   Gunther HS, 2008, ONCOGENE, V27, P2897, DOI 10.1038/sj.onc.1210949
   Hasegawa K, 2006, CLIN CANCER RES, V12, P1868, DOI 10.1158/1078-0432.CCR-05-1803
   Hashizume R, 2008, NEURO-ONCOLOGY, V10, P112, DOI 10.1215/15228517-2007-052
   Huynh GH, 2006, J CONTROL RELEASE, V110, P236, DOI 10.1016/j.jconrel.2005.09.053
   Jallo GI, 2006, CHILD NERV SYST, V22, P1519, DOI 10.1007/s00381-006-0174-8
   Jallo GI, 2005, CHILD NERV SYST, V21, P399, DOI 10.1007/s00381-004-1100-6
   Kang SH, 2006, CANCER RES, V66, P11991, DOI 10.1158/0008-5472.CAN-06-1320
   Lee James, 2005, Neurosurg Focus, V18, pE11
   Ozawa T, 2004, NEURO-ONCOLOGY, V6, P218, DOI 10.1215/S1152851704000055
   Ozawa T, 2002, IN VIVO, V16, P55
   Ozawa T, 2006, INT J RADIAT ONCOL, V66, P263, DOI 10.1016/j.ijrobp.2006.05.010
   Ozawa T, 2004, MOL PHARMACEUT, V1, P368, DOI 10.1021/mp049933i
   Oztopcu P, 2004, ACTA NEUROL BELG, V104, P106
   Recinos PF, 2007, PEDIATR NEUROSURG, V43, P192, DOI 10.1159/000098831
   Rehemtulla A, 2000, NEOPLASIA, V2, P491, DOI 10.1038/sj.neo.7900121
   Sarkaria JN, 2007, MOL CANCER THER, V6, P1167, DOI 10.1158/1535-7163.MCT-06-0691
   Thorne RG, 2004, NEUROSCIENCE, V127, P481, DOI 10.1016/j.neuroscience.2004.05.029
   TOKUDA K, 1988, CANCER RES, V48, P3100
   Wu Q, 2002, VET PATHOL, V39, P293, DOI 10.1354/vp.39-3-293
   Yaz G, 2004, AM J CLIN ONCOL-CANC, V27, P384, DOI 10.1097/01.COC.0000071945.15623.C6
NR 29
TC 47
Z9 52
U1 0
U2 6
PU SPRINGER
PI NEW YORK
PA 233 SPRING ST, NEW YORK, NY 10013 USA
SN 0167-594X
EI 1573-7373
J9 J NEURO-ONCOL
JI J. Neuro-Oncol.
PD JAN
PY 2010
VL 96
IS 2
BP 151
EP 159
DI 10.1007/s11060-009-9954-9
PG 9
WC Oncology; Clinical Neurology
WE Science Citation Index Expanded (SCI-EXPANDED)
SC Oncology; Neurosciences & Neurology
GA 546DK
UT WOS:000273788600002
PM 19585223
OA Green Published, Green Accepted, Green Submitted, hybrid
DA 2023-08-27
ER

PT J
AU Zagzag, D
   Miller, DC
   Knopp, E
   Farmer, JP
   Lee, M
   Biria, S
   Pellicer, A
   Epstein, FJ
   Allen, JC
AF Zagzag, D
   Miller, DC
   Knopp, E
   Farmer, JP
   Lee, M
   Biria, S
   Pellicer, A
   Epstein, FJ
   Allen, JC
TI Primitive neuroectodermal tumors of the brainstem: Investigation of
   seven cases
SO PEDIATRICS
LA English
DT Article
DE primitive neuroectodermal tumors; brainstem; children; p53 mutation
ID CENTRAL-NERVOUS-SYSTEM; P53 GENE-MUTATIONS; PEDIATRIC-ONCOLOGY-GROUP;
   HYPERFRACTIONATED RADIATION-THERAPY; STEM GLIOMAS; MEDULLOBLASTOMA;
   CHILDREN; CHILDHOOD; DIFFERENTIATION; CHEMOTHERAPY
AB Objective. We discuss the clinical aspects, pathology, and molecular genetics of 7 patients with primitive neuroectodermal tumors (PNETs) arising in the brainstem that were treated at our institution from 1986 through 1995. Most neuro-oncologists avoid performing biopsies in children with pontine tumors. This article raises the question as to whether biopsies should be performed, because treatment recommendations might differ if a PNET was diagnosed rather than a pontine glioma.
   Patients and Methods. We reviewed the clinical neuro-oncology database and the files of the Division of Neuropathology at New York University Medical Center from 1986 through 1995 and identified 7 histologically confirmed PNETs arising in the brainstem among 146 pediatric brainstem tumors. The clinical, neuroradiological, and neuropathological data were reviewed. Postmortem examinations were performed in 2 cases. Formalin-fixed, paraffin-embedded tumor tissues were also available in 6 of 7 patients that were tested for p53 gene mutations using single-strand conformation polymorphism analysis. We also tested 9 cerebellar PNETs, 9 brainstem gliomas, and 3 normal brains for p53 gene mutations as controls.
   Results. All 7 patients presented with focal cranial nerve deficits, and 2 were also hemiparetic. The median age at diagnosis was 2.7 (1-8 years). Magnetic resonance imaging (MRI) characteristics included a focal intrinsic exophytic nonenhancing brainstem lesion that had low T1-weighted and high T2-weighted signals. Hydrocephalus was present in 5 patients at diagnosis, 3 of whom had leptomeningeal dissemination. Meningeal dissemination occurred later in the course of the disease in 3 other patients. Five children required shunts at diagnosis and another 2 at recurrence. Despite therapy, all 7 PNET patients died within 17 months of diagnosis with a mean survival of 8 (4-17) months. No mutation in the p53 gene was detected.
   Conclusions. Brainstem PNETs tend to arise at a younger age than brainstem gliomas and medulloblastomas. The MRI pattern suggests a localized rather than a diffuse intrinsic nonenhancing brainstem tumor. Like other PNETs, brainstem PNETs have a high predilection to disseminate within the central nervous system. The absence of p53 mutations is similar to other PNETs. Despite their origin close to the cerebellum, brainstem PNETs exhibit a more aggressive behavior and result in worse clinical outcomes than do cerebellar PNETs.
C1 NYU, Sch Med,Med Ctr, Dept Pathol, Div Neuropathol, New York, NY 10016 USA.
   NYU, Med Ctr, Dept Neurosurg, New York, NY 10016 USA.
   NYU, Med Ctr, Dept Radiol, New York, NY 10016 USA.
   NYU, Med Ctr, Kaplan Canc Ctr, New York, NY 10016 USA.
   Montreal Childrens Hosp, Dept Neurosurg, Montreal, PQ H3H 1P3, Canada.
   Med Coll Georgia, Childrens Med Ctr, Dept Neurosurg, Augusta, GA 30912 USA.
   Beth Israel Med Ctr, Inst Neurol & Neurosurg, New York, NY 10003 USA.
C3 New York University; New York University; New York University; New York
   University; McGill University; University System of Georgia; Augusta
   University; Harvard University; Beth Israel Deaconess Medical Center
RP Zagzag, D (通讯作者)，NYU, Sch Med,Med Ctr, Dept Pathol, Div Neuropathol, 550 1st Ave, New York, NY 10016 USA.
RI MARTINEZ, ANTONIO PELLICER/C-4832-2015
OI Allen, Jeffrey/0000-0001-9957-6664; Pellicer, Angel/0000-0002-5062-0692
FU NCI NIH HHS [CA 50434] Funding Source: Medline
CR ALBRIGHT AL, 1983, CANCER, V52, P2313, DOI 10.1002/1097-0142(19831215)52:12<2313::AID-CNCR2820521226>3.0.CO;2-I
   ALTMAN N, 1985, AM J NEURORADIOL, V6, P15
   Bailey P, 1925, ARCH NEURO PSYCHIATR, V14, P192, DOI 10.1001/archneurpsyc.1925.02200140055002
   BARKOVICH AJ, 1995, PEDIAT NEUROIMAGING, P325
   BARKOVICH AJ, 1991, PEDIATR NEUROSURG, V16, P73, DOI DOI 10.1159/000120511]
   BECKER LE, 1983, HUM PATHOL, V14, P538, DOI 10.1016/S0046-8177(83)80006-9
   Behnke J, 1996, CHILD NERV SYST, V12, P125, DOI 10.1007/BF00266812
   BERGER MS, 1983, NEUROSURGERY, V12, P298, DOI 10.1227/00006123-198303000-00008
   BIEGEL JA, 1989, GENE CHROMOSOME CANC, V1, P139, DOI 10.1002/gcc.2870010206
   Bigner SH, 1990, BRAIN PATHOL, V1, P12, DOI 10.1111/j.1750-3639.1990.tb00633.x
   BIGNER SH, 1988, CANCER GENET CYTOGEN, V30, P91, DOI 10.1016/0165-4608(88)90096-9
   BILANIUK LT, 1980, RADIOLOGY, V134, P89, DOI 10.1148/radiology.134.1.7350640
   BONNIN JM, 1984, CANCER-AM CANCER SOC, V54, P2137, DOI 10.1002/1097-0142(19841115)54:10<2137::AID-CNCR2820541014>3.0.CO;2-D
   BORIT A, 1980, CANCER-AM CANCER SOC, V45, P1408, DOI 10.1002/1097-0142(19800315)45:6<1408::AID-CNCR2820450619>3.0.CO;2-0
   CAPUTY AJ, 1987, J NEUROSURG, V66, P80, DOI 10.3171/jns.1987.66.1.0080
   CHATTY EM, 1971, CANCER-AM CANCER SOC, V28, P977, DOI 10.1002/1097-0142(1971)28:4<977::AID-CNCR2820280422>3.0.CO;2-7
   DEUTSCH M, 1980, CANCER, V45, P2194, DOI 10.1002/1097-0142(19800415)45:8<2194::AID-CNCR2820450831>3.0.CO;2-M
   EPSTEIN F, 1986, J NEUROSURG, V64, P11, DOI 10.3171/jns.1986.64.1.0011
   EVANS AE, 1990, J NEUROSURG, V72, P572, DOI 10.3171/jns.1990.72.4.0572
   FARWELL JR, 1977, CANCER, V40, P3123, DOI 10.1002/1097-0142(197712)40:6<3123::AID-CNCR2820400656>3.0.CO;2-6
   Feigin I, 1983, J Neurooncol, V1, P95
   FRANKEL RH, 1992, CANCER RES, V52, P1427
   FREEMAN CR, 1991, CANCER-AM CANCER SOC, V68, P474, DOI 10.1002/1097-0142(19910801)68:3<474::AID-CNCR2820680305>3.0.CO;2-7
   GAIDANO G, 1991, P NATL ACAD SCI USA, V88, P5413, DOI 10.1073/pnas.88.12.5413
   HART MN, 1973, CANCER, V32, P890, DOI 10.1002/1097-0142(197310)32:4<890::AID-CNCR2820320421>3.0.CO;2-O
   HERRICK MK, 1979, BRAIN, V102, P289, DOI 10.1093/brain/102.2.289
   HOLLSTEIN M, 1991, SCIENCE, V253, P49, DOI 10.1126/science.1905840
   HORTEN BC, 1976, BRAIN, V99, P735, DOI 10.1093/brain/99.4.735
   HOSOI G, 1994, CANCER, V73, P3087, DOI 10.1002/1097-0142(19940615)73:12<3087::AID-CNCR2820731230>3.0.CO;2-9
   KELLY PJ, 1987, J NEUROSURG, V66, P865, DOI 10.3171/jns.1987.66.6.0865
   KORF H, 1987, CANCER, V60, P1763, DOI 10.1002/1097-0142(19871015)60:8<1763::AID-CNCR2820600815>3.0.CO;2-3
   KOSNIK EJ, 1978, J NEUROSURG, V48, P741, DOI 10.3171/jns.1978.48.5.0741
   KRETSCHMAR CS, 1993, CANCER-AM CANCER SOC, V72, P1404, DOI 10.1002/1097-0142(19930815)72:4<1404::AID-CNCR2820720441>3.0.CO;2-G
   LITTMAN P, 1980, CANCER, V45, P2787, DOI 10.1002/1097-0142(19800601)45:11<2787::AID-CNCR2820451113>3.0.CO;2-V
   LOUIS DN, 1993, J NEUROPATHOL EXP NE, V52, P505
   MALKIN D, 1993, PRINCIPLES PRACTICE, P7
   MASHIYAMA S, 1991, ONCOGENE, V6, P1313
   MEYERS SP, 1992, AM J ROENTGENOL, V158, P859, DOI 10.2214/ajr.158.4.1546606
   Molloy PT, 1996, J NEUROSURG, V84, P430, DOI 10.3171/jns.1996.84.3.0430
   MOSIJCZUK AD, 1993, CANCER-AM CANCER SOC, V72, P2755, DOI 10.1002/1097-0142(19931101)72:9<2755::AID-CNCR2820720937>3.0.CO;2-V
   NORTH C, 1985, AM J NEURORADIOL, V6, P11
   OHGAKI H, 1991, CANCER RES, V51, P6202
   PACKER RJ, 1984, J NEUROSURG, V61, P296, DOI 10.3171/jns.1984.61.2.0296
   PACKER RJ, 1985, NEUROLOGY, V35, P397, DOI 10.1212/WNL.35.3.397
   PACKER RJ, 1983, ANN NEUROL, V14, P177, DOI 10.1002/ana.410140204
   PACKER RJ, 1994, J NEUROSURG, V81, P690, DOI 10.3171/jns.1994.81.5.0690
   PACKER RJ, 1985, ANN NEUROL, V18, P217, DOI 10.1002/ana.410180209
   PACKER RJ, 1984, PEDIATRICS, V74, P97
   PACKER RJ, 1994, CANCER-AM CANCER SOC, V74, P1827, DOI 10.1002/1097-0142(19940915)74:6<1827::AID-CNCR2820740628>3.0.CO;2-Q
   PARK TS, 1983, J NEUROSURG, V58, P543, DOI 10.3171/jns.1983.58.4.0543
   Perez MI, 1997, ARCH DERMATOL, V133, P189
   RAFFEL C, 1993, NEUROSURGERY, V33, P301, DOI 10.1227/00006123-199308000-00018
   Raffel C, 1993, NEUROSURGERY, V33, P305
   Rorke LB, 1996, J NEUROSURG, V85, P56, DOI 10.3171/jns.1996.85.1.0056
   RORKE LB, 1983, J NEUROPATH EXP NEUR, V42, P1, DOI 10.1097/00005072-198301000-00001
   Rorke LB, 1997, BRAIN PATHOL, V7, P765, DOI 10.1111/j.1750-3639.1997.tb01063.x
   RORKE LB, 1985, CANCER-AM CANCER SOC, V56, P1869, DOI 10.1002/1097-0142(19851001)56:7+<1869::AID-CNCR2820561330>3.0.CO;2-0
   RUBINSTEIN LJ, 1972, J NEUROPATH EXP NEUR, V31, P7, DOI 10.1097/00005072-197201000-00002
   RUBINSTEIN LJ, 1975, CANCER, V31, P7
   RUSSELL DS, 1989, PATHOL TUMOURS NER, P277
   RUSSELL DS, 1989, PATHOL TUMOURS NER, P261
   SAYLORS RL, 1991, CANCER RES, V51, P4721
   STROINK AR, 1987, NEUROSURGERY, V20, P439, DOI 10.1227/00006123-198703000-00014
   STROINK AR, 1986, J NEUROSURG, V65, P745, DOI 10.3171/jns.1986.65.6.0745
   WIKSTRAND CJ, 1991, INVAS METAST, V11, P310
   WRIGHT DK, 1994, PCR PROTOCOLS GUIDE, P153
   ZAGZAG D, 1989, AM J NEURORADIOL, V10, P529
   ZAR JH, 1974, BIOSTAT ANAL, P291
   ZHANG SJ, 1993, BIOCHEM BIOPH RES CO, V196, P851, DOI 10.1006/bbrc.1993.2327
   Zimmerman RA, 1996, PEDIATR NEUROSURG, V25, P83, DOI 10.1159/000121102
   ZIMMERMAN RA, 1992, PEDIATR NEUROSURG, V18, P58, DOI 10.1159/000120644
NR 71
TC 51
Z9 53
U1 0
U2 1
PU AMER ACAD PEDIATRICS
PI ELK GROVE VILLAGE
PA 141 NORTH-WEST POINT BLVD,, ELK GROVE VILLAGE, IL 60007-1098 USA
SN 0031-4005
J9 PEDIATRICS
JI Pediatrics
PD NOV
PY 2000
VL 106
IS 5
BP 1045
EP 1053
DI 10.1542/peds.106.5.1045
PN 1
PG 9
WC Pediatrics
WE Science Citation Index Expanded (SCI-EXPANDED)
SC Pediatrics
GA 369ED
UT WOS:000165052300015
PM 11061774
DA 2023-08-27
ER

PT J
AU Lim, YG
   Tan, EEK
   Looi, WS
   Wong, RX
   Chang, KTE
   Low, DCY
   Seow, WT
   Low, SYY
AF Lim, Yuan Guang
   Tan, Enrica E. K.
   Looi, Wen Shen
   Wong, Ru Xin
   Chang, Kenneth T. E.
   Low, David C. Y.
   Seow, Wan Tew
   Low, Sharon Y. Y.
TI Biopsy of paediatric brainstem intrinsic tumours: Experience from a
   Singapore Children's Hospital
SO JOURNAL OF CLINICAL NEUROSCIENCE
LA English
DT Article
DE Brainstem biopsy; Brainstem tumour; Paediatric brainstem tumour
ID PONTINE GLIOMA; INTRAOPERATIVE MRI; SURGICAL BIOPSY; TEMOZOLOMIDE;
   FEASIBILITY; DIAGNOSIS; LESIONS; SAFETY
AB Background: Biopsy of intrinsic brainstem tumours presumed to be diffuse midline gliomas (previously known as DIPG) is controversial. Surgery has risks of injury to the eloquent brainstem and may not have direct benefit to the patient. Technological improvements in operative adjuncts have allowed the role of biopsy for paediatric brainstem lesions to be revisited with new insights. This study aims to evaluate our institutional experience in brainstem biopsy. Methods: This is an ethics-approved retrospective study based in KK Women's and Children's Hospital. Patients diagnosed with intrinsic brainstem tumours and managed by the Neurosurgical Service were included. Variables of interest included patient demographics, neuroimaging features, type of surgery, histological and molecular diagnosis, treatment, and outcomes.Results: From 2006 to 2021, a total of 27 brainstem intrinsic tumours were referred to the Neurosurgical Service. Eleven (40.7 %) patients underwent stereotactic biopsy and 10 (37 %) had open biopsies. Histologically, 10 (37 %) were confirmed to be high grade gliomas, eight (29.6 %) were low grade gliomas and 3 (11.1 %) were malignant embryonal tumours. No negative diagnostic results or permanent postoperative complications were encountered. Five patients went on to have their tumours interrogated via next-generation sequencing to look for targetable mutations. The remaining 6 (22.2 %) patients did not undergo biopsy, whereby 1 of them is still alive after 6 years.Conclusion: Biopsy of paediatric brainstem intrinsic tumours is a safe procedure that concurrs with accurate tissue diagnosis. This option can be offered to affected patients, especially to identify relevant markers for targeted therapy.
C1 [Lim, Yuan Guang; Low, David C. Y.; Seow, Wan Tew; Low, Sharon Y. Y.] Natl Neurosci Inst, Dept Neurosurg, 11 Jalan Tan Tock Seng, Singapore 308433, Singapore.
   [Tan, Enrica E. K.] KK Womens & Childrens Hosp, Paediat Haematol Oncol Serv, 100 Bukit Timah Rd, Singapore 229899, Singapore.
   [Looi, Wen Shen; Wong, Ru Xin] Natl Canc Ctr, Div Radiat Oncol, 31 Third Hosp Ave, Singapore 168753, Singapore.
   [Chang, Kenneth T. E.] KK Womens & Childrens Hosp, Dept Pathol & Lab Med, 100 Bukit Timah Rd, Singapore 229899, Singapore.
   [Low, David C. Y.; Seow, Wan Tew; Low, Sharon Y. Y.] KK Womens & Childrens Hosp, Neurosurg Serv, 100 Bukit Timah Rd, Singapore 229899, Singapore.
   [Low, David C. Y.; Seow, Wan Tew; Low, Sharon Y. Y.] SingHealth Duke NUS Neurosci Acad Clin Program, 11 Jalan Tan Tock Seng, Singapore 308433, Singapore.
   [Tan, Enrica E. K.; Low, Sharon Y. Y.] SingHlth Duke NUS Paediat Acad Clin Program, 100 Bukit Timah Rd, Singapore 229899, Singapore.
   [Low, Sharon Y. Y.] KK Womens & Childrens Hosp, Neurosurg Serv, 100 Bukit Timah Rd, Singapore 229899, Singapore.
C3 National Neuroscience Institute (NNI); Tan Tock Seng Hospital; KK
   Women's & Children's Hospital; National Cancer Centre Singapore (NCCS);
   KK Women's & Children's Hospital; KK Women's & Children's Hospital; Tan
   Tock Seng Hospital; KK Women's & Children's Hospital
RP Low, SYY (通讯作者)，KK Womens & Childrens Hosp, Neurosurg Serv, 100 Bukit Timah Rd, Singapore 229899, Singapore.
EM
CR Albright AL, 1996, PEDIATR NEUROSURG, V24, P252
   ALBRIGHT AL, 1993, NEUROSURGERY, V33, P1026
   Almojuela A, 2020, CAN J NEUROL SCI, V47, P578, DOI 10.1017/cjn.2020.52
   AlRayahi J, 2018, RADIOGRAPHICS, V38, P2102, DOI 10.1148/rg.2018180109
   [Anonymous], 2007, WHO HISTOLOGICAL CLA
   Azad TD, 2020, NEUROSURG FOCUS, V48, DOI 10.3171/2019.9.FOCUS19699
   Bugiani M, 2017, ONCOTARGET, V8, P60447, DOI 10.18632/oncotarget.19726
   Cage TA, 2013, CHILD NERV SYST, V29, P1313, DOI 10.1007/s00381-013-2101-0
   Cartmill M, 1999, CHILD NERV SYST, V15, P235
   Choudhri AF, 2015, PEDIATR RADIOL, V45, P397, DOI 10.1007/s00247-015-3322-z
   Coppola A, 2016, CHILD NERV SYST, V32, P1849, DOI 10.1007/s00381-016-3180-5
   Dellaretti M, 2011, J NEUROSURG-PEDIATR, V8, P539, DOI 10.3171/2011.9.PEDS1167
   Goker B, 2019, CHILD NERV SYST, V35, P429, DOI 10.1007/s00381-018-04037-4
   Goodman AM, 2017, JCO PRECIS ONCOL, V1, DOI 10.1200/PO.16.00004
   Green AL, 2015, CURR ONCOL REP, V17, DOI 10.1007/s11912-014-0436-7
   Green AL., 2020, NEUROONCOL ADV, V2, pvdaa021
   Guillamo JS, 2001, CURR OPIN NEUROL, V14, P711, DOI 10.1097/00019052-200112000-00006
   Gupta A, 2018, CHRON RESP DIS, V16, DOI 10.1177/1479972318769763
   Gupta N, 2018, NEURO-ONCOLOGY, V20, P1547, DOI 10.1093/neuonc/noy070
   Hankinson TC, 2011, J NEUROSURG-PEDIATR, V8, P97, DOI 10.3171/2011.4.PEDS1180
   Hegi ME, 2005, NEW ENGL J MED, V352, P997, DOI 10.1056/NEJMoa043331
   Hersh DS, 2020, J NEUROSURG-PEDIATR, V26, P552, DOI 10.3171/2020.4.PEDS2092
   Himes BT, 2019, FRONT ONCOL, V9, DOI 10.3389/fonc.2019.00031
   Kuzan-Fischer CM, 2019, J NEUROSURG-PEDIATR, V24, P611, DOI 10.3171/2019.5.PEDS18376
   Lai MY, 2020, J CLIN ONCOL, V38
   Louis DN., 2016, WHO CLASSIFICATION T
   Low SYY, 2020, WORLD NEUROSURG, V135, DOI 10.1016/j.wneu.2019.10.083
   Low SYY, 2018, CHILD NERV SYST, V34, P19, DOI 10.1007/s00381-017-3627-3
   Lu VM, 2019, J NEUROSURG-PEDIATR, V24, P593, DOI 10.3171/2019.6.PEDS19259
   La Madrid AM, 2015, FRONT ONCOL, V5, DOI 10.3389/fonc.2015.00148
   Mount CW, 2018, NAT MED, V24, P572, DOI 10.1038/s41591-018-0006-x
   Norska A, 2010, NEUROIMAG CLIN N AM, V20, P293, DOI 10.1016/j.nic.2010.04.003
   Orillac C, 2021, NEUROSURGERY, V89, P727, DOI 10.1093/neuros/nyaa475
   Orringer DA, 2012, EXPERT REV MED DEVIC, V9, P491, DOI [10.1586/ERD.12.42, 10.1586/erd.12.42]
   Parenrengi MA, 2022, CHILD NERV SYST, V38, P1643, DOI 10.1007/s00381-022-05482-y
   Pfaff E, 2019, EUR J CANCER, V114, P27, DOI 10.1016/j.ejca.2019.03.019
   Prabhu SP, 2011, CHILD NERV SYST, V27, P11, DOI 10.1007/s00381-010-1323-7
   Puget S, 2015, CHILD NERV SYST, V31, P1773, DOI 10.1007/s00381-015-2832-1
   Puget Stephanie, 2012, Am Soc Clin Oncol Educ Book, P629, DOI 10.14694/EdBook_AM.2012.32.629
   Recinos PF, 2007, PEDIATR NEUROSURG, V43, P192, DOI 10.1159/000098831
   Risholm P, 2011, NEUROSURG CLIN N AM, V22, P197, DOI 10.1016/j.nec.2010.12.001
   Rizzo D, 2015, J CHEMOTHERAPY, V27, P106, DOI 10.1179/1973947814Y.0000000228
   Roujeau T, 2007, J NEUROSURG, V107, P1, DOI 10.3171/PED-07/07/001
   Sala F, 2015, J NEUROSURG SCI, V59, P373
   Shi S, 2021, WORLD NEUROSURG, V148, pE565, DOI 10.1016/j.wneu.2021.01.024
   Singh DK, 2023, BRIT J NEUROSURG, V37, P82, DOI 10.1080/02688697.2021.2016615
   Srikanthan Dilakshan, 2021, Chin Neurosurg J, V7, P6, DOI 10.1186/s41016-020-00218-w
   STECK J, 1995, SURG NEUROL, V43, P563, DOI 10.1016/0090-3019(95)00156-5
   Synhaeve NE, 2018, ACTA NEUROPATHOL COM, V6, DOI 10.1186/s40478-018-0633-y
   Tisnado J, 2016, J CHILD NEUROL, V31, P1386, DOI 10.1177/0883073816634855
   Vitanza NA, 2019, CURR TREAT OPTION NE, V21, DOI 10.1007/s11940-019-0577-y
   Wadden J, 2022, FRONT IMMUNOL, V13, DOI 10.3389/fimmu.2022.882452
   Walker DA, 2013, NEURO-ONCOLOGY, V15, P462, DOI 10.1093/neuonc/nos330
   Wang ZHJ, 2015, PEDIATR BLOOD CANCER, V62, P163, DOI 10.1002/pbc.25224
   Warren KE, 2012, FRONT ONCOL, V2, DOI 10.3389/fonc.2012.00205
   WHO, 2021, CONF PROB COVID 19 C, P2021
   Zarifi M, 2016, PEDIATR RADIOL, V46, P952, DOI 10.1007/s00247-016-3547-5
NR 57
TC 0
Z9 0
U1 1
U2 2
PU ELSEVIER SCI LTD
PI OXFORD
PA THE BOULEVARD, LANGFORD LANE, KIDLINGTON, OXFORD OX5 1GB, OXON, ENGLAND
SN 0967-5868
EI 1532-2653
J9 J CLIN NEUROSCI
JI J. Clin. Neurosci.
PD DEC
PY 2022
VL 106
BP 8
EP 13
DI 10.1016/j.jocn.2022.09.021
EA OCT 2022
PG 6
WC Clinical Neurology; Neurosciences
WE Science Citation Index Expanded (SCI-EXPANDED)
SC Neurosciences & Neurology
GA 5I3YB
UT WOS:000868295200002
PM 36228504
DA 2023-08-27
ER

PT J
AU Fukuoka, K
   Yanagisawa, T
   Watanabe, Y
   Suzuki, T
   Shirahata, M
   Adachi, J
   Mishima, K
   Fujimaki, T
   Matsutani, M
   Wada, S
   Sasaki, A
   Nishikawa, R
AF Fukuoka, Kohei
   Yanagisawa, Takaaki
   Watanabe, Yuko
   Suzuki, Tomonari
   Shirahata, Mitsuaki
   Adachi, Jun-ichi
   Mishima, Kazuhiko
   Fujimaki, Takamitsu
   Matsutani, Masao
   Wada, Satoru
   Sasaki, Atsushi
   Nishikawa, Ryo
TI Brainstem oligodendroglial tumors in children: two case reports and
   review of literatures
SO CHILDS NERVOUS SYSTEM
LA English
DT Article
DE Oligodendroglial tumor; Brainstem tumor; Pediatric brain tumor;
   Pathological diagnosis; Histone H 3.3 mutation
ID ANAPLASTIC OLIGODENDROGLIOMA; PEDIATRIC OLIGODENDROGLIOMAS; VINCRISTINE
   CHEMOTHERAPY; K27M MUTATION; GLIOMAS; 19Q; 1P
AB There is little information on pediatric oligodendroglial tumor located in the brainstem because of its rarity.
   Here, we present two pediatric cases of pontine oligodendroglial tumors with radiological findings atypical for diffuse intrinsic pontine glioma.
   The first patient was an 8-year-old boy. Brain magnetic resonance imaging (MRI) demonstrated diffuse high-intensity changes in the pons, left middle cerebellar peduncle, and part of the left cerebellar hemisphere on T2-weighted and fluid-attenuated inversion recovery images, with an enhanced spot lesion in the left cerebellar hemisphere. The pathological diagnosis was anaplastic oligodendroglioma, and we identified a mutation in histone H3.3 in the tumor specimen. He succumbed to massive disseminated relapse 7 months from diagnosis despite local radiation therapy. The second patient, a 2-year-old girl, was diagnosed with oligoastrocytoma. Brain MRI revealed a large mass in her rostral pons extended to the fourth ventricle with obstructive hydrocephalus. The tumor recurred with intracranial dissemination 56 months post-surgery.
   Pediatric brainstem oligodendroglial tumors can include histone H3.3-mutated tumors and have a tendency to disseminate throughout the neuroaxis at the time of relapse.
C1 [Fukuoka, Kohei; Yanagisawa, Takaaki; Watanabe, Yuko; Suzuki, Tomonari] Saitama Med Univ, Int Med Ctr, Dept Neurooncol Neurosurg, Div Pediat Neurooncol, Hidaka, Saitama 3501298, Japan.
   [Shirahata, Mitsuaki; Adachi, Jun-ichi; Mishima, Kazuhiko; Matsutani, Masao; Nishikawa, Ryo] Saitama Med Univ, Int Med Ctr, Dept Neurooncol Neurosurg, Hidaka, Saitama 3501298, Japan.
   [Fujimaki, Takamitsu] Saitama Med Univ Hosp, Dept Neurosurg, Moroyama, Saitama, Japan.
   [Wada, Satoru] Saitama Med Univ, Int Med Ctr, Dept Breast Oncol, Hidaka, Saitama 3501298, Japan.
   [Sasaki, Atsushi] Saitama Med Univ, Dept Pathol, Moroyama, Saitama, Japan.
C3 Saitama Medical University; Saitama Medical University; Saitama Medical
   University; Saitama Medical University; Saitama Medical University
RP Fukuoka, K (通讯作者)，Saitama Med Univ, Int Med Ctr, Dept Neurooncol Neurosurg, Div Pediat Neurooncol, Yamane 1397-1, Hidaka, Saitama 3501298, Japan.
EM
CR Adachi J, 2012, J NEURO-ONCOL, V107, P147, DOI 10.1007/s11060-011-0721-3
   ALBRIGHT AL, 1993, NEUROSURGERY, V33, P1026
   Broniscer A, 2008, CANCER, V113, P566, DOI 10.1002/cncr.23584
   DOHRMANN GJ, 1978, SURG NEUROL, V10, P21
   Khuong-Quang DA, 2012, ACTA NEUROPATHOL, V124, P439, DOI 10.1007/s00401-012-0998-0
   Gielen GH, 2013, AM J CLIN PATHOL, V139, P345, DOI 10.1309/AJCPABOHBC33FVMO
   Hyder DJ, 2007, J NEURO-ONCOL, V83, P1, DOI 10.1007/s11060-006-9299-6
   Kreiger PA, 2005, ACTA NEUROPATHOL, V109, P387, DOI 10.1007/s00401-004-0976-2
   KRUEGER EG, 1952, NEUROLOGY, V2, P461, DOI 10.1212/WNL.2.11-12.461
   Lesniak MS, 2003, PEDIATR NEUROSURG, V39, P314, DOI 10.1159/000075260
   Mittelbronn M, 2005, HUM PATHOL, V36, P854, DOI 10.1016/j.humpath.2005.05.017
   MORK SJ, 1985, J NEUROSURG, V63, P881, DOI 10.3171/jns.1985.63.6.0881
   Ogiwara H, 2013, CHILD NERV SYST, V29, P833, DOI 10.1007/s00381-013-2042-7
   Packer RJ, 1997, J NEUROSURG, V86, P747, DOI 10.3171/jns.1997.86.5.0747
   PACKER RJ, 1985, CANCER, V56, P195, DOI 10.1002/1097-0142(19850701)56:1<195::AID-CNCR2820560133>3.0.CO;2-3
   Peters O, 2004, PEDIATR BLOOD CANCER, V43, P250, DOI 10.1002/pbc.20111
   Pollack IF, 2003, PEDIATR NEUROSURG, V39, P114, DOI 10.1159/000071647
   Raghavan R, 2003, J NEUROPATH EXP NEUR, V62, P530, DOI 10.1093/jnen/62.5.530
   REIFENBERGER G, 2007, WHO CLASSIFICATION T, P54
   Ronghe M, 2010, ONCOLOGY OF CNS TUMORS, SECOND EDITION, P453, DOI 10.1007/978-3-642-02874-8_29
   Sanai N, 2010, PEDIAT CNS TUMORS, P49
   van den Bent MJ, 2013, J CLIN ONCOL, V31, P344, DOI 10.1200/JCO.2012.43.2229
   Wang Kyu-Chang, 1993, Journal of Korean Medical Science, V8, P110
NR 23
TC 8
Z9 9
U1 0
U2 5
PU SPRINGER
PI NEW YORK
PA 233 SPRING ST, NEW YORK, NY 10013 USA
SN 0256-7040
EI 1433-0350
J9 CHILD NERV SYST
JI Childs Nerv. Syst.
PD MAR
PY 2015
VL 31
IS 3
BP 449
EP 455
DI 10.1007/s00381-014-2563-8
PG 7
WC Clinical Neurology; Pediatrics; Surgery
WE Science Citation Index Expanded (SCI-EXPANDED)
SC Neurosciences & Neurology; Pediatrics; Surgery
GA CB1CM
UT WOS:000349364200013
PM 25281433
DA 2023-08-27
ER

PT J
AU Yoo, TW
   Park, ES
   Kwon, DH
   Kim, CJ
AF Yoo, Tae Won
   Park, Eun Suk
   Kwon, Do Hoon
   Kim, Chang Jin
TI Gamma Knife Radiosurgery for Brainstem Metastasis
SO JOURNAL OF KOREAN NEUROSURGICAL SOCIETY
LA English
DT Article
DE Brainstem tumor; Gamma knife radiosurgery; Metastasis; Stereotactic
   radiosurgery
ID RECURSIVE PARTITIONING ANALYSIS; STEREOTACTIC RADIOSURGERY;
   PROGNOSTIC-FACTORS; RADIATION-THERAPY; RADIOTHERAPY; SURGERY; SURVIVAL;
   TUMORS
AB Objective : Brainstem metastases are rarely operable and generally unresponsive to conventional radiation therapy or chemotherapy. Recently, Gamma Knife Radiosurgery (GKRS) was used as feasible treatment option for brainstem metastasis. The present study evaluated our experience of brainstem metastasis which was treated with GKRS.
   Methods : Between November 1992 and June 2010, 32 patients (23 men and 9 women, mean age 56.1 years, range 39-73) were treated with GKRS for brainstem metastases. There were metastatic lesions in pons in 23, the midbrain in 6, and the medulla oblongata in 3 patients, respectively. The primary tumor site was lung in 21, breast in 3, kidney in 2 and other locations in 6 patients. The mean tumor volume was 1,517 mm(3) (range, 9-6,000), and the mean marginal dose was 15.9 Gy (range, 6-23). Magnetic Resonance Imaging (MRI) was obtained every 2-3 months following GKRS. Follow-up MRI was possible in 24 patients at a mean follow-up duration of 12.0 months (range, 1-45). Kaplan-Meier survival analysis was used to evaluate the prognostic factors.
   Results : Follow-up MRI showed tumor disappearance in 6, tumor shrinkage in 14, no change in tumor size in 1, and tumor growth in 3 patients, which translated into a local tumor control rate of 87.5% (21 of 24 tumors). The mean progression free survival was 12.2 months (range, 2-45) after GKRS. Nine patients were alive at the completion of the study, and the overall mean survival time after GKRS was 7.7 months (range, 1-22). One patient with metastatic melanoma experienced intratumoral hemorrhage during the follow-up period. Survival was found to be associated with score of more than 70 on Kamofsky performance status and low recursive partitioning analysis class (class 1 or 2), in terms of favorable prognostic factors.
   Conclusion : GKRS was found to be safe and effective for management of brainstem metastasis. The integral clinical status of patient seems to be important in determining the overall survival time.
C1 [Yoo, Tae Won; Park, Eun Suk; Kwon, Do Hoon; Kim, Chang Jin] Univ Ulsan, Coll Med, Asan Med Ctr, Dept Neurosurg, Seoul 138736, South Korea.
C3 University of Ulsan; Asan Medical Center
RP Kwon, DH (通讯作者)，Univ Ulsan, Coll Med, Asan Med Ctr, Dept Neurosurg, 88 Olymp Ro 43 Gil, Seoul 138736, South Korea.
EM
RI Park, Eun Suk/E-9024-2017
OI Park, Eun Suk/0000-0002-5090-5284
CR ADLER JR, 1992, J NEUROSURG, V76, P444, DOI 10.3171/jns.1992.76.3.0444
   ALEXANDER E, 1995, JNCI-J NATL CANCER I, V87, P34, DOI 10.1093/jnci/87.1.34
   Andrews DW, 2004, LANCET, V363, P1665, DOI 10.1016/S0140-6736(04)16250-8
   Aoyama H, 2006, JAMA-J AM MED ASSOC, V295, P2483, DOI 10.1001/jama.295.21.2483
   Auchter RM, 1996, INT J RADIAT ONCOL, V35, P27, DOI 10.1016/S0360-3016(96)85008-5
   Bindal AK, 1996, J NEUROSURG, V84, P748, DOI 10.3171/jns.1996.84.5.0748
   CAIRNCROSS JG, 1980, ANN NEUROL, V7, P529, DOI 10.1002/ana.410070606
   CHASON JL, 1963, CANCER, V16, P781, DOI 10.1002/1097-0142(196306)16:6<781::AID-CNCR2820160614>3.0.CO;2-M
   Chidel MA, 2000, INT J RADIAT ONCOL, V47, P993, DOI 10.1016/S0360-3016(00)00527-7
   FLICKINGER JC, 1994, INT J RADIAT ONCOL, V28, P797, DOI 10.1016/0360-3016(94)90098-1
   Flickinger JC, 1996, NEUROSURG CLIN N AM, V7, P497
   Fuentes S, 2006, NEUROSURGERY, V58, P37, DOI 10.1227/01.NEU.0000190655.95669.5C
   Gaspar L, 1997, INT J RADIAT ONCOL, V37, P745, DOI 10.1016/S0360-3016(96)00619-0
   Goodman KA, 2001, INT J RADIAT ONCOL, V50, P139, DOI 10.1016/S0360-3016(00)01584-4
   Huang CF, 1999, J NEUROSURG, V91, P563, DOI 10.3171/jns.1999.91.4.0563
   Hussain A, 2007, INT J RADIAT ONCOL, V67, P521, DOI 10.1016/j.ijrobp.2006.08.081
   Johnson JD, 1996, NEUROSURG CLIN N AM, V7, P337
   Kased N, 2008, J NEURO-ONCOL, V86, P195, DOI 10.1007/s11060-007-9458-4
   Kondziolka D, 1999, INT J RADIAT ONCOL, V45, P427, DOI 10.1016/S0360-3016(99)00198-4
   Lagerwaard FJ, 1999, INT J RADIAT ONCOL, V43, P795, DOI 10.1016/S0360-3016(98)00442-8
   LANG EF, 1964, SURG CLIN N AM, V44, P865
   Lorenzoni JG, 2009, SURG NEUROL, V71, P188, DOI 10.1016/j.surneu.2008.01.029
   MARKESBERY WR, 1978, ARCH NEUROL-CHICAGO, V35, P754, DOI 10.1001/archneur.1978.00500350058012
   Muacevic A, 1999, J NEUROSURG, V91, P35, DOI 10.3171/jns.1999.91.1.0035
   PATCHELL RA, 1990, NEW ENGL J MED, V322, P494, DOI 10.1056/NEJM199002223220802
   Shuto T, 2003, ACTA NEUROCHIR, V145, P755, DOI 10.1007/s00701-003-0034-1
   Simonova G, 2000, RADIOTHER ONCOL, V57, P207, DOI 10.1016/S0167-8140(00)00267-X
   SMALLEY SR, 1992, J NEUROSURG, V77, P531, DOI 10.3171/jns.1992.77.4.0531
   Sneed PK, 1999, INT J RADIAT ONCOL, V43, P549, DOI 10.1016/S0360-3016(98)00447-7
   Soffietti R, 2002, J NEUROL, V249, P1357, DOI 10.1007/s00415-002-0870-6
   Tosoni A, 2004, CRIT REV ONCOL HEMAT, V52, P199, DOI 10.1016/j.critrevonc.2004.08.006
   Valery CA, 2011, INT J, V80, P362
   VECHT CJ, 1993, ANN NEUROL, V33, P583, DOI 10.1002/ana.410330605
   Weltman E, 2000, INT J RADIAT ONCOL, V46, P1155, DOI 10.1016/S0360-3016(99)00549-0
   Yen CP, 2006, J NEUROSURG, V105, P213, DOI 10.3171/jns.2006.105.2.213
NR 35
TC 27
Z9 29
U1 0
U2 2
PU KOREAN NEUROSURGICAL SOC
PI SEOUL
PA BLDG B, #402, 27, CHILPAE-RO, JUNG-GU, SEOUL, 04511, SOUTH KOREA
SN 2005-3711
EI 1598-7876
J9 J KOREAN NEUROSURG S
JI J. Korean. Neurosurg. Soc.
PD OCT
PY 2011
VL 50
IS 4
BP 299
EP 303
DI 10.3340/jkns.2011.50.4.299
PG 5
WC Clinical Neurology; Surgery
WE Science Citation Index Expanded (SCI-EXPANDED)
SC Neurosciences & Neurology; Surgery
GA 843NF
UT WOS:000296678000004
PM 22200010
OA Green Published, gold
DA 2023-08-27
ER

PT J
AU Zuo, PC
   Li, YP
   He, C
   Wang, TT
   Zheng, X
   Liu, H
   Wu, Z
   Zhang, JT
   Liao, XB
   Zhang, LW
AF Zuo, Pengcheng
   Li, Yaopeng
   He, Chi
   Wang, Tantan
   Zheng, Xu
   Liu, Hao
   Wu, Zhen
   Zhang, Junting
   Liao, Xuebin
   Zhang, Liwei
TI Anti-tumor efficacy of anti-GD2 CAR NK-92 cells in diffuse intrinsic
   pontine gliomas
SO FRONTIERS IN IMMUNOLOGY
LA English
DT Article
DE GD2; CARNK; anti-tumor efficacy; diffuse intrinsic pontine gliomas;
   pediatric
ID BRAIN-STEM GLIOMA; ACTIVATION; RECEPTOR
AB BackgroundDiffuse intrinsic pontine gliomas (DIPGs) are rare and fatal pediatric brainstem gliomas with no cure. Chimeric antigen receptor (CAR)-engineered natural killer (NK) cells have been proven effective in treating glioblastoma (GBM) in preclinical studies. However, there are no relevant studies on the CAR-NK treatment for DIPG. Our study is the first to evaluate the anti-tumor activity and safety of GD2-CAR NK-92 cells treatment for DIPG. MethodsFive patient-derived DIPG cells and primary pontine neural progenitor cell (PPC) were used to access disialoganglioside GD2 expression. Cell killing activity of GD2-CAR NK-92 cells was analyzed by in vitro cytotoxicity assays. Two DIPG patient-derived xenograft models were established to detect the anti-tumor efficacy of GD2-CAR NK-92 cells in vivo. ResultsAmong the five patient-derived DIPG cells, four had high GD2 expression, and one had low GD2 expression. In in vitro assays, GD2-CAR NK-92 cells could effectively kill DIPG cells with high GD2 expression while having limited activity against DIPG cells with low GD2 expression. In in vivo assays, GD2-CAR NK-92 cells could inhibit tumor growth in TT150630 DIPG patient-derived xenograft mice (high GD2 expression) and prolong the overall survival of the mice. However, GD2-CAR NK-92 showed limited anti-tumor activity for TT190326DIPG patient-derived xenograft mice (low GD2 expression). ConclusionOur study demonstrates the potential and safety of GD2-CAR NK-92 cells for adoptive immunotherapy of DIPG. The safety and anti-tumor effect of this therapy need to be further demonstrated in future clinical trials.
C1 [Zuo, Pengcheng; Wu, Zhen; Zhang, Junting; Zhang, Liwei] Capital Med Univ, Beijing Tiantan Hosp, Dept Neurosurg, Beijing, Peoples R China.
   [Li, Yaopeng; He, Chi; Wang, Tantan; Liu, Hao; Liao, Xuebin] Tsinghua Univ, Peking Tsinghua Ctr Life Sci, Sch Pharmaceut Sci, Key Lab Bioorgan Phosphorus Chem & Chem Biol,Minis, Beijing, Peoples R China.
   [Zheng, Xu] Yufan Biotechnol Beijing Co LTD, Beijing, Peoples R China.
   [Liao, Xuebin] Capital Med Univ, Beijing Tiantan Hosp, Adv Innovat Ctr Human Brain Protect, Beijing, Peoples R China.
   [Zhang, Liwei] China Natl Clin Res Ctr Neurol Dis NCRC ND, Beijing, Peoples R China.
C3 Capital Medical University; Tsinghua University; Capital Medical
   University
RP Zhang, LW (通讯作者)，Capital Med Univ, Beijing Tiantan Hosp, Dept Neurosurg, Beijing, Peoples R China.; Liao, XB (通讯作者)，Tsinghua Univ, Peking Tsinghua Ctr Life Sci, Sch Pharmaceut Sci, Key Lab Bioorgan Phosphorus Chem & Chem Biol,Minis, Beijing, Peoples R China.; Liao, XB (通讯作者)，Capital Med Univ, Beijing Tiantan Hosp, Adv Innovat Ctr Human Brain Protect, Beijing, Peoples R China.; Zhang, LW (通讯作者)，China Natl Clin Res Ctr Neurol Dis NCRC ND, Beijing, Peoples R China.
EM
FU Multicenter clinical big data study and multi-path tumorigenesis
   mechanisms and precision treatment research on brainstem glioma
   [JINGYIYAN2018-7]
FX This study was funded by Multicenter clinical big data study and
   multi-path tumorigenesis mechanisms and precision treatment research on
   brainstem glioma (JINGYIYAN2018-7).
CR Brudno JN, 2019, BLOOD REV, V34, P45, DOI 10.1016/j.blre.2018.11.002
   Buczkowicz P, 2015, FRONT ONCOL, V5, DOI 10.3389/fonc.2015.00147
   Buczkowicz P, 2014, ACTA NEUROPATHOL, V128, P573, DOI 10.1007/s00401-014-1319-6
   Chang YH, 2013, CANCER RES, V73, P1777, DOI 10.1158/0008-5472.CAN-12-3558
   Donaldson SS, 2006, J CLIN ONCOL, V24, P1266, DOI 10.1200/JCO.2005.04.6599
   Khuong-Quang DA, 2012, ACTA NEUROPATHOL, V124, P439, DOI 10.1007/s00401-012-0998-0
   Esser R, 2012, J CELL MOL MED, V16, P569, DOI 10.1111/j.1582-4934.2011.01343.x
   Han JF, 2015, SCI REP-UK, V5, DOI 10.1038/srep11483
   Hargrove D, 2006, LANCET ONCOL, V7, P241, DOI 10.1016/S1470-2045(06)70615-5
   Jiang H, 2014, MOL ONCOL, V8, P297, DOI 10.1016/j.molonc.2013.12.001
   Majzner RG, 2022, NATURE, V603, P934, DOI 10.1038/s41586-022-04489-4
   MARIA BL, 1993, J CHILD NEUROL, V8, P112, DOI 10.1177/088307389300800203
   Montagner IM, 2020, CELLS-BASEL, V9, DOI 10.3390/cells9061382
   Mount CW, 2018, NAT MED, V24, P572, DOI 10.1038/s41591-018-0006-x
   Oshio K, 2004, FASEB J, V18, P76, DOI 10.1096/fj.04-1711fje
   Robison NJ, 2014, J NEURO-ONCOL, V119, P7, DOI 10.1007/s11060-014-1448-8
   Romanski A, 2016, J CELL MOL MED, V20, P1287, DOI 10.1111/jcmm.12810
   Sharifzad F, 2020, INT J MOL SCI, V21, DOI 10.3390/ijms21072263
   Suzuki M, 2015, EXPERT OPIN THER TAR, V19, P349, DOI 10.1517/14728222.2014.986459
   Tang XW, 2018, AM J CANCER RES, V8, P1083
   Xie GZ, 2020, EBIOMEDICINE, V59, DOI 10.1016/j.ebiom.2020.102975
   Xu C, 2017, ONCOTARGET, V8, P76644, DOI 10.18632/oncotarget.19656
   Zhang CC, 2016, JNCI-J NATL CANCER I, V108, DOI 10.1093/jnci/djv375
NR 23
TC 0
Z9 0
U1 3
U2 3
PU FRONTIERS MEDIA SA
PI LAUSANNE
PA AVENUE DU TRIBUNAL FEDERAL 34, LAUSANNE, CH-1015, SWITZERLAND
SN 1664-3224
J9 FRONT IMMUNOL
JI Front. Immunol.
PD MAY 12
PY 2023
VL 14
AR 1145706
DI 10.3389/fimmu.2023.1145706
PG 9
WC Immunology
WE Science Citation Index Expanded (SCI-EXPANDED)
SC Immunology
GA H2VL2
UT WOS:000994594400001
PM 37251413
OA gold, Green Published
DA 2023-08-27
ER

PT J
AU Hashizume, R
   Gupta, N
AF Hashizume, Rintaro
   Gupta, Nalin
TI Patient-derived Tumor Models for Diffuse Intrinsic Pontine Gliomas
SO CURRENT NEUROPHARMACOLOGY
LA English
DT Review
DE Bioluminescent imaging; brainstem glioma; DIPG; H3K27M mutation;
   infiltrating astrocytoma; xenograft model
ID ACTIVATING ACVR1 MUTATIONS; BRAIN-STEM GLIOMA; MOLECULAR SUBGROUPS;
   DISTINCT SUBGROUPS; XENOGRAFT MODEL; GLIOBLASTOMA; MEDULLOBLASTOMA;
   THERAPY; DISEASE; CELL
AB Background: Diffuse intrinsic pontine gliomas represent a unique subtype of primary brain tumors occuring in a specific location and age. Their growth demonstrates early invasion and, following diagnosis, rapid growth not responsive to common therapies. Until recently, the genetic and cellular basis of these tumors was unknown. Genetic evidence implicates mutations in the histone genes in the origin of these tumors.
   Methods: Surgical biopsies performed on selected patients have resulted in the establishment of anatomically accurate mouse models that have been used to examine patterns of growth and response to new therapeutic agents.
   Results: Human derived pontine glioma models recapitulate the invasive patterns of growth. The grade of the original tumor affects the latency of tumor growth after implantation.
   Conclusion: The use of human-derived xenograft models allows for improved pre-clinical testing of new therapeutic targets in a tumor-and organ-specific manner.
C1 [Hashizume, Rintaro] Northwestern Univ, Dept Neurol Surg, Chicago, IL 60611 USA.
   [Gupta, Nalin] Univ Calif San Francisco, Dept Neurol Surg, Brain Tumor Res Ctr, San Francisco, CA 94143 USA.
C3 Northwestern University; University of California System; University of
   California San Francisco
RP Gupta, N (通讯作者)，Univ Calif San Francisco, Dept Neurol Surg, 505 Parnassus Ave,Room M779,550 16th St,4th Floor, San Francisco, CA 94143 USA.
EM
FU Pediatric Brain Tumor Foundation Institute Award
FX This work was supported by the Pediatric Brain Tumor Foundation
   Institute Award to the University of California San Francisco.
CR Aoki Y, 2012, J NEURO-ONCOL, V108, P29, DOI 10.1007/s11060-011-0796-x
   BARKOVICH AJ, 1991, PEDIATR NEUROSURG, V16, P73, DOI DOI 10.1159/000120511]
   Becher OJ, 2010, CANCER RES, V70, P2548, DOI 10.1158/0008-5472.CAN-09-2503
   Buczkowicz P, 2014, NAT GENET, V46, P451, DOI 10.1038/ng.2936
   Chan KM, 2013, GENE DEV, V27, P985, DOI 10.1101/gad.217778.113
   Dinca EB, 2007, J NEUROSURG, V107, P610, DOI 10.3171/JNS-07/09/0610
   Donaldson SS, 2006, J CLIN ONCOL, V24, P1266, DOI 10.1200/JCO.2005.04.6599
   Khuong-Quang DA, 2012, ACTA NEUROPATHOL, V124, P439, DOI 10.1007/s00401-012-0998-0
   Fontebasso AM, 2014, NAT GENET, V46, P462, DOI 10.1038/ng.2950
   Giannini C, 2005, NEURO-ONCOLOGY, V7, P164, DOI 10.1215/S1152851704000821
   Hashizume R, 2014, NAT MED, V20, P1394, DOI 10.1038/nm.3716
   Hashizume R, 2012, J NEURO-ONCOL, V110, P305, DOI 10.1007/s11060-012-0973-6
   Hashizume R, 2010, J NEURO-ONCOL, V96, P151, DOI 10.1007/s11060-009-9954-9
   Jones DTW, 2012, NATURE, V488, P100, DOI 10.1038/nature11284
   Lau J, 2012, CHILD NERV SYST, V28, P521, DOI 10.1007/s00381-012-1704-1
   Masui K, 2010, GLIA, V58, P1050, DOI 10.1002/glia.20986
   Monje M, 2011, P NATL ACAD SCI USA, V108, P4453, DOI 10.1073/pnas.1101657108
   Northcott PA, 2012, NATURE, V488, P49, DOI 10.1038/nature11327
   Paugh BS, 2010, J CLIN ONCOL, V28, P3061, DOI 10.1200/JCO.2009.26.7252
   Phillips HS, 2006, CANCER CELL, V9, P157, DOI 10.1016/j.ccr.2006.02.019
   Saito R, 2004, CANCER RES, V64, P2572, DOI 10.1158/0008-5472.CAN-03-3631
   Sandhu GS, 2010, WIRES SYST BIOL MED, V2, P398, DOI 10.1002/wsbm.71
   Schwartzentruber J, 2012, NATURE, V482, P226, DOI [10.1038/nature10833, 10.1007/s13311-012-0137-6]
   Silver DJ, 2013, J NEUROSCI, V33, P15603, DOI 10.1523/JNEUROSCI.3004-12.2013
   Sturm D, 2014, NAT REV CANCER, V14, P92, DOI 10.1038/nrc3655
   Sturm D, 2012, CANCER CELL, V22, P425, DOI 10.1016/j.ccr.2012.08.024
   Taylor KR, 2014, NAT GENET, V46, P457, DOI 10.1038/ng.2925
   Thirant C, 2011, PLOS ONE, V6, DOI 10.1371/journal.pone.0016375
   Truffaux N, 2014, NEUROONCOL
   Witt H, 2011, CANCER CELL, V20, P143, DOI 10.1016/j.ccr.2011.07.007
   Wu G, 2012, NAT GENET, V44, P251, DOI 10.1038/ng.1102
   Zarghooni M, 2010, J CLIN ONCOL, V28, P1337, DOI 10.1200/JCO.2009.25.5463
NR 32
TC 9
Z9 9
U1 0
U2 9
PU BENTHAM SCIENCE PUBL LTD
PI SHARJAH
PA EXECUTIVE STE Y-2, PO BOX 7917, SAIF ZONE, 1200 BR SHARJAH, U ARAB
   EMIRATES
SN 1570-159X
EI 1875-6190
J9 CURR NEUROPHARMACOL
JI Curr. Neuropharmacol.
PY 2017
VL 15
IS 1
BP 98
EP 103
DI 10.2174/1570159X14666160523144117
PG 6
WC Neurosciences; Pharmacology & Pharmacy
WE Science Citation Index Expanded (SCI-EXPANDED)
SC Neurosciences & Neurology; Pharmacology & Pharmacy
GA EH6AX
UT WOS:000391855800011
PM 27212051
OA Green Published
DA 2023-08-27
ER

PT J
AU Goncalves-Ferreira, AJ
   Herculano-Carvalho, M
   Pimentel, J
   Kelly, PJ
   Friedman, WA
AF Goncalves-Ferreira, AJ
   Herculano-Carvalho, M
   Pimentel, J
   Kelly, PJ
   Friedman, WA
TI Stereotactic biopsies of focal brainstem lesions
SO SURGICAL NEUROLOGY
LA English
DT Article
DE brain tumor; brainstem; stereotactic biopsy; focal brain lesions
ID SUBOCCIPITAL TRANSCEREBELLAR BIOPSY; POSTERIOR-FOSSA LESIONS; MASS
   LESIONS; TOMOGRAPHY; CHILDREN; SURGERY; TUMORS; EXPERIENCE; ASPIRATION;
   DIAGNOSIS
AB BACKGROUND Stereotactic biopsies of the brainstem (SBB) are a selected group of stereotactic operations owing to the lower incidence of brainstem lesions requiring biopsy, the greater complexity, and the higher risks of these procedures. Usually, the lower the lesion in the brainstem, the greater the risks involved. The approach of the different target locations by distinct routes, transcerebral and trans cerebellar, is still a matter of debate. Moreover, pure medullary lesions are seldom biopsied, whereas diffuse brainstem lesions, typical of children's brainstem gliomas and rather frequent in most published series, depend less and less on the histopathological confirmation for treatment. In this study, the authors present their own experience on SBB aiming to discuss mainly their indications, approach routes choice, and procedure techniques.
   METHODS The authors reviewed a series of 30 SBB (27 adults and 3 children) out of 450 stereotactic biopsies of the central nervous system performed for the last 10 years. All cases but one presented as focal brainstem masses, 19 mainly in the pons and 10 in the midbrain. Eleven lesions (10 in the midbrain and one midbrain-pontine) were approached by a transfrontal route. All the others were approached by a suboccipital transcerebellar route.
   RESULTS Twenty-six (87%) out of the 28 cases (93%) where a positive histopathological result was obtained had a specific diagnosis: 18 tumors (14 astrocytomas, 2 primary brain lymphomas, 1 oligodendroglioma, and 1 ganglioglioma), 2 toxoplasmosis, 2 sarcoidosis, 1 aspergillus abscess, 1 vasculitis, 1 acute inflammatory demyelinating disease, and 1 progressive multifocal leucoencephalopathy. Four nontumoral lesions arose in patients with AIDS. Morbidity was restricted to 2 cases consisting of transient cranial nerve deficits.
   CONCLUSIONS Stereotactic biopsies are specially useful for the diagnosis of focal brainstem lesions. Midbrain and midline lesions should be approached through the cerebrum, whereas most of the pontine lesions through the cerebellum. Overall, this procedure should be performed with "functional-like" precision and require some technical refinements to reach maximal accuracy with minimal morbidity. (C) 2003 Elsevier Inc. All rights reserved.
C1 Univ Lisbon, Hosp Santa Maria, Inst Anat, Fac Med Lisboa Piso 1,Dept Neurosurg, P-1649028 Lisbon, Portugal.
   Univ Lisbon, Hosp Santa Maria, Dept Anat, P-1649028 Lisbon, Portugal.
   Univ Lisbon, Hosp Santa Maria, Neuropathol Lab, P-1649028 Lisbon, Portugal.
C3 Universidade de Lisboa; Hospital Santa Maria; Universidade de Lisboa;
   Hospital Santa Maria; Universidade de Lisboa; Hospital Santa Maria
RP Goncalves-Ferreira, AJ (通讯作者)，Univ Lisbon, Hosp Santa Maria, Inst Anat, Fac Med Lisboa Piso 1,Dept Neurosurg, Av Prof Egas Moniz, P-1649028 Lisbon, Portugal.
RI Kelly, Patrick Joseph/E-8056-2017; Friedman, William/GLR-1220-2022
OI Kelly, Patrick Joseph/0000-0002-8540-7488; Pimentel,
   Jose/0000-0002-6101-6650; Ferreira, Antonio Jose de Carvalho
   Goncalves/0000-0002-1933-0304
CR ABERNATHEY CD, 1989, J NEUROSURG, V70, P195, DOI 10.3171/jns.1989.70.2.0195
   ALBRIGHT AL, 1993, NEUROSURGERY, V33, P1026
   APUZZO MLJ, 1987, NEUROSURGERY, V20, P930, DOI 10.1227/00006123-198706000-00019
   BEATTY RM, 1983, NEUROSURGERY, V13, P204, DOI 10.1227/00006123-198308000-00019
   Blond S, 1991, Acta Neurochir Suppl (Wien), V52, P75
   BOSCH DA, 1985, J NEUROSURG, V62, P153, DOI 10.3171/jns.1985.62.1.0153
   BROGGI G, 1983, ITAL J NEUROL SCI S, V2, P51
   Cartmill M, 1999, CHILD NERV SYST, V15, P235
   COFFEY RJ, 1985, APPL NEUROPHYSIOL, V48, P467
   DAUMASDUPORT C, 1988, CANCER-AM CANCER SOC, V62, P2152, DOI 10.1002/1097-0142(19881115)62:10<2152::AID-CNCR2820621015>3.0.CO;2-T
   EPSTEIN F, 1986, J NEUROSURG, V64, P11, DOI 10.3171/jns.1986.64.1.0011
   Frank F, 1988, Acta Neurochir Suppl (Wien), V42, P177
   Galanda M, 1984, ACTA NEUROCHIR S, V33, P213
   Giunta F, 1989, Acta Neurochir Suppl (Wien), V46, P86
   GONCALVESFERREIRA A, 1991, ACTA NEUROCHIR, V113, P149, DOI 10.1007/BF01403201
   GUTHRIE BL, 1989, J NEUROSURG, V70, P649, DOI 10.3171/jns.1989.70.4.0649
   HOOD TW, 1986, J NEUROSURG, V65, P172, DOI 10.3171/jns.1986.65.2.0172
   KONDZIOLKA D, 1995, SURG NEUROL, V43, P558, DOI 10.1016/0090-3019(95)00009-7
   KONOVALOV AN, 1990, J NEUROSURG, V73, P181, DOI 10.3171/jns.1990.73.2.0181
   KRATIMENOS GP, 1992, ACTA NEUROCHIR, V116, P164, DOI 10.1007/BF01540871
   LOBATO RD, 1989, J NEUROSURG, V71, P466
   Massager N, 2000, J NEUROSURG, V93, P951, DOI 10.3171/jns.2000.93.6.0951
   MATHISEN JR, 1987, SURG NEUROL, V28, P100, DOI 10.1016/0090-3019(87)90080-2
   NAUTA HJW, 1987, NEUROSURGERY, V20, P476, DOI 10.1227/00006123-198703000-00022
   NEAL JH, 1993, NEUROSURGERY, V32, P473, DOI 10.1227/00006123-199303000-00024
   Nicolato A, 1997, SURG NEUROL, V48, P267, DOI 10.1016/S0090-3019(97)00123-7
   OSTERTAG CB, 1980, SURG NEUROL, V14, P275
   PATTISAPU JV, 1989, PEDIATR NEUROSCI, V15, P62, DOI 10.1159/000120444
   POOL JL, 1968, J NEUROSURG, V29, P164, DOI 10.3171/jns.1968.29.2.0164
   REIGEL DH, 1979, CHILD BRAIN, V5, P329
   RYKEN TC, 1992, STEREOT FUNCT NEUROS, V59, P111, DOI 10.1159/000098926
   SAHNI KS, 1985, APPL NEUROPHYSIOL, V48, P488
   Sawin PD, 1998, SURG NEUROL, V49, P640, DOI 10.1016/S0090-3019(97)00435-7
   Selvapandian S, 1999, ACTA NEUROCHIR, V141, P721, DOI 10.1007/s007010050367
   STECK J, 1995, SURG NEUROL, V43, P563, DOI 10.1016/0090-3019(95)00156-5
   Thomas D G, 1988, Br J Neurosurg, V2, P235, DOI 10.3109/02688698808992674
   Valdes-Gorcia J, 1998, ACTA NEUROCHIR, V140, P899, DOI 10.1007/s007010050191
NR 37
TC 50
Z9 52
U1 0
U2 4
PU ELSEVIER SCIENCE INC
PI NEW YORK
PA 360 PARK AVE SOUTH, NEW YORK, NY 10010-1710 USA
SN 0090-3019
J9 SURG NEUROL
JI Surg. Neurol.
PD OCT
PY 2003
VL 60
IS 4
BP 311
EP 320
DI 10.1016/S0090-3019(03)00379-3
PG 10
WC Clinical Neurology; Surgery
WE Science Citation Index Expanded (SCI-EXPANDED)
SC Neurosciences & Neurology; Surgery
GA 724DT
UT WOS:000185472400008
PM 14505847
DA 2023-08-27
ER

PT J
AU Siu, IM
   Tyler, BM
   Chen, JX
   Eberhart, CG
   Thomale, UW
   Olivi, A
   Jallo, GI
   Riggins, GJ
   Gallia, GL
AF Siu, I-Mei
   Tyler, Betty M.
   Chen, James X.
   Eberhart, Charles G.
   Thomale, Ulrich-Wilhelm
   Olivi, Alessandro
   Jallo, George I.
   Riggins, Gregory J.
   Gallia, Gary L.
TI Establishment of a human glioblastoma stemlike brainstem rodent tumor
   model Laboratory investigation
SO JOURNAL OF NEUROSURGERY-PEDIATRICS
LA English
DT Article
DE brainstem; animal model; human; glioma; glioblastoma; neurospheres;
   stemlike cells
ID GLIOMAS; CELLS
AB Object. Diffuse brainstem tumors are the most difficult type of pediatric CNS malignancy to treat. These inoperable lesions are treated with radiation alone or in combination with chemotherapy, and the survival rate is less than 10%. It is therefore essential to develop a reliable animal model to screen new therapeutic agents for the treatment of this type of tumor.
   Methods. A multipotent human glioblastoma stemlike neurosphere line, 060919, was established from a surgically resected glioblastoma specimen; when cells were implanted intracranially into athymic nude mice, they formed invasive, vascular tumors that exhibited the features of glioblastoma. Ten female Fischer 344 rats received an injection of 75,000 F98 rat glioma cells and 10 female athymic nude rats received an injection of 75,000 060919 human glioblastoma stemlike cells in the pontine tegmentum of the brainstem. A control group of 5 female Fischer rats received an injection of saline in the same location as the animals in the tumor groups. Kaplan-Meier curves were generated for survival, and brains were processed postmortem for histopathological investigation.
   Results. Both F98 cells and 060919 cells grew in 100% of the animals injected. Median survival of animals injected with F98 was 15 days, consistent with the authors' previous reports on the establishment of the brainstem tumor model using the F98 rat glioma line. Median survival of animals injected with 060919 was 31 days. Histopathological analysis of the tumors confirmed the presence of brainstem lesions in animals that received brainstem injections of F98 and in animals that received brainstem injections of 060919. The 060919 brainstem tumors histologically resembled glioblastoma.
   Conclusions. Tumor take and median survival were consistent for animals injected in the brainstem with either the established F98 rat glioma cell line or the 060919 human glioblastoma stemlike neurosphere line. Histopathological features of the 060919 brainstem tumors resembled glioblastoma. Establishment of this human glioblastoma stemlike brainstem animal model will improve the evaluation and identification of more efficacious agents for the treatment of high-grade brainstem tumors. (DOI: 10.3171/2010.3.PEDS09366)
C1 [Siu, I-Mei; Tyler, Betty M.; Chen, James X.; Olivi, Alessandro; Jallo, George I.; Riggins, Gregory J.; Gallia, Gary L.] Johns Hopkins Sch Med, Dept Neurosurg, Baltimore, MD USA.
   [Eberhart, Charles G.] Johns Hopkins Sch Med, Dept Pathol, Baltimore, MD USA.
   [Thomale, Ulrich-Wilhelm] Univ Med Berlin, Campus Virchow Klinikum, Charite, Selbstandiger Arbeitsbereich Padiatr Neurochirurg, Berlin, Germany.
C3 Johns Hopkins University; Johns Hopkins Medicine; Johns Hopkins
   University; Johns Hopkins Medicine; Free University of Berlin; Humboldt
   University of Berlin; Charite Universitatsmedizin Berlin
RP Gallia, GL (通讯作者)，Johns Hopkins Univ Hosp, Dept Neurosurg & Oncol, Baltimore, MD 21287 USA.
EM
RI Olivi, Alessandro/K-6531-2016
OI Olivi, Alessandro/0000-0002-4489-7564
FU Brain Tumor Funder
FX This work was supported by funding from the Brain Tumor Funders'
   Collaborative to Dr. Riggins.
CR Barth RF, 1998, J NEURO-ONCOL, V36, P91, DOI 10.1023/A:1005805203044
   Frazier JL, 2009, J NEUROSURG-PEDIATR, V3, P259, DOI 10.3171/2008.11.PEDS08281
   Galli R, 2004, CANCER RES, V64, P7011, DOI 10.1158/0008-5472.CAN-04-1364
   Jallo GI, 2006, CHILD NERV SYST, V22, P1519, DOI 10.1007/s00381-006-0174-8
   Lee J, 2006, CANCER CELL, V9, P391, DOI 10.1016/j.ccr.2006.03.030
   Lee KM, 2005, TRIBOL LETT, V18, P1, DOI 10.1007/s11249-004-1698-2
   Parsons DW, 2008, SCIENCE, V321, P1807, DOI 10.1126/science.1164382
   Singh SK, 2004, NATURE, V432, P396, DOI 10.1038/nature03128
   Thomale UW, 2009, CHILD NERV SYST, V25, P433, DOI 10.1007/s00381-008-0767-5
NR 9
TC 15
Z9 15
U1 0
U2 5
PU AMER ASSOC NEUROLOGICAL SURGEONS
PI ROLLING MEADOWS
PA 5550 MEADOWBROOK DRIVE, ROLLING MEADOWS, IL 60008 USA
SN 1933-0707
J9 J NEUROSURG-PEDIATR
JI J. Neurosurg.-Pediatr.
PD JUL
PY 2010
VL 6
IS 1
BP 92
EP 97
DI 10.3171/2010.3.PEDS09366
PG 6
WC Clinical Neurology; Pediatrics; Surgery
WE Science Citation Index Expanded (SCI-EXPANDED)
SC Neurosciences & Neurology; Pediatrics; Surgery
GA 615BV
UT WOS:000279107100015
PM 20593994
DA 2023-08-27
ER

PT J
AU Li, D
   Hao, SY
   Wu, Z
   Zhang, LW
   Zhang, JT
AF Li, Da
   Hao, Shu-Yu
   Wu, Zhen
   Zhang, Li-Wei
   Zhang, Jun-Ting
TI Primary medulla oblongata teratomas Report of 2 cases
SO JOURNAL OF NEUROSURGERY-PEDIATRICS
LA English
DT Article
DE brainstem; germ cell tumor; medulla oblongata; teratoma; oncology
ID INTRACRANIAL IMMATURE TERATOMA; OF-THE-LITERATURE; NEOADJUVANT
   CHEMOTHERAPY; MIDBRAIN
AB Medulla oblongata teratomas are rare. The authors report 2 new cases of teratomas that occurred exclusively in the medulla oblongata. The first case was in a 9-year-old boy who presented with a 6-month history of neck pain and repeated paroxysmal vomiting. Based on preoperative radiographic findings, the initial diagnosis was of an intraaxial medulla oblongata hemangioblastoma. Intraoperatively, the cystic component of the tumor was gray, gelatinous, and soft in consistency. The solid component was light pink, rubbery, and nodular in appearance, with an identifiable boundary. The lesion was completely removed. Histopathological investigation revealed a mature teratoma. Postoperatively, the patient was supported with ventilator assistance and received a tracheotomy, but died of intracranial infection. The second case was in a 10-year-old boy with intermittent headache for 1 month. Radiographs revealed an exophytic cystic and solid lesion with dorsal involvement of the medulla oblongata. The lesion was predominantly solid, pinkish gray, tenacious, and moderately vascularized, with clearly delineated surgical dissection planes. The histopathological examination confirmed a diagnosis of immature teratoma. Total resection was achieved, followed by postoperative chemotherapy. He was alive without recurrence of the lesion or symptoms at 59 months after surgery.
   Resection of medulla oblongata teratoma is challenging, with inherent surgical risks that are contingent on the tumor growth pattern. Teratomas should be considered in the differential diagnosis of brainstem lesions. Chemotherapy has been suggested for immature teratomas. Long-term follow-up and larger studies of teratomas in unusual locations are required to improve practitioners' understanding of this disease's treatment and outcomes.
C1 [Li, Da; Hao, Shu-Yu; Wu, Zhen; Zhang, Li-Wei; Zhang, Jun-Ting] Capital Med Univ, Beijing Tiantan Hosp, Dept Neurosurg, Beijing, Peoples R China.
C3 Capital Medical University
RP Zhang, JT (通讯作者)，Beijing Tiantan Hosp, Dept Neurosurg, Tiantan Xili 6, Beijing 100050, Peoples R China.
EM
CR Arslan E, 2007, PEDIATR NEUROSURG, V43, P338, DOI 10.1159/000103319
   Fukuoka K, 2014, J NEUROSURG-PEDIATR, V13, P38, DOI 10.3171/2013.9.PEDS1347
   Furtado SV, 2011, J CLIN NEUROSCI, V18, P296, DOI 10.1016/j.jocn.2010.03.061
   Goyal N, 2013, CHILD NERV SYST, V29, P2035, DOI 10.1007/s00381-013-2091-y
   Hao SY, 2013, WORLD J SURG ONCOL, V11, DOI 10.1186/1477-7819-11-274
   Kochi M, 2003, J NEUROSURG, V99, P106, DOI 10.3171/jns.2003.99.1.0106
   Koh EJ, 2009, J NEUROSURG-PEDIATR, V4, P137, DOI 10.3171/2009.3.PEDS08290
   Matsumoto K, 1998, CLIN NEUROL NEUROSUR, V100, P292, DOI 10.1016/S0303-8467(98)00050-X
   Tsuzuki N, 2001, ACTA NEUROCHIR, V143, P1303, DOI 10.1007/PL00010098
NR 9
TC 4
Z9 4
U1 0
U2 0
PU AMER ASSOC NEUROLOGICAL SURGEONS
PI ROLLING MEADOWS
PA 5550 MEADOWBROOK DRIVE, ROLLING MEADOWS, IL 60008 USA
SN 1933-0707
EI 1933-0715
J9 J NEUROSURG-PEDIATR
JI J. Neurosurg.-Pediatr.
PD SEP
PY 2014
VL 14
IS 3
BP 296
EP 300
DI 10.3171/2014.6.PEDS1423
PG 5
WC Clinical Neurology; Pediatrics; Surgery
WE Science Citation Index Expanded (SCI-EXPANDED)
SC Neurosciences & Neurology; Pediatrics; Surgery
GA AN6EX
UT WOS:000340688800014
PM 24995819
OA Bronze
DA 2023-08-27
ER

PT J
AU Hashizume, R
   Smirnov, I
   Liu, S
   Phillips, JJ
   Hyer, J
   McKnight, TR
   Wendland, M
   Prados, M
   Banerjee, A
   Nicolaides, T
   Mueller, S
   James, CD
   Gupta, N
AF Hashizume, Rintaro
   Smirnov, Ivan
   Liu, Sharon
   Phillips, Joanna J.
   Hyer, Jeanette
   McKnight, Tracy R.
   Wendland, Michael
   Prados, Michael
   Banerjee, Anu
   Nicolaides, Theodore
   Mueller, Sabine
   James, Charles D.
   Gupta, Nalin
TI Characterization of a diffuse intrinsic pontine glioma cell line:
   implications for future investigations and treatment
SO JOURNAL OF NEURO-ONCOLOGY
LA English
DT Article
DE Brainstem glioma; Animal model; Astrocytoma
ID HIGH-GRADE GLIOMAS; BRAIN-STEM GLIOMA; MODEL; XENOGRAFT; DISEASE; TUMORS
AB Diffuse intrinsic pontine gliomas arise almost exclusively in children, and despite advances in treatment, the majority of patients die within 2 years after initial diagnosis. Because of their infiltrative nature and anatomic location in an eloquent area of the brain, most pontine gliomas are treated without a surgical biopsy. The corresponding lack of tissue samples has resulted in a limited understanding of the underlying genetic and molecular biologic abnormalities associated with pontine gliomas, and is a substantial obstacle for the preclinical testing of targeted therapeutic agents for these tumors. We have established a human glioma cell line that originated from surgical biopsy performed on a patient with a pontine glioma. To insure sustainable in vitro propagation, tumor cells were modified with hTERT (human telomerase ribonucleoprotein reverse transcriptase), and with a luciferase reporter to enable non-invasive bioluminescence imaging. The hTERT modified cells are tumorigenic in athymic rodents, and produce brainstem tumors that recapitulate the infiltrative growth of brainstem gliomas in patients.
C1 [Hashizume, Rintaro; Smirnov, Ivan; Liu, Sharon; Phillips, Joanna J.; Hyer, Jeanette; McKnight, Tracy R.; Prados, Michael; Banerjee, Anu; Nicolaides, Theodore; Mueller, Sabine; James, Charles D.; Gupta, Nalin] Univ Calif San Francisco, Dept Neurol Surg, Brain Tumor Res Ctr, San Francisco, CA 94143 USA.
   [Phillips, Joanna J.] Univ Calif San Francisco, Dept Pathol, San Francisco, CA 94143 USA.
   [McKnight, Tracy R.; Wendland, Michael] Univ Calif San Francisco, Dept Radiol & Biomed Imaging, San Francisco, CA 94143 USA.
   [Banerjee, Anu; Nicolaides, Theodore; Mueller, Sabine; Gupta, Nalin] Univ Calif San Francisco, Dept Pediat, San Francisco, CA 94143 USA.
   [Mueller, Sabine] Univ Calif San Francisco, Dept Neurol, San Francisco, CA 94143 USA.
C3 University of California System; University of California San Francisco;
   University of California System; University of California San Francisco;
   University of California System; University of California San Francisco;
   University of California System; University of California San Francisco;
   University of California System; University of California San Francisco
RP Gupta, N (通讯作者)，Univ Calif San Francisco, Dept Neurol Surg, Brain Tumor Res Ctr, 505 Parnassus Ave,Room M779, San Francisco, CA 94143 USA.
EM
RI Prados, Michael/N-5220-2019
OI Nicolaides, Theodore/0000-0002-0637-1086
FU Pediatric Brain Tumor Foundation Institute; Timmy's Rainbow Foundation
   for Brainstem Tumor Research
FX This study was supported by the Pediatric Brain Tumor Foundation
   Institute Award to the University of California San Francisco, and
   Timmy's Rainbow Foundation for Brainstem Tumor Research. The authors
   also thank Dr. Susan Baker (St. Jude's Children's Hospital and Medical
   Center) for allowing permission to examine copy number data from a
   published data set of pediatric astrocytic tumors.
CR Aoki Y, 2012, J NEURO-ONCOL, V108, P29, DOI 10.1007/s11060-011-0796-x
   Becher OJ, 2010, CANCER RES, V70, P2548, DOI 10.1158/0008-5472.CAN-09-2503
   Caretti V, 2011, BRAIN PATHOL, V21, P441, DOI 10.1111/j.1750-3639.2010.00468.x
   Dinca EB, 2007, J NEUROSURG, V107, P610, DOI 10.3171/JNS-07/09/0610
   Donaldson SS, 2006, J CLIN ONCOL, V24, P1266, DOI 10.1200/JCO.2005.04.6599
   Finlay JL, 2005, J NEURO-ONCOL, V75, P253, DOI 10.1007/s11060-005-6747-7
   Gunther HS, 2008, ONCOGENE, V27, P2897, DOI 10.1038/sj.onc.1210949
   Hashizume R, 2010, J NEURO-ONCOL, V96, P151, DOI 10.1007/s11060-009-9954-9
   Huse JT, 2011, GLIA, V59, P1190, DOI 10.1002/glia.21165
   Johnson RA, 2010, NATURE, V466, P632, DOI 10.1038/nature09173
   Masui K, 2010, GLIA, V58, P1050, DOI 10.1002/glia.20986
   Monje M, 2011, P NATL ACAD SCI USA, V108, P4453, DOI 10.1073/pnas.1101657108
   Paugh BS, 2011, J CLIN ONCOL, V29, P3999, DOI 10.1200/JCO.2011.35.5677
   Paugh BS, 2010, J CLIN ONCOL, V28, P3061, DOI 10.1200/JCO.2009.26.7252
   Phillips HS, 2006, CANCER CELL, V9, P157, DOI 10.1016/j.ccr.2006.02.019
   Recinos PF, 2007, PEDIATR NEUROSURG, V43, P192, DOI 10.1159/000098831
   Sanai N, 2008, NEUROSURGERY, V63, P460, DOI 10.1227/01.NEU.0000324731.68843.74
   Sharp JR, 2010, EUR J CANCER, V46, P3271, DOI 10.1016/j.ejca.2010.06.115
   Wang K, 2007, GENOME RES, V17, P1665, DOI 10.1101/gr.6861907
   Wen VW, 2006, CANCER RES, V66, P10691, DOI 10.1158/0008-5472.CAN-06-0979
   Wu G, 2012, NAT GENET, V44, P251, DOI 10.1038/ng.1102
   Zarghooni M, 2010, J CLIN ONCOL, V28, P1337, DOI 10.1200/JCO.2009.25.5463
NR 22
TC 54
Z9 57
U1 0
U2 14
PU SPRINGER
PI NEW YORK
PA 233 SPRING ST, NEW YORK, NY 10013 USA
SN 0167-594X
J9 J NEURO-ONCOL
JI J. Neuro-Oncol.
PD DEC
PY 2012
VL 110
IS 3
BP 305
EP 313
DI 10.1007/s11060-012-0973-6
PG 9
WC Oncology; Clinical Neurology
WE Science Citation Index Expanded (SCI-EXPANDED)
SC Oncology; Neurosciences & Neurology
GA 038XV
UT WOS:000311208300001
PM 22983601
OA Green Submitted
DA 2023-08-27
ER

PT J
AU Buus-Gehrig, C
   Lehrnbecher, T
   Porto, L
   Becker, M
   Freiman, T
   Mittelbronn, M
   Bochennek, K
AF Buus-Gehrig, Constanze
   Lehrnbecher, Thomas
   Porto, Luciana
   Becker, Martina
   Freiman, Thomas
   Mittelbronn, Michel
   Bochennek, Konrad
TI Pontine tumor in a neonate: case report and analysis of the current
   literature
SO JOURNAL OF NEUROSURGERY-PEDIATRICS
LA English
DT Article
DE spontaneous regression; pontine tumor; newborn; biopsy; oncology
ID CEREBELLAR PILOCYTIC ASTROCYTOMAS; BRAIN-STEM LESION; SPONTANEOUS
   REGRESSION; SURVIVAL RATES; DIFFUSE; GLIOMA; REMISSION; CHILDREN; CANCER
AB Tumors of the central nervous system represent the largest group of solid tumors found in pediatric patients. Pilocytic astrocytoma is the most common pediatric glioma, mostly located in the posterior fossa. The majority of brainstem tumors, however, are classified as highly aggressive diffuse intrinsic pontine gliomas (DIPGs) and their prognosis is dismal.
   The authors report on the case of a neonate in whom MRI and neuropathological assessment were used to diagnose DIPG. Before initiation of the planned chemotherapy, the tumor regressed spontaneously, and the newborn exhibited a normal neurological development. Meanwhile, Illumina Human Methylation450 BeadChip analysis reclassified the tumor as pilocytic astrocytoma of the posterior fossa.
   In conclusion, the authors advocate not initiating immediate intensive therapy in newborns with brain tumors, even with classical appearance of a DIPG; rather, they would like to encourage a biopsy to define the best individual therapeutic approach and avoid ineffective chemotherapy.
C1 [Buus-Gehrig, Constanze; Lehrnbecher, Thomas; Becker, Martina; Bochennek, Konrad] Goethe Univ, Pediat Hematol & Oncol, Frankfurt, Germany.
   [Porto, Luciana] Goethe Univ, Dept Neuroradiol, Frankfurt, Germany.
   [Freiman, Thomas] Goethe Univ, Dept Neurosurg, Frankfurt, Germany.
   [Mittelbronn, Michel] Goethe Univ Frankfurt, Inst Neurol, Edinger Inst, Frankfurt, Germany.
   [Mittelbronn, Michel] Univ Luxembourg, LCSB, Luxembourg, Luxembourg.
   [Mittelbronn, Michel] Luxembourg Inst Hlth, NORLUX Neurooncol Lab, Luxembourg, Luxembourg.
   [Mittelbronn, Michel] LNS, Dudelenge, Luxembourg.
   [Mittelbronn, Michel] LCNP, Dudelenge, Luxembourg.
C3 Goethe University Frankfurt; Goethe University Frankfurt; Goethe
   University Frankfurt; Goethe University Frankfurt; University of
   Luxembourg; Luxembourg Institute of Health
RP Bochennek, K (通讯作者)，Goethe Univ, Frankfurt, Germany.
EM
RI Porto, Luciana/AAF-6355-2020; Freiman, Thomas/AAV-8494-2021
OI Porto, Luciana/0000-0003-0554-2493; Freiman, Thomas/0000-0002-2591-2829;
   Mittelbronn, Michel/0000-0002-2998-052X
FU Luxembourg National Research Fund (FNR) [FNR PEARL P16/BM/11192868]
FX Dr. Mittelbronnn would like to thank the Luxembourg National Research
   Fund (FNR) for their support (FNR PEARL P16/BM/11192868 grant).
CR Airewele G, 2007, J NEUROSURG, V107, P80, DOI 10.3171/PED-07/07/080
   Bilginer B, 2011, TURK NEUROSURG, V21, P22
   Buder T, 2015, PLOS COMPUT BIOL, V11, DOI 10.1371/journal.pcbi.1004662
   Burkhard C, 2003, J NEUROSURG, V98, P1170, DOI 10.3171/jns.2003.98.6.1170
   Capper D, 2018, NATURE, V555, P469, DOI 10.1038/nature26000
   CLARK GB, 1985, CANCER, V56, P1128, DOI 10.1002/1097-0142(19850901)56:5<1128::AID-CNCR2820560529>3.0.CO;2-J
   Cohen KJ, 2017, NEURO-ONCOLOGY, V19, P1025, DOI 10.1093/neuonc/nox021
   Collins VP, 2015, ACTA NEUROPATHOL, V129, P775, DOI 10.1007/s00401-015-1410-7
   Danielsson A, 2015, CLIN EPIGENETICS, V7, DOI 10.1186/s13148-015-0103-3
   Khuong-Quang DA, 2012, ACTA NEUROPATHOL, V124, P439, DOI 10.1007/s00401-012-0998-0
   Dunham C, 2015, CHILD NERV SYST, V31, P1729, DOI 10.1007/s00381-015-2735-1
   Esteller M, 2008, NEW ENGL J MED, V358, P1148, DOI 10.1056/NEJMra072067
   Frazier JL, 2009, J NEUROSURG-PEDIATR, V3, P259, DOI 10.3171/2008.11.PEDS08281
   Gaudino S, 2012, NEURORADIOLOGY, V54, P989, DOI 10.1007/s00234-012-1010-3
   Gnekow AK, 2012, NEURO-ONCOLOGY, V14, P1265, DOI 10.1093/neuonc/nos202
   Gunny RS, 2005, PEDIATR RADIOL, V35, P1086, DOI 10.1007/s00247-005-1546-z
   Hargrove D, 2006, LANCET ONCOL, V7, P241, DOI 10.1016/S1470-2045(06)70615-5
   Hobohm U, 2001, CANCER IMMUNOL IMMUN, V50, P391
   Lenard HG, 1998, NEUROPEDIATRICS, V29, P328, DOI 10.1055/s-2007-973589
   Loh JK, 2013, CHILD NERV SYST, V29, P2051, DOI 10.1007/s00381-013-2113-9
   Mager DL, 2006, J TRANSL MED, V4, DOI 10.1186/1479-5876-4-14
   Murray RD, 2011, J COMPUT ASSIST TOMO, V35, P495, DOI 10.1097/RCT.0b013e3182237cbf
   Ohgaki H, 2005, J NEUROPATH EXP NEUR, V64, P479, DOI 10.1093/jnen/64.6.479
   Poretti A, 2012, J MAGN RESON IMAGING, V35, P32, DOI 10.1002/jmri.22722
   Puget S, 2015, CHILD NERV SYST, V31, P1773, DOI 10.1007/s00381-015-2832-1
   Robison NJ, 2014, J NEURO-ONCOL, V119, P7, DOI 10.1007/s11060-014-1448-8
   Samadian M, 2016, NEUROSURG REV, V39, P335, DOI 10.1007/s10143-015-0672-9
   Schomerus L, 2007, PEDIATR RADIOL, V37, P399, DOI 10.1007/s00247-007-0424-2
   Sexton-Oates A, 2015, ANN ONCOL, V26, P1314, DOI 10.1093/annonc/mdv024
   Thompson WD, 2005, J NEUROSURG, V102, P65, DOI 10.3171/ped.2005.102.1.0065
   Vanan MI, 2015, FRONT ONCOL, V5, DOI 10.3389/fonc.2015.00237
   2011, NEUROSURGERY, V68, P1548, DOI DOI 10.1227/NEU.0B013E318214A66E
   2013, CLIN NEUROL NEUROSUR, V115, P1220, DOI DOI 10.1016/J.CLINEURO.2012.11.009
   2016, CHILD NERV SYST, V32, P1163, DOI DOI 10.1007/S00381-016-3045-Y
   2008, CANCER, V113, P566, DOI DOI 10.1002/CNCR.23584
   2014, PEDIAT BLOOD CANC, V61, P1180, DOI DOI 10.1002/PBC.24958
NR 36
TC 1
Z9 1
U1 0
U2 1
PU AMER ASSOC NEUROLOGICAL SURGEONS
PI ROLLING MEADOWS
PA 5550 MEADOWBROOK DRIVE, ROLLING MEADOWS, IL 60008 USA
SN 1933-0707
EI 1933-0715
J9 J NEUROSURG-PEDIATR
JI J. Neurosurg.-Pediatr.
PD MAY
PY 2019
VL 23
IS 5
BP 606
EP 612
DI 10.3171/2018.10.PEDS18215
PG 7
WC Clinical Neurology; Pediatrics; Surgery
WE Science Citation Index Expanded (SCI-EXPANDED)
SC Neurosciences & Neurology; Pediatrics; Surgery
GA HW1JT
UT WOS:000466439900010
PM 30771760
DA 2023-08-27
ER

PT J
AU Lesniak, MS
   Klem, JM
   Weingart, J
   Carson, BS
AF Lesniak, MS
   Klem, JM
   Weingart, J
   Carson, BS
TI Surgical outcome following resection of contrast-enhanced pediatric
   brainstem gliomas
SO PEDIATRIC NEUROSURGERY
LA English
DT Article
DE pediatric brainstem tumors; radiology; pathology; surgery
ID GAMMA-KNIFE RADIOSURGERY; HIGH-DOSE-CHEMOTHERAPY; STEREOTACTIC
   RADIOSURGERY; PILOCYTIC ASTROCYTOMA; HYPOTHALAMIC GLIOMAS; CHILDREN;
   TUMORS; RADIOTHERAPY; CARBOPLATIN; CLASSIFICATION
AB Background: The role of surgery in the management of gadolinium-enhancing pediatric brainstem lesions on magnetic resonance imaging (MRI) has been a matter of open debate. This clinical series correlates radiological and pathological findings to assess the role of contrast enhancement as an indication for surgery with respect to clinical outcome. Methods: We retrospectively reviewed the charts of all pediatric patients admitted to the Johns Hopkins Hospital with a diagnosis of a brainstem tumor between January 1985 and December 2000. Results: There were a total of 89 patients who met the inclusion criteria. Fifty-seven patients (64.0%) underwent surgical resection while 32 (36%) were treated with radiation and/ or chemotherapy. Of the surgical candidates, 57 (100%) had an accompanying MRI scan significant for an enhancing lesion in the midbrain, pons or the medulla. The pathology was consistent with juvenile pilocytic astrocytoma in 30 patients (52.6%) and glioblastoma multiforme in 12 patients (21.1%). The remaining cases consisted of 10 patients (17.5%) with fibrillary astrocytomas, 3 (5.3%) with gangliogliomas, 1 (1.8%) with an oligodendroglioma and 1 (1.8%) with a primitive neuroectodermal tumor. A total of 29 patients had a total surgical resection, 8 a near total resection (>90%), 15 a subtotal resection ( 50-90%) and 5 a partial resection (<50%). The progression-free survival of all patients was 71.9% at 3 years and 45.6% at 5 years. Conclusions: This case series illustrates that contrast-enhanced MRI has positive prognostic value in the management of pediatric brainstem gliomas. In our study, the majority of enhancing tumors were low grade and amenable to surgical intervention. Consequently, we recommend surgical resection and pathological diagnosis of all enhancing brainstem tumors with adjuvant therapy reserved for recurrent or unresectable cases. Copyright (C) 2003 S. Karger AG, Basel.
C1 Johns Hopkins Univ Hosp, Dept Neurol Surg, Baltimore, MD 21287 USA.
C3 Johns Hopkins University; Johns Hopkins Medicine
RP Lesniak, MS (通讯作者)，Johns Hopkins Univ Hosp, Dept Neurol Surg, 600 N Wolfe St,Meyer 8-161, Baltimore, MD 21287 USA.
EM
CR Abbott R, 1999, PRINCIPLES AND PRACTICE OF PEDIATRIC NEUROSURGERY, P629
   ALBRIGHT AL, 1993, NEUROSURGERY, V33, P1026
   *AM ASS NEUR SURG, 1982, PEDIAT NEUROSURGERY, P367
   BARKOVICH AJ, 1991, PEDIATR NEUROSURG, V16, P73, DOI 10.1159/000120511
   Bouffet E, 2000, CANCER, V88, P685, DOI 10.1002/(SICI)1097-0142(20000201)88:3<685::AID-CNCR27>3.0.CO;2-K
   Boviatsis EJ, 2001, MINIM INVAS NEUROSUR, V44, P226, DOI 10.1055/s-2001-19930
   Bowers DC, 2001, PEDIATR NEUROSURG, V34, P229, DOI 10.1159/000056027
   Broniscer A, 2000, J CLIN ONCOL, V18, P1246, DOI 10.1200/JCO.2000.18.6.1246
   Cartmill M, 1999, CHILD NERV SYST, V15, P235
   Cirillo S, 1988, Radiol Med, V76, P390
   DAVIS PC, 1991, AM J NEURORADIOL, V12, P293
   Dunkel IJ, 1996, PEDIATR NEUROSURG, V24, P263, DOI 10.1159/000121049
   Earnest F 4th, 1985, Clin Neurosurg, V32, P540
   EPSTEIN F, 1986, J NEUROSURG, V64, P11, DOI 10.3171/jns.1986.64.1.0011
   Epstein F, 1996, PEDIATR NEUROSURG, V24, P24, DOI 10.1159/000121011
   ETTINGER LJ, 1985, J NEURO-ONCOL, V3, P263, DOI 10.1007/BF00165188
   Fischbein NJ, 1996, PEDIATR NEUROSURG, V24, P9, DOI 10.1159/000121010
   Fisher PG, 2000, CANCER, V89, P1569, DOI 10.1002/1097-0142(20001001)89:7<1569::AID-CNCR22>3.0.CO;2-0
   Fisher PG, 1996, CONT NEUROSURG, V18, P1
   Freeman CR, 2000, INT J RADIAT ONCOL, V47, P561, DOI 10.1016/S0360-3016(00)00471-5
   Fuchs I, 2002, ACT NEUR S, V84, P85
   Hadjipanayis CG, 2002, STEREOT FUNCT NEUROS, V79, P13, DOI 10.1159/000069500
   Hadjipanayis CG, 2002, J NEUROSURG, V97, P56, DOI 10.3171/jns.2002.97.1.0056
   HIBI T, 1991, NEUROSURGERY, V31, P543
   HIRATO M, 1995, STEREOT FUNCT NEUROS, V64, P32, DOI 10.1159/000098762
   Huang CF, 1999, J NEUROSURG, V91, P563, DOI 10.3171/jns.1999.91.4.0563
   KAPLAN EL, 1958, J AM STAT ASSOC, V53, P457, DOI 10.2307/2281868
   Kato T, 1998, J NEURO-ONCOL, V37, P263, DOI 10.1023/A:1005866021835
   Lesser GJ, 2001, SEMIN RADIAT ONCOL, V11, P138, DOI 10.1053/srao.2001.21422
   Lewis J, 1997, INT J RADIAT ONCOL, V38, P925, DOI 10.1016/S0360-3016(97)00134-X
   Liu YM, 1998, JPN J CLIN ONCOL, V28, P474, DOI 10.1093/jjco/28.8.474
   Lopez-Aguilar E, 2000, ARCH MED RES, V31, P186, DOI 10.1016/S0188-4409(00)00051-5
   McLendon RE., 2000, PATHOLOGY TUMORS CEN
   Moghrabi A, 1995, PEDIATR NEUROSURG, V23, P293, DOI 10.1159/000120974
   Mulne AF, 2000, J PEDIAT HEMATOL ONC, V22, P41, DOI 10.1097/00043426-200001000-00008
   Osztie E, 2001, AM J NEURORADIOL, V22, P818
   POLLACK IF, 1993, J NEUROSURG, V78, P859, DOI 10.3171/jns.1993.78.6.0859
   REDDY A, 2000, NEUROONCOLOGY ESSENT, P352
   ROBERTSON PL, 1995, PEDIATR NEUROSURG, V22, P65, DOI 10.1159/000120879
   Schild SE, 1998, J NEURO-ONCOL, V40, P171, DOI 10.1023/A:1006193306286
   Silva MM, 2000, PEDIATR NEUROSURG, V33, P151, DOI 10.1159/000028996
   Valdes-Gorcia J, 1998, ACTA NEUROCHIR, V140, P899, DOI 10.1007/s007010050191
   Walter AW, 1998, MED PEDIATR ONCOL, V30, P28, DOI 10.1002/(SICI)1096-911X(199801)30:1<28::AID-MPO9>3.0.CO;2-2
   Walter AW, 2000, J PEDIAT HEMATOL ONC, V22, P247, DOI 10.1097/00043426-200005000-00010
   Weiner HL, 1997, PEDIATR NEUROSURG, V27, P12, DOI 10.1159/000121219
   YOUMANS JR, 1990, NEUROLOGICAL SURG, P3032
   Zagzag D, 2000, PEDIATRICS, V106, P1045, DOI 10.1542/peds.106.5.1045
NR 47
TC 45
Z9 55
U1 0
U2 2
PU KARGER
PI BASEL
PA ALLSCHWILERSTRASSE 10, CH-4009 BASEL, SWITZERLAND
SN 1016-2291
EI 1423-0305
J9 PEDIATR NEUROSURG
JI Pediatr. Neurosurg.
PY 2003
VL 39
IS 6
BP 314
EP 322
DI 10.1159/000075260
PG 9
WC Clinical Neurology; Pediatrics; Surgery
WE Science Citation Index Expanded (SCI-EXPANDED)
SC Neurosciences & Neurology; Pediatrics; Surgery
GA 765CL
UT WOS:000188249500006
PM 14734866
DA 2023-08-27
ER

PT J
AU Mursch, K
   Halatsch, ME
   Markakis, E
   Behnke-Mursch, J
AF Mursch, K
   Halatsch, ME
   Markakis, E
   Behnke-Mursch, J
TI Intrinsic brainstem tumours in adults: results of microneurosurgical
   treatment of 16 consecutive patients
SO BRITISH JOURNAL OF NEUROSURGERY
LA English
DT Article
DE adults; brainstem tumour; intrinsic lesion; microneurosurgery; pons
   tumour; prognosis
ID INTRAAXIAL ENDOPHYTIC TUMORS; AUDITORY EVOKED-POTENTIALS; AND/OR
   MEDULLA-OBLONGATA; PROGNOSTIC FACTORS; SURGICAL INDICATIONS;
   SPINAL-CORD; GLIOMAS; CHILDREN; CHILDHOOD; LESIONS
AB Intrinsic brainstem tumours in adults have a poor prognosis and surgical resection is rarely performed. Encouraged by successful operations on children performed in our department, we began a more aggressive strategy of open operations. Between 1986 and 1997, we operated upon 16 consecutive patients over 16 years of age (five female, 11 male, mean age 36.9 years) who were suffering from intrinsic tumours located in the pons and/or medulla oblongata. The extent of first open resection was 80 - 100% in two of the cases and more than 50% in nine cases. The mean survival time after the first occurrence of symptoms was 88.1 ( median 34.5) months, and 39.9 ( median 11) months after the first open operation. The rate of 5-year survival from the first occurrence of symptoms was 37.5% (25% after the first open surgical procedure). Thirteen out of 16 patients died within the follow-up period of at least 6.3 years, two of them within the immediate postoperative period. Eleven patients experienced a postoperative deterioration of symptoms from which only four recovered. Eight patients had from WHO grade II astrocytoma and a similar course as patients with higher-grade gliomas (n=4). Our results indicate that open microneurosurgery for intrinsic brainstem tumours is of questionable benefit for the patient. Although surgery offers the advantages of reliable confirmation of histopathology and may be associated with prolonged survival, neurological deterioration was common and, unlike in paediatric patients, often irreversible.
C1 Univ Gottingen, Dept Neurosurg, D-3400 Gottingen, Germany.
C3 University of Gottingen
RP Mursch, K (通讯作者)，Zent Klin, Dept Neurosurg, Robert Koch Allee 9, D-99438 Bad Berka, Germany.
EM
RI Halatsch, Marc-Eric/B-7807-2008
OI Halatsch, Marc-Eric/0000-0002-6981-2280
CR Abbott R, 1996, PEDIATR NEUROSURG, V25, P41, DOI 10.1159/000121095
   AHYAI A, 1990, NEUROSURG REV, V13, P45, DOI 10.1007/BF00638893
   ALBRIGHT AL, 1993, NEUROSURGERY, V33, P1026
   Alvarez JA, 1996, J NEUROSURG, V85, P1165, DOI 10.3171/jns.1996.85.6.1165
   ALVISI C, 1985, ACTA NEUROCHIR, V76, P12, DOI 10.1007/BF01403823
   Behnke J, 1996, CHILD NERV SYST, V12, P125, DOI 10.1007/BF00266812
   BEHNKE J, 1994, ADV NEUROS, V22, P65
   Behnke J, 1997, CHILD NERV SYST, V13, P122, DOI 10.1007/s003810050058
   Behnke J, 1997, CHILD NERV SYST, V13, P135, DOI 10.1007/s003810050059
   Bricolo A, 1995, Adv Tech Stand Neurosurg, V22, P261
   EPSTEIN CM, 1988, NEUROL CLIN, V6, P771, DOI 10.1016/S0733-8619(18)30842-9
   EPSTEIN F, 1985, CANCER, V56, P1804, DOI 10.1002/1097-0142(19851001)56:7+<1804::AID-CNCR2820561316>3.0.CO;2-5
   EPSTEIN F, 1988, J NEURO-ONCOL, V6, P309
   EPSTEIN F, 1986, J NEUROSURG, V64, P11, DOI 10.3171/jns.1986.64.1.0011
   EPSTEIN F, 1987, J NEUROSURG, V67, P483, DOI 10.3171/jns.1987.67.4.0483
   Frank F, 1988, Acta Neurochir Suppl (Wien), V42, P177
   GRIGSBY PW, 1989, CANCER-AM CANCER SOC, V63, P2124, DOI 10.1002/1097-0142(19890601)63:11<2124::AID-CNCR2820631109>3.0.CO;2-9
   Guillamo JS, 2001, BRAIN, V124, P2528, DOI 10.1093/brain/124.12.2528
   HARSH GR, 1990, NEUROLOGICAL SURG, P3094
   HEFFEZ DS, 1990, NEUROSURGERY, V27, P789, DOI 10.1227/00006123-199011000-00018
   HOFFMAN HJ, 1980, NEUROSURGERY, V7, P243, DOI 10.1227/00006123-198009000-00007
   NISHIO S, 1991, British Journal of Neurosurgery, V5, P265, DOI 10.3109/02688699109005186
   NISHIO S, 1988, J NEURO-ONCOL, V6, P245, DOI 10.1007/BF00163708
   Pell M. F., 1993, Annals Academy of Medicine Singapore, V22, P447
   PIERREKAHN A, 1993, J NEUROSURG, V79, P845, DOI 10.3171/jns.1993.79.6.0845
   RYOO MC, 1979, RADIOLOGY, V13, P503
   Schulz-Ertner D, 2000, RADIOTHER ONCOL, V57, P215, DOI 10.1016/S0167-8140(00)00230-9
   Thomas D G, 1988, Br J Neurosurg, V2, P235, DOI 10.3109/02688698808992674
   TOKURIKI Y, 1986, ACTA NEUROCHIR, V79, P67, DOI 10.1007/BF01407447
   WALSER H, 1982, SURG NEUROL, V18, P405, DOI 10.1016/0090-3019(82)90172-0
   WHITE HH, 1963, NEUROLOGY, V13, P292, DOI 10.1212/WNL.13.4.292
   Xu QW, 1997, SURG NEUROL, V48, P30, DOI 10.1016/S0090-3019(97)00125-0
   ZENTNER J, 1990, EUR ARCH PSY CLIN N, V239, P285, DOI 10.1007/BF01735052
   ZILELI M, 1988, J NEUROSURG, V69, P660, DOI 10.3171/jns.1988.69.5.0660
NR 34
TC 18
Z9 20
U1 0
U2 0
PU TAYLOR & FRANCIS LTD
PI ABINGDON
PA 2-4 PARK SQUARE, MILTON PARK, ABINGDON OR14 4RN, OXON, ENGLAND
SN 0268-8697
EI 1360-046X
J9 BRIT J NEUROSURG
JI Br. J. Neurosurg.
PD APR
PY 2005
VL 19
IS 2
BP 128
EP 136
DI 10.1080/02688690500145530
PG 9
WC Clinical Neurology; Surgery
WE Science Citation Index Expanded (SCI-EXPANDED)
SC Neurosciences & Neurology; Surgery
GA 958WK
UT WOS:000231478700004
PM 16120515
DA 2023-08-27
ER

PT J
AU Misuraca, KL
   Hu, G
   Barton, KL
   Chung, A
   Becher, OJ
AF Misuraca, Katherine L.
   Hu, Guo
   Barton, Kelly L.
   Chung, Alexander
   Becher, Oren J.
TI A Novel Mouse Model of Diffuse Intrinsic Pontine Glioma Initiated in
   Pax3-Expressing Cells
SO NEOPLASIA
LA English
DT Article
ID POSTERIOR NONAXIAL MESODERM; ACTIVATING ACVR1 MUTATIONS; HIGH-GRADE
   GLIOMAS; PEDIATRIC GLIOBLASTOMA; PROGENITOR CELLS; HISTONE H3.3; NEURAL
   PLATE; BRAIN; STEM; SUBGROUPS
AB Diffuse intrinsic pontine glioma (DIPG) is a rare and incurable brain tumor that arises predominately in children and involves the pons, a structure that along with the midbrain and medulla makes up the brainstem. We have previously developed genetically engineered mouse models of brainstem glioma using the RCAS/Tv-a system by targeting PDGF-B overexpression, p53 loss, and H3.3K27M mutation to Nestin-expressing brainstem progenitor cells of the neonatal mouse. Here we describe a novel mouse model targeting these same genetic alterations to Pax3-expressing cells, which in the neonatal mouse pons consist of a Pax3+/Nestin+/Sox2+ population lining the fourth ventricle and a Pax3+/NeuN+ parenchymal population. Injection of RCAS-PDGF-B into the brainstem of Pax3-Tv-a mice at postnatal day 3 results in 40% of mice developing asymptomatic low-grade glioma. A mixture of low-and high-grade glioma results from injection of Pax3-Tv-a;p53(fl/fl) mice with RCAS-PDGF-B and RCAS-Cre, with or without RCAS-H3.3K27M. These tumors are Ki67+, Nestin+, Olig2+, and largely GFAP- and can arise anywhere within the brainstem, including the classic DIPG location of the ventral pons. Expression of the H3.3K27M mutation reduces overall H3K27me3 as compared with tumors without the mutation, similar to what has been previously shown in human and mouse tumors. Thus, we have generated a novel genetically engineered mouse model of DIPG, which faithfully recapitulates the human disease and represents a novel platform with which to study the biology and treatment of this deadly disease.
C1 [Misuraca, Katherine L.; Hu, Guo; Barton, Kelly L.; Chung, Alexander; Becher, Oren J.] Duke Univ, Med Ctr, Div Pediat Hematol Oncol, Durham, NC 27710 USA.
   [Hu, Guo; Barton, Kelly L.; Chung, Alexander; Becher, Oren J.] Duke Univ, Med Ctr, Preston Robert Tisch Brain Tumor Ctr, Durham, NC 27710 USA.
   [Becher, Oren J.] Duke Univ, Med Ctr, Dept Pathol, Durham, NC 27710 USA.
C3 Duke University; Duke University; Duke University
RP Becher, OJ (通讯作者)，Duke Univ, Med Ctr, Dept Pediat, 308 Res Dr,LSRC B362, Durham, NC 27710 USA.
EM
RI Becher, Oren/GXN-3269-2022; Becher, Oren/AAI-7495-2020
OI Becher, Oren/0000-0003-4016-8492; Hu, Guo/0000-0003-0597-208X
FU Damon Runyon Cancer Research Foundation; U.S. Department of Defense;
   Rory David Deutsch Foundation; Pediatric Brain Tumor Foundation;
   National Institute of Neurological Disorders and Stroke of the National
   Institutes of Health [K02NS086917]
FX We would like to thank William Pavan, PhD, and Stacie Loftus, PhD, at
   the National Human Genome Research Institute for providing us with the
   Pax3-Tv-a mice, and Grigori Enikolopov, PhD, for the Nestin-CFPnuc
   transgenic mice. O. J. B. is supported by the Damon Runyon Cancer
   Research Foundation, U.S. Department of Defense, Rory David Deutsch
   Foundation, and Pediatric Brain Tumor Foundation. Research reported in
   this publication was supported by the National Institute of Neurological
   Disorders and Stroke of the National Institutes of Health under award
   number K02NS086917.
CR [Anonymous], 2013, ALL DEV MOUS BRAIN A
   Ballester LY, 2013, AM J SURG PATHOL, V37, P1357, DOI 10.1097/PAS.0b013e318294e817
   Bang AG, 1997, DEVELOPMENT, V124, P2075
   Bang AG, 1999, DEV BIOL, V212, P366, DOI 10.1006/dbio.1999.9319
   Barrow J, 2011, NEURO-ONCOLOGY, V13, P212, DOI 10.1093/neuonc/noq158
   Barton KL, 2013, PLOS ONE, V8, DOI 10.1371/journal.pone.0077639
   Becher OJ, 2010, GENE DEV, V24, P2233, DOI 10.1101/gad.1988010
   Becher OJ, 2010, CANCER RES, V70, P2548, DOI 10.1158/0008-5472.CAN-09-2503
   Buczkowicz P, 2014, NAT GENET, V46, P451, DOI 10.1038/ng.2936
   Castel D, 2015, ACTA NEUROPATHOL, V130, P815, DOI 10.1007/s00401-015-1478-0
   Chan KM, 2013, GENE DEV, V27, P985, DOI 10.1101/gad.217778.113
   Donaldson SS, 2006, J CLIN ONCOL, V24, P1266, DOI 10.1200/JCO.2005.04.6599
   Khuong-Quang DA, 2012, ACTA NEUROPATHOL, V124, P439, DOI 10.1007/s00401-012-0998-0
   Encinas JM, 2006, P NATL ACAD SCI USA, V103, P8233, DOI 10.1073/pnas.0601992103
   Fischbein NJ, 1996, PEDIATR NEUROSURG, V24, P9, DOI 10.1159/000121010
   Fontebasso AM, 2014, NAT GENET, V46, P462, DOI 10.1038/ng.2950
   Funato K, 2014, SCIENCE, V346, P1529, DOI 10.1126/science.1253799
   Hargrove D, 2006, LANCET ONCOL, V7, P241, DOI 10.1016/S1470-2045(06)70615-5
   Hoffman RM, 2015, NAT REV CANCER, V15, P451, DOI 10.1038/nrc3972
   Hou L, 2004, DEV DYNAM, V229, P54, DOI 10.1002/dvdy.10468
   Lewis PW, 2013, SCIENCE, V340, P857, DOI 10.1126/science.1232245
   Misuraca KL, 2015, FRONT ONCOL, V5, DOI 10.3389/fonc.2015.00172
   Misuraca KL, 2014, ACTA NEUROPATHOL COM, V2, DOI 10.1186/s40478-014-0134-6
   Monje M, 2011, P NATL ACAD SCI USA, V108, P4453, DOI 10.1073/pnas.1101657108
   Pani L, 2002, GENE DEV, V16, P676, DOI 10.1101/gad.969302
   Paugh BS, 2013, CANCER RES, V73, P6219, DOI 10.1158/0008-5472.CAN-13-1491
   Paugh BS, 2011, J CLIN ONCOL, V29, P3999, DOI 10.1200/JCO.2011.35.5677
   Paugh BS, 2010, J CLIN ONCOL, V28, P3061, DOI 10.1200/JCO.2009.26.7252
   Puget S, 2012, PLOS ONE, V7, DOI 10.1371/journal.pone.0030313
   Schwartzentruber J, 2012, NATURE, V482, P226, DOI [10.1038/nature10833, 10.1007/s13311-012-0137-6]
   Sharma MK, 2007, CANCER RES, V67, P890, DOI 10.1158/0008-5472.CAN-06-0973
   Sturm D, 2012, CANCER CELL, V22, P425, DOI 10.1016/j.ccr.2012.08.024
   Subashi E, 2015, J NEUROONCOL
   Tanabe Y, 1996, SCIENCE, V274, P1115, DOI 10.1126/science.274.5290.1115
   Tate MC, 2015, J COMP NEUROL, V523, P449, DOI 10.1002/cne.23690
   Taylor KR, 2014, NAT GENET, V46, P457, DOI 10.1038/ng.2925
   Wu G, 2014, NAT GENET, V46, P444, DOI 10.1038/ng.2938
   Wu G, 2012, NAT GENET, V44, P251, DOI 10.1038/ng.1102
   Zarghooni M, 2010, J CLIN ONCOL, V28, P1337, DOI 10.1200/JCO.2009.25.5463
NR 39
TC 36
Z9 40
U1 0
U2 12
PU ELSEVIER SCIENCE INC
PI NEW YORK
PA 360 PARK AVE SOUTH, NEW YORK, NY 10010-1710 USA
SN 1476-5586
J9 NEOPLASIA
JI Neoplasia
PD JAN
PY 2016
VL 18
IS 1
BP 60
EP 70
DI 10.1016/j.neo.2015.12.002
PG 11
WC Oncology
WE Science Citation Index Expanded (SCI-EXPANDED)
SC Oncology
GA DB8YK
UT WOS:000368802900006
PM 26806352
OA gold, Green Published
DA 2023-08-27
ER

PT J
AU Anderson, RCE
   Kennedy, B
   Yanes, CL
   Garvin, J
   Needle, M
   Canoll, P
   Feldstein, NA
   Bruce, JN
AF Anderson, Richard C. E.
   Kennedy, Benjamin
   Yanes, Candix L.
   Garvin, James
   Needle, Michael
   Canoll, Peter
   Feldstein, Neil A.
   Bruce, Jeffrey N.
TI Convection-enhanced delivery of topotecan into diffuse intrinsic
   brainstem tumors in children
SO JOURNAL OF NEUROSURGERY-PEDIATRICS
LA English
DT Article
DE convection-enhanced delivery; topotecan; pediatric neurosurgery;
   brainstem tumor; pontine glioma; oncology
ID RECURRENT MALIGNANT GLIOMA; RAT-BRAIN; PSEUDOMONAS EXOTOXIN;
   INTRACEREBRAL CLYSIS; FUTURE STRATEGIES; DRUG-DELIVERY; GROWTH-FACTOR;
   REAL-TIME; GLIOBLASTOMA; CHEMOTHERAPY
AB Convection-enhanced delivery (CED) for the treatment of malignant gliomas is a technique that can deliver chemotherapeutic agents directly into the tumor and the surrounding interstitium through sustained, low-grade positive-pressure infusion. This allows for high local concentrations of drug within the tumor while minimizing systemic levels that often lead to dose-limiting toxicity. Diffuse intrinsic pontine gliomas (DIPGs) are universally fatal childhood tumors for which there is currently no effective treatment. In this report the authors describe CED of the topoisomerase inhibitor topotecan for the treatment of DIPG in 2 children.
   As part of a pilot feasibility study, the authors treated 2 pediatric patients with DIPG. Stereotactic biopsy with frozen section confirmation of glial tumor was followed by placement of bilateral catheters for CED of topotecan during the same procedure. The first patient underwent CED 210 days after initial diagnosis, after radiation therapy and at the time of tumor recurrence, with a total dose of 0.403 mg in 6.04 ml over 100 hours. Her Karnofsky Performance Status (KPS) score was 60 before CED and 50 posttreatment. Serial MRI initially demonstrated a modest reduction in tumor size and edema, but the tumor progressed and the patient died 49 days after treatment. The second patient was treated 24 days after the initial diagnosis prior to radiation with a total dose of 0.284 mg in 5.30 ml over 100 hours. Her KPS score was 70 before CED and 50 posttreatment. Serial MRI shnilarly demonstrated an initial modest reduction in tumor size. The patient subsequently underwent fractionated radiation therapy, but the tumor progressed and she died 120 days after treatment.
   Topotecan delivered by prolonged CED into the brainstem in children with DIPG is technically feasible. In both patients, high infusion rates (> 0.12 ml/hr) and high infusion volumes (> 2.8 ml) resulted in new neurological deficits and reduction in the ICPS score, but lower infusion rates (< 0.04 ml/hr) were well tolerated. While serial MRI showed moderate treatment effect, CED did not prolong survival in these 2 patients. More studies are needed to improve patient selection and determine the optimal flow rates for CED of chemotherapeutic agents into DIPG to maximize safety and efficacy. Clinical trial registration no.: NCT00324844. (http://thejns.org/doi/abs/10.3171/2012.10.PEDS12142)
C1 [Anderson, Richard C. E.; Kennedy, Benjamin; Yanes, Candix L.; Feldstein, Neil A.; Bruce, Jeffrey N.] Columbia Univ, Coll Phys & Surg, Dept Neurosurg, New York, NY USA.
   [Garvin, James; Needle, Michael] Columbia Univ, Coll Phys & Surg, Dept Oncol, New York, NY USA.
   [Canoll, Peter] Columbia Univ, Coll Phys & Surg, Dept Pathol & Cell Biol, New York, NY USA.
C3 Columbia University; Columbia University; Columbia University
RP Anderson, RCE (通讯作者)，Neurol Inst, 710 West 168th St,Room 213, New York, NY 10032 USA.
EM
RI Bruce, Jeffrey N/AAC-5307-2022
FU NIH [5R01CA89395]; National Center for Research Resources (NCRR), a
   component of the NIH [UL1 RR024156]; NIH Roadmap for Medical Research
FX This study was supported by NIH grant 5R01CA89395. This publication was
   made possible by grant number UL1 RR024156 from the National Center for
   Research Resources (NCRR), a component of the NIH, and the NIH Roadmap
   for Medical Research. Its contents are solely the responsibility of the
   authors and do not necessarily represent the official view of NCRR or
   NIH. Information on NCRR is available at the NCRR website. Information
   on Re-engineering the Clinical Research Enterprise can be obtained from
   the NIH Roadmap website.
CR ALBRIGHT AL, 1983, CANCER, V52, P2313, DOI 10.1002/1097-0142(19831215)52:12<2313::AID-CNCR2820521226>3.0.CO;2-I
   Allen JC, 1996, PEDIATR NEUROSURG, V24, P98, DOI 10.1159/000121024
   BOBO RH, 1994, P NATL ACAD SCI USA, V91, P2076, DOI 10.1073/pnas.91.6.2076
   Bruce JN, 2011, NEUROSURGERY, V69, P1272, DOI 10.1227/NEU.0b013e3182233e24
   Bruce JN, 2000, NEUROSURGERY, V46, P683, DOI 10.1097/00006123-200003000-00031
   Frazier JL, 2009, J NEUROSURG-PEDIATR, V3, P259, DOI 10.3171/2008.11.PEDS08281
   Friedman HS, 1999, CANCER-AM CANCER SOC, V85, P1160, DOI 10.1002/(SICI)1097-0142(19990301)85:5<1160::AID-CNCR21>3.0.CO;2-F
   Gerstner ER, 2007, J CLIN ONCOL, V25, P2306, DOI 10.1200/JCO.2006.10.0677
   Groothuis DR, 2000, BRAIN RES, V856, P281, DOI 10.1016/S0006-8993(99)02089-2
   Hargrove D, 2006, LANCET ONCOL, V7, P241, DOI 10.1016/S1470-2045(06)70615-5
   Iyer RR, 2011, J NEUROSURG, V115, P474, DOI 10.3171/2011.5.JNS11246
   Kaiser MG, 2000, NEUROSURGERY, V47, P1391, DOI 10.1097/00006123-200012000-00026
   Khatua S, 2011, CHILD NERV SYST, V27, P1391, DOI 10.1007/s00381-011-1468-z
   Kunwar S, 2003, ACT NEUR S, V88, P105
   LANGER R, 1990, SCIENCE, V249, P1527, DOI 10.1126/science.2218494
   Laske DW, 1997, NAT MED, V3, P1362, DOI 10.1038/nm1297-1362
   Lonser RR, 2007, J NEUROSURG, V107, P190, DOI 10.3171/JNS-07/07/0190
   Morrison PF, 1999, AM J PHYSIOL-REG I, V277, pR1218, DOI 10.1152/ajpregu.1999.277.4.R1218
   Murad GJA, 2007, J NEUROSURG, V106, P351, DOI 10.3171/jns.2007.106.2.351
   Murad GJA, 2006, CLIN CANCER RES, V12, P3145, DOI 10.1158/1078-0432.CCR-05-2583
   Occhiogrosso G, 2003, NEUROSURGERY, V52, P388, DOI 10.1227/01.NEU.0000043696.83722.8D
   Pardridge WM, 1997, J CEREBR BLOOD F MET, V17, P713, DOI 10.1097/00004647-199707000-00001
   Patel SJ, 2005, NEUROSURGERY, V56, P1243, DOI 10.1227/01.NEU.0000159649.71890.30
   PIERREKAHN A, 1993, J NEUROSURG, V79, P845, DOI 10.3171/jns.1993.79.6.0845
   Popperl G, 2005, EUR J NUCL MED MOL I, V32, P1018, DOI 10.1007/s00259-005-1819-7
   Rand RW, 2000, CLIN CANCER RES, V6, P2157
   Saito R, 2011, J NEUROSURG-PEDIATR, V7, P522, DOI 10.3171/2011.2.PEDS10407
   Sampson JH, 2003, J NEURO-ONCOL, V65, P27, DOI 10.1023/A:1026290315809
   Sandberg DI, 2002, J NEUROSURG, V96, P885, DOI 10.3171/jns.2002.96.5.0885
   Thomale UW, 2009, CHILD NERV SYST, V25, P433, DOI 10.1007/s00381-008-0767-5
   Thomale UW, 2009, CHILD NERV SYST, V25, P21, DOI 10.1007/s00381-008-0684-7
   Voges J, 2003, ANN NEUROL, V54, P479, DOI 10.1002/ana.10688
   Walker DA, 1999, ARCH DIS CHILD, V80, P558, DOI 10.1136/adc.80.6.558
   Weber FW, 2003, ACT NEUR S, V88, P93
   Yang WL, 2004, APPL RADIAT ISOTOPES, V61, P981, DOI 10.1016/j.apradiso.2004.05.071
   Yin DL, 2010, J NEUROSURG, V113, P240, DOI 10.3171/2010.2.JNS091744
NR 36
TC 57
Z9 58
U1 0
U2 19
PU AMER ASSOC NEUROLOGICAL SURGEONS
PI ROLLING MEADOWS
PA 5550 MEADOWBROOK DRIVE, ROLLING MEADOWS, IL 60008 USA
SN 1933-0707
J9 J NEUROSURG-PEDIATR
JI J. Neurosurg.-Pediatr.
PD MAR
PY 2013
VL 11
IS 3
BP 289
EP 295
DI 10.3171/2012.10.PEDS12142
PG 7
WC Clinical Neurology; Pediatrics; Surgery
WE Science Citation Index Expanded (SCI-EXPANDED)
SC Neurosciences & Neurology; Pediatrics; Surgery
GA 094DV
UT WOS:000315244100009
PM 23240851
OA Green Accepted
DA 2023-08-27
ER

PT J
AU Fonseca, A
   Afzal, S
   Bowes, L
   Crooks, B
   Larouche, V
   Jabado, N
   Perreault, S
   Johnston, DL
   Zelcer, S
   Fleming, A
   Scheinemann, K
   Silva, M
   Vanan, MI
   Mpofu, C
   Wilson, B
   Eisenstat, DD
   Lafay-Cousin, L
   Hukin, J
   Hawkins, C
   Bartels, U
   Bouffet, E
AF Fonseca, Adriana
   Afzal, Samina
   Bowes, Lynette
   Crooks, Bruce
   Larouche, Valerie
   Jabado, Nada
   Perreault, Sebastien
   Johnston, Donna L.
   Zelcer, Shayna
   Fleming, Adam
   Scheinemann, Katrin
   Silva, Mariana
   Vanan, Magimairajan Issai
   Mpofu, Chris
   Wilson, Beverly
   Eisenstat, David D.
   Lafay-Cousin, Lucie
   Hukin, Juliette
   Hawkins, Cynthia
   Bartels, Ute
   Bouffet, Eric
TI Pontine gliomas a 10-year population-based study: a report from The
   Canadian Paediatric Brain Tumour Consortium (CPBTC)
SO JOURNAL OF NEURO-ONCOLOGY
LA English
DT Article
DE Diffuse intrinsic pontine glioma (DIPG); Epidemiology; Brainstem tumors
ID FIELD RADIATION-THERAPY; LONG-TERM SURVIVORS; STEM TUMORS;
   HYPERFRACTIONATED RADIOTHERAPY; MOTEXAFIN-GADOLINIUM; STEREOTACTIC
   BIOPSY; HISTONE H3.3; DIFFUSE; CHILDREN; CHILDHOOD
AB Background Diffuse intrinsic pontine gliomas (DIPG) are midline gliomas that arise from the pons and the majority are lethal within a few months after diagnosis. Due to the lack of histological diagnosis the epidemiology of DIPG is not completely understood. The aim of this report is to provide population-based data to characterize the descriptive epidemiology of this condition in Canadian children. Patients and methods A national retrospective study of children and adolescents diagnosed with DIPG between 2000 and 2010 was undertaken. All cases underwent central review to determine clinical and radiological diagnostic characteristics. Crude incidence figures were calculated using age-adjusted (0-17 year) population data from Statistics Canada. Survival analyses were performed using the Kaplan-Meier method. Results A total of 163 patients with pontine lesions were identified. Central review determined one-hundred and forty-three patients who met clinical, radiological and/or histological criteria for diagnosis. We estimate an incidence rate of 1.9 DIPG/1,000,000 children/year in the Canadian population over a 10 years period. Median age at diagnosis was 6.8 years and 50.3% of patients were female. Most patients presented with cranial nerve palsies (76%) and ataxia (66%). Despite typical clinical (i.e. long length of symptoms) and radiological characteristics, (i.e. focal tumors, tumors with exophytic components or cystic components) histological confirmation reported three lesions to be low-grade gliomas and three were diagnosed as CNS embryonal tumor not otherwise specified (NOS). Conclusions Our study highlights the challenges associated with epidemiology studies on DIPG and the importance of central review for incidence rate estimations. It emphasizes that tissue biopsies are required for accurate histological and molecular diagnosis in patients presenting with pontine lesions and reinforces the limitations of radiological and clinical diagnosis in DIPG. Likewise, it underscores the urgent need to increase the availability and accessibility to clinical trials.
C1 [Fonseca, Adriana; Hawkins, Cynthia; Bartels, Ute; Bouffet, Eric] Univ Toronto, Hosp Sick Children, Div Haematol Oncol, 555 Univ Ave, Toronto, ON M5G 1X8, Canada.
   [Afzal, Samina; Crooks, Bruce] Dalhousie Univ, IWK Hlth Ctr, Halifax, NS B3K 6R8, Canada.
   [Afzal, Samina] Al Jalila Childrens Specialty Hosp, Dubai, U Arab Emirates.
   [Bowes, Lynette] Mem Univ, Janeway Child Hlth Ctr, St John, NF A1B 3V6, Canada.
   [Larouche, Valerie] Laval Univ, CHU Quebec Univ Laval, Quebec City, PQ G1V 4G2, Canada.
   [Jabado, Nada] McGill Univ, Montreal Childrens Hosp, Montreal, PQ H4A 3J1, Canada.
   [Perreault, Sebastien] Univ Montreal, Ctr Hosp Univ St Justine, Montreal, PQ H3T 1C5, Canada.
   [Johnston, Donna L.] Univ Ottawa, Childrens Hosp Eastern Ontario, Ottawa, ON K1H 8L1, Canada.
   [Zelcer, Shayna] Western Univ, London Hlth Sci Childrens Hosp, London, ON N6A 5A5, Canada.
   [Fleming, Adam; Scheinemann, Katrin] McMaster Univ, McMaster Childrens Hosp, Hamilton L8P 1H1, New Zealand.
   [Scheinemann, Katrin] Kantonsspital Aarau, Dept Pediat, Aarau, Switzerland.
   [Scheinemann, Katrin] Univ Basel, Univ Childrens Hosp Basel, Basel, Switzerland.
   [Silva, Mariana] Queens Univ, Kingston Gen Hosp, Kingston, ON K7L 2V7, Canada.
   [Vanan, Magimairajan Issai] Univ Manitoba, Canc Care Manitoba, Pediat & Child Hlth, Winnipeg, MB R3E 0V9, Canada.
   [Mpofu, Chris] Univ Saskatchewan, Jim Pattison Childrens Hosp, Saskatoon, SK S7N 0W8, Canada.
   [Wilson, Beverly; Eisenstat, David D.] Univ Alberta, Stollery Childrens Hosp, Edmonton, AB T6G 2B7, Canada.
   [Lafay-Cousin, Lucie] Univ Calgary, Alberta Childrens Hosp, Calgary, AB T3B 6A8, Canada.
   [Hukin, Juliette] Univ British Columbia, BC Childrens Hosp, Vancouver, BC V6H 3N1, Canada.
C3 University of Toronto; Hospital for Sick Children (SickKids); Dalhousie
   University; Memorial University Newfoundland; Laval University; McGill
   University; Universite de Montreal; Centre Hospitalier Universitaire
   Sainte-Justine; University of Ottawa; Children's Hospital of Eastern
   Ontario; Western University (University of Western Ontario);
   Kantonsspital Aarau AG (KSA); University of Basel; Queens University -
   Canada; University of Manitoba; University of Saskatchewan; Stollery
   Children's Hospital; University of Alberta; Alberta Childrens Hospital;
   University of Calgary; BC Childrens Hospital; University of British
   Columbia
RP Fonseca, A (通讯作者)，Univ Toronto, Hosp Sick Children, Div Haematol Oncol, 555 Univ Ave, Toronto, ON M5G 1X8, Canada.
EM
RI Eisenstat, David/HZH-9682-2023; Fonseca, Adriana/K-3730-2019; Fleming,
   Adam J/ABB-2202-2021; Jabado, Nada/AAN-4026-2020; Bouffet,
   Eric/AAB-8999-2020; Eisenstat, David/AAA-4653-2020
OI Eisenstat, David/0000-0002-5976-0798; Fonseca,
   Adriana/0000-0002-6382-6833; Fleming, Adam J/0000-0002-7194-3220; Hukin,
   Juliette/0000-0002-8727-8456; Crooks, Bruce/0000-0003-4715-4536
FU "We love you Connie Foundation". Toronto, Canada
FX AF is supported by "We love you Connie Foundation". Toronto, Canada.
CR ALBRIGHT AL, 1983, CANCER, V52, P2313, DOI 10.1002/1097-0142(19831215)52:12<2313::AID-CNCR2820521226>3.0.CO;2-I
   Albright AL, 1996, PEDIATR NEUROSURG, V24, P252
   ALBRIGHT AL, 1993, NEUROSURGERY, V33, P1026
   Aquino-Parsons C, 2008, PEDIATR BLOOD CANCER, V50, P397, DOI 10.1002/pbc.21057
   BARKOVICH AJ, 1991, PEDIATR NEUROSURG, V16, P73, DOI 10.1159/000120511
   Bradley KA, 2013, INT J RADIAT ONCOL, V85, pE55, DOI 10.1016/j.ijrobp.2012.09.004
   Bradley KA, 2008, NEURO-ONCOLOGY, V10, P752, DOI 10.1215/15228517-2008-043
   Broniscer A, 2008, CANCER, V113, P566, DOI 10.1002/cncr.23584
   Buczkowicz P, 2014, ACTA NEUROPATHOL, V128, P573, DOI 10.1007/s00401-014-1319-6
   Buczkowicz P, 2014, NAT GENET, V46, P451, DOI 10.1038/ng.2936
   Cage TA, 2013, CHILD NERV SYST, V29, P1313, DOI 10.1007/s00381-013-2101-0
   Chiang J, 2020, ACTA NEUROPATHOL COM, V8, DOI 10.1186/s40478-020-00930-9
   Cohen KJ, 2011, NEURO-ONCOLOGY, V13, P410, DOI 10.1093/neuonc/noq205
   Khuong-Quang DA, 2012, ACTA NEUROPATHOL, V124, P439, DOI 10.1007/s00401-012-0998-0
   Freeman CR, 1996, CANCER-AM CANCER SOC, V77, P555
   Freeman CR, 1996, PEDIATR NEUROSURG, V24, P103, DOI 10.1159/000121025
   Hamisch C, 2017, J NEUROSURG-PEDIATR, V20, P261, DOI 10.3171/2017.2.PEDS1665
   Hargrove D, 2006, LANCET ONCOL, V7, P241, DOI 10.1016/S1470-2045(06)70615-5
   Hoffman LM, 2018, J CLIN ONCOL, V36, P1963, DOI 10.1200/JCO.2017.75.9308
   Jackson S, 2013, J NEURO-ONCOL, V114, P339, DOI 10.1007/s11060-013-1189-0
   Johnson KJ, 2014, CANCER EPIDEM BIOMAR, V23, P2716, DOI 10.1158/1055-9965.EPI-14-0207
   Korones DN, 2008, PEDIATR BLOOD CANCER, V50, P227, DOI 10.1002/pbc.21154
   Leach JL, 2020, NEURO-ONCOLOGY, DOI [10.1093/neuonc/noaa140, DOI 10.1093/NEUONC/NOAA140]
   Louis DN, 2016, ACTA NEUROPATHOL, V131, P803, DOI 10.1007/s00401-016-1545-1
   Ostrom QT, 2018, NEURO-ONCOLOGY, V20, P1, DOI 10.1093/neuonc/noy131
   Puget S, 2015, CHILD NERV SYST, V31, P1773, DOI 10.1007/s00381-015-2832-1
   Puget Stephanie, 2012, Am Soc Clin Oncol Educ Book, P629, DOI 10.14694/EdBook_AM.2012.32.629
   Roujeau T, 2007, J NEUROSURG, V107, P1, DOI 10.3171/PED-07/07/001
   Schwartzentruber J, 2012, NATURE, V482, P226, DOI [10.1038/nature10833, 10.1007/s13311-012-0137-6]
   Sharp JR, 2010, EUR J CANCER, V46, P3271, DOI 10.1016/j.ejca.2010.06.115
   Smith MA, 1998, J NATL CANCER I, V90, P1269, DOI 10.1093/jnci/90.17.1269
   Statistics Canada, 2019, CAN POP EST AG SEX
   Sufit A, 2012, J NEUROSURG-PEDIATR, V10, P81, DOI 10.3171/2012.3.PEDS11316
   Surveillance EaERSP, 2017, INC SEER 18, P18
   van Zanten SEMV, 2015, EXPERT REV ANTICANC, V15, P157, DOI 10.1586/14737140.2015.974563
   Wu G, 2012, NAT GENET, V44, P251, DOI 10.1038/ng.1102
NR 36
TC 8
Z9 8
U1 1
U2 7
PU SPRINGER
PI NEW YORK
PA ONE NEW YORK PLAZA, SUITE 4600, NEW YORK, NY, UNITED STATES
SN 0167-594X
EI 1573-7373
J9 J NEURO-ONCOL
JI J. Neuro-Oncol.
PD AUG
PY 2020
VL 149
IS 1
BP 45
EP 54
DI 10.1007/s11060-020-03568-8
EA JUL 2020
PG 10
WC Oncology; Clinical Neurology
WE Science Citation Index Expanded (SCI-EXPANDED)
SC Oncology; Neurosciences & Neurology
GA NF3CC
UT WOS:000545882500001
PM 32632896
DA 2023-08-27
ER

PT J
AU Kornreich, L
   Schwarz, M
   Karmazyn, B
   Cohen, IJ
   Shuper, A
   Michovitz, S
   Yaniv, I
   Fenig, E
   Horev, G
AF Kornreich, L
   Schwarz, M
   Karmazyn, B
   Cohen, IJ
   Shuper, A
   Michovitz, S
   Yaniv, I
   Fenig, E
   Horev, G
TI Role of MRI in the management of children with diffuse pontine tumors: a
   study of 15 patients and review of the literature
SO PEDIATRIC RADIOLOGY
LA English
DT Article
DE pontine tumors; MR; prognosis
ID BRAIN-STEM GLIOMAS; INTRAAXIAL ENDOPHYTIC TUMORS; AND/OR
   MEDULLA-OBLONGATA; FOLLOW-UP; CLASSIFICATION
AB Background: Pontine tumors carry the worst prognosis of all brain tumors. In most cases, the diagnosis is based solely on MR imaging, without biopsy. Objective: To describe the MR findings of pontine tumors at diagnosis and during follow-up and correlate those with prognosis and to assess the value of MR imaging in patient management compared to clinical evaluation. Materials and methods: Ninety-one MR scans of 15 children with diffuse pontine tumors were reviewed at diagnosis and during follow-up. The parameters analyzed were as follows: tumor extent, area, and volume; encasement of the basilar artery; presence of exophytic component; necrosis; cysts; hydrocephalus; and intensity and enhancement. Findings were correlated to length of progress] on-free and overall survival. Trends of amelioration or worsening on imaging were compared with the clinical findings. Results: Median length of progression-free survival was 10 months, and median survival was 20 months. Only hydrocephalus at presentation was associated with shorter progression-free survival (P=0.02). On the last examination of each patient, the craniocaudal diameter was significantly greater than at diagnosis (P=0.03). The concordance between the imaging and the clinical findings was good. Conclusion: MR is the mainstay for the diagnosis and management of pontine tumors. Cranial growth seems to be an ominous sign. However, the prognostic value of MR is limited. MR findings correlate well with the clinical examination.
C1 Tel Aviv Univ, Sackler Fac Med, IL-69978 Tel Aviv, Israel.
   Schneider Childrens Med Ctr, Dept Imaging, Petah Tiqwa, Israel.
   Schneider Childrens Med Ctr, Dept Hematol Oncol, Petah Tiqwa, Israel.
   Schneider Childrens Med Ctr, Dept Neurosurg, Petah Tiqwa, Israel.
   Rabin Med Ctr, Inst Oncol, Radiotherapy Unit, Petah Tiqwa, Israel.
C3 Tel Aviv University; Sackler Faculty of Medicine; Rabin Medical Center
RP Kornreich, L (通讯作者)，Tel Aviv Univ, Sackler Fac Med, IL-69978 Tel Aviv, Israel.
EM
RI Cohen, Ian/H-3358-2013
CR ALBRIGHT AL, 1993, NEUROSURGERY, V33, P1026
   BARKOVICH AJ, 1991, PEDIATR NEUROSURG, V16, P73, DOI 10.1159/000120511
   Behnke J, 1997, CHILD NERV SYST, V13, P122, DOI 10.1007/s003810050058
   Behnke J, 1997, CHILD NERV SYST, V13, P135, DOI 10.1007/s003810050059
   Benesch M, 2001, PEDIATR NEUROSURG, V35, P173, DOI 10.1159/000050418
   Bouffet E, 2000, CANCER, V88, P685, DOI 10.1002/(SICI)1097-0142(20000201)88:3<685::AID-CNCR27>3.0.CO;2-K
   Burger PC, 1996, PEDIATR NEUROSURG, V24, P35, DOI 10.1159/000121012
   Cartmill M, 1999, CHILD NERV SYST, V15, P235
   Curless RG, 2002, PEDIATR NEUROL, V26, P374, DOI 10.1016/S0887-8994(01)00418-0
   DIXON WJ, 1992, BMDP STAT SOFTWARE
   EPSTEIN FJ, 1993, J NEUROSURG, V78, P408, DOI 10.3171/jns.1993.78.3.0408
   Farmer JP, 2001, PEDIATR NEUROSURG, V34, P206, DOI 10.1159/000056021
   Fischbein NJ, 1996, PEDIATR NEUROSURG, V24, P9, DOI 10.1159/000121010
   Fisher PG, 2000, CANCER, V89, P1569, DOI 10.1002/1097-0142(20001001)89:7<1569::AID-CNCR22>3.0.CO;2-0
   Freeman CR, 1998, INT J RADIAT ONCOL, V40, P265, DOI 10.1016/S0360-3016(97)00572-5
   Galanis E, 2000, J NEUROSURG, V93, P201, DOI 10.3171/jns.2000.93.2.0201
   Guillamo JS, 2001, CURR OPIN NEUROL, V14, P711, DOI 10.1097/00019052-200112000-00006
   Hoffman HJ, 1996, PEDIATR NEUROSURG, V24, P256, DOI 10.1159/000121048
   Jallo GI, 2004, CHILD NERV SYST, V20, P143, DOI 10.1007/s00381-003-0870-6
   Kaplan AM, 1996, PEDIATR NEUROSURG, V24, P185, DOI 10.1159/000121036
   Kashimura H, 2003, NEUROSURGERY, V52, P1226, DOI 10.1227/01.NEU.0000058025.94734.A3
   KHATIB ZA, 1994, PEDIATR NEUROSURG, V20, P2, DOI 10.1159/000120759
   NELSON MD, 1994, RADIOLOGY, V191, P279, DOI 10.1148/radiology.191.1.8134588
   PACKER RJ, 1993, CANCER-AM CANCER SOC, V71, P2666, DOI 10.1002/1097-0142(19930415)71:8<2666::AID-CNCR2820710836>3.0.CO;2-K
   Walker DA, 1999, ARCH DIS CHILD, V80, P558, DOI 10.1136/adc.80.6.558
   Wiegell MR, 2000, RADIOLOGY, V217, P897, DOI 10.1148/radiology.217.3.r00nv43897
   Zagzag D, 2000, PEDIATRICS, V106, P1045, DOI 10.1542/peds.106.5.1045
NR 27
TC 25
Z9 28
U1 0
U2 1
PU SPRINGER
PI NEW YORK
PA ONE NEW YORK PLAZA, SUITE 4600, NEW YORK, NY, UNITED STATES
SN 0301-0449
EI 1432-1998
J9 PEDIATR RADIOL
JI Pediatr. Radiol.
PD SEP
PY 2005
VL 35
IS 9
BP 872
EP 879
DI 10.1007/s00247-005-1502-y
PG 8
WC Pediatrics; Radiology, Nuclear Medicine & Medical Imaging
WE Science Citation Index Expanded (SCI-EXPANDED)
SC Pediatrics; Radiology, Nuclear Medicine & Medical Imaging
GA 956QA
UT WOS:000231313700007
PM 15918050
DA 2023-08-27
ER

PT J
AU Roessler, K
   Heynold, E
   Coras, R
   Lucking, H
   Buchfelder, M
AF Roessler, Karl
   Heynold, Elisabeth
   Coras, Roland
   Luecking, Hannes
   Buchfelder, Michael
TI Successful Surgery of Exophytic Brainstem Glioma Mimicking
   Cerebellar-Pontine Angle Tumor: Case Report and Review of Literature
SO WORLD NEUROSURGERY
LA English
DT Review
DE Cerebellar-pontine angle; Complete resection; Pilocytic astrocytoma;
   Vestibular schwannoma hearing preservation
ID CEREBELLOPONTINE ANGLE; PILOCYTIC ASTROCYTOMA
AB BACKGROUND: Nontectal plate exophytic brainstem gliomas with pilocytic histology are rare and occur mainly in children. Because of their eloquent location, therapy usually consists of bioptic histologic verification and radio-therapy in case of progression.
   CASE DESCRIPTION: We report on a 43-year-old patient who presented with intermittent tinnitus and reduced hearing in his left ear, as well as a slight left-sided ataxia. Magnetic resonance imaging revealed a left-sided solid cystic cerebellar-pontine angle (CPA) tumor completely obliterating the CPA. Radiologically, a solid cystic vestibular schwannoma was diagnosed. Intraoperatively, a glossy-grayish, intensively bleeding tumor without any capsule or delineation to the brainstem or cranial nerves was resected using electrophysiologic monitoring. Postoperative histology revealed a pilocytic astrocytoma. Three months postoperatively, hearing was preserved (pure tone average-35 dB) and ataxia was equal to what it was preoperatively, gradually becoming better. Postoperative magnetic resonance scans demonstrated a complete resection of the tumor with completely restored brainstem contours.
   CONCLUSIONS: Exophytic brainstem gliomas may occur in the CPA and mimic vestibular schwannoma. Complete resection even with preserved hearing without neurologic deterioration may be feasible.
C1 [Roessler, Karl; Heynold, Elisabeth; Coras, Roland; Luecking, Hannes; Buchfelder, Michael] Univ Hosp Erlangen, Erlangen Univ Clin, Neurosurg Dept, Neuroradiol Dept, Erlangen, Germany.
C3 University of Erlangen Nuremberg
RP Roessler, K (通讯作者)，Univ Hosp Erlangen, Erlangen Univ Clin, Neurosurg Dept, Neuroradiol Dept, Erlangen, Germany.
EM
RI Rössler, Karl/AAN-5096-2021
OI Rössler, Karl/0000-0002-9249-8082
CR BEUTLER AS, 1995, NEUROSURGERY, V37, P125, DOI 10.1227/00006123-199507000-00019
   Dutta G, 2017, BRIT J NEUROSURG, P1
   Gass D, 2015, J NEURO-ONCOL, V122, P169, DOI 10.1007/s11060-014-1700-2
   KASANTIKUL V, 1980, ARCH OTOLARYNGOL, V106, P456
   KERNOHAN JW, 1948, J NEUROPATH EXP NEUR, V7, P349, DOI 10.1097/00005072-194810000-00001
   MILLEN SJ, 1985, AM J OTOL, V6, P503
   Mirone G, 2009, CHILD NERV SYST, V25, P247, DOI 10.1007/s00381-008-0690-9
   Recinos PF, 2007, PEDIATR NEUROSURG, V43, P192, DOI 10.1159/000098831
   SMIRNIOTOPOULOS JG, 1993, RADIOGRAPHICS, V13, P1131, DOI 10.1148/radiographics.13.5.8210595
   von Bueren AO, 2018, ACTA NEUROPATHOL, V136, P661, DOI 10.1007/s00401-018-1896-x
NR 10
TC 1
Z9 1
U1 0
U2 5
PU ELSEVIER SCIENCE INC
PI NEW YORK
PA STE 800, 230 PARK AVE, NEW YORK, NY 10169 USA
SN 1878-8750
EI 1878-8769
J9 WORLD NEUROSURG
JI World Neurosurg.
PD AUG
PY 2019
VL 128
BP 202
EP 205
DI 10.1016/j.wneu.2019.05.053
PG 4
WC Clinical Neurology; Surgery
WE Science Citation Index Expanded (SCI-EXPANDED)
SC Neurosciences & Neurology; Surgery
GA IJ4SX
UT WOS:000475895100168
PM 31100526
DA 2023-08-27
ER

PT J
AU Pan, CC
   Diplas, BH
   Chen, X
   Wu, YL
   Xiao, X
   Jiang, LP
   Geng, YB
   Xu, C
   Sun, Y
   Zhang, P
   Wu, WH
   Wang, Y
   Wu, Z
   Zhang, JT
   Jiao, YC
   Yan, H
   Zhang, LW
AF Pan, Changcun
   Diplas, Bill H.
   Chen, Xin
   Wu, Yuliang
   Xiao, Xiong
   Jiang, Liping
   Geng, Yibo
   Xu, Cheng
   Sun, Yu
   Zhang, Peng
   Wu, Wenhao
   Wang, Yu
   Wu, Zhen
   Zhang, Junting
   Jiao, Yuchen
   Yan, Hai
   Zhang, Liwei
TI Molecular profiling of tumors of the brainstem by sequencing of
   CSF-derived circulating tumor DNA
SO ACTA NEUROPATHOLOGICA
LA English
DT Article
DE Liquid biopsy; ctDNA; DIPG; Brainstem; H3K27M mutation
ID DIFFUSE INTRINSIC PONTINE; HIGH-GRADE; CEREBROSPINAL-FLUID; GLIOMA;
   MUTATIONS; CHILDREN; BIOPSY; ACVR1; MANAGEMENT; SUBGROUPS
AB Brainstem gliomas are molecularly heterogeneous diseases, many of which are difficult to safely surgically resect and have limited treatment options due to their eloquent location. These constraints pose challenges to biopsy, which limits the use of routine molecular profiling and identification of personalized therapies. Here, we explored the potential of sequencing of circulating tumor DNA (ctDNA) isolated from the cerebrospinal fluid (CSF) of brainstem glioma patients as a less invasive approach for tumor molecular profiling. CSF was obtained from patients either intraoperatively (91.2%, 52/57), from ventricular-peritoneal shunt (3.5%, 2/57), or by lumbar puncture (5.3%, 3/57), all prior to surgical manipulation of the tumor. Deep sequencing of glioma-associated genes was performed on CSF-derived ctDNA and, where available, matched blood and tumor DNA from 57 patients, including nine medullary and 23 diffuse intrinsic pontine gliomas (DIPG). At least one tumor-specific mutation was detected in over 82.5% of CSF ctDNA samples (47/57). In cases with primary tumors harboring at least one mutation, alterations were identified in the CSF ctDNA of 97.3% of cases (36/37). In over 83% (31/37) of cases, all primary tumor alterations were detected in the CSF, and in 91.9% (34/37) of cases, at least half of the alterations were identified. Among ten patients found to have primary tumors negative for mutations, 30% (3/10) had detectable somatic alterations in the CSF. Finally, mutation detection using plasma ctDNA was less sensitive than sequencing the CSF ctDNA (38% vs. 100%, respectively). Our study indicates that deep sequencing of CSF ctDNA is a reliable technique for detecting tumor-specific alterations in brainstem tumors. This approach may offer an alternative approach to stereotactic biopsy for molecular profiling of brainstem tumors.
C1 [Pan, Changcun; Chen, Xin; Wu, Yuliang; Xiao, Xiong; Geng, Yibo; Xu, Cheng; Sun, Yu; Zhang, Peng; Wu, Wenhao; Wang, Yu; Wu, Zhen; Zhang, Junting; Zhang, Liwei] Capital Med Univ, Beijing Tiantan Hosp, Dept Neurosurg, Nan Si Huan Xi Lu 119, Beijing 100070, Peoples R China.
   [Diplas, Bill H.; Yan, Hai] Duke Univ, Med Ctr, Dept Pathol, Preston Robert Tisch Brain Tumor Ctr, Durham, NC 27710 USA.
   [Jiang, Liping] Genetron Hlth Beijing Co Ltd, Beijing, Peoples R China.
   [Jiao, Yuchen] Chinese Acad Med Sci, Canc Hosp, Natl Canc Ctr, State Key Lab Mol Oncol,Lab Cell & Mol Biol, Beijing, Peoples R China.
   [Jiao, Yuchen] Peking Union Med Coll, Collaborat Innovat Ctr Canc Med, Beijing, Peoples R China.
   [Zhang, Liwei] China Natl Clin Res Ctr Neurol Dis, Nan Si Huan Xi Lu 119, Beijing 100070, Peoples R China.
C3 Capital Medical University; Duke University; Chinese Academy of Medical
   Sciences - Peking Union Medical College; Cancer Institute & Hospital -
   CAMS; Chinese Academy of Medical Sciences - Peking Union Medical
   College; Peking Union Medical College
RP Zhang, LW (通讯作者)，Capital Med Univ, Beijing Tiantan Hosp, Dept Neurosurg, Nan Si Huan Xi Lu 119, Beijing 100070, Peoples R China.; Yan, H (通讯作者)，Duke Univ, Med Ctr, Dept Pathol, Preston Robert Tisch Brain Tumor Ctr, Durham, NC 27710 USA.; Zhang, LW (通讯作者)，China Natl Clin Res Ctr Neurol Dis, Nan Si Huan Xi Lu 119, Beijing 100070, Peoples R China.
EM
RI jiao, yuchen/H-6313-2011
OI Xiao, Xiong/0000-0001-5137-8538; Diplas, Bill/0000-0001-9248-7351; Pan,
   Changcun/0000-0002-7183-9220
FU National Key Technology Research and Development Program of the Ministry
   of Science and Technology of China [2014BAI04B01, 2015BAI12B04]; Beijing
   Municipal Administration of Hospitals Clinical Medicine Development of
   Special Funding Support [ZYLX201608]; Beijing Municipal Bureau of Human
   Resources and Social Security [2017-ZZ-117]
FX Financial support was provided by the National Key Technology Research
   and Development Program of the Ministry of Science and Technology of
   China (Grant nos. 2014BAI04B01 and 2015BAI12B04), Beijing Municipal
   Administration of Hospitals Clinical Medicine Development of Special
   Funding Support (Grant no. ZYLX201608), and Grant for CP from Beijing
   Municipal Bureau of Human Resources and Social Security (Grant no.
   2017-ZZ-117).
CR Bettegowda C, 2014, SCI TRANSL MED, V6, DOI 10.1126/scitranslmed.3007094
   Buczkowicz P, 2014, NAT GENET, V46, P451, DOI 10.1038/ng.2936
   Bugiani M, 2017, ONCOTARGET, V8, P60447, DOI 10.18632/oncotarget.19726
   Castel D, 2015, ACTA NEUROPATHOL, V130, P815, DOI 10.1007/s00401-015-1478-0
   Cracolici V, 2018, ENDOCR PATHOL, V29, P9, DOI 10.1007/s12022-017-9491-6
   De Mattos-Arruda L, 2015, NAT COMMUN, V6, DOI 10.1038/ncomms9839
   Diaz LA, 2014, J CLIN ONCOL, V32, P579, DOI 10.1200/JCO.2012.45.2011
   Fedorov A, 2012, MAGN RESON IMAGING, V30, P1323, DOI 10.1016/j.mri.2012.05.001
   Fisher BJ, 2015, INT J RADIAT ONCOL, V91, P497, DOI 10.1016/j.ijrobp.2014.11.012
   Fontebasso AM, 2014, NAT GENET, V46, P462, DOI 10.1038/ng.2950
   Frazier JL, 2009, J NEUROSURG-PEDIATR, V3, P259, DOI 10.3171/2008.11.PEDS08281
   Gerasimidis Konstantinos, 2016, BMC Res Notes, V9, P365, DOI 10.1186/s13104-016-2171-7
   Hargrove D, 2006, LANCET ONCOL, V7, P241, DOI 10.1016/S1470-2045(06)70615-5
   Hoffman LM, 2016, ACTA NEUROPATHOL COM, V4, DOI 10.1186/s40478-015-0269-0
   Huang TY, 2017, ACTA NEUROPATHOL COM, V5, DOI 10.1186/s40478-017-0436-6
   Jansen MH, 2015, NEURO-ONCOLOGY, V17, P160, DOI 10.1093/neuonc/nou104
   Jiang T, 2017, THERANOSTICS, V7, P4753, DOI 10.7150/thno.21687
   Johung TB, 2017, CURR NEUROPHARMACOL, V15, P88, DOI 10.2174/1570159X14666160509123229
   Kadri S, 2017, J MOL DIAGN, V19, P43, DOI 10.1016/j.jmoldx.2016.07.012
   Kieran MW, 2016, J NEUROSURG-PEDIATR, V18, P390, DOI 10.3171/2015.6.PEDS15374
   Klimo P, 2013, J NEUROSURG-PEDIATR, V11, P274, DOI 10.3171/2012.11.PEDS12317
   Long WY, 2017, FRONT PHARMACOL, V8, DOI 10.3389/fphar.2017.00495
   Mackay A, 2017, CANCER CELL, V32, P520, DOI 10.1016/j.ccell.2017.08.017
   Mandell LR, 1999, INT J RADIAT ONCOL, V43, P959, DOI 10.1016/S0360-3016(98)00501-X
   Martinez-Ricarte F, 2018, CLIN CANC RES
   Nikbakht H, 2016, NAT COMMUN, V7, DOI 10.1038/ncomms11185
   Olar A, 2015, ACTA NEUROPATHOL, V129, P585, DOI 10.1007/s00401-015-1398-z
   Pan CC, 2016, J NEURO-ONCOL, V128, P445, DOI 10.1007/s11060-016-2131-z
   Pentsova EI, 2016, J CLIN ONCOL, V34, P2404, DOI 10.1200/JCO.2016.66.6487
   Puget S, 2015, CHILD NERV SYST, V31, P1773, DOI 10.1007/s00381-015-2832-1
   Puget S, 2012, PLOS ONE, V7, DOI 10.1371/journal.pone.0030313
   Roujeau T, 2011, CHILD NERV SYST, V27, P1735, DOI 10.1007/s00381-011-1538-2
   Saratsis AM, 2014, ACTA NEUROPATHOL, V127, P881, DOI 10.1007/s00401-013-1218-2
   Taylor KR, 2014, NAT GENET, V46, P457, DOI 10.1038/ng.2925
   Vanan MI, 2015, FRONT ONCOL, V5, DOI 10.3389/fonc.2015.00237
   Veldhuijzen van Zanten SEM, 2017, J NEUROONCOL
   Wang YX, 2015, P NATL ACAD SCI USA, V112, P9704, DOI 10.1073/pnas.1511694112
   Wu G, 2014, NAT GENET, V46, P444, DOI 10.1038/ng.2938
   Wu G, 2012, NAT GENET, V44, P251, DOI 10.1038/ng.1102
   Zhang LW, 2014, NAT GENET, V46, P726, DOI 10.1038/ng.2995
NR 40
TC 86
Z9 90
U1 0
U2 30
PU SPRINGER
PI NEW YORK
PA 233 SPRING ST, NEW YORK, NY 10013 USA
SN 0001-6322
EI 1432-0533
J9 ACTA NEUROPATHOL
JI Acta Neuropathol.
PD FEB
PY 2019
VL 137
IS 2
BP 297
EP 306
DI 10.1007/s00401-018-1936-6
PG 10
WC Clinical Neurology; Neurosciences; Pathology
WE Science Citation Index Expanded (SCI-EXPANDED)
SC Neurosciences & Neurology; Pathology
GA HJ7FH
UT WOS:000457360100007
PM 30460397
OA Green Accepted
DA 2023-08-27
ER

PT J
AU Fuentes, S
   Delsanti, C
   Metellus, P
   Peragut, JC
   Grisoli, F
   Regis, J
AF Fuentes, S
   Delsanti, C
   Metellus, P
   Peragut, JC
   Grisoli, F
   Regis, J
TI Brainstem metastases: Management using gamma knife radiosurgery
SO NEUROSURGERY
LA English
DT Article
DE brainstem tumors; gamma-knife radiosurgery; metastasis
ID SINGLE METASTASES; RANDOMIZED-TRIAL; RADIOTHERAPY; RADIATION; SURGERY
AB OBJECTIVE: Brainstem metastasis is an uncommon complication of systemic cancer, generally considered to have a highly unfavorable prognosis. Surgical risks are high and standard radiation or chemotherapy have little effect. The purpose of this study is to evaluate our experience using Gamma Knife radiosurgery (GKRS) for the management of brainstem metastasis.
   METHODS: Between July 1992 and March 2001, we treated 28 patients with brainstem metastasis using GKRS. Lesions were located in the pons in 17 patients, midbrain in nine, and medulla oblongata in two. At time of the radiosurgery, eight patients presented with another supratentorial metastasis. The most frequent primary tumor site was the lung (13 cases) followed by the melanoma in four cases, the kidney in two, and other locations in six. Only six patients underwent fractionated whole-brain radiation therapy. Mean marginal radiation dose for GKRS was 19.6 Gy (range, 11-30). Mean maximum diameter was 17.2 mm (range, 10-30).
   RESULTS: No GKRS-related morbidity was observed. Local tumor control was achieved in 92% of patients. Twenty-six patients have died. Death was related to the progression of the brainstem lesion in two cases. Mean and median survival after GKRS were 10.2 and 12 months, respectively. Follow-up periods in the two surviving patients were 12 and 13 months.
   CONCLUSION: The results of this small series demonstrate that GKRS can be a valuable modality for safe and effective management of brain stem metastasis. Owing to the high risk of surgical resection and low efficacy of medical treatment, radiosurgery can be proposed upfront.
C1 Timone Univ Hosp Ctr, Neurosurg Serv, Marseille, France.
C3 UDICE-French Research Universities; Aix-Marseille Universite
RP Fuentes, S (通讯作者)，Hop Timone Adulte, Serv Neurochirurg, Rue St Pierre 13, F-385 Marseille, France.
EM
CR ADLER JR, 1992, J NEUROSURG, V76, P444, DOI 10.3171/jns.1992.76.3.0444
   Auchter RM, 1996, INT J RADIAT ONCOL, V35, P27, DOI 10.1016/S0360-3016(96)85008-5
   Bindal AK, 1996, J NEUROSURG, V84, P748, DOI 10.3171/jns.1996.84.5.0748
   FLICKINGER JC, 1994, INT J RADIAT ONCOL, V28, P797, DOI 10.1016/0360-3016(94)90098-1
   Flickinger JC, 1996, NEUROSURG CLIN N AM, V7, P497
   Gaspar L, 1997, INT J RADIAT ONCOL, V37, P745, DOI 10.1016/S0360-3016(96)00619-0
   Grob JJ, 1998, EUR J CANCER, V34, P1187, DOI 10.1016/S0959-8049(98)00026-4
   Hasegawa T, 2003, NEUROSURGERY, V52, P1318, DOI 10.1227/01.NEU.0000064569.18914.DE
   Huang CF, 1999, J NEUROSURG, V91, P563, DOI 10.3171/jns.1999.91.4.0563
   Johnson JD, 1996, NEUROSURG CLIN N AM, V7, P337
   Lang FF, 1996, NEUROSURG CLIN N AM, V7, P459
   Matsumoto K, 1998, J NEUROSURG, V88, P141, DOI 10.3171/jns.1998.88.1.0141
   MEHTA MP, 1992, INT J RADIAT ONCOL, V24, P619, DOI 10.1016/0360-3016(92)90706-N
   PATCHELL RA, 1990, NEW ENGL J MED, V322, P494, DOI 10.1056/NEJM199002223220802
   Patchell RA, 1998, JAMA-J AM MED ASSOC, V280, P1485, DOI 10.1001/jama.280.17.1485
   Regis J, 2002, J NEUROSURG, V97, P588, DOI 10.3171/jns.2002.97.supplement_5.0588
   SAMAYA R, 1995, BRAIN TUMORS, P923
   Shiau CY, 1997, INT J RADIAT ONCOL, V37, P375, DOI 10.1016/S0360-3016(96)00497-X
   SMALLEY SR, 1992, J NEUROSURG, V77, P531, DOI 10.3171/jns.1992.77.4.0531
   Young RF, 1998, SEMIN SURG ONCOL, V14, P70
NR 20
TC 61
Z9 61
U1 0
U2 4
PU LIPPINCOTT WILLIAMS & WILKINS
PI PHILADELPHIA
PA 530 WALNUT ST, PHILADELPHIA, PA 19106-3261 USA
SN 0148-396X
J9 NEUROSURGERY
JI Neurosurgery
PD JAN
PY 2006
VL 58
IS 1
BP 37
EP 40
DI 10.1227/01.NEU.0000190655.95669.5C
PG 4
WC Clinical Neurology; Surgery
WE Science Citation Index Expanded (SCI-EXPANDED)
SC Neurosciences & Neurology; Surgery
GA 000IF
UT WOS:000234453800007
PM 16385327
DA 2023-08-27
ER

PT J
AU Zimmerman, RA
AF Zimmerman, RA
TI Neuroimaging of primary brainstem gliomas: Diagnosis and course
SO PEDIATRIC NEUROSURGERY
LA English
DT Article
DE brainstem glioma; pediatric brain tumors; MR imaging
ID POSTERIOR-FOSSA TUMORS; STEM GLIOMAS
AB MR imaging is the procedure of choice for identifying brainstem gliomas and classifying them anatomically. Serial studies show a poor prognosis for diffuse pontine gliomas, better outcome for exophytic medullary gliomas and an indolent course for most midbrain gliomas.
C1 CHILDRENS HOSP PHILADELPHIA,PHILADELPHIA,PA 19104.
C3 University of Pennsylvania; Pennsylvania Medicine; Childrens Hospital of
   Philadelphia
CR BILANIUK LT, 1980, RADIOLOGY, V14, P89
   EDWARDS MSB, 1994, J NEUROSURG, V80, P20, DOI 10.3171/jns.1994.80.1.0020
   Kaplan AM, 1996, PEDIATR NEUROSURG, V24, P185, DOI 10.1159/000121036
   PACKER RJ, 1993, CANCER-AM CANCER SOC, V71, P2666, DOI 10.1002/1097-0142(19930415)71:8<2666::AID-CNCR2820710836>3.0.CO;2-K
   ROBERTSON PL, 1995, PEDIATR NEUROSURG, V22, P65, DOI 10.1159/000120879
   SMITH RR, 1990, NEURORADIOLOGY, V32, P265, DOI 10.1007/BF00593044
   SUTTON LN, 1994, J NEUROSURG, V81, P443, DOI 10.3171/jns.1994.81.3.0443
   ZIMMERMAN RA, 1992, PEDIATR NEUROSURG, V18, P58, DOI 10.1159/000120644
NR 8
TC 27
Z9 29
U1 0
U2 1
PU KARGER
PI BASEL
PA ALLSCHWILERSTRASSE 10, CH-4009 BASEL, SWITZERLAND
SN 1016-2291
J9 PEDIATR NEUROSURG
JI Pediatr. Neurosurg.
PD JUL
PY 1996
VL 25
IS 1
BP 45
EP 53
DI 10.1159/000121096
PG 9
WC Clinical Neurology; Pediatrics; Surgery
WE Science Citation Index Expanded (SCI-EXPANDED)
SC Neurosciences & Neurology; Pediatrics; Surgery
GA WF932
UT WOS:A1996WF93200014
PM 9055335
DA 2023-08-27
ER

PT J
AU Najjar, M
   Taylor, A
   Agrawal, S
   Fojo, T
   Merkler, AE
   Rosenblum, MK
   Lennihan, L
   Kluger, MD
AF Najjar, Marc
   Taylor, Andrew
   Agrawal, Surbhi
   Fojo, Tito
   Merkler, Alexander E.
   Rosenblum, Marc K.
   Lennihan, Laura
   Kluger, Michael D.
TI Anti-Hu paraneoplastic brainstem encephalitis caused by a pancreatic
   neuroendocrine tumor presenting with central hypoventilation
SO JOURNAL OF CLINICAL NEUROSCIENCE
LA English
DT Article
DE Paraneoplastic syndrome; Brainstem encephalitis; Pancreatic
   neuroendocrine tumor
ID ENCEPHALOMYELITIS
AB Paraneoplastic neurological syndromes are rare autoimmune manifestations of malignancies associated with specific antibodies. Anti-Hu associated brainstem encephalitis, a well-described syndrome, usually presents subacutely with preferential involvement of the medulla. Anti-Hu antibodies target intraneuronal antigens and are therefore highly correlated with neurological syndromes when present concomitantly with a neoplasm. Reported is a case of anti-Hu brainstem encephalitis associated with a pancreatic neuroendocrine tumor (PNET) presenting with central hypoventilation. This is the first described case of brainstem encephalitis associated with a well-differentiated PNET as well as the first case of Anti-Hu antibodies associated with a PNET. There are no standardized protocols for the treatment of paraneoplastic brainstem encephalitis however, as in the present case, surgical resection and ontological treatment of the tumor is the first line treatment. (C) 2017 Elsevier Ltd. All rights reserved.
C1 [Najjar, Marc; Agrawal, Surbhi; Kluger, Michael D.] Columbia Univ, Med Ctr, Dept Surg, New York, NY 10027 USA.
   [Taylor, Andrew; Merkler, Alexander E.; Lennihan, Laura] Columbia Univ, Med Ctr, Dept Neurol, New York, NY 10027 USA.
   [Fojo, Tito] Columbia Univ, Med Ctr, Dept Med, Div Hematol Oncol, New York, NY 10027 USA.
   [Rosenblum, Marc K.] Mem Sloan Kettering Canc Ctr, Dept Pathol, Div Neuropathol, 1275 York Ave, New York, NY 10021 USA.
C3 Columbia University; Columbia University; Columbia University; Memorial
   Sloan Kettering Cancer Center
RP Najjar, M (通讯作者)，Columbia Univ, Med Ctr, Dept Surg, New York, NY 10027 USA.
EM
RI Agrawal, Surbhi/AAD-7469-2019
OI Agrawal, Surbhi/0000-0002-1155-491X; Kluger, Michael/0000-0003-4311-078X
CR Berger B, 2015, J NEUROL SCI, V352, P58, DOI 10.1016/j.jns.2015.03.027
   Brizzi MP, 2015, BMC CANCER, V15, DOI 10.1186/s12885-015-1923-4
   DALMAU J, 1992, MEDICINE, V71, P59, DOI 10.1097/00005792-199203000-00001
   Gomez-Choco MJ, 2007, J NEUROL NEUROSUR PS, V78, P1143, DOI 10.1136/jnnp.2007.117994
   Graus F, 2001, BRAIN, V124, P1138, DOI 10.1093/brain/124.6.1138
   O'Grady HL, 2008, EJSO-EUR J SURG ONC, V34, P324, DOI 10.1016/j.ejso.2007.07.209
   Saiz A, 2009, J NEUROL NEUROSUR PS, V80, P404, DOI 10.1136/jnnp.2008.158246
NR 7
TC 6
Z9 6
U1 0
U2 2
PU ELSEVIER SCI LTD
PI OXFORD
PA THE BOULEVARD, LANGFORD LANE, KIDLINGTON, OXFORD OX5 1GB, OXON, ENGLAND
SN 0967-5868
EI 1532-2653
J9 J CLIN NEUROSCI
JI J. Clin. Neurosci.
PD JUN
PY 2017
VL 40
BP 72
EP 73
DI 10.1016/j.jocn.2017.02.015
PG 2
WC Clinical Neurology; Neurosciences
WE Science Citation Index Expanded (SCI-EXPANDED)
SC Neurosciences & Neurology
GA FB1DR
UT WOS:000405884400019
PM 28256369
DA 2023-08-27
ER

PT J
AU Watanabe, T
   Sato, T
   Kishida, Y
   Ito, E
   Ichikawa, M
   Sakuma, J
   Nagatani, T
   Saito, K
AF Watanabe, Tadashi
   Sato, Taku
   Kishida, Yugo
   Ito, Eiji
   Ichikawa, Masahiro
   Sakuma, Jun
   Nagatani, Tetsuya
   Saito, Kiyoshi
TI Endoscopic resection of cystic pontine tumour: three case reports and a
   proposal for minimally invasive dual-endoscopic surgery
SO ACTA NEUROCHIRURGICA
LA English
DT Article
DE Dual-endoscopic; Endoscopic resection; Pontine cystic tumour;
   Subtemporal approach
ID SUPRASELLAR; REMOVAL
AB We describe a new endoscopic technique, the dual-endoscopic method, in which two independent endoscopic systems are simultaneously used in the subtemporal space and intraparenchymal field for patients with pontine cystic tumours. Tumours were successfully removed through a sheath placed in the brainstem via the subtemporal keyhole approach, with safety ensured by the second subtemporal endoscope.
   A purely endoscopic subtemporal approach was performed in three cases of pontine cystic tumour, including two low-grade gliomas and a recurrent chordoma. The gliomas showed nodules. Recurrent chordoma consisted of a mucinous component.
   Using a keyhole subtemporal approach and a 4.0-mm rigid endoscope with a high-definition camera, a transparent puncture needle containing a 2.7-mm rigid endoscope was inserted into the cyst. The tumour was removed using suction, biopsy forceps, and scissors simultaneously placed with the 2.7-mm endoscope in a transparent sheath.
   Since the 4.0-mm endoscope placed in the subtemporal space monitored all procedures of the 2.7-mm endoscope, tumour removal was performed safely without damaging the pons, cranial nerves, or vessels. We safely and successfully removed a cystic pontine tumour using a dual-endoscopic method. This new method ensures the safety of deep-seated endoscopic procedures via a minimally invasive transcranial approach.
C1 [Watanabe, Tadashi; Nagatani, Tetsuya] Nagoya Daini Red Cross Hosp, Dept Neurosurg, Showa Ku, Nagoya, Aichi 4668650, Japan.
   [Sato, Taku; Kishida, Yugo; Ichikawa, Masahiro; Sakuma, Jun; Saito, Kiyoshi] Fukushima Med Univ Sch Med, Dept Neurosurg, Fukushima, Japan.
   [Ito, Eiji] Chubu Rosai Hosp, Dept Neurosurg, Nagoya, Aichi, Japan.
C3 Japanese Red Cross Nagoya Daini Hospital; Fukushima Medical University
RP Watanabe, T (通讯作者)，Nagoya Daini Red Cross Hosp, Dept Neurosurg, Showa Ku, 2-9 Myoken Cho, Nagoya, Aichi 4668650, Japan.
EM
RI Watanabe, Tadashi/AAE-8965-2019
CR Horvath Z, 2000, ACTA NEUROCHIR, V142, P539, DOI 10.1007/s007010050466
   Jho HD, 1997, J NEUROSURG, V87, P44, DOI 10.3171/jns.1997.87.1.0044
   Kabil MS, 2005, J CRANIOFAC SURG, V16, P1070, DOI 10.1097/01.scs.0000198624.46501.e0
   Kassam Amin B, 2005, Neurosurg Focus, V19, pE6
   Kassam AB, 2009, J NEUROSURG, V110, P116, DOI 10.3171/2008.7.JNS08226
   Nishihara T, 2000, J NEUROSURG, V92, P1053, DOI 10.3171/jns.2000.92.6.1053
   OTSUKI T, 1994, NEUROSURGERY, V35, P751, DOI 10.1227/00006123-199410000-00026
   Taniguchi M, 1997, NEUROSURGERY, V41, P592, DOI 10.1097/00006123-199709000-00017
   Teo C, 2005, CHILD NERV SYST, V21, P696, DOI 10.1007/s00381-005-1204-7
   Watanabe T, 2011, NEUROL MED-CHIR, V51, P684, DOI 10.2176/nmc.51.684
NR 10
TC 13
Z9 15
U1 0
U2 4
PU SPRINGER WIEN
PI WIEN
PA SACHSENPLATZ 4-6, PO BOX 89, A-1201 WIEN, AUSTRIA
SN 0001-6268
EI 0942-0940
J9 ACTA NEUROCHIR
JI Acta Neurochir.
PD JUN
PY 2014
VL 156
IS 6
BP 1145
EP 1150
DI 10.1007/s00701-014-2085-x
PG 6
WC Clinical Neurology; Surgery
WE Science Citation Index Expanded (SCI-EXPANDED)
SC Neurosciences & Neurology; Surgery
GA AH8TR
UT WOS:000336411600014
PM 24740348
DA 2023-08-27
ER

PT J
AU Magdoom, KN
   Delgado, F
   Bohorquez, AC
   Brown, AC
   Carney, PR
   Rinaldi, C
   Mareci, TH
   Ewing, JR
   Sarntinoranont, M
AF Magdoom, Kulam Najmudeen
   Delgado, Francisco
   Bohorquez, Ana C.
   Brown, Alec C.
   Carney, Paul R.
   Rinaldi, Carlos
   Mareci, Thomas H.
   Ewing, James R.
   Sarntinoranont, Malisa
TI Longitudinal evaluation of tumor microenvironment in rat focal brainstem
   glioma using diffusion and perfusion MRI
SO JOURNAL OF MAGNETIC RESONANCE IMAGING
LA English
DT Article
DE tumor leakiness; tumor volume; dynamic contrast-enhanced MRI; brainstem
   glioma; diffusion imaging
ID BARRIER PERMEABILITY; MODEL SELECTION; IN-VIVO; PARAMETERS; CHILDREN;
   VOLUME
AB Background Brainstem gliomas are aggressive and difficult to treat. Growth of these tumors may be characterized with MRI methods. Purpose To visualize longitudinal changes in tumor volume, vascular leakiness, and tissue microstructure in an animal model of brainstem glioma. Study Type Prospective animal model. Animal Model Male Sprague-Dawley rats (n = 9) were imaged with 9L gliosarcoma cells infused into the pontine reticular formation of the brainstem. The MRI tumor microenvironment was studied at 3 and 10 days postimplantation of tumor cells. Field Strength/Sequence Diffusion tensor imaging (DTI) and dynamic contrast-enhanced (DCE)-MRI were performed at 4.7T using spin-echo multislice echo planar imaging and gradient echo multislice imaging, respectively. Assessment Tumor leakiness was assessed by the forward volumetric transfer constant, K-trans, estimated from DCE-MRI data. Tumor structure was evaluated with fractional anisotropy (FA) obtained from DTI. Tumor volumes, delineated by a T-1 map, T-2-weighted image, FA, and DCE signal enhancement were compared. Statistical Tests Changes in the assessed parameters within and across the groups (ie, rats 3 and 10 days post tumor cell implantation) were evaluated with Wilcoxon rank-sum tests. Results Day 3 tumors were visible mainly on contrast-enhanced images, while day 10 tumors were visible in both contrast-enhanced and diffusion-weighted images. Mean K-trans at day 10 was 41% lower than at day 3 (P = 0.23). In day 10 tumors, FA was regionally lower in the tumor compared to normal tissue (P = 0.0004), and tumor volume, segmented based on FA map, was significantly smaller (P <= 0.05) than that obtained from other contrasts. Data Conclusion Contrast-enhanced MRI was found to be more sensitive in detecting early-stage tumor boundaries than other contrasts. Areas of the tumor outlined by DCE-MRI and DTI were significantly different. Over the observed period of tumor growth, average vessel leakiness decreased with tumor progression.
C1 [Magdoom, Kulam Najmudeen; Sarntinoranont, Malisa] Univ Florida, Dept Mech & Aerosp Engn, Gainesville, FL 32611 USA.
   [Delgado, Francisco; Bohorquez, Ana C.; Rinaldi, Carlos; Mareci, Thomas H.; Sarntinoranont, Malisa] J Crayton Pruitt Family Dept Biomed Engn, Gainesville, FL USA.
   [Brown, Alec C.; Mareci, Thomas H.] Univ Florida, Dept Biochem & Mol Biol, Gainesville, FL 32610 USA.
   [Carney, Paul R.] Univ N Carolina, Dept Neurol, Chapel Hill, NC 27515 USA.
   [Carney, Paul R.] Univ N Carolina, Neurosci Ctr, Chapel Hill, NC 27515 USA.
   [Rinaldi, Carlos] Univ Florida, Dept Chem Engn, Gainesville, FL 32611 USA.
   [Ewing, James R.] Henry Ford Hosp, Dept Neurol, Detroit, MI 48202 USA.
C3 State University System of Florida; University of Florida; State
   University System of Florida; University of Florida; University of North
   Carolina; University of North Carolina Chapel Hill; University of North
   Carolina; University of North Carolina Chapel Hill; State University
   System of Florida; University of Florida; Henry Ford Health System;
   Henry Ford Hospital
RP Sarntinoranont, M (通讯作者)，Univ Florida, Dept Mech & Aerosp Engn, Gainesville, FL 32611 USA.
EM
RI Rinaldi, Carlos/C-9409-2011; Bohorquez, Ana/G-8694-2012
OI Rinaldi, Carlos/0000-0001-8886-5612; Bohorquez, Ana/0000-0001-9654-5619
FU National Science Foundation [DMR-1157490]; State of Florida; U.S.
   Department of Energy
FX We thank Dr. Rintaro Hashizume, Northwestern University, for providing
   the 9L gliosarcoma cell line used in this study. A portion of this work
   was performed in the Advanced MRI/S (AMRIS) Facility at the McKnight
   Brain Institute of the University of Florida, which is part of the
   National High Magnetic Field Laboratory (supported by National Science
   Foundation Cooperative Agreement DMR-1157490, the State of Florida, and
   the U.S. Department of Energy).
CR [Anonymous], 2004, TRANSPORT PHENOMENA
   Aryal MP, 2014, MAGN RESON MED, V71, P2206, DOI 10.1002/mrm.24873
   Bagher-Ebadian H, 2012, MAGN RESON MED, V68, P241, DOI 10.1002/mrm.23211
   Barth RF, 2009, J NEURO-ONCOL, V94, P299, DOI 10.1007/s11060-009-9875-7
   Beppu T, 2003, J NEURO-ONCOL, V63, P109, DOI 10.1023/A:1023977520909
   BERECZKI D, 1993, AM J PHYSIOL, V264, pH1360, DOI 10.1152/ajpheart.1993.264.5.H1360
   Burnham KP., 2004, MODEL SELECTION MULT, DOI [10.1007/b97636, DOI 10.1007/B97636, DOI 10.1016/J.ECOLMODEL.2003.11.004]
   Chen X, 2007, ACTA NEUROCHIR, V149, P1117, DOI 10.1007/s00701-007-1282-2
   Chen XM, 2008, MAGN RESON IMAGING, V26, P1433, DOI 10.1016/j.mri.2008.04.011
   Donaldson SS, 2006, J CLIN ONCOL, V24, P1266, DOI 10.1200/JCO.2005.04.6599
   Ewing JR, 2013, NMR BIOMED, V26, P1028, DOI 10.1002/nbm.2996
   FENSTERMACHER J, 1988, ANN NY ACAD SCI, V531, P29, DOI 10.1111/j.1749-6632.1988.tb51416.x
   Foottit GOC, 2011, P 19 ANN M ISMRM MON, P2038
   Haacke EM, 1999, MAGNETIC RESONANCE I
   Helton KJ, 2006, AM J NEURORADIOL, V27, P786
   Huszthy PC, 2012, NEURO-ONCOLOGY, V14, P979, DOI 10.1093/neuonc/nos135
   Jallo GI, 2006, CHILD NERV SYST, V22, P1519, DOI 10.1007/s00381-006-0174-8
   JohansenBerg H, 2009, DIFFUSION MRI: FROM QUANTITATIVE MEASUREMENT TO IN VIVO NEUROANATOMY, P1
   Jones DK, 1999, MAGNET RESON MED, V42, P515, DOI 10.1002/(SICI)1522-2594(199909)42:3<515::AID-MRM14>3.0.CO;2-Q
   LOGAN J, 1990, J CEREBR BLOOD F MET, V10, P740, DOI 10.1038/jcbfm.1990.127
   Magdoom KN, 2014, PLOS ONE, V9, DOI 10.1371/journal.pone.0089594
   Markwardt CB, 2009, ASTR SOC P, V411, P251
   Murad GJA, 2007, J NEUROSURG, V106, P351, DOI 10.3171/jns.2007.106.2.351
   Pickup S, 2005, MAGN RESON MED, V53, P35, DOI 10.1002/mrm.20336
   Rohrer M, 2005, INVEST RADIOL, V40, P715, DOI 10.1097/01.rli.0000184756.66360.d3
   Sarntinoranont M, 2003, ANN BIOMED ENG, V31, P327, DOI 10.1114/1.1554923
   SHOCKLEY RP, 1988, BRAIN RES, V454, P170, DOI 10.1016/0006-8993(88)90816-5
   Sinha S, 2002, AM J NEURORADIOL, V23, P520
   Smith MA, 1998, J NATL CANCER I, V90, P1269, DOI 10.1093/jnci/90.17.1269
   Stegman LD, 2000, GENE THER, V7, P1005, DOI 10.1038/sj.gt.3301199
   Stojiljkovic M, 2003, J NEURO-ONCOL, V63, P1, DOI 10.1023/A:1023732619651
   Subashi E, 2016, J NEURO-ONCOL, V126, P243, DOI 10.1007/s11060-015-1969-9
   Teo C, 2008, CHILD NERV SYST, V24, P1307, DOI 10.1007/s00381-008-0647-z
   Tofts PS, 1997, JMRI-J MAGN RESON IM, V7, P91, DOI 10.1002/jmri.1880070113
   TOFTS PS, 1991, MAGNET RESON MED, V17, P357, DOI 10.1002/mrm.1910170208
   Weinmann HJ, 1984, CHARACTERISTICS GADO, P619
   Wu Q, 2002, VET PATHOL, V39, P293, DOI 10.1354/vp.39-3-293
   Yankeelov TE, 2007, MAGN RESON IMAGING, V25, P1, DOI 10.1016/j.mri.2006.09.006
   Yen CP, 2007, J NEUROSURG, V106, P8, DOI 10.3171/jns.2007.106.1.8
   Zhang H, 2010, BIOPHYS J, V99, P1284, DOI 10.1016/j.bpj.2010.06.023
NR 40
TC 2
Z9 2
U1 2
U2 20
PU WILEY
PI HOBOKEN
PA 111 RIVER ST, HOBOKEN 07030-5774, NJ USA
SN 1053-1807
EI 1522-2586
J9 J MAGN RESON IMAGING
JI J. Magn. Reson. Imaging
PD MAY
PY 2019
VL 49
IS 5
BP 1322
EP 1332
DI 10.1002/jmri.26315
PG 11
WC Radiology, Nuclear Medicine & Medical Imaging
WE Science Citation Index Expanded (SCI-EXPANDED)
SC Radiology, Nuclear Medicine & Medical Imaging
GA HT2JY
UT WOS:000464390200012
PM 30318760
OA Green Accepted
DA 2023-08-27
ER

PT J
AU Roonprapunt, C
   Abbott, R
AF Roonprapunt, C
   Abbott, R
TI Surgical treatment of Brainstem gliomas in children
SO NEUROSURGERY QUARTERLY
LA English
DT Article
DE brainstem glioma; midbrain tumors; tectal tumors; exophytic;
   cervicomedullary
ID HYPERFRACTIONATED RADIOTHERAPY; PEDIATRIC-PATIENTS; INTRAAXIAL TUMORS;
   TECTAL TUMORS; CHILDHOOD; CHEMOTHERAPY; MANAGEMENT; NEUROFIBROMATOSIS;
   DIAGNOSIS; SYMPTOMS
AB Brainstem gliomas have historically been some of the most difficult pediatric cancers to treat. Tumors arising in the brainstem were once uniformly discounted as surgically unapproachable lesions. The advent of magnetic resonance imaging and sophisticated neurophysiologic monitoring techniques has significantly advanced the surgical treatment of these precarious lesions. Brainstem gliomas are now recognized as a heterogenous group of tumors. They have been broadly classified in four categories: diffuse, focal, exophytic, and cervicomedullary. This classification system has provided a framework to predict growth patterns, surgical resectability, and overall prognosis. Although most remain unresectable, there are certain brainstem gliomas that are readily amenable to surgical therapy.
C1 Beth Israel Deaconess Med Ctr, Inst Neurol & Neurosurg, North Div, Dept Neurosurg, New York, NY 10128 USA.
C3 Harvard University; Beth Israel Deaconess Medical Center
EM
CR Abbott R, 1993, Neurosurg Clin N Am, V4, P519
   Abbott R, 1996, PEDIATR NEUROSURG, V25, P41, DOI 10.1159/000121095
   ABBOTT R, 1992, PEDIATR NEUROSURG, V18, P153, DOI 10.1159/000120656
   ABBOTT R, 1992, PEDIATR NEUROSURG, V17, P239, DOI 10.1159/000120604
   ALBRIGHT AL, 1986, J NEUROSURG, V65, P751, DOI 10.3171/jns.1986.65.6.0751
   ALBRIGHT AL, 1993, NEUROSURGERY, V33, P1026
   ALBRIGHT AL, 1996, NEUROLOGICAL SURG, P2603
   Allen J., 1983, NEUROL NEUROSURG UPD, V4, P2
   Allen JC, 1996, PEDIATR NEUROSURG, V24, P98, DOI 10.1159/000121024
   ALVISI C, 1985, ACTA NEUROCHIR, V76, P12, DOI 10.1007/BF01403823
   [Anonymous], 2000, OPERATIVE TECHNIQUES
   Behnke J, 1997, CHILD NERV SYST, V13, P122, DOI 10.1007/s003810050058
   Beltramello A, 2000, OPERATIVE TECHNIQUES, V3, P87
   BERGER MS, 1983, NEUROSURGERY, V12, P298, DOI 10.1227/00006123-198303000-00008
   Blaney S, 2001, CLIN CANCER RES, V7, P32
   Bowers DC, 2000, PEDIATR NEUROSURG, V32, P24, DOI 10.1159/000028893
   Bowers DC, 2001, PEDIATR NEUROSURG, V34, P229, DOI 10.1159/000056027
   BOYDSTON WR, 1992, PEDIATR NEUROSURG, V17, P234, DOI 10.1159/000120603
   Bricolo A, 2000, OPER TECH NEUROSURG, V3, P137, DOI DOI 10.1053/oy.2000.6559
   Broniscer A, 1997, NEUROSURGERY, V40, P331, DOI 10.1097/00006123-199702000-00018
   Broniscer A, 2000, J CLIN ONCOL, V18, P1246, DOI 10.1200/JCO.2000.18.6.1246
   Choux M, 2000, PEDIAT NEUROSURGERY, P471
   Constantini S, 1996, J NEURO-ONCOL, V28, P193
   CONSTANTINI S, 1996, NEUROSURGERY, P2714
   Dechambre S, 1999, NEURORADIOLOGY, V41, P829, DOI 10.1007/s002340050850
   Deletis V, 2000, OPERATIVE TECHNIQUES, V3, P109, DOI DOI 10.1053/oy.2000.6562
   EPSTEIN F, 1988, J NEURO-ONCOL, V6, P309
   EPSTEIN F, 1986, J NEUROSURG, V64, P11, DOI 10.3171/jns.1986.64.1.0011
   EPSTEIN F, 1987, J NEUROSURG, V67, P483, DOI 10.3171/jns.1987.67.4.0483
   EPSTEIN FJ, 1993, J NEUROSURG, V78, P408, DOI 10.3171/jns.1993.78.3.0408
   EPSTEIN FJ, 1993, J NEUROSURG, V78, P505, DOI 10.3171/jns.1993.78.3.0505
   Fischbein NJ, 1996, PEDIATR NEUROSURG, V24, P9, DOI 10.1159/000121010
   Freeman CR, 1996, CANCER-AM CANCER SOC, V77, P555
   Freeman CR, 2000, INT J RADIAT ONCOL, V47, P561, DOI 10.1016/S0360-3016(00)00471-5
   HOFFMAN HJ, 1980, NEUROSURGERY, V7, P243, DOI 10.1227/00006123-198009000-00007
   Jallo GI, 2001, NEUROSURGERY, V48, P695, DOI 10.1097/00006123-200103000-00054
   JIMENEZ DF, 1998, NEUROSURGICAL TOPICS, P101
   Kaplan AM, 1996, PEDIATR NEUROSURG, V24, P185, DOI 10.1159/000121036
   KEDAR A, 1994, MED PEDIATR ONCOL, V23, P428, DOI 10.1002/mpo.2950230507
   KHATIB ZA, 1994, PEDIATR NEUROSURG, V20, P2, DOI 10.1159/000120759
   Kurpad SN, 1999, PEDIATR NEUROSURG, V31, P54, DOI 10.1159/000028833
   LAPRAS C, 1994, ACTA NEUROCHIR, V126, P76, DOI 10.1007/BF01476414
   LITTMAN P, 1980, CANCER, V45, P2787, DOI 10.1002/1097-0142(19800601)45:11<2787::AID-CNCR2820451113>3.0.CO;2-V
   Matson DD, 1969, NEUROSURGERY INFANCY, P469
   MILSTEIN JM, 1989, J NEURO-ONCOL, V7, P367, DOI 10.1007/BF02147094
   MOROTA N, 1995, NEUROSURGERY, V37, P922, DOI 10.1227/00006123-199511000-00011
   Murakami N, 1999, NEUROSURG REV, V22, P159
   NISHIO S, 1988, J NEURO-ONCOL, V6, P245, DOI 10.1007/BF00163708
   Oka K, 1999, J NEUROSURG, V91, P964, DOI 10.3171/jns.1999.91.6.0964
   POLLACK IF, 1995, NEUROSURGERY, V37, P885, DOI 10.1227/00006123-199511000-00006
   POLLACK IF, 1994, J NEUROSURG, V80, P681, DOI 10.3171/jns.1994.80.4.0681
   POLLACK IF, 1993, J NEUROSURG, V78, P859, DOI 10.3171/jns.1993.78.6.0859
   RAFFEL C, 1989, NEUROSURGERY, V25, P959, DOI 10.1227/00006123-198912000-00018
   Raffel C, 1996, J NEURO-ONCOL, V28, P121
   ROBERTSON PL, 1994, NEUROLOGY, V44, P1798, DOI 10.1212/WNL.44.10.1798
   ROSE E, 1998, AESTHETIC FACIAL RES
   ROUX FX, 1990, ACTA NEUROCHIR, V107, P161, DOI 10.1007/BF01405796
   SQUIRES LA, 1994, NEUROLOGY, V44, P953, DOI 10.1212/WNL.44.5.953
   STEIN BM, 1971, J NEUROSURG, V35, P197, DOI 10.3171/jns.1971.35.2.0197
   STROINK AR, 1987, NEUROSURGERY, V20, P439, DOI 10.1227/00006123-198703000-00014
   VANDERTOP WP, 1992, NEUROSURGERY, V31, P186, DOI 10.1227/00006123-199208000-00003
   Walker DA, 1999, ARCH DIS CHILD, V80, P558, DOI 10.1136/adc.80.6.558
   Walter AW, 1998, MED PEDIATR ONCOL, V30, P28, DOI 10.1002/(SICI)1096-911X(199801)30:1<28::AID-MPO9>3.0.CO;2-2
   Wang CC, 2000, SURG NEUROL, V53, P41, DOI 10.1016/S0090-3019(99)00165-2
   Weiner HL, 1997, PEDIATR NEUROSURG, V27, P12, DOI 10.1159/000121219
   Wisoff J H, 1984, Neurosurgery, V15, P707
   Zagzag D, 2000, PEDIATRICS, V106, P1045, DOI 10.1542/peds.106.5.1045
NR 67
TC 4
Z9 5
U1 0
U2 1
PU LIPPINCOTT WILLIAMS & WILKINS
PI PHILADELPHIA
PA TWO COMMERCE SQ, 2001 MARKET ST, PHILADELPHIA, PA 19103 USA
SN 1050-6438
EI 1534-4916
J9 NEUROSURG QUART
JI Neurosurg. Q.
PD JUN
PY 2002
VL 12
IS 2
BP 160
EP 170
DI 10.1097/00013414-200206000-00008
PG 11
WC Neurosciences; Surgery
WE Science Citation Index Expanded (SCI-EXPANDED)
SC Neurosciences & Neurology; Surgery
GA 555KW
UT WOS:000175794100008
DA 2023-08-27
ER

PT J
AU Lin, JK
   Feng, H
   Li, F
   Zhao, BM
   Guo, QN
AF Lin, JK
   Feng, H
   Li, F
   Zhao, BM
   Guo, QN
TI Intraparenchymal schwannoma of the medulla oblongata - Case report
SO JOURNAL OF NEUROSURGERY
LA English
DT Article
DE brainstem tumor; schwannoma; neuroma; medulla oblongata
ID GAMMA-KNIFE RADIOSURGERY; VESTIBULAR SCHWANNOMA; INTRAMEDULLARY
   SCHWANNOMA
AB A 48-year-old man presented with a rare intraparenchymal schwannoma of the medulla oblongata. After he underwent gamma knife surgery, the patient's condition deteriorated and the mass, which at the time was thought to be a glioma, became larger on magnetic resonance images. The mass was resected through a suboccipital craniectomy via the transcerebellomedullary fissure approach. The tumor, which was moderately firm but distinct from the surrounding parenchyma, was removed totally. Postoperative histological and immunohistochemical examinations confirmed the diagnosis of a benign schwannoma. Brainstem schwannomas can be cured by microneurosurgery. It is important to distinguish these tumors from glioma. The main theories on the cause(s) of this lesion are briefly reviewed.
C1 Third Mil Med Univ, SW Hosp, Dept Neurosurg, Chongqing 400038, Peoples R China.
   Third Mil Med Univ, SW Hosp, Dept Pathol, Chongqing 400038, Peoples R China.
C3 Army Medical University; Army Medical University
RP Lin, JK (通讯作者)，Third Mil Med Univ, SW Hosp, Dept Neurosurg, Chongqing 400038, Peoples R China.
CR ARYANPUR J, 1988, J NEUROSURG, V69, P446, DOI 10.3171/jns.1988.69.3.0446
   Beskonakli E, 1997, NEUROSURG REV, V20, P139, DOI 10.1007/BF01138200
   FRIM DM, 1992, PEDIATR NEUROSURG, V18, P190, DOI 10.1159/000120661
   GOKAY H, 1984, SURG NEUROL, V22, P69, DOI 10.1016/0090-3019(84)90232-5
   HERREGODTS P, 1991, J NEUROSURG, V74, P816, DOI 10.3171/jns.1991.74.5.0816
   Linskey ME, 2000, J NEUROSURG, V93, P90, DOI 10.3171/jns.2000.93.supplement_3.0090
   Matsushima T, 2001, J NEUROSURG, V94, P257, DOI 10.3171/jns.2001.94.2.0257
   Nakamura H, 2000, AM J NEURORADIOL, V21, P1540
   PERONE TP, 1984, NEUROSURGERY, V14, P71, DOI 10.1227/00006123-198401000-00016
   Pollock BE, 1998, J NEUROSURG, V89, P949, DOI 10.3171/jns.1998.89.6.0949
   PRAKASH B, 1980, J NEUROSURG, V53, P121, DOI 10.3171/jns.1980.53.1.0121
   ROSS DA, 1986, NEUROSURGERY, V19, P458, DOI 10.1227/00006123-198609000-00023
   ROUT D, 1983, J NEUROSURG, V58, P962, DOI 10.3171/jns.1983.58.6.0962
   SARKAR C, 1987, J NEUROSURG, V67, P120, DOI 10.3171/jns.1987.67.1.0120
   Shin M, 2002, LANCET, V360, P309, DOI 10.1016/S0140-6736(02)09521-1
   Tanabe Michiharu, 1996, Neurologia Medico-Chirurgica, V36, P880, DOI 10.2176/nmc.36.880
   TRANDINH HD, 1991, NEUROSURGERY, V29, P296, DOI 10.1227/00006123-199108000-00026
   VAILATI G, 1979, SURG NEUROL, V11, P60
   Zhang L, 2001, Zhonghua Yi Xue Za Zhi, V81, P788
NR 19
TC 13
Z9 14
U1 1
U2 4
PU AMER ASSOC NEUROLOGICAL SURGEONS
PI CHARLOTTESVILLE
PA UNIV VIRGINIA, 1224 WEST MAIN ST, STE 450, CHARLOTTESVILLE, VA 22903 USA
SN 0022-3085
J9 J NEUROSURG
JI J. Neurosurg.
PD MAR
PY 2003
VL 98
IS 3
BP 621
EP 624
DI 10.3171/jns.2003.98.3.0621
PG 4
WC Clinical Neurology; Surgery
WE Science Citation Index Expanded (SCI-EXPANDED)
SC Neurosciences & Neurology; Surgery
GA 653WW
UT WOS:000181460900026
PM 12650438
DA 2023-08-27
ER

PT J
AU Yang, DT
   Rozen, WM
   Rickert, CH
   Lo, PA
AF Yang, David Tran
   Rozen, Warren Matthew
   Rickert, Christian H.
   Lo, Patrick Anthony
TI Primary pontomedullary germinoma in a 12 year old boy
SO JOURNAL OF CLINICAL NEUROSCIENCE
LA English
DT Article
DE Brainstem; Pontine; Medullary; Germinoma; Pediatric brain tumor;
   Pediatric neurosurgery
ID PRIMARY INTRACRANIAL GERMINOMA; MEDULLA-OBLONGATA GERMINOMA;
   CEREBELLOPONTINE ANGLE; MIDBRAIN; MANAGEMENT; FOSSA
AB Primary intracranial germinomas are rare tumors, accounting for approximately 1-4% of all intracranial tumors. Intracranial germinomas are more commonly found in the suprasellar and pineal midline structures of the brain. Brainstem and posterior fossa germinomas are rarer still, with few reported cases in the literature, and little discussion of their presentation, management and clinical outcome. A unique case of pontomedullary germinoma, diagnosed in a 12-year-old boy, is described. Only six previous cases of lower brainstem germinoma have been reported, with varying modes of presentation and a lack of definitive management guidelines. Of these, all occurred in either females or a patient with Klinefelter's syndrome. We report the first case of a lower brainstem germinoma in a male without known genetic abnormality. Tumor remission was achieved with partial surgical resection, chemotherapy and radiotherapy. Crown Copyright (C) 2007 Published by Elsevier Ltd. All rights reserved.
C1 [Lo, Patrick Anthony] Royal Childrens Hosp, Dept Neurosurg, Parkville, Vic 3052, Australia.
   [Yang, David Tran; Rozen, Warren Matthew] Royal Melbourne Hosp, Dept Neurosurg, Parkville, Vic 3050, Australia.
   [Rickert, Christian H.] Royal Childrens Hosp, Dept Anat Pathol, Parkville, Vic 3052, Australia.
C3 Royal Children's Hospital Melbourne; Royal Melbourne Hospital; Royal
   Children's Hospital Melbourne
RP Lo, PA (通讯作者)，Laccon Neurosurg, POB 1108, Elwood, Vic 3184, Australia.
EM
RI Rozen, Warren Matthew/AAE-6296-2022
OI Rozen, Warren Matthew/0000-0002-4092-182X
CR Ben Amor S, 2004, BRIT J NEUROSURG, V18, P310, DOI 10.1080/02688690410001732832
   Buckner JC, 1999, J CLIN ONCOL, V17, P933, DOI 10.1200/JCO.1999.17.3.933
   Evanson EJ, 1997, NEURORADIOLOGY, V39, P716, DOI 10.1007/s002340050493
   Fujiwara K, 2002, NEUROL MED-CHIR, V42, P132, DOI 10.2176/nmc.42.132
   HASHIMOTO M, 1992, SURG NEUROL, V37, P384, DOI 10.1016/0090-3019(92)90009-C
   HOROWITZ MB, 1991, ARCH NEUROL-CHICAGO, V48, P652, DOI 10.1001/archneur.1991.00530180110026
   Kurtsoy A, 1996, NEUROSURG REV, V19, P127, DOI 10.1007/BF00418084
   Maiuri F, 2004, BRIT J NEUROSURG, V18, P284, DOI 10.1080/02688690410001732760
   Matsumoto K, 1998, CLIN NEUROL NEUROSUR, V100, P292, DOI 10.1016/S0303-8467(98)00050-X
   Matsutani M, 1998, Neurosurg Focus, V5, pe7
   Nakajima H, 2000, SURG NEUROL, V53, P448, DOI 10.1016/S0090-3019(00)00224-X
   Paulino AC, 1999, ONCOLOGY-NY, V13, P513
   PAULINO AC, 1999, ONCOLOGY HUNTINGT, V13, P528
   PAULINO AC, 1999, ONCOLOGY HUNTINGT, V13, P521
   Rogers SJ, 2005, LANCET ONCOL, V6, P509, DOI 10.1016/S1470-2045(05)70245-X
   Sawamura Y, 1997, J NEUROSURG, V87, P262, DOI 10.3171/jns.1997.87.2.0262
   Sugiyama K, 1994, Neurol Med Chir (Tokyo), V34, P291, DOI 10.2176/nmc.34.291
   Tashiro Takashi, 1993, Neurologia Medico-Chirurgica, V33, P251, DOI 10.2176/nmc.33.251
   Uchino M, 2006, J NEURO-ONCOL, V79, P255, DOI 10.1007/s11060-006-9138-9
   Yen PS, 2003, J NEURO-ONCOL, V62, P339, DOI 10.1023/A:1023331700995
NR 20
TC 8
Z9 8
U1 0
U2 0
PU ELSEVIER SCI LTD
PI OXFORD
PA THE BOULEVARD, LANGFORD LANE, KIDLINGTON, OXFORD OX5 1GB, OXON, ENGLAND
SN 0967-5868
EI 1532-2653
J9 J CLIN NEUROSCI
JI J. Clin. Neurosci.
PD FEB
PY 2009
VL 16
IS 2
BP 321
EP U350
DI 10.1016/j.jocn.2007.08.026
PG 5
WC Clinical Neurology; Neurosciences
WE Science Citation Index Expanded (SCI-EXPANDED)
SC Neurosciences & Neurology
GA 418XA
UT WOS:000264181900026
PM 19091568
DA 2023-08-27
ER

PT J
AU ROBERTSON, PL
   MURASZKO, KM
   BRUNBERG, JA
   AXTELL, RA
   DAUSER, RC
   TURRISI, AT
AF ROBERTSON, PL
   MURASZKO, KM
   BRUNBERG, JA
   AXTELL, RA
   DAUSER, RC
   TURRISI, AT
TI PEDIATRIC MIDBRAIN TUMORS - A BENIGN SUBGROUP OF BRAIN-STEM GLIOMAS
SO PEDIATRIC NEUROSURGERY
LA English
DT Article
DE MIDBRAIN; TECTAL REGION; NEOPLASM; GLIOMA
ID STEM GLIOMAS; CHILDHOOD; CHILDREN; TRIAL
AB The presentation, radiographic findings and course of 17 children with MRI-documented intrinsic midbrain lesions are reviewed. The anatomic centers of all the lesions were tectal, peritectal, or tegmental. Lesions centered at the pineal gland were excluded. Signs of increased intracranial pressure from hydrocephalus requiring shunt placement were present in 14 patients. Histopathological diagnosis was confirmed in three tumors; these were low grade astrocytomas and all received focal irradiation, as did one unbiopsied tumor. The remaining 13 patients with no histopathological diagnosis received no therapy other than shunt placement in 11. All but one of the lesions have remained clinically and radiographically stable, with a 4-year progression-free and total survival of 94 and 100%, respectively. We conclude that mass lesions originating in the upper midbrain are a subset of intrinsic brainstem tumors with a relatively benign course, usually presenting with hydrocephalus after infancy. They may remain stable for considerable periods and may require no further therapy after treatment of hydrocephalus. Surgical biopsy and/or resection can usually be reserved for progressive or atypical lesions which may also require further adjuvant therapy.
C1 UNIV MICHIGAN, MED CTR, DEPT NEUROL, ANN ARBOR, MI 48109 USA.
   UNIV MICHIGAN, MED CTR, DEPT NEUROSURG, ANN ARBOR, MI 48109 USA.
   UNIV MICHIGAN, MED CTR, DEPT NEURORADIOL, ANN ARBOR, MI 48109 USA.
   UNIV MICHIGAN, MED CTR, DEPT RADIAT ONCOL, ANN ARBOR, MI 48109 USA.
C3 University of Michigan System; University of Michigan; University of
   Michigan System; University of Michigan; University of Michigan System;
   University of Michigan; University of Michigan System; University of
   Michigan
RP ROBERTSON, PL (通讯作者)，UNIV MICHIGAN, MED CTR, DEPT PEDIAT, R 6060 KRESGE II, BOX 0570, ANN ARBOR, MI 48109 USA.
CR ALBRIGHT AL, 1986, J NEUROSURG, V65, P751, DOI 10.3171/jns.1986.65.6.0751
   BARKOVICH AJ, 1991, PEDIATR NEUROSURG, V16, P73, DOI 10.1159/000120511
   BOYDSTON WR, 1992, PEDIATR NEUROSURG, V17, P234, DOI 10.1159/000120603
   CHAPMAN PH, 1989, CONCEPTS PEDIATRIC N, V9, P97
   EDWARDS MSB, 1989, J NEUROSURG, V70, P691, DOI 10.3171/jns.1989.70.5.0691
   EPSTEIN F, 1988, J NEURO-ONCOL, V6, P309
   GUY G, 1989, NEUROCHIRURGIE, V35, P41
   HOFFMAN HJ, 1980, NEUROSURGERY, V7, P243, DOI 10.1227/00006123-198009000-00007
   JENKIN RDT, 1987, J NEUROSURG, V66, P227, DOI 10.3171/jns.1987.66.2.0227
   LEE BCP, 1985, AM J NEURORADIOL, V6, P159
   LITTMAN P, 1980, CANCER, V45, P2787, DOI 10.1002/1097-0142(19800601)45:11<2787::AID-CNCR2820451113>3.0.CO;2-V
   MAY PL, 1991, J NEUROSURG, V74, P867, DOI 10.3171/jns.1991.74.6.0867
   PACKER RJ, 1990, ANN NEUROL, V27, P167, DOI 10.1002/ana.410270212
   PENDL G, 1990, NEUROSURGERY, V26, P641, DOI 10.1227/00006123-199004000-00013
   RAFFEL C, 1988, PEDIATRICS, V82, P733
   SANFORD RA, 1982, J NEUROSURG, V57, P690, DOI 10.3171/jns.1982.57.5.0690
   SANFORD RA, 1988, SURG NEUROL, V30, P276, DOI 10.1016/0090-3019(88)90299-6
   STEINBOK P, 1987, CHILD NERV SYST, V3, P170, DOI 10.1007/BF00717895
   VANDERTOP WP, 1992, NEUROSURGERY, V31, P186, DOI 10.1227/00006123-199208000-00003
NR 19
TC 55
Z9 55
U1 0
U2 2
PU KARGER
PI BASEL
PA ALLSCHWILERSTRASSE 10, CH-4009 BASEL, SWITZERLAND
SN 1016-2291
EI 1423-0305
J9 PEDIATR NEUROSURG
JI Pediatr. Neurosurg.
PD FEB
PY 1995
VL 22
IS 2
BP 65
EP 73
DI 10.1159/000120879
PG 9
WC Clinical Neurology; Pediatrics; Surgery
WE Science Citation Index Expanded (SCI-EXPANDED)
SC Neurosciences & Neurology; Pediatrics; Surgery
GA QF262
UT WOS:A1995QF26200002
PM 7710975
DA 2023-08-27
ER

PT J
AU Aoki, Y
   Hashizume, R
   Ozawa, T
   Banerjee, A
   Prados, M
   James, CD
   Gupta, N
AF Aoki, Yasuyuki
   Hashizume, Rintaro
   Ozawa, Tomoko
   Banerjee, Anu
   Prados, Michael
   James, C. David
   Gupta, Nalin
TI An experimental xenograft mouse model of diffuse pontine glioma designed
   for therapeutic testing
SO JOURNAL OF NEURO-ONCOLOGY
LA English
DT Article
DE Brainstem glioma; Mouse model; Bioluminescence image
ID BRAIN-STEM GLIOMA; TUMORS; DELIVERY; GROWTH; INHIBITION; REVEALS
AB The prognosis for diffuse infiltrating pontine gliomas (DIPG) remains extremely poor, with the majority of patients surviving less than 2 years. Here, we have adapted standard xenograft techniques to study glioma growth in the mouse brainstem, and have utilized the mouse model for studying a relevant therapeutic for treating DIPGs. bioluminescence imaging monitoring revealed a progressive increase in signal following the injection of either of two tumor cell types into the brainstem. Mice with orthotopic GS2 tumors, and receiving a single 100 mg/kg dose of temozolomide showed a lengthy period of decreased tumor luminescence, with substantially increased survival relative to untreated mice (P < 0.001). A small molecule inhibitor that targets cdk4/6 was used to test AM-38 brainstem xenograft response to treatment. Drug treatment resulted in delayed tumor growth, and significantly extended survival. Our results demonstrate the feasibility of using an orthotopic brainstem tumor model in athymic mice, and for application to testing therapeutic agents in treating DIPG.
C1 [Aoki, Yasuyuki; Hashizume, Rintaro; Ozawa, Tomoko; Prados, Michael; James, C. David; Gupta, Nalin] Univ Calif San Francisco, Dept Neurol Surg, San Francisco, CA 94143 USA.
   [Aoki, Yasuyuki; Hashizume, Rintaro; Ozawa, Tomoko; Prados, Michael; James, C. David; Gupta, Nalin] Univ Calif San Francisco, Brain Tumor Res Ctr, San Francisco, CA 94143 USA.
   [Banerjee, Anu; Gupta, Nalin] Univ Calif San Francisco, Dept Pediat, San Francisco, CA 94143 USA.
C3 University of California System; University of California San Francisco;
   University of California System; University of California San Francisco;
   University of California System; University of California San Francisco
RP Gupta, N (通讯作者)，Univ Calif San Francisco, Dept Neurol Surg, 505 Parnassus Ave,Rm M779, San Francisco, CA 94143 USA.
EM
RI James, Charles D/E-2721-2012; Prados, Michael/N-5220-2019
OI James, Charles D/0000-0002-1027-203X; 
FU Pediatric Brain Tumor Foundation Institute
FX This work was supported by the Pediatric Brain Tumor Foundation
   Institute Award to the Brain Tumor Research Center at UCSF.
CR Becher OJ, 2010, CANCER RES, V70, P2548, DOI 10.1158/0008-5472.CAN-09-2503
   Caretti V, 2011, BRAIN PATHOL, V21, P441, DOI 10.1111/j.1750-3639.2010.00468.x
   Claes A, 2008, INT J CANCER, V122, P1981, DOI 10.1002/ijc.23306
   de Tayrac M, 2009, GENE CHROMOSOME CANC, V48, P55, DOI 10.1002/gcc.20618
   Dinca EB, 2007, J NEUROSURG, V107, P610, DOI 10.3171/JNS-07/09/0610
   DINSE GE, 1982, BIOMETRICS, V38, P921, DOI 10.2307/2529872
   Edinger M, 2002, EUR J CANCER, V38, P2128, DOI 10.1016/S0959-8049(02)00410-0
   Finlay JL, 2005, J NEURO-ONCOL, V75, P253, DOI 10.1007/s11060-005-6747-7
   Fry DW, 2004, MOL CANCER THER, V3, P1427
   Hargrove D, 2006, LANCET ONCOL, V7, P241, DOI 10.1016/S1470-2045(06)70615-5
   Hashizume R, 2008, NEURO-ONCOLOGY, V10, P112, DOI 10.1215/15228517-2007-052
   Hashizume R, 2010, J NEURO-ONCOL, V96, P151, DOI 10.1007/s11060-009-9954-9
   Huynh GH, 2006, J CONTROL RELEASE, V110, P236, DOI 10.1016/j.jconrel.2005.09.053
   Jallo GI, 2005, CHILD NERV SYST, V21, P399, DOI 10.1007/s00381-004-1100-6
   Lee James, 2005, Neurosurg Focus, V18, pE11
   Liu Q, 2008, J NEUROSURG, V109, P849, DOI 10.3171/JNS/2008/109/11/0849
   Malumbres M, 2009, NAT REV CANCER, V9, P153, DOI 10.1038/nrc2602
   Masui K, 2010, GLIA, V58, P1050, DOI 10.1002/glia.20986
   Michaud K, 2010, CANCER RES, V70, P3228, DOI 10.1158/0008-5472.CAN-09-4559
   Ozawa Tomoko, 2010, J Vis Exp, DOI 10.3791/1986
   Parsons DW, 2008, SCIENCE, V321, P1807, DOI 10.1126/science.1164382
   PETO R, 1972, J R STAT SOC SER A-G, V135, P185, DOI 10.2307/2344317
   Prasad G, 2011, NEURO-ONCOLOGY, V13, P384, DOI 10.1093/neuonc/noq193
   Sethi R, 2011, J NEURO-ONCOL, V102, P121, DOI 10.1007/s11060-010-0301-y
   Siu IM, 2010, J NEUROSURG-PEDIATR, V6, P92, DOI 10.3171/2010.3.PEDS09366
   Thorne RG, 2004, NEUROSCIENCE, V127, P481, DOI 10.1016/j.neuroscience.2004.05.029
NR 26
TC 40
Z9 43
U1 0
U2 11
PU SPRINGER
PI NEW YORK
PA 233 SPRING ST, NEW YORK, NY 10013 USA
SN 0167-594X
EI 1573-7373
J9 J NEURO-ONCOL
JI J. Neuro-Oncol.
PD MAY
PY 2012
VL 108
IS 1
BP 29
EP 35
DI 10.1007/s11060-011-0796-x
PG 7
WC Oncology; Clinical Neurology
WE Science Citation Index Expanded (SCI-EXPANDED)
SC Oncology; Neurosciences & Neurology
GA 934TM
UT WOS:000303469600003
PM 22231932
OA Green Accepted
DA 2023-08-27
ER

PT J
AU Jallo, GI
   Shiminski-Maher, T
   Velazquez, L
   Abbott, R
   Wisoff, J
   Epstein, F
AF Jallo, GI
   Shiminski-Maher, T
   Velazquez, L
   Abbott, R
   Wisoff, J
   Epstein, F
TI Recovery of lower cranial nerve function after surgery for medullary
   brainstem tumors
SO NEUROSURGERY
LA English
DT Article
DE brainstem; cranial nerve; medulla; postoperative deficits
ID LONG-TERM; GLIOMAS; CHILDHOOD
AB OBJECTIVE: Although optimal treatment for intrinsic focal tumors of the medulla remains controversial, many surgeons advocate radical surgery for patients with these tumors. Postoperative surgical morbidity may include loss of lower cranial nerve function and significant motor deficits. Recovery of lower cranial nerve dysfunction after radical surgery has not been reported previously.
   METHODS: Forty-one children and adolescents with tumors involving the medulla underwent operations between 1986 and 1997. Nineteen (46%) of these patients experienced loss of lower cranial nerve function requiring tracheostomy, ventilator support, and feeding gastrostomy. A retrospective analysis of this patient population and the time to cranial nerve recovery was undertaken.
   RESULTS: Thirteen (68%) of the 19 patients with loss of lower cranial nerve function had full recovery of lower cranial nerve function. Two patients (111%) have had significant improvement in their lower cranial nerve function, and four patients (21%) have remained without lower cranial nerve function.
   CONCLUSION: Lower cranial dysfunction is common after surgery for intrinsic medullary tumors. However, the majority of patients who require tracheostomy or gastrostomy tubes will recover cranial nerve function.
C1 Johns Hopkins Univ Hosp, Div Pediat Neurosurg, Baltimore, MD 21287 USA.
   Beth Israel Deaconess Med Ctr, Div Pediat Neurosurg, Inst Neurol & Neurosurg, New York, NY 10003 USA.
   NYU, Div Pediat Neurosurg, Sch Med, New York, NY 10016 USA.
C3 Johns Hopkins University; Johns Hopkins Medicine; Harvard University;
   Beth Israel Deaconess Medical Center; New York University
RP Jallo, GI (通讯作者)，Johns Hopkins Univ Hosp, Div Pediat Neurosurg, 600 N Wolfe St,Harvey 811, Baltimore, MD 21287 USA.
EM
RI Wisoff, Jeffrey/AAW-1500-2020
OI Wisoff, Jeffrey/0000-0003-3303-2220
CR Abbott R, 1996, PEDIATR NEUROSURG, V25, P41, DOI 10.1159/000121095
   ABBOTT R, 1992, PEDIATR NEUROSURG, V17, P239, DOI 10.1159/000120604
   ALVISI C, 1985, ACTA NEUROCHIR, V76, P12, DOI 10.1007/BF01403823
   [Anonymous], 2000, OPERATIVE TECHNIQUES
   BAILEY P, 1939, INTRACRANIAL TUMORS, P188
   Burger PC, 1996, PEDIATR NEUROSURG, V24, P35, DOI 10.1159/000121012
   Choux M, 2000, PEDIAT NEUROSURGERY, P471
   EPSTEIN F, 1985, CANCER, V56, P1804, DOI 10.1002/1097-0142(19851001)56:7+<1804::AID-CNCR2820561316>3.0.CO;2-5
   EPSTEIN F, 1986, J NEUROSURG, V64, P11, DOI 10.3171/jns.1986.64.1.0011
   Epstein F, 1990, Neurosurg Clin N Am, V1, P111
   Epstein F, 1996, PEDIATR NEUROSURG, V24, P24, DOI 10.1159/000121011
   Fischbein NJ, 1996, PEDIATR NEUROSURG, V24, P9, DOI 10.1159/000121010
   HOFFMAN HJ, 1980, NEUROSURGERY, V7, P243, DOI 10.1227/00006123-198009000-00007
   Jallo GI, 2003, ANN NEUROSURG, V3, P1
   Matson DD, 1969, NEUROSURGERY INFANCY, P469
   ShiminskiMaher T, 1996, PEDIATR NEUROSURG, V24, P267, DOI 10.1159/000121050
   Wang GH, 2001, NEUROSURG QUART, V11, P302, DOI 10.1097/00013414-200112000-00006
NR 17
TC 11
Z9 12
U1 0
U2 0
PU OXFORD UNIV PRESS INC
PI CARY
PA JOURNALS DEPT, 2001 EVANS RD, CARY, NC 27513 USA
SN 0148-396X
EI 1524-4040
J9 NEUROSURGERY
JI Neurosurgery
PD JAN
PY 2005
VL 56
IS 1
BP 74
EP 77
DI 10.1227/01.NEU.0000144782.39430.12
PG 4
WC Clinical Neurology; Surgery
WE Science Citation Index Expanded (SCI-EXPANDED)
SC Neurosciences & Neurology; Surgery
GA 884FD
UT WOS:000226069300027
PM 15617588
OA hybrid
DA 2023-08-27
ER

PT J
AU MOLLOY, PT
   BILANIUK, LT
   VAUGHAN, SN
   NEEDLE, MN
   LIU, GT
   ZACKAI, EH
   PHILLIPS, PC
AF MOLLOY, PT
   BILANIUK, LT
   VAUGHAN, SN
   NEEDLE, MN
   LIU, GT
   ZACKAI, EH
   PHILLIPS, PC
TI BRAIN-STEM TUMORS IN PATIENTS WITH NEUROFIBROMATOSIS TYPE-1 - A DISTINCT
   CLINICAL ENTITY
SO NEUROLOGY
LA English
DT Article
ID HYPERFRACTIONATED RADIATION-THERAPY; STEM TUMORS; PEDIATRIC-PATIENTS;
   IMAGING FINDINGS; MR FINDINGS; CHILDREN; GLIOMAS; LESIONS; SYSTEM;
   CHILDHOOD
AB The natural history and the clinical and neuroimaging features of brainstem tumors in neurofibromatosis type 1 (NF1) are poorly understood. Magnetic resonance imaging (MRI) has been useful in NF1 in detecting intracranial abnormalities, especially of the brainstem. Brainstem tumors in NF1 have been confused clinically with non-NF1 brainstem tumors and radiographically with the increased T-2 signal abnormalities, also known as ''unidentified bright objects'' (UBOs), which are common in NF1 and often located in the brainstem,This study, which evaluated 17 NF1 patients with brainstem tumors, is the largest series to date. Fifteen of 17 patients (88%) had neurologic signs and symptoms referable to brainstem dysfunction, including dysarthria, cranial neuropathies, and gross motor incoordination. Tumors were located primarily in the medulla in 14 of 17 NF1 patients (82%), in contrast to the pontine tumor location in the non-NF1 population. Seven NF1 patients (41%) required shunt placement for hydrocephalus at initial diagnosis, more frequent than in non-NF1 brainstem tumor patients. Six of 17 patients (35%) had evidence of radiographic tumor progression, but only three of them (18%) had correlative clinical progression. Two patients with progressive symptoms had partial surgical resection, and pathology revealed either fibrillary or anaplastic astrocytomas. Three patients were treated with radiation therapy, chemotherapy, or both, with two deaths. With a median follow-up of 52 months, 15 of 17 patients remain alive; 14 of them did not require adjuvant therapy. In our series, we describe NF1 brainstem tumors as a distinct clinical entity, much less aggressive than non-NF1 pontine tumors but more symptomatic than brainstem UBOs in NF1.
C1 CHILDRENS HOSP, DIV ONCOL, PHILADELPHIA, PA 19104 USA.
   CHILDRENS HOSP, DIV RADIOL, PHILADELPHIA, PA 19104 USA.
   CHILDRENS HOSP, DIV NEUROOPTHALMOL, PHILADELPHIA, PA 19104 USA.
   CHILDRENS HOSP, DIV GENET, PHILADELPHIA, PA 19104 USA.
C3 University of Pennsylvania; Pennsylvania Medicine; Childrens Hospital of
   Philadelphia; University of Pennsylvania; Pennsylvania Medicine;
   Childrens Hospital of Philadelphia; University of Pennsylvania;
   Pennsylvania Medicine; Childrens Hospital of Philadelphia; University of
   Pennsylvania; Pennsylvania Medicine; Childrens Hospital of Philadelphia
RP MOLLOY, PT (通讯作者)，CHILDRENS HOSP, DIV NEUROL, 324 S 34TH ST, PHILADELPHIA, PA 19104 USA.
CR ALBRIGHT AL, 1986, J NEUROSURG, V65, P751, DOI 10.3171/jns.1986.65.6.0751
   [Anonymous], 1988, ARCH NEUROL-CHICAGO, V45, P575, DOI 10.1097/00005537-199606000-00007
   AOKI S, 1989, RADIOLOGY, V172, P527, DOI 10.1148/radiology.172.2.2501822
   BARKOVICH AJ, 1991, PEDIATR NEUROSURG, V16, P73, DOI 10.1159/000120511
   BLATT J, 1986, CANCER, V57, P1225, DOI 10.1002/1097-0142(19860315)57:6<1225::AID-CNCR2820570627>3.0.CO;2-P
   BOGNANNO JR, 1988, AM J ROENTGENOL, V151, P381
   BRADAC GB, 1985, NEURORADIOLOGY, V27, P208, DOI 10.1007/BF00344489
   BRAFFMAN BH, 1990, SEMIN ROENTGENOL, V25, P198, DOI 10.1016/0037-198X(90)90049-A
   BROWN EW, 1987, AM J NEURORADIOL, V8, P1031
   Cohen B H, 1989, Oncology (Williston Park), V3, P23
   COHEN BH, 1991, PEDIATR NEUROSURG, V16, P66
   DIMARIO FJ, 1993, J CHILD NEUROL, V8, P32, DOI 10.1177/088307389300800105
   DIPAOLO DP, 1995, RADIOLOGY, V195, P721, DOI 10.1148/radiology.195.3.7754001
   EDWARDS MSB, 1989, J NEUROSURG, V70, P691, DOI 10.3171/jns.1989.70.5.0691
   EPSTEIN F, 1985, CANCER, V56, P1804, DOI 10.1002/1097-0142(19851001)56:7+<1804::AID-CNCR2820561316>3.0.CO;2-5
   EPSTEIN F, 1988, J NEURO-ONCOL, V6, P309
   EPSTEIN F, 1987, CONCEPTS PEDIATR NEU, V7, P117
   FERNER RE, 1993, J NEUROL NEUROSUR PS, V56, P492, DOI 10.1136/jnnp.56.5.492
   FREEMAN CR, 1993, INT J RADIAT ONCOL, V27, P197, DOI 10.1016/0360-3016(93)90228-N
   GOLDSTEIN SM, 1989, ARCH NEUROL-CHICAGO, V46, P1222, DOI 10.1001/archneur.1989.00520470086032
   GRAY J, 1987, PEDIATR NEUROL, V3, P335, DOI 10.1016/0887-8994(87)90004-X
   HANS JS, 1984, RADIOLOGY, V150, P705
   HURST RW, 1988, AM J NEURORADIOL, V9, P293
   LASSMAN LP, 1987, LANCET, V29, P913
   LEE BCP, 1985, AM J NEURORADIOL, V6, P159
   LISTERNICK R, 1994, J PEDIATR-US, V125, P63, DOI 10.1016/S0022-3476(94)70122-9
   LUND AM, 1991, EUR J PEDIATR, V150, P835, DOI 10.1007/BF01955002
   MENOR F, 1991, PEDIATR RADIOL, V21, P389, DOI 10.1007/BF02026665
   MILSTEIN JM, 1989, J NEURO-ONCOL, V7, P367, DOI 10.1007/BF02147094
   MIROWITZ SA, 1990, AM J ROENTGENOL, V154, P369, DOI 10.2214/ajr.154.2.2105032
   Packer R J, 1992, Neurosurg Clin N Am, V3, P863
   PACKER RJ, 1993, CANCER-AM CANCER SOC, V72, P1414, DOI 10.1002/1097-0142(19930815)72:4<1414::AID-CNCR2820720442>3.0.CO;2-C
   PETERMAN SB, 1985, NEURORADIOLOGY, V27, P202, DOI 10.1007/BF00344488
   PONT MS, 1992, AM J ROENTGENOL, V158, P1193, DOI 10.2214/ajr.158.6.1590106
   RAFFEL C, 1989, NEUROSURGERY, V25, P959, DOI 10.1227/00006123-198912000-00018
   RUBENSTEIN AE, 1990, NEUROFIBROMATOSIS HD, P51
   SEVICK RJ, 1992, AM J ROENTGENOL, V159, P171, DOI 10.2214/ajr.159.1.1609692
   SHU HH, 1993, AM J ROENTGENOL, V160, P159, DOI 10.2214/ajr.160.1.8416617
   STERN J, 1979, NEUROSURGERY, V4, P524, DOI 10.1227/00006123-197906000-00006
NR 39
TC 82
Z9 84
U1 0
U2 2
PU LIPPINCOTT WILLIAMS & WILKINS
PI PHILADELPHIA
PA TWO COMMERCE SQ, 2001 MARKET ST, PHILADELPHIA, PA 19103 USA
SN 0028-3878
EI 1526-632X
J9 NEUROLOGY
JI Neurology
PD OCT
PY 1995
VL 45
IS 10
BP 1897
EP 1902
DI 10.1212/WNL.45.10.1897
PG 6
WC Clinical Neurology
WE Science Citation Index Expanded (SCI-EXPANDED)
SC Neurosciences & Neurology
GA TA765
UT WOS:A1995TA76500019
PM 7477989
DA 2023-08-27
ER

PT J
AU Schuelke, MR
   Wongthida, P
   Thompson, J
   Kottke, T
   Driscoll, CB
   Huff, AL
   Shim, KG
   Coffey, M
   Pulido, J
   Evgin, L
   Vile, RG
AF Schuelke, Matthew R.
   Wongthida, Phonphimon
   Thompson, Jill
   Kottke, Timothy
   Driscoll, Christopher B.
   Huff, Amanda L.
   Shim, Kevin G.
   Coffey, Matt
   Pulido, Jose
   Evgin, Laura
   Vile, Richard G.
TI Diverse immunotherapies can effectively treat syngeneic brainstem tumors
   in the absence of overt toxicity
SO JOURNAL FOR IMMUNOTHERAPY OF CANCER
LA English
DT Article
DE Immunotherapy; Brainstem; Toxicity; DIPG
ID VESICULAR STOMATITIS-VIRUS; INTRINSIC PONTINE GLIOMA; IMMUNE-RESPONSE;
   GENE-THERAPY; CELL; REOVIRUS; MELANOMA; EFFICACY; TARGETS; STATE
AB Background: Immunotherapy has shown remarkable clinical promise in the treatment of various types of cancers. However, clinical benefits derive from a highly inflammatory mechanism of action. This presents unique challenges for use in pediatric brainstem tumors including diffuse intrinsic pontine glioma (DIPG), since treatment-related inflammation could cause catastrophic toxicity. Therefore, the goal of this study was to investigate whether inflammatory, immunebased therapies are likely to be too dangerous to pursue for the treatment of pediatric brainstem tumors.
   Methods: To complement previous immunotherapy studies using patient-derived xenografts in immunodeficient mice, we developed fully immunocompetent models of immunotherapy using transplantable, syngeneic tumors. These four models - HSVtk/GCV suicide gene immunotherapy, oncolytic viroimmunotherapy, adoptive T cell transfer, and CAR T cell therapy - have been optimized to treat tumors outside of the CNS and induce a broad spectrum of inflammatory profiles, maximizing the chances of observing brainstem toxicity.
   Results: All four models achieved anti-tumor efficacy in the absence of toxicity, with the exception of recombinant vaccinia virus expressing GMCSF, which demonstrated inflammatory toxicity. Histology, imaging, and flow cytometry confirmed the presence of brainstem inflammation in all models. Where used, the addition of immune checkpoint blockade did not introduce toxicity.
   Conclusions: It remains imperative to regard the brainstem with caution for immunotherapeutic intervention. Nonetheless, we show that further careful development of immunotherapies for pediatric brainstem tumors is warranted to harness the potential potency of anti-tumor immune responses, despite their possible toxicity within this anatomically sensitive location.
C1 [Schuelke, Matthew R.; Shim, Kevin G.; Vile, Richard G.] Mayo Clin, Dept Immunol, Rochester, MN 55905 USA.
   [Schuelke, Matthew R.; Shim, Kevin G.] Mayo Clin, Med Scientist Training Program, Rochester, MN 55905 USA.
   [Wongthida, Phonphimon; Thompson, Jill; Kottke, Timothy; Evgin, Laura; Vile, Richard G.] Mayo Clin, Dept Mol Med, Rochester, MN 55905 USA.
   [Driscoll, Christopher B.; Huff, Amanda L.] Mayo Clin, Virol & Gene Therapy Track, Rochester, MN 55905 USA.
   [Coffey, Matt] Oncolyt Biotech Inc, Calgary, AB T2N 1X7, Canada.
   [Pulido, Jose] Mayo Clin, Dept Ophthalmol, Rochester, MN 55905 USA.
   [Vile, Richard G.] Univ Leeds, St JamesUniv Hosp, Fac Med & Hlth, Leeds Canc Res UK Clin Ctr, Leeds, W Yorkshire, England.
C3 Mayo Clinic; Mayo Clinic; Mayo Clinic; Mayo Clinic; Oncolytics Biotech;
   Mayo Clinic; University of Leeds
RP Vile, RG (通讯作者)，Mayo Clin, Dept Immunol, Rochester, MN 55905 USA.; Vile, RG (通讯作者)，Mayo Clin, Dept Mol Med, Rochester, MN 55905 USA.; Vile, RG (通讯作者)，Univ Leeds, St JamesUniv Hosp, Fac Med & Hlth, Leeds Canc Res UK Clin Ctr, Leeds, W Yorkshire, England.
EM
OI Driscoll, Christopher/0000-0002-8629-1798; Evgin,
   Laura/0000-0002-0913-5038
FU European Research Council; Richard M. Schulze Family Foundation; Mayo
   Foundation; Cancer Research UK; National Institutes of Health
   [R01CA175386, R01CA108961, F30CA210334-02]; Shannon O'Hara Foundation;
   Hyundai Hope on Wheels; Mayo Clinic Medical Scientist Training Program
   [NIH 2T32GM065841-16]; Mayo Graduate School
FX The European Research Council; The Richard M. Schulze Family Foundation;
   Mayo Foundation; Cancer Research UK; National Institutes of Health
   (R01CA175386, R01CA108961, F30CA210334-02); Shannon O'Hara Foundation;
   Hyundai Hope on Wheels; Mayo Clinic Medical Scientist Training Program
   (NIH 2T32GM065841-16); Mayo Graduate School.
CR [Anonymous], 2015, FDA APPR TAL LAH TRE
   Askins MA, 2008, J CHILD NEUROL, V23, P1160, DOI 10.1177/0883073808321065
   Benarroch E. E., 2017, MAYO CLIN MED NEUROS
   Benitez-Ribas D, 2018, FRONT ONCOL, V8, DOI 10.3389/fonc.2018.00127
   Buczkowicz P, 2014, ACTA NEUROPATHOL, V128, P573, DOI 10.1007/s00401-014-1319-6
   Caretti V, 2011, BRAIN PATHOL, V21, P441, DOI 10.1111/j.1750-3639.2010.00468.x
   Chheda ZS, 2018, J EXP MED, V215, P141, DOI 10.1084/jem.20171046
   Chiou VL, 2015, J CLIN ONCOL, V33, P3541, DOI 10.1200/JCO.2015.61.6870
   Curtin Sally C, 2016, NCHS Data Brief, P1
   Davila ML, 2014, SCI TRANSL MED, V6, DOI 10.1126/scitranslmed.3008226
   Errington F, 2008, GENE THER, V15, P1257, DOI 10.1038/gt.2008.58
   Fernandez M, 2002, J VIROL, V76, P895, DOI 10.1128/JVI.76.2.895-904.2002
   Fried I, 2018, J NEURO-ONCOL, V136, P189, DOI 10.1007/s11060-017-2643-1
   Garcia LMC, 2016, J IMMUNOL METHODS, V439, P23, DOI 10.1016/j.jim.2016.09.002
   Gauthier J, 2018, CURR RES TRANSL MED, V66, P50, DOI 10.1016/j.retram.2018.03.003
   Green AL, 2015, CURR ONCOL REP, V17, DOI 10.1007/s11912-014-0436-7
   Gust J, 2018, CNS DRUGS, V32, P1091, DOI 10.1007/s40263-018-0582-9
   Heiss JD, 2019, J NEUROSURG-PEDIATR, V23, P333, DOI 10.3171/2018.9.PEDS17225
   Hermans IF, 2004, J IMMUNOL METHODS, V285, P25, DOI 10.1016/j.jim.2003.10.017
   Hofman F.M., 2013, CURR PROTOC IMMUNOL, V103, P4
   HOGQUIST KA, 1994, CELL, V76, P17, DOI 10.1016/0092-8674(94)90169-4
   Johnson DB, 2015, CANCER IMMUNOL RES, V3, P464, DOI 10.1158/2326-6066.CIR-14-0217
   Jubelt B, 2011, CURR NEUROL NEUROSCI, V11, P543, DOI 10.1007/s11910-011-0228-5
   Kicielinski KP, 2014, MOL THER, V22, P1056, DOI 10.1038/mt.2014.21
   Kumari A, 2016, FUTURE ONCOL, V12, P239, DOI 10.2217/fon.15.300
   Lieberman NAP, 2018, NEUROONCOLOGY
   Lin GL, 2018, ACTA NEUROPATHOL COM, V6, DOI 10.1186/s40478-018-0553-x
   Lun XQ, 2010, MOL THER, V18, P1927, DOI 10.1038/mt.2010.183
   Marelli G, 2018, FRONT IMMUNOL, V9, DOI 10.3389/fimmu.2018.00866
   Mitchell LA, 2017, NEURO-ONCOLOGY, V19, P930, DOI 10.1093/neuonc/nox037
   Mount CW, 2018, NAT MED, V24, P572, DOI 10.1038/s41591-018-0006-x
   Mount CW, 2018, NAT MED
   National Cancer Institute, 2017, CAR T CELL THER APPR
   Norelli M, 2018, NAT MED, V24, P739, DOI 10.1038/s41591-018-0036-4
   Ohlfest JR, 2013, J IMMUNOL, V190, P613, DOI 10.4049/jimmunol.1201557
   Overwijk WW, 2003, J EXP MED, V198, P569, DOI 10.1084/jem.20030590
   Park JH, 2018, NEW ENGL J MED, V378, P449, DOI 10.1056/NEJMoa1709919
   Parviainen S, 2015, INT J CANCER, V136, P1065, DOI 10.1002/ijc.29068
   Porter DL, 2015, SCI TRANSL MED, V7, DOI 10.1126/scitranslmed.aac5415
   Prestwich RJ, 2009, CLIN CANCER RES, V15, P4374, DOI 10.1158/1078-0432.CCR-09-0334
   Renner DN, 2016, NEUROTHERAPEUTICS, V13, P226, DOI 10.1007/s13311-015-0407-1
   Riccione K, 2015, JOVE-J VIS EXP, DOI 10.3791/52397
   Rommelfanger DM, 2012, CANCER RES, V72, P4753, DOI 10.1158/0008-5472.CAN-12-0600
   Rosenberg SA, 2011, NAT REV CLIN ONCOL, V8, P577, DOI 10.1038/nrclinonc.2011.116
   Russell SJ, 2012, NAT BIOTECHNOL, V30, P658, DOI 10.1038/nbt.2287
   Saha D, 2017, CANCER CELL, V32, P253, DOI 10.1016/j.ccell.2017.07.006
   Sampson JH, 2014, CLIN CANCER RES, V20, P972, DOI 10.1158/1078-0432.CCR-13-0709
   Samson A, 2018, SCI TRANSL MED, V10, DOI 10.1126/scitranslmed.aam7577
   Shim KG, 2017, MOL THER, V25, P962, DOI 10.1016/j.ymthe.2017.01.023
   Souweidane MM, 2018, LANCET ONCOL, V19, P1040, DOI 10.1016/S1470-2045(18)30322-X
   Suryadevara CM, 2018, ONCOIMMUNOLOGY, V7, DOI 10.1080/2162402X.2018.1434464
   Tejada S, 2018, FRONT ONCOL, V8, DOI 10.3389/fonc.2018.00061
   van Zanten SEMV, 2016, J NEURO-ONCOL, V128, P387, DOI 10.1007/s11060-016-2141-x
   Varela M, 2018, 18 INT S PED NEUR 20
   VILE RG, 1994, CANCER RES, V54, P6228
   Vile RG, 1997, INT J CANCER, V71, P267
   Warren KE, 2012, FRONT ONCOL, V2, DOI 10.3389/fonc.2012.00205
   Wongthida P, 2011, HUM GENE THER, V22, P1343, DOI 10.1089/hum.2010.216
   Wyatt Linda S, 2015, Curr Protoc Microbiol, V39, DOI 10.1002/9780471729259.mc14a04s39
NR 59
TC 8
Z9 8
U1 0
U2 3
PU BMC
PI LONDON
PA CAMPUS, 4 CRINAN ST, LONDON N1 9XW, ENGLAND
SN 2051-1426
J9 J IMMUNOTHER CANCER
JI J. Immunother. Cancer
PD JUL 17
PY 2019
VL 7
AR 188
DI 10.1186/s40425-019-0673-2
PG 13
WC Oncology; Immunology
WE Science Citation Index Expanded (SCI-EXPANDED)
SC Oncology; Immunology
GA IJ5CZ
UT WOS:000475921700001
PM 31315671
OA Green Published, gold
DA 2023-08-27
ER

PT J
AU Poussaint, TY
   Vajapeyam, S
   Ricci, KI
   Panigrahy, A
   Kocak, M
   Kun, LE
   Boyett, JM
   Pollack, IF
   Fouladi, M
AF Poussaint, Tina Young
   Vajapeyam, Sridhar
   Ricci, Kelsey I.
   Panigrahy, Ashok
   Kocak, Mehmet
   Kun, Larry E.
   Boyett, James M.
   Pollack, Ian F.
   Fouladi, Maryam
TI Apparent diffusion coefficient histogram metrics correlate with survival
   in diffuse intrinsic pontine glioma: a report from the Pediatric Brain
   Tumor Consortium
SO NEURO-ONCOLOGY
LA English
DT Article
DE ADC histogram; brain tumor; brainstem glioma; MR diffusion; pediatric
ID HYPERFRACTIONATED RADIATION-THERAPY; STEM GLIOMAS; CHILDREN;
   BEVACIZUMAB; SUBGROUPS; MARKER
AB Background. Diffuse intrinsic pontine glioma (DIPG) is associated with poor survival regardless of therapy. We used volumetric apparent diffusion coefficient (ADC) histogram metrics to determine associations with progression-free survival (PFS) and overall survival (OS) at baseline and after radiation therapy (RT).
   Methods. Baseline and post-RT quantitative ADC histograms were generated from fluid-attenuated inversion recovery (FLAIR) images and enhancement regions of interest. Metrics assessed included number of peaks (ie, unimodal or bimodal), mean and median ADC, standard deviation, mode, skewness, and kurtosis.
   Results. Based on FLAIR images, the majority of tumors had unimodal peaks with significantly shorter average survival. Pre-RT FLAIR mean, mode, and median values were significantly associated with decreased risk of progression; higher pre-RT ADC values had longer PFS on average. Pre-RT FLAIR skewness and standard deviation were significantly associated with increased risk of progression; higher pre-RT FLAIR skewness and standard deviation had shorter PFS. Nonenhancing tumors at baseline showed higher ADC FLAIR mean values, lower kurtosis, and higher PFS. For enhancing tumors at baseline, bimodal enhancement histograms had much worse PFS and OS than unimodal cases and significantly lower mean peak values. Enhancement in tumors only after RT led to significantly shorter PFS and OS than in patients with baseline or no baseline enhancement.
   Conclusions. ADC histogram metrics in DIPG demonstrate significant correlations between diffusion metrics and survival, with lower diffusion values (increased cellularity), increased skewness, and enhancement associated with shorter survival, requiring future investigations in large DIPG clinical trials.
C1 [Poussaint, Tina Young; Vajapeyam, Sridhar; Ricci, Kelsey I.] Boston Childrens Hosp, Dept Radiol, Boston, MA USA.
   [Panigrahy, Ashok] Univ Pittsburgh, Childrens Hosp Pittsburgh, Med Ctr, Dept Radiol, Pittsburgh, PA 15213 USA.
   [Kun, Larry E.] St Jude Childrens Res Hosp, Dept Diagnost Imaging, 332 N Lauderdale St, Memphis, TN 38105 USA.
   [Kocak, Mehmet; Boyett, James M.] St Jude Childrens Res Hosp, Dept Biostat, 332 N Lauderdale St, Memphis, TN 38105 USA.
   [Pollack, Ian F.] Childrens Hosp Pittsburgh, Dept Neurosurg, Pittsburgh, PA 15213 USA.
   [Fouladi, Maryam] Cincinnati Childrens Hosp, Neurooncol Program, Cincinnati, OH USA.
C3 Harvard University; Boston Children's Hospital; Pennsylvania
   Commonwealth System of Higher Education (PCSHE); University of
   Pittsburgh; St Jude Children's Research Hospital; St Jude Children's
   Research Hospital; Pennsylvania Commonwealth System of Higher Education
   (PCSHE); University of Pittsburgh; Cincinnati Children's Hospital
   Medical Center
RP Poussaint, TY (通讯作者)，Boston Childrens Hosp, Div Neuroradiol, Dept Radiol, 300 Longwood Ave, Boston, MA 02115 USA.
EM
OI Kocak, Mehmet/0000-0002-3386-1734; Vajapeyam,
   Sridhar/0000-0002-1782-1919
FU National Institutes of Health [U01 CA81457]; Pediatric Brain Tumor
   Consortium Foundation; Pediatric Brain Tumor Foundation of the United
   States; American Lebanese Syrian Associated Charities
FX This work was supported by the National Institutes of Health (U01
   CA81457); the Pediatric Brain Tumor Consortium Foundation; the Pediatric
   Brain Tumor Foundation of the United States; and the American Lebanese
   Syrian Associated Charities.
CR [Anonymous], 2009, NEOPLASIA, V11, P102, DOI 10.1593/neo.81328
   Buczkowicz P, 2014, ACTA NEUROPATHOL, V128, P573, DOI 10.1007/s00401-014-1319-6
   Buczkowicz P, 2014, NAT GENET, V46, P451, DOI 10.1038/ng.2936
   Bull JG, 2012, EUR RADIOL, V22, P447, DOI 10.1007/s00330-011-2255-7
   Chen HJ, 2010, AM J NEURORADIOL, V31, P1879, DOI 10.3174/ajnr.A2179
   Chenevert TL, 2009, NEUROIMAG CLIN N AM, V19, P559, DOI 10.1016/j.nic.2009.08.009
   Chenevert TL, 2000, JNCI-J NATL CANCER I, V92, P2029, DOI 10.1093/jnci/92.24.2029
   Conway AE, 2014, NEURORADIOLOGY, V56, P405, DOI 10.1007/s00234-014-1348-9
   Gutierrez DR, 2014, AM J NEURORADIOL, V35, P1009, DOI 10.3174/ajnr.A3784
   Hargrove D, 2006, LANCET ONCOL, V7, P241, DOI 10.1016/S1470-2045(06)70615-5
   Hipp SJ, 2011, NEURO-ONCOLOGY, V13, P904, DOI 10.1093/neuonc/nor076
   Jansen MH, 2015, NEURO-ONCOLOGY, V17, P160, DOI 10.1093/neuonc/nou104
   Jenkinson M, 2012, NEUROIMAGE, V62, P782, DOI 10.1016/j.neuroimage.2011.09.015
   Jennings MT, 1996, J NEURO-ONCOL, V28, P207
   Kaplan AM, 1996, PEDIATR NEUROSURG, V24, P185, DOI 10.1159/000121036
   Lober RM, 2014, J NEURO-ONCOL, V117, P175, DOI 10.1007/s11060-014-1375-8
   Lobel U, 2011, AM J NEURORADIOL, V32, P315, DOI 10.3174/ajnr.A2277
   Mulkern RV, 2008, ACAD RADIOL, V15, P1099, DOI 10.1016/j.acra.2008.04.004
   Mulkern RV, 2015, ACAD RADIOL, V22, P363, DOI 10.1016/j.acra.2014.10.006
   Nowosielski M, 2011, NEURORADIOLOGY, V53, P291, DOI 10.1007/s00234-010-0808-0
   PACKER RJ, 1993, CANCER-AM CANCER SOC, V72, P1414, DOI 10.1002/1097-0142(19930815)72:4<1414::AID-CNCR2820720442>3.0.CO;2-C
   PACKER RJ, 1987, INT J RADIAT ONCOL, V13, P1647, DOI 10.1016/0360-3016(87)90160-X
   Pope WB, 2011, AM J NEURORADIOL, V32, P882, DOI 10.3174/ajnr.A2385
   Pope WB, 2012, J NEURO-ONCOL, V108, P491, DOI 10.1007/s11060-012-0847-y
   Pope WB, 2009, RADIOLOGY, V252, P182, DOI 10.1148/radiol.2521081534
   Poussaint TY, 2011, NEURO-ONCOLOGY, V13, P417, DOI 10.1093/neuonc/noq200
   Roujeau T, 2007, J NEUROSURG, V107, P1, DOI 10.3171/PED-07/07/001
   Schindelin J, 2012, NAT METHODS, V9, P676, DOI [10.1038/NMETH.2019, 10.1038/nmeth.2019]
   SMITH RR, 1990, NEURORADIOLOGY, V32, P265, DOI 10.1007/BF00593044
   Sorensen AG, 2001, J CLIN ONCOL, V19, P551, DOI 10.1200/JCO.2001.19.2.551
   Steffen-Smith EA, 2014, BIOMED RES INT, V2014, DOI 10.1155/2014/647356
   Zukotynski KA, 2011, J NUCL MED, V52, P188, DOI 10.2967/jnumed.110.081463
NR 32
TC 48
Z9 50
U1 0
U2 8
PU OXFORD UNIV PRESS INC
PI CARY
PA JOURNALS DEPT, 2001 EVANS RD, CARY, NC 27513 USA
SN 1522-8517
EI 1523-5866
J9 NEURO-ONCOLOGY
JI Neuro-Oncology
PD MAY
PY 2016
VL 18
IS 5
BP 725
EP 734
DI 10.1093/neuonc/nov256
PG 10
WC Oncology; Clinical Neurology
WE Science Citation Index Expanded (SCI-EXPANDED)
SC Oncology; Neurosciences & Neurology
GA DM2BA
UT WOS:000376149700014
PM 26487690
OA Green Published, Bronze
DA 2023-08-27
ER

PT J
AU Foster, E
   McLean, C
   White, O
AF Foster, Emma
   McLean, Catriona
   White, Owen
TI Glioneuronal brainstem tumor - It's all in the eyes
SO JOURNAL OF CLINICAL NEUROSCIENCE
LA English
DT Article
DE Autoimmune diseases; Neuro-ophthalmology; Myasthenia; Primary brain
   tumor; Paraneoplastic syndrome
AB A previously well man presented with several months' history of neurological symptoms including diplopia and balance difficulties. Examination revealed fluctuating neurological deficits, fatigable weakness and slowed saccades. Extensive testing revealed mildly elevated cerebrospinal fluid protein, strongly positive single fiber electromyography and a dorsal pontine lesion at the floor of the 4th ventricle. An autoimmune process was felt to best account for the myasthenic presentation while the differential diagnoses for the brainstem lesion included glioma. Aggressive immunotherapy failed to halt clinical deterioration; over months he developed generalized weakness, aspiration pneumonia and died. Post-mortem analysis revealed glioneuronal tumor infiltration throughout the brainstem, cerebellum and along the meningeal surface. This is an unusual case of an infiltrative brainstem lesion, with the presentation suggesting a primary diagnosis of myasthenia gravis. The progressive nature of the illness, despite aggressive immune therapy, together with slow saccades, underscored a more sinister process. Cerebral imaging should be performed in patients with fluctuating neurological symptoms, progressive deterioration, and ocular, bulbar, respiratory, or pyramidal pattern deficits, and differentials for contrast-enhancing brain lesions should include primary brain tumors. In such cases, biopsy must proceed if the disease is of relatively recent onset, to facilitate diagnosis and maximize treatment opportunities. (C) 2018 Elsevier Ltd. All rights reserved.
C1 [Foster, Emma] Royal Melbourne Hosp, Dept Neurol, Melbourne, Vic, Australia.
   [Foster, Emma; White, Owen] Alfred Hosp, Dept Neurol, Melbourne, Vic, Australia.
   [McLean, Catriona] Alfred Hosp, Dept Anat Pathol, Melbourne, Vic, Australia.
C3 Royal Melbourne Hospital; Florey Institute of Neuroscience & Mental
   Health; Florey Institute of Neuroscience & Mental Health
RP Foster, E (通讯作者)，Royal Melbourne Hosp, 300 Grattan St, Melbourne, Vic 3050, Australia.
EM
OI McLean, Catriona/0000-0002-0302-5727; White, Owen/0000-0002-2836-7344;
   Foster, Emma/0000-0001-8958-3844
CR [Anonymous], 2016, WHO CLASSIFICATION T
   DIRR LY, 1989, NEUROLOGY, V39, P865, DOI 10.1212/WNL.39.6.865
   Guillamo JS, 2001, BRAIN, V124, P2528, DOI 10.1093/brain/124.12.2528
   Lakhan Shaheen E, 2009, J Med Case Rep, V3, P87, DOI 10.1186/1752-1947-3-87
   MCCUE MP, 1993, NEUROLOGY, V43, P377, DOI 10.1212/WNL.43.2.377
NR 5
TC 0
Z9 0
U1 0
U2 2
PU ELSEVIER SCI LTD
PI OXFORD
PA THE BOULEVARD, LANGFORD LANE, KIDLINGTON, OXFORD OX5 1GB, OXON, ENGLAND
SN 0967-5868
EI 1532-2653
J9 J CLIN NEUROSCI
JI J. Clin. Neurosci.
PD FEB
PY 2019
VL 60
BP 151
EP 153
DI 10.1016/j.jocn.2018.10.084
PG 3
WC Clinical Neurology; Neurosciences
WE Science Citation Index Expanded (SCI-EXPANDED)
SC Neurosciences & Neurology
GA HI9CT
UT WOS:000456753600029
PM 30366780
DA 2023-08-27
ER

PT J
AU Green, AL
   Kieran, MW
AF Green, Adam L.
   Kieran, Mark W.
TI Pediatric Brainstem Gliomas: New Understanding Leads to Potential New
   Treatments for Two Very Different Tumors
SO CURRENT ONCOLOGY REPORTS
LA English
DT Article
DE Focal brainstem glioma; Diffuse intrinsic pontine glioma; BRAF; Histone
   H3; ACVR1
ID INTRINSIC PONTINE GLIOMA; LOW-GRADE GLIOMAS; CHILDRENS ONCOLOGY GROUP;
   BEVACIZUMAB PLUS IRINOTECAN; ACTIVATING ACVR1 MUTATIONS; MAPK PATHWAY
   ACTIVATION; ANDERSON CANCER CENTER; PHASE-II TRIAL; PILOCYTIC
   ASTROCYTOMAS; STEREOTACTIC BIOPSY
AB Pediatric brainstem gliomas include low-grade focal brainstemgliomas (FBSG) and high-grade diffuse intrinsic pontine gliomas (DIPG). These tumors share a crucial and eloquent area of the brain as their location, which carries common challenges for treatment. Otherwise, though, these two diseases are very different in terms of presentation, biology, treatment, and prognosis. FBSG usually present with greater than 3 months of symptoms, while DIPG are usually diagnosed within 3 months of symptom onset. Surgery remains the preferred initial treatment for FBSG, with chemotherapy used for persistent, recurrent, or inoperable disease; conversely, radiation is the only known effective treatment for DIPG. Recent developments in biological understanding of both tumors have led to new treatment possibilities. In FBSG, two genetic changes related to BRAF characterize the majority of tumors, and key differences in their biological effects are informing strategies for targeted chemotherapy use. In DIPG, widespread histone H3 and ACVR1 mutations have led to new hope for effective targeted treatments. FBSG has an excellent prognosis, while the long-term survival rate of DIPG tragically remains near zero. In this review, we cover the epidemiology, biology, presentation, imaging characteristics, multimodality treatment, and prognosis of FBSG and DIPG, with a focus on recent biological discoveries.
C1 [Green, Adam L.] Univ Colorado Anschutz Med Campus, Ctr Canc & Blood Disorders, Univ Colorado Sch Med, Pediat Neurooncol,Dept Pediat, Aurora, CO 80045 USA.
   [Green, Adam L.] Univ Colorado Anschutz Med Campus, Childrens Hosp Colorado, Univ Colorado Sch Med, Aurora, CO 80045 USA.
   [Kieran, Mark W.] Harvard Univ, Dana Farber Boston Childrens Canc & Blood Disorde, Div Pediat Neurooncol, Sch Med, Boston, MA 02215 USA.
C3 University of Colorado System; University of Colorado Anschutz Medical
   Campus; Children's Hospital Colorado; University of Colorado System;
   University of Colorado Anschutz Medical Campus; Harvard University;
   Boston Children's Hospital; Dana-Farber Cancer Institute; Harvard
   Medical School
RP Green, AL (通讯作者)，Univ Colorado Anschutz Med Campus, Ctr Canc & Blood Disorders, Univ Colorado Sch Med, Pediat Neurooncol,Dept Pediat, Mail Stop 8302 12800 E 19th Ave,RC1-N, Aurora, CO 80045 USA.
EM
RI Green, Adam/T-9377-2019
OI Kieran, Mark/0000-0003-2184-7692
FU Novartis; GSK
FX Mark W. Kieran has received consultancy fees from Novartis and GSK and
   paid travel accommodations from Novartis.
CR ALBRIGHT AL, 1993, NEUROSURGERY, V33, P1026
   Angelini P, 2011, J NEURO-ONCOL, V101, P75, DOI 10.1007/s11060-010-0224-7
   Astrakas LG, 2004, CLIN CANCER RES, V10, P8220, DOI 10.1158/1078-0432.CCR-04-0603
   Ater JL, 2012, J CLIN ONCOL, V30, P2641, DOI 10.1200/JCO.2011.36.6054
   Bailey S, 2013, EUR J CANCER, V49, P3856, DOI 10.1016/j.ejca.2013.08.006
   Bandopadhayay P, 2014, PEDIATR BLOOD CANCER, V61, P1173, DOI 10.1002/pbc.24958
   Bar EE, 2008, J NEUROPATH EXP NEUR, V67, P878, DOI 10.1097/NEN.0b013e3181845622
   BARKOVICH AJ, 1991, PEDIATR NEUROSURG, V16, P73, DOI 10.1159/000120511
   Barrow J, 2011, NEURO-ONCOLOGY, V13, P212, DOI 10.1093/neuonc/noq158
   Barua NU, 2013, ACTA NEUROCHIR, V155, P1459, DOI 10.1007/s00701-013-1700-6
   Bouffet E, 2012, J CLIN ONCOL, V30, P1358, DOI 10.1200/JCO.2011.34.5843
   Bowers DC, 2000, PEDIATR NEUROSURG, V32, P24, DOI 10.1159/000028893
   Broniscer A, 2006, CANCER, V106, P1364, DOI 10.1002/cncr.21749
   Broniscer A, 2005, CANCER-AM CANCER SOC, V103, P133, DOI 10.1002/cncr.20741
   Broniscer A, 2013, CLIN CANCER RES, V19, P3050, DOI 10.1158/1078-0432.CCR-13-0306
   Broniscer A, 2010, CANCER-AM CANCER SOC, V116, P4632, DOI 10.1002/cncr.25405
   Buczkowicz P, 2014, NAT GENET, V46, P451, DOI 10.1038/ng.2936
   Buczkowicz P, 2013, BRAIN PATHOL, V23, P244, DOI 10.1111/j.1750-3639.2012.00633.x
   Caretti V, 2013, NEUROPATH APPL NEURO, V39, P426, DOI 10.1111/j.1365-2990.2012.01294.x
   Chassot A, 2012, J NEURO-ONCOL, V106, P399, DOI 10.1007/s11060-011-0681-7
   Chen HJ, 2010, AM J NEURORADIOL, V31, P1879, DOI 10.3174/ajnr.A2179
   Chiang KL, 2010, CHILD NERV SYST, V26, P1035, DOI 10.1007/s00381-010-1106-1
   Cohen KJ, 2011, NEURO-ONCOLOGY, V13, P410, DOI 10.1093/neuonc/noq205
   Darken RS, 2009, J PEDIAT HEMATOL ONC, V31, P972, DOI 10.1097/MPH.0b013e3181b8701f
   Di Maio S, 2009, CHILD NERV SYST, V25, P1401, DOI 10.1007/s00381-009-0956-x
   Dias-Santagata D, 2011, PLOS ONE, V6, DOI 10.1371/journal.pone.0017948
   Khuong-Quang DA, 2012, ACTA NEUROPATHOL, V124, P439, DOI 10.1007/s00401-012-0998-0
   EPSTEIN FJ, 1993, J NEUROSURG, V78, P408, DOI 10.3171/jns.1993.78.3.0408
   Fisher PG, 2000, CANCER, V89, P1569, DOI 10.1002/1097-0142(20001001)89:7<1569::AID-CNCR22>3.0.CO;2-0
   Fontanilla HP, 2012, AM J CLIN ONCOL-CANC, V35, P51, DOI 10.1097/COC.0b013e318201a2b7
   Fontebasso AM, 2014, NAT GENET, V46, P462, DOI 10.1038/ng.2950
   Fouladi M, 2007, CANCER-AM CANCER SOC, V110, P2535, DOI 10.1002/cncr.23078
   Geoerger B, 2011, NEURO-ONCOLOGY, V13, P109, DOI 10.1093/neuonc/noq141
   Grill J, 2012, PEDIATR BLOOD CANCER, V58, P489, DOI 10.1002/pbc.24060
   Gururangan S, 2002, J CLIN ONCOL, V20, P2951, DOI 10.1200/JCO.2002.12.008
   Gururangan S, 2014, NEURO-ONCOLOGY, V16, P310, DOI 10.1093/neuonc/not154
   Gururangan S, 2010, J CLIN ONCOL, V28, P3069, DOI 10.1200/JCO.2009.26.8789
   Haas-Kogan DA, 2008, NEURO-ONCOLOGY, V10, P341, DOI 10.1215/15228517-2008-004
   Hankinson TC, 2011, J NEUROSURG-PEDIATR, V8, P97, DOI 10.3171/2011.4.PEDS1180
   Hargrave D, 2008, J NEURO-ONCOL, V86, P313, DOI 10.1007/s11060-007-9473-5
   Hashizume R, 2008, NEURO-ONCOLOGY, V10, P112, DOI 10.1215/15228517-2007-052
   Hashizume R, 2012, J NEURO-ONCOL, V110, P305, DOI 10.1007/s11060-012-0973-6
   Hwang EI, 2013, PEDIATR BLOOD CANCER, V60, P776, DOI 10.1002/pbc.24297
   Jackson S, 2013, J NEURO-ONCOL, V114, P339, DOI 10.1007/s11060-013-1189-0
   Jacob K, 2009, BRIT J CANCER, V101, P722, DOI 10.1038/sj.bjc.6605179
   Jalali R, 2010, INT J RADIAT ONCOL, V77, P113, DOI 10.1016/j.ijrobp.2009.04.031
   Janssens GO, 2013, INT J RADIAT ONCOL, V85, P315, DOI 10.1016/j.ijrobp.2012.04.006
   Janssens GORJ, 2009, INT J RADIAT ONCOL, V73, P722, DOI 10.1016/j.ijrobp.2008.05.030
   Jeuken JWM, 2010, J PATHOL, V222, P324, DOI 10.1002/path.2780
   Joshi BH, 2008, NEURO-ONCOLOGY, V10, P265, DOI 10.1215/15228517-2007-066
   Karajannis MA, 2014, NEURO-ONCOLOGY, V16, P1408, DOI 10.1093/neuonc/nou059
   Khaw SL, 2007, PEDIATR BLOOD CANCER, V49, P808, DOI 10.1002/pbc.21270
   Kieran M, 2014, NEURO-ONCOLOGY, V16, P56
   Kieran MW, 2015, PEDIATR BLOOD CANCER, V62, P3, DOI 10.1002/pbc.25266
   Kieran Mark W, 2014, Am Soc Clin Oncol Educ Book, pe436, DOI 10.14694/EdBook_AM.2014.34.e436
   Ko C, 2012, INT J RADIAT ONCOL, V83, P1252, DOI 10.1016/j.ijrobp.2011.09.046
   Lazareff JA, 1998, NEUROSURGERY, V43, P809, DOI 10.1097/00006123-199810000-00053
   Lewis PW, 2013, SCIENCE, V340, P857, DOI 10.1126/science.1232245
   Li Khan W, 2005, Neurosurg Focus, V18, pE2
   Lobel U, 2011, AM J NEURORADIOL, V32, P315, DOI 10.3174/ajnr.A2277
   Lobel U, 2010, NEURORADIOLOGY, V52, P1167, DOI 10.1007/s00234-010-0771-9
   Lui YW, 2007, NEUROSURGERY, V61, P1199, DOI 10.1227/01.neu.0000306098.38141.81
   MacConaill LE, 2009, PLOS ONE, V4, DOI 10.1371/journal.pone.0007887
   Massimino M, 2008, J NEURO-ONCOL, V87, P355, DOI 10.1007/s11060-008-9525-5
   Michalski A, 2010, J NEURO-ONCOL, V100, P81, DOI 10.1007/s11060-010-0141-9
   Negretti L, 2011, J NEURO-ONCOL, V104, P773, DOI 10.1007/s11060-011-0542-4
   Nicholson HS, 2007, CANCER, V110, P1542, DOI 10.1002/cncr.22961
   Ogiwara H, 2013, CHILD NERV SYST, V29, P833, DOI 10.1007/s00381-013-2042-7
   Ohno M, 2009, PEDIATR BLOOD CANCER, V53, P37, DOI 10.1002/pbc.21987
   PACKER RJ, 1993, J CLIN ONCOL, V11, P850, DOI 10.1200/JCO.1993.11.5.850
   Packer RJ, 1997, J NEUROSURG, V86, P747, DOI 10.3171/jns.1997.86.5.0747
   Panandiker ASP, 2014, PEDIATR BLOOD CANCER, V61, P1180, DOI [10.1002/pbc.24958, 10.1002/pbc.24971]
   Park KY, 2007, J NEUROSURG, V107, P251, DOI 10.3171/PED-07/09/251
   Patankar AP, 2013, BRIT J NEUROSURG, V27, P256, DOI 10.3109/02688697.2012.741738
   Paugh BS, 2011, J CLIN ONCOL, V29, P3999, DOI 10.1200/JCO.2011.35.5677
   Perez-Gomez JL, 2010, CHILD NERV SYST, V26, P29, DOI 10.1007/s00381-009-1000-x
   Pfister S, 2008, J CLIN INVEST, V118, P1739, DOI 10.1172/JCI33656
   PIERREKAHN A, 1993, J NEUROSURG, V79, P845, DOI 10.3171/jns.1993.79.6.0845
   Pirotte BJM, 2007, J NEUROSURG, V107, P392, DOI 10.3171/PED-07/11/392
   Pollack IF, 2011, NEURO-ONCOLOGY, V13, P290, DOI 10.1093/neuonc/noq199
   Poulikakos PI, 2010, NATURE, V464, P427, DOI 10.1038/nature08902
   Poussaint TY, 2011, NEURO-ONCOLOGY, V13, P417, DOI 10.1093/neuonc/noq200
   Prabhu SP, 2011, CHILD NERV SYST, V27, P11, DOI 10.1007/s00381-010-1323-7
   Prados MD, 1997, J NEURO-ONCOL, V32, P235, DOI 10.1023/A:1005736104205
   Puget S, 2012, PLOS ONE, V7, DOI 10.1371/journal.pone.0030313
   Rajshekhar V, 2010, STEREOT FUNCT NEUROS, V88, P360, DOI 10.1159/000319044
   Recinos PF, 2007, PEDIATR NEUROSURG, V43, P192, DOI 10.1159/000098831
   ROBERTSON PL, 1995, PEDIATR NEUROSURG, V22, P65, DOI 10.1159/000120879
   ROBERTSON PL, 1994, NEUROLOGY, V44, P1798, DOI 10.1212/WNL.44.10.1798
   Robison NJ, 2014, PEDIATR BLOOD CANCER, V61, P636, DOI 10.1002/pbc.24794
   Ronghe M, 2010, PEDIATR BLOOD CANCER, V55, P471, DOI 10.1002/pbc.22557
   Saratsis AM, 2012, NEURO-ONCOLOGY, V14, P547, DOI 10.1093/neuonc/nos067
   Schindler G, 2011, ACTA NEUROPATHOL, V121, P397, DOI 10.1007/s00401-011-0802-6
   Schwartzentruber J, 2012, NATURE, V482, P226, DOI [10.1038/nature10833, 10.1007/s13311-012-0137-6]
   Sedlacik J, 2013, AM J NEURORADIOL, V34, P1450, DOI 10.3174/ajnr.A3421
   Sethi R, 2011, J NEURO-ONCOL, V102, P121, DOI 10.1007/s11060-010-0301-y
   Sharp JR, 2010, EUR J CANCER, V46, P3271, DOI 10.1016/j.ejca.2010.06.115
   Sievert AJ, 2013, P NATL ACAD SCI USA, V110, P5957, DOI 10.1073/pnas.1219232110
   Siu IM, 2010, J NEUROSURG-PEDIATR, V6, P92, DOI 10.3171/2010.3.PEDS09366
   Steffen-Smith EA, 2012, INT J RADIAT ONCOL, V84, P774, DOI 10.1016/j.ijrobp.2012.01.032
   Steffen-Smith EA, 2011, J NEURO-ONCOL, V105, P365, DOI 10.1007/s11060-011-0601-x
   Sun B, 1999, NEURORADIOLOGY, V41, P158, DOI 10.1007/s002340050723
   Taylor KR, 2014, NAT GENET, V46, P457, DOI 10.1038/ng.2925
   Teo C, 2008, CHILD NERV SYST, V24, P1307, DOI 10.1007/s00381-008-0647-z
   Ternier J, 2006, J NEUROSURG, V104, P369, DOI 10.3171/ped.2006.104.6.369
   Tronnier VM, 1997, NEUROSURGERY, V40, P891, DOI 10.1097/00006123-199705000-00001
   Van Poppel K, 2009, PEDIATR BLOOD CANCER, V53, P1355, DOI 10.1002/pbc.22232
   Veringa SJE, 2013, PLOS ONE, V8, DOI 10.1371/journal.pone.0061512
   Walker DA, 2013, NEURO-ONCOLOGY, V15, P462, DOI 10.1093/neuonc/nos330
   Wang ZHJ, 2015, PEDIATR BLOOD CANCER, V62, P163, DOI 10.1002/pbc.25224
   Warren K, 2012, CANCER-AM CANCER SOC, V118, P3607, DOI 10.1002/cncr.26659
   Warren KE, 2012, J NEURO-ONCOL, V106, P643, DOI 10.1007/s11060-011-0709-z
   Warren KE, 2000, J CLIN ONCOL, V18, P1020, DOI 10.1200/JCO.2000.18.5.1020
   Wisoff JH, 2011, NEUROSURGERY, V68, P1548, DOI 10.1227/NEU.0b013e318214a66e
   Wolff JE, 2012, J NEURO-ONCOL, V106, P391, DOI 10.1007/s11060-011-0677-3
   Wolff JE, 2011, ANTICANCER RES, V31, P2265
   Wu G, 2014, NAT GENET, V46, P444, DOI 10.1038/ng.2938
   Wu G, 2012, NAT GENET, V44, P251, DOI 10.1038/ng.1102
   Yamasaki F, 2011, NEURO-ONCOLOGY, V13, P791, DOI 10.1093/neuonc/nor038
   Yin LX, 2013, CAN J NEUROL SCI, V40, P348, DOI 10.1017/S0317167100014293
   Yoshimura J, 2003, NEUROL MED-CHIR, V43, P375, DOI 10.2176/nmc.43.375
   Zaghloul MS, 2014, RADIOTHER ONCOL, V111, P35, DOI 10.1016/j.radonc.2014.01.013
   Zhou ZP, 2013, J NEURO-ONCOL, V111, P257, DOI 10.1007/s11060-012-1021-2
NR 123
TC 36
Z9 40
U1 0
U2 11
PU SPRINGER
PI NEW YORK
PA 233 SPRING ST, NEW YORK, NY 10013 USA
SN 1523-3790
EI 1534-6269
J9 CURR ONCOL REP
JI Curr. Oncol. Rep.
PD MAR
PY 2015
VL 17
IS 3
AR 12
DI 10.1007/s11912-014-0436-7
PG 11
WC Oncology
WE Science Citation Index Expanded (SCI-EXPANDED)
SC Oncology
GA CC3GQ
UT WOS:000350235200006
PM 25702179
DA 2023-08-27
ER

PT J
AU Ogiwara, H
   Morota, N
AF Ogiwara, Hideki
   Morota, Nobuhito
TI The efficacy of a biopsy of intrinsic brainstem lesions for decision
   making of the treatments
SO CHILDS NERVOUS SYSTEM
LA English
DT Article
DE Pediatric brainstem tumors; Open biopsy; Diffuse intrinsic pontine
   glioma
ID PRIMITIVE NEUROECTODERMAL TUMORS; STEREOTACTIC BIOPSY; DIAGNOSIS;
   SURGERY; GLIOMAS
AB Object The diagnosis of diffuse intrinsic pontine gliomas (DIPGs) has been generally made mainly by magnetic resonance imaging (MRI) and clinical course. However, the accuracy of MRI-based diagnosis has not been fully confirmed yet. Our aim was to review efficacy of biopsy for decision making of the treatments.
   Methods We retrospectively analyzed pediatric patients undergoing biopsy for intrinsic brainstem lesions which were considered atypical for DIPGs by MRI findings. The lesion was evaluated atypical when it extended beyond the pons or it had a well-margined localized enhancing portion.
   Results Seven patients underwent biopsy. Preoperative MRI revealed a lesion extending beyond the pons in five patients and a focal enhancing lesion in four. Two patients had both of these. Open biopsy was performed via midline suboccipital approach in six patients and retrosigmoid approach in one. No intraoperative complications were observed. Histopathological examination revealed diffuse brainstem glioma in five patients, primitive neuroectodermal tumor (PNET) in one, and pilocytic astrocytoma in one. In the case with PNET, chemotherapy and radiotherapy were effective and the patient had been stable for 12 months without recurrence. The patient with pilocytic astrocytoma did not undergo radiotherapy and has been stable without regrowth of the tumor for 9 months.
   Conclusions Open biopsy of intrinsic brainstem lesions is considered to be safe and effective for selecting an appropriate course of therapy. Patients with intrinsic pontine lesions which extend beyond the pons or with localized enhancing portion seem to be benefited from the biopsy.
C1 [Ogiwara, Hideki; Morota, Nobuhito] Natl Ctr Child Hlth & Dev, Div Neurosurg, Setagaya Ku, Tokyo 1578535, Japan.
C3 National Center for Child Health & Development - Japan
RP Ogiwara, H (通讯作者)，Natl Ctr Child Hlth & Dev, Div Neurosurg, Setagaya Ku, Okura 2-10-1, Tokyo 1578535, Japan.
EM
CR ALBRIGHT AL, 1993, NEUROSURGERY, V33, P1026
   Blond S, 1991, Acta Neurochir Suppl (Wien), V52, P75
   Cartmill M, 1999, CHILD NERV SYST, V15, P235
   COFFEY RJ, 1985, APPL NEUROPHYSIOL, V48, P467
   de Leon FCP, 2003, CHILD NERV SYST, V19, P305, DOI 10.1007/s00381-003-0737-x
   Dellaretti M, 2011, J NEUROSURG-PEDIATR, V8, P539, DOI 10.3171/2011.9.PEDS1167
   Epstein F, 1996, PEDIATR NEUROSURG, V24, P24, DOI 10.1159/000121011
   Frank F, 1988, Acta Neurochir Suppl (Wien), V42, P177
   Franzini A, 1988, Acta Neurochir Suppl (Wien), V42, P170
   Giunta F, 1989, Acta Neurochir Suppl (Wien), V46, P86
   GODANO U, 1987, CHILD NERV SYST, V3, P85, DOI 10.1007/BF00271129
   Goncalves-Ferreira AJ, 2003, SURG NEUROL, V60, P311, DOI 10.1016/S0090-3019(03)00379-3
   MOROTA N, 1995, NEUROSURGERY, V37, P922, DOI 10.1227/00006123-199511000-00011
   Morota N, 2010, CHILD NERV SYST, V26, P513, DOI 10.1007/s00381-009-1080-7
   Pincus DW, 2006, J NEUROSURG, V104, P108, DOI 10.3171/ped.2006.104.2.108
   Rachinger W, 2009, J NEUROL NEUROSUR PS, V80, P1134, DOI 10.1136/jnnp.2009.174250
   Roujeau T, 2007, J NEUROSURG, V107, P1, DOI 10.3171/PED-07/07/001
   St George EJ, 2004, CHILD NERV SYST, V20, P163, DOI 10.1007/s00381-003-0897-8
   STECK J, 1995, SURG NEUROL, V43, P563, DOI 10.1016/0090-3019(95)00156-5
   Sufit A, 2012, J NEUROSURG-PEDIATR, V10, P81, DOI 10.3171/2012.3.PEDS11316
   Valdes-Gorcia J, 1998, ACTA NEUROCHIR, V140, P899, DOI 10.1007/s007010050191
   Zagzag D, 2000, PEDIATRICS, V106, P1045, DOI 10.1542/peds.106.5.1045
NR 22
TC 13
Z9 13
U1 0
U2 9
PU SPRINGER
PI NEW YORK
PA 233 SPRING ST, NEW YORK, NY 10013 USA
SN 0256-7040
EI 1433-0350
J9 CHILD NERV SYST
JI Childs Nerv. Syst.
PD MAY
PY 2013
VL 29
IS 5
BP 833
EP 837
DI 10.1007/s00381-013-2042-7
PG 5
WC Clinical Neurology; Pediatrics; Surgery
WE Science Citation Index Expanded (SCI-EXPANDED)
SC Neurosciences & Neurology; Pediatrics; Surgery
GA 148EZ
UT WOS:000319227200019
PM 23377758
DA 2023-08-27
ER

PT J
AU Jung, IH
   Chang, KW
   Park, SH
   Moon, JH
   Kim, EH
   Jung, HH
   Kang, SG
   Chang, JH
   Chang, JW
   Chang, WS
AF Jung, In-Ho
   Chang, Kyung W.
   Park, So H.
   Moon, Ju H.
   Kim, Eui H.
   Jung, Hyun H.
   Kang, Seok-Gu
   Chang, Jong H.
   Chang, Jin W.
   Chang, Won S.
TI Stereotactic biopsy for adult brainstem lesions: A surgical approach and
   its diagnostic value according to the 2016 World Health Organization
   Classification
SO CANCER MEDICINE
LA English
DT Article
DE brainstem; diffuse midline glioma; medulla oblongata; midbrain; pons;
   Stereotactic biopsy
ID MAGNETIC-RESONANCE-SPECTROSCOPY; INTRINSIC PONTINE GLIOMA;
   CENTRAL-NERVOUS-SYSTEM; HISTONE H3.3; TUMORS; CHILDHOOD; CHILDREN;
   SAFETY; METAANALYSIS; SERIES
AB Background The brainstem has the critical role of regulating cardiac and respiratory function and it also provides motor and sensory function to the face via the cranial nerves. Despite the observation of a brainstem lesion in a radiological examination, it is difficult to obtain tissues for a pathological diagnosis because of the location and small volume of the brainstem. Thus, we aimed to share our 6-year experience with stereotactic biopsies from brainstem lesions and confirm the value and safety of stereotactic biopsy on this highly eloquent area in this study. Methods We retrospectively reviewed the medical records of 42 adult patients who underwent stereotactic biopsy on brainstem lesions from 2015 to 2020. The radiological findings, surgical records, pathological diagnosis, and postoperative complications of all patients were analyzed. Results Histopathological diagnoses were made in 40 (95.2%) patients. Astrocytic tumors were diagnosed in 29 (69.0%) patients, diffuse large B cell lymphoma in 5 (11.9%) patients, demyelinating disease in 4 (9.5%) patients, germinoma in 1 (2.4%) patient, and radiation necrosis in 1 (2.4%) patient. In the 40 patients with successful stereotactic biopsy, 10 (25.0%) patients had inconsistent preoperative radiological diagnosis and postoperative pathological diagnosis. In addition, there was a difference between the treatments prescribed by the radiological and pathological diagnoses in 8 out of 10 patients whose diagnoses changed after biopsy. There was no operative mortality among the 42 patients. Conclusions A pathological diagnosis can be made safely and efficiently in brainstem lesions using stereotactic biopsy. This pathological diagnosis will enable patients to receive appropriate treatment.
C1 [Jung, In-Ho; Chang, Kyung W.; Park, So H.; Jung, Hyun H.; Chang, Jin W.; Chang, Won S.] Yonsei Univ, Brain Res Inst, Dept Neurosurg, Coll Med, 50-1 Yonsei Ro, Seoul 03722, South Korea.
   [Jung, In-Ho; Moon, Ju H.; Kim, Eui H.; Kang, Seok-Gu; Chang, Jong H.] Yonsei Univ, Brain Tumor Ctr, Dept Neurosurg, Coll Med, Seoul, South Korea.
C3 Yonsei University; Yonsei University Health System; Yonsei University;
   Yonsei University Health System
RP Chang, WS (通讯作者)，Yonsei Univ, Brain Res Inst, Dept Neurosurg, Coll Med, 50-1 Yonsei Ro, Seoul 03722, South Korea.
EM
OI Chang, Won Seok/0000-0003-3145-4016; Chang, Jong
   Hee/0000-0003-1509-9800; Kim, Eui Hyun/0000-0002-2523-7122; Park, So
   Hee/0000-0003-0114-8587; Chang, Jin Woo/0000-0002-2717-0101; Kang,
   Seok-Gu/0000-0001-5676-2037
FU Samsung Research Funding & Incubation Center of Samsung Electronics
   [SRFC-IT2001-01]
FX Samsung Research Funding & Incubation Center of Samsung Electronics,
   Grant/Award Number: SRFC-IT2001-01
CR Aker FV, 2005, NEUROPATHOLOGY, V25, P207, DOI 10.1111/j.1440-1789.2005.00634.x
   Al-Saiari S, 2020, CUREUS J MED SCIENCE, V12, DOI 10.7759/cureus.12002
   ALBRIGHT AL, 1993, NEUROSURGERY, V33, P1026
   Amundson EW, 2005, J NEUROSURG, V102, P565, DOI 10.3171/jns.2005.102.3.0565
   Buczkowicz P, 2014, ACTA NEUROPATHOL, V128, P573, DOI 10.1007/s00401-014-1319-6
   Cartmill M, 1999, CHILD NERV SYST, V15, P235
   Castel D, 2016, ACTA NEUROPATHOL, V131, P795, DOI 10.1007/s00401-016-1568-7
   Cavalheiro S, 2015, CHILD NERV SYST, V31, P1815, DOI 10.1007/s00381-015-2799-y
   Chang WS, 2011, ACTA NEUROCHIR, V153, P1087, DOI 10.1007/s00701-011-0953-1
   Chen SY, 2011, J CHIN MED ASSOC, V74, P110, DOI 10.1016/j.jcma.2011.01.024
   Chen X, 2007, ACTA NEUROCHIR, V149, P1117, DOI 10.1007/s00701-007-1282-2
   Curless RG, 2002, PEDIATR NEUROL, V26, P219, DOI 10.1016/S0887-8994(01)00363-0
   Davison JE, 2011, J CHILD NEUROL, V26, P356, DOI 10.1177/0883073810381279
   Dellaretti M, 2012, STEREOT FUNCT NEUROS, V90, P79, DOI 10.1159/000335502
   Dellaretti M, 2011, J NEUROSURG-PEDIATR, V8, P539, DOI 10.3171/2011.9.PEDS1167
   Khuong-Quang DA, 2012, ACTA NEUROPATHOL, V124, P439, DOI 10.1007/s00401-012-0998-0
   Guillamo JS, 2001, CURR OPIN NEUROL, V14, P711, DOI 10.1097/00019052-200112000-00006
   Hamisch C, 2017, J NEUROSURG-PEDIATR, V20, P261, DOI 10.3171/2017.2.PEDS1665
   Hankinson TC, 2011, J NEUROSURG-PEDIATR, V8, P97, DOI 10.3171/2011.4.PEDS1180
   Hersh DS, 2020, J NEUROSURG-PEDIATR, V26, P552, DOI 10.3171/2020.4.PEDS2092
   HOOD TW, 1986, J NEUROSURG, V65, P172, DOI 10.3171/jns.1986.65.2.0172
   Jadhav V, 2007, SURG NEUROL, V67, P15, DOI 10.1016/j.surneu.2006.07.014
   Kickingereder P, 2013, NEUROSURGERY, V72, P873, DOI 10.1227/NEU.0b013e31828bf445
   Komori T, 2017, NEUROL MED-CHIR, V57, P301, DOI 10.2176/nmc.ra.2017-0010
   Kornreich L, 2005, PEDIATR RADIOL, V35, P872, DOI 10.1007/s00247-005-1502-y
   Louis DN, 2016, ACTA NEUROPATHOL, V131, P803, DOI 10.1007/s00401-016-1545-1
   Manoj N, 2014, J NEUROSCI RURAL PRA, V5, P32, DOI 10.4103/0976-3147.127869
   Minchev G, 2020, OPER NEUROSURG, V19, P292, DOI 10.1093/ons/opaa123
   Porto L, 2007, CHILD NERV SYST, V23, P305, DOI 10.1007/s00381-006-0221-5
   Puget S, 2015, CHILD NERV SYST, V31, P1773, DOI 10.1007/s00381-015-2832-1
   Rachinger W, 2009, J NEUROL NEUROSUR PS, V80, P1134, DOI 10.1136/jnnp.2009.174250
   Schumacher M, 2007, J NEUROSURG, V106, P111, DOI 10.3171/ped.2007.106.2.111
   Schwartzentruber J, 2012, NATURE, V482, P226, DOI [10.1038/nature10833, 10.1007/s13311-012-0137-6]
   Smith JK, 2002, NEURORADIOLOGY, V44, P825, DOI 10.1007/s00234-002.0821-z
   STECK J, 1995, SURG NEUROL, V43, P563, DOI 10.1016/0090-3019(95)00156-5
   Taweesomboonyat C, 2019, J NEUROSCI RURAL PRA, V10, P78, DOI 10.4103/jnrp.jnrp_166_18
   Toma AK, 2011, NEUROSURGERY, V68, P245, DOI 10.1227/NEU.0b013e318214a809
   Wu G, 2012, NAT GENET, V44, P251, DOI 10.1038/ng.1102
   Wu SQ, 2021, BRIT J NEUROSURG, V35, P319, DOI 10.1080/02688697.2020.1812519
   Xie S, 2021, NEUROSURG REV, V44, P423, DOI 10.1007/s10143-019-01230-0
   Zhou CH, 2021, J CLIN NEUROL, V17, P220, DOI 10.3988/jcn.2021.17.2.220
NR 41
TC 5
Z9 5
U1 1
U2 2
PU WILEY
PI HOBOKEN
PA 111 RIVER ST, HOBOKEN 07030-5774, NJ USA
SN 2045-7634
J9 CANCER MED-US
JI Cancer Med.
PD NOV
PY 2021
VL 10
IS 21
BP 7514
EP 7524
DI 10.1002/cam4.4272
EA SEP 2021
PG 11
WC Oncology
WE Science Citation Index Expanded (SCI-EXPANDED)
SC Oncology
GA WP6WI
UT WOS:000694978800001
PM 34510820
OA Green Published
DA 2023-08-27
ER

PT J
AU Grimm, SA
   Chamberlain, MC
AF Grimm, Sean A.
   Chamberlain, Marc C.
TI Brainstem Glioma: A Review
SO CURRENT NEUROLOGY AND NEUROSCIENCE REPORTS
LA English
DT Review
DE Brainstem glioma; Pontine glioma; Diffuse intrinsic pontine glioma
ID INTRINSIC PONTINE GLIOMA; PEDIATRIC-ONCOLOGY-GROUP; PHASE-I TRIAL;
   HYPERFRACTIONATED RADIOTHERAPY; RADIATION-THERAPY; CHILDREN; TUMORS;
   CHEMOTHERAPY; MANAGEMENT; TEMOZOLOMIDE
AB Brainstem gliomas (BGs) are a heterogenous group of gliomas that occur predominately in children. They can be separated into groups on the basis of anatomy and clinical behavior: diffuse intrinsic pontine glioma (DIPG), exophytic medullary glioma, and tectal glioma. DIPG is the commonest BG. Median age at onset is 6.5 years and median survival is less than 1 year. Adults with DIPG survive longer, suggesting a less aggressive and biologically different tumor from that in children. Patients present with cranial nerve dysfunction, long tract signs, or ataxia, either in isolation or in combination. Magnetic resonance imaging shows an infiltrative lesion occupying most of the pons and contrast enhancement is usually not prominent. Standard treatment is fractionated radiotherapy. Platelet-derived growth factor receptor alpha and epidermal growth factor receptor mutations have been identified. Inhibitors of these growth factor receptors are being evaluated in clinical trials. Exophytic medullary and tectal gliomas are relatively indolent tumors that can often be followed closely without treatment.
C1 [Grimm, Sean A.] Univ Minnesota, Sch Med, Dept Neurol, Minneapolis, MN 55455 USA.
   [Chamberlain, Marc C.] Univ Washington, Fred Hutchinson Canc Res Ctr, Seattle Canc Care Alliance, Dept Neurol & Neurol Surg, Seattle, WA 98109 USA.
C3 University of Minnesota System; University of Minnesota Twin Cities;
   Fred Hutchinson Cancer Center; Seattle Cancer Care Alliance; University
   of Washington; University of Washington Seattle
RP Chamberlain, MC (通讯作者)，Univ Washington, Fred Hutchinson Canc Res Ctr, Seattle Canc Care Alliance, Dept Neurol & Neurol Surg, 825 Eastlake Ave E,POB 19023,MS-G4940, Seattle, WA 98109 USA.
EM
CR Aguilar LK, 2012, CURR TREAT OPTION ON, V13, P437, DOI 10.1007/s11864-012-0208-2
   Allen J, 1999, CANCER, V86, P1064, DOI 10.1002/(SICI)1097-0142(19990915)86:6<1064::AID-CNCR24>3.0.CO;2-1
   Bartels U, 2011, J NEURO-ONCOL, V105, P119, DOI 10.1007/s11060-011-0704-4
   Bernier-Chastagner V, 2005, CANCER-AM CANCER SOC, V104, P2792, DOI 10.1002/cncr.21534
   BOGNAR L, 1994, ACTA NEUROCHIR, V127, P48, DOI 10.1007/BF01808546
   Bowers DC, 2000, PEDIATR NEUROSURG, V32, P24, DOI 10.1159/000028893
   Bradley KA, 2013, INT J RADIAT ONCOL, V85, pE55, DOI 10.1016/j.ijrobp.2012.09.004
   Broniscer A, 2010, J CLIN ONCOL, V28, P4762, DOI 10.1200/JCO.2010.30.3545
   Chassot A, 2012, J NEURO-ONCOL, V106, P399, DOI 10.1007/s11060-011-0681-7
   Cohen KJ, 2011, NEURO-ONCOLOGY, V13, P410, DOI 10.1093/neuonc/noq205
   Dolecek TA, 2012, NEURO-ONCOLOGY, V14, pv1, DOI 10.1093/neuonc/nos218
   Donaldson SS, 2006, J CLIN ONCOL, V24, P1266, DOI 10.1200/JCO.2005.04.6599
   Fangusaro J, 2009, J CHILD NEUROL, V24, P1409, DOI 10.1177/0883073809338960
   Fisher PG, 2000, CANCER, V89, P1569, DOI 10.1002/1097-0142(20001001)89:7<1569::AID-CNCR22>3.0.CO;2-0
   Fouladi M, 2010, J CLIN ONCOL, V28, P4221, DOI 10.1200/JCO.2010.28.4687
   Freeman CR, 1999, CHILD NERV SYST, V15, P545, DOI 10.1007/s003810050542
   Freeman CR, 2000, INT J RADIAT ONCOL, V47, P561, DOI 10.1016/S0360-3016(00)00471-5
   Freeman CR, 1998, INT J RADIAT ONCOL, V40, P265, DOI 10.1016/S0360-3016(97)00572-5
   Geoerger B, 2011, NEURO-ONCOLOGY, V13, P109, DOI 10.1093/neuonc/noq141
   Geyer JR, 2010, EUR J CANCER, V46, P3287, DOI 10.1016/j.ejca.2010.07.005
   Gilbertson RJ, 2003, CLIN CANCER RES, V9, P3620
   Guillamo JS, 2001, BRAIN, V124, P2528, DOI 10.1093/brain/124.12.2528
   Gururangan S, 2010, J CLIN ONCOL, V28, P3069, DOI 10.1200/JCO.2009.26.8789
   Haas-Kogan DA, 2011, NEURO-ONCOLOGY, V13, P298, DOI 10.1093/neuonc/noq202
   Hargrove D, 2006, LANCET ONCOL, V7, P241, DOI 10.1016/S1470-2045(06)70615-5
